# Association of childhood experiences with wellbeing and flourishing outcomes in Indian adults: Evidence from the India Sample of the Global Flourishing Study (GFS)

**DOI:** 10.64898/2026.10.09.26365161

**Authors:** Yiren Hou, Tami Sullivan, Emma Zang, Shekhar Saxena, Madhu Gupta, Nandini Chatterjee Singh, Bhramar Mukherjee

**Affiliations:** Department of Biostatistics, Yale School of Public Health, New Haven, CT 06510, USA; Department of Psychiatry, Yale University School of Medicine, New Haven, CT 06511, USA; Department of Sociology, Yale University, New Haven, CT 06511, USA; Department of Global Health and Population, Harvard T.H. Chan School of Public Health, Boston, MA 02115, USA; Department of Community Medicine and School of Public Health, PGIMER, Chandigarh 160012, India; Department of Psychology and Cognitive Sciences, Ashoka University, Sonipat, Haryana 131021, India; Department of Chronic Disease Epidemiology, Yale School of Public Health, New Haven, CT 06510, USA; Department of Statistics and Data Science, Yale University, New Haven, CT06511, USA

**Author notes:** <u>Correspondence to</u>: Bhramar Mukherjee, PhD, Department of Biostatistics and Chronic Disease Epidemiology Yale School of Public Health, 350 George Street, Rm 320 New Haven, CT 06511, USA.

**Keywords:** abuse, adult outcomes, adverse childhood experiences, flourishing, gender differences, Global Flourishing Study, health outcomes, India, wellbeing

## Abstract

**Background:** Childhood experiences contribute to health and wellbeing across the life course, yet research from low- and middle-income countries has largely focused on a few outcomes rather than broader patterns of adult wellbeing, behaviors, and social and financial circumstances.

**Methods:** In this cross-sectional analysis, we used a nationally representative sample of adults in India (N = 12,765; aged ≥18 years) from the Global Flourishing Study (GFS) Wave 1 (2023) to examine associations between eight retrospectively reported childhood experience items, 45 adult outcomes, and two composite flourishing indices. Survey-weighted linear, logistic, and proportional-odds models adjusted for age, gender, education, and residential setting, with Bonferroni correction were used to assess associations and effect modification by gender.

**Findings:** Although the breadth and strength of associations differed across childhood experiences, less favorable childhood experiences were generally associated with poorer wellbeing and flourishing, greater odds of adverse health outcomes, and reduced access to resources and safety in adult life. However, outcomes related to prosocial behaviors showed a different pattern: childhood abuse (physical or sexual) was associated with greater odds of volunteering (OR 1·31, 95% confidence interval or CI [1·13,1·51]), feeling like an outsider in one’s family with greater odds of donating (OR 1·80, 95% CI [1·52,2·13]), volunteering (OR 1·53, 95% CI [1·28,1·82]), and helping a stranger (OR 1·49, 95% CI [1·27,1·74]). Associations were broadly similar between females and males, with little evidence of gender differences.

**Interpretation:** Associations of childhood adversity with poorer health and increased prosocial behavior in adult life may reflect unmeasured social, cultural, or dispositional factors and heterogeneous life-course responses. Nevertheless, these findings support strengthening child-protection, financial assistance programs, and mental health services in India.

## Introduction

Childhood experiences are associated with physical and mental health, health behaviors, social relationships, and subjective wellbeing across the life course.^1–5^ A substantial literature, developed predominantly in North America and Europe, has linked childhood abuse, family and relational adversity, poor childhood health, and socioeconomic hardship with adult outcomes including depression, anxiety, substance use, chronic disease, and social difficulties.^2,3,5^ Child adversity has also been linked with more severe outcomes, such as suicidal ideation, suicide attempts, criminal offending, arrest, and incarceration.^6,7^ Growing evidence from South and Southeast Asia similarly has shown that adverse childhood experiences are associated with poorer adult outcomes.^8–11^ A life-course perspective provides a useful framework for these associations: childhood experiences may have enduring implications for later health and wellbeing through developmental processes, accumulation of risk, and chains of social and behavioral exposures.^12^ Childhood experiences are also embedded within family and broader social contexts, so their meaning and consequences may vary across settings and over the life course.^13^

India, with approximately 1·45 billion residents (nearly 18% of the world’s population) and a median age of approximately 28 years, represents an important setting in the Global South for examining the long-term associations between childhood experiences and adult outcomes.^14^ Nationally representative analyses from the Longitudinal Ageing Study in India (LASI) have linked retrospectively reported childhood socioeconomic disadvantage and poor childhood health with later-life depressive symptoms, cognitive impairment, and adverse self-rated health trajectories among middle-aged and older adults.^15–17^ Developmental and birth-cohort studies in India have likewise examined associations between early-life adversity and subsequent health and neurodevelopment.^18,19^ Other studies from India have frequently focused on individual childhood experiences or selected outcomes, particularly depression, anxiety, stress, and substance use, among adolescents, young adults, clinical populations, or geographically restricted samples.^8,9,11,20^ Less is known about how multiple dimensions of childhood experience are associated with a broad range of adult outcomes across the full adult age range in a nationally representative Indian population, and whether these associations differ by gender. Although females experience a higher prevalence of certain adversities, particularly sexual abuse, and more frequent co-occurrence of multiple adversities, evidence on gender differences in these associations is mixed: for adult depression and anxiety, some studies report stronger associations among females and others comparable or stronger associations among males, depending on the type and accumulation of adversity.^21–23^ Within India, the few studies considering gender have largely relied on sex-stratified or gender-adjusted analyses rather than formal tests for interaction.^15,16,20^

The Global Flourishing Study (GFS), a longitudinal panel study with population-based samples and a common survey instrument assessing multiple dimensions of flourishing and other adult outcomes across diverse settings, provides an opportunity to address this gap.^24–27^ Previous Wave 1 GFS analyses have evaluated childhood predictors across 22 countries, each focusing on a specific adult outcome, including physical health, pain, suffering, financial wellbeing, optimism, belonging, social support, volunteering, charitable giving, helping others, smoking, inner peace, and sense of mastery.^28–40^ These studies synthesized country-specific associations through random-effects meta-analysis, with India-specific results reported in supplementary analyses, and established that associations vary across childhood exposures and national contexts. They do not, however, provide an integrated India-focused assessment across the breadth of GFS outcomes, including gender differences.

In this study, we used Wave 1 (2023) GFS data from a nationally representative sample of adults aged 18 years or older in India (N = 12,765) to examine associations between eight retrospectively reported childhood experiences and 45 adult outcomes (see eTable 1 for detailed information about the survey questions, responses, and analysis recodes). For descriptive organization, we grouped the eight childhood experiences into four domains based on the aspect of childhood they primarily represented: *adverse experiences* (childhood abuse and feeling like an outsider in one’s family while growing up), *emotional experiences* (not feeling loved by one’s mother or father and reporting a somewhat bad or very bad relationship with one’s mother or father), *health* (poor self-rated health while growing up), and *structural conditions* (perceiving one’s family financial situation as very difficult while growing up). The 45 adult outcomes were organized into four broad groupings of wellbeing and flourishing, adverse health outcomes, character and prosocial behaviors, and social, civic, and material resources. The GFS does not define a standard grouping of adult outcomes; therefore, these groupings draw on the GFS flourishing domains and prior GFS analytic domains.^1,41,42^ Both sets of groupings organize the presentation and interpretation of findings and are not mutually exclusive causal constructs. We also examined two GFS composite flourishing indices: the mean of 10 items assessing happiness, health, meaning, character, and social relationships, and the mean of these items plus two financial-stability items.^26^ All component items were measured on 0-to-10 scales, with higher values indicating greater flourishing (eTable 2).

To broaden the range of adult experiences considered, we also conducted a supplementary analysis using the 2024 India GFS mid-year survey to examine childhood abuse in relation to 14 additional psychosocial, attitude-related, and behavioral outcomes, including but not limited to connection with nature, appreciation of beauty, diligence, and the perceived importance of health, happiness, and relationships (eTable 3). These outcomes were not included in the primary association analyses because they were collected in a separate mid-year questionnaire administered to a smaller subset of respondents.

Our specific objectives were to: (1) characterize associations between the eight childhood experiences and adult outcomes; (2) identify whether patterns of association differed across domains of childhood experience and adult outcomes; and (3) assess whether these associations differed between female and male respondents using formal predictor-by-gender interaction tests. We *a priori* hypothesized that less favorable childhood experiences would generally be associated with poorer adult outcomes, although the pattern of associations across outcome groupings and the strength of associations might differ across childhood experiences. Based on the mixed prior literature and limited evidence from India, we *a priori* hypothesized that gender differences, if present, would vary across childhood experiences and adult outcomes rather than consistently indicate stronger associations among either females or males.

## Methods

### Data source, study population, and survey design

Data were drawn from Wave 1 of the Global Flourishing Study (GFS), an ongoing longitudinal panel study developed by Harvard University, Baylor University, Gallup, and the Center for Open Science to examine human flourishing across cultural and geographic settings.^24–27^ Wave 1 includes more than 200,000 participants from 22 countries and was designed to yield approximately nationally representative estimates for adults aged 18 years or older, with measures of wellbeing, health, and social, economic, religious, psychological, and demographic characteristics.^25–27^

In India (N = 12,765), participants were recruited through probability-based face-to-face sampling with computer-assisted personal interviewing, and the Wave 1 annual survey was completed by telephone (n = 12,549) or web (n = 216) between April 25 and December 8, 2023.^27^ Fieldwork yielded 28,644 intake-survey and 12,765 annual-survey completions, with intake and annual response rates of 68·9% and 44·6%, respectively. Coverage was approximately 93%; most northeastern states, remote islands, and Jammu and Kashmir were excluded from the sampling frame (state distributions in eTable 4).^27^ The questionnaire was administered in 11 languages (Assamese, Bengali, Gujarati, Hindi, Kannada, Malayalam, Marathi, Odia, Punjabi, Tamil, and Telugu), with translations following a modified TRAPD (translation, review, adjudication, pretesting, and documentation) process and local pilot testing.^25,27^ Gallup constructed sampling weights from selection probabilities, with post-stratification and annual-survey nonresponse adjustment, to project the sample to the target adult population.^27^ All analyses used the Wave 1 cross-sectional annual weight, strata, and primary sampling unit (PSU) variables.

### Measures

Item wording and coding for childhood predictors and outcomes are provided in eTable 1, flourishing domains and indicators in eTable 2, and the full questionnaire and codebook through the Center for Open Science.^24–26^ Non-substantive responses (“does not apply,” “saw, skipped,” “don’t know,” “refused,” and other missing codes) were excluded from analyses, but their unweighted sample sizes and weighted proportions are reported in the descriptive tables.

#### Demographic variables

All adjusted models included age (continuous, in years), gender (female or male; no other responses were recorded), education (elementary or less [up to eight years of basic education], secondary [9-15 years of education], or tertiary [four years of education beyond high school and/or a 4-year college degree]), and residential setting (rural area or farm, small town or village, large city, or suburb of a large city). These covariates were specified *a priori* for adjustment to account for demographic characteristics that could be related to both retrospectively reported childhood experiences and adult outcomes.

#### Childhood predictors

Eight retrospectively reported experiences from when respondents were growing up were examined. *Childhood abuse* (ever physically or sexually abused) and *feeling like an outsider in one’s family* were coded yes versus no (reference). *Feeling loved by mother and by father* were coded no versus yes (reference). *Relationship quality with mother and with father*, originally rated very good, somewhat good, somewhat bad, or very bad, were modeled as very good (reference), somewhat good, and somewhat or very bad, combining the last two because of small sample sizes. *Self-rated childhood health* was modeled on its five-point scale (excellent [reference], very good, good, fair, poor), and *perceived family financial situation* at approximately age 12 on its four-point scale (lived comfortably [reference], got by, found it difficult, found it very difficult).

#### Outcome variables and flourishing composite indices

The 45 adult outcomes covered happiness and life satisfaction, physical and mental health, meaning and purpose, character and virtue, close social relationships, financial stability, civic engagement, and health behaviors. Their groupings are shown in Figure 1. Twenty-two 0-10 items were analyzed as continuous variables, seven as binary variables (1 = yes, 0 = no), and 16 as ordinal variables, with higher categories representing higher levels of the named construct (see eTable 1 for item wording, response categories, and recodes). The two flourishing indices, with and without financial indicators, were calculated as the mean of their GFS component items before any reverse coding of standalone outcomes, following GFS scoring (eTable 2).^26^

**Figure 1.**
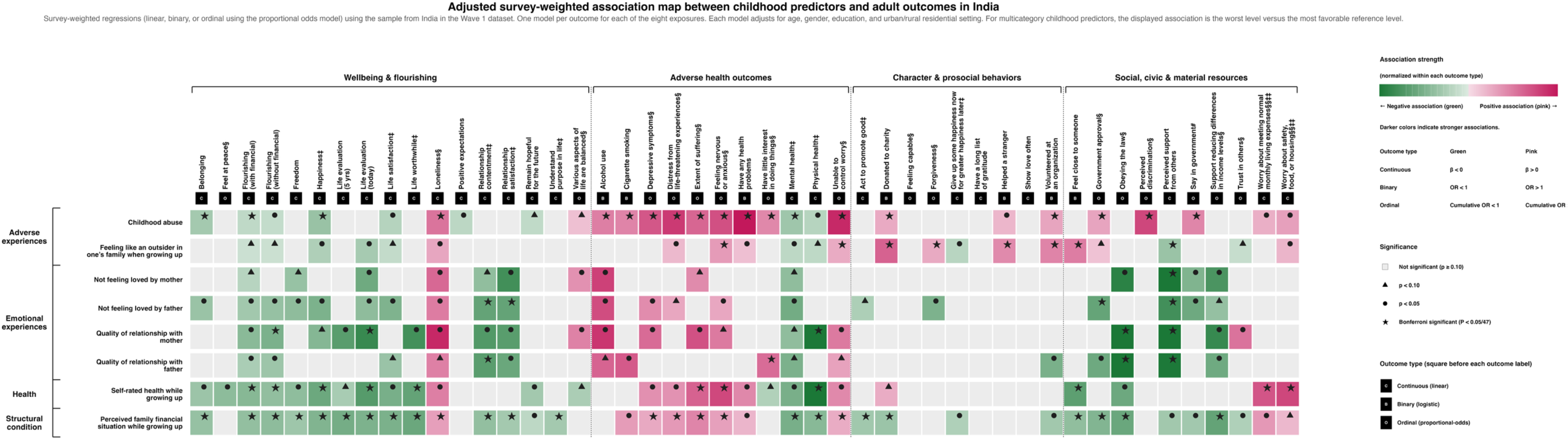
Adjusted survey-weighted association map between eight childhood predictors and 45 adult outcomes and two composite flourishing indices in India using the Wave 1 GFS dataset. ***Notes***: Each cell summarizes the adjusted association between one childhood experience and one adult outcome estimated from a survey-weighted regression model using Wave 1 data from India (N = 12,765). Continuous, binary, and ordinal outcomes were analyzed using linear, logistic, and proportional-odds models, respectively. Models adjusted for age, gender, educational attainment, and residential setting. Cell color indicates the direction and relative magnitude of the association, and symbols denote statistical significance. • Cell color indicates the direction and relative magnitude of the association. A pink cell indicates a positive association, and a green cell indicates a negative association. Color intensity is normalized within each outcome type (which the dark grey squares label either continuous [C], binary [B], or ordinal [O]), with darker shades indicating larger absolute effect sizes; therefore, color intensity should not be compared across outcome types. • Symbols indicating statistical significance include a star for Bonferroni significant (*P* < 0·05/47 ≈ 0·0011), a circle for *P* < 0·05, and a triangle for *P* < 0·1. • The subscript **^‡^** indicates that this outcome is included in the flourishing composite (without financial indicators). The subscript **^‡‡^** indicates that this is a financial indicator included in the flourishing composite (with financial indicators). • The outcome variables marked with **^§^** were reverse coded in the association analysis. • The outcome variable *say in government* was recoded in the order by level 1 = disagree, level 2 = unsure, and level 3 = agree. • The outcome variables (*worry about meeting normal monthly living expenses* and *worry about safety, food, or housing*) marked with **^§§^** were reverse coded in the association analysis, and they were reverse coded for the table standalone reporting (eTable 1 and Table 2). Within flourishing index calculations, *worry about meeting normal monthly living expenses* and *worry about safety, food, or housing* are on original scales. • Binary outcomes, as stated in Methods, are coded where No = 0 and Yes = 1.

### Statistical analysis

All analyses were conducted in R (version 4.5.1) using the survey package for design-based estimation.^43–45^ The Wave 1 cross-sectional annual sampling weights, strata, and primary sampling unit variables were incorporated into all analyses, with variance estimated by Taylor-series linearization and lonely primary sampling units handled using the centered adjustment. ^27,41^ All point estimates, standard errors, and 95% confidence intervals (CIs) are design based; unweighted sample sizes and weighted proportions are reported in the Results and Tables. To describe relationships among variables before the association analyses, we estimated survey-weighted Pearson and Spearman correlations (the latter based on weighted mid-ranks) among the eight childhood predictors and 45 outcomes, using pairwise complete observations (eFigures 1-2; eTables 5-6).

For each childhood predictor, separate models were fitted for each of the 45 outcomes and two indices, adjusting for all covariates: survey-weighted linear regression for continuous outcomes, logistic regression for binary outcomes, and cumulative-logit proportional-odds models for ordinal outcomes. Multicategory predictors were entered as indicator terms for each non-reference category; figures display the least versus most favorable category, and all category-specific coefficients are provided in eTables 7-14.

For ordinal outcomes, we used survey-weighted cumulative-logit proportional-odds models. For an ordinal outcome with *K* ordered response categories, the model estimated the log odds of being at or below category *k*, for each threshold *k* = 1, …, *K* – 1. The linear predictor,denoted by n_ii_, included the childhood predictor and covariates for respondent *i*:

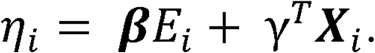

The proportional-odds cumulative logit model can be written as

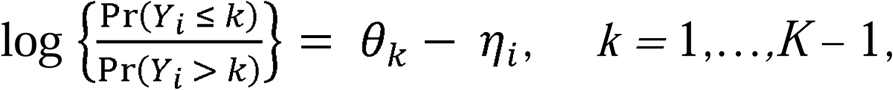

where *γ_i_* is the ordinal outcome; *θ_k_* is the threshold-specific intercept; *E_i_* is the childhood predictor term or set of indicator terms; and X_i_ is the covariate vector. Because the svyolr function parametrization uses logit{ Pr(*γi* ≤ *k*)} = *θk* - *n_i_*, positive predictor coefficients indicate a shift toward higher outcome categories. Exponentiated coefficients are reported as cumulative odds ratios for being in a higher outcome category among exposed versus reference respondents. The proportional-odds assumption was assessed with a Wald chi-squared test comparing exposure coefficients from binary logistic models fitted at each cumulative cut-point. Models with evidence of violation (*P* < 0·05) were flagged in the figures and retained for transparency and should be interpreted cautiously.

Gender-specific estimates were derived from full-sample models including predictor-by-gender interaction terms (one per non-reference category), rather than from stratified models; covariate associations were thus assumed common across genders while predictor associations could differ. With males as reference, the male-specific estimate was the predictor coefficient and the female-specific estimate the sum of the predictor and interaction coefficients. Gender differences were tested using the interaction coefficient, representing the female-minus-male difference on the mean-difference, log-odds, or log cumulative-odds scale, with design-based two-sided P values and 95% CIs.

To account for multiple testing, significance was evaluated at a Bonferroni-adjusted threshold of *P* < 0·0011 (0·05/47, for 45 outcomes and two indices),^46^ applied separately for each childhood predictor because each represented a distinct scientific question regarding its pattern of associations across adult outcomes. Figures additionally indicate *P* < 0·05 and *P* < 0·10. Complete-case analysis was conducted for each predictor-outcome pair, so analytic sample sizes varied across models due to missing data or nonresponse in predictors, outcomes, and covariates. Item nonresponse is reported in the descriptive tables (Tables 1-2). Additional details for the India mid-year survey and sensitivity analyses and their results are provided in Results and eResults 2.

**Table 1.**
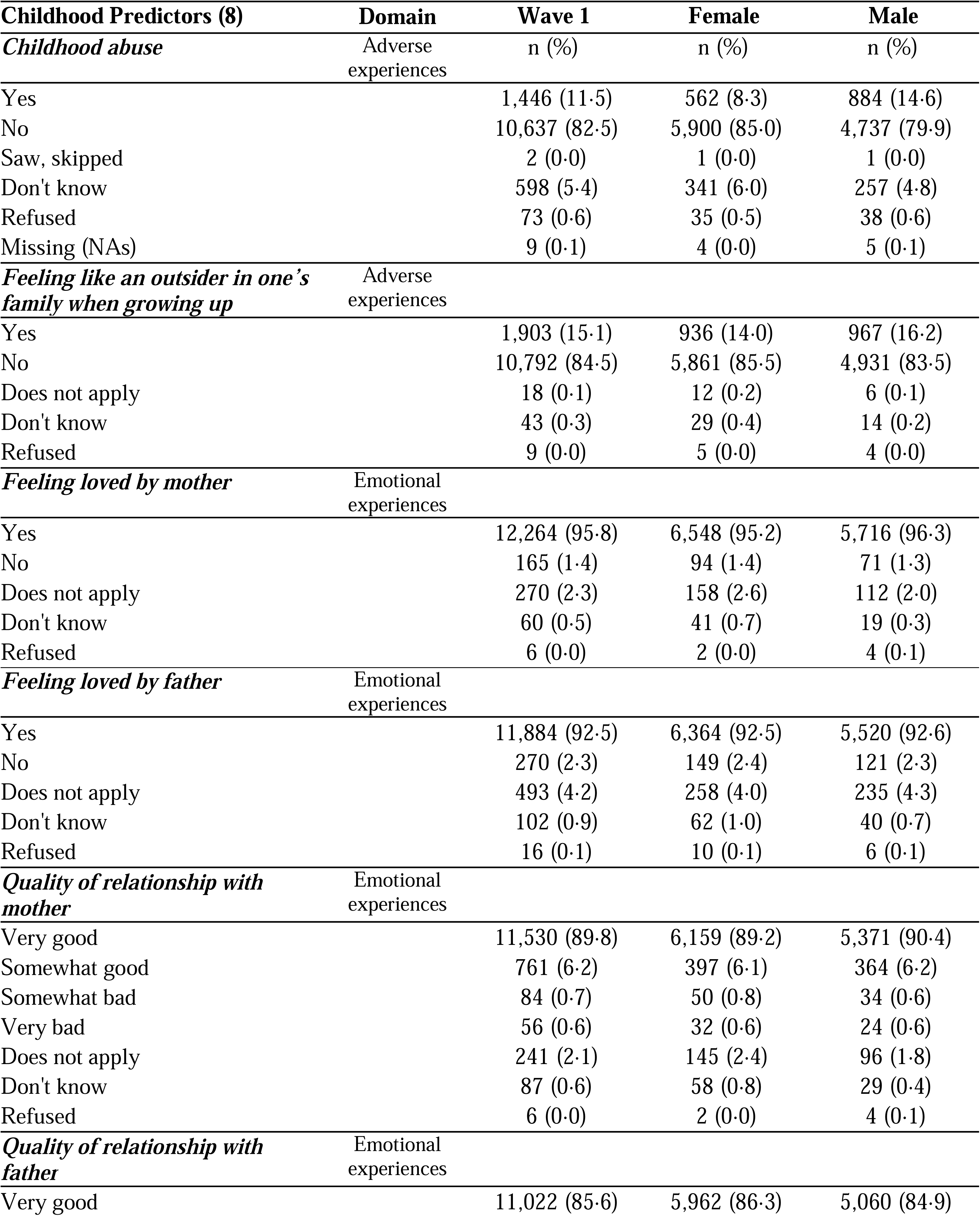
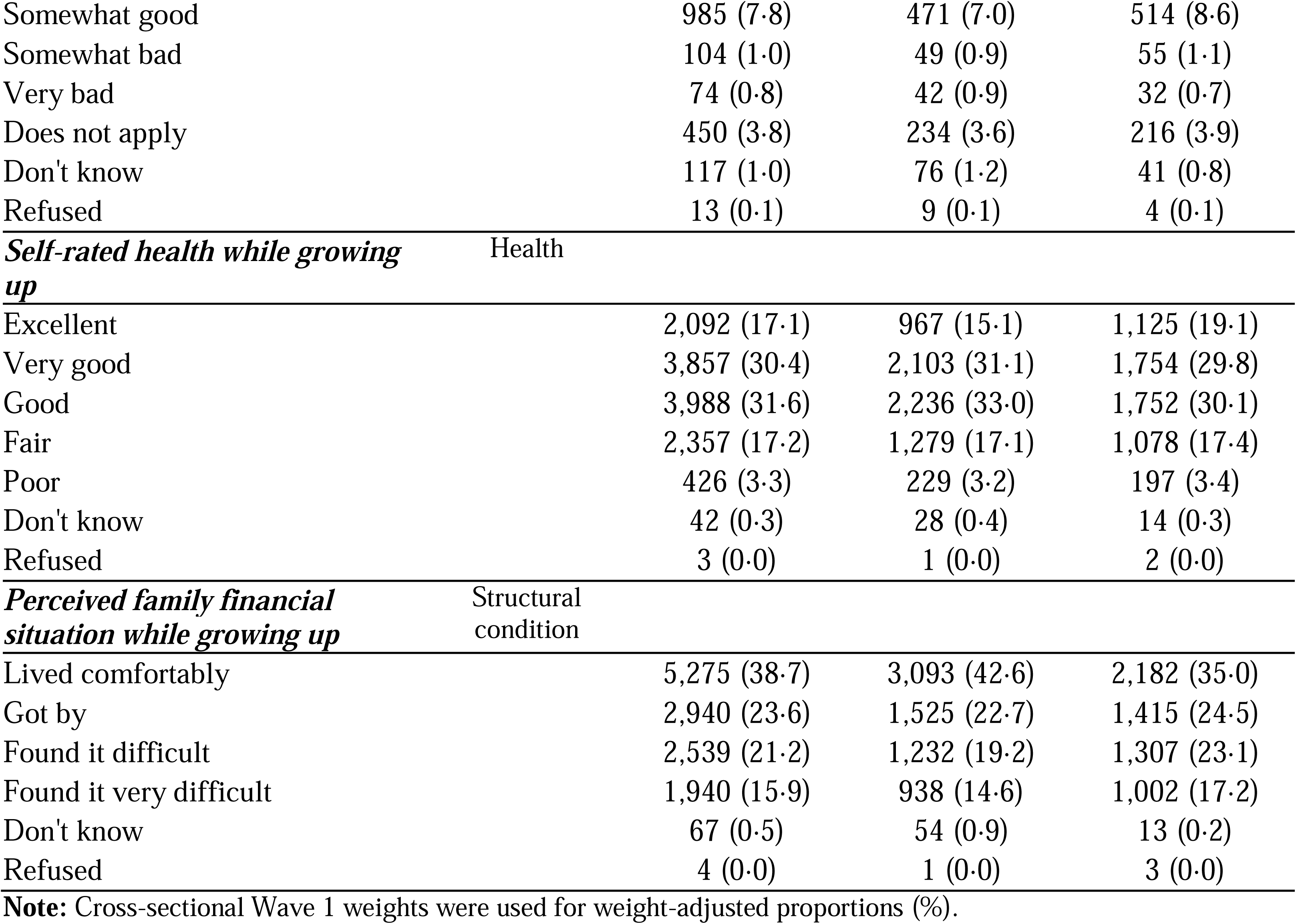
Distribution of eight childhood predictors collected in the Wave 1 survey (2023) in India and stratified by gender (N = 12,765). Unweighted sample sizes (n) and weighted proportions (%) are reported.

**Table 2.**
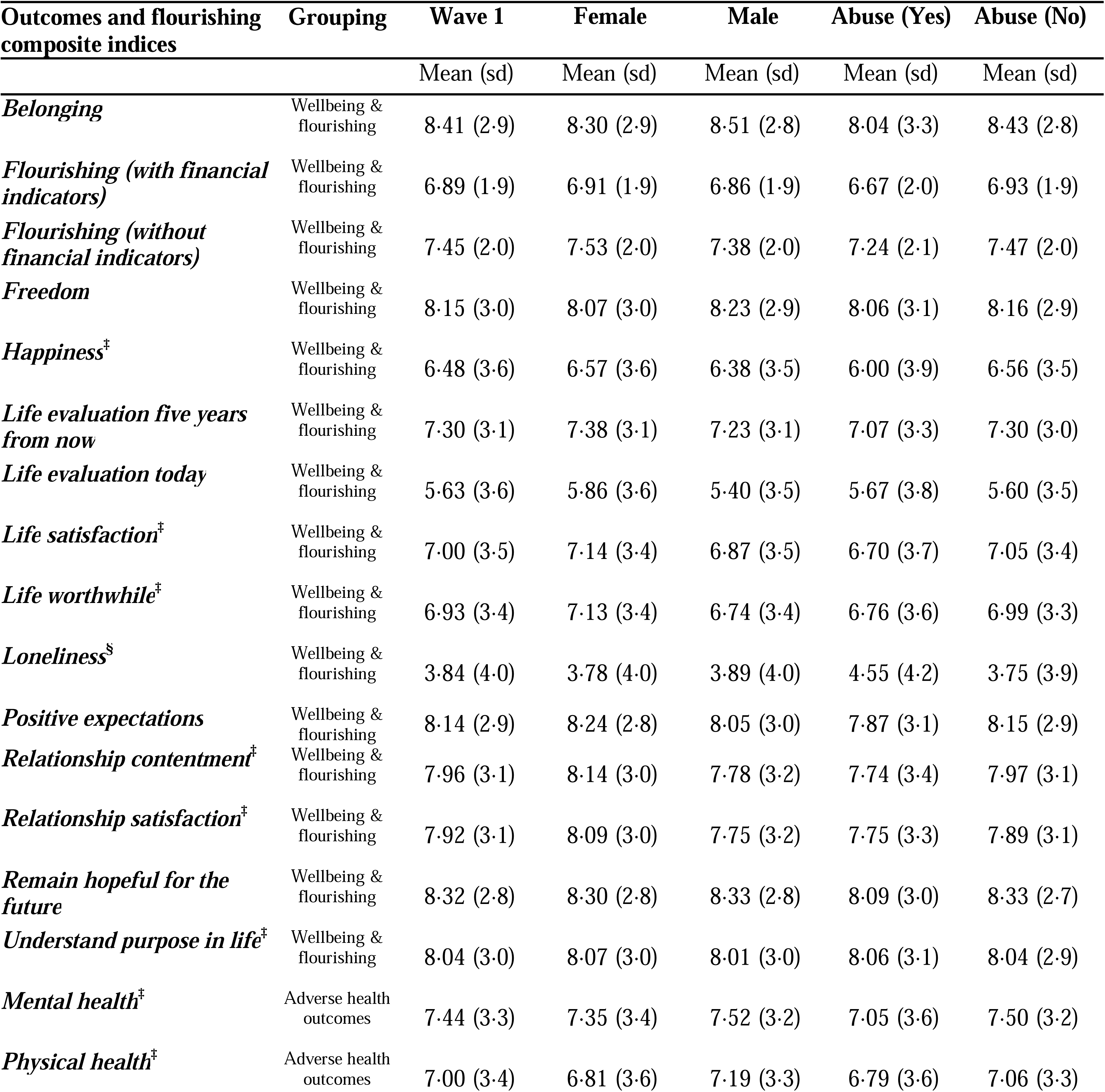
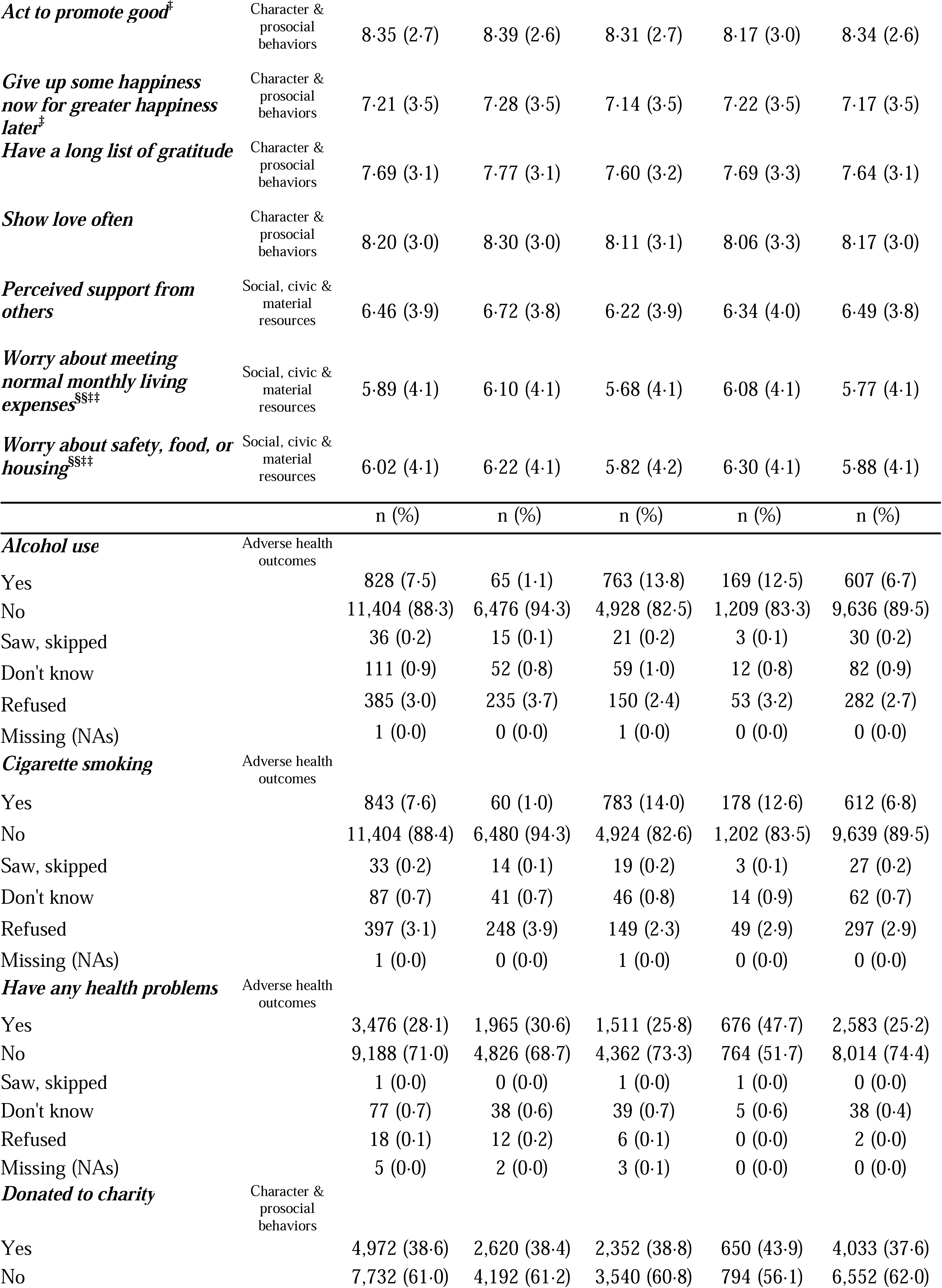
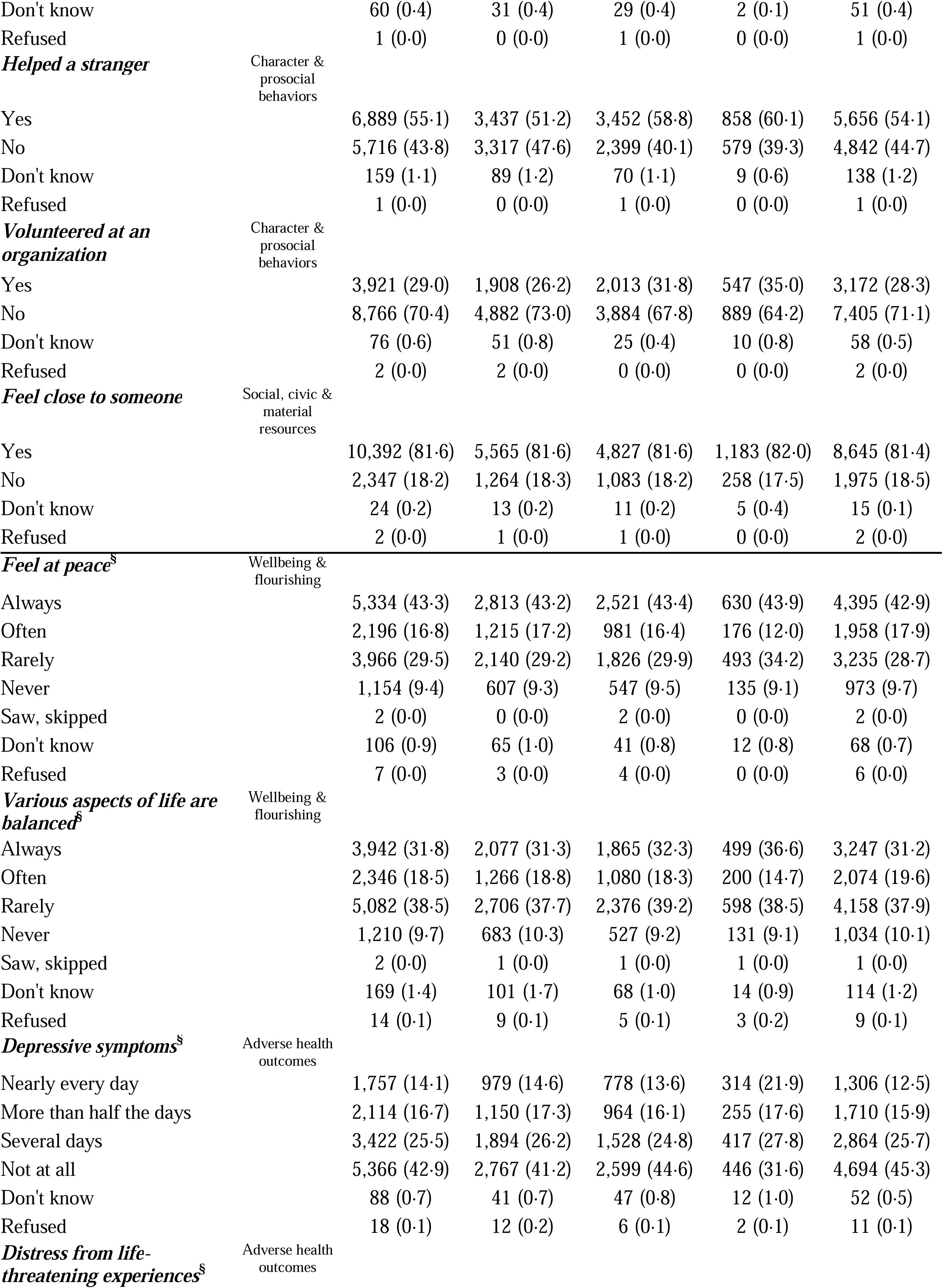
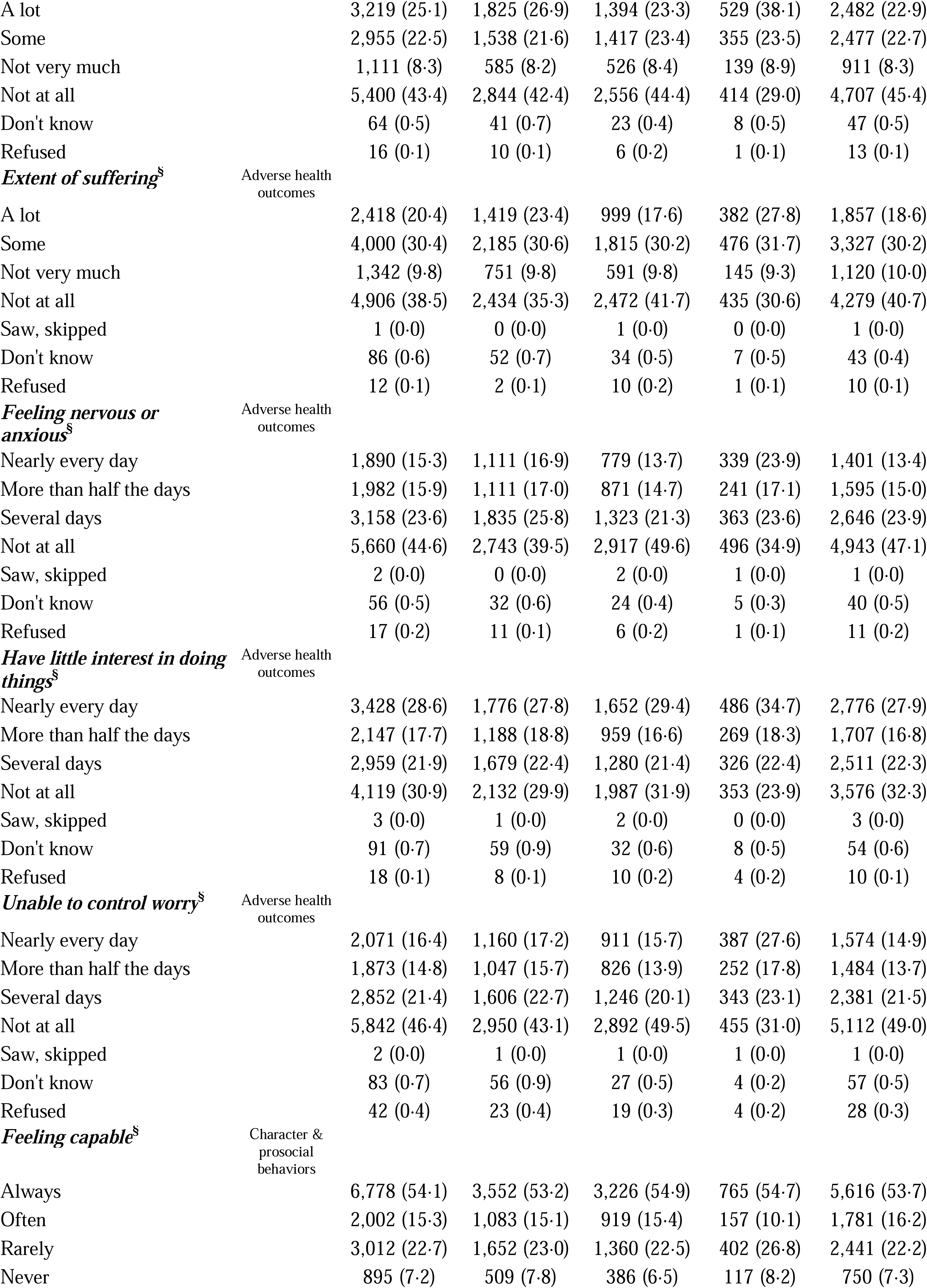
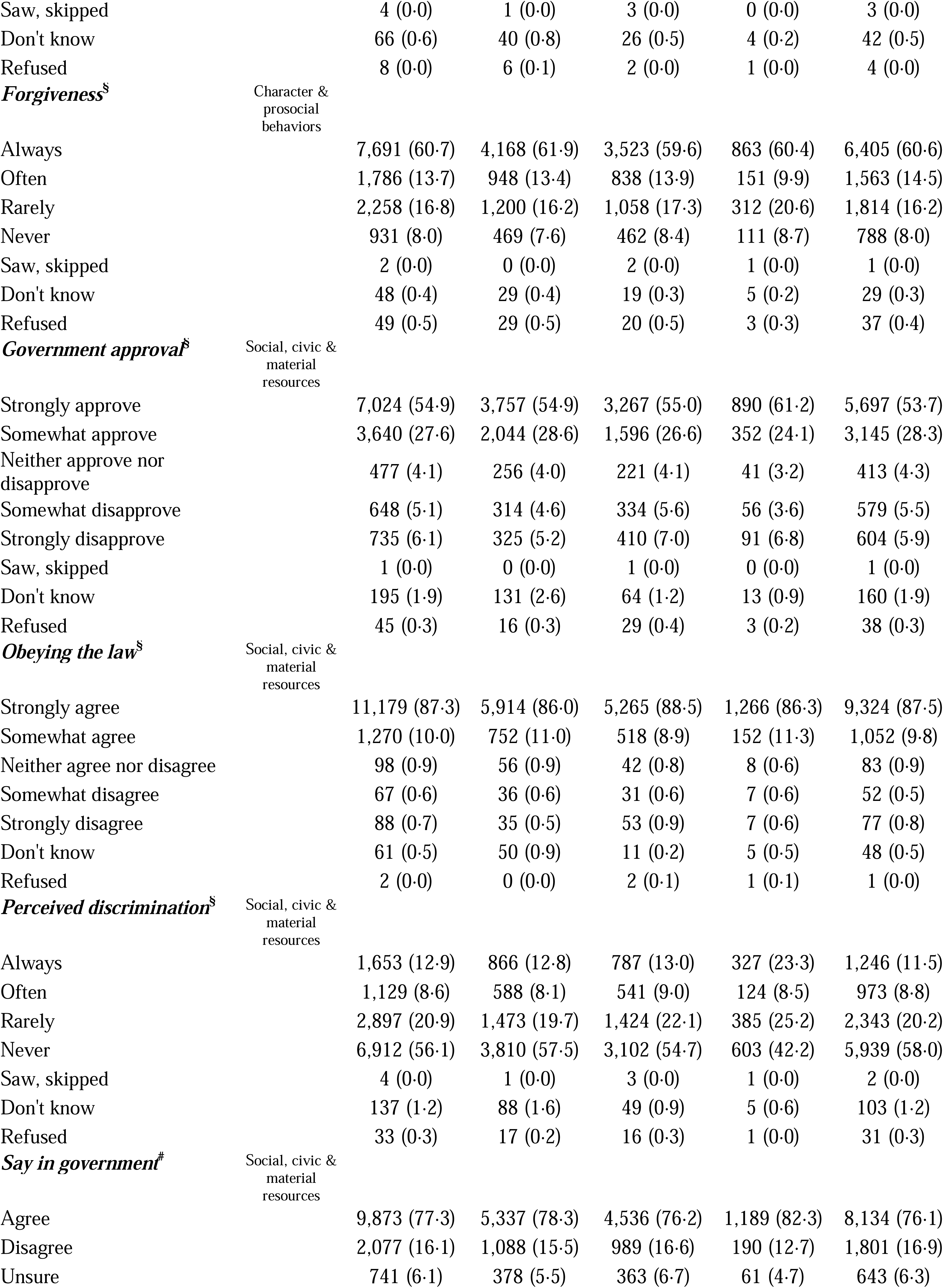
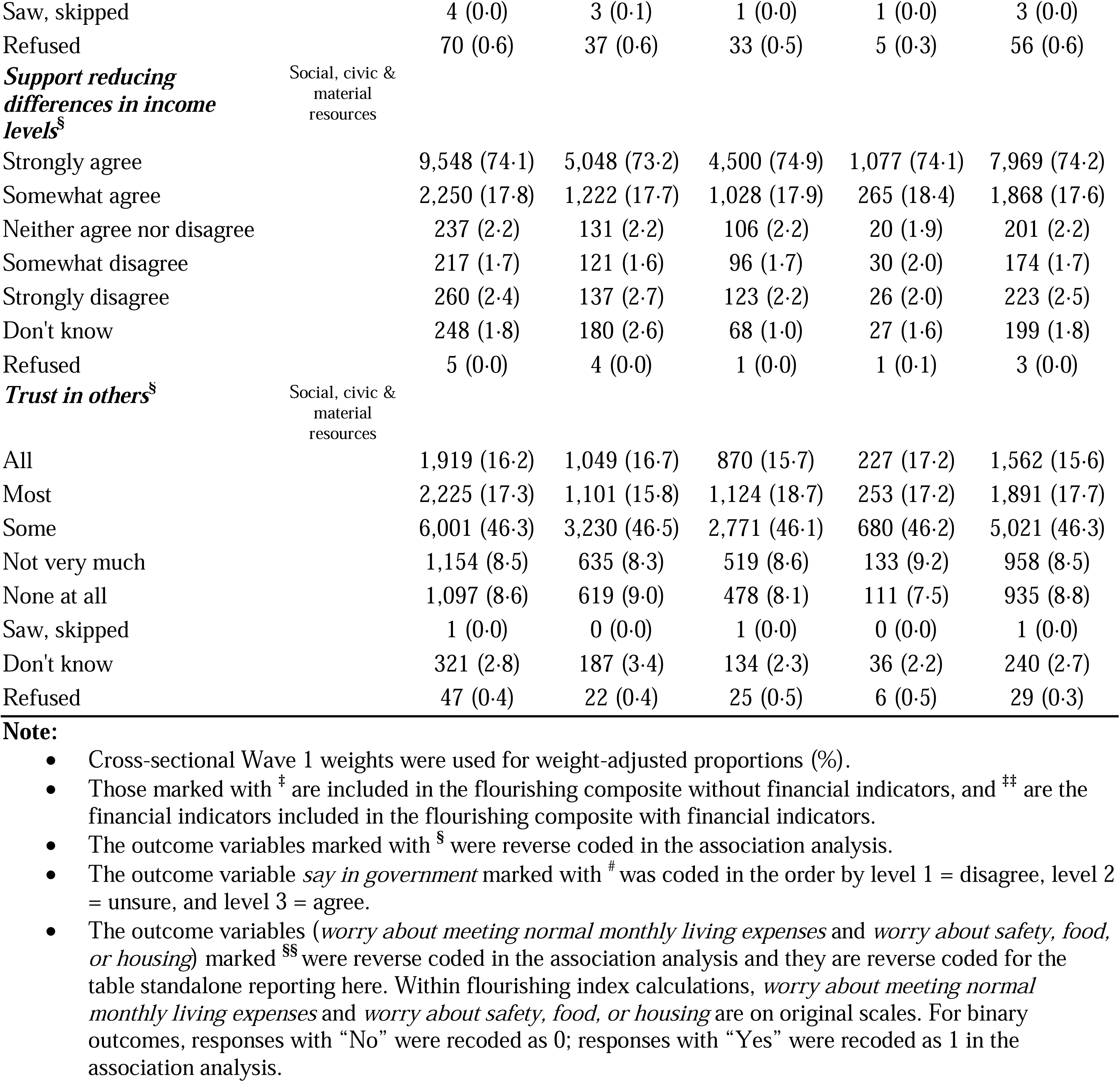
Distribution of 45 outcome variables collected in Wave 1 survey (2023) in India and two flourishing composite indices, stratified by gender and predictor childhood abuse (N = 12,765). *Subheader*: For continuous outcomes, we reported weighted mean and standard deviation (sd); all continuous measures and both composite flourishing indices are scored 0-10, with higher values indicating more of the named construct. For binary and ordinal variables, we reported unweighted sample size (n) and weighted proportion (%); binary outcomes are coded Yes/No, and ordinal outcomes retain their original 3-, 4-, or 5-point response categories. Full item wording, response options, and coding for all outcomes are provided in eTable 1.

### Ethics approval

This study used de-identified, publicly available data from the Global Flourishing Study and did not require ethics approval. We completed the data sharing protocols of GFS to download the data.

## Results

### Descriptive statistics

The GFS India Wave 1 analytic sample comprised 12,765 respondents (6,843 [49·3%] female and 5,922 [50·7%] male; unweighted n and weighted %). Most respondents were married (10,082 [77·1%]), born in India (12,637 [98·9%]), and completed the survey by telephone (12,549 [98·3%]). Most lived in a small town or village (5,181 [41·4%]) or a rural area or on a farm (5,003 [34·9%]), with 1,996 (18·3%) in a large city and 585 (5·4%) in a suburb. Educational attainment was generally low, with 7,684 (68·6%) having completed elementary education or less (up to eight years), and 5,510 (47·4%) were employed or self-employed (eTable 15).

The prevalence of childhood experiences varied across items (Table 1). Overall, 1,446 (11·5%) respondents reported physical or sexual abuse during childhood (884 [14·6%] males, 562 [8·3%] females), and 1,903 (15·1%) reported feeling like an outsider in the family (967 [16·2%] males, 936 [14·0%] females). Most respondents reported positive parent-child relationships: 12,264 (95·8%) felt loved by their mother and 11,884 (92·5%) by their father, and 11,530 (89·8%) and 11,022 (85·6%) reported a very good relationship with their mother and father, respectively. Only 426 (3·3%) reported poor health while growing up. Perceived childhood financial hardship, however, was relatively common: 2,539 (21·2%) reported that their family found it difficult financially and 1,940 (15·9%) very difficult.

Respondents generally reported favorable levels of wellbeing and flourishing, close social relationships, and civic engagement, whereas current cigarette smoking (843 [7·6%]) and alcohol use in the seven days before the survey (828 [7·5%]) were relatively uncommon (Table 2, overall and by gender and childhood abuse).

### Correlation analyses

Survey-weighted Pearson and Spearman correlations among childhood predictors were generally low to moderate, whereas adult outcomes clustered strongly within the four conceptual groupings, particularly the wellbeing and flourishing measures. Correlations between childhood predictors and adult outcomes were generally smaller than those within either set of variables (eFigures 1-2; eTables 5-6). A detailed description is provided in eResults 1.

### Associations between childhood predictors and adult outcomes, reported by conceptual domain of childhood experiences

Survey-weighted regression models were adjusted for age, gender, educational attainment, and residential setting. Effect estimates are adjusted mean differences (*β̂*) for continuous outcomes, adjusted odds ratios (ORs) for binary outcomes, and cumulative odds ratios (cumulative ORs) for ordinal outcomes, with 95% Wald confidence intervals (CIs) in brackets. For the multicategory childhood predictors, the text and figures present the least favorable versus the most favorable (reference) category: somewhat bad or very bad versus very good relationship with one’s mother or father, poor versus excellent self-rated health, and very difficult versus comfortable family financial situation. Estimates for all other predictor levels and for model covariates are provided in eTables 7-14. Associations meeting the Bonferroni-adjusted threshold of *P* < 0·0011 (0·05/47, reflecting 45 adult outcomes and two flourishing indices tested for each childhood predictor) are summarized below. Associations with nominal significance (*P* < 0·05 or *P* < 0·10) are shown in the main and supplementary figures. We applied the correction separately for each childhood predictor because each represented a distinct scientific question regarding its pattern of associations across multiple adult outcomes. Gender differences were assessed by adding a predictor-by-gender interaction term to each model, assuming common covariate associations across genders, and one predictor-by-gender interaction met the Bonferroni threshold. Overall, gender-specific, and interaction estimates are presented in Figure 2 and eFigures 3-9, and details on category coding, interaction models, and multiple-testing correction are provided in Methods.

**Figure 2.**
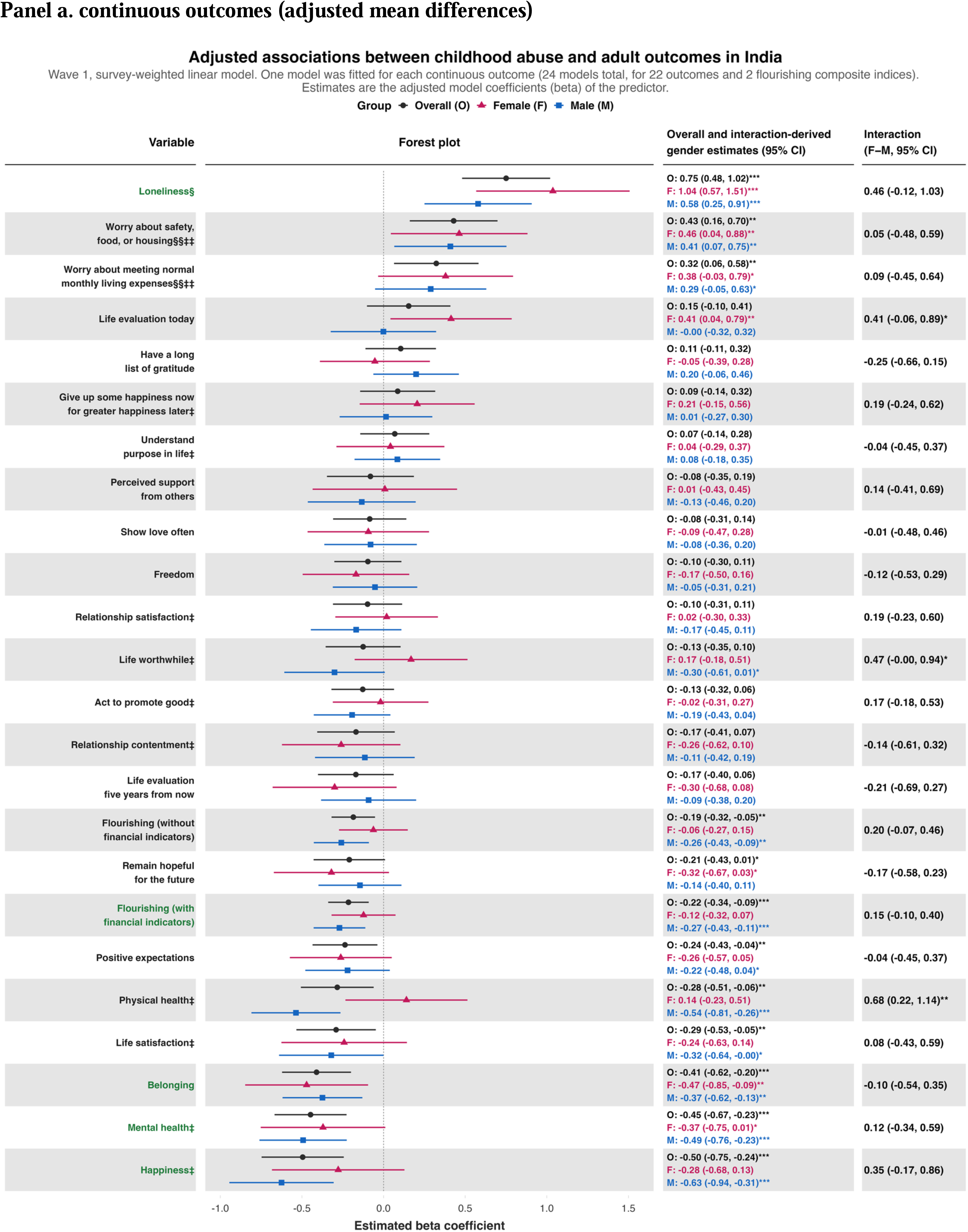
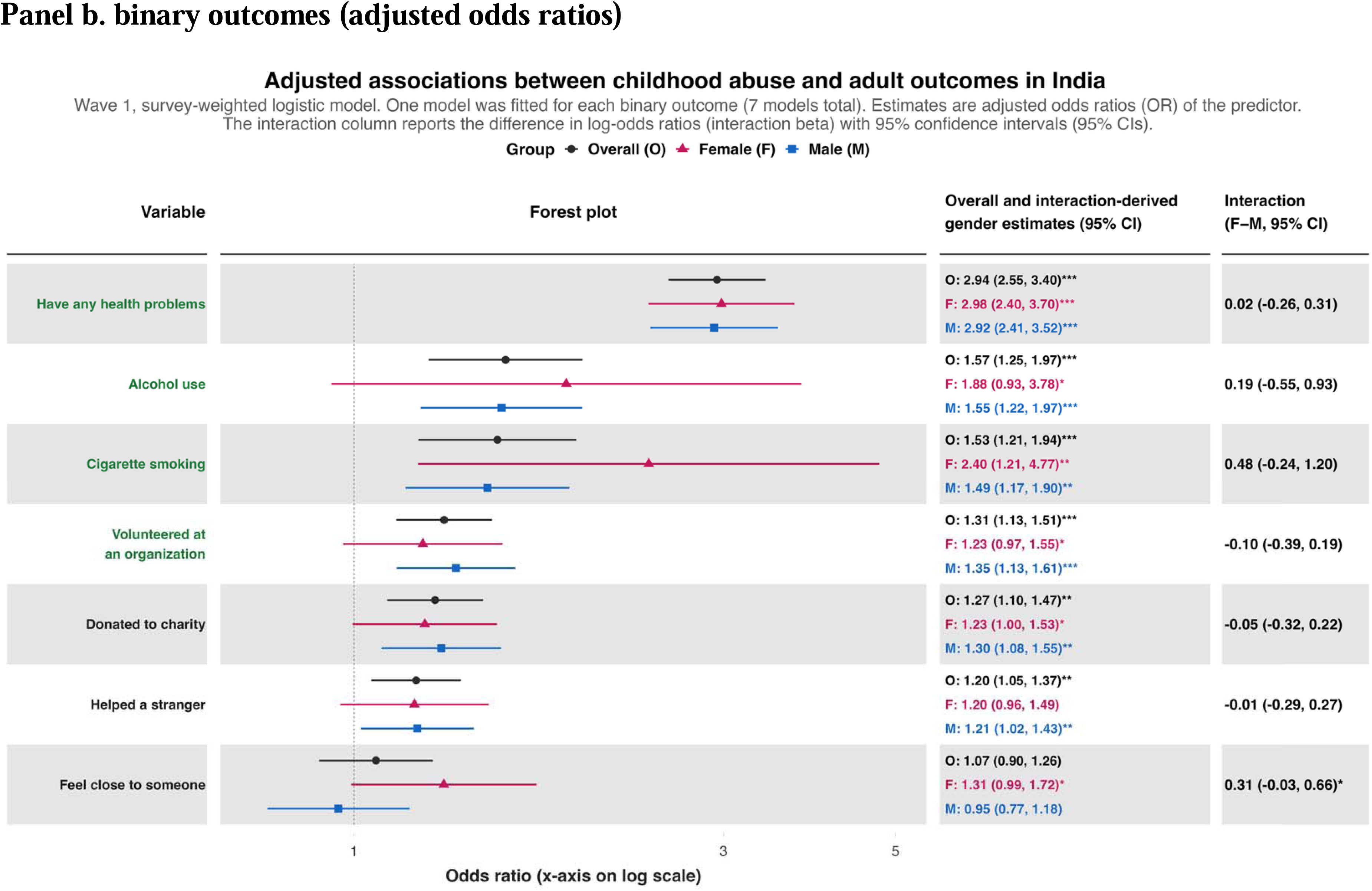
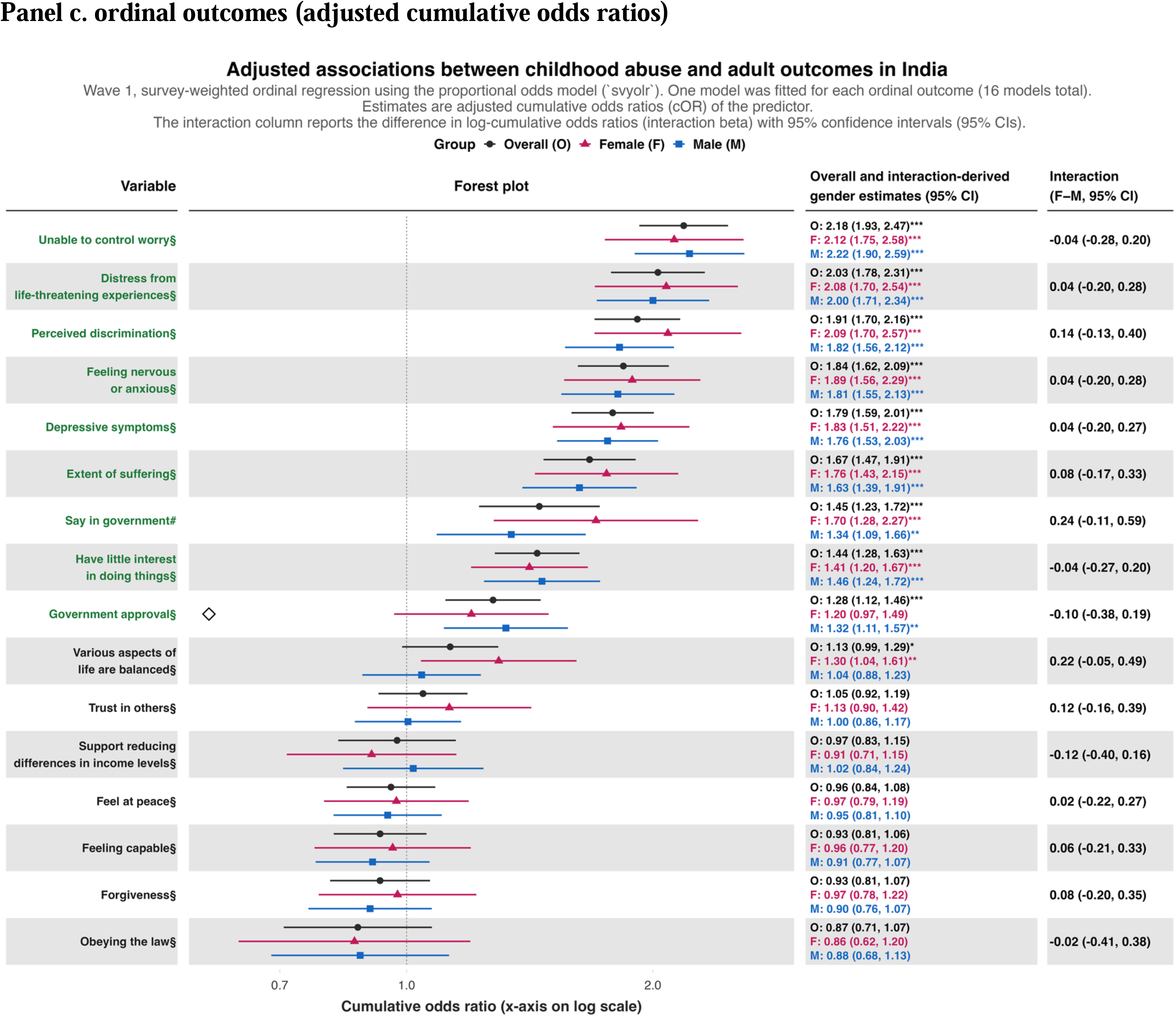
panels. Forest plots of associations between childhood abuse and 45 outcomes and two flourishing composite indices using samples from India in the Wave 1 GFS dataset. Each panel shows the associations between ever experiencing physical or sexual abuse while growing up and each of the continuous outcomes (**Panel a**), binary outcomes (**Panel b**), and ordinal outcomes (**Panel c**).Notes: • Outcome labels in bold green are significant after Bonferroni correction (0·05/47 ≈ 0·0011). • Stars next to each estimate indicate: *** *P* < Bonferroni significant (0·05/47 ≈ 0·0011), ** *P* < 0·05, * *P* < 0·1. • The subscript **^‡^**indicates that this outcome is included in the flourishing composite (without financial indicators). The subscript **^‡‡^** indicates that this is a financial indicator included in the flourishing composite (with financial indicators). • The outcome variables marked with **^§^** are reverse coded in the association analysis. • The outcome variable *say in government* is recoded in the order by level 1 = disagree, level 2 = unsure, and level 3 = agree. • The outcome variables (*worry about meeting normal monthly living expenses* and *worry about safety, food, or housing*) marked with **^§§^** are reverse coded in the association analysis, and they are reverse coded for the table standalone reporting (eTable 1 and Table 2). Within flourishing index calculations, *worry about meeting normal monthly living expenses* and *worry about safety, food, or housing* are on original scales. • Binary outcomes, as stated in Methods, are coded where No = 0 and Yes = 1. • In proportional-odds model subfigures, black open diamonds identify estimates for which the proportional-odds assumption was violated by the Wald chi-squared test (P < 0·05). For the overall estimate, the assumption was assessed in the adjusted main-effects model; for female- and male-specific estimates, it was assessed in the full-sample adjusted predictor-by-gender interaction model. When the interaction model violated the assumption, both gender-specific estimates are marked. • Each model is adjusted for age, gender, education, and residential setting. The reported female- and male-specific estimates are derived from one full-sample predictor-by-gender interaction model; no gender-stratified models produced those estimates.

#### Patterns of association between childhood predictors and composite flourishing indices

The two flourishing indices largely mirrored the patterns seen among wellbeing and flourishing outcomes (Figure 1). Childhood financial hardship was associated with lower flourishing both with (*β̂* = −0·54, 95% CI [−0·68,-0·40]) and without financial indicators (*β̂* = −0·57, 95% CI [−0·71,-0·43]), as was poor self-rated childhood health (with: *β̂* = −0·63, 95% CI [−0·87,-0·39]; without: *β̂* = −0·54, 95% CI [−0·80,-0·29]). Childhood abuse was associated with lower flourishing with financial indicators only (*β̂* = −0·22, 95% CI [−0·34,-0·09]), and poorer relationship with mother was associated with lower flourishing without financial indicators only (*β̂* = −0·71, 95% CI [−1·13,-0·29]). Other childhood experiences were associated with either or both flourishing indices only at nominal significance.

#### Adverse childhood experience: Childhood abuse

*Childhood abuse* had one of the broadest patterns of significant association across the adult outcome groupings (Figure 1 and Figure 2). Within the wellbeing and flourishing grouping, respondents reporting childhood abuse had greater loneliness (*β̂* = 0·75, 95% CI [0·48,1·02]), lower happiness (*β̂* = −0·50, 95% CI [−0·75,-0·24]) and feeling of belonging (*β̂* = −0·41, 95% CI [−0·62,-0·20]).

Within the adverse health outcomes grouping, childhood abuse was associated with poorer mental health (*β̂* = −0·45, 95% CI [−0·67,-0·23]), with higher odds of activity-limiting health problems (OR = 2·94, 95% CI [2·55,3·40]), alcohol use in the seven days before the survey (OR = 1·57, 95% CI [1·25,1·97]), and cigarette smoking (OR = 1·53, 95% CI [1·21,1·94]), and with higher cumulative odds of inability to control worry (cumulative OR = 2·18, 95% CI [1·93,2·47]), distress from life-threatening experiences (cumulative OR = 2·03, 95% CI [1·78,2·31]), nervousness or anxiousness (cumulative OR = 1·84, 95% CI [1·62,2·09]), depressive symptoms (cumulative OR = 1·79, 95% CI [1·59,2·01]), suffering (cumulative OR = 1·67, 95% CI [1·47,1·91]), and little interest or pleasure in activities (cumulative OR = 1·44, 95% CI [1·28,1·63]).

In addition to these associations with adverse outcomes, childhood abuse was associated with higher odds of volunteering for an organization (OR = 1·31, 95% CI [1·13,1·51]) within the character and prosocial behaviors grouping. Within the social, civic, and material resources grouping, it was associated with higher cumulative odds of agreeing that people like oneself have a say in government (cumulative OR = 1·45, 95% CI [1·23,1·72]) and of feeling discriminated against (cumulative OR = 1·91, 95% CI [1·70,2·16]).

#### Adverse childhood experience: Feeling like an outsider in the family

*Feeling like an outsider in one’s family* showed a more selective pattern of associations than childhood abuse (Figure 1 and eFigure 3). None of its associations with wellbeing and flourishing outcomes met the Bonferroni threshold. Within the adverse health outcomes grouping, it was associated with higher cumulative odds of nervousness or anxiousness (cumulative OR = 1·33, 95% CI [1·18,1·50]) and inability to control worry (cumulative OR = 1·24, 95% CI [1·10,1·39]). A different pattern was observed for character and prosocial behaviors: respondents who reported feeling like an outsider in their family had higher odds of donating to charity (OR = 1·80, 95% CI [1·52,2·13]), volunteering (OR = 1·53, 95% CI [1·28,1·82]), and helping a stranger (OR = 1·49, 95% CI [1·27,1·74]), and higher cumulative odds of forgiveness (cumulative OR = 1·25, 95% CI [1·11,1·40]). Within the social, civic, and material resources grouping, feeling like an outsider was associated with higher odds of having someone they felt close to (OR = 1·58, 95% CI [1·30,1·93]) but lower perceived support from others (*β̂* = −0·49, 95% CI [−0·73,-0·24]).

#### Emotional childhood experience: Not feeling loved by mother and by father

*Not feeling loved by one’s mother* showed relatively few associations across the adult outcome groupings (Figure 1 and eFigure 4), and the only association meeting the Bonferroni threshold was with lower perceived support from others (*β̂* = −1·42, 95% CI [−2·23,-0·62]). *Not feeling loved by one’s father* showed a broader pattern of associations than *not feeling loved by one’s mother* (Figure 1 and eFigure 5). Within the wellbeing and flourishing grouping, not feeling loved by one’s father was associated with lower satisfaction with relationships (*β̂* = −0·88, 95% CI [−1·38,-0·37]) and lower relationship contentment (*β̂* = −0·88, 95% CI [−1·36,-0·39]). Within the social, civic, and material resources grouping, it was also associated with lower perceived support from others (*β̂* = −1·12, 95% CI [−1·76,-0·48]) and lower cumulative odds of approving the national government (cumulative OR = 0·59, 95% CI [0·45,0·77]).

#### Emotional childhood experience: Poorer relationship quality with mother and with father

Because few respondents reported bad parental relationships, “somewhat bad” and “very bad” responses were combined into a single category and compared with “very good” (reference), with “somewhat good” retained separately (see Methods). *Poorer relationship quality with one’s mother* showed associations primarily within the wellbeing and flourishing, adverse health outcomes, and social, civic, and material resources groupings (Figure 1 and eFigure 6). Respondents reporting a somewhat bad or very bad relationship with their mother had poorer physical health (*β̂* = −1·40, 95% CI [−2·11,-0·70]), lower life evaluation at the time of the survey (*β̂* = −1·20, 95% CI [−1·91,-0·49]), and they reported lower perceived support from others (*β̂* = −1·61, 95% CI [−2·37,-0·85]) and lower cumulative odds of agreeing that the law should always be obeyed (cumulative OR = 0·31, 95% CI [0·19,0·52]).

*Poorer relationship quality with one’s father* showed a similar but more limited pattern (Figure 1 and eFigure 7), with lower relationship contentment (*β̂* = −0·95, 95% CI [−1·49,-0·41]) within the wellbeing and flourishing grouping; little interest or pleasure in activities (cumulative OR = 1·63, 95% CI [1·22,2·18]) within the adverse health outcomes grouping; and lower perceived support from others (*β̂* = −1·39, 95% CI [−2·13,-0·64]) and lower cumulative odds of agreeing that the law should always be obeyed (cumulative OR = 0·42, 95% CI [0·26,0·69]) within the social, civic, and material resources groupings.

#### Health and structural childhood conditions

*Poor self-rated childhood health* was primarily associated with outcomes in the wellbeing and flourishing, adverse health outcomes, and social, civic, and material resources groupings (Figure 1 and eFigure 8). Compared with respondents reporting excellent childhood health, those reporting poor childhood health had greater worry about safety, food, or housing (*β̂* = 1·07, 95% CI [0·55,1·59]) and about meeting normal monthly expenses (*β̂* = 0·97, 95% CI [0·42,1·51]), poorer physical health (*β̂* = −1·41, 95% CI [−1·83,-0·98]), lower happiness (*β̂* = −0·82, 95% CI [−1·27,-0·36]) and feeling life is worthwhile (*β̂* = −0·88, 95% CI [−1·36,-0·40]), lower life evaluation at the time of survey (*β̂* = −0·98, 95% CI [−1·50,-0·45]), higher cumulative odds of nervousness or anxiousness (cumulative OR = 1·90, 95% CI [1·51,2·40]) and suffering (cumulative OR = 1·70, 95% CI [1·32,2·19]), and lower odds of feeling close to someone (OR = 0·53, 95% CI [0·39,0·74]). Poor self-rated childhood health was the only childhood predictor with a Bonferroni-significant gender difference. It was associated with lower scores on acting to promote good among females (*β̂*f= −0·64, 95% CI [−1·16,-0·12]) but higher scores among males (*β̂*_i_ = 0·44, 95% CI [0·05,0·84]; interaction *β̂* = −1·08, 95% CI [−1·70,-0·46]).

*Perceived childhood financial hardship* showed one of the broadest patterns of association, mostly spanning the wellbeing and flourishing, adverse health outcomes, and social, civic, and material resources groupings (Figure 1 and eFigure 9). Notable associations included lower life satisfaction (*β̂* = −0·71, 95% CI [−0·94,-0·49]), poorer mental health (*β̂* = −0·73, 95% CI [−0·94,-0·52]), higher cumulative odds of suffering (cumulative OR = 1·46, 95% CI [1·29,1·66]), distress from life-threatening experiences (cumulative OR = 1·34, 95% CI [1·18,1·51]), and lower odds of feeling close to someone (OR = 0·69, 95% CI [0·57,0·84]).

### Association analyses using mid-year survey responses available in India

We additionally examined associations between childhood abuse and 14 additional outcomes from the India mid-year survey from February to May, 2024 (eTable 3), using a Bonferroni-adjusted threshold of *P* < 0·0036 (0·05/14). Respondents reporting childhood abuse rated it as less important to be healthy (*β̂* = −0·57, 95% CI [−0·82,-0·32]), to have good relationships (*β̂* = −0·48, 95% CI [−0·77,-0·20]), and to be happy (*β̂* = −0·44, 95% CI [−0·70,-0·18]). Survey questions, response options, and association estimates for the 14 outcomes are presented in eTable 3 and 16 and eFigure 10.

## Discussion

In this nationally representative, India-focused analysis, different childhood experiences unveiled distinct patterns of association across adult outcome groupings spanning wellbeing and flourishing, adverse health outcomes, character and prosocial behaviors, and social, civic, and material resources. Childhood abuse and perceived childhood financial hardship showed some of the broadest patterns of association, whereas feeling like an outsider, emotional childhood experiences, and poor childhood health showed more outcome-specific patterns. Table 3 summarizes these patterns and their potential policy implications across the four outcome groupings. For example, the associations of childhood abuse with poorer mental health and greater psychological distress emphasize the relevance of childhood protection initiatives such as Mission Vatsalya,^47,48^ alongside accessible mental health services through the National Mental Health Programme and Tele-MANAS.^53,54^

**Table 3.**
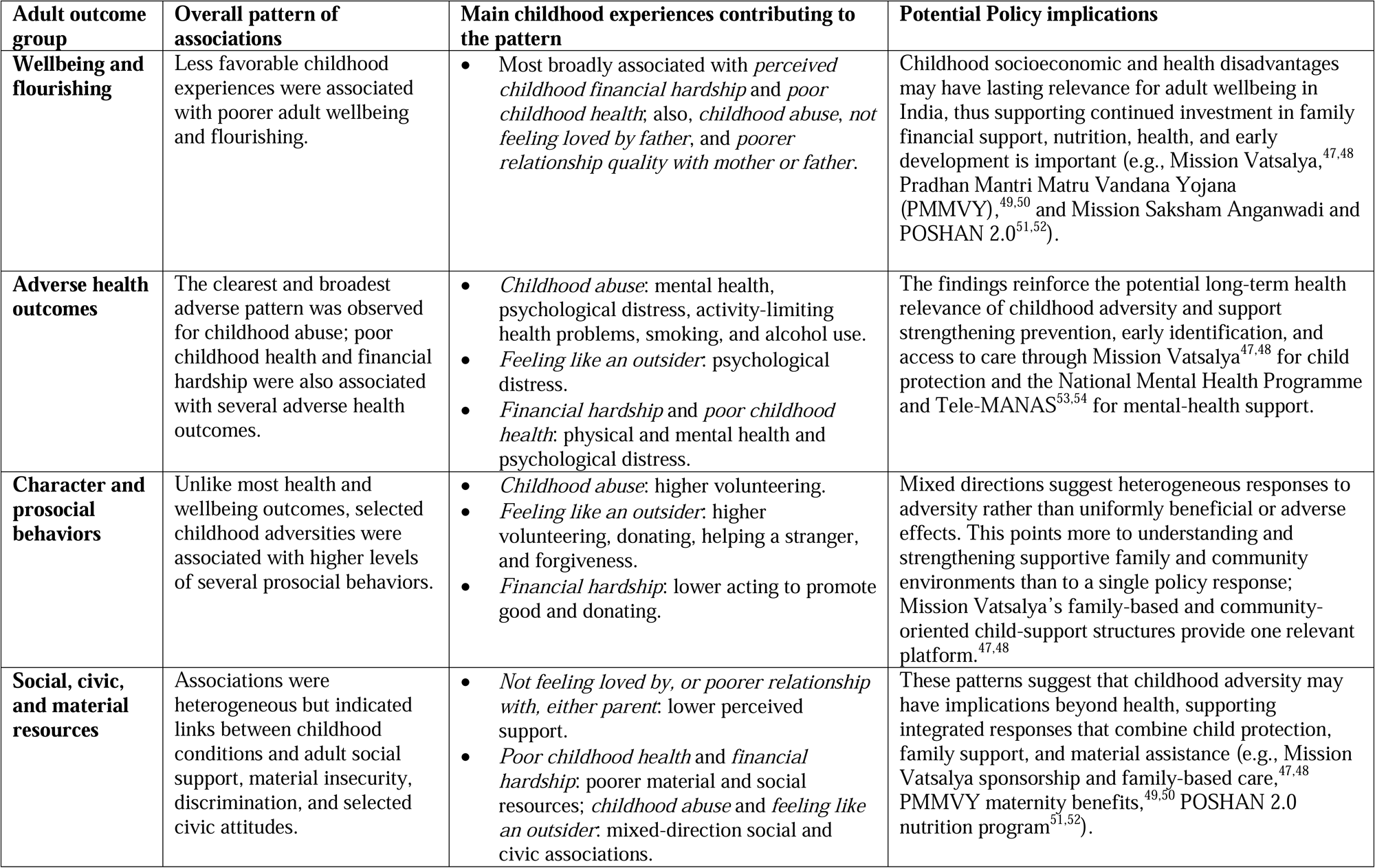
Summary of observed association patterns and potential implications of childhood experiences across adult outcome groups.

| Adult outcome group | Overall pattern of associations | Main childhood experiences contributing to the pattern | Potential Policy implications |
| --- | --- | --- | --- |
| **Wellbeing and flourishing** | Less favorable childhood experiences were associated with poorer adult wellbeing and flourishing. | Most broadly associated with *perceived childhood financial hardship* and *poor childhood health*; also, *childhood abuse*, *not feeling loved by father*, and *poorer relationship quality with mother or father*. | Childhood socioeconomic and health disadvantages may have lasting relevance for adult wellbeing in India, thus supporting continued investment in family financial support, nutrition, health, and early development is important (e.g., Mission Vatsalya,<sup>47,48</sup> Pradhan Mantri Matru Vandana Yojana (PMMVY),<sup>49,50</sup> and Mission Saksham Anganwadi and POSHAN 2.0<sup>51,52</sup>). |
| **Adverse health outcomes** | The clearest and broadest adverse pattern was observed for childhood abuse; poor childhood health and financial hardship were also associated with several adverse health outcomes. | *Childhood abuse*: mental health, psychological distress, activity-limiting health problems, smoking, and alcohol use.<br>*Feeling like an outsider*: psychological distress.<br>*Financial hardship* and *poor childhood health*: physical and mental health and psychological distress. | The findings reinforce the potential long-term health relevance of childhood adversity and support strengthening prevention, early identification, and access to care through Mission Vatsalya<sup>47,48</sup> for child protection and the National Mental Health Programme and Tele-MANAS<sup>53,54</sup> for mental-health support. |
| **Character and prosocial behaviors** | Unlike most health and wellbeing outcomes, selected childhood adversities were associated with higher levels of several prosocial behaviors. | *Childhood abuse*: higher volunteering.<br>*Feeling like an outsider*: higher volunteering, donating, helping a stranger, and forgiveness.<br>*Financial hardship*: lower acting to promote good and donating. | Mixed directions suggest heterogeneous responses to adversity rather than uniformly beneficial or adverse effects. This points more to understanding and strengthening supportive family and community environments than to a single policy response; Mission Vatsalya's family-based and community-oriented child-support structures provide one relevant platform.<sup>47,48</sup> |
| **Social, civic, and material resources** | Associations were heterogeneous but indicated links between childhood conditions and adult social support, material insecurity, discrimination, and selected civic attitudes. | *Not feeling loved by, or poorer relationship with, either parent*: lower perceived support.<br>*Poor childhood health* and *financial hardship*: poorer material and social resources; *childhood abuse* and *feeling like an outsider*: mixed-direction social and civic associations. | These patterns suggest that childhood adversity may have implications beyond health, supporting integrated responses that combine child protection, family support, and material assistance (e.g., Mission Vatsalya sponsorship and family-based care,<sup>47,48</sup> PMMVY maternity benefits,<sup>49,50</sup> POSHAN 2.0 nutrition program<sup>51,52</sup>). |

Associations were broadly similar between females and males, and the only Bonferroni-significant gender interaction, in which poor childhood health was associated with lower self-reported acting to promote good among females but higher among males, may reflect differences in self-perception and gendered norms rather than actual behavior,^55^ and warrants cautious interpretation. Modest gender differences may nevertheless exist for selected childhood experiences and warrant further study (eFigures 3-9). The higher reported prevalence of childhood abuse among males should also be interpreted cautiously because the questionnaire combined physical and sexual abuse and may reflect gender differences in the types of abuse experienced or underreporting among females where stigma, privacy concerns, or disclosure barriers exist.^56,57^ Together, these findings suggest that childhood experiences should be viewed not as a single dimension of adversity but as distinguishable domains relating to different aspects of adult life, paralleling the multidimensional framework underlying the GFS.^25–27,41^

A somewhat surprising finding was that childhood abuse was associated with higher odds of volunteering and feeling like an outsider in one’s family with higher odds of volunteering, charitable giving, and helping a stranger. Previous GFS analyses have similarly reported associations between childhood experiences and adult prosocial behaviors, although these vary in magnitude and direction across countries.^38,39^ These positive-direction associations should not be interpreted as indicating that childhood adversity is beneficial or resilience-promoting, as a recent meta-analysis found a modest overall negative association between childhood maltreatment and prosocial behavior, with heterogeneity across maltreatment types and studies.^58^ Greater past adversity has, in some settings, been associated with empathy-mediated compassion and helping,^59^ and responses to adversity can follow heterogeneous trajectories, though these pathways were not measured here.^60^ In India, prosocial activities may also be shaped by social, community, religious, and family contexts, including among economically disadvantaged groups.^61^ The observed associations may therefore reflect unmeasured cultural, social, religious, or dispositional factors, supportive relationships or other protective resources, or heterogeneous life-course responses, rather than direct consequences of childhood adversity.

Several limitations warrant caution. First, retrospective reports of childhood experiences may be subject to recall and reporting error, which could differ by gender and other social or cultural factors and be influenced by current mental health and life circumstances.^62,63^ For example, respondents with poorer current mental health could be more likely to recall or report experiencing childhood adversities, which could overstate the associations, whereas underreporting of abuse because of stigma would be expected to bias associations towards the null. Second, combining ordinal scales may have obscured differences in severity, and the childhood measures did not capture the duration or accumulation of experiences. Third, potentially important factors were not measured or included, such as caste, religion, childhood family structure, personality, coping, resilience, social support, and self-efficacy, which may shape childhood experiences, their reporting, and adult outcomes, contributing to residual confounding and heterogeneity. Finally, survey attrition, selective survival, and item nonresponse, particularly “don’t know” responses to the abuse item, may have introduced bias, although sensitivity analyses recoding non-substantive responses yielded broadly similar findings and can be found in eResults 2 and eFigures 11-16. We also explored potential heterogeneity by age and residential setting (descriptive distributions in eTables 17-20) and did not observe consistent gender patterns across these subgroups; however, these exploratory findings should be interpreted cautiously (eFigures 17-18).

Despite these limitations, this study is, to our knowledge, one of the most comprehensive examinations of childhood experiences and adult outcomes in a nationally representative sample from India, evaluating eight experiences across 45 outcomes with survey-weighted models and formal gender-interaction tests. Additional sensitivity analyses using repeated observations across survey waves via mixed linear models yielded broadly similar associations (eFigure 19).

The findings extend predominantly Western evidence to a large South Asian sample across the breadth of the flourishing framework. Smoking and alcohol use, most clearly elevated among adults reporting abuse, are one possible behavioral pathway to poorer long-term health, whereas more severe outcomes, such as suicidality and criminal behavior, were beyond the scope of the GFS, so the full range of adverse outcomes could be understated. The positive-direction prosocial associations highlight the need to examine why individuals with similar childhood experiences follow different adult trajectories. Longitudinal and ecological momentary assessment studies could identify when and for whom supportive relationships, positive childhood experiences, and other resources are associated with more favorable outcomes, and whether associations vary by gender, age cohort, socioeconomic position, and cultural context.^4,64,65^ Although India has established child-protection, social-protection, and mental health programs, there still are gaps in implementation, coordination, and access that require consolidated policy work.

## Data Sharing Statement

This study used de-identified individual participant data from Wave 1 (2023) and the 2024 mid-year survey of the Global Flourishing Study (GFS). These data, together with the codebook (data dictionary) and questionnaire, are publicly available through the Center for Open Science (https://www.cos.io/gfs-access-data). Non-sensitive data have been available without restriction since April 8, 2026. Preregistration protocol was developed for this secondary analysis.

## Contributors

**Yiren Hou:** Methodology, Software, Validation, Formal Analysis, Investigation, Data Curation, Writing-Original Draft, Writing-Review & Editing, Visualization; **Tami Sullivan:** Investigation, Writing-Review & Editing; **Emma Zang**: Investigation, Writing-Review & Editing; **Shekhar Saxena:** Conceptualization, Writing-Review & Editing; **Madhu Gupta:** Writing-Review & Editing; **Nandini Chatterjee Singh:** Investigation, Writing-Review & Editing; **Bhramar Mukherjee:** Conceptualization, Methodology, Investigation, Supervision, Writing-Review & Editing; Visualization; Funding acquisition. **YH** and **BM** accessed and verified the underlying data. All authors had full access to all the data in the study and had final responsibility for the decision to submit for publication.

## Acknowledgements/Funding

This work was funded by the Yale School of Public Health. The funder had no role in study design, data collection, data analysis and interpretation, writing of the report, or the decision to submit the paper for publication.

## AI Declaration

During the preparation of this work, GPT-5·6 Sol was used to assist with organizing the supplementary materials, including formatting and arranging the supplementary tables. The authors reviewed, edited, and verified all output and take full responsibility for the content of the publication.

