## Supplementary Materials for "Association of childhood experiences with wellbeing and flourishing outcomes in Indian adults: Evidence from the India Sample of the Global Flourishing Study (GFS)"

**eFigure 1.** Weighted Pearson correlation of 45 adult outcomes and eight childhood variables.

**eFigure 2.** Weighted Spearman's rank correlation of 45 adult outcomes and eight childhood variables.

**eFigure 3.** Adjusted associations between feeling like an outsider in one's family as predictor and 45 outcomes of interest and two flourishing composite indices.

**eFigure 4.** Adjusted associations between not feeling loved by mother as predictor and 45 outcomes of interest and two flourishing composite indices.

**eFigure 5.** Adjusted associations between not feeling loved by father as predictor and 45 outcomes of interest and two flourishing composite indices.

**eFigure 6.** Adjusted associations between bad or very bad quality of relationship with mother as predictor (very good quality as reference category) and 45 outcomes of interest and two flourishing composite indices.

**eFigure 7.** Adjusted associations between bad or very bad quality of relationship with father as predictor (very good quality as reference category) and 45 outcomes of interest and two flourishing composite indices.

**eFigure 8.** Adjusted associations between poorly self-rated childhood health as predictor (excellent self-rated childhood health as reference category) and 45 outcomes of interest and two flourishing composite indices.

**eFigure 9.** Adjusted associations between finding it very difficult in perceived family financial situation as predictor (comfortable perceived family financial situation reference category) and 45 outcomes of interest and two flourishing composite indices.

**eFigure 10.** Adjusted associations between childhood abuse and 14 outcomes from India in the mid-year survey (February-May 2024).

**eFigure 11.** Adjusted associations between childhood abuse and 45 outcomes of interest and two flourishing composite indices, where "don't know" and "refused" responses are counted towards the exposed group.

**eFigure 12.** Adjusted associations between childhood abuse and 45 outcomes of interest and two flourishing composite indices, where "don't know" and "refused" responses are counted towards the unexposed group.

**eFigure 13.** Adjusted associations between not feeling loved by mother and 45 outcomes of interest and two flourishing composite indices, where "does not apply" responses are counted towards the adverse exposure.

**eFigure 14.** Adjusted associations between not feeling loved by father and 45 outcomes of interest and two flourishing composite indices, where "does not apply" responses are counted towards the adverse exposure.

**eFigure 15.** Adjusted associations between bad or very bad relationship with mother and 45 outcomes of interest and two flourishing composite indices, where "does not apply" responses are counted towards the adverse exposure.

**eFigure 16.** Adjusted associations between bad or very bad relationship with father and 45 outcomes of interest and two flourishing composite indices, where "does not apply" responses are counted towards the adverse exposure.

**eFigure 17.** Adjusted associations between childhood predictors and 45 outcomes of interest and two flourishing composite indices for categorized residential location (Rural area or on a farm, small town or village, large city or suburb of a large city).

**eFigure 18.** Adjusted associations between childhood predictors and 22 continuous outcomes of interest and two flourishing composite indices for each age group, 18 to 24 inclusive, 25 to 29 inclusive, 30 to 39 inclusive, 40 to 49 inclusive, 50 to 59 inclusive, and 60+.

**eFigure 19.** Comparison with linear mixed model results using Waves 1 and 2 data.

**eFigure 20.** Weighted correlation between childhood variables and covariate age and current educational attainment.

**eResults 1.** Correlation results.

**eResults 2.** Sensitivity analyses results for item nonresponse in questions regarding childhood abuse, feeling not loved by mother and by father, and quality of relationship with mother and with father.

**eTable 1.** Description of eight childhood predictors and 45 outcomes of interest.

**eTable 2.** Flourishing domain, its indicators, questions, and responses, in the Global Flourishing Study.

**eTable 3.** Description of 8 childhood predictors and 14 outcomes of interest from mid-year items fielded between Year 1 and 2.

**eTable 4.** Descriptive statistics of states and union territories in India (N = 12,765) in Wave 1 Global Flourishing Data.

**eTable 5.** Weighted Pearson correlation of 45 adult outcomes and eight childhood variables.

**eTable 6.** Weighted Spearman's rank correlation of 45 adult outcomes and eight childhood variables.

**eTable 7.** Table of coefficients for adjusted associations between childhood abuse as predictor and 45 outcomes of interest and two flourishing composite indices.

**eTable 8.** Table of coefficients for adjusted associations between feeling like an outsider in one's family as predictor and 45 outcomes of interest and two flourishing composite indices.

**eTable 9.** Table of coefficients for adjusted associations between not feeling loved by mother as predictor and 45 outcomes of interest and two flourishing composite indices.

**eTable 10.** Table of coefficients for adjusted associations between not feeling loved by father as predictor and 45 outcomes of interest and two flourishing composite indices.

**eTable 11.** Table of coefficients for adjusted associations between bad or very bad quality of relationship with mother as predictor (very good quality as reference category) and 45 outcomes of interest and two flourishing composite indices.

**eTable 12.** Table of coefficients for adjusted associations between bad or very bad quality of relationship with father as predictor (very good quality as reference category) and 45 outcomes of interest and two flourishing composite indices.

**eTable 13.** Table of coefficients for adjusted associations between poorly self-rated childhood health as predictor (excellent self-rated childhood health as reference category) and 45 outcomes of interest and two flourishing composite indices.

**eTable 14.** Table of coefficients for adjusted associations between finding it very difficult in perceived family financial situation as predictor (comfortable perceived family financial situation reference category) and 45 outcomes of interest and two flourishing composite indices.

**eTable 15.** Demographic statistics of unweighted sample sizes (n) and weighted proportions (%) using responses collected in the Wave 1 survey (2023) in India and stratified by gender and survey mode (N = 12,765).

**eTable 16.** Table of coefficients for adjusted associations between childhood abuse and 14 outcomes from India in the mid-year survey (February-May 2024).

**eTable 17.** Distribution of childhood predictors collected in Wave 1 survey (2023) in India and stratified by survey mode and residential location (N = 12,765).

**eTable 18.** Distribution of outcome variables collected in Wave 1 survey (2023) in India and stratified by survey mode and residential location (N = 12,765).

**eTable 19.** Distribution of childhood predictors collected in Wave 1 survey (2023) in India and stratified by age groups 18 to 24 inclusive, 25 to 29 inclusive, 30 to 39 inclusive, 40 to 49 inclusive, 50 to 59 inclusive, and 60+ (N = 12,765).

**eTable 20.** Distribution of outcome variables collected in Wave 1 survey (2023) in India and stratified by age groups 18 to 24 inclusive, 25 to 29 inclusive, 30 to 39 inclusive, 40 to 49 inclusive, 50 to 59 inclusive, and 60+ (N = 12,765).

**eTable 21.** Parental marital status at approximately age 12 among respondents selecting "does not apply" for parental experience items, reporting unweighted sample size and weighted proportions.

**eFigure 1.** Weighted Pearson correlation of 45 adult outcomes and eight childhood variables.

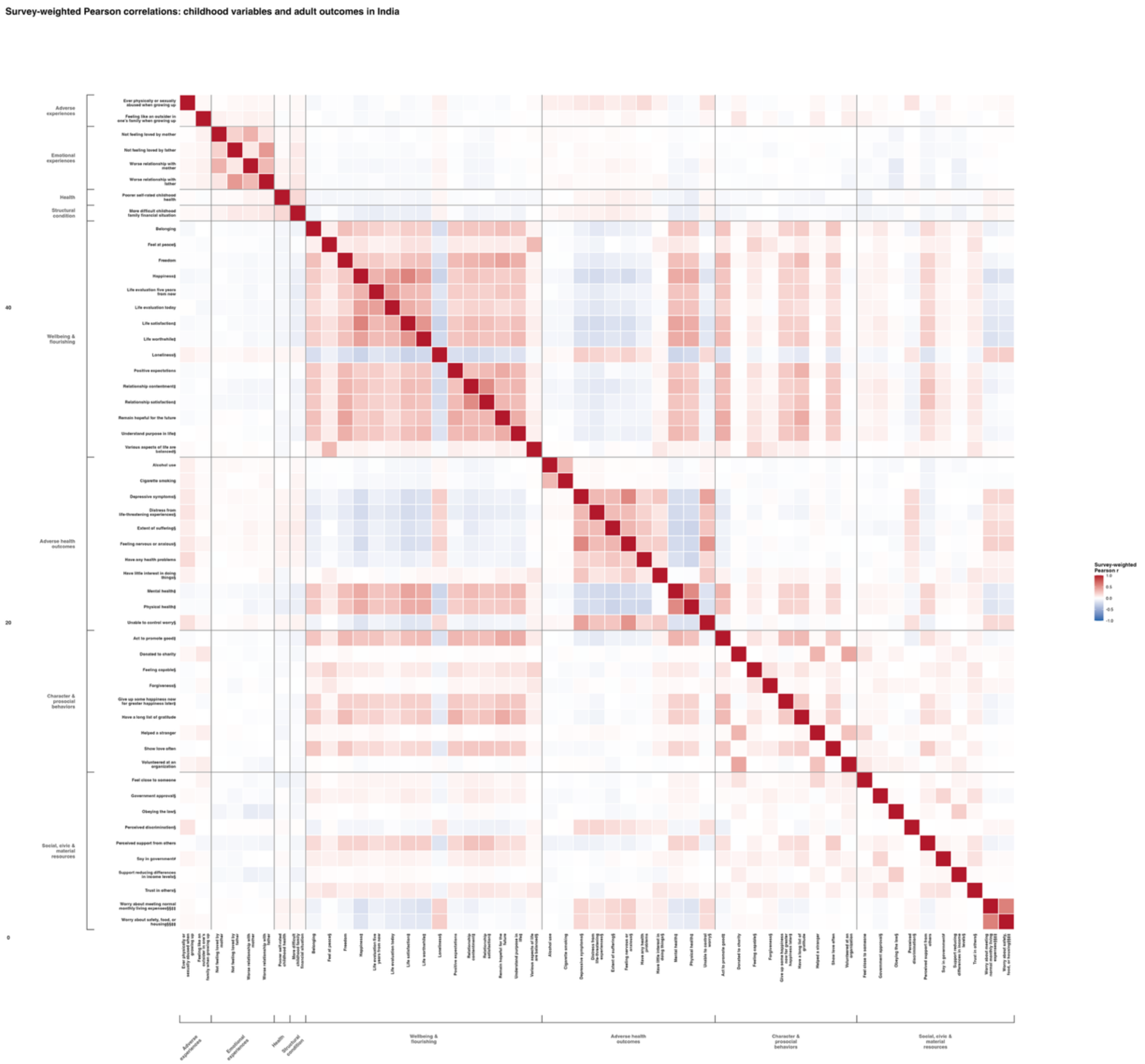

**eFigure 2.** Weighted Spearman's rank correlation of 45 adult outcomes and eight childhood variables.

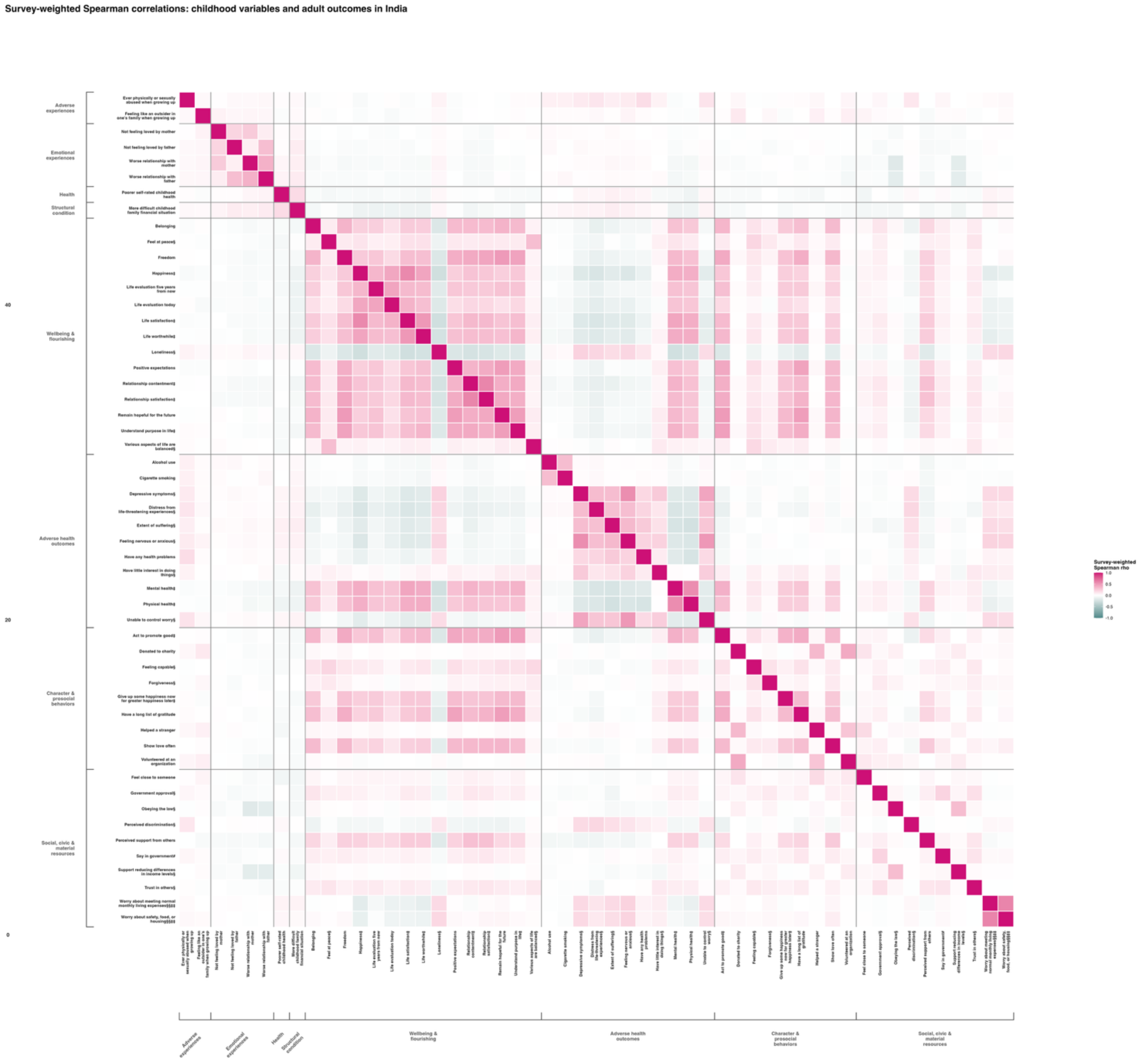

**eFigure 3.** Adjusted associations between feeling like an outsider in one's family as predictor and 45 outcomes of interest and two flourishing composite indices.

Each panel shows the associations between the predictor and each of the continuous outcomes (**Panel A**), binary outcomes (**Panel B**), and ordinal outcomes (**Panel C**).

**Panel A**

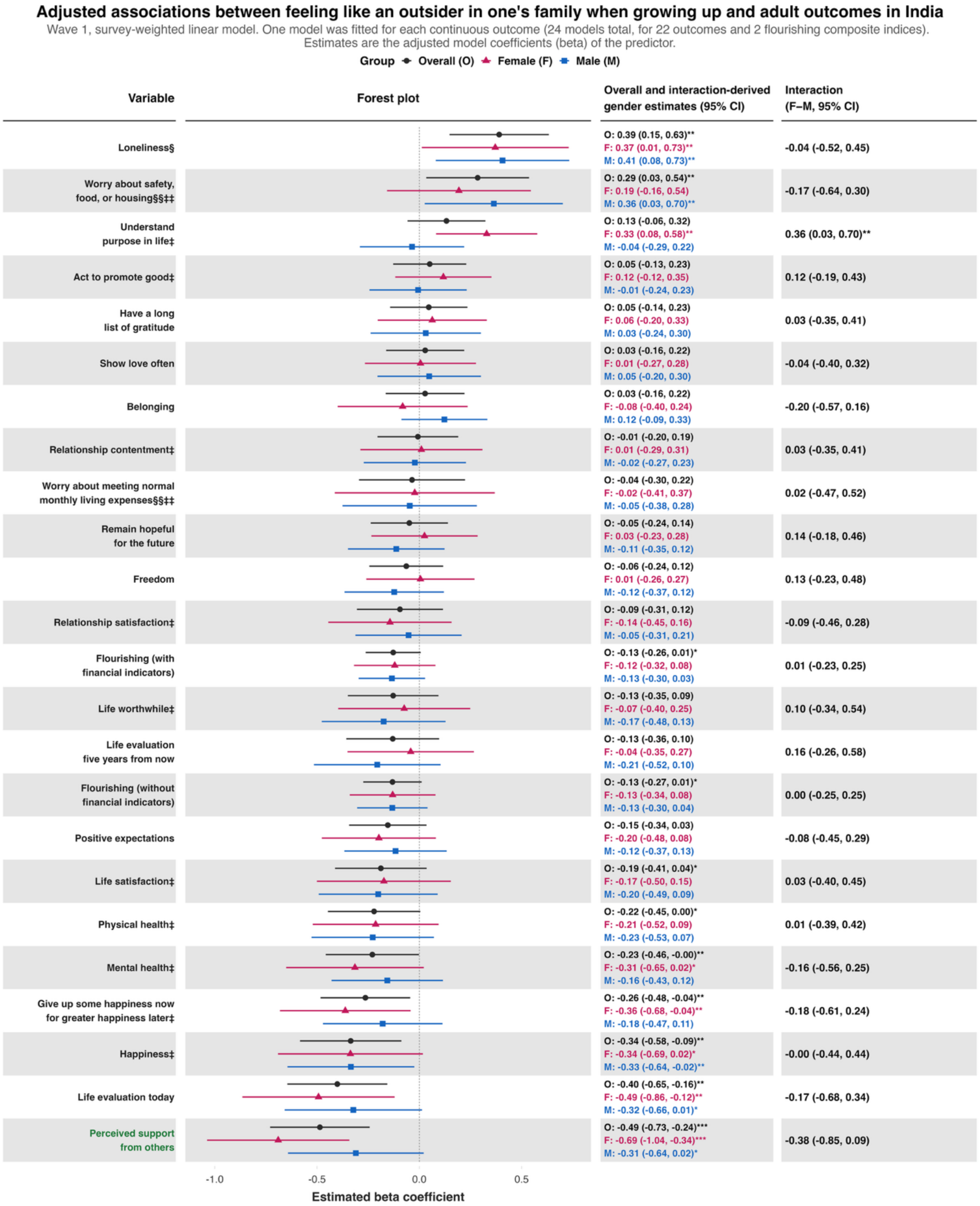

**Panel B**

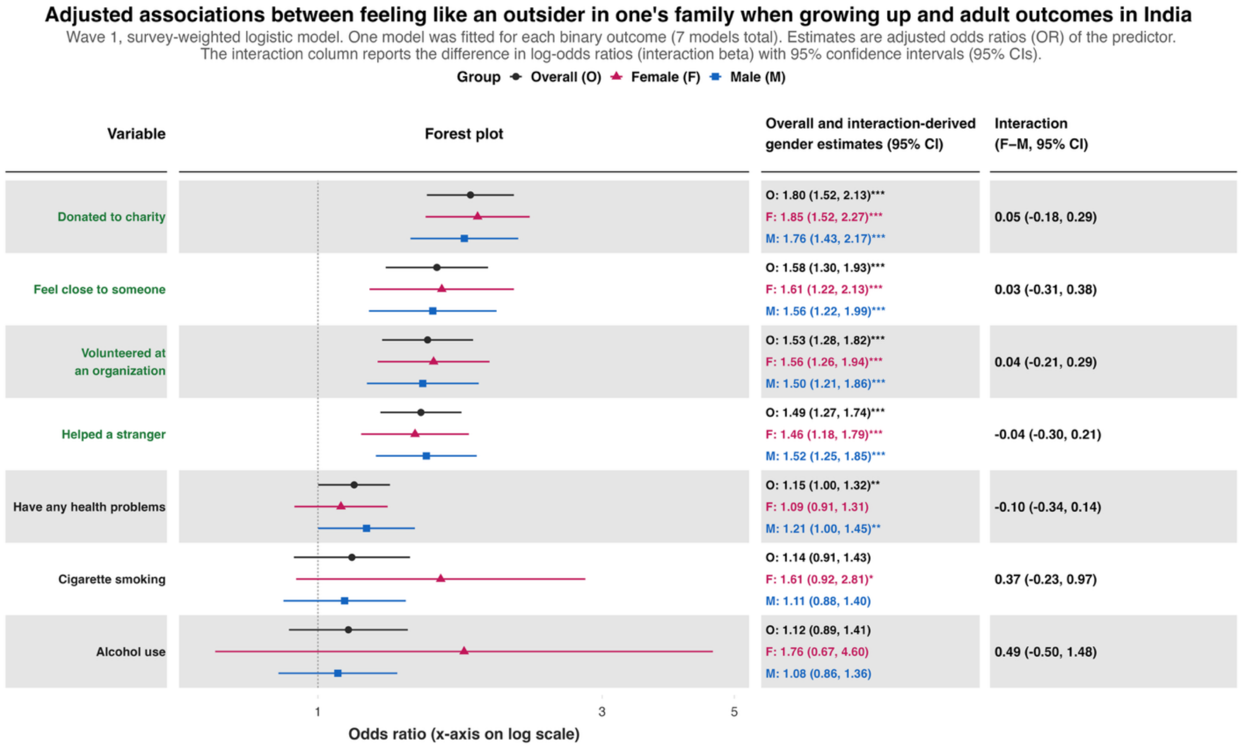
Adjusted associations between feeling like an outsider in one's family when growing up and adult outcomes in India
Wave 1, survey-weighted logistic model. One model was fitted for each binary outcome (7 models total). Estimates are adjusted odds ratios (OR) of the predictor.
The interaction column reports the difference in log-odds ratios (interaction beta) with 95% confidence intervals (95% CIs).
Group Overall (O) Female (F) Male (M)
Variable
Forest plot
Overall and interaction-derived gender estimates (95% CI)
Interaction (F–M, 95% CI)
Donated to charity
O: 1.80 (1.52, 2.13)***
F: 1.85 (1.52, 2.27)***
M: 1.76 (1.43, 2.17)***
0.05 (-0.18, 0.29)
Feel close to someone
O: 1.58 (1.30, 1.93)***
F: 1.61 (1.22, 2.13)***
M: 1.56 (1.22, 1.99)***
0.03 (-0.31, 0.38)
Volunteered at an organization
O: 1.53 (1.28, 1.82)***
F: 1.56 (1.26, 1.94)***
M: 1.50 (1.21, 1.86)***
0.04 (-0.21, 0.29)
Helped a stranger
O: 1.49 (1.27, 1.74)***
F: 1.46 (1.18, 1.79)***
M: 1.52 (1.25, 1.85)***
-0.04 (-0.30, 0.21)
Have any health problems
O: 1.15 (1.00, 1.32)**
F: 1.09 (0.91, 1.31)
M: 1.21 (1.00, 1.45)**
-0.10 (-0.34, 0.14)
Cigarette smoking
O: 1.14 (0.91, 1.43)
F: 1.61 (0.92, 2.81)*
M: 1.11 (0.88, 1.40)
0.37 (-0.23, 0.97)
Alcohol use
O: 1.12 (0.89, 1.41)
F: 1.76 (0.67, 4.60)
M: 1.08 (0.86, 1.36)
0.49 (-0.50, 1.48)
1
3
5
Odds ratio (x-axis on log scale)

**Panel C**

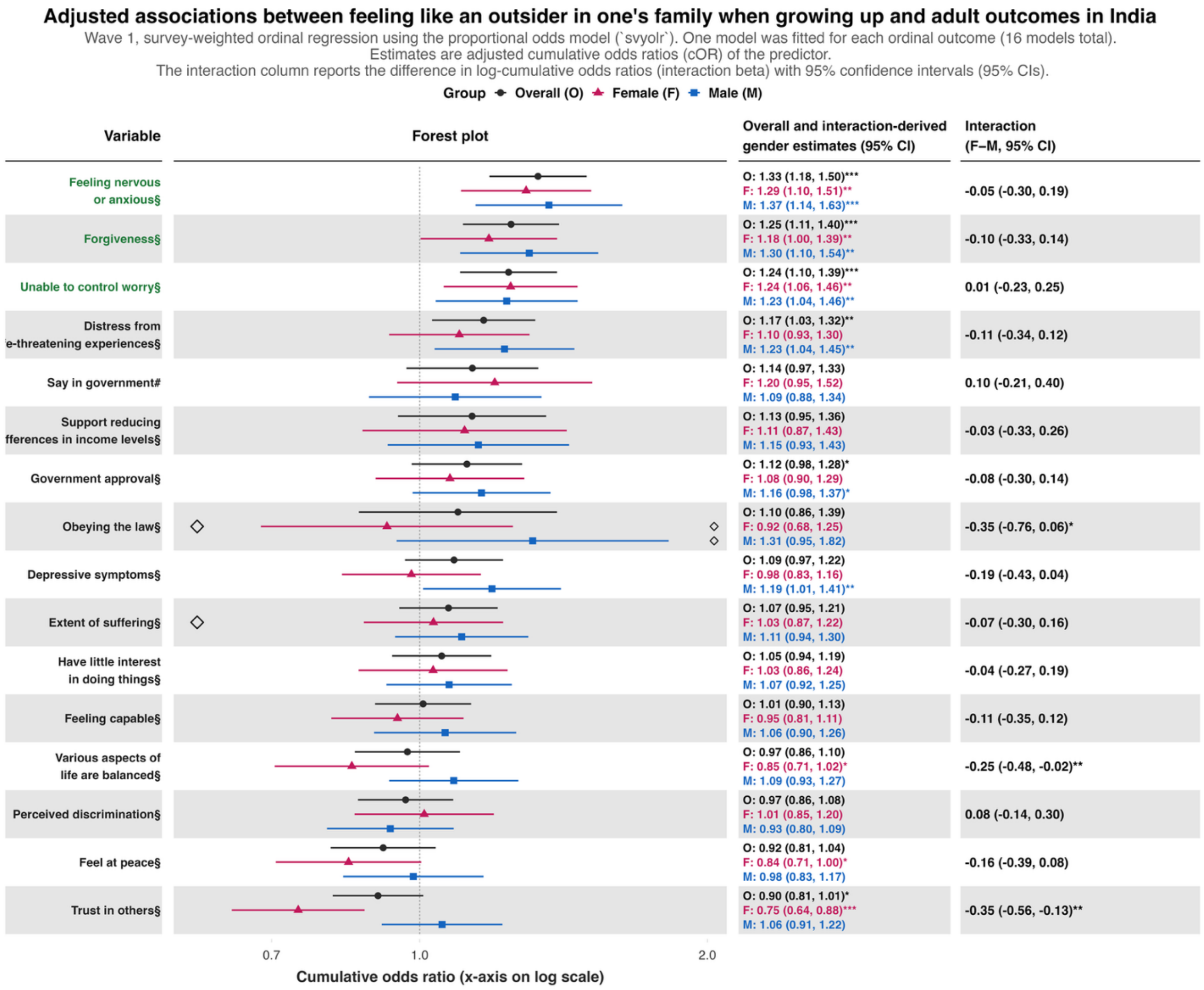
Adjusted associations between feeling like an outsider in one's family when growing up and adult outcomes in India
Wave 1, survey-weighted ordinal regression using the proportional odds model (`svyolr`). One model was fitted for each ordinal outcome (16 models total).
Estimates are adjusted cumulative odds ratios (cOR) of the predictor.
The interaction column reports the difference in log-cumulative odds ratios (interaction beta) with 95% confidence intervals (95% CIs).
Group Overall (O) Female (F) Male (M)
Variable
Forest plot
Overall and interaction-derived gender estimates (95% CI)
Interaction (F–M, 95% CI)
Feeling nervous or anxious§
O: 1.33 (1.18, 1.50)*** F: 1.29 (1.10, 1.51)** M: 1.37 (1.14, 1.63)***
-0.05 (-0.30, 0.19)
Forgiveness§
O: 1.25 (1.11, 1.40)*** F: 1.18 (1.00, 1.39)** M: 1.30 (1.10, 1.54)**
-0.10 (-0.33, 0.14)
Unable to control worry§
O: 1.24 (1.10, 1.39)*** F: 1.24 (1.06, 1.46)** M: 1.23 (1.04, 1.46)**
0.01 (-0.23, 0.25)
Distress from e-threatening experiences§
O: 1.17 (1.03, 1.32)** F: 1.10 (0.93, 1.30) M: 1.23 (1.04, 1.45)**
-0.11 (-0.34, 0.12)
Say in government#
O: 1.14 (0.97, 1.33) F: 1.20 (0.95, 1.52) M: 1.09 (0.88, 1.34)
0.10 (-0.21, 0.40)
Support reducing fferences in income levels§
O: 1.13 (0.95, 1.36) F: 1.11 (0.87, 1.43) M: 1.15 (0.93, 1.43)
-0.03 (-0.33, 0.26)
Government approval§
O: 1.12 (0.98, 1.28)* F: 1.08 (0.90, 1.29) M: 1.16 (0.98, 1.37)*
-0.08 (-0.30, 0.14)
Obeying the law§
O: 1.10 (0.86, 1.39) F: 0.92 (0.68, 1.25) M: 1.31 (0.95, 1.82)
-0.35 (-0.76, 0.06)*
Depressive symptoms§
O: 1.09 (0.97, 1.22) F: 0.98 (0.83, 1.16) M: 1.19 (1.01, 1.41)**
-0.19 (-0.43, 0.04)
Extent of suffering§
O: 1.07 (0.95, 1.21) F: 1.03 (0.87, 1.22) M: 1.11 (0.94, 1.30)
-0.07 (-0.30, 0.16)
Have little interest in doing things§
O: 1.05 (0.94, 1.19) F: 1.03 (0.86, 1.24) M: 1.07 (0.92, 1.25)
-0.04 (-0.27, 0.19)
Feeling capable§
O: 1.01 (0.90, 1.13) F: 0.95 (0.81, 1.11) M: 1.06 (0.90, 1.26)
-0.11 (-0.35, 0.12)
Various aspects of life are balanced§
O: 0.97 (0.86, 1.10) F: 0.85 (0.71, 1.02)* M: 1.09 (0.93, 1.27)
-0.25 (-0.48, -0.02)**
Perceived discrimination§
O: 0.97 (0.86, 1.08) F: 1.01 (0.85, 1.20) M: 0.93 (0.80, 1.09)
0.08 (-0.14, 0.30)
Feel at peace§
O: 0.92 (0.81, 1.04) F: 0.84 (0.71, 1.00)* M: 0.98 (0.83, 1.17)
-0.16 (-0.39, 0.08)
Trust in others§
O: 0.90 (0.81, 1.01)* F: 0.75 (0.64, 0.88)*** M: 1.06 (0.91, 1.22)
-0.35 (-0.56, -0.13)**
0.7
1.0
2.0
Cumulative odds ratio (x-axis on log scale)

**eFigure 4.** Adjusted associations between not feeling loved by mother as predictor and 45 outcomes of interest and two flourishing composite indices.

Each panel shows the associations between the predictor and each of the continuous outcomes (**Panel A**), binary outcomes (**Panel B**), and ordinal outcomes (**Panel C**).

**Panel A**

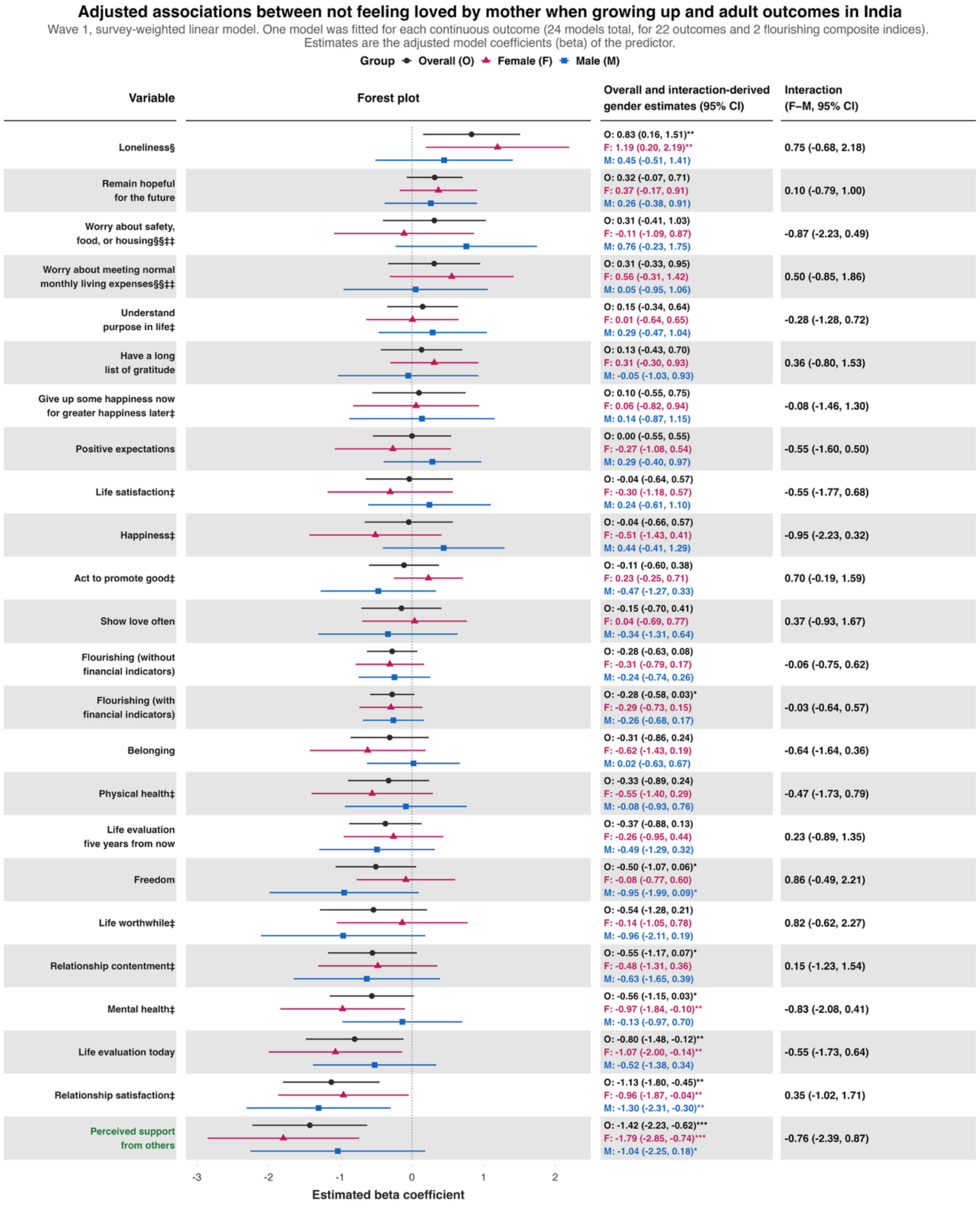

**Panel B**

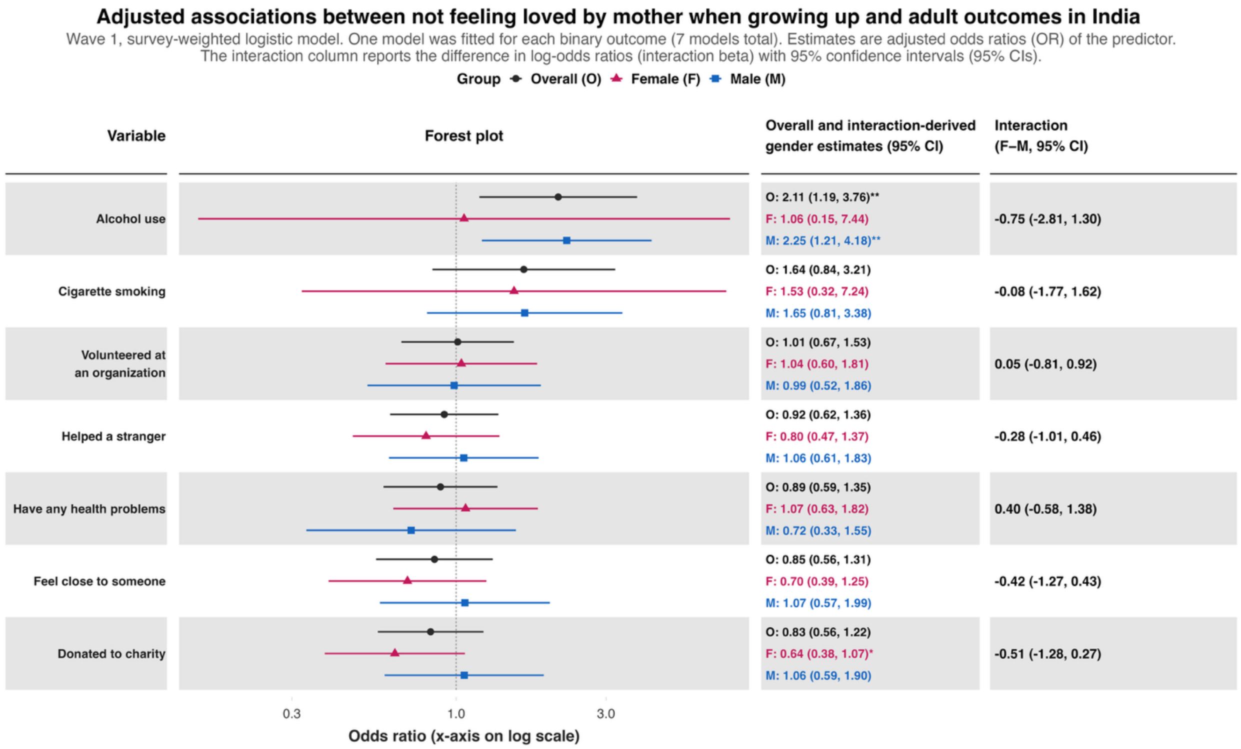
Adjusted associations between not feeling loved by mother when growing up and adult outcomes in India
Wave 1, survey-weighted logistic model. One model was fitted for each binary outcome (7 models total). Estimates are adjusted odds ratios (OR) of the predictor.
The interaction column reports the difference in log-odds ratios (interaction beta) with 95% confidence intervals (95% CIs).
Group Overall (O) Female (F) Male (M)
Variable
Forest plot
Overall and interaction-derived gender estimates (95% CI)
Interaction (F–M, 95% CI)
Alcohol use
O: 2.11 (1.19, 3.76)**
F: 1.06 (0.15, 7.44)
M: 2.25 (1.21, 4.18)**
-0.75 (-2.81, 1.30)
Cigarette smoking
O: 1.64 (0.84, 3.21)
F: 1.53 (0.32, 7.24)
M: 1.65 (0.81, 3.38)
-0.08 (-1.77, 1.62)
Volunteered at an organization
O: 1.01 (0.67, 1.53)
F: 1.04 (0.60, 1.81)
M: 0.99 (0.52, 1.86)
0.05 (-0.81, 0.92)
Helped a stranger
O: 0.92 (0.62, 1.36)
F: 0.80 (0.47, 1.37)
M: 1.06 (0.61, 1.83)
-0.28 (-1.01, 0.46)
Have any health problems
O: 0.89 (0.59, 1.35)
F: 1.07 (0.63, 1.82)
M: 0.72 (0.33, 1.55)
0.40 (-0.58, 1.38)
Feel close to someone
O: 0.85 (0.56, 1.31)
F: 0.70 (0.39, 1.25)
M: 1.07 (0.57, 1.99)
-0.42 (-1.27, 0.43)
Donated to charity
O: 0.83 (0.56, 1.22)
F: 0.64 (0.38, 1.07)*
M: 1.06 (0.59, 1.90)
-0.51 (-1.28, 0.27)
0.3
1.0
3.0
Odds ratio (x-axis on log scale)

**Panel C**

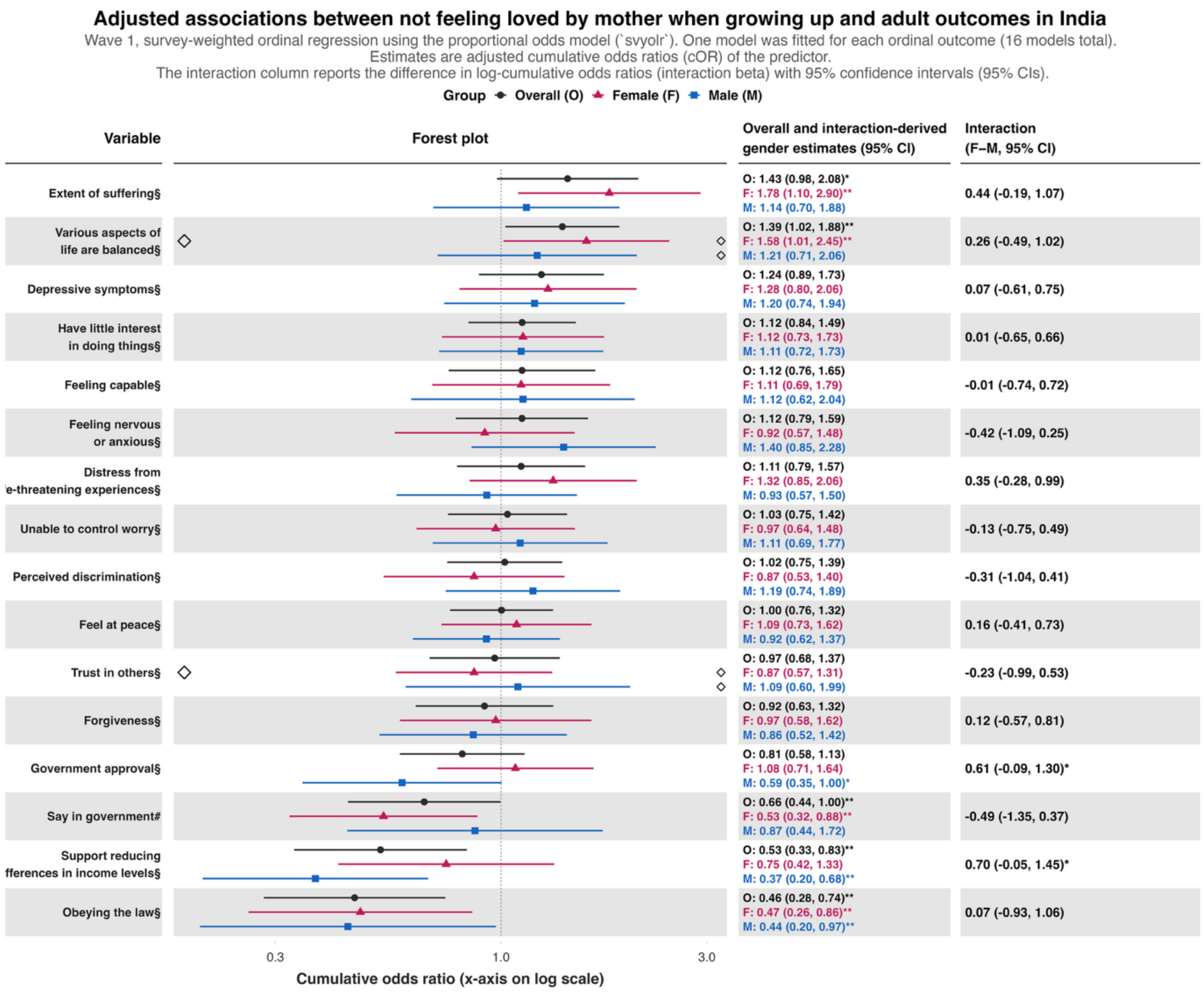
Adjusted associations between not feeling loved by mother when growing up and adult outcomes in India
Wave 1, survey-weighted ordinal regression using the proportional odds model (`svyolr`). One model was fitted for each ordinal outcome (16 models total).
Estimates are adjusted cumulative odds ratios (cOR) of the predictor.
The interaction column reports the difference in log-cumulative odds ratios (interaction beta) with 95% confidence intervals (95% CIs).
Group Overall (O) Female (F) Male (M)
Variable
Forest plot
Overall and interaction-derived gender estimates (95% CI)
Interaction (F–M, 95% CI)
Extent of suffering§
O: 1.43 (0.98, 2.08)*
F: 1.78 (1.10, 2.90)**
M: 1.14 (0.70, 1.88)
0.44 (-0.19, 1.07)
Various aspects of life are balanced§
O: 1.39 (1.02, 1.88)**
F: 1.58 (1.01, 2.45)**
M: 1.21 (0.71, 2.06)
0.26 (-0.49, 1.02)
Depressive symptoms§
O: 1.24 (0.89, 1.73)
F: 1.28 (0.80, 2.06)
M: 1.20 (0.74, 1.94)
0.07 (-0.61, 0.75)
Have little interest in doing things§
O: 1.12 (0.84, 1.49)
F: 1.12 (0.73, 1.73)
M: 1.11 (0.72, 1.73)
0.01 (-0.65, 0.66)
Feeling capable§
O: 1.12 (0.76, 1.65)
F: 1.11 (0.69, 1.79)
M: 1.12 (0.62, 2.04)
-0.01 (-0.74, 0.72)
Feeling nervous or anxious§
O: 1.12 (0.79, 1.59)
F: 0.92 (0.57, 1.48)
M: 1.40 (0.85, 2.28)
-0.42 (-1.09, 0.25)
Distress from e-threatening experiences§
O: 1.11 (0.79, 1.57)
F: 1.32 (0.85, 2.06)
M: 0.93 (0.57, 1.50)
0.35 (-0.28, 0.99)
Unable to control worry§
O: 1.03 (0.75, 1.42)
F: 0.97 (0.64, 1.48)
M: 1.11 (0.69, 1.77)
-0.13 (-0.75, 0.49)
Perceived discrimination§
O: 1.02 (0.75, 1.39)
F: 0.87 (0.53, 1.40)
M: 1.19 (0.74, 1.89)
-0.31 (-1.04, 0.41)
Feel at peace§
O: 1.00 (0.76, 1.32)
F: 1.09 (0.73, 1.62)
M: 0.92 (0.62, 1.37)
0.16 (-0.41, 0.73)
Trust in others§
O: 0.97 (0.68, 1.37)
F: 0.87 (0.57, 1.31)
M: 1.09 (0.60, 1.99)
-0.23 (-0.99, 0.53)
Forgiveness§
O: 0.92 (0.63, 1.32)
F: 0.97 (0.58, 1.62)
M: 0.86 (0.52, 1.42)
0.12 (-0.57, 0.81)
Government approval§
O: 0.81 (0.58, 1.13)
F: 1.08 (0.71, 1.64)
M: 0.59 (0.35, 1.00)*
0.61 (-0.09, 1.30)*
Say in government#
O: 0.66 (0.44, 1.00)**
F: 0.53 (0.32, 0.88)**
M: 0.87 (0.44, 1.72)
-0.49 (-1.35, 0.37)
Support reducing fferences in income levels§
O: 0.53 (0.33, 0.83)**
F: 0.75 (0.42, 1.33)
M: 0.37 (0.20, 0.68)**
0.70 (-0.05, 1.45)*
Obeying the law§
O: 0.46 (0.28, 0.74)**
F: 0.47 (0.26, 0.86)**
M: 0.44 (0.20, 0.97)**
0.07 (-0.93, 1.06)
0.3
1.0
3.0
Cumulative odds ratio (x-axis on log scale)

**eFigure 5.** Adjusted associations between not feeling loved by father as predictor and 45 outcomes of interest and two flourishing composite indices.

Each panel shows the associations between the predictor and each of the continuous outcomes (**Panel A**), binary outcomes (**Panel B**), and ordinal outcomes (**Panel C**).

**Panel A**

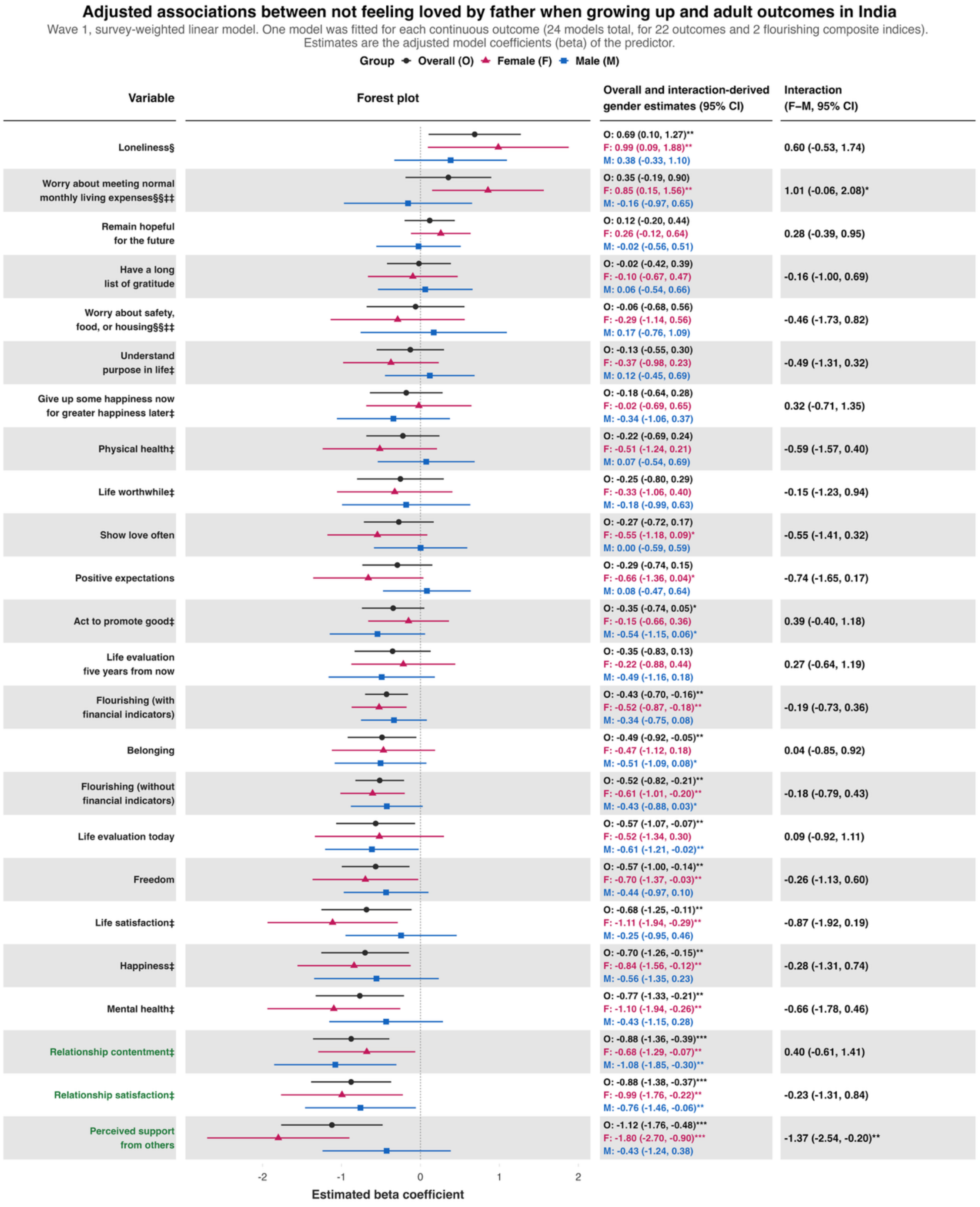

**Panel B**

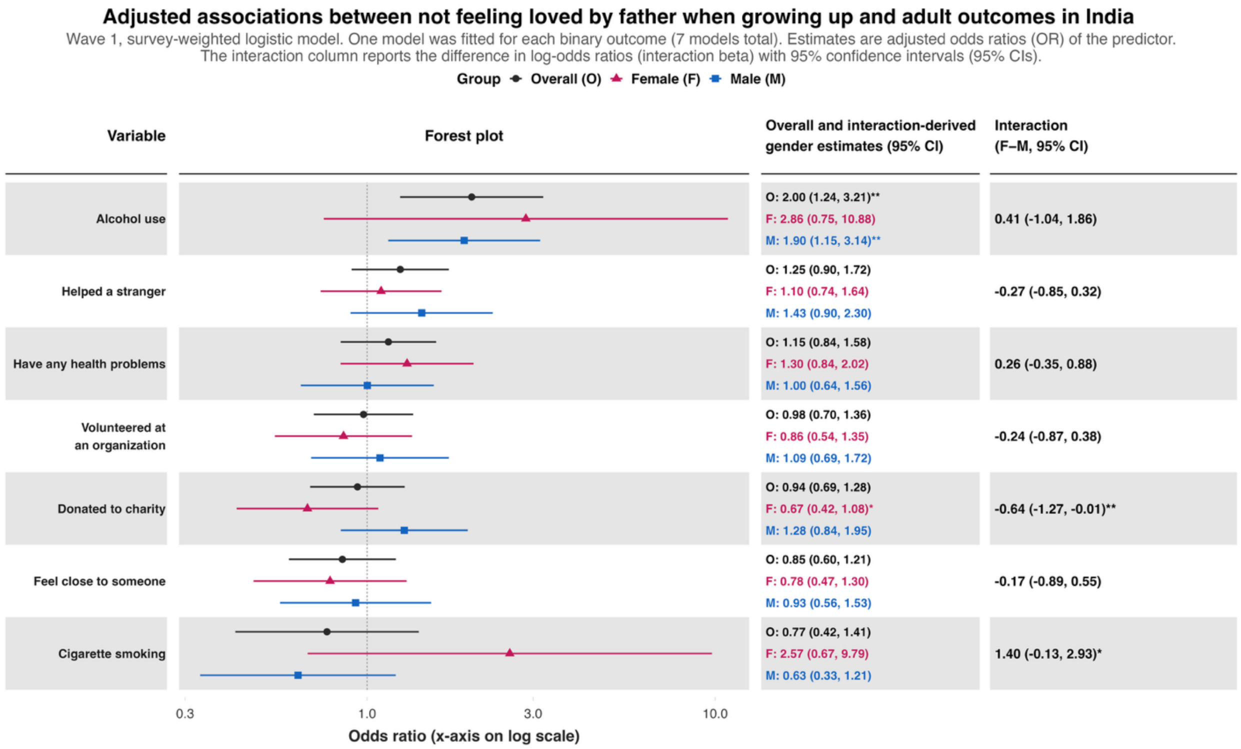
Adjusted associations between not feeling loved by father when growing up and adult outcomes in India
Wave 1, survey-weighted logistic model. One model was fitted for each binary outcome (7 models total). Estimates are adjusted odds ratios (OR) of the predictor.
The interaction column reports the difference in log-odds ratios (interaction beta) with 95% confidence intervals (95% CIs).
Group
Overall (O)
Female (F)
Male (M)
Variable
Forest plot
Overall and interaction-derived gender estimates (95% CI)
Interaction (F–M, 95% CI)
Alcohol use
O: 2.00 (1.24, 3.21)**
F: 2.86 (0.75, 10.88)
M: 1.90 (1.15, 3.14)**
0.41 (-1.04, 1.86)
Helped a stranger
O: 1.25 (0.90, 1.72)
F: 1.10 (0.74, 1.64)
M: 1.43 (0.90, 2.30)
-0.27 (-0.85, 0.32)
Have any health problems
O: 1.15 (0.84, 1.58)
F: 1.30 (0.84, 2.02)
M: 1.00 (0.64, 1.56)
0.26 (-0.35, 0.88)
Volunteered at an organization
O: 0.98 (0.70, 1.36)
F: 0.86 (0.54, 1.35)
M: 1.09 (0.69, 1.72)
-0.24 (-0.87, 0.38)
Donated to charity
O: 0.94 (0.69, 1.28)
F: 0.67 (0.42, 1.08)*
M: 1.28 (0.84, 1.95)
-0.64 (-1.27, -0.01)**
Feel close to someone
O: 0.85 (0.60, 1.21)
F: 0.78 (0.47, 1.30)
M: 0.93 (0.56, 1.53)
-0.17 (-0.89, 0.55)
Cigarette smoking
O: 0.77 (0.42, 1.41)
F: 2.57 (0.67, 9.79)
M: 0.63 (0.33, 1.21)
1.40 (-0.13, 2.93)*
0.3
1.0
3.0
10.0
Odds ratio (x-axis on log scale)

**Panel C**

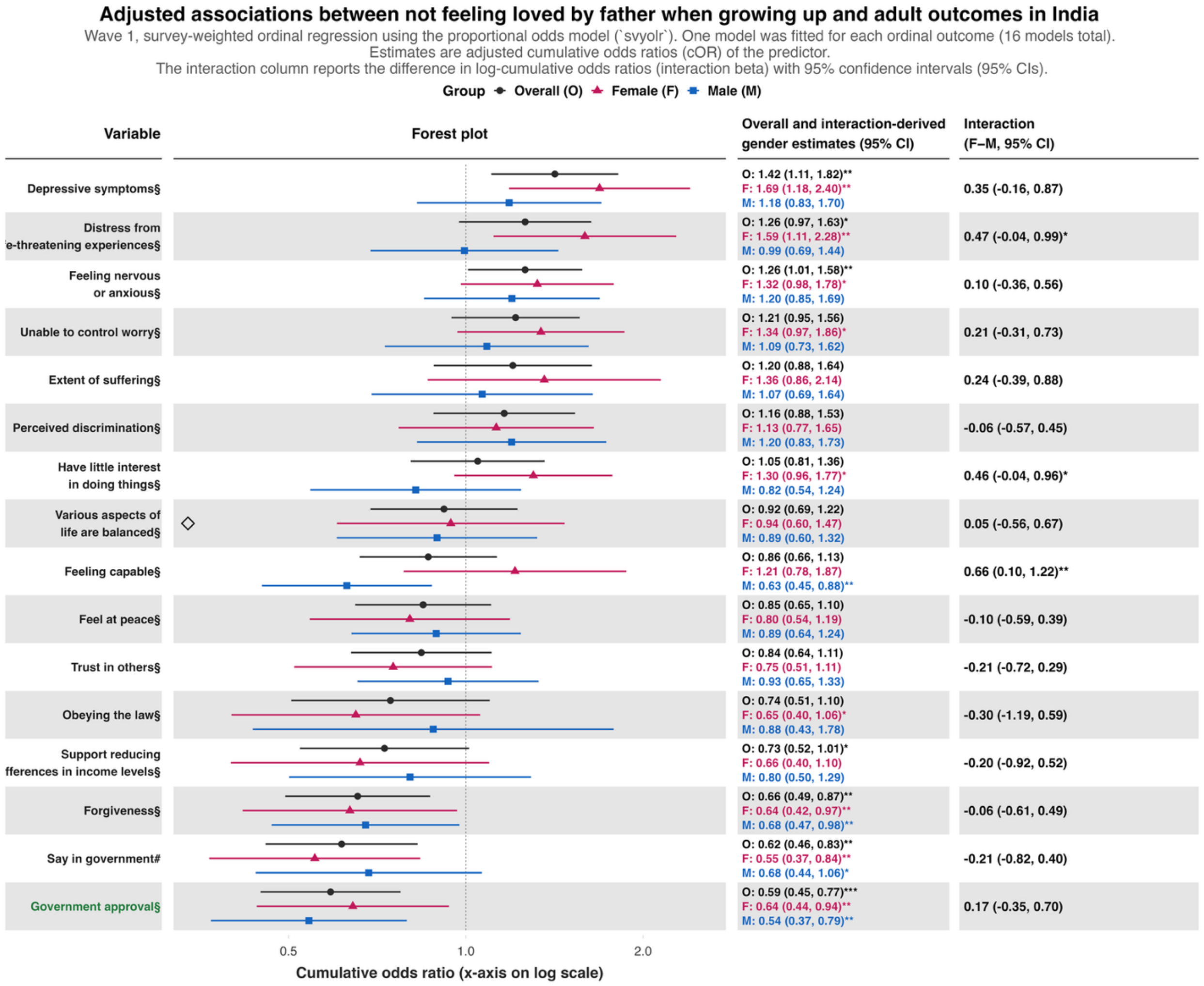
Adjusted associations between not feeling loved by father when growing up and adult outcomes in India
Wave 1, survey-weighted ordinal regression using the proportional odds model (`svyolr`). One model was fitted for each ordinal outcome (16 models total).
Estimates are adjusted cumulative odds ratios (cOR) of the predictor.
The interaction column reports the difference in log-cumulative odds ratios (interaction beta) with 95% confidence intervals (95% CIs).
Group Overall (O) Female (F) Male (M)
Variable
Forest plot
Overall and interaction-derived gender estimates (95% CI)
Interaction (F–M, 95% CI)
Depressive symptoms§
O: 1.42 (1.11, 1.82)**
F: 1.69 (1.18, 2.40)**
M: 1.18 (0.83, 1.70)
0.35 (-0.16, 0.87)
Distress from e-threatening experiences§
O: 1.26 (0.97, 1.63)*
F: 1.59 (1.11, 2.28)**
M: 0.99 (0.69, 1.44)
0.47 (-0.04, 0.99)*
Feeling nervous or anxious§
O: 1.26 (1.01, 1.58)**
F: 1.32 (0.98, 1.78)*
M: 1.20 (0.85, 1.69)
0.10 (-0.36, 0.56)
Unable to control worry§
O: 1.21 (0.95, 1.56)
F: 1.34 (0.97, 1.86)*
M: 1.09 (0.73, 1.62)
0.21 (-0.31, 0.73)
Extent of suffering§
O: 1.20 (0.88, 1.64)
F: 1.36 (0.86, 2.14)
M: 1.07 (0.69, 1.64)
0.24 (-0.39, 0.88)
Perceived discrimination§
O: 1.16 (0.88, 1.53)
F: 1.13 (0.77, 1.65)
M: 1.20 (0.83, 1.73)
-0.06 (-0.57, 0.45)
Have little interest in doing things§
O: 1.05 (0.81, 1.36)
F: 1.30 (0.96, 1.77)*
M: 0.82 (0.54, 1.24)
0.46 (-0.04, 0.96)*
Various aspects of life are balanced§
O: 0.92 (0.69, 1.22)
F: 0.94 (0.60, 1.47)
M: 0.89 (0.60, 1.32)
0.05 (-0.56, 0.67)
Feeling capable§
O: 0.86 (0.66, 1.13)
F: 1.21 (0.78, 1.87)
M: 0.63 (0.45, 0.88)**
0.66 (0.10, 1.22)**
Feel at peace§
O: 0.85 (0.65, 1.10)
F: 0.80 (0.54, 1.19)
M: 0.89 (0.64, 1.24)
-0.10 (-0.59, 0.39)
Trust in others§
O: 0.84 (0.64, 1.11)
F: 0.75 (0.51, 1.11)
M: 0.93 (0.65, 1.33)
-0.21 (-0.72, 0.29)
Obeying the law§
O: 0.74 (0.51, 1.10)
F: 0.65 (0.40, 1.06)*
M: 0.88 (0.43, 1.78)
-0.30 (-1.19, 0.59)
Support reducing fferences in income levels§
O: 0.73 (0.52, 1.01)*
F: 0.66 (0.40, 1.10)
M: 0.80 (0.50, 1.29)
-0.20 (-0.92, 0.52)
Forgiveness§
O: 0.66 (0.49, 0.87)**
F: 0.64 (0.42, 0.97)**
M: 0.68 (0.47, 0.98)**
-0.06 (-0.61, 0.49)
Say in government#
O: 0.62 (0.46, 0.83)**
F: 0.55 (0.37, 0.84)**
M: 0.68 (0.44, 1.06)*
-0.21 (-0.82, 0.40)
Government approval§
O: 0.59 (0.45, 0.77)***
F: 0.64 (0.44, 0.94)**
M: 0.54 (0.37, 0.79)**
0.17 (-0.35, 0.70)
0.5
1.0
2.0
Cumulative odds ratio (x-axis on log scale)

**eFigure 6.** Adjusted associations between bad or very bad quality of relationship with mother as predictor (very good quality as reference category) and 45 outcomes of interest and two flourishing composite indices.

Each panel shows the associations between the predictor and each of the continuous outcomes (**Panel A**), binary outcomes (**Panel B**), and ordinal outcomes (**Panel C**).

**Panel A**

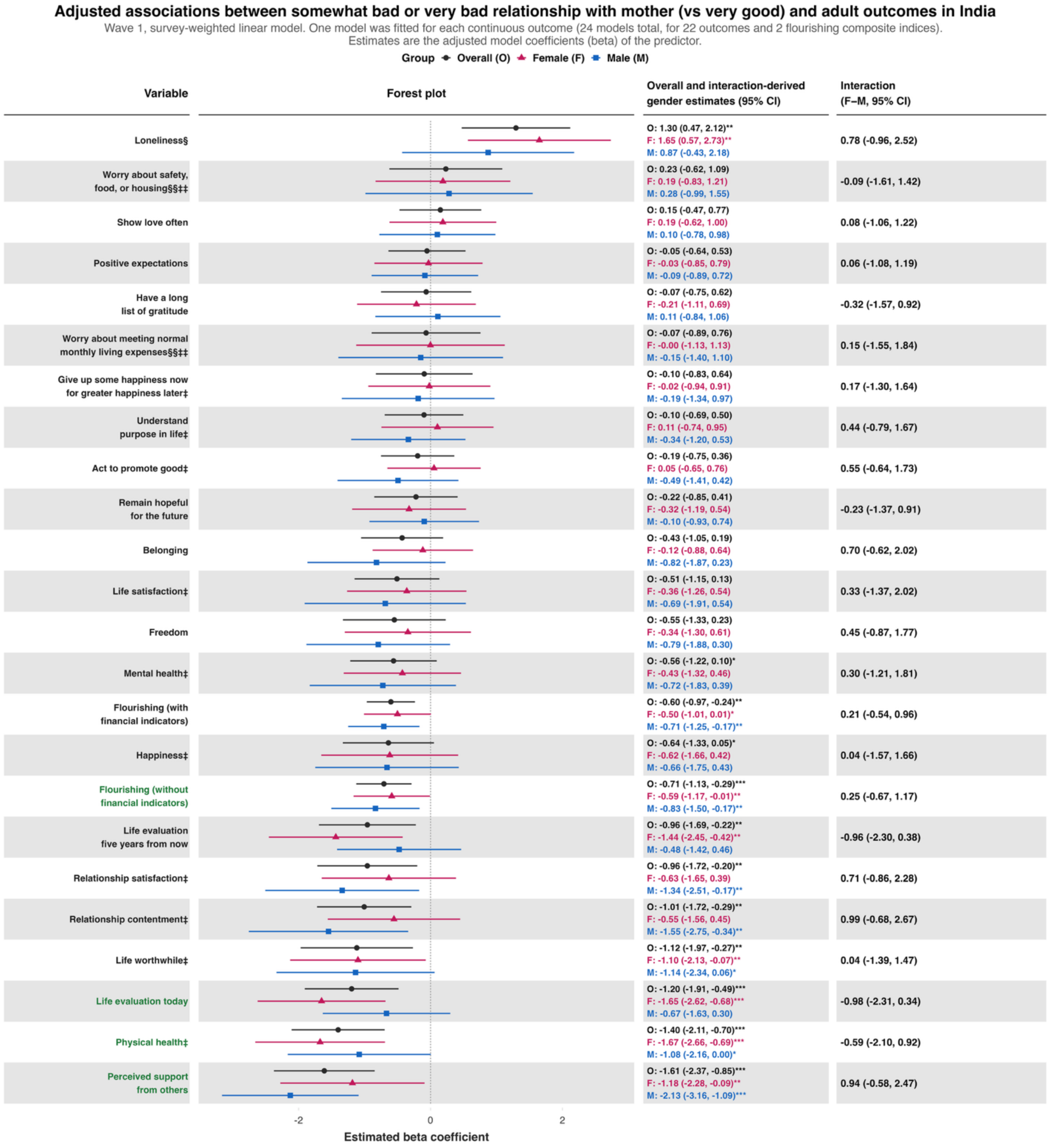

**Panel B**

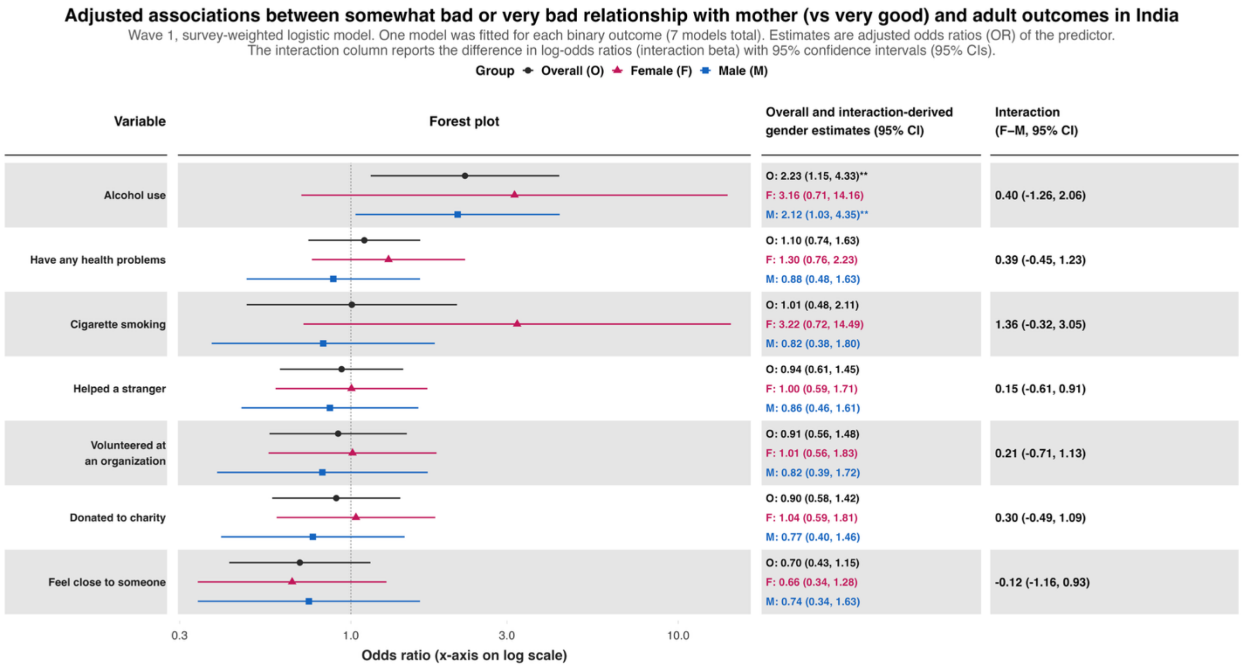
Adjusted associations between somewhat bad or very bad relationship with mother (vs very good) and adult outcomes in India
Wave 1, survey-weighted logistic model. One model was fitted for each binary outcome (7 models total). Estimates are adjusted odds ratios (OR) of the predictor.
The interaction column reports the difference in log-odds ratios (interaction beta) with 95% confidence intervals (95% CIs).
Group Overall (O) Female (F) Male (M)
Variable
Forest plot
Overall and interaction-derived gender estimates (95% CI)
Interaction (F–M, 95% CI)
Alcohol use
O: 2.23 (1.15, 4.33)**
F: 3.16 (0.71, 14.16)
M: 2.12 (1.03, 4.35)**
0.40 (-1.26, 2.06)
Have any health problems
O: 1.10 (0.74, 1.63)
F: 1.30 (0.76, 2.23)
M: 0.88 (0.48, 1.63)
0.39 (-0.45, 1.23)
Cigarette smoking
O: 1.01 (0.48, 2.11)
F: 3.22 (0.72, 14.49)
M: 0.82 (0.38, 1.80)
1.36 (-0.32, 3.05)
Helped a stranger
O: 0.94 (0.61, 1.45)
F: 1.00 (0.59, 1.71)
M: 0.86 (0.46, 1.61)
0.15 (-0.61, 0.91)
Volunteered at an organization
O: 0.91 (0.56, 1.48)
F: 1.01 (0.56, 1.83)
M: 0.82 (0.39, 1.72)
0.21 (-0.71, 1.13)
Donated to charity
O: 0.90 (0.58, 1.42)
F: 1.04 (0.59, 1.81)
M: 0.77 (0.40, 1.46)
0.30 (-0.49, 1.09)
Feel close to someone
O: 0.70 (0.43, 1.15)
F: 0.66 (0.34, 1.28)
M: 0.74 (0.34, 1.63)
-0.12 (-1.16, 0.93)
0.3
1.0
3.0
10.0
Odds ratio (x-axis on log scale)

**Panel C**

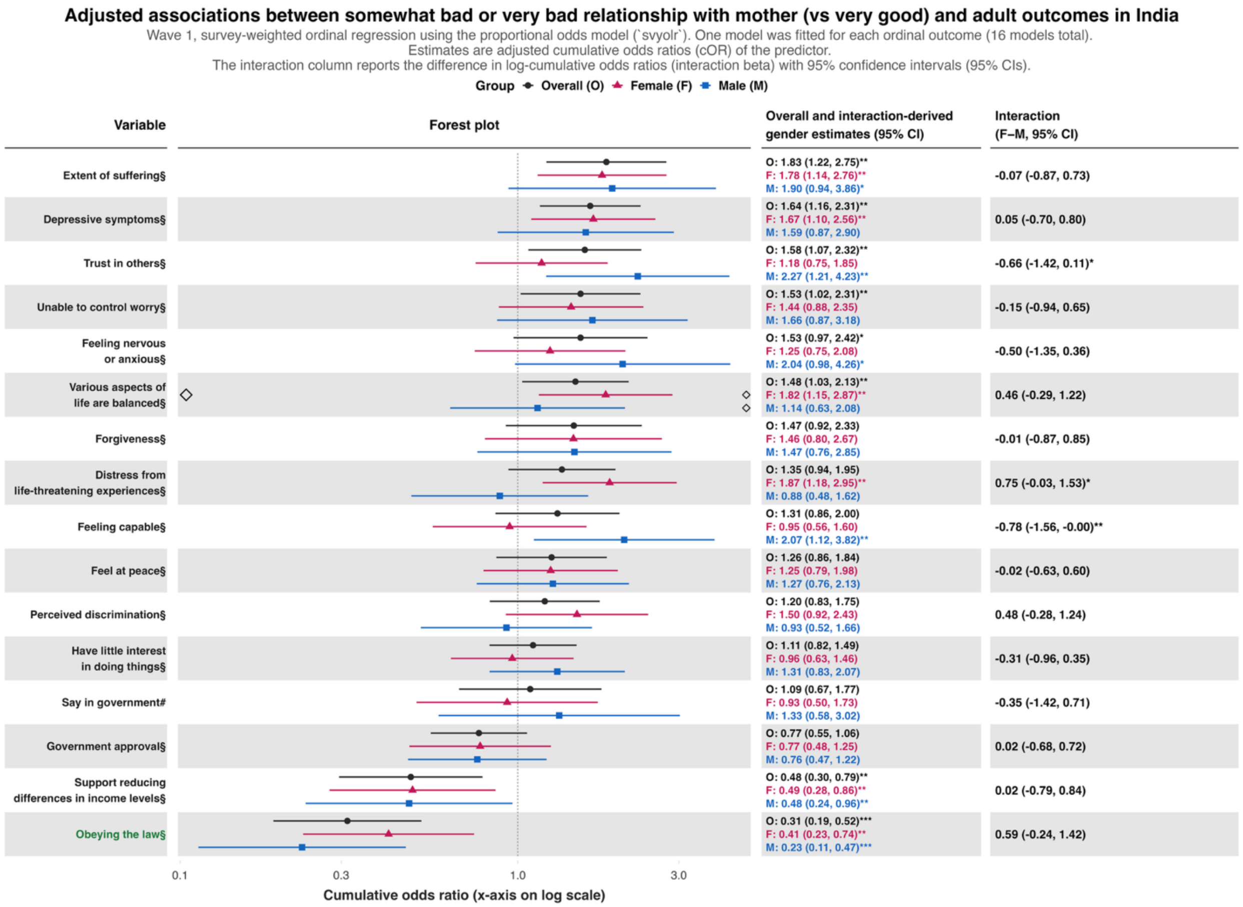
Adjusted associations between somewhat bad or very bad relationship with mother (vs very good) and adult outcomes in India
Wave 1, survey-weighted ordinal regression using the proportional odds model (`svyolr`). One model was fitted for each ordinal outcome (16 models total).
Estimates are adjusted cumulative odds ratios (cOR) of the predictor.
The interaction column reports the difference in log-cumulative odds ratios (interaction beta) with 95% confidence intervals (95% CIs).
Group
Overall (O)
Female (F)
Male (M)
Variable
Forest plot
Overall and interaction-derived gender estimates (95% CI)
Interaction (F–M, 95% CI)
Extent of suffering§
O: 1.83 (1.22, 2.75)**
F: 1.78 (1.14, 2.76)**
M: 1.90 (0.94, 3.86)*
-0.07 (-0.87, 0.73)
Depressive symptoms§
O: 1.64 (1.16, 2.31)**
F: 1.67 (1.10, 2.56)**
M: 1.59 (0.87, 2.90)
0.05 (-0.70, 0.80)
Trust in others§
O: 1.58 (1.07, 2.32)**
F: 1.18 (0.75, 1.85)
M: 2.27 (1.21, 4.23)**
-0.66 (-1.42, 0.11)*
Unable to control worry§
O: 1.53 (1.02, 2.31)**
F: 1.44 (0.88, 2.35)
M: 1.66 (0.87, 3.18)
-0.15 (-0.94, 0.65)
Feeling nervous or anxious§
O: 1.53 (0.97, 2.42)*
F: 1.25 (0.75, 2.08)
M: 2.04 (0.98, 4.26)*
-0.50 (-1.35, 0.36)
Various aspects of life are balanced§
O: 1.48 (1.03, 2.13)**
F: 1.82 (1.15, 2.87)**
M: 1.14 (0.63, 2.08)
0.46 (-0.29, 1.22)
Forgiveness§
O: 1.47 (0.92, 2.33)
F: 1.46 (0.80, 2.67)
M: 1.47 (0.76, 2.85)
-0.01 (-0.87, 0.85)
Distress from life-threatening experiences§
O: 1.35 (0.94, 1.95)
F: 1.87 (1.18, 2.95)**
M: 0.88 (0.48, 1.62)
0.75 (-0.03, 1.53)*
Feeling capable§
O: 1.31 (0.86, 2.00)
F: 0.95 (0.56, 1.60)
M: 2.07 (1.12, 3.82)**
-0.78 (-1.56, -0.00)**
Feel at peace§
O: 1.26 (0.86, 1.84)
F: 1.25 (0.79, 1.98)
M: 1.27 (0.76, 2.13)
-0.02 (-0.63, 0.60)
Perceived discrimination§
O: 1.20 (0.83, 1.75)
F: 1.50 (0.92, 2.43)
M: 0.93 (0.52, 1.66)
0.48 (-0.28, 1.24)
Have little interest in doing things§
O: 1.11 (0.82, 1.49)
F: 0.96 (0.63, 1.46)
M: 1.31 (0.83, 2.07)
-0.31 (-0.96, 0.35)
Say in government#
O: 1.09 (0.67, 1.77)
F: 0.93 (0.50, 1.73)
M: 1.33 (0.58, 3.02)
-0.35 (-1.42, 0.71)
Government approval§
O: 0.77 (0.55, 1.06)
F: 0.77 (0.48, 1.25)
M: 0.76 (0.47, 1.22)
0.02 (-0.68, 0.72)
Support reducing differences in income levels§
O: 0.48 (0.30, 0.79)**
F: 0.49 (0.28, 0.86)**
M: 0.48 (0.24, 0.96)**
0.02 (-0.79, 0.84)
Obeying the law§
O: 0.31 (0.19, 0.52)***
F: 0.41 (0.23, 0.74)**
M: 0.23 (0.11, 0.47)***
0.59 (-0.24, 1.42)
0.1
0.3
1.0
3.0
Cumulative odds ratio (x-axis on log scale)

**eFigure 7.** Adjusted associations between bad or very bad quality of relationship with father as predictor (very good quality as reference category) and 45 outcomes of interest and two flourishing composite indices.

Each panel shows the associations between the predictor and each of the continuous outcomes (**Panel A**), binary outcomes (**Panel B**), and ordinal outcomes (**Panel C**).

### Panel A

**Adjusted associations between somewhat bad or very bad relationship with father (vs very good) and adult outcomes in India**

Wave 1, survey-weighted linear model. One model was fitted for each continuous outcome (24 models total, for 22 outcomes and 2 flourishing composite indices).
Estimates are the adjusted model coefficients (beta) of the predictor.

Group: Overall (O), Female (F), Male (M)

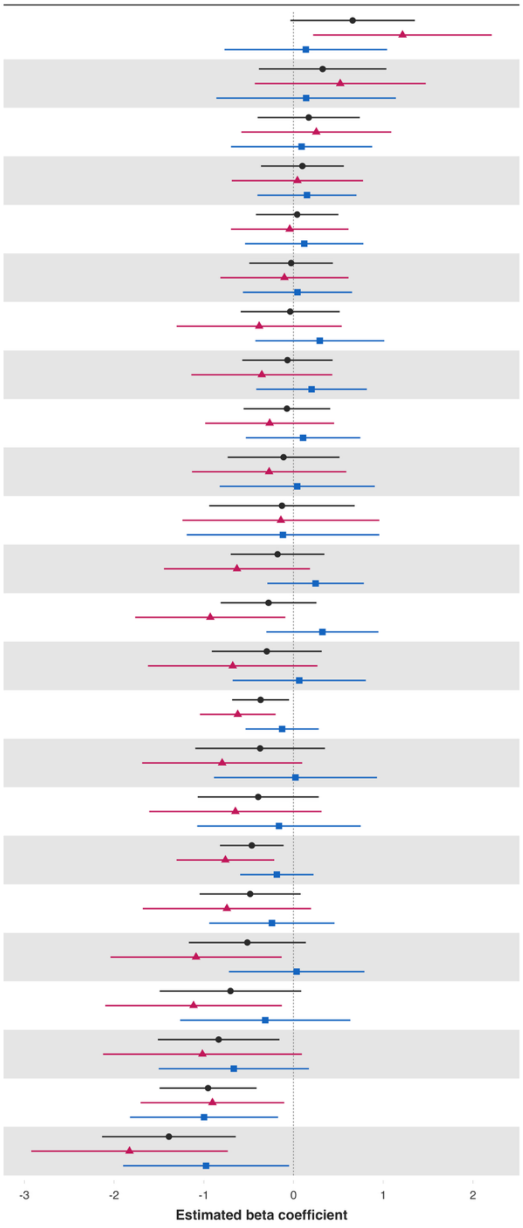


| Variable | Overall and interaction-derived gender estimates (95% CI) | Interaction (F–M, 95% CI) |
|---|---|---|
| Loneliness§ | O: 0.66 (-0.03, 1.35)*<br>F: 1.21 (0.22, 2.21)**<br>M: 0.14 (-0.77, 1.04) | 1.08 (-0.25, 2.41) |
| Worry about meeting normal monthly living expenses§§‡‡ | O: 0.32 (-0.39, 1.03)<br>F: 0.52 (-0.43, 1.47)<br>M: 0.14 (-0.86, 1.14) | 0.38 (-0.95, 1.71) |
| Give up some happiness now for greater happiness later‡ | O: 0.17 (-0.40, 0.74)<br>F: 0.25 (-0.58, 1.09)<br>M: 0.09 (-0.70, 0.88) | 0.16 (-0.99, 1.32) |
| Remain hopeful for the future | O: 0.10 (-0.36, 0.56)<br>F: 0.04 (-0.69, 0.77)<br>M: 0.15 (-0.40, 0.70) | -0.11 (-1.01, 0.79) |
| Show love often | O: 0.04 (-0.42, 0.50)<br>F: -0.04 (-0.70, 0.61)<br>M: 0.12 (-0.54, 0.78) | -0.16 (-1.11, 0.78) |
| Belonging | O: -0.03 (-0.49, 0.44)<br>F: -0.10 (-0.82, 0.61)<br>M: 0.04 (-0.56, 0.65) | -0.15 (-1.09, 0.79) |
| Life evaluation five years from now | O: -0.04 (-0.59, 0.52)<br>F: -0.38 (-1.30, 0.54)<br>M: 0.29 (-0.43, 1.01) | -0.68 (-1.90, 0.55) |
| Positive expectations | O: -0.07 (-0.57, 0.44)<br>F: -0.35 (-1.14, 0.43)<br>M: 0.20 (-0.42, 0.82) | -0.55 (-1.55, 0.44) |
| Act to promote good‡ | O: -0.07 (-0.56, 0.41)<br>F: -0.27 (-0.98, 0.45)<br>M: 0.11 (-0.53, 0.75) | -0.37 (-1.33, 0.58) |
| Have a long list of gratitude | O: -0.11 (-0.73, 0.51)<br>F: -0.27 (-1.13, 0.59)<br>M: 0.04 (-0.82, 0.91) | -0.31 (-1.50, 0.87) |
| Worry about safety, food, or housing§§‡‡ | O: -0.13 (-0.94, 0.68)<br>F: -0.14 (-1.24, 0.96)<br>M: -0.12 (-1.19, 0.96) | -0.02 (-1.47, 1.42) |
| Freedom | O: -0.18 (-0.70, 0.34)<br>F: -0.63 (-1.44, 0.18)<br>M: 0.25 (-0.29, 0.78) | -0.88 (-1.79, 0.04)* |
| Understand purpose in life‡ | O: -0.28 (-0.81, 0.26)<br>F: -0.93 (-1.77, -0.09)**<br>M: 0.32 (-0.30, 0.95) | -1.25 (-2.29, -0.21)** |
| Happiness‡ | O: -0.30 (-0.91, 0.31)<br>F: -0.68 (-1.62, 0.27)<br>M: 0.06 (-0.68, 0.81) | -0.74 (-1.92, 0.44) |
| Flourishing (with financial indicators) | O: -0.37 (-0.68, -0.05)**<br>F: -0.62 (-1.04, -0.20)**<br>M: -0.13 (-0.53, 0.28) | -0.49 (-1.04, 0.05)* |
| Life evaluation today | O: -0.37 (-1.09, 0.35)<br>F: -0.80 (-1.69, 0.10)*<br>M: 0.02 (-0.89, 0.93) | -0.82 (-1.91, 0.28) |
| Life worthwhile‡ | O: -0.39 (-1.07, 0.28)<br>F: -0.65 (-1.61, 0.31)<br>M: -0.16 (-1.07, 0.75) | -0.49 (-1.80, 0.83) |
| Flourishing (without financial indicators) | O: -0.47 (-0.82, -0.11)**<br>F: -0.76 (-1.30, -0.22)**<br>M: -0.19 (-0.60, 0.22) | -0.57 (-1.25, 0.10)* |
| Life satisfaction‡ | O: -0.48 (-1.05, 0.08)*<br>F: -0.74 (-1.68, 0.20)<br>M: -0.24 (-0.94, 0.46) | -0.50 (-1.70, 0.70) |
| Physical health‡ | O: -0.51 (-1.17, 0.14)<br>F: -1.09 (-2.04, -0.13)**<br>M: 0.04 (-0.72, 0.79) | -1.12 (-2.28, 0.04)* |
| Mental health‡ | O: -0.70 (-1.49, 0.09)*<br>F: -1.11 (-2.10, -0.13)**<br>M: -0.32 (-1.26, 0.63) | -0.80 (-1.93, 0.33) |
| Relationship satisfaction‡ | O: -0.83 (-1.51, -0.16)**<br>F: -1.02 (-2.12, 0.09)*<br>M: -0.67 (-1.50, 0.17) | -0.35 (-1.77, 1.07) |
| Relationship contentment‡ | O: -0.95 (-1.49, -0.41)***<br>F: -0.90 (-1.70, -0.10)**<br>M: -1.00 (-1.82, -0.17)** | 0.09 (-1.12, 1.31) |
| Perceived support from others | O: -1.39 (-2.13, -0.64)***<br>F: -1.83 (-2.92, -0.73)**<br>M: -0.97 (-1.90, -0.05)** | -0.85 (-2.23, 0.52) |

**Panel B**

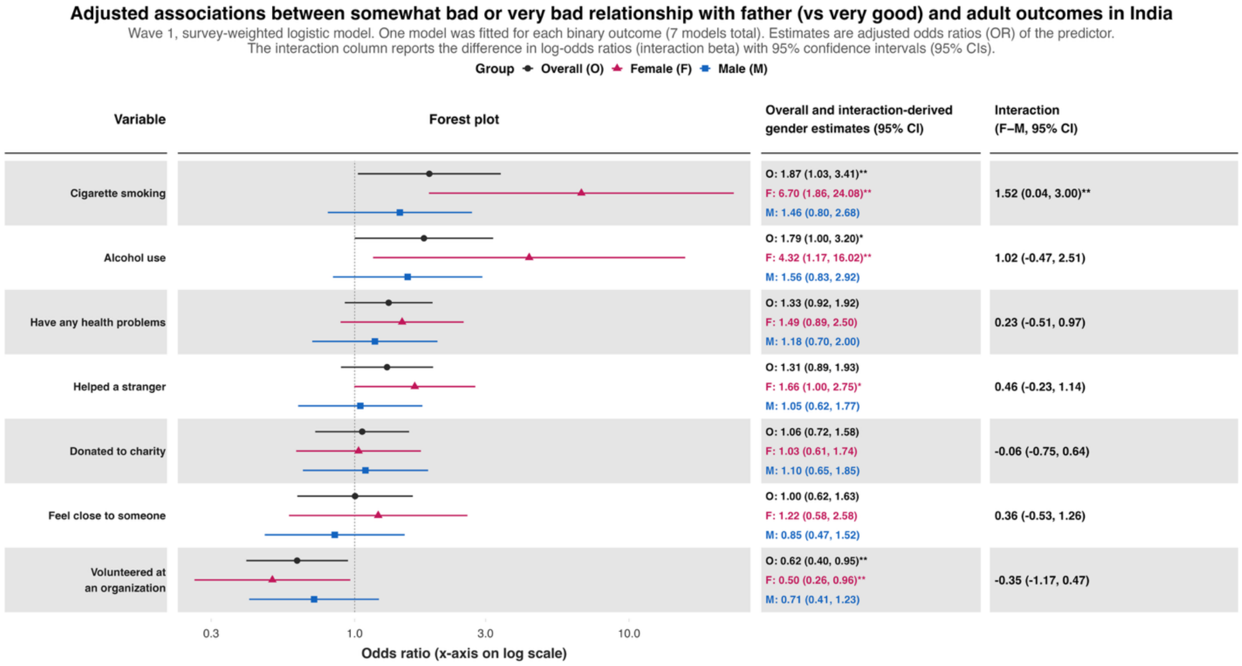
Adjusted associations between somewhat bad or very bad relationship with father (vs very good) and adult outcomes in India
Wave 1, survey-weighted logistic model. One model was fitted for each binary outcome (7 models total). Estimates are adjusted odds ratios (OR) of the predictor.
The interaction column reports the difference in log-odds ratios (interaction beta) with 95% confidence intervals (95% CIs).
Group Overall (O) Female (F) Male (M)
Variable
Forest plot
Overall and interaction-derived gender estimates (95% CI)
Interaction (F−M, 95% CI)
Cigarette smoking
O: 1.87 (1.03, 3.41)**
F: 6.70 (1.86, 24.08)**
M: 1.46 (0.80, 2.68)
1.52 (0.04, 3.00)**
Alcohol use
O: 1.79 (1.00, 3.20)*
F: 4.32 (1.17, 16.02)**
M: 1.56 (0.83, 2.92)
1.02 (-0.47, 2.51)
Have any health problems
O: 1.33 (0.92, 1.92)
F: 1.49 (0.89, 2.50)
M: 1.18 (0.70, 2.00)
0.23 (-0.51, 0.97)
Helped a stranger
O: 1.31 (0.89, 1.93)
F: 1.66 (1.00, 2.75)*
M: 1.05 (0.62, 1.77)
0.46 (-0.23, 1.14)
Donated to charity
O: 1.06 (0.72, 1.58)
F: 1.03 (0.61, 1.74)
M: 1.10 (0.65, 1.85)
-0.06 (-0.75, 0.64)
Feel close to someone
O: 1.00 (0.62, 1.63)
F: 1.22 (0.58, 2.58)
M: 0.85 (0.47, 1.52)
0.36 (-0.53, 1.26)
Volunteered at an organization
O: 0.62 (0.40, 0.95)**
F: 0.50 (0.26, 0.96)**
M: 0.71 (0.41, 1.23)
-0.35 (-1.17, 0.47)
0.3
1.0
3.0
10.0
Odds ratio (x-axis on log scale)

**Panel C**

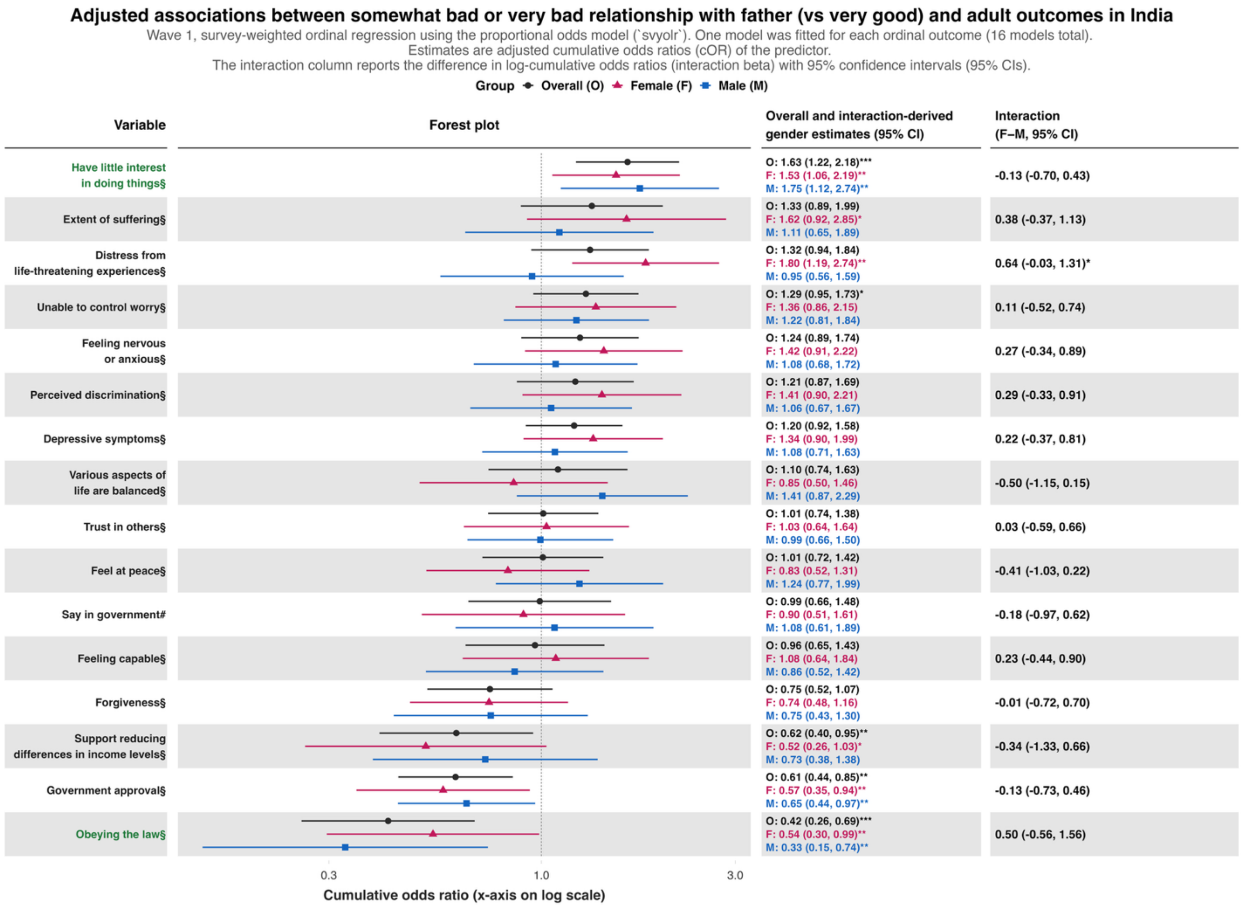
Adjusted associations between somewhat bad or very bad relationship with father (vs very good) and adult outcomes in India
Wave 1, survey-weighted ordinal regression using the proportional odds model (`svyolr`). One model was fitted for each ordinal outcome (16 models total).
Estimates are adjusted cumulative odds ratios (cOR) of the predictor.
The interaction column reports the difference in log-cumulative odds ratios (interaction beta) with 95% confidence intervals (95% CIs).
Group
Overall (O)
Female (F)
Male (M)
Variable
Forest plot
Overall and interaction-derived gender estimates (95% CI)
Interaction (F–M, 95% CI)
Have little interest in doing things§
O: 1.63 (1.22, 2.18)***
F: 1.53 (1.06, 2.19)**
M: 1.75 (1.12, 2.74)**
-0.13 (-0.70, 0.43)
Extent of suffering§
O: 1.33 (0.89, 1.99)
F: 1.62 (0.92, 2.85)*
M: 1.11 (0.65, 1.89)
0.38 (-0.37, 1.13)
Distress from life-threatening experiences§
O: 1.32 (0.94, 1.84)
F: 1.80 (1.19, 2.74)**
M: 0.95 (0.56, 1.59)
0.64 (-0.03, 1.31)*
Unable to control worry§
O: 1.29 (0.95, 1.73)*
F: 1.36 (0.86, 2.15)
M: 1.22 (0.81, 1.84)
0.11 (-0.52, 0.74)
Feeling nervous or anxious§
O: 1.24 (0.89, 1.74)
F: 1.42 (0.91, 2.22)
M: 1.08 (0.68, 1.72)
0.27 (-0.34, 0.89)
Perceived discrimination§
O: 1.21 (0.87, 1.69)
F: 1.41 (0.90, 2.21)
M: 1.06 (0.67, 1.67)
0.29 (-0.33, 0.91)
Depressive symptoms§
O: 1.20 (0.92, 1.58)
F: 1.34 (0.90, 1.99)
M: 1.08 (0.71, 1.63)
0.22 (-0.37, 0.81)
Various aspects of life are balanced§
O: 1.10 (0.74, 1.63)
F: 0.85 (0.50, 1.46)
M: 1.41 (0.87, 2.29)
-0.50 (-1.15, 0.15)
Trust in others§
O: 1.01 (0.74, 1.38)
F: 1.03 (0.64, 1.64)
M: 0.99 (0.66, 1.50)
0.03 (-0.59, 0.66)
Feel at peace§
O: 1.01 (0.72, 1.42)
F: 0.83 (0.52, 1.31)
M: 1.24 (0.77, 1.99)
-0.41 (-1.03, 0.22)
Say in government#
O: 0.99 (0.66, 1.48)
F: 0.90 (0.51, 1.61)
M: 1.08 (0.61, 1.89)
-0.18 (-0.97, 0.62)
Feeling capable§
O: 0.96 (0.65, 1.43)
F: 1.08 (0.64, 1.84)
M: 0.86 (0.52, 1.42)
0.23 (-0.44, 0.90)
Forgiveness§
O: 0.75 (0.52, 1.07)
F: 0.74 (0.48, 1.16)
M: 0.75 (0.43, 1.30)
-0.01 (-0.72, 0.70)
Support reducing differences in income levels§
O: 0.62 (0.40, 0.95)**
F: 0.52 (0.26, 1.03)*
M: 0.73 (0.38, 1.38)
-0.34 (-1.33, 0.66)
Government approval§
O: 0.61 (0.44, 0.85)**
F: 0.57 (0.35, 0.94)**
M: 0.65 (0.44, 0.97)**
-0.13 (-0.73, 0.46)
Obeying the law§
O: 0.42 (0.26, 0.69)***
F: 0.54 (0.30, 0.99)**
M: 0.33 (0.15, 0.74)**
0.50 (-0.56, 1.56)
0.3
1.0
3.0
Cumulative odds ratio (x-axis on log scale)

**eFigure 8.** Adjusted associations between poorly self-rated childhood health as predictor (excellent self-rated childhood health as reference category) and 45 outcomes of interest and two flourishing composite indices.

Each panel shows the associations between the predictor and each of the continuous outcomes (**Panel A**), binary outcomes (**Panel B**), and ordinal outcomes (**Panel C**).

**Panel A**

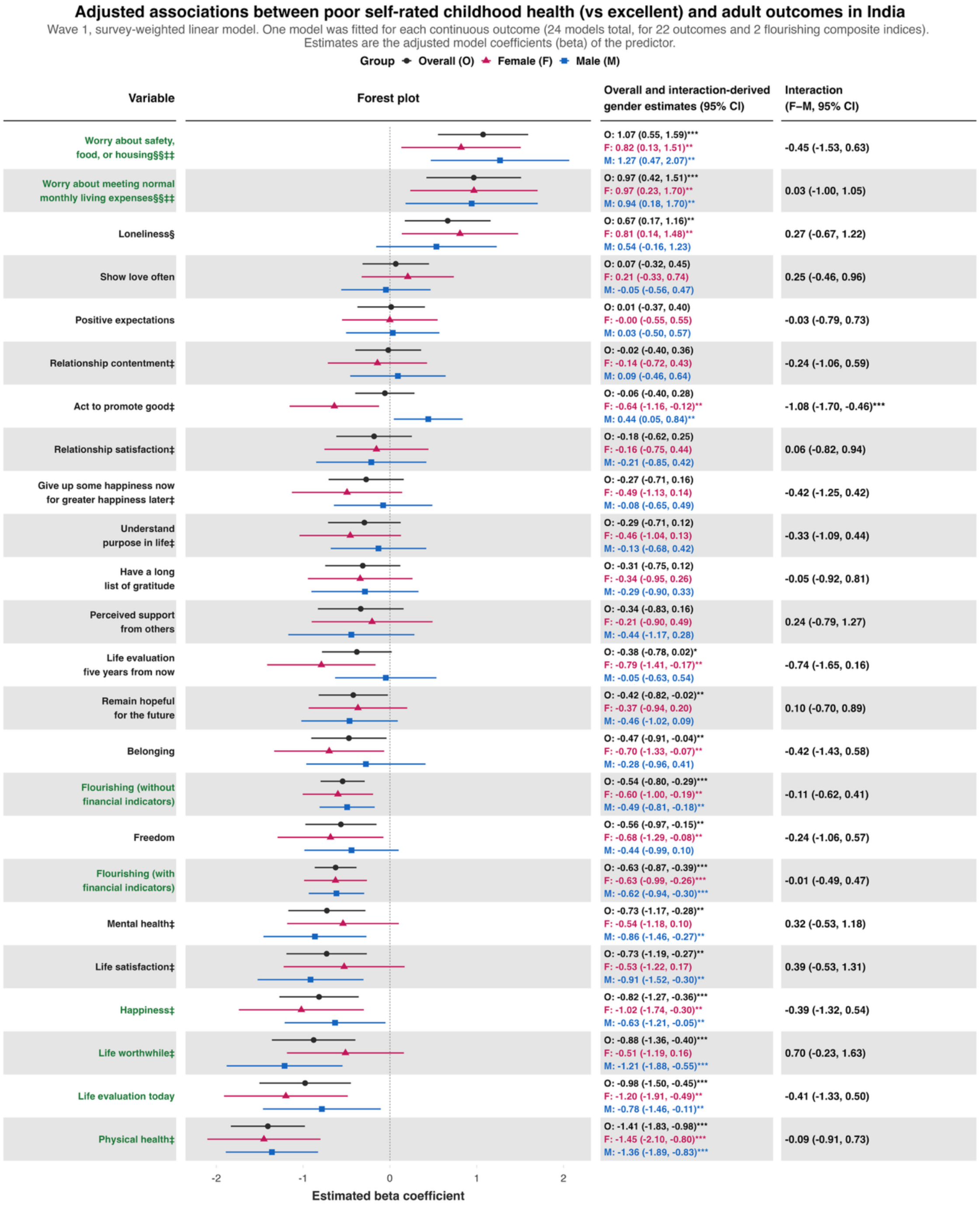

**Panel B**

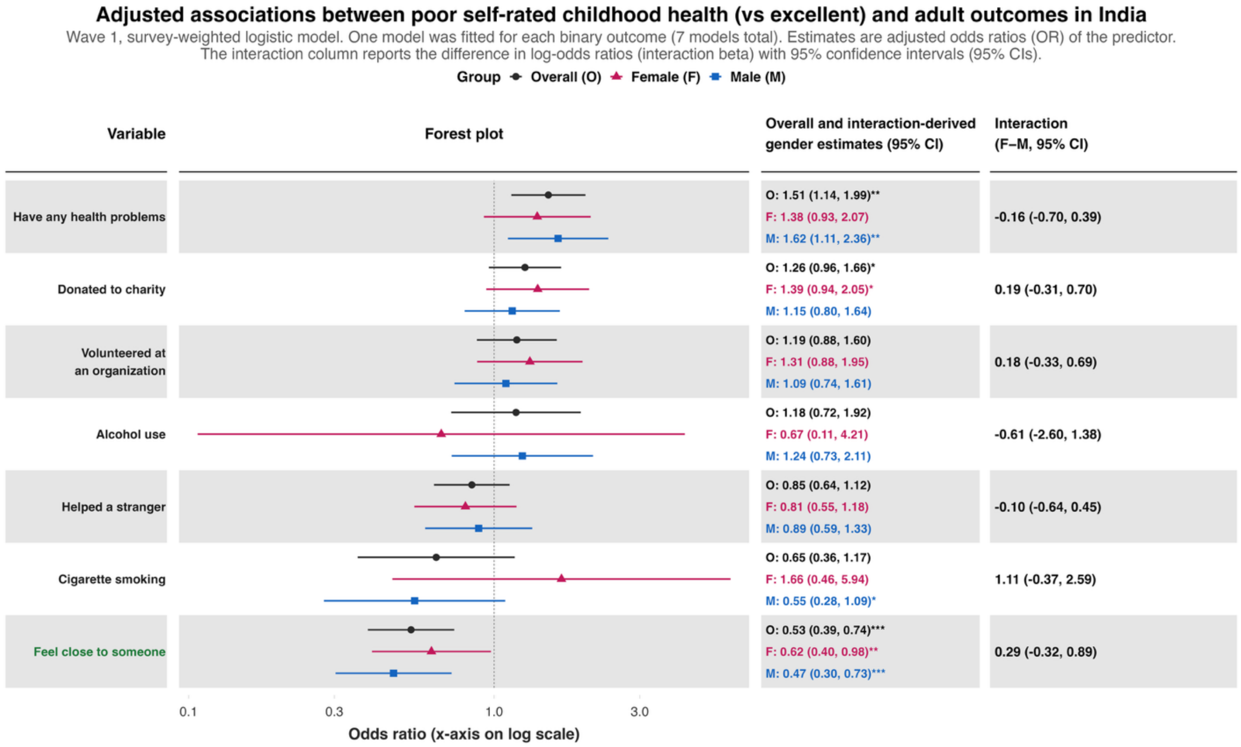

Adjusted associations between poor self-rated childhood health (vs excellent) and adult outcomes in India
Wave 1, survey-weighted logistic model. One model was fitted for each binary outcome (7 models total). Estimates are adjusted odds ratios (OR) of the predictor.
The interaction column reports the difference in log-odds ratios (interaction beta) with 95% confidence intervals (95% CIs).
Group Overall (O) Female (F) Male (M)
Variable
Forest plot
Overall and interaction-derived gender estimates (95% CI)
Interaction (F–M, 95% CI)
Have any health problems
O: 1.51 (1.14, 1.99)**
F: 1.38 (0.93, 2.07)
M: 1.62 (1.11, 2.36)**
-0.16 (-0.70, 0.39)
Donated to charity
O: 1.26 (0.96, 1.66)*
F: 1.39 (0.94, 2.05)*
M: 1.15 (0.80, 1.64)
0.19 (-0.31, 0.70)
Volunteered at an organization
O: 1.19 (0.88, 1.60)
F: 1.31 (0.88, 1.95)
M: 1.09 (0.74, 1.61)
0.18 (-0.33, 0.69)
Alcohol use
O: 1.18 (0.72, 1.92)
F: 0.67 (0.11, 4.21)
M: 1.24 (0.73, 2.11)
-0.61 (-2.60, 1.38)
Helped a stranger
O: 0.85 (0.64, 1.12)
F: 0.81 (0.55, 1.18)
M: 0.89 (0.59, 1.33)
-0.10 (-0.64, 0.45)
Cigarette smoking
O: 0.65 (0.36, 1.17)
F: 1.66 (0.46, 5.94)
M: 0.55 (0.28, 1.09)*
1.11 (-0.37, 2.59)
Feel close to someone
O: 0.53 (0.39, 0.74)***
F: 0.62 (0.40, 0.98)**
M: 0.47 (0.30, 0.73)***
0.29 (-0.32, 0.89)
0.1
0.3
1.0
3.0
Odds ratio (x-axis on log scale)

**Panel C**

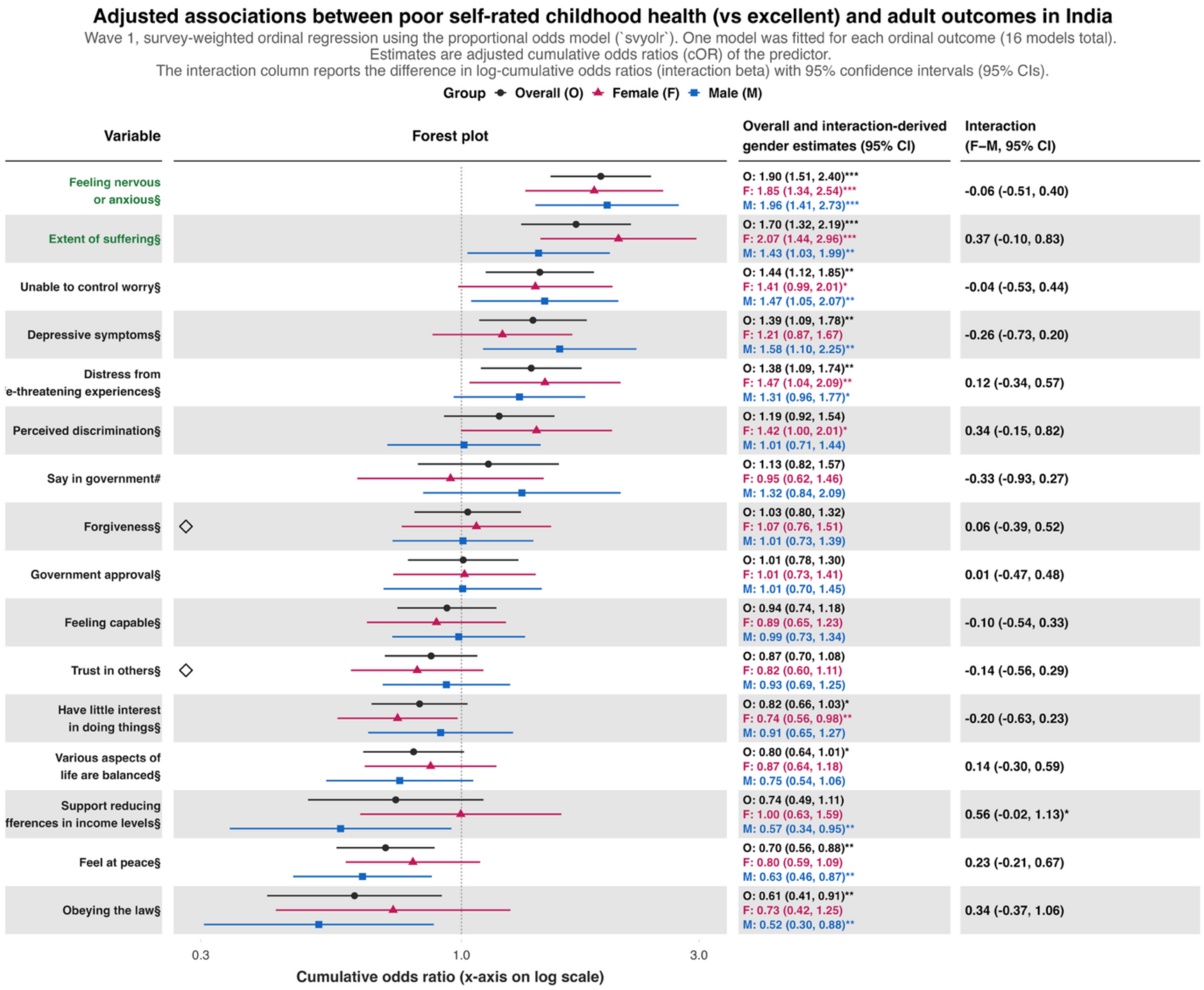

Adjusted associations between poor self-rated childhood health (vs excellent) and adult outcomes in India
Wave 1, survey-weighted ordinal regression using the proportional odds model (`svyolr`). One model was fitted for each ordinal outcome (16 models total).
Estimates are adjusted cumulative odds ratios (cOR) of the predictor.
The interaction column reports the difference in log-cumulative odds ratios (interaction beta) with 95% confidence intervals (95% CIs).
Group Overall (O) Female (F) Male (M)
Variable
Forest plot
Overall and interaction-derived gender estimates (95% CI)
Interaction (F–M, 95% CI)
Feeling nervous or anxious§
O: 1.90 (1.51, 2.40)***
F: 1.85 (1.34, 2.54)***
M: 1.96 (1.41, 2.73)***
-0.06 (-0.51, 0.40)
Extent of suffering§
O: 1.70 (1.32, 2.19)***
F: 2.07 (1.44, 2.96)***
M: 1.43 (1.03, 1.99)**
0.37 (-0.10, 0.83)
Unable to control worry§
O: 1.44 (1.12, 1.85)**
F: 1.41 (0.99, 2.01)*
M: 1.47 (1.05, 2.07)**
-0.04 (-0.53, 0.44)
Depressive symptoms§
O: 1.39 (1.09, 1.78)**
F: 1.21 (0.87, 1.67)
M: 1.58 (1.10, 2.25)**
-0.26 (-0.73, 0.20)
Distress from
e-threatening experiences§
O: 1.38 (1.09, 1.74)**
F: 1.47 (1.04, 2.09)**
M: 1.31 (0.96, 1.77)*
0.12 (-0.34, 0.57)
Perceived discrimination§
O: 1.19 (0.92, 1.54)
F: 1.42 (1.00, 2.01)*
M: 1.01 (0.71, 1.44)
0.34 (-0.15, 0.82)
Say in government#
O: 1.13 (0.82, 1.57)
F: 0.95 (0.62, 1.46)
M: 1.32 (0.84, 2.09)
-0.33 (-0.93, 0.27)
Forgiveness§
O: 1.03 (0.80, 1.32)
F: 1.07 (0.76, 1.51)
M: 1.01 (0.73, 1.39)
0.06 (-0.39, 0.52)
Government approval§
O: 1.01 (0.78, 1.30)
F: 1.01 (0.73, 1.41)
M: 1.01 (0.70, 1.45)
0.01 (-0.47, 0.48)
Feeling capable§
O: 0.94 (0.74, 1.18)
F: 0.89 (0.65, 1.23)
M: 0.99 (0.73, 1.34)
-0.10 (-0.54, 0.33)
Trust in others§
O: 0.87 (0.70, 1.08)
F: 0.82 (0.60, 1.11)
M: 0.93 (0.69, 1.25)
-0.14 (-0.56, 0.29)
Have little interest in doing things§
O: 0.82 (0.66, 1.03)*
F: 0.74 (0.56, 0.98)**
M: 0.91 (0.65, 1.27)
-0.20 (-0.63, 0.23)
Various aspects of life are balanced§
O: 0.80 (0.64, 1.01)*
F: 0.87 (0.64, 1.18)
M: 0.75 (0.54, 1.06)
0.14 (-0.30, 0.59)
Support reducing
fferences in income levels§
O: 0.74 (0.49, 1.11)
F: 1.00 (0.63, 1.59)
M: 0.57 (0.34, 0.95)**
0.56 (-0.02, 1.13)*
Feel at peace§
O: 0.70 (0.56, 0.88)**
F: 0.80 (0.59, 1.09)
M: 0.63 (0.46, 0.87)**
0.23 (-0.21, 0.67)
Obeying the law§
O: 0.61 (0.41, 0.91)**
F: 0.73 (0.42, 1.25)
M: 0.52 (0.30, 0.88)**
0.34 (-0.37, 1.06)
0.3
1.0
3.0
Cumulative odds ratio (x-axis on log scale)

**eFigure 9.** Adjusted associations between finding it very difficult in perceived family financial situation as predictor (comfortable perceived family financial situation reference category) and 45 outcomes of interest and two flourishing composite indices.

Each panel shows the associations between the predictor and each of the continuous outcomes (**Panel A**), binary outcomes (**Panel B**), and ordinal outcomes (**Panel C**).

**Panel A**

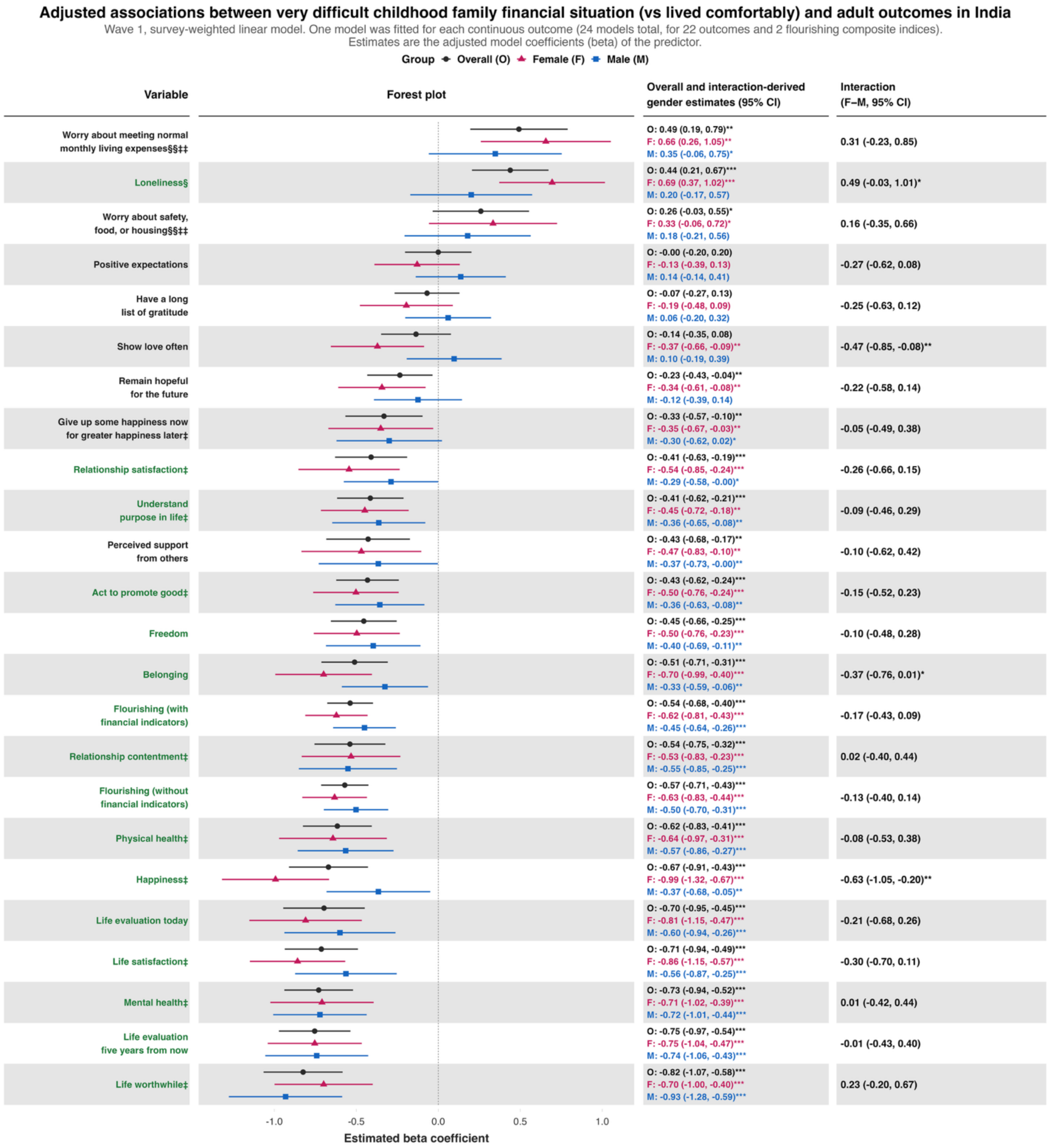

**Panel B**

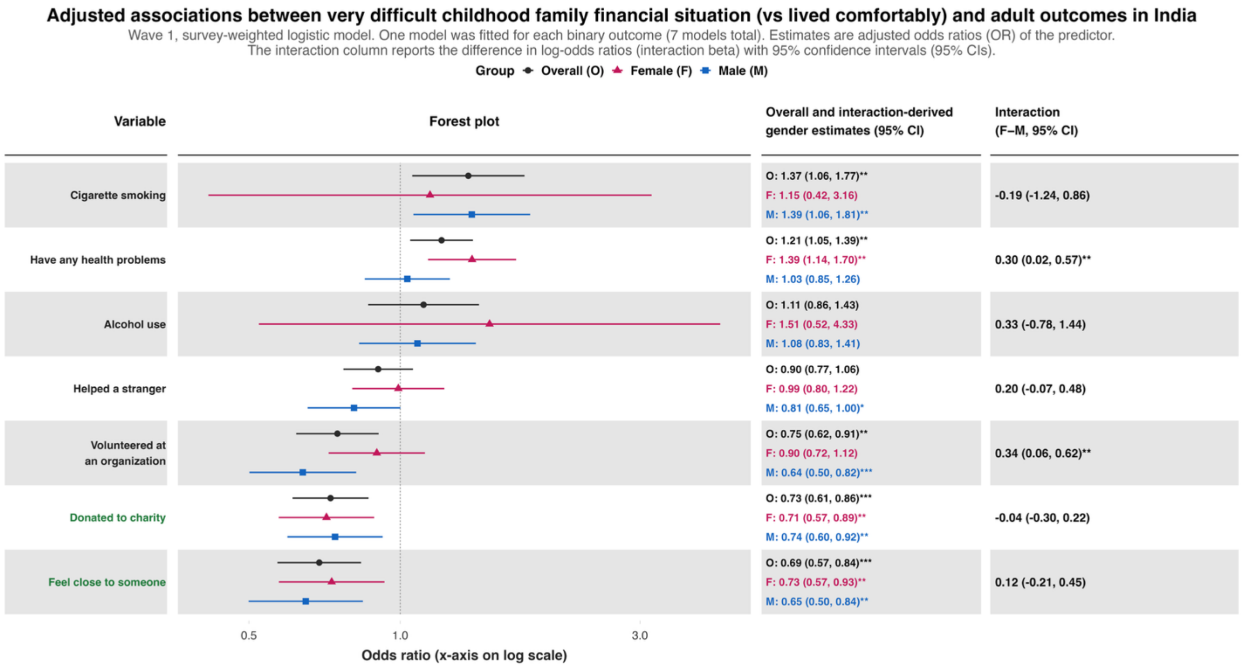
Adjusted associations between very difficult childhood family financial situation (vs lived comfortably) and adult outcomes in India
Wave 1, survey-weighted logistic model. One model was fitted for each binary outcome (7 models total). Estimates are adjusted odds ratios (OR) of the predictor.
The interaction column reports the difference in log-odds ratios (interaction beta) with 95% confidence intervals (95% CIs).
Group Overall (O) Female (F) Male (M)
Variable
Forest plot
Overall and interaction-derived gender estimates (95% CI)
Interaction (F–M, 95% CI)
Cigarette smoking
O: 1.37 (1.06, 1.77)**
F: 1.15 (0.42, 3.16)
M: 1.39 (1.06, 1.81)**
-0.19 (-1.24, 0.86)
Have any health problems
O: 1.21 (1.05, 1.39)**
F: 1.39 (1.14, 1.70)**
M: 1.03 (0.85, 1.26)
0.30 (0.02, 0.57)**
Alcohol use
O: 1.11 (0.86, 1.43)
F: 1.51 (0.52, 4.33)
M: 1.08 (0.83, 1.41)
0.33 (-0.78, 1.44)
Helped a stranger
O: 0.90 (0.77, 1.06)
F: 0.99 (0.80, 1.22)
M: 0.81 (0.65, 1.00)*
0.20 (-0.07, 0.48)
Volunteered at an organization
O: 0.75 (0.62, 0.91)**
F: 0.90 (0.72, 1.12)
M: 0.64 (0.50, 0.82)***
0.34 (0.06, 0.62)**
Donated to charity
O: 0.73 (0.61, 0.86)***
F: 0.71 (0.57, 0.89)**
M: 0.74 (0.60, 0.92)**
-0.04 (-0.30, 0.22)
Feel close to someone
O: 0.69 (0.57, 0.84)***
F: 0.73 (0.57, 0.93)**
M: 0.65 (0.50, 0.84)**
0.12 (-0.21, 0.45)
0.5
1.0
3.0
Odds ratio (x-axis on log scale)

**Panel C**

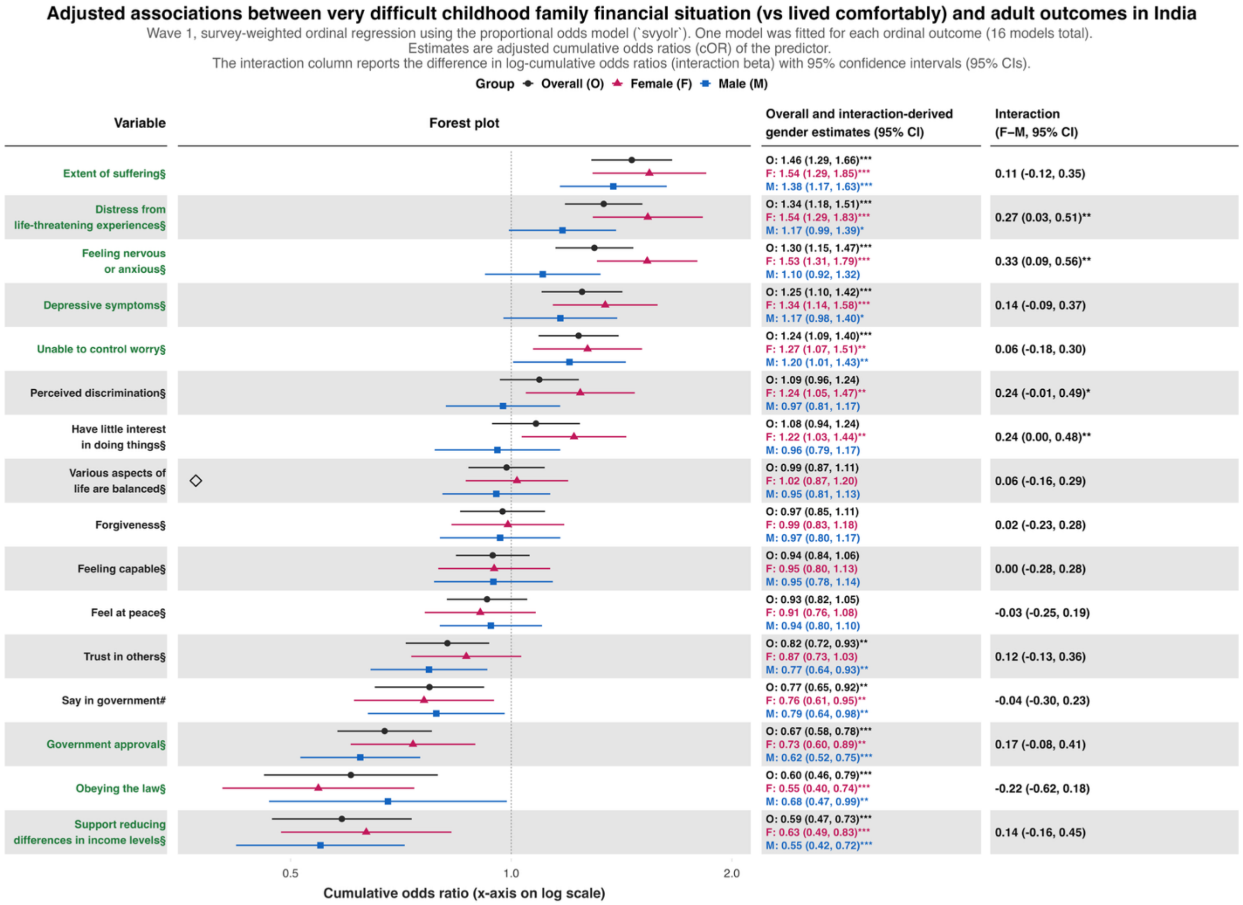

Adjusted associations between very difficult childhood family financial situation (vs lived comfortably) and adult outcomes in India
Wave 1, survey-weighted ordinal regression using the proportional odds model (`svyolr`). One model was fitted for each ordinal outcome (16 models total).
Estimates are adjusted cumulative odds ratios (cOR) of the predictor.
The interaction column reports the difference in log-cumulative odds ratios (interaction beta) with 95% confidence intervals (95% CIs).
Group Overall (O) Female (F) Male (M)
Variable
Forest plot
Overall and interaction-derived gender estimates (95% CI)
Interaction (F–M, 95% CI)
Extent of suffering§
O: 1.46 (1.29, 1.66)*** F: 1.54 (1.29, 1.85)*** M: 1.38 (1.17, 1.63)***
0.11 (-0.12, 0.35)
Distress from life-threatening experiences§
O: 1.34 (1.18, 1.51)*** F: 1.54 (1.29, 1.83)*** M: 1.17 (0.99, 1.39)*
0.27 (0.03, 0.51)**
Feeling nervous or anxious§
O: 1.30 (1.15, 1.47)*** F: 1.53 (1.31, 1.79)*** M: 1.10 (0.92, 1.32)
0.33 (0.09, 0.56)**
Depressive symptoms§
O: 1.25 (1.10, 1.42)*** F: 1.34 (1.14, 1.58)*** M: 1.17 (0.98, 1.40)*
0.14 (-0.09, 0.37)
Unable to control worry§
O: 1.24 (1.09, 1.40)*** F: 1.27 (1.07, 1.51)** M: 1.20 (1.01, 1.43)**
0.06 (-0.18, 0.30)
Perceived discrimination§
O: 1.09 (0.96, 1.24) F: 1.24 (1.05, 1.47)** M: 0.97 (0.81, 1.17)
0.24 (-0.01, 0.49)*
Have little interest in doing things§
O: 1.08 (0.94, 1.24) F: 1.22 (1.03, 1.44)** M: 0.96 (0.79, 1.17)
0.24 (0.00, 0.48)**
Various aspects of life are balanced§
O: 0.99 (0.87, 1.11) F: 1.02 (0.87, 1.20) M: 0.95 (0.81, 1.13)
0.06 (-0.16, 0.29)
Forgiveness§
O: 0.97 (0.85, 1.11) F: 0.99 (0.83, 1.18) M: 0.97 (0.80, 1.17)
0.02 (-0.23, 0.28)
Feeling capable§
O: 0.94 (0.84, 1.06) F: 0.95 (0.80, 1.13) M: 0.95 (0.78, 1.14)
0.00 (-0.28, 0.28)
Feel at peace§
O: 0.93 (0.82, 1.05) F: 0.91 (0.76, 1.08) M: 0.94 (0.80, 1.10)
-0.03 (-0.25, 0.19)
Trust in others§
O: 0.82 (0.72, 0.93)** F: 0.87 (0.73, 1.03) M: 0.77 (0.64, 0.93)**
0.12 (-0.13, 0.36)
Say in government#
O: 0.77 (0.65, 0.92)** F: 0.76 (0.61, 0.95)** M: 0.79 (0.64, 0.98)**
-0.04 (-0.30, 0.23)
Government approval§
O: 0.67 (0.58, 0.78)*** F: 0.73 (0.60, 0.89)** M: 0.62 (0.52, 0.75)***
0.17 (-0.08, 0.41)
Obeying the law§
O: 0.60 (0.46, 0.79)*** F: 0.55 (0.40, 0.74)*** M: 0.68 (0.47, 0.99)**
-0.22 (-0.62, 0.18)
Support reducing differences in income levels§
O: 0.59 (0.47, 0.73)*** F: 0.63 (0.49, 0.83)*** M: 0.55 (0.42, 0.72)***
0.14 (-0.16, 0.45)
0.5
1.0
2.0
Cumulative odds ratio (x-axis on log scale)

**eFigure 10.** Adjusted associations between childhood abuse and 14 outcomes from India in the mid-year survey (February-May 2024).

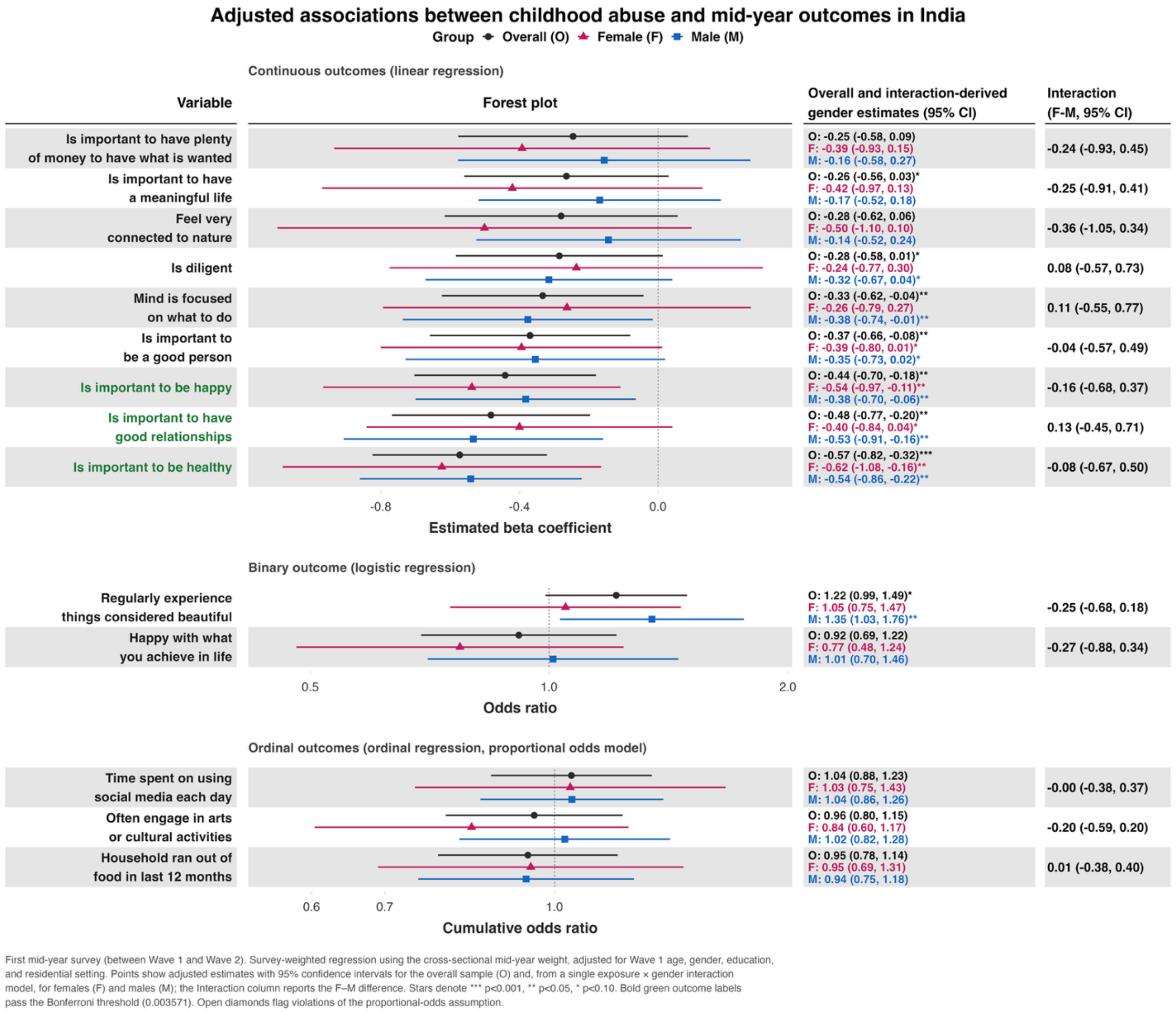

**eFigure 11.** Adjusted associations between childhood abuse and 45 outcomes of interest and two flourishing composite indices, where “don’t know” and “refused” responses are counted towards the exposed group.

Each panel shows the associations between the predictor and each of the continuous outcomes (**Panel A**), binary outcomes (**Panel B**), and ordinal outcomes (**Panel C**).

**Panel A**

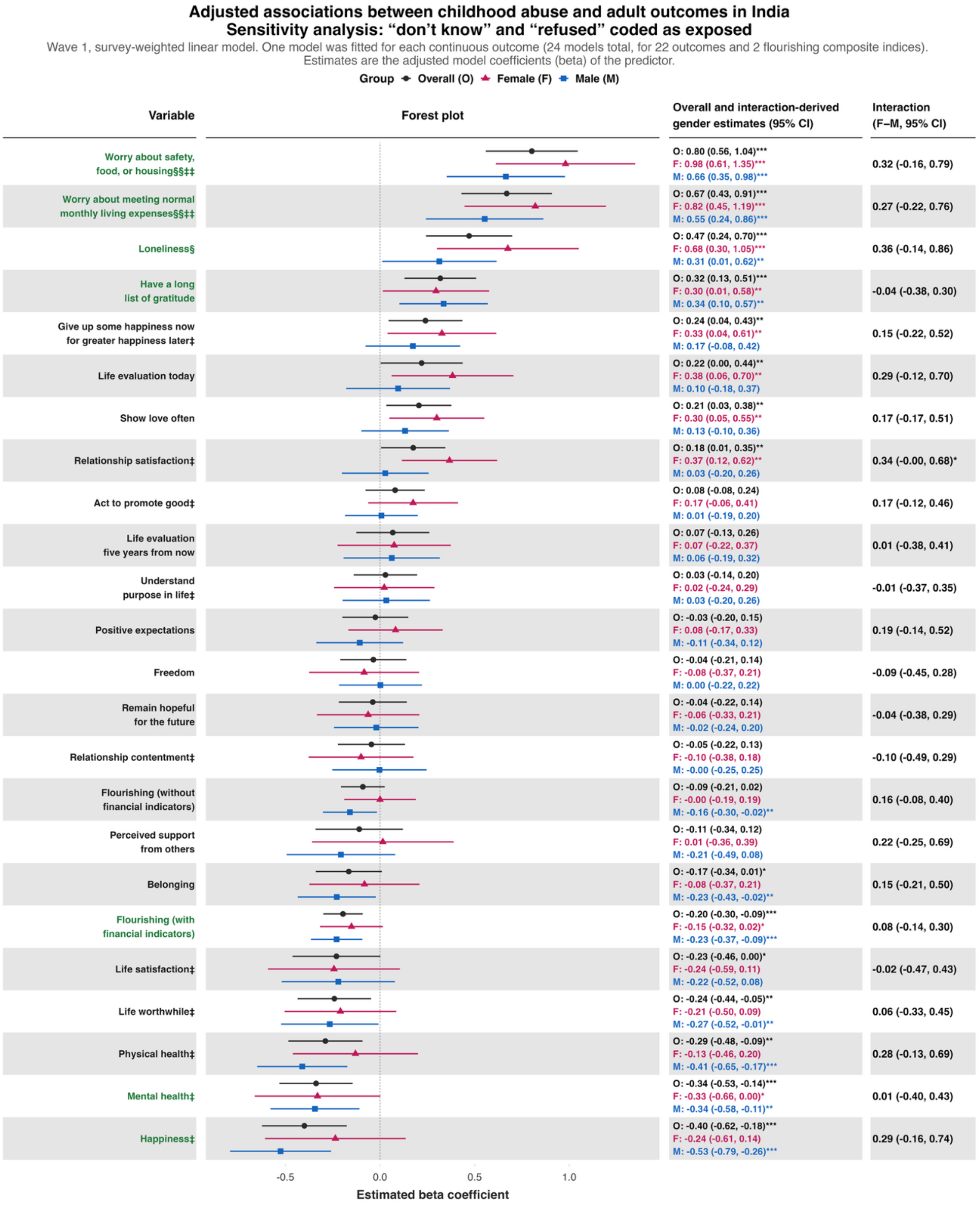

**Panel B**

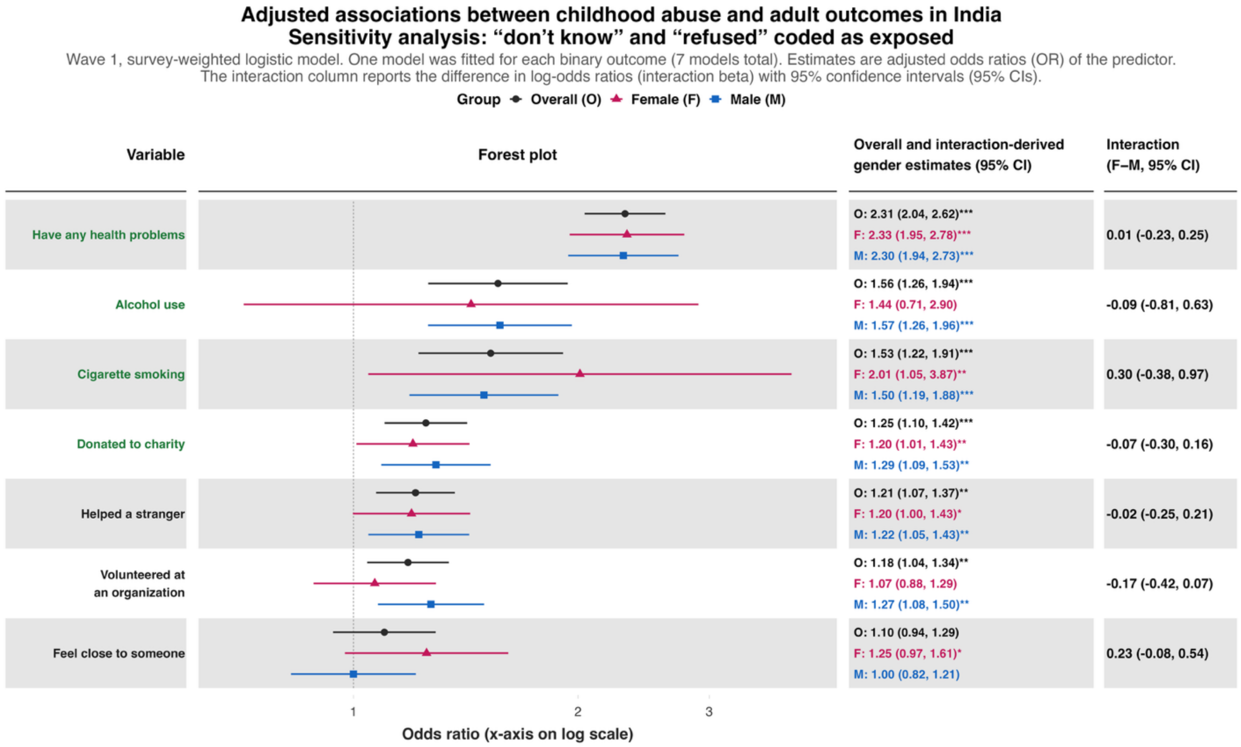
Adjusted associations between childhood abuse and adult outcomes in India
Sensitivity analysis: “don’t know” and “refused” coded as exposed
Wave 1, survey-weighted logistic model. One model was fitted for each binary outcome (7 models total). Estimates are adjusted odds ratios (OR) of the predictor.
The interaction column reports the difference in log-odds ratios (interaction beta) with 95% confidence intervals (95% CIs).
Group Overall (O) Female (F) Male (M)
Variable
Forest plot
Overall and interaction-derived gender estimates (95% CI)
Interaction (F–M, 95% CI)
Have any health problems
O: 2.31 (2.04, 2.62)***
F: 2.33 (1.95, 2.78)***
M: 2.30 (1.94, 2.73)***
0.01 (-0.23, 0.25)
Alcohol use
O: 1.56 (1.26, 1.94)***
F: 1.44 (0.71, 2.90)
M: 1.57 (1.26, 1.96)***
-0.09 (-0.81, 0.63)
Cigarette smoking
O: 1.53 (1.22, 1.91)***
F: 2.01 (1.05, 3.87)**
M: 1.50 (1.19, 1.88)***
0.30 (-0.38, 0.97)
Donated to charity
O: 1.25 (1.10, 1.42)***
F: 1.20 (1.01, 1.43)**
M: 1.29 (1.09, 1.53)**
-0.07 (-0.30, 0.16)
Helped a stranger
O: 1.21 (1.07, 1.37)**
F: 1.20 (1.00, 1.43)*
M: 1.22 (1.05, 1.43)**
-0.02 (-0.25, 0.21)
Volunteered at an organization
O: 1.18 (1.04, 1.34)**
F: 1.07 (0.88, 1.29)
M: 1.27 (1.08, 1.50)**
-0.17 (-0.42, 0.07)
Feel close to someone
O: 1.10 (0.94, 1.29)
F: 1.25 (0.97, 1.61)*
M: 1.00 (0.82, 1.21)
0.23 (-0.08, 0.54)
1
2
3
Odds ratio (x-axis on log scale)

**Panel C**

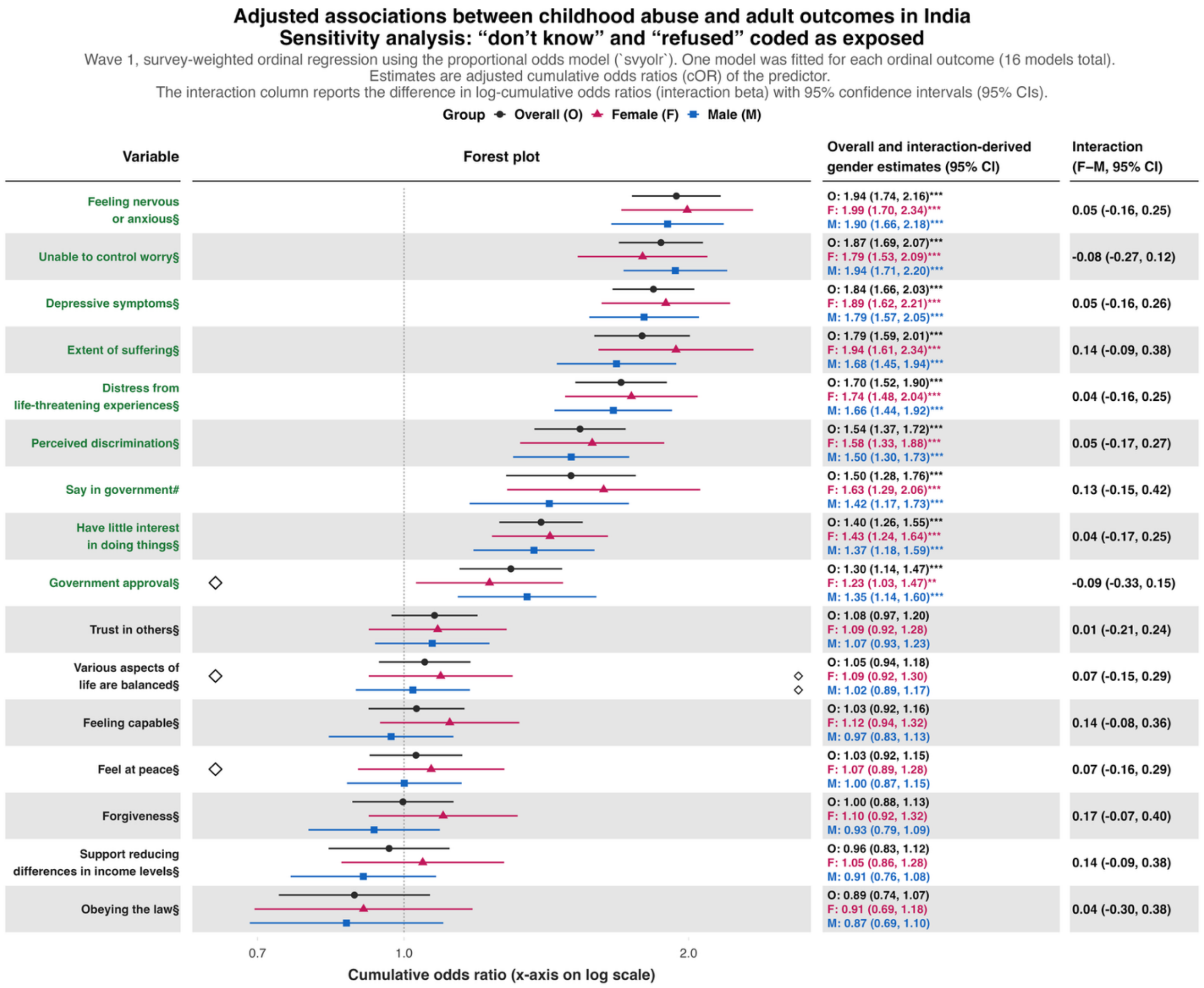
Adjusted associations between childhood abuse and adult outcomes in India
Sensitivity analysis: “don’t know” and “refused” coded as exposed
Wave 1, survey-weighted ordinal regression using the proportional odds model (`svyolr`). One model was fitted for each ordinal outcome (16 models total).
Estimates are adjusted cumulative odds ratios (cOR) of the predictor.
The interaction column reports the difference in log-cumulative odds ratios (interaction beta) with 95% confidence intervals (95% CIs).
Group Overall (O) Female (F) Male (M)
Variable
Forest plot
Overall and interaction-derived gender estimates (95% CI)
Interaction (F–M, 95% CI)
Feeling nervous or anxious§
O: 1.94 (1.74, 2.16)***
F: 1.99 (1.70, 2.34)***
M: 1.90 (1.66, 2.18)***
0.05 (-0.16, 0.25)
Unable to control worry§
O: 1.87 (1.69, 2.07)***
F: 1.79 (1.53, 2.09)***
M: 1.94 (1.71, 2.20)***
-0.08 (-0.27, 0.12)
Depressive symptoms§
O: 1.84 (1.66, 2.03)***
F: 1.89 (1.62, 2.21)***
M: 1.79 (1.57, 2.05)***
0.05 (-0.16, 0.26)
Extent of suffering§
O: 1.79 (1.59, 2.01)***
F: 1.94 (1.61, 2.34)***
M: 1.68 (1.45, 1.94)***
0.14 (-0.09, 0.38)
Distress from life-threatening experiences§
O: 1.70 (1.52, 1.90)***
F: 1.74 (1.48, 2.04)***
M: 1.66 (1.44, 1.92)***
0.04 (-0.16, 0.25)
Perceived discrimination§
O: 1.54 (1.37, 1.72)***
F: 1.58 (1.33, 1.88)***
M: 1.50 (1.30, 1.73)***
0.05 (-0.17, 0.27)
Say in government#
O: 1.50 (1.28, 1.76)***
F: 1.63 (1.29, 2.06)***
M: 1.42 (1.17, 1.73)***
0.13 (-0.15, 0.42)
Have little interest in doing things§
O: 1.40 (1.26, 1.55)***
F: 1.43 (1.24, 1.64)***
M: 1.37 (1.18, 1.59)***
0.04 (-0.17, 0.25)
Government approval§
O: 1.30 (1.14, 1.47)***
F: 1.23 (1.03, 1.47)**
M: 1.35 (1.14, 1.60)***
-0.09 (-0.33, 0.15)
Trust in others§
O: 1.08 (0.97, 1.20)
F: 1.09 (0.92, 1.28)
M: 1.07 (0.93, 1.23)
0.01 (-0.21, 0.24)
Various aspects of life are balanced§
O: 1.05 (0.94, 1.18)
F: 1.09 (0.92, 1.30)
M: 1.02 (0.89, 1.17)
0.07 (-0.15, 0.29)
Feeling capable§
O: 1.03 (0.92, 1.16)
F: 1.12 (0.94, 1.32)
M: 0.97 (0.83, 1.13)
0.14 (-0.08, 0.36)
Feel at peace§
O: 1.03 (0.92, 1.15)
F: 1.07 (0.89, 1.28)
M: 1.00 (0.87, 1.15)
0.07 (-0.16, 0.29)
Forgiveness§
O: 1.00 (0.88, 1.13)
F: 1.10 (0.92, 1.32)
M: 0.93 (0.79, 1.09)
0.17 (-0.07, 0.40)
Support reducing differences in income levels§
O: 0.96 (0.83, 1.12)
F: 1.05 (0.86, 1.28)
M: 0.91 (0.76, 1.08)
0.14 (-0.09, 0.38)
Obeying the law§
O: 0.89 (0.74, 1.07)
F: 0.91 (0.69, 1.18)
M: 0.87 (0.69, 1.10)
0.04 (-0.30, 0.38)
0.7
1.0
2.0
Cumulative odds ratio (x-axis on log scale)

**eFigure 12.** Adjusted associations between childhood abuse and 45 outcomes of interest and two flourishing composite indices, where “don’t know” and “refused” responses are counted towards the unexposed group.

Each panel shows the associations between the predictor and each of the continuous outcomes (**Panel A**), binary outcomes (**Panel B**), and ordinal outcomes (**Panel C**).

**Panel A**

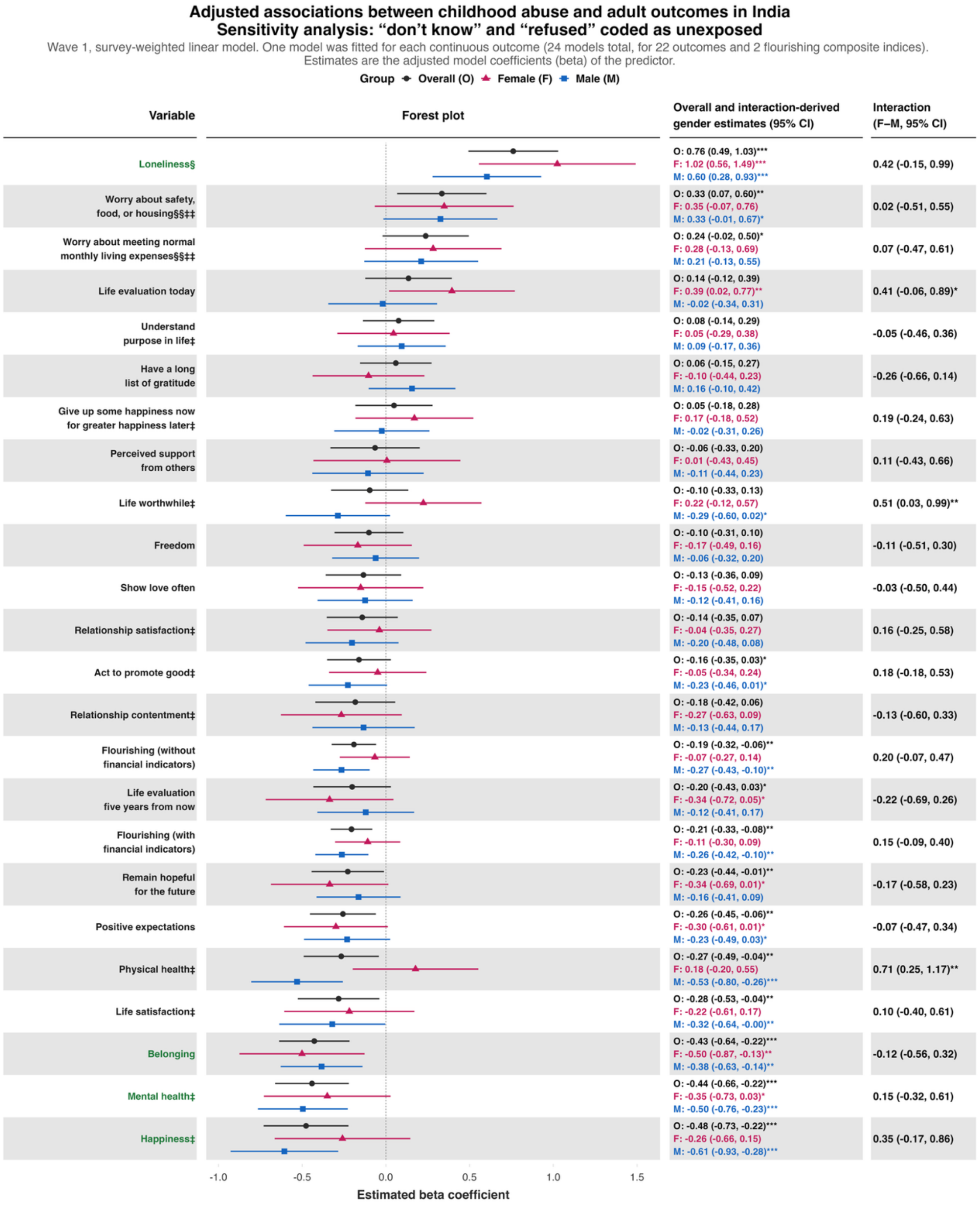

**Panel B**

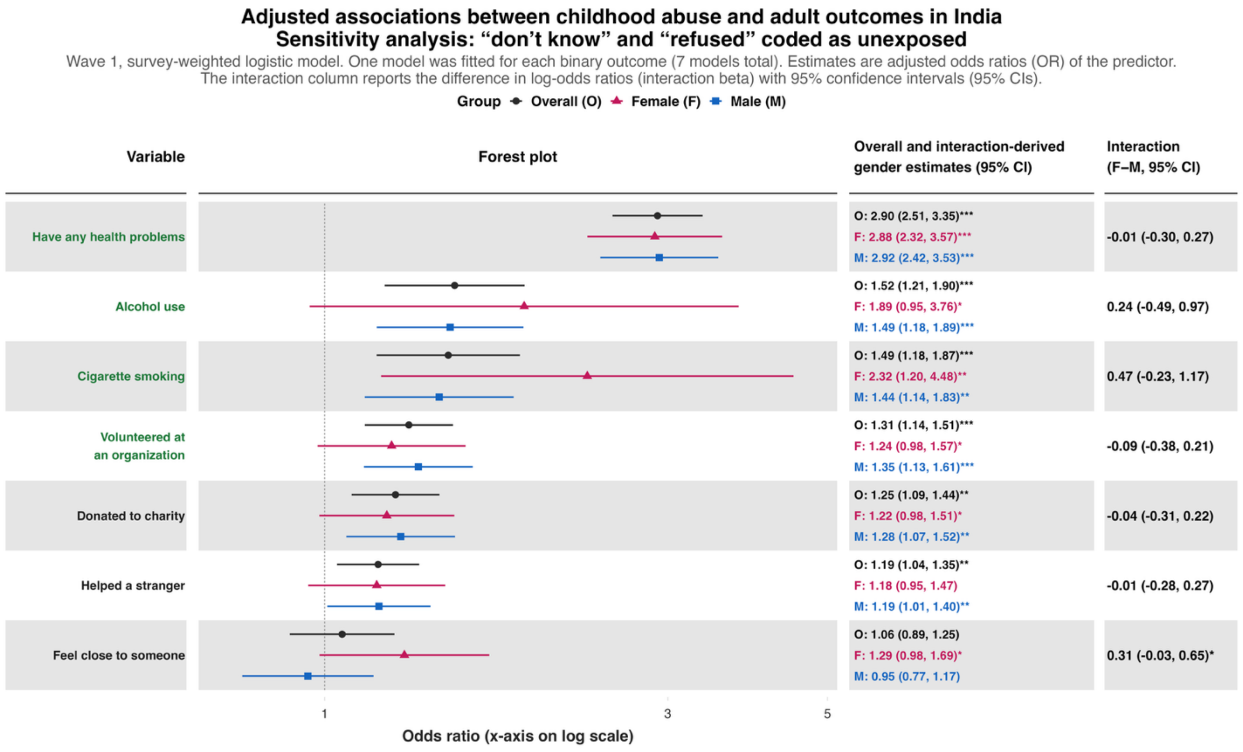
Adjusted associations between childhood abuse and adult outcomes in India
Sensitivity analysis: “don’t know” and “refused” coded as unexposed
Wave 1, survey-weighted logistic model. One model was fitted for each binary outcome (7 models total). Estimates are adjusted odds ratios (OR) of the predictor.
The interaction column reports the difference in log-odds ratios (interaction beta) with 95% confidence intervals (95% CIs).
Group Overall (O) Female (F) Male (M)
Variable
Forest plot
Overall and interaction-derived gender estimates (95% CI)
Interaction (F–M, 95% CI)
Have any health problems
O: 2.90 (2.51, 3.35)***
F: 2.88 (2.32, 3.57)***
M: 2.92 (2.42, 3.53)***
-0.01 (-0.30, 0.27)
Alcohol use
O: 1.52 (1.21, 1.90)***
F: 1.89 (0.95, 3.76)*
M: 1.49 (1.18, 1.89)***
0.24 (-0.49, 0.97)
Cigarette smoking
O: 1.49 (1.18, 1.87)***
F: 2.32 (1.20, 4.48)**
M: 1.44 (1.14, 1.83)**
0.47 (-0.23, 1.17)
Volunteered at an organization
O: 1.31 (1.14, 1.51)***
F: 1.24 (0.98, 1.57)*
M: 1.35 (1.13, 1.61)***
-0.09 (-0.38, 0.21)
Donated to charity
O: 1.25 (1.09, 1.44)**
F: 1.22 (0.98, 1.51)*
M: 1.28 (1.07, 1.52)**
-0.04 (-0.31, 0.22)
Helped a stranger
O: 1.19 (1.04, 1.35)**
F: 1.18 (0.95, 1.47)
M: 1.19 (1.01, 1.40)**
-0.01 (-0.28, 0.27)
Feel close to someone
O: 1.06 (0.89, 1.25)
F: 1.29 (0.98, 1.69)*
M: 0.95 (0.77, 1.17)
0.31 (-0.03, 0.65)*
1
3
5
Odds ratio (x-axis on log scale)

**Panel C**

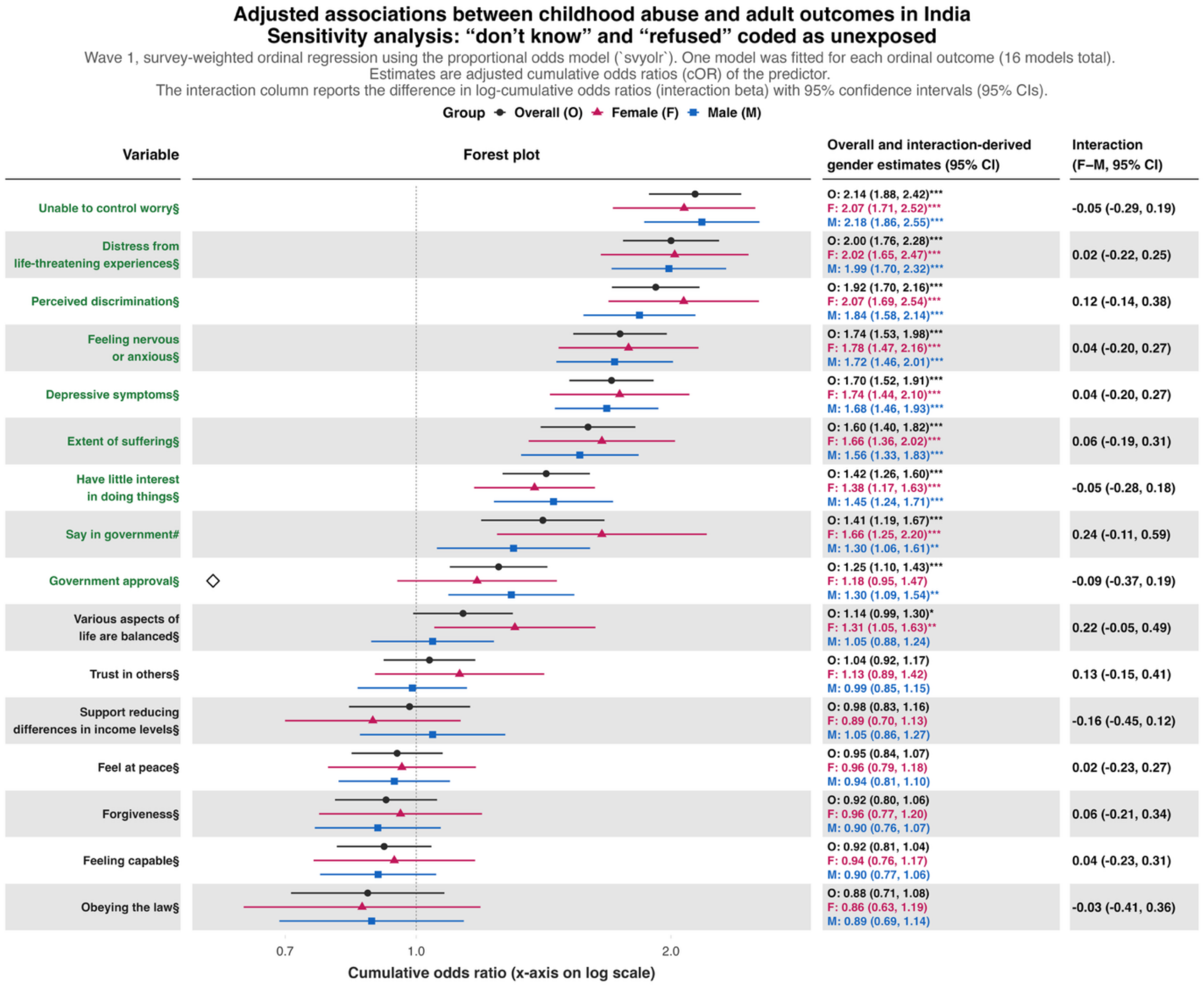
Adjusted associations between childhood abuse and adult outcomes in India
Sensitivity analysis: "don't know" and "refused" coded as unexposed
Wave 1, survey-weighted ordinal regression using the proportional odds model (`svyolr`). One model was fitted for each ordinal outcome (16 models total).
Estimates are adjusted cumulative odds ratios (cOR) of the predictor.
The interaction column reports the difference in log-cumulative odds ratios (interaction beta) with 95% confidence intervals (95% CIs).
Group Overall (O) Female (F) Male (M)
Variable
Forest plot
Overall and interaction-derived gender estimates (95% CI)
Interaction (F–M, 95% CI)
Unable to control worry§
O: 2.14 (1.88, 2.42)***
F: 2.07 (1.71, 2.52)***
M: 2.18 (1.86, 2.55)***
-0.05 (-0.29, 0.19)
Distress from life-threatening experiences§
O: 2.00 (1.76, 2.28)***
F: 2.02 (1.65, 2.47)***
M: 1.99 (1.70, 2.32)***
0.02 (-0.22, 0.25)
Perceived discrimination§
O: 1.92 (1.70, 2.16)***
F: 2.07 (1.69, 2.54)***
M: 1.84 (1.58, 2.14)***
0.12 (-0.14, 0.38)
Feeling nervous or anxious§
O: 1.74 (1.53, 1.98)***
F: 1.78 (1.47, 2.16)***
M: 1.72 (1.46, 2.01)***
0.04 (-0.20, 0.27)
Depressive symptoms§
O: 1.70 (1.52, 1.91)***
F: 1.74 (1.44, 2.10)***
M: 1.68 (1.46, 1.93)***
0.04 (-0.20, 0.27)
Extent of suffering§
O: 1.60 (1.40, 1.82)***
F: 1.66 (1.36, 2.02)***
M: 1.56 (1.33, 1.83)***
0.06 (-0.19, 0.31)
Have little interest in doing things§
O: 1.42 (1.26, 1.60)***
F: 1.38 (1.17, 1.63)***
M: 1.45 (1.24, 1.71)***
-0.05 (-0.28, 0.18)
Say in government#
O: 1.41 (1.19, 1.67)***
F: 1.66 (1.25, 2.20)***
M: 1.30 (1.06, 1.61)**
0.24 (-0.11, 0.59)
Government approval§
O: 1.25 (1.10, 1.43)***
F: 1.18 (0.95, 1.47)
M: 1.30 (1.09, 1.54)**
-0.09 (-0.37, 0.19)
Various aspects of life are balanced§
O: 1.14 (0.99, 1.30)*
F: 1.31 (1.05, 1.63)**
M: 1.05 (0.88, 1.24)
0.22 (-0.05, 0.49)
Trust in others§
O: 1.04 (0.92, 1.17)
F: 1.13 (0.89, 1.42)
M: 0.99 (0.85, 1.15)
0.13 (-0.15, 0.41)
Support reducing differences in income levels§
O: 0.98 (0.83, 1.16)
F: 0.89 (0.70, 1.13)
M: 1.05 (0.86, 1.27)
-0.16 (-0.45, 0.12)
Feel at peace§
O: 0.95 (0.84, 1.07)
F: 0.96 (0.79, 1.18)
M: 0.94 (0.81, 1.10)
0.02 (-0.23, 0.27)
Forgiveness§
O: 0.92 (0.80, 1.06)
F: 0.96 (0.77, 1.20)
M: 0.90 (0.76, 1.07)
0.06 (-0.21, 0.34)
Feeling capable§
O: 0.92 (0.81, 1.04)
F: 0.94 (0.76, 1.17)
M: 0.90 (0.77, 1.06)
0.04 (-0.23, 0.31)
Obeying the law§
O: 0.88 (0.71, 1.08)
F: 0.86 (0.63, 1.19)
M: 0.89 (0.69, 1.14)
-0.03 (-0.41, 0.36)
0.7
1.0
2.0
Cumulative odds ratio (x-axis on log scale)

**eFigure 13.** Adjusted associations between not feeling loved by mother and 45 outcomes of interest and two flourishing composite indices, where "does not apply" responses are counted towards the adverse exposure.

Each panel shows the associations between the predictor and each of the continuous outcomes (**Panel A**), binary outcomes (**Panel B**), and ordinal outcomes (**Panel C**).

**Panel A**

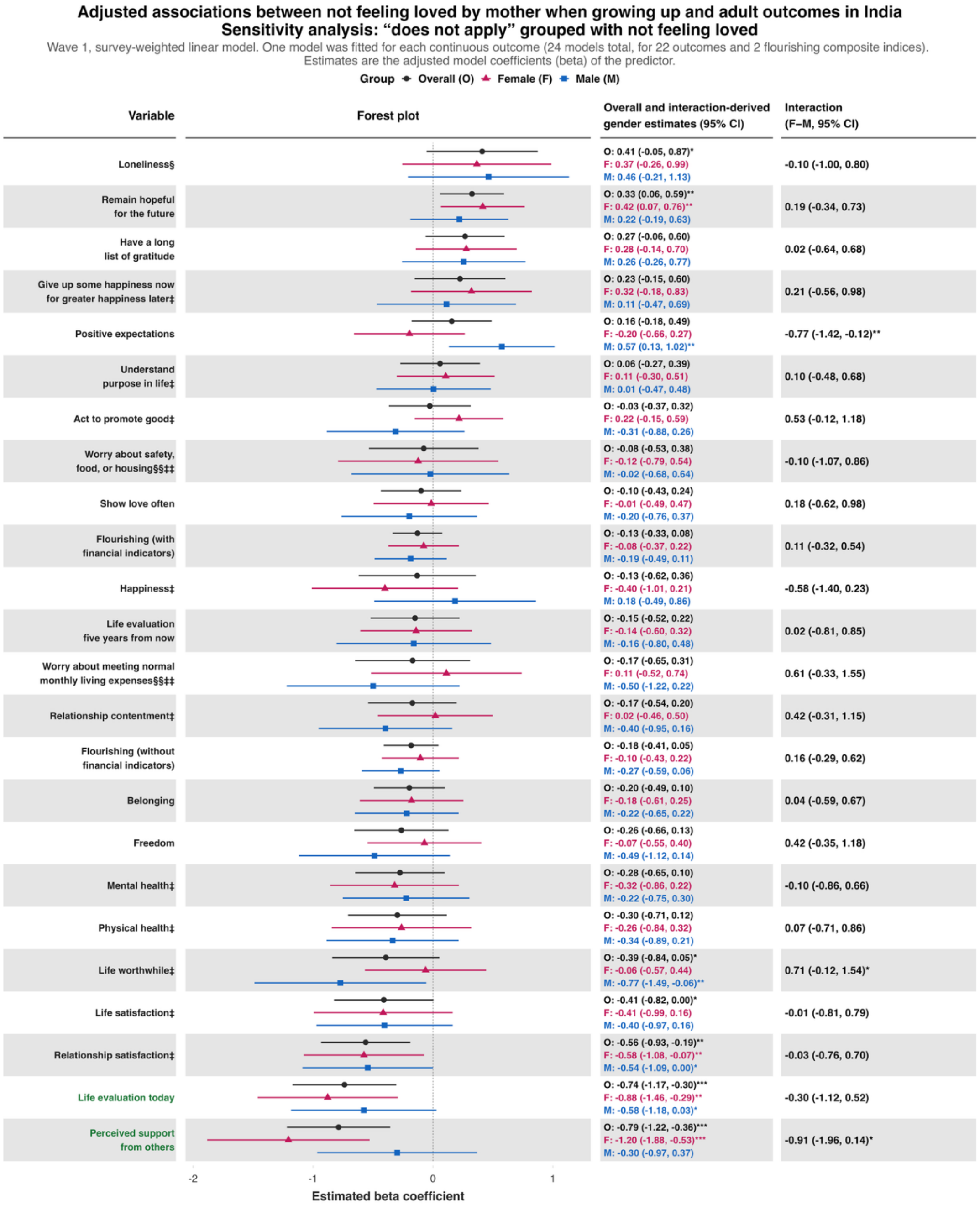

**Panel B**

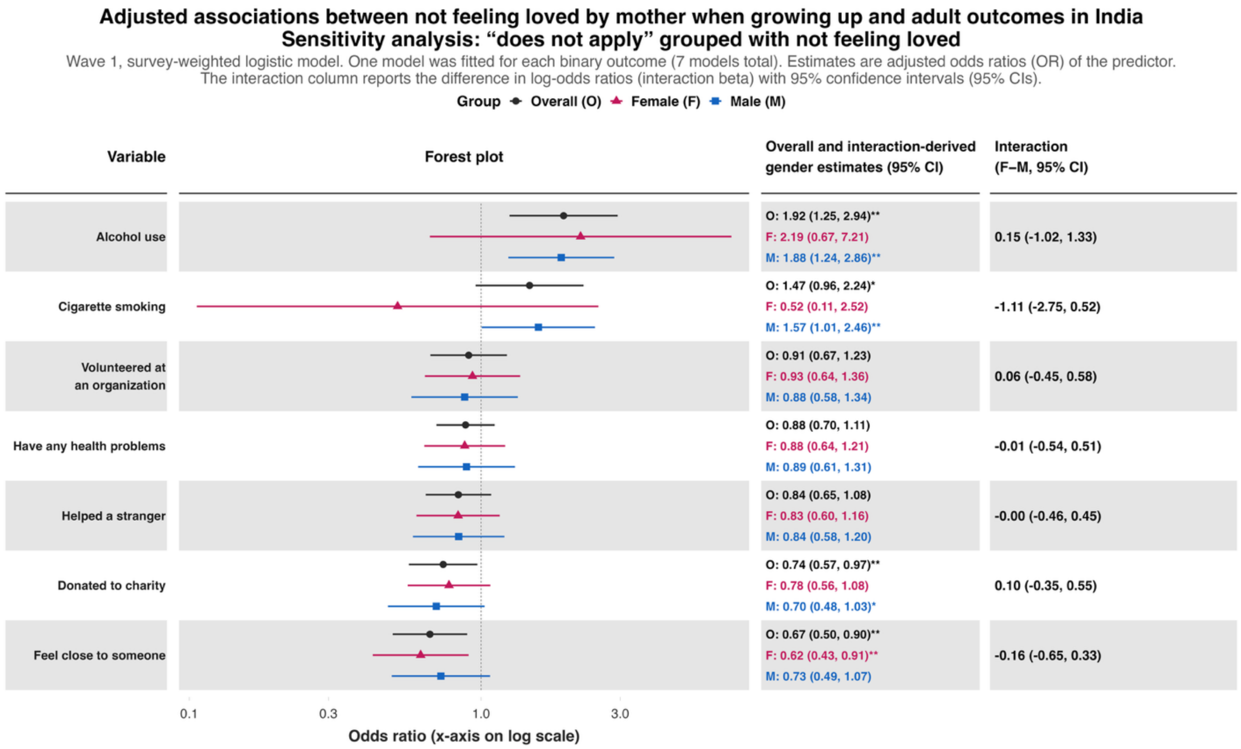

Adjusted associations between not feeling loved by mother when growing up and adult outcomes in India
Sensitivity analysis: "does not apply" grouped with not feeling loved
Wave 1, survey-weighted logistic model. One model was fitted for each binary outcome (7 models total). Estimates are adjusted odds ratios (OR) of the predictor.
The interaction column reports the difference in log-odds ratios (interaction beta) with 95% confidence intervals (95% CIs).
Group Overall (O) Female (F) Male (M)
Variable
Forest plot
Overall and interaction-derived gender estimates (95% CI)
Interaction (F–M, 95% CI)
Alcohol use
O: 1.92 (1.25, 2.94)**
F: 2.19 (0.67, 7.21)
M: 1.88 (1.24, 2.86)**
0.15 (-1.02, 1.33)
Cigarette smoking
O: 1.47 (0.96, 2.24)*
F: 0.52 (0.11, 2.52)
M: 1.57 (1.01, 2.46)**
-1.11 (-2.75, 0.52)
Volunteered at an organization
O: 0.91 (0.67, 1.23)
F: 0.93 (0.64, 1.36)
M: 0.88 (0.58, 1.34)
0.06 (-0.45, 0.58)
Have any health problems
O: 0.88 (0.70, 1.11)
F: 0.88 (0.64, 1.21)
M: 0.89 (0.61, 1.31)
-0.01 (-0.54, 0.51)
Helped a stranger
O: 0.84 (0.65, 1.08)
F: 0.83 (0.60, 1.16)
M: 0.84 (0.58, 1.20)
-0.00 (-0.46, 0.45)
Donated to charity
O: 0.74 (0.57, 0.97)**
F: 0.78 (0.56, 1.08)
M: 0.70 (0.48, 1.03)*
0.10 (-0.35, 0.55)
Feel close to someone
O: 0.67 (0.50, 0.90)**
F: 0.62 (0.43, 0.91)**
M: 0.73 (0.49, 1.07)
-0.16 (-0.65, 0.33)
0.1
0.3
1.0
3.0
Odds ratio (x-axis on log scale)

**Panel C**

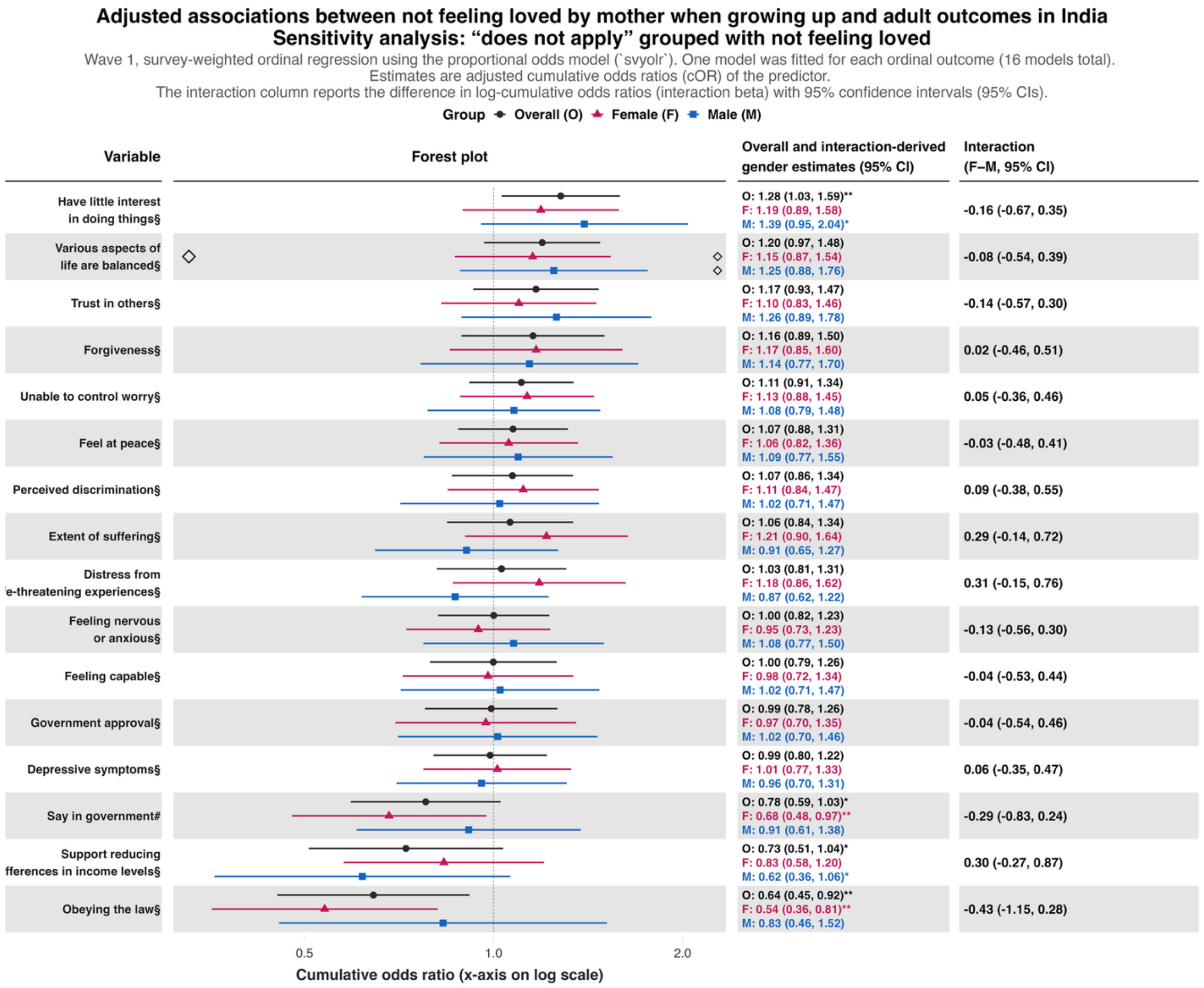
Adjusted associations between not feeling loved by mother when growing up and adult outcomes in India
Sensitivity analysis: “does not apply” grouped with not feeling loved
Wave 1, survey-weighted ordinal regression using the proportional odds model (`svyolr`). One model was fitted for each ordinal outcome (16 models total).
Estimates are adjusted cumulative odds ratios (cOR) of the predictor.
The interaction column reports the difference in log-cumulative odds ratios (interaction beta) with 95% confidence intervals (95% CIs).
Group Overall (O) Female (F) Male (M)
Variable
Forest plot
Overall and interaction-derived gender estimates (95% CI)
Interaction (F–M, 95% CI)
Have little interest in doing things§
O: 1.28 (1.03, 1.59)** F: 1.19 (0.89, 1.58) M: 1.39 (0.95, 2.04)*
-0.16 (-0.67, 0.35)
Various aspects of life are balanced§
O: 1.20 (0.97, 1.48) F: 1.15 (0.87, 1.54) M: 1.25 (0.88, 1.76)
-0.08 (-0.54, 0.39)
Trust in others§
O: 1.17 (0.93, 1.47) F: 1.10 (0.83, 1.46) M: 1.26 (0.89, 1.78)
-0.14 (-0.57, 0.30)
Forgiveness§
O: 1.16 (0.89, 1.50) F: 1.17 (0.85, 1.60) M: 1.14 (0.77, 1.70)
0.02 (-0.46, 0.51)
Unable to control worry§
O: 1.11 (0.91, 1.34) F: 1.13 (0.88, 1.45) M: 1.08 (0.79, 1.48)
0.05 (-0.36, 0.46)
Feel at peace§
O: 1.07 (0.88, 1.31) F: 1.06 (0.82, 1.36) M: 1.09 (0.77, 1.55)
-0.03 (-0.48, 0.41)
Perceived discrimination§
O: 1.07 (0.86, 1.34) F: 1.11 (0.84, 1.47) M: 1.02 (0.71, 1.47)
0.09 (-0.38, 0.55)
Extent of suffering§
O: 1.06 (0.84, 1.34) F: 1.21 (0.90, 1.64) M: 0.91 (0.65, 1.27)
0.29 (-0.14, 0.72)
Distress from 'e-threatening experiences§
O: 1.03 (0.81, 1.31) F: 1.18 (0.86, 1.62) M: 0.87 (0.62, 1.22)
0.31 (-0.15, 0.76)
Feeling nervous or anxious§
O: 1.00 (0.82, 1.23) F: 0.95 (0.73, 1.23) M: 1.08 (0.77, 1.50)
-0.13 (-0.56, 0.30)
Feeling capable§
O: 1.00 (0.79, 1.26) F: 0.98 (0.72, 1.34) M: 1.02 (0.71, 1.47)
-0.04 (-0.53, 0.44)
Government approval§
O: 0.99 (0.78, 1.26) F: 0.97 (0.70, 1.35) M: 1.02 (0.70, 1.46)
-0.04 (-0.54, 0.46)
Depressive symptoms§
O: 0.99 (0.80, 1.22) F: 1.01 (0.77, 1.33) M: 0.96 (0.70, 1.31)
0.06 (-0.35, 0.47)
Say in government#
O: 0.78 (0.59, 1.03)* F: 0.68 (0.48, 0.97)** M: 0.91 (0.61, 1.38)
-0.29 (-0.83, 0.24)
Support reducing fferences in income levels§
O: 0.73 (0.51, 1.04)* F: 0.83 (0.58, 1.20) M: 0.62 (0.36, 1.06)*
0.30 (-0.27, 0.87)
Obeying the law§
O: 0.64 (0.45, 0.92)** F: 0.54 (0.36, 0.81)** M: 0.83 (0.46, 1.52)
-0.43 (-1.15, 0.28)
0.5
1.0
2.0
Cumulative odds ratio (x-axis on log scale)

**eFigure 14.** Adjusted associations between not feeling loved by father and 45 outcomes of interest and two flourishing composite indices, where “does not apply” responses are counted towards the adverse exposure.

Each panel shows the associations between the predictor and each of the continuous outcomes (**Panel A**), binary outcomes (**Panel B**), and ordinal outcomes (**Panel C**).

**Panel A**

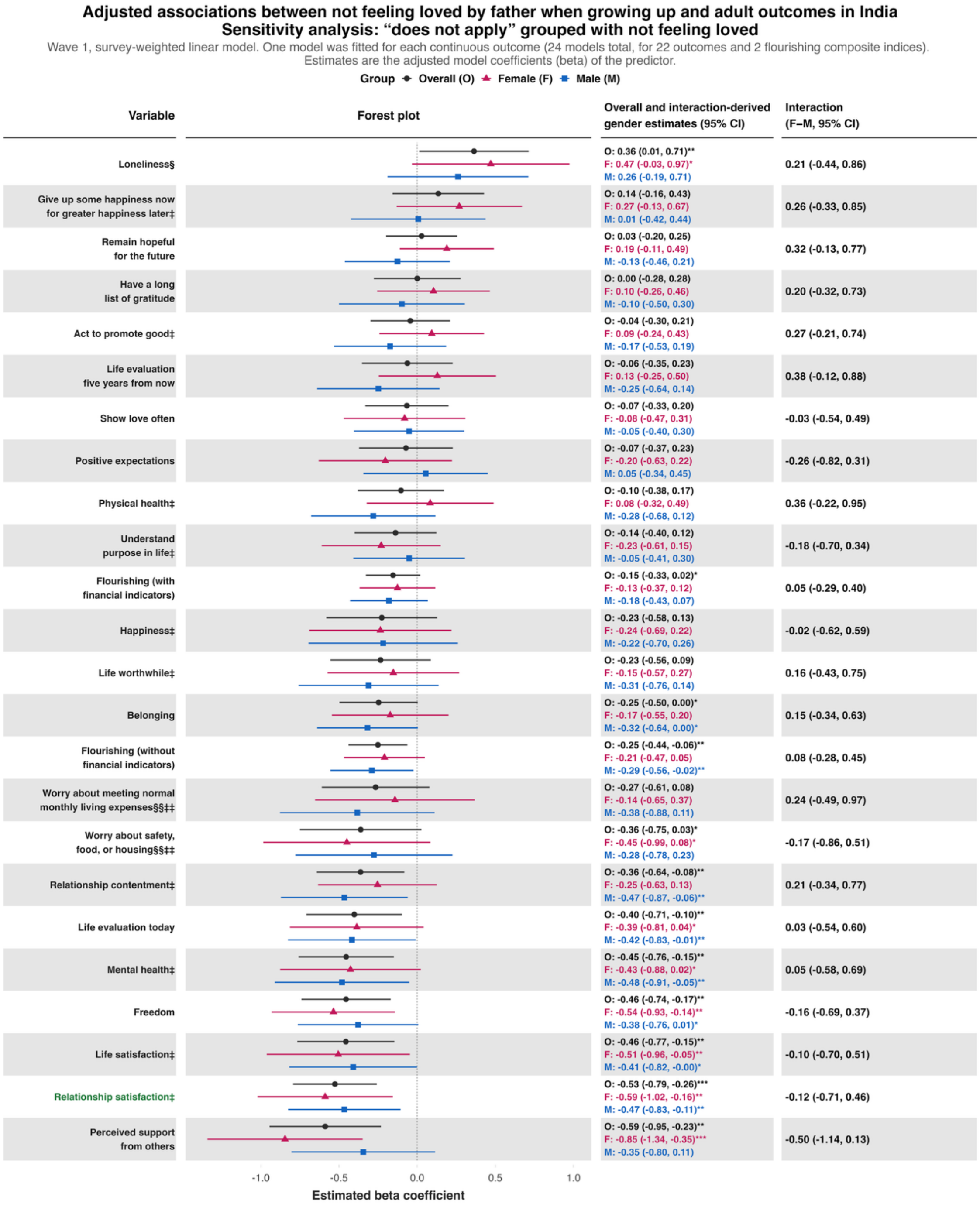

**Panel B**

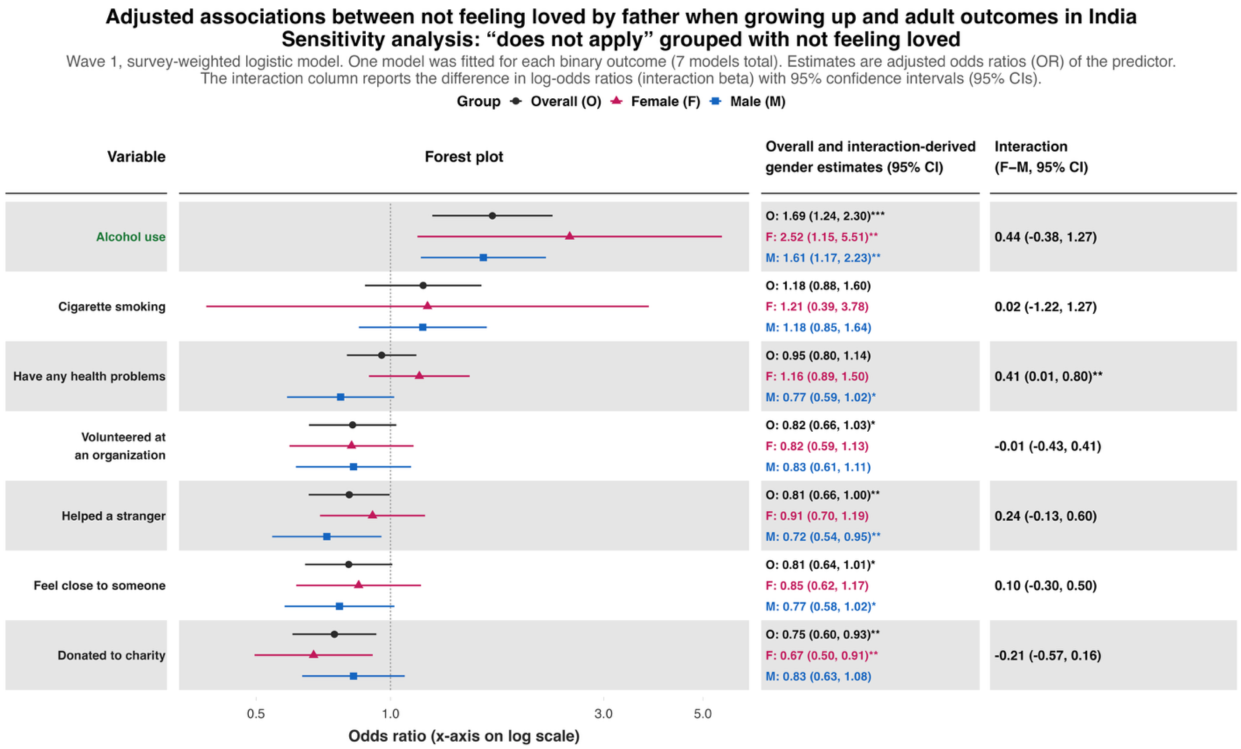

Adjusted associations between not feeling loved by father when growing up and adult outcomes in India
Sensitivity analysis: “does not apply” grouped with not feeling loved
Wave 1, survey-weighted logistic model. One model was fitted for each binary outcome (7 models total). Estimates are adjusted odds ratios (OR) of the predictor.
The interaction column reports the difference in log-odds ratios (interaction beta) with 95% confidence intervals (95% CIs).
Group
Overall (O)
Female (F)
Male (M)
Variable
Forest plot
Overall and interaction-derived gender estimates (95% CI)
Interaction (F–M, 95% CI)
Alcohol use
O: 1.69 (1.24, 2.30)***
F: 2.52 (1.15, 5.51)**
M: 1.61 (1.17, 2.23)**
0.44 (-0.38, 1.27)
Cigarette smoking
O: 1.18 (0.88, 1.60)
F: 1.21 (0.39, 3.78)
M: 1.18 (0.85, 1.64)
0.02 (-1.22, 1.27)
Have any health problems
O: 0.95 (0.80, 1.14)
F: 1.16 (0.89, 1.50)
M: 0.77 (0.59, 1.02)*
0.41 (0.01, 0.80)**
Volunteered at an organization
O: 0.82 (0.66, 1.03)*
F: 0.82 (0.59, 1.13)
M: 0.83 (0.61, 1.11)
-0.01 (-0.43, 0.41)
Helped a stranger
O: 0.81 (0.66, 1.00)**
F: 0.91 (0.70, 1.19)
M: 0.72 (0.54, 0.95)**
0.24 (-0.13, 0.60)
Feel close to someone
O: 0.81 (0.64, 1.01)*
F: 0.85 (0.62, 1.17)
M: 0.77 (0.58, 1.02)*
0.10 (-0.30, 0.50)
Donated to charity
O: 0.75 (0.60, 0.93)**
F: 0.67 (0.50, 0.91)**
M: 0.83 (0.63, 1.08)
-0.21 (-0.57, 0.16)
0.5
1.0
3.0
5.0
Odds ratio (x-axis on log scale)

**Panel C**

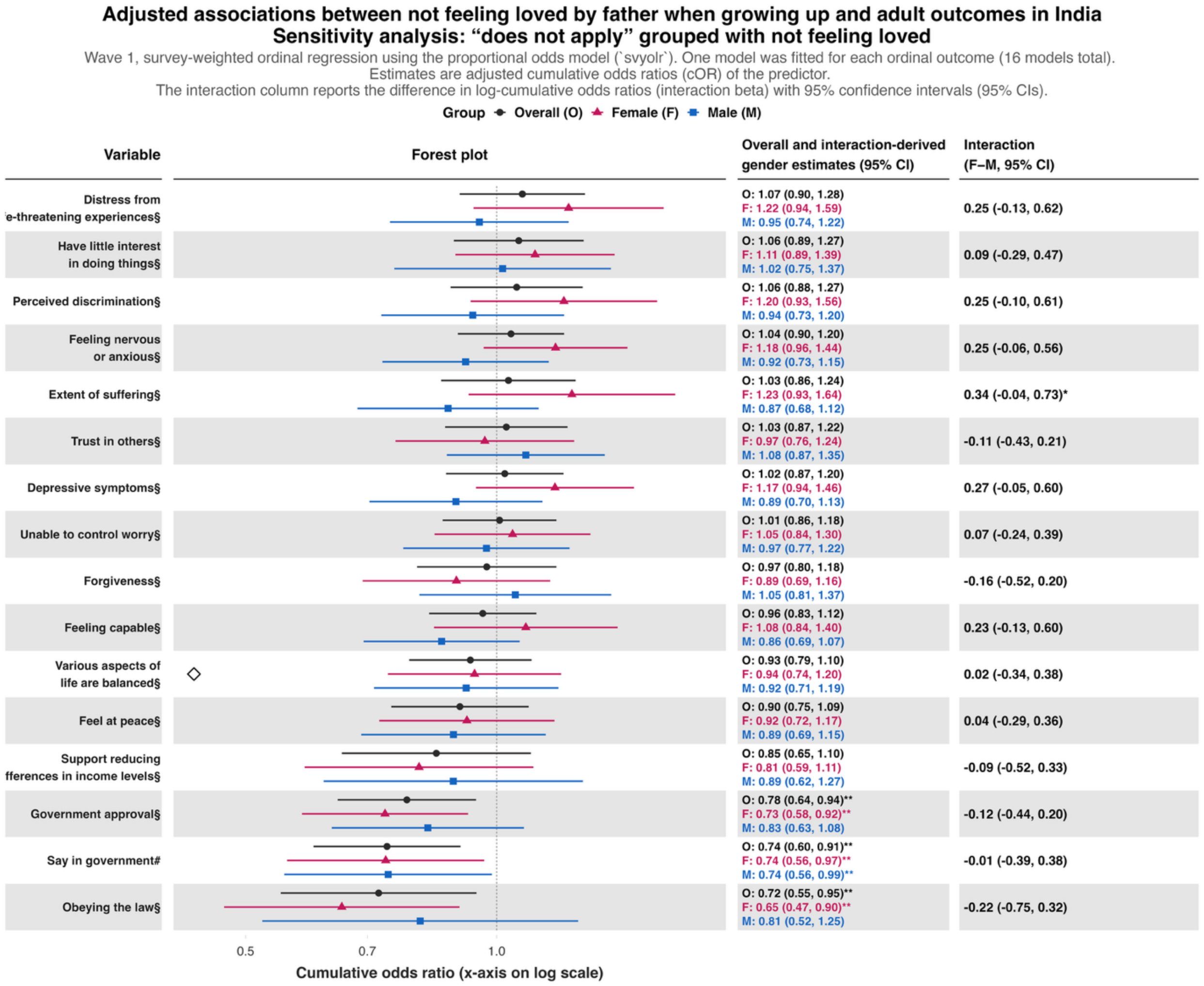

Adjusted associations between not feeling loved by father when growing up and adult outcomes in India
Sensitivity analysis: “does not apply” grouped with not feeling loved
Wave 1, survey-weighted ordinal regression using the proportional odds model (`svyolr`). One model was fitted for each ordinal outcome (16 models total).
Estimates are adjusted cumulative odds ratios (cOR) of the predictor.
The interaction column reports the difference in log-cumulative odds ratios (interaction beta) with 95% confidence intervals (95% CIs).
Group Overall (O) Female (F) Male (M)
Variable
Forest plot
Overall and interaction-derived gender estimates (95% CI)
Interaction (F–M, 95% CI)
Distress from
'e-threatening experiences§
O: 1.07 (0.90, 1.28)
F: 1.22 (0.94, 1.59)
M: 0.95 (0.74, 1.22)
0.25 (-0.13, 0.62)
Have little interest in doing things§
O: 1.06 (0.89, 1.27)
F: 1.11 (0.89, 1.39)
M: 1.02 (0.75, 1.37)
0.09 (-0.29, 0.47)
Perceived discrimination§
O: 1.06 (0.88, 1.27)
F: 1.20 (0.93, 1.56)
M: 0.94 (0.73, 1.20)
0.25 (-0.10, 0.61)
Feeling nervous or anxious§
O: 1.04 (0.90, 1.20)
F: 1.18 (0.96, 1.44)
M: 0.92 (0.73, 1.15)
0.25 (-0.06, 0.56)
Extent of suffering§
O: 1.03 (0.86, 1.24)
F: 1.23 (0.93, 1.64)
M: 0.87 (0.68, 1.12)
0.34 (-0.04, 0.73)*
Trust in others§
O: 1.03 (0.87, 1.22)
F: 0.97 (0.76, 1.24)
M: 1.08 (0.87, 1.35)
-0.11 (-0.43, 0.21)
Depressive symptoms§
O: 1.02 (0.87, 1.20)
F: 1.17 (0.94, 1.46)
M: 0.89 (0.70, 1.13)
0.27 (-0.05, 0.60)
Unable to control worry§
O: 1.01 (0.86, 1.18)
F: 1.05 (0.84, 1.30)
M: 0.97 (0.77, 1.22)
0.07 (-0.24, 0.39)
Forgiveness§
O: 0.97 (0.80, 1.18)
F: 0.89 (0.69, 1.16)
M: 1.05 (0.81, 1.37)
-0.16 (-0.52, 0.20)
Feeling capable§
O: 0.96 (0.83, 1.12)
F: 1.08 (0.84, 1.40)
M: 0.86 (0.69, 1.07)
0.23 (-0.13, 0.60)
Various aspects of life are balanced§
O: 0.93 (0.79, 1.10)
F: 0.94 (0.74, 1.20)
M: 0.92 (0.71, 1.19)
0.02 (-0.34, 0.38)
Feel at peace§
O: 0.90 (0.75, 1.09)
F: 0.92 (0.72, 1.17)
M: 0.89 (0.69, 1.15)
0.04 (-0.29, 0.36)
Support reducing
fferences in income levels§
O: 0.85 (0.65, 1.10)
F: 0.81 (0.59, 1.11)
M: 0.89 (0.62, 1.27)
-0.09 (-0.52, 0.33)
Government approval§
O: 0.78 (0.64, 0.94)**
F: 0.73 (0.58, 0.92)**
M: 0.83 (0.63, 1.08)
-0.12 (-0.44, 0.20)
Say in government#
O: 0.74 (0.60, 0.91)**
F: 0.74 (0.56, 0.97)**
M: 0.74 (0.56, 0.99)**
-0.01 (-0.39, 0.38)
Obeying the law§
O: 0.72 (0.55, 0.95)**
F: 0.65 (0.47, 0.90)**
M: 0.81 (0.52, 1.25)
-0.22 (-0.75, 0.32)
0.5
0.7
1.0
Cumulative odds ratio (x-axis on log scale)

**eFigure 15.** Adjusted associations between bad or very bad relationship with mother and 45 outcomes of interest and two flourishing composite indices, where "does not apply" responses are counted towards the adverse exposure.

Each panel shows the associations between the predictor and each of the continuous outcomes (**Panel A**), binary outcomes (**Panel B**), and ordinal outcomes (**Panel C**).

**Panel A**

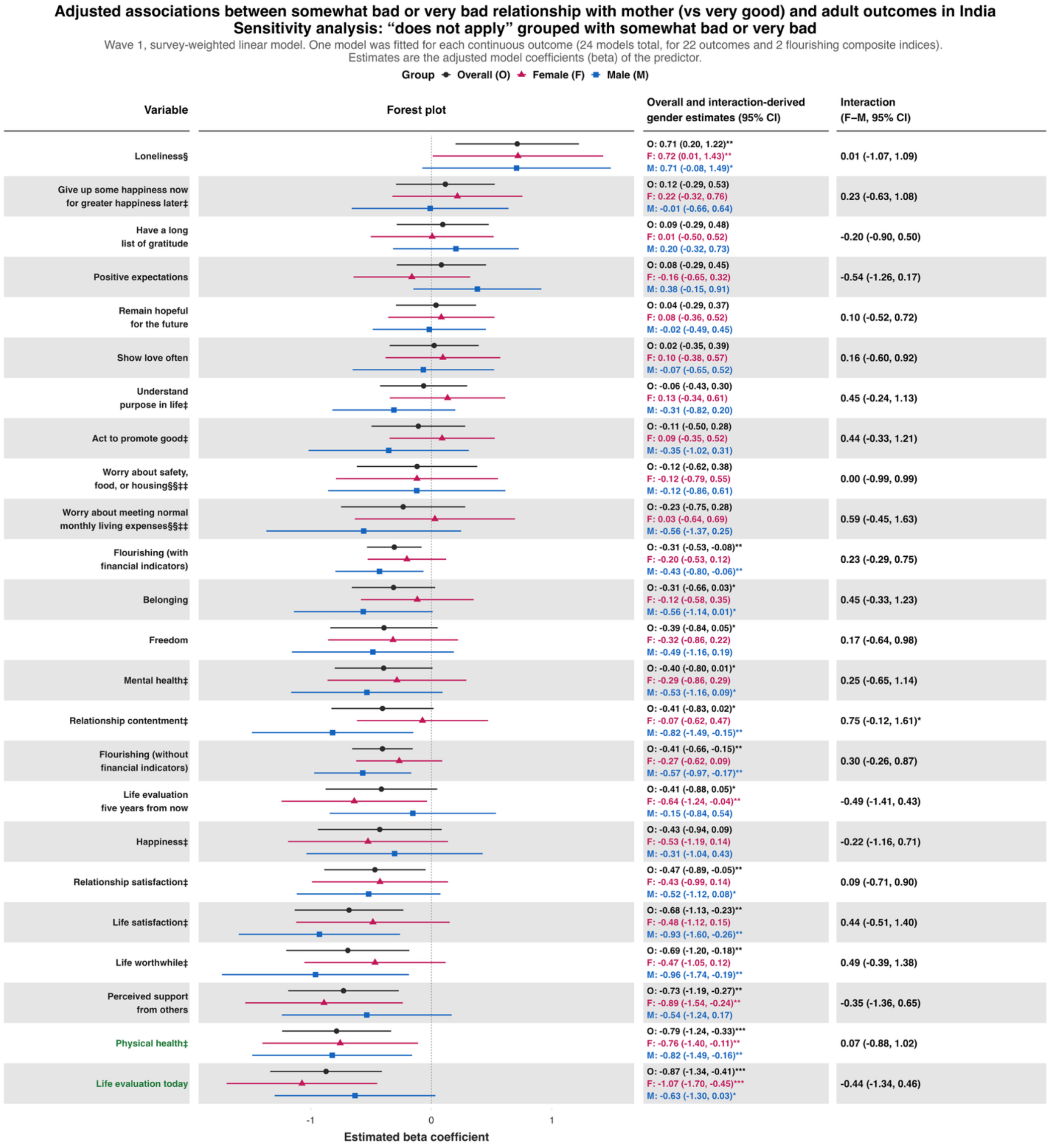

**Panel B**

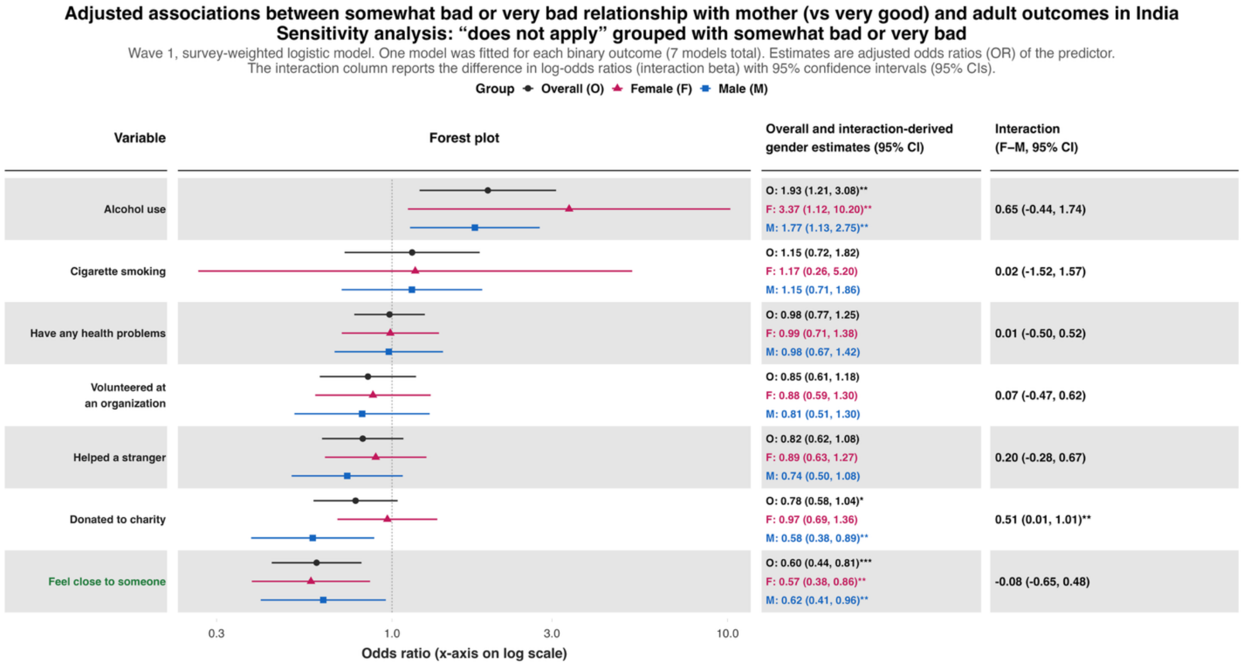
Adjusted associations between somewhat bad or very bad relationship with mother (vs very good) and adult outcomes in India
Sensitivity analysis: "does not apply" grouped with somewhat bad or very bad
Wave 1, survey-weighted logistic model. One model was fitted for each binary outcome (7 models total). Estimates are adjusted odds ratios (OR) of the predictor.
The interaction column reports the difference in log-odds ratios (interaction beta) with 95% confidence intervals (95% CIs).
Group Overall (O) Female (F) Male (M)
Variable
Forest plot
Overall and interaction-derived gender estimates (95% CI)
Interaction (F–M, 95% CI)
Alcohol use
O: 1.93 (1.21, 3.08)**
F: 3.37 (1.12, 10.20)**
M: 1.77 (1.13, 2.75)**
0.65 (-0.44, 1.74)
Cigarette smoking
O: 1.15 (0.72, 1.82)
F: 1.17 (0.26, 5.20)
M: 1.15 (0.71, 1.86)
0.02 (-1.52, 1.57)
Have any health problems
O: 0.98 (0.77, 1.25)
F: 0.99 (0.71, 1.38)
M: 0.98 (0.67, 1.42)
0.01 (-0.50, 0.52)
Volunteered at an organization
O: 0.85 (0.61, 1.18)
F: 0.88 (0.59, 1.30)
M: 0.81 (0.51, 1.30)
0.07 (-0.47, 0.62)
Helped a stranger
O: 0.82 (0.62, 1.08)
F: 0.89 (0.63, 1.27)
M: 0.74 (0.50, 1.08)
0.20 (-0.28, 0.67)
Donated to charity
O: 0.78 (0.58, 1.04)*
F: 0.97 (0.69, 1.36)
M: 0.58 (0.38, 0.89)**
0.51 (0.01, 1.01)**
Feel close to someone
O: 0.60 (0.44, 0.81)***
F: 0.57 (0.38, 0.86)**
M: 0.62 (0.41, 0.96)**
-0.08 (-0.65, 0.48)
0.3
1.0
3.0
10.0
Odds ratio (x-axis on log scale)

**Panel C**

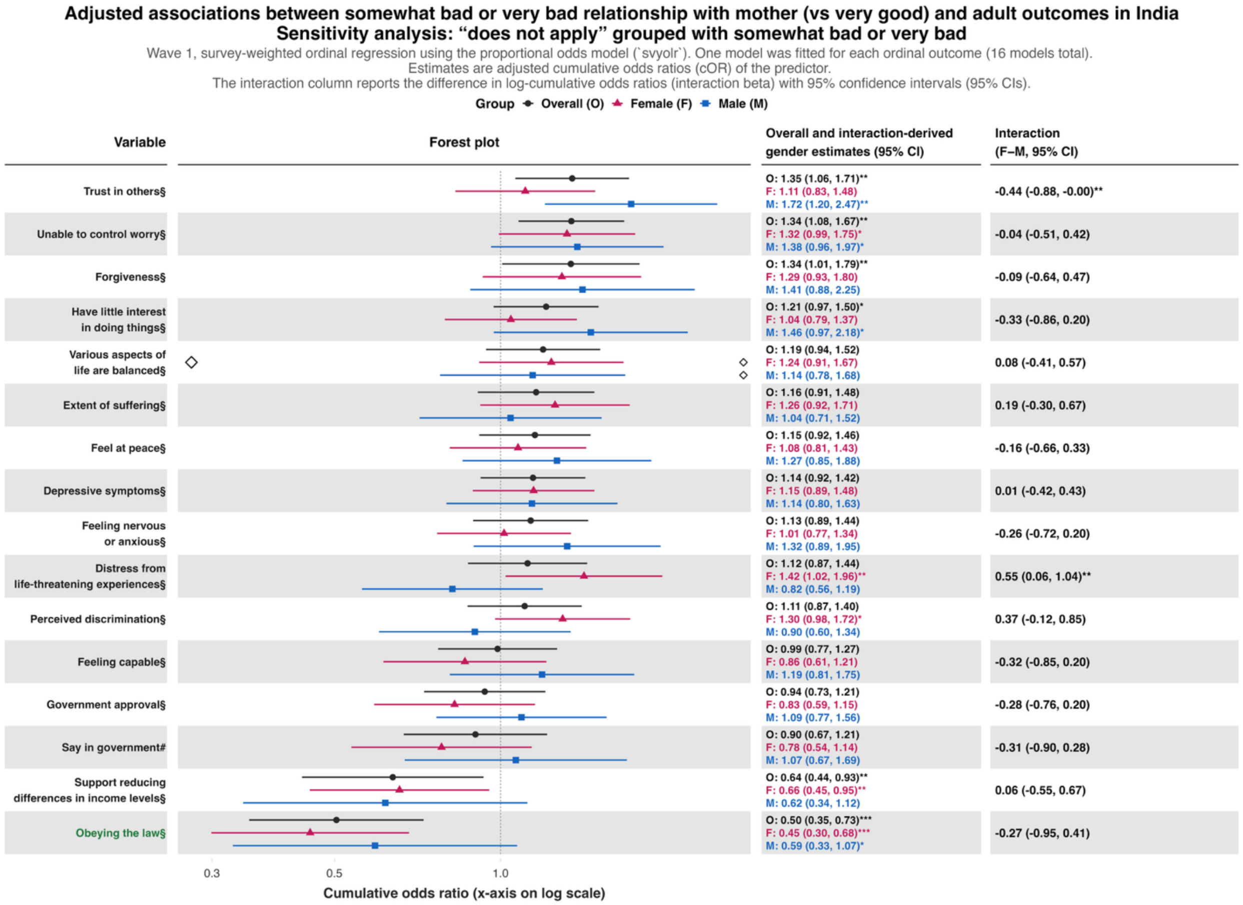


| Variable | Overall and interaction-derived gender estimates (95% CI) | Interaction (F–M, 95% CI) |
|---|---|---|
| Trust in others§ | O: 1.35 (1.06, 1.71)**<br>F: 1.11 (0.83, 1.48)<br>M: 1.72 (1.20, 2.47)** | -0.44 (-0.88, -0.00)** |
| Unable to control worry§ | O: 1.34 (1.08, 1.67)**<br>F: 1.32 (0.99, 1.75)*<br>M: 1.38 (0.96, 1.97)* | -0.04 (-0.51, 0.42) |
| Forgiveness§ | O: 1.34 (1.01, 1.79)**<br>F: 1.29 (0.93, 1.80)<br>M: 1.41 (0.88, 2.25) | -0.09 (-0.64, 0.47) |
| Have little interest in doing things§ | O: 1.21 (0.97, 1.50)*<br>F: 1.04 (0.79, 1.37)<br>M: 1.46 (0.97, 2.18)* | -0.33 (-0.86, 0.20) |
| Various aspects of life are balanced§ | O: 1.19 (0.94, 1.52)<br>F: 1.24 (0.91, 1.67)<br>M: 1.14 (0.78, 1.68) | 0.08 (-0.41, 0.57) |
| Extent of suffering§ | O: 1.16 (0.91, 1.48)<br>F: 1.26 (0.92, 1.71)<br>M: 1.04 (0.71, 1.52) | 0.19 (-0.30, 0.67) |
| Feel at peace§ | O: 1.15 (0.92, 1.46)<br>F: 1.08 (0.81, 1.43)<br>M: 1.27 (0.85, 1.88) | -0.16 (-0.66, 0.33) |
| Depressive symptoms§ | O: 1.14 (0.92, 1.42)<br>F: 1.15 (0.89, 1.48)<br>M: 1.14 (0.80, 1.63) | 0.01 (-0.42, 0.43) |
| Feeling nervous or anxious§ | O: 1.13 (0.89, 1.44)<br>F: 1.01 (0.77, 1.34)<br>M: 1.32 (0.89, 1.95) | -0.26 (-0.72, 0.20) |
| Distress from life-threatening experiences§ | O: 1.12 (0.87, 1.44)<br>F: 1.42 (1.02, 1.96)**<br>M: 0.82 (0.56, 1.19) | 0.55 (0.06, 1.04)** |
| Perceived discrimination§ | O: 1.11 (0.87, 1.40)<br>F: 1.30 (0.98, 1.72)*<br>M: 0.90 (0.60, 1.34) | 0.37 (-0.12, 0.85) |
| Feeling capable§ | O: 0.99 (0.77, 1.27)<br>F: 0.86 (0.61, 1.21)<br>M: 1.19 (0.81, 1.75) | -0.32 (-0.85, 0.20) |
| Government approval§ | O: 0.94 (0.73, 1.21)<br>F: 0.83 (0.59, 1.15)<br>M: 1.09 (0.77, 1.56) | -0.28 (-0.76, 0.20) |
| Say in government# | O: 0.90 (0.67, 1.21)<br>F: 0.78 (0.54, 1.14)<br>M: 1.07 (0.67, 1.69) | -0.31 (-0.90, 0.28) |
| Support reducing differences in income levels§ | O: 0.64 (0.44, 0.93)**<br>F: 0.66 (0.45, 0.95)**<br>M: 0.62 (0.34, 1.12) | 0.06 (-0.55, 0.67) |
| Obeying the law§ | O: 0.50 (0.35, 0.73)***<br>F: 0.45 (0.30, 0.68)***<br>M: 0.59 (0.33, 1.07)* | -0.27 (-0.95, 0.41) |

**eFigure 16.** Adjusted associations between bad or very bad relationship with father and 45 outcomes of interest and two flourishing composite indices, where "does not apply" responses are counted towards the adverse exposure.

Each panel shows the associations between the predictor and each of the continuous outcomes (**Panel A**), binary outcomes (**Panel B**), and ordinal outcomes (**Panel C**).

**Panel A**

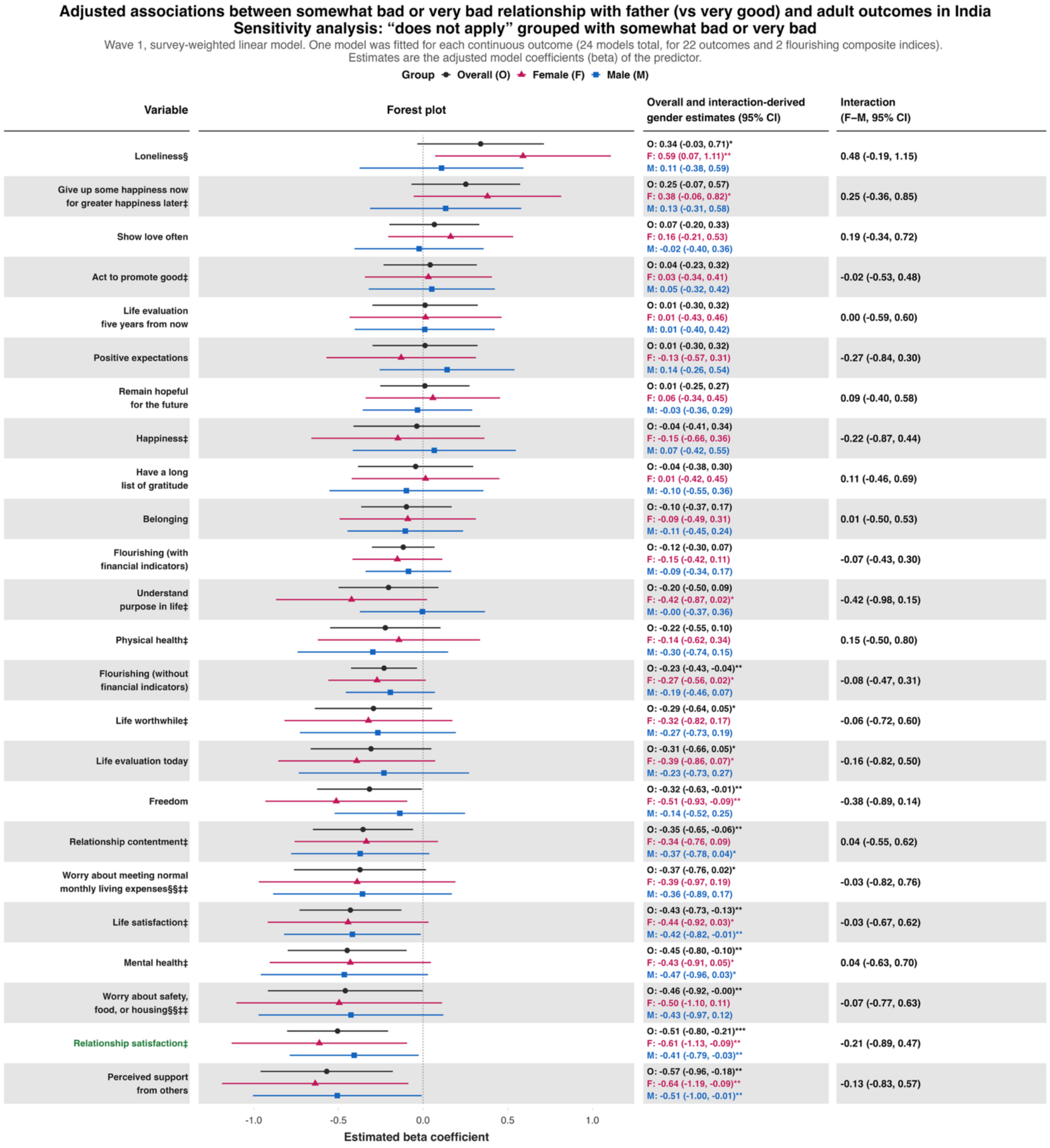

**Panel B**

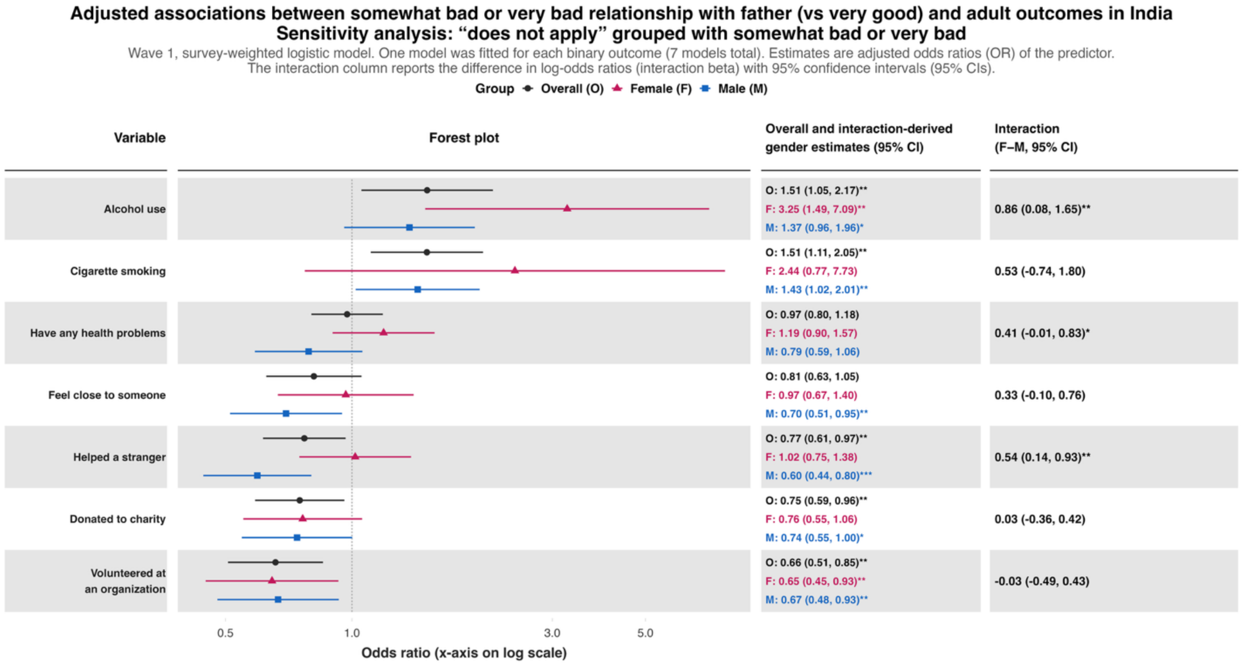
Adjusted associations between somewhat bad or very bad relationship with father (vs very good) and adult outcomes in India
Sensitivity analysis: "does not apply" grouped with somewhat bad or very bad
Wave 1, survey-weighted logistic model. One model was fitted for each binary outcome (7 models total). Estimates are adjusted odds ratios (OR) of the predictor.
The interaction column reports the difference in log-odds ratios (interaction beta) with 95% confidence intervals (95% CIs).
Group Overall (O) Female (F) Male (M)
Variable
Forest plot
Overall and interaction-derived gender estimates (95% CI)
Interaction (F–M, 95% CI)
Alcohol use
O: 1.51 (1.05, 2.17)**
F: 3.25 (1.49, 7.09)**
M: 1.37 (0.96, 1.96)*
0.86 (0.08, 1.65)**
Cigarette smoking
O: 1.51 (1.11, 2.05)**
F: 2.44 (0.77, 7.73)
M: 1.43 (1.02, 2.01)**
0.53 (-0.74, 1.80)
Have any health problems
O: 0.97 (0.80, 1.18)
F: 1.19 (0.90, 1.57)
M: 0.79 (0.59, 1.06)
0.41 (-0.01, 0.83)*
Feel close to someone
O: 0.81 (0.63, 1.05)
F: 0.97 (0.67, 1.40)
M: 0.70 (0.51, 0.95)**
0.33 (-0.10, 0.76)
Helped a stranger
O: 0.77 (0.61, 0.97)**
F: 1.02 (0.75, 1.38)
M: 0.60 (0.44, 0.80)***
0.54 (0.14, 0.93)**
Donated to charity
O: 0.75 (0.59, 0.96)**
F: 0.76 (0.55, 1.06)
M: 0.74 (0.55, 1.00)*
0.03 (-0.36, 0.42)
Volunteered at an organization
O: 0.66 (0.51, 0.85)**
F: 0.65 (0.45, 0.93)**
M: 0.67 (0.48, 0.93)**
-0.03 (-0.49, 0.43)
0.5
1.0
3.0
5.0
Odds ratio (x-axis on log scale)

**Panel C**

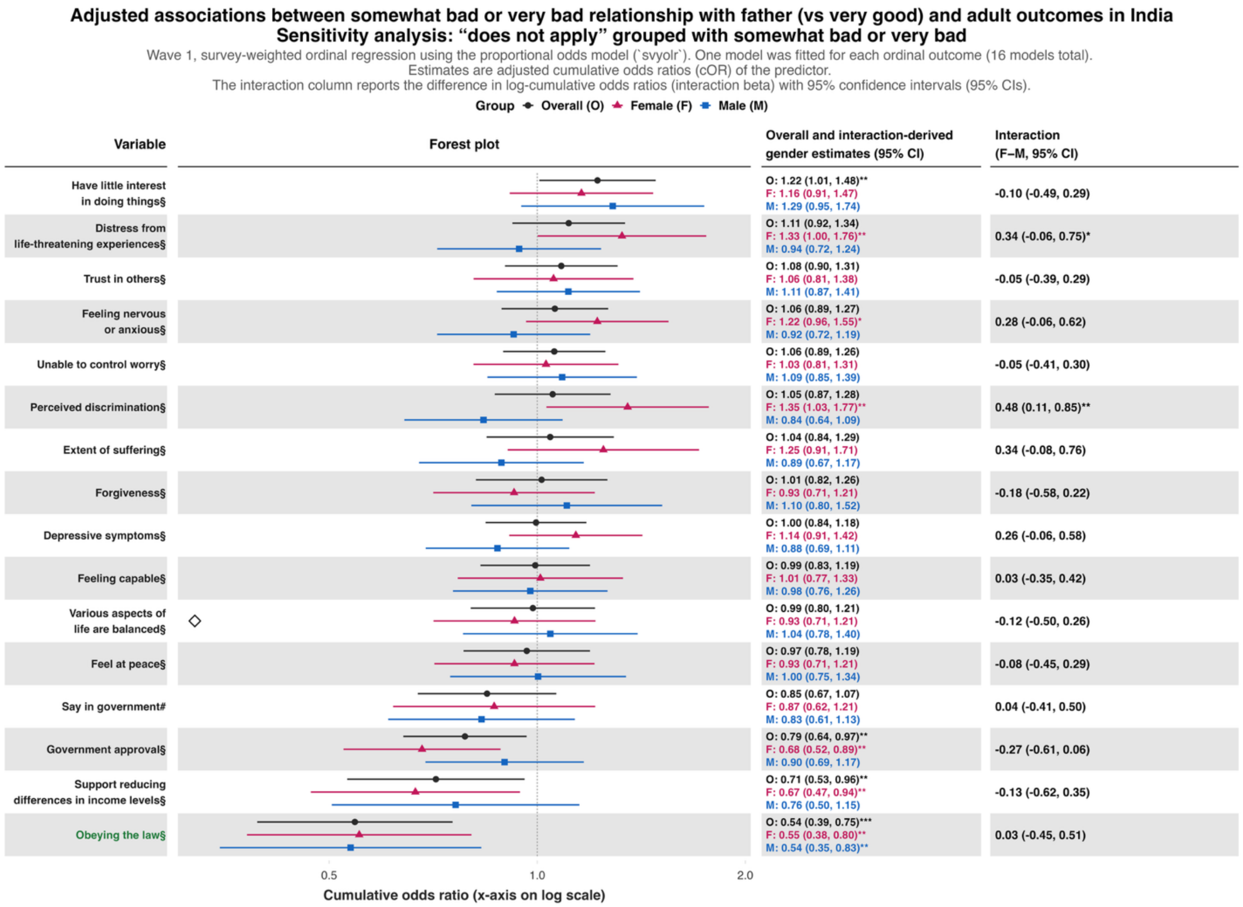

Adjusted associations between somewhat bad or very bad relationship with father (vs very good) and adult outcomes in India
Sensitivity analysis: “does not apply” grouped with somewhat bad or very bad
Wave 1, survey-weighted ordinal regression using the proportional odds model (`svyolr`). One model was fitted for each ordinal outcome (16 models total).
Estimates are adjusted cumulative odds ratios (cOR) of the predictor.
The interaction column reports the difference in log-cumulative odds ratios (interaction beta) with 95% confidence intervals (95% CIs).
Group Overall (O) Female (F) Male (M)
Variable
Forest plot
Overall and interaction-derived gender estimates (95% CI)
Interaction (F–M, 95% CI)
Have little interest in doing things§
O: 1.22 (1.01, 1.48)**
F: 1.16 (0.91, 1.47)
M: 1.29 (0.95, 1.74)
-0.10 (-0.49, 0.29)
Distress from life-threatening experiences§
O: 1.11 (0.92, 1.34)
F: 1.33 (1.00, 1.76)**
M: 0.94 (0.72, 1.24)
0.34 (-0.06, 0.75)*
Trust in others§
O: 1.08 (0.90, 1.31)
F: 1.06 (0.81, 1.38)
M: 1.11 (0.87, 1.41)
-0.05 (-0.39, 0.29)
Feeling nervous or anxious§
O: 1.06 (0.89, 1.27)
F: 1.22 (0.96, 1.55)*
M: 0.92 (0.72, 1.19)
0.28 (-0.06, 0.62)
Unable to control worry§
O: 1.06 (0.89, 1.26)
F: 1.03 (0.81, 1.31)
M: 1.09 (0.85, 1.39)
-0.05 (-0.41, 0.30)
Perceived discrimination§
O: 1.05 (0.87, 1.28)
F: 1.35 (1.03, 1.77)**
M: 0.84 (0.64, 1.09)
0.48 (0.11, 0.85)**
Extent of suffering§
O: 1.04 (0.84, 1.29)
F: 1.25 (0.91, 1.71)
M: 0.89 (0.67, 1.17)
0.34 (-0.08, 0.76)
Forgiveness§
O: 1.01 (0.82, 1.26)
F: 0.93 (0.71, 1.21)
M: 1.10 (0.80, 1.52)
-0.18 (-0.58, 0.22)
Depressive symptoms§
O: 1.00 (0.84, 1.18)
F: 1.14 (0.91, 1.42)
M: 0.88 (0.69, 1.11)
0.26 (-0.06, 0.58)
Feeling capable§
O: 0.99 (0.83, 1.19)
F: 1.01 (0.77, 1.33)
M: 0.98 (0.76, 1.26)
0.03 (-0.35, 0.42)
Various aspects of life are balanced§
O: 0.99 (0.80, 1.21)
F: 0.93 (0.71, 1.21)
M: 1.04 (0.78, 1.40)
-0.12 (-0.50, 0.26)
Feel at peace§
O: 0.97 (0.78, 1.19)
F: 0.93 (0.71, 1.21)
M: 1.00 (0.75, 1.34)
-0.08 (-0.45, 0.29)
Say in government#
O: 0.85 (0.67, 1.07)
F: 0.87 (0.62, 1.21)
M: 0.83 (0.61, 1.13)
0.04 (-0.41, 0.50)
Government approval§
O: 0.79 (0.64, 0.97)**
F: 0.68 (0.52, 0.89)**
M: 0.90 (0.69, 1.17)
-0.27 (-0.61, 0.06)
Support reducing differences in income levels§
O: 0.71 (0.53, 0.96)**
F: 0.67 (0.47, 0.94)**
M: 0.76 (0.50, 1.15)
-0.13 (-0.62, 0.35)
Obeying the law§
O: 0.54 (0.39, 0.75)***
F: 0.55 (0.38, 0.80)**
M: 0.54 (0.35, 0.83)**
0.03 (-0.45, 0.51)
0.5
1.0
2.0
Cumulative odds ratio (x-axis on log scale)

**eFigure 17.** Adjusted associations between childhood predictors and 45 outcomes of interest and two flourishing composite indices for categorized residential location (Rural area or on a farm, small town or village, large city or suburb of a large city).

***Note:*** Estimates are from survey-weighted models fitted separately within each residence group, using the same outcome coding, covariates (excluding residential setting), and exposure-by-gender interaction as the primary analyses. Stratification reduces sample sizes, particularly for female respondents and for rare binary outcomes such as cigarette smoking and alcohol use. Where very few or no exposed participants in a subgroup reported the outcome, models could not estimate the association reliably. These estimates appear as odds ratios near 0 or extremely large, with implausibly very wide 95% CIs

**Predictor:** ***Childhood abuse***
***Childhood abuse – Rural area or on a farm***

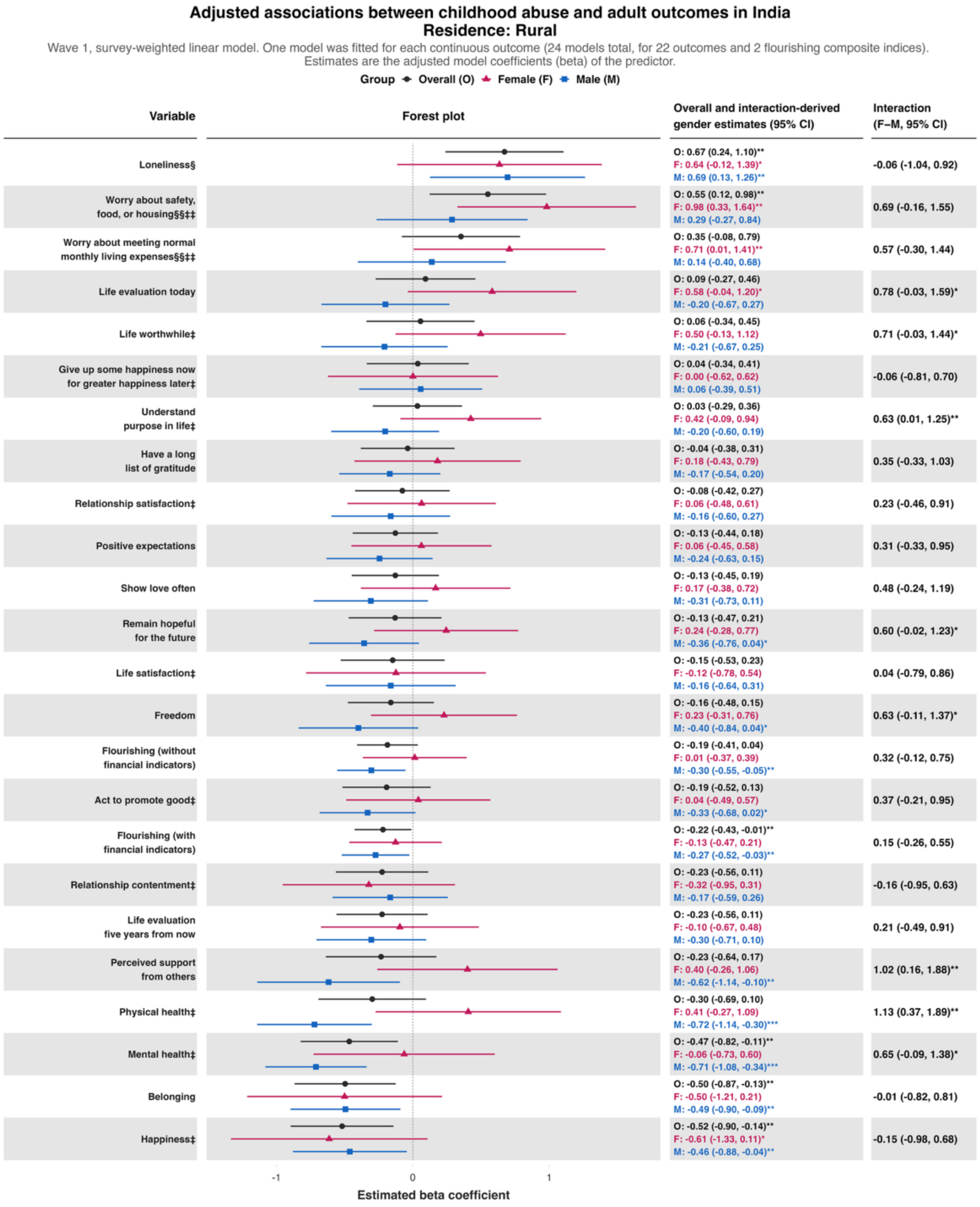

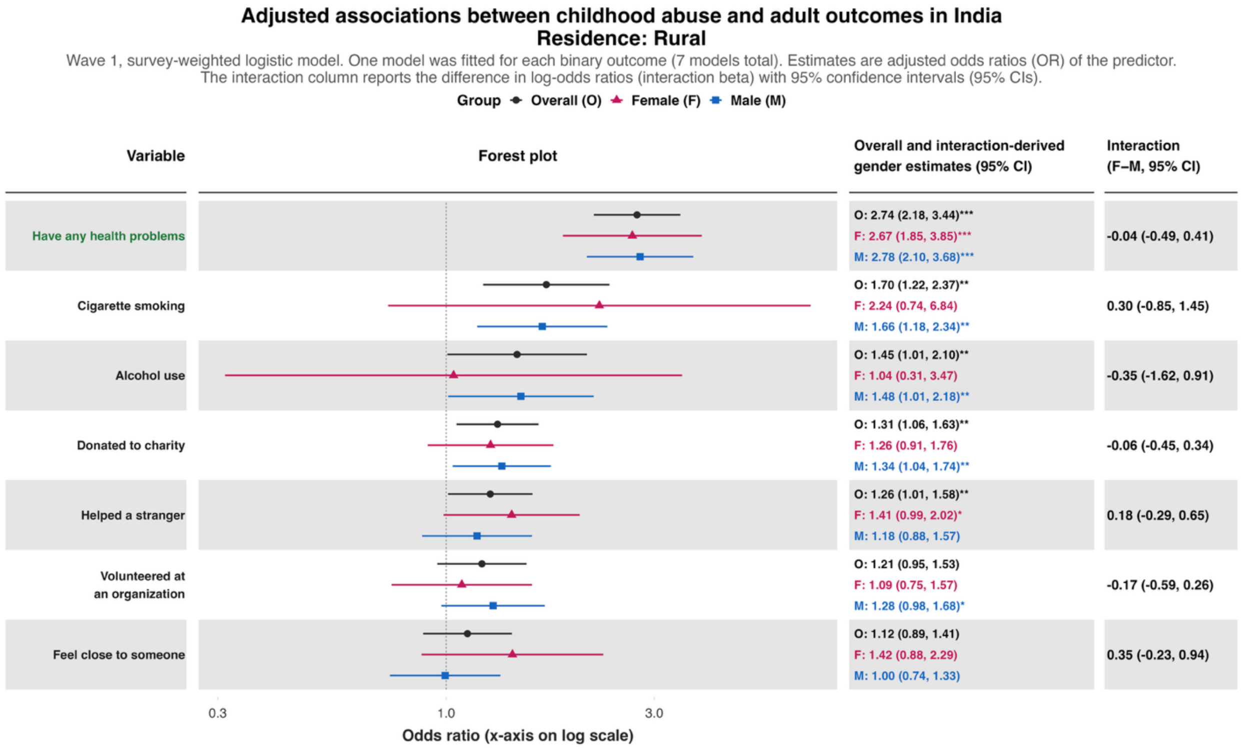
Adjusted associations between childhood abuse and adult outcomes in India
Residence: Rural
Wave 1, survey-weighted logistic model. One model was fitted for each binary outcome (7 models total). Estimates are adjusted odds ratios (OR) of the predictor.
The interaction column reports the difference in log-odds ratios (interaction beta) with 95% confidence intervals (95% CIs).
Group Overall (O) Female (F) Male (M)
Variable
Forest plot
Overall and interaction-derived gender estimates (95% CI)
Interaction (F–M, 95% CI)
Have any health problems
O: 2.74 (2.18, 3.44)***
F: 2.67 (1.85, 3.85)***
M: 2.78 (2.10, 3.68)***
-0.04 (-0.49, 0.41)
Cigarette smoking
O: 1.70 (1.22, 2.37)**
F: 2.24 (0.74, 6.84)
M: 1.66 (1.18, 2.34)**
0.30 (-0.85, 1.45)
Alcohol use
O: 1.45 (1.01, 2.10)**
F: 1.04 (0.31, 3.47)
M: 1.48 (1.01, 2.18)**
-0.35 (-1.62, 0.91)
Donated to charity
O: 1.31 (1.06, 1.63)**
F: 1.26 (0.91, 1.76)
M: 1.34 (1.04, 1.74)**
-0.06 (-0.45, 0.34)
Helped a stranger
O: 1.26 (1.01, 1.58)**
F: 1.41 (0.99, 2.02)*
M: 1.18 (0.88, 1.57)
0.18 (-0.29, 0.65)
Volunteered at an organization
O: 1.21 (0.95, 1.53)
F: 1.09 (0.75, 1.57)
M: 1.28 (0.98, 1.68)*
-0.17 (-0.59, 0.26)
Feel close to someone
O: 1.12 (0.89, 1.41)
F: 1.42 (0.88, 2.29)
M: 1.00 (0.74, 1.33)
0.35 (-0.23, 0.94)
0.3
1.0
3.0
Odds ratio (x-axis on log scale)

**Adjusted associations between childhood abuse and adult outcomes in India**
**Residence: Rural**

Wave 1, survey-weighted ordinal regression using the proportional odds model (`svyolr`). One model was fitted for each ordinal outcome (16 models total).
Estimates are adjusted cumulative odds ratios (cOR) of the predictor.
The interaction column reports the difference in log-cumulative odds ratios (interaction beta) with 95% confidence intervals (95% CIs).

Group: Overall (O), Female (F), Male (M)

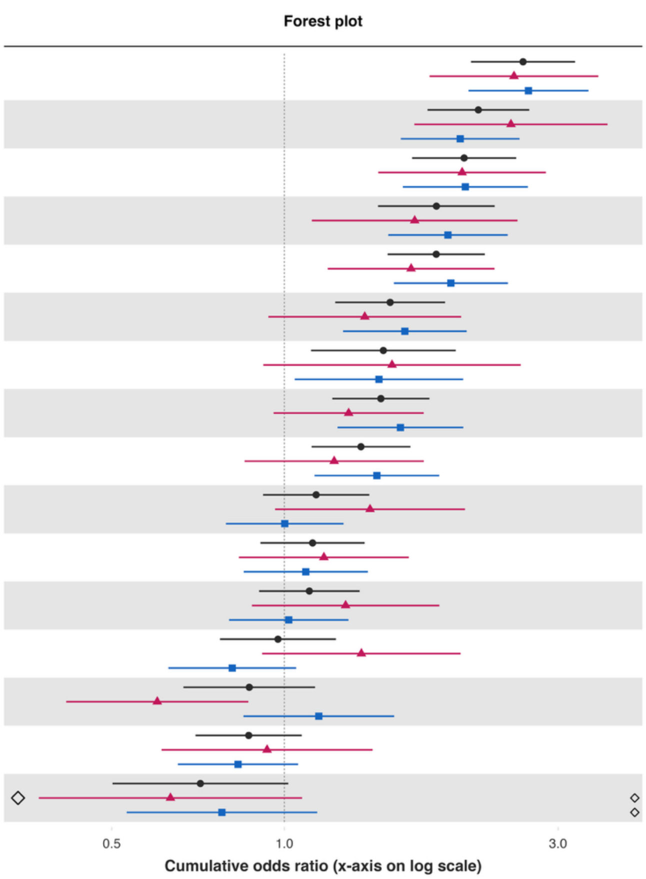


| Variable | Overall and interaction-derived gender estimates (95% CI) | Interaction (F–M, 95% CI) |
|---|---|---|
| Unable to control worry§ | O: 2.61 (2.11, 3.21)***<br>F: 2.51 (1.79, 3.53)***<br>M: 2.66 (2.09, 3.39)*** | -0.06 (-0.45, 0.33) |
| Perceived discrimination§ | O: 2.18 (1.78, 2.67)***<br>F: 2.48 (1.69, 3.66)***<br>M: 2.03 (1.60, 2.57)*** | 0.20 (-0.26, 0.66) |
| Feeling nervous or anxious§ | O: 2.06 (1.67, 2.54)***<br>F: 2.04 (1.46, 2.86)***<br>M: 2.07 (1.61, 2.66)*** | -0.01 (-0.42, 0.39) |
| Distress from life-threatening experiences§ | O: 1.84 (1.46, 2.33)***<br>F: 1.69 (1.12, 2.55)**<br>M: 1.93 (1.52, 2.45)*** | -0.13 (-0.57, 0.30) |
| Depressive symptoms§ | O: 1.84 (1.51, 2.24)***<br>F: 1.66 (1.19, 2.32)**<br>M: 1.95 (1.55, 2.45)*** | -0.16 (-0.56, 0.24) |
| Extent of suffering§ | O: 1.53 (1.23, 1.91)***<br>F: 1.38 (0.94, 2.03)<br>M: 1.62 (1.27, 2.08)*** | -0.16 (-0.60, 0.28) |
| Say in government# | O: 1.49 (1.11, 1.99)**<br>F: 1.54 (0.92, 2.58)<br>M: 1.46 (1.04, 2.05)** | 0.05 (-0.55, 0.65) |
| Have little interest in doing things§ | O: 1.47 (1.21, 1.79)***<br>F: 1.29 (0.96, 1.75)*<br>M: 1.59 (1.24, 2.05)*** | -0.21 (-0.60, 0.18) |
| Government approval§ | O: 1.36 (1.12, 1.66)**<br>F: 1.22 (0.85, 1.75)<br>M: 1.45 (1.13, 1.86)** | -0.17 (-0.63, 0.28) |
| Various aspects of life are balanced§ | O: 1.14 (0.92, 1.41)<br>F: 1.41 (0.96, 2.07)*<br>M: 1.00 (0.79, 1.27) | 0.34 (-0.09, 0.77) |
| Feel at peace§ | O: 1.12 (0.91, 1.38)<br>F: 1.17 (0.83, 1.65)<br>M: 1.09 (0.85, 1.40) | 0.07 (-0.33, 0.48) |
| Trust in others§ | O: 1.11 (0.90, 1.35)<br>F: 1.28 (0.88, 1.86)<br>M: 1.02 (0.80, 1.29) | 0.23 (-0.22, 0.68) |
| Forgiveness§ | O: 0.97 (0.77, 1.23)<br>F: 1.36 (0.91, 2.03)<br>M: 0.81 (0.63, 1.05) | 0.52 (0.08, 0.96)** |
| Support reducing differences in income levels§ | O: 0.87 (0.67, 1.13)<br>F: 0.60 (0.42, 0.87)**<br>M: 1.15 (0.85, 1.55) | -0.65 (-1.06, -0.24)** |
| Feeling capable§ | O: 0.87 (0.70, 1.07)<br>F: 0.93 (0.61, 1.42)<br>M: 0.83 (0.65, 1.06) | 0.12 (-0.38, 0.61) |
| Obeying the law§ | O: 0.71 (0.50, 1.02)*<br>F: 0.63 (0.37, 1.07)*<br>M: 0.78 (0.53, 1.14) | -0.21 (-0.78, 0.37) |

#### *Childhood abuse – Small town or village*

**Adjusted associations between childhood abuse and adult outcomes in India**
**Residence: Small town**

Wave 1, survey-weighted linear model. One model was fitted for each continuous outcome (24 models total, for 22 outcomes and 2 flourishing composite indices).
Estimates are the adjusted model coefficients (beta) of the predictor.

Group ◆ Overall (O) ▲ Female (F) ■ Male (M)

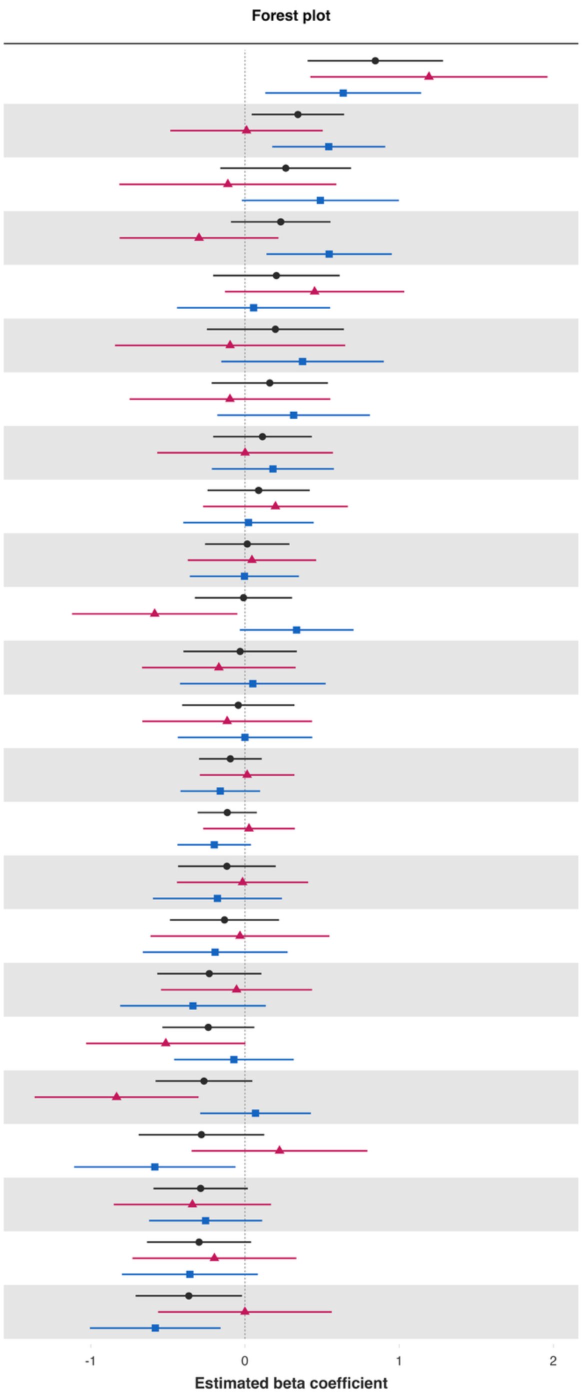


| Variable | Overall and interaction-derived gender estimates (95% CI) | Interaction (F–M, 95% CI) |
|---|---|---|
| Loneliness§ | O: 0.85 (0.41, 1.28)***<br>F: 1.19 (0.42, 1.96)**<br>M: 0.64 (0.13, 1.14)** | 0.56 (-0.34, 1.46) |
| Understand purpose in life‡ | O: 0.34 (0.04, 0.64)**<br>F: 0.01 (-0.48, 0.50)<br>M: 0.54 (0.18, 0.91)** | -0.53 (-1.14, 0.08)* |
| Worry about safety, food, or housing§§‡‡ | O: 0.26 (-0.16, 0.69)<br>F: -0.11 (-0.81, 0.59)<br>M: 0.49 (-0.02, 1.00)* | -0.60 (-1.45, 0.25) |
| Have a long list of gratitude | O: 0.23 (-0.09, 0.55)<br>F: -0.30 (-0.81, 0.22)<br>M: 0.55 (0.14, 0.95)** | -0.84 (-1.49, -0.19)** |
| Life evaluation today | O: 0.20 (-0.21, 0.61)<br>F: 0.45 (-0.13, 1.03)<br>M: 0.06 (-0.44, 0.55) | 0.40 (-0.30, 1.09) |
| Perceived support from others | O: 0.20 (-0.25, 0.64)<br>F: -0.10 (-0.84, 0.65)<br>M: 0.37 (-0.15, 0.90) | -0.47 (-1.36, 0.42) |
| Worry about meeting normal monthly living expenses§§‡‡ | O: 0.16 (-0.22, 0.54)<br>F: -0.10 (-0.75, 0.55)<br>M: 0.32 (-0.18, 0.81) | -0.41 (-1.26, 0.44) |
| Show love often | O: 0.11 (-0.21, 0.43)<br>F: 0.00 (-0.57, 0.57)<br>M: 0.18 (-0.21, 0.58) | -0.18 (-0.89, 0.53) |
| Give up some happiness now for greater happiness later‡ | O: 0.09 (-0.24, 0.42)<br>F: 0.20 (-0.27, 0.67)<br>M: 0.02 (-0.40, 0.45) | 0.17 (-0.43, 0.78) |
| Act to promote good‡ | O: 0.01 (-0.26, 0.29)<br>F: 0.05 (-0.37, 0.46)<br>M: -0.00 (-0.36, 0.35) | 0.05 (-0.49, 0.59) |
| Freedom | O: -0.01 (-0.33, 0.31)<br>F: -0.59 (-1.12, -0.05)**<br>M: 0.34 (-0.03, 0.70)* | -0.92 (-1.57, -0.28)** |
| Relationship contentment‡ | O: -0.03 (-0.40, 0.34)<br>F: -0.17 (-0.67, 0.33)<br>M: 0.05 (-0.42, 0.52) | -0.22 (-0.87, 0.43) |
| Life evaluation five years from now | O: -0.04 (-0.41, 0.32)<br>F: -0.12 (-0.67, 0.43)<br>M: 0.00 (-0.44, 0.44) | -0.12 (-0.77, 0.54) |
| Flourishing (without financial indicators) | O: -0.09 (-0.30, 0.11)<br>F: 0.01 (-0.29, 0.32)<br>M: -0.16 (-0.42, 0.10) | 0.17 (-0.22, 0.57) |
| Flourishing (with financial indicators) | O: -0.11 (-0.31, 0.08)<br>F: 0.03 (-0.27, 0.32)<br>M: -0.20 (-0.44, 0.04) | 0.23 (-0.14, 0.59) |
| Relationship satisfaction‡ | O: -0.12 (-0.43, 0.20)<br>F: -0.02 (-0.44, 0.41)<br>M: -0.18 (-0.60, 0.24) | 0.16 (-0.41, 0.74) |
| Life satisfaction‡ | O: -0.13 (-0.49, 0.22)<br>F: -0.03 (-0.61, 0.55)<br>M: -0.19 (-0.66, 0.28) | 0.16 (-0.61, 0.93) |
| Life worthwhile‡ | O: -0.23 (-0.57, 0.11)<br>F: -0.05 (-0.54, 0.43)<br>M: -0.34 (-0.81, 0.14) | 0.28 (-0.42, 0.98) |
| Positive expectations | O: -0.24 (-0.54, 0.06)<br>F: -0.51 (-1.03, 0.00)*<br>M: -0.07 (-0.46, 0.32) | -0.44 (-1.11, 0.23) |
| Remain hopeful for the future | O: -0.27 (-0.58, 0.05)*<br>F: -0.83 (-1.36, -0.30)**<br>M: 0.07 (-0.29, 0.43) | -0.90 (-1.53, -0.28)** |
| Happiness‡ | O: -0.28 (-0.69, 0.12)<br>F: 0.22 (-0.35, 0.79)<br>M: -0.58 (-1.11, -0.06)** | 0.81 (0.06, 1.56)** |
| Belonging | O: -0.29 (-0.59, 0.02)*<br>F: -0.34 (-0.85, 0.17)<br>M: -0.26 (-0.62, 0.11) | -0.09 (-0.70, 0.53) |
| Mental health‡ | O: -0.30 (-0.64, 0.04)*<br>F: -0.20 (-0.73, 0.33)<br>M: -0.36 (-0.80, 0.08) | 0.16 (-0.54, 0.85) |
| Physical health‡ | O: -0.36 (-0.71, -0.02)**<br>F: -0.00 (-0.56, 0.56)<br>M: -0.58 (-1.01, -0.16)** | 0.58 (-0.11, 1.28) |

**Adjusted associations between childhood abuse and adult outcomes in India**
**Residence: Small town**

Wave 1, survey-weighted logistic model. One model was fitted for each binary outcome (7 models total). Estimates are adjusted odds ratios (OR) of the predictor.
The interaction column reports the difference in log-odds ratios (interaction beta) with 95% confidence intervals (95% CIs).

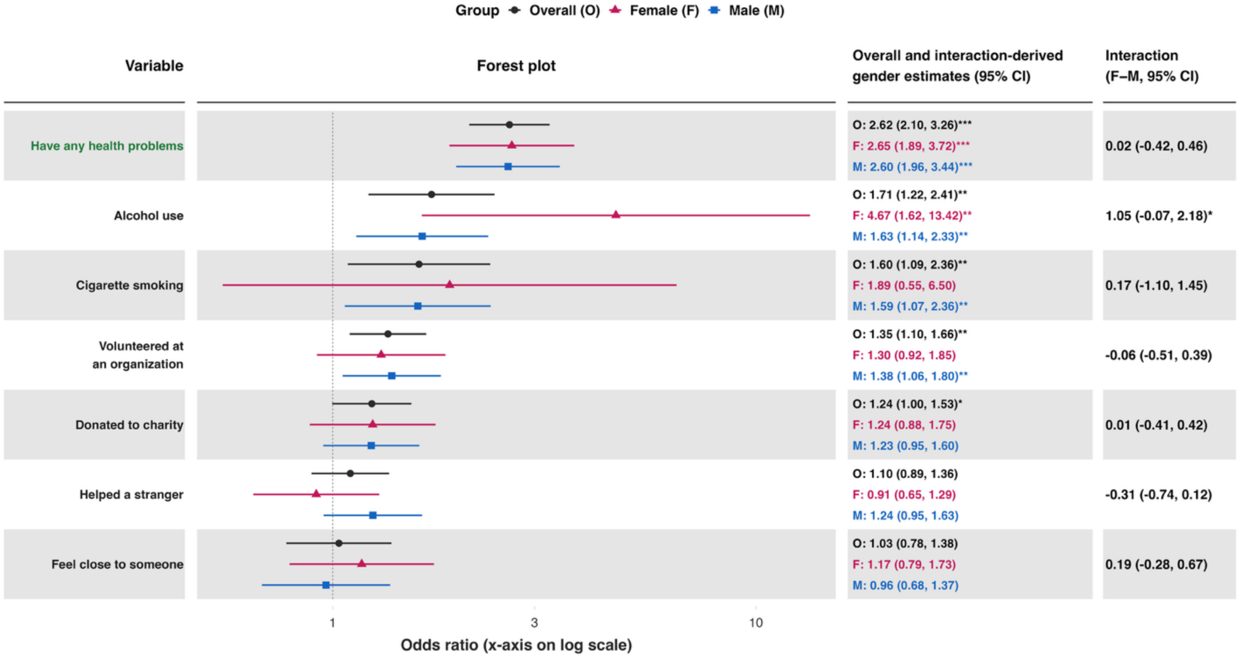

### Adjusted associations between childhood abuse and adult outcomes in India
### Residence: Small town

Wave 1, survey-weighted ordinal regression using the proportional odds model (`svyolr`). One model was fitted for each ordinal outcome (16 models total).
Estimates are adjusted cumulative odds ratios (cOR) of the predictor.
The interaction column reports the difference in log-cumulative odds ratios (interaction beta) with 95% confidence intervals (95% CIs).

Group ◆ Overall (O) ▲ Female (F) ■ Male (M)

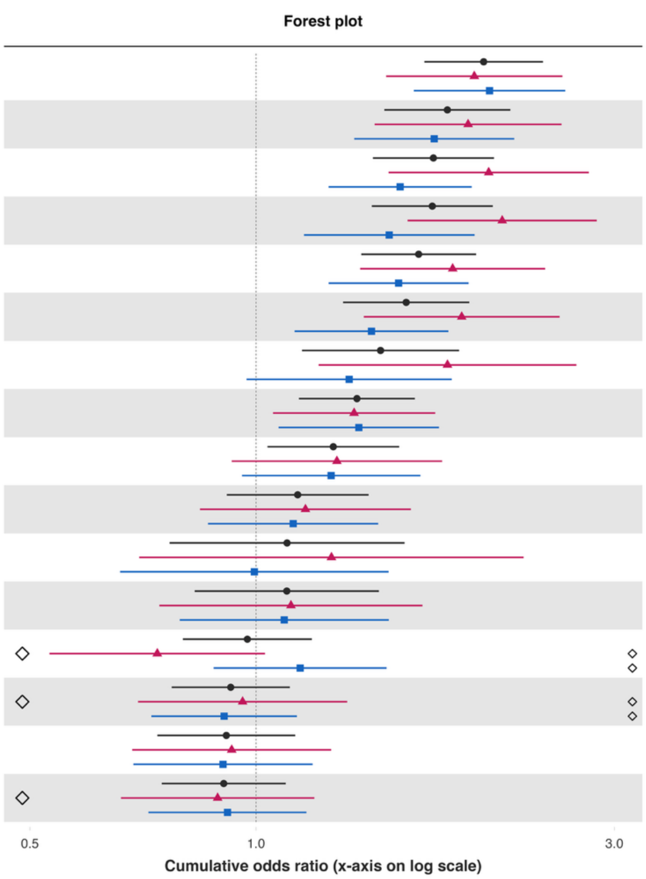


| Variable | Overall and interaction-derived gender estimates (95% CI) | Interaction (F–M, 95% CI) |
|---|---|---|
| Distress from life-threatening experiences§ | O: 2.01 (1.68, 2.41)***<br>F: 1.95 (1.49, 2.56)***<br>M: 2.05 (1.62, 2.58)*** | -0.05 (-0.39, 0.30) |
| Unable to control worry§ | O: 1.80 (1.48, 2.18)***<br>F: 1.92 (1.44, 2.55)***<br>M: 1.73 (1.35, 2.21)*** | 0.10 (-0.26, 0.47) |
| Depressive symptoms§ | O: 1.72 (1.43, 2.07)***<br>F: 2.04 (1.50, 2.77)***<br>M: 1.56 (1.25, 1.94)*** | 0.27 (-0.10, 0.64) |
| Perceived discrimination§ | O: 1.72 (1.43, 2.07)***<br>F: 2.13 (1.59, 2.84)***<br>M: 1.50 (1.16, 1.95)** | 0.35 (-0.07, 0.76)* |
| Extent of suffering§ | O: 1.65 (1.38, 1.96)***<br>F: 1.83 (1.38, 2.43)***<br>M: 1.55 (1.25, 1.92)*** | 0.17 (-0.18, 0.51) |
| Feeling nervous or anxious§ | O: 1.58 (1.31, 1.92)***<br>F: 1.88 (1.39, 2.54)***<br>M: 1.42 (1.13, 1.80)** | 0.28 (-0.09, 0.64) |
| Say in government# | O: 1.46 (1.15, 1.86)**<br>F: 1.80 (1.21, 2.67)**<br>M: 1.33 (0.97, 1.82)* | 0.30 (-0.22, 0.82) |
| Have little interest in doing things§ | O: 1.36 (1.14, 1.63)***<br>F: 1.35 (1.05, 1.73)**<br>M: 1.37 (1.07, 1.75)** | -0.01 (-0.36, 0.34) |
| Government approval§ | O: 1.27 (1.04, 1.55)**<br>F: 1.28 (0.93, 1.77)<br>M: 1.26 (0.96, 1.65)* | 0.02 (-0.42, 0.46) |
| Various aspects of life are balanced§ | O: 1.14 (0.91, 1.41)<br>F: 1.16 (0.84, 1.61)<br>M: 1.12 (0.86, 1.45) | 0.04 (-0.35, 0.42) |
| Obeying the law§ | O: 1.10 (0.77, 1.58)<br>F: 1.26 (0.70, 2.27)<br>M: 0.99 (0.66, 1.50) | 0.24 (-0.44, 0.91) |
| Support reducing differences in income levels§ | O: 1.10 (0.83, 1.46)<br>F: 1.11 (0.74, 1.67)<br>M: 1.09 (0.79, 1.50) | 0.02 (-0.42, 0.46) |
| Forgiveness§ | O: 0.97 (0.80, 1.19)<br>F: 0.74 (0.53, 1.03)*<br>M: 1.14 (0.88, 1.49) | -0.44 (-0.88, 0.01)* |
| Trust in others§ | O: 0.93 (0.77, 1.11)<br>F: 0.96 (0.70, 1.32)<br>M: 0.91 (0.73, 1.13) | 0.06 (-0.34, 0.45) |
| Feeling capable§ | O: 0.91 (0.74, 1.13)<br>F: 0.93 (0.68, 1.26)<br>M: 0.90 (0.69, 1.19) | 0.03 (-0.37, 0.43) |
| Feel at peace§ | O: 0.91 (0.75, 1.10)<br>F: 0.89 (0.66, 1.20)<br>M: 0.92 (0.72, 1.17) | -0.03 (-0.41, 0.35) |

### *Childhood abuse – Large city or suburb of a large city*

**Adjusted associations between childhood abuse and adult outcomes in India**
**Residence: Large city or suburb**

Wave 1, survey-weighted linear model. One model was fitted for each continuous outcome (24 models total, for 22 outcomes and 2 flourishing composite indices).
Estimates are the adjusted model coefficients (beta) of the predictor.

**Group** ● Overall (O) ▲ Female (F) ■ Male (M)

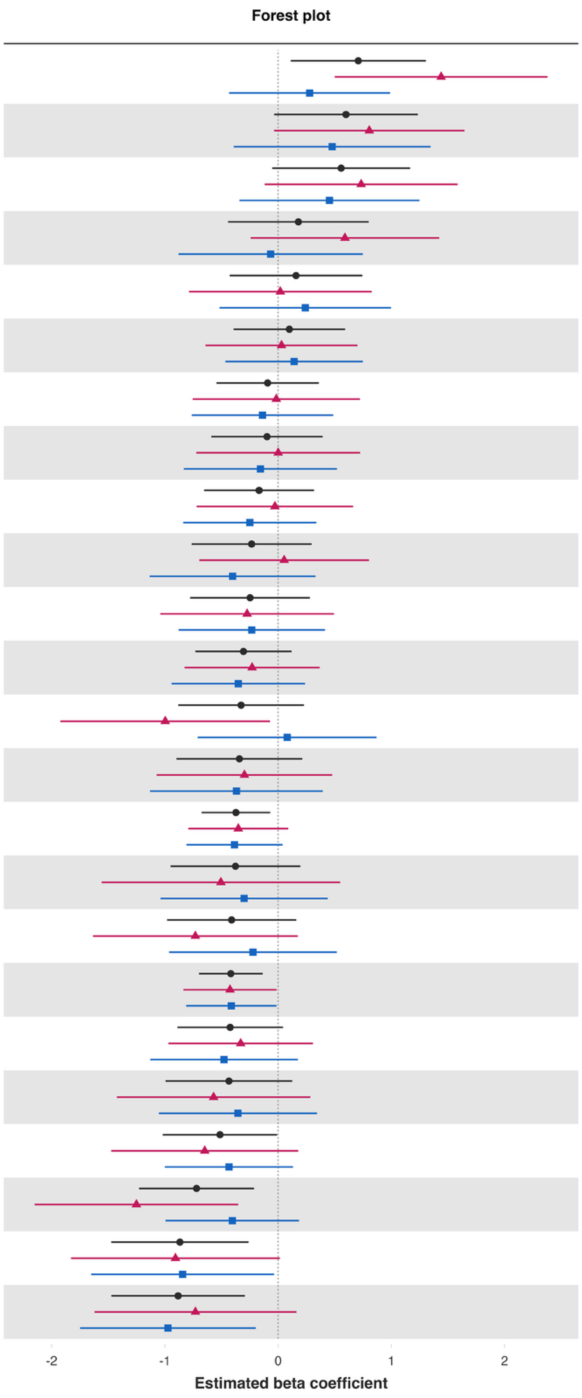


| Variable | Overall and interaction-derived gender estimates (95% CI) | Interaction (F–M, 95% CI) |
|---|---|---|
| Loneliness§ | O: 0.71 (0.11, 1.30)**<br>F: 1.44 (0.50, 2.38)**<br>M: 0.28 (-0.43, 0.99) | 1.16 (0.03, 2.30)** |
| Worry about meeting normal monthly living expenses§§‡‡ | O: 0.60 (-0.04, 1.23)*<br>F: 0.80 (-0.04, 1.65)*<br>M: 0.48 (-0.39, 1.35) | 0.33 (-0.88, 1.53) |
| Worry about safety, food, or housing§§‡‡ | O: 0.56 (-0.05, 1.17)*<br>F: 0.73 (-0.12, 1.59)*<br>M: 0.45 (-0.34, 1.25) | 0.28 (-0.85, 1.41) |
| Give up some happiness now for greater happiness later‡ | O: 0.18 (-0.44, 0.80)<br>F: 0.59 (-0.24, 1.42)<br>M: -0.07 (-0.88, 0.75) | 0.66 (-0.45, 1.77) |
| Life evaluation today | O: 0.16 (-0.43, 0.74)<br>F: 0.02 (-0.79, 0.83)<br>M: 0.24 (-0.52, 1.00) | -0.22 (-1.28, 0.84) |
| Have a long list of gratitude | O: 0.10 (-0.39, 0.59)<br>F: 0.03 (-0.64, 0.70)<br>M: 0.14 (-0.47, 0.75) | -0.11 (-0.93, 0.71) |
| Physical health‡ | O: -0.09 (-0.54, 0.36)<br>F: -0.02 (-0.75, 0.72)<br>M: -0.14 (-0.76, 0.49) | 0.12 (-0.90, 1.14) |
| Relationship satisfaction‡ | O: -0.10 (-0.59, 0.39)<br>F: 0.00 (-0.72, 0.72)<br>M: -0.16 (-0.83, 0.52) | 0.16 (-0.86, 1.17) |
| Freedom | O: -0.17 (-0.65, 0.32)<br>F: -0.03 (-0.72, 0.66)<br>M: -0.25 (-0.84, 0.34) | 0.22 (-0.60, 1.05) |
| Life worthwhile‡ | O: -0.23 (-0.76, 0.30)<br>F: 0.05 (-0.70, 0.80)<br>M: -0.40 (-1.14, 0.33) | 0.46 (-0.62, 1.53) |
| Remain hopeful for the future | O: -0.25 (-0.78, 0.28)<br>F: -0.27 (-1.04, 0.49)<br>M: -0.23 (-0.88, 0.41) | -0.04 (-0.97, 0.89) |
| Act to promote good‡ | O: -0.31 (-0.73, 0.12)<br>F: -0.23 (-0.83, 0.37)<br>M: -0.35 (-0.94, 0.24) | 0.12 (-0.73, 0.98) |
| Life evaluation five years from now | O: -0.33 (-0.88, 0.23)<br>F: -1.00 (-1.92, -0.07)**<br>M: 0.08 (-0.71, 0.87) | -1.08 (-2.40, 0.25) |
| Relationship contentment‡ | O: -0.34 (-0.90, 0.21)<br>F: -0.30 (-1.07, 0.48)<br>M: -0.37 (-1.13, 0.39) | 0.07 (-1.03, 1.17) |
| Flourishing (without financial indicators) | O: -0.37 (-0.68, -0.07)**<br>F: -0.35 (-0.79, 0.09)<br>M: -0.39 (-0.81, 0.04)* | 0.03 (-0.60, 0.67) |
| Perceived support from others | O: -0.38 (-0.95, 0.20)<br>F: -0.51 (-1.56, 0.55)<br>M: -0.30 (-1.04, 0.44) | -0.20 (-1.56, 1.15) |
| Show love often | O: -0.41 (-0.98, 0.16)<br>F: -0.73 (-1.64, 0.17)<br>M: -0.22 (-0.96, 0.52) | -0.51 (-1.69, 0.67) |
| Flourishing (with financial indicators) | O: -0.42 (-0.70, -0.14)**<br>F: -0.43 (-0.84, -0.01)**<br>M: -0.41 (-0.81, -0.01)** | -0.01 (-0.61, 0.59) |
| Positive expectations | O: -0.42 (-0.89, 0.04)*<br>F: -0.33 (-0.97, 0.31)<br>M: -0.48 (-1.13, 0.17) | 0.15 (-0.78, 1.07) |
| Understand purpose in life‡ | O: -0.43 (-0.99, 0.13)<br>F: -0.57 (-1.42, 0.28)<br>M: -0.36 (-1.05, 0.34) | -0.22 (-1.28, 0.85) |
| Belonging | O: -0.51 (-1.02, -0.01)**<br>F: -0.65 (-1.47, 0.18)<br>M: -0.43 (-1.00, 0.13) | -0.21 (-1.14, 0.71) |
| Mental health‡ | O: -0.72 (-1.23, -0.21)**<br>F: -1.25 (-2.15, -0.35)**<br>M: -0.41 (-0.99, 0.18) | -0.85 (-1.91, 0.22) |
| Life satisfaction‡ | O: -0.87 (-1.47, -0.26)**<br>F: -0.91 (-1.83, 0.02)*<br>M: -0.84 (-1.65, -0.04)** | -0.06 (-1.30, 1.18) |
| Happiness‡ | O: -0.88 (-1.47, -0.29)**<br>F: -0.73 (-1.62, 0.16)<br>M: -0.97 (-1.75, -0.20)** | 0.24 (-0.94, 1.42) |

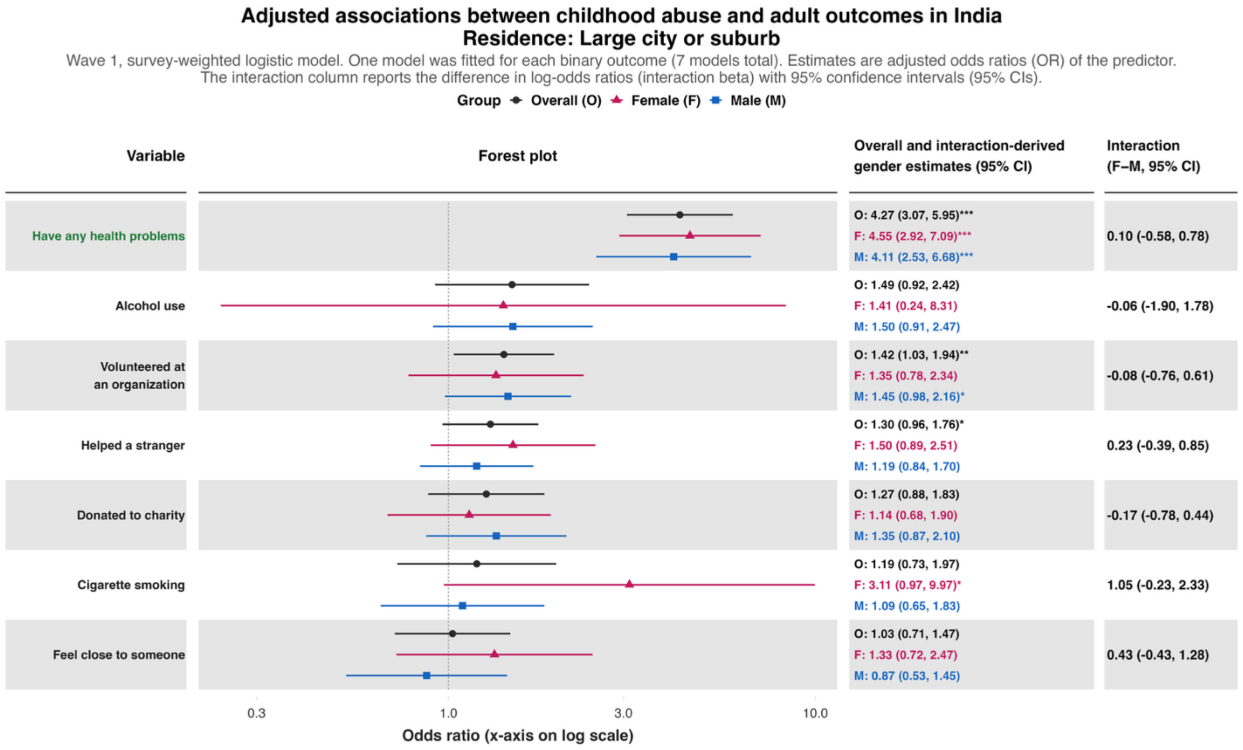
Adjusted associations between childhood abuse and adult outcomes in India
Residence: Large city or suburb
Wave 1, survey-weighted logistic model. One model was fitted for each binary outcome (7 models total). Estimates are adjusted odds ratios (OR) of the predictor.
The interaction column reports the difference in log-odds ratios (interaction beta) with 95% confidence intervals (95% CIs).
Group Overall (O) Female (F) Male (M)
Variable
Forest plot
Overall and interaction-derived gender estimates (95% CI)
Interaction (F–M, 95% CI)
Have any health problems
O: 4.27 (3.07, 5.95)***
F: 4.55 (2.92, 7.09)***
M: 4.11 (2.53, 6.68)***
0.10 (-0.58, 0.78)
Alcohol use
O: 1.49 (0.92, 2.42)
F: 1.41 (0.24, 8.31)
M: 1.50 (0.91, 2.47)
-0.06 (-1.90, 1.78)
Volunteered at an organization
O: 1.42 (1.03, 1.94)**
F: 1.35 (0.78, 2.34)
M: 1.45 (0.98, 2.16)*
-0.08 (-0.76, 0.61)
Helped a stranger
O: 1.30 (0.96, 1.76)*
F: 1.50 (0.89, 2.51)
M: 1.19 (0.84, 1.70)
0.23 (-0.39, 0.85)
Donated to charity
O: 1.27 (0.88, 1.83)
F: 1.14 (0.68, 1.90)
M: 1.35 (0.87, 2.10)
-0.17 (-0.78, 0.44)
Cigarette smoking
O: 1.19 (0.73, 1.97)
F: 3.11 (0.97, 9.97)*
M: 1.09 (0.65, 1.83)
1.05 (-0.23, 2.33)
Feel close to someone
O: 1.03 (0.71, 1.47)
F: 1.33 (0.72, 2.47)
M: 0.87 (0.53, 1.45)
0.43 (-0.43, 1.28)
0.3
1.0
3.0
10.0
Odds ratio (x-axis on log scale)

### Adjusted associations between childhood abuse and adult outcomes in India
### Residence: Large city or suburb

Wave 1, survey-weighted ordinal regression using the proportional odds model (`svyolr`). One model was fitted for each ordinal outcome (16 models total).
Estimates are adjusted cumulative odds ratios (cOR) of the predictor.
The interaction column reports the difference in log-cumulative odds ratios (interaction beta) with 95% confidence intervals (95% CIs).

Group: Overall (O), Female (F), Male (M)

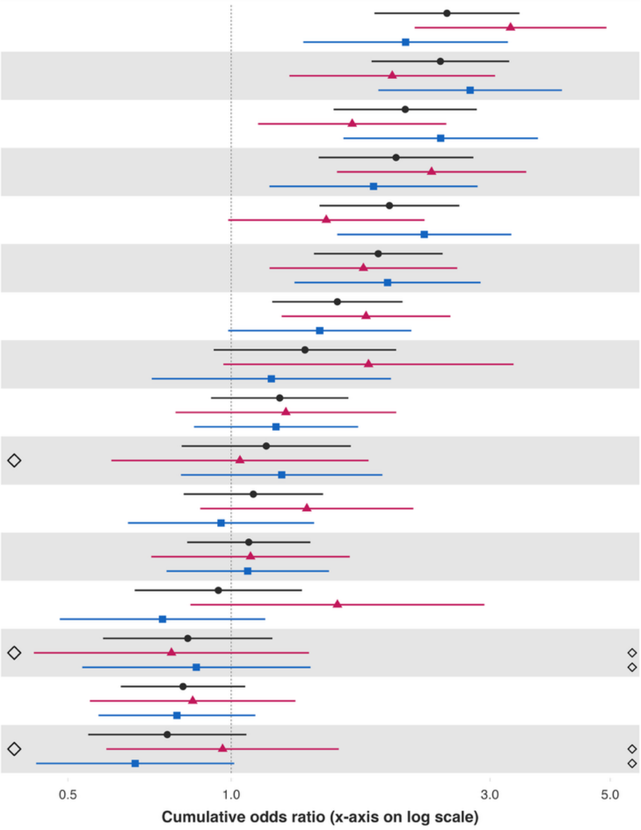


| Variable | Overall and interaction-derived gender estimates (95% CI) | Interaction (F–M, 95% CI) |
|---|---|---|
| Distress from life-threatening experiences§ | O: 2.50 (1.84, 3.40)***<br>F: 3.27 (2.18, 4.92)***<br>M: 2.10 (1.36, 3.24)*** | 0.45 (-0.16, 1.05) |
| Unable to control worry§ | O: 2.43 (1.81, 3.26)***<br>F: 1.98 (1.28, 3.07)**<br>M: 2.76 (1.87, 4.07)*** | -0.33 (-0.92, 0.26) |
| Feeling nervous or anxious§ | O: 2.09 (1.54, 2.84)***<br>F: 1.67 (1.12, 2.49)**<br>M: 2.43 (1.61, 3.68)*** | -0.38 (-0.93, 0.18) |
| Extent of suffering§ | O: 2.01 (1.45, 2.80)***<br>F: 2.34 (1.57, 3.50)***<br>M: 1.83 (1.18, 2.85)** | 0.25 (-0.31, 0.81) |
| Perceived discrimination§ | O: 1.96 (1.46, 2.63)***<br>F: 1.50 (0.99, 2.27)*<br>M: 2.27 (1.57, 3.28)*** | -0.42 (-0.93, 0.10) |
| Depressive symptoms§ | O: 1.87 (1.42, 2.45)***<br>F: 1.75 (1.18, 2.61)**<br>M: 1.94 (1.31, 2.88)*** | -0.10 (-0.69, 0.49) |
| Have little interest in doing things§ | O: 1.57 (1.19, 2.07)**<br>F: 1.77 (1.24, 2.54)**<br>M: 1.46 (0.99, 2.15)* | 0.20 (-0.34, 0.73) |
| Say in government# | O: 1.37 (0.93, 2.01)<br>F: 1.79 (0.97, 3.32)*<br>M: 1.19 (0.71, 1.97) | 0.41 (-0.39, 1.22) |
| Trust in others§ | O: 1.23 (0.92, 1.64)<br>F: 1.26 (0.79, 2.02)<br>M: 1.21 (0.85, 1.71) | 0.04 (-0.52, 0.60) |
| Government approval§ | O: 1.16 (0.81, 1.66)<br>F: 1.04 (0.60, 1.79)<br>M: 1.24 (0.81, 1.90) | -0.18 (-0.82, 0.47) |
| Various aspects of life are balanced§ | O: 1.10 (0.82, 1.48)<br>F: 1.38 (0.88, 2.17)<br>M: 0.96 (0.65, 1.42) | 0.36 (-0.24, 0.97) |
| Feeling capable§ | O: 1.08 (0.83, 1.40)<br>F: 1.09 (0.71, 1.65)<br>M: 1.07 (0.76, 1.51) | 0.01 (-0.54, 0.57) |
| Support reducing differences in income levels§ | O: 0.95 (0.66, 1.35)<br>F: 1.57 (0.84, 2.93)<br>M: 0.75 (0.48, 1.16) | 0.74 (-0.02, 1.51)* |
| Obeying the law§ | O: 0.83 (0.58, 1.19)<br>F: 0.78 (0.43, 1.39)<br>M: 0.86 (0.53, 1.40) | -0.11 (-0.90, 0.69) |
| Feel at peace§ | O: 0.82 (0.63, 1.06)<br>F: 0.85 (0.55, 1.31)<br>M: 0.79 (0.57, 1.11) | 0.07 (-0.48, 0.62) |
| Forgiveness§ | O: 0.76 (0.54, 1.07)<br>F: 0.96 (0.59, 1.58)<br>M: 0.67 (0.44, 1.01)* | 0.37 (-0.24, 0.99) |

**Predictor:** ***Feeling like an outsider in one's family when growing up***

***Feeling like an outsider in one's family when growing up – Rural area or on a farm***

#### Adjusted associations between feeling like an outsider in one's family when growing up and adult outcomes in India
#### Residence: Rural

Wave 1, survey-weighted linear model. One model was fitted for each continuous outcome (24 models total, for 22 outcomes and 2 flourishing composite indices). Estimates are the adjusted model coefficients (beta) of the predictor.

Group ◆ Overall (O) ▲ Female (F) ■ Male (M)

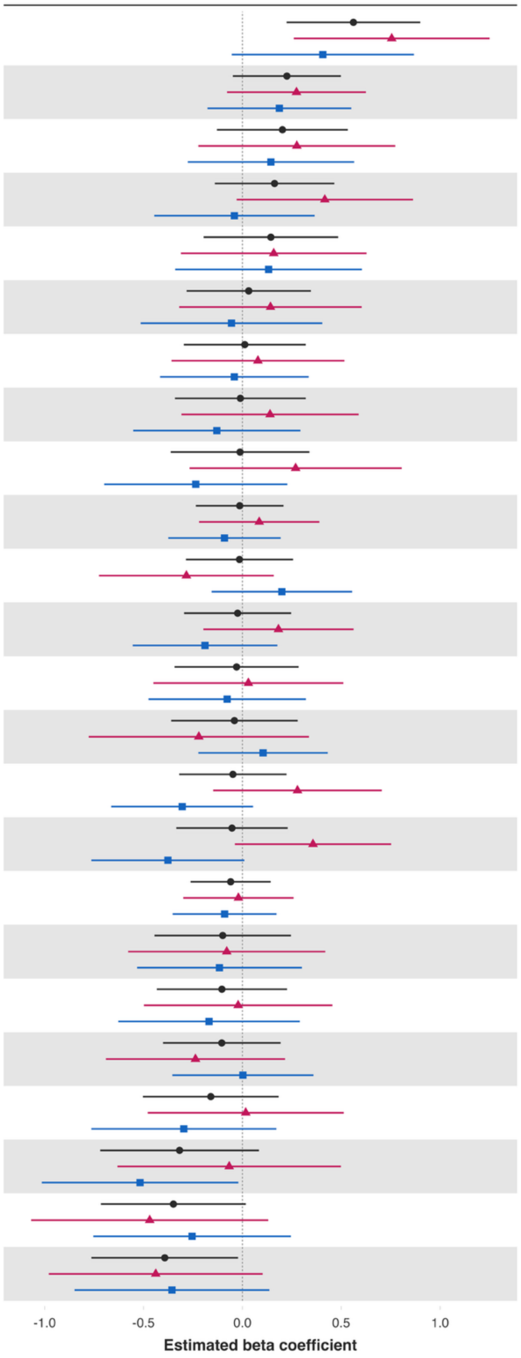


| Variable | Overall and interaction-derived gender estimates (95% CI) | Interaction (F–M, 95% CI) |
|---|---|---|
| Worry about safety, food, or housing§§‡‡ | O: 0.56 (0.22, 0.90)**<br>F: 0.76 (0.26, 1.25)**<br>M: 0.41 (-0.05, 0.87)* | 0.35 (-0.33, 1.02) |
| Act to promote good‡ | O: 0.23 (-0.05, 0.50)<br>F: 0.27 (-0.08, 0.62)<br>M: 0.19 (-0.18, 0.55) | 0.09 (-0.38, 0.55) |
| Loneliness§ | O: 0.20 (-0.13, 0.53)<br>F: 0.28 (-0.22, 0.77)<br>M: 0.14 (-0.28, 0.56) | 0.13 (-0.50, 0.76) |
| Have a long list of gratitude | O: 0.16 (-0.14, 0.47)<br>F: 0.42 (-0.03, 0.86)*<br>M: -0.04 (-0.45, 0.37) | 0.46 (-0.14, 1.06) |
| Life satisfaction‡ | O: 0.14 (-0.20, 0.48)<br>F: 0.16 (-0.31, 0.63)<br>M: 0.13 (-0.34, 0.61) | 0.03 (-0.63, 0.68) |
| Life worthwhile‡ | O: 0.03 (-0.28, 0.35)<br>F: 0.14 (-0.32, 0.60)<br>M: -0.06 (-0.51, 0.40) | 0.20 (-0.48, 0.88) |
| Relationship satisfaction‡ | O: 0.01 (-0.30, 0.32)<br>F: 0.08 (-0.36, 0.52)<br>M: -0.04 (-0.42, 0.34) | 0.12 (-0.41, 0.65) |
| Mental health‡ | O: -0.01 (-0.34, 0.32)<br>F: 0.14 (-0.31, 0.59)<br>M: -0.13 (-0.55, 0.29) | 0.27 (-0.30, 0.84) |
| Worry about meeting normal monthly living expenses§§‡‡ | O: -0.01 (-0.36, 0.34)<br>F: 0.27 (-0.27, 0.81)<br>M: -0.24 (-0.70, 0.23) | 0.51 (-0.20, 1.21) |
| Flourishing (without financial indicators) | O: -0.01 (-0.24, 0.21)<br>F: 0.09 (-0.22, 0.39)<br>M: -0.09 (-0.37, 0.19) | 0.18 (-0.21, 0.56) |
| Show love often | O: -0.01 (-0.29, 0.26)<br>F: -0.28 (-0.72, 0.16)<br>M: 0.20 (-0.16, 0.55) | -0.48 (-1.06, 0.10) |
| Remain hopeful for the future | O: -0.02 (-0.29, 0.25)<br>F: 0.18 (-0.20, 0.56)<br>M: -0.19 (-0.55, 0.18) | 0.37 (-0.14, 0.88) |
| Relationship contentment‡ | O: -0.03 (-0.34, 0.28)<br>F: 0.03 (-0.45, 0.51)<br>M: -0.08 (-0.47, 0.32) | 0.11 (-0.50, 0.72) |
| Belonging | O: -0.04 (-0.36, 0.28)<br>F: -0.22 (-0.78, 0.34)<br>M: 0.10 (-0.22, 0.43) | -0.32 (-0.95, 0.30) |
| Understand purpose in life‡ | O: -0.05 (-0.32, 0.22)<br>F: 0.28 (-0.15, 0.71)<br>M: -0.30 (-0.66, 0.05)* | 0.58 (0.02, 1.14)** |
| Freedom | O: -0.05 (-0.33, 0.23)<br>F: 0.36 (-0.04, 0.75)*<br>M: -0.38 (-0.76, 0.01)* | 0.73 (0.18, 1.29)** |
| Flourishing (with financial indicators) | O: -0.06 (-0.26, 0.14)<br>F: -0.02 (-0.30, 0.26)<br>M: -0.09 (-0.35, 0.17) | 0.07 (-0.29, 0.43) |
| Give up some happiness now for greater happiness later‡ | O: -0.10 (-0.44, 0.25)<br>F: -0.08 (-0.58, 0.42)<br>M: -0.12 (-0.53, 0.30) | 0.04 (-0.56, 0.63) |
| Physical health‡ | O: -0.10 (-0.43, 0.23)<br>F: -0.02 (-0.50, 0.45)<br>M: -0.17 (-0.63, 0.29) | 0.15 (-0.52, 0.81) |
| Positive expectations | O: -0.10 (-0.40, 0.19)<br>F: -0.24 (-0.69, 0.22)<br>M: 0.00 (-0.35, 0.36) | -0.24 (-0.79, 0.31) |
| Life evaluation five years from now | O: -0.16 (-0.50, 0.18)<br>F: 0.02 (-0.48, 0.51)<br>M: -0.30 (-0.76, 0.17) | 0.31 (-0.36, 0.99) |
| Happiness‡ | O: -0.32 (-0.72, 0.08)<br>F: -0.07 (-0.63, 0.50)<br>M: -0.52 (-1.01, -0.02)** | 0.45 (-0.24, 1.14) |
| Life evaluation today | O: -0.35 (-0.72, 0.02)*<br>F: -0.47 (-1.07, 0.13)<br>M: -0.25 (-0.75, 0.25) | -0.21 (-1.03, 0.60) |
| Perceived support from others | O: -0.39 (-0.76, -0.02)**<br>F: -0.44 (-0.98, 0.10)<br>M: -0.36 (-0.85, 0.14) | -0.08 (-0.80, 0.64) |

**Adjusted associations between feeling like an outsider in one's family when growing up and adult outcomes in India**
**Residence: Rural**

Wave 1, survey-weighted logistic model. One model was fitted for each binary outcome (7 models total). Estimates are adjusted odds ratios (OR) of the predictor.
The interaction column reports the difference in log-odds ratios (interaction beta) with 95% confidence intervals (95% CIs).

Group • Overall (O) ▲ Female (F) ■ Male (M)

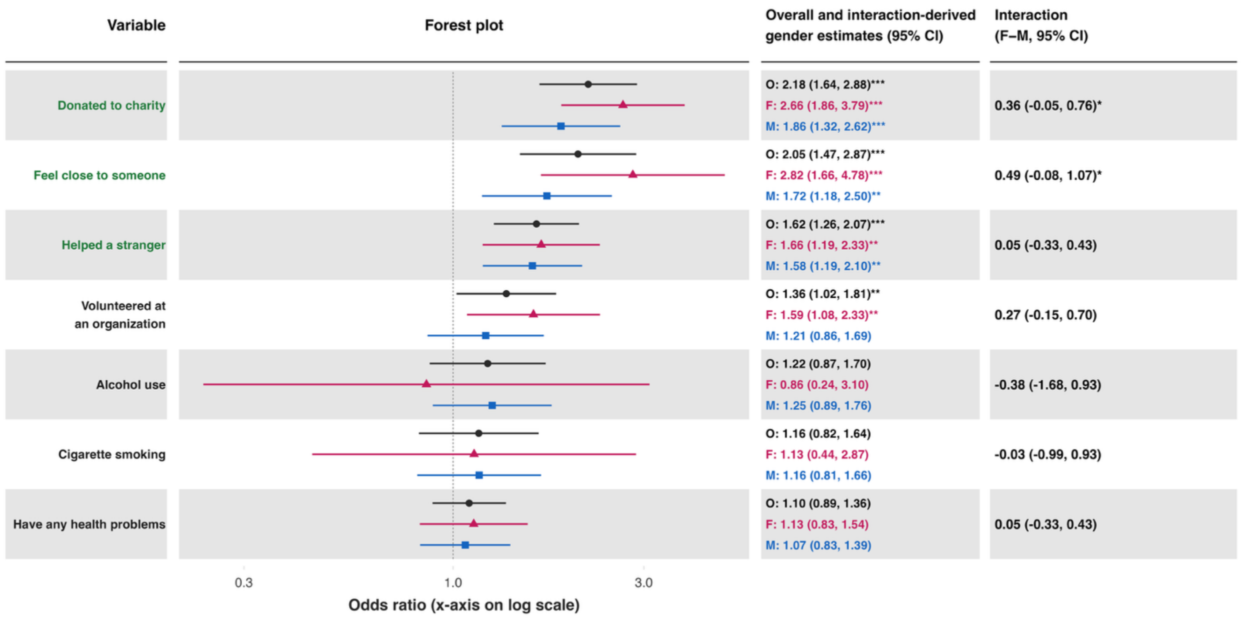

### Adjusted associations between feeling like an outsider in one's family when growing up and adult outcomes in India
### Residence: Rural

Wave 1, survey-weighted ordinal regression using the proportional odds model (`svyolr`). One model was fitted for each ordinal outcome (16 models total).
Estimates are adjusted cumulative odds ratios (cOR) of the predictor.
The interaction column reports the difference in log-cumulative odds ratios (interaction beta) with 95% confidence intervals (95% CIs).

**Group** ● Overall (O) ▲ Female (F) ■ Male (M)

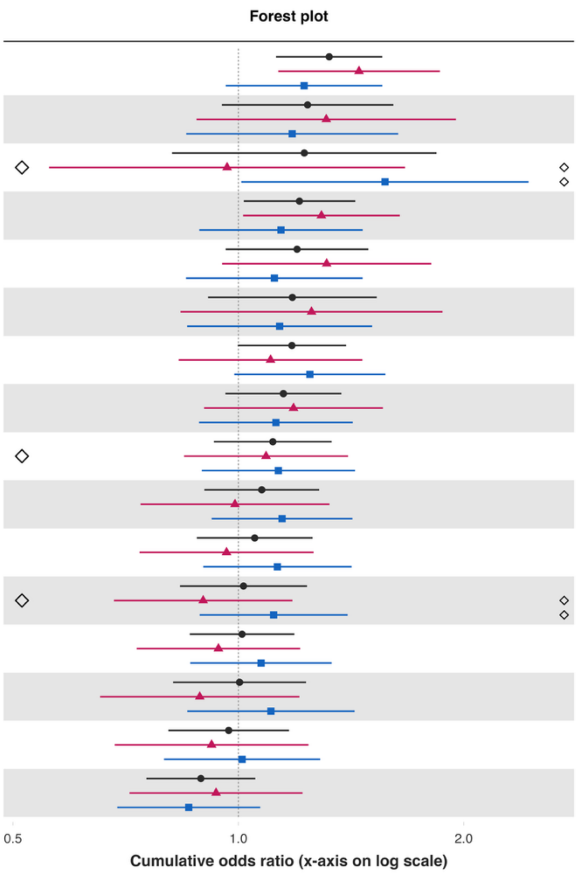


| Variable | Overall and interaction-derived gender estimates (95% CI) | Interaction (F–M, 95% CI) |
|---|---|---|
| Feeling nervous or anxious§ | O: 1.32 (1.12, 1.56)***<br>F: 1.45 (1.13, 1.86)**<br>M: 1.22 (0.96, 1.56) | 0.17 (-0.20, 0.53) |
| Support reducing fferences in income levels§ | O: 1.24 (0.95, 1.61)<br>F: 1.31 (0.88, 1.95)<br>M: 1.18 (0.85, 1.63) | 0.10 (-0.39, 0.60) |
| Obeying the law§ | O: 1.22 (0.82, 1.84)<br>F: 0.97 (0.56, 1.67)<br>M: 1.57 (1.01, 2.44)** | -0.49 (-1.07, 0.10) |
| Unable to control worry§ | O: 1.21 (1.02, 1.43)**<br>F: 1.29 (1.01, 1.64)**<br>M: 1.14 (0.89, 1.47) | 0.12 (-0.23, 0.48) |
| Government approval§ | O: 1.20 (0.96, 1.49)<br>F: 1.31 (0.95, 1.81)*<br>M: 1.12 (0.85, 1.46) | 0.16 (-0.23, 0.56) |
| Say in government# | O: 1.18 (0.91, 1.53)<br>F: 1.25 (0.84, 1.87)<br>M: 1.14 (0.85, 1.51) | 0.10 (-0.34, 0.54) |
| Depressive symptoms§ | O: 1.18 (1.00, 1.39)*<br>F: 1.10 (0.83, 1.46)<br>M: 1.25 (0.99, 1.57)* | -0.12 (-0.51, 0.27) |
| Forgiveness§ | O: 1.15 (0.96, 1.37)<br>F: 1.18 (0.90, 1.56)<br>M: 1.12 (0.89, 1.42) | 0.05 (-0.31, 0.42) |
| Extent of suffering§ | O: 1.11 (0.93, 1.33)<br>F: 1.09 (0.85, 1.40)<br>M: 1.13 (0.89, 1.43) | -0.04 (-0.36, 0.29) |
| Trust in others§ | O: 1.07 (0.90, 1.28)<br>F: 0.99 (0.74, 1.32)<br>M: 1.14 (0.92, 1.42) | -0.14 (-0.51, 0.22) |
| Feeling capable§ | O: 1.05 (0.88, 1.26)<br>F: 0.96 (0.74, 1.26)<br>M: 1.13 (0.90, 1.42) | -0.16 (-0.50, 0.19) |
| Various aspects of life are balanced§ | O: 1.02 (0.84, 1.23)<br>F: 0.90 (0.68, 1.18)<br>M: 1.11 (0.89, 1.40) | -0.22 (-0.54, 0.10) |
| Have little interest in doing things§ | O: 1.01 (0.86, 1.19)<br>F: 0.94 (0.73, 1.21)<br>M: 1.07 (0.86, 1.33) | -0.13 (-0.47, 0.21) |
| Distress from e-threatening experiences§ | O: 1.00 (0.82, 1.23)<br>F: 0.89 (0.65, 1.21)<br>M: 1.11 (0.85, 1.43) | -0.22 (-0.61, 0.17) |
| Feel at peace§ | O: 0.97 (0.81, 1.17)<br>F: 0.92 (0.68, 1.24)<br>M: 1.01 (0.80, 1.29) | -0.09 (-0.48, 0.29) |
| Perceived discrimination§ | O: 0.89 (0.75, 1.05)<br>F: 0.93 (0.72, 1.22)<br>M: 0.86 (0.69, 1.07) | 0.08 (-0.27, 0.43) |

*Feeling like an outsider in one's family when growing up – Small town or village*

**Adjusted associations between feeling like an outsider in one's family when growing up and adult outcomes in India**
**Residence: Small town**

Wave 1, survey-weighted linear model. One model was fitted for each continuous outcome (24 models total, for 22 outcomes and 2 flourishing composite indices).
Estimates are the adjusted model coefficients (beta) of the predictor.

**Group** Overall (O) Female (F) Male (M)

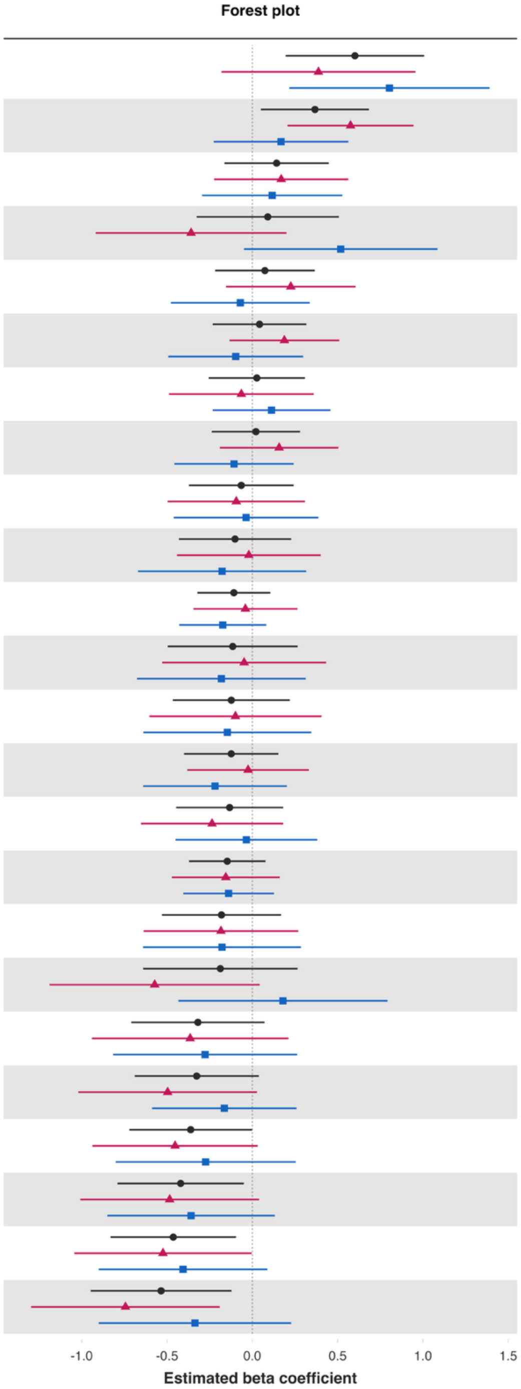


| Variable | Overall and interaction-derived gender estimates (95% CI) | Interaction (F–M, 95% CI) |
|---|---|---|
| Loneliness§ | O: 0.60 (0.20, 1.01)**<br>F: 0.39 (-0.18, 0.96)<br>M: 0.80 (0.22, 1.39)** | -0.42 (-1.24, 0.41) |
| Understand purpose in life‡ | O: 0.37 (0.05, 0.68)**<br>F: 0.58 (0.21, 0.95)**<br>M: 0.17 (-0.23, 0.56) | 0.41 (-0.03, 0.84)* |
| Relationship contentment‡ | O: 0.14 (-0.16, 0.45)<br>F: 0.17 (-0.22, 0.56)<br>M: 0.12 (-0.29, 0.53) | 0.05 (-0.47, 0.58) |
| Worry about safety, food, or housing§§‡‡ | O: 0.09 (-0.33, 0.51)<br>F: -0.36 (-0.92, 0.20)<br>M: 0.52 (-0.05, 1.09)* | -0.88 (-1.65, -0.11)** |
| Show love often | O: 0.07 (-0.22, 0.37)<br>F: 0.23 (-0.15, 0.61)<br>M: -0.07 (-0.48, 0.34) | 0.30 (-0.23, 0.83) |
| Remain hopeful for the future | O: 0.04 (-0.23, 0.32)<br>F: 0.19 (-0.13, 0.51)<br>M: -0.10 (-0.49, 0.30) | 0.29 (-0.19, 0.76) |
| Belonging | O: 0.03 (-0.26, 0.31)<br>F: -0.06 (-0.49, 0.36)<br>M: 0.11 (-0.23, 0.46) | -0.18 (-0.70, 0.35) |
| Act to promote good‡ | O: 0.02 (-0.24, 0.28)<br>F: 0.16 (-0.19, 0.50)<br>M: -0.11 (-0.46, 0.24) | 0.26 (-0.21, 0.73) |
| Relationship satisfaction‡ | O: -0.06 (-0.37, 0.24)<br>F: -0.09 (-0.50, 0.31)<br>M: -0.04 (-0.46, 0.39) | -0.06 (-0.61, 0.50) |
| Have a long list of gratitude | O: -0.10 (-0.43, 0.23)<br>F: -0.02 (-0.44, 0.40)<br>M: -0.18 (-0.67, 0.32) | 0.16 (-0.48, 0.80) |
| Flourishing (with financial indicators) | O: -0.11 (-0.32, 0.11)<br>F: -0.04 (-0.35, 0.26)<br>M: -0.17 (-0.43, 0.08) | 0.13 (-0.23, 0.50) |
| Life evaluation five years from now | O: -0.12 (-0.50, 0.27)<br>F: -0.05 (-0.53, 0.43)<br>M: -0.18 (-0.68, 0.31) | 0.13 (-0.48, 0.74) |
| Life worthwhile‡ | O: -0.12 (-0.47, 0.22)<br>F: -0.10 (-0.60, 0.41)<br>M: -0.15 (-0.64, 0.35) | 0.05 (-0.68, 0.77) |
| Positive expectations | O: -0.12 (-0.40, 0.15)<br>F: -0.02 (-0.38, 0.33)<br>M: -0.22 (-0.64, 0.20) | 0.19 (-0.36, 0.75) |
| Freedom | O: -0.13 (-0.45, 0.18)<br>F: -0.24 (-0.65, 0.18)<br>M: -0.04 (-0.45, 0.38) | -0.20 (-0.75, 0.34) |
| Flourishing (without financial indicators) | O: -0.15 (-0.37, 0.08)<br>F: -0.16 (-0.47, 0.16)<br>M: -0.14 (-0.41, 0.13) | -0.02 (-0.39, 0.36) |
| Physical health‡ | O: -0.18 (-0.53, 0.17)<br>F: -0.18 (-0.64, 0.27)<br>M: -0.18 (-0.64, 0.28) | -0.01 (-0.60, 0.59) |
| Worry about meeting normal monthly living expenses§§‡‡ | O: -0.19 (-0.64, 0.27)<br>F: -0.57 (-1.19, 0.04)*<br>M: 0.18 (-0.43, 0.79) | -0.75 (-1.60, 0.10)* |
| Life evaluation today | O: -0.32 (-0.71, 0.07)<br>F: -0.36 (-0.94, 0.21)<br>M: -0.28 (-0.82, 0.26) | -0.09 (-0.88, 0.71) |
| Mental health‡ | O: -0.33 (-0.69, 0.04)*<br>F: -0.50 (-1.02, 0.03)*<br>M: -0.16 (-0.59, 0.26) | -0.33 (-0.95, 0.28) |
| Give up some happiness now for greater happiness later‡ | O: -0.36 (-0.72, -0.00)**<br>F: -0.45 (-0.94, 0.03)*<br>M: -0.27 (-0.80, 0.25) | -0.18 (-0.89, 0.53) |
| Happiness‡ | O: -0.42 (-0.79, -0.05)**<br>F: -0.48 (-1.01, 0.04)*<br>M: -0.36 (-0.85, 0.13) | -0.13 (-0.82, 0.57) |
| Life satisfaction‡ | O: -0.46 (-0.83, -0.10)**<br>F: -0.52 (-1.04, -0.00)**<br>M: -0.41 (-0.90, 0.09) | -0.12 (-0.81, 0.58) |
| Perceived support from others | O: -0.53 (-0.95, -0.12)**<br>F: -0.74 (-1.30, -0.19)**<br>M: -0.34 (-0.90, 0.23) | -0.41 (-1.16, 0.35) |

### Adjusted associations between feeling like an outsider in one's family when growing up and adult outcomes in India
### Residence: Small town

Wave 1, survey-weighted logistic model. One model was fitted for each binary outcome (7 models total). Estimates are adjusted odds ratios (OR) of the predictor.
The interaction column reports the difference in log-odds ratios (interaction beta) with 95% confidence intervals (95% CIs).

Group ● Overall (O) ▲ Female (F) ■ Male (M)

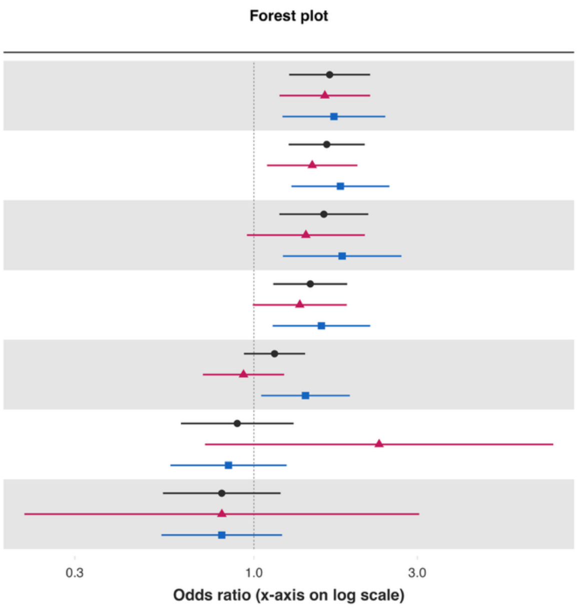


| Variable | Overall and interaction-derived gender estimates (95% CI) | Interaction (F–M, 95% CI) |
|---|---|---|
| Volunteered at an organization | O: 1.66 (1.27, 2.18)***<br>F: 1.61 (1.19, 2.18)**<br>M: 1.71 (1.21, 2.42)** | -0.06 (-0.42, 0.30) |
| Donated to charity | O: 1.63 (1.26, 2.11)***<br>F: 1.48 (1.09, 2.01)**<br>M: 1.79 (1.29, 2.49)*** | -0.19 (-0.56, 0.18) |
| Feel close to someone | O: 1.60 (1.19, 2.16)**<br>F: 1.42 (0.95, 2.11)*<br>M: 1.81 (1.22, 2.70)** | -0.24 (-0.76, 0.28) |
| Helped a stranger | O: 1.46 (1.14, 1.87)**<br>F: 1.36 (0.99, 1.87)*<br>M: 1.57 (1.13, 2.19)** | -0.15 (-0.55, 0.26) |
| Have any health problems | O: 1.15 (0.94, 1.41)<br>F: 0.93 (0.71, 1.23)<br>M: 1.41 (1.05, 1.91)** | -0.42 (-0.82, -0.02)** |
| Cigarette smoking | O: 0.89 (0.61, 1.31)<br>F: 2.32 (0.72, 7.48)<br>M: 0.84 (0.57, 1.25) | 1.01 (-0.20, 2.23) |
| Alcohol use | O: 0.81 (0.54, 1.20)<br>F: 0.81 (0.21, 3.04)<br>M: 0.81 (0.54, 1.21) | 0.00 (-1.36, 1.36) |

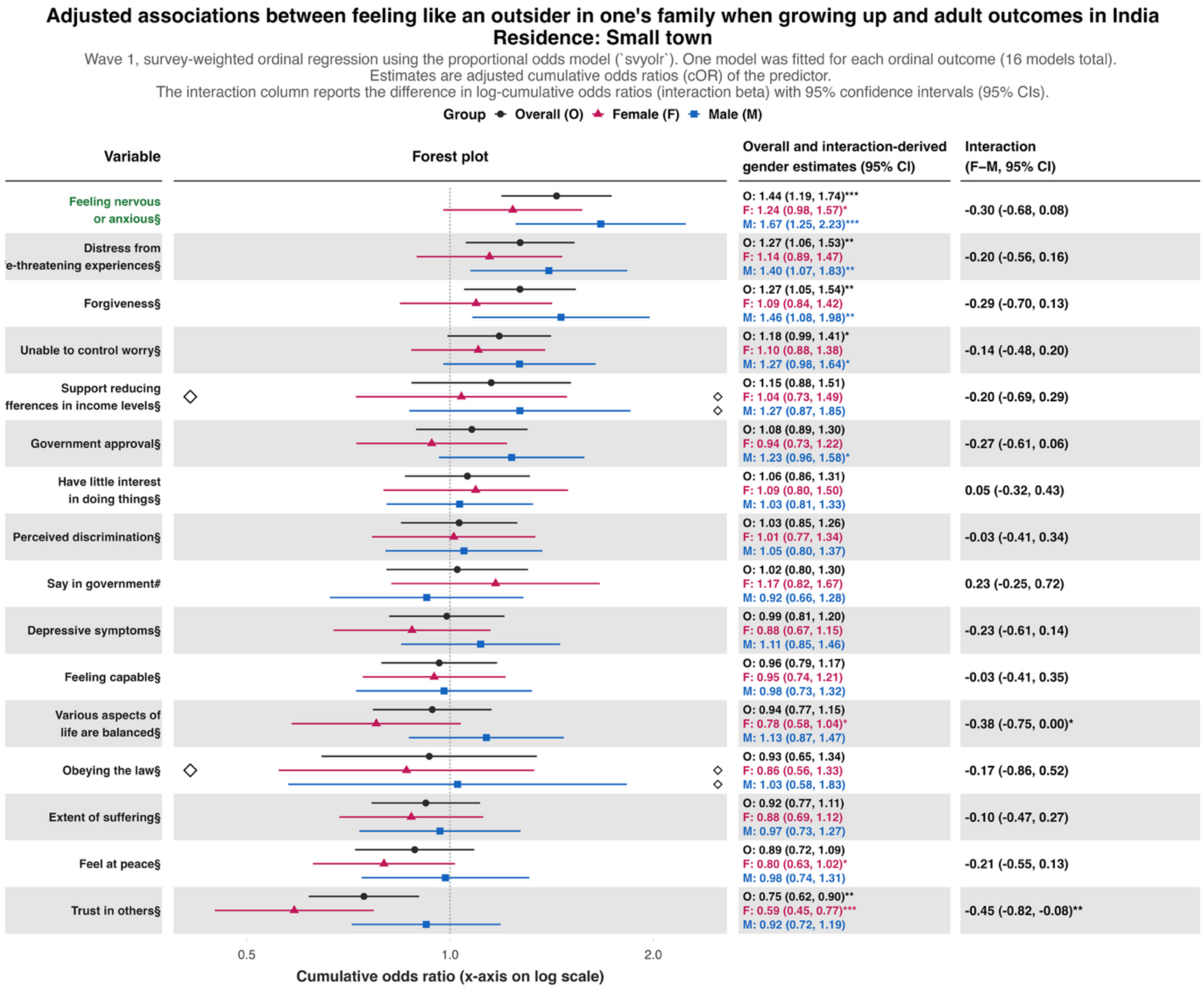

Adjusted associations between feeling like an outsider in one's family when growing up and adult outcomes in India
Residence: Small town
Wave 1, survey-weighted ordinal regression using the proportional odds model (`svyolr`). One model was fitted for each ordinal outcome (16 models total).
Estimates are adjusted cumulative odds ratios (cOR) of the predictor.
The interaction column reports the difference in log-cumulative odds ratios (interaction beta) with 95% confidence intervals (95% CIs).
Group Overall (O) Female (F) Male (M)
Variable
Forest plot
Overall and interaction-derived gender estimates (95% CI)
Interaction (F–M, 95% CI)
Feeling nervous or anxious§
O: 1.44 (1.19, 1.74)***
F: 1.24 (0.98, 1.57)*
M: 1.67 (1.25, 2.23)***
-0.30 (-0.68, 0.08)
Distress from e-threatening experiences§
O: 1.27 (1.06, 1.53)**
F: 1.14 (0.89, 1.47)
M: 1.40 (1.07, 1.83)**
-0.20 (-0.56, 0.16)
Forgiveness§
O: 1.27 (1.05, 1.54)**
F: 1.09 (0.84, 1.42)
M: 1.46 (1.08, 1.98)**
-0.29 (-0.70, 0.13)
Unable to control worry§
O: 1.18 (0.99, 1.41)*
F: 1.10 (0.88, 1.38)
M: 1.27 (0.98, 1.64)*
-0.14 (-0.48, 0.20)
Support reducing fferences in income levels§
O: 1.15 (0.88, 1.51)
F: 1.04 (0.73, 1.49)
M: 1.27 (0.87, 1.85)
-0.20 (-0.69, 0.29)
Government approval§
O: 1.08 (0.89, 1.30)
F: 0.94 (0.73, 1.22)
M: 1.23 (0.96, 1.58)*
-0.27 (-0.61, 0.06)
Have little interest in doing things§
O: 1.06 (0.86, 1.31)
F: 1.09 (0.80, 1.50)
M: 1.03 (0.81, 1.33)
0.05 (-0.32, 0.43)
Perceived discrimination§
O: 1.03 (0.85, 1.26)
F: 1.01 (0.77, 1.34)
M: 1.05 (0.80, 1.37)
-0.03 (-0.41, 0.34)
Say in government#
O: 1.02 (0.80, 1.30)
F: 1.17 (0.82, 1.67)
M: 0.92 (0.66, 1.28)
0.23 (-0.25, 0.72)
Depressive symptoms§
O: 0.99 (0.81, 1.20)
F: 0.88 (0.67, 1.15)
M: 1.11 (0.85, 1.46)
-0.23 (-0.61, 0.14)
Feeling capable§
O: 0.96 (0.79, 1.17)
F: 0.95 (0.74, 1.21)
M: 0.98 (0.73, 1.32)
-0.03 (-0.41, 0.35)
Various aspects of life are balanced§
O: 0.94 (0.77, 1.15)
F: 0.78 (0.58, 1.04)*
M: 1.13 (0.87, 1.47)
-0.38 (-0.75, 0.00)*
Obeying the law§
O: 0.93 (0.65, 1.34)
F: 0.86 (0.56, 1.33)
M: 1.03 (0.58, 1.83)
-0.17 (-0.86, 0.52)
Extent of suffering§
O: 0.92 (0.77, 1.11)
F: 0.88 (0.69, 1.12)
M: 0.97 (0.73, 1.27)
-0.10 (-0.47, 0.27)
Feel at peace§
O: 0.89 (0.72, 1.09)
F: 0.80 (0.63, 1.02)*
M: 0.98 (0.74, 1.31)
-0.21 (-0.55, 0.13)
Trust in others§
O: 0.75 (0.62, 0.90)**
F: 0.59 (0.45, 0.77)***
M: 0.92 (0.72, 1.19)
-0.45 (-0.82, -0.08)**
0.5
1.0
2.0
Cumulative odds ratio (x-axis on log scale)

*Feeling like an outsider in one's family when growing up – Large city or suburb of a large city*

**Adjusted associations between feeling like an outsider in one's family when growing up and adult outcomes in India**
**Residence: Large city or suburb**

Wave 1, survey-weighted linear model. One model was fitted for each continuous outcome (24 models total, for 22 outcomes and 2 flourishing composite indices). Estimates are the adjusted model coefficients (beta) of the predictor.

Group: Overall (O) · Female (F) · Male (M)

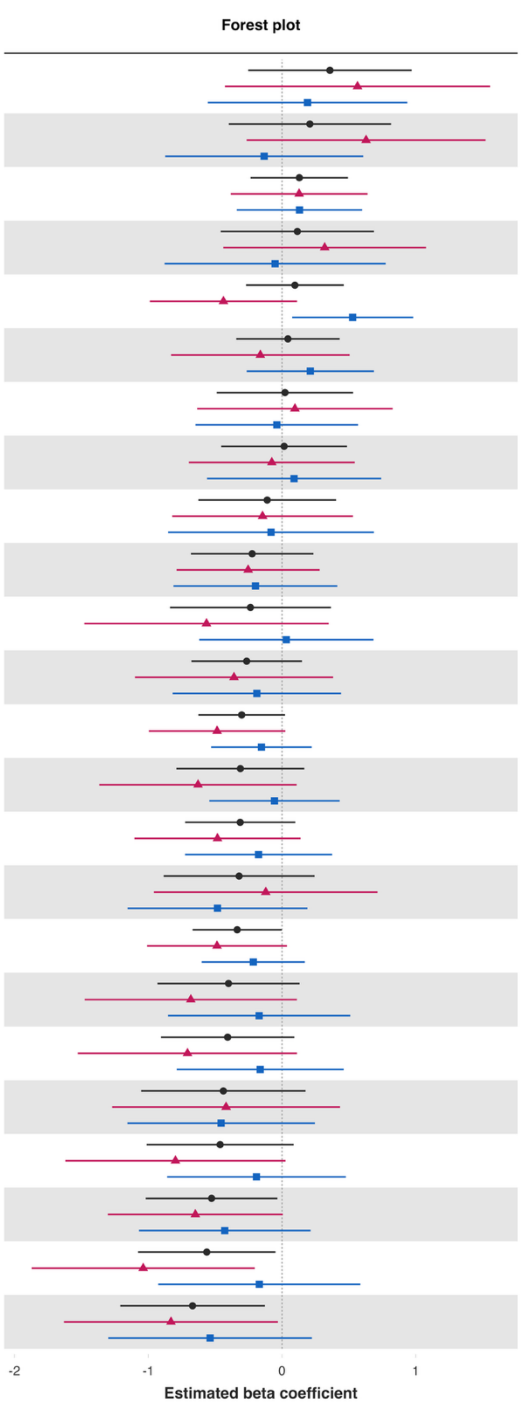


| Variable | Overall and interaction-derived gender estimates (95% CI) | Interaction (F–M, 95% CI) |
|---|---|---|
| Loneliness§ | O: 0.36 (-0.25, 0.97)<br>F: 0.56 (-0.43, 1.56)<br>M: 0.19 (-0.56, 0.94) | 0.37 (-0.86, 1.61) |
| Worry about meeting normal monthly living expenses§§‡‡ | O: 0.21 (-0.40, 0.81)<br>F: 0.63 (-0.26, 1.52)<br>M: -0.13 (-0.87, 0.61) | 0.76 (-0.33, 1.86) |
| Belonging | O: 0.13 (-0.24, 0.49)<br>F: 0.13 (-0.38, 0.64)<br>M: 0.13 (-0.34, 0.60) | -0.00 (-0.66, 0.65) |
| Worry about safety, food, or housing§§‡‡ | O: 0.11 (-0.46, 0.69)<br>F: 0.32 (-0.44, 1.08)<br>M: -0.05 (-0.88, 0.78) | 0.37 (-0.74, 1.48) |
| Have a long list of gratitude | O: 0.10 (-0.27, 0.46)<br>F: -0.44 (-0.99, 0.11)<br>M: 0.53 (0.08, 0.98)** | -0.97 (-1.64, -0.29)** |
| Freedom | O: 0.04 (-0.34, 0.43)<br>F: -0.16 (-0.83, 0.51)<br>M: 0.21 (-0.26, 0.69) | -0.37 (-1.21, 0.46) |
| Show love often | O: 0.02 (-0.49, 0.53)<br>F: 0.10 (-0.63, 0.83)<br>M: -0.04 (-0.65, 0.57) | 0.14 (-0.73, 1.00) |
| Understand purpose in life‡ | O: 0.02 (-0.45, 0.49)<br>F: -0.08 (-0.70, 0.54)<br>M: 0.09 (-0.56, 0.74) | -0.17 (-1.03, 0.69) |
| Life evaluation five years from now | O: -0.11 (-0.63, 0.40)<br>F: -0.15 (-0.82, 0.53)<br>M: -0.08 (-0.85, 0.69) | -0.06 (-1.10, 0.98) |
| Act to promote good‡ | O: -0.22 (-0.68, 0.23)<br>F: -0.25 (-0.79, 0.28)<br>M: -0.20 (-0.81, 0.41) | -0.05 (-0.77, 0.66) |
| Happiness‡ | O: -0.24 (-0.84, 0.37)<br>F: -0.56 (-1.48, 0.35)<br>M: 0.03 (-0.62, 0.68) | -0.60 (-1.60, 0.41) |
| Relationship contentment‡ | O: -0.26 (-0.68, 0.15)<br>F: -0.36 (-1.10, 0.38)<br>M: -0.19 (-0.82, 0.44) | -0.17 (-1.26, 0.92) |
| Flourishing (with financial indicators) | O: -0.30 (-0.63, 0.02)*<br>F: -0.49 (-1.00, 0.03)*<br>M: -0.15 (-0.53, 0.22) | -0.33 (-0.93, 0.27) |
| Remain hopeful for the future | O: -0.31 (-0.79, 0.17)<br>F: -0.63 (-1.37, 0.11)*<br>M: -0.06 (-0.54, 0.43) | -0.57 (-1.35, 0.21) |
| Positive expectations | O: -0.31 (-0.72, 0.10)<br>F: -0.48 (-1.10, 0.14)<br>M: -0.18 (-0.73, 0.37) | -0.31 (-1.13, 0.52) |
| Life satisfaction‡ | O: -0.32 (-0.89, 0.24)<br>F: -0.12 (-0.96, 0.71)<br>M: -0.48 (-1.15, 0.19) | 0.36 (-0.63, 1.35) |
| Flourishing (without financial indicators) | O: -0.34 (-0.67, -0.00)*<br>F: -0.49 (-1.01, 0.04)*<br>M: -0.21 (-0.60, 0.17) | -0.27 (-0.89, 0.34) |
| Give up some happiness now for greater happiness later‡ | O: -0.40 (-0.93, 0.13)<br>F: -0.68 (-1.48, 0.11)*<br>M: -0.17 (-0.85, 0.51) | -0.51 (-1.52, 0.50) |
| Relationship satisfaction‡ | O: -0.41 (-0.90, 0.09)<br>F: -0.71 (-1.53, 0.11)*<br>M: -0.16 (-0.79, 0.46) | -0.54 (-1.58, 0.50) |
| Life worthwhile‡ | O: -0.44 (-1.05, 0.18)<br>F: -0.42 (-1.27, 0.43)<br>M: -0.46 (-1.16, 0.25) | 0.04 (-0.90, 0.98) |
| Mental health‡ | O: -0.46 (-1.01, 0.09)<br>F: -0.80 (-1.62, 0.03)*<br>M: -0.19 (-0.86, 0.48) | -0.61 (-1.62, 0.41) |
| Physical health‡ | O: -0.53 (-1.02, -0.03)**<br>F: -0.65 (-1.30, 0.01)*<br>M: -0.43 (-1.07, 0.21) | -0.22 (-1.06, 0.62) |
| Perceived support from others | O: -0.56 (-1.08, -0.05)**<br>F: -1.04 (-1.87, -0.20)**<br>M: -0.17 (-0.92, 0.59) | -0.87 (-2.08, 0.34) |
| Life evaluation today | O: -0.67 (-1.21, -0.13)**<br>F: -0.83 (-1.63, -0.03)**<br>M: -0.54 (-1.30, 0.22) | -0.29 (-1.42, 0.84) |

### Adjusted associations between feeling like an outsider in one's family when growing up and adult outcomes in India
### Residence: Large city or suburb

Wave 1, survey-weighted logistic model. One model was fitted for each binary outcome (7 models total). Estimates are adjusted odds ratios (OR) of the predictor. The interaction column reports the difference in log-odds ratios (interaction beta) with 95% confidence intervals (95% CIs).

**Group** Overall (O) Female (F) Male (M)

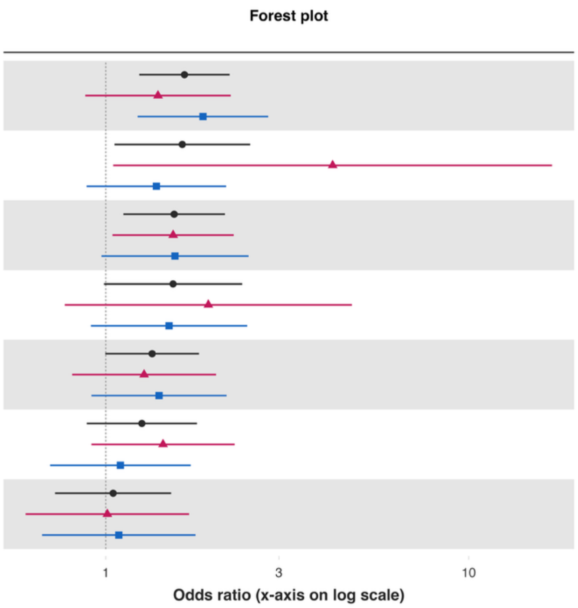


| Variable | Overall and interaction-derived gender estimates (95% CI) | Interaction (F–M, 95% CI) |
|---|---|---|
| Volunteered at an organization | O: 1.65 (1.24, 2.19)***<br>F: 1.39 (0.88, 2.21)<br>M: 1.85 (1.22, 2.80)** | -0.29 (-0.95, 0.38) |
| Alcohol use | O: 1.62 (1.06, 2.50)**<br>F: 4.22 (1.05, 16.97)**<br>M: 1.38 (0.89, 2.15) | 1.12 (-0.45, 2.68) |
| Donated to charity | O: 1.54 (1.12, 2.13)**<br>F: 1.53 (1.04, 2.25)**<br>M: 1.55 (0.97, 2.47)* | -0.01 (-0.59, 0.56) |
| Cigarette smoking | O: 1.53 (0.99, 2.37)*<br>F: 1.92 (0.77, 4.77)<br>M: 1.49 (0.91, 2.45) | 0.25 (-0.87, 1.37) |
| Helped a stranger | O: 1.34 (1.00, 1.81)*<br>F: 1.27 (0.81, 2.01)<br>M: 1.40 (0.91, 2.15) | -0.09 (-0.75, 0.56) |
| Have any health problems | O: 1.26 (0.89, 1.78)<br>F: 1.44 (0.91, 2.26)<br>M: 1.10 (0.70, 1.71) | 0.27 (-0.29, 0.83) |
| Feel close to someone | O: 1.05 (0.72, 1.51)<br>F: 1.01 (0.60, 1.70)<br>M: 1.09 (0.67, 1.77) | -0.07 (-0.76, 0.61) |

**Adjusted associations between feeling like an outsider in one's family when growing up and adult outcomes in India**
**Residence: Large city or suburb**

Wave 1, survey-weighted ordinal regression using the proportional odds model (`svyolr`). One model was fitted for each ordinal outcome (16 models total).
Estimates are adjusted cumulative odds ratios (cOR) of the predictor.
The interaction column reports the difference in log-cumulative odds ratios (interaction beta) with 95% confidence intervals (95% CIs).

Group: Overall (O), Female (F), Male (M)

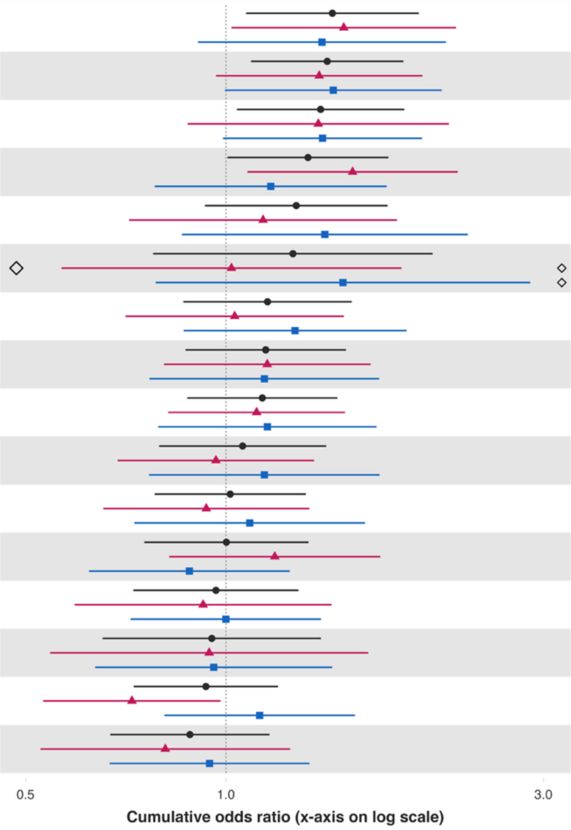


| Variable | Overall and interaction-derived gender estimates (95% CI) | Interaction (F–M, 95% CI) |
|---|---|---|
| Unable to control worry§ | O: 1.45 (1.07, 1.95)**<br>F: 1.50 (1.02, 2.22)**<br>M: 1.39 (0.91, 2.14) | 0.08 (-0.49, 0.64) |
| Forgiveness§ | O: 1.42 (1.09, 1.85)**<br>F: 1.38 (0.97, 1.97)*<br>M: 1.45 (1.00, 2.11)* | -0.05 (-0.57, 0.47) |
| Extent of suffering§ | O: 1.39 (1.04, 1.85)**<br>F: 1.38 (0.88, 2.16)<br>M: 1.40 (0.99, 1.97)* | -0.01 (-0.56, 0.53) |
| Distress from e-threatening experiences§ | O: 1.33 (1.01, 1.75)**<br>F: 1.55 (1.08, 2.23)**<br>M: 1.17 (0.78, 1.74) | 0.28 (-0.25, 0.82) |
| Say in government# | O: 1.28 (0.93, 1.75)<br>F: 1.14 (0.72, 1.81)<br>M: 1.41 (0.86, 2.31) | -0.21 (-0.94, 0.51) |
| Obeying the law§ | O: 1.26 (0.78, 2.04)<br>F: 1.02 (0.57, 1.84)<br>M: 1.50 (0.78, 2.87) | -0.39 (-1.15, 0.38) |
| Depressive symptoms§ | O: 1.15 (0.86, 1.55)<br>F: 1.03 (0.71, 1.50)<br>M: 1.27 (0.86, 1.87) | -0.21 (-0.70, 0.29) |
| Feeling nervous or anxious§ | O: 1.15 (0.87, 1.51)<br>F: 1.15 (0.81, 1.65)<br>M: 1.14 (0.77, 1.70) | 0.01 (-0.51, 0.53) |
| Have little interest in doing things§ | O: 1.13 (0.87, 1.47)<br>F: 1.11 (0.82, 1.51)<br>M: 1.15 (0.79, 1.68) | -0.04 (-0.49, 0.42) |
| Government approval§ | O: 1.06 (0.79, 1.41)<br>F: 0.97 (0.69, 1.36)<br>M: 1.14 (0.77, 1.70) | -0.17 (-0.64, 0.31) |
| Feeling capable§ | O: 1.02 (0.78, 1.32)<br>F: 0.93 (0.65, 1.33)<br>M: 1.09 (0.73, 1.62) | -0.15 (-0.71, 0.41) |
| Perceived discrimination§ | O: 1.00 (0.75, 1.33)<br>F: 1.18 (0.82, 1.71)<br>M: 0.88 (0.62, 1.25) | 0.30 (-0.13, 0.72) |
| Various aspects of life are balanced§ | O: 0.97 (0.73, 1.29)<br>F: 0.92 (0.59, 1.44)<br>M: 1.00 (0.72, 1.39) | -0.08 (-0.60, 0.44) |
| Support reducing fferences in income levels§ | O: 0.95 (0.65, 1.39)<br>F: 0.94 (0.54, 1.64)<br>M: 0.96 (0.64, 1.44) | -0.02 (-0.60, 0.57) |
| Trust in others§ | O: 0.93 (0.73, 1.20)<br>F: 0.72 (0.53, 0.98)**<br>M: 1.12 (0.81, 1.56) | -0.44 (-0.86, -0.03)** |
| Feel at peace§ | O: 0.88 (0.67, 1.16)<br>F: 0.81 (0.53, 1.25)<br>M: 0.94 (0.67, 1.33) | -0.15 (-0.70, 0.39) |

**Predictor:** ***Not feeling loved by mother***
***Not feeling loved by mother – Rural area or on a farm***

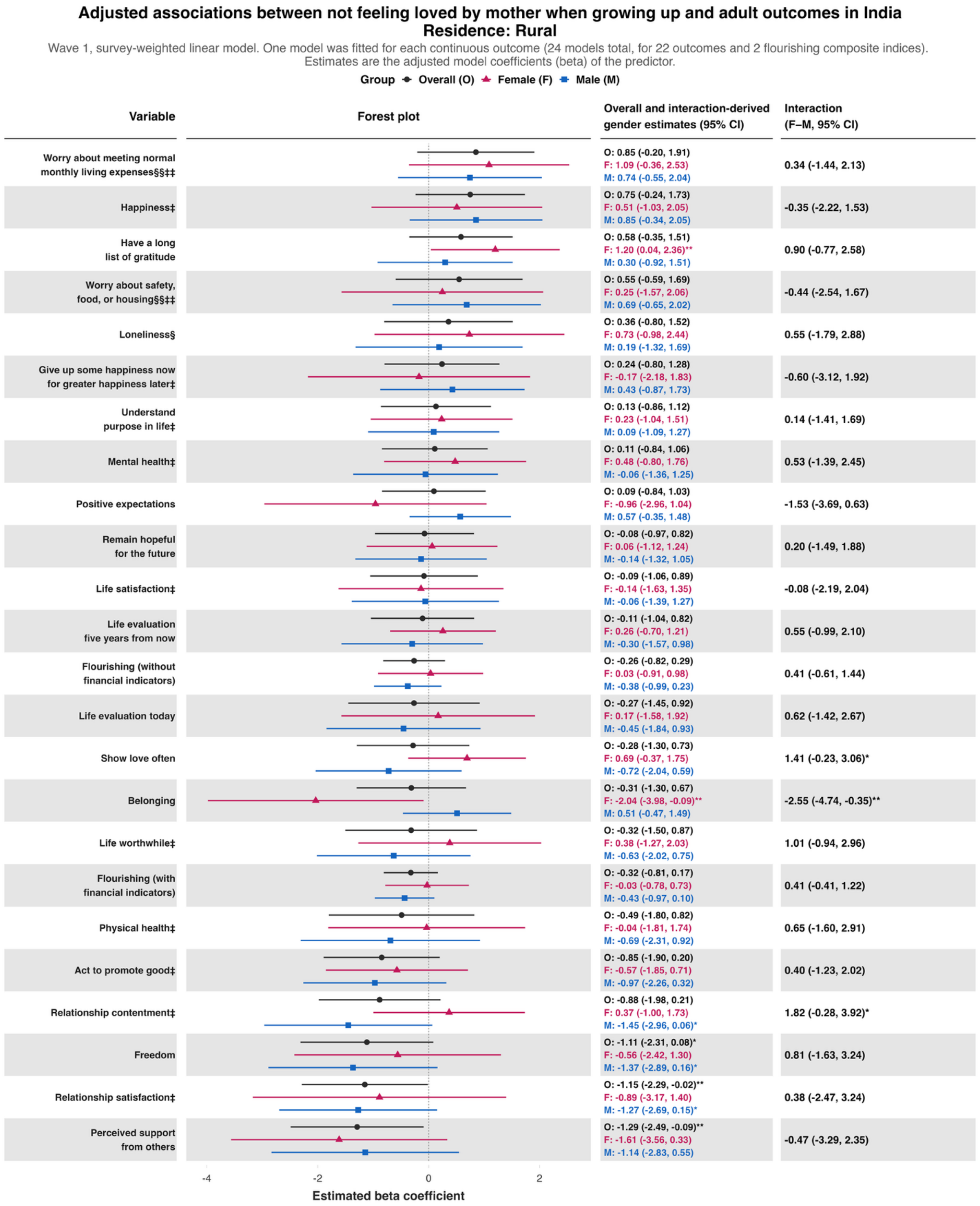

**Adjusted associations between not feeling loved by mother when growing up and adult outcomes in India**
**Residence: Rural**

Wave 1, survey-weighted logistic model. One model was fitted for each binary outcome (7 models total). Estimates are adjusted odds ratios (OR) of the predictor.
The interaction column reports the difference in log-odds ratios (interaction beta) with 95% confidence intervals (95% CIs).

Group: Overall (O), Female (F), Male (M)

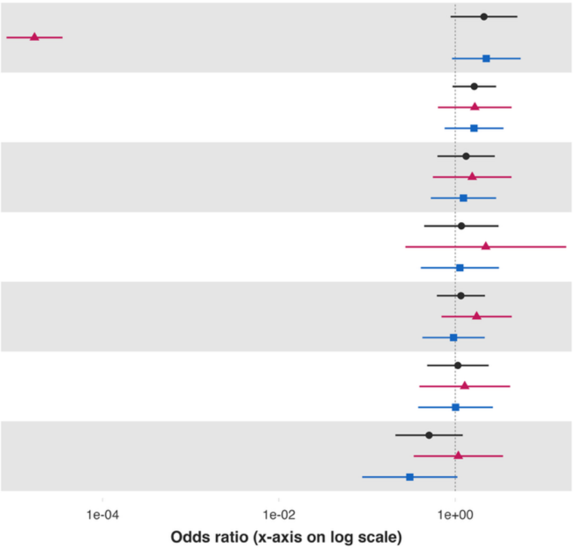


| Variable | Overall and interaction-derived gender estimates (95% CI) | Interaction (F–M, 95% CI) |
|---|---|---|
| Alcohol use | O: 2.12 (0.89, 5.06)*<br>F: 0.00 (0.00, 0.00)***<br>M: 2.25 (0.92, 5.52)* | -11.80 (-13.00, -10.59)*** |
| Donated to charity | O: 1.64 (0.93, 2.90)*<br>F: 1.67 (0.64, 4.36)<br>M: 1.63 (0.76, 3.52) | 0.02 (-1.30, 1.34) |
| Helped a stranger | O: 1.33 (0.63, 2.81)<br>F: 1.56 (0.56, 4.35)<br>M: 1.24 (0.53, 2.90) | 0.23 (-0.90, 1.36) |
| Cigarette smoking | O: 1.17 (0.44, 3.10)<br>F: 2.22 (0.27, 18.15)<br>M: 1.13 (0.41, 3.13) | 0.68 (-1.62, 2.97) |
| Volunteered at an organization | O: 1.16 (0.62, 2.17)<br>F: 1.75 (0.70, 4.39)<br>M: 0.96 (0.42, 2.16) | 0.60 (-0.60, 1.80) |
| Feel close to someone | O: 1.07 (0.48, 2.39)<br>F: 1.28 (0.39, 4.19)<br>M: 1.01 (0.38, 2.67) | 0.24 (-1.27, 1.75) |
| Have any health problems | O: 0.51 (0.21, 1.22)<br>F: 1.08 (0.34, 3.48)<br>M: 0.31 (0.09, 1.06)* | 1.27 (-0.36, 2.90) |

**Adjusted associations between not feeling loved by mother when growing up and adult outcomes in India**
**Residence: Rural**

Wave 1, survey-weighted ordinal regression using the proportional odds model (`svyolr`). One model was fitted for each ordinal outcome (16 models total).
Estimates are adjusted cumulative odds ratios (cOR) of the predictor.
The interaction column reports the difference in log-cumulative odds ratios (interaction beta) with 95% confidence intervals (95% CIs).

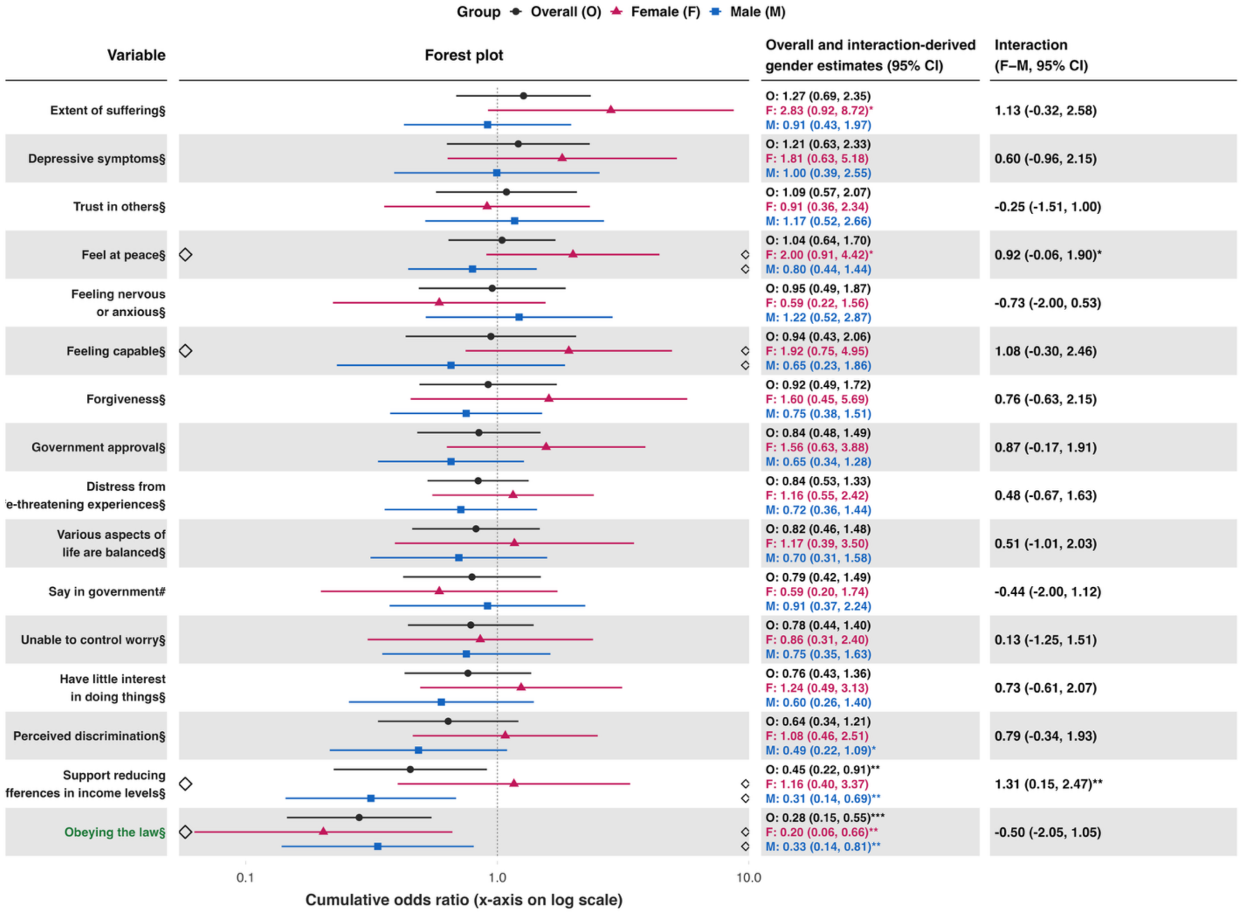

*Not feeling loved by mother – Small town or village*

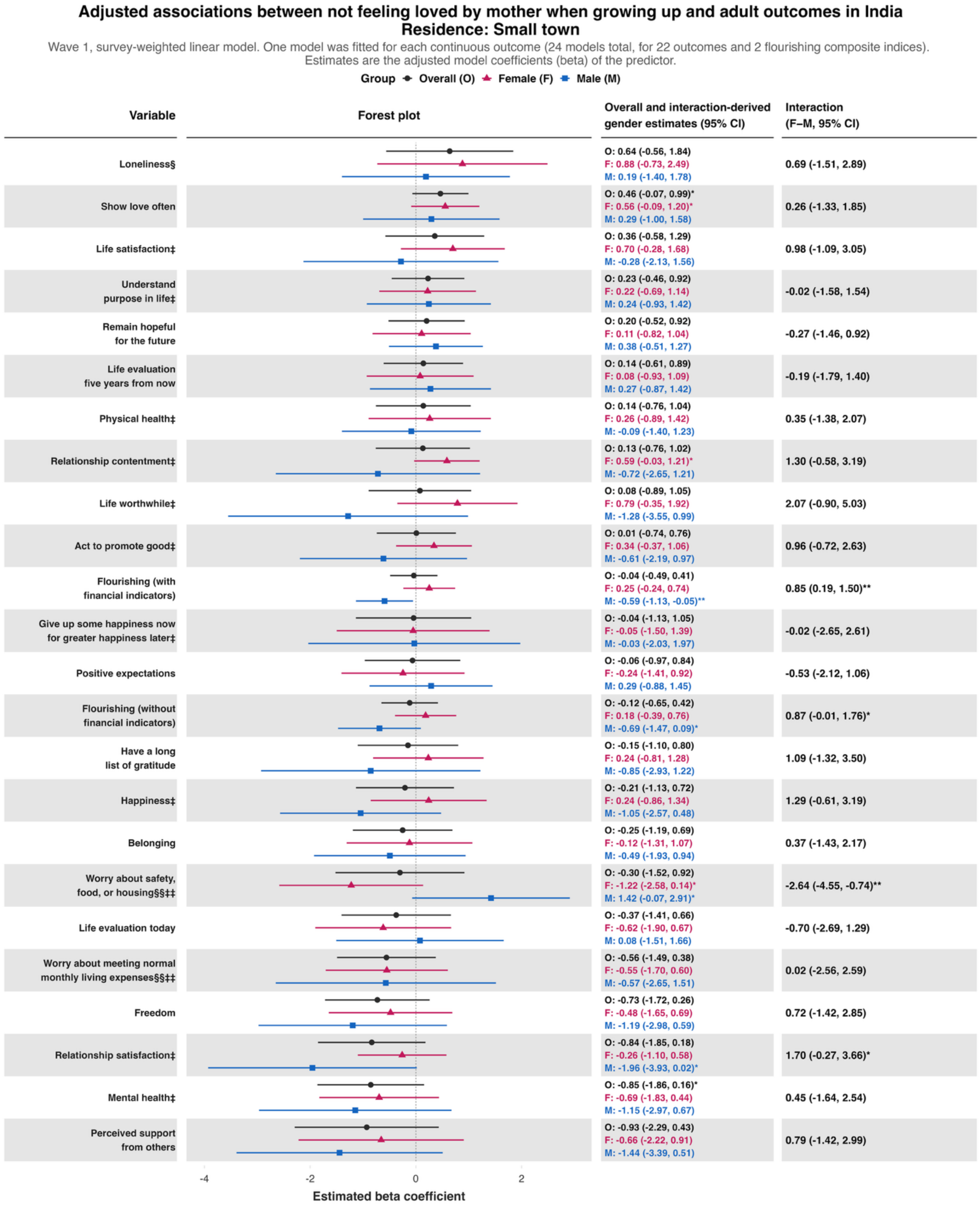

**Adjusted associations between not feeling loved by mother when growing up and adult outcomes in India**
**Residence: Small town**

Wave 1, survey-weighted logistic model. One model was fitted for each binary outcome (7 models total). Estimates are adjusted odds ratios (OR) of the predictor.
The interaction column reports the difference in log-odds ratios (interaction beta) with 95% confidence intervals (95% CIs).

Group: Overall (O), Female (F), Male (M)

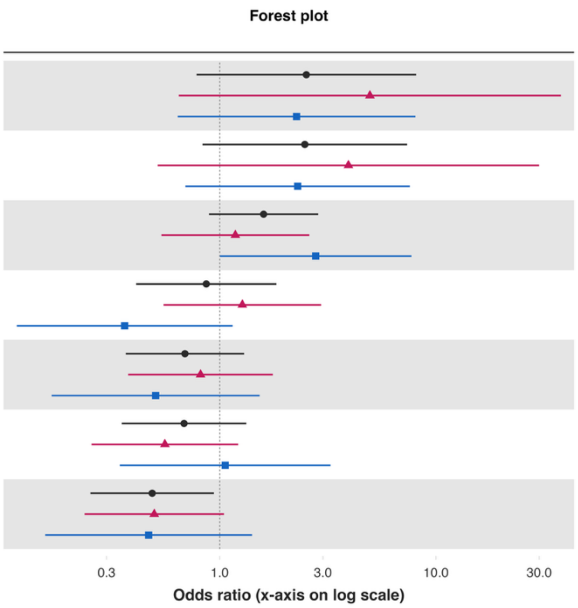


| Variable | Overall and interaction-derived gender estimates (95% CI) | Interaction (F–M, 95% CI) |
|---|---|---|
| Cigarette smoking | O: 2.51 (0.78, 8.09)<br>F: 4.95 (0.65, 37.79)<br>M: 2.27 (0.64, 8.03) | 0.78 (-1.55, 3.11) |
| Alcohol use | O: 2.47 (0.83, 7.35)<br>F: 3.94 (0.52, 29.96)<br>M: 2.29 (0.69, 7.57) | 0.54 (-1.79, 2.87) |
| Have any health problems | O: 1.60 (0.89, 2.85)<br>F: 1.18 (0.54, 2.60)<br>M: 2.78 (1.00, 7.69)* | -0.85 (-2.20, 0.49) |
| Volunteered at an organization | O: 0.87 (0.41, 1.83)<br>F: 1.27 (0.55, 2.94)<br>M: 0.36 (0.12, 1.15)* | 1.25 (-0.12, 2.63)* |
| Helped a stranger | O: 0.69 (0.37, 1.30)<br>F: 0.82 (0.38, 1.76)<br>M: 0.51 (0.17, 1.53) | 0.48 (-0.90, 1.85) |
| Feel close to someone | O: 0.68 (0.35, 1.33)<br>F: 0.56 (0.26, 1.22)<br>M: 1.06 (0.35, 3.25) | -0.64 (-1.98, 0.70) |
| Donated to charity | O: 0.49 (0.25, 0.94)**<br>F: 0.50 (0.24, 1.05)*<br>M: 0.47 (0.16, 1.41) | 0.06 (-1.19, 1.31) |

### Adjusted associations between not feeling loved by mother when growing up and adult outcomes in India
### Residence: Small town

Wave 1, survey-weighted ordinal regression using the proportional odds model (`svyolr`). One model was fitted for each ordinal outcome (16 models total).
Estimates are adjusted cumulative odds ratios (cOR) of the predictor.
The interaction column reports the difference in log-cumulative odds ratios (interaction beta) with 95% confidence intervals (95% CIs).

**Group** Overall (O) Female (F) Male (M)

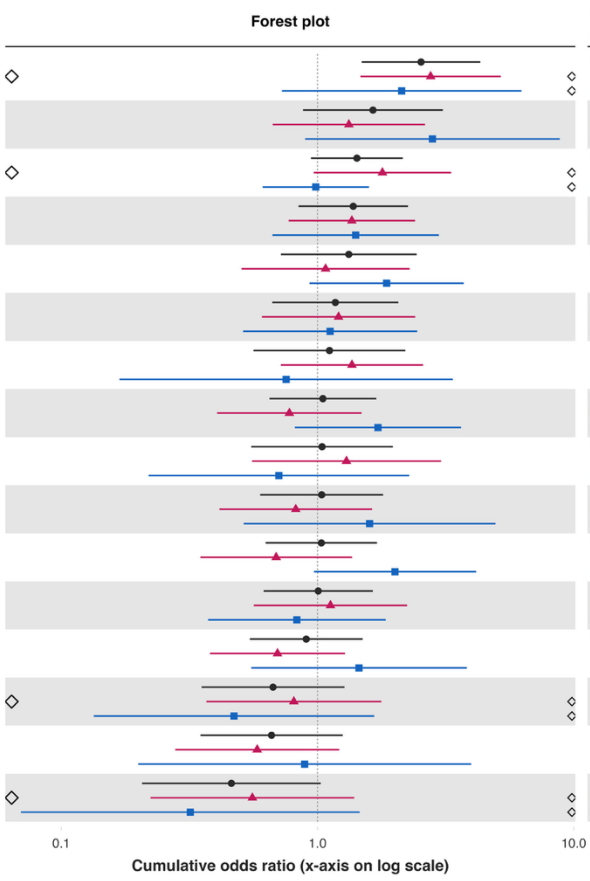


| Variable | Overall and interaction-derived gender estimates (95% CI) | Interaction (F–M, 95% CI) |
|---|---|---|
| Various aspects of life are balanced§ | O: 2.54 (1.49, 4.32)***<br>F: 2.76 (1.47, 5.19)**<br>M: 2.13 (0.73, 6.25) | 0.26 (-1.01, 1.53) |
| Feeling capable§ | O: 1.65 (0.88, 3.09)<br>F: 1.33 (0.67, 2.63)<br>M: 2.81 (0.89, 8.82)* | -0.75 (-2.01, 0.51) |
| Have little interest in doing things§ | O: 1.42 (0.94, 2.15)*<br>F: 1.79 (0.97, 3.32)*<br>M: 0.98 (0.61, 1.59) | 0.60 (-0.22, 1.42) |
| Trust in others§ | O: 1.38 (0.84, 2.25)<br>F: 1.36 (0.77, 2.40)<br>M: 1.41 (0.67, 2.98) | -0.03 (-0.89, 0.82) |
| Extent of suffering§ | O: 1.32 (0.72, 2.44)<br>F: 1.07 (0.51, 2.29)<br>M: 1.86 (0.93, 3.72)* | -0.55 (-1.45, 0.36) |
| Distress from e-threatening experiences§ | O: 1.17 (0.67, 2.07)<br>F: 1.21 (0.61, 2.41)<br>M: 1.12 (0.51, 2.45) | 0.08 (-0.86, 1.01) |
| Forgiveness§ | O: 1.11 (0.56, 2.20)<br>F: 1.36 (0.72, 2.58)<br>M: 0.75 (0.17, 3.38) | 0.59 (-0.97, 2.16) |
| Depressive symptoms§ | O: 1.05 (0.65, 1.70)<br>F: 0.78 (0.41, 1.49)<br>M: 1.72 (0.82, 3.63) | -0.80 (-1.90, 0.30) |
| Government approval§ | O: 1.04 (0.55, 1.97)<br>F: 1.30 (0.55, 3.03)<br>M: 0.71 (0.22, 2.28) | 0.61 (-0.95, 2.16) |
| Perceived discrimination§ | O: 1.04 (0.60, 1.80)<br>F: 0.82 (0.41, 1.63)<br>M: 1.60 (0.52, 4.95) | -0.66 (-2.10, 0.77) |
| Feeling nervous or anxious§ | O: 1.03 (0.63, 1.71)<br>F: 0.69 (0.35, 1.36)<br>M: 2.01 (0.97, 4.16)* | -1.07 (-2.11, -0.02)** |
| Feel at peace§ | O: 1.01 (0.62, 1.64)<br>F: 1.12 (0.56, 2.24)<br>M: 0.83 (0.37, 1.85) | 0.30 (-0.84, 1.45) |
| Unable to control worry§ | O: 0.90 (0.54, 1.50)<br>F: 0.70 (0.38, 1.28)<br>M: 1.45 (0.55, 3.83) | -0.73 (-1.96, 0.49) |
| Support reducing fferences in income levels§ | O: 0.67 (0.35, 1.27)<br>F: 0.81 (0.37, 1.77)<br>M: 0.47 (0.13, 1.66) | 0.54 (-1.02, 2.10) |
| Say in government# | O: 0.66 (0.35, 1.25)<br>F: 0.58 (0.28, 1.21)<br>M: 0.89 (0.20, 3.98) | -0.43 (-2.18, 1.33) |
| Obeying the law§ | O: 0.46 (0.21, 1.03)*<br>F: 0.56 (0.22, 1.39)<br>M: 0.32 (0.07, 1.46) | 0.56 (-1.24, 2.35) |

*Not feeling loved by mother – Large city or suburb of a large city*

**Adjusted associations between not feeling loved by mother when growing up and adult outcomes in India**
**Residence: Large city or suburb**

Wave 1, survey-weighted linear model. One model was fitted for each continuous outcome (24 models total, for 22 outcomes and 2 flourishing composite indices). Estimates are the adjusted model coefficients (beta) of the predictor.

Group: Overall (O), Female (F), Male (M)

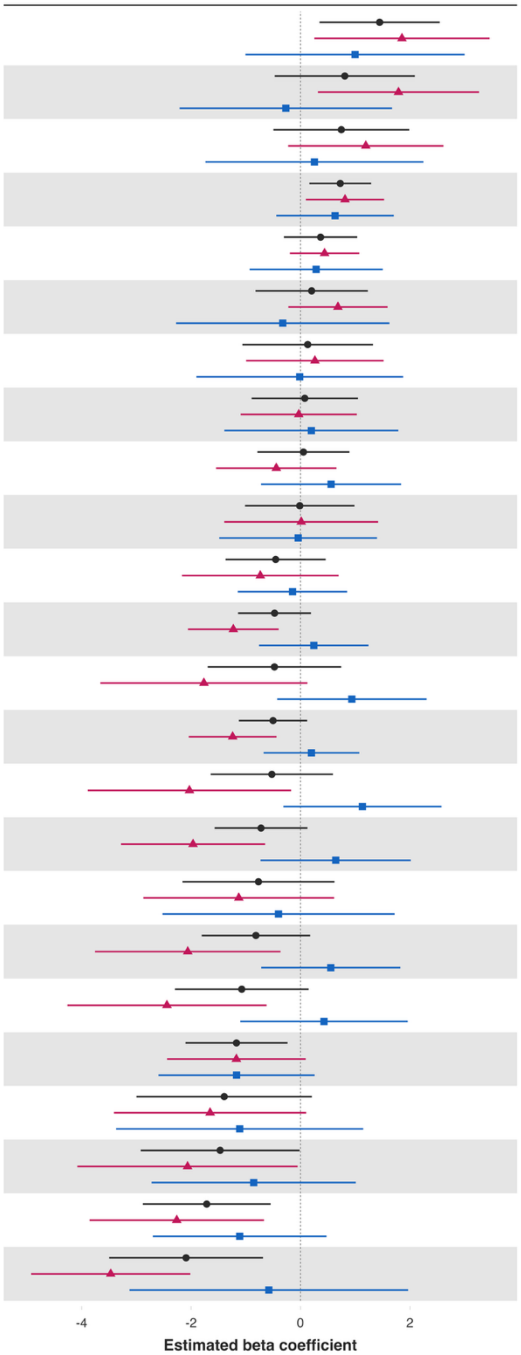


| Variable | Overall and interaction-derived gender estimates (95% CI) | Interaction (F–M, 95% CI) |
|---|---|---|
| Loneliness§ | O: 1.45 (0.35, 2.54)**<br>F: 1.85 (0.25, 3.45)**<br>M: 1.00 (-1.00, 3.00) | 0.86 (-2.04, 3.75) |
| Worry about meeting normal monthly living expenses§§‡‡ | O: 0.81 (-0.47, 2.09)<br>F: 1.79 (0.32, 3.26)**<br>M: -0.27 (-2.21, 1.67) | 2.06 (-0.23, 4.35)* |
| Worry about safety, food, or housing§§‡‡ | O: 0.75 (-0.50, 1.99)<br>F: 1.19 (-0.23, 2.61)<br>M: 0.25 (-1.74, 2.25) | 0.94 (-1.46, 3.33) |
| Remain hopeful for the future | O: 0.73 (0.16, 1.29)**<br>F: 0.81 (0.10, 1.53)**<br>M: 0.63 (-0.44, 1.70) | 0.18 (-1.24, 1.61) |
| Act to promote good‡ | O: 0.37 (-0.30, 1.04)<br>F: 0.44 (-0.19, 1.07)<br>M: 0.29 (-0.93, 1.50) | 0.15 (-1.21, 1.52) |
| Freedom | O: 0.20 (-0.82, 1.23)<br>F: 0.68 (-0.22, 1.59)<br>M: -0.32 (-2.27, 1.62) | 1.01 (-1.28, 3.29) |
| Give up some happiness now for greater happiness later‡ | O: 0.13 (-1.06, 1.32)<br>F: 0.26 (-0.99, 1.52)<br>M: -0.01 (-1.90, 1.87) | 0.28 (-1.82, 2.38) |
| Have a long list of gratitude | O: 0.08 (-0.89, 1.05)<br>F: -0.03 (-1.09, 1.03)<br>M: 0.20 (-1.39, 1.79) | -0.23 (-2.07, 1.61) |
| Understand purpose in life‡ | O: 0.05 (-0.79, 0.89)<br>F: -0.44 (-1.54, 0.66)<br>M: 0.56 (-0.72, 1.84) | -1.00 (-2.74, 0.73) |
| Positive expectations | O: -0.01 (-1.01, 0.98)<br>F: 0.01 (-1.39, 1.42)<br>M: -0.04 (-1.48, 1.40) | 0.06 (-1.97, 2.08) |
| Belonging | O: -0.45 (-1.37, 0.46)<br>F: -0.73 (-2.17, 0.70)<br>M: -0.15 (-1.14, 0.85) | -0.59 (-2.27, 1.10) |
| Flourishing (without financial indicators) | O: -0.47 (-1.14, 0.19)<br>F: -1.23 (-2.06, -0.40)**<br>M: 0.24 (-0.76, 1.24) | -1.47 (-2.83, -0.11)** |
| Life satisfaction‡ | O: -0.48 (-1.70, 0.74)<br>F: -1.77 (-3.66, 0.13)*<br>M: 0.94 (-0.43, 2.30) | -2.70 (-4.96, -0.45)** |
| Flourishing (with financial indicators) | O: -0.50 (-1.13, 0.12)<br>F: -1.24 (-2.04, -0.44)**<br>M: 0.20 (-0.67, 1.07) | -1.44 (-2.66, -0.22)** |
| Happiness‡ | O: -0.52 (-1.64, 0.59)<br>F: -2.03 (-3.89, -0.17)**<br>M: 1.13 (-0.31, 2.57) | -3.16 (-5.71, -0.61)** |
| Physical health‡ | O: -0.72 (-1.57, 0.13)*<br>F: -1.96 (-3.28, -0.65)**<br>M: 0.64 (-0.73, 2.01) | -2.61 (-4.73, -0.48)** |
| Show love often | O: -0.77 (-2.16, 0.62)<br>F: -1.13 (-2.87, 0.61)<br>M: -0.40 (-2.52, 1.72) | -0.73 (-3.46, 2.00) |
| Mental health‡ | O: -0.81 (-1.80, 0.17)<br>F: -2.06 (-3.76, -0.37)**<br>M: 0.55 (-0.72, 1.82) | -2.61 (-4.93, -0.29)** |
| Relationship contentment‡ | O: -1.07 (-2.29, 0.15)*<br>F: -2.44 (-4.26, -0.62)**<br>M: 0.43 (-1.10, 1.96) | -2.87 (-5.45, -0.29)** |
| Life evaluation five years from now | O: -1.17 (-2.10, -0.24)**<br>F: -1.17 (-2.44, 0.09)*<br>M: -1.17 (-2.59, 0.26) | -0.00 (-1.99, 1.98) |
| Life worthwhile‡ | O: -1.39 (-3.00, 0.21)*<br>F: -1.65 (-3.41, 0.10)*<br>M: -1.11 (-3.37, 1.15) | -0.54 (-2.99, 1.91) |
| Relationship satisfaction‡ | O: -1.47 (-2.92, -0.02)**<br>F: -2.06 (-4.08, -0.05)**<br>M: -0.86 (-2.72, 1.01) | -1.21 (-3.86, 1.44) |
| Life evaluation today | O: -1.71 (-2.88, -0.55)**<br>F: -2.26 (-3.85, -0.67)**<br>M: -1.11 (-2.70, 0.48) | -1.15 (-3.39, 1.09) |
| Perceived support from others | O: -2.09 (-3.50, -0.69)**<br>F: -3.47 (-4.92, -2.01)***<br>M: -0.58 (-3.12, 1.97) | -2.89 (-6.06, 0.28)* |

### Adjusted associations between not feeling loved by mother when growing up and adult outcomes in India
### Residence: Large city or suburb

Wave 1, survey-weighted logistic model. One model was fitted for each binary outcome (7 models total). Estimates are adjusted odds ratios (OR) of the predictor.
The interaction column reports the difference in log-odds ratios (interaction beta) with 95% confidence intervals (95% CIs).

**Group** Overall (O) Female (F) Male (M)

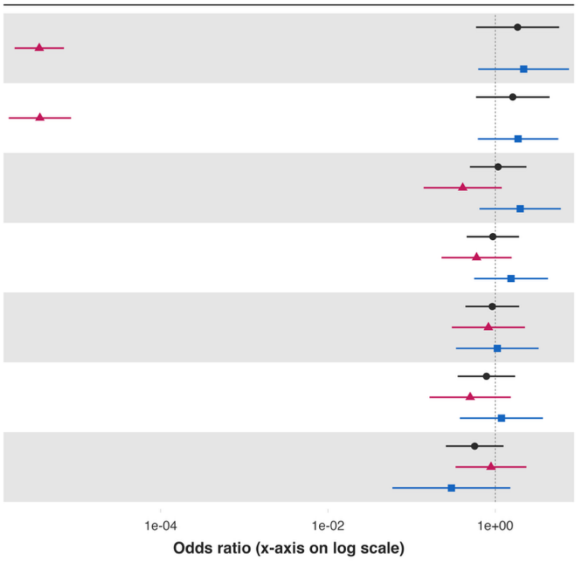


| Variable | Overall and interaction-derived gender estimates (95% CI) | Interaction (F–M, 95% CI) |
|---|---|---|
| Alcohol use | O: 1.85 (0.59, 5.81)<br>F: 0.00 (0.00, 0.00)***<br>M: 2.18 (0.63, 7.61) | -13.33 (-14.81, -11.85)*** |
| Cigarette smoking | O: 1.62 (0.59, 4.45)<br>F: 0.00 (0.00, 0.00)***<br>M: 1.88 (0.62, 5.67) | -13.16 (-14.59, -11.73)*** |
| Volunteered at an organization | O: 1.08 (0.50, 2.36)<br>F: 0.41 (0.14, 1.20)<br>M: 1.99 (0.65, 6.08) | -1.58 (-3.08, -0.08)** |
| Helped a stranger | O: 0.94 (0.45, 1.92)<br>F: 0.60 (0.23, 1.57)<br>M: 1.54 (0.56, 4.27) | -0.95 (-2.23, 0.33) |
| Feel close to someone | O: 0.92 (0.44, 1.93)<br>F: 0.83 (0.30, 2.27)<br>M: 1.06 (0.34, 3.30) | -0.25 (-1.80, 1.31) |
| Donated to charity | O: 0.78 (0.36, 1.73)<br>F: 0.50 (0.16, 1.53)<br>M: 1.18 (0.38, 3.71) | -0.86 (-2.43, 0.71) |
| Have any health problems | O: 0.57 (0.26, 1.26)<br>F: 0.89 (0.34, 2.35)<br>M: 0.30 (0.06, 1.51) | 1.09 (-0.85, 3.03) |

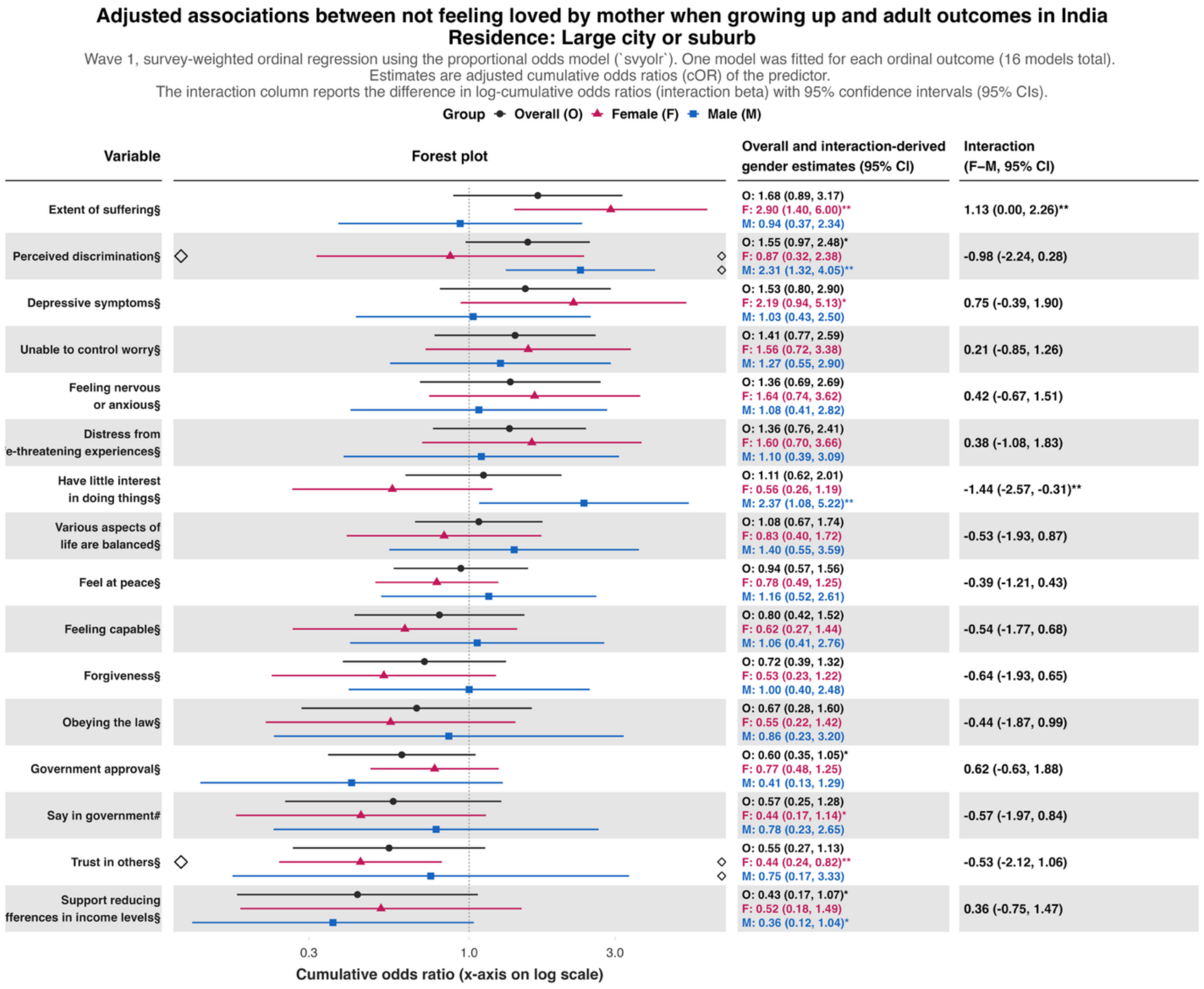
Adjusted associations between not feeling loved by mother when growing up and adult outcomes in India
Residence: Large city or suburb
Wave 1, survey-weighted ordinal regression using the proportional odds model (`svyolr`). One model was fitted for each ordinal outcome (16 models total).
Estimates are adjusted cumulative odds ratios (cOR) of the predictor.
The interaction column reports the difference in log-cumulative odds ratios (interaction beta) with 95% confidence intervals (95% CIs).
Group Overall (O) Female (F) Male (M)
Variable
Forest plot
Overall and interaction-derived gender estimates (95% CI)
Interaction (F–M, 95% CI)
Extent of suffering§
O: 1.68 (0.89, 3.17)
F: 2.90 (1.40, 6.00)**
M: 0.94 (0.37, 2.34)
1.13 (0.00, 2.26)**
Perceived discrimination§
O: 1.55 (0.97, 2.48)*
F: 0.87 (0.32, 2.38)
M: 2.31 (1.32, 4.05)**
-0.98 (-2.24, 0.28)
Depressive symptoms§
O: 1.53 (0.80, 2.90)
F: 2.19 (0.94, 5.13)*
M: 1.03 (0.43, 2.50)
0.75 (-0.39, 1.90)
Unable to control worry§
O: 1.41 (0.77, 2.59)
F: 1.56 (0.72, 3.38)
M: 1.27 (0.55, 2.90)
0.21 (-0.85, 1.26)
Feeling nervous or anxious§
O: 1.36 (0.69, 2.69)
F: 1.64 (0.74, 3.62)
M: 1.08 (0.41, 2.82)
0.42 (-0.67, 1.51)
Distress from e-threatening experiences§
O: 1.36 (0.76, 2.41)
F: 1.60 (0.70, 3.66)
M: 1.10 (0.39, 3.09)
0.38 (-1.08, 1.83)
Have little interest in doing things§
O: 1.11 (0.62, 2.01)
F: 0.56 (0.26, 1.19)
M: 2.37 (1.08, 5.22)**
-1.44 (-2.57, -0.31)**
Various aspects of life are balanced§
O: 1.08 (0.67, 1.74)
F: 0.83 (0.40, 1.72)
M: 1.40 (0.55, 3.59)
-0.53 (-1.93, 0.87)
Feel at peace§
O: 0.94 (0.57, 1.56)
F: 0.78 (0.49, 1.25)
M: 1.16 (0.52, 2.61)
-0.39 (-1.21, 0.43)
Feeling capable§
O: 0.80 (0.42, 1.52)
F: 0.62 (0.27, 1.44)
M: 1.06 (0.41, 2.76)
-0.54 (-1.77, 0.68)
Forgiveness§
O: 0.72 (0.39, 1.32)
F: 0.53 (0.23, 1.22)
M: 1.00 (0.40, 2.48)
-0.64 (-1.93, 0.65)
Obeying the law§
O: 0.67 (0.28, 1.60)
F: 0.55 (0.22, 1.42)
M: 0.86 (0.23, 3.20)
-0.44 (-1.87, 0.99)
Government approval§
O: 0.60 (0.35, 1.05)*
F: 0.77 (0.48, 1.25)
M: 0.41 (0.13, 1.29)
0.62 (-0.63, 1.88)
Say in government#
O: 0.57 (0.25, 1.28)
F: 0.44 (0.17, 1.14)*
M: 0.78 (0.23, 2.65)
-0.57 (-1.97, 0.84)
Trust in others§
O: 0.55 (0.27, 1.13)
F: 0.44 (0.24, 0.82)**
M: 0.75 (0.17, 3.33)
-0.53 (-2.12, 1.06)
Support reducing fferences in income levels§
O: 0.43 (0.17, 1.07)*
F: 0.52 (0.18, 1.49)
M: 0.36 (0.12, 1.04)*
0.36 (-0.75, 1.47)
0.3
1.0
3.0
Cumulative odds ratio (x-axis on log scale)

**Predictor:** ***Not feeling loved by father***

***Not feeling loved by father – Rural area or on a farm***

**Adjusted associations between not feeling loved by father when growing up and adult outcomes in India**
**Residence: Rural**

Wave 1, survey-weighted linear model. One model was fitted for each continuous outcome (24 models total, for 22 outcomes and 2 flourishing composite indices).
Estimates are the adjusted model coefficients (beta) of the predictor.

Group ● Overall (O) ▲ Female (F) ■ Male (M)

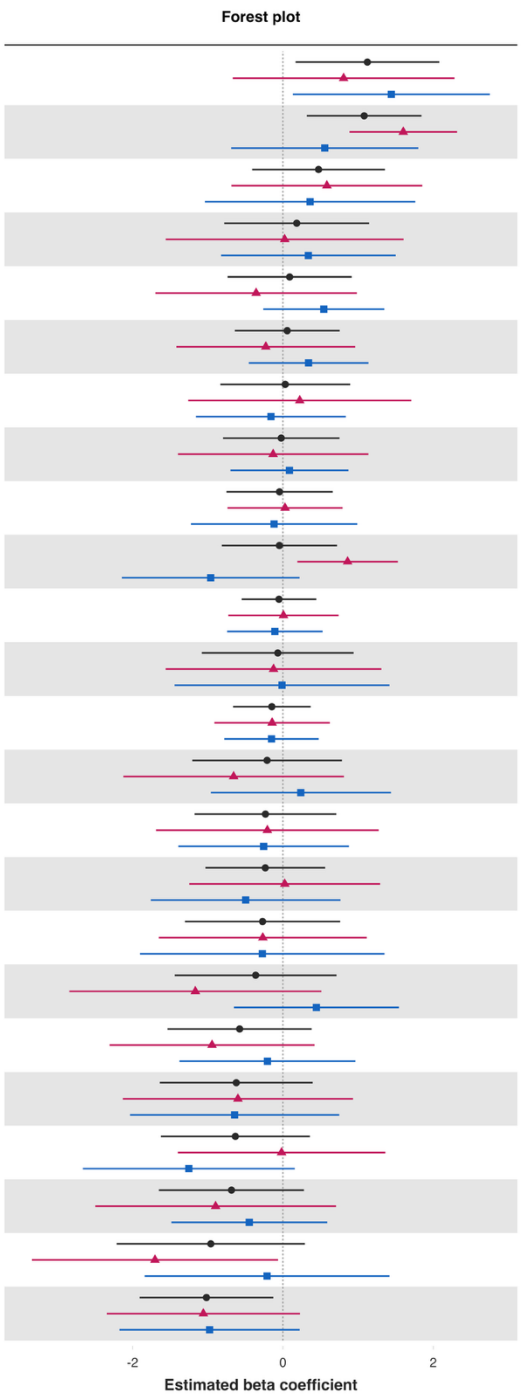


| Variable | Overall and interaction-derived gender estimates (95% CI) | Interaction (F–M, 95% CI) |
|---|---|---|
| Loneliness§ | O: 1.12 (0.17, 2.08)**<br>F: 0.81 (-0.67, 2.28)<br>M: 1.44 (0.13, 2.75)** | -0.64 (-2.66, 1.39) |
| Life worthwhile‡ | O: 1.08 (0.32, 1.84)**<br>F: 1.60 (0.89, 2.32)***<br>M: 0.56 (-0.69, 1.80) | 1.05 (-0.28, 2.37) |
| Worry about meeting normal monthly living expenses§§‡‡ | O: 0.47 (-0.41, 1.36)<br>F: 0.58 (-0.69, 1.85)<br>M: 0.36 (-1.04, 1.76) | 0.22 (-1.79, 2.24) |
| Physical health‡ | O: 0.18 (-0.78, 1.15)<br>F: 0.02 (-1.56, 1.60)<br>M: 0.34 (-0.82, 1.50) | -0.32 (-2.32, 1.69) |
| Positive expectations | O: 0.09 (-0.74, 0.91)<br>F: -0.36 (-1.70, 0.98)<br>M: 0.54 (-0.26, 1.35) | -0.90 (-2.40, 0.59) |
| Understand purpose in life‡ | O: 0.06 (-0.64, 0.75)<br>F: -0.23 (-1.42, 0.96)<br>M: 0.34 (-0.45, 1.14) | -0.57 (-2.05, 0.91) |
| Life evaluation today | O: 0.03 (-0.83, 0.89)<br>F: 0.22 (-1.26, 1.71)<br>M: -0.16 (-1.16, 0.84) | 0.38 (-1.45, 2.22) |
| Act to promote good‡ | O: -0.02 (-0.80, 0.75)<br>F: -0.13 (-1.40, 1.14)<br>M: 0.09 (-0.70, 0.87) | -0.22 (-1.65, 1.22) |
| Remain hopeful for the future | O: -0.05 (-0.75, 0.66)<br>F: 0.03 (-0.74, 0.79)<br>M: -0.12 (-1.22, 0.99) | 0.15 (-1.13, 1.42) |
| Show love often | O: -0.05 (-0.81, 0.72)<br>F: 0.86 (0.19, 1.53)**<br>M: -0.96 (-2.14, 0.22) | 1.82 (0.55, 3.09)** |
| Flourishing (with financial indicators) | O: -0.05 (-0.55, 0.44)<br>F: 0.01 (-0.73, 0.74)<br>M: -0.11 (-0.74, 0.53) | 0.11 (-0.82, 1.05) |
| Happiness‡ | O: -0.07 (-1.08, 0.94)<br>F: -0.13 (-1.56, 1.31)<br>M: -0.01 (-1.44, 1.42) | -0.11 (-2.16, 1.93) |
| Flourishing (without financial indicators) | O: -0.15 (-0.66, 0.37)<br>F: -0.14 (-0.91, 0.62)<br>M: -0.15 (-0.78, 0.48) | 0.01 (-0.94, 0.95) |
| Life satisfaction‡ | O: -0.21 (-1.21, 0.79)<br>F: -0.66 (-2.12, 0.81)<br>M: 0.24 (-0.96, 1.44) | -0.89 (-2.77, 0.99) |
| Life evaluation five years from now | O: -0.23 (-1.18, 0.71)<br>F: -0.21 (-1.69, 1.27)<br>M: -0.26 (-1.39, 0.88) | 0.05 (-1.79, 1.89) |
| Give up some happiness now for greater happiness later‡ | O: -0.24 (-1.03, 0.56)<br>F: 0.02 (-1.25, 1.29)<br>M: -0.50 (-1.76, 0.77) | 0.52 (-1.43, 2.47) |
| Worry about safety, food, or housing§§‡‡ | O: -0.27 (-1.30, 0.76)<br>F: -0.27 (-1.65, 1.12)<br>M: -0.28 (-1.90, 1.35) | 0.01 (-2.20, 2.21) |
| Mental health‡ | O: -0.36 (-1.44, 0.71)<br>F: -1.17 (-2.84, 0.51)<br>M: 0.45 (-0.65, 1.54) | -1.61 (-3.61, 0.39) |
| Have a long list of gratitude | O: -0.58 (-1.54, 0.38)<br>F: -0.94 (-2.31, 0.42)<br>M: -0.21 (-1.38, 0.96) | -0.74 (-2.45, 0.97) |
| Relationship satisfaction‡ | O: -0.62 (-1.64, 0.40)<br>F: -0.60 (-2.13, 0.93)<br>M: -0.64 (-2.04, 0.75) | 0.04 (-2.06, 2.15) |
| Relationship contentment‡ | O: -0.63 (-1.62, 0.36)<br>F: -0.02 (-1.40, 1.36)<br>M: -1.25 (-2.66, 0.16)* | 1.23 (-0.69, 3.16) |
| Belonging | O: -0.68 (-1.65, 0.28)<br>F: -0.90 (-2.50, 0.71)<br>M: -0.45 (-1.49, 0.59) | -0.45 (-2.40, 1.50) |
| Perceived support from others | O: -0.96 (-2.21, 0.29)<br>F: -1.70 (-3.34, -0.06)**<br>M: -0.21 (-1.84, 1.42) | -1.49 (-3.70, 0.71) |
| Freedom | O: -1.02 (-1.91, -0.13)**<br>F: -1.06 (-2.34, 0.23)<br>M: -0.98 (-2.18, 0.22) | -0.08 (-1.82, 1.66) |

**Adjusted associations between not feeling loved by father when growing up and adult outcomes in India**
**Residence: Rural**

Wave 1, survey-weighted ordinal regression using the proportional odds model (`svyolr`). One model was fitted for each ordinal outcome (16 models total).
Estimates are adjusted cumulative odds ratios (cOR) of the predictor.
The interaction column reports the difference in log-cumulative odds ratios (interaction beta) with 95% confidence intervals (95% CIs).

Group ● Overall (O) ▲ Female (F) ■ Male (M)

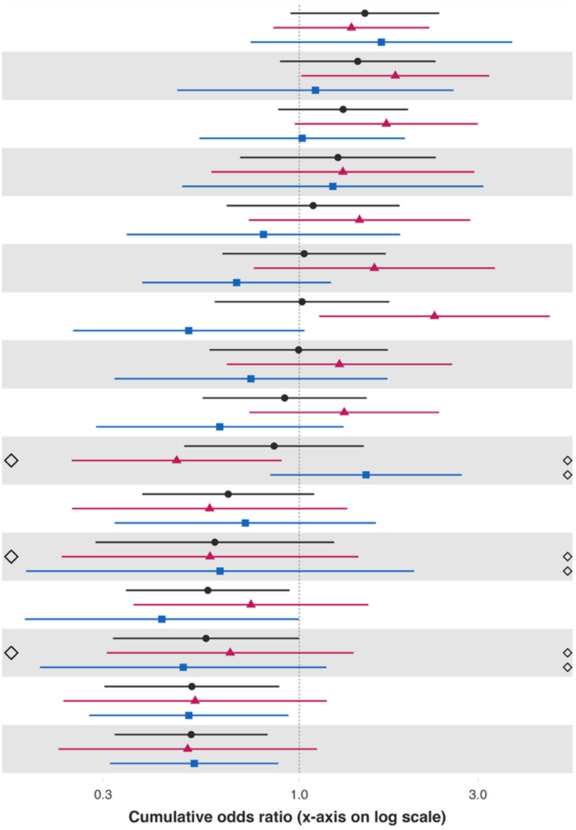


| Variable | Overall and interaction-derived gender estimates (95% CI) | Interaction (F–M, 95% CI) |
|---|---|---|
| Feeling nervous or anxious§ | O: 1.50 (0.95, 2.36)*<br>F: 1.38 (0.85, 2.22)<br>M: 1.66 (0.74, 3.70) | -0.18 (-1.10, 0.73) |
| Depressive symptoms§ | O: 1.43 (0.89, 2.31)<br>F: 1.80 (1.01, 3.21)**<br>M: 1.10 (0.47, 2.58) | 0.49 (-0.59, 1.57) |
| Distress from 'e-threatening experiences§ | O: 1.31 (0.88, 1.95)<br>F: 1.71 (0.97, 3.00)*<br>M: 1.02 (0.54, 1.91) | 0.52 (-0.37, 1.41) |
| Extent of suffering§ | O: 1.27 (0.70, 2.31)<br>F: 1.31 (0.58, 2.93)<br>M: 1.23 (0.49, 3.10) | 0.06 (-1.18, 1.31) |
| Have little interest in doing things§ | O: 1.09 (0.64, 1.85)<br>F: 1.45 (0.73, 2.86)<br>M: 0.80 (0.35, 1.86) | 0.59 (-0.48, 1.66) |
| Feel at peace§ | O: 1.03 (0.62, 1.70)<br>F: 1.59 (0.76, 3.33)<br>M: 0.68 (0.38, 1.22) | 0.85 (-0.06, 1.75)* |
| Feeling capable§ | O: 1.02 (0.60, 1.74)<br>F: 2.29 (1.13, 4.66)**<br>M: 0.51 (0.25, 1.03)* | 1.51 (0.54, 2.48)** |
| Unable to control worry§ | O: 1.00 (0.58, 1.72)<br>F: 1.28 (0.64, 2.56)<br>M: 0.74 (0.32, 1.72) | 0.54 (-0.52, 1.61) |
| Perceived discrimination§ | O: 0.91 (0.55, 1.51)<br>F: 1.32 (0.74, 2.36)<br>M: 0.61 (0.29, 1.31) | 0.76 (-0.16, 1.68) |
| Trust in others§ | O: 0.86 (0.49, 1.49)<br>F: 0.47 (0.25, 0.90)**<br>M: 1.51 (0.84, 2.71) | -1.16 (-1.96, -0.37)** |
| Say in government# | O: 0.65 (0.38, 1.10)<br>F: 0.58 (0.25, 1.34)<br>M: 0.72 (0.32, 1.60) | -0.22 (-1.49, 1.05) |
| Obeying the law§ | O: 0.60 (0.29, 1.24)<br>F: 0.58 (0.23, 1.44)<br>M: 0.61 (0.19, 2.02) | -0.06 (-1.58, 1.45) |
| Various aspects of life are balanced§ | O: 0.57 (0.35, 0.94)**<br>F: 0.74 (0.36, 1.53)<br>M: 0.43 (0.19, 1.00)** | 0.55 (-0.68, 1.77) |
| Support reducing fferences in income levels§ | O: 0.56 (0.32, 1.00)**<br>F: 0.65 (0.31, 1.40)<br>M: 0.49 (0.20, 1.18) | 0.29 (-0.92, 1.49) |
| Forgiveness§ | O: 0.52 (0.30, 0.88)**<br>F: 0.53 (0.23, 1.18)<br>M: 0.51 (0.28, 0.94)** | 0.04 (-0.90, 0.98) |
| Government approval§ | O: 0.51 (0.32, 0.82)**<br>F: 0.50 (0.23, 1.12)*<br>M: 0.52 (0.31, 0.88)** | -0.04 (-0.97, 0.89) |

*Not feeling loved by father – Small town or village*

**Adjusted associations between not feeling loved by father when growing up and adult outcomes in India**
**Residence: Small town**

Wave 1, survey-weighted linear model. One model was fitted for each continuous outcome (24 models total, for 22 outcomes and 2 flourishing composite indices).
Estimates are the adjusted model coefficients (beta) of the predictor.

Group ● Overall (O) ▲ Female (F) ■ Male (M)

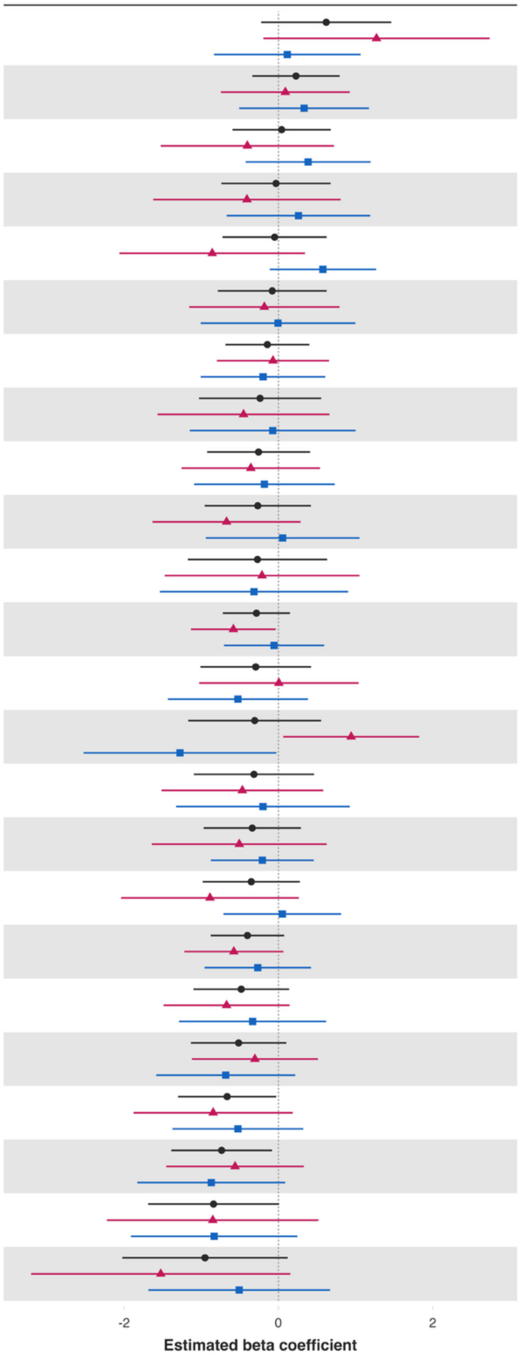


| Variable | Overall and interaction-derived gender estimates (95% CI) | Interaction (F–M, 95% CI) |
|---|---|---|
| Loneliness§ | O: 0.62 (-0.22, 1.46)<br>F: 1.27 (-0.19, 2.74)*<br>M: 0.12 (-0.83, 1.06) | 1.16 (-0.59, 2.90) |
| Have a long list of gratitude | O: 0.23 (-0.34, 0.79)<br>F: 0.09 (-0.75, 0.92)<br>M: 0.33 (-0.51, 1.17) | -0.24 (-1.49, 1.00) |
| Give up some happiness now for greater happiness later‡ | O: 0.04 (-0.59, 0.68)<br>F: -0.40 (-1.53, 0.72)<br>M: 0.39 (-0.42, 1.19) | -0.79 (-2.24, 0.66) |
| Physical health‡ | O: -0.03 (-0.74, 0.68)<br>F: -0.41 (-1.62, 0.81)<br>M: 0.26 (-0.67, 1.19) | -0.67 (-2.29, 0.95) |
| Show love often | O: -0.05 (-0.72, 0.63)<br>F: -0.86 (-2.06, 0.34)<br>M: 0.58 (-0.11, 1.27) | -1.43 (-2.85, -0.02)** |
| Life evaluation today | O: -0.08 (-0.79, 0.63)<br>F: -0.18 (-1.16, 0.79)<br>M: -0.00 (-1.01, 1.00) | -0.18 (-1.59, 1.23) |
| Remain hopeful for the future | O: -0.14 (-0.69, 0.40)<br>F: -0.07 (-0.80, 0.66)<br>M: -0.20 (-1.01, 0.61) | 0.13 (-0.98, 1.24) |
| Life worthwhile‡ | O: -0.24 (-1.03, 0.55)<br>F: -0.45 (-1.57, 0.66)<br>M: -0.07 (-1.15, 1.00) | -0.38 (-1.90, 1.15) |
| Understand purpose in life‡ | O: -0.26 (-0.92, 0.41)<br>F: -0.36 (-1.26, 0.54)<br>M: -0.18 (-1.09, 0.73) | -0.18 (-1.40, 1.05) |
| Happiness‡ | O: -0.27 (-0.96, 0.42)<br>F: -0.67 (-1.63, 0.29)<br>M: 0.05 (-0.94, 1.05) | -0.73 (-2.16, 0.70) |
| Worry about safety, food, or housing§§‡‡ | O: -0.27 (-1.17, 0.63)<br>F: -0.21 (-1.47, 1.05)<br>M: -0.32 (-1.54, 0.90) | 0.10 (-1.61, 1.82) |
| Flourishing (with financial indicators) | O: -0.28 (-0.72, 0.15)<br>F: -0.58 (-1.13, -0.03)**<br>M: -0.06 (-0.71, 0.59) | -0.53 (-1.40, 0.35) |
| Life evaluation five years from now | O: -0.29 (-1.01, 0.42)<br>F: 0.01 (-1.03, 1.04)<br>M: -0.52 (-1.43, 0.38) | 0.53 (-0.78, 1.84) |
| Worry about meeting normal monthly living expenses§§‡‡ | O: -0.31 (-1.17, 0.55)<br>F: 0.94 (0.06, 1.82)**<br>M: -1.28 (-2.53, -0.03)** | 2.22 (0.69, 3.75)** |
| Life satisfaction‡ | O: -0.32 (-1.10, 0.46)<br>F: -0.47 (-1.52, 0.58)<br>M: -0.20 (-1.33, 0.92) | -0.27 (-1.81, 1.28) |
| Freedom | O: -0.34 (-0.97, 0.29)<br>F: -0.51 (-1.64, 0.63)<br>M: -0.21 (-0.88, 0.46) | -0.30 (-1.60, 1.00) |
| Positive expectations | O: -0.35 (-0.98, 0.28)<br>F: -0.89 (-2.04, 0.27)<br>M: 0.05 (-0.71, 0.81) | -0.94 (-2.39, 0.51) |
| Flourishing (without financial indicators) | O: -0.40 (-0.88, 0.07)*<br>F: -0.58 (-1.22, 0.06)*<br>M: -0.27 (-0.96, 0.42) | -0.31 (-1.27, 0.65) |
| Relationship satisfaction‡ | O: -0.48 (-1.10, 0.14)<br>F: -0.67 (-1.49, 0.15)<br>M: -0.33 (-1.29, 0.62) | -0.34 (-1.66, 0.98) |
| Belonging | O: -0.52 (-1.13, 0.10)<br>F: -0.30 (-1.12, 0.51)<br>M: -0.68 (-1.58, 0.22) | 0.38 (-0.84, 1.60) |
| Relationship contentment‡ | O: -0.66 (-1.30, -0.03)**<br>F: -0.85 (-1.88, 0.19)<br>M: -0.52 (-1.37, 0.32) | -0.32 (-1.70, 1.06) |
| Act to promote good‡ | O: -0.74 (-1.39, -0.08)**<br>F: -0.56 (-1.45, 0.33)<br>M: -0.87 (-1.83, 0.09)* | 0.31 (-1.03, 1.64) |
| Mental health‡ | O: -0.84 (-1.69, 0.01)*<br>F: -0.85 (-2.22, 0.52)<br>M: -0.83 (-1.91, 0.25) | -0.02 (-1.77, 1.73) |
| Perceived support from others | O: -0.95 (-2.02, 0.12)*<br>F: -1.53 (-3.21, 0.15)*<br>M: -0.51 (-1.69, 0.67) | -1.02 (-2.92, 0.89) |

### Adjusted associations between not feeling loved by father when growing up and adult outcomes in India
### Residence: Small town

Wave 1, survey-weighted logistic model. One model was fitted for each binary outcome (7 models total). Estimates are adjusted odds ratios (OR) of the predictor.
The interaction column reports the difference in log-odds ratios (interaction beta) with 95% confidence intervals (95% CIs).

**Group** Overall (O) Female (F) Male (M)

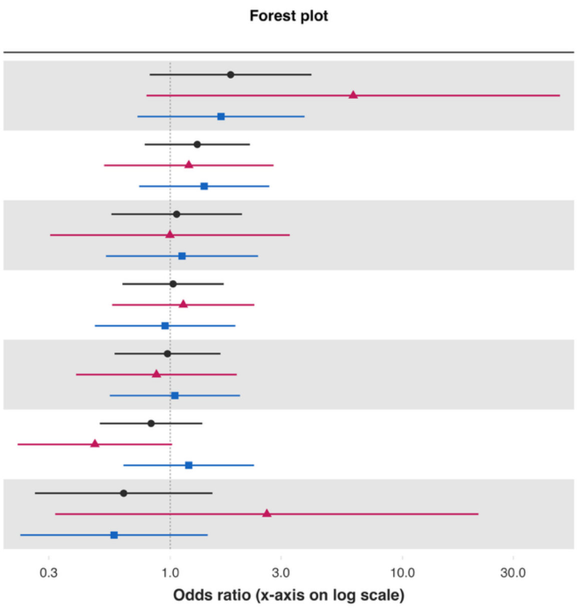


| Variable | Overall and interaction-derived gender estimates (95% CI) | Interaction (F–M, 95% CI) |
|---|---|---|
| Alcohol use | O: 1.82 (0.82, 4.05)<br>F: 6.14 (0.79, 47.65)*<br>M: 1.65 (0.72, 3.79) | 1.31 (-0.91, 3.53) |
| Have any health problems | O: 1.31 (0.78, 2.20)<br>F: 1.20 (0.52, 2.79)<br>M: 1.40 (0.73, 2.67) | -0.15 (-1.20, 0.90) |
| Feel close to someone | O: 1.07 (0.56, 2.04)<br>F: 1.00 (0.30, 3.27)<br>M: 1.12 (0.53, 2.39) | -0.12 (-1.59, 1.35) |
| Helped a stranger | O: 1.03 (0.62, 1.70)<br>F: 1.14 (0.56, 2.30)<br>M: 0.95 (0.47, 1.91) | 0.18 (-0.80, 1.16) |
| Volunteered at an organization | O: 0.97 (0.58, 1.64)<br>F: 0.87 (0.39, 1.93)<br>M: 1.05 (0.55, 2.00) | -0.18 (-1.15, 0.78) |
| Donated to charity | O: 0.83 (0.50, 1.37)<br>F: 0.47 (0.22, 1.02)*<br>M: 1.20 (0.63, 2.29) | -0.93 (-1.91, 0.05)* |
| Cigarette smoking | O: 0.63 (0.26, 1.52)<br>F: 2.61 (0.32, 21.24)<br>M: 0.57 (0.23, 1.45) | 1.51 (-0.77, 3.80) |

### Adjusted associations between not feeling loved by father when growing up and adult outcomes in India
### Residence: Small town

Wave 1, survey-weighted ordinal regression using the proportional odds model (`svyolr`). One model was fitted for each ordinal outcome (16 models total).
Estimates are adjusted cumulative odds ratios (cOR) of the predictor.
The interaction column reports the difference in log-cumulative odds ratios (interaction beta) with 95% confidence intervals (95% CIs).

Group: Overall (O), Female (F), Male (M)

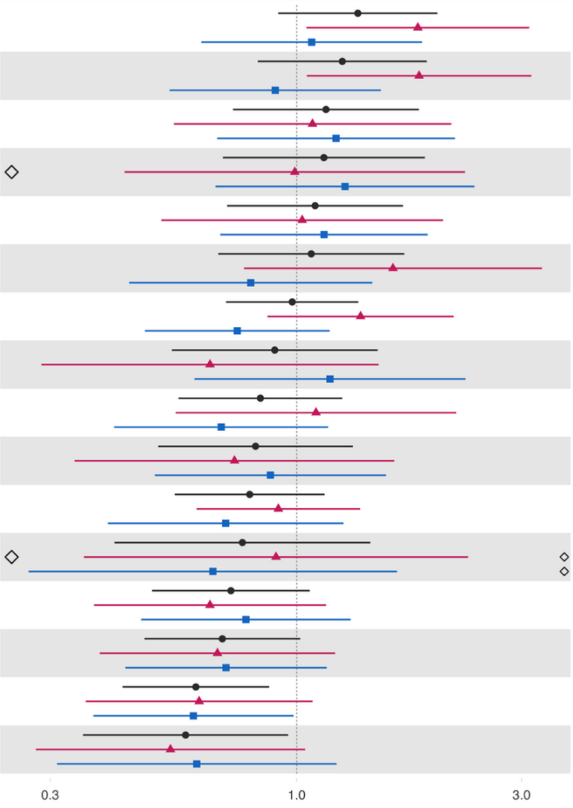


| Variable | Overall and interaction-derived gender estimates (95% CI) | Interaction (F–M, 95% CI) |
|---|---|---|
| Depressive symptoms§ | O: 1.35 (0.91, 1.99)<br>F: 1.81 (1.05, 3.11)**<br>M: 1.08 (0.63, 1.84) | 0.52 (-0.24, 1.28) |
| Unable to control worry§ | O: 1.25 (0.83, 1.89)<br>F: 1.82 (1.05, 3.14)**<br>M: 0.90 (0.54, 1.51) | 0.70 (-0.01, 1.42)* |
| Perceived discrimination§ | O: 1.15 (0.73, 1.82)<br>F: 1.08 (0.55, 2.13)<br>M: 1.21 (0.68, 2.17) | -0.12 (-0.98, 0.75) |
| Various aspects of life are balanced§ | O: 1.14 (0.70, 1.87)<br>F: 0.99 (0.43, 2.27)<br>M: 1.27 (0.67, 2.38) | -0.25 (-1.31, 0.82) |
| Extent of suffering§ | O: 1.09 (0.71, 1.68)<br>F: 1.03 (0.52, 2.04)<br>M: 1.14 (0.69, 1.90) | -0.11 (-0.92, 0.71) |
| Distress from e-threatening experiences§ | O: 1.07 (0.68, 1.69)<br>F: 1.60 (0.77, 3.31)<br>M: 0.80 (0.44, 1.45) | 0.69 (-0.26, 1.65) |
| Feeling nervous or anxious§ | O: 0.98 (0.71, 1.35)<br>F: 1.37 (0.87, 2.15)<br>M: 0.75 (0.48, 1.18) | 0.60 (-0.05, 1.25)* |
| Support reducing fferences in income levels§ | O: 0.90 (0.54, 1.48)<br>F: 0.65 (0.29, 1.49)<br>M: 1.18 (0.61, 2.28) | -0.59 (-1.68, 0.51) |
| Feeling capable§ | O: 0.84 (0.56, 1.25)<br>F: 1.10 (0.55, 2.18)<br>M: 0.69 (0.41, 1.17) | 0.46 (-0.44, 1.36) |
| Forgiveness§ | O: 0.82 (0.51, 1.32)<br>F: 0.74 (0.34, 1.61)<br>M: 0.88 (0.50, 1.55) | -0.18 (-1.12, 0.77) |
| Have little interest in doing things§ | O: 0.79 (0.55, 1.15)<br>F: 0.91 (0.61, 1.36)<br>M: 0.71 (0.40, 1.26) | 0.26 (-0.43, 0.95) |
| Obeying the law§ | O: 0.77 (0.41, 1.43)<br>F: 0.90 (0.35, 2.31)<br>M: 0.66 (0.27, 1.63) | 0.31 (-1.05, 1.67) |
| Trust in others§ | O: 0.73 (0.49, 1.07)<br>F: 0.65 (0.37, 1.15)<br>M: 0.78 (0.47, 1.30) | -0.18 (-0.93, 0.58) |
| Feel at peace§ | O: 0.70 (0.48, 1.02)*<br>F: 0.68 (0.38, 1.21)<br>M: 0.71 (0.43, 1.16) | -0.04 (-0.78, 0.70) |
| Government approval§ | O: 0.61 (0.43, 0.87)**<br>F: 0.62 (0.36, 1.08)*<br>M: 0.60 (0.37, 0.98)** | 0.03 (-0.73, 0.78) |
| Say in government# | O: 0.58 (0.35, 0.96)**<br>F: 0.54 (0.28, 1.04)*<br>M: 0.61 (0.31, 1.22) | -0.13 (-1.04, 0.78) |

*Not feeling loved by father – Large city or suburb of a large city*

**Adjusted associations between not feeling loved by father when growing up and adult outcomes in India**
**Residence: Large city or suburb**

Wave 1, survey-weighted linear model. One model was fitted for each continuous outcome (24 models total, for 22 outcomes and 2 flourishing composite indices).
Estimates are the adjusted model coefficients (beta) of the predictor.

Group: Overall (O), Female (F), Male (M)

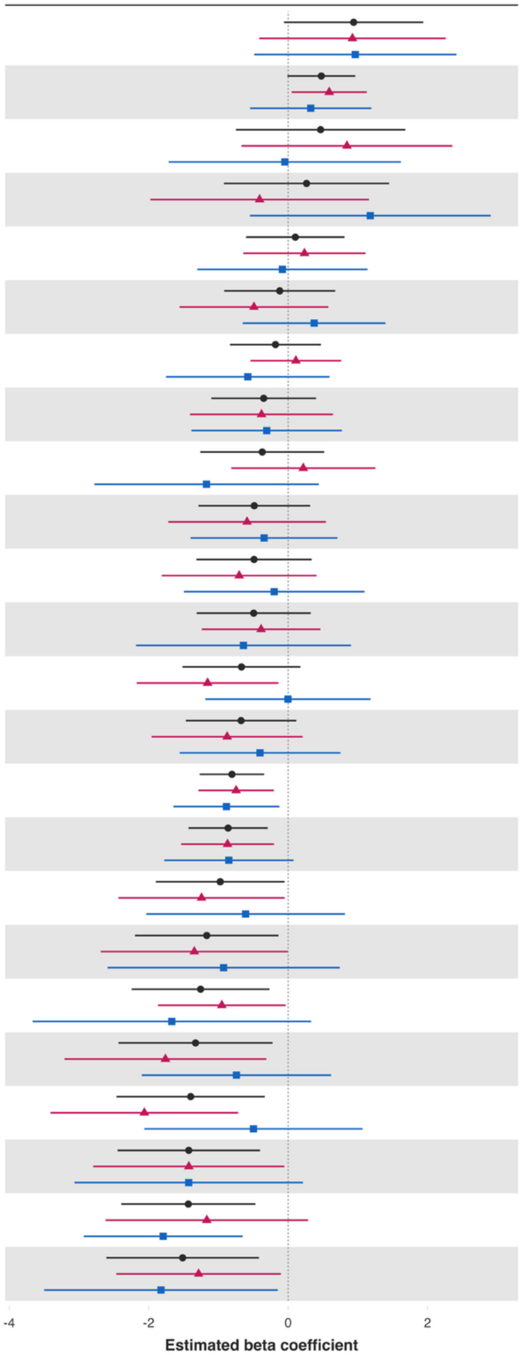


| Variable | Overall and interaction-derived gender estimates (95% CI) | Interaction (F–M, 95% CI) |
| --- | --- | --- |
| Worry about meeting normal monthly living expenses§§‡‡ | O: 0.94 (-0.06, 1.94)*<br>F: 0.92 (-0.41, 2.26)<br>M: 0.96 (-0.49, 2.41) | -0.04 (-1.99, 1.91) |
| Remain hopeful for the future | O: 0.48 (-0.01, 0.96)*<br>F: 0.59 (0.05, 1.13)**<br>M: 0.32 (-0.55, 1.19) | 0.26 (-0.75, 1.28) |
| Loneliness§ | O: 0.47 (-0.75, 1.68)<br>F: 0.84 (-0.67, 2.36)<br>M: -0.05 (-1.71, 1.62) | 0.89 (-1.21, 2.99) |
| Worry about safety, food, or housing§§‡‡ | O: 0.26 (-0.92, 1.45)<br>F: -0.41 (-1.98, 1.16)<br>M: 1.18 (-0.55, 2.90) | -1.59 (-3.93, 0.76) |
| Have a long list of gratitude | O: 0.10 (-0.60, 0.81)<br>F: 0.23 (-0.64, 1.11)<br>M: -0.08 (-1.30, 1.14) | 0.32 (-1.20, 1.84) |
| Understand purpose in life‡ | O: -0.12 (-0.91, 0.68)<br>F: -0.49 (-1.56, 0.58)<br>M: 0.37 (-0.65, 1.40) | -0.86 (-2.30, 0.57) |
| Act to promote good‡ | O: -0.18 (-0.83, 0.47)<br>F: 0.11 (-0.54, 0.76)<br>M: -0.58 (-1.75, 0.59) | 0.69 (-0.61, 1.99) |
| Belonging | O: -0.35 (-1.10, 0.40)<br>F: -0.38 (-1.41, 0.64)<br>M: -0.31 (-1.39, 0.77) | -0.07 (-1.56, 1.41) |
| Give up some happiness now for greater happiness later‡ | O: -0.37 (-1.26, 0.52)<br>F: 0.22 (-0.82, 1.25)<br>M: -1.17 (-2.78, 0.44) | 1.39 (-0.58, 3.35) |
| Freedom | O: -0.49 (-1.29, 0.32)<br>F: -0.59 (-1.72, 0.54)<br>M: -0.34 (-1.40, 0.71) | -0.24 (-1.78, 1.29) |
| Positive expectations | O: -0.49 (-1.31, 0.34)<br>F: -0.70 (-1.81, 0.41)<br>M: -0.20 (-1.49, 1.10) | -0.50 (-2.25, 1.25) |
| Life evaluation five years from now | O: -0.49 (-1.31, 0.32)<br>F: -0.39 (-1.24, 0.46)<br>M: -0.64 (-2.18, 0.90) | 0.25 (-1.50, 2.01) |
| Show love often | O: -0.67 (-1.51, 0.18)<br>F: -1.15 (-2.17, -0.14)**<br>M: -0.00 (-1.19, 1.18) | -1.15 (-2.64, 0.33) |
| Physical health‡ | O: -0.67 (-1.47, 0.12)*<br>F: -0.87 (-1.96, 0.21)<br>M: -0.40 (-1.56, 0.75) | -0.47 (-2.05, 1.11) |
| Flourishing (with financial indicators) | O: -0.81 (-1.27, -0.34)***<br>F: -0.75 (-1.29, -0.21)**<br>M: -0.88 (-1.64, -0.13)** | 0.14 (-0.77, 1.05) |
| Flourishing (without financial indicators) | O: -0.86 (-1.43, -0.29)**<br>F: -0.87 (-1.53, -0.20)**<br>M: -0.85 (-1.78, 0.08)* | -0.02 (-1.13, 1.09) |
| Mental health‡ | O: -0.97 (-1.90, -0.05)**<br>F: -1.24 (-2.43, -0.05)**<br>M: -0.61 (-2.03, 0.82) | -0.63 (-2.49, 1.23) |
| Life worthwhile‡ | O: -1.17 (-2.20, -0.14)**<br>F: -1.35 (-2.69, -0.00)*<br>M: -0.92 (-2.59, 0.74) | -0.42 (-2.59, 1.75) |
| Relationship contentment‡ | O: -1.25 (-2.24, -0.27)**<br>F: -0.95 (-1.87, -0.04)**<br>M: -1.67 (-3.67, 0.33) | 0.72 (-1.51, 2.95) |
| Life satisfaction‡ | O: -1.33 (-2.43, -0.22)**<br>F: -1.76 (-3.21, -0.31)**<br>M: -0.74 (-2.10, 0.62) | -1.02 (-2.84, 0.80) |
| Perceived support from others | O: -1.40 (-2.46, -0.33)**<br>F: -2.06 (-3.41, -0.72)**<br>M: -0.50 (-2.06, 1.07) | -1.57 (-3.58, 0.45) |
| Relationship satisfaction‡ | O: -1.42 (-2.45, -0.40)**<br>F: -1.42 (-2.79, -0.05)**<br>M: -1.43 (-3.06, 0.21)* | 0.00 (-2.19, 2.20) |
| Life evaluation today | O: -1.43 (-2.39, -0.47)**<br>F: -1.17 (-2.62, 0.29)<br>M: -1.79 (-2.93, -0.65)** | 0.63 (-1.23, 2.48) |
| Happiness‡ | O: -1.51 (-2.61, -0.42)**<br>F: -1.28 (-2.46, -0.10)**<br>M: -1.82 (-3.50, -0.15)** | 0.54 (-1.27, 2.35) |

**Adjusted associations between not feeling loved by father when growing up and adult outcomes in India**
**Residence: Large city or suburb**

Wave 1, survey-weighted logistic model. One model was fitted for each binary outcome (7 models total). Estimates are adjusted odds ratios (OR) of the predictor.
The interaction column reports the difference in log-odds ratios (interaction beta) with 95% confidence intervals (95% CIs).

Group: Overall (O), Female (F), Male (M)

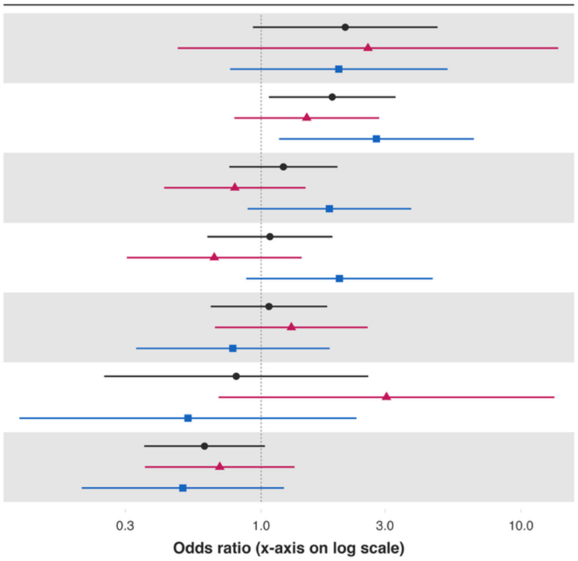


| Variable | Overall and interaction-derived gender estimates (95% CI) | Interaction (F–M, 95% CI) |
|---|---|---|
| Alcohol use | O: 2.11 (0.93, 4.78)*<br>F: 2.58 (0.48, 13.92)<br>M: 1.99 (0.76, 5.22) | 0.26 (-1.77, 2.28) |
| Helped a stranger | O: 1.88 (1.07, 3.29)**<br>F: 1.50 (0.79, 2.85)<br>M: 2.78 (1.17, 6.59)** | -0.62 (-1.59, 0.36) |
| Volunteered at an organization | O: 1.22 (0.75, 1.97)<br>F: 0.79 (0.42, 1.48)<br>M: 1.83 (0.89, 3.78) | -0.84 (-1.79, 0.12)* |
| Donated to charity | O: 1.08 (0.62, 1.88)<br>F: 0.66 (0.30, 1.43)<br>M: 2.00 (0.88, 4.57) | -1.11 (-2.27, 0.06)* |
| Have any health problems | O: 1.07 (0.64, 1.80)<br>F: 1.31 (0.66, 2.57)<br>M: 0.78 (0.33, 1.84) | 0.52 (-0.58, 1.62) |
| Cigarette smoking | O: 0.80 (0.25, 2.59)<br>F: 3.04 (0.69, 13.45)<br>M: 0.52 (0.12, 2.33) | 1.76 (-0.57, 4.09) |
| Feel close to someone | O: 0.61 (0.35, 1.03)*<br>F: 0.69 (0.36, 1.35)<br>M: 0.50 (0.20, 1.23) | 0.33 (-0.79, 1.45) |

**Adjusted associations between not feeling loved by father when growing up and adult outcomes in India**
**Residence: Large city or suburb**

Wave 1, survey-weighted ordinal regression using the proportional odds model (`svyolr`). One model was fitted for each ordinal outcome (16 models total).
Estimates are adjusted cumulative odds ratios (cOR) of the predictor.
The interaction column reports the difference in log-cumulative odds ratios (interaction beta) with 95% confidence intervals (95% CIs).

**Group** ● Overall (O) ▲ Female (F) ■ Male (M)

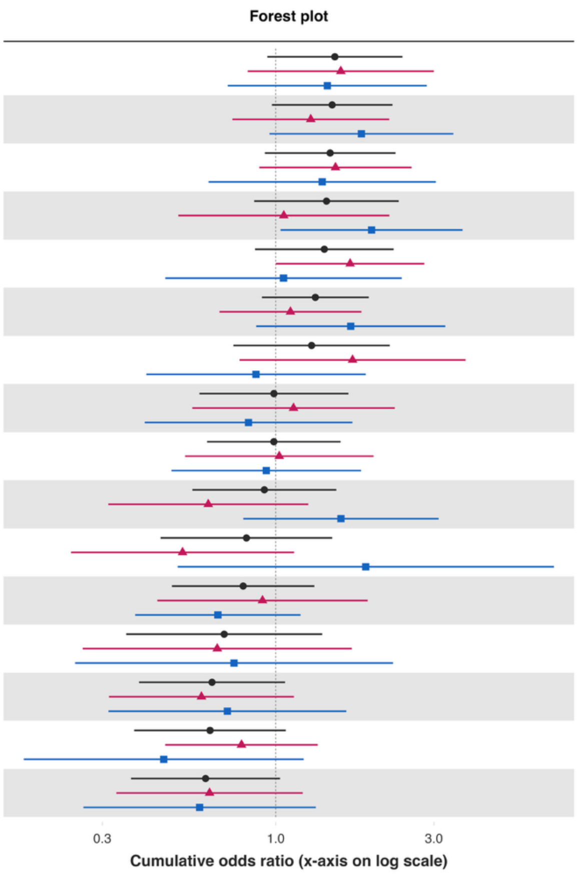


| Variable | Overall and interaction-derived gender estimates (95% CI) | Interaction (F–M, 95% CI) |
|---|---|---|
| Depressive symptoms§ | O: 1.51 (0.94, 2.41)*<br>F: 1.57 (0.82, 3.00)<br>M: 1.43 (0.72, 2.86) | 0.09 (-0.86, 1.05) |
| Feeling nervous or anxious§ | O: 1.48 (0.97, 2.25)*<br>F: 1.28 (0.74, 2.20)<br>M: 1.81 (0.96, 3.43)* | -0.35 (-1.20, 0.50) |
| Distress from e-threatening experiences§ | O: 1.46 (0.93, 2.30)<br>F: 1.51 (0.89, 2.57)<br>M: 1.38 (0.63, 3.04) | 0.09 (-0.85, 1.03) |
| Perceived discrimination§ | O: 1.42 (0.86, 2.35)<br>F: 1.06 (0.51, 2.20)<br>M: 1.95 (1.03, 3.66)** | -0.61 (-1.56, 0.34) |
| Have little interest in doing things§ | O: 1.40 (0.87, 2.27)<br>F: 1.68 (1.00, 2.81)**<br>M: 1.06 (0.46, 2.40) | 0.46 (-0.44, 1.36) |
| Unable to control worry§ | O: 1.32 (0.91, 1.91)<br>F: 1.11 (0.68, 1.81)<br>M: 1.68 (0.87, 3.25) | -0.42 (-1.30, 0.46) |
| Extent of suffering§ | O: 1.28 (0.75, 2.21)<br>F: 1.70 (0.78, 3.74)<br>M: 0.87 (0.41, 1.87) | 0.67 (-0.44, 1.78) |
| Trust in others§ | O: 0.99 (0.59, 1.66)<br>F: 1.13 (0.56, 2.29)<br>M: 0.83 (0.40, 1.70) | 0.31 (-0.67, 1.29) |
| Various aspects of life are balanced§ | O: 0.99 (0.62, 1.57)<br>F: 1.02 (0.53, 1.97)<br>M: 0.94 (0.48, 1.81) | 0.09 (-0.85, 1.03) |
| Feel at peace§ | O: 0.92 (0.56, 1.52)<br>F: 0.63 (0.31, 1.25)<br>M: 1.57 (0.80, 3.10) | -0.92 (-1.85, 0.01)* |
| Obeying the law§ | O: 0.82 (0.45, 1.48)<br>F: 0.52 (0.24, 1.14)<br>M: 1.87 (0.51, 6.90) | -1.27 (-2.93, 0.39) |
| Feeling capable§ | O: 0.80 (0.49, 1.31)<br>F: 0.91 (0.44, 1.89)<br>M: 0.67 (0.38, 1.19) | 0.31 (-0.59, 1.21) |
| Support reducing fferences in income levels§ | O: 0.70 (0.35, 1.38)<br>F: 0.67 (0.26, 1.70)<br>M: 0.75 (0.25, 2.26) | -0.12 (-1.63, 1.40) |
| Say in government# | O: 0.64 (0.39, 1.07)*<br>F: 0.60 (0.31, 1.13)<br>M: 0.71 (0.31, 1.63) | -0.18 (-1.22, 0.86) |
| Government approval§ | O: 0.63 (0.37, 1.07)*<br>F: 0.79 (0.46, 1.34)<br>M: 0.46 (0.17, 1.21) | 0.54 (-0.51, 1.59) |
| Forgiveness§ | O: 0.61 (0.37, 1.03)*<br>F: 0.63 (0.33, 1.21)<br>M: 0.59 (0.26, 1.32) | 0.07 (-0.94, 1.08) |

**Predictor:** ***Poorer relationship quality with mother (bad or very bad relationship)***
***Poorer relationship quality with mother (bad or very bad relationship) – Rural area or on a farm***

**Adjusted associations between somewhat bad or very bad relationship with mother (vs very good) and adult outcomes in India**
**Residence: Rural**

Wave 1, survey-weighted linear model. One model was fitted for each continuous outcome (24 models total, for 22 outcomes and 2 flourishing composite indices).
Estimates are the adjusted model coefficients (beta) of the predictor.

Group: Overall (O), Female (F), Male (M)

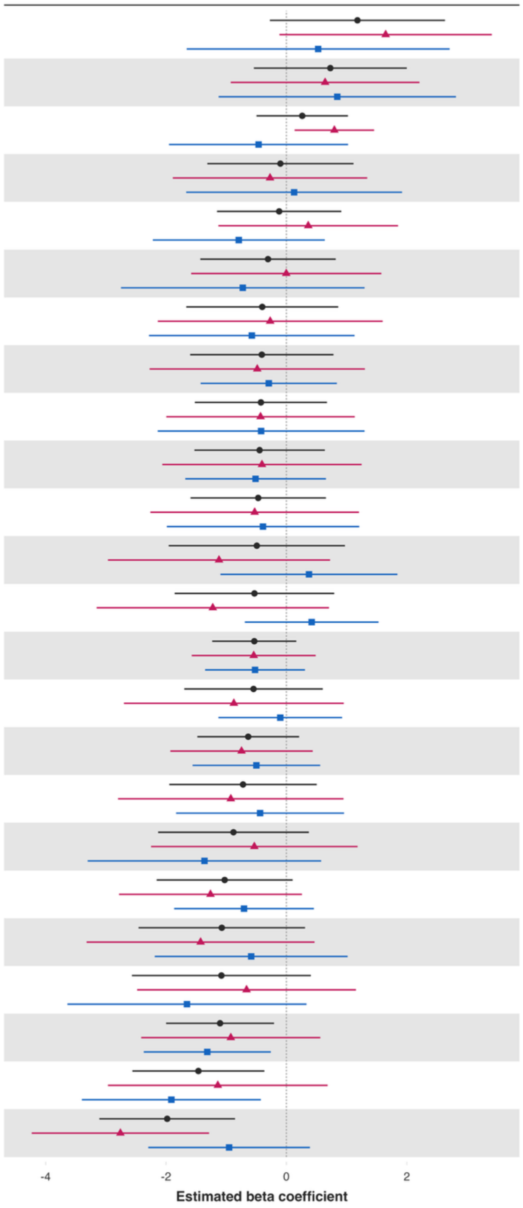


| Variable | Overall and interaction-derived gender estimates (95% CI) | Interaction (F–M, 95% CI) |
|---|---|---|
| Loneliness§ | O: 1.18 (-0.27, 2.63)<br>F: 1.65 (-0.12, 3.41)*<br>M: 0.53 (-1.66, 2.71) | 1.12 (-1.68, 3.92) |
| Worry about safety, food, or housing§§‡‡ | O: 0.73 (-0.54, 2.00)<br>F: 0.64 (-0.93, 2.21)<br>M: 0.84 (-1.13, 2.82) | -0.20 (-2.64, 2.23) |
| Show love often | O: 0.26 (-0.50, 1.02)<br>F: 0.80 (0.14, 1.46)**<br>M: -0.46 (-1.95, 1.02) | 1.26 (-0.27, 2.79) |
| Give up some happiness now for greater happiness later‡ | O: -0.10 (-1.31, 1.11)<br>F: -0.27 (-1.89, 1.34)<br>M: 0.13 (-1.67, 1.92) | -0.40 (-2.81, 2.01) |
| Mental health‡ | O: -0.12 (-1.15, 0.91)<br>F: 0.36 (-1.13, 1.85)<br>M: -0.79 (-2.22, 0.64) | 1.15 (-0.94, 3.24) |
| Life satisfaction‡ | O: -0.31 (-1.43, 0.82)<br>F: -0.00 (-1.58, 1.58)<br>M: -0.72 (-2.75, 1.30) | 0.72 (-2.02, 3.46) |
| Understand purpose in life‡ | O: -0.40 (-1.66, 0.86)<br>F: -0.27 (-2.14, 1.60)<br>M: -0.58 (-2.28, 1.13) | 0.31 (-2.27, 2.88) |
| Have a long list of gratitude | O: -0.41 (-1.60, 0.78)<br>F: -0.48 (-2.27, 1.30)<br>M: -0.29 (-1.43, 0.84) | -0.19 (-2.19, 1.81) |
| Act to promote good‡ | O: -0.42 (-1.52, 0.67)<br>F: -0.43 (-2.00, 1.14)<br>M: -0.42 (-2.14, 1.30) | -0.01 (-2.45, 2.44) |
| Positive expectations | O: -0.45 (-1.53, 0.64)<br>F: -0.41 (-2.06, 1.25)<br>M: -0.51 (-1.68, 0.65) | 0.11 (-1.91, 2.12) |
| Belonging | O: -0.47 (-1.59, 0.65)<br>F: -0.53 (-2.26, 1.20)<br>M: -0.39 (-1.99, 1.21) | -0.14 (-2.69, 2.41) |
| Life worthwhile‡ | O: -0.49 (-1.96, 0.97)<br>F: -1.12 (-2.96, 0.72)<br>M: 0.37 (-1.10, 1.84) | -1.49 (-3.42, 0.44) |
| Worry about meeting normal monthly living expenses§§‡‡ | O: -0.53 (-1.86, 0.79)<br>F: -1.22 (-3.15, 0.71)<br>M: 0.42 (-0.69, 1.53) | -1.64 (-3.71, 0.42) |
| Flourishing (with financial indicators) | O: -0.53 (-1.23, 0.16)<br>F: -0.54 (-1.57, 0.48)<br>M: -0.52 (-1.35, 0.31) | -0.02 (-1.30, 1.26) |
| Happiness‡ | O: -0.55 (-1.70, 0.60)<br>F: -0.87 (-2.70, 0.95)<br>M: -0.10 (-1.13, 0.92) | -0.77 (-2.89, 1.34) |
| Flourishing (without financial indicators) | O: -0.64 (-1.48, 0.21)<br>F: -0.75 (-1.93, 0.44)<br>M: -0.50 (-1.56, 0.56) | -0.25 (-1.78, 1.28) |
| Relationship satisfaction‡ | O: -0.72 (-1.95, 0.50)<br>F: -0.92 (-2.80, 0.95)<br>M: -0.44 (-1.83, 0.96) | -0.49 (-2.85, 1.87) |
| Relationship contentment‡ | O: -0.88 (-2.13, 0.37)<br>F: -0.53 (-2.25, 1.18)<br>M: -1.36 (-3.30, 0.58) | 0.83 (-1.79, 3.44) |
| Life evaluation today | O: -1.03 (-2.16, 0.10)*<br>F: -1.26 (-2.78, 0.26)<br>M: -0.70 (-1.87, 0.46) | -0.56 (-2.18, 1.07) |
| Remain hopeful for the future | O: -1.07 (-2.46, 0.31)<br>F: -1.43 (-3.32, 0.47)<br>M: -0.59 (-2.19, 1.02) | -0.84 (-3.14, 1.46) |
| Freedom | O: -1.08 (-2.57, 0.41)<br>F: -0.66 (-2.48, 1.15)<br>M: -1.65 (-3.64, 0.33) | 0.99 (-1.37, 3.34) |
| Life evaluation five years from now | O: -1.10 (-2.00, -0.21)**<br>F: -0.92 (-2.41, 0.56)<br>M: -1.32 (-2.37, -0.26)** | 0.39 (-1.48, 2.26) |
| Perceived support from others | O: -1.46 (-2.56, -0.37)**<br>F: -1.14 (-2.96, 0.68)<br>M: -1.91 (-3.40, -0.43)** | 0.77 (-1.86, 3.41) |
| Physical health‡ | O: -1.98 (-3.11, -0.85)***<br>F: -2.76 (-4.23, -1.28)***<br>M: -0.95 (-2.29, 0.39) | -1.81 (-3.70, 0.09)* |

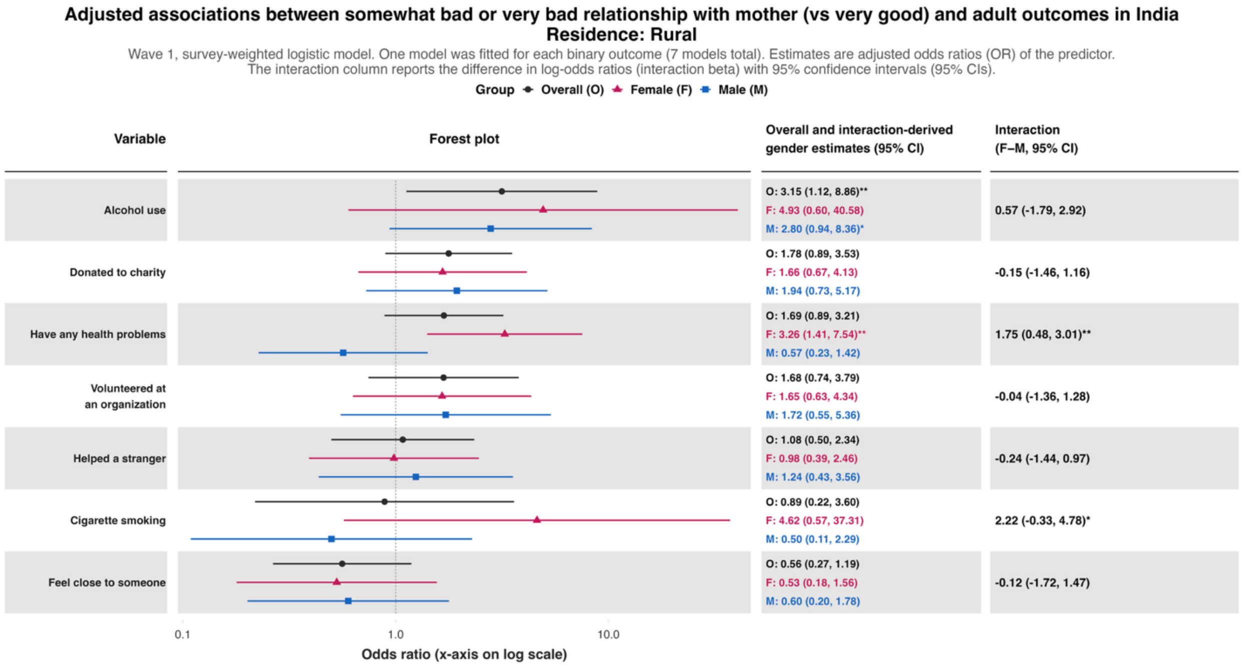
Adjusted associations between somewhat bad or very bad relationship with mother (vs very good) and adult outcomes in India
Residence: Rural
Wave 1, survey-weighted logistic model. One model was fitted for each binary outcome (7 models total). Estimates are adjusted odds ratios (OR) of the predictor.
The interaction column reports the difference in log-odds ratios (interaction beta) with 95% confidence intervals (95% CIs).
Group Overall (O) Female (F) Male (M)
Variable
Forest plot
Overall and interaction-derived gender estimates (95% CI)
Interaction (F–M, 95% CI)
Alcohol use
O: 3.15 (1.12, 8.86)**
F: 4.93 (0.60, 40.58)
M: 2.80 (0.94, 8.36)*
0.57 (-1.79, 2.92)
Donated to charity
O: 1.78 (0.89, 3.53)
F: 1.66 (0.67, 4.13)
M: 1.94 (0.73, 5.17)
-0.15 (-1.46, 1.16)
Have any health problems
O: 1.69 (0.89, 3.21)
F: 3.26 (1.41, 7.54)**
M: 0.57 (0.23, 1.42)
1.75 (0.48, 3.01)**
Volunteered at an organization
O: 1.68 (0.74, 3.79)
F: 1.65 (0.63, 4.34)
M: 1.72 (0.55, 5.36)
-0.04 (-1.36, 1.28)
Helped a stranger
O: 1.08 (0.50, 2.34)
F: 0.98 (0.39, 2.46)
M: 1.24 (0.43, 3.56)
-0.24 (-1.44, 0.97)
Cigarette smoking
O: 0.89 (0.22, 3.60)
F: 4.62 (0.57, 37.31)
M: 0.50 (0.11, 2.29)
2.22 (-0.33, 4.78)*
Feel close to someone
O: 0.56 (0.27, 1.19)
F: 0.53 (0.18, 1.56)
M: 0.60 (0.20, 1.78)
-0.12 (-1.72, 1.47)
0.1
1.0
10.0
Odds ratio (x-axis on log scale)

**Adjusted associations between somewhat bad or very bad relationship with mother (vs very good) and adult outcomes in India**
**Residence: Rural**

Wave 1, survey-weighted ordinal regression using the proportional odds model (`svyolr`). One model was fitted for each ordinal outcome (16 models total).
Estimates are adjusted cumulative odds ratios (cOR) of the predictor.
The interaction column reports the difference in log-cumulative odds ratios (interaction beta) with 95% confidence intervals (95% CIs).

**Group** ● Overall (O) ▲ Female (F) ■ Male (M)

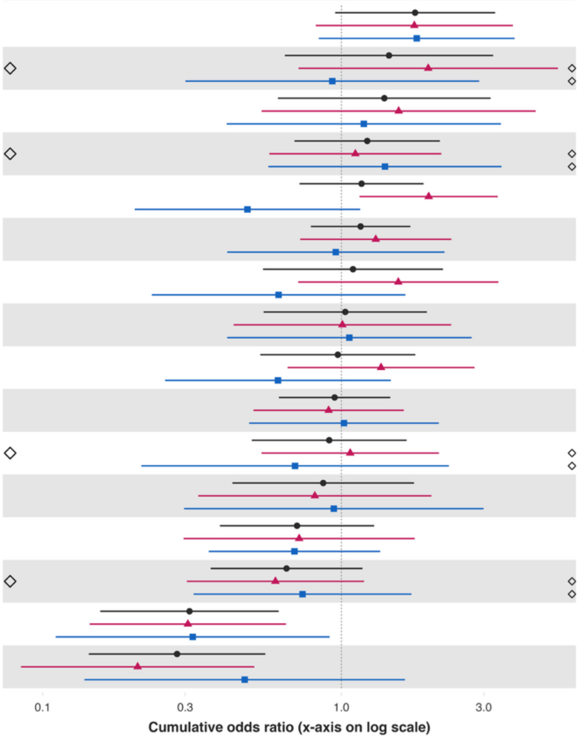


| Variable | Overall and interaction-derived gender estimates (95% CI) | Interaction (F–M, 95% CI) |
|---|---|---|
| Feel at peace§ | O: 1.77 (0.95, 3.27)*<br>F: 1.76 (0.82, 3.75)<br>M: 1.79 (0.84, 3.80) | -0.02 (-0.91, 0.87) |
| Forgiveness§ | O: 1.44 (0.65, 3.22)<br>F: 1.95 (0.72, 5.31)<br>M: 0.93 (0.30, 2.89) | 0.74 (-0.57, 2.05) |
| Feeling nervous or anxious§ | O: 1.39 (0.61, 3.16)<br>F: 1.56 (0.54, 4.47)<br>M: 1.19 (0.41, 3.42) | 0.27 (-1.11, 1.65) |
| Trust in others§ | O: 1.22 (0.70, 2.14)<br>F: 1.11 (0.57, 2.16)<br>M: 1.40 (0.57, 3.44) | -0.23 (-1.31, 0.85) |
| Distress from life-threatening experiences§ | O: 1.17 (0.72, 1.88)<br>F: 1.96 (1.15, 3.34)**<br>M: 0.48 (0.20, 1.16) | 1.40 (0.31, 2.49)** |
| Depressive symptoms§ | O: 1.16 (0.79, 1.70)<br>F: 1.30 (0.73, 2.33)<br>M: 0.96 (0.41, 2.21) | 0.31 (-0.86, 1.48) |
| Extent of suffering§ | O: 1.09 (0.55, 2.19)<br>F: 1.55 (0.72, 3.36)<br>M: 0.62 (0.23, 1.64) | 0.92 (-0.14, 1.99)* |
| Unable to control worry§ | O: 1.03 (0.55, 1.93)<br>F: 1.01 (0.44, 2.33)<br>M: 1.06 (0.41, 2.73) | -0.05 (-1.31, 1.20) |
| Perceived discrimination§ | O: 0.97 (0.54, 1.77)<br>F: 1.36 (0.66, 2.79)<br>M: 0.61 (0.26, 1.46) | 0.79 (-0.20, 1.79) |
| Have little interest in doing things§ | O: 0.95 (0.62, 1.46)<br>F: 0.91 (0.51, 1.62)<br>M: 1.02 (0.49, 2.12) | -0.12 (-1.10, 0.87) |
| Various aspects of life are balanced§ | O: 0.91 (0.50, 1.65)<br>F: 1.07 (0.54, 2.12)<br>M: 0.70 (0.21, 2.29) | 0.42 (-0.96, 1.81) |
| Say in government# | O: 0.87 (0.43, 1.75)<br>F: 0.81 (0.33, 2.00)<br>M: 0.94 (0.30, 2.99) | -0.15 (-1.64, 1.35) |
| Government approval§ | O: 0.71 (0.39, 1.29)<br>F: 0.72 (0.30, 1.76)<br>M: 0.70 (0.36, 1.35) | 0.04 (-1.04, 1.11) |
| Feeling capable§ | O: 0.65 (0.36, 1.18)<br>F: 0.60 (0.30, 1.19)<br>M: 0.74 (0.32, 1.72) | -0.21 (-1.19, 0.77) |
| Support reducing differences in income levels§ | O: 0.31 (0.16, 0.62)***<br>F: 0.31 (0.14, 0.65)**<br>M: 0.32 (0.11, 0.91)** | -0.04 (-1.20, 1.12) |
| Obeying the law§ | O: 0.28 (0.14, 0.56)***<br>F: 0.21 (0.08, 0.51)***<br>M: 0.47 (0.14, 1.63) | -0.83 (-2.36, 0.71) |

*Poorer relationship quality with mother (bad or very bad relationship) – Small town or village*

**Adjusted associations between somewhat bad or very bad relationship with mother (vs very good) and adult outcomes in India**
**Residence: Small town**

Wave 1, survey-weighted linear model. One model was fitted for each continuous outcome (24 models total, for 22 outcomes and 2 flourishing composite indices).
Estimates are the adjusted model coefficients (beta) of the predictor.

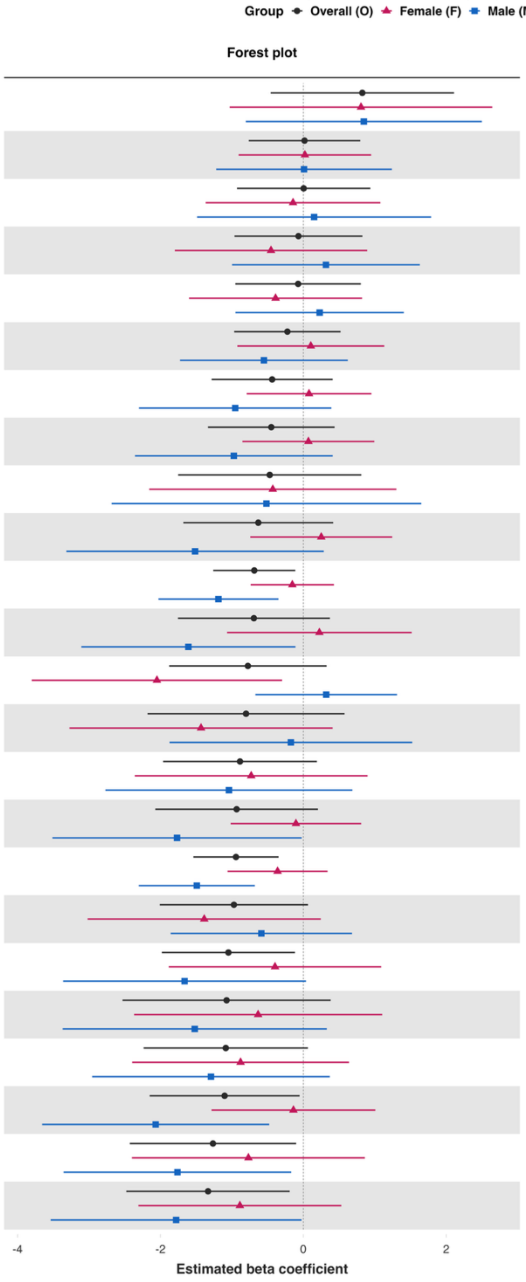


| Variable | Overall and interaction-derived gender estimates (95% CI) | Interaction (F–M, 95% CI) |
|---|---|---|
| Loneliness§ | O: 0.83 (-0.46, 2.11)<br>F: 0.81 (-1.03, 2.65)<br>M: 0.85 (-0.81, 2.50) | -0.04 (-2.42, 2.34) |
| Remain hopeful for the future | O: 0.02 (-0.76, 0.80)<br>F: 0.02 (-0.91, 0.95)<br>M: 0.01 (-1.22, 1.24) | 0.01 (-1.50, 1.53) |
| Have a long list of gratitude | O: 0.00 (-0.93, 0.94)<br>F: -0.14 (-1.37, 1.08)<br>M: 0.15 (-1.49, 1.79) | -0.30 (-2.51, 1.92) |
| Show love often | O: -0.07 (-0.96, 0.83)<br>F: -0.45 (-1.80, 0.89)<br>M: 0.32 (-1.00, 1.63) | -0.77 (-2.77, 1.24) |
| Positive expectations | O: -0.07 (-0.95, 0.81)<br>F: -0.39 (-1.60, 0.82)<br>M: 0.23 (-0.95, 1.41) | -0.62 (-2.27, 1.04) |
| Understand purpose in life‡ | O: -0.22 (-0.97, 0.52)<br>F: 0.10 (-0.92, 1.13)<br>M: -0.55 (-1.73, 0.62) | 0.66 (-0.98, 2.29) |
| Act to promote good‡ | O: -0.44 (-1.28, 0.41)<br>F: 0.08 (-0.79, 0.95)<br>M: -0.95 (-2.30, 0.39) | 1.03 (-0.52, 2.59) |
| Belonging | O: -0.45 (-1.34, 0.44)<br>F: 0.07 (-0.85, 0.99)<br>M: -0.97 (-2.36, 0.41) | 1.04 (-0.58, 2.67) |
| Worry about meeting normal monthly living expenses§§‡‡ | O: -0.47 (-1.75, 0.81)<br>F: -0.43 (-2.16, 1.30)<br>M: -0.52 (-2.68, 1.65) | 0.09 (-2.87, 3.04) |
| Relationship contentment‡ | O: -0.63 (-1.68, 0.42)<br>F: 0.25 (-0.74, 1.24)<br>M: -1.52 (-3.32, 0.29) | 1.77 (-0.28, 3.81)* |
| Flourishing (with financial indicators) | O: -0.69 (-1.26, -0.11)**<br>F: -0.15 (-0.74, 0.43)<br>M: -1.19 (-2.03, -0.35)** | 1.03 (0.03, 2.03)** |
| Happiness‡ | O: -0.69 (-1.76, 0.37)<br>F: 0.22 (-1.07, 1.52)<br>M: -1.61 (-3.11, -0.11)** | 1.83 (-0.13, 3.80)* |
| Life evaluation five years from now | O: -0.78 (-1.88, 0.33)<br>F: -2.05 (-3.80, -0.30)**<br>M: 0.32 (-0.67, 1.31) | -2.37 (-4.41, -0.33)** |
| Worry about safety, food, or housing§§‡‡ | O: -0.80 (-2.18, 0.58)<br>F: -1.43 (-3.27, 0.41)<br>M: -0.17 (-1.87, 1.52) | -1.26 (-3.53, 1.01) |
| Give up some happiness now for greater happiness later‡ | O: -0.89 (-1.96, 0.19)<br>F: -0.73 (-2.36, 0.90)<br>M: -1.04 (-2.77, 0.69) | 0.31 (-2.26, 2.88) |
| Relationship satisfaction‡ | O: -0.93 (-2.07, 0.20)<br>F: -0.10 (-1.02, 0.81)<br>M: -1.77 (-3.51, -0.02)** | 1.66 (-0.06, 3.39)* |
| Flourishing (without financial indicators) | O: -0.94 (-1.54, -0.35)**<br>F: -0.36 (-1.06, 0.34)<br>M: -1.49 (-2.30, -0.68)*** | 1.13 (0.09, 2.17)** |
| Life evaluation today | O: -0.97 (-2.01, 0.07)*<br>F: -1.39 (-3.02, 0.24)*<br>M: -0.59 (-1.86, 0.68) | -0.80 (-2.86, 1.26) |
| Life worthwhile‡ | O: -1.05 (-1.98, -0.12)**<br>F: -0.40 (-1.88, 1.09)<br>M: -1.66 (-3.36, 0.04)* | 1.27 (-1.40, 3.93) |
| Perceived support from others | O: -1.07 (-2.53, 0.38)<br>F: -0.63 (-2.37, 1.10)<br>M: -1.52 (-3.37, 0.33) | 0.89 (-1.26, 3.03) |
| Freedom | O: -1.09 (-2.24, 0.06)*<br>F: -0.88 (-2.40, 0.64)<br>M: -1.29 (-2.96, 0.37) | 0.41 (-1.80, 2.63) |
| Life satisfaction‡ | O: -1.10 (-2.15, -0.05)**<br>F: -0.14 (-1.29, 1.01)<br>M: -2.07 (-3.66, -0.48)** | 1.93 (-0.03, 3.88)* |
| Physical health‡ | O: -1.27 (-2.43, -0.10)**<br>F: -0.77 (-2.40, 0.86)<br>M: -1.76 (-3.35, -0.17)** | 0.99 (-1.30, 3.29) |
| Mental health‡ | O: -1.33 (-2.48, -0.19)**<br>F: -0.89 (-2.31, 0.53)<br>M: -1.78 (-3.54, -0.03)** | 0.89 (-1.41, 3.20) |

**Adjusted associations between somewhat bad or very bad relationship with mother (vs very good) and adult outcomes in India**
**Residence: Small town**

Wave 1, survey-weighted logistic model. One model was fitted for each binary outcome (7 models total). Estimates are adjusted odds ratios (OR) of the predictor.
The interaction column reports the difference in log-odds ratios (interaction beta) with 95% confidence intervals (95% CIs).

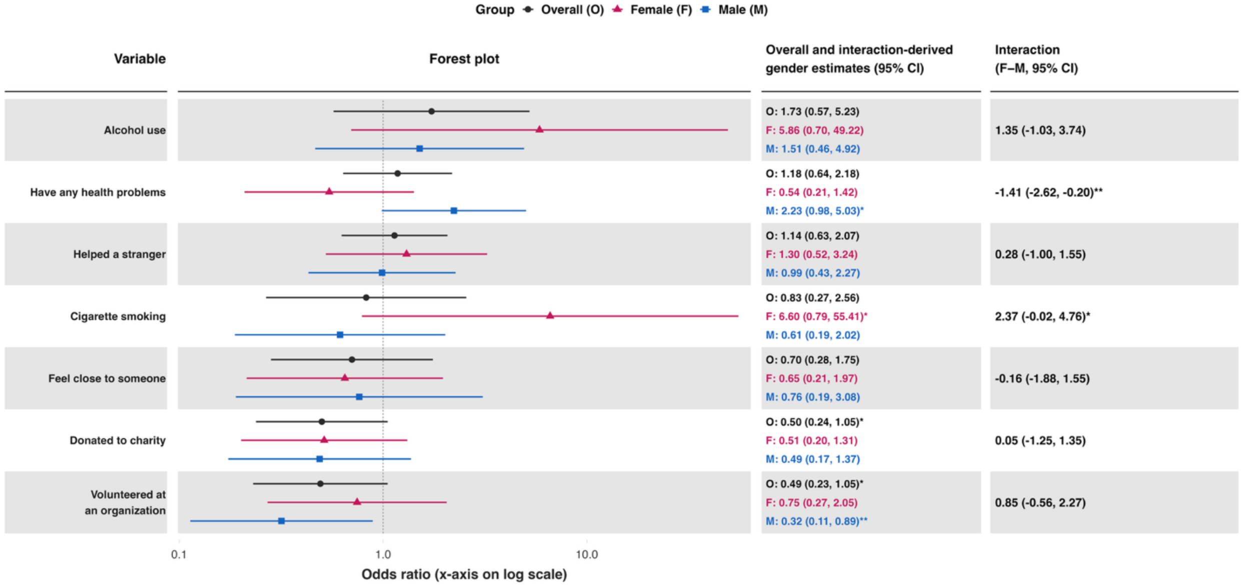

### Adjusted associations between somewhat bad or very bad relationship with mother (vs very good) and adult outcomes in India
#### Residence: Small town

Wave 1, survey-weighted ordinal regression using the proportional odds model (`svyolr`). One model was fitted for each ordinal outcome (16 models total).
Estimates are adjusted cumulative odds ratios (cOR) of the predictor.
The interaction column reports the difference in log-cumulative odds ratios (interaction beta) with 95% confidence intervals (95% CIs).

**Group** Overall (O) Female (F) Male (M)

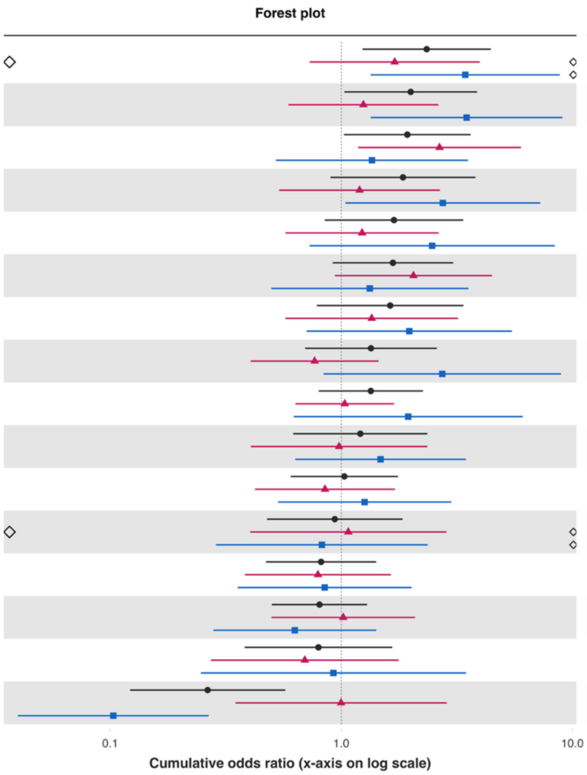


| Variable | Overall and interaction-derived gender estimates (95% CI) | Interaction (F–M, 95% CI) |
|---|---|---|
| Feeling capable§ | O: 2.34 (1.23, 4.44)**<br>F: 1.70 (0.73, 3.97)<br>M: 3.44 (1.34, 8.82)** | -0.70 (-1.96, 0.55) |
| Extent of suffering§ | O: 2.00 (1.03, 3.87)**<br>F: 1.25 (0.59, 2.63)<br>M: 3.48 (1.34, 9.06)** | -1.03 (-2.17, 0.12)* |
| Various aspects of life are balanced§ | O: 1.93 (1.03, 3.63)**<br>F: 2.66 (1.18, 5.99)**<br>M: 1.36 (0.52, 3.53) | 0.67 (-0.55, 1.90) |
| Trust in others§ | O: 1.85 (0.90, 3.81)*<br>F: 1.20 (0.54, 2.67)<br>M: 2.75 (1.04, 7.26)** | -0.83 (-1.95, 0.29) |
| Unable to control worry§ | O: 1.69 (0.85, 3.37)<br>F: 1.23 (0.57, 2.64)<br>M: 2.47 (0.73, 8.38) | -0.70 (-2.18, 0.79) |
| Perceived discrimination§ | O: 1.67 (0.92, 3.05)*<br>F: 2.05 (0.94, 4.49)*<br>M: 1.33 (0.50, 3.55) | 0.43 (-0.86, 1.73) |
| Forgiveness§ | O: 1.63 (0.78, 3.37)<br>F: 1.35 (0.57, 3.20)<br>M: 1.97 (0.71, 5.48) | -0.37 (-1.58, 0.83) |
| Feeling nervous or anxious§ | O: 1.34 (0.70, 2.59)<br>F: 0.77 (0.41, 1.45)<br>M: 2.73 (0.84, 8.92)* | -1.27 (-2.64, 0.10)* |
| Depressive symptoms§ | O: 1.34 (0.80, 2.26)<br>F: 1.03 (0.63, 1.69)<br>M: 1.95 (0.62, 6.10) | -0.63 (-1.95, 0.69) |
| Distress from life-threatening experiences§ | O: 1.21 (0.62, 2.36)<br>F: 0.98 (0.41, 2.36)<br>M: 1.48 (0.63, 3.46) | -0.41 (-1.51, 0.68) |
| Have little interest in doing things§ | O: 1.03 (0.60, 1.76)<br>F: 0.85 (0.42, 1.71)<br>M: 1.26 (0.53, 2.99) | -0.39 (-1.57, 0.78) |
| Support reducing differences in income levels§ | O: 0.94 (0.48, 1.84)<br>F: 1.07 (0.40, 2.85)<br>M: 0.82 (0.29, 2.37) | 0.26 (-1.26, 1.79) |
| Feel at peace§ | O: 0.82 (0.47, 1.41)<br>F: 0.79 (0.38, 1.64)<br>M: 0.85 (0.36, 2.02) | -0.07 (-1.22, 1.09) |
| Government approval§ | O: 0.80 (0.50, 1.29)<br>F: 1.02 (0.50, 2.09)<br>M: 0.63 (0.28, 1.42) | 0.48 (-0.71, 1.68) |
| Say in government# | O: 0.80 (0.38, 1.66)<br>F: 0.69 (0.27, 1.77)<br>M: 0.92 (0.25, 3.46) | -0.29 (-2.02, 1.45) |
| Obeying the law§ | O: 0.26 (0.12, 0.57)***<br>F: 1.00 (0.35, 2.86)<br>M: 0.10 (0.04, 0.27)*** | 2.27 (0.85, 3.69)** |

*Poorer relationship quality with mother (bad or very bad relationship) – Large city or suburb of a large city*

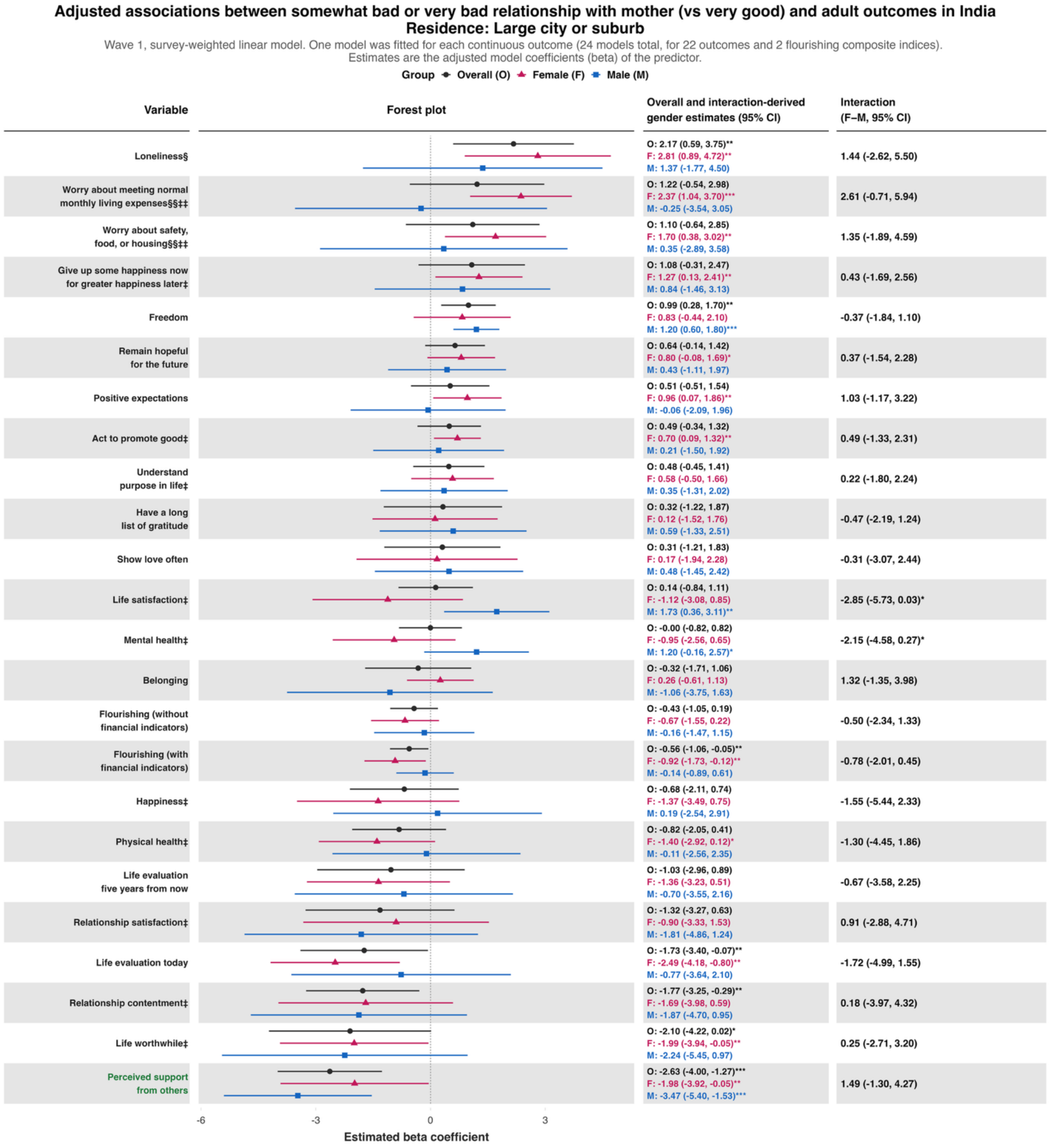

**Adjusted associations between somewhat bad or very bad relationship with mother (vs very good) and adult outcomes in India**
**Residence: Large city or suburb**

Wave 1, survey-weighted logistic model. One model was fitted for each binary outcome (7 models total). Estimates are adjusted odds ratios (OR) of the predictor.
The interaction column reports the difference in log-odds ratios (interaction beta) with 95% confidence intervals (95% CIs).

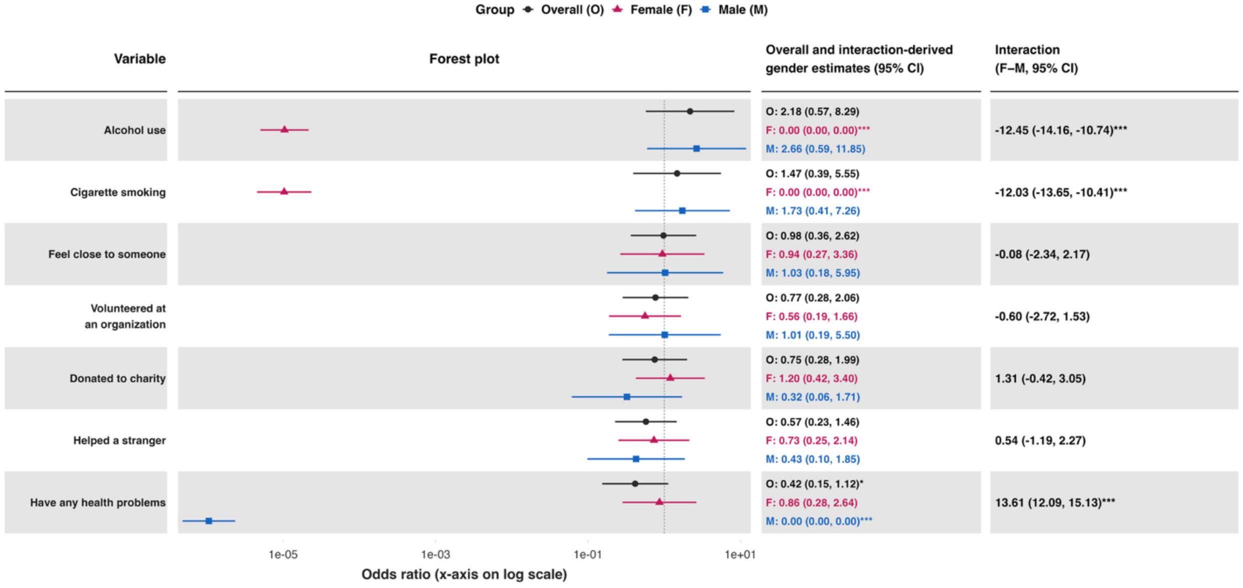

### Adjusted associations between somewhat bad or very bad relationship with mother (vs very good) and adult outcomes in India
### Residence: Large city or suburb

Wave 1, survey-weighted ordinal regression using the proportional odds model (`svyolr`). One model was fitted for each ordinal outcome (16 models total).
Estimates are adjusted cumulative odds ratios (cOR) of the predictor.
The interaction column reports the difference in log-cumulative odds ratios (interaction beta) with 95% confidence intervals (95% CIs).

**Group** ● Overall (O) ▲ Female (F) ■ Male (M)

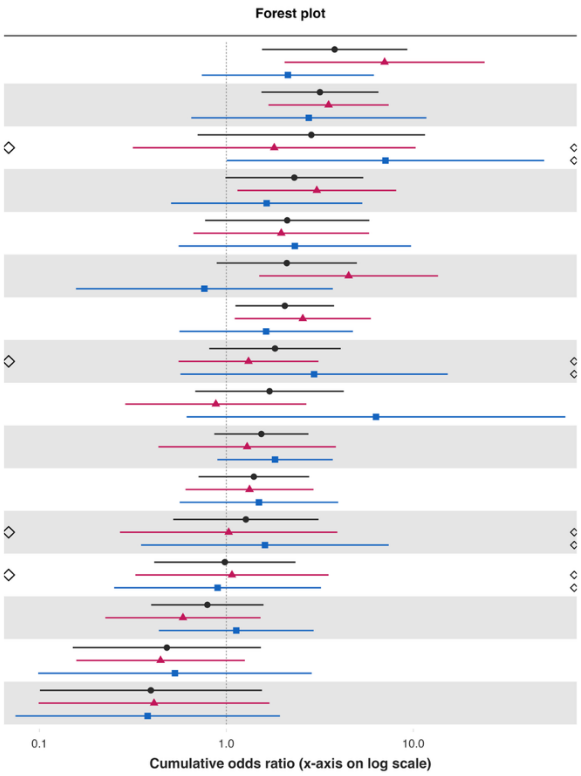


| Variable | Overall and interaction-derived gender estimates (95% CI) | Interaction (F–M, 95% CI) |
|---|---|---|
| Depressive symptoms§ | O: 3.79 (1.55, 9.29)**<br>F: 7.02 (2.05, 24.05)**<br>M: 2.14 (0.74, 6.16) | 1.19 (-0.15, 2.53)* |
| Extent of suffering§ | O: 3.17 (1.54, 6.51)**<br>F: 3.52 (1.68, 7.39)***<br>M: 2.76 (0.65, 11.75) | 0.24 (-1.43, 1.92) |
| Say in government# ◇ | O: 2.85 (0.70, 11.55)<br>◇ F: 1.80 (0.32, 10.29)<br>◇ M: 7.10 (1.01, 50.13)** | -1.37 (-4.14, 1.40) |
| Unable to control worry§ | O: 2.31 (0.99, 5.39)*<br>F: 3.05 (1.15, 8.10)**<br>M: 1.64 (0.51, 5.34) | 0.62 (-0.67, 1.91) |
| Feeling nervous or anxious§ | O: 2.12 (0.77, 5.81)<br>F: 1.97 (0.67, 5.80)<br>M: 2.33 (0.56, 9.73) | -0.17 (-1.68, 1.35) |
| Distress from life-threatening experiences§ | O: 2.11 (0.89, 4.99)*<br>F: 4.51 (1.50, 13.56)**<br>M: 0.76 (0.16, 3.71) | 1.78 (-0.31, 3.87)* |
| Various aspects of life are balanced§ | O: 2.05 (1.12, 3.77)**<br>F: 2.56 (1.11, 5.93)**<br>M: 1.63 (0.56, 4.75) | 0.45 (-1.04, 1.94) |
| Trust in others§ ◇ | O: 1.82 (0.81, 4.09)<br>◇ F: 1.31 (0.56, 3.11)<br>◇ M: 2.95 (0.57, 15.29) | -0.81 (-2.70, 1.09) |
| Feeling capable§ | O: 1.70 (0.68, 4.25)<br>F: 0.88 (0.29, 2.68)<br>M: 6.32 (0.61, 65.06) | -1.97 (-4.55, 0.60) |
| Have little interest in doing things§ | O: 1.54 (0.86, 2.75)<br>F: 1.29 (0.43, 3.85)<br>M: 1.82 (0.90, 3.71)* | -0.34 (-1.78, 1.09) |
| Feel at peace§ | O: 1.40 (0.71, 2.77)<br>F: 1.33 (0.61, 2.93)<br>M: 1.49 (0.56, 3.97) | -0.12 (-1.23, 1.00) |
| Forgiveness§ ◇ | O: 1.27 (0.52, 3.11)<br>◇ F: 1.03 (0.27, 3.93)<br>◇ M: 1.61 (0.35, 7.39) | -0.45 (-2.68, 1.79) |
| Perceived discrimination§ ◇ | O: 0.98 (0.41, 2.34)<br>◇ F: 1.07 (0.33, 3.52)<br>◇ M: 0.90 (0.25, 3.21) | 0.18 (-1.57, 1.92) |
| Government approval§ | O: 0.79 (0.40, 1.58)<br>F: 0.59 (0.23, 1.52)<br>M: 1.13 (0.44, 2.93) | -0.66 (-1.91, 0.60) |
| Obeying the law§ | O: 0.48 (0.15, 1.53)<br>F: 0.44 (0.16, 1.26)<br>M: 0.53 (0.10, 2.86) | -0.18 (-1.58, 1.23) |
| Support reducing differences in income levels§ | O: 0.39 (0.10, 1.55)<br>F: 0.41 (0.10, 1.70)<br>M: 0.38 (0.07, 1.94) | 0.08 (-1.28, 1.43) |

**Predictor:** ***Poorer relationship quality with father (bad or very bad relationship)***

***Poorer relationship quality with father (bad or very bad relationship) – Rural area or on a farm***

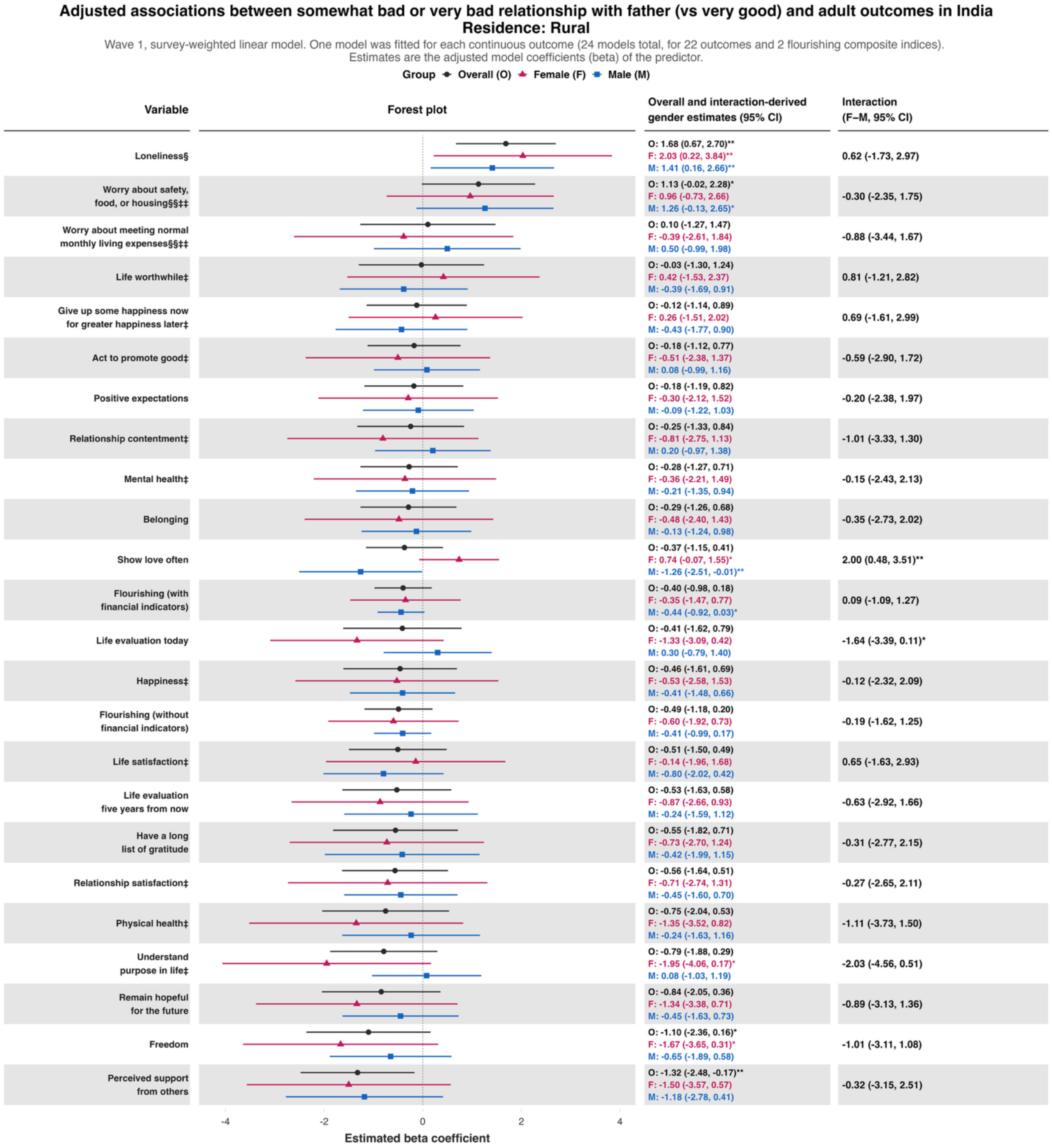

**Adjusted associations between somewhat bad or very bad relationship with father (vs very good) and adult outcomes in India**
**Residence: Rural**

Wave 1, survey-weighted logistic model. One model was fitted for each binary outcome (7 models total). Estimates are adjusted odds ratios (OR) of the predictor.
The interaction column reports the difference in log-odds ratios (interaction beta) with 95% confidence intervals (95% CIs).

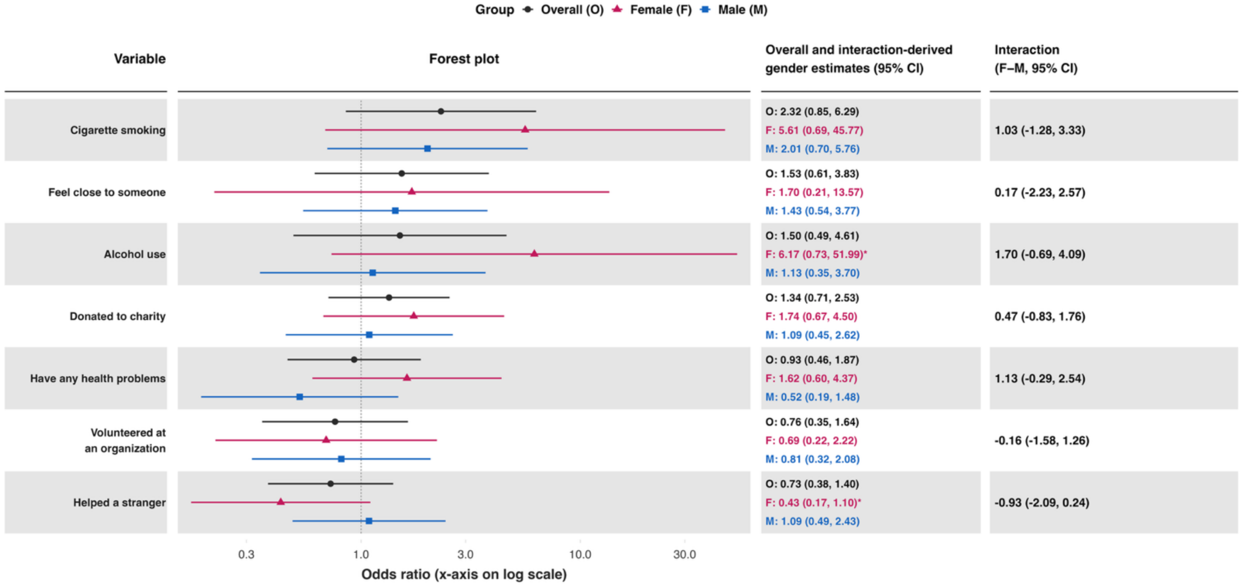

### Adjusted associations between somewhat bad or very bad relationship with father (vs very good) and adult outcomes in India
### Residence: Rural

Wave 1, survey-weighted ordinal regression using the proportional odds model (`svyolr`). One model was fitted for each ordinal outcome (16 models total).
Estimates are adjusted cumulative odds ratios (cOR) of the predictor.
The interaction column reports the difference in log-cumulative odds ratios (interaction beta) with 95% confidence intervals (95% CIs).

Group: Overall (O), Female (F), Male (M)

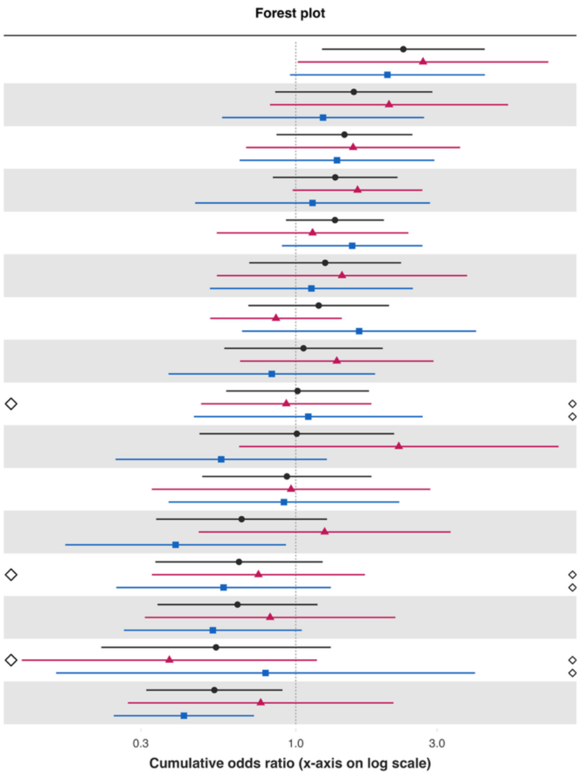


| Variable | Overall and interaction-derived gender estimates (95% CI) | Interaction (F–M, 95% CI) |
|---|---|---|
| Feel at peace§ | O: 2.31 (1.23, 4.34)**<br>F: 2.69 (1.02, 7.12)**<br>M: 2.04 (0.96, 4.35)* | 0.28 (-0.88, 1.43) |
| Feeling nervous or anxious§ | O: 1.57 (0.85, 2.89)<br>F: 2.06 (0.82, 5.20)<br>M: 1.24 (0.56, 2.71) | 0.51 (-0.69, 1.72) |
| Have little interest in doing things§ | O: 1.46 (0.86, 2.48)<br>F: 1.56 (0.68, 3.59)<br>M: 1.38 (0.65, 2.94) | 0.13 (-1.06, 1.31) |
| Distress from life-threatening experiences§ | O: 1.36 (0.84, 2.21)<br>F: 1.62 (0.98, 2.68)*<br>M: 1.14 (0.46, 2.84) | 0.35 (-0.76, 1.46) |
| Depressive symptoms§ | O: 1.36 (0.93, 1.98)<br>F: 1.14 (0.54, 2.40)<br>M: 1.55 (0.90, 2.68) | -0.31 (-1.35, 0.73) |
| Unable to control worry§ | O: 1.26 (0.70, 2.27)<br>F: 1.43 (0.54, 3.79)<br>M: 1.13 (0.51, 2.48) | 0.24 (-1.07, 1.54) |
| Trust in others§ | O: 1.20 (0.69, 2.06)<br>F: 0.86 (0.51, 1.43)<br>M: 1.64 (0.66, 4.06) | -0.65 (-1.70, 0.40) |
| Perceived discrimination§ | O: 1.06 (0.57, 1.96)<br>F: 1.37 (0.65, 2.91)<br>M: 0.83 (0.37, 1.85) | 0.50 (-0.50, 1.50) |
| Various aspects of life are balanced§ | O: 1.01 (0.58, 1.77)<br>F: 0.93 (0.48, 1.80)<br>M: 1.10 (0.45, 2.68) | -0.17 (-1.30, 0.96) |
| Extent of suffering§ | O: 1.01 (0.47, 2.15)<br>F: 2.23 (0.64, 7.70)<br>M: 0.56 (0.25, 1.27) | 1.38 (-0.05, 2.81)* |
| Say in government# | O: 0.93 (0.48, 1.80)<br>F: 0.96 (0.33, 2.84)<br>M: 0.91 (0.37, 2.23) | 0.06 (-1.41, 1.52) |
| Feeling capable§ | O: 0.66 (0.34, 1.27)<br>F: 1.25 (0.47, 3.33)<br>M: 0.39 (0.17, 0.93)** | 1.16 (-0.10, 2.41)* |
| Support reducing differences in income levels§ | O: 0.64 (0.34, 1.23)<br>F: 0.75 (0.33, 1.71)<br>M: 0.57 (0.25, 1.31) | 0.27 (-0.79, 1.33) |
| Forgiveness§ | O: 0.64 (0.34, 1.18)<br>F: 0.82 (0.31, 2.17)<br>M: 0.52 (0.26, 1.05)* | 0.44 (-0.64, 1.53) |
| Obeying the law§ | O: 0.54 (0.22, 1.31)<br>F: 0.37 (0.12, 1.18)*<br>M: 0.79 (0.16, 4.03) | -0.75 (-2.78, 1.28) |
| Government approval§ | O: 0.53 (0.31, 0.90)**<br>F: 0.76 (0.27, 2.14)<br>M: 0.42 (0.24, 0.72)** | 0.60 (-0.57, 1.76) |

*Poorer relationship quality with father (bad or very bad relationship) – Small town or village*

**Adjusted associations between somewhat bad or very bad relationship with father (vs very good) and adult outcomes in India**
**Residence: Small town**

Wave 1, survey-weighted linear model. One model was fitted for each continuous outcome (24 models total, for 22 outcomes and 2 flourishing composite indices).
Estimates are the adjusted model coefficients (beta) of the predictor.

Group ● Overall (O) ▲ Female (F) ■ Male (M)

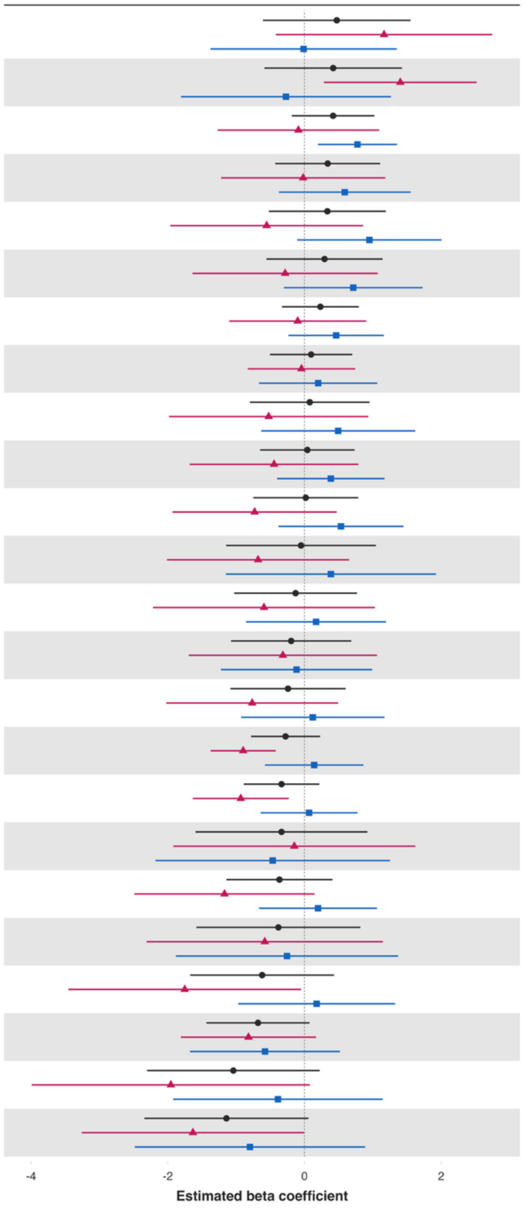


| Variable | Overall and interaction-derived gender estimates (95% CI) | Interaction (F–M, 95% CI) |
|---|---|---|
| Loneliness§ | O: 0.47 (-0.61, 1.55)<br>F: 1.16 (-0.42, 2.75)<br>M: -0.01 (-1.38, 1.35) | 1.18 (-0.84, 3.20) |
| Worry about meeting normal monthly living expenses§§‡‡ | O: 0.42 (-0.59, 1.43)<br>F: 1.40 (0.28, 2.52)**<br>M: -0.27 (-1.81, 1.27) | 1.67 (-0.23, 3.57)* |
| Freedom | O: 0.42 (-0.19, 1.02)<br>F: -0.09 (-1.27, 1.09)<br>M: 0.78 (0.20, 1.35)** | -0.86 (-2.17, 0.44) |
| Show love often | O: 0.34 (-0.43, 1.11)<br>F: -0.02 (-1.22, 1.18)<br>M: 0.59 (-0.37, 1.55) | -0.61 (-2.16, 0.95) |
| Have a long list of gratitude | O: 0.33 (-0.52, 1.19)<br>F: -0.55 (-1.96, 0.86)<br>M: 0.95 (-0.11, 2.01)* | -1.50 (-3.28, 0.28)* |
| Happiness‡ | O: 0.29 (-0.56, 1.14)<br>F: -0.28 (-1.64, 1.07)<br>M: 0.71 (-0.30, 1.73) | -1.00 (-2.69, 0.70) |
| Remain hopeful for the future | O: 0.23 (-0.33, 0.80)<br>F: -0.10 (-1.10, 0.90)<br>M: 0.46 (-0.24, 1.16) | -0.56 (-1.85, 0.72) |
| Belonging | O: 0.10 (-0.50, 0.70)<br>F: -0.04 (-0.83, 0.74)<br>M: 0.20 (-0.66, 1.06) | -0.24 (-1.42, 0.93) |
| Give up some happiness now for greater happiness later‡ | O: 0.08 (-0.80, 0.95)<br>F: -0.52 (-1.98, 0.93)<br>M: 0.49 (-0.63, 1.62) | -1.02 (-2.97, 0.94) |
| Positive expectations | O: 0.04 (-0.65, 0.73)<br>F: -0.45 (-1.68, 0.79)<br>M: 0.39 (-0.40, 1.17) | -0.83 (-2.34, 0.68) |
| Understand purpose in life‡ | O: 0.02 (-0.75, 0.79)<br>F: -0.73 (-1.93, 0.47)<br>M: 0.53 (-0.38, 1.45) | -1.26 (-2.72, 0.20)* |
| Life evaluation today | O: -0.05 (-1.15, 1.05)<br>F: -0.68 (-2.01, 0.65)<br>M: 0.39 (-1.15, 1.92) | -1.07 (-3.03, 0.90) |
| Life evaluation five years from now | O: -0.13 (-1.03, 0.77)<br>F: -0.59 (-2.22, 1.03)<br>M: 0.17 (-0.85, 1.19) | -0.76 (-2.68, 1.16) |
| Life satisfaction‡ | O: -0.19 (-1.07, 0.69)<br>F: -0.32 (-1.69, 1.06)<br>M: -0.12 (-1.22, 0.99) | -0.20 (-1.95, 1.54) |
| Relationship satisfaction‡ | O: -0.24 (-1.08, 0.60)<br>F: -0.76 (-2.02, 0.49)<br>M: 0.12 (-0.93, 1.17) | -0.89 (-2.49, 0.72) |
| Flourishing (with financial indicators) | O: -0.28 (-0.78, 0.23)<br>F: -0.90 (-1.37, -0.42)***<br>M: 0.14 (-0.58, 0.86) | -1.04 (-1.90, -0.18)** |
| Flourishing (without financial indicators) | O: -0.34 (-0.89, 0.22)<br>F: -0.93 (-1.63, -0.23)**<br>M: 0.07 (-0.64, 0.78) | -1.00 (-1.99, -0.01)** |
| Worry about safety, food, or housing§§‡‡ | O: -0.34 (-1.59, 0.92)<br>F: -0.15 (-1.92, 1.62)<br>M: -0.46 (-2.18, 1.25) | 0.32 (-2.11, 2.74) |
| Act to promote good‡ | O: -0.37 (-1.14, 0.41)<br>F: -1.17 (-2.49, 0.15)*<br>M: 0.20 (-0.66, 1.06) | -1.37 (-2.97, 0.23)* |
| Life worthwhile‡ | O: -0.38 (-1.58, 0.82)<br>F: -0.58 (-2.31, 1.15)<br>M: -0.26 (-1.88, 1.37) | -0.33 (-2.69, 2.04) |
| Physical health‡ | O: -0.62 (-1.67, 0.43)<br>F: -1.75 (-3.45, -0.05)**<br>M: 0.18 (-0.97, 1.33) | -1.93 (-3.94, 0.08)* |
| Relationship contentment‡ | O: -0.68 (-1.43, 0.07)*<br>F: -0.82 (-1.81, 0.17)<br>M: -0.58 (-1.67, 0.52) | -0.24 (-1.74, 1.25) |
| Perceived support from others | O: -1.04 (-2.30, 0.22)<br>F: -1.95 (-3.99, 0.08)*<br>M: -0.39 (-1.92, 1.15) | -1.57 (-4.03, 0.90) |
| Mental health‡ | O: -1.14 (-2.34, 0.06)*<br>F: -1.63 (-3.26, -0.01)*<br>M: -0.80 (-2.48, 0.89) | -0.83 (-3.17, 1.50) |

**Adjusted associations between somewhat bad or very bad relationship with father (vs very good) and adult outcomes in India**
**Residence: Small town**

Wave 1, survey-weighted logistic model. One model was fitted for each binary outcome (7 models total). Estimates are adjusted odds ratios (OR) of the predictor.
The interaction column reports the difference in log-odds ratios (interaction beta) with 95% confidence intervals (95% CIs).

Group: Overall (O), Female (F), Male (M)

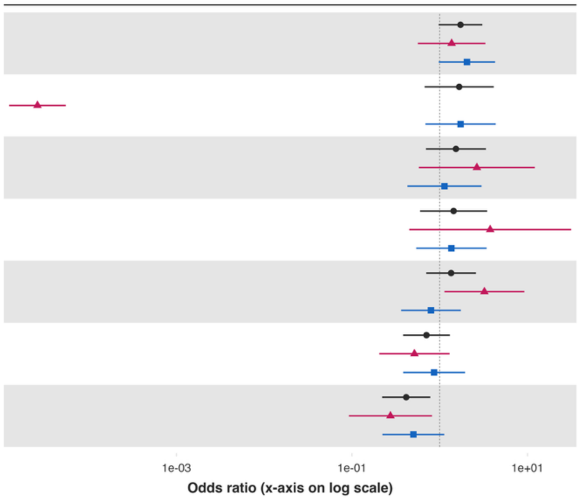


| Variable | Overall and interaction-derived gender estimates (95% CI) | Interaction (F–M, 95% CI) |
|---|---|---|
| Have any health problems | O: 1.73 (0.97, 3.06)*<br>F: 1.37 (0.56, 3.32)<br>M: 2.05 (0.98, 4.29)* | -0.40 (-1.54, 0.73) |
| Alcohol use | O: 1.67 (0.67, 4.13)<br>F: 0.00 (0.00, 0.00)***<br>M: 1.73 (0.69, 4.35) | -11.10 (-12.28, -9.92)*** |
| Feel close to someone | O: 1.53 (0.70, 3.36)<br>F: 2.65 (0.58, 12.16)<br>M: 1.14 (0.43, 3.00) | 0.85 (-1.00, 2.70) |
| Cigarette smoking | O: 1.44 (0.60, 3.48)<br>F: 3.76 (0.45, 31.48)<br>M: 1.36 (0.54, 3.42) | 1.02 (-1.29, 3.33) |
| Helped a stranger | O: 1.35 (0.70, 2.59)<br>F: 3.23 (1.13, 9.22)**<br>M: 0.80 (0.36, 1.74) | 1.40 (0.11, 2.69)** |
| Donated to charity | O: 0.71 (0.38, 1.31)<br>F: 0.52 (0.20, 1.30)<br>M: 0.86 (0.38, 1.95) | -0.52 (-1.71, 0.68) |
| Volunteered at an organization | O: 0.42 (0.22, 0.78)**<br>F: 0.27 (0.09, 0.82)**<br>M: 0.50 (0.22, 1.13)* | -0.60 (-1.98, 0.78) |

**Adjusted associations between somewhat bad or very bad relationship with father (vs very good) and adult outcomes in India**
**Residence: Small town**

Wave 1, survey-weighted ordinal regression using the proportional odds model (`svyolr`). One model was fitted for each ordinal outcome (16 models total).
Estimates are adjusted cumulative odds ratios (cOR) of the predictor.
The interaction column reports the difference in log-cumulative odds ratios (interaction beta) with 95% confidence intervals (95% CIs).

Group ● Overall (O) ▲ Female (F) ■ Male (M)

Forest plot

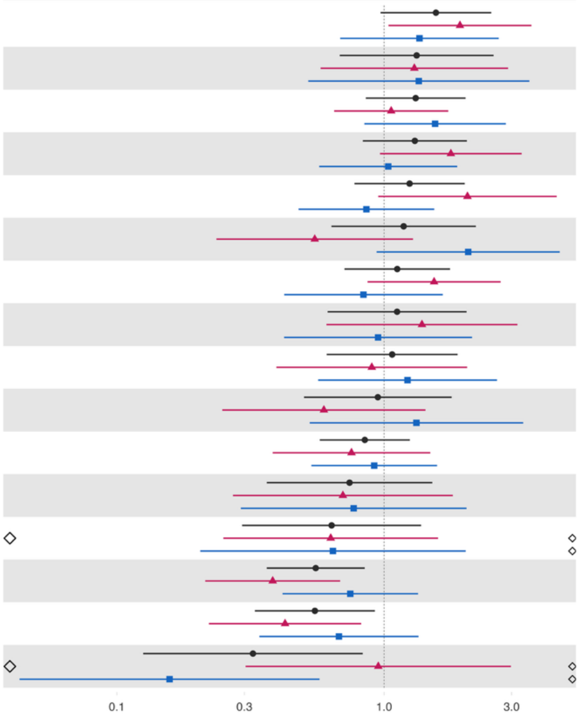


| Variable | Overall and interaction-derived gender estimates (95% CI) | Interaction (F–M, 95% CI) |
|---|---|---|
| Perceived discrimination§ | O: 1.56 (0.97, 2.52)*<br>F: 1.92 (1.04, 3.56)**<br>M: 1.36 (0.68, 2.68) | 0.35 (-0.57, 1.27) |
| Extent of suffering§ | O: 1.32 (0.68, 2.57)<br>F: 1.30 (0.58, 2.91)<br>M: 1.35 (0.52, 3.50) | -0.04 (-1.24, 1.17) |
| Have little interest in doing things§ | O: 1.31 (0.85, 2.02)<br>F: 1.06 (0.65, 1.74)<br>M: 1.55 (0.84, 2.85) | -0.38 (-1.12, 0.36) |
| Feeling nervous or anxious§ | O: 1.30 (0.83, 2.04)<br>F: 1.78 (0.97, 3.27)*<br>M: 1.04 (0.57, 1.88) | 0.54 (-0.27, 1.35) |
| Unable to control worry§ | O: 1.25 (0.77, 2.00)<br>F: 2.05 (0.95, 4.42)*<br>M: 0.86 (0.48, 1.54) | 0.87 (-0.10, 1.85)* |
| Various aspects of life are balanced§ | O: 1.18 (0.63, 2.20)<br>F: 0.55 (0.24, 1.28)<br>M: 2.06 (0.94, 4.54)* | -1.32 (-2.45, -0.20)** |
| Depressive symptoms§ | O: 1.12 (0.71, 1.76)<br>F: 1.54 (0.87, 2.73)<br>M: 0.84 (0.42, 1.66) | 0.61 (-0.30, 1.51) |
| Distress from life-threatening experiences§ | O: 1.12 (0.61, 2.03)<br>F: 1.39 (0.61, 3.15)<br>M: 0.95 (0.42, 2.13) | 0.38 (-0.76, 1.52) |
| Feeling capable§ | O: 1.07 (0.61, 1.88)<br>F: 0.90 (0.40, 2.05)<br>M: 1.22 (0.57, 2.64) | -0.31 (-1.43, 0.82) |
| Say in government# | O: 0.95 (0.50, 1.79)<br>F: 0.60 (0.25, 1.43)<br>M: 1.32 (0.53, 3.31) | -0.80 (-2.09, 0.49) |
| Trust in others§ | O: 0.85 (0.57, 1.25)<br>F: 0.75 (0.38, 1.49)<br>M: 0.92 (0.53, 1.58) | -0.20 (-1.13, 0.74) |
| Forgiveness§ | O: 0.74 (0.36, 1.51)<br>F: 0.70 (0.27, 1.81)<br>M: 0.77 (0.29, 2.03) | -0.09 (-1.42, 1.23) |
| Support reducing differences in income levels§ | O: 0.64 (0.29, 1.37)<br>F: 0.63 (0.25, 1.59)<br>M: 0.64 (0.21, 2.02) | -0.02 (-1.49, 1.45) |
| Government approval§ | O: 0.55 (0.36, 0.85)**<br>F: 0.38 (0.21, 0.68)**<br>M: 0.75 (0.42, 1.34) | -0.67 (-1.49, 0.15) |
| Feel at peace§ | O: 0.55 (0.33, 0.92)**<br>F: 0.43 (0.22, 0.82)**<br>M: 0.68 (0.34, 1.34) | -0.46 (-1.32, 0.39) |
| Obeying the law§ | O: 0.32 (0.13, 0.83)**<br>F: 0.95 (0.30, 2.98)<br>M: 0.16 (0.04, 0.57)** | 1.80 (0.08, 3.52)** |

*Poorer relationship quality with father (bad or very bad relationship) – Large city or suburb of a large city*

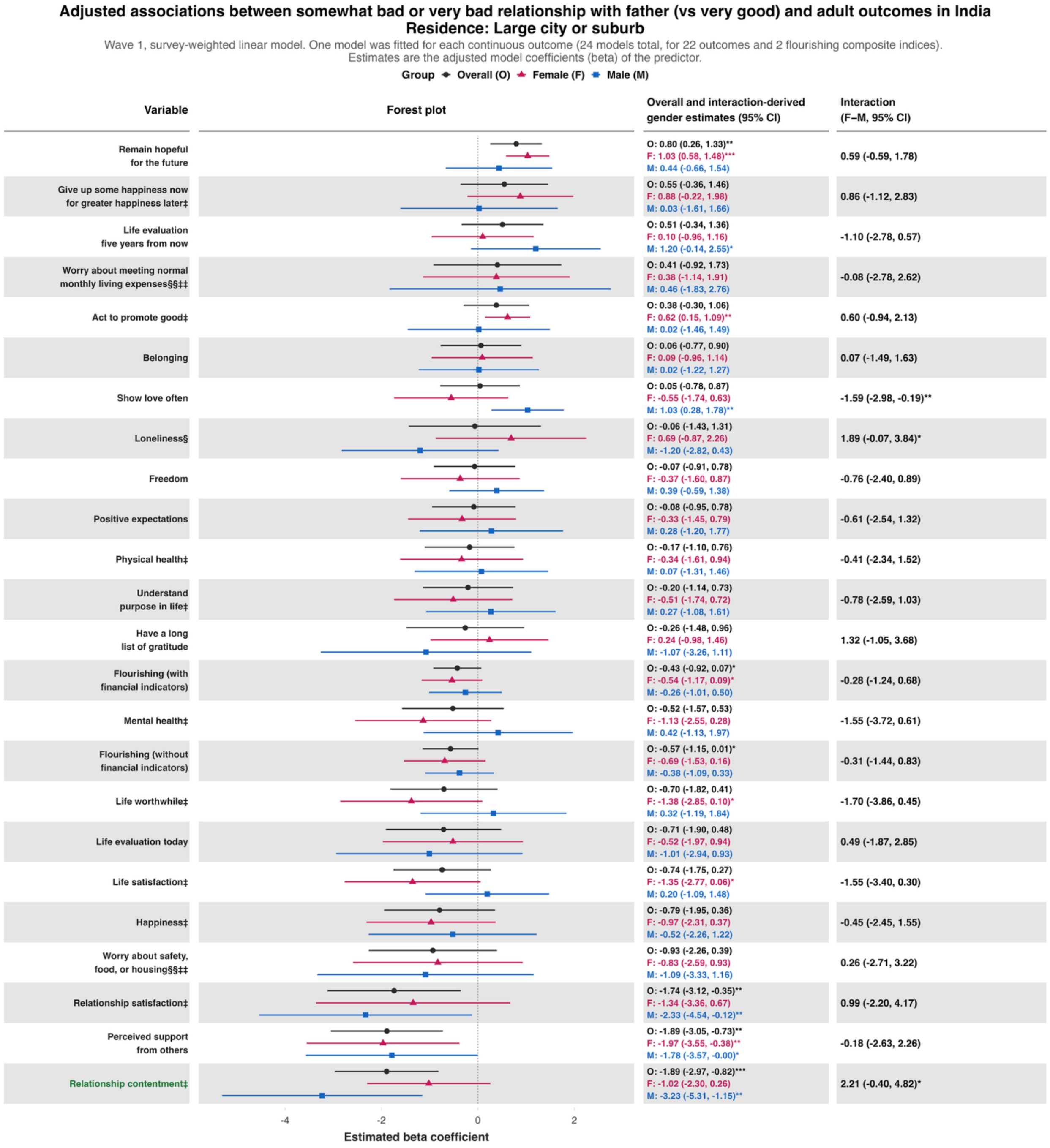

**Adjusted associations between somewhat bad or very bad relationship with father (vs very good) and adult outcomes in India**
**Residence: Large city or suburb**

Wave 1, survey-weighted logistic model. One model was fitted for each binary outcome (7 models total). Estimates are adjusted odds ratios (OR) of the predictor.
The interaction column reports the difference in log-odds ratios (interaction beta) with 95% confidence intervals (95% CIs).

Group — Overall (O) — Female (F) — Male (M)

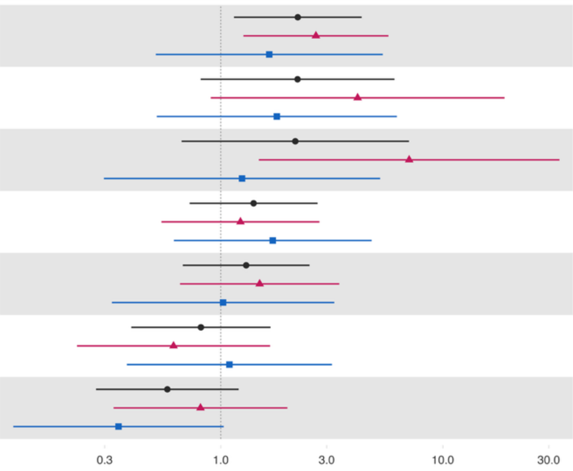


| Variable | Overall and interaction-derived gender estimates (95% CI) | Interaction (F–M, 95% CI) |
|---|---|---|
| Helped a stranger | O: 2.22 (1.15, 4.31)**<br>F: 2.68 (1.26, 5.69)**<br>M: 1.65 (0.51, 5.36) | 0.48 (-0.88, 1.85) |
| Alcohol use | O: 2.22 (0.81, 6.05)<br>F: 4.13 (0.90, 18.95)*<br>M: 1.79 (0.51, 6.21) | 0.84 (-1.26, 2.93) |
| Cigarette smoking | O: 2.17 (0.67, 7.04)<br>F: 7.05 (1.48, 33.52)**<br>M: 1.25 (0.30, 5.22) | 1.73 (-0.74, 4.20) |
| Donated to charity | O: 1.41 (0.72, 2.73)<br>F: 1.23 (0.54, 2.78)<br>M: 1.71 (0.62, 4.78) | -0.33 (-1.61, 0.94) |
| Have any health problems | O: 1.30 (0.67, 2.51)<br>F: 1.50 (0.65, 3.42)<br>M: 1.02 (0.32, 3.24) | 0.38 (-1.05, 1.81) |
| Volunteered at an organization | O: 0.81 (0.40, 1.67)<br>F: 0.61 (0.23, 1.67)<br>M: 1.09 (0.38, 3.16) | -0.58 (-2.05, 0.89) |
| Feel close to someone | O: 0.57 (0.27, 1.20)<br>F: 0.81 (0.33, 1.99)<br>M: 0.35 (0.12, 1.03)* | 0.85 (-0.44, 2.14) |

### Adjusted associations between somewhat bad or very bad relationship with father (vs very good) and adult outcomes in India
### Residence: Large city or suburb

Wave 1, survey-weighted ordinal regression using the proportional odds model (`svyolr`). One model was fitted for each ordinal outcome (16 models total).
Estimates are adjusted cumulative odds ratios (cOR) of the predictor.
The interaction column reports the difference in log-cumulative odds ratios (interaction beta) with 95% confidence intervals (95% CIs).

Group ● Overall (O) ▲ Female (F) ■ Male (M)

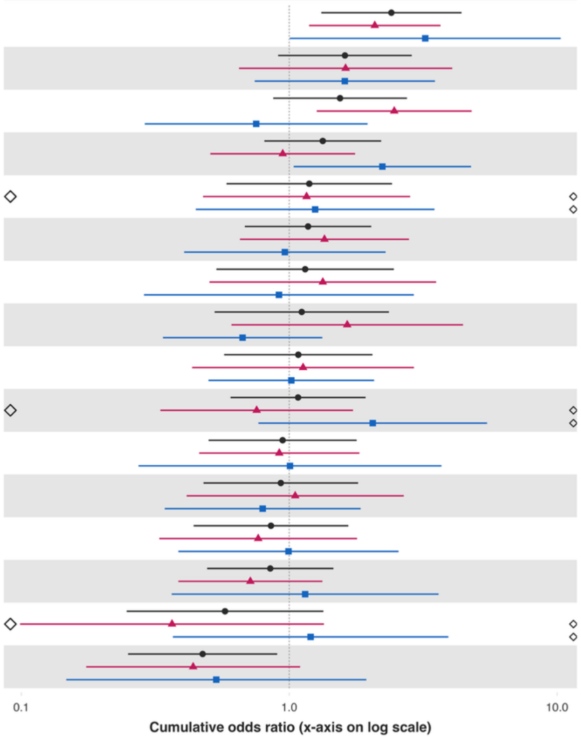


| Variable | Overall and interaction-derived gender estimates (95% CI) | Interaction (F–M, 95% CI) |
|---|---|---|
| Have little interest in doing things§ | O: 2.41 (1.32, 4.39)**<br>F: 2.09 (1.19, 3.67)**<br>M: 3.22 (1.01, 10.31)** | -0.43 (-1.59, 0.72) |
| Extent of suffering§ | O: 1.61 (0.91, 2.87)<br>F: 1.62 (0.65, 4.06)<br>M: 1.61 (0.74, 3.50) | 0.01 (-1.29, 1.30) |
| Distress from life-threatening experiences§ | O: 1.55 (0.87, 2.75)<br>F: 2.47 (1.27, 4.80)**<br>M: 0.75 (0.29, 1.96) | 1.19 (0.06, 2.31)** |
| Unable to control worry§ | O: 1.33 (0.81, 2.20)<br>F: 0.95 (0.51, 1.76)<br>M: 2.23 (1.04, 4.77)** | -0.86 (-1.89, 0.18) |
| Feeling capable§ | O: 1.19 (0.58, 2.42)<br>F: 1.16 (0.48, 2.83)<br>M: 1.25 (0.45, 3.48) | -0.07 (-1.34, 1.20) |
| Depressive symptoms§ | O: 1.18 (0.68, 2.02)<br>F: 1.35 (0.65, 2.80)<br>M: 0.96 (0.41, 2.29) | 0.34 (-0.81, 1.49) |
| Say in government# | O: 1.15 (0.53, 2.46)<br>F: 1.33 (0.50, 3.53)<br>M: 0.92 (0.29, 2.92) | 0.38 (-1.13, 1.88) |
| Trust in others§ | O: 1.11 (0.53, 2.36)<br>F: 1.65 (0.61, 4.45)<br>M: 0.67 (0.34, 1.33) | 0.90 (-0.17, 1.97) |
| Various aspects of life are balanced§ | O: 1.08 (0.57, 2.04)<br>F: 1.13 (0.43, 2.92)<br>M: 1.02 (0.50, 2.07) | 0.10 (-1.10, 1.30) |
| Feel at peace§ | O: 1.08 (0.60, 1.93)<br>F: 0.76 (0.33, 1.73)<br>M: 2.05 (0.77, 5.48) | -1.00 (-2.29, 0.30) |
| Feeling nervous or anxious§ | O: 0.94 (0.50, 1.78)<br>F: 0.92 (0.46, 1.83)<br>M: 1.01 (0.27, 3.70) | -0.09 (-1.58, 1.39) |
| Perceived discrimination§ | O: 0.93 (0.48, 1.81)<br>F: 1.05 (0.41, 2.68)<br>M: 0.80 (0.34, 1.85) | 0.28 (-0.92, 1.47) |
| Government approval§ | O: 0.85 (0.44, 1.66)<br>F: 0.77 (0.33, 1.79)<br>M: 0.99 (0.39, 2.56) | -0.26 (-1.49, 0.97) |
| Forgiveness§ | O: 0.85 (0.49, 1.46)<br>F: 0.72 (0.39, 1.33)<br>M: 1.15 (0.36, 3.61) | -0.47 (-1.80, 0.86) |
| Support reducing differences in income levels§ | O: 0.58 (0.25, 1.34)<br>F: 0.37 (0.10, 1.35)<br>M: 1.20 (0.37, 3.93) | -1.19 (-3.19, 0.80) |
| Obeying the law§ | O: 0.48 (0.25, 0.90)**<br>F: 0.44 (0.17, 1.10)*<br>M: 0.53 (0.15, 1.94) | -0.20 (-1.98, 1.58) |

**Predictor:** ***Poor self-rated health while growing up***

***Poor self-rated health while growing up – Rural area or on a farm***

### Adjusted associations between poor self-rated childhood health (vs excellent) and adult outcomes in India
### Residence: Rural

Wave 1, survey-weighted linear model. One model was fitted for each continuous outcome (24 models total, for 22 outcomes and 2 flourishing composite indices). Estimates are the adjusted model coefficients (beta) of the predictor.

Group ◆ Overall (O) ▲ Female (F) ■ Male (M)

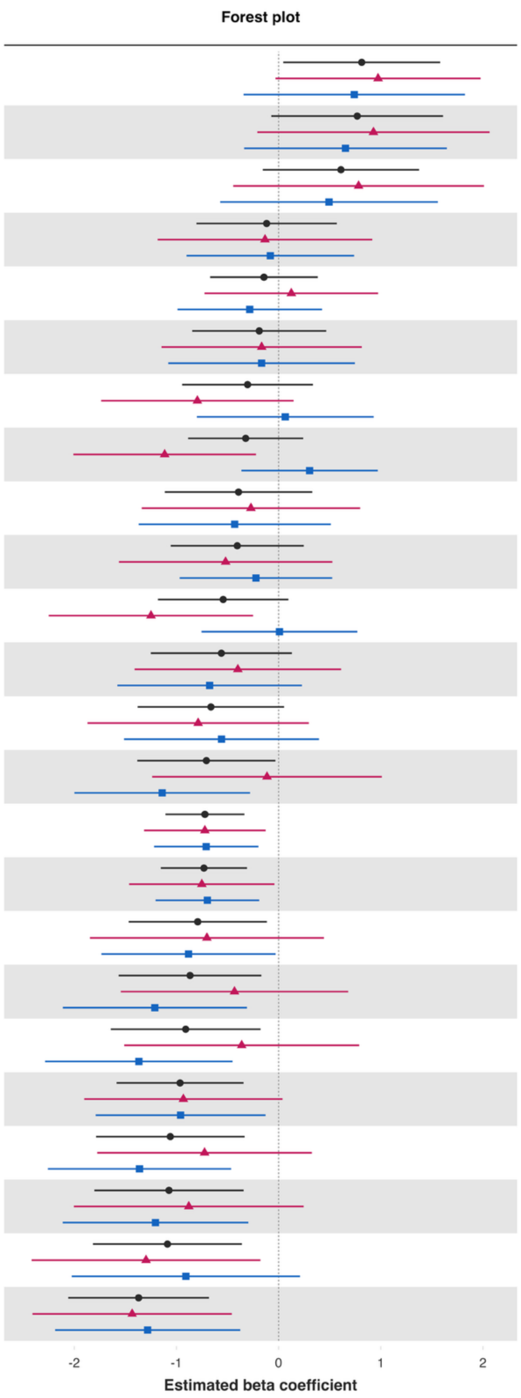


| Variable | Overall and interaction-derived gender estimates (95% CI) | Interaction (F–M, 95% CI) |
|---|---|---|
| Loneliness§ | O: 0.81 (0.05, 1.58)**<br>F: 0.97 (-0.03, 1.98)*<br>M: 0.74 (-0.34, 1.82) | 0.23 (-1.20, 1.67) |
| Worry about meeting normal monthly living expenses§§‡‡ | O: 0.77 (-0.07, 1.61)*<br>F: 0.93 (-0.21, 2.06)<br>M: 0.65 (-0.34, 1.65) | 0.27 (-1.01, 1.56) |
| Worry about safety, food, or housing§§‡‡ | O: 0.61 (-0.15, 1.37)<br>F: 0.78 (-0.44, 2.01)<br>M: 0.49 (-0.57, 1.56) | 0.29 (-1.40, 1.97) |
| Positive expectations | O: -0.12 (-0.80, 0.57)<br>F: -0.13 (-1.18, 0.92)<br>M: -0.08 (-0.90, 0.74) | -0.05 (-1.30, 1.20) |
| Show love often | O: -0.14 (-0.67, 0.38)<br>F: 0.12 (-0.73, 0.97)<br>M: -0.28 (-0.99, 0.42) | 0.41 (-0.74, 1.55) |
| Perceived support from others | O: -0.19 (-0.85, 0.47)<br>F: -0.17 (-1.15, 0.81)<br>M: -0.17 (-1.08, 0.75) | 0.00 (-1.36, 1.36) |
| Life evaluation five years from now | O: -0.30 (-0.94, 0.34)<br>F: -0.80 (-1.74, 0.15)*<br>M: 0.06 (-0.80, 0.93) | -0.86 (-2.14, 0.42) |
| Act to promote good‡ | O: -0.32 (-0.89, 0.24)<br>F: -1.12 (-2.01, -0.22)**<br>M: 0.30 (-0.36, 0.97) | -1.42 (-2.54, -0.30)** |
| Have a long list of gratitude | O: -0.39 (-1.11, 0.33)<br>F: -0.27 (-1.34, 0.80)<br>M: -0.43 (-1.37, 0.51) | 0.16 (-1.24, 1.55) |
| Understand purpose in life‡ | O: -0.40 (-1.06, 0.25)<br>F: -0.52 (-1.56, 0.53)<br>M: -0.22 (-0.97, 0.52) | -0.30 (-1.54, 0.95) |
| Relationship contentment‡ | O: -0.54 (-1.18, 0.09)*<br>F: -1.25 (-2.25, -0.25)**<br>M: 0.01 (-0.75, 0.77) | -1.26 (-2.50, -0.02)** |
| Give up some happiness now for greater happiness later‡ | O: -0.56 (-1.25, 0.13)<br>F: -0.40 (-1.41, 0.61)<br>M: -0.68 (-1.58, 0.23) | 0.28 (-1.06, 1.61) |
| Relationship satisfaction‡ | O: -0.66 (-1.38, 0.05)*<br>F: -0.79 (-1.87, 0.30)<br>M: -0.56 (-1.51, 0.40) | -0.23 (-1.66, 1.21) |
| Mental health‡ | O: -0.71 (-1.38, -0.03)**<br>F: -0.11 (-1.24, 1.01)<br>M: -1.14 (-2.00, -0.28)** | 1.03 (-0.42, 2.48) |
| Flourishing (with financial indicators) | O: -0.72 (-1.11, -0.34)***<br>F: -0.72 (-1.32, -0.13)**<br>M: -0.71 (-1.22, -0.20)** | -0.01 (-0.80, 0.77) |
| Flourishing (without financial indicators) | O: -0.73 (-1.15, -0.31)***<br>F: -0.75 (-1.46, -0.04)**<br>M: -0.70 (-1.20, -0.19)** | -0.05 (-0.93, 0.82) |
| Happiness‡ | O: -0.79 (-1.47, -0.12)**<br>F: -0.70 (-1.85, 0.44)<br>M: -0.88 (-1.73, -0.03)** | 0.18 (-1.28, 1.63) |
| Life evaluation today | O: -0.87 (-1.57, -0.17)**<br>F: -0.43 (-1.55, 0.68)<br>M: -1.21 (-2.11, -0.31)** | 0.78 (-0.66, 2.21) |
| Life satisfaction‡ | O: -0.91 (-1.64, -0.18)**<br>F: -0.36 (-1.51, 0.79)<br>M: -1.37 (-2.29, -0.45)** | 1.01 (-0.44, 2.46) |
| Remain hopeful for the future | O: -0.96 (-1.59, -0.34)**<br>F: -0.93 (-1.90, 0.04)*<br>M: -0.96 (-1.79, -0.13)** | 0.03 (-1.28, 1.34) |
| Physical health‡ | O: -1.06 (-1.79, -0.33)**<br>F: -0.72 (-1.78, 0.33)<br>M: -1.36 (-2.26, -0.46)** | 0.64 (-0.66, 1.93) |
| Life worthwhile‡ | O: -1.07 (-1.80, -0.34)**<br>F: -0.88 (-2.00, 0.25)<br>M: -1.21 (-2.11, -0.30)** | 0.33 (-1.08, 1.73) |
| Belonging | O: -1.09 (-1.82, -0.36)**<br>F: -1.30 (-2.42, -0.18)**<br>M: -0.91 (-2.02, 0.21) | -0.39 (-2.09, 1.31) |
| Freedom | O: -1.37 (-2.06, -0.68)***<br>F: -1.43 (-2.41, -0.46)**<br>M: -1.28 (-2.19, -0.38)** | -0.15 (-1.44, 1.14) |

### Adjusted associations between poor self-rated childhood health (vs excellent) and adult outcomes in India
### Residence: Rural

Wave 1, survey-weighted logistic model. One model was fitted for each binary outcome (7 models total). Estimates are adjusted odds ratios (OR) of the predictor.
The interaction column reports the difference in log-odds ratios (interaction beta) with 95% confidence intervals (95% CIs).

**Group** Overall (O) Female (F) Male (M)

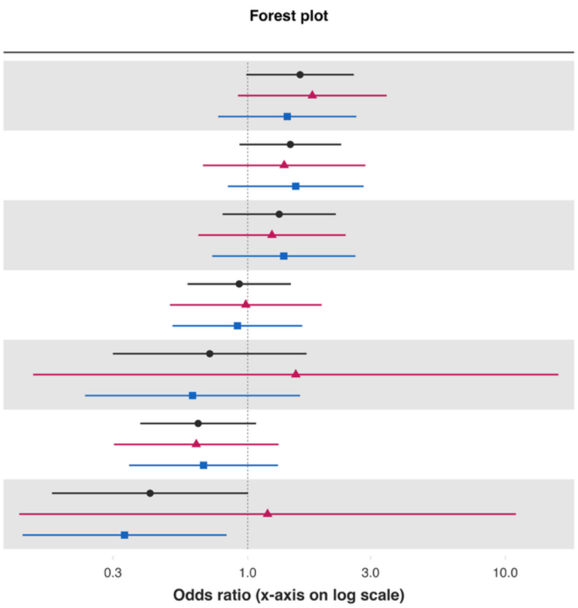


| Variable | Overall and interaction-derived gender estimates (95% CI) | Interaction (F–M, 95% CI) |
|---|---|---|
| Donated to charity | O: 1.60 (0.99, 2.58)*<br>F: 1.78 (0.92, 3.47)*<br>M: 1.42 (0.77, 2.64) | 0.22 (-0.61, 1.05) |
| Have any health problems | O: 1.46 (0.93, 2.31)<br>F: 1.39 (0.67, 2.86)<br>M: 1.54 (0.84, 2.82) | -0.10 (-1.06, 0.85) |
| Volunteered at an organization | O: 1.33 (0.80, 2.20)<br>F: 1.24 (0.64, 2.40)<br>M: 1.38 (0.73, 2.62) | -0.11 (-0.91, 0.70) |
| Helped a stranger | O: 0.93 (0.58, 1.47)<br>F: 0.98 (0.50, 1.94)<br>M: 0.91 (0.51, 1.63) | 0.07 (-0.76, 0.91) |
| Alcohol use | O: 0.71 (0.30, 1.69)<br>F: 1.54 (0.15, 16.09)<br>M: 0.61 (0.23, 1.60) | 0.92 (-1.70, 3.55) |
| Feel close to someone | O: 0.64 (0.38, 1.08)*<br>F: 0.63 (0.30, 1.32)<br>M: 0.67 (0.35, 1.31) | -0.07 (-1.00, 0.87) |
| Cigarette smoking | O: 0.42 (0.17, 1.00)*<br>F: 1.19 (0.13, 11.00)<br>M: 0.33 (0.13, 0.83)** | 1.28 (-1.16, 3.72) |

### Adjusted associations between poor self-rated childhood health (vs excellent) and adult outcomes in India
### Residence: Rural

Wave 1, survey-weighted ordinal regression using the proportional odds model (`svyolr`). One model was fitted for each ordinal outcome (16 models total).
Estimates are adjusted cumulative odds ratios (cOR) of the predictor.
The interaction column reports the difference in log-cumulative odds ratios (interaction beta) with 95% confidence intervals (95% CIs).

**Group** ● Overall (O) ▲ Female (F) ■ Male (M)

**Forest plot**

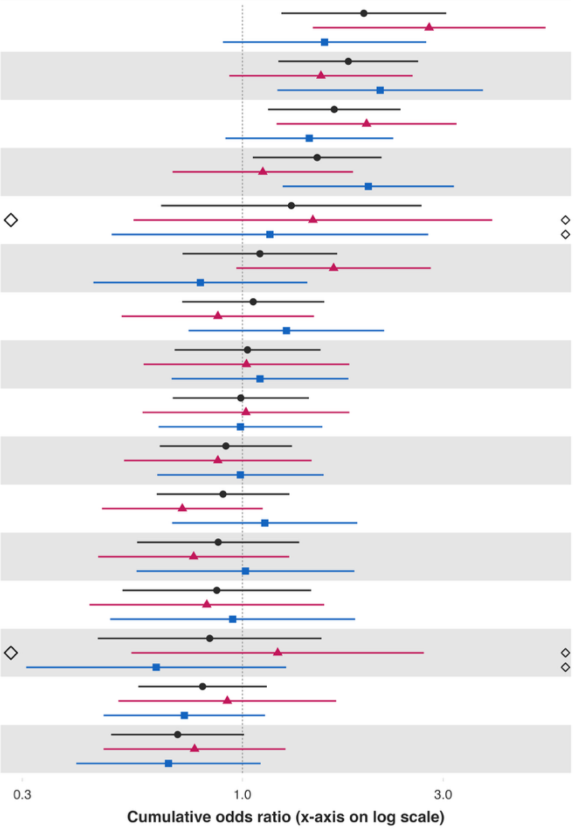


| Variable | Overall and interaction-derived gender estimates (95% CI) | Interaction (F–M, 95% CI) |
|---|---|---|
| Extent of suffering§ | O: 1.94 (1.24, 3.05)**<br>F: 2.78 (1.47, 5.25)**<br>M: 1.57 (0.90, 2.73) | 0.57 (-0.21, 1.36) |
| Feeling nervous or anxious§ | O: 1.79 (1.22, 2.62)**<br>F: 1.54 (0.93, 2.54)*<br>M: 2.13 (1.21, 3.73)** | -0.32 (-1.08, 0.43) |
| Distress from e-threatening experiences§ | O: 1.65 (1.15, 2.37)**<br>F: 1.97 (1.21, 3.23)**<br>M: 1.44 (0.91, 2.28) | 0.31 (-0.31, 0.93) |
| Depressive symptoms§ | O: 1.51 (1.06, 2.14)**<br>F: 1.12 (0.68, 1.83)<br>M: 1.99 (1.25, 3.18)** | -0.58 (-1.24, 0.08)* |
| Obeying the law§ | O: 1.31 (0.64, 2.66)<br>F: 1.47 (0.55, 3.92)<br>M: 1.16 (0.49, 2.76) | 0.24 (-0.93, 1.40) |
| Perceived discrimination§ | O: 1.10 (0.72, 1.68)<br>F: 1.65 (0.97, 2.80)*<br>M: 0.79 (0.44, 1.43) | 0.73 (-0.02, 1.48)* |
| Unable to control worry§ | O: 1.06 (0.72, 1.56)<br>F: 0.87 (0.52, 1.48)<br>M: 1.27 (0.74, 2.17) | -0.37 (-1.11, 0.36) |
| Forgiveness§ | O: 1.03 (0.69, 1.53)<br>F: 1.02 (0.58, 1.79)<br>M: 1.10 (0.68, 1.79) | -0.07 (-0.74, 0.59) |
| Feeling capable§ | O: 0.99 (0.68, 1.44)<br>F: 1.02 (0.58, 1.79)<br>M: 0.99 (0.63, 1.55) | 0.03 (-0.65, 0.71) |
| Various aspects of life are balanced§ | O: 0.91 (0.64, 1.31)<br>F: 0.87 (0.52, 1.46)<br>M: 0.99 (0.63, 1.56) | -0.12 (-0.78, 0.53) |
| Have little interest in doing things§ | O: 0.90 (0.63, 1.29)<br>F: 0.72 (0.46, 1.12)<br>M: 1.13 (0.68, 1.88) | -0.45 (-1.08, 0.18) |
| Government approval§ | O: 0.88 (0.56, 1.37)<br>F: 0.77 (0.45, 1.29)<br>M: 1.02 (0.56, 1.85) | -0.28 (-0.99, 0.42) |
| Say in government# | O: 0.87 (0.52, 1.46)<br>F: 0.82 (0.43, 1.56)<br>M: 0.95 (0.48, 1.85) | -0.14 (-0.95, 0.67) |
| Support reducing fferences in income levels§ | O: 0.84 (0.45, 1.54)<br>F: 1.21 (0.54, 2.70)<br>M: 0.62 (0.31, 1.27) | 0.66 (-0.23, 1.56) |
| Trust in others§ | O: 0.80 (0.57, 1.14)<br>F: 0.92 (0.51, 1.67)<br>M: 0.73 (0.47, 1.13) | 0.24 (-0.51, 0.98) |
| Feel at peace§ | O: 0.70 (0.49, 1.01)*<br>F: 0.77 (0.47, 1.27)<br>M: 0.67 (0.40, 1.10) | 0.14 (-0.55, 0.84) |

*Poor self-rated health while growing up – Small town or village*

**Adjusted associations between poor self-rated childhood health (vs excellent) and adult outcomes in India**
**Residence: Small town**

Wave 1, survey-weighted linear model. One model was fitted for each continuous outcome (24 models total, for 22 outcomes and 2 flourishing composite indices).
Estimates are the adjusted model coefficients (beta) of the predictor.

**Group** ● Overall (O) ▲ Female (F) ■ Male (M)

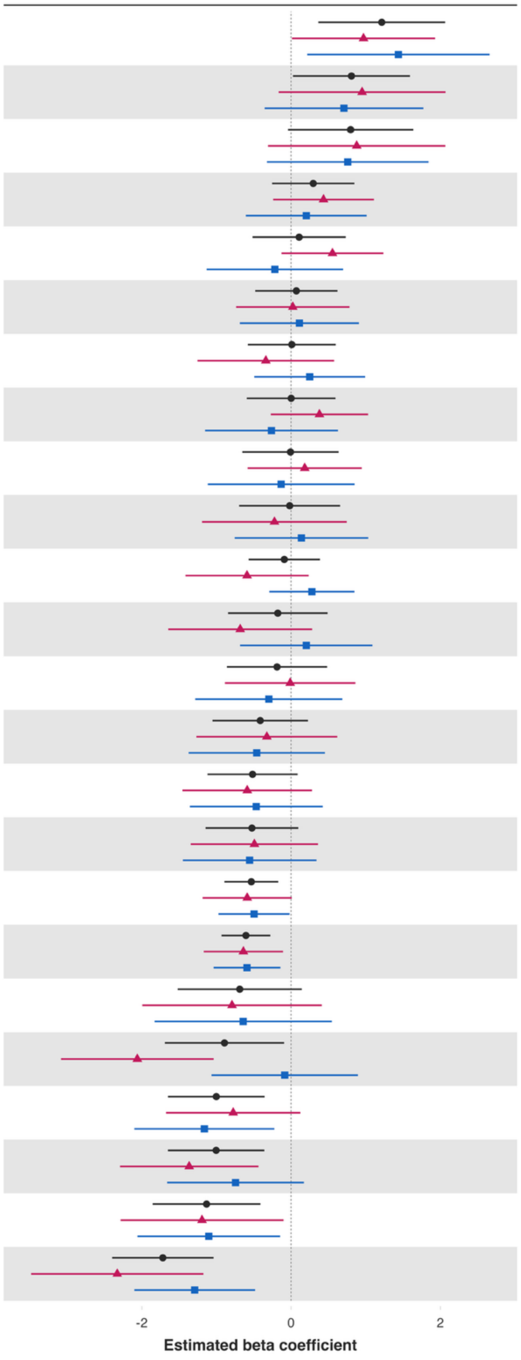


| Variable | Overall and interaction-derived gender estimates (95% CI) | Interaction (F–M, 95% CI) |
|---|---|---|
| Worry about safety, food, or housing§§‡‡ | O: 1.22 (0.37, 2.06)**<br>F: 0.97 (0.01, 1.93)**<br>M: 1.44 (0.22, 2.66)** | -0.47 (-1.93, 1.00) |
| Loneliness§ | O: 0.81 (0.02, 1.59)**<br>F: 0.95 (-0.17, 2.07)*<br>M: 0.71 (-0.35, 1.77) | 0.24 (-1.29, 1.77) |
| Worry about meeting normal monthly living expenses§§‡‡ | O: 0.80 (-0.04, 1.64)*<br>F: 0.88 (-0.31, 2.07)<br>M: 0.76 (-0.32, 1.84) | 0.12 (-1.41, 1.65) |
| Relationship contentment‡ | O: 0.30 (-0.25, 0.85)<br>F: 0.43 (-0.24, 1.11)<br>M: 0.20 (-0.61, 1.01) | 0.23 (-0.82, 1.28) |
| Show love often | O: 0.11 (-0.52, 0.73)<br>F: 0.55 (-0.13, 1.24)<br>M: -0.22 (-1.13, 0.70) | 0.77 (-0.34, 1.89) |
| Positive expectations | O: 0.07 (-0.48, 0.62)<br>F: 0.02 (-0.74, 0.78)<br>M: 0.11 (-0.69, 0.91) | -0.09 (-1.22, 1.04) |
| Freedom | O: 0.01 (-0.58, 0.60)<br>F: -0.34 (-1.25, 0.58)<br>M: 0.25 (-0.49, 0.99) | -0.59 (-1.78, 0.61) |
| Remain hopeful for the future | O: 0.00 (-0.59, 0.59)<br>F: 0.38 (-0.27, 1.03)<br>M: -0.26 (-1.15, 0.63) | 0.64 (-0.46, 1.74) |
| Relationship satisfaction‡ | O: -0.01 (-0.65, 0.64)<br>F: 0.18 (-0.58, 0.95)<br>M: -0.13 (-1.12, 0.85) | 0.32 (-0.96, 1.60) |
| Give up some happiness now for greater happiness later‡ | O: -0.02 (-0.70, 0.66)<br>F: -0.22 (-1.19, 0.75)<br>M: 0.14 (-0.76, 1.03) | -0.36 (-1.65, 0.92) |
| Act to promote good‡ | O: -0.09 (-0.57, 0.39)<br>F: -0.59 (-1.42, 0.24)<br>M: 0.28 (-0.29, 0.85) | -0.87 (-1.87, 0.13)* |
| Belonging | O: -0.18 (-0.85, 0.49)<br>F: -0.68 (-1.65, 0.28)<br>M: 0.20 (-0.68, 1.09) | -0.88 (-2.19, 0.42) |
| Have a long list of gratitude | O: -0.19 (-0.86, 0.48)<br>F: -0.01 (-0.89, 0.86)<br>M: -0.30 (-1.28, 0.69) | 0.29 (-1.07, 1.64) |
| Life evaluation five years from now | O: -0.41 (-1.05, 0.23)<br>F: -0.32 (-1.27, 0.62)<br>M: -0.46 (-1.37, 0.45) | 0.13 (-1.23, 1.50) |
| Life satisfaction‡ | O: -0.52 (-1.12, 0.09)*<br>F: -0.59 (-1.46, 0.28)<br>M: -0.47 (-1.36, 0.42) | -0.12 (-1.43, 1.18) |
| Understand purpose in life‡ | O: -0.52 (-1.15, 0.10)*<br>F: -0.49 (-1.34, 0.36)<br>M: -0.56 (-1.45, 0.34) | 0.06 (-1.20, 1.33) |
| Flourishing (without financial indicators) | O: -0.53 (-0.89, -0.17)**<br>F: -0.59 (-1.19, 0.01)*<br>M: -0.50 (-0.97, -0.02)** | -0.09 (-0.88, 0.70) |
| Flourishing (with financial indicators) | O: -0.61 (-0.93, -0.28)***<br>F: -0.64 (-1.17, -0.11)**<br>M: -0.59 (-1.04, -0.14)** | -0.05 (-0.77, 0.68) |
| Perceived support from others | O: -0.69 (-1.52, 0.14)<br>F: -0.79 (-1.99, 0.41)<br>M: -0.64 (-1.83, 0.55) | -0.15 (-1.90, 1.60) |
| Life evaluation today | O: -0.89 (-1.69, -0.09)**<br>F: -2.06 (-3.08, -1.04)***<br>M: -0.09 (-1.07, 0.90) | -1.98 (-3.30, -0.65)** |
| Life worthwhile‡ | O: -1.00 (-1.65, -0.36)**<br>F: -0.78 (-1.68, 0.12)*<br>M: -1.16 (-2.10, -0.22)** | 0.39 (-0.95, 1.72) |
| Happiness‡ | O: -1.00 (-1.65, -0.36)**<br>F: -1.37 (-2.29, -0.44)**<br>M: -0.74 (-1.66, 0.17) | -0.62 (-1.96, 0.72) |
| Mental health‡ | O: -1.13 (-1.85, -0.41)**<br>F: -1.19 (-2.29, -0.10)**<br>M: -1.10 (-2.06, -0.15)** | -0.09 (-1.53, 1.35) |
| Physical health‡ | O: -1.72 (-2.40, -1.04)***<br>F: -2.33 (-3.48, -1.18)***<br>M: -1.29 (-2.10, -0.48)** | -1.04 (-2.45, 0.38) |

### Adjusted associations between poor self-rated childhood health (vs excellent) and adult outcomes in India
### Residence: Small town

Wave 1, survey-weighted logistic model. One model was fitted for each binary outcome (7 models total). Estimates are adjusted odds ratios (OR) of the predictor.
The interaction column reports the difference in log-odds ratios (interaction beta) with 95% confidence intervals (95% CIs).

**Group** ● Overall (O) ▲ Female (F) ■ Male (M)

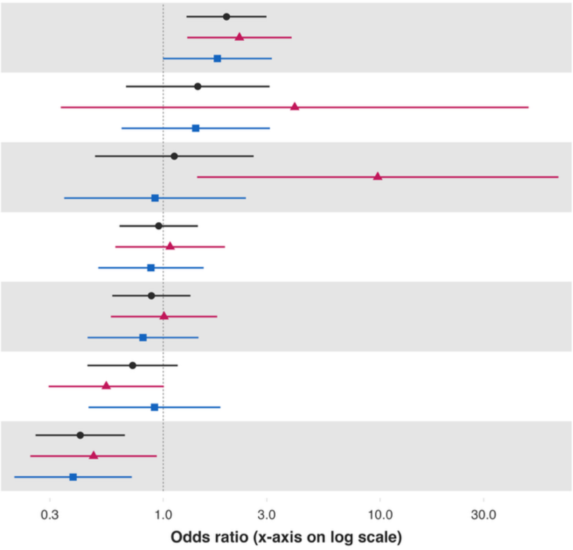


| Variable | Overall and interaction-derived gender estimates (95% CI) | Interaction (F–M, 95% CI) |
|---|---|---|
| Have any health problems | O: 1.96 (1.28, 2.99)**<br>F: 2.24 (1.29, 3.90)**<br>M: 1.78 (1.00, 3.17)* | 0.23 (-0.52, 0.99) |
| Alcohol use | O: 1.44 (0.67, 3.09)<br>F: 4.04 (0.34, 48.29)<br>M: 1.41 (0.64, 3.10) | 1.05 (-1.58, 3.68) |
| Cigarette smoking | O: 1.12 (0.48, 2.61)<br>F: 9.74 (1.43, 66.27)**<br>M: 0.92 (0.35, 2.40) | 2.36 (0.14, 4.58)** |
| Volunteered at an organization | O: 0.95 (0.63, 1.44)<br>F: 1.08 (0.60, 1.92)<br>M: 0.88 (0.50, 1.53) | 0.20 (-0.59, 1.00) |
| Donated to charity | O: 0.88 (0.58, 1.34)<br>F: 1.01 (0.57, 1.77)<br>M: 0.81 (0.45, 1.45) | 0.22 (-0.58, 1.03) |
| Helped a stranger | O: 0.72 (0.45, 1.17)<br>F: 0.55 (0.30, 1.01)*<br>M: 0.91 (0.45, 1.83) | -0.51 (-1.43, 0.40) |
| Feel close to someone | O: 0.41 (0.26, 0.67)***<br>F: 0.48 (0.24, 0.93)**<br>M: 0.38 (0.21, 0.72)** | 0.22 (-0.69, 1.12) |

### Adjusted associations between poor self-rated childhood health (vs excellent) and adult outcomes in India
### Residence: Small town

Wave 1, survey-weighted ordinal regression using the proportional odds model (`svyolr`). One model was fitted for each ordinal outcome (16 models total).
Estimates are adjusted cumulative odds ratios (cOR) of the predictor.
The interaction column reports the difference in log-cumulative odds ratios (interaction beta) with 95% confidence intervals (95% CIs).

**Group** ● Overall (O) ▲ Female (F) ■ Male (M)

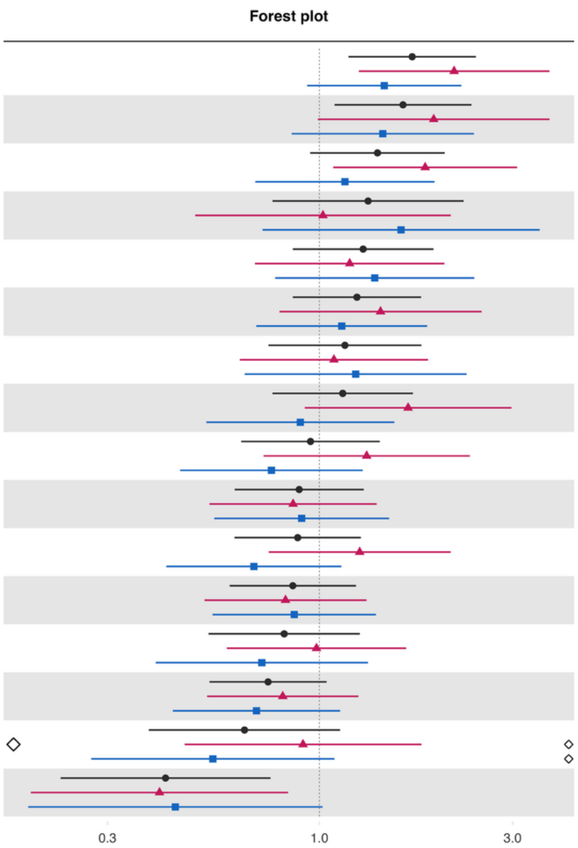


| Variable | Overall and interaction-derived gender estimates (95% CI) | Interaction (F–M, 95% CI) |
|---|---|---|
| Feeling nervous or anxious§ | O: 1.69 (1.18, 2.43)**<br>F: 2.15 (1.25, 3.70)**<br>M: 1.45 (0.93, 2.24)* | 0.40 (-0.25, 1.04) |
| Unable to control worry§ | O: 1.61 (1.09, 2.37)**<br>F: 1.92 (0.99, 3.70)*<br>M: 1.43 (0.85, 2.41) | 0.29 (-0.58, 1.15) |
| Extent of suffering§ | O: 1.39 (0.95, 2.04)*<br>F: 1.83 (1.08, 3.08)**<br>M: 1.16 (0.69, 1.93) | 0.46 (-0.24, 1.15) |
| Say in government# | O: 1.32 (0.77, 2.27)<br>F: 1.02 (0.49, 2.11)<br>M: 1.59 (0.72, 3.50) | -0.44 (-1.51, 0.62) |
| Perceived discrimination§ | O: 1.28 (0.86, 1.91)<br>F: 1.19 (0.69, 2.04)<br>M: 1.37 (0.78, 2.41) | -0.14 (-0.92, 0.64) |
| Distress from e-threatening experiences§ | O: 1.24 (0.86, 1.78)<br>F: 1.42 (0.80, 2.52)<br>M: 1.14 (0.70, 1.85) | 0.22 (-0.54, 0.99) |
| Depressive symptoms§ | O: 1.16 (0.75, 1.79)<br>F: 1.09 (0.64, 1.86)<br>M: 1.23 (0.65, 2.31) | -0.12 (-0.92, 0.68) |
| Forgiveness§ | O: 1.14 (0.77, 1.70)<br>F: 1.66 (0.92, 2.98)*<br>M: 0.90 (0.53, 1.53) | 0.61 (-0.19, 1.42) |
| Government approval§ | O: 0.95 (0.64, 1.41)<br>F: 1.31 (0.73, 2.35)<br>M: 0.76 (0.45, 1.28) | 0.54 (-0.24, 1.33) |
| Feeling capable§ | O: 0.89 (0.62, 1.29)<br>F: 0.86 (0.54, 1.39)<br>M: 0.90 (0.55, 1.49) | -0.05 (-0.69, 0.60) |
| Feel at peace§ | O: 0.88 (0.62, 1.27)<br>F: 1.26 (0.75, 2.11)<br>M: 0.69 (0.42, 1.13) | 0.60 (-0.12, 1.32) |
| Trust in others§ | O: 0.86 (0.60, 1.23)<br>F: 0.82 (0.52, 1.31)<br>M: 0.87 (0.54, 1.38) | -0.05 (-0.66, 0.56) |
| Various aspects of life are balanced§ | O: 0.82 (0.53, 1.26)<br>F: 0.98 (0.59, 1.64)<br>M: 0.72 (0.39, 1.32) | 0.31 (-0.43, 1.05) |
| Have little interest in doing things§ | O: 0.75 (0.54, 1.04)*<br>F: 0.81 (0.53, 1.25)<br>M: 0.70 (0.43, 1.13) | 0.15 (-0.48, 0.78) |
| Support reducing fferences in income levels§ | O: 0.65 (0.38, 1.13)<br>F: 0.91 (0.46, 1.79)<br>M: 0.55 (0.27, 1.09)* | 0.51 (-0.39, 1.42) |
| Obeying the law§ | O: 0.42 (0.23, 0.76)**<br>F: 0.40 (0.19, 0.84)**<br>M: 0.44 (0.19, 1.02)* | -0.09 (-1.16, 0.98) |

*Poor self-rated health while growing up – Large city or suburb of a large city*

**Adjusted associations between poor self-rated childhood health (vs excellent) and adult outcomes in India**
**Residence: Large city or suburb**

Wave 1, survey-weighted linear model. One model was fitted for each continuous outcome (24 models total, for 22 outcomes and 2 flourishing composite indices). Estimates are the adjusted model coefficients (beta) of the predictor.

**Group** ● Overall (O) ▲ Female (F) ■ Male (M)

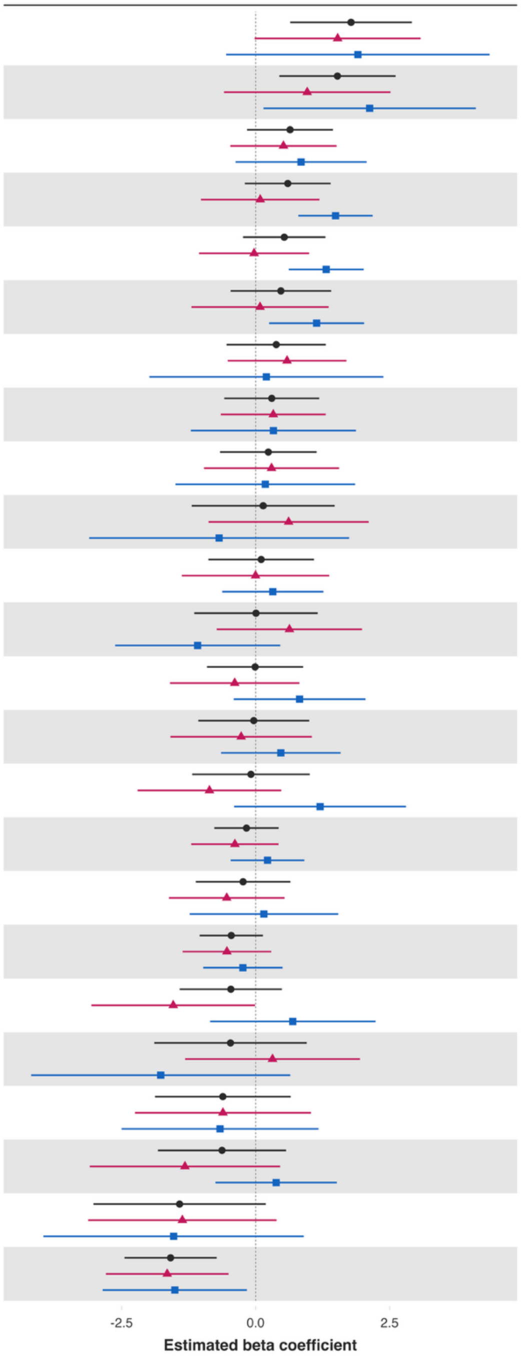


| Variable | Overall and interaction-derived gender estimates (95% CI) | Interaction (F–M, 95% CI) |
|---|---|---|
| Worry about meeting normal monthly living expenses§§‡‡ | O: 1.78 (0.64, 2.92)**<br>F: 1.53 (-0.02, 3.08)*<br>M: 1.91 (-0.55, 4.37) | -0.38 (-3.60, 2.85) |
| Worry about safety, food, or housing§§‡‡ | O: 1.53 (0.44, 2.61)**<br>F: 0.96 (-0.59, 2.51)<br>M: 2.13 (0.15, 4.11)** | -1.17 (-3.90, 1.57) |
| Relationship satisfaction‡ | O: 0.64 (-0.16, 1.44)<br>F: 0.52 (-0.47, 1.51)<br>M: 0.85 (-0.38, 2.07) | -0.33 (-1.83, 1.17) |
| Understand purpose in life‡ | O: 0.60 (-0.20, 1.40)<br>F: 0.09 (-1.02, 1.19)<br>M: 1.49 (0.80, 2.18)*** | -1.40 (-2.65, -0.16)** |
| Act to promote good‡ | O: 0.53 (-0.24, 1.30)<br>F: -0.03 (-1.06, 1.00)<br>M: 1.32 (0.62, 2.02)*** | -1.35 (-2.51, -0.18)** |
| Show love often | O: 0.47 (-0.47, 1.41)<br>F: 0.08 (-1.20, 1.36)<br>M: 1.14 (0.25, 2.02)** | -1.06 (-2.55, 0.44) |
| Relationship contentment‡ | O: 0.38 (-0.54, 1.31)<br>F: 0.59 (-0.52, 1.69)<br>M: 0.20 (-1.98, 2.38) | 0.39 (-2.31, 3.08) |
| Positive expectations | O: 0.30 (-0.59, 1.19)<br>F: 0.33 (-0.65, 1.31)<br>M: 0.33 (-1.21, 1.87) | -0.00 (-1.74, 1.74) |
| Belonging | O: 0.24 (-0.66, 1.14)<br>F: 0.30 (-0.96, 1.56)<br>M: 0.18 (-1.50, 1.86) | 0.12 (-2.18, 2.41) |
| Perceived support from others | O: 0.14 (-1.19, 1.47)<br>F: 0.62 (-0.88, 2.11)<br>M: -0.68 (-3.11, 1.74) | 1.30 (-1.54, 4.13) |
| Freedom | O: 0.10 (-0.88, 1.09)<br>F: -0.00 (-1.38, 1.37)<br>M: 0.32 (-0.62, 1.26) | -0.32 (-1.91, 1.27) |
| Loneliness§ | O: 0.01 (-1.14, 1.16)<br>F: 0.63 (-0.73, 1.99)<br>M: -1.08 (-2.62, 0.46) | 1.71 (-0.09, 3.51)* |
| Mental health‡ | O: -0.01 (-0.91, 0.89)<br>F: -0.39 (-1.60, 0.82)<br>M: 0.82 (-0.41, 2.05) | -1.21 (-2.96, 0.54) |
| Remain hopeful for the future | O: -0.04 (-1.07, 1.00)<br>F: -0.27 (-1.59, 1.05)<br>M: 0.47 (-0.65, 1.59) | -0.74 (-2.30, 0.82) |
| Give up some happiness now for greater happiness later‡ | O: -0.09 (-1.18, 1.01)<br>F: -0.86 (-2.20, 0.48)<br>M: 1.20 (-0.40, 2.81) | -2.07 (-4.14, 0.01)* |
| Flourishing (without financial indicators) | O: -0.17 (-0.77, 0.43)<br>F: -0.39 (-1.20, 0.43)<br>M: 0.22 (-0.47, 0.91) | -0.61 (-1.64, 0.43) |
| Have a long list of gratitude | O: -0.23 (-1.12, 0.65)<br>F: -0.54 (-1.62, 0.54)<br>M: 0.15 (-1.23, 1.54) | -0.69 (-2.41, 1.02) |
| Flourishing (with financial indicators) | O: -0.46 (-1.05, 0.14)<br>F: -0.54 (-1.36, 0.29)<br>M: -0.24 (-0.98, 0.50) | -0.30 (-1.43, 0.83) |
| Life evaluation five years from now | O: -0.46 (-1.42, 0.49)<br>F: -1.54 (-3.06, -0.01)*<br>M: 0.69 (-0.85, 2.24) | -2.23 (-4.70, 0.24)* |
| Life worthwhile‡ | O: -0.47 (-1.89, 0.95)<br>F: 0.31 (-1.32, 1.95)<br>M: -1.77 (-4.19, 0.65) | 2.09 (-0.68, 4.86) |
| Life satisfaction‡ | O: -0.61 (-1.88, 0.65)<br>F: -0.61 (-2.25, 1.03)<br>M: -0.66 (-2.50, 1.17) | 0.05 (-2.32, 2.43) |
| Happiness‡ | O: -0.63 (-1.82, 0.57)<br>F: -1.32 (-3.10, 0.46)<br>M: 0.38 (-0.75, 1.52) | -1.70 (-3.85, 0.45) |
| Life evaluation today | O: -1.42 (-3.03, 0.19)*<br>F: -1.37 (-3.13, 0.39)<br>M: -1.53 (-3.96, 0.90) | 0.16 (-2.43, 2.76) |
| Physical health‡ | O: -1.59 (-2.45, -0.73)***<br>F: -1.65 (-2.80, -0.51)**<br>M: -1.51 (-2.86, -0.16)** | -0.14 (-1.93, 1.64) |

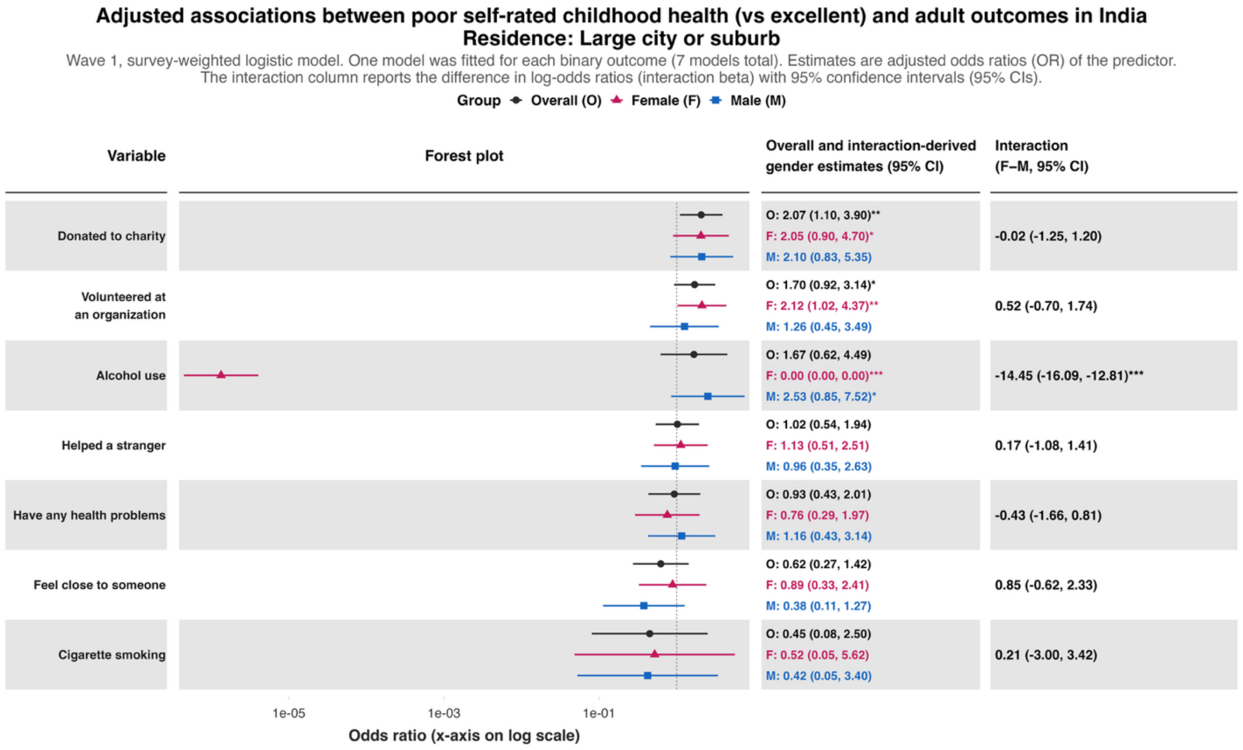
Adjusted associations between poor self-rated childhood health (vs excellent) and adult outcomes in India
Residence: Large city or suburb
Wave 1, survey-weighted logistic model. One model was fitted for each binary outcome (7 models total). Estimates are adjusted odds ratios (OR) of the predictor.
The interaction column reports the difference in log-odds ratios (interaction beta) with 95% confidence intervals (95% CIs).
Group Overall (O) Female (F) Male (M)
Variable
Forest plot
Overall and interaction-derived gender estimates (95% CI)
Interaction (F–M, 95% CI)
Donated to charity
O: 2.07 (1.10, 3.90)**
F: 2.05 (0.90, 4.70)*
M: 2.10 (0.83, 5.35)
-0.02 (-1.25, 1.20)
Volunteered at an organization
O: 1.70 (0.92, 3.14)*
F: 2.12 (1.02, 4.37)**
M: 1.26 (0.45, 3.49)
0.52 (-0.70, 1.74)
Alcohol use
O: 1.67 (0.62, 4.49)
F: 0.00 (0.00, 0.00)***
M: 2.53 (0.85, 7.52)*
-14.45 (-16.09, -12.81)***
Helped a stranger
O: 1.02 (0.54, 1.94)
F: 1.13 (0.51, 2.51)
M: 0.96 (0.35, 2.63)
0.17 (-1.08, 1.41)
Have any health problems
O: 0.93 (0.43, 2.01)
F: 0.76 (0.29, 1.97)
M: 1.16 (0.43, 3.14)
-0.43 (-1.66, 0.81)
Feel close to someone
O: 0.62 (0.27, 1.42)
F: 0.89 (0.33, 2.41)
M: 0.38 (0.11, 1.27)
0.85 (-0.62, 2.33)
Cigarette smoking
O: 0.45 (0.08, 2.50)
F: 0.52 (0.05, 5.62)
M: 0.42 (0.05, 3.40)
0.21 (-3.00, 3.42)
1e-05
1e-03
1e-01
Odds ratio (x-axis on log scale)

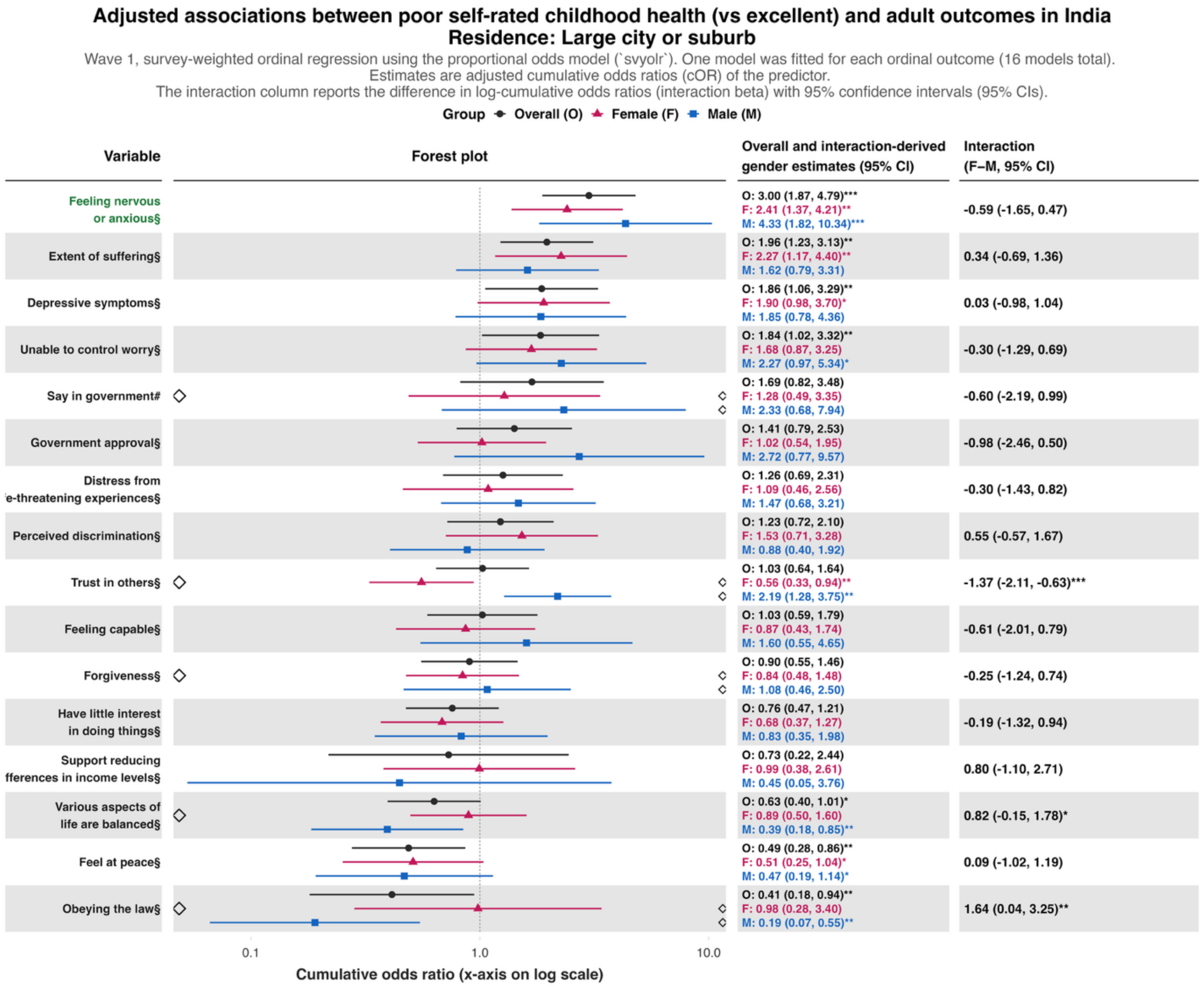
Adjusted associations between poor self-rated childhood health (vs excellent) and adult outcomes in India
Residence: Large city or suburb
Wave 1, survey-weighted ordinal regression using the proportional odds model (`svyolr`). One model was fitted for each ordinal outcome (16 models total).
Estimates are adjusted cumulative odds ratios (cOR) of the predictor.
The interaction column reports the difference in log-cumulative odds ratios (interaction beta) with 95% confidence intervals (95% CIs).
Group Overall (O) Female (F) Male (M)
Variable
Forest plot
Overall and interaction-derived gender estimates (95% CI)
Interaction (F–M, 95% CI)
Feeling nervous or anxious§
O: 3.00 (1.87, 4.79)***
F: 2.41 (1.37, 4.21)**
M: 4.33 (1.82, 10.34)***
-0.59 (-1.65, 0.47)
Extent of suffering§
O: 1.96 (1.23, 3.13)**
F: 2.27 (1.17, 4.40)**
M: 1.62 (0.79, 3.31)
0.34 (-0.69, 1.36)
Depressive symptoms§
O: 1.86 (1.06, 3.29)**
F: 1.90 (0.98, 3.70)*
M: 1.85 (0.78, 4.36)
0.03 (-0.98, 1.04)
Unable to control worry§
O: 1.84 (1.02, 3.32)**
F: 1.68 (0.87, 3.25)
M: 2.27 (0.97, 5.34)*
-0.30 (-1.29, 0.69)
Say in government#
O: 1.69 (0.82, 3.48)
F: 1.28 (0.49, 3.35)
M: 2.33 (0.68, 7.94)
-0.60 (-2.19, 0.99)
Government approval§
O: 1.41 (0.79, 2.53)
F: 1.02 (0.54, 1.95)
M: 2.72 (0.77, 9.57)
-0.98 (-2.46, 0.50)
Distress from
'e-threatening experiences§
O: 1.26 (0.69, 2.31)
F: 1.09 (0.46, 2.56)
M: 1.47 (0.68, 3.21)
-0.30 (-1.43, 0.82)
Perceived discrimination§
O: 1.23 (0.72, 2.10)
F: 1.53 (0.71, 3.28)
M: 0.88 (0.40, 1.92)
0.55 (-0.57, 1.67)
Trust in others§
O: 1.03 (0.64, 1.64)
F: 0.56 (0.33, 0.94)**
M: 2.19 (1.28, 3.75)**
-1.37 (-2.11, -0.63)***
Feeling capable§
O: 1.03 (0.59, 1.79)
F: 0.87 (0.43, 1.74)
M: 1.60 (0.55, 4.65)
-0.61 (-2.01, 0.79)
Forgiveness§
O: 0.90 (0.55, 1.46)
F: 0.84 (0.48, 1.48)
M: 1.08 (0.46, 2.50)
-0.25 (-1.24, 0.74)
Have little interest in doing things§
O: 0.76 (0.47, 1.21)
F: 0.68 (0.37, 1.27)
M: 0.83 (0.35, 1.98)
-0.19 (-1.32, 0.94)
Support reducing
fferences in income levels§
O: 0.73 (0.22, 2.44)
F: 0.99 (0.38, 2.61)
M: 0.45 (0.05, 3.76)
0.80 (-1.10, 2.71)
Various aspects of life are balanced§
O: 0.63 (0.40, 1.01)*
F: 0.89 (0.50, 1.60)
M: 0.39 (0.18, 0.85)**
0.82 (-0.15, 1.78)*
Feel at peace§
O: 0.49 (0.28, 0.86)**
F: 0.51 (0.25, 1.04)*
M: 0.47 (0.19, 1.14)*
0.09 (-1.02, 1.19)
Obeying the law§
O: 0.41 (0.18, 0.94)**
F: 0.98 (0.28, 3.40)
M: 0.19 (0.07, 0.55)**
1.64 (0.04, 3.25)**
0.1
1.0
10.0
Cumulative odds ratio (x-axis on log scale)

**Predictor:** ***Perceived family financial situation as very difficult while growing up***

***Perceived family financial situation as very difficult while growing up – Rural area or on a farm***

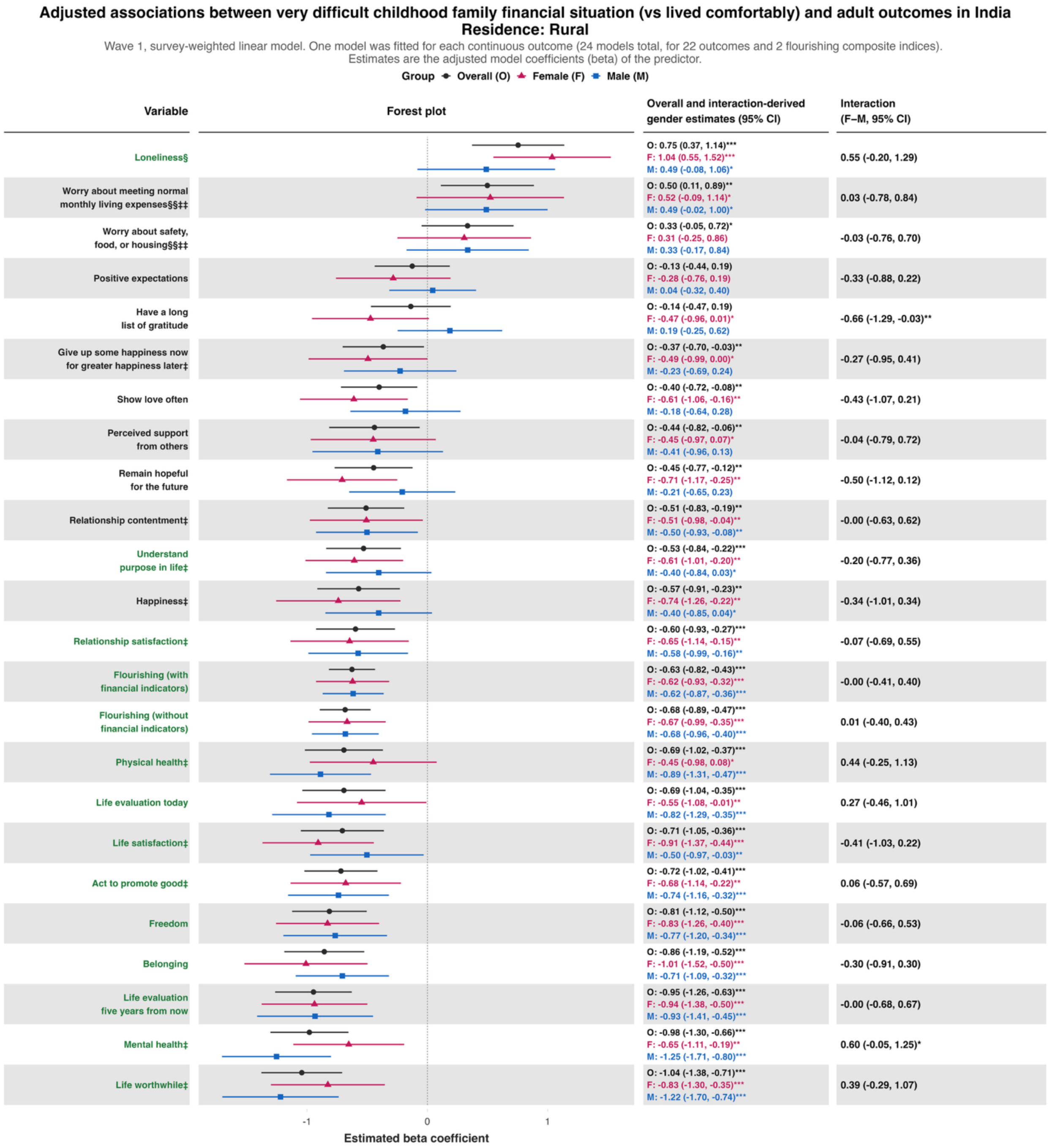

**Adjusted associations between very difficult childhood family financial situation (vs lived comfortably) and adult outcomes in India**
**Residence: Rural**

Wave 1, survey-weighted logistic model. One model was fitted for each binary outcome (7 models total). Estimates are adjusted odds ratios (OR) of the predictor.
The interaction column reports the difference in log-odds ratios (interaction beta) with 95% confidence intervals (95% CIs).

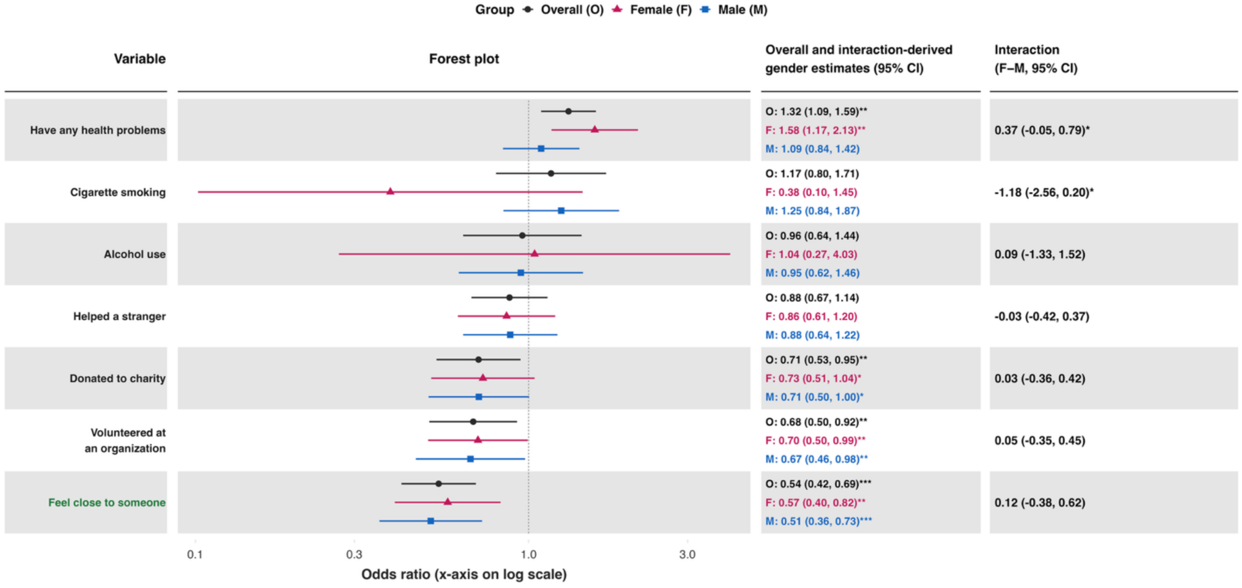

### Adjusted associations between very difficult childhood family financial situation (vs lived comfortably) and adult outcomes in India
### Residence: Rural

Wave 1, survey-weighted ordinal regression using the proportional odds model (`svyolr`). One model was fitted for each ordinal outcome (16 models total).
Estimates are adjusted cumulative odds ratios (cOR) of the predictor.
The interaction column reports the difference in log-cumulative odds ratios (interaction beta) with 95% confidence intervals (95% CIs).

**Group** ● Overall (O) ▲ Female (F) ■ Male (M)

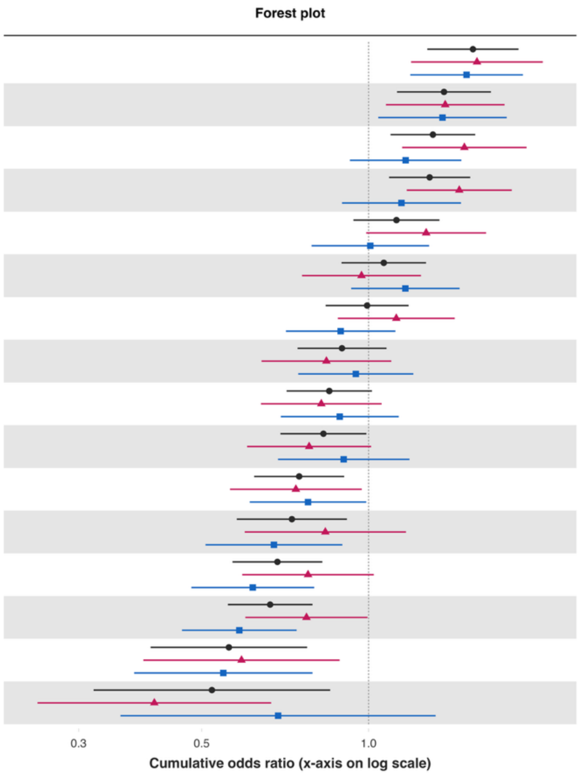


| Variable | Overall and interaction-derived gender estimates (95% CI) | Interaction (F–M, 95% CI) |
|---|---|---|
| Extent of suffering§ | O: 1.54 (1.28, 1.86)***<br>F: 1.57 (1.19, 2.06)**<br>M: 1.50 (1.19, 1.90)*** | 0.04 (-0.29, 0.38) |
| Depressive symptoms§ | O: 1.37 (1.13, 1.66)**<br>F: 1.37 (1.07, 1.76)**<br>M: 1.36 (1.04, 1.77)** | 0.01 (-0.32, 0.35) |
| Distress from life-threatening experiences§ | O: 1.31 (1.10, 1.56)**<br>F: 1.49 (1.15, 1.93)**<br>M: 1.17 (0.93, 1.47) | 0.24 (-0.09, 0.58) |
| Feeling nervous or anxious§ | O: 1.29 (1.09, 1.53)**<br>F: 1.46 (1.17, 1.81)***<br>M: 1.15 (0.90, 1.47) | 0.24 (-0.08, 0.56) |
| Perceived discrimination§ | O: 1.12 (0.94, 1.34)<br>F: 1.27 (0.99, 1.63)*<br>M: 1.01 (0.79, 1.29) | 0.23 (-0.11, 0.57) |
| Unable to control worry§ | O: 1.07 (0.89, 1.27)<br>F: 0.97 (0.76, 1.24)<br>M: 1.17 (0.93, 1.46) | -0.18 (-0.50, 0.13) |
| Have little interest in doing things§ | O: 0.99 (0.84, 1.18)<br>F: 1.12 (0.88, 1.43)<br>M: 0.89 (0.71, 1.12) | 0.23 (-0.09, 0.55) |
| Feel at peace§ | O: 0.90 (0.74, 1.08)<br>F: 0.84 (0.64, 1.10)<br>M: 0.95 (0.75, 1.20) | -0.12 (-0.47, 0.22) |
| Various aspects of life are balanced§ | O: 0.85 (0.71, 1.01)*<br>F: 0.82 (0.64, 1.06)<br>M: 0.89 (0.69, 1.13) | -0.08 (-0.42, 0.27) |
| Feeling capable§ | O: 0.83 (0.69, 0.99)**<br>F: 0.78 (0.60, 1.01)*<br>M: 0.90 (0.69, 1.19) | -0.14 (-0.54, 0.25) |
| Forgiveness§ | O: 0.75 (0.62, 0.90)**<br>F: 0.74 (0.56, 0.97)**<br>M: 0.78 (0.61, 0.99)** | -0.05 (-0.40, 0.30) |
| Say in government# | O: 0.73 (0.58, 0.91)**<br>F: 0.84 (0.60, 1.17)<br>M: 0.67 (0.51, 0.90)** | 0.21 (-0.19, 0.62) |
| Government approval§ | O: 0.68 (0.57, 0.83)***<br>F: 0.78 (0.59, 1.02)*<br>M: 0.62 (0.48, 0.80)*** | 0.23 (-0.14, 0.60) |
| Trust in others§ | O: 0.66 (0.56, 0.79)***<br>F: 0.77 (0.60, 1.00)**<br>M: 0.58 (0.46, 0.74)*** | 0.28 (-0.07, 0.62) |
| Support reducing differences in income levels§ | O: 0.56 (0.40, 0.77)***<br>F: 0.59 (0.39, 0.89)**<br>M: 0.55 (0.38, 0.79)** | 0.08 (-0.35, 0.50) |
| Obeying the law§ | O: 0.52 (0.32, 0.85)**<br>F: 0.41 (0.25, 0.67)***<br>M: 0.69 (0.36, 1.32) | -0.51 (-1.10, 0.07)* |

### *Perceived family financial situation as very difficult while growing up – Small town or village*

#### Adjusted associations between very difficult childhood family financial situation (vs lived comfortably) and adult outcomes in India
#### Residence: Small town

Wave 1, survey-weighted linear model. One model was fitted for each continuous outcome (24 models total, for 22 outcomes and 2 flourishing composite indices).
Estimates are the adjusted model coefficients (beta) of the predictor.

Group — Overall (O) — Female (F) — Male (M)

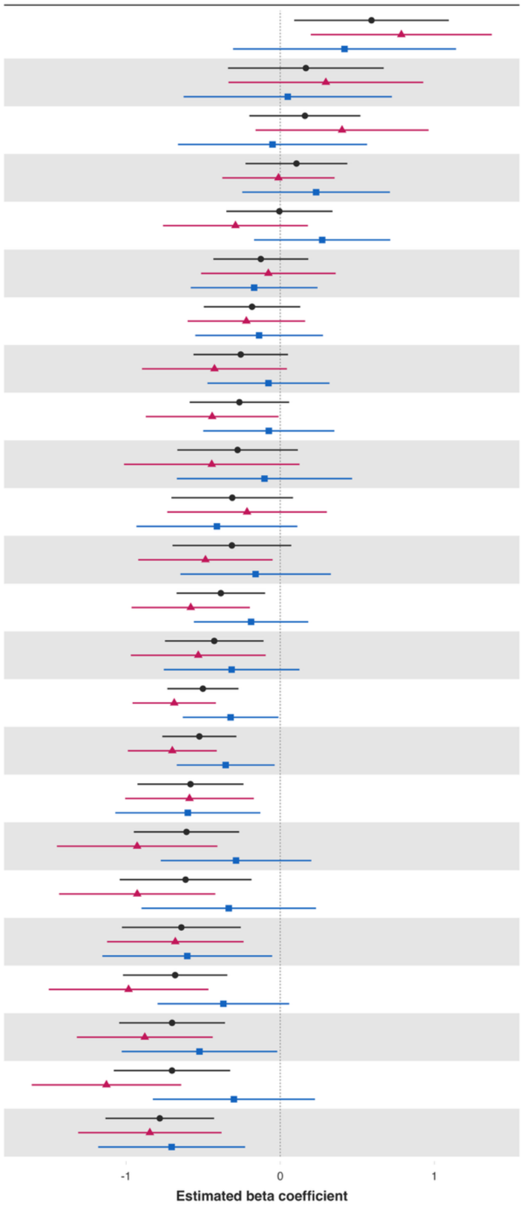


| Variable | Overall and interaction-derived gender estimates (95% CI) | Interaction (F–M, 95% CI) |
|---|---|---|
| Worry about meeting normal monthly living expenses§§‡‡ | O: 0.59 (0.09, 1.09)**<br>F: 0.79 (0.20, 1.37)**<br>M: 0.42 (-0.31, 1.14) | 0.37 (-0.50, 1.23) |
| Worry about safety, food, or housing§§‡‡ | O: 0.17 (-0.34, 0.67)<br>F: 0.30 (-0.34, 0.93)<br>M: 0.05 (-0.63, 0.72) | 0.25 (-0.59, 1.08) |
| Loneliness§ | O: 0.16 (-0.20, 0.52)<br>F: 0.40 (-0.16, 0.96)<br>M: -0.05 (-0.66, 0.56) | 0.45 (-0.48, 1.38) |
| Positive expectations | O: 0.11 (-0.23, 0.44)<br>F: -0.01 (-0.38, 0.35)<br>M: 0.23 (-0.25, 0.71) | -0.24 (-0.78, 0.30) |
| Show love often | O: -0.01 (-0.35, 0.34)<br>F: -0.29 (-0.76, 0.18)<br>M: 0.27 (-0.17, 0.71) | -0.56 (-1.17, 0.04)* |
| Have a long list of gratitude | O: -0.13 (-0.43, 0.18)<br>F: -0.08 (-0.51, 0.36)<br>M: -0.17 (-0.58, 0.24) | 0.09 (-0.49, 0.67) |
| Remain hopeful for the future | O: -0.18 (-0.50, 0.13)<br>F: -0.22 (-0.60, 0.16)<br>M: -0.14 (-0.55, 0.28) | -0.08 (-0.58, 0.41) |
| Belonging | O: -0.26 (-0.56, 0.05)<br>F: -0.43 (-0.90, 0.04)*<br>M: -0.08 (-0.47, 0.32) | -0.35 (-0.96, 0.26) |
| Freedom | O: -0.26 (-0.59, 0.06)<br>F: -0.44 (-0.87, -0.01)**<br>M: -0.07 (-0.50, 0.35) | -0.37 (-0.93, 0.20) |
| Perceived support from others | O: -0.28 (-0.67, 0.11)<br>F: -0.44 (-1.01, 0.13)<br>M: -0.10 (-0.67, 0.47) | -0.34 (-1.17, 0.49) |
| Give up some happiness now for greater happiness later‡ | O: -0.31 (-0.71, 0.08)<br>F: -0.22 (-0.73, 0.30)<br>M: -0.41 (-0.93, 0.11) | 0.20 (-0.48, 0.87) |
| Relationship satisfaction‡ | O: -0.31 (-0.70, 0.07)<br>F: -0.49 (-0.92, -0.05)**<br>M: -0.16 (-0.65, 0.33) | -0.33 (-0.85, 0.20) |
| Act to promote good‡ | O: -0.39 (-0.67, -0.10)**<br>F: -0.58 (-0.96, -0.20)**<br>M: -0.19 (-0.56, 0.18) | -0.39 (-0.88, 0.10) |
| Understand purpose in life‡ | O: -0.43 (-0.75, -0.11)**<br>F: -0.53 (-0.97, -0.09)**<br>M: -0.32 (-0.76, 0.12) | -0.22 (-0.82, 0.38) |
| Flourishing (with financial indicators) | O: -0.50 (-0.73, -0.27)***<br>F: -0.69 (-0.96, -0.42)***<br>M: -0.32 (-0.63, -0.01)** | -0.37 (-0.73, -0.00)* |
| Flourishing (without financial indicators) | O: -0.52 (-0.76, -0.28)***<br>F: -0.70 (-0.99, -0.41)***<br>M: -0.35 (-0.67, -0.04)** | -0.35 (-0.72, 0.03)* |
| Relationship contentment‡ | O: -0.58 (-0.93, -0.24)***<br>F: -0.59 (-1.01, -0.17)**<br>M: -0.60 (-1.07, -0.13)** | 0.01 (-0.56, 0.58) |
| Physical health‡ | O: -0.61 (-0.95, -0.27)***<br>F: -0.93 (-1.45, -0.41)***<br>M: -0.29 (-0.77, 0.20) | -0.64 (-1.38, 0.10)* |
| Life evaluation today | O: -0.61 (-1.04, -0.19)**<br>F: -0.93 (-1.43, -0.42)***<br>M: -0.33 (-0.90, 0.23) | -0.59 (-1.24, 0.06)* |
| Life worthwhile‡ | O: -0.64 (-1.03, -0.26)**<br>F: -0.68 (-1.12, -0.24)**<br>M: -0.60 (-1.15, -0.05)** | -0.08 (-0.72, 0.57) |
| Mental health‡ | O: -0.68 (-1.02, -0.34)***<br>F: -0.98 (-1.50, -0.47)***<br>M: -0.37 (-0.80, 0.06)* | -0.61 (-1.28, 0.05)* |
| Life satisfaction‡ | O: -0.70 (-1.04, -0.36)***<br>F: -0.88 (-1.32, -0.44)***<br>M: -0.52 (-1.03, -0.02)** | -0.35 (-1.01, 0.30) |
| Happiness‡ | O: -0.70 (-1.08, -0.33)***<br>F: -1.13 (-1.61, -0.64)***<br>M: -0.30 (-0.83, 0.22) | -0.83 (-1.51, -0.14)** |
| Life evaluation five years from now | O: -0.78 (-1.13, -0.43)***<br>F: -0.85 (-1.31, -0.38)***<br>M: -0.70 (-1.18, -0.23)** | -0.14 (-0.76, 0.48) |

### Adjusted associations between very difficult childhood family financial situation (vs lived comfortably) and adult outcomes in India
### Residence: Small town

Wave 1, survey-weighted logistic model. One model was fitted for each binary outcome (7 models total). Estimates are adjusted odds ratios (OR) of the predictor.
The interaction column reports the difference in log-odds ratios (interaction beta) with 95% confidence intervals (95% CIs).

**Group** — Overall (O) — Female (F) — Male (M)

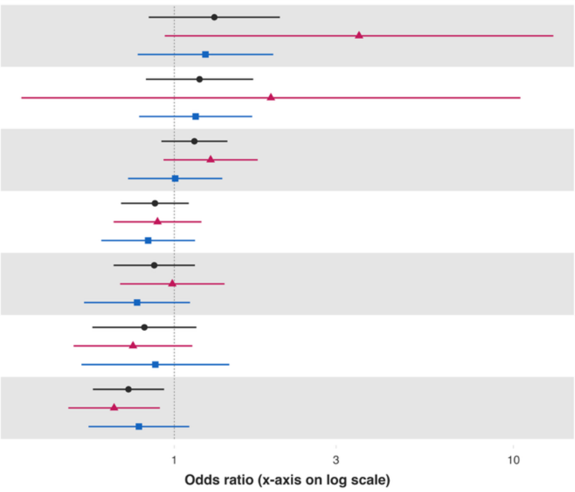


| Variable | Overall and interaction-derived gender estimates (95% CI) | Interaction (F–M, 95% CI) |
|---|---|---|
| Cigarette smoking | O: 1.31 (0.84, 2.05)<br>F: 3.51 (0.94, 13.12)*<br>M: 1.24 (0.78, 1.96) | 1.04 (-0.34, 2.42) |
| Alcohol use | O: 1.19 (0.82, 1.71)<br>F: 1.93 (0.35, 10.48)<br>M: 1.16 (0.79, 1.70) | 0.51 (-1.27, 2.29) |
| Have any health problems | O: 1.15 (0.92, 1.43)<br>F: 1.28 (0.93, 1.76)<br>M: 1.01 (0.73, 1.39) | 0.24 (-0.21, 0.70) |
| Helped a stranger | O: 0.88 (0.70, 1.10)<br>F: 0.89 (0.66, 1.20)<br>M: 0.84 (0.61, 1.15) | 0.06 (-0.34, 0.47) |
| Volunteered at an organization | O: 0.87 (0.66, 1.15)<br>F: 0.99 (0.69, 1.41)<br>M: 0.78 (0.54, 1.11) | 0.24 (-0.22, 0.70) |
| Feel close to someone | O: 0.82 (0.57, 1.16)<br>F: 0.76 (0.50, 1.13)<br>M: 0.88 (0.53, 1.45) | -0.15 (-0.72, 0.42) |
| Donated to charity | O: 0.73 (0.58, 0.93)**<br>F: 0.66 (0.49, 0.91)**<br>M: 0.79 (0.56, 1.11) | -0.17 (-0.61, 0.27) |

### Adjusted associations between very difficult childhood family financial situation (vs lived comfortably) and adult outcomes in India
### Residence: Small town

Wave 1, survey-weighted ordinal regression using the proportional odds model (`svyolr`). One model was fitted for each ordinal outcome (16 models total).
Estimates are adjusted cumulative odds ratios (cOR) of the predictor.
The interaction column reports the difference in log-cumulative odds ratios (interaction beta) with 95% confidence intervals (95% CIs).

**Group** ● Overall (O) ▲ Female (F) ■ Male (M)

Forest plot

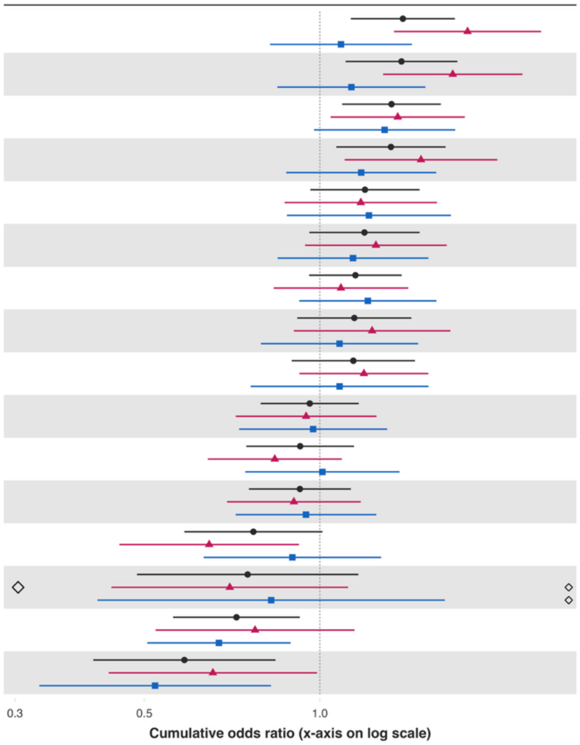


| Variable | Overall and interaction-derived gender estimates (95% CI) | Interaction (F–M, 95% CI) |
|---|---|---|
| Distress from life-threatening experiences§ | O: 1.39 (1.13, 1.70)**<br>F: 1.79 (1.34, 2.40)***<br>M: 1.09 (0.82, 1.44) | 0.50 (0.10, 0.90)** |
| Feeling nervous or anxious§ | O: 1.38 (1.11, 1.72)**<br>F: 1.69 (1.28, 2.23)***<br>M: 1.13 (0.85, 1.52) | 0.40 (0.05, 0.76)** |
| Extent of suffering§ | O: 1.33 (1.09, 1.61)**<br>F: 1.36 (1.04, 1.77)**<br>M: 1.29 (0.98, 1.71)* | 0.05 (-0.33, 0.43) |
| Unable to control worry§ | O: 1.32 (1.07, 1.64)**<br>F: 1.49 (1.10, 2.02)**<br>M: 1.18 (0.88, 1.58) | 0.24 (-0.18, 0.65) |
| Forgiveness§ | O: 1.19 (0.96, 1.48)<br>F: 1.18 (0.87, 1.59)<br>M: 1.21 (0.88, 1.68) | -0.03 (-0.49, 0.42) |
| Depressive symptoms§ | O: 1.19 (0.96, 1.48)<br>F: 1.25 (0.94, 1.65)<br>M: 1.14 (0.85, 1.54) | 0.09 (-0.29, 0.47) |
| Various aspects of life are balanced§ | O: 1.15 (0.96, 1.38)<br>F: 1.09 (0.83, 1.42)<br>M: 1.21 (0.92, 1.58) | -0.11 (-0.50, 0.28) |
| Perceived discrimination§ | O: 1.15 (0.91, 1.44)<br>F: 1.23 (0.90, 1.67)<br>M: 1.08 (0.79, 1.47) | 0.13 (-0.30, 0.56) |
| Have little interest in doing things§ | O: 1.14 (0.89, 1.46)<br>F: 1.19 (0.92, 1.53)<br>M: 1.08 (0.76, 1.54) | 0.10 (-0.28, 0.47) |
| Feeling capable§ | O: 0.96 (0.79, 1.17)<br>F: 0.95 (0.72, 1.25)<br>M: 0.97 (0.73, 1.30) | -0.03 (-0.45, 0.39) |
| Trust in others§ | O: 0.93 (0.75, 1.14)<br>F: 0.84 (0.64, 1.09)<br>M: 1.01 (0.74, 1.37) | -0.19 (-0.57, 0.19) |
| Feel at peace§ | O: 0.92 (0.76, 1.13)<br>F: 0.90 (0.69, 1.18)<br>M: 0.95 (0.72, 1.25) | -0.05 (-0.41, 0.31) |
| Say in government# | O: 0.77 (0.59, 1.01)*<br>F: 0.65 (0.45, 0.92)**<br>M: 0.90 (0.63, 1.27) | -0.33 (-0.77, 0.12) |
| Obeying the law§ | O: 0.75 (0.49, 1.16)<br>F: 0.70 (0.44, 1.12)<br>M: 0.83 (0.42, 1.64) | -0.16 (-0.92, 0.59) |
| Government approval§ | O: 0.72 (0.56, 0.92)**<br>F: 0.77 (0.52, 1.15)<br>M: 0.67 (0.51, 0.89)** | 0.14 (-0.31, 0.60) |
| Support reducing differences in income levels§ | O: 0.59 (0.41, 0.84)**<br>F: 0.66 (0.43, 0.99)**<br>M: 0.52 (0.33, 0.82)** | 0.23 (-0.27, 0.72) |

### *Perceived family financial situation as very difficult while growing up – Large city or suburb of a large city*

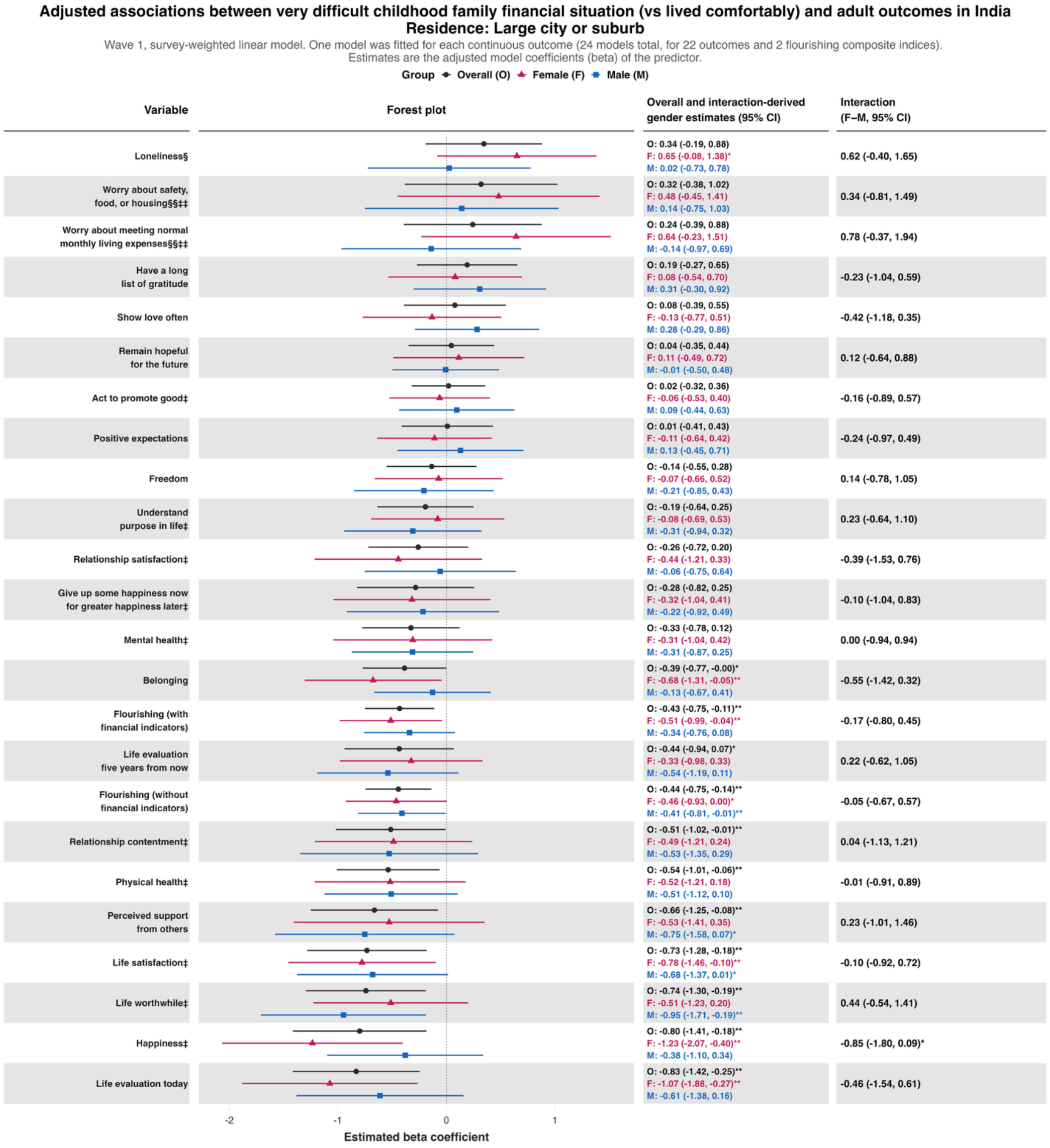

**Adjusted associations between very difficult childhood family financial situation (vs lived comfortably) and adult outcomes in India**
**Residence: Large city or suburb**

Wave 1, survey-weighted logistic model. One model was fitted for each binary outcome (7 models total). Estimates are adjusted odds ratios (OR) of the predictor.
The interaction column reports the difference in log-odds ratios (interaction beta) with 95% confidence intervals (95% CIs).

Group: Overall (O), Female (F), Male (M)

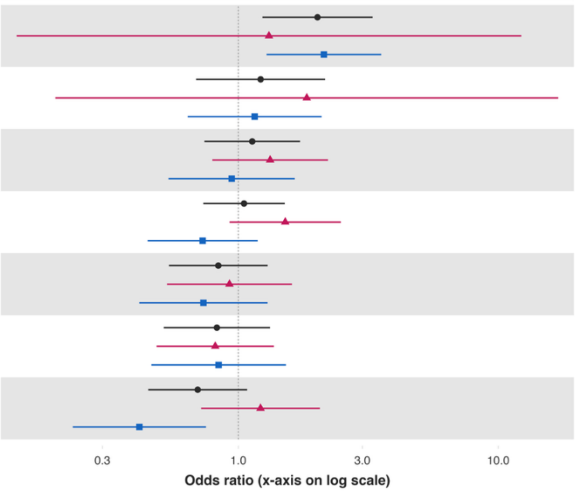


| Variable | Overall and interaction-derived gender estimates (95% CI) | Interaction (F–M, 95% CI) |
|---|---|---|
| Cigarette smoking | O: 2.02 (1.24, 3.28)**<br>F: 1.31 (0.14, 12.26)<br>M: 2.13 (1.28, 3.55)** | -0.49 (-2.80, 1.83) |
| Alcohol use | O: 1.22 (0.69, 2.16)<br>F: 1.83 (0.20, 17.00)<br>M: 1.16 (0.64, 2.09) | 0.46 (-1.91, 2.83) |
| Have any health problems | O: 1.13 (0.74, 1.73)<br>F: 1.33 (0.79, 2.21)<br>M: 0.94 (0.54, 1.65) | 0.34 (-0.31, 0.99) |
| Helped a stranger | O: 1.05 (0.73, 1.51)<br>F: 1.51 (0.93, 2.48)<br>M: 0.73 (0.45, 1.19) | 0.73 (0.06, 1.40)** |
| Feel close to someone | O: 0.84 (0.54, 1.30)<br>F: 0.92 (0.53, 1.61)<br>M: 0.73 (0.42, 1.30) | 0.23 (-0.47, 0.94) |
| Donated to charity | O: 0.83 (0.52, 1.32)<br>F: 0.81 (0.48, 1.37)<br>M: 0.84 (0.46, 1.52) | -0.03 (-0.64, 0.58) |
| Volunteered at an organization | O: 0.70 (0.45, 1.08)<br>F: 1.22 (0.72, 2.06)<br>M: 0.42 (0.23, 0.75)** | 1.07 (0.33, 1.81)** |

**Adjusted associations between very difficult childhood family financial situation (vs lived comfortably) and adult outcomes in India**
**Residence: Large city or suburb**

Wave 1, survey-weighted ordinal regression using the proportional odds model (`svyolr`). One model was fitted for each ordinal outcome (16 models total).
Estimates are adjusted cumulative odds ratios (cOR) of the predictor.
The interaction column reports the difference in log-cumulative odds ratios (interaction beta) with 95% confidence intervals (95% CIs).

**Group** ● Overall (O) ▲ Female (F) ■ Male (M)

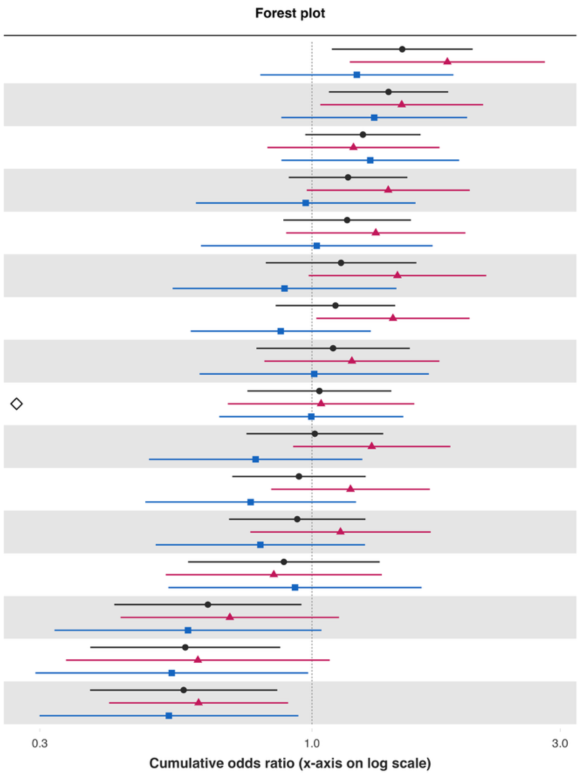


| Variable | Overall and interaction-derived gender estimates (95% CI) | Interaction (F–M, 95% CI) |
|---|---|---|
| Extent of suffering§ | O: 1.49 (1.09, 2.04)**<br>F: 1.82 (1.18, 2.80)**<br>M: 1.22 (0.80, 1.87) | 0.40 (-0.19, 0.99) |
| Unable to control worry§ | O: 1.40 (1.08, 1.83)**<br>F: 1.49 (1.04, 2.13)**<br>M: 1.32 (0.87, 1.99) | 0.12 (-0.45, 0.69) |
| Distress from life-threatening experiences§ | O: 1.25 (0.97, 1.62)*<br>F: 1.20 (0.82, 1.76)<br>M: 1.29 (0.87, 1.92) | -0.07 (-0.65, 0.51) |
| Feeling nervous or anxious§ | O: 1.17 (0.90, 1.53)<br>F: 1.40 (0.98, 2.01)*<br>M: 0.97 (0.60, 1.58) | 0.37 (-0.31, 1.04) |
| Feeling capable§ | O: 1.17 (0.88, 1.55)<br>F: 1.33 (0.89, 1.97)<br>M: 1.02 (0.61, 1.71) | 0.26 (-0.46, 0.98) |
| Have little interest in doing things§ | O: 1.14 (0.82, 1.59)<br>F: 1.46 (0.99, 2.16)*<br>M: 0.89 (0.54, 1.45) | 0.50 (-0.10, 1.10) |
| Depressive symptoms§ | O: 1.11 (0.85, 1.45)<br>F: 1.43 (1.02, 2.01)**<br>M: 0.87 (0.58, 1.30) | 0.50 (-0.03, 1.02)* |
| Forgiveness§ | O: 1.10 (0.78, 1.54)<br>F: 1.19 (0.81, 1.76)<br>M: 1.01 (0.61, 1.68) | 0.17 (-0.44, 0.78) |
| Feel at peace§ | O: 1.03 (0.75, 1.42)<br>F: 1.04 (0.69, 1.57)<br>M: 1.00 (0.66, 1.50) | 0.04 (-0.46, 0.55) |
| Various aspects of life are balanced§ | O: 1.01 (0.75, 1.37)<br>F: 1.30 (0.92, 1.84)<br>M: 0.78 (0.49, 1.25) | 0.51 (-0.06, 1.08)* |
| Perceived discrimination§ | O: 0.94 (0.70, 1.27)<br>F: 1.19 (0.84, 1.68)<br>M: 0.76 (0.48, 1.22) | 0.44 (-0.15, 1.03) |
| Trust in others§ | O: 0.94 (0.69, 1.27)<br>F: 1.13 (0.76, 1.69)<br>M: 0.80 (0.50, 1.26) | 0.35 (-0.28, 0.99) |
| Say in government# | O: 0.88 (0.58, 1.35)<br>F: 0.84 (0.52, 1.36)<br>M: 0.93 (0.53, 1.62) | -0.09 (-0.70, 0.52) |
| Support reducing differences in income levels§ | O: 0.63 (0.42, 0.95)**<br>F: 0.70 (0.43, 1.13)<br>M: 0.58 (0.32, 1.04)* | 0.19 (-0.53, 0.90) |
| Obeying the law§ | O: 0.57 (0.38, 0.87)**<br>F: 0.60 (0.34, 1.08)*<br>M: 0.54 (0.29, 0.98)** | 0.12 (-0.72, 0.95) |
| Government approval§ | O: 0.57 (0.37, 0.86)**<br>F: 0.61 (0.41, 0.90)**<br>M: 0.53 (0.30, 0.94)** | 0.13 (-0.40, 0.66) |

**eFigure 18.** Adjusted associations between childhood predictors and 22 continuous outcomes of interest and two flourishing composite indices for each age group, 18 to 24 inclusive, 25 to 29 inclusive, 30 to 39 inclusive, 40 to 49 inclusive, 50 to 59 inclusive, and 60+.

***Note:*** Estimates are from survey-weighted models fitted separately within each age group, using the same outcome coding, covariates (excluding age), and exposure-by-gender interaction as the primary analyses.

**Predictor:** ***Childhood abuse***
***Childhood abuse – Ages 18 to 24 inclusive***

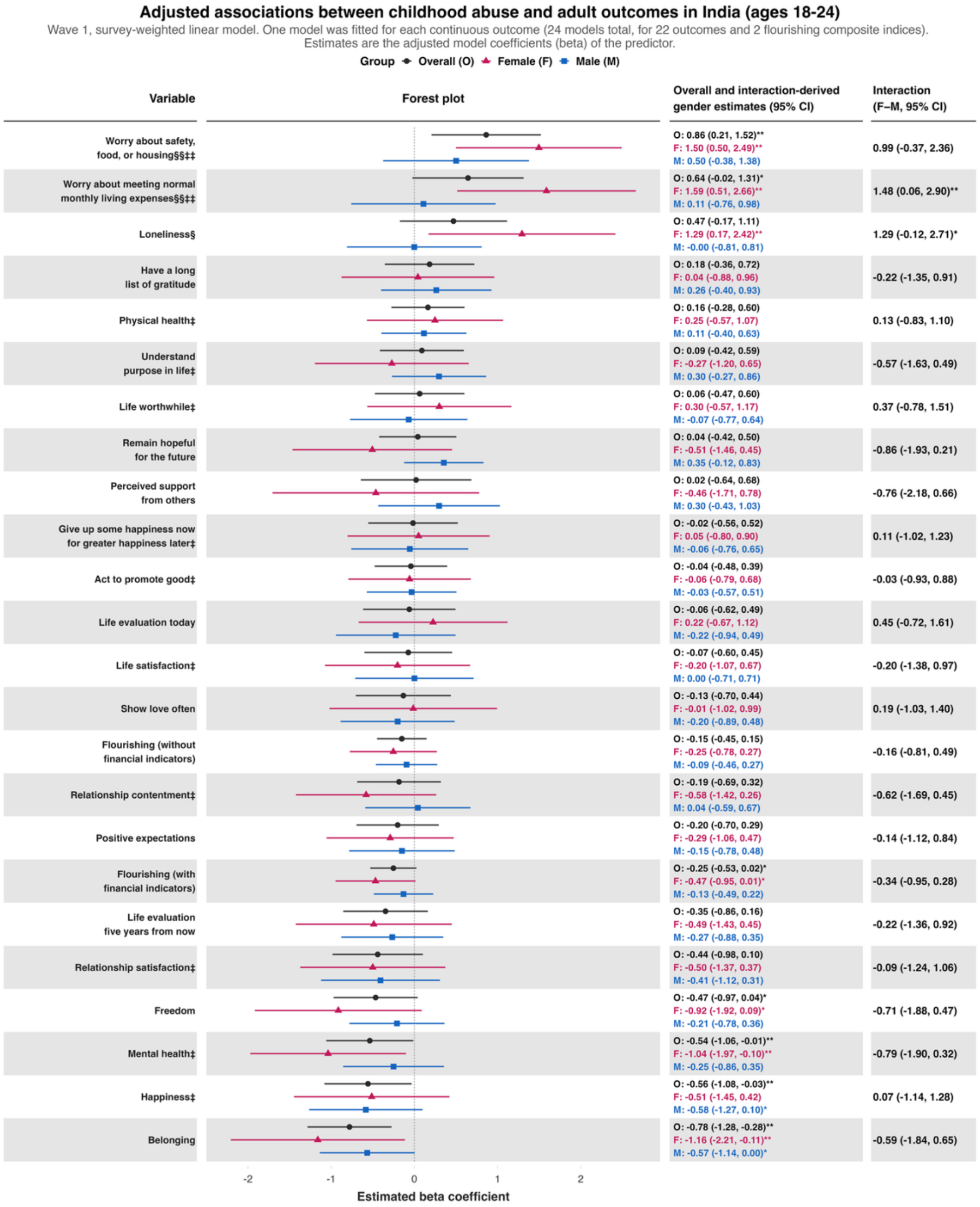

*Childhood abuse – Ages 25 to 29 inclusive*

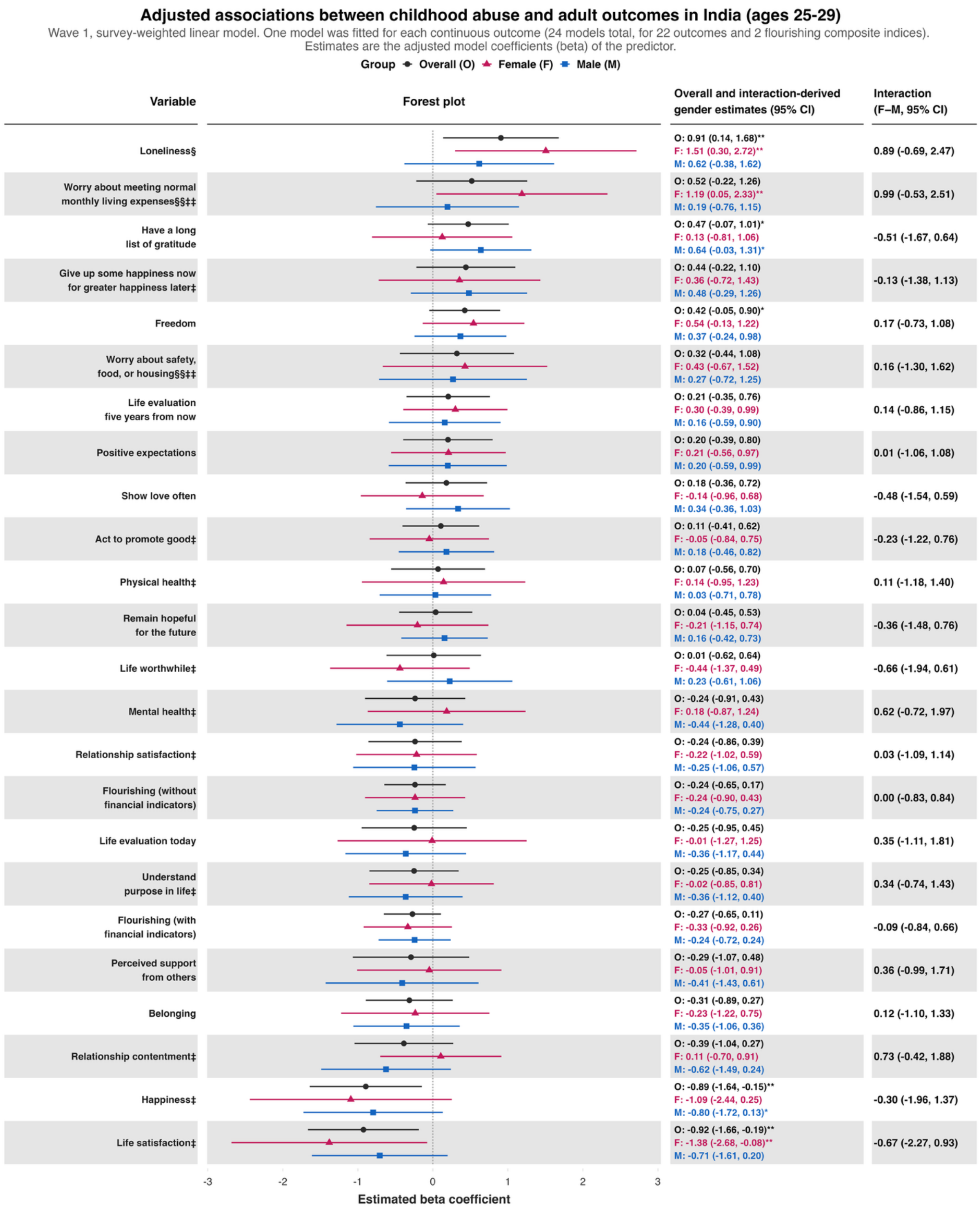

### *Childhood abuse – Ages 30 to 39 inclusive*

**Adjusted associations between childhood abuse and adult outcomes in India (ages 30-39)**

Wave 1, survey-weighted linear model. One model was fitted for each continuous outcome (24 models total, for 22 outcomes and 2 flourishing composite indices).
Estimates are the adjusted model coefficients (beta) of the predictor.

**Group** ● Overall (O) ▲ Female (F) ■ Male (M)

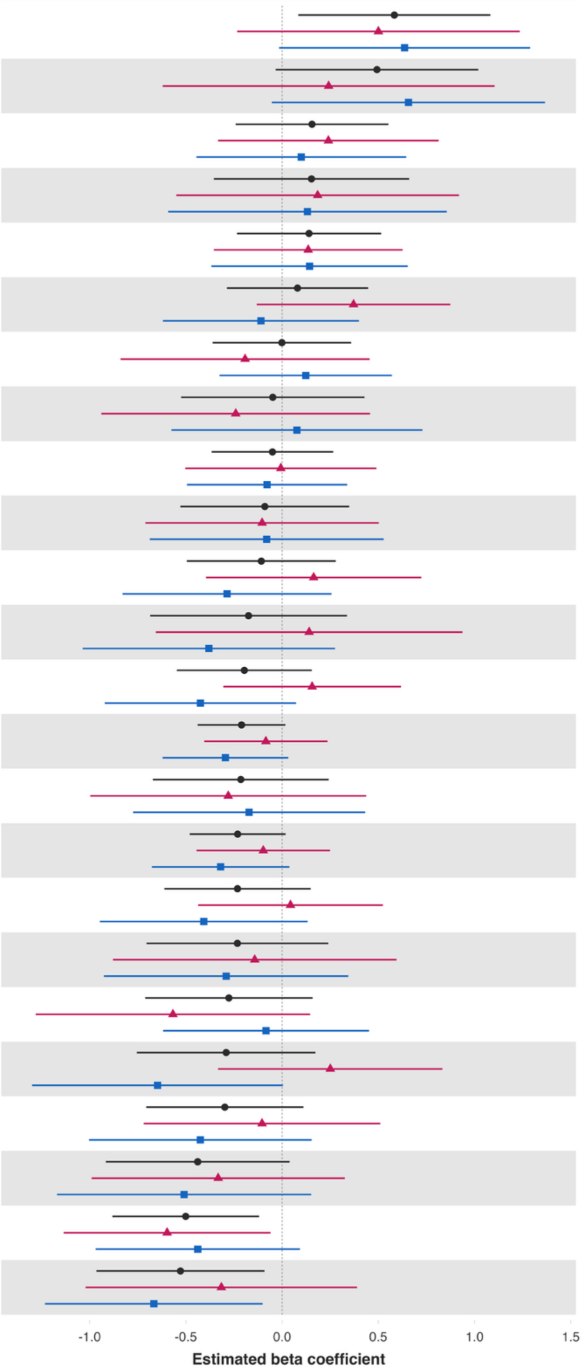


| Variable | Overall and interaction-derived gender estimates (95% CI) | Interaction (F–M, 95% CI) |
|---|---|---|
| Life evaluation today | O: 0.58 (0.08, 1.08)**<br>F: 0.50 (-0.23, 1.23)<br>M: 0.64 (-0.02, 1.29)* | -0.14 (-1.10, 0.83) |
| Loneliness§ | O: 0.49 (-0.03, 1.02)*<br>F: 0.24 (-0.62, 1.10)<br>M: 0.66 (-0.05, 1.37)* | -0.41 (-1.58, 0.75) |
| Relationship satisfaction‡ | O: 0.16 (-0.24, 0.55)<br>F: 0.24 (-0.33, 0.81)<br>M: 0.10 (-0.45, 0.64) | 0.14 (-0.66, 0.94) |
| Worry about safety, food, or housing§§‡‡ | O: 0.15 (-0.35, 0.66)<br>F: 0.18 (-0.55, 0.92)<br>M: 0.13 (-0.59, 0.86) | 0.05 (-1.02, 1.12) |
| Have a long list of gratitude | O: 0.14 (-0.23, 0.51)<br>F: 0.14 (-0.35, 0.63)<br>M: 0.14 (-0.37, 0.65) | -0.01 (-0.69, 0.68) |
| Show love often | O: 0.08 (-0.29, 0.45)<br>F: 0.37 (-0.13, 0.87)<br>M: -0.11 (-0.62, 0.40) | 0.48 (-0.23, 1.20) |
| Understand purpose in life‡ | O: -0.00 (-0.36, 0.36)<br>F: -0.19 (-0.84, 0.46)<br>M: 0.12 (-0.33, 0.57) | -0.32 (-1.13, 0.50) |
| Worry about meeting normal monthly living expenses§§‡‡ | O: -0.05 (-0.52, 0.43)<br>F: -0.24 (-0.94, 0.46)<br>M: 0.08 (-0.58, 0.73) | -0.32 (-1.29, 0.65) |
| Belonging | O: -0.05 (-0.37, 0.27)<br>F: -0.01 (-0.50, 0.49)<br>M: -0.08 (-0.49, 0.34) | 0.07 (-0.58, 0.73) |
| Relationship contentment‡ | O: -0.09 (-0.53, 0.35)<br>F: -0.10 (-0.71, 0.50)<br>M: -0.08 (-0.69, 0.53) | -0.02 (-0.88, 0.83) |
| Freedom | O: -0.11 (-0.50, 0.28)<br>F: 0.16 (-0.40, 0.72)<br>M: -0.29 (-0.83, 0.26) | 0.45 (-0.35, 1.25) |
| Perceived support from others | O: -0.17 (-0.69, 0.34)<br>F: 0.14 (-0.66, 0.94)<br>M: -0.38 (-1.04, 0.28) | 0.52 (-0.50, 1.54) |
| Act to promote good‡ | O: -0.20 (-0.55, 0.15)<br>F: 0.16 (-0.31, 0.62)<br>M: -0.42 (-0.92, 0.07)* | 0.58 (-0.10, 1.26)* |
| Flourishing (with financial indicators) | O: -0.21 (-0.44, 0.02)*<br>F: -0.08 (-0.40, 0.24)<br>M: -0.29 (-0.62, 0.03)* | 0.21 (-0.26, 0.68) |
| Life satisfaction‡ | O: -0.21 (-0.67, 0.24)<br>F: -0.28 (-1.00, 0.44)<br>M: -0.17 (-0.77, 0.43) | -0.11 (-1.06, 0.84) |
| Flourishing (without financial indicators) | O: -0.23 (-0.48, 0.02)*<br>F: -0.10 (-0.44, 0.25)<br>M: -0.32 (-0.68, 0.04)* | 0.22 (-0.29, 0.73) |
| Remain hopeful for the future | O: -0.23 (-0.61, 0.15)<br>F: 0.04 (-0.44, 0.52)<br>M: -0.41 (-0.95, 0.13) | 0.45 (-0.27, 1.17) |
| Happiness‡ | O: -0.23 (-0.70, 0.24)<br>F: -0.14 (-0.88, 0.59)<br>M: -0.29 (-0.93, 0.34) | 0.15 (-0.84, 1.14) |
| Life evaluation five years from now | O: -0.28 (-0.71, 0.16)<br>F: -0.57 (-1.28, 0.15)<br>M: -0.08 (-0.62, 0.45) | -0.48 (-1.37, 0.40) |
| Give up some happiness now for greater happiness later‡ | O: -0.29 (-0.75, 0.17)<br>F: 0.25 (-0.33, 0.83)<br>M: -0.65 (-1.30, 0.00)* | 0.90 (0.03, 1.76)** |
| Mental health‡ | O: -0.30 (-0.71, 0.11)<br>F: -0.10 (-0.72, 0.51)<br>M: -0.42 (-1.00, 0.15) | 0.32 (-0.56, 1.20) |
| Life worthwhile‡ | O: -0.44 (-0.92, 0.04)*<br>F: -0.33 (-0.99, 0.33)<br>M: -0.51 (-1.17, 0.15) | 0.18 (-0.75, 1.11) |
| Positive expectations | O: -0.50 (-0.88, -0.12)**<br>F: -0.60 (-1.13, -0.06)**<br>M: -0.44 (-0.97, 0.09) | -0.16 (-0.92, 0.60) |
| Physical health‡ | O: -0.53 (-0.96, -0.09)**<br>F: -0.32 (-1.02, 0.39)<br>M: -0.67 (-1.23, -0.10)** | 0.35 (-0.56, 1.26) |

### *Childhood abuse – Ages 40 to 49 inclusive*

#### Adjusted associations between childhood abuse and adult outcomes in India (ages 40-49)

Wave 1, survey-weighted linear model. One model was fitted for each continuous outcome (24 models total, for 22 outcomes and 2 flourishing composite indices). Estimates are the adjusted model coefficients (beta) of the predictor.

**Group** ● Overall (O) ▲ Female (F) ■ Male (M)

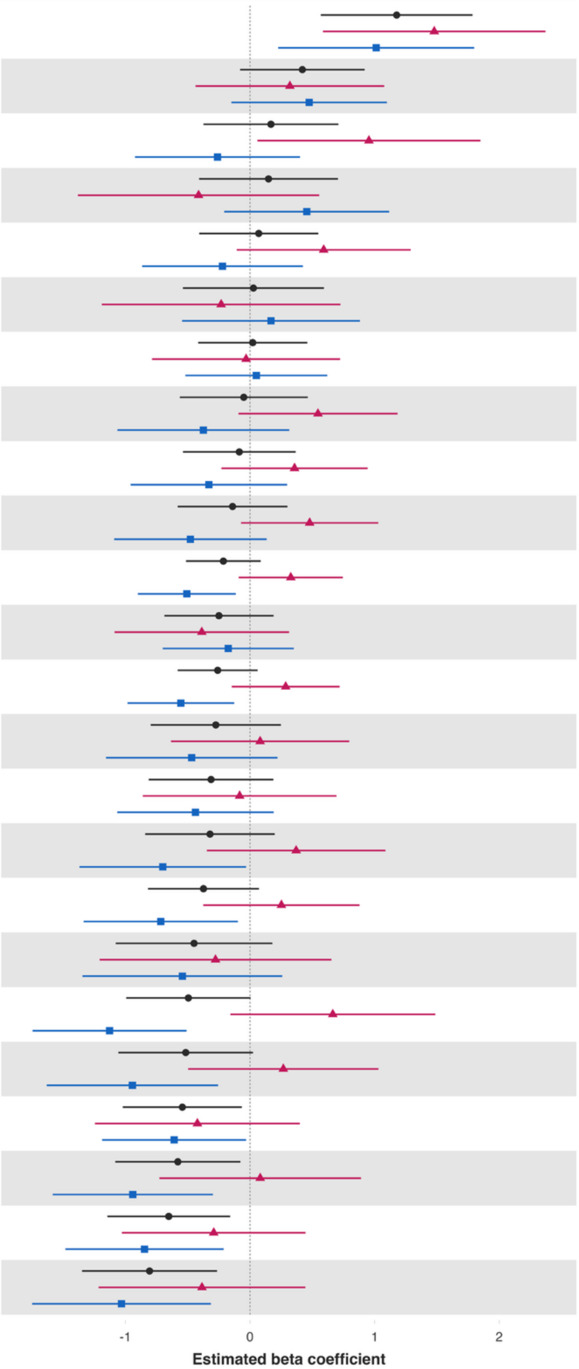


| Variable | Overall and interaction-derived gender estimates (95% CI) | Interaction (F–M, 95% CI) |
|---|---|---|
| Loneliness§ | O: 1.18 (0.57, 1.79)***<br>F: 1.48 (0.59, 2.37)**<br>M: 1.01 (0.23, 1.80)** | 0.47 (-0.70, 1.63) |
| Give up some happiness now for greater happiness later‡ | O: 0.42 (-0.08, 0.92)*<br>F: 0.32 (-0.44, 1.08)<br>M: 0.48 (-0.15, 1.10) | -0.16 (-1.10, 0.79) |
| Life evaluation today | O: 0.17 (-0.37, 0.71)<br>F: 0.95 (0.06, 1.85)**<br>M: -0.26 (-0.92, 0.40) | 1.22 (0.11, 2.32)** |
| Worry about meeting normal monthly living expenses§§‡‡ | O: 0.15 (-0.41, 0.71)<br>F: -0.41 (-1.38, 0.56)<br>M: 0.46 (-0.21, 1.12) | -0.87 (-2.03, 0.30) |
| Life evaluation five years from now | O: 0.07 (-0.41, 0.55)<br>F: 0.59 (-0.11, 1.29)*<br>M: -0.22 (-0.87, 0.42) | 0.81 (-0.16, 1.78) |
| Worry about safety, food, or housing§§‡‡ | O: 0.03 (-0.54, 0.59)<br>F: -0.23 (-1.19, 0.73)<br>M: 0.17 (-0.54, 0.88) | -0.40 (-1.61, 0.81) |
| Have a long list of gratitude | O: 0.02 (-0.42, 0.46)<br>F: -0.03 (-0.79, 0.72)<br>M: 0.05 (-0.52, 0.62) | -0.08 (-1.06, 0.90) |
| Relationship satisfaction‡ | O: -0.05 (-0.56, 0.46)<br>F: 0.55 (-0.09, 1.18)*<br>M: -0.37 (-1.06, 0.31) | 0.92 (0.00, 1.84)** |
| Act to promote good‡ | O: -0.09 (-0.54, 0.37)<br>F: 0.36 (-0.23, 0.94)<br>M: -0.33 (-0.96, 0.30) | 0.69 (-0.18, 1.56) |
| Understand purpose in life‡ | O: -0.14 (-0.58, 0.30)<br>F: 0.48 (-0.07, 1.03)*<br>M: -0.48 (-1.09, 0.13) | 0.96 (0.12, 1.79)** |
| Flourishing (with financial indicators) | O: -0.21 (-0.51, 0.09)<br>F: 0.33 (-0.09, 0.75)<br>M: -0.51 (-0.90, -0.11)** | 0.83 (0.26, 1.41)** |
| Freedom | O: -0.25 (-0.69, 0.19)<br>F: -0.39 (-1.09, 0.31)<br>M: -0.17 (-0.70, 0.35) | -0.21 (-1.05, 0.62) |
| Flourishing (without financial indicators) | O: -0.26 (-0.58, 0.06)<br>F: 0.29 (-0.15, 0.72)<br>M: -0.55 (-0.98, -0.13)** | 0.84 (0.23, 1.45)** |
| Relationship contentment‡ | O: -0.27 (-0.80, 0.25)<br>F: 0.08 (-0.63, 0.80)<br>M: -0.47 (-1.16, 0.22) | 0.55 (-0.42, 1.52) |
| Show love often | O: -0.31 (-0.81, 0.19)<br>F: -0.08 (-0.86, 0.69)<br>M: -0.44 (-1.07, 0.19) | 0.35 (-0.63, 1.34) |
| Life worthwhile‡ | O: -0.32 (-0.84, 0.20)<br>F: 0.37 (-0.35, 1.09)<br>M: -0.70 (-1.37, -0.03)** | 1.07 (0.12, 2.02)** |
| Positive expectations | O: -0.37 (-0.82, 0.07)<br>F: 0.25 (-0.37, 0.88)<br>M: -0.72 (-1.34, -0.10)** | 0.97 (0.05, 1.88)** |
| Perceived support from others | O: -0.45 (-1.08, 0.18)<br>F: -0.28 (-1.21, 0.65)<br>M: -0.54 (-1.34, 0.26) | 0.27 (-0.93, 1.46) |
| Physical health‡ | O: -0.49 (-0.99, 0.00)*<br>F: 0.66 (-0.16, 1.49)<br>M: -1.13 (-1.74, -0.51)*** | 1.79 (0.73, 2.86)*** |
| Life satisfaction‡ | O: -0.52 (-1.06, 0.02)*<br>F: 0.27 (-0.50, 1.03)<br>M: -0.94 (-1.63, -0.26)** | 1.21 (0.21, 2.21)** |
| Remain hopeful for the future | O: -0.54 (-1.02, -0.07)**<br>F: -0.42 (-1.24, 0.40)<br>M: -0.61 (-1.19, -0.03)** | 0.19 (-0.81, 1.18) |
| Mental health‡ | O: -0.58 (-1.08, -0.08)**<br>F: 0.08 (-0.73, 0.89)<br>M: -0.94 (-1.58, -0.30)** | 1.02 (-0.02, 2.07)* |
| Belonging | O: -0.65 (-1.14, -0.16)**<br>F: -0.29 (-1.03, 0.45)<br>M: -0.85 (-1.48, -0.21)** | 0.56 (-0.42, 1.53) |
| Happiness‡ | O: -0.81 (-1.35, -0.26)**<br>F: -0.39 (-1.22, 0.44)<br>M: -1.03 (-1.75, -0.31)** | 0.65 (-0.47, 1.76) |

### *Childhood abuse – Ages 50 to 59 inclusive*

#### Adjusted associations between childhood abuse and adult outcomes in India (ages 50-59)

Wave 1, survey-weighted linear model. One model was fitted for each continuous outcome (24 models total, for 22 outcomes and 2 flourishing composite indices). Estimates are the adjusted model coefficients (beta) of the predictor.

Group ● Overall (O) ▲ Female (F) ■ Male (M)

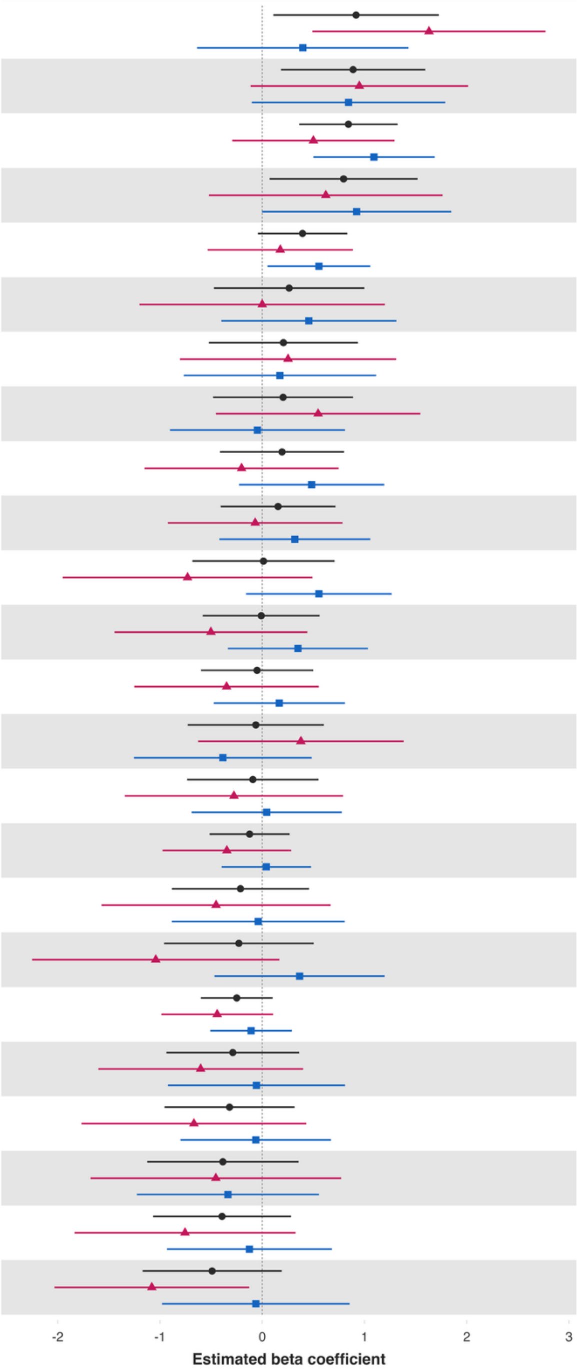


| Variable | Overall and interaction-derived gender estimates (95% CI) | Interaction (F–M, 95% CI) |
|---|---|---|
| Loneliness§ | O: 0.92 (0.11, 1.73)**<br>F: 1.63 (0.49, 2.77)**<br>M: 0.40 (-0.64, 1.43) | 1.23 (-0.24, 2.71) |
| Worry about meeting normal monthly living expenses§§‡‡ | O: 0.89 (0.18, 1.59)**<br>F: 0.95 (-0.11, 2.01)*<br>M: 0.84 (-0.10, 1.79)* | 0.11 (-1.32, 1.53) |
| Understand purpose in life‡ | O: 0.84 (0.36, 1.32)***<br>F: 0.50 (-0.29, 1.29)<br>M: 1.09 (0.50, 1.69)*** | -0.59 (-1.58, 0.39) |
| Worry about safety, food, or housing§§‡‡ | O: 0.80 (0.07, 1.52)**<br>F: 0.62 (-0.52, 1.76)<br>M: 0.92 (-0.00, 1.85)* | -0.30 (-1.76, 1.16) |
| Positive expectations | O: 0.39 (-0.04, 0.83)*<br>F: 0.18 (-0.53, 0.89)<br>M: 0.55 (0.05, 1.06)** | -0.38 (-1.21, 0.46) |
| Perceived support from others | O: 0.26 (-0.47, 1.00)<br>F: -0.00 (-1.20, 1.20)<br>M: 0.46 (-0.40, 1.31) | -0.46 (-1.87, 0.96) |
| Life evaluation today | O: 0.21 (-0.52, 0.94)<br>F: 0.25 (-0.80, 1.31)<br>M: 0.17 (-0.77, 1.11) | 0.08 (-1.28, 1.44) |
| Life worthwhile‡ | O: 0.20 (-0.48, 0.89)<br>F: 0.55 (-0.45, 1.55)<br>M: -0.05 (-0.90, 0.81) | 0.59 (-0.66, 1.84) |
| Have a long list of gratitude | O: 0.19 (-0.41, 0.80)<br>F: -0.20 (-1.15, 0.75)<br>M: 0.48 (-0.23, 1.19) | -0.69 (-1.79, 0.42) |
| Freedom | O: 0.15 (-0.41, 0.72)<br>F: -0.07 (-0.92, 0.79)<br>M: 0.32 (-0.42, 1.06) | -0.39 (-1.52, 0.74) |
| Show love often | O: 0.01 (-0.68, 0.71)<br>F: -0.73 (-1.95, 0.49)<br>M: 0.55 (-0.16, 1.27) | -1.28 (-2.67, 0.10)* |
| Act to promote good‡ | O: -0.01 (-0.58, 0.56)<br>F: -0.50 (-1.45, 0.44)<br>M: 0.35 (-0.34, 1.03) | -0.85 (-2.00, 0.29) |
| Remain hopeful for the future | O: -0.05 (-0.60, 0.50)<br>F: -0.35 (-1.25, 0.55)<br>M: 0.17 (-0.47, 0.81) | -0.52 (-1.58, 0.55) |
| Physical health‡ | O: -0.06 (-0.73, 0.60)<br>F: 0.38 (-0.63, 1.38)<br>M: -0.39 (-1.26, 0.49) | 0.76 (-0.56, 2.09) |
| Give up some happiness now for greater happiness later‡ | O: -0.09 (-0.73, 0.55)<br>F: -0.28 (-1.34, 0.79)<br>M: 0.04 (-0.69, 0.78) | -0.32 (-1.57, 0.93) |
| Flourishing (without financial indicators) | O: -0.12 (-0.51, 0.27)<br>F: -0.35 (-0.98, 0.28)<br>M: 0.04 (-0.40, 0.48) | -0.39 (-1.10, 0.33) |
| Life satisfaction‡ | O: -0.21 (-0.88, 0.46)<br>F: -0.45 (-1.57, 0.67)<br>M: -0.04 (-0.89, 0.81) | -0.41 (-1.84, 1.01) |
| Relationship contentment‡ | O: -0.23 (-0.96, 0.50)<br>F: -1.04 (-2.25, 0.17)*<br>M: 0.37 (-0.47, 1.20) | -1.41 (-2.83, 0.01)* |
| Flourishing (with financial indicators) | O: -0.25 (-0.60, 0.10)<br>F: -0.44 (-0.99, 0.11)<br>M: -0.11 (-0.51, 0.29) | -0.33 (-0.96, 0.29) |
| Relationship satisfaction‡ | O: -0.29 (-0.94, 0.36)<br>F: -0.60 (-1.60, 0.40)<br>M: -0.06 (-0.92, 0.81) | -0.55 (-1.88, 0.79) |
| Mental health‡ | O: -0.32 (-0.96, 0.32)<br>F: -0.67 (-1.77, 0.43)<br>M: -0.06 (-0.80, 0.67) | -0.60 (-1.91, 0.70) |
| Happiness‡ | O: -0.39 (-1.13, 0.36)<br>F: -0.45 (-1.68, 0.77)<br>M: -0.33 (-1.23, 0.56) | -0.12 (-1.61, 1.38) |
| Belonging | O: -0.39 (-1.07, 0.28)<br>F: -0.76 (-1.84, 0.32)<br>M: -0.13 (-0.93, 0.68) | -0.63 (-1.94, 0.68) |
| Life evaluation five years from now | O: -0.49 (-1.17, 0.19)<br>F: -1.08 (-2.03, -0.13)**<br>M: -0.06 (-0.98, 0.85) | -1.02 (-2.32, 0.28) |

### *Childhood abuse – Ages 60+*

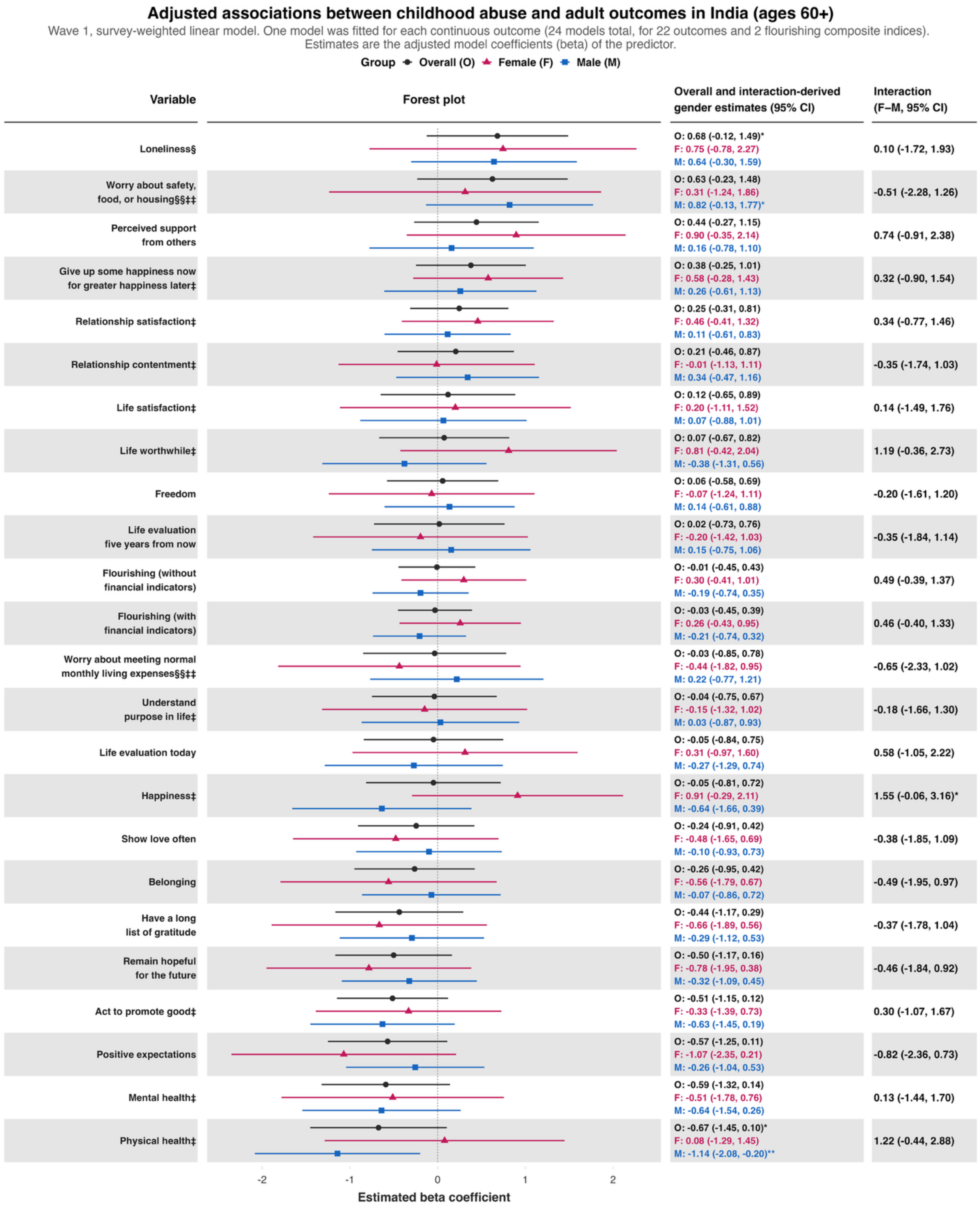

**Predictor:** ***Feeling like an outsider in one's family when growing up***
***Feeling like an outsider in one's family when growing up – Ages 18 to 24 inclusive***

**Adjusted associations between feeling like an outsider in one's family when growing up and adult outcomes in India (ages 18-24)**

Wave 1, survey-weighted linear model. One model was fitted for each continuous outcome (24 models total, for 22 outcomes and 2 flourishing composite indices).
Estimates are the adjusted model coefficients (beta) of the predictor.

Group: Overall (O), Female (F), Male (M)

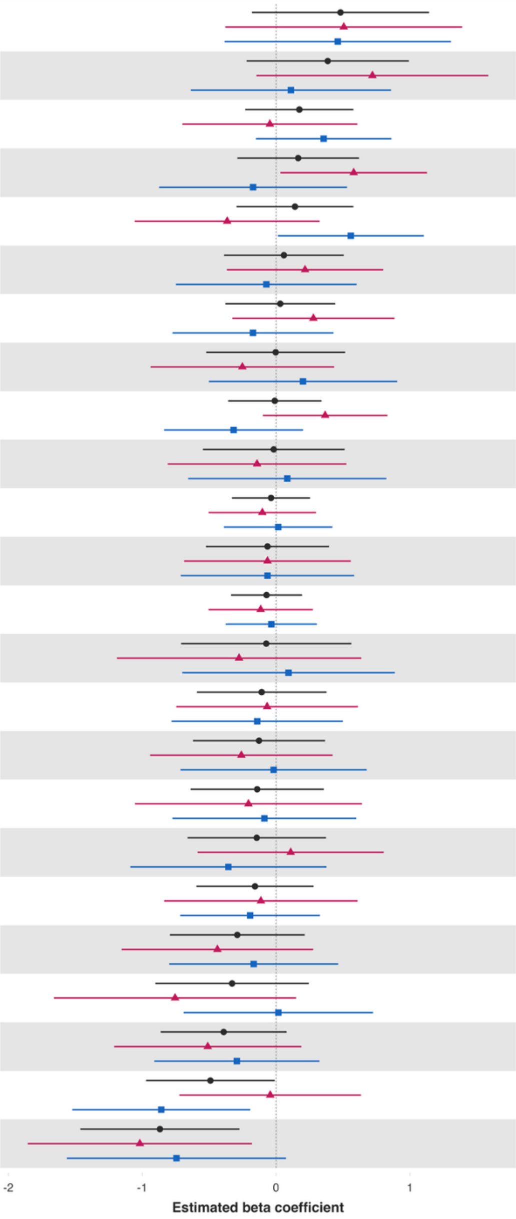


| Variable | Overall and interaction-derived gender estimates (95% CI) | Interaction (F–M, 95% CI) |
|---|---|---|
| Worry about safety, food, or housing§§‡‡ | O: 0.48 (-0.18, 1.14)<br>F: 0.51 (-0.38, 1.39)<br>M: 0.46 (-0.39, 1.31) | 0.04 (-1.07, 1.16) |
| Loneliness§ | O: 0.39 (-0.22, 0.99)<br>F: 0.72 (-0.15, 1.59)<br>M: 0.11 (-0.64, 0.86) | 0.61 (-0.44, 1.66) |
| Show love often | O: 0.17 (-0.23, 0.58)<br>F: -0.05 (-0.70, 0.61)<br>M: 0.36 (-0.15, 0.86) | -0.40 (-1.23, 0.43) |
| Have a long list of gratitude | O: 0.16 (-0.29, 0.62)<br>F: 0.58 (0.03, 1.13)**<br>M: -0.17 (-0.87, 0.53) | 0.75 (-0.16, 1.66) |
| Relationship satisfaction‡ | O: 0.14 (-0.30, 0.58)<br>F: -0.37 (-1.06, 0.33)<br>M: 0.56 (0.01, 1.10)** | -0.92 (-1.81, -0.04)** |
| Understand purpose in life‡ | O: 0.06 (-0.39, 0.51)<br>F: 0.22 (-0.37, 0.80)<br>M: -0.07 (-0.75, 0.60) | 0.29 (-0.62, 1.20) |
| Freedom | O: 0.03 (-0.38, 0.44)<br>F: 0.28 (-0.33, 0.88)<br>M: -0.17 (-0.77, 0.43) | 0.45 (-0.44, 1.34) |
| Happiness‡ | O: -0.00 (-0.52, 0.52)<br>F: -0.25 (-0.94, 0.43)<br>M: 0.20 (-0.50, 0.90) | -0.45 (-1.39, 0.49) |
| Act to promote good‡ | O: -0.01 (-0.36, 0.34)<br>F: 0.37 (-0.10, 0.83)<br>M: -0.32 (-0.84, 0.20) | 0.68 (-0.03, 1.39)* |
| Life worthwhile‡ | O: -0.02 (-0.55, 0.51)<br>F: -0.14 (-0.81, 0.52)<br>M: 0.08 (-0.66, 0.82) | -0.23 (-1.17, 0.72) |
| Flourishing (without financial indicators) | O: -0.04 (-0.33, 0.25)<br>F: -0.10 (-0.50, 0.30)<br>M: 0.02 (-0.39, 0.42) | -0.12 (-0.68, 0.44) |
| Physical health‡ | O: -0.06 (-0.52, 0.40)<br>F: -0.06 (-0.69, 0.56)<br>M: -0.06 (-0.71, 0.58) | -0.00 (-0.89, 0.88) |
| Flourishing (with financial indicators) | O: -0.07 (-0.34, 0.19)<br>F: -0.11 (-0.50, 0.27)<br>M: -0.04 (-0.38, 0.31) | -0.08 (-0.58, 0.42) |
| Worry about meeting normal monthly living expenses§§‡‡ | O: -0.07 (-0.71, 0.56)<br>F: -0.28 (-1.19, 0.64)<br>M: 0.09 (-0.70, 0.89) | -0.37 (-1.51, 0.77) |
| Mental health‡ | O: -0.11 (-0.59, 0.38)<br>F: -0.07 (-0.75, 0.61)<br>M: -0.14 (-0.78, 0.50) | 0.07 (-0.82, 0.97) |
| Life satisfaction‡ | O: -0.13 (-0.62, 0.37)<br>F: -0.26 (-0.94, 0.42)<br>M: -0.02 (-0.71, 0.68) | -0.24 (-1.21, 0.73) |
| Give up some happiness now for greater happiness later‡ | O: -0.14 (-0.64, 0.36)<br>F: -0.21 (-1.05, 0.64)<br>M: -0.09 (-0.77, 0.60) | -0.12 (-1.28, 1.04) |
| Life evaluation five years from now | O: -0.14 (-0.66, 0.37)<br>F: 0.11 (-0.59, 0.80)<br>M: -0.36 (-1.09, 0.38) | 0.47 (-0.53, 1.46) |
| Belonging | O: -0.16 (-0.60, 0.28)<br>F: -0.11 (-0.84, 0.61)<br>M: -0.19 (-0.72, 0.33) | 0.08 (-0.80, 0.97) |
| Relationship contentment‡ | O: -0.29 (-0.79, 0.21)<br>F: -0.44 (-1.15, 0.28)<br>M: -0.17 (-0.80, 0.46) | -0.27 (-1.16, 0.62) |
| Life evaluation today | O: -0.33 (-0.90, 0.24)<br>F: -0.75 (-1.66, 0.15)<br>M: 0.02 (-0.69, 0.72) | -0.77 (-1.90, 0.36) |
| Positive expectations | O: -0.39 (-0.86, 0.08)<br>F: -0.51 (-1.21, 0.19)<br>M: -0.29 (-0.91, 0.32) | -0.22 (-1.13, 0.70) |
| Remain hopeful for the future | O: -0.49 (-0.97, -0.01)**<br>F: -0.04 (-0.72, 0.63)<br>M: -0.86 (-1.52, -0.19)** | 0.81 (-0.13, 1.76)* |
| Perceived support from others | O: -0.87 (-1.46, -0.27)**<br>F: -1.02 (-1.86, -0.18)**<br>M: -0.74 (-1.56, 0.07)* | -0.27 (-1.43, 0.88) |

*Feeling like an outsider in one's family when growing up – Ages 25 to 29 inclusive*

**Adjusted associations between feeling like an outsider in one's family when growing up and adult outcomes in India (ages 25-29)**

Wave 1, survey-weighted linear model. One model was fitted for each continuous outcome (24 models total, for 22 outcomes and 2 flourishing composite indices).
Estimates are the adjusted model coefficients (beta) of the predictor.

Group: Overall (O), Female (F), Male (M)

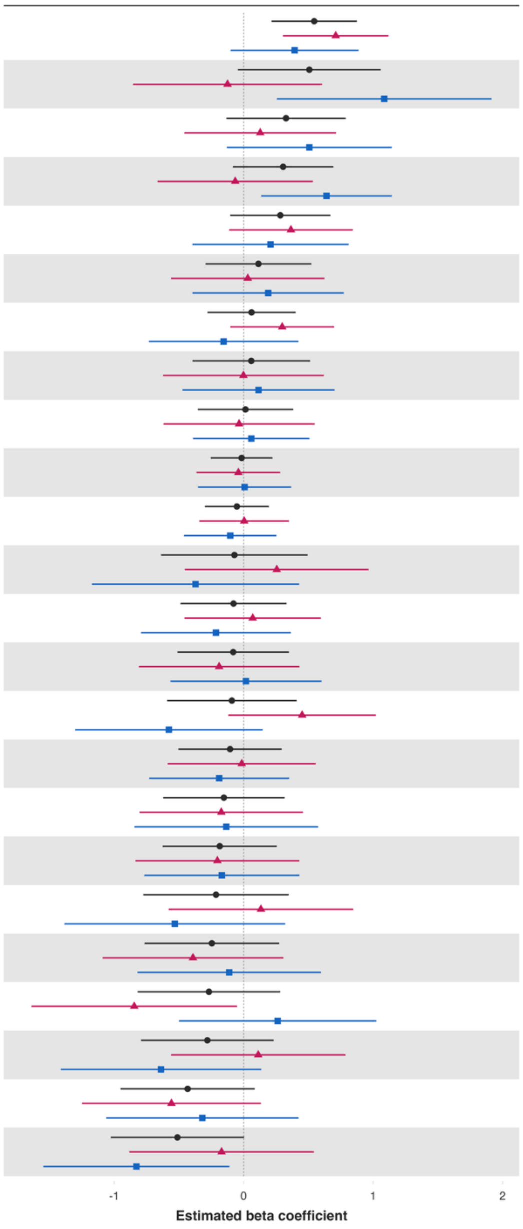


| Variable | Overall and interaction-derived gender estimates (95% CI) | Interaction (F–M, 95% CI) |
|---|---|---|
| Understand purpose in life‡ | O: 0.54 (0.21, 0.87)**<br>F: 0.71 (0.30, 1.12)***<br>M: 0.39 (-0.10, 0.89) | 0.32 (-0.31, 0.94) |
| Loneliness§ | O: 0.51 (-0.04, 1.06)*<br>F: -0.12 (-0.85, 0.61)<br>M: 1.09 (0.26, 1.91)** | -1.21 (-2.34, -0.08)** |
| Have a long list of gratitude | O: 0.33 (-0.13, 0.79)<br>F: 0.13 (-0.46, 0.71)<br>M: 0.51 (-0.13, 1.14) | -0.38 (-1.19, 0.43) |
| Relationship contentment‡ | O: 0.30 (-0.08, 0.69)<br>F: -0.07 (-0.67, 0.53)<br>M: 0.64 (0.14, 1.14)** | -0.71 (-1.50, 0.09)* |
| Positive expectations | O: 0.28 (-0.10, 0.67)<br>F: 0.36 (-0.11, 0.84)<br>M: 0.21 (-0.40, 0.81) | 0.16 (-0.61, 0.93) |
| Freedom | O: 0.11 (-0.29, 0.52)<br>F: 0.03 (-0.56, 0.62)<br>M: 0.19 (-0.40, 0.77) | -0.16 (-1.00, 0.69) |
| Act to promote good‡ | O: 0.06 (-0.28, 0.40)<br>F: 0.30 (-0.10, 0.70)<br>M: -0.16 (-0.73, 0.42) | 0.45 (-0.28, 1.19) |
| Show love often | O: 0.06 (-0.40, 0.51)<br>F: -0.00 (-0.62, 0.62)<br>M: 0.11 (-0.47, 0.70) | -0.12 (-0.91, 0.68) |
| Remain hopeful for the future | O: 0.01 (-0.35, 0.38)<br>F: -0.04 (-0.62, 0.55)<br>M: 0.06 (-0.39, 0.51) | -0.09 (-0.82, 0.64) |
| Flourishing (with financial indicators) | O: -0.02 (-0.25, 0.22)<br>F: -0.04 (-0.36, 0.28)<br>M: 0.01 (-0.35, 0.37) | -0.05 (-0.54, 0.45) |
| Flourishing (without financial indicators) | O: -0.05 (-0.30, 0.19)<br>F: 0.00 (-0.34, 0.35)<br>M: -0.10 (-0.46, 0.25) | 0.11 (-0.39, 0.61) |
| Worry about meeting normal monthly living expenses§§‡‡ | O: -0.07 (-0.64, 0.49)<br>F: 0.26 (-0.45, 0.96)<br>M: -0.37 (-1.17, 0.43) | 0.63 (-0.39, 1.64) |
| Belonging | O: -0.08 (-0.49, 0.33)<br>F: 0.07 (-0.46, 0.60)<br>M: -0.21 (-0.79, 0.36) | 0.28 (-0.46, 1.03) |
| Mental health‡ | O: -0.08 (-0.51, 0.35)<br>F: -0.19 (-0.81, 0.43)<br>M: 0.02 (-0.57, 0.60) | -0.21 (-1.05, 0.63) |
| Life worthwhile‡ | O: -0.09 (-0.59, 0.41)<br>F: 0.45 (-0.12, 1.02)<br>M: -0.58 (-1.30, 0.15) | 1.03 (0.16, 1.90)** |
| Life evaluation five years from now | O: -0.11 (-0.50, 0.29)<br>F: -0.02 (-0.59, 0.56)<br>M: -0.19 (-0.73, 0.35) | 0.17 (-0.60, 0.95) |
| Relationship satisfaction‡ | O: -0.15 (-0.62, 0.32)<br>F: -0.17 (-0.80, 0.46)<br>M: -0.13 (-0.84, 0.57) | -0.04 (-1.00, 0.93) |
| Physical health‡ | O: -0.19 (-0.63, 0.26)<br>F: -0.20 (-0.84, 0.43)<br>M: -0.17 (-0.77, 0.43) | -0.03 (-0.89, 0.82) |
| Worry about safety, food, or housing§§‡‡ | O: -0.21 (-0.78, 0.35)<br>F: 0.13 (-0.58, 0.85)<br>M: -0.53 (-1.38, 0.32) | 0.67 (-0.45, 1.78) |
| Give up some happiness now for greater happiness later‡ | O: -0.25 (-0.77, 0.27)<br>F: -0.39 (-1.09, 0.31)<br>M: -0.11 (-0.82, 0.60) | -0.28 (-1.23, 0.67) |
| Perceived support from others | O: -0.27 (-0.82, 0.28)<br>F: -0.85 (-1.64, -0.05)**<br>M: 0.26 (-0.50, 1.02) | -1.11 (-2.22, 0.00)* |
| Life satisfaction‡ | O: -0.28 (-0.79, 0.23)<br>F: 0.11 (-0.56, 0.79)<br>M: -0.64 (-1.41, 0.14) | 0.75 (-0.29, 1.79) |
| Life evaluation today | O: -0.43 (-0.95, 0.09)<br>F: -0.56 (-1.25, 0.13)<br>M: -0.32 (-1.06, 0.42) | -0.24 (-1.24, 0.76) |
| Happiness‡ | O: -0.51 (-1.03, 0.00)*<br>F: -0.17 (-0.88, 0.54)<br>M: -0.83 (-1.55, -0.11)** | 0.66 (-0.34, 1.65) |

*Feeling like an outsider in one's family when growing up – Ages 30 to 39 inclusive*

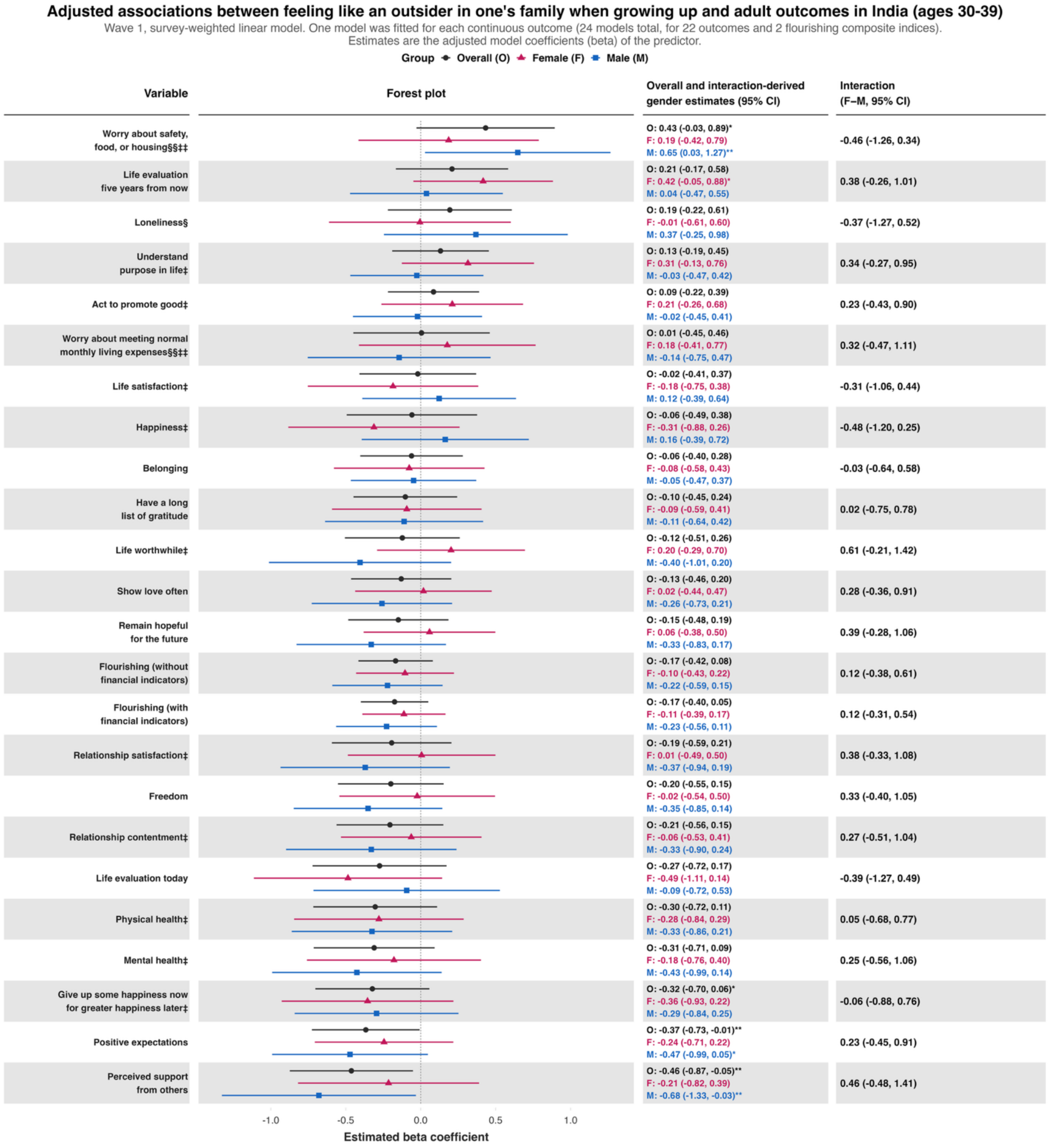

### *Feeling like an outsider in one's family when growing up – Ages 40 to 49 inclusive*

#### Adjusted associations between feeling like an outsider in one's family when growing up and adult outcomes in India (ages 40-49)

Wave 1, survey-weighted linear model. One model was fitted for each continuous outcome (24 models total, for 22 outcomes and 2 flourishing composite indices). Estimates are the adjusted model coefficients (beta) of the predictor.

Group: Overall (O), Female (F), Male (M)

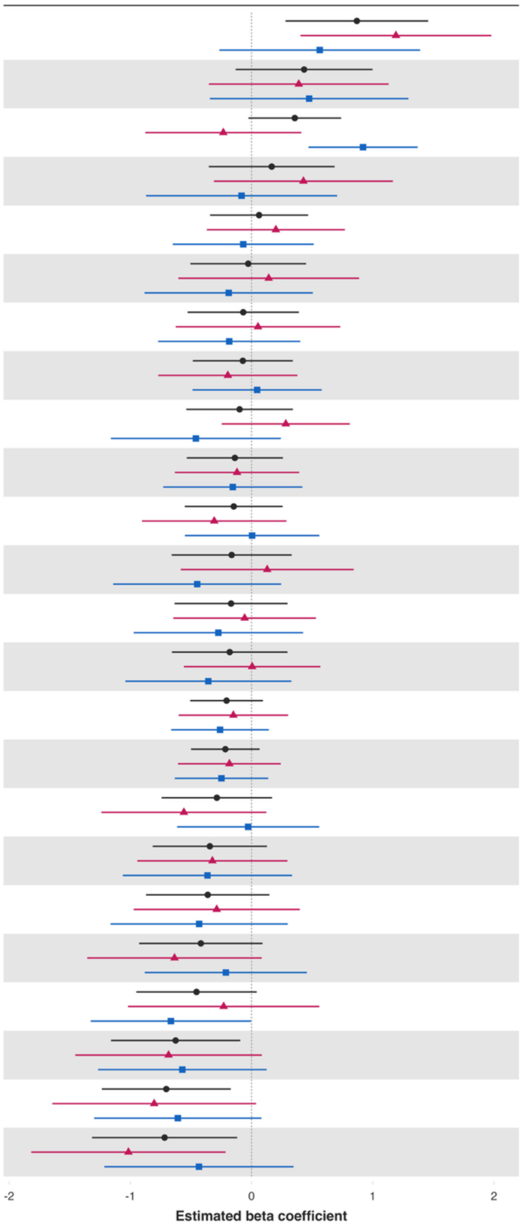


| Variable | Overall and interaction-derived gender estimates (95% CI) | Interaction (F–M, 95% CI) |
|---|---|---|
| Loneliness§ | O: 0.87 (0.28, 1.46)**<br>F: 1.19 (0.40, 1.98)**<br>M: 0.56 (-0.27, 1.39) | 0.63 (-0.49, 1.74) |
| Worry about safety, food, or housing§§‡‡ | O: 0.43 (-0.13, 1.00)<br>F: 0.39 (-0.35, 1.13)<br>M: 0.48 (-0.34, 1.29) | -0.09 (-1.17, 1.00) |
| Belonging | O: 0.36 (-0.03, 0.74)*<br>F: -0.23 (-0.88, 0.41)<br>M: 0.92 (0.47, 1.37)*** | -1.15 (-1.95, -0.35)** |
| Worry about meeting normal monthly living expenses§§‡‡ | O: 0.17 (-0.35, 0.69)<br>F: 0.43 (-0.31, 1.17)<br>M: -0.08 (-0.87, 0.71) | 0.51 (-0.61, 1.63) |
| Have a long list of gratitude | O: 0.06 (-0.34, 0.47)<br>F: 0.20 (-0.37, 0.77)<br>M: -0.07 (-0.65, 0.51) | 0.27 (-0.55, 1.09) |
| Life satisfaction‡ | O: -0.03 (-0.50, 0.45)<br>F: 0.14 (-0.60, 0.89)<br>M: -0.19 (-0.88, 0.51) | 0.33 (-0.75, 1.41) |
| Physical health‡ | O: -0.07 (-0.53, 0.39)<br>F: 0.05 (-0.63, 0.73)<br>M: -0.18 (-0.77, 0.40) | 0.24 (-0.64, 1.11) |
| Remain hopeful for the future | O: -0.07 (-0.48, 0.34)<br>F: -0.20 (-0.77, 0.38)<br>M: 0.05 (-0.49, 0.58) | -0.24 (-0.98, 0.49) |
| Relationship contentment‡ | O: -0.10 (-0.54, 0.34)<br>F: 0.28 (-0.25, 0.81)<br>M: -0.46 (-1.16, 0.24) | 0.74 (-0.14, 1.63) |
| Positive expectations | O: -0.14 (-0.53, 0.26)<br>F: -0.12 (-0.63, 0.39)<br>M: -0.15 (-0.73, 0.42) | 0.04 (-0.71, 0.78) |
| Act to promote good‡ | O: -0.15 (-0.55, 0.26)<br>F: -0.31 (-0.90, 0.29)<br>M: 0.00 (-0.55, 0.56) | -0.31 (-1.13, 0.51) |
| Life worthwhile‡ | O: -0.16 (-0.66, 0.33)<br>F: 0.13 (-0.58, 0.84)<br>M: -0.45 (-1.14, 0.24) | 0.58 (-0.43, 1.58) |
| Understand purpose in life‡ | O: -0.17 (-0.64, 0.30)<br>F: -0.06 (-0.65, 0.53)<br>M: -0.27 (-0.97, 0.43) | 0.22 (-0.69, 1.12) |
| Show love often | O: -0.18 (-0.66, 0.30)<br>F: 0.00 (-0.56, 0.57)<br>M: -0.36 (-1.04, 0.33) | 0.36 (-0.46, 1.18) |
| Flourishing (without financial indicators) | O: -0.21 (-0.51, 0.09)<br>F: -0.15 (-0.60, 0.30)<br>M: -0.26 (-0.66, 0.14) | 0.11 (-0.50, 0.72) |
| Flourishing (with financial indicators) | O: -0.22 (-0.50, 0.07)<br>F: -0.18 (-0.61, 0.24)<br>M: -0.25 (-0.63, 0.14) | 0.07 (-0.51, 0.65) |
| Freedom | O: -0.29 (-0.74, 0.17)<br>F: -0.56 (-1.24, 0.12)<br>M: -0.03 (-0.61, 0.56) | -0.53 (-1.41, 0.35) |
| Relationship satisfaction‡ | O: -0.34 (-0.82, 0.13)<br>F: -0.32 (-0.94, 0.30)<br>M: -0.36 (-1.06, 0.33) | 0.04 (-0.89, 0.97) |
| Life evaluation five years from now | O: -0.36 (-0.87, 0.15)<br>F: -0.29 (-0.97, 0.40)<br>M: -0.43 (-1.16, 0.30) | 0.15 (-0.84, 1.13) |
| Mental health‡ | O: -0.42 (-0.93, 0.09)<br>F: -0.64 (-1.36, 0.08)*<br>M: -0.21 (-0.88, 0.46) | -0.42 (-1.37, 0.52) |
| Happiness‡ | O: -0.45 (-0.95, 0.04)*<br>F: -0.23 (-1.02, 0.56)<br>M: -0.67 (-1.33, -0.00)** | 0.44 (-0.63, 1.50) |
| Life evaluation today | O: -0.63 (-1.16, -0.09)**<br>F: -0.68 (-1.45, 0.09)*<br>M: -0.57 (-1.27, 0.12) | -0.11 (-1.12, 0.89) |
| Give up some happiness now for greater happiness later‡ | O: -0.70 (-1.24, -0.17)**<br>F: -0.80 (-1.64, 0.04)*<br>M: -0.61 (-1.30, 0.08)* | -0.20 (-1.30, 0.91) |
| Perceived support from others | O: -0.72 (-1.32, -0.12)**<br>F: -1.02 (-1.82, -0.21)**<br>M: -0.43 (-1.21, 0.35) | -0.58 (-1.62, 0.45) |

*Feeling like an outsider in one's family when growing up – Ages 50 to 59 inclusive*

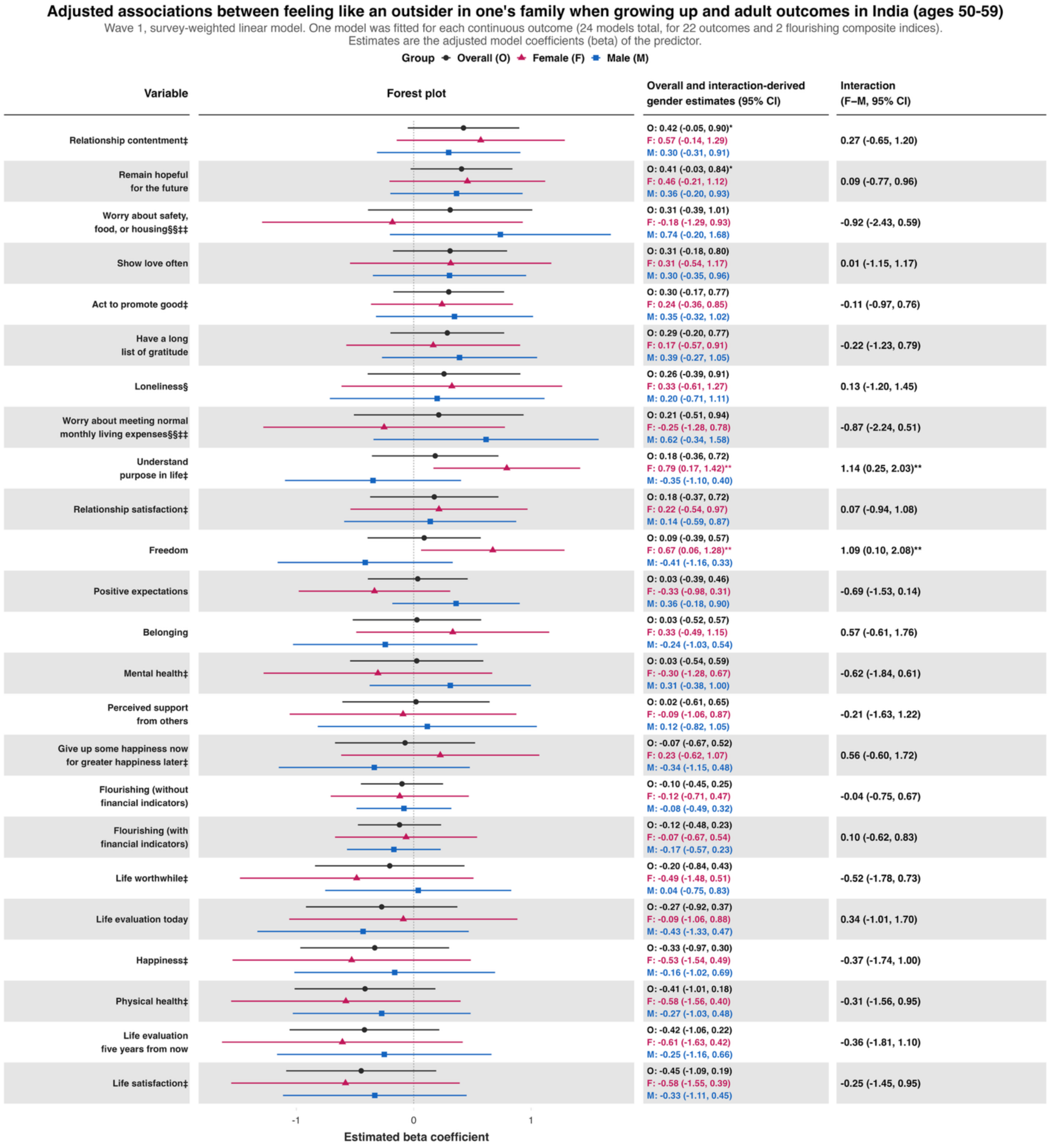


| Variable | Overall and interaction-derived gender estimates (95% CI) | Interaction (F–M, 95% CI) |
|---|---|---|
| Relationship contentment‡ | O: 0.42 (-0.05, 0.90)*<br>F: 0.57 (-0.14, 1.29)<br>M: 0.30 (-0.31, 0.91) | 0.27 (-0.65, 1.20) |
| Remain hopeful for the future | O: 0.41 (-0.03, 0.84)*<br>F: 0.46 (-0.21, 1.12)<br>M: 0.36 (-0.20, 0.93) | 0.09 (-0.77, 0.96) |
| Worry about safety, food, or housing§§‡‡ | O: 0.31 (-0.39, 1.01)<br>F: -0.18 (-1.29, 0.93)<br>M: 0.74 (-0.20, 1.68) | -0.92 (-2.43, 0.59) |
| Show love often | O: 0.31 (-0.18, 0.80)<br>F: 0.31 (-0.54, 1.17)<br>M: 0.30 (-0.35, 0.96) | 0.01 (-1.15, 1.17) |
| Act to promote good‡ | O: 0.30 (-0.17, 0.77)<br>F: 0.24 (-0.36, 0.85)<br>M: 0.35 (-0.32, 1.02) | -0.11 (-0.97, 0.76) |
| Have a long list of gratitude | O: 0.29 (-0.20, 0.77)<br>F: 0.17 (-0.57, 0.91)<br>M: 0.39 (-0.27, 1.05) | -0.22 (-1.23, 0.79) |
| Loneliness§ | O: 0.26 (-0.39, 0.91)<br>F: 0.33 (-0.61, 1.27)<br>M: 0.20 (-0.71, 1.11) | 0.13 (-1.20, 1.45) |
| Worry about meeting normal monthly living expenses§§‡‡ | O: 0.21 (-0.51, 0.94)<br>F: -0.25 (-1.28, 0.78)<br>M: 0.62 (-0.34, 1.58) | -0.87 (-2.24, 0.51) |
| Understand purpose in life‡ | O: 0.18 (-0.36, 0.72)<br>F: 0.79 (0.17, 1.42)**<br>M: -0.35 (-1.10, 0.40) | 1.14 (0.25, 2.03)** |
| Relationship satisfaction‡ | O: 0.18 (-0.37, 0.72)<br>F: 0.22 (-0.54, 0.97)<br>M: 0.14 (-0.59, 0.87) | 0.07 (-0.94, 1.08) |
| Freedom | O: 0.09 (-0.39, 0.57)<br>F: 0.67 (0.06, 1.28)**<br>M: -0.41 (-1.16, 0.33) | 1.09 (0.10, 2.08)** |
| Positive expectations | O: 0.03 (-0.39, 0.46)<br>F: -0.33 (-0.98, 0.31)<br>M: 0.36 (-0.18, 0.90) | -0.69 (-1.53, 0.14) |
| Belonging | O: 0.03 (-0.52, 0.57)<br>F: 0.33 (-0.49, 1.15)<br>M: -0.24 (-1.03, 0.54) | 0.57 (-0.61, 1.76) |
| Mental health‡ | O: 0.03 (-0.54, 0.59)<br>F: -0.30 (-1.28, 0.67)<br>M: 0.31 (-0.38, 1.00) | -0.62 (-1.84, 0.61) |
| Perceived support from others | O: 0.02 (-0.61, 0.65)<br>F: -0.09 (-1.06, 0.87)<br>M: 0.12 (-0.82, 1.05) | -0.21 (-1.63, 1.22) |
| Give up some happiness now for greater happiness later‡ | O: -0.07 (-0.67, 0.52)<br>F: 0.23 (-0.62, 1.07)<br>M: -0.34 (-1.15, 0.48) | 0.56 (-0.60, 1.72) |
| Flourishing (without financial indicators) | O: -0.10 (-0.45, 0.25)<br>F: -0.12 (-0.71, 0.47)<br>M: -0.08 (-0.49, 0.32) | -0.04 (-0.75, 0.67) |
| Flourishing (with financial indicators) | O: -0.12 (-0.48, 0.23)<br>F: -0.07 (-0.67, 0.54)<br>M: -0.17 (-0.57, 0.23) | 0.10 (-0.62, 0.83) |
| Life worthwhile‡ | O: -0.20 (-0.84, 0.43)<br>F: -0.49 (-1.48, 0.51)<br>M: 0.04 (-0.75, 0.83) | -0.52 (-1.78, 0.73) |
| Life evaluation today | O: -0.27 (-0.92, 0.37)<br>F: -0.09 (-1.06, 0.88)<br>M: -0.43 (-1.33, 0.47) | 0.34 (-1.01, 1.70) |
| Happiness‡ | O: -0.33 (-0.97, 0.30)<br>F: -0.53 (-1.54, 0.49)<br>M: -0.16 (-1.02, 0.69) | -0.37 (-1.74, 1.00) |
| Physical health‡ | O: -0.41 (-1.01, 0.18)<br>F: -0.58 (-1.56, 0.40)<br>M: -0.27 (-1.03, 0.48) | -0.31 (-1.56, 0.95) |
| Life evaluation five years from now | O: -0.42 (-1.06, 0.22)<br>F: -0.61 (-1.63, 0.42)<br>M: -0.25 (-1.16, 0.66) | -0.36 (-1.81, 1.10) |
| Life satisfaction‡ | O: -0.45 (-1.09, 0.19)<br>F: -0.58 (-1.55, 0.39)<br>M: -0.33 (-1.11, 0.45) | -0.25 (-1.45, 0.95) |

### *Feeling like an outsider in one's family when growing up – Ages 60+*

#### Adjusted associations between feeling like an outsider in one's family when growing up and adult outcomes in India (ages 60+)

Wave 1, survey-weighted linear model. One model was fitted for each continuous outcome (24 models total, for 22 outcomes and 2 flourishing composite indices).
Estimates are the adjusted model coefficients (beta) of the predictor.

Group ● Overall (O) ▲ Female (F) ■ Male (M)

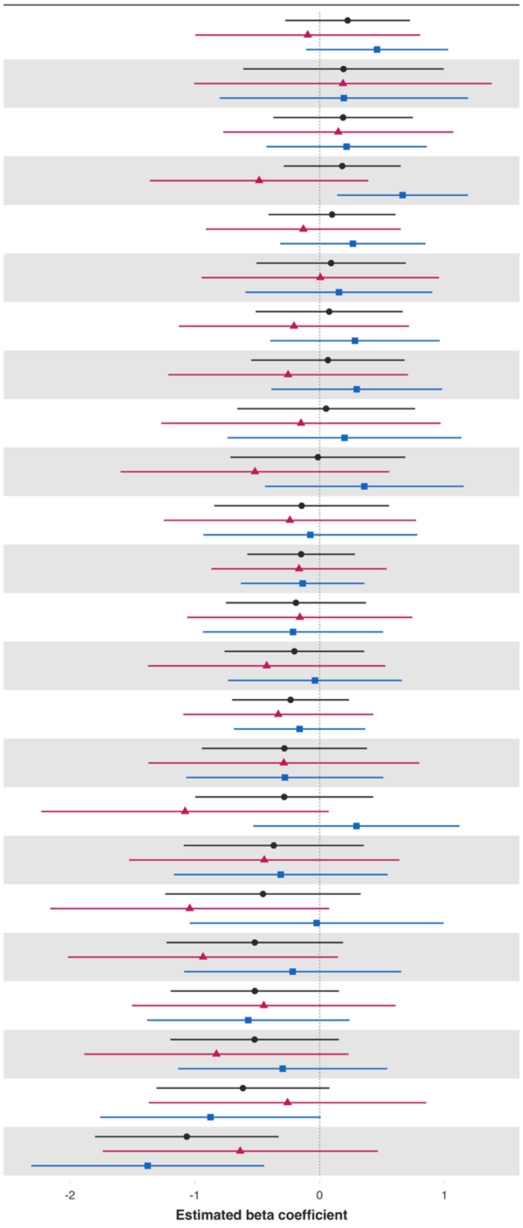


| Variable | Overall and interaction-derived gender estimates (95% CI) | Interaction (F–M, 95% CI) |
|---|---|---|
| Remain hopeful for the future | O: 0.22 (-0.28, 0.72)<br>F: -0.10 (-1.00, 0.80)<br>M: 0.46 (-0.11, 1.03) | -0.56 (-1.64, 0.53) |
| Loneliness§ | O: 0.19 (-0.61, 0.99)<br>F: 0.19 (-1.01, 1.38)<br>M: 0.19 (-0.80, 1.19) | -0.01 (-1.48, 1.46) |
| Understand purpose in life‡ | O: 0.19 (-0.37, 0.75)<br>F: 0.15 (-0.77, 1.07)<br>M: 0.22 (-0.43, 0.86) | -0.07 (-1.15, 1.02) |
| Belonging | O: 0.18 (-0.29, 0.65)<br>F: -0.48 (-1.36, 0.39)<br>M: 0.66 (0.14, 1.19)** | -1.15 (-2.20, -0.09)** |
| Act to promote good‡ | O: 0.10 (-0.41, 0.61)<br>F: -0.13 (-0.91, 0.65)<br>M: 0.27 (-0.32, 0.85) | -0.40 (-1.30, 0.50) |
| Relationship contentment‡ | O: 0.09 (-0.51, 0.69)<br>F: 0.01 (-0.95, 0.96)<br>M: 0.15 (-0.59, 0.90) | -0.15 (-1.35, 1.05) |
| Show love often | O: 0.08 (-0.51, 0.66)<br>F: -0.21 (-1.13, 0.72)<br>M: 0.28 (-0.40, 0.96) | -0.49 (-1.57, 0.59) |
| Freedom | O: 0.07 (-0.55, 0.68)<br>F: -0.25 (-1.21, 0.71)<br>M: 0.30 (-0.39, 0.98) | -0.55 (-1.64, 0.54) |
| Worry about safety, food, or housing§§‡‡ | O: 0.05 (-0.66, 0.76)<br>F: -0.15 (-1.27, 0.97)<br>M: 0.20 (-0.74, 1.14) | -0.35 (-1.82, 1.12) |
| Give up some happiness now for greater happiness later‡ | O: -0.01 (-0.71, 0.69)<br>F: -0.52 (-1.59, 0.56)<br>M: 0.36 (-0.44, 1.15) | -0.88 (-2.13, 0.38) |
| Life evaluation five years from now | O: -0.14 (-0.84, 0.56)<br>F: -0.24 (-1.25, 0.77)<br>M: -0.07 (-0.93, 0.78) | -0.16 (-1.39, 1.06) |
| Flourishing (with financial indicators) | O: -0.15 (-0.58, 0.28)<br>F: -0.17 (-0.87, 0.54)<br>M: -0.14 (-0.63, 0.36) | -0.03 (-0.85, 0.79) |
| Relationship satisfaction‡ | O: -0.19 (-0.75, 0.37)<br>F: -0.16 (-1.06, 0.74)<br>M: -0.21 (-0.93, 0.51) | 0.05 (-1.11, 1.21) |
| Positive expectations | O: -0.20 (-0.76, 0.36)<br>F: -0.42 (-1.37, 0.53)<br>M: -0.04 (-0.73, 0.66) | -0.39 (-1.59, 0.82) |
| Flourishing (without financial indicators) | O: -0.23 (-0.70, 0.23)<br>F: -0.33 (-1.09, 0.43)<br>M: -0.16 (-0.69, 0.37) | -0.17 (-1.05, 0.71) |
| Physical health‡ | O: -0.28 (-0.94, 0.38)<br>F: -0.29 (-1.37, 0.80)<br>M: -0.28 (-1.07, 0.51) | -0.01 (-1.32, 1.30) |
| Life worthwhile‡ | O: -0.28 (-1.00, 0.43)<br>F: -1.08 (-2.23, 0.07)*<br>M: 0.29 (-0.53, 1.12) | -1.37 (-2.77, 0.02)* |
| Mental health‡ | O: -0.37 (-1.09, 0.35)<br>F: -0.44 (-1.53, 0.64)<br>M: -0.31 (-1.17, 0.54) | -0.13 (-1.41, 1.15) |
| Perceived support from others | O: -0.45 (-1.24, 0.33)<br>F: -1.04 (-2.16, 0.08)*<br>M: -0.02 (-1.04, 0.99) | -1.02 (-2.48, 0.45) |
| Worry about meeting normal monthly living expenses§§‡‡ | O: -0.52 (-1.23, 0.19)<br>F: -0.93 (-2.02, 0.15)*<br>M: -0.22 (-1.08, 0.65) | -0.72 (-2.05, 0.61) |
| Life satisfaction‡ | O: -0.52 (-1.19, 0.15)<br>F: -0.45 (-1.50, 0.61)<br>M: -0.57 (-1.38, 0.24) | 0.12 (-1.15, 1.39) |
| Have a long list of gratitude | O: -0.52 (-1.20, 0.15)<br>F: -0.83 (-1.89, 0.23)<br>M: -0.30 (-1.13, 0.54) | -0.53 (-1.86, 0.80) |
| Life evaluation today | O: -0.61 (-1.31, 0.08)*<br>F: -0.26 (-1.37, 0.85)<br>M: -0.87 (-1.76, 0.01)* | 0.62 (-0.80, 2.04) |
| Happiness‡ | O: -1.06 (-1.80, -0.33)**<br>F: -0.64 (-1.74, 0.47)<br>M: -1.38 (-2.31, -0.44)** | 0.74 (-0.65, 2.13) |

**Predictor:** *Not feeling loved by mother*

***Not feeling loved by mother – Ages 18 to 24 inclusive***

**Adjusted associations between not feeling loved by mother when growing up and adult outcomes in India (ages 18-24)**

Wave 1, survey-weighted linear model. One model was fitted for each continuous outcome (24 models total, for 22 outcomes and 2 flourishing composite indices). Estimates are the adjusted model coefficients (beta) of the predictor.

Group ◆ Overall (O) ▲ Female (F) ■ Male (M)

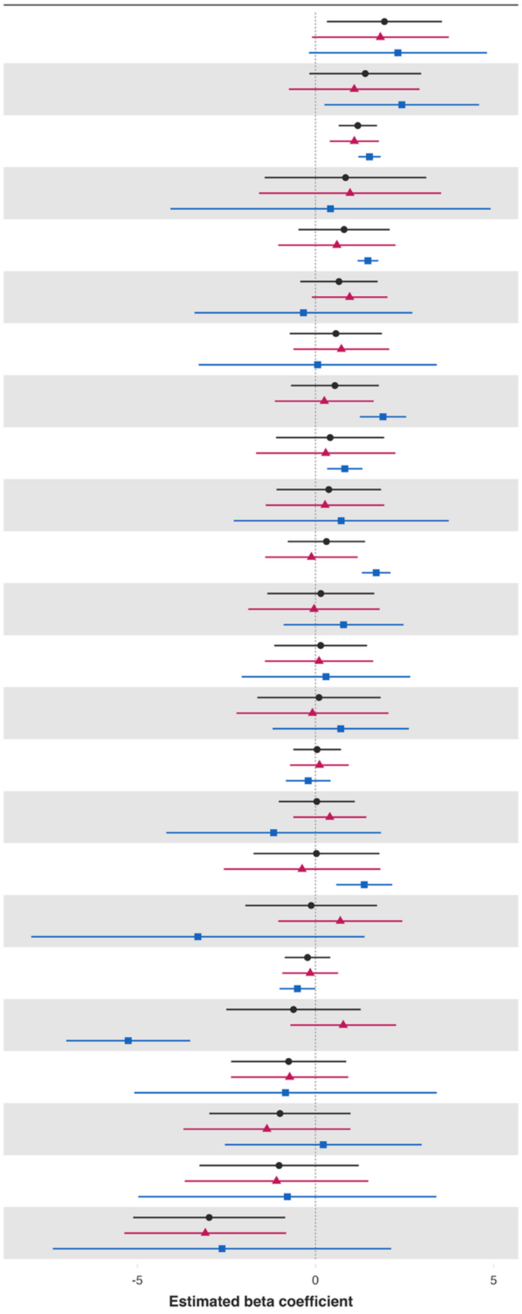


| Variable | Overall and interaction-derived gender estimates (95% CI) | Interaction (F–M, 95% CI) |
|---|---|---|
| Worry about meeting normal monthly living expenses§§‡‡ | O: 1.94 (0.32, 3.55)**<br>F: 1.82 (-0.10, 3.74)*<br>M: 2.31 (-0.19, 4.81)* | -0.49 (-3.63, 2.64) |
| Worry about safety, food, or housing§§‡‡ | O: 1.40 (-0.17, 2.96)*<br>F: 1.09 (-0.75, 2.92)<br>M: 2.42 (0.25, 4.59)** | -1.34 (-4.14, 1.47) |
| Remain hopeful for the future | O: 1.19 (0.65, 1.73)***<br>F: 1.09 (0.40, 1.78)**<br>M: 1.51 (1.20, 1.83)*** | -0.42 (-1.17, 0.32) |
| Loneliness§ | O: 0.84 (-1.42, 3.11)<br>F: 0.97 (-1.58, 3.53)<br>M: 0.42 (-4.07, 4.92) | 0.55 (-4.56, 5.66) |
| Freedom | O: 0.80 (-0.48, 2.08)<br>F: 0.60 (-1.05, 2.25)<br>M: 1.47 (1.18, 1.76)*** | -0.87 (-2.55, 0.80) |
| Give up some happiness now for greater happiness later‡ | O: 0.66 (-0.43, 1.74)<br>F: 0.96 (-0.10, 2.02)*<br>M: -0.34 (-3.39, 2.72) | 1.30 (-1.93, 4.52) |
| Life satisfaction‡ | O: 0.57 (-0.72, 1.86)<br>F: 0.73 (-0.62, 2.07)<br>M: 0.06 (-3.28, 3.40) | 0.66 (-2.93, 4.26) |
| Life evaluation five years from now | O: 0.55 (-0.69, 1.78)<br>F: 0.25 (-1.14, 1.63)<br>M: 1.90 (1.25, 2.55)*** | -1.65 (-3.16, -0.14)** |
| Belonging | O: 0.41 (-1.11, 1.93)<br>F: 0.29 (-1.67, 2.24)<br>M: 0.82 (0.33, 1.32)** | -0.53 (-2.57, 1.50) |
| Have a long list of gratitude | O: 0.37 (-1.09, 1.84)<br>F: 0.27 (-1.40, 1.93)<br>M: 0.72 (-2.29, 3.74) | -0.45 (-3.90, 2.99) |
| Understand purpose in life‡ | O: 0.31 (-0.78, 1.39)<br>F: -0.11 (-1.41, 1.18)<br>M: 1.70 (1.30, 2.11)*** | -1.82 (-3.16, -0.48)** |
| Positive expectations | O: 0.15 (-1.35, 1.65)<br>F: -0.04 (-1.89, 1.80)<br>M: 0.79 (-0.89, 2.47) | -0.83 (-3.33, 1.67) |
| Mental health‡ | O: 0.15 (-1.16, 1.45)<br>F: 0.10 (-1.42, 1.62)<br>M: 0.30 (-2.07, 2.66) | -0.19 (-2.98, 2.59) |
| Relationship contentment‡ | O: 0.10 (-1.63, 1.83)<br>F: -0.08 (-2.22, 2.05)<br>M: 0.71 (-1.20, 2.62) | -0.79 (-3.64, 2.05) |
| Flourishing (without financial indicators) | O: 0.05 (-0.62, 0.72)<br>F: 0.11 (-0.71, 0.93)<br>M: -0.20 (-0.83, 0.42) | 0.31 (-0.71, 1.34) |
| Act to promote good‡ | O: 0.04 (-1.03, 1.10)<br>F: 0.40 (-0.62, 1.43)<br>M: -1.17 (-4.19, 1.84) | 1.58 (-1.61, 4.76) |
| Show love often | O: 0.03 (-1.74, 1.80)<br>F: -0.38 (-2.57, 1.82)<br>M: 1.37 (0.58, 2.15)*** | -1.74 (-4.07, 0.58) |
| Life worthwhile‡ | O: -0.12 (-1.97, 1.73)<br>F: 0.70 (-1.05, 2.44)<br>M: -3.30 (-7.98, 1.38) | 4.00 (-1.03, 9.02) |
| Flourishing (with financial indicators) | O: -0.22 (-0.86, 0.42)<br>F: -0.15 (-0.93, 0.63)<br>M: -0.51 (-1.01, -0.01)** | 0.36 (-0.57, 1.29) |
| Happiness‡ | O: -0.62 (-2.50, 1.27)<br>F: 0.78 (-0.70, 2.26)<br>M: -5.25 (-6.99, -3.51)*** | 6.03 (3.73, 8.34)*** |
| Physical health‡ | O: -0.75 (-2.37, 0.86)<br>F: -0.73 (-2.37, 0.92)<br>M: -0.84 (-5.09, 3.40) | 0.12 (-4.41, 4.64) |
| Relationship satisfaction‡ | O: -1.00 (-2.98, 0.98)<br>F: -1.36 (-3.71, 0.98)<br>M: 0.22 (-2.54, 2.98) | -1.58 (-5.18, 2.02) |
| Life evaluation today | O: -1.02 (-3.25, 1.21)<br>F: -1.09 (-3.67, 1.48)<br>M: -0.79 (-4.97, 3.39) | -0.30 (-5.16, 4.55) |
| Perceived support from others | O: -2.98 (-5.12, -0.85)**<br>F: -3.09 (-5.36, -0.82)**<br>M: -2.62 (-7.37, 2.12) | -0.47 (-5.63, 4.69) |

*Not feeling loved by mother – Ages 25 to 29 inclusive*

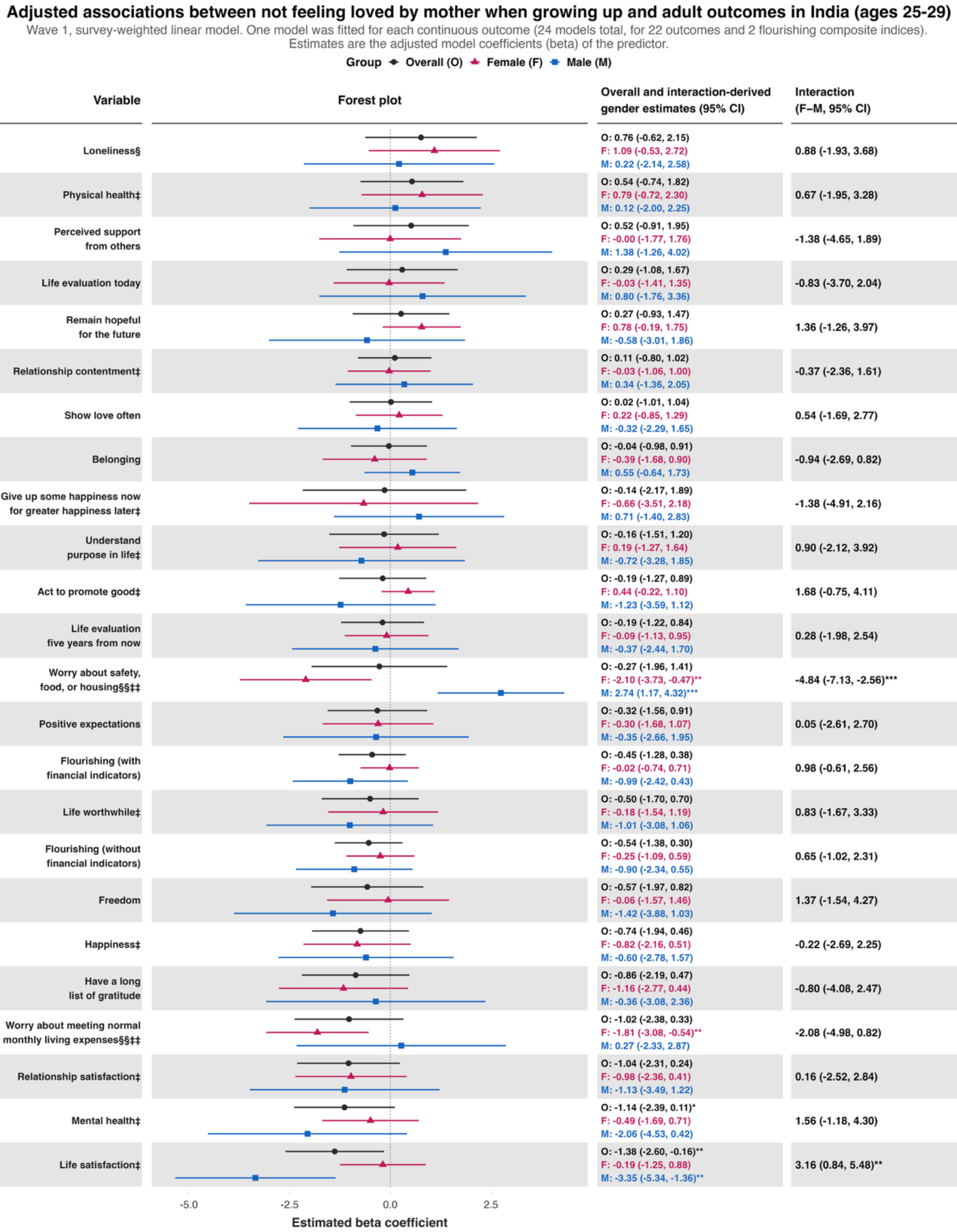


| Variable | Overall and interaction-derived gender estimates (95% CI) | Interaction (F–M, 95% CI) |
|---|---|---|
| Loneliness§ | O: 0.76 (-0.62, 2.15)<br>F: 1.09 (-0.53, 2.72)<br>M: 0.22 (-2.14, 2.58) | 0.88 (-1.93, 3.68) |
| Physical health‡ | O: 0.54 (-0.74, 1.82)<br>F: 0.79 (-0.72, 2.30)<br>M: 0.12 (-2.00, 2.25) | 0.67 (-1.95, 3.28) |
| Perceived support from others | O: 0.52 (-0.91, 1.95)<br>F: -0.00 (-1.77, 1.76)<br>M: 1.38 (-1.26, 4.02) | -1.38 (-4.65, 1.89) |
| Life evaluation today | O: 0.29 (-1.08, 1.67)<br>F: -0.03 (-1.41, 1.35)<br>M: 0.80 (-1.76, 3.36) | -0.83 (-3.70, 2.04) |
| Remain hopeful for the future | O: 0.27 (-0.93, 1.47)<br>F: 0.78 (-0.19, 1.75)<br>M: -0.58 (-3.01, 1.86) | 1.36 (-1.26, 3.97) |
| Relationship contentment‡ | O: 0.11 (-0.80, 1.02)<br>F: -0.03 (-1.06, 1.00)<br>M: 0.34 (-1.36, 2.05) | -0.37 (-2.36, 1.61) |
| Show love often | O: 0.02 (-1.01, 1.04)<br>F: 0.22 (-0.85, 1.29)<br>M: -0.32 (-2.29, 1.65) | 0.54 (-1.69, 2.77) |
| Belonging | O: -0.04 (-0.98, 0.91)<br>F: -0.39 (-1.68, 0.90)<br>M: 0.55 (-0.64, 1.73) | -0.94 (-2.69, 0.82) |
| Give up some happiness now for greater happiness later‡ | O: -0.14 (-2.17, 1.89)<br>F: -0.66 (-3.51, 2.18)<br>M: 0.71 (-1.40, 2.83) | -1.38 (-4.91, 2.16) |
| Understand purpose in life‡ | O: -0.16 (-1.51, 1.20)<br>F: 0.19 (-1.27, 1.64)<br>M: -0.72 (-3.28, 1.85) | 0.90 (-2.12, 3.92) |
| Act to promote good‡ | O: -0.19 (-1.27, 0.89)<br>F: 0.44 (-0.22, 1.10)<br>M: -1.23 (-3.59, 1.12) | 1.68 (-0.75, 4.11) |
| Life evaluation five years from now | O: -0.19 (-1.22, 0.84)<br>F: -0.09 (-1.13, 0.95)<br>M: -0.37 (-2.44, 1.70) | 0.28 (-1.98, 2.54) |
| Worry about safety, food, or housing§§‡‡ | O: -0.27 (-1.96, 1.41)<br>F: -2.10 (-3.73, -0.47)**<br>M: 2.74 (1.17, 4.32)*** | -4.84 (-7.13, -2.56)*** |
| Positive expectations | O: -0.32 (-1.56, 0.91)<br>F: -0.30 (-1.68, 1.07)<br>M: -0.35 (-2.66, 1.95) | 0.05 (-2.61, 2.70) |
| Flourishing (with financial indicators) | O: -0.45 (-1.28, 0.38)<br>F: -0.02 (-0.74, 0.71)<br>M: -0.99 (-2.42, 0.43) | 0.98 (-0.61, 2.56) |
| Life worthwhile‡ | O: -0.50 (-1.70, 0.70)<br>F: -0.18 (-1.54, 1.19)<br>M: -1.01 (-3.08, 1.06) | 0.83 (-1.67, 3.33) |
| Flourishing (without financial indicators) | O: -0.54 (-1.38, 0.30)<br>F: -0.25 (-1.09, 0.59)<br>M: -0.90 (-2.34, 0.55) | 0.65 (-1.02, 2.31) |
| Freedom | O: -0.57 (-1.97, 0.82)<br>F: -0.06 (-1.57, 1.46)<br>M: -1.42 (-3.88, 1.03) | 1.37 (-1.54, 4.27) |
| Happiness‡ | O: -0.74 (-1.94, 0.46)<br>F: -0.82 (-2.16, 0.51)<br>M: -0.60 (-2.78, 1.57) | -0.22 (-2.69, 2.25) |
| Have a long list of gratitude | O: -0.86 (-2.19, 0.47)<br>F: -1.16 (-2.77, 0.44)<br>M: -0.36 (-3.08, 2.36) | -0.80 (-4.08, 2.47) |
| Worry about meeting normal monthly living expenses§§‡‡ | O: -1.02 (-2.38, 0.33)<br>F: -1.81 (-3.08, -0.54)**<br>M: 0.27 (-2.33, 2.87) | -2.08 (-4.98, 0.82) |
| Relationship satisfaction‡ | O: -1.04 (-2.31, 0.24)<br>F: -0.98 (-2.36, 0.41)<br>M: -1.13 (-3.49, 1.22) | 0.16 (-2.52, 2.84) |
| Mental health‡ | O: -1.14 (-2.39, 0.11)*<br>F: -0.49 (-1.69, 0.71)<br>M: -2.06 (-4.53, 0.42) | 1.56 (-1.18, 4.30) |
| Life satisfaction‡ | O: -1.38 (-2.60, -0.16)**<br>F: -0.19 (-1.25, 0.88)<br>M: -3.35 (-5.34, -1.36)** | 3.16 (0.84, 5.48)** |

*Not feeling loved by mother – Ages 30 to 39 inclusive*

**Adjusted associations between not feeling loved by mother when growing up and adult outcomes in India (ages 30-39)**

Wave 1, survey-weighted linear model. One model was fitted for each continuous outcome (24 models total, for 22 outcomes and 2 flourishing composite indices). Estimates are the adjusted model coefficients (beta) of the predictor.

Group: Overall (O), Female (F), Male (M)

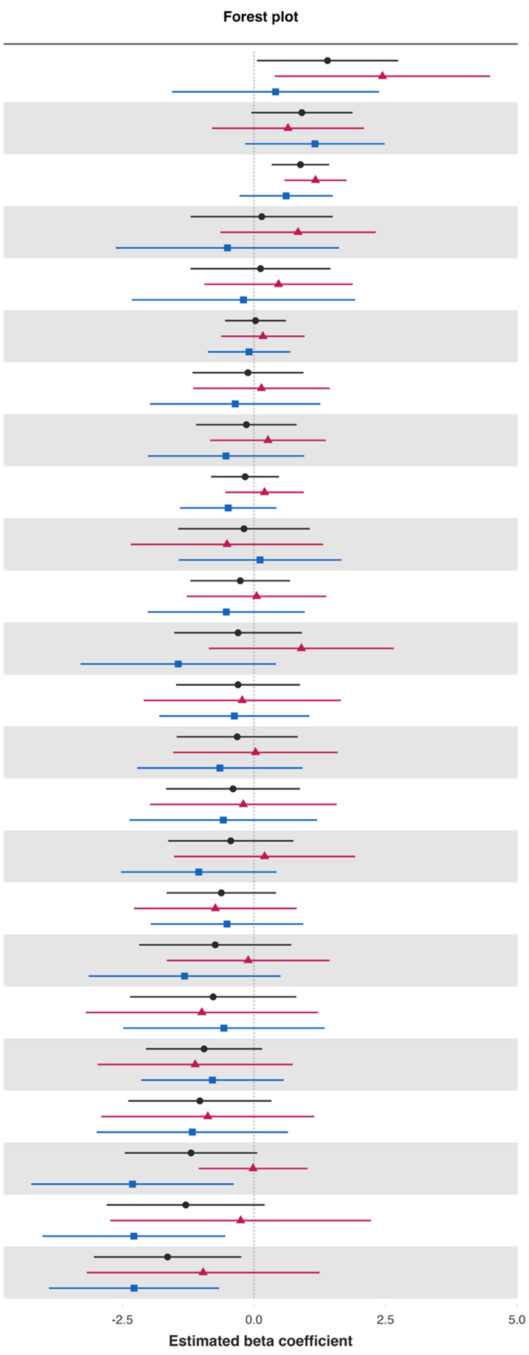


| Variable | Overall and interaction-derived gender estimates (95% CI) | Interaction (F–M, 95% CI) |
|---|---|---|
| Loneliness§ | O: 1.40 (0.06, 2.74)**<br>F: 2.44 (0.40, 4.49)**<br>M: 0.41 (-1.56, 2.38) | 2.03 (-0.93, 4.99) |
| Happiness‡ | O: 0.91 (-0.05, 1.87)*<br>F: 0.65 (-0.80, 2.09)<br>M: 1.16 (-0.17, 2.49)* | -0.51 (-2.50, 1.47) |
| Show love often | O: 0.88 (0.34, 1.43)**<br>F: 1.17 (0.58, 1.76)***<br>M: 0.61 (-0.27, 1.50) | 0.56 (-0.49, 1.61) |
| Life satisfaction‡ | O: 0.15 (-1.20, 1.50)<br>F: 0.84 (-0.64, 2.31)<br>M: -0.50 (-2.63, 1.62) | 1.34 (-1.28, 3.96) |
| Life worthwhile‡ | O: 0.12 (-1.20, 1.45)<br>F: 0.47 (-0.94, 1.88)<br>M: -0.20 (-2.32, 1.92) | 0.67 (-1.84, 3.17) |
| Flourishing (with financial indicators) | O: 0.03 (-0.55, 0.61)<br>F: 0.17 (-0.62, 0.96)<br>M: -0.09 (-0.88, 0.69) | 0.26 (-0.83, 1.35) |
| Remain hopeful for the future | O: -0.11 (-1.17, 0.94)<br>F: 0.14 (-1.16, 1.44)<br>M: -0.36 (-1.97, 1.26) | 0.50 (-1.56, 2.56) |
| Understand purpose in life‡ | O: -0.14 (-1.10, 0.81)<br>F: 0.27 (-0.83, 1.36)<br>M: -0.53 (-2.02, 0.96) | 0.80 (-1.01, 2.61) |
| Flourishing (without financial indicators) | O: -0.17 (-0.81, 0.48)<br>F: 0.20 (-0.54, 0.94)<br>M: -0.49 (-1.40, 0.43) | 0.69 (-0.44, 1.82) |
| Relationship contentment‡ | O: -0.19 (-1.44, 1.06)<br>F: -0.51 (-2.34, 1.32)<br>M: 0.12 (-1.43, 1.66) | -0.63 (-2.93, 1.67) |
| Life evaluation five years from now | O: -0.26 (-1.21, 0.69)<br>F: 0.05 (-1.28, 1.38)<br>M: -0.52 (-2.02, 0.97) | 0.58 (-1.57, 2.72) |
| Worry about meeting normal monthly living expenses§§‡‡ | O: -0.30 (-1.51, 0.91)<br>F: 0.90 (-0.86, 2.66)<br>M: -1.44 (-3.29, 0.42) | 2.34 (-0.31, 4.99)* |
| Relationship satisfaction‡ | O: -0.30 (-1.48, 0.87)<br>F: -0.22 (-2.10, 1.65)<br>M: -0.37 (-1.80, 1.06) | 0.15 (-2.17, 2.47) |
| Mental health‡ | O: -0.32 (-1.47, 0.83)<br>F: 0.03 (-1.54, 1.60)<br>M: -0.65 (-2.22, 0.92) | 0.68 (-1.50, 2.86) |
| Positive expectations | O: -0.40 (-1.67, 0.87)<br>F: -0.20 (-1.97, 1.57)<br>M: -0.58 (-2.36, 1.20) | 0.38 (-2.11, 2.87) |
| Physical health‡ | O: -0.44 (-1.63, 0.75)<br>F: 0.20 (-1.52, 1.92)<br>M: -1.05 (-2.53, 0.43) | 1.25 (-0.94, 3.44) |
| Life evaluation today | O: -0.62 (-1.66, 0.42)<br>F: -0.73 (-2.28, 0.81)<br>M: -0.51 (-1.96, 0.94) | -0.22 (-2.37, 1.93) |
| Have a long list of gratitude | O: -0.74 (-2.18, 0.71)<br>F: -0.11 (-1.66, 1.44)<br>M: -1.32 (-3.14, 0.50) | 1.21 (-0.63, 3.05) |
| Give up some happiness now for greater happiness later‡ | O: -0.77 (-2.35, 0.80)<br>F: -0.99 (-3.20, 1.22)<br>M: -0.57 (-2.48, 1.34) | -0.42 (-3.09, 2.26) |
| Belonging | O: -0.95 (-2.05, 0.16)*<br>F: -1.12 (-2.97, 0.74)<br>M: -0.79 (-2.14, 0.57) | -0.33 (-2.69, 2.03) |
| Freedom | O: -1.03 (-2.39, 0.33)<br>F: -0.88 (-2.90, 1.15)<br>M: -1.17 (-2.99, 0.65) | 0.30 (-2.43, 3.02) |
| Act to promote good‡ | O: -1.19 (-2.45, 0.06)*<br>F: -0.02 (-1.05, 1.02)<br>M: -2.31 (-4.23, -0.38)** | 2.29 (0.16, 4.42)** |
| Worry about safety, food, or housing§§‡‡ | O: -1.30 (-2.80, 0.21)*<br>F: -0.25 (-2.73, 2.23)<br>M: -2.28 (-4.02, -0.55)** | 2.03 (-1.03, 5.09) |
| Perceived support from others | O: -1.64 (-3.04, -0.24)**<br>F: -0.96 (-3.18, 1.25)<br>M: -2.28 (-3.90, -0.66)** | 1.31 (-1.38, 4.01) |

*Not feeling loved by mother – Ages 40 to 49 inclusive*

**Adjusted associations between not feeling loved by mother when growing up and adult outcomes in India (ages 40-49)**

Wave 1, survey-weighted linear model. One model was fitted for each continuous outcome (24 models total, for 22 outcomes and 2 flourishing composite indices).
Estimates are the adjusted model coefficients (beta) of the predictor.

Group ◆ Overall (O) ▲ Female (F) ■ Male (M)

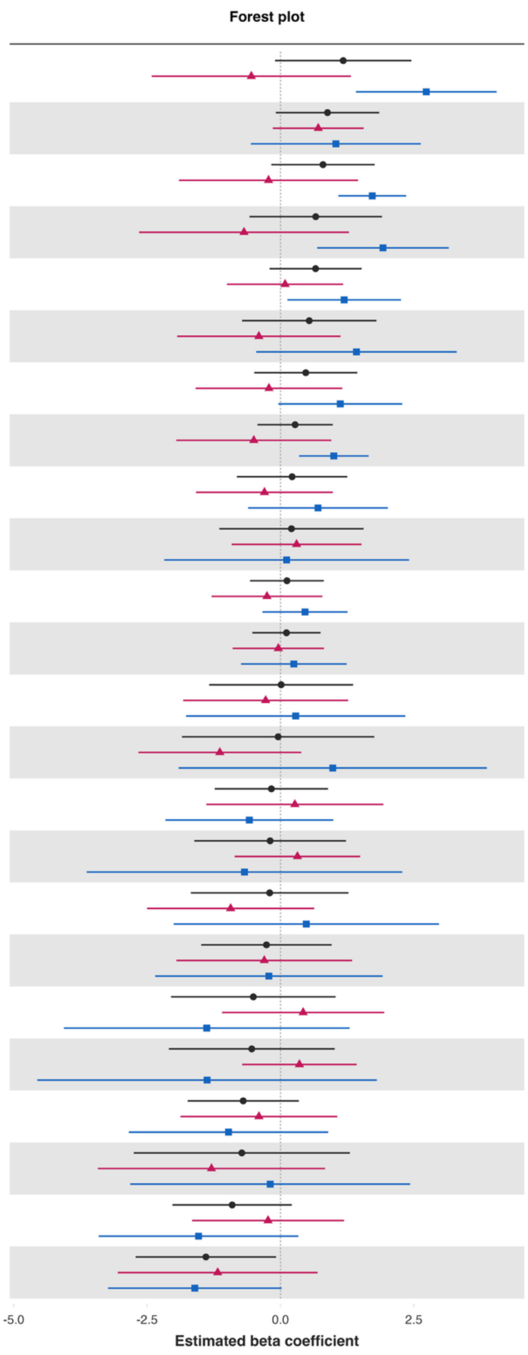


| Variable | Overall and interaction-derived gender estimates (95% CI) | Interaction (F–M, 95% CI) |
|---|---|---|
| Happiness‡ | O: 1.18 (-0.10, 2.46)*<br>F: -0.55 (-2.41, 1.32)<br>M: 2.73 (1.42, 4.05)*** | -3.28 (-5.50, -1.06)** |
| Have a long list of gratitude | O: 0.88 (-0.09, 1.85)*<br>F: 0.71 (-0.14, 1.56)<br>M: 1.04 (-0.55, 2.63) | -0.33 (-2.09, 1.43) |
| Understand purpose in life‡ | O: 0.80 (-0.17, 1.77)<br>F: -0.22 (-1.90, 1.46)<br>M: 1.72 (1.09, 2.36)*** | -1.94 (-3.73, -0.16)** |
| Life satisfaction‡ | O: 0.66 (-0.58, 1.90)<br>F: -0.68 (-2.65, 1.28)<br>M: 1.92 (0.69, 3.16)** | -2.61 (-4.86, -0.35)** |
| Positive expectations | O: 0.66 (-0.20, 1.52)<br>F: 0.09 (-1.00, 1.18)<br>M: 1.19 (0.13, 2.26)** | -1.11 (-2.52, 0.31) |
| Give up some happiness now for greater happiness later‡ | O: 0.54 (-0.72, 1.80)<br>F: -0.40 (-1.94, 1.13)<br>M: 1.43 (-0.45, 3.31) | -1.83 (-4.35, 0.69) |
| Belonging | O: 0.47 (-0.49, 1.44)<br>F: -0.22 (-1.59, 1.16)<br>M: 1.12 (-0.04, 2.28)* | -1.34 (-3.11, 0.44) |
| Remain hopeful for the future | O: 0.28 (-0.43, 0.98)<br>F: -0.50 (-1.95, 0.95)<br>M: 1.00 (0.35, 1.65)** | -1.50 (-3.17, 0.17)* |
| Act to promote good‡ | O: 0.22 (-0.82, 1.25)<br>F: -0.30 (-1.58, 0.98)<br>M: 0.70 (-0.61, 2.01) | -1.00 (-2.71, 0.70) |
| Relationship contentment‡ | O: 0.21 (-1.15, 1.56)<br>F: 0.30 (-0.91, 1.52)<br>M: 0.12 (-2.18, 2.41) | 0.19 (-2.33, 2.71) |
| Flourishing (without financial indicators) | O: 0.12 (-0.57, 0.81)<br>F: -0.25 (-1.29, 0.79)<br>M: 0.46 (-0.34, 1.26) | -0.71 (-1.99, 0.56) |
| Flourishing (with financial indicators) | O: 0.11 (-0.53, 0.75)<br>F: -0.04 (-0.90, 0.82)<br>M: 0.25 (-0.74, 1.24) | -0.29 (-1.66, 1.07) |
| Mental health‡ | O: 0.01 (-1.34, 1.36)<br>F: -0.28 (-1.82, 1.27)<br>M: 0.29 (-1.77, 2.34) | -0.56 (-2.99, 1.87) |
| Worry about safety, food, or housing§§‡‡ | O: -0.04 (-1.85, 1.76)<br>F: -1.14 (-2.66, 0.39)<br>M: 0.98 (-1.91, 3.87) | -2.12 (-5.22, 0.98) |
| Life evaluation today | O: -0.17 (-1.23, 0.89)<br>F: 0.27 (-1.39, 1.93)<br>M: -0.58 (-2.16, 0.99) | 0.85 (-1.50, 3.21) |
| Freedom | O: -0.19 (-1.62, 1.23)<br>F: 0.32 (-0.86, 1.50)<br>M: -0.67 (-3.63, 2.28) | 0.99 (-2.46, 4.44) |
| Worry about meeting normal monthly living expenses§§‡‡ | O: -0.20 (-1.68, 1.28)<br>F: -0.93 (-2.50, 0.63)<br>M: 0.48 (-2.00, 2.97) | -1.42 (-4.35, 1.52) |
| Life evaluation five years from now | O: -0.26 (-1.49, 0.96)<br>F: -0.30 (-1.95, 1.34)<br>M: -0.22 (-2.35, 1.91) | -0.09 (-2.99, 2.82) |
| Life worthwhile‡ | O: -0.51 (-2.05, 1.04)<br>F: 0.43 (-1.09, 1.95)<br>M: -1.38 (-4.06, 1.30) | 1.81 (-1.46, 5.08) |
| Show love often | O: -0.54 (-2.09, 1.02)<br>F: 0.36 (-0.72, 1.43)<br>M: -1.38 (-4.56, 1.81) | 1.73 (-1.80, 5.26) |
| Physical health‡ | O: -0.70 (-1.74, 0.34)<br>F: -0.40 (-1.87, 1.07)<br>M: -0.97 (-2.84, 0.89) | 0.57 (-2.05, 3.19) |
| Perceived support from others | O: -0.72 (-2.75, 1.30)<br>F: -1.29 (-3.42, 0.84)<br>M: -0.19 (-2.82, 2.43) | -1.10 (-3.78, 1.58) |
| Relationship satisfaction‡ | O: -0.91 (-2.02, 0.21)<br>F: -0.23 (-1.66, 1.19)<br>M: -1.54 (-3.41, 0.34) | 1.30 (-1.14, 3.75) |
| Loneliness§ | O: -1.40 (-2.71, -0.08)**<br>F: -1.18 (-3.05, 0.70)<br>M: -1.61 (-3.23, 0.02)* | 0.43 (-1.89, 2.75) |

*Not feeling loved by mother – Ages 50 to 59 inclusive*

**Adjusted associations between not feeling loved by mother when growing up and adult outcomes in India (ages 50-59)**

Wave 1, survey-weighted linear model. One model was fitted for each continuous outcome (24 models total, for 22 outcomes and 2 flourishing composite indices).
Estimates are the adjusted model coefficients (beta) of the predictor.

Group ◆ Overall (O) ▲ Female (F) ■ Male (M)

Forest plot

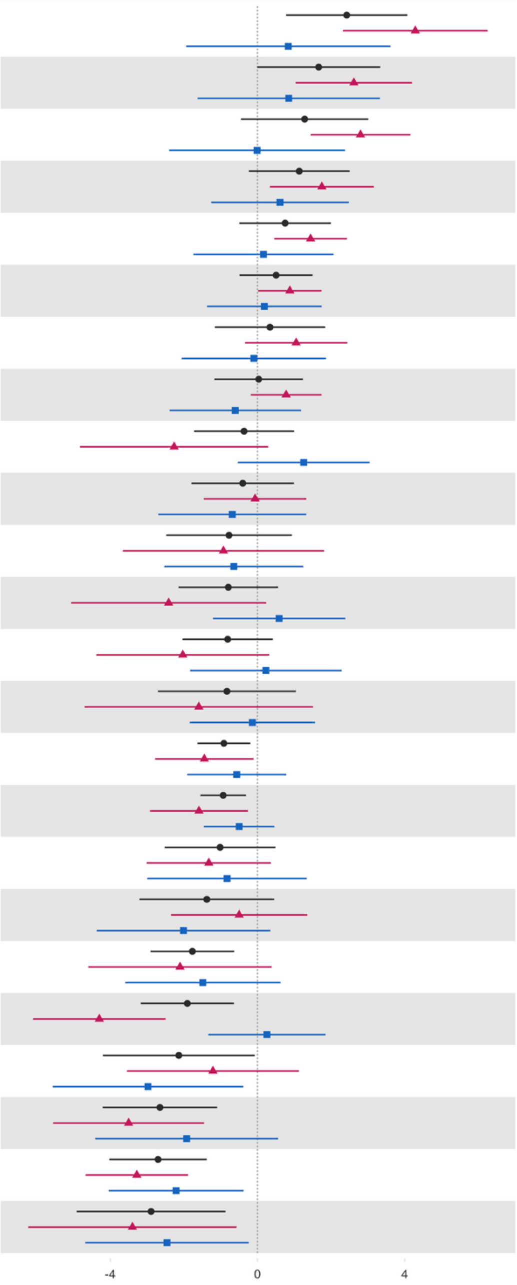

| Variable | Overall and interaction-derived gender estimates (95% CI) | Interaction (F–M, 95% CI) |
|---|---|---|
| Loneliness§ | O: 2.42 (0.78, 4.07)**<br>F: 4.29 (2.32, 6.25)***<br>M: 0.84 (-1.94, 3.61) | 3.45 (-0.22, 7.13)* |
| Worry about safety, food, or housing§§‡‡ | O: 1.66 (-0.01, 3.34)*<br>F: 2.62 (1.04, 4.20)**<br>M: 0.85 (-1.63, 3.33) | 1.77 (-0.97, 4.51) |
| Worry about meeting normal monthly living expenses§§‡‡ | O: 1.28 (-0.45, 3.01)<br>F: 2.80 (1.45, 4.15)***<br>M: -0.01 (-2.40, 2.38) | 2.81 (0.46, 5.17)** |
| Give up some happiness now for greater happiness later‡ | O: 1.14 (-0.23, 2.51)<br>F: 1.75 (0.34, 3.16)**<br>M: 0.61 (-1.26, 2.48) | 1.14 (-0.94, 3.21) |
| Have a long list of gratitude | O: 0.75 (-0.49, 2.00)<br>F: 1.44 (0.45, 2.43)**<br>M: 0.16 (-1.74, 2.07) | 1.28 (-0.75, 3.31) |
| Act to promote good‡ | O: 0.51 (-0.49, 1.50)<br>F: 0.88 (0.02, 1.74)**<br>M: 0.19 (-1.37, 1.74) | 0.69 (-1.00, 2.38) |
| Understand purpose in life‡ | O: 0.34 (-1.16, 1.84)<br>F: 1.05 (-0.34, 2.44)<br>M: -0.10 (-2.06, 1.86) | 1.15 (-0.99, 3.30) |
| Remain hopeful for the future | O: 0.03 (-1.17, 1.24)<br>F: 0.78 (-0.19, 1.74)<br>M: -0.60 (-2.39, 1.18) | 1.38 (-0.49, 3.25) |
| Life satisfaction‡ | O: -0.36 (-1.72, 0.99)<br>F: -2.26 (-4.82, 0.29)*<br>M: 1.26 (-0.53, 3.05) | -3.52 (-6.96, -0.08)** |
| Freedom | O: -0.40 (-1.79, 0.99)<br>F: -0.07 (-1.46, 1.32)<br>M: -0.69 (-2.70, 1.32) | 0.62 (-1.54, 2.78) |
| Belonging | O: -0.77 (-2.48, 0.94)<br>F: -0.93 (-3.66, 1.81)<br>M: -0.64 (-2.53, 1.24) | -0.28 (-3.43, 2.86) |
| Physical health‡ | O: -0.79 (-2.14, 0.56)<br>F: -2.41 (-5.06, 0.24)*<br>M: 0.59 (-1.21, 2.39) | -3.00 (-6.52, 0.51)* |
| Happiness‡ | O: -0.81 (-2.04, 0.42)<br>F: -2.03 (-4.38, 0.32)*<br>M: 0.23 (-1.83, 2.29) | -2.26 (-5.89, 1.37) |
| Positive expectations | O: -0.83 (-2.71, 1.05)<br>F: -1.59 (-4.70, 1.51)<br>M: -0.14 (-1.84, 1.56) | -1.46 (-4.74, 1.83) |
| Flourishing (without financial indicators) | O: -0.91 (-1.63, -0.20)**<br>F: -1.44 (-2.78, -0.10)**<br>M: -0.56 (-1.91, 0.78) | -0.88 (-3.12, 1.36) |
| Flourishing (with financial indicators) | O: -0.93 (-1.55, -0.31)**<br>F: -1.59 (-2.92, -0.26)**<br>M: -0.50 (-1.46, 0.46) | -1.09 (-2.94, 0.76) |
| Life evaluation five years from now | O: -1.02 (-2.52, 0.49)<br>F: -1.32 (-3.01, 0.36)<br>M: -0.83 (-3.00, 1.34) | -0.49 (-3.23, 2.24) |
| Show love often | O: -1.38 (-3.21, 0.45)<br>F: -0.50 (-2.35, 1.35)<br>M: -2.01 (-4.37, 0.35)* | 1.51 (-0.93, 3.95) |
| Relationship contentment‡ | O: -1.77 (-2.91, -0.63)**<br>F: -2.11 (-4.60, 0.39)*<br>M: -1.48 (-3.59, 0.63) | -0.62 (-4.56, 3.32) |
| Mental health‡ | O: -1.91 (-3.17, -0.64)**<br>F: -4.30 (-6.10, -2.50)***<br>M: 0.25 (-1.34, 1.85) | -4.55 (-7.23, -1.87)*** |
| Relationship satisfaction‡ | O: -2.14 (-4.20, -0.07)**<br>F: -1.21 (-3.55, 1.12)<br>M: -2.98 (-5.56, -0.39)** | 1.76 (-1.15, 4.68) |
| Perceived support from others | O: -2.65 (-4.21, -1.10)***<br>F: -3.50 (-5.55, -1.45)***<br>M: -1.93 (-4.41, 0.56) | -1.58 (-5.09, 1.94) |
| Life evaluation today | O: -2.70 (-4.03, -1.38)***<br>F: -3.28 (-4.67, -1.88)***<br>M: -2.21 (-4.04, -0.38)** | -1.07 (-3.09, 0.95) |
| Life worthwhile‡ | O: -2.89 (-4.91, -0.87)**<br>F: -3.40 (-6.23, -0.57)**<br>M: -2.46 (-4.68, -0.24)** | -0.94 (-4.00, 2.12) |

Estimated beta coefficient

*Not feeling loved by mother – Ages 60+*

**Adjusted associations between not feeling loved by mother when growing up and adult outcomes in India (ages 60+)**

Wave 1, survey-weighted linear model. One model was fitted for each continuous outcome (24 models total, for 22 outcomes and 2 flourishing composite indices).
Estimates are the adjusted model coefficients (beta) of the predictor.

Group ● Overall (O) ▲ Female (F) ■ Male (M)

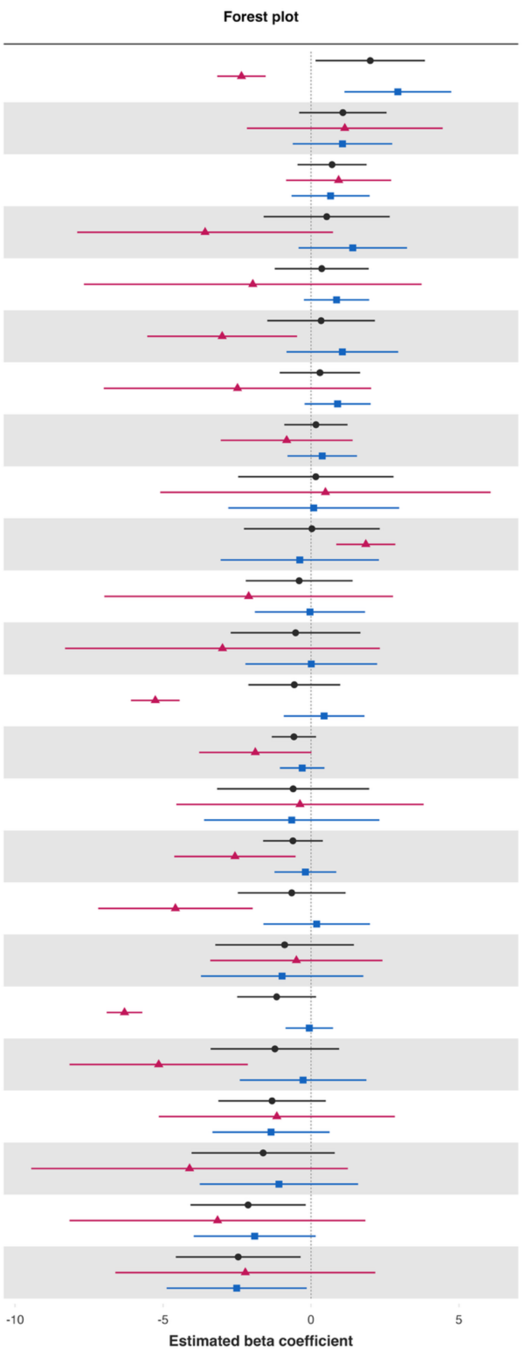


| Variable | Overall and interaction-derived gender estimates (95% CI) | Interaction (F–M, 95% CI) |
|---|---|---|
| Loneliness§ | O: 2.00 (0.15, 3.85)**<br>F: -2.35 (-3.17, -1.53)***<br>M: 2.93 (1.13, 4.74)** | -5.29 (-7.10, -3.47)*** |
| Life worthwhile‡ | O: 1.08 (-0.40, 2.55)<br>F: 1.14 (-2.17, 4.45)<br>M: 1.06 (-0.62, 2.74) | 0.08 (-3.70, 3.86) |
| Positive expectations | O: 0.71 (-0.46, 1.88)<br>F: 0.93 (-0.84, 2.71)<br>M: 0.66 (-0.66, 1.98) | 0.27 (-1.79, 2.33) |
| Worry about safety, food, or housing§§‡‡ | O: 0.53 (-1.60, 2.66)<br>F: -3.58 (-7.90, 0.74)<br>M: 1.41 (-0.42, 3.24) | -4.99 (-9.55, -0.43)** |
| Remain hopeful for the future | O: 0.36 (-1.22, 1.95)<br>F: -1.97 (-7.67, 3.74)<br>M: 0.86 (-0.24, 1.97) | -2.83 (-8.58, 2.91) |
| Physical health‡ | O: 0.34 (-1.48, 2.16)<br>F: -3.00 (-5.53, -0.47)**<br>M: 1.06 (-0.83, 2.94) | -4.06 (-7.22, -0.90)** |
| Show love often | O: 0.30 (-1.06, 1.66)<br>F: -2.49 (-7.00, 2.03)<br>M: 0.90 (-0.22, 2.01) | -3.39 (-8.08, 1.31) |
| Act to promote good‡ | O: 0.17 (-0.90, 1.23)<br>F: -0.82 (-3.05, 1.40)<br>M: 0.38 (-0.80, 1.55) | -1.20 (-3.76, 1.35) |
| Worry about meeting normal monthly living expenses§§‡‡ | O: 0.16 (-2.46, 2.78)<br>F: 0.49 (-5.09, 6.07)<br>M: 0.09 (-2.80, 2.98) | 0.40 (-5.77, 6.56) |
| Have a long list of gratitude | O: 0.03 (-2.26, 2.32)<br>F: 1.85 (0.85, 2.85)***<br>M: -0.38 (-3.05, 2.29) | 2.23 (-0.59, 5.05) |
| Understand purpose in life‡ | O: -0.40 (-2.21, 1.40)<br>F: -2.11 (-6.98, 2.77)<br>M: -0.03 (-1.90, 1.83) | -2.07 (-7.26, 3.12) |
| Life satisfaction‡ | O: -0.52 (-2.71, 1.67)<br>F: -2.99 (-8.31, 2.33)<br>M: 0.01 (-2.22, 2.23) | -3.00 (-8.51, 2.51) |
| Life evaluation today | O: -0.57 (-2.12, 0.99)<br>F: -5.26 (-6.09, -4.44)***<br>M: 0.44 (-0.92, 1.81) | -5.70 (-7.24, -4.17)*** |
| Flourishing (with financial indicators) | O: -0.58 (-1.33, 0.17)<br>F: -1.88 (-3.78, 0.01)*<br>M: -0.30 (-1.05, 0.46) | -1.59 (-3.58, 0.41) |
| Perceived support from others | O: -0.60 (-3.17, 1.97)<br>F: -0.37 (-4.55, 3.80)<br>M: -0.65 (-3.62, 2.31) | 0.28 (-4.80, 5.36) |
| Flourishing (without financial indicators) | O: -0.61 (-1.62, 0.40)<br>F: -2.57 (-4.62, -0.52)**<br>M: -0.19 (-1.24, 0.85) | -2.38 (-4.62, -0.14)** |
| Mental health‡ | O: -0.65 (-2.47, 1.17)<br>F: -4.58 (-7.19, -1.97)***<br>M: 0.19 (-1.61, 1.99) | -4.77 (-8.00, -1.55)** |
| Give up some happiness now for greater happiness later‡ | O: -0.89 (-3.23, 1.45)<br>F: -0.50 (-3.40, 2.41)<br>M: -0.98 (-3.72, 1.76) | 0.48 (-3.48, 4.45) |
| Happiness‡ | O: -1.16 (-2.49, 0.17)*<br>F: -6.30 (-6.91, -5.70)***<br>M: -0.06 (-0.86, 0.74) | -6.24 (-7.26, -5.23)*** |
| Belonging | O: -1.22 (-3.39, 0.95)<br>F: -5.15 (-8.16, -2.14)***<br>M: -0.27 (-2.41, 1.87) | -4.88 (-8.54, -1.22)** |
| Life evaluation five years from now | O: -1.32 (-3.13, 0.50)<br>F: -1.16 (-5.15, 2.83)<br>M: -1.35 (-3.33, 0.63) | 0.20 (-4.16, 4.55) |
| Relationship satisfaction‡ | O: -1.62 (-4.04, 0.80)<br>F: -4.11 (-9.45, 1.24)<br>M: -1.08 (-3.76, 1.59) | -3.02 (-8.87, 2.83) |
| Freedom | O: -2.13 (-4.07, -0.18)**<br>F: -3.16 (-8.16, 1.84)<br>M: -1.90 (-3.97, 0.16)* | -1.26 (-6.63, 4.12) |
| Relationship contentment‡ | O: -2.46 (-4.57, -0.35)**<br>F: -2.22 (-6.61, 2.17)<br>M: -2.51 (-4.88, -0.15)** | 0.29 (-4.69, 5.28) |

**Predictor:** *Not feeling loved by father*
*Not feeling loved by father – Ages 18 to 24 inclusive*

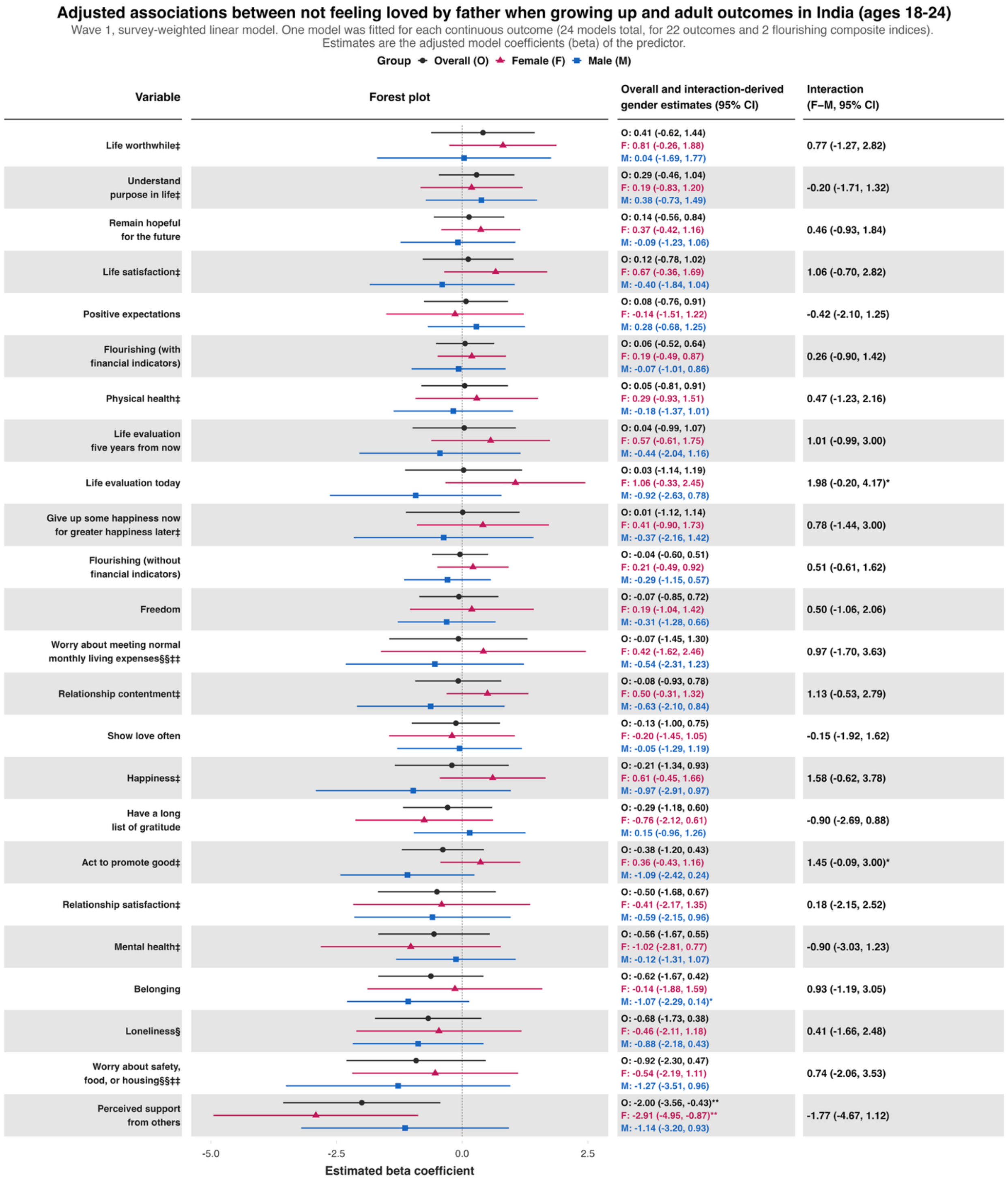

*Not feeling loved by father – Ages 25 to 29 inclusive*

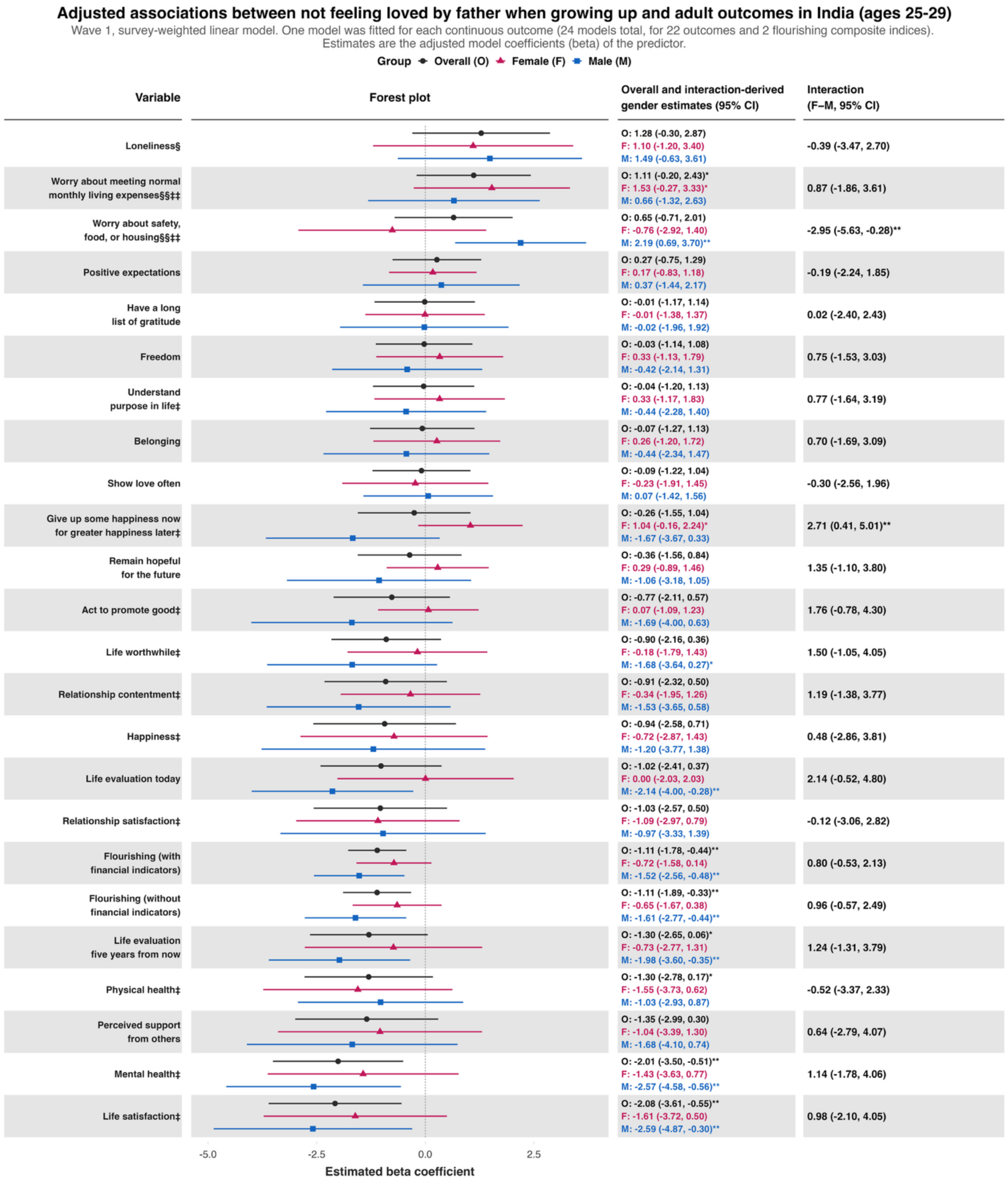

*Not feeling loved by father – Ages 30 to 39 inclusive*

**Adjusted associations between not feeling loved by father when growing up and adult outcomes in India (ages 30-39)**

Wave 1, survey-weighted linear model. One model was fitted for each continuous outcome (24 models total, for 22 outcomes and 2 flourishing composite indices).
Estimates are the adjusted model coefficients (beta) of the predictor.

Group: Overall (O), Female (F), Male (M)

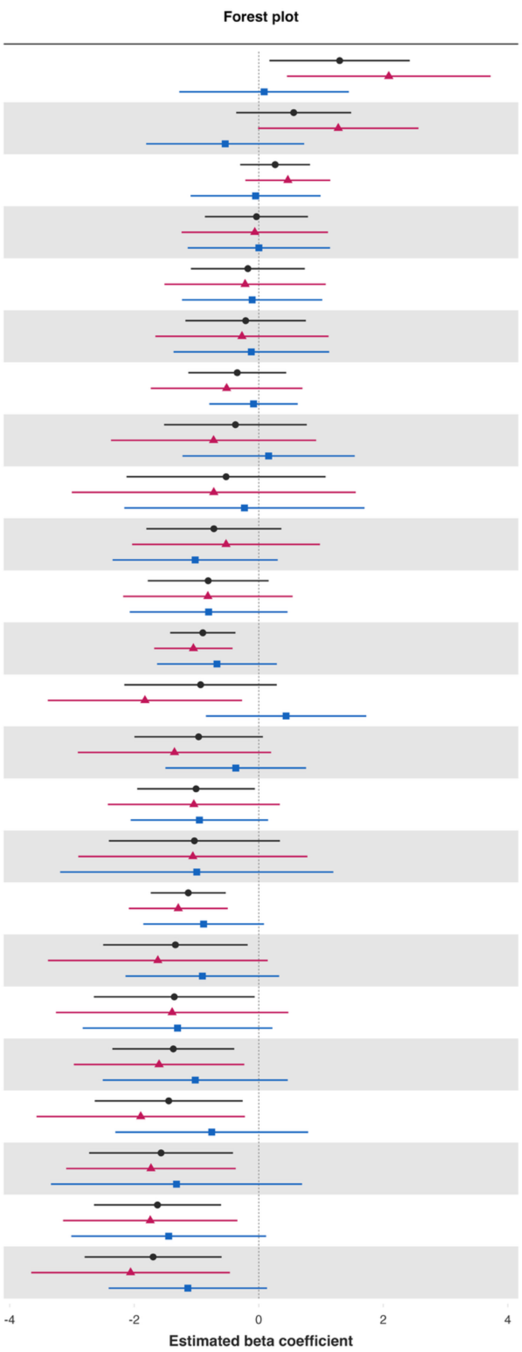


| Variable | Overall and interaction-derived gender estimates (95% CI) | Interaction (F–M, 95% CI) |
| --- | --- | --- |
| Loneliness§ | O: 1.30 (0.17, 2.43)**<br>F: 2.09 (0.45, 3.72)**<br>M: 0.09 (-1.28, 1.45) | 2.00 (-0.16, 4.17)* |
| Worry about meeting normal monthly living expenses§§‡‡ | O: 0.56 (-0.36, 1.48)<br>F: 1.28 (-0.01, 2.57)*<br>M: -0.54 (-1.81, 0.73) | 1.82 (-0.04, 3.67)* |
| Remain hopeful for the future | O: 0.26 (-0.30, 0.82)<br>F: 0.47 (-0.21, 1.15)<br>M: -0.05 (-1.09, 0.99) | 0.52 (-0.78, 1.82) |
| Life evaluation five years from now | O: -0.04 (-0.86, 0.79)<br>F: -0.06 (-1.24, 1.11)<br>M: 0.00 (-1.14, 1.15) | -0.07 (-1.73, 1.60) |
| Show love often | O: -0.17 (-1.09, 0.74)<br>F: -0.22 (-1.51, 1.08)<br>M: -0.11 (-1.23, 1.02) | -0.11 (-1.81, 1.58) |
| Have a long list of gratitude | O: -0.21 (-1.18, 0.76)<br>F: -0.27 (-1.66, 1.12)<br>M: -0.12 (-1.37, 1.13) | -0.15 (-2.04, 1.74) |
| Understand purpose in life‡ | O: -0.35 (-1.13, 0.44)<br>F: -0.52 (-1.73, 0.70)<br>M: -0.08 (-0.79, 0.63) | -0.43 (-1.87, 1.00) |
| Life evaluation today | O: -0.37 (-1.52, 0.77)<br>F: -0.73 (-2.37, 0.92)<br>M: 0.16 (-1.22, 1.54) | -0.88 (-3.06, 1.29) |
| Worry about safety, food, or housing§§‡‡ | O: -0.53 (-2.12, 1.07)<br>F: -0.72 (-3.00, 1.56)<br>M: -0.23 (-2.16, 1.70) | -0.49 (-3.45, 2.47) |
| Give up some happiness now for greater happiness later‡ | O: -0.72 (-1.81, 0.36)<br>F: -0.53 (-2.03, 0.98)<br>M: -1.02 (-2.35, 0.30) | 0.50 (-1.46, 2.45) |
| Belonging | O: -0.81 (-1.78, 0.16)<br>F: -0.82 (-2.18, 0.54)<br>M: -0.80 (-2.07, 0.46) | -0.01 (-1.86, 1.83) |
| Flourishing (with financial indicators) | O: -0.90 (-1.42, -0.37)***<br>F: -1.05 (-1.68, -0.42)**<br>M: -0.67 (-1.63, 0.29) | -0.38 (-1.55, 0.80) |
| Perceived support from others | O: -0.93 (-2.16, 0.29)<br>F: -1.83 (-3.39, -0.27)**<br>M: 0.44 (-0.85, 1.73) | -2.27 (-4.12, -0.41)** |
| Physical health‡ | O: -0.97 (-2.00, 0.07)*<br>F: -1.35 (-2.91, 0.20)*<br>M: -0.37 (-1.50, 0.76) | -0.98 (-2.95, 0.98) |
| Act to promote good‡ | O: -1.01 (-1.95, -0.06)**<br>F: -1.04 (-2.42, 0.34)<br>M: -0.95 (-2.06, 0.15)* | -0.09 (-1.85, 1.67) |
| Life worthwhile‡ | O: -1.03 (-2.41, 0.34)<br>F: -1.06 (-2.90, 0.78)<br>M: -1.00 (-3.19, 1.20) | -0.06 (-3.00, 2.87) |
| Flourishing (without financial indicators) | O: -1.13 (-1.73, -0.53)***<br>F: -1.29 (-2.09, -0.50)**<br>M: -0.89 (-1.85, 0.08)* | -0.41 (-1.69, 0.88) |
| Mental health‡ | O: -1.34 (-2.50, -0.18)**<br>F: -1.62 (-3.39, 0.14)*<br>M: -0.91 (-2.14, 0.33) | -0.72 (-2.93, 1.50) |
| Relationship satisfaction‡ | O: -1.36 (-2.65, -0.06)**<br>F: -1.39 (-3.26, 0.47)<br>M: -1.30 (-2.83, 0.22)* | -0.09 (-2.47, 2.30) |
| Happiness‡ | O: -1.37 (-2.35, -0.39)**<br>F: -1.60 (-2.97, -0.23)**<br>M: -1.02 (-2.51, 0.47) | -0.58 (-2.68, 1.52) |
| Life satisfaction‡ | O: -1.45 (-2.63, -0.26)**<br>F: -1.90 (-3.57, -0.22)**<br>M: -0.76 (-2.30, 0.79) | -1.14 (-3.46, 1.18) |
| Relationship contentment‡ | O: -1.57 (-2.72, -0.41)**<br>F: -1.73 (-3.09, -0.37)**<br>M: -1.32 (-3.34, 0.70) | -0.41 (-2.82, 2.00) |
| Positive expectations | O: -1.63 (-2.65, -0.60)**<br>F: -1.74 (-3.14, -0.34)**<br>M: -1.45 (-3.01, 0.12)* | -0.30 (-2.44, 1.84) |
| Freedom | O: -1.70 (-2.80, -0.60)**<br>F: -2.06 (-3.65, -0.46)**<br>M: -1.14 (-2.41, 0.13)* | -0.92 (-2.98, 1.14) |

*Not feeling loved by father – Ages 40 to 49 inclusive*

**Adjusted associations between not feeling loved by father when growing up and adult outcomes in India (ages 40-49)**

Wave 1, survey-weighted linear model. One model was fitted for each continuous outcome (24 models total, for 22 outcomes and 2 flourishing composite indices). Estimates are the adjusted model coefficients (beta) of the predictor.

Group: Overall (O), Female (F), Male (M)

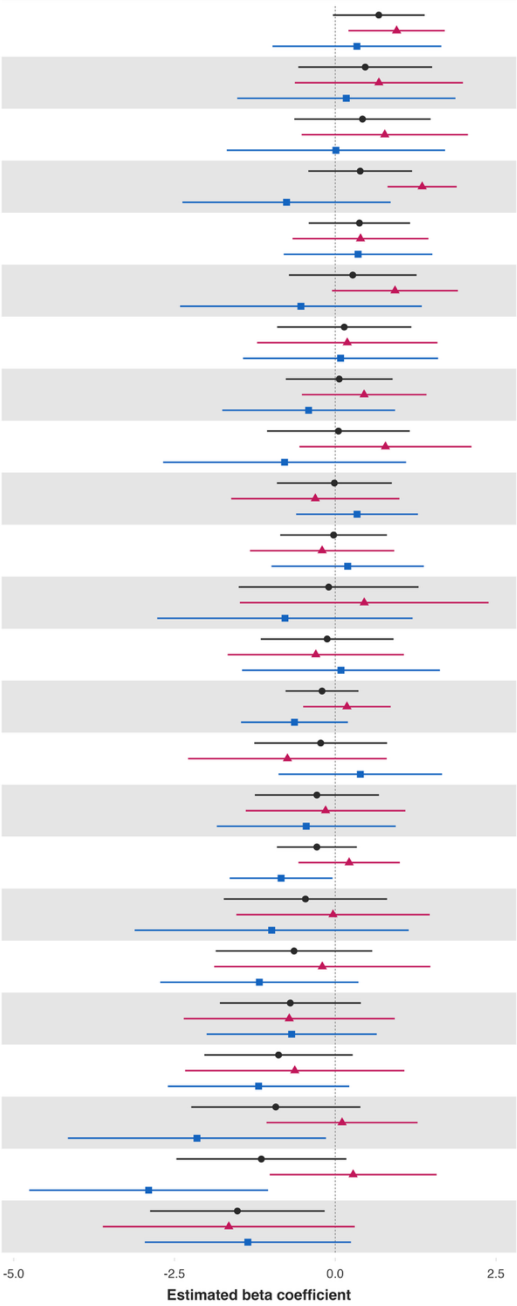


| Variable | Overall and interaction-derived gender estimates (95% CI) | Interaction (F–M, 95% CI) |
|---|---|---|
| Show love often | O: 0.68 (-0.04, 1.39)*<br>F: 0.95 (0.21, 1.70)**<br>M: 0.34 (-0.98, 1.65) | 0.62 (-0.91, 2.14) |
| Life evaluation five years from now | O: 0.47 (-0.57, 1.51)<br>F: 0.68 (-0.63, 1.99)<br>M: 0.17 (-1.52, 1.87) | 0.51 (-1.62, 2.63) |
| Life worthwhile‡ | O: 0.42 (-0.64, 1.48)<br>F: 0.77 (-0.52, 2.06)<br>M: 0.01 (-1.69, 1.71) | 0.76 (-1.34, 2.86) |
| Have a long list of gratitude | O: 0.39 (-0.42, 1.19)<br>F: 1.35 (0.81, 1.89)***<br>M: -0.76 (-2.38, 0.86) | 2.11 (0.39, 3.82)** |
| Positive expectations | O: 0.38 (-0.41, 1.16)<br>F: 0.39 (-0.66, 1.45)<br>M: 0.35 (-0.80, 1.51) | 0.04 (-1.51, 1.59) |
| Mental health‡ | O: 0.27 (-0.72, 1.27)<br>F: 0.93 (-0.05, 1.91)*<br>M: -0.53 (-2.41, 1.35) | 1.46 (-0.68, 3.61) |
| Worry about meeting normal monthly living expenses§§‡‡ | O: 0.14 (-0.90, 1.18)<br>F: 0.19 (-1.22, 1.59)<br>M: 0.08 (-1.43, 1.60) | 0.10 (-1.94, 2.15) |
| Freedom | O: 0.06 (-0.77, 0.89)<br>F: 0.45 (-0.52, 1.42)<br>M: -0.41 (-1.76, 0.93) | 0.86 (-0.75, 2.47) |
| Physical health‡ | O: 0.05 (-1.06, 1.16)<br>F: 0.78 (-0.56, 2.12)<br>M: -0.79 (-2.68, 1.10) | 1.57 (-0.79, 3.93) |
| Remain hopeful for the future | O: -0.01 (-0.91, 0.88)<br>F: -0.31 (-1.62, 1.00)<br>M: 0.34 (-0.61, 1.29) | -0.65 (-2.19, 0.89) |
| Act to promote good‡ | O: -0.03 (-0.86, 0.80)<br>F: -0.20 (-1.33, 0.92)<br>M: 0.19 (-0.99, 1.38) | -0.40 (-2.01, 1.22) |
| Loneliness§ | O: -0.10 (-1.50, 1.30)<br>F: 0.45 (-1.48, 2.39)<br>M: -0.78 (-2.77, 1.20) | 1.23 (-1.60, 4.06) |
| Belonging | O: -0.13 (-1.16, 0.91)<br>F: -0.30 (-1.67, 1.07)<br>M: 0.09 (-1.45, 1.63) | -0.39 (-2.44, 1.66) |
| Flourishing (with financial indicators) | O: -0.21 (-0.77, 0.36)<br>F: 0.18 (-0.50, 0.86)<br>M: -0.63 (-1.47, 0.20) | 0.82 (-0.23, 1.86) |
| Understand purpose in life‡ | O: -0.23 (-1.26, 0.81)<br>F: -0.74 (-2.29, 0.80)<br>M: 0.39 (-0.88, 1.66) | -1.13 (-3.14, 0.87) |
| Give up some happiness now for greater happiness later‡ | O: -0.29 (-1.25, 0.68)<br>F: -0.15 (-1.39, 1.09)<br>M: -0.45 (-1.84, 0.94) | 0.30 (-1.49, 2.09) |
| Flourishing (without financial indicators) | O: -0.29 (-0.91, 0.34)<br>F: 0.22 (-0.57, 1.00)<br>M: -0.84 (-1.64, -0.04)** | 1.06 (-0.03, 2.14)* |
| Worry about safety, food, or housing§§‡‡ | O: -0.46 (-1.73, 0.81)<br>F: -0.03 (-1.54, 1.47)<br>M: -0.99 (-3.12, 1.14) | 0.95 (-1.73, 3.64) |
| Happiness‡ | O: -0.64 (-1.86, 0.58)<br>F: -0.20 (-1.88, 1.48)<br>M: -1.18 (-2.72, 0.36) | 0.98 (-1.20, 3.16) |
| Life satisfaction‡ | O: -0.70 (-1.79, 0.40)<br>F: -0.72 (-2.36, 0.93)<br>M: -0.68 (-2.00, 0.64) | -0.04 (-2.11, 2.04) |
| Life evaluation today | O: -0.88 (-2.04, 0.27)<br>F: -0.63 (-2.33, 1.08)<br>M: -1.19 (-2.60, 0.22)* | 0.56 (-1.66, 2.79) |
| Relationship satisfaction‡ | O: -0.92 (-2.24, 0.39)<br>F: 0.11 (-1.07, 1.28)<br>M: -2.15 (-4.16, -0.14)** | 2.26 (-0.01, 4.52)* |
| Relationship contentment‡ | O: -1.15 (-2.47, 0.17)*<br>F: 0.28 (-1.02, 1.58)<br>M: -2.90 (-4.76, -1.04)** | 3.18 (0.89, 5.47)** |
| Perceived support from others | O: -1.52 (-2.88, -0.16)**<br>F: -1.65 (-3.61, 0.30)*<br>M: -1.36 (-2.96, 0.24)* | -0.30 (-2.71, 2.12) |

*Not feeling loved by father – Ages 50 to 59 inclusive*

**Adjusted associations between not feeling loved by father when growing up and adult outcomes in India (ages 50-59)**

Wave 1, survey-weighted linear model. One model was fitted for each continuous outcome (24 models total, for 22 outcomes and 2 flourishing composite indices).
Estimates are the adjusted model coefficients (beta) of the predictor.

Group ● Overall (O) ▲ Female (F) ■ Male (M)

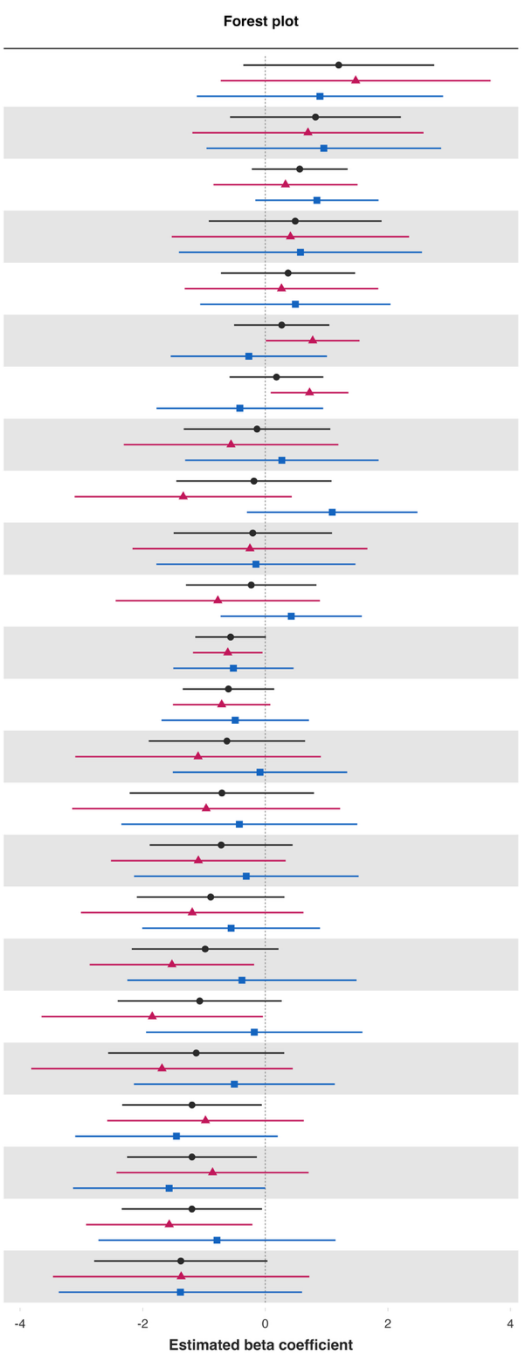


| Variable | Overall and interaction-derived gender estimates (95% CI) | Interaction (F–M, 95% CI) |
|---|---|---|
| Loneliness§ | O: 1.20 (-0.36, 2.76)<br>F: 1.48 (-0.73, 3.68)<br>M: 0.89 (-1.12, 2.90) | 0.58 (-2.31, 3.48) |
| Worry about safety, food, or housing§§‡‡ | O: 0.82 (-0.57, 2.21)<br>F: 0.70 (-1.19, 2.58)<br>M: 0.96 (-0.96, 2.87) | -0.26 (-2.84, 2.33) |
| Have a long list of gratitude | O: 0.56 (-0.22, 1.34)<br>F: 0.33 (-0.85, 1.51)<br>M: 0.84 (-0.16, 1.85) | -0.51 (-2.06, 1.04) |
| Worry about meeting normal monthly living expenses§§‡‡ | O: 0.49 (-0.92, 1.90)<br>F: 0.41 (-1.53, 2.35)<br>M: 0.57 (-1.41, 2.56) | -0.16 (-2.88, 2.56) |
| Physical health‡ | O: 0.37 (-0.72, 1.47)<br>F: 0.26 (-1.31, 1.84)<br>M: 0.49 (-1.06, 2.04) | -0.23 (-2.47, 2.02) |
| Remain hopeful for the future | O: 0.27 (-0.51, 1.05)<br>F: 0.77 (0.01, 1.54)**<br>M: -0.27 (-1.54, 1.01) | 1.04 (-0.44, 2.53) |
| Act to promote good‡ | O: 0.18 (-0.58, 0.95)<br>F: 0.72 (0.09, 1.36)**<br>M: -0.42 (-1.78, 0.94) | 1.14 (-0.35, 2.63) |
| Understand purpose in life‡ | O: -0.14 (-1.33, 1.06)<br>F: -0.56 (-2.31, 1.19)<br>M: 0.27 (-1.31, 1.85) | -0.83 (-3.15, 1.49) |
| Life satisfaction‡ | O: -0.19 (-1.45, 1.08)<br>F: -1.34 (-3.11, 0.43)<br>M: 1.09 (-0.30, 2.48) | -2.43 (-4.70, -0.16)** |
| Give up some happiness now for greater happiness later‡ | O: -0.20 (-1.50, 1.09)<br>F: -0.25 (-2.17, 1.67)<br>M: -0.15 (-1.78, 1.47) | -0.10 (-2.56, 2.36) |
| Belonging | O: -0.23 (-1.29, 0.83)<br>F: -0.77 (-2.44, 0.89)<br>M: 0.42 (-0.73, 1.58) | -1.20 (-3.27, 0.88) |
| Flourishing (with financial indicators) | O: -0.57 (-1.14, 0.01)*<br>F: -0.61 (-1.18, -0.05)**<br>M: -0.52 (-1.50, 0.46) | -0.09 (-1.21, 1.03) |
| Flourishing (without financial indicators) | O: -0.60 (-1.35, 0.15)<br>F: -0.71 (-1.51, 0.08)*<br>M: -0.49 (-1.69, 0.71) | -0.22 (-1.62, 1.18) |
| Positive expectations | O: -0.63 (-1.90, 0.65)<br>F: -1.10 (-3.10, 0.91)<br>M: -0.09 (-1.51, 1.34) | -1.01 (-3.47, 1.45) |
| Perceived support from others | O: -0.71 (-2.21, 0.80)<br>F: -0.96 (-3.15, 1.22)<br>M: -0.42 (-2.35, 1.50) | -0.54 (-3.36, 2.28) |
| Life evaluation five years from now | O: -0.72 (-1.89, 0.45)<br>F: -1.09 (-2.52, 0.33)<br>M: -0.31 (-2.14, 1.52) | -0.78 (-3.10, 1.54) |
| Freedom | O: -0.89 (-2.10, 0.31)<br>F: -1.19 (-3.01, 0.62)<br>M: -0.56 (-2.01, 0.89) | -0.64 (-2.89, 1.61) |
| Happiness‡ | O: -0.98 (-2.18, 0.22)<br>F: -1.52 (-2.86, -0.18)**<br>M: -0.38 (-2.25, 1.49) | -1.14 (-3.39, 1.11) |
| Mental health‡ | O: -1.07 (-2.41, 0.27)<br>F: -1.84 (-3.65, -0.04)**<br>M: -0.18 (-1.95, 1.58) | -1.66 (-4.15, 0.82) |
| Show love often | O: -1.13 (-2.56, 0.31)<br>F: -1.69 (-3.82, 0.45)<br>M: -0.51 (-2.15, 1.13) | -1.18 (-3.80, 1.44) |
| Relationship satisfaction‡ | O: -1.20 (-2.34, -0.06)**<br>F: -0.98 (-2.58, 0.63)<br>M: -1.45 (-3.10, 0.20)* | 0.47 (-1.86, 2.81) |
| Relationship contentment‡ | O: -1.20 (-2.26, -0.14)**<br>F: -0.86 (-2.43, 0.71)<br>M: -1.57 (-3.14, 0.00)* | 0.71 (-1.59, 3.01) |
| Life evaluation today | O: -1.20 (-2.34, -0.05)**<br>F: -1.57 (-2.92, -0.21)**<br>M: -0.79 (-2.72, 1.15) | -0.78 (-3.19, 1.63) |
| Life worthwhile‡ | O: -1.38 (-2.79, 0.04)*<br>F: -1.37 (-3.47, 0.72)<br>M: -1.39 (-3.37, 0.60) | 0.01 (-2.94, 2.96) |

*Not feeling loved by father – Ages 60+*

**Adjusted associations between not feeling loved by father when growing up and adult outcomes in India (ages 60+)**

Wave 1, survey-weighted linear model. One model was fitted for each continuous outcome (24 models total, for 22 outcomes and 2 flourishing composite indices). Estimates are the adjusted model coefficients (beta) of the predictor.

Group: Overall (O), Female (F), Male (M)

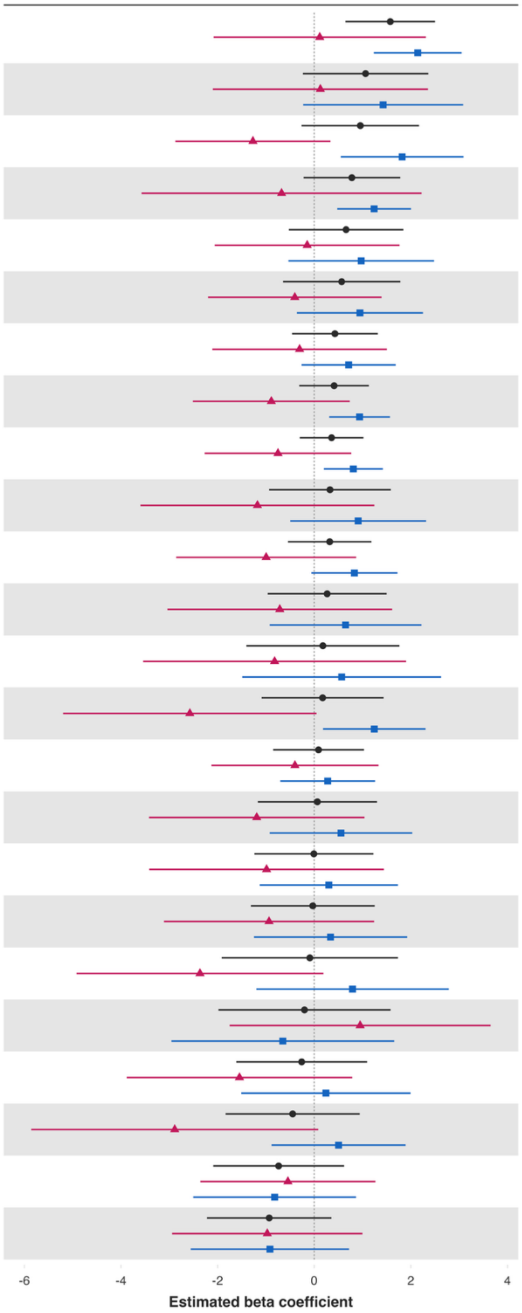


| Variable | Overall and interaction-derived gender estimates (95% CI) | Interaction (F–M, 95% CI) |
|---|---|---|
| Life worthwhile‡ | O: 1.57 (0.65, 2.50)***<br>F: 0.11 (-2.08, 2.31)<br>M: 2.14 (1.23, 3.05)*** | -2.03 (-4.46, 0.41) |
| Loneliness§ | O: 1.06 (-0.23, 2.36)<br>F: 0.13 (-2.10, 2.35)<br>M: 1.43 (-0.23, 3.08)* | -1.30 (-4.27, 1.67) |
| Physical health‡ | O: 0.95 (-0.26, 2.17)<br>F: -1.27 (-2.87, 0.34)<br>M: 1.82 (0.55, 3.09)** | -3.09 (-5.06, -1.12)** |
| Positive expectations | O: 0.78 (-0.22, 1.78)<br>F: -0.68 (-3.57, 2.22)<br>M: 1.24 (0.48, 2.00)** | -1.92 (-4.84, 1.01) |
| Give up some happiness now for greater happiness later‡ | O: 0.66 (-0.53, 1.84)<br>F: -0.15 (-2.06, 1.77)<br>M: 0.97 (-0.53, 2.48) | -1.12 (-3.62, 1.38) |
| Mental health‡ | O: 0.57 (-0.65, 1.78)<br>F: -0.40 (-2.20, 1.39)<br>M: 0.95 (-0.36, 2.25) | -1.35 (-3.34, 0.65) |
| Act to promote good‡ | O: 0.43 (-0.46, 1.32)<br>F: -0.30 (-2.11, 1.50)<br>M: 0.71 (-0.26, 1.69) | -1.02 (-3.09, 1.06) |
| Flourishing (without financial indicators) | O: 0.41 (-0.31, 1.13)<br>F: -0.89 (-2.51, 0.74)<br>M: 0.94 (0.31, 1.57)** | -1.83 (-3.56, -0.09)** |
| Flourishing (with financial indicators) | O: 0.36 (-0.30, 1.02)<br>F: -0.75 (-2.27, 0.77)<br>M: 0.81 (0.20, 1.42)** | -1.56 (-3.23, 0.11)* |
| Life satisfaction‡ | O: 0.33 (-0.93, 1.59)<br>F: -1.18 (-3.60, 1.25)<br>M: 0.91 (-0.50, 2.32) | -2.09 (-4.86, 0.69) |
| Remain hopeful for the future | O: 0.32 (-0.54, 1.18)<br>F: -0.99 (-2.86, 0.87)<br>M: 0.83 (-0.06, 1.72)* | -1.83 (-3.90, 0.25)* |
| Happiness‡ | O: 0.27 (-0.96, 1.50)<br>F: -0.71 (-3.04, 1.61)<br>M: 0.65 (-0.92, 2.22) | -1.36 (-4.31, 1.58) |
| Worry about safety, food, or housing§§‡‡ | O: 0.18 (-1.40, 1.76)<br>F: -0.82 (-3.54, 1.90)<br>M: 0.57 (-1.49, 2.63) | -1.39 (-5.03, 2.26) |
| Relationship satisfaction‡ | O: 0.17 (-1.09, 1.44)<br>F: -2.57 (-5.20, 0.05)*<br>M: 1.24 (0.18, 2.30)** | -3.82 (-6.65, -0.99)** |
| Freedom | O: 0.09 (-0.85, 1.03)<br>F: -0.40 (-2.13, 1.33)<br>M: 0.28 (-0.70, 1.26) | -0.68 (-2.54, 1.18) |
| Relationship contentment‡ | O: 0.07 (-1.17, 1.30)<br>F: -1.19 (-3.42, 1.04)<br>M: 0.55 (-0.92, 2.03) | -1.74 (-4.45, 0.96) |
| Life evaluation today | O: -0.01 (-1.24, 1.22)<br>F: -0.99 (-3.42, 1.44)<br>M: 0.30 (-1.13, 1.73) | -1.29 (-4.09, 1.51) |
| Understand purpose in life‡ | O: -0.03 (-1.31, 1.25)<br>F: -0.94 (-3.11, 1.24)<br>M: 0.34 (-1.25, 1.92) | -1.27 (-3.94, 1.39) |
| Perceived support from others | O: -0.09 (-1.91, 1.73)<br>F: -2.36 (-4.92, 0.19)*<br>M: 0.79 (-1.20, 2.78) | -3.16 (-6.00, -0.32)** |
| Worry about meeting normal monthly living expenses§§‡‡ | O: -0.20 (-1.98, 1.58)<br>F: 0.95 (-1.75, 3.65)<br>M: -0.65 (-2.96, 1.66) | 1.60 (-2.16, 5.36) |
| Have a long list of gratitude | O: -0.26 (-1.61, 1.09)<br>F: -1.55 (-3.88, 0.79)<br>M: 0.24 (-1.51, 1.99) | -1.79 (-4.81, 1.23) |
| Show love often | O: -0.45 (-1.83, 0.94)<br>F: -2.89 (-5.86, 0.08)*<br>M: 0.50 (-0.88, 1.89) | -3.39 (-6.77, -0.01)* |
| Belonging | O: -0.74 (-2.09, 0.62)<br>F: -0.54 (-2.36, 1.27)<br>M: -0.82 (-2.50, 0.87) | 0.27 (-2.06, 2.60) |
| Life evaluation five years from now | O: -0.93 (-2.22, 0.36)<br>F: -0.97 (-2.94, 1.00)<br>M: -0.92 (-2.56, 0.72) | -0.06 (-2.67, 2.56) |

**Predictor:** ***Poorer relationship quality with mother (bad or very bad relationship)***
***Poorer relationship quality with mother (bad or very bad relationship) – Ages 18 to 24 inclusive***

**Adjusted associations between somewhat bad or very bad relationship with mother (vs very good) and adult outcomes in India (ages 18-24)**

Wave 1, survey-weighted linear model. One model was fitted for each continuous outcome (24 models total, for 22 outcomes and 2 flourishing composite indices).
Estimates are the adjusted model coefficients (beta) of the predictor.

Group: Overall (O), Female (F), Male (M)

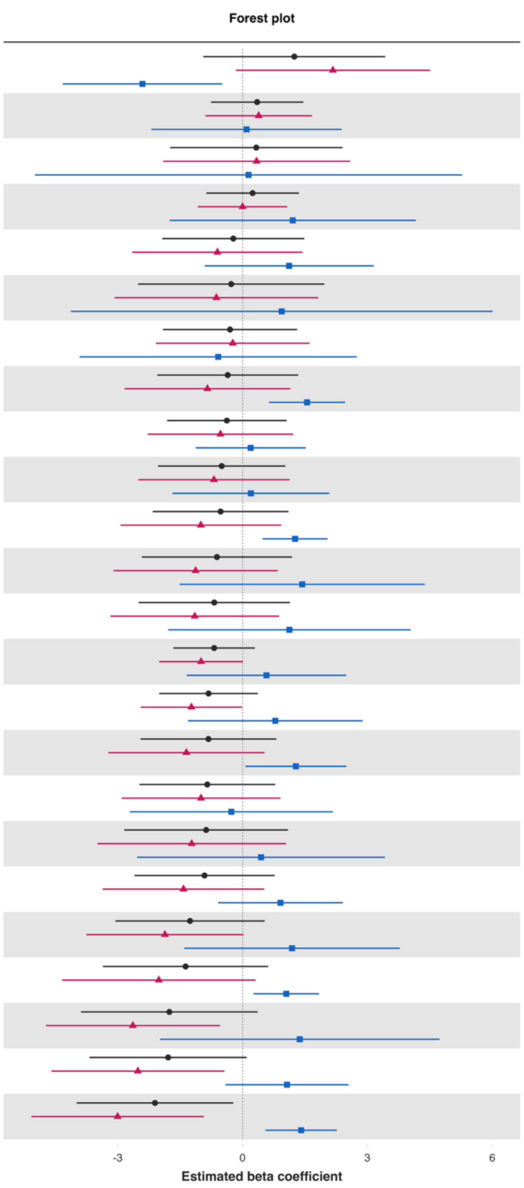


| Variable | Overall and interaction-derived gender estimates (95% CI) | Interaction (F–M, 95% CI) |
|---|---|---|
| Loneliness§ | O: 1.24 (-0.94, 3.43)<br>F: 2.17 (-0.16, 4.50)*<br>M: -2.41 (-4.32, -0.49)** | 4.58 (1.54, 7.62)** |
| Mental health‡ | O: 0.35 (-0.76, 1.46)<br>F: 0.39 (-0.89, 1.67)<br>M: 0.09 (-2.19, 2.38) | 0.29 (-2.35, 2.93) |
| Worry about safety, food, or housing§§‡‡ | O: 0.33 (-1.74, 2.40)<br>F: 0.34 (-1.91, 2.58)<br>M: 0.14 (-4.99, 5.28) | 0.19 (-5.40, 5.79) |
| Life satisfaction‡ | O: 0.24 (-0.87, 1.35)<br>F: -0.00 (-1.08, 1.07)<br>M: 1.21 (-1.76, 4.17) | -1.21 (-4.35, 1.94) |
| Perceived support from others | O: -0.22 (-1.93, 1.48)<br>F: -0.61 (-2.65, 1.44)<br>M: 1.12 (-0.91, 3.15) | -1.72 (-4.59, 1.14) |
| Worry about meeting normal monthly living expenses§§‡‡ | O: -0.27 (-2.51, 1.96)<br>F: -0.63 (-3.08, 1.82)<br>M: 0.94 (-4.13, 6.01) | -1.57 (-7.19, 4.05) |
| Belonging | O: -0.30 (-1.91, 1.31)<br>F: -0.24 (-2.08, 1.61)<br>M: -0.59 (-3.92, 2.74) | 0.35 (-3.46, 4.16) |
| Show love often | O: -0.36 (-2.05, 1.34)<br>F: -0.85 (-2.84, 1.15)<br>M: 1.55 (0.63, 2.46)*** | -2.40 (-4.57, -0.22)** |
| Act to promote good‡ | O: -0.38 (-1.81, 1.06)<br>F: -0.53 (-2.28, 1.22)<br>M: 0.19 (-1.13, 1.52) | -0.73 (-2.93, 1.47) |
| Positive expectations | O: -0.50 (-2.03, 1.03)<br>F: -0.69 (-2.51, 1.13)<br>M: 0.20 (-1.68, 2.08) | -0.89 (-3.51, 1.73) |
| Remain hopeful for the future | O: -0.53 (-2.16, 1.10)<br>F: -1.00 (-2.94, 0.93)<br>M: 1.26 (0.48, 2.04)** | -2.26 (-4.34, -0.18)** |
| Happiness‡ | O: -0.62 (-2.42, 1.19)<br>F: -1.13 (-3.10, 0.84)<br>M: 1.43 (-1.51, 4.38) | -2.56 (-6.13, 1.00) |
| Give up some happiness now for greater happiness later‡ | O: -0.68 (-2.50, 1.14)<br>F: -1.15 (-3.18, 0.88)<br>M: 1.12 (-1.79, 4.04) | -2.28 (-5.83, 1.28) |
| Flourishing (with financial indicators) | O: -0.68 (-1.66, 0.30)<br>F: -1.00 (-2.01, 0.01)*<br>M: 0.57 (-1.34, 2.49) | -1.57 (-3.74, 0.60) |
| Flourishing (without financial indicators) | O: -0.82 (-2.00, 0.37)<br>F: -1.23 (-2.45, -0.01)**<br>M: 0.79 (-1.31, 2.88) | -2.02 (-4.45, 0.41) |
| Have a long list of gratitude | O: -0.82 (-2.45, 0.81)<br>F: -1.35 (-3.23, 0.53)<br>M: 1.28 (0.07, 2.49)** | -2.63 (-4.87, -0.39)** |
| Understand purpose in life‡ | O: -0.85 (-2.48, 0.78)<br>F: -1.00 (-2.91, 0.91)<br>M: -0.27 (-2.71, 2.17) | -0.73 (-3.82, 2.36) |
| Relationship satisfaction‡ | O: -0.88 (-2.84, 1.09)<br>F: -1.22 (-3.49, 1.04)<br>M: 0.44 (-2.54, 3.42) | -1.67 (-5.40, 2.07) |
| Life evaluation five years from now | O: -0.92 (-2.60, 0.77)<br>F: -1.42 (-3.36, 0.52)<br>M: 0.91 (-0.59, 2.41) | -2.33 (-4.80, 0.14)* |
| Life worthwhile‡ | O: -1.26 (-3.06, 0.53)<br>F: -1.87 (-3.76, 0.02)*<br>M: 1.19 (-1.40, 3.78) | -3.05 (-6.27, 0.16)* |
| Freedom | O: -1.37 (-3.36, 0.61)<br>F: -2.01 (-4.34, 0.31)*<br>M: 1.05 (0.26, 1.84)** | -3.06 (-5.53, -0.60)** |
| Life evaluation today | O: -1.76 (-3.88, 0.36)<br>F: -2.64 (-4.73, -0.54)**<br>M: 1.37 (-1.98, 4.73) | -4.01 (-7.98, -0.04)** |
| Relationship contentment‡ | O: -1.79 (-3.68, 0.10)*<br>F: -2.52 (-4.60, -0.44)**<br>M: 1.07 (-0.41, 2.55) | -3.58 (-6.13, -1.04)** |
| Physical health‡ | O: -2.11 (-3.99, -0.22)**<br>F: -3.00 (-5.08, -0.93)**<br>M: 1.41 (0.55, 2.26)** | -4.41 (-6.67, -2.16)*** |

***Poorer relationship quality with mother (bad or very bad relationship) – Ages 25 to 29 inclusive***

**Adjusted associations between somewhat bad or very bad relationship with mother (vs very good) and adult outcomes in India (ages 25-29)**

Wave 1, survey-weighted linear model. One model was fitted for each continuous outcome (24 models total, for 22 outcomes and 2 flourishing composite indices).
Estimates are the adjusted model coefficients (beta) of the predictor.

Group: Overall (O), Female (F), Male (M)

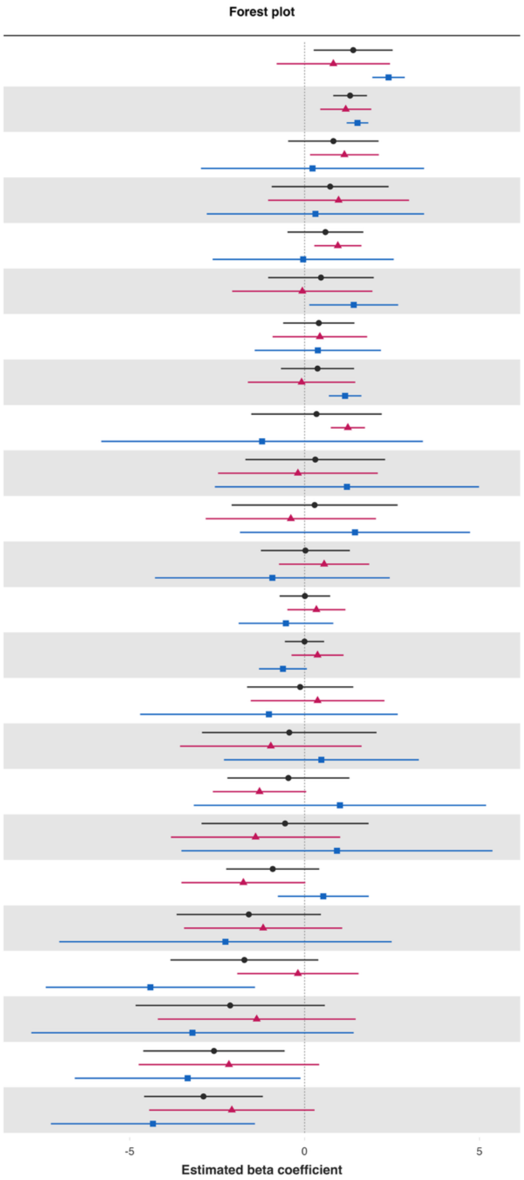


| Variable | Overall and interaction-derived gender estimates (95% CI) | Interaction (F–M, 95% CI) |
|---|---|---|
| Give up some happiness now for greater happiness later‡ | O: 1.39 (0.26, 2.52)**<br>F: 0.82 (-0.80, 2.44)<br>M: 2.40 (1.94, 2.86)*** | -1.58 (-3.28, 0.12)* |
| Understand purpose in life‡ | O: 1.30 (0.82, 1.78)***<br>F: 1.18 (0.45, 1.91)**<br>M: 1.51 (1.20, 1.82)*** | -0.34 (-1.13, 0.46) |
| Mental health‡ | O: 0.82 (-0.47, 2.11)<br>F: 1.14 (0.16, 2.12)**<br>M: 0.23 (-2.96, 3.42) | 0.91 (-2.46, 4.29) |
| Happiness‡ | O: 0.73 (-0.94, 2.40)<br>F: 0.97 (-1.04, 2.99)<br>M: 0.31 (-2.79, 3.42) | 0.66 (-3.08, 4.40) |
| Act to promote good‡ | O: 0.60 (-0.49, 1.68)<br>F: 0.95 (0.28, 1.62)**<br>M: -0.04 (-2.63, 2.55) | 0.99 (-1.65, 3.63) |
| Physical health‡ | O: 0.47 (-1.04, 1.98)<br>F: -0.06 (-2.07, 1.94)<br>M: 1.40 (0.13, 2.67)** | -1.47 (-3.73, 0.79) |
| Freedom | O: 0.41 (-0.61, 1.43)<br>F: 0.44 (-0.91, 1.79)<br>M: 0.38 (-1.42, 2.18) | 0.06 (-2.32, 2.44) |
| Remain hopeful for the future | O: 0.37 (-0.67, 1.42)<br>F: -0.09 (-1.62, 1.45)<br>M: 1.16 (0.69, 1.62)*** | -1.24 (-2.85, 0.36) |
| Show love often | O: 0.34 (-1.52, 2.20)<br>F: 1.24 (0.75, 1.73)***<br>M: -1.21 (-5.81, 3.38) | 2.45 (-2.13, 7.04) |
| Worry about meeting normal monthly living expenses§§‡‡ | O: 0.31 (-1.69, 2.30)<br>F: -0.19 (-2.47, 2.09)<br>M: 1.21 (-2.56, 4.99) | -1.40 (-5.98, 3.18) |
| Life evaluation today | O: 0.29 (-2.08, 2.66)<br>F: -0.39 (-2.83, 2.04)<br>M: 1.44 (-1.85, 4.73) | -1.83 (-5.17, 1.50) |
| Positive expectations | O: 0.02 (-1.25, 1.29)<br>F: 0.56 (-0.73, 1.85)<br>M: -0.92 (-4.28, 2.43) | 1.48 (-2.33, 5.29) |
| Flourishing (without financial indicators) | O: 0.01 (-0.71, 0.73)<br>F: 0.34 (-0.49, 1.17)<br>M: -0.53 (-1.89, 0.82) | 0.87 (-0.72, 2.46) |
| Flourishing (with financial indicators) | O: -0.00 (-0.56, 0.56)<br>F: 0.37 (-0.37, 1.11)<br>M: -0.62 (-1.30, 0.07)* | 0.99 (0.01, 1.97)** |
| Life satisfaction‡ | O: -0.13 (-1.64, 1.39)<br>F: 0.37 (-1.54, 2.28)<br>M: -1.02 (-4.70, 2.66) | 1.39 (-3.09, 5.87) |
| Have a long list of gratitude | O: -0.44 (-2.93, 2.06)<br>F: -0.97 (-3.56, 1.63)<br>M: 0.48 (-2.31, 3.26) | -1.45 (-3.82, 0.93) |
| Loneliness§ | O: -0.46 (-2.21, 1.28)<br>F: -1.29 (-2.62, 0.05)*<br>M: 1.01 (-3.17, 5.19) | -2.30 (-6.69, 2.09) |
| Worry about safety, food, or housing§§‡‡ | O: -0.56 (-2.94, 1.83)<br>F: -1.40 (-3.82, 1.02)<br>M: 0.93 (-3.52, 5.37) | -2.33 (-7.18, 2.52) |
| Belonging | O: -0.91 (-2.24, 0.42)<br>F: -1.75 (-3.52, 0.02)*<br>M: 0.54 (-0.76, 1.83) | -2.29 (-4.53, -0.04)** |
| Relationship satisfaction‡ | O: -1.59 (-3.66, 0.47)<br>F: -1.18 (-3.44, 1.08)<br>M: -2.26 (-7.01, 2.49) | 1.08 (-4.51, 6.67) |
| Relationship contentment‡ | O: -1.72 (-3.83, 0.39)<br>F: -0.19 (-1.92, 1.54)<br>M: -4.41 (-7.40, -1.42)** | 4.22 (1.04, 7.39)** |
| Life evaluation five years from now | O: -2.13 (-4.83, 0.58)<br>F: -1.37 (-4.20, 1.46)<br>M: -3.20 (-7.81, 1.40) | 1.84 (-3.50, 7.17) |
| Life worthwhile‡ | O: -2.59 (-4.61, -0.57)**<br>F: -2.16 (-4.74, 0.42)<br>M: -3.34 (-6.57, -0.12)** | 1.18 (-3.06, 5.42) |
| Perceived support from others | O: -2.89 (-4.59, -1.19)***<br>F: -2.08 (-4.44, 0.28)*<br>M: -4.33 (-7.25, -1.42)** | 2.26 (-2.20, 6.71) |

*Poorer relationship quality with mother (bad or very bad relationship) – Ages 30 to 39 inclusive*

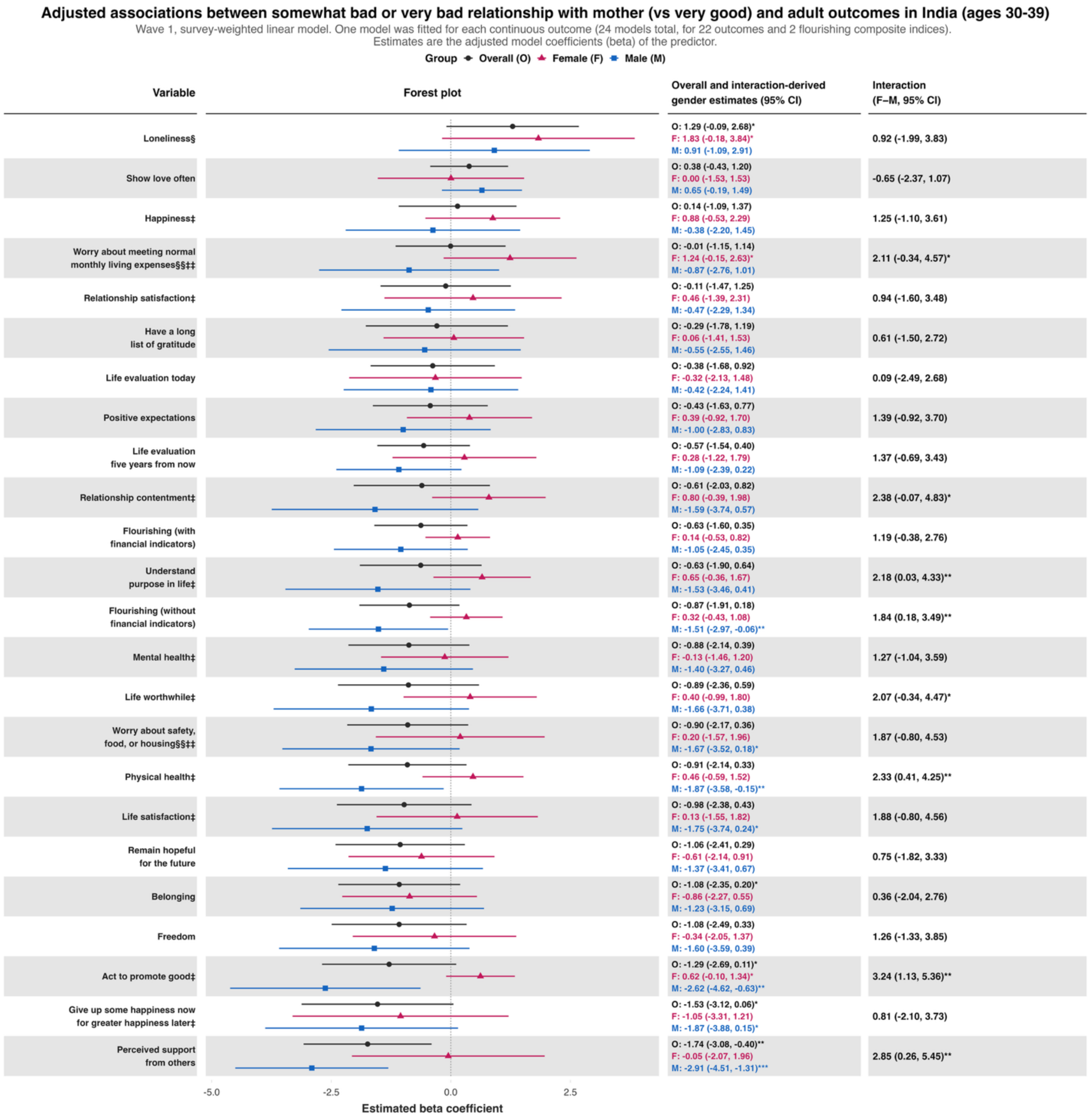

#### *Poorer relationship quality with mother (bad or very bad relationship) – Ages 40 to 49 inclusive*

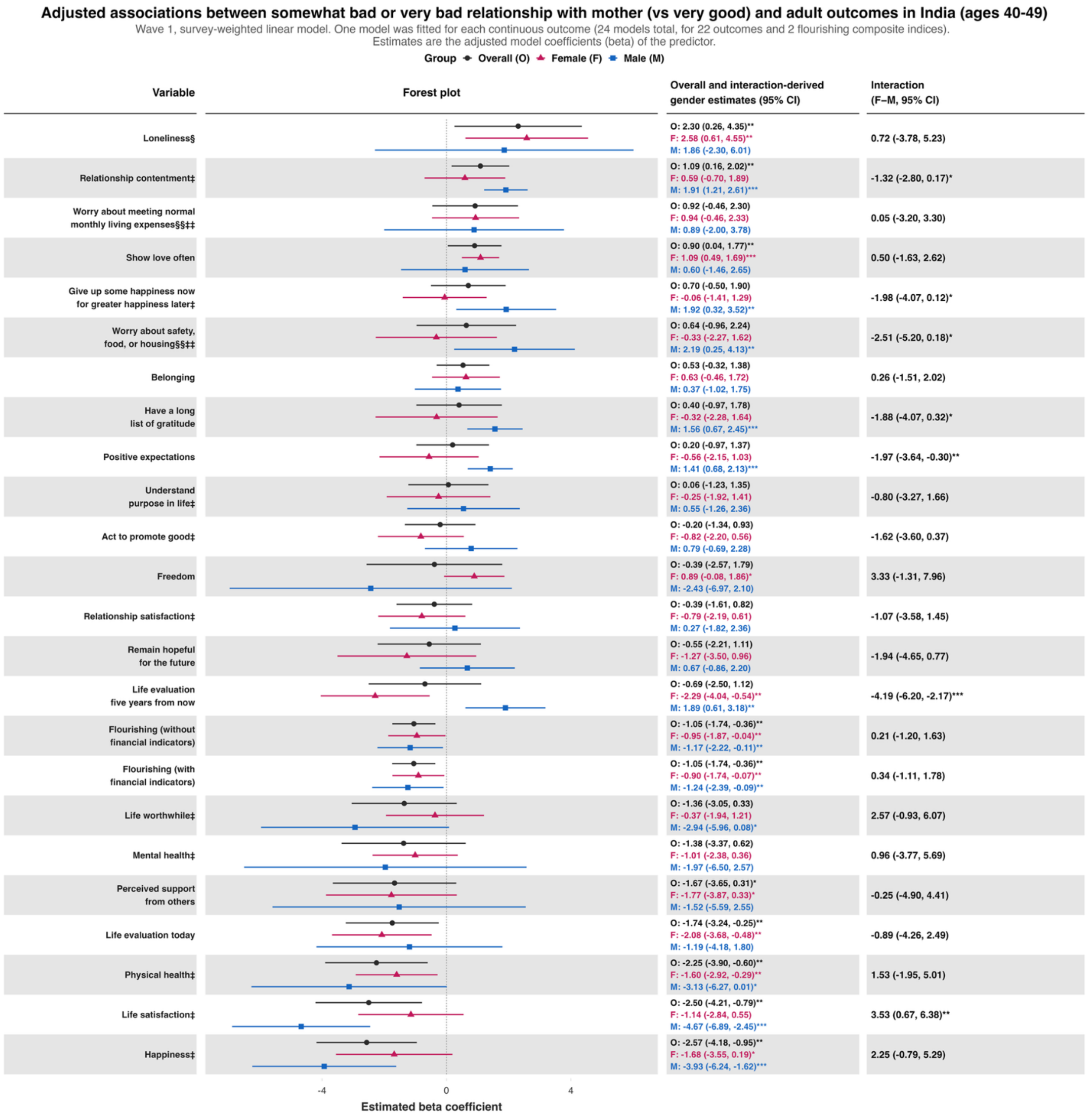

***Poorer relationship quality with mother (bad or very bad relationship) – Ages 50 to 59 inclusive***

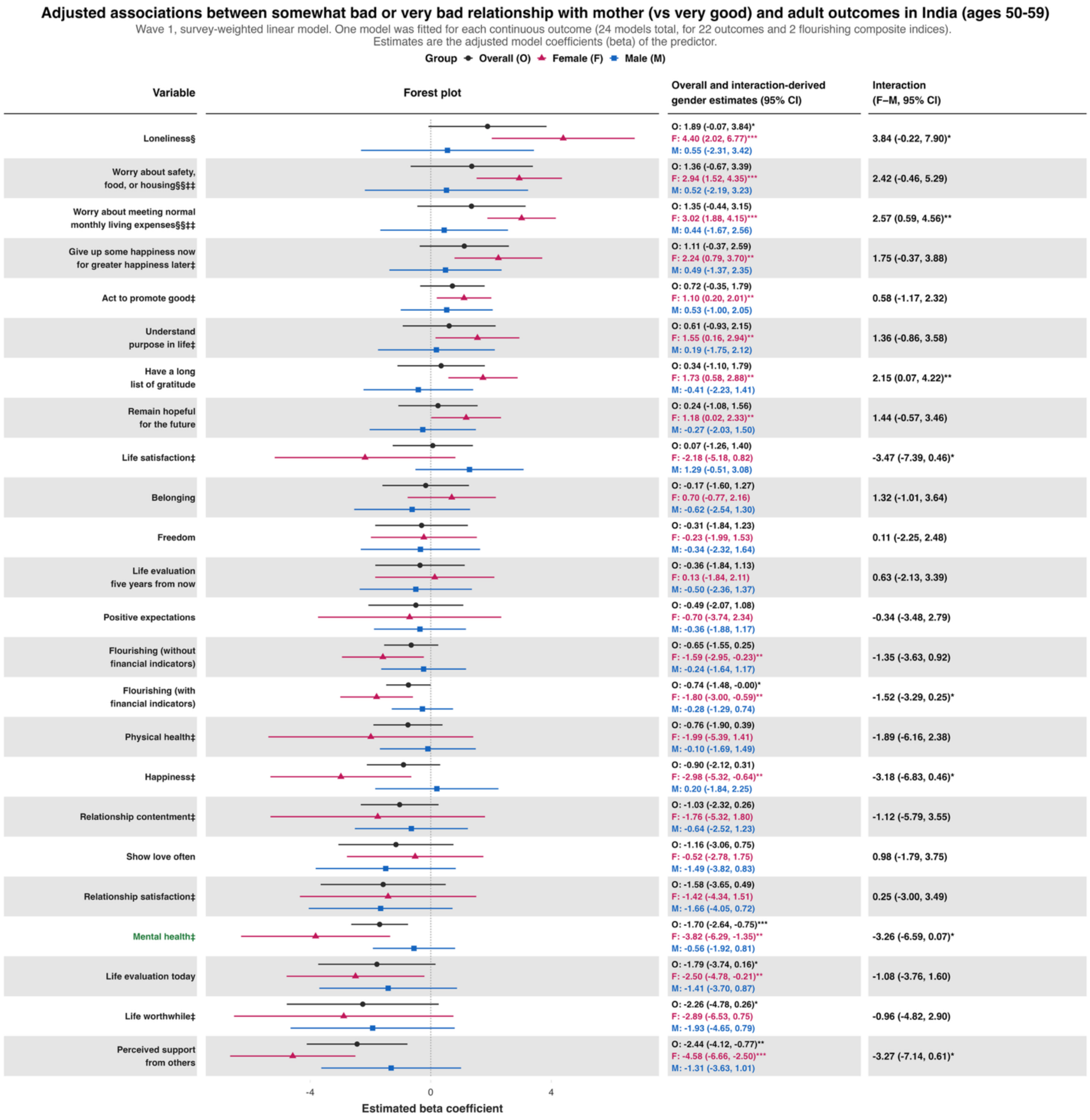

### *Poorer relationship quality with mother (bad or very bad relationship) – Ages 60+*

**Adjusted associations between somewhat bad or very bad relationship with mother (vs very good) and adult outcomes in India (ages 60+)**

Wave 1, survey-weighted linear model. One model was fitted for each continuous outcome (24 models total, for 22 outcomes and 2 flourishing composite indices). Estimates are the adjusted model coefficients (beta) of the predictor.

Group: Overall (O), Female (F), Male (M)

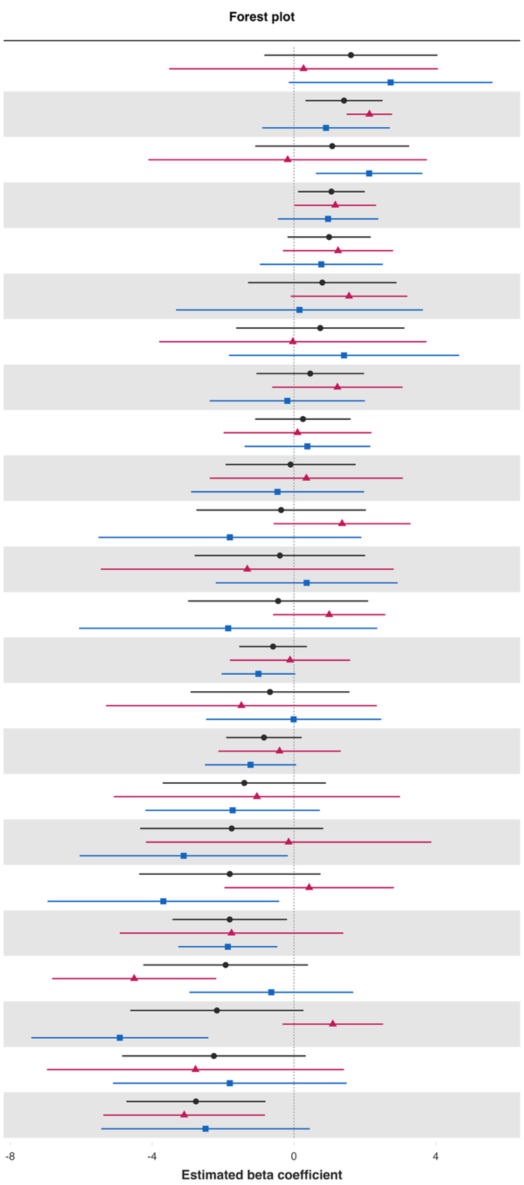


| Variable | Overall and interaction-derived gender estimates (95% CI) | Interaction (F–M, 95% CI) |
|---|---|---|
| Loneliness§ | O: 1.61 (-0.83, 4.05)<br>F: 0.27 (-3.52, 4.06)<br>M: 2.73 (-0.15, 5.60)* | -2.46 (-7.21, 2.30) |
| Positive expectations | O: 1.41 (0.32, 2.50)**<br>F: 2.13 (1.49, 2.77)***<br>M: 0.90 (-0.90, 2.70) | 1.22 (-0.68, 3.13) |
| Life worthwhile‡ | O: 1.08 (-1.09, 3.25)<br>F: -0.18 (-4.10, 3.75)<br>M: 2.12 (0.62, 3.63)** | -2.30 (-6.48, 1.88) |
| Show love often | O: 1.06 (0.11, 2.00)**<br>F: 1.17 (0.01, 2.32)**<br>M: 0.96 (-0.45, 2.38) | 0.20 (-1.60, 2.01) |
| Remain hopeful for the future | O: 0.99 (-0.18, 2.17)*<br>F: 1.24 (-0.31, 2.80)<br>M: 0.77 (-0.96, 2.51) | 0.47 (-1.88, 2.82) |
| Have a long list of gratitude | O: 0.80 (-1.29, 2.89)<br>F: 1.55 (-0.09, 3.20)*<br>M: 0.16 (-3.33, 3.64) | 1.40 (-2.50, 5.29) |
| Worry about safety, food, or housing§§‡‡ | O: 0.74 (-1.63, 3.12)<br>F: -0.03 (-3.80, 3.74)<br>M: 1.41 (-1.83, 4.66) | -1.44 (-6.48, 3.59) |
| Freedom | O: 0.46 (-1.06, 1.97)<br>F: 1.23 (-0.61, 3.06)<br>M: -0.19 (-2.38, 2.00) | 1.41 (-1.46, 4.29) |
| Act to promote good‡ | O: 0.25 (-1.09, 1.60)<br>F: 0.10 (-1.99, 2.19)<br>M: 0.38 (-1.39, 2.15) | -0.28 (-3.03, 2.48) |
| Understand purpose in life‡ | O: -0.09 (-1.93, 1.74)<br>F: 0.35 (-2.37, 3.07)<br>M: -0.47 (-2.91, 1.97) | 0.81 (-2.86, 4.49) |
| Give up some happiness now for greater happiness later‡ | O: -0.36 (-2.75, 2.03)<br>F: 1.36 (-0.58, 3.29)<br>M: -1.81 (-5.51, 1.90) | 3.16 (-1.12, 7.44) |
| Life satisfaction‡ | O: -0.40 (-2.80, 2.01)<br>F: -1.32 (-5.45, 2.81)<br>M: 0.36 (-2.21, 2.92) | -1.68 (-6.58, 3.23) |
| Belonging | O: -0.45 (-2.99, 2.09)<br>F: 0.99 (-0.59, 2.58)<br>M: -1.86 (-6.06, 2.35) | 2.85 (-1.59, 7.28) |
| Flourishing (with financial indicators) | O: -0.59 (-1.54, 0.37)<br>F: -0.11 (-1.80, 1.59)<br>M: -1.00 (-2.04, 0.04)* | 0.89 (-1.13, 2.92) |
| Mental health‡ | O: -0.68 (-2.92, 1.57)<br>F: -1.48 (-5.30, 2.33)<br>M: -0.01 (-2.48, 2.46) | -1.47 (-6.09, 3.14) |
| Flourishing (without financial indicators) | O: -0.85 (-1.91, 0.22)<br>F: -0.41 (-2.13, 1.32)<br>M: -1.22 (-2.51, 0.06)* | 0.82 (-1.37, 3.01) |
| Happiness‡ | O: -1.40 (-3.70, 0.90)<br>F: -1.04 (-5.08, 2.99)<br>M: -1.73 (-4.19, 0.73) | 0.68 (-4.02, 5.39) |
| Perceived support from others | O: -1.76 (-4.34, 0.83)<br>F: -0.15 (-4.17, 3.87)<br>M: -3.11 (-6.05, -0.17)** | 2.96 (-2.01, 7.93) |
| Relationship satisfaction‡ | O: -1.81 (-4.37, 0.75)<br>F: 0.43 (-1.96, 2.82)<br>M: -3.68 (-6.95, -0.42)** | 4.11 (0.05, 8.17)** |
| Life evaluation today | O: -1.81 (-3.43, -0.20)**<br>F: -1.76 (-4.91, 1.39)<br>M: -1.87 (-3.26, -0.47)** | 0.11 (-3.36, 3.58) |
| Life evaluation five years from now | O: -1.93 (-4.25, 0.39)<br>F: -4.51 (-6.82, -2.19)***<br>M: -0.64 (-2.95, 1.67) | -3.87 (-7.15, -0.59)** |
| Relationship contentment‡ | O: -2.17 (-4.62, 0.27)*<br>F: 1.09 (-0.32, 2.51)<br>M: -4.91 (-7.41, -2.41)*** | 6.01 (3.12, 8.89)*** |
| Worry about meeting normal monthly living expenses§§‡‡ | O: -2.26 (-4.85, 0.33)*<br>F: -2.78 (-6.97, 1.41)<br>M: -1.81 (-5.10, 1.49) | -0.97 (-6.31, 4.37) |
| Physical health‡ | O: -2.76 (-4.73, -0.80)**<br>F: -3.10 (-5.38, -0.82)**<br>M: -2.49 (-5.43, 0.45)* | -0.60 (-4.27, 3.06) |

**Predictor:** ***Poorer relationship quality with father (bad or very bad relationship)***
***Poorer relationship quality with father (bad or very bad relationship) – Ages 18 to 24 inclusive***

**Adjusted associations between somewhat bad or very bad relationship with father (vs very good) and adult outcomes in India (ages 18-24)**

Wave 1, survey-weighted linear model. One model was fitted for each continuous outcome (24 models total, for 22 outcomes and 2 flourishing composite indices).
Estimates are the adjusted model coefficients (beta) of the predictor.

**Group** ● Overall (O) ▲ Female (F) ■ Male (M)

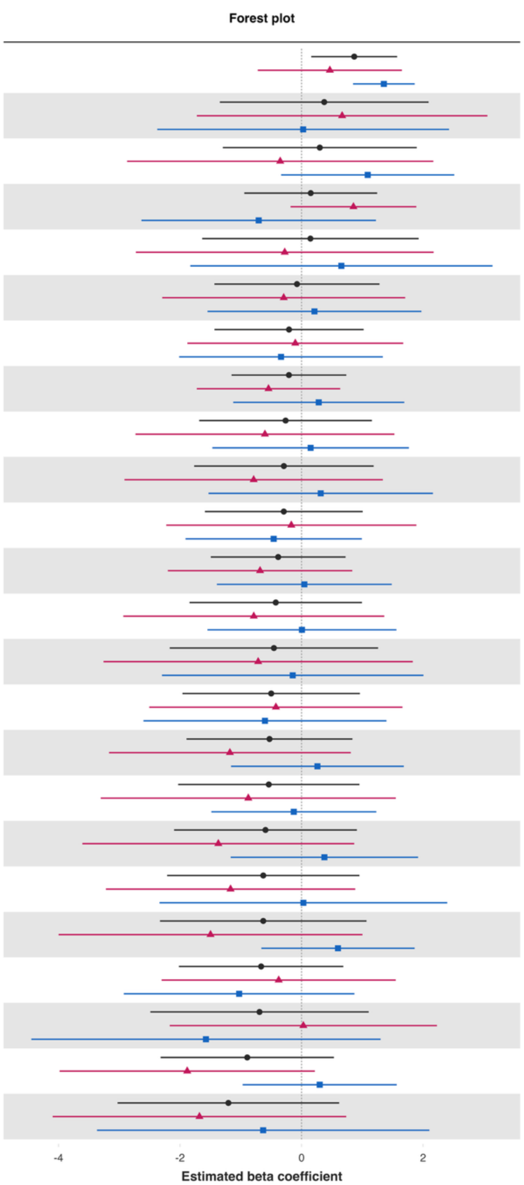


| Variable | Overall and interaction-derived gender estimates (95% CI) | Interaction (F–M, 95% CI) |
|---|---|---|
| Show love often | O: 0.87 (0.16, 1.57)**<br>F: 0.47 (-0.72, 1.65)<br>M: 1.35 (0.85, 1.86)*** | -0.89 (-2.18, 0.41) |
| Loneliness§ | O: 0.37 (-1.34, 2.09)<br>F: 0.67 (-1.72, 3.06)<br>M: 0.03 (-2.37, 2.43) | 0.64 (-2.72, 4.01) |
| Give up some happiness now for greater happiness later‡ | O: 0.30 (-1.30, 1.90)<br>F: -0.35 (-2.87, 2.17)<br>M: 1.09 (-0.34, 2.51) | -1.44 (-4.33, 1.45) |
| Life satisfaction‡ | O: 0.15 (-0.94, 1.24)<br>F: 0.85 (-0.18, 1.89)<br>M: -0.71 (-2.64, 1.22) | 1.56 (-0.62, 3.74) |
| Worry about meeting normal monthly living expenses§§‡‡ | O: 0.15 (-1.64, 1.93)<br>F: -0.28 (-2.73, 2.17)<br>M: 0.66 (-1.83, 3.14) | -0.93 (-4.44, 2.57) |
| Life evaluation five years from now | O: -0.08 (-1.43, 1.28)<br>F: -0.29 (-2.29, 1.71)<br>M: 0.21 (-1.55, 1.97) | -0.51 (-3.22, 2.21) |
| Mental health‡ | O: -0.21 (-1.43, 1.02)<br>F: -0.10 (-1.88, 1.67)<br>M: -0.34 (-2.01, 1.34) | 0.23 (-2.21, 2.68) |
| Flourishing (with financial indicators) | O: -0.21 (-1.15, 0.74)<br>F: -0.54 (-1.72, 0.64)<br>M: 0.28 (-1.12, 1.69) | -0.83 (-2.66, 1.00) |
| Relationship satisfaction‡ | O: -0.26 (-1.68, 1.16)<br>F: -0.60 (-2.73, 1.53)<br>M: 0.15 (-1.47, 1.77) | -0.75 (-3.42, 1.91) |
| Life worthwhile‡ | O: -0.29 (-1.77, 1.19)<br>F: -0.79 (-2.91, 1.34)<br>M: 0.32 (-1.53, 2.16) | -1.10 (-3.90, 1.69) |
| Belonging | O: -0.29 (-1.59, 1.01)<br>F: -0.17 (-2.23, 1.89)<br>M: -0.46 (-1.91, 0.99) | 0.29 (-2.22, 2.80) |
| Flourishing (without financial indicators) | O: -0.38 (-1.49, 0.72)<br>F: -0.68 (-2.20, 0.83)<br>M: 0.05 (-1.39, 1.49) | -0.73 (-2.81, 1.36) |
| Remain hopeful for the future | O: -0.42 (-1.84, 0.99)<br>F: -0.79 (-2.93, 1.36)<br>M: 0.01 (-1.55, 1.56) | -0.79 (-3.42, 1.83) |
| Life evaluation today | O: -0.46 (-2.17, 1.26)<br>F: -0.71 (-3.26, 1.83)<br>M: -0.15 (-2.30, 2.01) | -0.57 (-3.88, 2.75) |
| Happiness‡ | O: -0.50 (-1.96, 0.96)<br>F: -0.42 (-2.51, 1.66)<br>M: -0.60 (-2.60, 1.40) | 0.18 (-2.69, 3.05) |
| Positive expectations | O: -0.53 (-1.89, 0.84)<br>F: -1.18 (-3.17, 0.81)<br>M: 0.26 (-1.16, 1.68) | -1.44 (-3.88, 0.99) |
| Freedom | O: -0.54 (-2.03, 0.95)<br>F: -0.88 (-3.31, 1.55)<br>M: -0.13 (-1.48, 1.23) | -0.75 (-3.53, 2.03) |
| Understand purpose in life‡ | O: -0.59 (-2.10, 0.91)<br>F: -1.37 (-3.61, 0.87)<br>M: 0.38 (-1.17, 1.92) | -1.75 (-4.47, 0.97) |
| Have a long list of gratitude | O: -0.63 (-2.21, 0.95)<br>F: -1.17 (-3.22, 0.88)<br>M: 0.03 (-2.34, 2.40) | -1.20 (-4.31, 1.92) |
| Physical health‡ | O: -0.63 (-2.33, 1.07)<br>F: -1.50 (-4.00, 1.00)<br>M: 0.60 (-0.66, 1.86) | -2.10 (-4.88, 0.68) |
| Act to promote good‡ | O: -0.67 (-2.02, 0.69)<br>F: -0.38 (-2.30, 1.55)<br>M: -1.03 (-2.93, 0.87) | 0.65 (-2.04, 3.34) |
| Worry about safety, food, or housing§§‡‡ | O: -0.69 (-2.49, 1.10)<br>F: 0.03 (-2.17, 2.23)<br>M: -1.57 (-4.45, 1.30) | 1.60 (-2.01, 5.22) |
| Relationship contentment‡ | O: -0.89 (-2.32, 0.53)<br>F: -1.88 (-3.98, 0.22)*<br>M: 0.30 (-0.97, 1.57) | -2.18 (-4.63, 0.27)* |
| Perceived support from others | O: -1.20 (-3.03, 0.62)<br>F: -1.68 (-4.10, 0.74)<br>M: -0.63 (-3.37, 2.10) | -1.05 (-4.69, 2.60) |

*Poorer relationship quality with father (bad or very bad relationship) – Ages 25 to 29 inclusive*

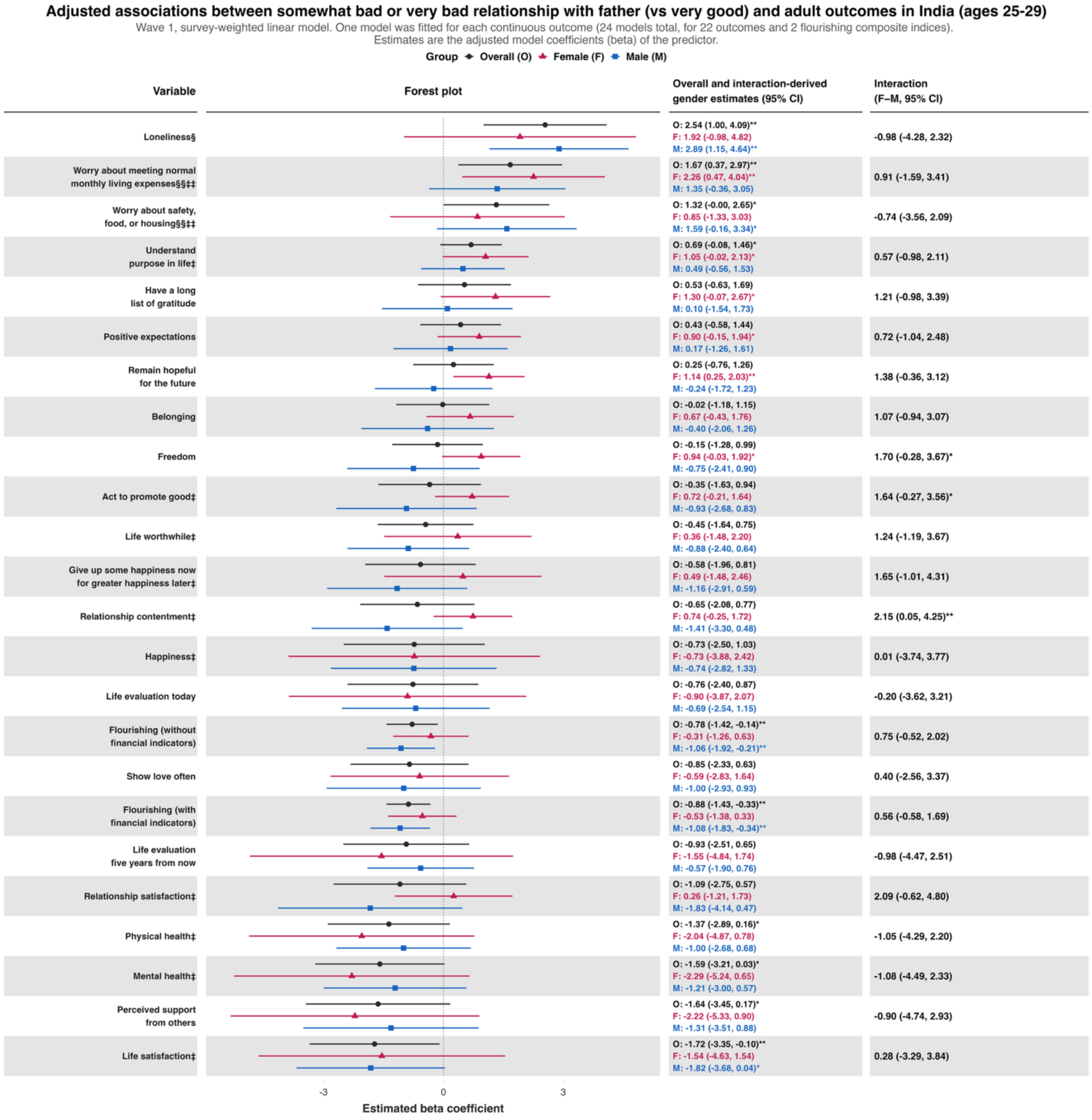

*Poorer relationship quality with father (bad or very bad relationship) – Ages 30 to 39 inclusive*

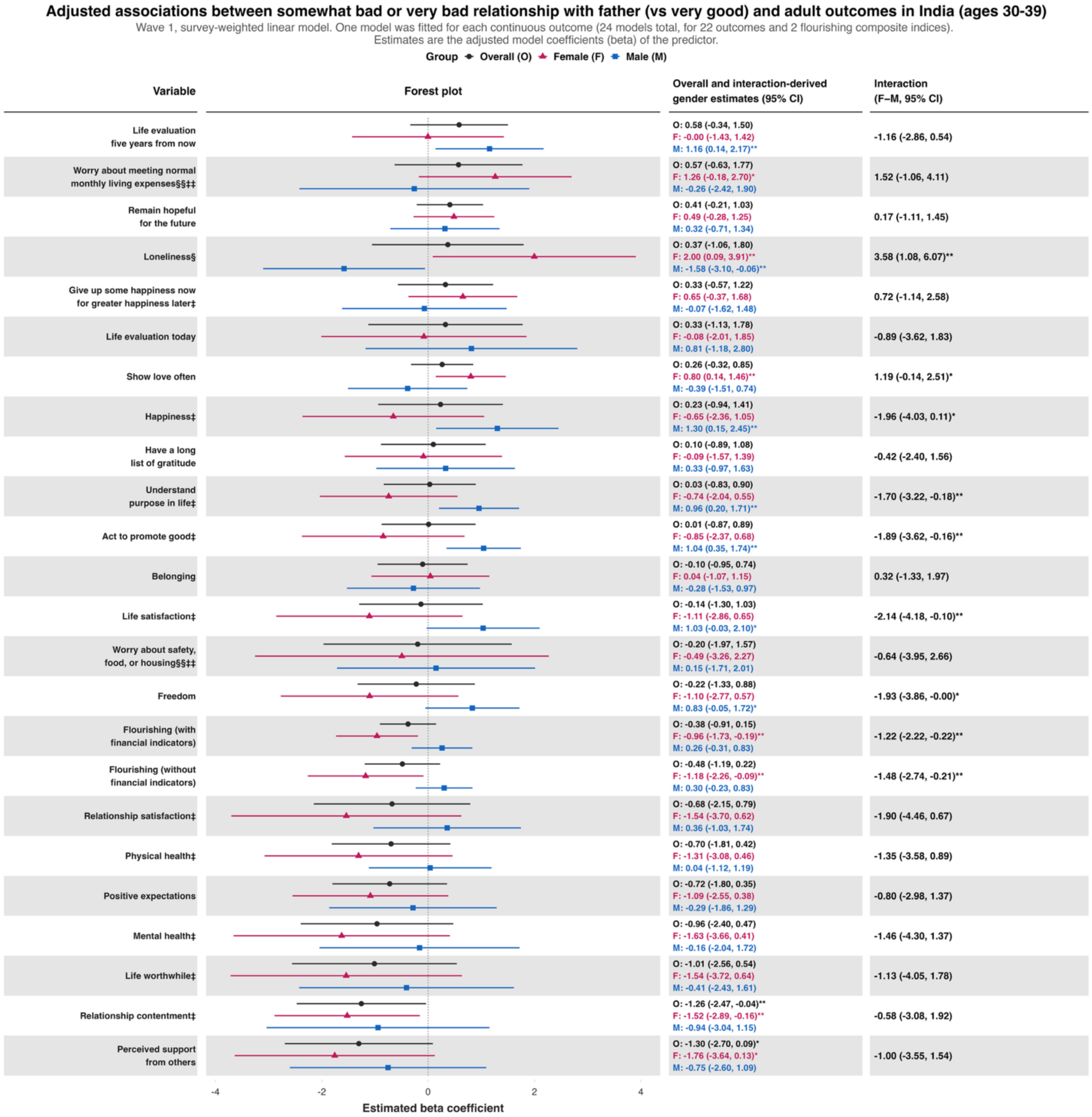

### *Poorer relationship quality with father (bad or very bad relationship) – Ages 40 to 49 inclusive*

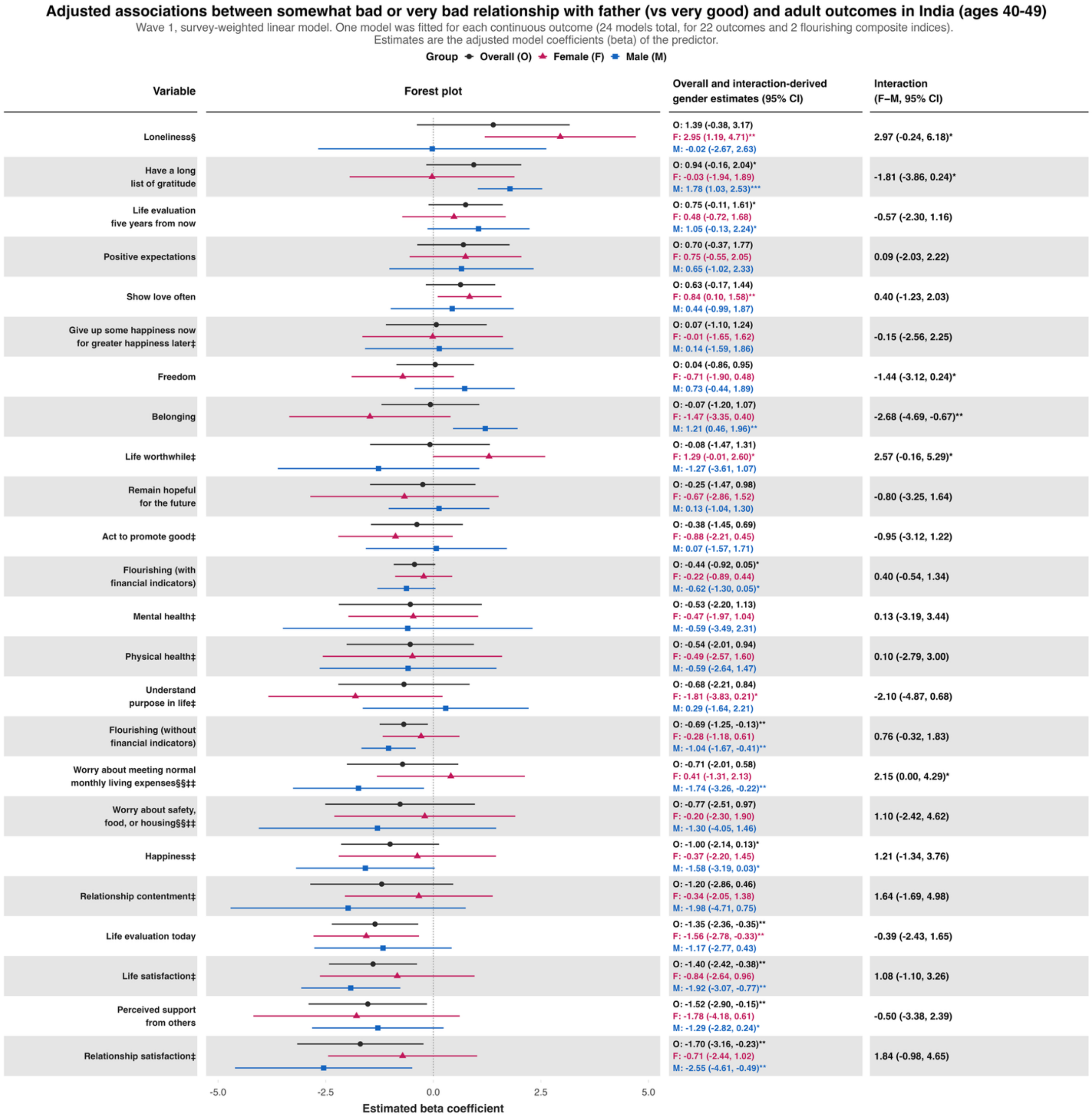

*Poorer relationship quality with father (bad or very bad relationship) – Ages 50 to 59 inclusive*

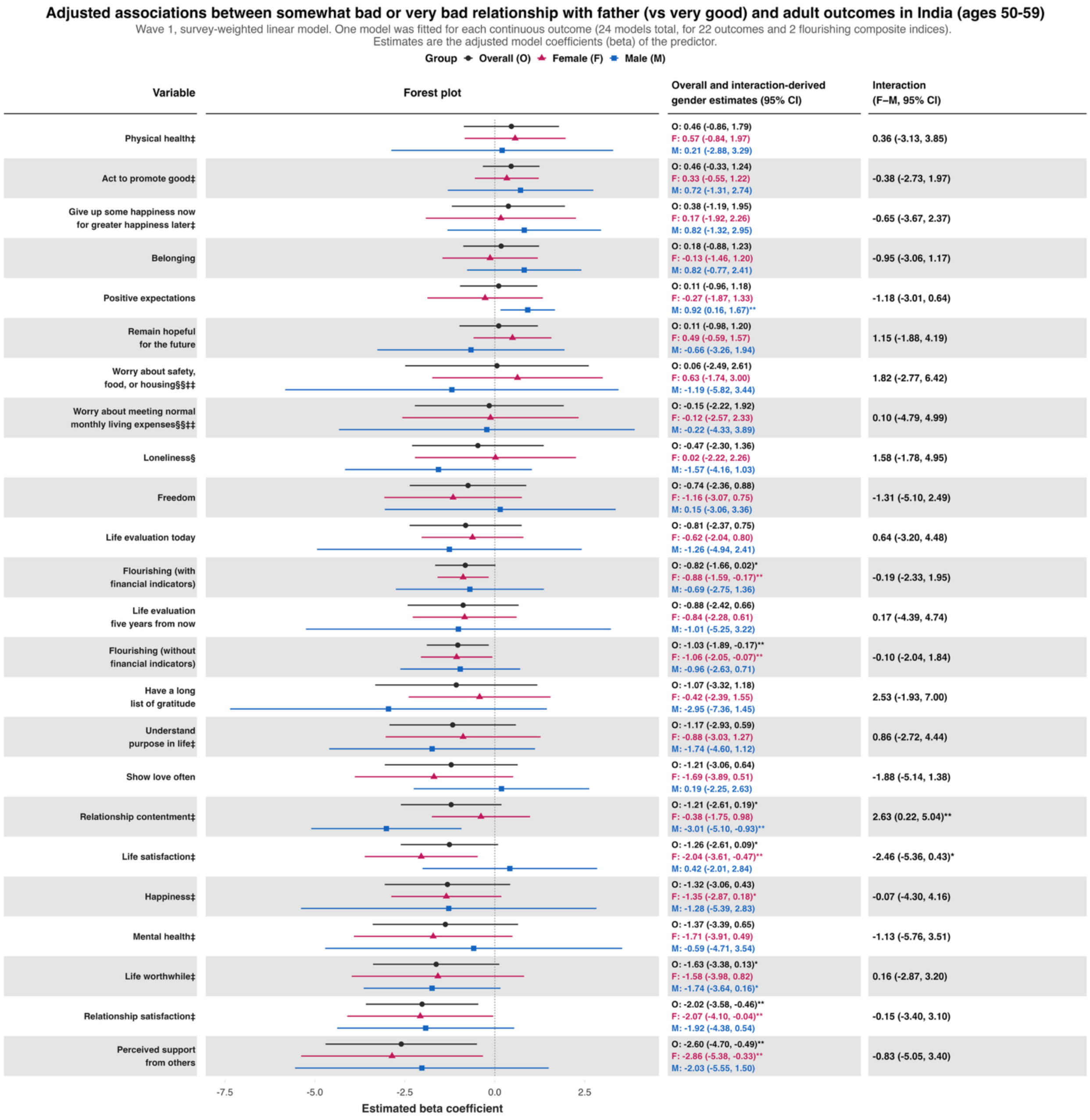

***Poorer relationship quality with father (bad or very bad relationship) – Ages 60+***

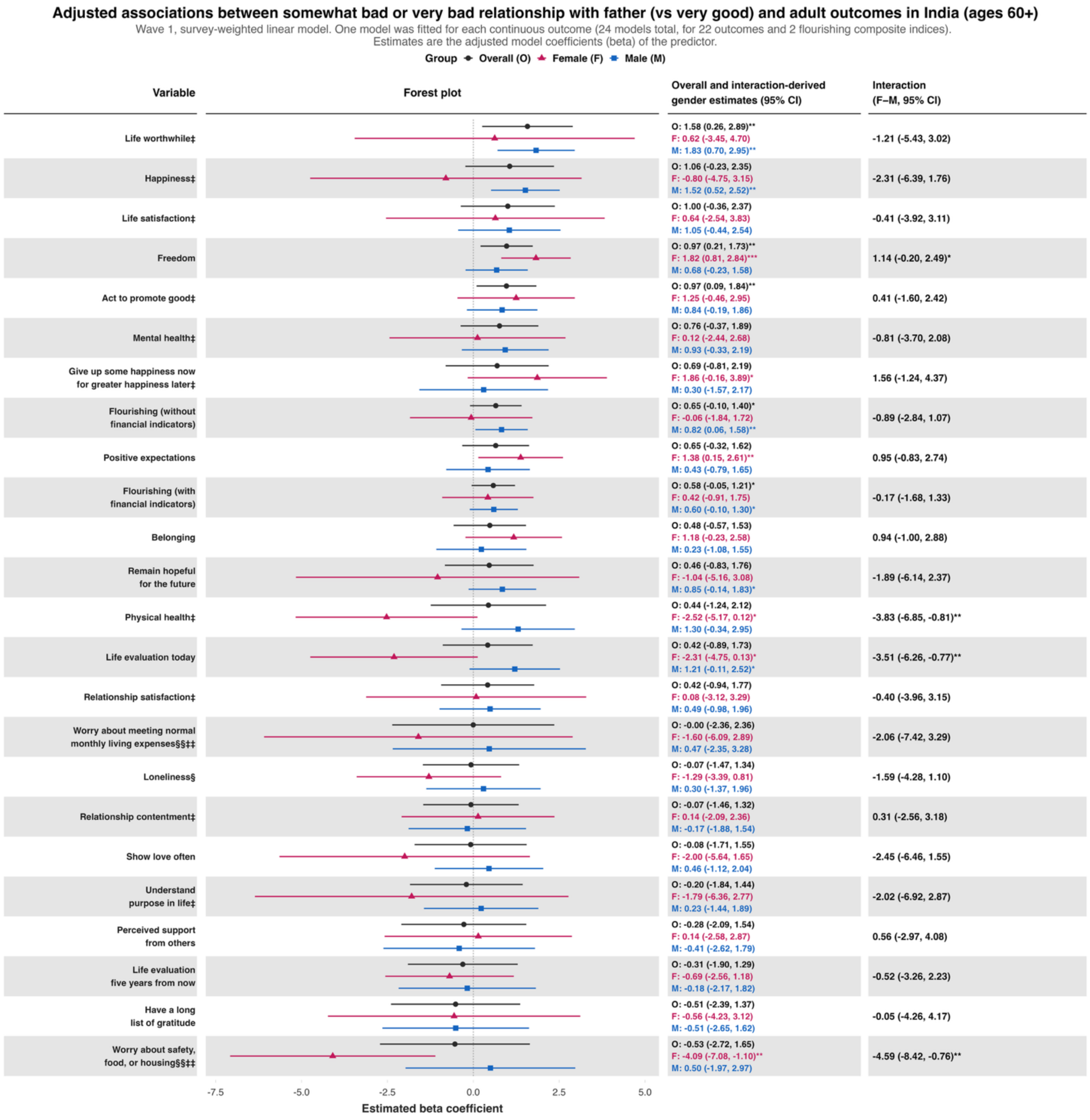

**Predictor:** ***Poor self-rated health while growing up***
***Poor self-rated health while growing up – Ages 18 to 24 inclusive***

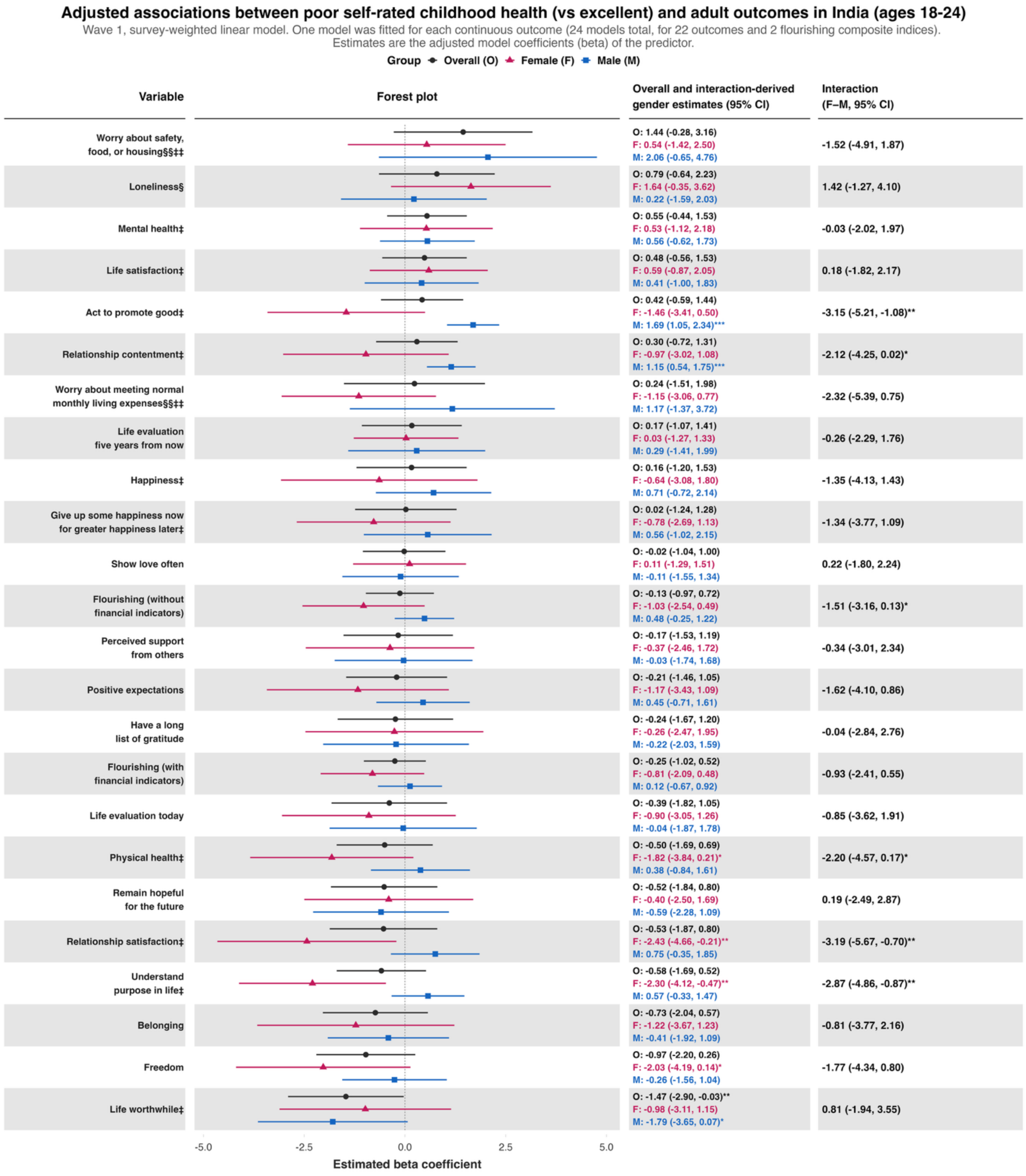

***Poor self-rated health while growing up – Ages 25 to 29 inclusive***

**Adjusted associations between poor self-rated childhood health (vs excellent) and adult outcomes in India (ages 25-29)**

Wave 1, survey-weighted linear model. One model was fitted for each continuous outcome (24 models total, for 22 outcomes and 2 flourishing composite indices).
Estimates are the adjusted model coefficients (beta) of the predictor.

Group: Overall (O), Female (F), Male (M)

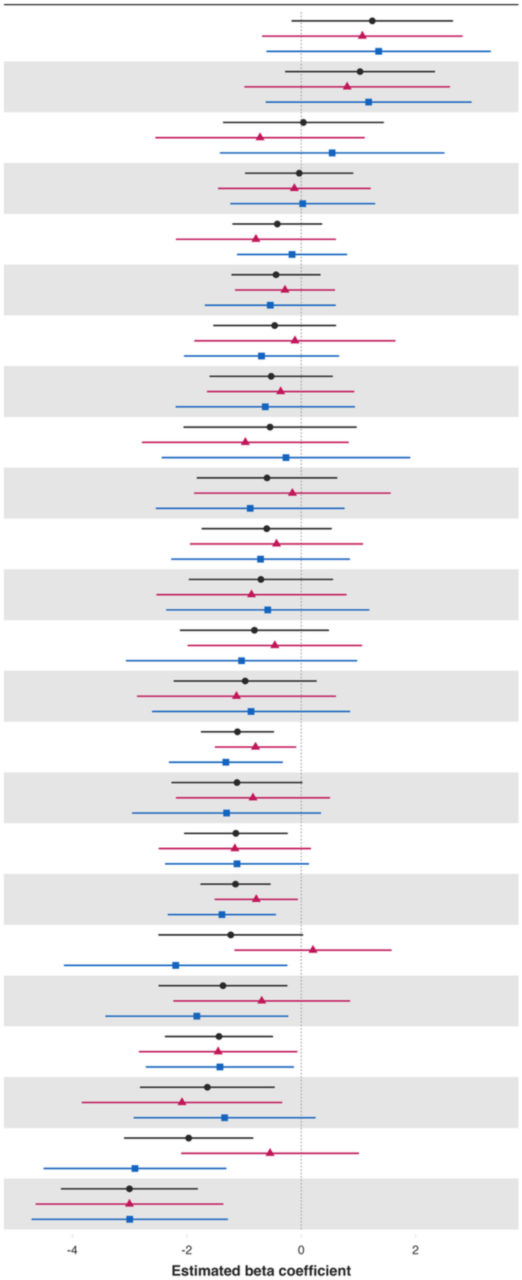


| Variable | Overall and interaction-derived gender estimates (95% CI) | Interaction (F–M, 95% CI) |
|---|---|---|
| Worry about meeting normal monthly living expenses§§‡‡ | O: 1.24 (-0.17, 2.65)*<br>F: 1.07 (-0.68, 2.82)<br>M: 1.35 (-0.61, 3.32) | -0.29 (-2.91, 2.33) |
| Worry about safety, food, or housing§§‡‡ | O: 1.03 (-0.28, 2.34)<br>F: 0.80 (-0.99, 2.60)<br>M: 1.18 (-0.62, 2.98) | -0.38 (-2.87, 2.12) |
| Loneliness§ | O: 0.04 (-1.37, 1.44)<br>F: -0.72 (-2.55, 1.11)<br>M: 0.54 (-1.42, 2.50) | -1.26 (-3.88, 1.35) |
| Positive expectations | O: -0.03 (-0.98, 0.91)<br>F: -0.12 (-1.46, 1.22)<br>M: 0.03 (-1.24, 1.29) | -0.15 (-1.97, 1.68) |
| Understand purpose in life‡ | O: -0.42 (-1.20, 0.37)<br>F: -0.79 (-2.19, 0.61)<br>M: -0.16 (-1.12, 0.80) | -0.63 (-2.35, 1.09) |
| Act to promote good‡ | O: -0.44 (-1.22, 0.34)<br>F: -0.28 (-1.16, 0.59)<br>M: -0.54 (-1.69, 0.60) | 0.26 (-1.17, 1.68) |
| Show love often | O: -0.46 (-1.54, 0.61)<br>F: -0.11 (-1.87, 1.65)<br>M: -0.69 (-2.05, 0.66) | 0.58 (-1.64, 2.81) |
| Relationship contentment‡ | O: -0.53 (-1.60, 0.55)<br>F: -0.36 (-1.65, 0.93)<br>M: -0.63 (-2.20, 0.94) | 0.27 (-1.75, 2.29) |
| Perceived support from others | O: -0.54 (-2.06, 0.97)<br>F: -0.98 (-2.78, 0.83)<br>M: -0.27 (-2.44, 1.91) | -0.71 (-3.56, 2.14) |
| Life evaluation today | O: -0.60 (-1.83, 0.63)<br>F: -0.15 (-1.87, 1.56)<br>M: -0.89 (-2.54, 0.76) | 0.74 (-1.59, 3.06) |
| Life worthwhile‡ | O: -0.60 (-1.74, 0.54)<br>F: -0.43 (-1.94, 1.08)<br>M: -0.71 (-2.27, 0.85) | 0.28 (-1.86, 2.42) |
| Belonging | O: -0.70 (-1.97, 0.56)<br>F: -0.87 (-2.53, 0.79)<br>M: -0.59 (-2.36, 1.19) | -0.28 (-2.73, 2.16) |
| Relationship satisfaction‡ | O: -0.82 (-2.12, 0.48)<br>F: -0.46 (-1.99, 1.06)<br>M: -1.04 (-3.06, 0.98) | 0.58 (-2.07, 3.23) |
| Give up some happiness now for greater happiness later‡ | O: -0.98 (-2.23, 0.27)<br>F: -1.13 (-2.87, 0.61)<br>M: -0.88 (-2.61, 0.85) | -0.26 (-2.71, 2.20) |
| Flourishing (with financial indicators) | O: -1.11 (-1.75, -0.48)***<br>F: -0.80 (-1.51, -0.09)**<br>M: -1.32 (-2.31, -0.32)** | 0.52 (-0.72, 1.76) |
| Freedom | O: -1.12 (-2.27, 0.02)*<br>F: -0.84 (-2.19, 0.50)<br>M: -1.31 (-2.96, 0.35) | 0.46 (-1.64, 2.57) |
| Life evaluation five years from now | O: -1.14 (-2.05, -0.24)**<br>F: -1.16 (-2.49, 0.17)*<br>M: -1.12 (-2.38, 0.14)* | -0.04 (-1.90, 1.83) |
| Flourishing (without financial indicators) | O: -1.15 (-1.76, -0.53)***<br>F: -0.79 (-1.51, -0.06)**<br>M: -1.39 (-2.33, -0.44)** | 0.60 (-0.62, 1.82) |
| Life satisfaction‡ | O: -1.23 (-2.50, 0.03)*<br>F: 0.21 (-1.17, 1.58)<br>M: -2.19 (-4.15, -0.24)** | 2.40 (0.01, 4.79)** |
| Happiness‡ | O: -1.37 (-2.49, -0.24)**<br>F: -0.69 (-2.23, 0.85)<br>M: -1.82 (-3.42, -0.23)** | 1.13 (-1.11, 3.38) |
| Remain hopeful for the future | O: -1.44 (-2.38, -0.49)**<br>F: -1.45 (-2.84, -0.07)**<br>M: -1.42 (-2.72, -0.13)** | -0.03 (-1.95, 1.88) |
| Have a long list of gratitude | O: -1.64 (-2.82, -0.46)**<br>F: -2.08 (-3.84, -0.33)**<br>M: -1.34 (-2.93, 0.25)* | -0.75 (-3.17, 1.68) |
| Mental health‡ | O: -1.97 (-3.10, -0.84)***<br>F: -0.54 (-2.10, 1.01)<br>M: -2.91 (-4.50, -1.31)*** | 2.36 (0.08, 4.64)** |
| Physical health‡ | O: -3.00 (-4.20, -1.81)***<br>F: -3.00 (-4.64, -1.36)***<br>M: -3.00 (-4.71, -1.28)*** | -0.01 (-2.43, 2.42) |

*Poor self-rated health while growing up – Ages 30 to 39 inclusive*

### Adjusted associations between poor self-rated childhood health (vs excellent) and adult outcomes in India (ages 30-39)

Wave 1, survey-weighted linear model. One model was fitted for each continuous outcome (24 models total, for 22 outcomes and 2 flourishing composite indices).
Estimates are the adjusted model coefficients (beta) of the predictor.

**Group** ● Overall (O) ▲ Female (F) ■ Male (M)

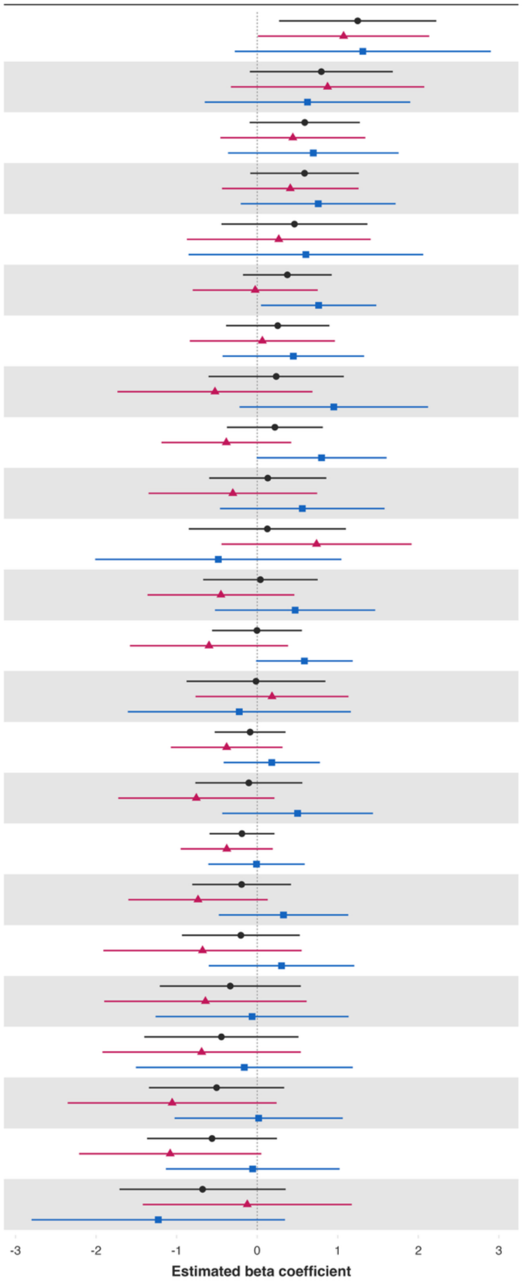


| Variable | Overall and interaction-derived gender estimates (95% CI) | Interaction (F–M, 95% CI) |
|---|---|---|
| Worry about safety, food, or housing§§‡‡ | O: 1.25 (0.27, 2.22)**<br>F: 1.07 (0.01, 2.14)**<br>M: 1.31 (-0.28, 2.90) | -0.24 (-2.10, 1.63) |
| Worry about meeting normal monthly living expenses§§‡‡ | O: 0.80 (-0.09, 1.68)*<br>F: 0.87 (-0.33, 2.07)<br>M: 0.62 (-0.65, 1.90) | 0.25 (-1.48, 1.98) |
| Relationship satisfaction‡ | O: 0.59 (-0.09, 1.27)*<br>F: 0.44 (-0.46, 1.34)<br>M: 0.70 (-0.36, 1.76) | -0.25 (-1.67, 1.16) |
| Have a long list of gratitude | O: 0.59 (-0.09, 1.26)*<br>F: 0.41 (-0.44, 1.26)<br>M: 0.76 (-0.20, 1.72) | -0.35 (-1.57, 0.87) |
| Perceived support from others | O: 0.46 (-0.44, 1.37)<br>F: 0.27 (-0.87, 1.41)<br>M: 0.60 (-0.85, 2.06) | -0.34 (-2.23, 1.56) |
| Freedom | O: 0.37 (-0.18, 0.93)<br>F: -0.03 (-0.80, 0.75)<br>M: 0.76 (0.05, 1.48)** | -0.79 (-1.82, 0.25) |
| Show love often | O: 0.25 (-0.39, 0.90)<br>F: 0.06 (-0.84, 0.96)<br>M: 0.45 (-0.43, 1.33) | -0.38 (-1.62, 0.85) |
| Give up some happiness now for greater happiness later‡ | O: 0.24 (-0.60, 1.07)<br>F: -0.52 (-1.74, 0.69)<br>M: 0.95 (-0.22, 2.12) | -1.48 (-3.19, 0.24)* |
| Remain hopeful for the future | O: 0.22 (-0.38, 0.81)<br>F: -0.38 (-1.19, 0.42)<br>M: 0.80 (-0.01, 1.61)* | -1.18 (-2.33, -0.03)** |
| Mental health‡ | O: 0.13 (-0.60, 0.86)<br>F: -0.30 (-1.35, 0.74)<br>M: 0.56 (-0.46, 1.58) | -0.86 (-2.34, 0.61) |
| Loneliness§ | O: 0.13 (-0.85, 1.10)<br>F: 0.74 (-0.44, 1.92)<br>M: -0.48 (-2.01, 1.05) | 1.22 (-0.70, 3.14) |
| Life evaluation five years from now | O: 0.04 (-0.67, 0.75)<br>F: -0.45 (-1.36, 0.46)<br>M: 0.47 (-0.52, 1.47) | -0.92 (-2.21, 0.37) |
| Belonging | O: -0.00 (-0.56, 0.55)<br>F: -0.60 (-1.58, 0.38)<br>M: 0.59 (-0.01, 1.18)* | -1.18 (-2.39, 0.02)* |
| Relationship contentment‡ | O: -0.02 (-0.88, 0.85)<br>F: 0.18 (-0.77, 1.13)<br>M: -0.22 (-1.61, 1.16) | 0.41 (-1.24, 2.05) |
| Flourishing (without financial indicators) | O: -0.09 (-0.53, 0.35)<br>F: -0.38 (-1.07, 0.31)<br>M: 0.18 (-0.42, 0.78) | -0.56 (-1.51, 0.39) |
| Positive expectations | O: -0.10 (-0.77, 0.56)<br>F: -0.76 (-1.72, 0.21)<br>M: 0.50 (-0.43, 1.44) | -1.26 (-2.66, 0.14)* |
| Flourishing (with financial indicators) | O: -0.19 (-0.59, 0.21)<br>F: -0.38 (-0.95, 0.19)<br>M: -0.01 (-0.61, 0.59) | -0.37 (-1.21, 0.48) |
| Act to promote good‡ | O: -0.19 (-0.81, 0.42)<br>F: -0.73 (-1.60, 0.13)*<br>M: 0.33 (-0.48, 1.13) | -1.06 (-2.22, 0.10)* |
| Understand purpose in life‡ | O: -0.20 (-0.93, 0.53)<br>F: -0.68 (-1.91, 0.55)<br>M: 0.30 (-0.60, 1.20) | -0.98 (-2.57, 0.61) |
| Life satisfaction‡ | O: -0.33 (-1.21, 0.54)<br>F: -0.64 (-1.90, 0.61)<br>M: -0.06 (-1.26, 1.14) | -0.58 (-2.31, 1.15) |
| Life evaluation today | O: -0.44 (-1.40, 0.51)<br>F: -0.69 (-1.92, 0.54)<br>M: -0.16 (-1.51, 1.19) | -0.53 (-2.27, 1.21) |
| Happiness‡ | O: -0.50 (-1.34, 0.34)<br>F: -1.06 (-2.35, 0.24)<br>M: 0.02 (-1.03, 1.06) | -1.07 (-2.73, 0.59) |
| Physical health‡ | O: -0.56 (-1.37, 0.24)<br>F: -1.08 (-2.21, 0.05)*<br>M: -0.06 (-1.13, 1.02) | -1.02 (-2.55, 0.50) |
| Life worthwhile‡ | O: -0.68 (-1.71, 0.35)<br>F: -0.12 (-1.42, 1.17)<br>M: -1.23 (-2.80, 0.34) | 1.10 (-0.95, 3.16) |

### *Poor self-rated health while growing up – Ages 40 to 49 inclusive*

#### Adjusted associations between poor self-rated childhood health (vs excellent) and adult outcomes in India (ages 40-49)

Wave 1, survey-weighted linear model. One model was fitted for each continuous outcome (24 models total, for 22 outcomes and 2 flourishing composite indices). Estimates are the adjusted model coefficients (beta) of the predictor.

Group: Overall (O), Female (F), Male (M)

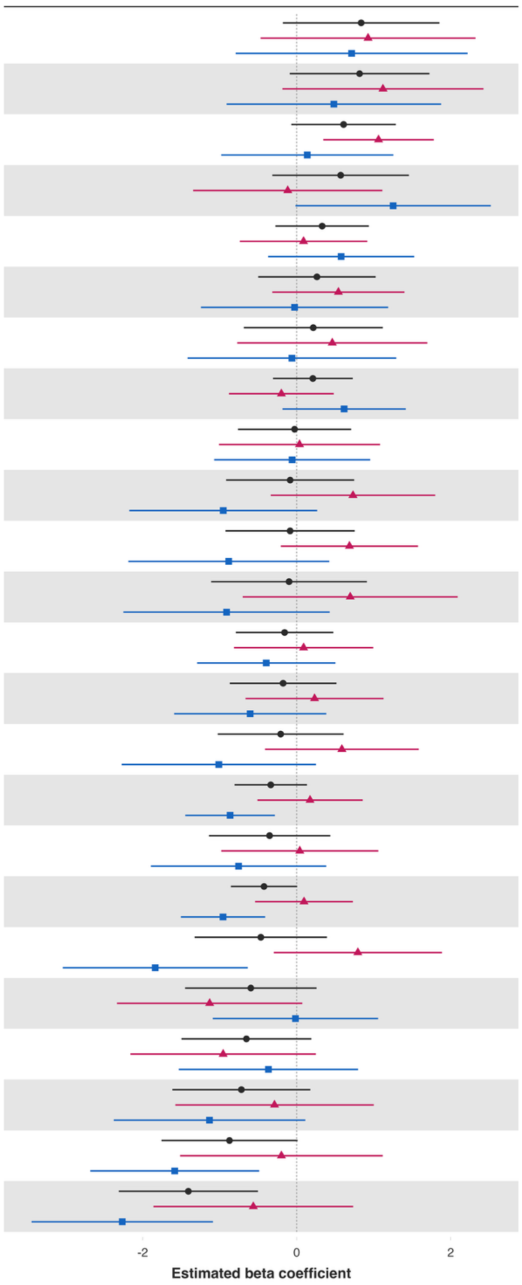


| Variable | Overall and interaction-derived gender estimates (95% CI) | Interaction (F–M, 95% CI) |
|---|---|---|
| Worry about meeting normal monthly living expenses§§‡‡ | O: 0.84 (-0.18, 1.86)<br>F: 0.93 (-0.47, 2.32)<br>M: 0.71 (-0.79, 2.22) | 0.21 (-1.88, 2.31) |
| Loneliness§ | O: 0.82 (-0.09, 1.72)*<br>F: 1.12 (-0.19, 2.43)*<br>M: 0.48 (-0.91, 1.88) | 0.64 (-1.36, 2.63) |
| Show love often | O: 0.61 (-0.07, 1.29)*<br>F: 1.06 (0.35, 1.78)**<br>M: 0.14 (-0.98, 1.26) | 0.93 (-0.35, 2.20) |
| Worry about safety, food, or housing§§‡‡ | O: 0.57 (-0.32, 1.46)<br>F: -0.12 (-1.34, 1.11)<br>M: 1.25 (-0.02, 2.52)* | -1.37 (-3.12, 0.38) |
| Belonging | O: 0.33 (-0.28, 0.94)<br>F: 0.09 (-0.74, 0.92)<br>M: 0.58 (-0.37, 1.53) | -0.49 (-1.80, 0.82) |
| Relationship contentment‡ | O: 0.26 (-0.50, 1.03)<br>F: 0.54 (-0.32, 1.40)<br>M: -0.03 (-1.24, 1.19) | 0.57 (-0.88, 2.02) |
| Give up some happiness now for greater happiness later‡ | O: 0.22 (-0.69, 1.12)<br>F: 0.46 (-0.77, 1.70)<br>M: -0.06 (-1.42, 1.29) | 0.52 (-1.32, 2.36) |
| Act to promote good‡ | O: 0.21 (-0.31, 0.73)<br>F: -0.20 (-0.88, 0.48)<br>M: 0.62 (-0.18, 1.42) | -0.82 (-1.89, 0.26) |
| Understand purpose in life‡ | O: -0.03 (-0.76, 0.71)<br>F: 0.04 (-1.01, 1.08)<br>M: -0.06 (-1.07, 0.96) | 0.10 (-1.36, 1.55) |
| Life worthwhile‡ | O: -0.08 (-0.92, 0.75)<br>F: 0.73 (-0.34, 1.80)<br>M: -0.95 (-2.17, 0.27) | 1.69 (0.04, 3.33)** |
| Relationship satisfaction‡ | O: -0.09 (-0.92, 0.75)<br>F: 0.69 (-0.21, 1.58)<br>M: -0.88 (-2.19, 0.42) | 1.57 (-0.02, 3.16)* |
| Perceived support from others | O: -0.10 (-1.11, 0.91)<br>F: 0.70 (-0.70, 2.09)<br>M: -0.91 (-2.25, 0.43) | 1.61 (-0.28, 3.50)* |
| Remain hopeful for the future | O: -0.16 (-0.79, 0.48)<br>F: 0.09 (-0.81, 0.99)<br>M: -0.39 (-1.29, 0.50) | 0.48 (-0.80, 1.77) |
| Freedom | O: -0.18 (-0.87, 0.52)<br>F: 0.23 (-0.66, 1.13)<br>M: -0.60 (-1.59, 0.38) | 0.84 (-0.45, 2.13) |
| Positive expectations | O: -0.21 (-1.03, 0.61)<br>F: 0.59 (-0.41, 1.59)<br>M: -1.01 (-2.27, 0.25) | 1.60 (0.00, 3.19)* |
| Flourishing (without financial indicators) | O: -0.34 (-0.81, 0.13)<br>F: 0.17 (-0.51, 0.86)<br>M: -0.86 (-1.45, -0.28)** | 1.04 (0.15, 1.93)** |
| Have a long list of gratitude | O: -0.35 (-1.14, 0.44)<br>F: 0.04 (-0.98, 1.06)<br>M: -0.75 (-1.89, 0.38) | 0.79 (-0.67, 2.26) |
| Flourishing (with financial indicators) | O: -0.42 (-0.86, 0.01)*<br>F: 0.10 (-0.54, 0.73)<br>M: -0.96 (-1.50, -0.41)*** | 1.05 (0.22, 1.89)** |
| Life satisfaction‡ | O: -0.47 (-1.32, 0.39)<br>F: 0.79 (-0.30, 1.89)<br>M: -1.84 (-3.04, -0.63)** | 2.63 (1.02, 4.25)** |
| Life evaluation today | O: -0.60 (-1.45, 0.26)<br>F: -1.13 (-2.33, 0.07)*<br>M: -0.02 (-1.09, 1.06) | -1.11 (-2.70, 0.47) |
| Life evaluation five years from now | O: -0.65 (-1.50, 0.19)<br>F: -0.95 (-2.16, 0.25)<br>M: -0.37 (-1.53, 0.80) | -0.59 (-2.28, 1.10) |
| Mental health‡ | O: -0.72 (-1.61, 0.18)<br>F: -0.29 (-1.58, 1.00)<br>M: -1.13 (-2.38, 0.11)* | 0.84 (-0.95, 2.64) |
| Happiness‡ | O: -0.87 (-1.76, 0.01)*<br>F: -0.20 (-1.51, 1.12)<br>M: -1.58 (-2.68, -0.49)** | 1.39 (-0.29, 3.06) |
| Physical health‡ | O: -1.41 (-2.31, -0.50)**<br>F: -0.56 (-1.86, 0.73)<br>M: -2.27 (-3.44, -1.09)*** | 1.70 (-0.04, 3.44)* |

### *Poor self-rated health while growing up – Ages 50 to 59 inclusive*

#### Adjusted associations between poor self-rated childhood health (vs excellent) and adult outcomes in India (ages 50-59)

Wave 1, survey-weighted linear model. One model was fitted for each continuous outcome (24 models total, for 22 outcomes and 2 flourishing composite indices).
Estimates are the adjusted model coefficients (beta) of the predictor.

Group — Overall (O) — Female (F) — Male (M)

Forest plot

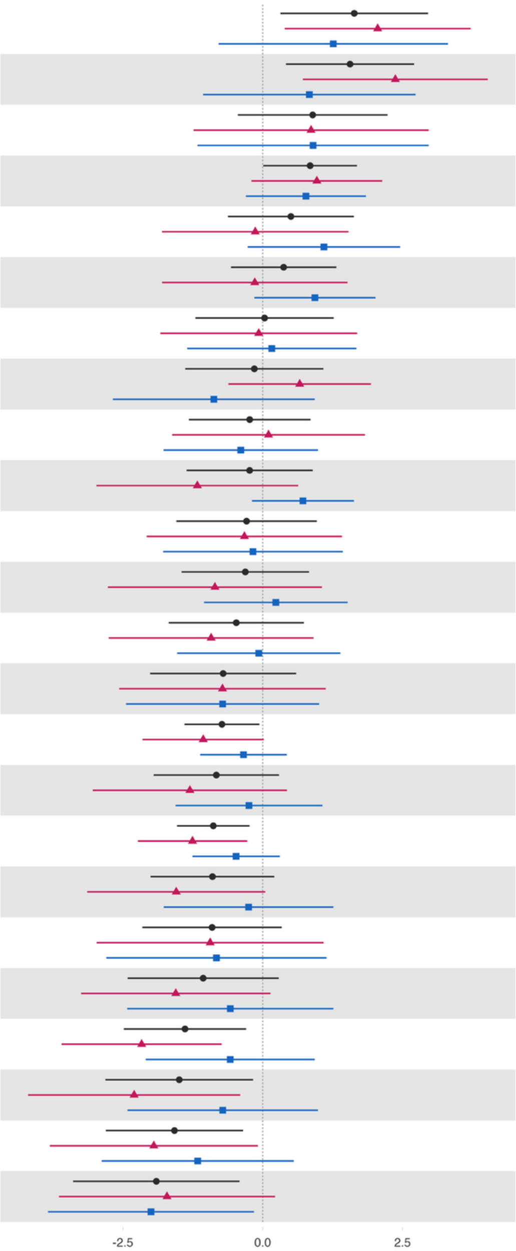


| Variable | Overall and interaction-derived gender estimates (95% CI) | Interaction (F–M, 95% CI) |
|---|---|---|
| Worry about meeting normal monthly living expenses§§‡‡ | O: 1.64 (0.32, 2.96)**<br>F: 2.05 (0.39, 3.72)**<br>M: 1.26 (-0.79, 3.31) | 0.79 (-1.85, 3.44) |
| Worry about safety, food, or housing§§‡‡ | O: 1.56 (0.42, 2.71)**<br>F: 2.37 (0.72, 4.03)**<br>M: 0.83 (-1.07, 2.73) | 1.54 (-1.19, 4.26) |
| Loneliness§ | O: 0.89 (-0.44, 2.23)<br>F: 0.86 (-1.24, 2.97)<br>M: 0.90 (-1.17, 2.97) | -0.04 (-3.22, 3.15) |
| Positive expectations | O: 0.85 (0.01, 1.69)**<br>F: 0.97 (-0.20, 2.14)<br>M: 0.77 (-0.30, 1.85) | 0.20 (-1.27, 1.66) |
| Have a long list of gratitude | O: 0.50 (-0.62, 1.63)<br>F: -0.13 (-1.80, 1.53)<br>M: 1.09 (-0.27, 2.46) | -1.23 (-3.27, 0.81) |
| Relationship contentment‡ | O: 0.37 (-0.57, 1.32)<br>F: -0.14 (-1.80, 1.52)<br>M: 0.93 (-0.15, 2.02)* | -1.08 (-3.13, 0.98) |
| Show love often | O: 0.03 (-1.20, 1.27)<br>F: -0.07 (-1.83, 1.69)<br>M: 0.16 (-1.35, 1.67) | -0.23 (-2.37, 1.90) |
| Understand purpose in life‡ | O: -0.15 (-1.39, 1.09)<br>F: 0.66 (-0.61, 1.93)<br>M: -0.87 (-2.68, 0.93) | 1.54 (-0.40, 3.47) |
| Give up some happiness now for greater happiness later‡ | O: -0.23 (-1.32, 0.86)<br>F: 0.10 (-1.62, 1.83)<br>M: -0.39 (-1.77, 0.99) | 0.50 (-1.73, 2.72) |
| Act to promote good‡ | O: -0.23 (-1.36, 0.89)<br>F: -1.17 (-2.97, 0.63)<br>M: 0.72 (-0.19, 1.63) | -1.89 (-3.72, -0.06)** |
| Remain hopeful for the future | O: -0.29 (-1.55, 0.97)<br>F: -0.33 (-2.07, 1.42)<br>M: -0.18 (-1.78, 1.43) | -0.15 (-2.34, 2.04) |
| Life evaluation five years from now | O: -0.31 (-1.45, 0.83)<br>F: -0.86 (-2.77, 1.06)<br>M: 0.23 (-1.05, 1.52) | -1.09 (-3.37, 1.19) |
| Relationship satisfaction‡ | O: -0.47 (-1.68, 0.74)<br>F: -0.92 (-2.75, 0.91)<br>M: -0.07 (-1.53, 1.39) | -0.85 (-3.07, 1.36) |
| Belonging | O: -0.71 (-2.01, 0.60)<br>F: -0.72 (-2.57, 1.13)<br>M: -0.72 (-2.44, 1.01) | -0.00 (-2.44, 2.43) |
| Flourishing (without financial indicators) | O: -0.73 (-1.40, -0.06)**<br>F: -1.06 (-2.15, 0.02)*<br>M: -0.34 (-1.12, 0.43) | -0.72 (-2.03, 0.59) |
| Freedom | O: -0.83 (-1.95, 0.29)<br>F: -1.30 (-3.04, 0.43)<br>M: -0.25 (-1.56, 1.07) | -1.06 (-3.16, 1.05) |
| Flourishing (with financial indicators) | O: -0.88 (-1.53, -0.24)**<br>F: -1.26 (-2.23, -0.28)**<br>M: -0.48 (-1.26, 0.30) | -0.78 (-1.98, 0.42) |
| Happiness‡ | O: -0.90 (-2.01, 0.21)<br>F: -1.55 (-3.14, 0.05)*<br>M: -0.25 (-1.77, 1.26) | -1.29 (-3.50, 0.91) |
| Perceived support from others | O: -0.91 (-2.15, 0.34)<br>F: -0.94 (-2.97, 1.09)<br>M: -0.83 (-2.80, 1.14) | -0.11 (-3.24, 3.01) |
| Life worthwhile‡ | O: -1.07 (-2.42, 0.29)<br>F: -1.56 (-3.25, 0.14)*<br>M: -0.58 (-2.42, 1.26) | -0.98 (-3.32, 1.37) |
| Physical health‡ | O: -1.39 (-2.48, -0.30)**<br>F: -2.17 (-3.60, -0.73)**<br>M: -0.58 (-2.09, 0.93) | -1.58 (-3.55, 0.39) |
| Life satisfaction‡ | O: -1.49 (-2.81, -0.17)**<br>F: -2.30 (-4.20, -0.40)**<br>M: -0.72 (-2.42, 0.99) | -1.58 (-4.00, 0.83) |
| Mental health‡ | O: -1.58 (-2.81, -0.35)**<br>F: -1.95 (-3.81, -0.09)**<br>M: -1.16 (-2.88, 0.56) | -0.79 (-3.34, 1.76) |
| Life evaluation today | O: -1.90 (-3.39, -0.41)**<br>F: -1.71 (-3.64, 0.22)*<br>M: -2.00 (-3.84, -0.16)** | 0.29 (-2.06, 2.63) |

*Poor self-rated health while growing up – Ages 60+*

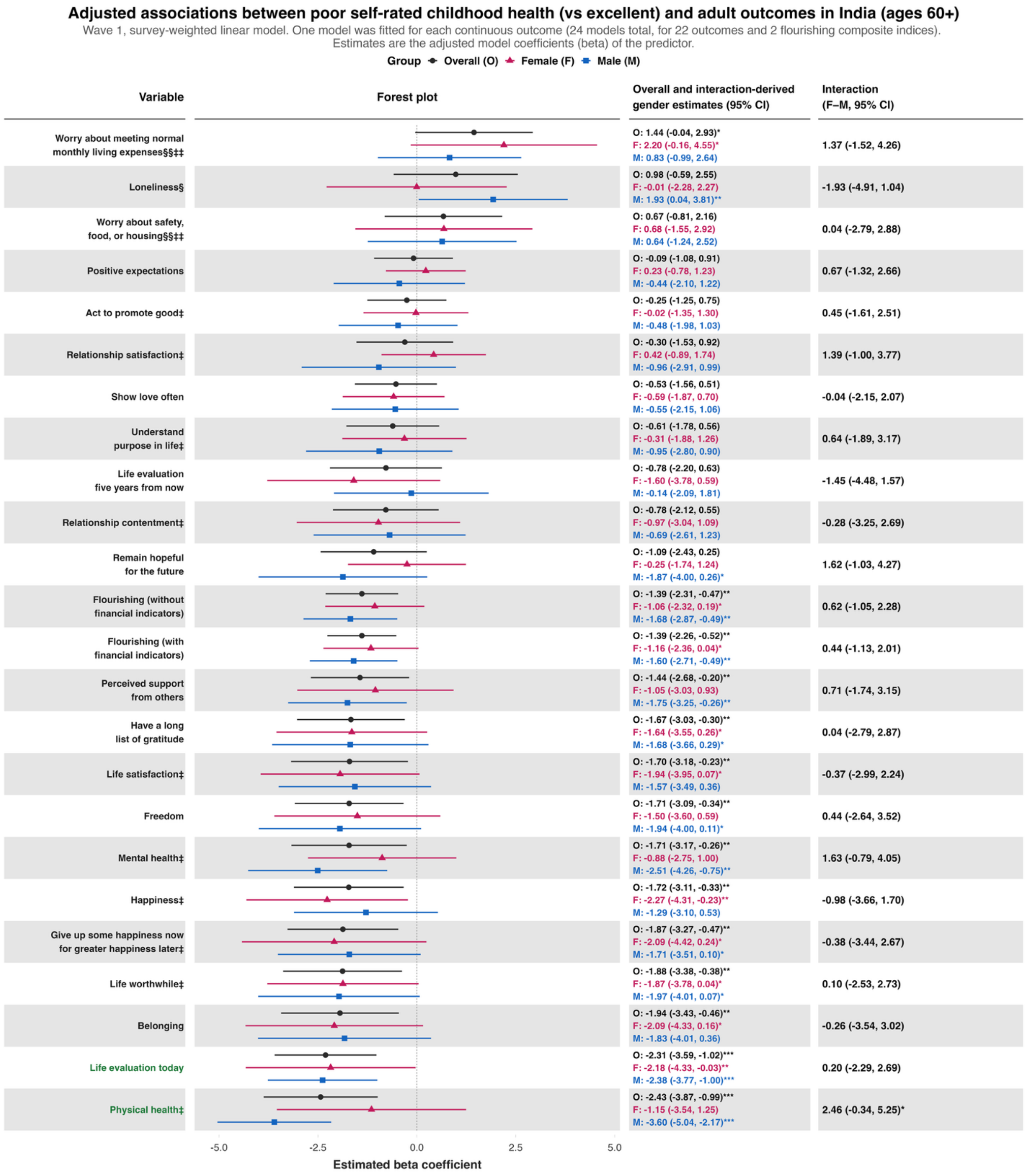

**Predictor:** ***Perceived family financial situation as very difficult while growing up***
***Perceived family financial situation as very difficult while growing up – Ages 18 to 24 inclusive***

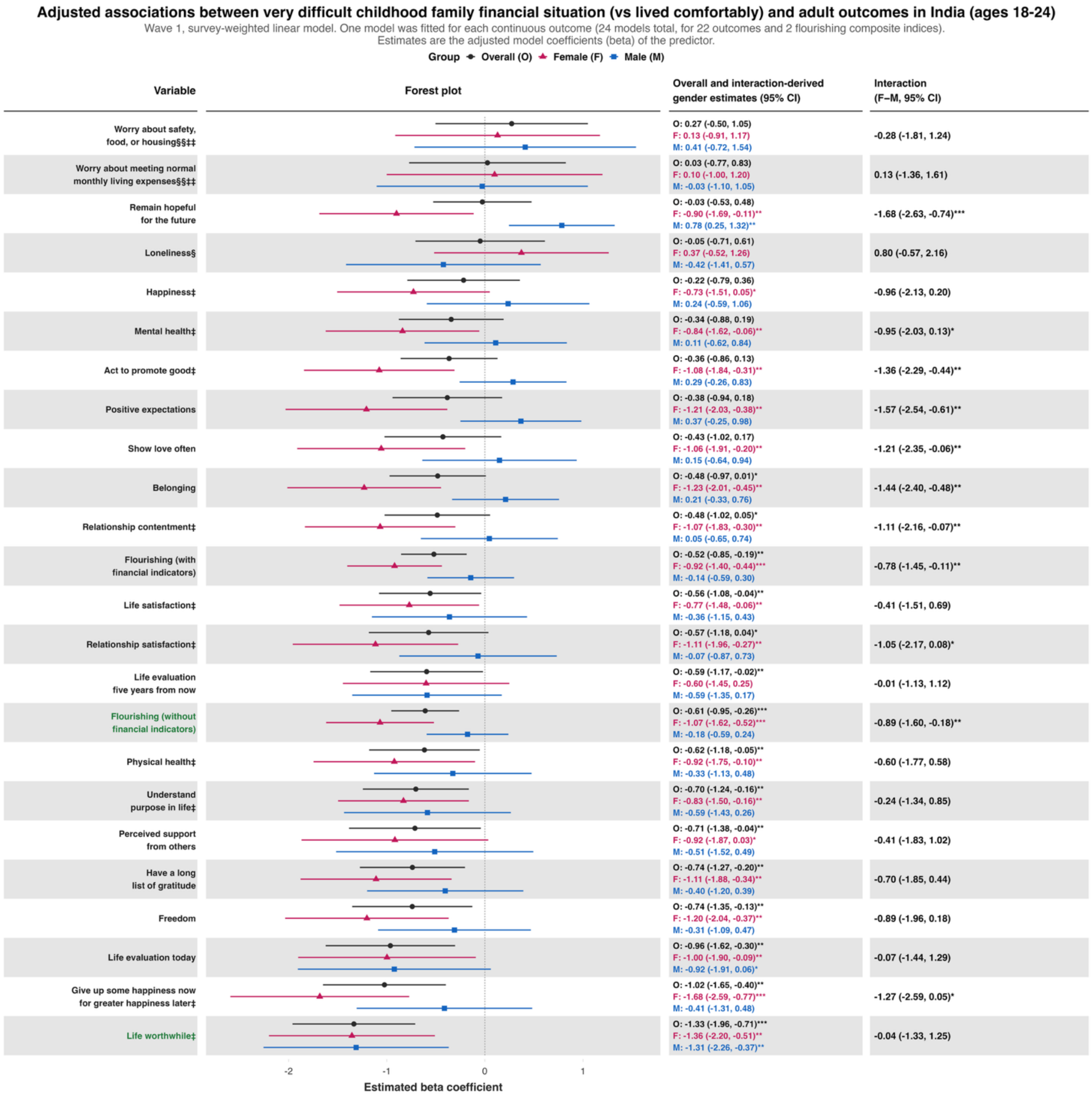

### *Perceived family financial situation as very difficult while growing up – Ages 25 to 29 inclusive*

**Adjusted associations between very difficult childhood family financial situation (vs lived comfortably) and adult outcomes in India (ages 25-29)**

Wave 1, survey-weighted linear model. One model was fitted for each continuous outcome (24 models total, for 22 outcomes and 2 flourishing composite indices).
Estimates are the adjusted model coefficients (beta) of the predictor.

Group ◆ Overall (O) ▲ Female (F) ■ Male (M)

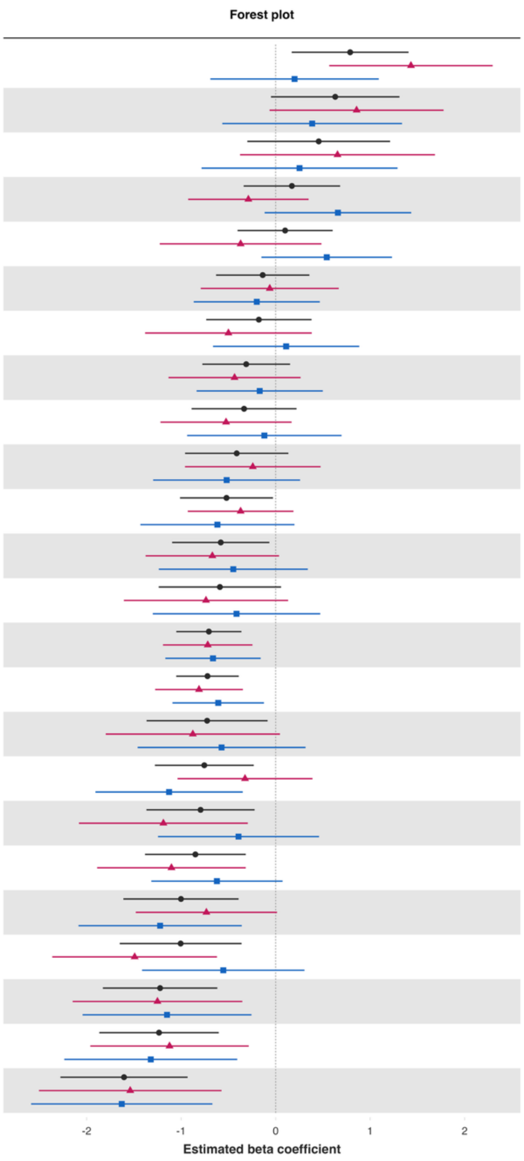


| Variable | Overall and interaction-derived gender estimates (95% CI) | Interaction (F–M, 95% CI) |
|---|---|---|
| Worry about meeting normal monthly living expenses§§‡‡ | O: 0.79 (0.17, 1.41)**<br>F: 1.43 (0.57, 2.30)**<br>M: 0.20 (-0.69, 1.09) | 1.23 (-0.02, 2.48)* |
| Worry about safety, food, or housing§§‡‡ | O: 0.63 (-0.05, 1.31)*<br>F: 0.86 (-0.06, 1.78)*<br>M: 0.39 (-0.56, 1.34) | 0.47 (-0.82, 1.76) |
| Loneliness§ | O: 0.46 (-0.30, 1.21)<br>F: 0.65 (-0.38, 1.68)<br>M: 0.25 (-0.78, 1.29) | 0.40 (-1.00, 1.80) |
| Positive expectations | O: 0.17 (-0.34, 0.68)<br>F: -0.29 (-0.93, 0.35)<br>M: 0.66 (-0.12, 1.43)* | -0.95 (-1.93, 0.04)* |
| Show love often | O: 0.10 (-0.40, 0.60)<br>F: -0.37 (-1.23, 0.48)<br>M: 0.54 (-0.15, 1.23) | -0.91 (-2.07, 0.24) |
| Remain hopeful for the future | O: -0.14 (-0.63, 0.36)<br>F: -0.06 (-0.79, 0.67)<br>M: -0.20 (-0.87, 0.47) | 0.14 (-0.85, 1.12) |
| Have a long list of gratitude | O: -0.18 (-0.73, 0.38)<br>F: -0.50 (-1.38, 0.38)<br>M: 0.11 (-0.66, 0.88) | -0.61 (-1.84, 0.62) |
| Belonging | O: -0.31 (-0.78, 0.15)<br>F: -0.43 (-1.13, 0.26)<br>M: -0.17 (-0.84, 0.50) | -0.27 (-1.26, 0.73) |
| Relationship satisfaction‡ | O: -0.33 (-0.89, 0.22)<br>F: -0.52 (-1.22, 0.17)<br>M: -0.12 (-0.94, 0.70) | -0.41 (-1.44, 0.63) |
| Understand purpose in life‡ | O: -0.41 (-0.96, 0.13)<br>F: -0.24 (-0.96, 0.48)<br>M: -0.52 (-1.30, 0.26) | 0.28 (-0.75, 1.31) |
| Act to promote good‡ | O: -0.52 (-1.01, -0.03)**<br>F: -0.37 (-0.93, 0.19)<br>M: -0.62 (-1.43, 0.20) | 0.25 (-0.77, 1.26) |
| Relationship contentment‡ | O: -0.58 (-1.10, -0.07)**<br>F: -0.67 (-1.38, 0.04)*<br>M: -0.45 (-1.24, 0.34) | -0.22 (-1.32, 0.87) |
| Physical health‡ | O: -0.59 (-1.24, 0.06)*<br>F: -0.74 (-1.61, 0.13)*<br>M: -0.41 (-1.30, 0.47) | -0.32 (-1.50, 0.85) |
| Flourishing (without financial indicators) | O: -0.71 (-1.05, -0.36)***<br>F: -0.72 (-1.19, -0.25)**<br>M: -0.66 (-1.17, -0.16)** | -0.06 (-0.75, 0.64) |
| Flourishing (with financial indicators) | O: -0.72 (-1.05, -0.39)***<br>F: -0.81 (-1.28, -0.35)***<br>M: -0.61 (-1.09, -0.12)** | -0.20 (-0.88, 0.47) |
| Give up some happiness now for greater happiness later‡ | O: -0.73 (-1.37, -0.09)**<br>F: -0.88 (-1.80, 0.05)*<br>M: -0.57 (-1.46, 0.32) | -0.30 (-1.57, 0.96) |
| Life worthwhile‡ | O: -0.76 (-1.28, -0.23)**<br>F: -0.32 (-1.04, 0.39)<br>M: -1.13 (-1.91, -0.35)** | 0.80 (-0.28, 1.88) |
| Life satisfaction‡ | O: -0.79 (-1.37, -0.22)**<br>F: -1.19 (-2.08, -0.29)**<br>M: -0.39 (-1.25, 0.46) | -0.79 (-2.11, 0.52) |
| Life evaluation five years from now | O: -0.85 (-1.38, -0.32)**<br>F: -1.10 (-1.89, -0.32)**<br>M: -0.62 (-1.32, 0.07)* | -0.48 (-1.51, 0.54) |
| Freedom | O: -1.00 (-1.61, -0.39)**<br>F: -0.73 (-1.48, 0.01)*<br>M: -1.22 (-2.09, -0.36)** | 0.49 (-0.60, 1.58) |
| Life evaluation today | O: -1.01 (-1.65, -0.36)**<br>F: -1.49 (-2.36, -0.62)***<br>M: -0.55 (-1.41, 0.30) | -0.94 (-2.10, 0.22) |
| Mental health‡ | O: -1.22 (-1.83, -0.62)***<br>F: -1.25 (-2.15, -0.35)**<br>M: -1.15 (-2.04, -0.26)** | -0.10 (-1.42, 1.22) |
| Happiness‡ | O: -1.23 (-1.87, -0.60)***<br>F: -1.12 (-1.96, -0.28)**<br>M: -1.32 (-2.24, -0.41)** | 0.20 (-1.03, 1.43) |
| Perceived support from others | O: -1.60 (-2.28, -0.93)***<br>F: -1.54 (-2.50, -0.57)**<br>M: -1.63 (-2.59, -0.67)*** | 0.09 (-1.29, 1.47) |

### *Perceived family financial situation as very difficult while growing up – Ages 30 to 39 inclusive*

**Adjusted associations between very difficult childhood family financial situation (vs lived comfortably) and adult outcomes in India (ages 30-39)**

Wave 1, survey-weighted linear model. One model was fitted for each continuous outcome (24 models total, for 22 outcomes and 2 flourishing composite indices).
Estimates are the adjusted model coefficients (beta) of the predictor.

**Group** ◆ Overall (O) ▲ Female (F) ■ Male (M)

Forest plot

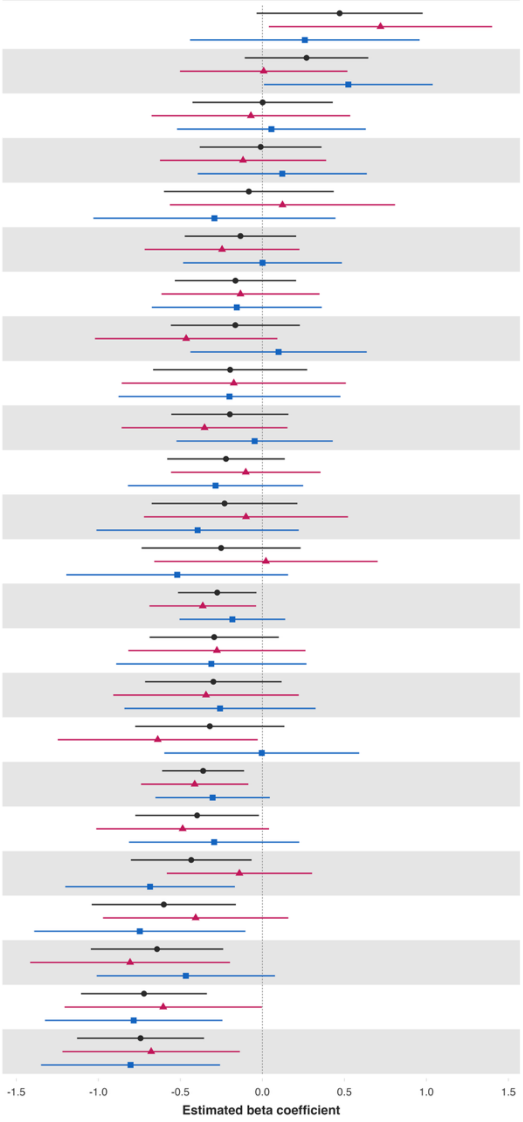


| Variable | Overall and interaction-derived gender estimates (95% CI) | Interaction (F–M, 95% CI) |
|---|---|---|
| Loneliness§ | O: 0.47 (-0.04, 0.98)*<br>F: 0.72 (0.04, 1.40)**<br>M: 0.26 (-0.44, 0.96) | 0.46 (-0.47, 1.39) |
| Have a long list of gratitude | O: 0.27 (-0.11, 0.65)<br>F: 0.01 (-0.50, 0.52)<br>M: 0.52 (0.01, 1.04)** | -0.52 (-1.21, 0.17) |
| Give up some happiness now for greater happiness later‡ | O: 0.00 (-0.43, 0.43)<br>F: -0.07 (-0.68, 0.54)<br>M: 0.05 (-0.52, 0.63) | -0.13 (-0.94, 0.69) |
| Positive expectations | O: -0.01 (-0.38, 0.36)<br>F: -0.12 (-0.62, 0.39)<br>M: 0.12 (-0.39, 0.64) | -0.24 (-0.94, 0.46) |
| Worry about meeting normal monthly living expenses§§‡‡ | O: -0.08 (-0.60, 0.43)<br>F: 0.12 (-0.56, 0.81)<br>M: -0.29 (-1.03, 0.45) | 0.41 (-0.56, 1.39) |
| Show love often | O: -0.13 (-0.47, 0.21)<br>F: -0.25 (-0.72, 0.23)<br>M: 0.00 (-0.48, 0.48) | -0.25 (-0.92, 0.43) |
| Freedom | O: -0.16 (-0.53, 0.21)<br>F: -0.13 (-0.62, 0.35)<br>M: -0.16 (-0.67, 0.36) | 0.02 (-0.65, 0.70) |
| Relationship satisfaction‡ | O: -0.16 (-0.56, 0.23)<br>F: -0.46 (-1.02, 0.09)<br>M: 0.10 (-0.44, 0.64) | -0.56 (-1.32, 0.19) |
| Perceived support from others | O: -0.20 (-0.67, 0.27)<br>F: -0.17 (-0.86, 0.51)<br>M: -0.20 (-0.88, 0.48) | 0.03 (-0.95, 1.01) |
| Belonging | O: -0.20 (-0.56, 0.16)<br>F: -0.35 (-0.86, 0.15)<br>M: -0.05 (-0.52, 0.43) | -0.31 (-0.97, 0.36) |
| Remain hopeful for the future | O: -0.22 (-0.58, 0.14)<br>F: -0.10 (-0.56, 0.35)<br>M: -0.29 (-0.82, 0.25) | 0.18 (-0.51, 0.88) |
| Life evaluation today | O: -0.23 (-0.67, 0.21)<br>F: -0.10 (-0.72, 0.52)<br>M: -0.39 (-1.01, 0.22) | 0.30 (-0.57, 1.16) |
| Worry about safety, food, or housing§§‡‡ | O: -0.25 (-0.74, 0.23)<br>F: 0.02 (-0.66, 0.70)<br>M: -0.52 (-1.20, 0.16) | 0.54 (-0.39, 1.47) |
| Flourishing (with financial indicators) | O: -0.28 (-0.51, -0.04)**<br>F: -0.36 (-0.69, -0.04)**<br>M: -0.18 (-0.51, 0.14) | -0.18 (-0.61, 0.25) |
| Relationship contentment‡ | O: -0.29 (-0.69, 0.10)<br>F: -0.28 (-0.82, 0.26)<br>M: -0.31 (-0.89, 0.27) | 0.03 (-0.76, 0.83) |
| Life satisfaction‡ | O: -0.30 (-0.71, 0.12)<br>F: -0.34 (-0.91, 0.22)<br>M: -0.26 (-0.84, 0.32) | -0.09 (-0.87, 0.70) |
| Happiness‡ | O: -0.32 (-0.78, 0.13)<br>F: -0.64 (-1.25, -0.03)**<br>M: -0.00 (-0.60, 0.59) | -0.63 (-1.41, 0.15) |
| Flourishing (without financial indicators) | O: -0.36 (-0.61, -0.11)**<br>F: -0.41 (-0.74, -0.09)**<br>M: -0.30 (-0.65, 0.04)* | -0.11 (-0.57, 0.35) |
| Understand purpose in life‡ | O: -0.40 (-0.77, -0.02)**<br>F: -0.49 (-1.01, 0.04)*<br>M: -0.29 (-0.81, 0.22) | -0.19 (-0.91, 0.53) |
| Act to promote good‡ | O: -0.43 (-0.80, -0.07)**<br>F: -0.14 (-0.58, 0.30)<br>M: -0.68 (-1.20, -0.17)** | 0.54 (-0.09, 1.18)* |
| Life worthwhile‡ | O: -0.60 (-1.04, -0.16)**<br>F: -0.41 (-0.97, 0.16)<br>M: -0.75 (-1.39, -0.10)** | 0.34 (-0.50, 1.18) |
| Physical health‡ | O: -0.64 (-1.05, -0.24)**<br>F: -0.81 (-1.42, -0.20)**<br>M: -0.47 (-1.01, 0.08)* | -0.34 (-1.16, 0.48) |
| Mental health‡ | O: -0.72 (-1.11, -0.34)***<br>F: -0.60 (-1.21, -0.00)**<br>M: -0.78 (-1.33, -0.24)** | 0.18 (-0.66, 1.02) |
| Life evaluation five years from now | O: -0.74 (-1.13, -0.36)***<br>F: -0.68 (-1.22, -0.14)**<br>M: -0.80 (-1.35, -0.26)** | 0.13 (-0.64, 0.89) |

### *Perceived family financial situation as very difficult while growing up – Ages 40 to 49 inclusive*

**Adjusted associations between very difficult childhood family financial situation (vs lived comfortably) and adult outcomes in India (ages 40-49)**

Wave 1, survey-weighted linear model. One model was fitted for each continuous outcome (24 models total, for 22 outcomes and 2 flourishing composite indices).
Estimates are the adjusted model coefficients (beta) of the predictor.

**Group** ● Overall (O) ▲ Female (F) ■ Male (M)

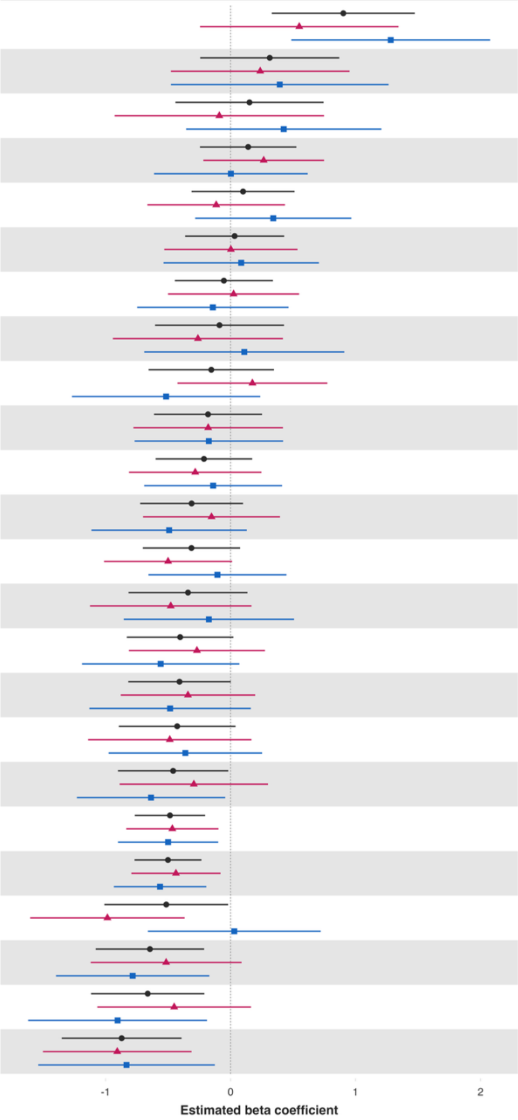


| Variable | Overall and interaction-derived gender estimates (95% CI) | Interaction (F–M, 95% CI) |
|---|---|---|
| Worry about meeting normal monthly living expenses§§‡‡ | O: 0.90 (0.33, 1.47)**<br>F: 0.55 (-0.24, 1.34)<br>M: 1.28 (0.49, 2.08)** | -0.73 (-1.84, 0.38) |
| Loneliness§ | O: 0.31 (-0.24, 0.87)<br>F: 0.24 (-0.48, 0.95)<br>M: 0.39 (-0.48, 1.26) | -0.16 (-1.29, 0.97) |
| Worry about safety, food, or housing§§‡‡ | O: 0.15 (-0.44, 0.74)<br>F: -0.09 (-0.93, 0.75)<br>M: 0.43 (-0.36, 1.21) | -0.52 (-1.63, 0.60) |
| Positive expectations | O: 0.14 (-0.25, 0.53)<br>F: 0.26 (-0.22, 0.75)<br>M: 0.00 (-0.61, 0.62) | 0.26 (-0.52, 1.05) |
| Show love often | O: 0.10 (-0.31, 0.51)<br>F: -0.12 (-0.67, 0.44)<br>M: 0.34 (-0.28, 0.97) | -0.46 (-1.29, 0.38) |
| Have a long list of gratitude | O: 0.03 (-0.36, 0.43)<br>F: 0.00 (-0.53, 0.54)<br>M: 0.09 (-0.54, 0.71) | -0.08 (-0.92, 0.75) |
| Understand purpose in life‡ | O: -0.05 (-0.45, 0.34)<br>F: 0.02 (-0.50, 0.55)<br>M: -0.14 (-0.75, 0.46) | 0.17 (-0.65, 0.98) |
| Perceived support from others | O: -0.09 (-0.60, 0.43)<br>F: -0.26 (-0.94, 0.42)<br>M: 0.11 (-0.69, 0.91) | -0.37 (-1.43, 0.69) |
| Give up some happiness now for greater happiness later‡ | O: -0.15 (-0.66, 0.35)<br>F: 0.17 (-0.42, 0.77)<br>M: -0.52 (-1.27, 0.24) | 0.69 (-0.22, 1.60) |
| Freedom | O: -0.18 (-0.61, 0.25)<br>F: -0.18 (-0.78, 0.42)<br>M: -0.17 (-0.77, 0.42) | -0.01 (-0.83, 0.82) |
| Remain hopeful for the future | O: -0.21 (-0.60, 0.17)<br>F: -0.28 (-0.81, 0.25)<br>M: -0.14 (-0.69, 0.41) | -0.14 (-0.90, 0.61) |
| Life evaluation five years from now | O: -0.31 (-0.72, 0.10)<br>F: -0.15 (-0.70, 0.40)<br>M: -0.49 (-1.11, 0.13) | 0.34 (-0.49, 1.17) |
| Act to promote good‡ | O: -0.31 (-0.70, 0.08)<br>F: -0.50 (-1.01, 0.01)*<br>M: -0.11 (-0.66, 0.45) | -0.39 (-1.12, 0.33) |
| Life evaluation today | O: -0.34 (-0.82, 0.14)<br>F: -0.48 (-1.12, 0.17)<br>M: -0.17 (-0.85, 0.51) | -0.31 (-1.23, 0.62) |
| Life worthwhile‡ | O: -0.40 (-0.83, 0.02)*<br>F: -0.27 (-0.81, 0.27)<br>M: -0.56 (-1.19, 0.07)* | 0.29 (-0.52, 1.10) |
| Belonging | O: -0.41 (-0.82, 0.00)*<br>F: -0.34 (-0.88, 0.20)<br>M: -0.48 (-1.13, 0.16) | 0.14 (-0.71, 1.00) |
| Relationship satisfaction‡ | O: -0.43 (-0.89, 0.04)*<br>F: -0.49 (-1.14, 0.17)<br>M: -0.36 (-0.98, 0.25) | -0.12 (-0.99, 0.74) |
| Mental health‡ | O: -0.46 (-0.90, -0.02)**<br>F: -0.29 (-0.89, 0.30)<br>M: -0.64 (-1.23, -0.04)** | 0.34 (-0.45, 1.14) |
| Flourishing (without financial indicators) | O: -0.48 (-0.77, -0.20)***<br>F: -0.47 (-0.83, -0.10)**<br>M: -0.50 (-0.90, -0.10)** | 0.03 (-0.49, 0.56) |
| Flourishing (with financial indicators) | O: -0.50 (-0.77, -0.23)***<br>F: -0.44 (-0.79, -0.08)**<br>M: -0.56 (-0.93, -0.19)** | 0.13 (-0.36, 0.62) |
| Happiness‡ | O: -0.51 (-1.01, -0.02)**<br>F: -0.99 (-1.60, -0.37)**<br>M: 0.03 (-0.66, 0.72) | -1.02 (-1.87, -0.16)** |
| Physical health‡ | O: -0.65 (-1.08, -0.21)**<br>F: -0.52 (-1.12, 0.09)*<br>M: -0.78 (-1.40, -0.17)** | 0.27 (-0.59, 1.13) |
| Relationship contentment‡ | O: -0.66 (-1.12, -0.21)**<br>F: -0.45 (-1.07, 0.16)<br>M: -0.90 (-1.62, -0.19)** | 0.45 (-0.52, 1.42) |
| Life satisfaction‡ | O: -0.87 (-1.35, -0.39)***<br>F: -0.91 (-1.50, -0.31)**<br>M: -0.83 (-1.54, -0.13)** | -0.07 (-0.95, 0.80) |

***Perceived family financial situation as very difficult while growing up – Ages 50 to 59 inclusive***

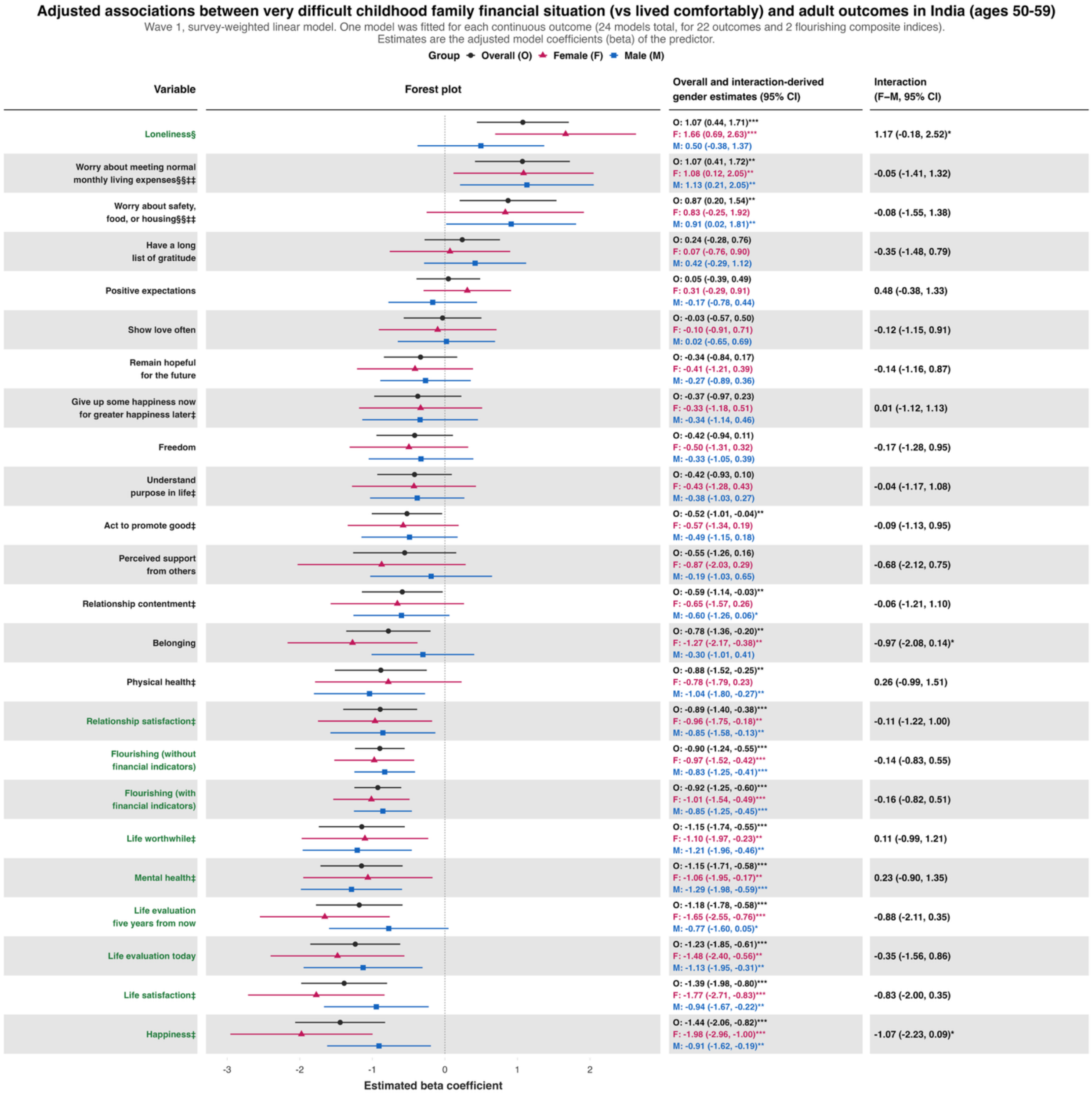

### *Perceived family financial situation as very difficult while growing up – Ages 60+*

**Adjusted associations between very difficult childhood family financial situation (vs lived comfortably) and adult outcomes in India (ages 60+)**

Wave 1, survey-weighted linear model. One model was fitted for each continuous outcome (24 models total, for 22 outcomes and 2 flourishing composite indices).
Estimates are the adjusted model coefficients (beta) of the predictor.

Group ◆ Overall (O) ▲ Female (F) ■ Male (M)

| Variable | Overall and interaction-derived gender estimates (95% CI) | Interaction (F–M, 95% CI) |
|---|---|---|
| Worry about meeting normal monthly living expenses§§‡‡ | O: 0.64 (-0.11, 1.39)*<br>F: 1.11 (0.05, 2.17)**<br>M: 0.34 (-0.64, 1.33) | 0.77 (-0.62, 2.16) |
| Loneliness§ | O: 0.38 (-0.29, 1.05)<br>F: 0.30 (-0.66, 1.26)<br>M: 0.37 (-0.49, 1.23) | -0.07 (-1.29, 1.15) |
| Worry about safety, food, or housing§§‡‡ | O: 0.32 (-0.40, 1.05)<br>F: 0.71 (-0.45, 1.88)<br>M: -0.09 (-1.03, 0.85) | 0.80 (-0.73, 2.34) |
| Positive expectations | O: 0.17 (-0.42, 0.75)<br>F: 0.14 (-0.66, 0.93)<br>M: 0.21 (-0.54, 0.96) | -0.07 (-1.09, 0.95) |
| Have a long list of gratitude | O: 0.11 (-0.50, 0.72)<br>F: 0.26 (-0.57, 1.09)<br>M: 0.03 (-0.78, 0.84) | 0.23 (-0.85, 1.32) |
| Perceived support from others | O: 0.10 (-0.62, 0.81)<br>F: 0.61 (-0.37, 1.59)<br>M: -0.26 (-1.25, 0.72) | 0.87 (-0.49, 2.24) |
| Give up some happiness now for greater happiness later‡ | O: -0.02 (-0.55, 0.51)<br>F: 0.15 (-0.72, 1.02)<br>M: -0.07 (-0.79, 0.66) | 0.22 (-0.97, 1.41) |
| Relationship satisfaction‡ | O: -0.02 (-0.56, 0.51)<br>F: 0.47 (-0.28, 1.22)<br>M: -0.53 (-1.27, 0.21) | 1.00 (-0.07, 2.07)* |
| Act to promote good‡ | O: -0.23 (-0.79, 0.32)<br>F: -0.34 (-1.11, 0.43)<br>M: -0.05 (-0.83, 0.73) | -0.29 (-1.37, 0.79) |
| Show love often | O: -0.28 (-0.79, 0.24)<br>F: -0.42 (-1.16, 0.31)<br>M: -0.04 (-0.75, 0.67) | -0.38 (-1.38, 0.61) |
| Remain hopeful for the future | O: -0.32 (-0.85, 0.21)<br>F: -0.24 (-1.05, 0.56)<br>M: -0.41 (-1.07, 0.26) | 0.16 (-0.86, 1.19) |
| Understand purpose in life‡ | O: -0.38 (-0.92, 0.17)<br>F: -0.78 (-1.65, 0.08)*<br>M: 0.06 (-0.66, 0.79) | -0.85 (-1.99, 0.30) |
| Flourishing (with financial indicators) | O: -0.39 (-0.78, -0.00)**<br>F: -0.23 (-0.83, 0.37)<br>M: -0.49 (-0.98, 0.00)* | 0.26 (-0.50, 1.02) |
| Flourishing (without financial indicators) | O: -0.41 (-0.81, -0.01)**<br>F: -0.12 (-0.74, 0.50)<br>M: -0.60 (-1.10, -0.10)** | 0.47 (-0.31, 1.25) |
| Freedom | O: -0.43 (-0.99, 0.13)<br>F: -0.54 (-1.31, 0.23)<br>M: -0.30 (-1.05, 0.45) | -0.24 (-1.26, 0.78) |
| Physical health‡ | O: -0.45 (-1.09, 0.18)<br>F: 0.01 (-1.00, 1.03)<br>M: -0.71 (-1.46, 0.04)* | 0.73 (-0.49, 1.94) |
| Life satisfaction‡ | O: -0.46 (-1.11, 0.20)<br>F: 0.01 (-0.98, 1.00)<br>M: -0.81 (-1.61, -0.02)** | 0.82 (-0.38, 2.03) |
| Relationship contentment‡ | O: -0.48 (-1.06, 0.09)<br>F: 0.02 (-0.78, 0.82)<br>M: -0.90 (-1.68, -0.13)** | 0.93 (-0.17, 2.02)* |
| Life evaluation today | O: -0.58 (-1.16, -0.01)**<br>F: -0.40 (-1.40, 0.60)<br>M: -0.67 (-1.41, 0.07)* | 0.27 (-1.02, 1.57) |
| Happiness‡ | O: -0.66 (-1.28, -0.03)**<br>F: -0.38 (-1.44, 0.68)<br>M: -0.93 (-1.66, -0.20)** | 0.55 (-0.71, 1.82) |
| Life worthwhile‡ | O: -0.67 (-1.33, -0.01)**<br>F: -0.54 (-1.56, 0.49)<br>M: -0.78 (-1.57, 0.01)* | 0.24 (-0.98, 1.47) |
| Mental health‡ | O: -0.71 (-1.29, -0.14)**<br>F: -0.44 (-1.31, 0.43)<br>M: -0.75 (-1.49, -0.01)** | 0.31 (-0.80, 1.42) |
| Belonging | O: -0.84 (-1.42, -0.25)**<br>F: -0.63 (-1.51, 0.26)<br>M: -0.97 (-1.63, -0.32)** | 0.35 (-0.66, 1.36) |
| Life evaluation five years from now | O: -0.90 (-1.54, -0.26)**<br>F: -0.71 (-1.50, 0.09)*<br>M: -0.80 (-1.74, 0.14)* | 0.09 (-1.13, 1.31) |

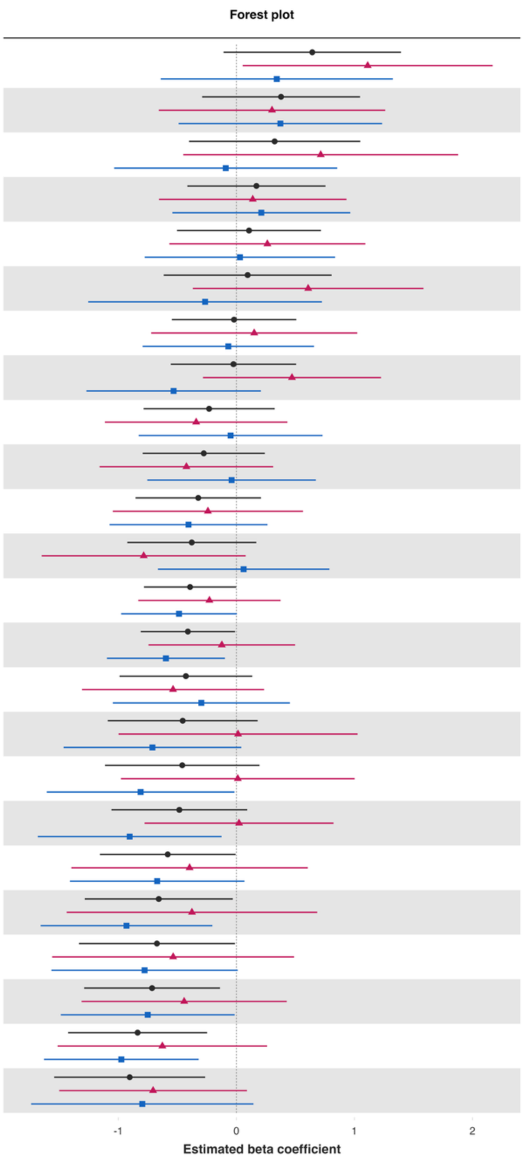

**eFigure 19.** Comparison with linear mixed model results using Waves 1 and 2 data.

***Note:*** Estimates are shown for the 22 continuous adult outcomes, recoded the same way as in the Wave 1 association analyses. Pooled estimates (green circles) come from linear mixed models fit to Waves 1 and 2 combined, with a random intercept for each respondent. The childhood predictor was measured once and applied to both waves, and the models were adjusted for Wave 1 age, gender, education, and residential setting. These models were not survey-weighted and do not account for the complex survey design; therefore, they are a model-based sensitivity analysis. Wave 1 primary estimates (orange triangles) are the survey-weighted adjusted regression coefficients from the main association analyses. Horizontal bars show 95% confidence intervals.

**Predictor:** ***Childhood abuse***

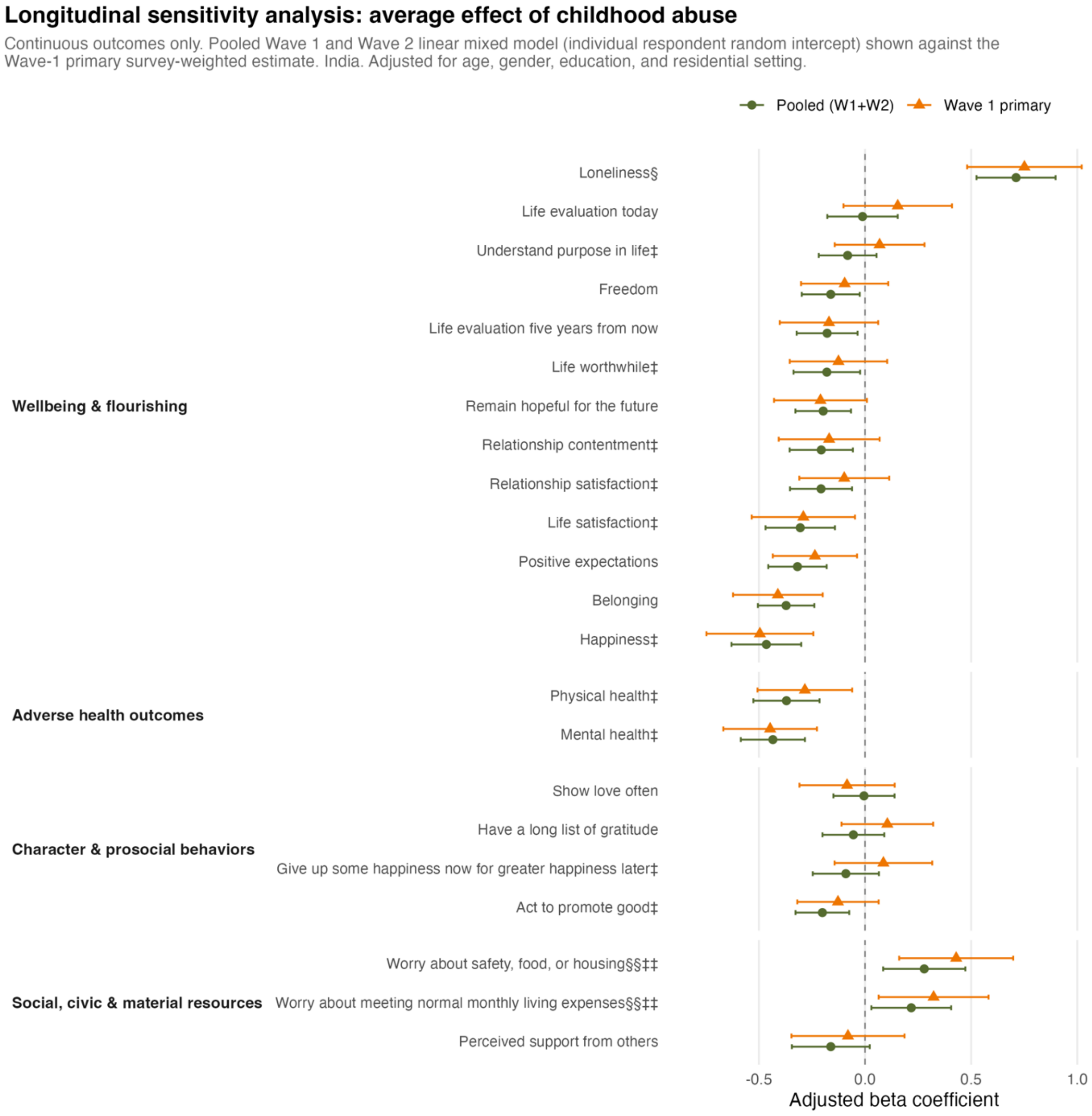

**Predictor:** ***Feeling like an outsider in one's family when growing up***

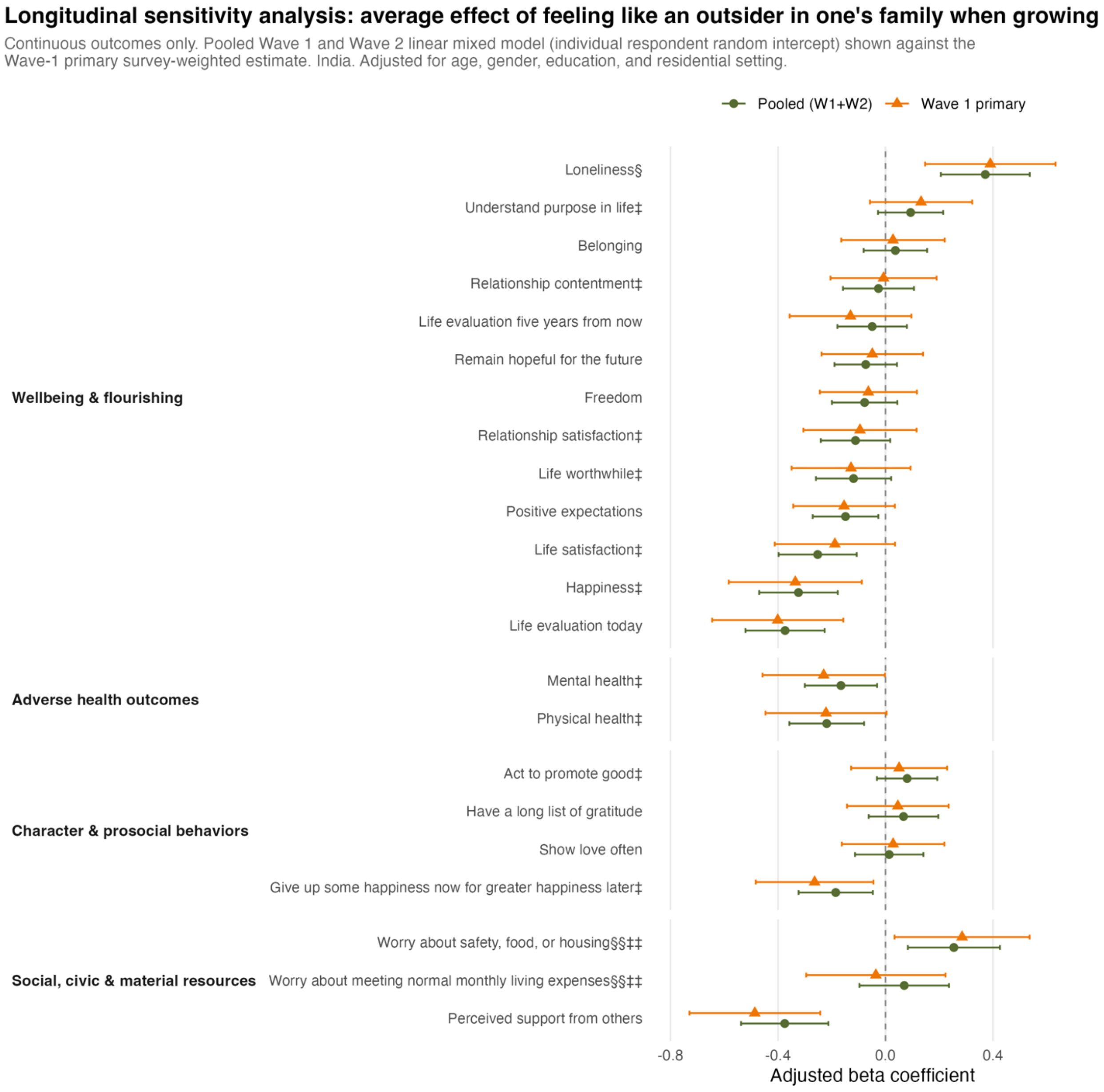

**Predictor:** ***Not feeling loved by mother***

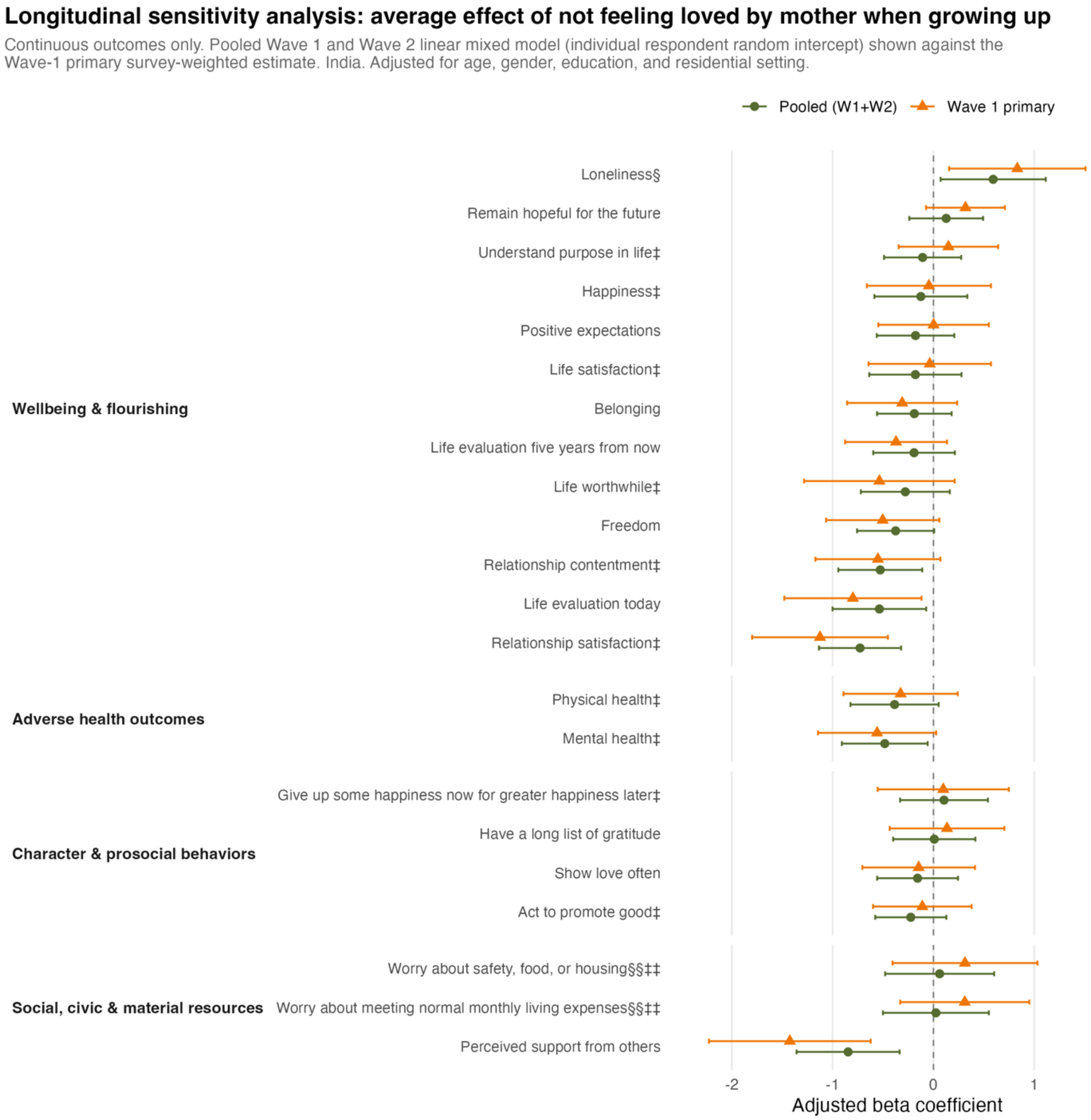

**Predictor:** ***Not feeling loved by father***

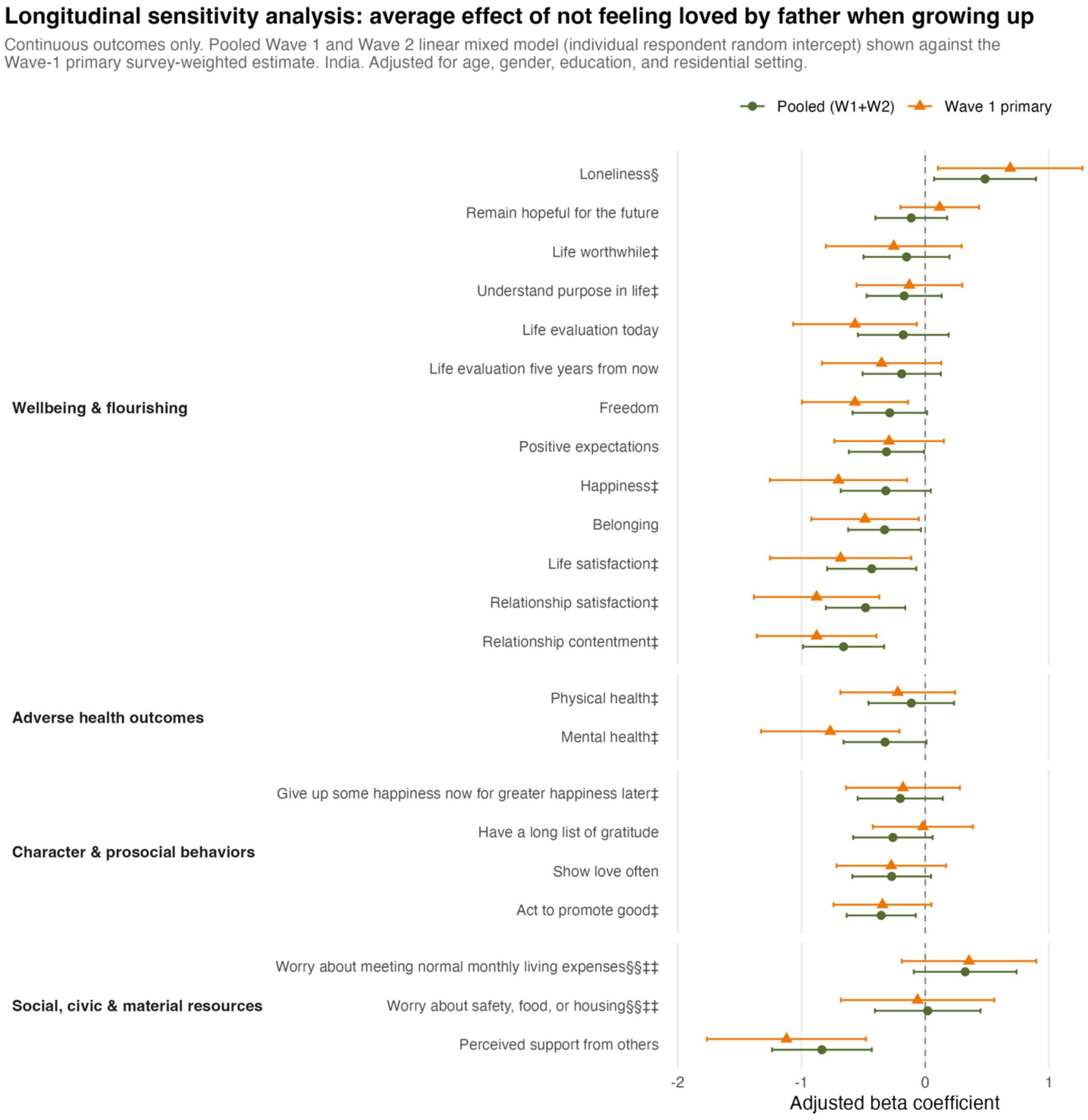

**Predictor:** ***Poorer relationship quality with mother (bad or very bad relationship)***

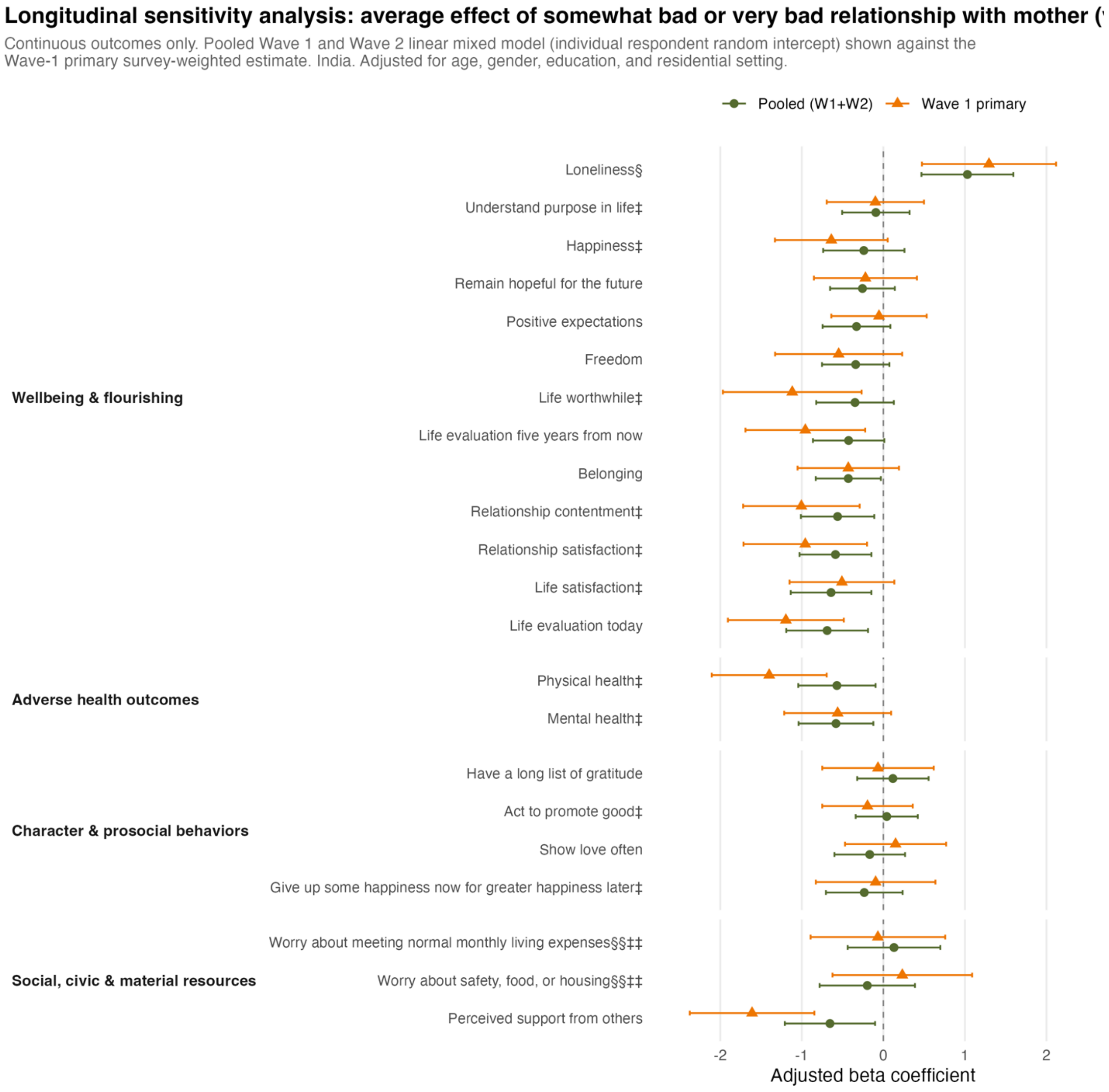

**Predictor:** ***Poorer relationship quality with father (bad or very bad relationship)***

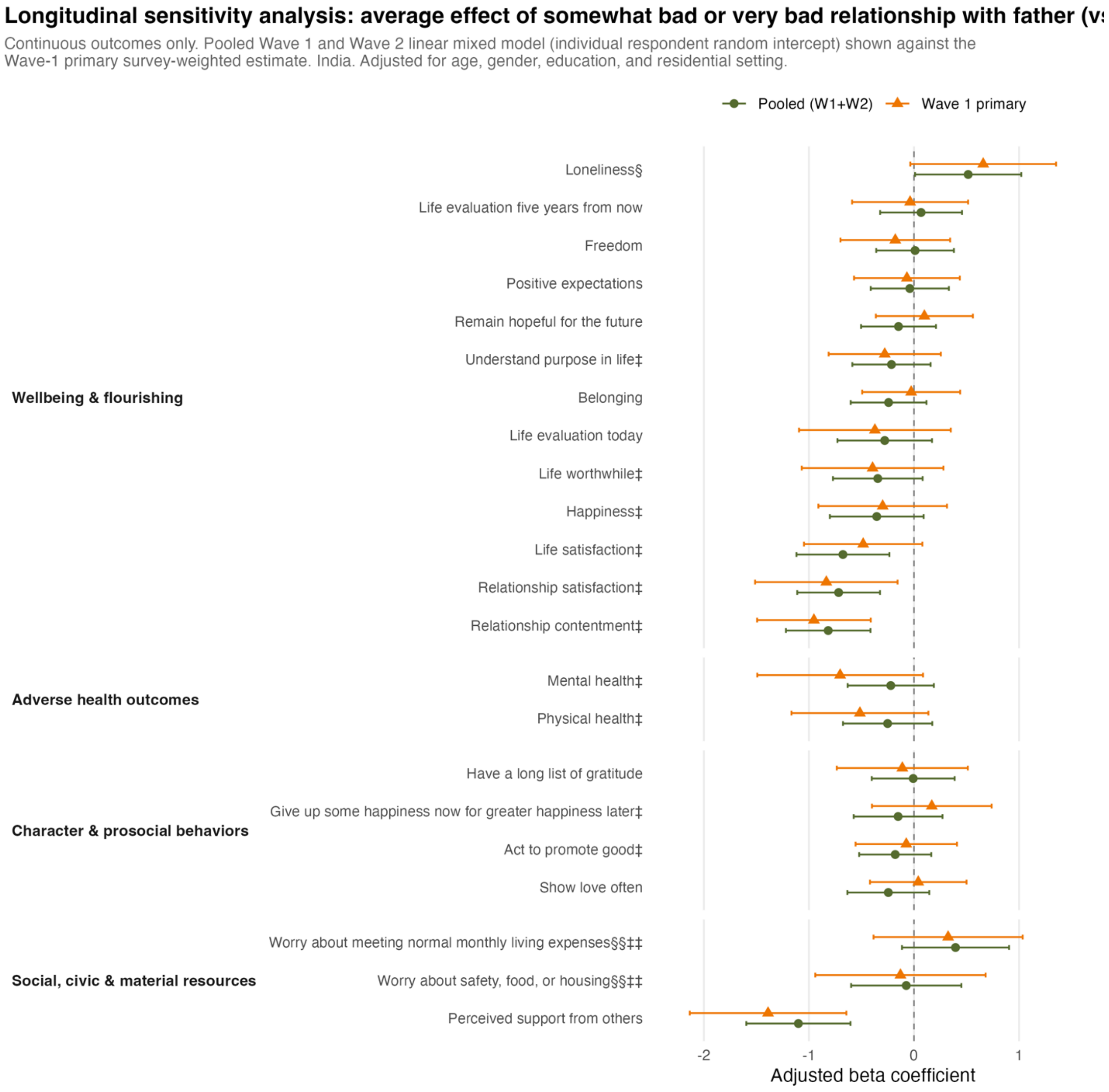

**Predictor:** ***Poor self-rated health while growing up***

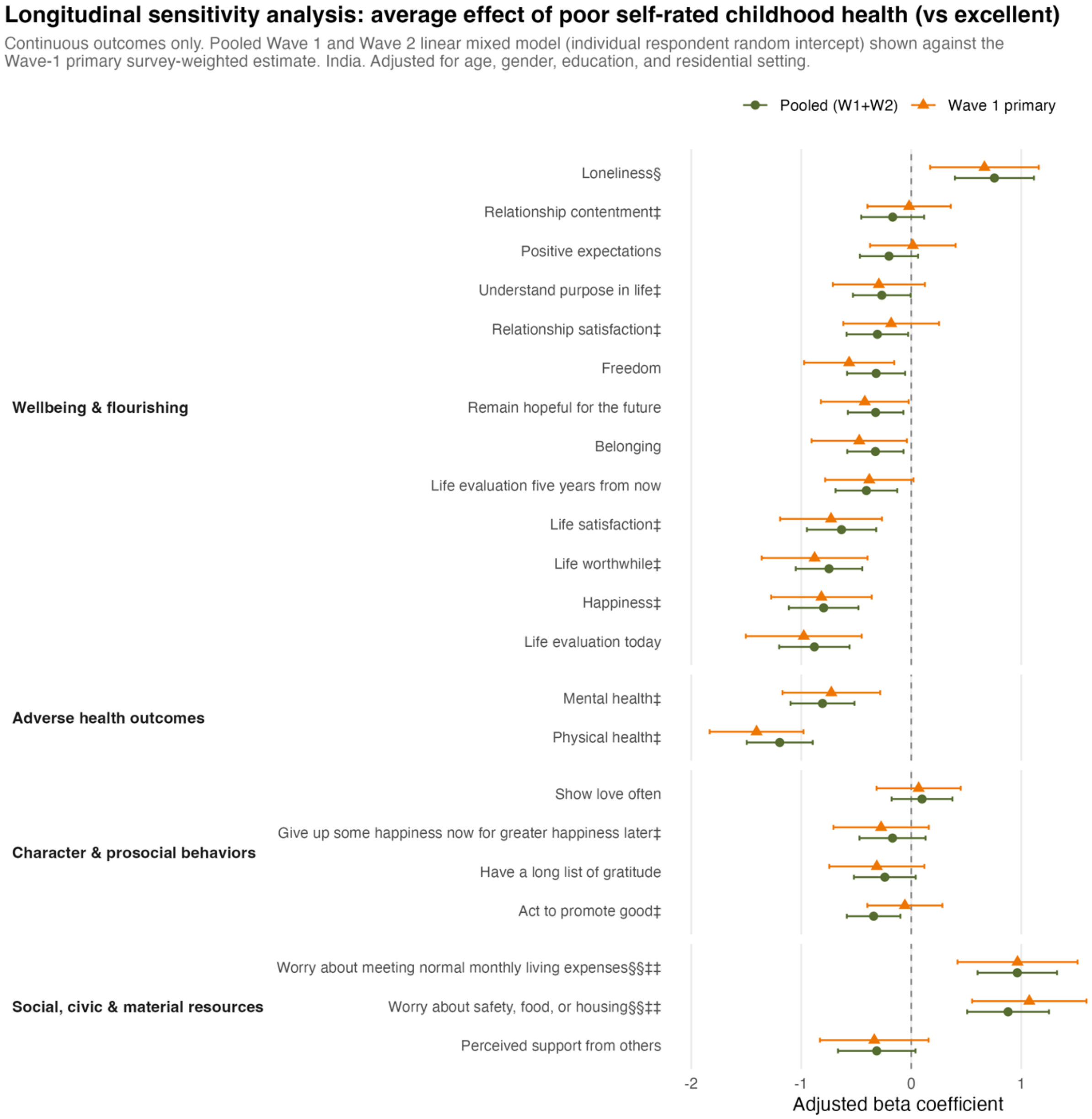

**Predictor:** ***Perceived family financial situation as very difficult while growing up***

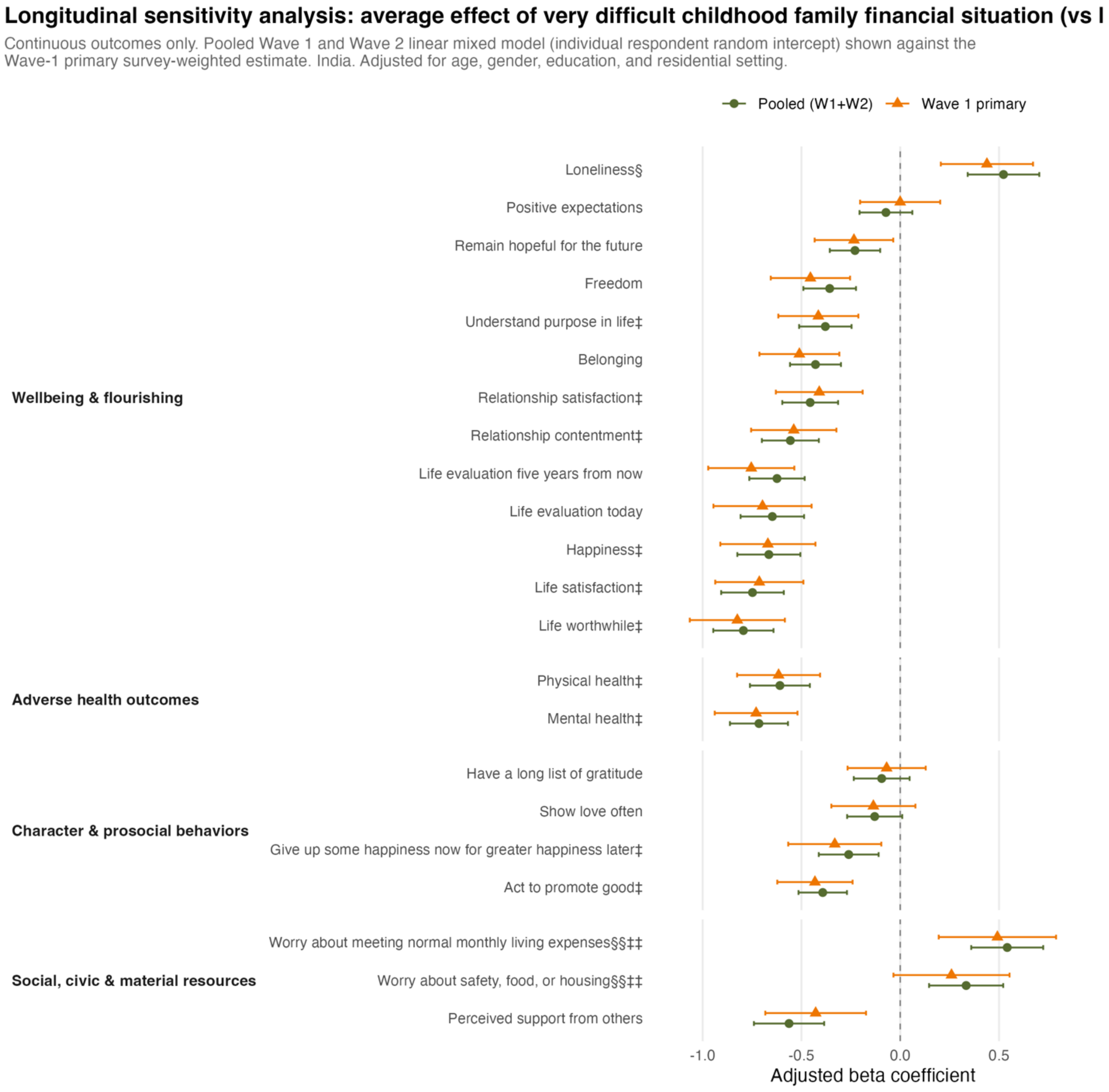

**eFigure 20.** Weighted correlation between childhood variables and covariate age and current educational attainment.

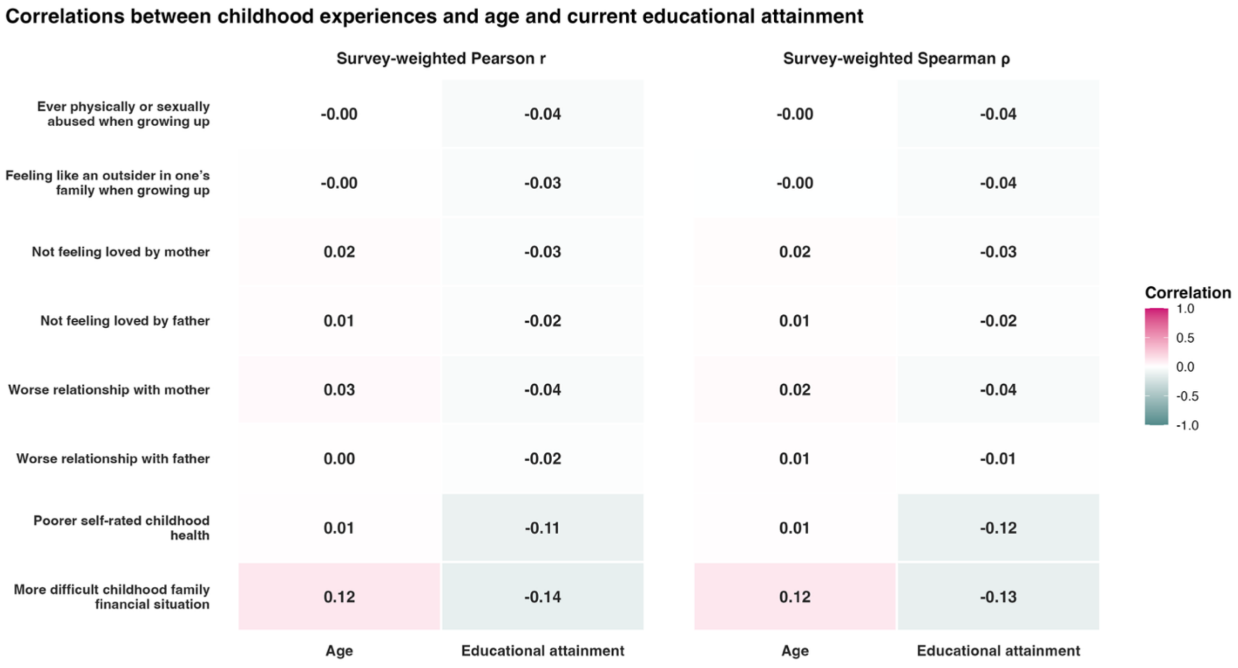

**eResults 1.** Correlation results.

Pairwise correlations among the eight predictors and 45 outcomes were estimated as survey-weighted Pearson correlations and as survey-weighted Spearman correlations of weighted mid-ranks, using pairwise complete observations (eFigures 1-2; eTables 5-6). For these analyses, childhood abuse, feeling like an outsider, and not feeling loved by one's mother or father were coded as binary indicators (1 = exposed), and relationship quality with one's mother and father, childhood self-rated health, and childhood family financial situation were analyzed on their full ordinal scales, with higher values indicating more adverse experiences.

***<u>Correlations between childhood experience and adult outcomes</u>***

Survey-weighted Pearson and Spearman correlation matrices were used to examine correlations within the eight childhood predictors, within the 45 adult outcomes, and between childhood predictors and adult outcomes (eFigures 1-2; eTables 5-6). Overall, correlations among childhood predictors were generally low to moderate. Adult outcomes tended to cluster within the four conceptual groupings, most clearly among the well-being and flourishing measures and the adverse health outcomes. Correlations between childhood predictors and adult outcomes were generally smaller than correlations observed within either set of variables.

Among the childhood experience predictors, the strongest correlations were observed between feeling loved by one's father and relationship quality with one's father ($r$ = 0.47), between feeling loved by one's mother and relationship quality with one's mother ($r$ = 0.36), and between relationship quality with one's mother and with one's father ($r$ = 0.33). Childhood abuse was weakly correlated with all other childhood predictors ($|r| \leq 0.04$). Childhood health and perceived family financial situation were modestly correlated with each other ($r$ = 0.18), and each showed only weak correlations with the remaining childhood predictors ($|r| \leq 0.11$).

Among adult outcomes, the strongest correlations generally occurred within the well-being and flourishing and adverse health outcomes groupings. Notable correlations included those between happiness and life satisfaction ($r$ = 0.58), depressive symptoms and feeling nervous or anxious ($r$ = 0.56), and mental and physical health ($r$ = 0.55). The strongest correlation overall was between worry about meeting monthly living expenses and worry about safety, food, or housing ($r$ = 0.59), both within the social, civic, and material resources grouping. The strongest correlation across groupings was between life satisfaction and mental health ($r$ = 0.42). Among the prosocial behavior outcomes, donating to charity and volunteering were moderately correlated ($r$ = 0.42).

Correlations between childhood predictors and adult outcomes were smaller in magnitude (all $|r| \leq 0.17$). Childhood abuse showed the strongest Pearson correlations, with adult health problems ($r$ = 0.17) and inability to control worry ($r$ = 0.14). Although the prosocial behavior outcomes were correlated with one another, their correlations with childhood predictors were generally small. The largest was between feeling like an outsider while growing up and donating to charity ($r$ = 0.11).

***<u>Correlations between childhood experience, age, and current educational attainment</u>***

Survey-weighted Pearson and Spearman correlations between reported childhood experiences, age, and current educational attainment were generally weak and yielded similar patterns. The strongest correlation was between childhood financial hardship and lower current educational attainment ($r$ = -0.14) (eFigure 20).

**eResults 2.** Sensitivity analyses results for item nonresponse in questions regarding childhood abuse, feeling not loved by mother and by father, and quality of relationship with mother and with father.

Because analyses in this study used only available observations for each predictor-outcome pair, analytic sample sizes differed across models, which may have introduced bias because item nonresponse was not incorporated into the primary analyses. This concern is particularly relevant for the childhood abuse question, for which 5.4% of respondents selected "don't know" and 0.6% refused to respond; such responses may not be missing at random, and the direction of bias is uncertain. Although multiple imputation could be considered, it may not fully address this concern if "don't know" or refusal responses are related to unobserved factors or to respondents' underlying abuse experiences. Sensitivity analyses classifying "don't know" and "refused" responses as either exposed or unexposed yielded broadly similar results, with no significant abuse-by-gender interaction after Bonferroni correction (eFigures 11-12). Similarly, sensitivity analyses grouping "does not apply" responses for questions on feeling loved by and relationship quality with mother and father with the adverse category yielded broadly similar overall and gender-interaction findings (eFigures 13-16); parental marital-status histories among respondents are shown in eTable 21.

**eTable 1.** Description of eight childhood predictors and 45 outcomes of interest.

| **Childhood predictors [8]**<br>***(Labels used in this paper)*** | **Outcomes [45]**<br>***(Labels used in this paper and figures)*** | | | | |
|---|---|---|---|---|---|
| **Abused**<br>***(Childhood abuse)***<br>**[Organized domain: Adverse experiences]**<br>Were you ever physically or sexually abused when you were growing up?<br>*1 = Yes, 2 = No* | **Approve government[§]**<br>***(Government approval)***<br>How much do you approve or disapprove of the job performance of the national government of this country?<br>*1 = Strongly approve,*<br>*2 = Somewhat approve,*<br>*3 = Neither approve nor disapprove,*<br>*4 = Somewhat disapprove,*<br>*5 = Strongly disapprove* | **Donated**<br>***(Donated to charity)***<br>In the past month, have you donated money to a charity?<br>*1 = Yes, 2 = No* | **Happy[‡]**<br>***(Happiness)***<br>In general, how happy or unhappy do you usually feel?<br>*0 = Extremely unhappy to*<br>*10 = Extremely happy* | **Life purpose[‡]**<br>***(Understand purpose in life)***<br>I understand my purpose in life.<br>*0 = Strongly disagree to*<br>*10 = Strongly agree* | **Satisfaction with relationship[‡]**<br>***(Relationship satisfaction)***<br>My relationships are as satisfying as I would want them to be.<br>*0 = Strongly disagree to*<br>*10 = Strongly agree* |
| **Outsider**<br>***(Feeling like an outsider in one's family when growing up)***<br>**[Organized domain: Adverse experiences]**<br>When you were growing up, did you feel like an outsider in your family?<br>*1 = Yes, 2 = No* | **Belonging**<br>***(Belonging in this country)***<br>How would you describe your sense of belonging in your country?<br>*0 = Very weak to 10 = Very strong* | **Drinks**<br>***(Alcohol use)***<br>Approximately how many full drinks of any kind of alcoholic beverage did you drink in the past seven days.<br>*Recoded to 1 = None,*<br>*2 = Had at least one drink in the past seven days* | **Health problem**<br>***(Have any health problems)***<br>Do you have any health problems that prevent you from doing any of the things people your age normally can do?<br>*1 = Yes, 2 = No* | **Life satisfaction[‡]**<br>***(Life satisfaction)***<br>Overall, how satisfied are you with life as a whole these days?<br>*0 = Not at all satisfied with your life to 10 = Completely satisfied with your life* | **Say in government[#]**<br>***(Say in government)***<br>Do you agree or disagree with the following statement? People like me have a say about what the government does?<br>*1 = Agree, 2 = Disagree,*<br>*3 = Unsure* |
| **Mother loved**<br>***(Feeling loved by mother)***<br>**[Organized domain: Emotional experiences]**<br>In general, did you feel loved by your mother when you were growing up?<br>*1 = Yes, 2 = No* | **Capable[§]**<br>***(Feeling capable of things in life)***<br>How often do you feel very capable in most things you do in life?<br>*1 = Always, 2 = Often,*<br>*3 = Rarely, 4 = Never* | **Expect good**<br>***(Positive expectations)***<br>Overall, I expect more good things to happen to me than bad.<br>*0 = Strongly disagree to*<br>*10 = Strongly agree* | **Help stranger**<br>***(Helped a stranger in past month)***<br>In the past month, have you helped a stranger or someone you didn't know who needed help?<br>*1 = Yes, 2 = No* | **Lonely[§]**<br>***(Loneliness)***<br>How often do you feel lonely?<br>*0 = Always to 10 = Never* | **Show love**<br>***(Show love often)***<br>How often do you show someone in your life that you love or care for them?<br>*0 = Never to 10 = Always* |
| **Father loved**<br>***(Feeling loved by father)***<br>**[Organized domain: Emotional experiences]**<br>In general, did you feel loved by your father when you were growing up?<br>*1 = Yes, 2 = No* | **Cigarettes**<br>***(Cigarette smoking)***<br>About how many cigarettes do you smoke each day, if any?<br>*Recoded to 1 = None, 2 = Smoke at least one cigarette each day* | **Expenses[§§‡‡]**<br>***(Worry about meeting normal monthly living expenses)***<br>How often do you worry about being able to meet normal monthly living expenses?<br>*0 = Worry all of the time to*<br>*10 = Do not ever worry* | **Hope for future**<br>***(Remain hopeful for the future)***<br>Despite challenges, I always remain hopeful about the future.<br>*0 = Strongly disagree to*<br>*10 = Strongly agree* | **Mental health[‡]**<br>***(Mental health)***<br>How would you rate your overall mental health?<br>*0 = Poor mental health to*<br>*10 = Excellent mental health* | **Suffering[§]**<br>***(Extent of suffering)***<br>To what extent are you suffering? This can be any type of physical or mental suffering?<br>*1 = A lot, 2 = Some,*<br>*3 = Not very much,*<br>*4 = Not at all* |
| **Mother relationship**<br>***(Quality of relationship with mother)***<br>**[Organized domain: Emotional experiences]**<br>Please think about your relationship with your mother when you were growing up. In general, would you say that relationship was very good, somewhat good, somewhat bad, or very bad? | **Close to someone**<br>***(Feel close to someone)***<br>Is there any one special person you know that you feel very close to?<br>*1 = Yes, 2 = No* | **Feel anxious[§]**<br>***(Feeling nervous or anxious)***<br>Feeling nervous, anxious or on edge.<br>*1 = Nearly every day,*<br>*2 = More than half the days,*<br>*3 = Several days,*<br>*4 = Not at all* | **Income difference[§]**<br>***(Support reducing differences in income levels)***<br>The government should take measures to reduce differences in income levels.<br>*1 = Strongly agree,*<br>*2 = Somewhat agree,*<br>*3 = Neither agree nor disagree,*<br>*4 = Somewhat disagree,* | **Obey law[§]**<br>***(Obeying the law)***<br>Whatever the circumstances, the law should always be obeyed.<br>*1 = Strongly agree,*<br>*2 = Somewhat agree,*<br>*3 = Neither agree nor disagree,*<br>*4 = Somewhat disagree,*<br>*5 = Strongly disagree* | **Threat in life[§]**<br>***(Distress from life threat)***<br>Think about the biggest threat to life you've ever witnessed or experienced first-hand during your life. In the past month, how much have you been bothered by this experience?<br>*1 = A lot, 2 = Some,*<br>*3 = Not very much,* |

| Predictors (Childhood Variables) | Outcomes (continued) | | | | |
|---|---|---|---|---|---|
| *1 = Very good, 2 = Somewhat good, 3 = Somewhat bad, 4 = Very bad* | | | *5 = Strongly disagree* | | *4 = Not at all* |
| **Father relationship**<br>***(Quality of relationship with father)***<br>**[Organized domain: Emotional experiences]**<br>Please think about your relationship with your father when you were growing up. In general, would you say that relationship was very good, somewhat good, somewhat bad, or very bad?<br>*1 = Very good, 2 = Somewhat good, 3 = Somewhat bad, 4 = Very bad* | **Content‡**<br>***(Relationship contentment)***<br>I am content with my friendships and relationships.<br>*0 = Strongly disagree to 10 = Strongly agree* | **Forgive§**<br>***(Forgiveness)***<br>How often have you forgiven those who have hurt you?<br>*1 = Always, 2 = Often, 3 = Rarely, 4 = Never* | **Interest§**<br>***(Have little interest in doing things)***<br>Little interest or pleasure in doing things.<br>*1 = Nearly every day, 2 = More than half the days, 3 = Several days, 4 = Not at all* | **Peace§**<br>***(Feeling at peace)***<br>In general, how often do you feel at peace with your thoughts and feelings?<br>*1 = Always, 2 = Often, 3 = Rarely, 4 = Never* | **Trust§**<br>***(Trust in others in this country)***<br>How many people in this country trust one another?<br>*1 = All, 2 = Most, 3 = Some, 4 = Not very many, 5 = None* |
| **Health growing up**<br>***(Self-rated health while growing up)***<br>**[Organized domain: Health]**<br>In general, how was your health when you were growing up?<br>*1 = Excellent, 2 = Very good, 3 = Good, 4 = Fair, 5 = Poor* | **Control worry§**<br>***(Inability to control worry)***<br>Not being able to stop or control worrying.<br>*1 = Nearly every day, 2 = More than half the days, 3 = Several days, 4 = Not at all* | **Freedom**<br>***(Freedom in life to pursue things of importance)***<br>I have the freedom in my life to pursue the things that are most important to me.<br>*0 = Strongly disagree to 10 = Strongly agree* | **Life balance§**<br>***(Various aspects of life are balanced)***<br>In general, how often are the various aspects of your life in balance?<br>*1 = Always, 2 = Often, 3 = Rarely, 4 = Never* | **People help**<br>***(Perceived support from others when oneself is in trouble)***<br>If you were in trouble, how often could you count on people in your life, like relatives or friends, to help you whenever you need them?<br>*0 = Never to 10 = Always* | **Volunteered**<br>***(Volunteered at an organization)***<br>In the past month, have you volunteered your time to an organization?<br>*1 = Yes, 2 = No* |
| **Household income at the age of 12 years old**<br>***(Perceived family financial situation while growing up)***<br>**[Organized domain: Structural conditions]**<br>Which one of these phrases comes closest to your own feelings about your family's household income when you were growing up, such as when you were around 12 years old?<br>*1 = Lived comfortably, 2 = Got by, 3 = Found it difficult, 4 = Found it very difficult* | **Depressed§**<br>***(Depressive symptoms)***<br>Feeling down, depressed or hopeless.<br>*1 = Nearly every day, 2 = More than half the days, 3 = Several days, 4 = Not at all* | **Give up‡**<br>***(Give up some happiness now for greater happiness later)***<br>I am always able to give up some happiness now for greater happiness later.<br>*0 = Not true of you at all to 10 = Completely true of you* | **Life evaluation five years from now**<br>***(Life evaluation five years from now)***<br>Just your best guess, on which step do you think you will stand in the future, say about five years from now?<br>*0 = Worst possible to 10 = Best possible* | **Physical health‡**<br>***(Physical health)***<br>In general, how would you rate your physical health?<br>*0 = Poor physical health to 10 = Excellent physical health* | **Worry safety§§‡‡**<br>***(Worry about safety, food, or housing)***<br>How often do you worry about safety, food, or housing?<br>*0 = Worry all of the time to 10 = Do not ever worry* |
| | **Discriminated§**<br>***(Perceived discrimination)***<br>How often do you feel discriminated against because of any group you are a part of?<br>*1 = Always, 2 = Often, 3 = Rarely, 4 = Never* | **Grateful**<br>***(Have a long list of gratitude)***<br>If I had to list everything that I felt grateful for, it would be a very long list.<br>*0 = Strongly disagree to 10 = Strongly agree* | **Life evaluation today**<br>***(Life evaluation today)***<br>On which step of the ladder would you say you personally feel you stand at this time?<br>*0 = Worst possible to 10 = Best possible* | **Promote good‡**<br>***(Act to promote good)***<br>I always act to promote good in all circumstances, even in difficult and challenging situations.<br>*0 = Not true of you at all to 10 = Completely true of you* | **Worthwhile‡**<br>***(Things done in life are worthwhile)***<br>Overall, to what extent do you feel the things you do in your life are worthwhile?<br>*0 = Not at all worthwhile to 10 = Completely worthwhile* |

***Note:*** Those marked with ‡ are included in the flourishing composite without financial indicators, and ‡‡ are the financial indicators included in the flourishing composite with financial indicators. The outcome variables marked with § were reverse coded in the association analysis. The outcome variables (*worry about meeting normal monthly living expenses* and *worry about safety, food, or housing*) marked §§ were reverse coded in the association analysis. Within flourishing index calculations, *worry about meeting normal monthly living expenses* and *worry about safety, food, or housing* are on original scales. For binary outcomes, responses with "No" were recoded as 0; responses with "Yes" were recoded as 1 in the association analysis. The outcome variable *say in government* marked with # was coded in the order by level 1 = disagree, level 2 = unsure, and level 3 = agree.

**eTable 2.** Flourishing domain, its indicators, questions, and responses, in the Global Flourishing Study.

| Domain | Flourishing indicators | Questions | Response |
|---|---|---|---|
| **D1. Happiness** | Happy | In general, how happy or unhappy do you usually feel? | *0 = Extremely unhappy to 10 = Extremely happy* |
| | Life satisfaction | Overall, how satisfied are you with life as a whole these days? | *0 = Not at all satisfied with your life to 10 = Completely satisfied with your life* |
| **D2. Health** | Mental health | How would you rate your overall mental health? | *0 = Poor mental health to 10 = Excellent mental health* |
| | Physical health | In general, how would you rate your physical health? | *0 = Poor physical health to 10 = Excellent physical health* |
| **D3. Meaning** | Life purpose | I understand my purpose in life. | *0 = Strongly disagree to 10 = Strongly agree* |
| | Worthwhile | Overall, to what extent do you feel the things you do in your life are worthwhile? | *0 = Not at all worthwhile to 10 = Completely worthwhile* |
| **D4. Character** | Give up | I am always able to give up some happiness now for greater happiness later. | *0 = Not true of you at all to 10 = Completely true of you* |
| | Promote good | I always act to promote good in all circumstances, even in difficult and challenging situations. | *0 = Not true of you at all to 10 = Completely true of you* |
| **D5. Relationships** | Content | I am content with my friendships and relationships. | *0 = Strongly disagree to 10 = Strongly agree* |
| | Satisfied with relationship | My relationships are as satisfying as I would want them to be. | *0 = Strongly disagree to 10 = Strongly agree* |
| **D6. Financial stability** | Expenses | How often do you worry about being able to meet normal monthly living expenses? | *0 = Worry all of the time to 10 = Do not ever worry* |
| | Worry safety | How often do you worry about safety, food, or housing? | *0 = Worry all of the time to 10 Do not ever worry* |

**eTable 3.** Description of 8 childhood predictors and 14 outcomes of interest from mid-year items fielded between Year 1 and 2.

| **Childhood predictors [8]**<br>***(Labels used in this paper)*** | | **Outcomes of interest from mid-year items fielded between Year 1 and 2 [14]**<br>***(Labels used in this paper and figures)*** | | | |
|---|---|---|---|---|---|
| **Abused**<br>***(Childhood abuse)***<br>**[Organized domain: Adverse experiences]**<br>Were you ever physically or sexually abused when you were growing up?<br>*1 = Yes, 2 = No* | **Mother relationship**<br>***(Quality of relationship with mother)***<br>**[Organized domain: Emotional experiences]**<br>Please think about your relationship with your mother when you were growing up. In general, would you say that relationship was very good, somewhat good, somewhat bad, or very bad?<br>*1 = Very good, 2 = Somewhat good, 3 = Somewhat bad, 4 = Very bad* | **Achieving**<br>***(Happy with what you achieve in life)***<br>Are you happy with what you are achieving in life?<br>*1 = Yes, 2 = No* | **Diligent**<br>***(Is diligent)***<br>I am diligent.<br>*0 = Strongly disagree to 10 = Strongly agree* | **Healthy**<br>***(Is important to be healthy)***<br>Being healthy.<br>*0 = Not at all important to 10 = Extremely important* | **Money**<br>***(Is important to have plenty of money to have what is wanted)***<br>Having plenty of money to have what I want.<br>*0 = Not at all important to 10 = Extremely important* |
| **Outsider**<br>***(Feeling like an outsider in one's family when growing up)***<br>**[Organized domain: Adverse experiences]**<br>When you were growing up, did you feel like an outsider in your family?<br>*1 = Yes, 2 = No* | **Father relationship**<br>***(Quality of relationship with father)***<br>**[Organized domain: Emotional experiences]**<br>Please think about your relationship with your father when you were growing up. In general, would you say that relationship was very good, somewhat good, somewhat bad, or very bad?<br>*1 = Very good, 2 = Somewhat good, 3 = Somewhat bad, 4 = Very bad* | **Beauty**<br>***(Regularly experience things considered beautiful)***<br>Do you regularly experience things that you consider beautiful? This may include physical beauty or abstract beauty like that found in music, art, or nature.<br>*1 = Yes, 2 = No* | **Good person**<br>***(Is important to be a good person)***<br>Being a good person.<br>*0 = Not at all important to 10 = Extremely important* | **Meaningful**<br>***(Is important to have a meaningful life)***<br>Having a meaningful life.<br>*0 = Not at all important to 10 = Extremely important* | **Nature**<br>***(Feel very connected to nature)***<br>I feel very connected to nature.<br>*0 = Strong disagree to 10 = Strongly agree* |
| **Mother loved**<br>***(Feeling loved by mother)***<br>**[Organized domain: Emotional experiences]**<br>In general, did you feel loved by your mother when you were growing up?<br>*1 = Yes, 2 = No* | **Health growing up**<br>***(Self-rated health while growing up)***<br>**[Organized domain: Health]**<br>In general, how was your health when you were growing up?<br>*1 = Excellent, 2 = Very good, 3 = Good, 4 = Fair, 5 = Poor* | **Engage arts**[§]<br>***(Often engage in arts or cultural activities)***<br>How often do you engage in an arts or cultural activity, such as singing, painting, playing a musical instrument, drawing, dancing, creative writing, photography, or visiting a museum, theatre, or concert hall? Please think of all the times you engage in these activities, whether it is for enjoyment or you receive payment in return.<br>*1 = More than once a week,*<br>*2 = Once a week,*<br>*3 = One to three times a month,*<br>*4 = A few times a year,*<br>*5 = Never* | **Good relation**<br>***(Is important to have good relationships)***<br>Having good relationships.<br>*0 = Not at all important to 10 = Extremely important* | **Mind focused**<br>***(Mind is focused on what to do)***<br>My mind is focused on what I do.<br>*0 = Never to 10 = Always* | **Time media**<br>***(Time spent on using social media each day)***<br>On average, how much time do you spend each day using social media platforms, such as [EXAMPLE]?<br>*1 = None/I don't use social media,*<br>*2 = Less than 30 minutes,*<br>*3 = 30 minutes or less than an hour,*<br>*4 = 1 to 2 hours,*<br>*5 = More than 2 hours* |
| **Father loved**<br>***(Feeling loved by father)***<br>**[Organized domain: Emotional experiences]**<br>In general, did you feel loved by your father when you were growing up?<br>*1 = Yes, 2 = No* | **Household income at the age of 12 years old**<br>***(Perceived family financial situation while growing up)***<br>**[Organized domain: Structural conditions]**<br>Which one of these phrases comes closest to your own feelings about your family's household income when you were growing up, such as when you were around 12 years old?<br>*1 = Lived comfortably, 2 = Got by, 3 = Found it difficult,*<br>*4 = Found it very difficult* | **Food insecure**[§]<br>***(Household ran out of food in last 12 months)***<br>In the last 12 months, how frequently did your household run out of food because of a lack of money or other resources?<br>*1 = Often,*<br>*2 = Sometimes,*<br>*3 = Never* | **Happy important**<br>***(Is important to be happy)***<br>Being happy.<br>*0 = Not at all important to 10 = Extremely important* | | |

**Notes:** The outcome variables marked with § will be reverse coded in the association analysis. For binary outcomes, responses with "No" will be recoded as 0; responses with "Yes" will be recoded as 1 in the association analysis.

**eTable 4.** Descriptive statistics of states and union territories in India (N = 12,765) in Wave 1 Global Flourishing Data.

| Region Code | Region | n (%) |
|---|---|---|
| 601 | Andhra Pradesh | 460 (5.5) |
| 603 | Assam | 355 (2.9) |
| 604 | Bihar | 1,122 (6.8) |
| 605 | Chandigarh | 22 (0.2) |
| 606 | Chhattisgarh | 193 (2.1) |
| 607 | Delhi | 140 (1.4) |
| 608 | Goa | 23 (0.2) |
| 609 | Gujarat | 755 (6.4) |
| 610 | Haryana | 299 (2.3) |
| 611 | Himachal Pradesh | 53 (0.4) |
| 613 | Jharkhand | 264 (1.8) |
| 614 | Karnataka | 363 (3.4) |
| 615 | Kerala | 87 (1.2) |
| 616 | Madhya Pradesh | 627 (5.6) |
| 617 | Maharashtra | 1,203 (9.1) |
| 622 | Odisha (Orissa) | 620 (4.1) |
| 624 | Punjab | 244 (2.0) |
| 625 | Rajasthan | 763 (4.9) |
| 627 | Tamil Nadu | 607 (7.1) |
| 629 | Uttar Pradesh | 2,497 (17.3) |
| 630 | Uttarakhand | 105 (0.8) |
| 631 | West Bengal | 1,623 (10.3) |
| 632 | Telangana | 340 (4.3) |

***Note:*** Cross-sectional Wave 1 weights were used for weight-adjusted proportions (%).

**eTable 5.** Weighted Pearson correlation of 45 adult outcomes and eight childhood variables.

| Variables | r | Variables | r | Variables | r |
|---|---|---|---|---|---|
| Worry about meeting normal monthly living expenses§§‡‡ - Worry about safety, food, or housing§§‡‡ | 0.590 | Happiness‡ - Life satisfaction‡ | 0.579 | Depressive symptoms§ - Feeling nervous or anxious§ | 0.560 |
| Mental health‡ - Physical health‡ | 0.548 | Relationship contentment‡ - Relationship satisfaction‡ | 0.542 | Feeling nervous or anxious§ - Unable to control worry§ | 0.502 |
| Life satisfaction‡ - Life worthwhile‡ | 0.494 | Not feeling loved by father - Worse relationship with father | 0.474 | Life evaluation five years from now - Life evaluation today | 0.442 |
| Happiness‡ - Life worthwhile‡ | 0.439 | Happiness‡ - Life evaluation today | 0.437 | Depressive symptoms§ - Unable to control worry§ | 0.435 |
| Freedom - Remain hopeful for the future | 0.429 | Life satisfaction‡ - Mental health‡ | 0.425 | Donated to charity - Volunteered at an organization | 0.417 |
| Happiness‡ - Mental health‡ | 0.394 | Remain hopeful for the future - Act to promote good‡ | 0.393 | Remain hopeful for the future - Have a long list of gratitude | 0.392 |
| Positive expectations - Remain hopeful for the future | 0.392 | Life worthwhile‡ - Mental health‡ | 0.388 | Life worthwhile‡ - Physical health‡ | 0.387 |
| Happiness‡ - Physical health‡ | 0.386 | Understand purpose in life‡ - Act to promote good‡ | 0.381 | Life satisfaction‡ - Physical health‡ | 0.381 |
| Life evaluation today - Life satisfaction‡ | 0.380 | Remain hopeful for the future - Understand purpose in life‡ | 0.379 | Freedom - Understand purpose in life‡ | 0.369 |
| Freedom - Relationship satisfaction‡ | 0.369 | Positive expectations - Have a long list of gratitude | 0.360 | Not feeling loved by mother - Worse relationship with mother | 0.358 |
| Happiness‡ - Life evaluation five years from now | 0.354 | Mental health‡ - Act to promote good‡ | 0.351 | Donated to charity - Helped a stranger | 0.346 |
| Freedom - Relationship contentment‡ | 0.346 | Relationship contentment‡ - Understand purpose in life‡ | 0.340 | Extent of suffering§ - Feeling nervous or anxious§ | 0.339 |
| Positive expectations - Act to promote good‡ | 0.336 | Relationship satisfaction‡ - Remain hopeful for the future | 0.334 | Understand purpose in life‡ - Mental health‡ | 0.328 |
| Freedom - Act to promote good‡ | 0.326 | Worse relationship with mother - Worse relationship with father | 0.326 | Alcohol use - Cigarette smoking | 0.326 |
| Life evaluation five years from now - Life satisfaction‡ | 0.325 | Depressive symptoms§ - Distress from life-threatening experiences§ | 0.324 | Feel at peace§ - Various aspects of life are balanced§ | 0.324 |
| Life evaluation today - Life worthwhile‡ | 0.321 | Act to promote good‡ - Have a long list of gratitude | 0.321 | Depressive symptoms§ - Extent of suffering§ | 0.319 |
| Positive expectations - Understand purpose in life‡ | 0.319 | Life worthwhile‡ - Understand purpose in life‡ | 0.319 | Relationship contentment‡ - Mental health‡ | 0.319 |
| Positive expectations - Relationship satisfaction‡ | 0.318 | Life worthwhile‡ - Act to promote good‡ | 0.315 | Freedom - Positive expectations | 0.314 |
| Distress from life-threatening experiences§ - Feeling nervous or anxious§ | 0.313 | Freedom - Have a long list of gratitude | 0.313 | Relationship contentment‡ - Remain hopeful for the future | 0.311 |
| Life satisfaction‡ - Relationship contentment‡ | 0.311 | Distress from life-threatening experiences§ - Extent of suffering§ | 0.310 | Life evaluation five years from now - Life worthwhile‡ | 0.310 |
| Understand purpose in life‡ - Have a long list of gratitude | 0.308 | Relationship satisfaction‡ - Understand purpose in life‡ | 0.305 | Relationship satisfaction‡ - Act to promote good‡ | 0.302 |
| Act to promote good‡ - Give up some happiness now for greater happiness later‡ | 0.300 | Life satisfaction‡ - Act to promote good‡ | 0.300 | Life satisfaction‡ - Relationship satisfaction‡ | 0.299 |
| Relationship contentment‡ - Act to promote good‡ | 0.298 | Freedom - Mental health‡ | 0.297 | Belonging - Remain hopeful for the future | 0.295 |
| Life worthwhile‡ - Relationship contentment‡ | 0.294 | Belonging - Freedom | 0.294 | Life satisfaction‡ - Understand purpose in life‡ | 0.291 |
| Life evaluation today - Mental health‡ | 0.291 | Remain hopeful for the future - Mental health‡ | 0.288 | Freedom - Life worthwhile‡ | 0.287 |
| Relationship contentment‡ - Perceived support from others | 0.286 | Positive expectations - Relationship contentment‡ | 0.285 | Helped a stranger - Volunteered at an organization | 0.284 |
| Physical health‡ - Act to promote good‡ | 0.281 | Relationship satisfaction‡ - Have a long list of gratitude | 0.279 | Happiness‡ - Relationship contentment‡ | 0.279 |
| Relationship satisfaction‡ - Perceived support from others | 0.278 | Life evaluation today - Physical health‡ | 0.278 | Freedom - Life satisfaction‡ | 0.277 |
| Understand purpose in life‡ - Physical health‡ | 0.276 | Distress from life-threatening experiences§ - Unable to control worry§ | 0.275 | Life evaluation five years from now - Act to promote good‡ | 0.275 |
| Life worthwhile‡ - Relationship satisfaction‡ | 0.275 | Life evaluation five years from now - Mental health‡ | 0.273 | Life worthwhile‡ - Remain hopeful for the future | 0.273 |
| Relationship satisfaction‡ - Mental health‡ | 0.270 | Happiness‡ - Act to promote good‡ | 0.269 | Life worthwhile‡ - Positive expectations | 0.269 |
| Belonging - Act to promote good‡ | 0.268 | Extent of suffering§ - Unable to control worry§ | 0.268 | Life satisfaction‡ - Loneliness§ | -0.267 |
| Life evaluation five years from now - Physical health‡ | 0.267 | Freedom - Physical health‡ | 0.266 | Depressive symptoms§ - Have little interest in doing things§ | 0.265 |
| Happiness‡ - Loneliness§ | -0.265 | Relationship satisfaction‡ - Show love often | 0.263 | Belonging - Show love often | 0.262 |
| Relationship contentment‡ - Physical health‡ | 0.262 | Belonging - Relationship contentment‡ | 0.261 | Extent of suffering§ - Physical health‡ | -0.261 |
| Belonging - Mental health‡ | 0.260 | Extent of suffering§ - Have any health problems | 0.260 | Positive expectations - Mental health‡ | 0.260 |
| Understand purpose in life‡ - Give up some happiness now for greater happiness later‡ | 0.258 | Remain hopeful for the future - Give up some happiness now for greater happiness later‡ | 0.257 | Life evaluation five years from now - Remain hopeful for the future | 0.257 |
| Happiness‡ - Understand purpose in life‡ | 0.257 | Remain hopeful for the future - Show love often | 0.256 | Belonging - Understand purpose in life‡ | 0.255 |
| Belonging - Relationship satisfaction‡ | 0.255 | Relationship contentment‡ - Show love often | 0.254 | Freedom - Show love often | 0.254 |
| Life satisfaction‡ - Positive expectations | 0.252 | Loneliness§ - Mental health‡ | -0.252 | Life worthwhile‡ - Have a long list of gratitude | 0.250 |

| Variables | r | Variables | r | Variables | r |
|---|---|---|---|---|---|
| Life satisfaction‡ - Remain hopeful for the future | 0.250 | Belonging - Positive expectations | 0.250 | Happiness‡ - Relationship satisfaction‡ | 0.250 |
| Life evaluation five years from now - Understand purpose in life‡ | 0.249 | Positive expectations - Give up some happiness now for greater happiness later‡ | 0.247 | Give up some happiness now for greater happiness later‡ - Have a long list of gratitude | 0.247 |
| Have little interest in doing things§ - Unable to control worry§ | 0.247 | Belonging - Physical health‡ | 0.246 | Remain hopeful for the future - Physical health‡ | 0.246 |
| Have any health problems - Physical health‡ | -0.244 | Relationship contentment‡ - Have a long list of gratitude | 0.243 | Positive expectations - Show love often | 0.242 |
| Freedom - Happiness‡ | 0.241 | Freedom - Life evaluation five years from now | 0.239 | Belonging - Happiness‡ | 0.238 |
| Act to promote good‡ - Show love often | 0.237 | Extent of suffering§ - Mental health‡ | -0.237 | Belonging - Have a long list of gratitude | 0.235 |
| Life worthwhile‡ - Loneliness§ | -0.235 | Relationship satisfaction‡ - Physical health‡ | 0.234 | Feeling nervous or anxious§ - Physical health‡ | -0.233 |
| Life satisfaction‡ - Perceived support from others | 0.233 | Life evaluation five years from now - Positive expectations | 0.230 | Loneliness§ - Worry about safety, food, or housing§§‡‡ | 0.230 |
| Positive expectations - Physical health‡ | 0.229 | Loneliness§ - Relationship contentment‡ | -0.228 | Obeying the law§ - Support reducing differences in income levels§ | 0.228 |
| Loneliness§ - Depressive symptoms§ | 0.228 | Show love often - Perceived support from others | 0.227 | Belonging - Life satisfaction‡ | 0.227 |
| Understand purpose in life‡ - Show love often | 0.227 | Feeling nervous or anxious§ - Have little interest in doing things§ | 0.226 | Distress from life-threatening experiences§ - Mental health‡ | -0.226 |
| Life worthwhile‡ - Perceived support from others | 0.226 | Happiness‡ - Remain hopeful for the future | 0.225 | Life evaluation five years from now - Relationship contentment‡ | 0.224 |
| Life satisfaction‡ - Give up some happiness now for greater happiness later‡ | 0.223 | Happiness‡ - Positive expectations | 0.223 | Feeling nervous or anxious§ - Mental health‡ | -0.221 |
| Have a long list of gratitude - Show love often | 0.220 | Feeling nervous or anxious§ - Have any health problems | 0.220 | Happiness‡ - Distress from life-threatening experiences§ | -0.219 |
| Mental health‡ - Have a long list of gratitude | 0.218 | Feeling nervous or anxious§ - Worry about meeting normal monthly living expenses§§‡‡ | 0.218 | Life worthwhile‡ - Give up some happiness now for greater happiness later‡ | 0.218 |
| Belonging - Life worthwhile‡ | 0.217 | Loneliness§ - Physical health‡ | -0.217 | Freedom - Give up some happiness now for greater happiness later‡ | 0.216 |
| Life satisfaction‡ - Have a long list of gratitude | 0.215 | Life satisfaction‡ - Distress from life-threatening experiences§ | -0.215 | Happiness‡ - Give up some happiness now for greater happiness later‡ | 0.215 |
| Life evaluation five years from now - Relationship satisfaction‡ | 0.214 | Relationship contentment‡ - Give up some happiness now for greater happiness later‡ | 0.213 | Happiness‡ - Feeling nervous or anxious§ | -0.213 |
| Feeling nervous or anxious§ - Worry about safety, food, or housing§§‡‡ | 0.210 | Belonging - Life evaluation five years from now | 0.210 | Distress from life-threatening experiences§ - Physical health‡ | -0.210 |
| Loneliness§ - Feeling nervous or anxious§ | 0.208 | Not feeling loved by mother - Not feeling loved by father | 0.208 | Life evaluation five years from now - Have a long list of gratitude | 0.207 |
| Loneliness§ - Relationship satisfaction‡ | -0.207 | Happiness‡ - Perceived support from others | 0.207 | Loneliness§ - Worry about meeting normal monthly living expenses§§‡‡ | 0.207 |
| Distress from life-threatening experiences§ - Have any health problems | 0.206 | Life evaluation today - Relationship contentment‡ | 0.204 | Depressive symptoms§ - Physical health‡ | -0.204 |
| Belonging - Give up some happiness now for greater happiness later‡ | 0.203 | Relationship satisfaction‡ - Give up some happiness now for greater happiness later‡ | 0.203 | Depressive symptoms§ - Mental health‡ | -0.202 |
| Mental health‡ - Give up some happiness now for greater happiness later‡ | 0.202 | Freedom - Perceived support from others | 0.201 | Physical health‡ - Give up some happiness now for greater happiness later‡ | 0.200 |
| Mental health‡ - Perceived support from others | 0.200 | Life satisfaction‡ - Depressive symptoms§ | -0.200 | Loneliness§ - Distress from life-threatening experiences§ | 0.200 |
| Feel at peace§ - Feeling capable§ | 0.200 | Life evaluation today - Understand purpose in life‡ | 0.199 | Life evaluation today - Relationship satisfaction‡ | 0.199 |
| Belonging - Loneliness§ | -0.198 | Life satisfaction‡ - Feeling nervous or anxious§ | -0.197 | Life evaluation five years from now - Give up some happiness now for greater happiness later‡ | 0.197 |
| Happiness‡ - Depressive symptoms§ | -0.197 | Life evaluation today - Act to promote good‡ | 0.196 | Depressive symptoms§ - Have any health problems | 0.194 |
| Give up some happiness now for greater happiness later‡ - Show love often | 0.194 | Loneliness§ - Positive expectations | -0.194 | Mental health‡ - Show love often | 0.194 |
| Various aspects of life are balanced§ - Feeling capable§ | 0.192 | Distress from life-threatening experiences§ - Perceived discrimination§ | 0.192 | Physical health‡ - Have a long list of gratitude | 0.191 |
| Happiness‡ - Have a long list of gratitude | 0.191 | Life evaluation today - Loneliness§ | -0.189 | Government approval§ - Say in government# | 0.189 |
| Physical health‡ - Perceived support from others | 0.188 | Life worthwhile‡ - Show love often | 0.187 | Positive expectations - Perceived support from others | 0.187 |
| Depressive symptoms§ - Worry about meeting normal monthly living expenses§§‡‡ | 0.187 | Happiness‡ - Worry about meeting normal monthly living expenses§§‡‡ | -0.187 | Freedom - Loneliness§ | -0.186 |
| Life worthwhile‡ - Distress from life-threatening experiences§ | -0.186 | Depressive symptoms§ - Worry about safety, food, or housing§§‡‡ | 0.185 | Life evaluation today - Perceived support from others | 0.185 |

| Variables | r | Variables | r | Variables | r |
|---|---|---|---|---|---|
| Life worthwhile‡ - Worry about meeting normal monthly living expenses§§‡‡ | -0.185 | Have any health problems - Mental health‡ | -0.184 | Freedom - Life evaluation today | 0.182 |
| Extent of suffering§ - Worry about meeting normal monthly living expenses§§‡‡ | 0.182 | Loneliness§ - Act to promote good‡ | -0.181 | Life satisfaction‡ - Extent of suffering§ | -0.180 |
| Poorer self-rated childhood health - More difficult childhood family financial situation | 0.180 | Life worthwhile‡ - Feeling nervous or anxious§ | -0.180 | Helped a stranger - Feel close to someone | 0.180 |
| Have any health problems - Unable to control worry§ | 0.179 | Life worthwhile‡ - Depressive symptoms§ | -0.178 | Physical health‡ - Unable to control worry§ | -0.176 |
| Unable to control worry§ - Worry about meeting normal monthly living expenses§§‡‡ | 0.176 | Loneliness§ - Unable to control worry§ | 0.176 | Life satisfaction‡ - Show love often | 0.175 |
| Loneliness§ - Understand purpose in life‡ | -0.175 | Life evaluation today - Give up some happiness now for greater happiness later‡ | 0.174 | Mental health‡ - Unable to control worry§ | -0.174 |
| Depressive symptoms§ - Perceived discrimination§ | 0.171 | Belonging - Perceived support from others | 0.170 | Life evaluation five years from now - Show love often | 0.169 |
| Belonging - Life evaluation today | 0.168 | Happiness‡ - Extent of suffering§ | -0.168 | Loneliness§ - Extent of suffering§ | 0.167 |
| Loneliness§ - Remain hopeful for the future | -0.167 | Life evaluation today - Positive expectations | 0.167 | Physical health‡ - Worry about meeting normal monthly living expenses§§‡‡ | -0.166 |
| Loneliness§ - Perceived support from others | -0.166 | Ever physically or sexually abused when growing up - Have any health problems | 0.165 | Happiness‡ - Unable to control worry§ | -0.164 |
| Happiness‡ - Show love often | 0.164 | Have a long list of gratitude - Perceived support from others | 0.162 | Unable to control worry§ - Worry about safety, food, or housing§§‡‡ | 0.162 |
| Physical health‡ - Show love often | 0.161 | Extent of suffering§ - Worry about safety, food, or housing§§‡‡ | 0.161 | Life satisfaction‡ - Unable to control worry§ | -0.160 |
| Life evaluation five years from now - Loneliness§ | -0.160 | Life evaluation five years from now - Perceived support from others | 0.159 | Happiness‡ - Worry about safety, food, or housing§§‡‡ | -0.158 |
| Remain hopeful for the future - Perceived support from others | 0.156 | Extent of suffering§ - Perceived discrimination§ | 0.156 | Life worthwhile‡ - Extent of suffering§ | -0.155 |
| Life satisfaction‡ - Worry about meeting normal monthly living expenses§§‡‡ | -0.154 | Life evaluation today - Remain hopeful for the future | 0.153 | Life evaluation today - Distress from life-threatening experiences§ | -0.151 |
| Understand purpose in life‡ - Perceived support from others | 0.150 | Life worthwhile‡ - Unable to control worry§ | -0.147 | Life worthwhile‡ - Worry about safety, food, or housing§§‡‡ | -0.146 |
| Unable to control worry§ - Perceived discrimination§ | 0.144 | Extent of suffering§ - Have little interest in doing things§ | 0.143 | Mental health‡ - Worry about meeting normal monthly living expenses§§‡‡ | -0.143 |
| Life evaluation today - Have a long list of gratitude | 0.142 | Give up some happiness now for greater happiness later‡ - Perceived support from others | 0.141 | Positive expectations - Feeling capable§ | 0.141 |
| Act to promote good‡ - Perceived support from others | 0.141 | Ever physically or sexually abused when growing up - Unable to control worry§ | 0.139 | Government approval§ - Trust in others§ | 0.138 |
| Relationship contentment‡ - Distress from life-threatening experiences§ | -0.138 | Life satisfaction‡ - Worry about safety, food, or housing§§‡‡ | -0.138 | Feel at peace§ - Life satisfaction‡ | 0.138 |
| Loneliness§ - Have a long list of gratitude | -0.137 | Belonging - Distress from life-threatening experiences§ | -0.137 | Distress from life-threatening experiences§ - Act to promote good‡ | -0.134 |
| Say in government# - Trust in others§ | 0.133 | Feeling capable§ - Forgiveness§ | 0.133 | Feel at peace§ - Life worthwhile‡ | 0.132 |
| Feeling nervous or anxious§ - Perceived discrimination§ | 0.132 | Loneliness§ - Show love often | -0.132 | Donated to charity - Feel close to someone | 0.131 |
| Remain hopeful for the future - Feeling capable§ | 0.131 | Distress from life-threatening experiences§ - Worry about meeting normal monthly living expenses§§‡‡ | 0.130 | Feel at peace§ - Trust in others§ | 0.130 |
| Physical health‡ - Worry about safety, food, or housing§§‡‡ | -0.129 | Life evaluation five years from now - Distress from life-threatening experiences§ | -0.129 | Happiness‡ - Have any health problems | -0.127 |
| Loneliness§ - Have any health problems | 0.126 | Relationship contentment‡ - Extent of suffering§ | -0.126 | Freedom - Distress from life-threatening experiences§ | -0.125 |
| Ever physically or sexually abused when growing up - Distress from life-threatening experiences§ | 0.125 | Understand purpose in life‡ - Feeling capable§ | 0.123 | Act to promote good‡ - Feeling capable§ | 0.123 |
| Life evaluation today - Show love often | 0.123 | Life evaluation today - Feeling nervous or anxious§ | -0.123 | Ever physically or sexually abused when growing up - Perceived discrimination§ | 0.123 |
| Relationship satisfaction‡ - Distress from life-threatening experiences§ | -0.121 | Feeling capable§ - Have a long list of gratitude | 0.121 | Perceived support from others - Trust in others§ | 0.120 |
| Worse relationship with mother - Obeying the law§ | -0.120 | Relationship satisfaction‡ - Trust in others§ | 0.119 | Life evaluation today - Worry about meeting normal monthly living expenses§§‡‡ | -0.119 |

| Variables | r | Variables | r | Variables | r |
|---|---|---|---|---|---|
| Feel at peace§ - Happiness‡ | 0.119 | Relationship contentment‡ - Depressive symptoms§ | -0.118 | Distress from life-threatening experiences§ - Have little interest in doing things§ | 0.117 |
| Various aspects of life are balanced§ - Trust in others§ | 0.117 | Life evaluation today - Extent of suffering§ | -0.117 | Loneliness§ - Perceived discrimination§ | 0.116 |
| Relationship contentment‡ - Trust in others§ | 0.115 | Life evaluation today - Depressive symptoms§ | -0.114 | Not feeling loved by mother - Worse relationship with father | 0.113 |
| Volunteered at an organization - Feel close to someone | 0.112 | Worse relationship with father - More difficult childhood family financial situation | 0.111 | Life evaluation today - Worry about safety, food, or housing§§‡‡ | -0.111 |
| Belonging - Feeling capable§ | 0.111 | Remain hopeful for the future - Distress from life-threatening experiences§ | -0.110 | Distress from life-threatening experiences§ - Worry about safety, food, or housing§§‡‡ | 0.110 |
| More difficult childhood family financial situation - Life evaluation five years from now | -0.110 | Have a long list of gratitude - Say in government# | 0.110 | Life satisfaction‡ - Have any health problems | -0.109 |
| Worse relationship with father - Obeying the law§ | -0.109 | Feeling like an outsider in one’s family when growing up - Donated to charity | 0.109 | Feeling capable§ - Trust in others§ | 0.109 |
| Not feeling loved by father - Worse relationship with mother | 0.108 | Various aspects of life are balanced§ - Have little interest in doing things§ | 0.108 | Have little interest in doing things§ - Trust in others§ | 0.108 |
| Feel at peace§ - Mental health‡ | 0.107 | Mental health‡ - Worry about safety, food, or housing§§‡‡ | -0.107 | Feel at peace§ - Forgiveness§ | 0.106 |
| Ever physically or sexually abused when growing up - Feeling nervous or anxious§ | 0.106 | Understand purpose in life‡ - Distress from life-threatening experiences§ | -0.106 | Ever physically or sexually abused when growing up - Depressive symptoms§ | 0.106 |
| Positive expectations - Distress from life-threatening experiences§ | -0.105 | Relationship satisfaction‡ - Say in government# | 0.104 | Belonging - Government approval§ | 0.104 |
| Various aspects of life are balanced§ - Forgiveness§ | 0.103 | Remain hopeful for the future - Trust in others§ | 0.103 | Belonging - Extent of suffering§ | -0.102 |
| Loneliness§ - Give up some happiness now for greater happiness later‡ | -0.102 | Freedom - Depressive symptoms§ | -0.101 | Life evaluation five years from now - Trust in others§ | 0.101 |
| More difficult childhood family financial situation - Life worthwhile‡ | -0.100 | More difficult childhood family financial situation - Mental health‡ | -0.100 | Poorer self-rated childhood health - Physical health‡ | -0.100 |
| Depressive symptoms§ - Act to promote good‡ | -0.100 | Life evaluation today - Unable to control worry§ | -0.100 | Not feeling loved by father - More difficult childhood family financial situation | 0.099 |
| Feel at peace§ - Act to promote good‡ | 0.098 | Freedom - Feeling capable§ | 0.098 | Happiness‡ - Feeling capable§ | 0.098 |
| More difficult childhood family financial situation - Happiness‡ | -0.098 | Life worthwhile‡ - Feeling capable§ | 0.098 | Relationship contentment‡ - Government approval§ | 0.098 |
| Life evaluation five years from now - Worry about meeting normal monthly living expenses§§‡‡ | -0.097 | Mental health‡ - Feeling capable§ | 0.096 | Feel at peace§ - Positive expectations | 0.096 |
| More difficult childhood family financial situation - Life satisfaction‡ | -0.096 | Relationship contentment‡ - Feeling capable§ | 0.096 | Have any health problems - Have little interest in doing things§ | 0.096 |
| More difficult childhood family financial situation - Physical health‡ | -0.095 | Understand purpose in life‡ - Depressive symptoms§ | -0.094 | Freedom - Extent of suffering§ | -0.094 |
| Life satisfaction‡ - Trust in others§ | 0.094 | Belonging - Trust in others§ | 0.094 | Relationship satisfaction‡ - Feeling capable§ | 0.094 |
| Relationship contentment‡ - Feeling nervous or anxious§ | -0.094 | Relationship contentment‡ - Unable to control worry§ | -0.093 | Relationship satisfaction‡ - Depressive symptoms§ | -0.093 |
| More difficult childhood family financial situation - Life evaluation today | -0.093 | Relationship contentment‡ - Say in government# | 0.092 | Belonging - Feeling nervous or anxious§ | -0.092 |
| Various aspects of life are balanced§ - Act to promote good‡ | 0.092 | Life satisfaction‡ - Feeling capable§ | 0.091 | Happiness‡ - Trust in others§ | 0.091 |
| Feel at peace§ - Freedom | 0.091 | Feel at peace§ - Remain hopeful for the future | 0.090 | Life evaluation five years from now - Depressive symptoms§ | -0.090 |
| Positive expectations - Trust in others§ | 0.089 | Have any health problems - Perceived discrimination§ | 0.089 | Forgiveness§ - Trust in others§ | 0.088 |
| Worse relationship with mother - More difficult childhood family financial situation | 0.088 | Life satisfaction‡ - Government approval§ | 0.088 | Relationship contentment‡ - Worry about meeting normal monthly living expenses§§‡‡ | -0.088 |
| Understand purpose in life‡ - Feeling nervous or anxious§ | -0.088 | Feel at peace§ - Understand purpose in life‡ | 0.088 | Freedom - Feeling nervous or anxious§ | -0.088 |
| Have a long list of gratitude - Trust in others§ | 0.087 | More difficult childhood family financial situation - Extent of suffering§ | 0.087 | Belonging - Worry about meeting normal monthly living expenses§§‡‡ | -0.087 |
| Belonging - Depressive symptoms§ | -0.087 | Understand purpose in life‡ - Have any health problems | -0.087 | Feel at peace§ - Relationship satisfaction‡ | 0.087 |
| Worse relationship with mother - Support reducing differences in income levels§ | -0.087 | Act to promote good‡ - Perceived discrimination§ | -0.087 | Feeling nervous or anxious§ - Act to promote good‡ | -0.087 |
| Have little interest in doing things§ - Worry about meeting normal monthly living expenses§§‡‡ | 0.087 | Depressive symptoms§ - Perceived support from others | -0.087 | Life evaluation five years from now - Extent of suffering§ | -0.086 |

| Variables | r | Variables | r | Variables | r |
|---|---|---|---|---|---|
| Feel at peace§ - Have a long list of gratitude | 0.086 | Remain hopeful for the future - Say in government# | 0.085 | Relationship satisfaction‡ - Government approval§ | 0.084 |
| Volunteered at an organization - Support reducing differences in income levels§ | 0.084 | Unable to control worry§ - Perceived support from others | -0.084 | Ever physically or sexually abused when growing up - Extent of suffering§ | 0.084 |
| Feel at peace§ - Relationship contentment‡ | 0.084 | Feel at peace§ - Show love often | 0.083 | Feeling capable§ - Say in government# | 0.083 |
| Feeling capable§ - Show love often | 0.083 | Feel at peace§ - Loneliness§ | -0.082 | Show love often - Say in government# | 0.082 |
| Remain hopeful for the future - Perceived discrimination§ | -0.082 | Donated to charity - Support reducing differences in income levels§ | 0.081 | Distress from life-threatening experiences§ - Perceived support from others | -0.081 |
| Forgiveness§ - Show love often | 0.081 | Physical health‡ - Feeling capable§ | 0.081 | Belonging - Say in government# | 0.081 |
| Relationship satisfaction‡ - Perceived discrimination§ | -0.081 | Feeling nervous or anxious§ - Perceived support from others | -0.081 | Remain hopeful for the future - Various aspects of life are balanced§ | 0.080 |
| Feel at peace§ - Physical health‡ | 0.080 | Positive expectations - Depressive symptoms§ | -0.080 | Freedom - Say in government# | 0.079 |
| Life evaluation five years from now - Feeling capable§ | 0.079 | Relationship satisfaction‡ - Feeling nervous or anxious§ | -0.079 | Feeling like an outsider in one's family when growing up - Volunteered at an organization | 0.078 |
| Belonging - Perceived discrimination§ | -0.078 | Relationship satisfaction‡ - Extent of suffering§ | -0.078 | Life satisfaction‡ - Various aspects of life are balanced§ | 0.078 |
| Feel at peace§ - Life evaluation today | 0.078 | Remain hopeful for the future - Depressive symptoms§ | -0.078 | Feel at peace§ - Have little interest in doing things§ | 0.077 |
| Various aspects of life are balanced§ - Have a long list of gratitude | 0.077 | Positive expectations - Various aspects of life are balanced§ | 0.077 | Have little interest in doing things§ - Worry about safety, food, or housing§§‡‡ | 0.077 |
| Belonging - Have any health problems | -0.077 | Life worthwhile‡ - Have any health problems | -0.077 | Poorer self-rated childhood health - Extent of suffering§ | 0.076 |
| Poorer self-rated childhood health - Feel close to someone | -0.075 | Loneliness§ - Feeling capable§ | -0.075 | Have little interest in doing things§ - Feeling capable§ | 0.075 |
| Ever physically or sexually abused when growing up - Alcohol use | 0.075 | Extent of suffering§ - Act to promote good‡ | -0.075 | Worse relationship with father - Support reducing differences in income levels§ | -0.075 |
| Have little interest in doing things§ - Perceived discrimination§ | 0.074 | Positive expectations - Say in government# | 0.074 | Have any health problems - Act to promote good‡ | -0.074 |
| Government approval§ - Perceived support from others | 0.074 | Belonging - Feel at peace§ | 0.074 | Freedom - Unable to control worry§ | -0.073 |
| Ever physically or sexually abused when growing up - Cigarette smoking | 0.073 | More difficult childhood family financial situation - Relationship contentment‡ | -0.073 | Mental health‡ - Trust in others§ | 0.073 |
| Freedom - Trust in others§ | 0.073 | Show love often - Trust in others§ | 0.072 | Feeling like an outsider in one's family when growing up - Helped a stranger | 0.072 |
| Relationship contentment‡ - Have any health problems | -0.072 | Belonging - Unable to control worry§ | -0.072 | Life worthwhile‡ - Trust in others§ | 0.072 |
| Freedom - Have any health problems | -0.071 | Life evaluation five years from now - Feeling nervous or anxious§ | -0.071 | More difficult childhood family financial situation - Support reducing differences in income levels§ | -0.071 |
| Understand purpose in life‡ - Trust in others§ | 0.071 | Life evaluation five years from now - Unable to control worry§ | -0.071 | Alcohol use - Perceived support from others | -0.071 |
| Relationship contentment‡ - Perceived discrimination§ | -0.070 | Relationship satisfaction‡ - Unable to control worry§ | -0.070 | Life worthwhile‡ - Various aspects of life are balanced§ | 0.070 |
| Perceived support from others - Worry about meeting normal monthly living expenses§§‡‡ | -0.070 | Feel at peace§ - Perceived support from others | 0.070 | Feel at peace§ - Distress from life-threatening experiences§ | -0.069 |
| Distress from life-threatening experiences§ - Have a long list of gratitude | -0.069 | Understand purpose in life‡ - Extent of suffering§ | -0.069 | Understand purpose in life‡ - Various aspects of life are balanced§ | 0.069 |
| More difficult childhood family financial situation - Belonging | -0.069 | Relationship contentment‡ - Feel close to someone | 0.069 | Understand purpose in life‡ - Unable to control worry§ | -0.069 |
| Act to promote good‡ - Say in government# | 0.069 | More difficult childhood family financial situation - Feel close to someone | -0.068 | Understand purpose in life‡ - Government approval§ | 0.068 |
| More difficult childhood family financial situation - Feeling nervous or anxious§ | 0.068 | Worse relationship with father - Perceived support from others | -0.068 | Freedom - Worry about meeting normal monthly living expenses§§‡‡ | -0.067 |
| More difficult childhood family financial situation - Understand purpose in life‡ | -0.067 | Life evaluation today - Trust in others§ | 0.067 | Forgiveness§ - Government approval§ | 0.067 |
| Physical health‡ - Trust in others§ | 0.067 | Life worthwhile‡ - Government approval§ | 0.066 | More difficult childhood family financial situation - Distress from life-threatening experiences§ | 0.066 |
| Ever physically or sexually abused when growing up - Loneliness§ | 0.066 | Understand purpose in life‡ - Say in government# | 0.066 | Feeling like an outsider in one's family when growing up - Worse relationship with mother | 0.066 |
| Loneliness§ - Trust in others§ | -0.066 | Give up some happiness now for greater happiness later‡ - Worry about meeting normal monthly living expenses§§‡‡ | -0.066 | Remain hopeful for the future - Have little interest in doing things§ | 0.066 |
| Act to promote good‡ - Trust in others§ | 0.065 | Feel at peace§ - Say in government# | 0.065 | Life evaluation five years from now - Have any health problems | -0.065 |

| Variables | r | Variables | r | Variables | r |
|---|---|---|---|---|---|
| Have any health problems - Worry about meeting normal monthly living expenses§§‡‡ | 0.065 | Ever physically or sexually abused when growing up - Have little interest in doing things§ | 0.065 | Feel at peace§ - Life evaluation five years from now | 0.065 |
| More difficult childhood family financial situation - Act to promote good‡ | -0.064 | Have little interest in doing things§ - Show love often | 0.064 | Life evaluation today - Have any health problems | -0.064 |
| Poorer self-rated childhood health - Feeling nervous or anxious§ | 0.064 | Feeling like an outsider in one's family when growing up - Not feeling loved by mother | 0.064 | Worse relationship with mother - Physical health‡ | -0.064 |
| Freedom - Government approval§ | 0.064 | Poorer self-rated childhood health - Life worthwhile‡ | -0.063 | Remain hopeful for the future - Feeling nervous or anxious§ | -0.063 |
| Relationship contentment‡ - Alcohol use | -0.063 | Poorer self-rated childhood health - Mental health‡ | -0.062 | Happiness‡ - Government approval§ | 0.062 |
| Understand purpose in life‡ - Worry about meeting normal monthly living expenses§§‡‡ | -0.062 | Loneliness§ - Government approval§ | -0.062 | Remain hopeful for the future - Have any health problems | -0.062 |
| Feeling like an outsider in one's family when growing up - Worse relationship with father | 0.062 | Distress from life-threatening experiences§ - Trust in others§ | -0.062 | Happiness‡ - Various aspects of life are balanced§ | 0.062 |
| Life evaluation five years from now - Worry about safety, food, or housing§§‡‡ | -0.062 | Volunteered at an organization - Obeying the law§ | 0.062 | Have a long list of gratitude - Worry about meeting normal monthly living expenses§§‡‡ | -0.061 |
| Relationship satisfaction‡ - Feel close to someone | 0.061 | Feeling capable§ - Perceived support from others | 0.061 | Worse relationship with mother - Poorer self-rated childhood health | 0.061 |
| Positive expectations - Perceived discrimination§ | -0.060 | Poorer self-rated childhood health - Worry about meeting normal monthly living expenses§§‡‡ | 0.060 | Cigarette smoking - Perceived support from others | -0.060 |
| More difficult childhood family financial situation - Freedom | -0.059 | Have little interest in doing things§ - Act to promote good‡ | 0.059 | Life satisfaction‡ - Say in government# | 0.059 |
| Have any health problems - Worry about safety, food, or housing§§‡‡ | 0.059 | Not feeling loved by mother - Obeying the law§ | -0.058 | More difficult childhood family financial situation - Depressive symptoms§ | 0.058 |
| More difficult childhood family financial situation - Relationship satisfaction‡ | -0.058 | Various aspects of life are balanced§ - Give up some happiness now for greater happiness later‡ | 0.058 | Worse relationship with father - Poorer self-rated childhood health | 0.058 |
| Unable to control worry§ - Act to promote good‡ | -0.058 | Happiness‡ - Perceived discrimination§ | -0.058 | Helped a stranger - Support reducing differences in income levels§ | 0.057 |
| Extent of suffering§ - Perceived support from others | -0.057 | More difficult childhood family financial situation - Worry about meeting normal monthly living expenses§§‡‡ | 0.057 | Various aspects of life are balanced§ - Mental health‡ | 0.057 |
| More difficult childhood family financial situation - Unable to control worry§ | 0.057 | Life worthwhile‡ - Cigarette smoking | -0.057 | Life evaluation five years from now - Perceived discrimination§ | -0.057 |
| Worse relationship with mother - Relationship contentment‡ | -0.057 | Understand purpose in life‡ - Feel close to someone | 0.057 | Feeling like an outsider in one's family when growing up - Feeling nervous or anxious§ | 0.057 |
| Positive expectations - Have little interest in doing things§ | 0.057 | Not feeling loved by mother - More difficult childhood family financial situation | 0.057 | Relationship satisfaction‡ - Forgiveness§ | 0.057 |
| Happiness‡ - Say in government# | 0.057 | Have little interest in doing things§ - Give up some happiness now for greater happiness later‡ | 0.057 | Feeling like an outsider in one's family when growing up - Feel close to someone | 0.056 |
| Remain hopeful for the future - Government approval§ | 0.056 | Poorer self-rated childhood health - Feel at peace§ | -0.056 | Poorer self-rated childhood health - Happiness‡ | -0.056 |
| Volunteered at an organization - Government approval§ | 0.056 | Worse relationship with father - Mental health‡ | -0.055 | Show love often - Feel close to someone | 0.055 |
| Mental health‡ - Perceived discrimination§ | -0.055 | Poorer self-rated childhood health - Life evaluation five years from now | -0.055 | More difficult childhood family financial situation - Perceived support from others | -0.055 |
| Extent of suffering§ - Government approval§ | -0.055 | Positive expectations - Worry about meeting normal monthly living expenses§§‡‡ | -0.055 | Belonging - Worry about safety, food, or housing§§‡‡ | -0.055 |
| Have little interest in doing things§ - Have a long list of gratitude | 0.055 | Various aspects of life are balanced§ - Unable to control worry§ | 0.054 | Remain hopeful for the future - Extent of suffering§ | -0.054 |
| Belonging - Various aspects of life are balanced§ | 0.054 | Life evaluation today - Say in government# | 0.054 | Life worthwhile‡ - Alcohol use | -0.054 |
| Relationship satisfaction‡ - Have any health problems | -0.054 | More difficult childhood family financial situation - Loneliness§ | 0.054 | Perceived support from others - Say in government# | 0.054 |
| Remain hopeful for the future - Unable to control worry§ | -0.054 | Mental health‡ - Say in government# | 0.054 | Worse relationship with mother - Loneliness§ | 0.053 |
| Life evaluation today - Various aspects of life are balanced§ | 0.053 | Give up some happiness now for greater happiness later‡ - Trust in others§ | 0.053 | Worse relationship with father - Volunteered at an organization | -0.053 |
| Poorer self-rated childhood health - Helped a stranger | -0.053 | Remain hopeful for the future - Forgiveness§ | 0.053 | Donated to charity - Obeying the law§ | 0.053 |
| Understand purpose in life‡ - Forgiveness§ | 0.053 | Poorer self-rated childhood health - Belonging | -0.053 | Worse relationship with mother - Relationship satisfaction‡ | -0.053 |
| Distress from life-threatening experiences§ - Give up some happiness now for greater happiness later‡ | -0.052 | Perceived support from others - Worry about safety, food, or housing§§‡‡ | -0.052 | Positive expectations - Government approval§ | 0.052 |

| Variables | r | Variables | r | Variables | r |
|---|---|---|---|---|---|
| More difficult childhood family financial situation - Cigarette smoking | 0.052 | Freedom - Perceived discrimination§ | -0.052 | Relationship contentment‡ - Various aspects of life are balanced§ | 0.052 |
| Poorer self-rated childhood health - Life satisfaction‡ | -0.052 | Life evaluation five years from now - Feel close to someone | 0.052 | Worse relationship with father - Relationship contentment‡ | -0.052 |
| Feel close to someone - Support reducing differences in income levels§ | 0.052 | Feel at peace§ - Government approval§ | 0.052 | Life worthwhile‡ - Perceived discrimination§ | -0.052 |
| Physical health‡ - Perceived discrimination§ | -0.051 | Give up some happiness now for greater happiness later‡ - Say in government# | 0.051 | Ever physically or sexually abused when growing up - Happiness‡ | -0.051 |
| More difficult childhood family financial situation - Have any health problems | 0.051 | Poorer self-rated childhood health - Distress from life-threatening experiences§ | 0.051 | Worse relationship with father - Relationship satisfaction‡ | -0.051 |
| Positive expectations - Unable to control worry§ | -0.051 | Positive expectations - Feel close to someone | 0.051 | Life evaluation today - Feel close to someone | 0.050 |
| Physical health‡ - Feel close to someone | 0.050 | Freedom - Various aspects of life are balanced§ | 0.050 | More difficult childhood family financial situation - Government approval§ | -0.050 |
| Various aspects of life are balanced§ - Physical health‡ | 0.050 | Distress from life-threatening experiences§ - Feeling capable§ | -0.050 | Show love often - Government approval§ | 0.050 |
| Depressive symptoms§ - Have a long list of gratitude | -0.050 | Understand purpose in life‡ - Have little interest in doing things§ | 0.050 | Relationship satisfaction‡ - Cigarette smoking | -0.050 |
| Happiness‡ - Feel close to someone | 0.050 | Poorer self-rated childhood health - Freedom | -0.049 | Act to promote good‡ - Worry about meeting normal monthly living expenses§§‡‡ | -0.049 |
| Life evaluation today - Government approval§ | 0.049 | Worse relationship with mother - Life evaluation five years from now | -0.049 | Worse relationship with mother - Perceived support from others | -0.049 |
| Feeling capable§ - Give up some happiness now for greater happiness later‡ | 0.049 | Mental health‡ - Feel close to someone | 0.049 | Ever physically or sexually abused when growing up - Volunteered at an organization | 0.049 |
| Worse relationship with mother - Extent of suffering§ | 0.049 | Poorer self-rated childhood health - Have any health problems | 0.049 | Poorer self-rated childhood health - Have little interest in doing things§ | -0.049 |
| Positive expectations - Feeling nervous or anxious§ | -0.049 | Worse relationship with mother - Mental health‡ | -0.049 | Act to promote good‡ - Feel close to someone | 0.049 |
| Physical health‡ - Government approval§ | 0.048 | Not feeling loved by father - Perceived support from others | -0.048 | Various aspects of life are balanced§ - Perceived discrimination§ | 0.048 |
| Worse relationship with mother - Life worthwhile‡ | -0.048 | Relationship satisfaction‡ - Alcohol use | -0.048 | Have little interest in doing things§ - Support reducing differences in income levels§ | -0.047 |
| Poorer self-rated childhood health - Remain hopeful for the future | -0.047 | Poorer self-rated childhood health - Worry about safety, food, or housing§§‡‡ | 0.047 | Act to promote good‡ - Forgiveness§ | 0.047 |
| Alcohol use - Mental health‡ | -0.047 | Feel at peace§ - Depressive symptoms§ | -0.047 | Life satisfaction‡ - Feel close to someone | 0.047 |
| Belonging - Feel close to someone | 0.047 | Life worthwhile‡ - Feel close to someone | 0.047 | More difficult childhood family financial situation - Obeying the law§ | -0.047 |
| Have little interest in doing things§ - Say in government# | 0.047 | Life evaluation today - Cigarette smoking | -0.047 | Relationship contentment‡ - Worry about safety, food, or housing§§‡‡ | -0.046 |
| Happiness‡ - Cigarette smoking | -0.046 | Life worthwhile‡ - Say in government# | 0.046 | Ever physically or sexually abused when growing up - Mental health‡ | -0.046 |
| Feeling like an outsider in one's family when growing up - Perceived support from others | -0.046 | Not feeling loved by mother - Perceived support from others | -0.046 | Life satisfaction‡ - Alcohol use | -0.046 |
| Not feeling loved by father - Relationship contentment‡ | -0.045 | Understand purpose in life‡ - Perceived discrimination§ | -0.045 | Mental health‡ - Government approval§ | 0.045 |
| Positive expectations - Forgiveness§ | 0.045 | More difficult childhood family financial situation - Donated to charity | -0.045 | Poorer self-rated childhood health - Depressive symptoms§ | 0.045 |
| Ever physically or sexually abused when growing up - Say in government# | 0.045 | Ever physically or sexually abused when growing up - Belonging | -0.045 | Forgiveness§ - Say in government# | 0.045 |
| Feeling like an outsider in one's family when growing up - Not feeling loved by father | 0.044 | Life satisfaction‡ - Perceived discrimination§ | -0.044 | Relationship satisfaction‡ - Various aspects of life are balanced§ | 0.044 |
| Feeling like an outsider in one's family when growing up - Unable to control worry§ | 0.044 | Donated to charity - Say in government# | 0.044 | Feeling like an outsider in one's family when growing up - Life evaluation today | -0.044 |
| Life satisfaction‡ - Cigarette smoking | -0.044 | Worse relationship with mother - Freedom | -0.044 | Positive expectations - Have any health problems | -0.044 |
| Poorer self-rated childhood health - Unable to control worry§ | 0.044 | Volunteered at an organization - Say in government# | 0.044 | Not feeling loved by father - Relationship satisfaction‡ | -0.043 |
| Life evaluation today - Alcohol use | -0.043 | Poorer self-rated childhood health - Various aspects of life are balanced§ | -0.043 | More difficult childhood family financial situation - Remain hopeful for the future | -0.043 |

| Variables | r | Variables | r | Variables | r |
|---|---|---|---|---|---|
| Poorer self-rated childhood health - Life evaluation today | -0.043 | Life worthwhile‡ - Have little interest in doing things§ | 0.043 | Forgiveness§ - Perceived discrimination§ | 0.043 |
| Not feeling loved by mother - Relationship satisfaction‡ | -0.043 | Freedom - Feel close to someone | 0.042 | Worse relationship with mother - Feeling nervous or anxious§ | 0.042 |
| Have any health problems - Perceived support from others | -0.042 | Worse relationship with mother - Happiness‡ | -0.042 | Belonging - Helped a stranger | 0.042 |
| Poorer self-rated childhood health - Loneliness§ | 0.042 | Have a long list of gratitude - Helped a stranger | 0.042 | Worse relationship with mother - Life evaluation today | -0.042 |
| Poorer self-rated childhood health - Act to promote good‡ | -0.041 | Understand purpose in life‡ - Worry about safety, food, or housing§§‡‡ | -0.041 | Belonging - Forgiveness§ | 0.041 |
| Poorer self-rated childhood health - Understand purpose in life‡ | -0.041 | Ever physically or sexually abused when growing up - Donated to charity | 0.041 | Feel close to someone - Perceived support from others | 0.041 |
| Have a long list of gratitude - Government approval§ | 0.041 | Life evaluation five years from now - Say in government# | 0.041 | More difficult childhood family financial situation - Give up some happiness now for greater happiness later‡ | -0.041 |
| Life evaluation five years from now - Helped a stranger | 0.041 | Forgiveness§ - Worry about meeting normal monthly living expenses§§‡‡ | 0.041 | Worse relationship with father - Loneliness§ | 0.041 |
| Worse relationship with father - Physical health‡ | -0.041 | Have a long list of gratitude - Perceived discrimination§ | -0.041 | More difficult childhood family financial situation - Volunteered at an organization | -0.041 |
| Poorer self-rated childhood health - Have a long list of gratitude | -0.040 | Ever physically or sexually abused when growing up - Worse relationship with mother | 0.040 | Not feeling loved by father - Government approval§ | -0.040 |
| Worse relationship with mother - Unable to control worry§ | 0.040 | Forgiveness§ - Obeying the law§ | 0.040 | Happiness‡ - Alcohol use | -0.040 |
| Perceived discrimination§ - Worry about safety, food, or housing§§‡‡ | 0.040 | Various aspects of life are balanced§ - Distress from life-threatening experiences§ | -0.040 | Perceived discrimination§ - Worry about meeting normal monthly living expenses§§‡‡ | 0.040 |
| Forgiveness§ - Worry about safety, food, or housing§§‡‡ | 0.040 | Loneliness§ - Have little interest in doing things§ | 0.040 | Alcohol use - Unable to control worry§ | 0.040 |
| Feel close to someone - Trust in others§ | 0.040 | Loneliness§ - Feel close to someone | -0.040 | Relationship contentment‡ - Cigarette smoking | -0.040 |
| Feeling like an outsider in one's family when growing up - Loneliness§ | 0.039 | Extent of suffering§ - Obeying the law§ | -0.039 | Loneliness§ - Say in government# | -0.039 |
| Cigarette smoking - Mental health‡ | -0.039 | Loneliness§ - Alcohol use | 0.039 | Have a long list of gratitude - Feel close to someone | 0.039 |
| Not feeling loved by mother - Support reducing differences in income levels§ | -0.039 | Freedom - Forgiveness§ | 0.039 | Feeling like an outsider in one's family when growing up - Happiness‡ | -0.039 |
| Freedom - Worry about safety, food, or housing§§‡‡ | -0.038 | Forgiveness§ - Have a long list of gratitude | 0.038 | Obeying the law§ - Perceived support from others | 0.038 |
| Relationship satisfaction‡ - Worry about meeting normal monthly living expenses§§‡‡ | -0.038 | Worse relationship with mother - Feel close to someone | -0.038 | Ever physically or sexually abused when growing up - Helped a stranger | 0.038 |
| Ever physically or sexually abused when growing up - Worse relationship with father | 0.038 | Worse relationship with father - Life satisfaction‡ | -0.038 | Worse relationship with mother - Life satisfaction‡ | -0.037 |
| Distress from life-threatening experiences§ - Show love often | -0.037 | Say in government# - Support reducing differences in income levels§ | 0.037 | Remain hopeful for the future - Worry about meeting normal monthly living expenses§§‡‡ | -0.037 |
| Not feeling loved by father - Mental health‡ | -0.037 | Feeling capable§ - Government approval§ | 0.037 | Obeying the law§ - Trust in others§ | 0.037 |
| Life evaluation five years from now - Government approval§ | 0.037 | Worse relationship with mother - Volunteered at an organization | -0.036 | Alcohol use - Obeying the law§ | -0.036 |
| Worse relationship with father - Have little interest in doing things§ | 0.036 | Worse relationship with father - Life evaluation today | -0.036 | Life evaluation five years from now - Cigarette smoking | -0.036 |
| Various aspects of life are balanced§ - Show love often | 0.035 | Government approval§ - Worry about safety, food, or housing§§‡‡ | -0.035 | Worse relationship with father - Cigarette smoking | 0.035 |
| Volunteered at an organization - Perceived discrimination§ | 0.035 | Poorer self-rated childhood health - Feeling capable§ | -0.035 | Give up some happiness now for greater happiness later‡ - Worry about safety, food, or housing§§‡‡ | -0.035 |
| Positive expectations - Worry about safety, food, or housing§§‡‡ | -0.035 | Alcohol use - Support reducing differences in income levels§ | -0.035 | Not feeling loved by father - Alcohol use | 0.034 |
| Mental health‡ - Obeying the law§ | 0.034 | Feeling like an outsider in one's family when growing up - Forgiveness§ | 0.034 | Government approval§ - Worry about meeting normal monthly living expenses§§‡‡ | -0.034 |
| Unable to control worry§ - Have a long list of gratitude | -0.034 | Remain hopeful for the future - Feel close to someone | 0.034 | Not feeling loved by father - Forgiveness§ | -0.034 |
| Not feeling loved by father - Say in government# | -0.034 | Give up some happiness now for greater happiness later‡ - Feel close to someone | 0.034 | Feel at peace§ - Feeling nervous or anxious§ | -0.034 |
| Depressive symptoms§ - Show love often | -0.034 | Freedom - Helped a stranger | 0.034 | Cigarette smoking - Government approval§ | -0.034 |
| Extent of suffering§ - Volunteered at an organization | -0.034 | Ever physically or sexually abused when growing up - Life satisfaction‡ | -0.034 | Ever physically or sexually abused when growing up - Worry about safety, food, or housing§§‡‡ | 0.034 |

| Variables | r | Variables | r | Variables | r |
|---|---|---|---|---|---|
| Give up some happiness now for greater happiness later‡ - Government approval§ | 0.033 | Cigarette smoking - Distress from life-threatening experiences§ | 0.033 | Ever physically or sexually abused when growing up - Feeling like an outsider in one's family when growing up | 0.033 |
| Government approval§ - Support reducing differences in income levels§ | 0.033 | Unable to control worry§ - Forgiveness§ | 0.033 | Feeling nervous or anxious§ - Obeying the law§ | -0.033 |
| Freedom - Donated to charity | 0.033 | More difficult childhood family financial situation - Worry about safety, food, or housing§§‡‡ | 0.033 | Worse relationship with father - Life worthwhile‡ | -0.033 |
| Life evaluation today - Feeling capable§ | 0.033 | Feel at peace§ - Give up some happiness now for greater happiness later‡ | 0.033 | Ever physically or sexually abused when growing up - Not feeling loved by father | 0.033 |
| Alcohol use - Distress from life-threatening experiences§ | 0.033 | Alcohol use - Forgiveness§ | -0.032 | Various aspects of life are balanced§ - Perceived support from others | 0.032 |
| Feeling nervous or anxious§ - Have a long list of gratitude | -0.032 | Distress from life-threatening experiences§ - Helped a stranger | 0.032 | Mental health‡ - Support reducing differences in income levels§ | 0.032 |
| Ever physically or sexually abused when growing up - More difficult childhood family financial situation | 0.032 | Ever physically or sexually abused when growing up - Positive expectations | -0.032 | Show love often - Perceived discrimination§ | -0.032 |
| Feeling like an outsider in one's family when growing up - Distress from life-threatening experiences§ | 0.032 | Alcohol use - Show love often | -0.032 | Worse relationship with father - Happiness‡ | -0.032 |
| Feeling capable§ - Helped a stranger | 0.032 | Worse relationship with mother - Belonging | -0.032 | Helped a stranger - Perceived discrimination§ | 0.032 |
| Relationship contentment‡ - Forgiveness§ | 0.032 | Positive expectations - Helped a stranger | 0.032 | Not feeling loved by father - Life satisfaction‡ | -0.032 |
| Give up some happiness now for greater happiness later‡ - Volunteered at an organization | -0.032 | Donated to charity - Worry about safety, food, or housing§§‡‡ | 0.032 | Not feeling loved by father - Happiness‡ | -0.031 |
| More difficult childhood family financial situation - Say in government# | -0.031 | Cigarette smoking - Feeling capable§ | -0.031 | More difficult childhood family financial situation - Alcohol use | 0.031 |
| Have little interest in doing things§ - Obeying the law§ | -0.031 | Unable to control worry§ - Obeying the law§ | -0.031 | Have any health problems - Support reducing differences in income levels§ | -0.031 |
| Remain hopeful for the future - Helped a stranger | 0.031 | Feeling like an outsider in one's family when growing up - More difficult childhood family financial situation | 0.031 | Have any health problems - Show love often | -0.031 |
| Life evaluation five years from now - Various aspects of life are balanced§ | 0.031 | Cigarette smoking - Unable to control worry§ | 0.031 | Not feeling loved by father - Support reducing differences in income levels§ | -0.031 |
| Not feeling loved by father - Obeying the law§ | -0.030 | Loneliness§ - Various aspects of life are balanced§ | -0.030 | Physical health‡ - Helped a stranger | 0.030 |
| Not feeling loved by father - Depressive symptoms§ | 0.030 | Cigarette smoking - Forgiveness§ | -0.030 | Feeling like an outsider in one's family when growing up - Give up some happiness now for greater happiness later‡ | -0.030 |
| Positive expectations - Volunteered at an organization | -0.030 | Relationship contentment‡ - Obeying the law§ | 0.030 | Worse relationship with father - Government approval§ | -0.029 |
| Extent of suffering§ - Feel close to someone | -0.029 | Worse relationship with father - Freedom | -0.029 | Not feeling loved by father - Freedom | -0.029 |
| Happiness‡ - Volunteered at an organization | -0.029 | Life evaluation today - Support reducing differences in income levels§ | 0.029 | Relationship contentment‡ - Have little interest in doing things§ | 0.029 |
| Feeling like an outsider in one's family when growing up - Mental health‡ | -0.029 | Alcohol use - Government approval§ | -0.029 | Extent of suffering§ - Helped a stranger | 0.029 |
| Not feeling loved by mother - Alcohol use | 0.029 | Forgiveness§ - Support reducing differences in income levels§ | 0.029 | Positive expectations - Alcohol use | -0.028 |
| Poorer self-rated childhood health - Give up some happiness now for greater happiness later‡ | -0.028 | Perceived support from others - Support reducing differences in income levels§ | 0.028 | Poorer self-rated childhood health - Donated to charity | -0.028 |
| Ever physically or sexually abused when growing up - Remain hopeful for the future | -0.028 | Life evaluation today - Have little interest in doing things§ | -0.028 | Feel at peace§ - Helped a stranger | 0.028 |
| Have a long list of gratitude - Worry about safety, food, or housing§§‡‡ | -0.028 | Cigarette smoking - Show love often | -0.028 | Worse relationship with mother - Depressive symptoms§ | 0.028 |
| Cigarette smoking - Physical health‡ | -0.028 | Act to promote good‡ - Worry about safety, food, or housing§§‡‡ | -0.028 | Ever physically or sexually abused when growing up - Government approval§ | 0.028 |
| Have little interest in doing things§ - Forgiveness§ | 0.028 | Poorer self-rated childhood health - Trust in others§ | -0.028 | Unable to control worry§ - Feeling capable§ | -0.028 |
| Helped a stranger - Obeying the law§ | 0.028 | Worse relationship with father - Extent of suffering§ | 0.028 | Positive expectations - Obeying the law§ | -0.027 |
| More difficult childhood family financial situation - Show love often | -0.027 | Feeling capable§ - Feel close to someone | 0.027 | Have any health problems - Feeling capable§ | -0.027 |

| Variables | r | Variables | r | Variables | r |
|---|---|---|---|---|---|
| More difficult childhood family financial situation - Have a long list of gratitude | -0.027 | Poorer self-rated childhood health - Perceived discrimination§ | 0.027 | Not feeling loved by mother - Life evaluation today | -0.027 |
| Feeling nervous or anxious§ - Feeling capable§ | -0.027 | Donated to charity - Forgiveness§ | 0.027 | Belonging - Support reducing differences in income levels§ | 0.027 |
| Worse relationship with mother - Donated to charity | -0.027 | Mental health‡ - Forgiveness§ | 0.027 | Understand purpose in life‡ - Helped a stranger | 0.026 |
| Government approval§ - Obeying the law§ | 0.026 | Worse relationship with mother - Distress from life-threatening experiences§ | 0.026 | Life satisfaction‡ - Forgiveness§ | 0.026 |
| Feeling like an outsider in one's family when growing up - Physical health‡ | -0.026 | Not feeling loved by father - Belonging | -0.026 | Ever physically or sexually abused when growing up - Physical health‡ | -0.026 |
| Not feeling loved by father - Loneliness§ | 0.026 | Feeling like an outsider in one's family when growing up - Worry about safety, food, or housing§§‡‡ | 0.026 | Poorer self-rated childhood health - Say in government# | 0.026 |
| Loneliness§ - Cigarette smoking | 0.026 | Remain hopeful for the future - Worry about safety, food, or housing§§‡‡ | -0.025 | Not feeling loved by father - Life evaluation today | -0.025 |
| Physical health‡ - Obeying the law§ | 0.025 | Various aspects of life are balanced§ - Say in government# | 0.025 | Feeling like an outsider in one's family when growing up - Positive expectations | -0.025 |
| Worse relationship with father - Trust in others§ | -0.025 | Extent of suffering§ - Support reducing differences in income levels§ | -0.025 | Life satisfaction‡ - Support reducing differences in income levels§ | 0.025 |
| Distress from life-threatening experiences§ - Feel close to someone | -0.025 | Relationship satisfaction‡ - Obeying the law§ | 0.025 | Ever physically or sexually abused when growing up - Worry about meeting normal monthly living expenses§§‡‡ | 0.025 |
| Alcohol use - Depressive symptoms§ | 0.025 | Not feeling loved by mother - Loneliness§ | 0.025 | Understand purpose in life‡ - Donated to charity | 0.025 |
| Worse relationship with father - Feeling nervous or anxious§ | 0.024 | Distress from life-threatening experiences§ - Say in government# | -0.024 | Poorer self-rated childhood health - Positive expectations | -0.024 |
| Act to promote good‡ - Government approval§ | 0.024 | Ever physically or sexually abused when growing up - Life evaluation five years from now | -0.024 | Physical health‡ - Say in government# | 0.024 |
| Mental health‡ - Helped a stranger | 0.024 | Not feeling loved by mother - Extent of suffering§ | 0.024 | Life satisfaction‡ - Obeying the law§ | 0.024 |
| Worse relationship with mother - Act to promote good‡ | -0.024 | Worse relationship with father - Feel at peace§ | -0.024 | Ever physically or sexually abused when growing up - Relationship contentment‡ | -0.024 |
| Donated to charity - Give up some happiness now for greater happiness later‡ | -0.024 | Act to promote good‡ - Helped a stranger | 0.024 | Extent of suffering§ - Donated to charity | -0.024 |
| Have any health problems - Feel close to someone | -0.024 | Worse relationship with father - Alcohol use | 0.024 | Have little interest in doing things§ - Donated to charity | -0.023 |
| Feeling like an outsider in one's family when growing up - Life satisfaction‡ | -0.023 | Feeling capable§ - Worry about safety, food, or housing§§‡‡ | 0.023 | Not feeling loved by mother - Poorer self-rated childhood health | 0.023 |
| Feeling like an outsider in one's family when growing up - Feel at peace§ | -0.023 | Worse relationship with father - Forgiveness§ | -0.023 | Forgiveness§ - Give up some happiness now for greater happiness later‡ | 0.023 |
| Positive expectations - Extent of suffering§ | -0.023 | Happiness‡ - Obeying the law§ | 0.023 | Not feeling loved by mother - Say in government# | -0.023 |
| Not feeling loved by mother - Relationship contentment‡ | -0.023 | Feeling like an outsider in one's family when growing up - Depressive symptoms§ | 0.023 | Belonging - Donated to charity | 0.023 |
| Relationship contentment‡ - Support reducing differences in income levels§ | 0.023 | Life evaluation five years from now - Have little interest in doing things§ | 0.022 | Not feeling loved by mother - Mental health‡ | -0.022 |
| Cigarette smoking - Depressive symptoms§ | 0.022 | Remain hopeful for the future - Obeying the law§ | -0.022 | Donated to charity - Trust in others§ | 0.022 |
| Worse relationship with father - Belonging | -0.022 | Feeling like an outsider in one's family when growing up - Government approval§ | 0.022 | Loneliness§ - Volunteered at an organization | 0.022 |
| Depressive symptoms§ - Forgiveness§ | 0.022 | Feeling like an outsider in one's family when growing up - Say in government# | 0.022 | Say in government# - Worry about safety, food, or housing§§‡‡ | 0.022 |
| Worse relationship with mother - Have any health problems | 0.022 | Have little interest in doing things§ - Helped a stranger | 0.022 | Freedom - Have little interest in doing things§ | 0.022 |
| Feeling nervous or anxious§ - Forgiveness§ | 0.022 | Feeling capable§ - Support reducing differences in income levels§ | 0.022 | Feeling like an outsider in one's family when growing up - Cigarette smoking | 0.022 |
| Poorer self-rated childhood health - Support reducing differences in income levels§ | -0.022 | Ever physically or sexually abused when growing up - Life worthwhile‡ | -0.022 | Feeling like an outsider in one's family when growing up - Have any health problems | 0.022 |
| Life evaluation today - Helped a stranger | 0.022 | Unable to control worry§ - Trust in others§ | 0.022 | Not feeling loved by mother - Cigarette smoking | 0.022 |
| Relationship satisfaction‡ - Support reducing differences in income levels§ | 0.022 | Depressive symptoms§ - Feeling capable§ | -0.022 | Relationship satisfaction‡ - Have little interest in doing things§ | 0.022 |

| Variables | r | Variables | r | Variables | r |
|---|---|---|---|---|---|
| Not feeling loved by father - Poorer self-rated childhood health | 0.022 | Ever physically or sexually abused when growing up - Various aspects of life are balanced[§] | 0.022 | Depressive symptoms[§] - Give up some happiness now for greater happiness later[‡] | -0.022 |
| Poorer self-rated childhood health - Obeying the law[§] | -0.021 | Say in government# - Worry about meeting normal monthly living expenses[§§‡‡] | 0.021 | Ever physically or sexually abused when growing up - Act to promote good[‡] | -0.021 |
| Feeling like an outsider in one's family when growing up - Life evaluation five years from now | -0.021 | Support reducing differences in income levels[§] - Trust in others[§] | 0.021 | Worse relationship with father - Distress from life-threatening experiences[§] | 0.021 |
| Life worthwhile[‡] - Obeying the law[§] | 0.021 | Extent of suffering[§] - Say in government# | 0.021 | Not feeling loved by mother - Freedom | -0.021 |
| Happiness[‡] - Support reducing differences in income levels[§] | 0.021 | Cigarette smoking - Worry about safety, food, or housing[§§‡‡] | -0.021 | More difficult childhood family financial situation - Have little interest in doing things[§] | 0.021 |
| Worse relationship with father - Have any health problems | 0.021 | Not feeling loved by mother - Various aspects of life are balanced[§] | 0.020 | Worse relationship with father - Feel close to someone | -0.020 |
| Feeling like an outsider in one's family when growing up - Life worthwhile[‡] | -0.020 | More difficult childhood family financial situation - Trust in others[§] | -0.020 | Feeling like an outsider in one's family when growing up - Poorer self-rated childhood health | -0.020 |
| Remain hopeful for the future - Volunteered at an organization | -0.020 | Worse relationship with father - Depressive symptoms[§] | 0.020 | Various aspects of life are balanced[§] - Worry about meeting normal monthly living expenses[§§‡‡] | 0.020 |
| Life satisfaction[‡] - Have little interest in doing things[§] | 0.020 | Distress from life-threatening experiences[§] - Government approval[§] | -0.020 | Donated to charity - Worry about meeting normal monthly living expenses[§§‡‡] | 0.020 |
| Worse relationship with father - Feeling capable[§] | -0.020 | Not feeling loved by mother - Life worthwhile[‡] | -0.020 | Belonging - Cigarette smoking | -0.020 |
| Have any health problems - Give up some happiness now for greater happiness later[‡] | -0.019 | Ever physically or sexually abused when growing up - Forgiveness[§] | -0.019 | Not feeling loved by father - Feeling nervous or anxious[§] | 0.019 |
| Not feeling loved by father - Distress from life-threatening experiences[§] | 0.019 | Poorer self-rated childhood health - Perceived support from others | -0.019 | Worse relationship with father - Understand purpose in life[‡] | -0.019 |
| Have little interest in doing things[§] - Government approval[§] | 0.019 | Worse relationship with mother - Understand purpose in life[‡] | -0.019 | Not feeling loved by father - Life evaluation five years from now | -0.019 |
| Cigarette smoking - Helped a stranger | 0.019 | Unable to control worry[§] - Show love often | -0.019 | Cigarette smoking - Have any health problems | 0.019 |
| Show love often - Worry about meeting normal monthly living expenses[§§‡‡] | -0.019 | Not feeling loved by father - Unable to control worry[§] | 0.019 | Cigarette smoking - Worry about meeting normal monthly living expenses[§§‡‡] | -0.019 |
| Feeling like an outsider in one's family when growing up - Alcohol use | 0.019 | Distress from life-threatening experiences[§] - Volunteered at an organization | 0.018 | Unable to control worry[§] - Helped a stranger | 0.018 |
| Remain hopeful for the future - Alcohol use | -0.018 | Not feeling loved by father - Act to promote good[‡] | -0.018 | Volunteered at an organization - Trust in others[§] | 0.018 |
| Alcohol use - Have little interest in doing things[§] | 0.018 | Worse relationship with mother - Remain hopeful for the future | -0.018 | Relationship contentment[‡] - Donated to charity | 0.018 |
| Belonging - Alcohol use | -0.018 | Worse relationship with mother - Alcohol use | 0.018 | Alcohol use - Have a long list of gratitude | -0.018 |
| Relationship satisfaction[‡] - Donated to charity | 0.018 | Ever physically or sexually abused when growing up - Feeling capable[§] | -0.018 | Life evaluation today - Perceived discrimination[§] | -0.018 |
| Not feeling loved by father - Extent of suffering[§] | 0.018 | More difficult childhood family financial situation - Helped a stranger | -0.018 | Donated to charity - Government approval[§] | 0.018 |
| Helped a stranger - Show love often | 0.018 | Feeling like an outsider in one's family when growing up - Trust in others[§] | -0.017 | Worse relationship with father - Unable to control worry[§] | 0.017 |
| Feel at peace[§] - Cigarette smoking | -0.017 | Understand purpose in life[‡] - Volunteered at an organization | 0.017 | Cigarette smoking - Feel close to someone | -0.017 |
| Worse relationship with father - Life evaluation five years from now | -0.017 | Happiness[‡] - Helped a stranger | 0.017 | Belonging - Obeying the law[§] | 0.017 |
| Not feeling loved by mother - Life evaluation five years from now | -0.017 | Life worthwhile[‡] - Helped a stranger | 0.017 | Have little interest in doing things[§] - Physical health[‡] | -0.017 |
| Cigarette smoking - Have little interest in doing things[§] | 0.017 | Various aspects of life are balanced[§] - Volunteered at an organization | -0.017 | Poorer self-rated childhood health - Relationship contentment[‡] | -0.017 |
| Various aspects of life are balanced[§] - Extent of suffering[§] | 0.016 | Not feeling loved by father - Trust in others[§] | -0.016 | Give up some happiness now for greater happiness later[‡] - Support reducing differences in income levels[§] | -0.016 |
| Depressive symptoms[§] - Government approval[§] | -0.016 | Positive expectations - Cigarette smoking | -0.016 | Not feeling loved by mother - Depressive symptoms[§] | 0.016 |
| Worse relationship with mother - Have a long list of gratitude | -0.016 | Ever physically or sexually abused when growing up - Poorer self-rated childhood health | 0.016 | More difficult childhood family financial situation - Positive expectations | -0.016 |
| Feeling nervous or anxious[§] - Give up some happiness now for greater happiness later[‡] | -0.016 | Life evaluation today - Forgiveness[§] | -0.016 | Donated to charity - Have a long list of gratitude | 0.016 |

| Variables | r | Variables | r | Variables | r |
|---|---|---|---|---|---|
| Remain hopeful for the future - Support reducing differences in income levels§ | -0.016 | Alcohol use - Feeling nervous or anxious§ | 0.016 | Alcohol use - Donated to charity | -0.016 |
| Various aspects of life are balanced§ - Have any health problems | -0.016 | Have any health problems - Say in government# | 0.016 | Alcohol use - Say in government# | -0.016 |
| Have any health problems - Have a long list of gratitude | -0.016 | Ever physically or sexually abused when growing up - Relationship satisfaction‡ | -0.015 | Donated to charity - Show love often | 0.015 |
| Belonging - Volunteered at an organization | 0.015 | Worse relationship with mother - Various aspects of life are balanced§ | 0.015 | Forgiveness§ - Feel close to someone | 0.015 |
| Understand purpose in life‡ - Cigarette smoking | -0.015 | Unable to control worry§ - Donated to charity | 0.015 | Worse relationship with mother - Government approval§ | -0.015 |
| Feel at peace§ - Have any health problems | -0.015 | Not feeling loved by mother - Physical health‡ | -0.015 | Feel at peace§ - Perceived discrimination§ | 0.015 |
| Life worthwhile‡ - Donated to charity | 0.015 | Forgiveness§ - Perceived support from others | 0.015 | Feeling like an outsider in one's family when growing up - Relationship satisfaction‡ | -0.015 |
| Life worthwhile‡ - Support reducing differences in income levels§ | 0.015 | Cigarette smoking - Donated to charity | 0.015 | Helped a stranger - Say in government# | 0.015 |
| Helped a stranger - Perceived support from others | 0.015 | Life evaluation five years from now - Support reducing differences in income levels§ | 0.015 | Cigarette smoking - Act to promote good‡ | -0.015 |
| Not feeling loved by father - Show love often | -0.015 | Remain hopeful for the future - Donated to charity | 0.015 | Unable to control worry§ - Give up some happiness now for greater happiness later‡ | -0.015 |
| Worse relationship with father - Donated to charity | -0.014 | Have any health problems - Forgiveness§ | -0.014 | Loneliness§ - Forgiveness§ | -0.014 |
| Have little interest in doing things§ - Perceived support from others | -0.014 | Helped a stranger - Government approval§ | -0.014 | Support reducing differences in income levels§ - Worry about safety, food, or housing§§‡‡ | 0.014 |
| Feel at peace§ - Support reducing differences in income levels§ | 0.014 | Not feeling loved by father - Positive expectations | -0.014 | Various aspects of life are balanced§ - Worry about safety, food, or housing§§‡‡ | 0.014 |
| Not feeling loved by father - Helped a stranger | 0.014 | Have little interest in doing things§ - Volunteered at an organization | -0.014 | Not feeling loved by mother - Belonging | -0.014 |
| Cigarette smoking - Trust in others§ | -0.014 | Worse relationship with mother - Positive expectations | -0.014 | Mental health‡ - Donated to charity | 0.014 |
| Distress from life-threatening experiences§ - Obeying the law§ | -0.014 | Give up some happiness now for greater happiness later‡ - Obeying the law§ | -0.013 | Act to promote good‡ - Donated to charity | 0.013 |
| Not feeling loved by father - Worry about meeting normal monthly living expenses§§‡‡ | 0.013 | More difficult childhood family financial situation - Feel at peace§ | -0.013 | Ever physically or sexually abused when growing up - Trust in others§ | 0.013 |
| Have little interest in doing things§ - Mental health‡ | -0.013 | Cigarette smoking - Give up some happiness now for greater happiness later‡ | -0.013 | Have any health problems - Trust in others§ | 0.013 |
| Feeling like an outsider in one's family when growing up - Remain hopeful for the future | -0.013 | Extent of suffering§ - Trust in others§ | 0.013 | Forgiveness§ - Volunteered at an organization | 0.013 |
| Alcohol use - Physical health‡ | -0.013 | Depressive symptoms§ - Helped a stranger | 0.013 | Life evaluation five years from now - Forgiveness§ | 0.013 |
| Cigarette smoking - Have a long list of gratitude | -0.012 | Show love often - Volunteered at an organization | -0.012 | Not feeling loved by father - Have any health problems | 0.012 |
| Understand purpose in life‡ - Obeying the law§ | -0.012 | Ever physically or sexually abused when growing up - Perceived support from others | -0.012 | Positive expectations - Support reducing differences in income levels§ | -0.012 |
| Not feeling loved by father - Feel close to someone | -0.012 | Cigarette smoking - Say in government# | -0.012 | Worse relationship with father - Perceived discrimination§ | 0.012 |
| Relationship satisfaction‡ - Worry about safety, food, or housing§§‡‡ | -0.012 | Physical health‡ - Donated to charity | 0.012 | Not feeling loved by mother - Remain hopeful for the future | 0.012 |
| Extent of suffering§ - Feeling capable§ | 0.012 | Ever physically or sexually abused when growing up - Show love often | -0.012 | Alcohol use - Worry about meeting normal monthly living expenses§§‡‡ | -0.012 |
| Not feeling loved by mother - Donated to charity | -0.012 | Not feeling loved by mother - Government approval§ | -0.012 | Worse relationship with mother - Show love often | -0.012 |
| More difficult childhood family financial situation - Feeling capable§ | -0.012 | Not feeling loved by mother - Feeling nervous or anxious§ | 0.012 | Have any health problems - Government approval§ | 0.012 |
| Feeling like an outsider in one's family when growing up - Freedom | -0.012 | Depressive symptoms§ - Volunteered at an organization | 0.011 | Give up some happiness now for greater happiness later‡ - Helped a stranger | 0.011 |
| Not feeling loved by father - Feel at peace§ | -0.011 | Various aspects of life are balanced§ - Cigarette smoking | 0.011 | Not feeling loved by father - Life worthwhile‡ | -0.011 |
| Extent of suffering§ - Have a long list of gratitude | -0.011 | Alcohol use - Have any health problems | 0.011 | Life worthwhile‡ - Forgiveness§ | 0.011 |
| Not feeling loved by father - Physical health‡ | -0.011 | Extent of suffering§ - Show love often | -0.011 | Feeling like an outsider in one's family when growing up - Support reducing differences in income levels§ | 0.011 |

| Variables | r | Variables | r | Variables | r |
|---|---|---|---|---|---|
| Cigarette smoking - Volunteered at an organization | 0.011 | Distress from life-threatening experiences§ - Support reducing differences in income levels§ | -0.011 | Belonging - Have little interest in doing things§ | 0.011 |
| Support reducing differences in income levels§ - Worry about meeting normal monthly living expenses§§‡‡ | 0.011 | Feel close to someone - Worry about meeting normal monthly living expenses§§‡‡ | -0.011 | Ever physically or sexually abused when growing up - Freedom | -0.011 |
| Alcohol use - Extent of suffering§ | 0.011 | Feeling like an outsider in one's family when growing up - Obeying the law§ | 0.011 | More difficult childhood family financial situation - Perceived discrimination§ | 0.011 |
| Freedom - Volunteered at an organization | 0.011 | Feeling like an outsider in one's family when growing up - Extent of suffering§ | 0.011 | Have any health problems - Obeying the law§ | -0.011 |
| Life evaluation five years from now - Volunteered at an organization | -0.011 | Worse relationship with mother - Perceived discrimination§ | 0.010 | Not feeling loved by father - Feeling capable§ | -0.010 |
| Not feeling loved by father - Perceived discrimination§ | 0.010 | Happiness‡ - Forgiveness§ | 0.010 | Various aspects of life are balanced§ - Support reducing differences in income levels§ | 0.010 |
| Feel at peace§ - Donated to charity | 0.010 | Physical health‡ - Support reducing differences in income levels§ | 0.010 | Poorer self-rated childhood health - Government approval§ | 0.010 |
| Remain hopeful for the future - Cigarette smoking | -0.010 | Mental health‡ - Volunteered at an organization | -0.010 | Various aspects of life are balanced§ - Feeling nervous or anxious§ | 0.010 |
| Perceived discrimination§ - Support reducing differences in income levels§ | 0.010 | Ever physically or sexually abused when growing up - Not feeling loved by mother | 0.010 | Loneliness§ - Donated to charity | 0.010 |
| Show love often - Worry about safety, food, or housing§§‡‡ | 0.010 | Worse relationship with mother - Forgiveness§ | 0.010 | Freedom - Support reducing differences in income levels§ | 0.010 |
| Not feeling loved by mother - Worry about meeting normal monthly living expenses§§‡‡ | 0.010 | Feeling like an outsider in one's family when growing up - Understand purpose in life‡ | 0.010 | Not feeling loved by mother - Worry about safety, food, or housing§§‡‡ | 0.010 |
| Feeling nervous or anxious§ - Helped a stranger | 0.010 | Life evaluation five years from now - Donated to charity | 0.010 | Not feeling loved by mother - Feel close to someone | -0.010 |
| Feeling nervous or anxious§ - Support reducing differences in income levels§ | -0.010 | Ever physically or sexually abused when growing up - Feel at peace§ | -0.010 | Alcohol use - Give up some happiness now for greater happiness later‡ | -0.010 |
| Poorer self-rated childhood health - Show love often | -0.010 | Life evaluation today - Volunteered at an organization | -0.010 | Distress from life-threatening experiences§ - Donated to charity | 0.009 |
| Various aspects of life are balanced§ - Alcohol use | 0.009 | Worse relationship with father - Worry about meeting normal monthly living expenses§§‡‡ | 0.009 | Worse relationship with mother - Feel at peace§ | -0.009 |
| Life evaluation today - Donated to charity | 0.009 | Life worthwhile‡ - Volunteered at an organization | -0.009 | Unable to control worry§ - Volunteered at an organization | 0.009 |
| Worse relationship with father - Have a long list of gratitude | -0.009 | Feeling nervous or anxious§ - Trust in others§ | 0.009 | Cigarette smoking - Perceived discrimination§ | 0.009 |
| Depressive symptoms§ - Donated to charity | -0.009 | Relationship contentment‡ - Volunteered at an organization | 0.009 | Extent of suffering§ - Forgiveness§ | -0.009 |
| Have any health problems - Donated to charity | 0.009 | Not feeling loved by father - Give up some happiness now for greater happiness later‡ | -0.009 | Worse relationship with mother - Cigarette smoking | 0.008 |
| Freedom - Cigarette smoking | -0.008 | Various aspects of life are balanced§ - Helped a stranger | 0.008 | Cigarette smoking - Feeling nervous or anxious§ | 0.008 |
| Have a long list of gratitude - Obeying the law§ | -0.008 | Cigarette smoking - Support reducing differences in income levels§ | -0.008 | Feel at peace§ - Worry about safety, food, or housing§§‡‡ | 0.008 |
| Volunteered at an organization - Perceived support from others | -0.008 | Loneliness§ - Obeying the law§ | -0.008 | Poorer self-rated childhood health - Relationship satisfaction‡ | -0.008 |
| Worse relationship with father - Act to promote good‡ | -0.008 | Freedom - Alcohol use | -0.008 | Cigarette smoking - Extent of suffering§ | 0.008 |
| Poorer self-rated childhood health - Alcohol use | 0.008 | Obeying the law§ - Say in government# | 0.008 | Not feeling loved by father - Donated to charity | -0.008 |
| Feel at peace§ - Worry about meeting normal monthly living expenses§§‡‡ | -0.008 | Life evaluation five years from now - Alcohol use | -0.008 | Feeling like an outsider in one's family when growing up - Feeling capable§ | -0.008 |
| Not feeling loved by mother - Have little interest in doing things§ | 0.007 | Loneliness§ - Support reducing differences in income levels§ | -0.007 | Ever physically or sexually abused when growing up - Feel close to someone | 0.007 |
| Not feeling loved by father - Volunteered at an organization | -0.007 | Feel at peace§ - Obeying the law§ | 0.007 | Physical health‡ - Forgiveness§ | 0.007 |
| Feeling nervous or anxious§ - Show love often | -0.007 | Various aspects of life are balanced§ - Government approval§ | 0.007 | Not feeling loved by father - Remain hopeful for the future | 0.007 |
| Not feeling loved by mother - Distress from life-threatening experiences§ | 0.007 | Not feeling loved by father - Cigarette smoking | -0.007 | Feeling nervous or anxious§ - Donated to charity | 0.007 |
| Distress from life-threatening experiences§ - Forgiveness§ | 0.007 | Poorer self-rated childhood health - Volunteered at an organization | -0.007 | Depressive symptoms§ - Trust in others§ | -0.007 |
| Feeling like an outsider in one's family when growing up - Have little interest in doing things§ | 0.007 | Feel at peace§ - Feel close to someone | 0.007 | Not feeling loved by mother - Helped a stranger | -0.007 |
| Helped a stranger - Trust in others§ | 0.007 | Feeling capable§ - Perceived discrimination§ | 0.007 | Ever physically or sexually abused when growing up - Life evaluation today | 0.007 |
| Worse relationship with mother - Helped a stranger | -0.007 | Not feeling loved by mother - Show love often | -0.007 | Worse relationship with mother - Say in government# | -0.007 |

| Variables | r | Variables | r | Variables | r |
|---|---|---|---|---|---|
| Life evaluation today - Obeying the law[§] | -0.007 | Feel close to someone - Obeying the law[§] | 0.007 | Positive expectations - Donated to charity | -0.007 |
| Act to promote good[‡] - Obeying the law[§] | -0.007 | Not feeling loved by father - Understand purpose in life[‡] | -0.007 | Not feeling loved by father - Various aspects of life are balanced[§] | -0.007 |
| Happiness[‡] - Donated to charity | -0.007 | Depressive symptoms[§] - Feel close to someone | -0.007 | Obeying the law[§] - Perceived discrimination[§] | 0.006 |
| Worse relationship with father - Positive expectations | -0.006 | Feel at peace[§] - Alcohol use | -0.006 | Act to promote good[‡] - Volunteered at an organization | -0.006 |
| Worse relationship with father - Give up some happiness now for greater happiness later[‡] | 0.006 | Act to promote good[‡] - Support reducing differences in income levels[§] | -0.006 | Feeling like an outsider in one’s family when growing up - Various aspects of life are balanced[§] | -0.006 |
| Unable to control worry[§] - Government approval[§] | 0.006 | Loneliness[§] - Helped a stranger | 0.006 | Understand purpose in life[‡] - Support reducing differences in income levels[§] | -0.006 |
| Relationship satisfaction[‡] - Volunteered at an organization | -0.006 | Various aspects of life are balanced[§] - Obeying the law[§] | -0.006 | Forgiveness[§] - Helped a stranger | 0.006 |
| Have a long list of gratitude - Volunteered at an organization | 0.006 | Not feeling loved by mother - Act to promote good[‡] | -0.006 | Have any health problems - Volunteered at an organization | 0.005 |
| Government approval[§] - Perceived discrimination[§] | -0.005 | Ever physically or sexually abused when growing up - Have a long list of gratitude | 0.005 | Worse relationship with father - Remain hopeful for the future | 0.005 |
| Show love often - Obeying the law[§] | 0.005 | Not feeling loved by mother - Forgiveness[§] | -0.005 | Not feeling loved by mother - Unable to control worry[§] | 0.005 |
| Alcohol use - Feel close to someone | -0.005 | Helped a stranger - Worry about meeting normal monthly living expenses[§§‡‡] | -0.005 | Life satisfaction[‡] - Volunteered at an organization | -0.005 |
| Ever physically or sexually abused when growing up - Give up some happiness now for greater happiness later[‡] | 0.005 | Donated to charity - Perceived support from others | -0.005 | Various aspects of life are balanced[§] - Feel close to someone | 0.005 |
| Not feeling loved by mother - Have a long list of gratitude | 0.005 | Not feeling loved by father - Have little interest in doing things[§] | 0.005 | Unable to control worry[§] - Say in government# | -0.005 |
| Perceived discrimination[§] - Say in government# | 0.005 | Trust in others[§] - Worry about meeting normal monthly living expenses[§§‡‡] | -0.005 | Helped a stranger - Worry about safety, food, or housing[§§‡‡] | 0.005 |
| Volunteered at an organization - Worry about meeting normal monthly living expenses[§§‡‡] | 0.005 | Relationship satisfaction[‡] - Helped a stranger | 0.005 | Feeling like an outsider in one’s family when growing up - Relationship contentment[‡] | -0.005 |
| Feel close to someone - Say in government# | 0.004 | Worse relationship with father - Show love often | -0.004 | Trust in others[§] - Worry about safety, food, or housing[§§‡‡] | -0.004 |
| Feel close to someone - Government approval[§] | 0.004 | Various aspects of life are balanced[§] - Donated to charity | 0.004 | Alcohol use - Volunteered at an organization | 0.004 |
| Worse relationship with mother - Have little interest in doing things[§] | 0.004 | Donated to charity - Perceived discrimination[§] | 0.004 | Poorer self-rated childhood health - Forgiveness[§] | 0.004 |
| Happiness[‡] - Have little interest in doing things[§] | 0.004 | Poorer self-rated childhood health - Cigarette smoking | -0.004 | Worse relationship with mother - Worry about meeting normal monthly living expenses[§§‡‡] | -0.004 |
| Alcohol use - Feeling capable[§] | -0.004 | Not feeling loved by mother - Understand purpose in life[‡] | 0.004 | Life satisfaction[‡] - Donated to charity | -0.004 |
| Ever physically or sexually abused when growing up - Obeying the law[§] | -0.004 | Depressive symptoms[§] - Obeying the law[§] | -0.004 | Not feeling loved by mother - Happiness[‡] | -0.004 |
| Alcohol use - Trust in others[§] | -0.004 | Not feeling loved by mother - Feeling capable[§] | 0.004 | Not feeling loved by mother - Feel at peace[§] | 0.003 |
| Ever physically or sexually abused when growing up - Support reducing differences in income levels[§] | 0.003 | Have any health problems - Helped a stranger | 0.003 | More difficult childhood family financial situation - Various aspects of life are balanced[§] | 0.003 |
| Worse relationship with father - Various aspects of life are balanced[§] | 0.003 | Alcohol use - Helped a stranger | 0.003 | Not feeling loved by mother - Trust in others[§] | -0.003 |
| Feeling nervous or anxious[§] - Feel close to someone | -0.003 | Show love often - Support reducing differences in income levels[§] | 0.003 | Not feeling loved by mother - Life satisfaction[‡] | -0.003 |
| Feeling nervous or anxious[§] - Say in government# | 0.003 | Not feeling loved by mother - Volunteered at an organization | -0.003 | Worse relationship with mother - Feeling capable[§] | -0.003 |
| Depressive symptoms[§] - Say in government# | 0.003 | Alcohol use - Act to promote good[‡] | -0.003 | Worse relationship with mother - Trust in others[§] | 0.003 |
| Feeling nervous or anxious[§] - Government approval[§] | -0.003 | More difficult childhood family financial situation - Forgiveness[§] | -0.003 | Not feeling loved by mother - Have any health problems | -0.003 |
| Feel close to someone - Worry about safety, food, or housing[§§‡‡] | 0.003 | Feeling nervous or anxious[§] - Volunteered at an organization | -0.003 | Perceived discrimination[§] - Trust in others[§] | 0.003 |
| Unable to control worry[§] - Support reducing differences in income levels[§] | -0.002 | Life satisfaction[‡] - Helped a stranger | -0.002 | Freedom - Obeying the law[§] | -0.002 |
| Alcohol use - Perceived discrimination[§] | 0.002 | Feel at peace[§] - Volunteered at an organization | 0.002 | Feeling capable[§] - Volunteered at an organization | -0.002 |
| Not feeling loved by mother - Give up some happiness now for greater happiness later[‡] | 0.002 | Cigarette smoking - Obeying the law[§] | 0.002 | Various aspects of life are balanced[§] - Depressive symptoms[§] | -0.002 |
| Extent of suffering[§] - Give up some happiness now for greater happiness later[‡] | -0.002 | Unable to control worry[§] - Feel close to someone | 0.002 | Perceived discrimination[§] - Perceived support from others | -0.002 |

| Variables | r | Variables | r | Variables | r |
|---|---|---|---|---|---|
| Feeling like an outsider in one's family when growing up - Worry about meeting normal monthly living expenses[§§‡‡] | -0.002 | Feeling like an outsider in one's family when growing up - Perceived discrimination[§] | -0.002 | Alcohol use - Worry about safety, food, or housing[§§‡‡] | -0.002 |
| Worse relationship with father - Say in government# | -0.002 | Understand purpose in life[‡] - Alcohol use | -0.002 | Donated to charity - Feeling capable[§] | -0.002 |
| Ever physically or sexually abused when growing up - Understand purpose in life[‡] | 0.002 | Not feeling loved by father - Worry about safety, food, or housing[§§‡‡] | -0.002 | Worse relationship with father - Worry about safety, food, or housing[§§‡‡] | 0.002 |
| Feel at peace[§] - Unable to control worry[§] | 0.002 | Feeling like an outsider in one's family when growing up - Act to promote good[‡] | -0.001 | Feeling capable[§] - Obeying the law[§] | 0.001 |
| Have a long list of gratitude - Support reducing differences in income levels[§] | -0.001 | Life evaluation five years from now - Obeying the law[§] | 0.001 | Not feeling loved by mother - Perceived discrimination[§] | 0.001 |
| Physical health[‡] - Volunteered at an organization | -0.001 | Relationship contentment[‡] - Helped a stranger | -0.001 | Give up some happiness now for greater happiness later[‡] - Perceived discrimination[§] | 0.001 |
| Volunteered at an organization - Worry about safety, food, or housing[§§‡‡] | 0.001 | Feeling like an outsider in one's family when growing up - Show love often | 0.001 | Feeling capable[§] - Worry about meeting normal monthly living expenses[§§‡‡] | -0.001 |
| Have little interest in doing things[§] - Feel close to someone | 0.001 | Feel close to someone - Perceived discrimination[§] | -0.001 | Obeying the law[§] - Worry about meeting normal monthly living expenses[§§‡‡] | 0.001 |
| Feeling like an outsider in one's family when growing up - Belonging | 0.000 | Worse relationship with mother - Worry about safety, food, or housing[§§‡‡] | -0.000 | Feeling like an outsider in one's family when growing up - Have a long list of gratitude | -0.000 |
| Worse relationship with father - Helped a stranger | 0.000 | Worse relationship with mother - Give up some happiness now for greater happiness later[‡] | -0.000 | Not feeling loved by father - Have a long list of gratitude | 0.000 |
| Obeying the law[§] - Worry about safety, food, or housing[§§‡‡] | -0.000 | Depressive symptoms[§] - Support reducing differences in income levels[§] | -0.000 | Not feeling loved by mother - Positive expectations | 0.000 |
| Feel at peace[§] - Extent of suffering[§] | -0.000 | | | | |

**eTable 6.** Weighted Spearman's rank correlation of 45 adult outcomes and eight childhood variables.

| Variables | rho | Variables | rho | Variables | rho |
|---|---|---|---|---|---|
| Worry about meeting normal monthly living expenses§§‡‡ - Worry about safety, food, or housing§§‡‡ | 0.597 | Relationship contentment‡ - Relationship satisfaction‡ | 0.594 | Happiness‡ - Life satisfaction‡ | 0.578 |
| Mental health‡ - Physical health‡ | 0.557 | Depressive symptoms§ - Feeling nervous or anxious§ | 0.556 | Feeling nervous or anxious§ - Unable to control worry§ | 0.508 |
| Life satisfaction‡ - Life worthwhile‡ | 0.506 | Freedom - Remain hopeful for the future | 0.496 | Remain hopeful for the future - Act to promote good‡ | 0.494 |
| Positive expectations - Remain hopeful for the future | 0.484 | Remain hopeful for the future - Have a long list of gratitude | 0.478 | Remain hopeful for the future - Understand purpose in life‡ | 0.477 |
| Understand purpose in life‡ - Act to promote good‡ | 0.466 | Positive expectations - Have a long list of gratitude | 0.453 | Freedom - Understand purpose in life‡ | 0.452 |
| Happiness‡ - Life worthwhile‡ | 0.446 | Freedom - Relationship satisfaction‡ | 0.444 | Depressive symptoms§ - Unable to control worry§ | 0.442 |
| Life evaluation five years from now - Life evaluation today | 0.441 | Life satisfaction‡ - Mental health‡ | 0.439 | Happiness‡ - Life evaluation today | 0.436 |
| Positive expectations - Act to promote good‡ | 0.430 | Relationship satisfaction‡ - Remain hopeful for the future | 0.425 | Freedom - Relationship contentment‡ | 0.420 |
| Relationship contentment‡ - Understand purpose in life‡ | 0.419 | Freedom - Act to promote good‡ | 0.419 | Donated to charity - Volunteered at an organization | 0.417 |
| Positive expectations - Understand purpose in life‡ | 0.417 | Act to promote good‡ - Have a long list of gratitude | 0.414 | Positive expectations - Relationship satisfaction‡ | 0.412 |
| Relationship satisfaction‡ - Act to promote good‡ | 0.408 | Life worthwhile‡ - Mental health‡ | 0.407 | Understand purpose in life‡ - Have a long list of gratitude | 0.405 |
| Life worthwhile‡ - Physical health‡ | 0.404 | Happiness‡ - Mental health‡ | 0.402 | Relationship contentment‡ - Remain hopeful for the future | 0.402 |
| Freedom - Positive expectations | 0.401 | Life satisfaction‡ - Physical health‡ | 0.398 | Freedom - Have a long list of gratitude | 0.398 |
| Relationship satisfaction‡ - Understand purpose in life‡ | 0.396 | Relationship contentment‡ - Act to promote good‡ | 0.396 | Happiness‡ - Physical health‡ | 0.392 |
| Mental health‡ - Act to promote good‡ | 0.388 | Worse relationship with mother - Worse relationship with father | 0.383 | Act to promote good‡ - Give up some happiness now for greater happiness later‡ | 0.380 |
| Belonging - Remain hopeful for the future | 0.377 | Relationship satisfaction‡ - Have a long list of gratitude | 0.376 | Understand purpose in life‡ - Mental health‡ | 0.375 |
| Life evaluation today - Life satisfaction‡ | 0.375 | Relationship contentment‡ - Mental health‡ | 0.373 | Positive expectations - Relationship contentment‡ | 0.370 |
| Belonging - Freedom | 0.368 | Remain hopeful for the future - Show love often | 0.365 | Belonging - Act to promote good‡ | 0.361 |
| Life worthwhile‡ - Understand purpose in life‡ | 0.360 | Relationship satisfaction‡ - Show love often | 0.359 | Happiness‡ - Life evaluation five years from now | 0.356 |
| Freedom - Show love often | 0.353 | Life worthwhile‡ - Act to promote good‡ | 0.352 | Freedom - Mental health‡ | 0.348 |
| Extent of suffering§ - Feeling nervous or anxious§ | 0.347 | Life satisfaction‡ - Relationship contentment‡ | 0.347 | Relationship contentment‡ - Show love often | 0.346 |
| Donated to charity - Helped a stranger | 0.346 | Life satisfaction‡ - Relationship satisfaction‡ | 0.344 | Remain hopeful for the future - Mental health‡ | 0.343 |
| Remain hopeful for the future - Give up some happiness now for greater happiness later‡ | 0.341 | Life evaluation five years from now - Life satisfaction‡ | 0.340 | Positive expectations - Show love often | 0.339 |
| Act to promote good‡ - Show love often | 0.339 | Understand purpose in life‡ - Give up some happiness now for greater happiness later‡ | 0.338 | Belonging - Understand purpose in life‡ | 0.338 |
| Belonging - Show love often | 0.337 | Give up some happiness now for greater happiness later‡ - Have a long list of gratitude | 0.337 | Relationship contentment‡ - Have a long list of gratitude | 0.337 |
| Relationship satisfaction‡ - Mental health‡ | 0.337 | Belonging - Relationship satisfaction‡ | 0.335 | Belonging - Relationship contentment‡ | 0.335 |
| Life satisfaction‡ - Act to promote good‡ | 0.334 | Belonging - Positive expectations | 0.333 | Life evaluation five years from now - Life worthwhile‡ | 0.333 |
| Depressive symptoms§ - Distress from life-threatening experiences§ | 0.333 | Life worthwhile‡ - Relationship contentment‡ | 0.328 | Depressive symptoms§ - Extent of suffering§ | 0.327 |
| Understand purpose in life‡ - Show love often | 0.326 | Alcohol use - Cigarette smoking | 0.326 | Positive expectations - Give up some happiness now for greater happiness later‡ | 0.325 |
| Obeying the law§ - Support reducing differences in income levels§ | 0.325 | Life worthwhile‡ - Relationship satisfaction‡ | 0.324 | Life satisfaction‡ - Understand purpose in life‡ | 0.323 |
| Feel at peace§ - Various aspects of life are balanced§ | 0.321 | Not feeling loved by father - Worse relationship with father | 0.321 | Distress from life-threatening experiences§ - Feeling nervous or anxious§ | 0.320 |
| Belonging - Have a long list of gratitude | 0.320 | Freedom - Life satisfaction‡ | 0.319 | Have a long list of gratitude - Show love often | 0.319 |
| Life evaluation today - Life worthwhile‡ | 0.316 | Freedom - Life worthwhile‡ | 0.316 | Life evaluation five years from now - Act to promote good‡ | 0.314 |
| Distress from life-threatening experiences§ - Extent of suffering§ | 0.313 | Physical health‡ - Act to promote good‡ | 0.312 | Life worthwhile‡ - Positive expectations | 0.312 |
| Relationship contentment‡ - Perceived support from others | 0.310 | Positive expectations - Mental health‡ | 0.309 | Life worthwhile‡ - Remain hopeful for the future | 0.309 |
| Understand purpose in life‡ - Physical health‡ | 0.306 | Relationship contentment‡ - Physical health‡ | 0.306 | Relationship satisfaction‡ - Perceived support from others | 0.305 |
| Freedom - Physical health‡ | 0.305 | Life evaluation five years from now - Mental health‡ | 0.304 | Belonging - Mental health‡ | 0.303 |

| Variables | rho | Variables | rho | Variables | rho |
|---|---|---|---|---|---|
| Life worthwhile‡ - Have a long list of gratitude | 0.301 | Freedom - Give up some happiness now for greater happiness later‡ | 0.299 | Life evaluation five years from now - Remain hopeful for the future | 0.297 |
| Happiness‡ - Relationship contentment‡ | 0.295 | Relationship satisfaction‡ - Give up some happiness now for greater happiness later‡ | 0.294 | Life satisfaction‡ - Remain hopeful for the future | 0.293 |
| Life evaluation today - Mental health‡ | 0.291 | Life evaluation five years from now - Understand purpose in life‡ | 0.290 | Distress from life-threatening experiences§ - Unable to control worry§ | 0.289 |
| Relationship satisfaction‡ - Physical health‡ | 0.288 | Relationship contentment‡ - Give up some happiness now for greater happiness later‡ | 0.285 | Life evaluation five years from now - Physical health‡ | 0.285 |
| Helped a stranger - Volunteered at an organization | 0.284 | Mental health‡ - Have a long list of gratitude | 0.284 | Life satisfaction‡ - Positive expectations | 0.283 |
| Extent of suffering§ - Unable to control worry§ | 0.282 | Remain hopeful for the future - Physical health‡ | 0.281 | Happiness‡ - Relationship satisfaction‡ | 0.279 |
| Life evaluation today - Physical health‡ | 0.278 | Freedom - Life evaluation five years from now | 0.277 | Happiness‡ - Act to promote good‡ | 0.275 |
| Life evaluation five years from now - Positive expectations | 0.275 | Happiness‡ - Understand purpose in life‡ | 0.273 | Life satisfaction‡ - Give up some happiness now for greater happiness later‡ | 0.272 |
| Happiness‡ - Loneliness§ | -0.270 | Life satisfaction‡ - Loneliness§ | -0.269 | Positive expectations - Physical health‡ | 0.269 |
| Give up some happiness now for greater happiness later‡ - Show love often | 0.268 | Life satisfaction‡ - Have a long list of gratitude | 0.268 | Belonging - Give up some happiness now for greater happiness later‡ | 0.267 |
| Belonging - Physical health‡ | 0.267 | Life worthwhile‡ - Give up some happiness now for greater happiness later‡ | 0.267 | Life evaluation five years from now - Relationship contentment‡ | 0.266 |
| Mental health‡ - Show love often | 0.263 | Depressive symptoms§ - Have little interest in doing things§ | 0.263 | Life evaluation five years from now - Have a long list of gratitude | 0.262 |
| Life evaluation five years from now - Relationship satisfaction‡ | 0.262 | Extent of suffering§ - Have any health problems | 0.262 | Loneliness§ - Mental health‡ | -0.260 |
| Show love often - Perceived support from others | 0.260 | Freedom - Happiness‡ | 0.259 | Not feeling loved by mother - Worse relationship with mother | 0.259 |
| Belonging - Life satisfaction‡ | 0.258 | Life evaluation five years from now - Give up some happiness now for greater happiness later‡ | 0.257 | Life satisfaction‡ - Perceived support from others | 0.256 |
| Mental health‡ - Give up some happiness now for greater happiness later‡ | 0.254 | Extent of suffering§ - Physical health‡ | -0.252 | Life worthwhile‡ - Perceived support from others | 0.250 |
| Have little interest in doing things§ - Unable to control worry§ | 0.245 | Life worthwhile‡ - Show love often | 0.245 | Belonging - Life worthwhile‡ | 0.245 |
| Physical health‡ - Give up some happiness now for greater happiness later‡ | 0.244 | Physical health‡ - Have a long list of gratitude | 0.244 | Happiness‡ - Give up some happiness now for greater happiness later‡ | 0.243 |
| Life worthwhile‡ - Loneliness§ | -0.243 | Happiness‡ - Positive expectations | 0.240 | Belonging - Life evaluation five years from now | 0.239 |
| Loneliness§ - Relationship contentment‡ | -0.238 | Happiness‡ - Remain hopeful for the future | 0.237 | Belonging - Happiness‡ | 0.234 |
| Life satisfaction‡ - Show love often | 0.231 | Freedom - Perceived support from others | 0.230 | Have any health problems - Physical health‡ | -0.227 |
| Loneliness§ - Relationship satisfaction‡ | -0.227 | Extent of suffering§ - Mental health‡ | -0.226 | Feeling nervous or anxious§ - Have little interest in doing things§ | 0.224 |
| Happiness‡ - Perceived support from others | 0.224 | Loneliness§ - Depressive symptoms§ | 0.224 | Mental health‡ - Perceived support from others | 0.224 |
| Positive expectations - Perceived support from others | 0.222 | Happiness‡ - Have a long list of gratitude | 0.221 | Loneliness§ - Physical health‡ | -0.221 |
| Feeling nervous or anxious§ - Worry about meeting normal monthly living expenses§§‡‡ | 0.219 | Physical health‡ - Show love often | 0.218 | Life evaluation five years from now - Show love often | 0.216 |
| Feeling nervous or anxious§ - Have any health problems | 0.216 | Happiness‡ - Distress from life-threatening experiences§ | -0.215 | Physical health‡ - Perceived support from others | 0.215 |
| Distress from life-threatening experiences§ - Mental health‡ | -0.214 | Feeling nervous or anxious§ - Worry about safety, food, or housing§§‡‡ | 0.213 | Government approval§ - Say in government# | 0.210 |
| Feeling nervous or anxious§ - Physical health‡ | -0.210 | Life satisfaction‡ - Distress from life-threatening experiences§ | -0.210 | Not feeling loved by mother - Not feeling loved by father | 0.208 |
| Distress from life-threatening experiences§ - Have any health problems | 0.208 | Have a long list of gratitude - Perceived support from others | 0.207 | Loneliness§ - Positive expectations | -0.205 |
| Loneliness§ - Feeling nervous or anxious§ | 0.204 | Loneliness§ - Distress from life-threatening experiences§ | 0.202 | Feel at peace§ - Feeling capable§ | 0.201 |
| Freedom - Loneliness§ | -0.200 | Belonging - Loneliness§ | -0.200 | Distress from life-threatening experiences§ - Physical health‡ | -0.200 |
| Happiness‡ - Feeling nervous or anxious§ | -0.199 | Distress from life-threatening experiences§ - Perceived discrimination§ | 0.199 | Belonging - Perceived support from others | 0.198 |
| Loneliness§ - Act to promote good‡ | -0.198 | Life evaluation today - Relationship contentment‡ | 0.197 | Loneliness§ - Worry about safety, food, or housing§§‡‡ | 0.195 |
| Remain hopeful for the future - Perceived support from others | 0.194 | Life evaluation today - Relationship satisfaction‡ | 0.193 | Loneliness§ - Understand purpose in life‡ | -0.192 |
| Depressive symptoms§ - Have any health problems | 0.191 | Life evaluation today - Understand purpose in life‡ | 0.190 | Happiness‡ - Show love often | 0.190 |

| Variables | rho | Variables | rho | Variables | rho |
|---|---|---|---|---|---|
| Understand purpose in life[‡] - Perceived support from others | 0.190 | Depressive symptoms[§] - Physical health[‡] | -0.190 | Life evaluation today - Perceived support from others | 0.190 |
| Depressive symptoms[§] - Worry about meeting normal monthly living expenses[§§‡‡] | 0.189 | Feeling nervous or anxious[§] - Mental health[‡] | -0.189 | Various aspects of life are balanced[§] - Feeling capable[§] | 0.188 |
| Life evaluation today - Loneliness[§] | -0.187 | Depressive symptoms[§] - Worry about safety, food, or housing[§§‡‡] | 0.187 | Happiness[‡] - Depressive symptoms[§] | -0.186 |
| Loneliness[§] - Remain hopeful for the future | -0.186 | Life satisfaction[‡] - Depressive symptoms[§] | -0.186 | Life worthwhile[‡] - Distress from life-threatening experiences[§] | -0.185 |
| Life evaluation five years from now - Perceived support from others | 0.184 | Life evaluation today - Act to promote good[‡] | 0.184 | Extent of suffering[§] - Worry about meeting normal monthly living expenses[§§‡‡] | 0.183 |
| Life evaluation today - Give up some happiness now for greater happiness later[‡] | 0.182 | Have any health problems - Unable to control worry[§] | 0.181 | Give up some happiness now for greater happiness later[‡] - Perceived support from others | 0.181 |
| Unable to control worry[§] - Worry about meeting normal monthly living expenses[§§‡‡] | 0.181 | Loneliness[§] - Worry about meeting normal monthly living expenses[§§‡‡] | 0.180 | Helped a stranger - Feel close to someone | 0.180 |
| Act to promote good[‡] - Perceived support from others | 0.180 | Freedom - Life evaluation today | 0.180 | Depressive symptoms[§] - Perceived discrimination[§] | 0.179 |
| Worse relationship with mother - Obeying the law[§] | -0.179 | Depressive symptoms[§] - Mental health[‡] | -0.178 | Life satisfaction[‡] - Feeling nervous or anxious[§] | -0.178 |
| Life satisfaction[‡] - Extent of suffering[§] | -0.177 | Loneliness[§] - Perceived support from others | -0.177 | Poorer self-rated childhood health - More difficult childhood family financial situation | 0.176 |
| Loneliness[§] - Unable to control worry[§] | 0.174 | Have any health problems - Mental health[‡] | -0.171 | Life evaluation five years from now - Loneliness[§] | -0.171 |
| Loneliness[§] - Extent of suffering[§] | 0.170 | Life worthwhile[‡] - Depressive symptoms[§] | -0.167 | Physical health[‡] - Unable to control worry[§] | -0.166 |
| Ever physically or sexually abused when growing up - Have any health problems | 0.165 | Unable to control worry[§] - Worry about safety, food, or housing[§§‡‡] | 0.165 | Life worthwhile[‡] - Feeling nervous or anxious[§] | -0.164 |
| Life evaluation today - Positive expectations | 0.164 | Happiness[‡] - Extent of suffering[§] | -0.163 | Extent of suffering[§] - Worry about safety, food, or housing[§§‡‡] | 0.163 |
| Loneliness[§] - Have a long list of gratitude | -0.163 | Mental health[‡] - Unable to control worry[§] | -0.160 | Positive expectations - Feeling capable[§] | 0.159 |
| Happiness[‡] - Unable to control worry[§] | -0.159 | Happiness[‡] - Worry about meeting normal monthly living expenses[§§‡‡] | -0.157 | Extent of suffering[§] - Perceived discrimination[§] | 0.157 |
| Life satisfaction[‡] - Unable to control worry[§] | -0.155 | Government approval[§] - Trust in others[§] | 0.155 | Worse relationship with mother - Support reducing differences in income levels[§] | -0.153 |
| Worse relationship with father - Obeying the law[§] | -0.152 | Loneliness[§] - Show love often | -0.152 | Belonging - Life evaluation today | 0.152 |
| Life evaluation today - Distress from life-threatening experiences[§] | -0.151 | Worse relationship with father - Support reducing differences in income levels[§] | -0.151 | Unable to control worry[§] - Perceived discrimination[§] | 0.148 |
| Extent of suffering[§] - Have little interest in doing things[§] | 0.148 | Life worthwhile[‡] - Extent of suffering[§] | -0.148 | Feel at peace[§] - Life satisfaction[‡] | 0.148 |
| Remain hopeful for the future - Feeling capable[§] | 0.148 | Life evaluation today - Remain hopeful for the future | 0.147 | Feel at peace[§] - Life worthwhile[‡] | 0.147 |
| Life evaluation today - Have a long list of gratitude | 0.146 | Feeling capable[§] - Have a long list of gratitude | 0.145 | Life worthwhile[‡] - Unable to control worry[§] | -0.145 |
| Understand purpose in life[‡] - Feeling capable[§] | 0.144 | Act to promote good[‡] - Feeling capable[§] | 0.143 | Feeling capable[§] - Forgiveness[§] | 0.142 |
| Life worthwhile[‡] - Worry about meeting normal monthly living expenses[§§‡‡] | -0.142 | Ever physically or sexually abused when growing up - Unable to control worry[§] | 0.137 | Belonging - Feeling capable[§] | 0.136 |
| Feeling nervous or anxious[§] - Perceived discrimination[§] | 0.133 | Feel at peace[§] - Happiness[‡] | 0.132 | Say in government# - Trust in others[§] | 0.131 |
| Donated to charity - Feel close to someone | 0.131 | Relationship contentment[‡] - Distress from life-threatening experiences[§] | -0.131 | Feel at peace[§] - Trust in others[§] | 0.130 |
| Belonging - Government approval[§] | 0.130 | Distress from life-threatening experiences[§] - Worry about meeting normal monthly living expenses[§§‡‡] | 0.130 | Feel at peace[§] - Mental health[‡] | 0.129 |
| Belonging - Distress from life-threatening experiences[§] | -0.128 | Happiness[‡] - Worry about safety, food, or housing[§§‡‡] | -0.128 | Life evaluation five years from now - Distress from life-threatening experiences[§] | -0.128 |
| Ever physically or sexually abused when growing up - Distress from life-threatening experiences[§] | 0.126 | Loneliness[§] - Perceived discrimination[§] | 0.125 | Distress from life-threatening experiences[§] - Act to promote good[‡] | -0.124 |
| Life evaluation today - Show love often | 0.124 | Loneliness[§] - Have any health problems | 0.123 | Freedom - Distress from life-threatening experiences[§] | -0.122 |
| Distress from life-threatening experiences[§] - Have little interest in doing things[§] | 0.122 | Perceived support from others - Trust in others[§] | 0.122 | Life worthwhile[‡] - Feeling capable[§] | 0.121 |
| Physical health[‡] - Worry about meeting normal monthly living expenses[§§‡‡] | -0.121 | Relationship satisfaction[‡] - Trust in others[§] | 0.121 | Freedom - Feeling capable[§] | 0.121 |
| Relationship contentment[‡] - Extent of suffering[§] | -0.120 | Ever physically or sexually abused when growing up - Perceived discrimination[§] | 0.120 | Happiness[‡] - Have any health problems | -0.119 |

| Variables | rho | Variables | rho | Variables | rho |
|---|---|---|---|---|---|
| Loneliness[§] - Give up some happiness now for greater happiness later[‡] | -0.119 | Relationship satisfaction[‡] - Feeling capable[§] | 0.118 | Relationship contentment[‡] - Government approval[§] | 0.118 |
| Life evaluation today - Feeling nervous or anxious[§] | -0.117 | Mental health[‡] - Feeling capable[§] | 0.117 | Have a long list of gratitude - Say in government# | 0.116 |
| Relationship satisfaction[‡] - Say in government# | 0.116 | Relationship satisfaction[‡] - Distress from life-threatening experiences[§] | -0.115 | Life evaluation today - Extent of suffering[§] | -0.115 |
| Various aspects of life are balanced[§] - Trust in others[§] | 0.114 | Volunteered at an organization - Feel close to someone | 0.112 | Life evaluation today - Depressive symptoms[§] | -0.112 |
| Life satisfaction[‡] - Worry about meeting normal monthly living expenses[§§‡‡] | -0.111 | Relationship contentment[‡] - Feeling capable[§] | 0.110 | Feel at peace[§] - Positive expectations | 0.110 |
| Distress from life-threatening experiences[§] - Worry about safety, food, or housing[§§‡‡] | 0.109 | Relationship satisfaction[‡] - Government approval[§] | 0.109 | Feeling capable[§] - Trust in others[§] | 0.109 |
| Feeling like an outsider in one's family when growing up - Donated to charity | 0.109 | Feel at peace[§] - Forgiveness[§] | 0.108 | Feel at peace[§] - Relationship satisfaction[‡] | 0.108 |
| Relationship contentment[‡] - Trust in others[§] | 0.107 | Feeling capable[§] - Show love often | 0.107 | Have little interest in doing things[§] - Trust in others[§] | 0.106 |
| Feel at peace[§] - Have a long list of gratitude | 0.106 | Relationship contentment[‡] - Say in government# | 0.106 | Relationship contentment[‡] - Depressive symptoms[§] | -0.106 |
| Act to promote good[‡] - Perceived discrimination[§] | -0.106 | Life evaluation today - Worry about meeting normal monthly living expenses[§§‡‡] | -0.105 | Feel at peace[§] - Act to promote good[‡] | 0.105 |
| Forgiveness[§] - Show love often | 0.105 | Forgiveness[§] - Government approval[§] | 0.105 | Life evaluation five years from now - Trust in others[§] | 0.105 |
| Life satisfaction[‡] - Feeling capable[§] | 0.105 | Various aspects of life are balanced[§] - Have little interest in doing things[§] | 0.105 | Feel at peace[§] - Freedom | 0.104 |
| Ever physically or sexually abused when growing up - Depressive symptoms[§] | 0.104 | Life satisfaction[‡] - Trust in others[§] | 0.103 | Feel at peace[§] - Remain hopeful for the future | 0.103 |
| Happiness[‡] - Feeling capable[§] | 0.103 | More difficult childhood family financial situation - Life evaluation five years from now | -0.102 | Ever physically or sexually abused when growing up - Feeling nervous or anxious[§] | 0.101 |
| Life worthwhile[‡] - Worry about safety, food, or housing[§§‡‡] | -0.101 | Life satisfaction[‡] - Have any health problems | -0.101 | Life satisfaction[‡] - Government approval[§] | 0.100 |
| Feel at peace[§] - Understand purpose in life[‡] | 0.100 | Remain hopeful for the future - Distress from life-threatening experiences[§] | -0.100 | Life evaluation today - Unable to control worry[§] | -0.100 |
| Feel at peace[§] - Show love often | 0.099 | More difficult childhood family financial situation - Happiness[‡] | -0.099 | Feel at peace[§] - Physical health[‡] | 0.099 |
| Worse relationship with father - More difficult childhood family financial situation | 0.098 | Belonging - Trust in others[§] | 0.098 | More difficult childhood family financial situation - Life worthwhile[‡] | -0.098 |
| Various aspects of life are balanced[§] - Forgiveness[§] | 0.098 | Remain hopeful for the future - Perceived discrimination[§] | -0.098 | Have any health problems - Have little interest in doing things[§] | 0.098 |
| Feel at peace[§] - Loneliness[§] | -0.097 | Happiness[‡] - Trust in others[§] | 0.097 | More difficult childhood family financial situation - Life satisfaction[‡] | -0.097 |
| Feel at peace[§] - Relationship contentment[‡] | 0.097 | Positive expectations - Distress from life-threatening experiences[§] | -0.097 | Remain hopeful for the future - Trust in others[§] | 0.096 |
| Understand purpose in life[‡] - Distress from life-threatening experiences[§] | -0.096 | More difficult childhood family financial situation - Mental health[‡] | -0.096 | Show love often - Say in government# | 0.096 |
| Belonging - Perceived discrimination[§] | -0.096 | Physical health[‡] - Feeling capable[§] | 0.096 | Have a long list of gratitude - Trust in others[§] | 0.096 |
| Volunteered at an organization - Support reducing differences in income levels[§] | 0.095 | Have little interest in doing things[§] - Have a long list of gratitude | 0.095 | Donated to charity - Support reducing differences in income levels[§] | 0.095 |
| More difficult childhood family financial situation - Life evaluation today | -0.094 | Not feeling loved by father - More difficult childhood family financial situation | 0.094 | Belonging - Extent of suffering[§] | -0.094 |
| Remain hopeful for the future - Have little interest in doing things[§] | 0.093 | Belonging - Feel at peace[§] | 0.093 | Life evaluation today - Worry about safety, food, or housing[§§‡‡] | -0.093 |
| Remain hopeful for the future - Various aspects of life are balanced[§] | 0.091 | Have little interest in doing things[§] - Worry about meeting normal monthly living expenses[§§‡‡] | 0.091 | Understand purpose in life[‡] - Government approval[§] | 0.091 |
| Mental health[‡] - Trust in others[§] | 0.091 | Relationship satisfaction[‡] - Perceived discrimination[§] | -0.091 | Belonging - Say in government# | 0.091 |
| Life satisfaction[‡] - Worry about safety, food, or housing[§§‡‡] | -0.091 | More difficult childhood family financial situation - Physical health[‡] | -0.090 | Poorer self-rated childhood health - Physical health[‡] | -0.090 |
| Positive expectations - Trust in others[§] | 0.089 | More difficult childhood family financial situation - Extent of suffering[§] | 0.089 | Not feeling loved by mother - Worse relationship with father | 0.089 |
| Life evaluation five years from now - Feeling capable[§] | 0.089 | Freedom - Extent of suffering[§] | -0.089 | Freedom - Say in government# | 0.089 |
| Have any health problems - Perceived discrimination[§] | 0.089 | Mental health[‡] - Government approval[§] | 0.088 | Feeling capable[§] - Say in government# | 0.088 |

| Variables | rho | Variables | rho | Variables | rho |
|---|---|---|---|---|---|
| Various aspects of life are balanced§ - Have a long list of gratitude | 0.087 | Loneliness§ - Feeling capable§ | -0.086 | Relationship contentment‡ - Unable to control worry§ | -0.086 |
| Life satisfaction‡ - Various aspects of life are balanced§ | 0.086 | Various aspects of life are balanced§ - Act to promote good‡ | 0.086 | Life worthwhile‡ - Trust in others§ | 0.086 |
| Life evaluation five years from now - Depressive symptoms§ | -0.086 | Positive expectations - Say in government# | 0.085 | Mental health‡ - Worry about meeting normal monthly living expenses§§‡‡ | -0.085 |
| Freedom - Depressive symptoms§ | -0.085 | Life worthwhile‡ - Government approval§ | 0.085 | Depressive symptoms§ - Perceived support from others | -0.085 |
| Relationship contentment‡ - Perceived discrimination§ | -0.085 | Government approval§ - Perceived support from others | 0.085 | Ever physically or sexually abused when growing up - Extent of suffering§ | 0.084 |
| Forgiveness§ - Trust in others§ | 0.084 | Have little interest in doing things§ - Act to promote good‡ | 0.084 | Physical health‡ - Trust in others§ | 0.083 |
| Distress from life-threatening experiences§ - Perceived support from others | -0.083 | Have little interest in doing things§ - Give up some happiness now for greater happiness later‡ | 0.083 | Have little interest in doing things§ - Show love often | 0.083 |
| Feel at peace§ - Perceived support from others | 0.082 | Understand purpose in life‡ - Have little interest in doing things§ | 0.082 | Depressive symptoms§ - Act to promote good‡ | -0.082 |
| Act to promote good‡ - Say in government# | 0.082 | Show love often - Worry about safety, food, or housing§§‡‡ | 0.081 | Feel at peace§ - Government approval§ | 0.081 |
| Positive expectations - Various aspects of life are balanced§ | 0.081 | Have little interest in doing things§ - Worry about safety, food, or housing§§‡‡ | 0.081 | Physical health‡ - Worry about safety, food, or housing§§‡‡ | -0.081 |
| Freedom - Government approval§ | 0.080 | Feel at peace§ - Life evaluation today | 0.080 | Positive expectations - Have little interest in doing things§ | 0.080 |
| Worse relationship with mother - More difficult childhood family financial situation | 0.079 | Understand purpose in life‡ - Various aspects of life are balanced§ | 0.079 | Not feeling loved by father - Worse relationship with mother | 0.079 |
| Life worthwhile‡ - Various aspects of life are balanced§ | 0.079 | Physical health‡ - Government approval§ | 0.079 | Remain hopeful for the future - Say in government# | 0.079 |
| Relationship satisfaction‡ - Depressive symptoms§ | -0.079 | Feeling like an outsider in one's family when growing up - Volunteered at an organization | 0.078 | More difficult childhood family financial situation - Support reducing differences in income levels§ | -0.078 |
| Relationship contentment‡ - Feeling nervous or anxious§ | -0.078 | Show love often - Government approval§ | 0.078 | Unable to control worry§ - Perceived support from others | -0.077 |
| Feel at peace§ - Life evaluation five years from now | 0.077 | Understand purpose in life‡ - Trust in others§ | 0.077 | Poorer self-rated childhood health - Feel close to someone | -0.076 |
| Remain hopeful for the future - Forgiveness§ | 0.076 | Extent of suffering§ - Act to promote good‡ | -0.076 | Understand purpose in life‡ - Say in government# | 0.076 |
| Poorer self-rated childhood health - Extent of suffering§ | 0.076 | Understand purpose in life‡ - Have any health problems | -0.075 | Have a long list of gratitude - Government approval§ | 0.075 |
| Understand purpose in life‡ - Depressive symptoms§ | -0.075 | Ever physically or sexually abused when growing up - Alcohol use | 0.075 | Life evaluation five years from now - Extent of suffering§ | -0.075 |
| Volunteered at an organization - Obeying the law§ | 0.075 | Donated to charity - Obeying the law§ | 0.074 | Positive expectations - Perceived discrimination§ | -0.074 |
| Feel at peace§ - Have little interest in doing things§ | 0.074 | Life satisfaction‡ - Say in government# | 0.073 | Feeling nervous or anxious§ - Perceived support from others | -0.073 |
| More difficult childhood family financial situation - Relationship contentment‡ | -0.073 | Have little interest in doing things§ - Feeling capable§ | 0.073 | Ever physically or sexually abused when growing up - Cigarette smoking | 0.073 |
| Feel at peace§ - Distress from life-threatening experiences§ | -0.073 | Feeling capable§ - Give up some happiness now for greater happiness later‡ | 0.073 | Relationship contentment‡ - Feel close to someone | 0.072 |
| Feel at peace§ - Say in government# | 0.072 | Volunteered at an organization - Government approval§ | 0.072 | Relationship satisfaction‡ - Forgiveness§ | 0.072 |
| Feeling like an outsider in one's family when growing up - Helped a stranger | 0.072 | Feeling capable§ - Perceived support from others | 0.072 | Belonging - Forgiveness§ | 0.072 |
| Distress from life-threatening experiences§ - Have a long list of gratitude | -0.072 | Extent of suffering§ - Government approval§ | -0.071 | Life evaluation today - Trust in others§ | 0.071 |
| More difficult childhood family financial situation - Feeling nervous or anxious§ | 0.071 | Freedom - Perceived discrimination§ | -0.070 | Remain hopeful for the future - Government approval§ | 0.070 |
| Freedom - Trust in others§ | 0.070 | Various aspects of life are balanced§ - Mental health‡ | 0.070 | Show love often - Trust in others§ | 0.070 |
| Alcohol use - Perceived support from others | -0.070 | Loneliness§ - Trust in others§ | -0.070 | Understand purpose in life‡ - Forgiveness§ | 0.070 |
| Freedom - Unable to control worry§ | -0.069 | Various aspects of life are balanced§ - Give up some happiness now for greater happiness later‡ | 0.069 | Mental health‡ - Say in government# | 0.069 |
| Happiness‡ - Various aspects of life are balanced§ | 0.069 | Relationship satisfaction‡ - Extent of suffering§ | -0.069 | Act to promote good‡ - Forgiveness§ | 0.069 |
| Positive expectations - Depressive symptoms§ | -0.068 | More difficult childhood family financial situation - Feel close to someone | -0.068 | Loneliness§ - Government approval§ | -0.068 |
| Feeling capable§ - Government approval§ | 0.068 | Life evaluation five years from now - Unable to control worry§ | -0.067 | Happiness‡ - Government approval§ | 0.067 |
| Life evaluation five years from now - Perceived discrimination§ | -0.067 | Happiness‡ - Say in government# | 0.067 | Act to promote good‡ - Trust in others§ | 0.067 |
| Mental health‡ - Perceived discrimination§ | -0.067 | Life worthwhile‡ - Have any health problems | -0.067 | Helped a stranger - Support reducing differences in income levels§ | 0.066 |

| Variables | rho | Variables | rho | Variables | rho |
|---|---|---|---|---|---|
| Positive expectations - Government approval§ | 0.066 | Give up some happiness now for greater happiness later‡ - Say in government# | 0.066 | Distress from life-threatening experiences§ - Trust in others§ | -0.066 |
| Freedom - Feeling nervous or anxious§ | -0.066 | Relationship contentment‡ - Have any health problems | -0.066 | Ever physically or sexually abused when growing up - Have little interest in doing things§ | 0.065 |
| Happiness‡ - Perceived discrimination§ | -0.065 | Poorer self-rated childhood health - Feeling nervous or anxious§ | 0.065 | Positive expectations - Forgiveness§ | 0.065 |
| More difficult childhood family financial situation - Distress from life-threatening experiences§ | 0.065 | Relationship contentment‡ - Alcohol use | -0.064 | Understand purpose in life‡ - Feeling nervous or anxious§ | -0.064 |
| Belonging - Various aspects of life are balanced§ | 0.064 | Freedom - Have any health problems | -0.064 | Feeling like an outsider in one’s family when growing up - Not feeling loved by mother | 0.064 |
| Give up some happiness now for greater happiness later‡ - Trust in others§ | 0.064 | More difficult childhood family financial situation - Understand purpose in life‡ | -0.064 | Remain hopeful for the future - Depressive symptoms§ | -0.063 |
| Feeling nervous or anxious§ - Act to promote good‡ | -0.063 | Life worthwhile‡ - Perceived discrimination§ | -0.063 | Belonging - Feeling nervous or anxious§ | -0.063 |
| Have little interest in doing things§ - Perceived discrimination§ | 0.063 | Perceived support from others - Say in government# | 0.063 | Have any health problems - Worry about meeting normal monthly living expenses§§‡‡ | 0.062 |
| Life worthwhile‡ - Cigarette smoking | -0.062 | Belonging - Depressive symptoms§ | -0.062 | Belonging - Have any health problems | -0.062 |
| Worse relationship with mother - Physical health‡ | -0.062 | Cigarette smoking - Perceived support from others | -0.062 | Life worthwhile‡ - Have little interest in doing things§ | 0.062 |
| More difficult childhood family financial situation - Unable to control worry§ | 0.061 | Life evaluation today - Have any health problems | -0.061 | Understand purpose in life‡ - Extent of suffering§ | -0.061 |
| Worse relationship with father - Volunteered at an organization | -0.061 | Relationship satisfaction‡ - Unable to control worry§ | -0.061 | Worse relationship with father - Perceived support from others | -0.061 |
| Freedom - Forgiveness§ | 0.061 | More difficult childhood family financial situation - Depressive symptoms§ | 0.061 | Worse relationship with father - Mental health‡ | -0.060 |
| Ever physically or sexually abused when growing up - Loneliness§ | 0.060 | More difficult childhood family financial situation - Belonging | -0.060 | Feel at peace§ - Depressive symptoms§ | -0.060 |
| Relationship satisfaction‡ - Feel close to someone | 0.059 | Understand purpose in life‡ - Perceived discrimination§ | -0.059 | More difficult childhood family financial situation - Worry about meeting normal monthly living expenses§§‡‡ | 0.059 |
| Show love often - Feel close to someone | 0.059 | Understand purpose in life‡ - Feel close to someone | 0.059 | Various aspects of life are balanced§ - Physical health‡ | 0.059 |
| Life evaluation five years from now - Worry about meeting normal monthly living expenses§§‡‡ | -0.059 | Life evaluation five years from now - Feeling nervous or anxious§ | -0.059 | Extent of suffering§ - Perceived support from others | -0.059 |
| Relationship contentment‡ - Various aspects of life are balanced§ | 0.059 | Poorer self-rated childhood health - Mental health‡ | -0.058 | Forgiveness§ - Have a long list of gratitude | 0.058 |
| Physical health‡ - Perceived discrimination§ | -0.058 | Poorer self-rated childhood health - Worry about meeting normal monthly living expenses§§‡‡ | 0.058 | Distress from life-threatening experiences§ - Feeling capable§ | -0.058 |
| Worse relationship with mother - Mental health‡ | -0.058 | More difficult childhood family financial situation - Perceived support from others | -0.058 | Distress from life-threatening experiences§ - Give up some happiness now for greater happiness later‡ | -0.058 |
| Understand purpose in life‡ - Unable to control worry§ | -0.057 | Freedom - Various aspects of life are balanced§ | 0.057 | Worse relationship with father - Poorer self-rated childhood health | 0.057 |
| Happiness‡ - Feel close to someone | 0.057 | Life evaluation five years from now - Feel close to someone | 0.057 | Have a long list of gratitude - Perceived discrimination§ | -0.057 |
| Life evaluation today - Say in government# | 0.057 | Act to promote good‡ - Government approval§ | 0.056 | Poorer self-rated childhood health - Life worthwhile‡ | -0.056 |
| Feeling like an outsider in one’s family when growing up - Feel close to someone | 0.056 | Have any health problems - Worry about safety, food, or housing§§‡‡ | 0.056 | Poorer self-rated childhood health - Helped a stranger | -0.056 |
| Worse relationship with mother - Loneliness§ | 0.056 | Life evaluation today - Various aspects of life are balanced§ | 0.056 | Relationship satisfaction‡ - Various aspects of life are balanced§ | 0.056 |
| Physical health‡ - Feel close to someone | 0.056 | Life evaluation five years from now - Say in government# | 0.055 | Have little interest in doing things§ - Support reducing differences in income levels§ | -0.055 |
| Life satisfaction‡ - Perceived discrimination§ | -0.055 | More difficult childhood family financial situation - Government approval§ | -0.055 | More difficult childhood family financial situation - Obeying the law§ | -0.055 |
| Feeling like an outsider in one’s family when growing up - Feeling nervous or anxious§ | 0.055 | Poorer self-rated childhood health - Feel at peace§ | -0.055 | Life evaluation five years from now - Have any health problems | -0.054 |
| Belonging - Unable to control worry§ | -0.054 | Relationship satisfaction‡ - Feeling nervous or anxious§ | -0.054 | Life worthwhile‡ - Alcohol use | -0.054 |
| Forgiveness§ - Obeying the law§ | 0.054 | Show love often - Perceived discrimination§ | -0.054 | Positive expectations - Feel close to someone | 0.053 |
| Have any health problems - Act to promote good‡ | -0.053 | More difficult childhood family financial situation - Relationship satisfaction‡ | -0.053 | Worse relationship with mother - Relationship contentment‡ | -0.053 |
| Not feeling loved by mother - More difficult childhood family financial situation | 0.052 | Life worthwhile‡ - Say in government# | 0.052 | Unable to control worry§ - Act to promote good‡ | -0.052 |

| Variables | rho | Variables | rho | Variables | rho |
|---|---|---|---|---|---|
| Mental health‡ - Feel close to someone | 0.052 | Poorer self-rated childhood health - Depressive symptoms§ | 0.052 | Remain hopeful for the future - Worry about safety, food, or housing§§‡‡ | 0.052 |
| Relationship satisfaction‡ - Worry about safety, food, or housing§§‡‡ | 0.051 | More difficult childhood family financial situation - Have any health problems | 0.051 | Relationship contentment‡ - Forgiveness§ | 0.051 |
| Life evaluation today - Feel close to someone | 0.051 | Poorer self-rated childhood health - Happiness‡ | -0.051 | Poorer self-rated childhood health - Distress from life-threatening experiences§ | 0.051 |
| Life satisfaction‡ - Feel close to someone | 0.051 | More difficult childhood family financial situation - Loneliness§ | 0.051 | Feel at peace§ - Give up some happiness now for greater happiness later‡ | 0.051 |
| More difficult childhood family financial situation - Freedom | -0.051 | Positive expectations - Unable to control worry§ | -0.050 | Remain hopeful for the future - Unable to control worry§ | -0.050 |
| Relationship satisfaction‡ - Have any health problems | -0.050 | More difficult childhood family financial situation - Act to promote good‡ | -0.050 | Feeling like an outsider in one's family when growing up - Worse relationship with mother | 0.050 |
| More difficult childhood family financial situation - Cigarette smoking | 0.050 | Mental health‡ - Support reducing differences in income levels§ | 0.050 | Worse relationship with mother - Freedom | -0.050 |
| Act to promote good‡ - Feel close to someone | 0.050 | Government approval§ - Support reducing differences in income levels§ | 0.049 | Ever physically or sexually abused when growing up - Volunteered at an organization | 0.049 |
| Poorer self-rated childhood health - Life evaluation five years from now | -0.049 | Relationship contentment‡ - Have little interest in doing things§ | 0.049 | Remain hopeful for the future - Have any health problems | -0.048 |
| Relationship satisfaction‡ - Have little interest in doing things§ | 0.048 | Forgiveness§ - Say in government# | 0.048 | Happiness‡ - Cigarette smoking | -0.048 |
| Freedom - Have little interest in doing things§ | 0.048 | Not feeling loved by father - Perceived support from others | -0.048 | Poorer self-rated childhood health - Have little interest in doing things§ | -0.048 |
| Worse relationship with father - Relationship satisfaction‡ | -0.048 | Worse relationship with mother - Poorer self-rated childhood health | 0.047 | Poorer self-rated childhood health - Life satisfaction‡ | -0.047 |
| Poorer self-rated childhood health - Have any health problems | 0.047 | Not feeling loved by father - Relationship contentment‡ | -0.047 | Life evaluation today - Government approval§ | 0.047 |
| Mental health‡ - Worry about safety, food, or housing§§‡‡ | -0.047 | Relationship satisfaction‡ - Cigarette smoking | -0.047 | Donated to charity - Say in government# | 0.047 |
| Worse relationship with mother - Relationship satisfaction‡ | -0.047 | Life satisfaction‡ - Alcohol use | -0.047 | Mental health‡ - Forgiveness§ | 0.047 |
| Forgiveness§ - Support reducing differences in income levels§ | 0.047 | Extent of suffering§ - Obeying the law§ | -0.047 | Life evaluation today - Cigarette smoking | -0.047 |
| Ever physically or sexually abused when growing up - Say in government# | 0.047 | Life worthwhile‡ - Feel close to someone | 0.046 | Belonging - Helped a stranger | 0.046 |
| Cigarette smoking - Mental health‡ | -0.046 | Worse relationship with father - Relationship contentment‡ | -0.046 | Life evaluation five years from now - Helped a stranger | 0.046 |
| Have little interest in doing things§ - Say in government# | 0.046 | Freedom - Feel close to someone | 0.046 | Worse relationship with mother - Belonging | -0.046 |
| Belonging - Feel close to someone | 0.046 | Alcohol use - Mental health‡ | -0.046 | Various aspects of life are balanced§ - Show love often | 0.046 |
| Government approval§ - Obeying the law§ | 0.046 | Poorer self-rated childhood health - Unable to control worry§ | 0.046 | Worse relationship with mother - Life worthwhile‡ | -0.046 |
| Volunteered at an organization - Say in government# | 0.046 | Have a long list of gratitude - Helped a stranger | 0.045 | Worse relationship with father - Physical health‡ | -0.045 |
| Depressive symptoms§ - Have a long list of gratitude | -0.045 | Worse relationship with mother - Volunteered at an organization | -0.045 | Loneliness§ - Say in government# | -0.045 |
| Life satisfaction‡ - Cigarette smoking | -0.045 | Act to promote good‡ - Worry about safety, food, or housing§§‡‡ | 0.045 | Poorer self-rated childhood health - Worry about safety, food, or housing§§‡‡ | 0.044 |
| Feeling like an outsider in one's family when growing up - Not feeling loved by father | 0.044 | Poorer self-rated childhood health - Various aspects of life are balanced§ | -0.044 | Not feeling loved by father - Government approval§ | -0.044 |
| Remain hopeful for the future - Feeling nervous or anxious§ | -0.044 | Give up some happiness now for greater happiness later‡ - Government approval§ | 0.044 | Various aspects of life are balanced§ - Unable to control worry§ | 0.044 |
| Distress from life-threatening experiences§ - Show love often | -0.044 | Poorer self-rated childhood health - Freedom | -0.044 | Worse relationship with mother - Life evaluation five years from now | -0.044 |
| More difficult childhood family financial situation - Give up some happiness now for greater happiness later‡ | -0.044 | Have a long list of gratitude - Feel close to someone | 0.044 | Happiness‡ - Alcohol use | -0.044 |
| Feel at peace§ - Feeling nervous or anxious§ | -0.044 | Not feeling loved by father - Relationship satisfaction‡ | -0.044 | More difficult childhood family financial situation - Donated to charity | -0.044 |
| Feeling like an outsider in one's family when growing up - Worse relationship with father | 0.043 | Show love often - Worry about meeting normal monthly living expenses§§‡‡ | 0.043 | Life evaluation today - Alcohol use | -0.043 |
| Mental health‡ - Obeying the law§ | 0.043 | Life evaluation five years from now - Various aspects of life are balanced§ | 0.043 | More difficult childhood family financial situation - Volunteered at an organization | -0.042 |

| Variables | rho | Variables | rho | Variables | rho |
|---|---|---|---|---|---|
| Freedom - Helped a stranger | 0.042 | Remain hopeful for the future - Extent of suffering§ | -0.042 | Feel close to someone - Perceived support from others | 0.042 |
| Not feeling loved by mother - Perceived support from others | -0.042 | Poorer self-rated childhood health - Remain hopeful for the future | -0.042 | Ever physically or sexually abused when growing up - Government approval§ | 0.042 |
| Belonging - Have little interest in doing things§ | 0.042 | Relationship satisfaction‡ - Alcohol use | -0.042 | Relationship contentment‡ - Cigarette smoking | -0.042 |
| Feeling like an outsider in one's family when growing up - Unable to control worry§ | 0.042 | Give up some happiness now for greater happiness later‡ - Feel close to someone | 0.042 | Worse relationship with mother - Happiness‡ | -0.042 |
| Various aspects of life are balanced§ - Distress from life-threatening experiences§ | -0.042 | Worse relationship with mother - Life satisfaction‡ | -0.042 | Life evaluation five years from now - Government approval§ | 0.042 |
| Forgiveness§ - Give up some happiness now for greater happiness later‡ | 0.042 | Forgiveness§ - Worry about safety, food, or housing§§‡‡ | 0.041 | Ever physically or sexually abused when growing up - Donated to charity | 0.041 |
| Perceived support from others - Worry about meeting normal monthly living expenses§§‡‡ | -0.041 | Feel close to someone - Support reducing differences in income levels§ | 0.041 | Loneliness§ - Various aspects of life are balanced§ | -0.041 |
| Extent of suffering§ - Support reducing differences in income levels§ | -0.041 | Ever physically or sexually abused when growing up - Happiness‡ | -0.041 | Say in government# - Support reducing differences in income levels§ | 0.041 |
| Perceived discrimination§ - Worry about meeting normal monthly living expenses§§‡‡ | 0.041 | Forgiveness§ - Worry about meeting normal monthly living expenses§§‡‡ | 0.041 | Have little interest in doing things§ - Obeying the law§ | -0.040 |
| Obeying the law§ - Trust in others§ | 0.040 | Poorer self-rated childhood health - Understand purpose in life‡ | -0.040 | Worse relationship with mother - Extent of suffering§ | 0.040 |
| Feeling like an outsider in one's family when growing up - Life evaluation today | -0.040 | Loneliness§ - Alcohol use | 0.040 | Act to promote good‡ - Helped a stranger | 0.040 |
| Loneliness§ - Feel close to someone | -0.039 | Freedom - Donated to charity | 0.039 | Not feeling loved by mother - Obeying the law§ | -0.039 |
| Have any health problems - Perceived support from others | -0.039 | Poorer self-rated childhood health - Act to promote good‡ | -0.039 | Ever physically or sexually abused when growing up - Worse relationship with father | 0.039 |
| Poorer self-rated childhood health - Have a long list of gratitude | -0.039 | Positive expectations - Helped a stranger | 0.039 | Various aspects of life are balanced§ - Perceived support from others | 0.039 |
| Poorer self-rated childhood health - Loneliness§ | 0.038 | Worse relationship with mother - Perceived support from others | -0.038 | Not feeling loved by father - Say in government# | -0.038 |
| Depressive symptoms§ - Feeling capable§ | -0.038 | Worse relationship with mother - Feel close to someone | -0.038 | Volunteered at an organization - Perceived discrimination§ | 0.038 |
| Feeling like an outsider in one's family when growing up - Perceived support from others | -0.038 | Poorer self-rated childhood health - Feeling capable§ | -0.038 | Belonging - Obeying the law§ | 0.038 |
| Ever physically or sexually abused when growing up - Helped a stranger | 0.038 | Feeling like an outsider in one's family when growing up - Forgiveness§ | 0.038 | Worse relationship with father - Cigarette smoking | 0.038 |
| Feeling nervous or anxious§ - Obeying the law§ | -0.038 | Not feeling loved by mother - Relationship satisfaction‡ | -0.037 | Physical health‡ - Helped a stranger | 0.037 |
| Remain hopeful for the future - Feel close to someone | 0.037 | Poorer self-rated childhood health - Donated to charity | -0.037 | Helped a stranger - Obeying the law§ | 0.037 |
| Have a long list of gratitude - Worry about safety, food, or housing§§‡‡ | 0.037 | Alcohol use - Unable to control worry§ | 0.037 | Worse relationship with father - Loneliness§ | 0.037 |
| Poorer self-rated childhood health - Belonging | -0.037 | Worse relationship with father - Life evaluation today | -0.037 | Feeling nervous or anxious§ - Feeling capable§ | -0.037 |
| Worse relationship with father - Life satisfaction‡ | -0.037 | Life evaluation five years from now - Have little interest in doing things§ | 0.037 | Feel close to someone - Trust in others§ | 0.037 |
| Perceived discrimination§ - Worry about safety, food, or housing§§‡‡ | 0.036 | Life satisfaction‡ - Forgiveness§ | 0.036 | Belonging - Donated to charity | 0.036 |
| Remain hopeful for the future - Helped a stranger | 0.036 | Have any health problems - Support reducing differences in income levels§ | -0.036 | Donated to charity - Government approval§ | 0.036 |
| Not feeling loved by mother - Support reducing differences in income levels§ | -0.036 | Feeling like an outsider in one's family when growing up - Loneliness§ | 0.036 | Physical health‡ - Say in government# | 0.035 |
| Not feeling loved by father - Mental health‡ | -0.035 | Life evaluation five years from now - Cigarette smoking | -0.035 | Poorer self-rated childhood health - Life evaluation today | -0.035 |
| Positive expectations - Worry about safety, food, or housing§§‡‡ | 0.035 | Life satisfaction‡ - Have little interest in doing things§ | 0.035 | Worse relationship with mother - Government approval§ | -0.034 |
| More difficult childhood family financial situation - Show love often | -0.034 | Not feeling loved by father - Alcohol use | 0.034 | Physical health‡ - Obeying the law§ | 0.034 |
| Worse relationship with father - Life worthwhile‡ | -0.034 | Worse relationship with father - Happiness‡ | -0.034 | Poorer self-rated childhood health - Perceived discrimination§ | 0.034 |
| Understand purpose in life‡ - Donated to charity | 0.034 | Ever physically or sexually abused when growing up - Worse relationship with mother | 0.034 | Unable to control worry§ - Feeling capable§ | -0.034 |

| Variables | rho | Variables | rho | Variables | rho |
|---|---|---|---|---|---|
| Feeling like an outsider in one's family when growing up - Distress from life-threatening experiences§ | 0.034 | Not feeling loved by father - Forgiveness§ | -0.034 | Helped a stranger - Perceived discrimination§ | 0.034 |
| Donated to charity - Worry about safety, food, or housing§§‡‡ | 0.034 | Worse relationship with father - Trust in others§ | -0.034 | Poorer self-rated childhood health - Obeying the law§ | -0.034 |
| Not feeling loved by father - Happiness‡ | -0.034 | Worse relationship with mother - Feeling nervous or anxious§ | 0.034 | Freedom - Worry about safety, food, or housing§§‡‡ | 0.033 |
| Say in government# - Worry about safety, food, or housing§§‡‡ | 0.033 | Ever physically or sexually abused when growing up - Feeling like an outsider in one's family when growing up | 0.033 | More difficult childhood family financial situation - Remain hopeful for the future | -0.033 |
| Ever physically or sexually abused when growing up - Worry about safety, food, or housing§§‡‡ | 0.033 | Worse relationship with father - Feel at peace§ | -0.033 | Cigarette smoking - Distress from life-threatening experiences§ | 0.033 |
| Positive expectations - Have any health problems | -0.033 | Extent of suffering§ - Volunteered at an organization | -0.033 | Not feeling loved by father - Life satisfaction‡ | -0.033 |
| Alcohol use - Show love often | -0.033 | Remain hopeful for the future - Worry about meeting normal monthly living expenses§§‡‡ | 0.033 | Ever physically or sexually abused when growing up - More difficult childhood family financial situation | 0.033 |
| Ever physically or sexually abused when growing up - Not feeling loved by father | 0.033 | More difficult childhood family financial situation - Alcohol use | 0.032 | Belonging - Support reducing differences in income levels§ | 0.032 |
| Unable to control worry§ - Obeying the law§ | -0.032 | Cigarette smoking - Physical health‡ | -0.032 | Life evaluation today - Feeling capable§ | 0.032 |
| Alcohol use - Distress from life-threatening experiences§ | 0.032 | Unable to control worry§ - Have a long list of gratitude | -0.032 | Say in government# - Worry about meeting normal monthly living expenses§§‡‡ | 0.032 |
| Donated to charity - Forgiveness§ | 0.032 | Worse relationship with father - Freedom | -0.032 | Worse relationship with mother - Donated to charity | -0.032 |
| Worse relationship with mother - Unable to control worry§ | 0.032 | More difficult childhood family financial situation - Worry about safety, food, or housing§§‡‡ | 0.032 | Cigarette smoking - Government approval§ | -0.032 |
| Positive expectations - Feeling nervous or anxious§ | -0.032 | Distress from life-threatening experiences§ - Helped a stranger | 0.032 | Poorer self-rated childhood health - Trust in others§ | -0.031 |
| Understand purpose in life‡ - Helped a stranger | 0.031 | Mental health‡ - Helped a stranger | 0.031 | Not feeling loved by father - Depressive symptoms§ | 0.031 |
| Donated to charity - Show love often | 0.031 | Cigarette smoking - Feeling capable§ | -0.031 | Not feeling loved by father - Freedom | -0.030 |
| Depressive symptoms§ - Show love often | -0.030 | More difficult childhood family financial situation - Say in government# | -0.030 | Worse relationship with mother - Life evaluation today | -0.030 |
| Feeling capable§ - Helped a stranger | 0.030 | Belonging - Worry about safety, food, or housing§§‡‡ | 0.030 | Poorer self-rated childhood health - Say in government# | 0.030 |
| Worse relationship with mother - Act to promote good‡ | -0.030 | Loneliness§ - Have little interest in doing things§ | 0.030 | Extent of suffering§ - Feel close to someone | -0.030 |
| Cigarette smoking - Unable to control worry§ | 0.030 | Understand purpose in life‡ - Worry about safety, food, or housing§§‡‡ | 0.029 | Worse relationship with father - Belonging | -0.029 |
| Relationship contentment‡ - Worry about meeting normal monthly living expenses§§‡‡ | -0.029 | Give up some happiness now for greater happiness later‡ - Volunteered at an organization | -0.029 | Relationship contentment‡ - Donated to charity | 0.029 |
| Physical health‡ - Support reducing differences in income levels§ | 0.029 | Depressive symptoms§ - Government approval§ | -0.029 | Not feeling loved by mother - Alcohol use | 0.029 |
| Feeling like an outsider in one's family when growing up - More difficult childhood family financial situation | 0.029 | Ever physically or sexually abused when growing up - Mental health‡ | -0.029 | Worse relationship with father - Government approval§ | -0.028 |
| Alcohol use - Forgiveness§ | -0.028 | Feeling like an outsider in one's family when growing up - Happiness‡ | -0.028 | Feeling capable§ - Feel close to someone | 0.028 |
| Various aspects of life are balanced§ - Government approval§ | 0.028 | Extent of suffering§ - Helped a stranger | 0.028 | Poorer self-rated childhood health - Support reducing differences in income levels§ | -0.028 |
| Act to promote good‡ - Donated to charity | 0.028 | Loneliness§ - Cigarette smoking | 0.028 | Feel at peace§ - Helped a stranger | 0.028 |
| Relationship contentment‡ - Obeying the law§ | 0.027 | Have any health problems - Feeling capable§ | -0.027 | Cigarette smoking - Show love often | -0.027 |
| Not feeling loved by father - Belonging | -0.027 | Worse relationship with father - Have little interest in doing things§ | 0.027 | Poorer self-rated childhood health - Give up some happiness now for greater happiness later‡ | -0.027 |
| Cigarette smoking - Forgiveness§ | -0.027 | Obeying the law§ - Perceived support from others | 0.027 | Relationship satisfaction‡ - Donated to charity | 0.026 |
| Worse relationship with mother - Have any health problems | 0.026 | Forgiveness§ - Perceived support from others | 0.026 | Feeling like an outsider in one's family when growing up - Worry about safety, food, or housing§§‡‡ | 0.026 |
| Feel at peace§ - Support reducing differences in income levels§ | 0.026 | Give up some happiness now for greater happiness later‡ - Worry about safety, food, or housing§§‡‡ | 0.026 | Not feeling loved by mother - Life evaluation today | -0.026 |
| Worse relationship with mother - Understand purpose in life‡ | -0.026 | Feeling capable§ - Worry about safety, food, or housing§§‡‡ | 0.026 | Have little interest in doing things§ - Forgiveness§ | 0.026 |
| Distress from life-threatening experiences§ - Feel close to someone | -0.026 | Worse relationship with father - Feeling capable§ | -0.026 | Have any health problems - Obeying the law§ | -0.026 |
| Helped a stranger - Show love often | 0.025 | Ever physically or sexually abused when growing up - Worry about meeting normal monthly living expenses§§‡‡ | 0.025 | Not feeling loved by father - Loneliness§ | 0.025 |

| Variables | rho | Variables | rho | Variables | rho |
|---|---|---|---|---|---|
| Have little interest in doing things§ - Government approval§ | 0.025 | Alcohol use - Obeying the law§ | -0.025 | Happiness‡ - Volunteered at an organization | -0.025 |
| Feeling like an outsider in one's family when growing up - Positive expectations | -0.025 | Feeling capable§ - Perceived discrimination§ | -0.025 | Have any health problems - Show love often | -0.025 |
| Not feeling loved by mother - Extent of suffering§ | 0.025 | Worse relationship with mother - Have a long list of gratitude | -0.025 | Have little interest in doing things§ - Donated to charity | -0.025 |
| Not feeling loved by mother - Loneliness§ | 0.025 | Loneliness§ - Volunteered at an organization | 0.024 | Worse relationship with father - Feel close to someone | -0.024 |
| Not feeling loved by father - Life evaluation today | -0.024 | More difficult childhood family financial situation - Have little interest in doing things§ | 0.024 | Worse relationship with mother - Feel at peace§ | -0.024 |
| Perceived support from others - Support reducing differences in income levels§ | 0.024 | Ever physically or sexually abused when growing up - Belonging | -0.024 | Life worthwhile‡ - Forgiveness§ | 0.024 |
| Feeling like an outsider in one's family when growing up - Give up some happiness now for greater happiness later‡ | -0.024 | Worse relationship with father - Extent of suffering§ | 0.024 | Feeling like an outsider in one's family when growing up - Government approval§ | 0.024 |
| Happiness‡ - Obeying the law§ | 0.024 | Not feeling loved by father - Feeling nervous or anxious§ | 0.024 | Life satisfaction‡ - Obeying the law§ | 0.024 |
| Worse relationship with mother - Distress from life-threatening experiences§ | 0.024 | Worse relationship with father - Donated to charity | -0.024 | Mental health‡ - Donated to charity | 0.024 |
| Have any health problems - Feel close to someone | -0.024 | Loneliness§ - Forgiveness§ | -0.023 | Belonging - Volunteered at an organization | 0.023 |
| Support reducing differences in income levels§ - Trust in others§ | 0.023 | Donated to charity - Trust in others§ | 0.023 | Unable to control worry§ - Forgiveness§ | 0.023 |
| Extent of suffering§ - Say in government# | 0.023 | Various aspects of life are balanced§ - Worry about meeting normal monthly living expenses§§‡‡ | 0.023 | Positive expectations - Alcohol use | -0.023 |
| Distress from life-threatening experiences§ - Say in government# | -0.023 | Extent of suffering§ - Donated to charity | -0.023 | More difficult childhood family financial situation - Have a long list of gratitude | -0.023 |
| Donated to charity - Have a long list of gratitude | 0.023 | Feeling like an outsider in one's family when growing up - Poorer self-rated childhood health | -0.023 | Extent of suffering§ - Show love often | -0.023 |
| Life evaluation today - Have little interest in doing things§ | -0.023 | Life evaluation five years from now - Forgiveness§ | 0.022 | Belonging - Cigarette smoking | -0.022 |
| Government approval§ - Perceived discrimination§ | -0.022 | Alcohol use - Depressive symptoms§ | 0.022 | Relationship contentment‡ - Worry about safety, food, or housing§§‡‡ | 0.022 |
| Not feeling loved by mother - Life evaluation five years from now | -0.022 | Life evaluation today - Helped a stranger | 0.022 | Not feeling loved by mother - Say in government# | -0.022 |
| Feeling like an outsider in one's family when growing up - Cigarette smoking | 0.022 | Feeling like an outsider in one's family when growing up - Have any health problems | 0.022 | Not feeling loved by mother - Cigarette smoking | 0.022 |
| Cigarette smoking - Depressive symptoms§ | 0.022 | Not feeling loved by father - Support reducing differences in income levels§ | -0.022 | Cigarette smoking - Worry about safety, food, or housing§§‡‡ | -0.022 |
| Worse relationship with mother - Depressive symptoms§ | 0.022 | Positive expectations - Volunteered at an organization | -0.022 | Belonging - Alcohol use | -0.022 |
| Give up some happiness now for greater happiness later‡ - Helped a stranger | 0.022 | Worse relationship with mother - Positive expectations | -0.022 | Remain hopeful for the future - Donated to charity | 0.022 |
| Ever physically or sexually abused when growing up - Life satisfaction‡ | -0.022 | Relationship contentment‡ - Support reducing differences in income levels§ | 0.022 | Worse relationship with father - Forgiveness§ | -0.022 |
| Various aspects of life are balanced§ - Say in government# | 0.021 | Worse relationship with father - Understand purpose in life‡ | -0.021 | Have little interest in doing things§ - Helped a stranger | 0.021 |
| Alcohol use - Support reducing differences in income levels§ | -0.021 | Worse relationship with mother - Show love often | -0.021 | Life worthwhile‡ - Helped a stranger | 0.021 |
| Life satisfaction‡ - Support reducing differences in income levels§ | 0.021 | Positive expectations - Obeying the law§ | -0.021 | Not feeling loved by mother - Relationship contentment‡ | -0.021 |
| Not feeling loved by father - Life evaluation five years from now | -0.021 | Not feeling loved by mother - Various aspects of life are balanced§ | 0.021 | Feeling like an outsider in one's family when growing up - Say in government# | 0.021 |
| Ever physically or sexually abused when growing up - Positive expectations | -0.021 | Ever physically or sexually abused when growing up - Various aspects of life are balanced§ | 0.021 | Life worthwhile‡ - Obeying the law§ | 0.021 |
| Ever physically or sexually abused when growing up - Have a long list of gratitude | 0.021 | Worse relationship with mother - Remain hopeful for the future | -0.021 | Feeling capable§ - Support reducing differences in income levels§ | 0.021 |
| Not feeling loved by mother - Mental health‡ | -0.020 | Happiness‡ - Support reducing differences in income levels§ | 0.020 | Distress from life-threatening experiences§ - Volunteered at an organization | 0.020 |
| Life worthwhile‡ - Donated to charity | 0.020 | Life worthwhile‡ - Support reducing differences in income levels§ | 0.020 | Not feeling loved by father - Act to promote good‡ | -0.020 |
| Government approval§ - Worry about meeting normal monthly living expenses§§‡‡ | -0.020 | Donated to charity - Worry about meeting normal monthly living expenses§§‡‡ | 0.020 | Life evaluation today - Perceived discrimination§ | -0.020 |

| Variables | rho | Variables | rho | Variables | rho |
|---|---|---|---|---|---|
| Life evaluation today - Forgiveness§ | -0.020 | Feeling nervous or anxious§ - Have a long list of gratitude | -0.020 | Feeling like an outsider in one's family when growing up - Physical health‡ | -0.020 |
| Distress from life-threatening experiences§ - Support reducing differences in income levels§ | -0.020 | Physical health‡ - Donated to charity | 0.020 | More difficult childhood family financial situation - Helped a stranger | -0.020 |
| Various aspects of life are balanced§ - Support reducing differences in income levels§ | 0.020 | Cigarette smoking - Perceived discrimination§ | 0.020 | Not feeling loved by father - Unable to control worry§ | 0.020 |
| Feeling like an outsider in one's family when growing up - Trust in others§ | -0.020 | Feeling like an outsider in one's family when growing up - Mental health‡ | -0.019 | Worse relationship with father - Show love often | -0.019 |
| Not feeling loved by father - Distress from life-threatening experiences§ | 0.019 | Poorer self-rated childhood health - Relationship contentment‡ | -0.019 | Physical health‡ - Forgiveness§ | 0.019 |
| Feeling like an outsider in one's family when growing up - Support reducing differences in income levels§ | 0.019 | Feeling like an outsider in one's family when growing up - Feel at peace§ | -0.019 | Distress from life-threatening experiences§ - Government approval§ | -0.019 |
| Relationship satisfaction‡ - Worry about meeting normal monthly living expenses§§‡‡ | 0.019 | Unable to control worry§ - Show love often | -0.019 | Cigarette smoking - Helped a stranger | 0.019 |
| Freedom - Volunteered at an organization | 0.019 | Cigarette smoking - Have any health problems | 0.019 | Remain hopeful for the future - Alcohol use | -0.019 |
| Various aspects of life are balanced§ - Perceived discrimination§ | 0.019 | Forgiveness§ - Volunteered at an organization | 0.019 | Feeling like an outsider in one's family when growing up - Alcohol use | 0.019 |
| Happiness‡ - Helped a stranger | 0.018 | Worse relationship with father - Alcohol use | 0.018 | Cigarette smoking - Give up some happiness now for greater happiness later‡ | -0.018 |
| Positive expectations - Extent of suffering§ | -0.018 | Remain hopeful for the future - Support reducing differences in income levels§ | -0.018 | Not feeling loved by father - Obeying the law§ | -0.018 |
| Alcohol use - Have little interest in doing things§ | 0.018 | Feeling like an outsider in one's family when growing up - Depressive symptoms§ | 0.018 | Worse relationship with father - Life evaluation five years from now | -0.018 |
| Worse relationship with mother - Trust in others§ | -0.018 | Various aspects of life are balanced§ - Volunteered at an organization | -0.018 | Life evaluation five years from now - Donated to charity | 0.018 |
| Volunteered at an organization - Trust in others§ | 0.018 | Feeling nervous or anxious§ - Support reducing differences in income levels§ | -0.018 | Cigarette smoking - Worry about meeting normal monthly living expenses§§‡‡ | -0.018 |
| Understand purpose in life‡ - Volunteered at an organization | 0.018 | Alcohol use - Government approval§ | -0.018 | Helped a stranger - Perceived support from others | 0.018 |
| Worse relationship with father - Feeling nervous or anxious§ | 0.018 | Depressive symptoms§ - Trust in others§ | -0.018 | Ever physically or sexually abused when growing up - Understand purpose in life‡ | 0.018 |
| Alcohol use - Say in government# | -0.018 | Perceived support from others - Worry about safety, food, or housing§§‡‡ | -0.018 | More difficult childhood family financial situation - Trust in others§ | -0.018 |
| Not feeling loved by father - Extent of suffering§ | 0.017 | Distress from life-threatening experiences§ - Obeying the law§ | -0.017 | Life evaluation five years from now - Worry about safety, food, or housing§§‡‡ | -0.017 |
| Alcohol use - Physical health‡ | -0.017 | Feel at peace§ - Cigarette smoking | -0.017 | Various aspects of life are balanced§ - Worry about safety, food, or housing§§‡‡ | 0.017 |
| Cigarette smoking - Feel close to someone | -0.017 | Not feeling loved by father - Poorer self-rated childhood health | 0.017 | Relationship satisfaction‡ - Obeying the law§ | 0.017 |
| Worse relationship with father - Have any health problems | 0.017 | Not feeling loved by mother - Poorer self-rated childhood health | 0.017 | Understand purpose in life‡ - Cigarette smoking | -0.017 |
| Feeling like an outsider in one's family when growing up - Understand purpose in life‡ | 0.017 | Not feeling loved by mother - Freedom | -0.017 | Feel at peace§ - Perceived discrimination§ | -0.017 |
| Relationship satisfaction‡ - Support reducing differences in income levels§ | 0.017 | Unable to control worry§ - Helped a stranger | 0.017 | Cigarette smoking - Have little interest in doing things§ | 0.017 |
| Poorer self-rated childhood health - Perceived support from others | -0.016 | Alcohol use - Have a long list of gratitude | -0.016 | Alcohol use - Feeling nervous or anxious§ | 0.016 |
| Government approval§ - Worry about safety, food, or housing§§‡‡ | -0.016 | Ever physically or sexually abused when growing up - Poorer self-rated childhood health | 0.016 | Various aspects of life are balanced§ - Extent of suffering§ | 0.016 |
| Worse relationship with father - Distress from life-threatening experiences§ | 0.016 | Happiness‡ - Forgiveness§ | 0.016 | Happiness‡ - Have little interest in doing things§ | 0.016 |
| Alcohol use - Donated to charity | -0.016 | Have any health problems - Say in government# | 0.016 | Depressive symptoms§ - Give up some happiness now for greater happiness later‡ | -0.016 |
| Worse relationship with mother - Feeling capable§ | -0.016 | Various aspects of life are balanced§ - Have any health problems | -0.016 | Unable to control worry§ - Support reducing differences in income levels§ | -0.016 |

| Variables | rho | Variables | rho | Variables | rho |
|---|---|---|---|---|---|
| Extent of suffering§ - Forgiveness§ | -0.015 | Forgiveness§ - Feel close to someone | 0.015 | Feeling nervous or anxious§ - Forgiveness§ | 0.015 |
| Cigarette smoking - Trust in others§ | -0.015 | Not feeling loved by mother - Life worthwhile‡ | -0.015 | Remain hopeful for the future - Cigarette smoking | -0.015 |
| Worse relationship with mother - Say in government# | -0.015 | Helped a stranger - Say in government# | 0.015 | Positive expectations - Cigarette smoking | -0.015 |
| Have any health problems - Forgiveness§ | -0.015 | Cigarette smoking - Donated to charity | 0.015 | Perceived discrimination§ - Perceived support from others | -0.015 |
| Have any health problems - Give up some happiness now for greater happiness later‡ | -0.015 | Worse relationship with father - Unable to control worry§ | 0.015 | Not feeling loved by mother - Depressive symptoms§ | 0.015 |
| Have little interest in doing things§ - Mental health‡ | 0.015 | Not feeling loved by mother - Physical health‡ | -0.015 | More difficult childhood family financial situation - Positive expectations | -0.015 |
| Feel close to someone - Government approval§ | 0.015 | Not feeling loved by father - Trust in others§ | -0.015 | Give up some happiness now for greater happiness later‡ - Worry about meeting normal monthly living expenses§§‡‡ | -0.014 |
| Life evaluation today - Obeying the law§ | -0.014 | Have little interest in doing things§ - Volunteered at an organization | -0.014 | Feeling like an outsider in one's family when growing up - Life satisfaction‡ | -0.014 |
| Not feeling loved by father - Physical health‡ | -0.014 | Unable to control worry§ - Trust in others§ | 0.014 | Forgiveness§ - Perceived discrimination§ | 0.014 |
| Not feeling loved by father - Helped a stranger | 0.014 | Unable to control worry§ - Give up some happiness now for greater happiness later‡ | -0.014 | Not feeling loved by father - Worry about meeting normal monthly living expenses§§‡‡ | 0.014 |
| More difficult childhood family financial situation - Feel at peace§ | -0.014 | Not feeling loved by father - Positive expectations | -0.014 | Poorer self-rated childhood health - Volunteered at an organization | -0.013 |
| Feeling like an outsider in one's family when growing up - Show love often | 0.013 | Not feeling loved by father - Give up some happiness now for greater happiness later‡ | -0.013 | Ever physically or sexually abused when growing up - Give up some happiness now for greater happiness later‡ | 0.013 |
| Feel at peace§ - Have any health problems | -0.013 | Depressive symptoms§ - Volunteered at an organization | 0.013 | Cigarette smoking - Say in government# | -0.013 |
| Not feeling loved by mother - Government approval§ | -0.013 | Not feeling loved by mother - Remain hopeful for the future | 0.013 | Ever physically or sexually abused when growing up - Forgiveness§ | -0.013 |
| Remain hopeful for the future - Volunteered at an organization | -0.013 | More difficult childhood family financial situation - Feeling capable§ | -0.013 | Feeling like an outsider in one's family when growing up - Act to promote good‡ | 0.013 |
| Feeling capable§ - Obeying the law§ | 0.013 | Remain hopeful for the future - Obeying the law§ | -0.013 | Poorer self-rated childhood health - Positive expectations | -0.013 |
| Feeling like an outsider in one's family when growing up - Obeying the law§ | 0.013 | Feeling like an outsider in one's family when growing up - Life worthwhile‡ | -0.013 | Various aspects of life are balanced§ - Depressive symptoms§ | -0.013 |
| Feel at peace§ - Donated to charity | 0.012 | Give up some happiness now for greater happiness later‡ - Obeying the law§ | -0.012 | Not feeling loved by father - Have any health problems | 0.012 |
| Worse relationship with father - Have a long list of gratitude | -0.012 | Not feeling loved by father - Feel close to someone | -0.012 | Life evaluation today - Donated to charity | 0.012 |
| Not feeling loved by father - Feel at peace§ | -0.012 | Relationship satisfaction‡ - Helped a stranger | 0.012 | Poorer self-rated childhood health - Show love often | -0.012 |
| Depressive symptoms§ - Forgiveness§ | 0.012 | Depressive symptoms§ - Helped a stranger | 0.012 | Feeling like an outsider in one's family when growing up - Belonging | 0.012 |
| Feel at peace§ - Obeying the law§ | 0.012 | Donated to charity - Give up some happiness now for greater happiness later‡ | -0.012 | Feel close to someone - Obeying the law§ | 0.012 |
| Not feeling loved by mother - Donated to charity | -0.012 | Ever physically or sexually abused when growing up - Remain hopeful for the future | -0.012 | Feeling like an outsider in one's family when growing up - Extent of suffering§ | 0.012 |
| Have any health problems - Trust in others§ | 0.011 | Loneliness§ - Support reducing differences in income levels§ | -0.011 | Ever physically or sexually abused when growing up - Feeling capable§ | -0.011 |
| Ever physically or sexually abused when growing up - Obeying the law§ | -0.011 | Life evaluation five years from now - Alcohol use | -0.011 | Alcohol use - Have any health problems | 0.011 |
| Have any health problems - Government approval§ | 0.011 | Not feeling loved by father - Perceived discrimination§ | 0.011 | Life worthwhile‡ - Volunteered at an organization | -0.011 |
| Ever physically or sexually abused when growing up - Physical health‡ | -0.011 | Not feeling loved by father - Show love often | -0.011 | Feel at peace§ - Unable to control worry§ | -0.011 |
| Cigarette smoking - Volunteered at an organization | 0.011 | Give up some happiness now for greater happiness later‡ - Perceived discrimination§ | -0.011 | Ever physically or sexually abused when growing up - Trust in others§ | 0.011 |
| Cigarette smoking - Have a long list of gratitude | -0.011 | Various aspects of life are balanced§ - Cigarette smoking | 0.011 | Distress from life-threatening experiences§ - Donated to charity | 0.011 |
| Give up some happiness now for greater happiness later‡ - Support reducing differences in income levels§ | -0.011 | Ever physically or sexually abused when growing up - Life worthwhile‡ | -0.011 | Ever physically or sexually abused when growing up - Life evaluation today | 0.011 |
| Not feeling loved by father - Feeling capable§ | -0.010 | Not feeling loved by mother - Feeling nervous or anxious§ | 0.010 | Alcohol use - Extent of suffering§ | 0.010 |

| Variables | rho | Variables | rho | Variables | rho |
|---|---|---|---|---|---|
| Life evaluation today - Support reducing differences in income levels[§] | 0.010 | Alcohol use - Worry about meeting normal monthly living expenses[§§‡‡] | -0.010 | Worse relationship with father - Remain hopeful for the future | 0.010 |
| Depressive symptoms[§] - Donated to charity | -0.010 | Freedom - Support reducing differences in income levels[§] | 0.010 | Ever physically or sexually abused when growing up - Not feeling loved by mother | 0.010 |
| Feeling like an outsider in one's family when growing up - Life evaluation five years from now | -0.010 | Feel close to someone - Worry about meeting normal monthly living expenses[§§‡‡] | -0.010 | Worse relationship with father - Depressive symptoms[§] | 0.010 |
| Alcohol use - Give up some happiness now for greater happiness later[‡] | -0.010 | Not feeling loved by father - Understand purpose in life[‡] | -0.010 | Worse relationship with father - Helped a stranger | -0.010 |
| Ever physically or sexually abused when growing up - Life evaluation five years from now | -0.010 | Not feeling loved by mother - Feel close to someone | -0.010 | Perceived discrimination[§] - Trust in others[§] | -0.010 |
| Have any health problems - Have a long list of gratitude | -0.010 | Not feeling loved by mother - Belonging | -0.010 | Support reducing differences in income levels[§] - Worry about safety, food, or housing[§§‡‡] | 0.009 |
| Not feeling loved by mother - Worry about meeting normal monthly living expenses[§§‡‡] | 0.009 | Cigarette smoking - Obeying the law[§] | 0.009 | Support reducing differences in income levels[§] - Worry about meeting normal monthly living expenses[§§‡‡] | 0.009 |
| Freedom - Cigarette smoking | -0.009 | Depressive symptoms[§] - Feel close to someone | -0.009 | Not feeling loved by father - Life worthwhile[‡] | -0.009 |
| Worse relationship with mother - Cigarette smoking | 0.009 | Not feeling loved by mother - Worry about safety, food, or housing[§§‡‡] | 0.009 | Helped a stranger - Trust in others[§] | 0.009 |
| Positive expectations - Worry about meeting normal monthly living expenses[§§‡‡] | 0.009 | Feel at peace[§] - Worry about safety, food, or housing[§§‡‡] | 0.009 | Belonging - Worry about meeting normal monthly living expenses[§§‡‡] | -0.009 |
| Act to promote good[‡] - Worry about meeting normal monthly living expenses[§§‡‡] | 0.009 | Life evaluation five years from now - Volunteered at an organization | -0.009 | Feeling nervous or anxious[§] - Helped a stranger | 0.009 |
| Have any health problems - Donated to charity | 0.009 | Various aspects of life are balanced[§] - Helped a stranger | 0.009 | Worse relationship with mother - Worry about safety, food, or housing[§§‡‡] | -0.009 |
| Cigarette smoking - Extent of suffering[§] | 0.009 | Not feeling loved by mother - Distress from life-threatening experiences[§] | 0.008 | Various aspects of life are balanced[§] - Alcohol use | 0.008 |
| Depressive symptoms[§] - Obeying the law[§] | -0.008 | Not feeling loved by mother - Have little interest in doing things[§] | 0.008 | Worse relationship with father - Perceived discrimination[§] | 0.008 |
| Life evaluation five years from now - Obeying the law[§] | 0.008 | Ever physically or sexually abused when growing up - Feel at peace[§] | -0.008 | Life evaluation today - Volunteered at an organization | -0.008 |
| Extent of suffering[§] - Have a long list of gratitude | -0.008 | Freedom - Alcohol use | -0.008 | Not feeling loved by mother - Have a long list of gratitude | 0.008 |
| Understand purpose in life[‡] - Obeying the law[§] | -0.008 | Worse relationship with father - Say in government# | -0.008 | Unable to control worry[§] - Donated to charity | 0.008 |
| Have a long list of gratitude - Volunteered at an organization | 0.008 | Not feeling loved by father - Donated to charity | -0.008 | Feel at peace[§] - Feel close to someone | 0.008 |
| Feel at peace[§] - Alcohol use | -0.008 | Poorer self-rated childhood health - Alcohol use | 0.008 | Worse relationship with mother - Perceived discrimination[§] | 0.007 |
| Ever physically or sexually abused when growing up - Feel close to someone | 0.007 | Not feeling loved by father - Volunteered at an organization | -0.007 | Perceived discrimination[§] - Say in government# | 0.007 |
| Cigarette smoking - Act to promote good[‡] | -0.007 | Feeling like an outsider in one's family when growing up - Various aspects of life are balanced[§] | -0.007 | More difficult childhood family financial situation - Various aspects of life are balanced[§] | 0.007 |
| Worse relationship with mother - Alcohol use | 0.007 | Not feeling loved by father - Cigarette smoking | -0.007 | Show love often - Obeying the law[§] | 0.007 |
| Unable to control worry[§] - Say in government# | -0.007 | Worse relationship with mother - Forgiveness[§] | 0.007 | Not feeling loved by mother - Feeling capable[§] | 0.007 |
| Not feeling loved by mother - Helped a stranger | -0.007 | Helped a stranger - Worry about safety, food, or housing[§§‡‡] | 0.007 | Perceived discrimination[§] - Support reducing differences in income levels[§] | 0.007 |
| Obeying the law[§] - Say in government# | 0.007 | Feeling nervous or anxious[§] - Say in government# | 0.007 | Alcohol use - Perceived discrimination[§] | 0.007 |
| Worse relationship with mother - Helped a stranger | -0.007 | Life satisfaction[‡] - Donated to charity | 0.007 | Alcohol use - Trust in others[§] | -0.007 |
| Worse relationship with father - Worry about meeting normal monthly living expenses[§§‡‡] | 0.006 | Loneliness[§] - Donated to charity | 0.006 | Depressive symptoms[§] - Support reducing differences in income levels[§] | -0.006 |
| Feeling like an outsider in one's family when growing up - Remain hopeful for the future | -0.006 | Relationship contentment[‡] - Helped a stranger | 0.006 | Feeling like an outsider in one's family when growing up - Have little interest in doing things[§] | 0.006 |
| Feeling nervous or anxious[§] - Donated to charity | 0.006 | Unable to control worry[§] - Volunteered at an organization | 0.006 | Freedom - Obeying the law[§] | 0.006 |
| Life evaluation five years from now - Support reducing differences in income levels[§] | -0.006 | Feel at peace[§] - Worry about meeting normal monthly living expenses[§§‡‡] | -0.006 | Loneliness[§] - Obeying the law[§] | -0.006 |
| Distress from life-threatening experiences[§] - Forgiveness[§] | 0.006 | Cigarette smoking - Feeling nervous or anxious[§] | 0.006 | Obeying the law[§] - Worry about safety, food, or housing[§§‡‡] | -0.006 |

| Variables | rho | Variables | rho | Variables | rho |
|---|---|---|---|---|---|
| More difficult childhood family financial situation - Perceived discrimination§ | 0.006 | Worse relationship with father - Act to promote good‡ | -0.006 | Act to promote good‡ - Obeying the law§ | 0.006 |
| Worse relationship with father - Give up some happiness now for greater happiness later‡ | 0.006 | Helped a stranger - Government approval§ | -0.006 | Not feeling loved by father - Various aspects of life are balanced§ | -0.006 |
| Relationship contentment‡ - Volunteered at an organization | 0.006 | Have any health problems - Volunteered at an organization | 0.005 | Feeling nervous or anxious§ - Government approval§ | -0.005 |
| Not feeling loved by mother - Give up some happiness now for greater happiness later‡ | 0.005 | Not feeling loved by father - Have little interest in doing things§ | 0.005 | Feeling like an outsider in one's family when growing up - Have a long list of gratitude | 0.005 |
| Ever physically or sexually abused when growing up - Relationship contentment‡ | -0.005 | Feel close to someone - Worry about safety, food, or housing§§‡‡ | 0.005 | Obeying the law§ - Worry about meeting normal monthly living expenses§§‡‡ | -0.005 |
| Not feeling loved by mother - Unable to control worry§ | 0.005 | Alcohol use - Feel close to someone | -0.005 | Depressive symptoms§ - Say in government# | 0.005 |
| Poorer self-rated childhood health - Government approval§ | 0.005 | Extent of suffering§ - Trust in others§ | 0.005 | Poorer self-rated childhood health - Relationship satisfaction‡ | -0.005 |
| Alcohol use - Act to promote good‡ | 0.005 | Feel at peace§ - Extent of suffering§ | -0.005 | Positive expectations - Donated to charity | -0.005 |
| Cigarette smoking - Support reducing differences in income levels§ | -0.005 | Not feeling loved by mother - Show love often | -0.005 | Worse relationship with mother - Various aspects of life are balanced§ | 0.005 |
| Forgiveness§ - Helped a stranger | 0.005 | Mental health‡ - Volunteered at an organization | -0.005 | Have a long list of gratitude - Support reducing differences in income levels§ | -0.005 |
| Show love often - Support reducing differences in income levels§ | 0.005 | Worse relationship with mother - Worry about meeting normal monthly living expenses§§‡‡ | -0.005 | Feel at peace§ - Volunteered at an organization | 0.005 |
| Not feeling loved by mother - Positive expectations | 0.004 | Not feeling loved by mother - Forgiveness§ | -0.004 | Have a long list of gratitude - Obeying the law§ | -0.004 |
| Various aspects of life are balanced§ - Feel close to someone | 0.004 | Feeling nervous or anxious§ - Feel close to someone | -0.004 | Alcohol use - Volunteered at an organization | 0.004 |
| Show love often - Volunteered at an organization | -0.004 | Helped a stranger - Worry about meeting normal monthly living expenses§§‡‡ | -0.004 | Loneliness§ - Helped a stranger | 0.004 |
| Volunteered at an organization - Perceived support from others | -0.004 | Feel close to someone - Say in government# | 0.004 | Not feeling loved by mother - Understand purpose in life‡ | 0.004 |
| Feeling nervous or anxious§ - Show love often | 0.004 | Not feeling loved by mother - Act to promote good‡ | -0.004 | Trust in others§ - Worry about safety, food, or housing§§‡‡ | -0.004 |
| Ever physically or sexually abused when growing up - Freedom | 0.004 | Worse relationship with father - Positive expectations | -0.004 | Volunteered at an organization - Worry about safety, food, or housing§§‡‡ | 0.004 |
| Not feeling loved by mother - Happiness‡ | -0.004 | Volunteered at an organization - Worry about meeting normal monthly living expenses§§‡‡ | 0.004 | Ever physically or sexually abused when growing up - Act to promote good‡ | 0.004 |
| Feeling nervous or anxious§ - Give up some happiness now for greater happiness later‡ | -0.003 | Have any health problems - Helped a stranger | 0.003 | Various aspects of life are balanced§ - Obeying the law§ | -0.003 |
| Trust in others§ - Worry about meeting normal monthly living expenses§§‡‡ | -0.003 | Understand purpose in life‡ - Support reducing differences in income levels§ | -0.003 | Alcohol use - Helped a stranger | 0.003 |
| Have little interest in doing things§ - Physical health‡ | 0.003 | Ever physically or sexually abused when growing up - Perceived support from others | -0.003 | Unable to control worry§ - Government approval§ | -0.003 |
| Poorer self-rated childhood health - Cigarette smoking | -0.003 | Various aspects of life are balanced§ - Donated to charity | 0.003 | Life satisfaction‡ - Helped a stranger | 0.003 |
| Not feeling loved by mother - Volunteered at an organization | -0.003 | Physical health‡ - Volunteered at an organization | 0.003 | Donated to charity - Feeling capable§ | 0.003 |
| Alcohol use - Feeling capable§ | -0.003 | Obeying the law§ - Perceived discrimination§ | -0.003 | Not feeling loved by mother - Have any health problems | -0.003 |
| Feeling like an outsider in one's family when growing up - Relationship satisfaction‡ | -0.003 | Understand purpose in life‡ - Worry about meeting normal monthly living expenses§§‡‡ | -0.003 | More difficult childhood family financial situation - Forgiveness§ | -0.003 |
| Feeling nervous or anxious§ - Volunteered at an organization | -0.003 | Understand purpose in life‡ - Alcohol use | -0.003 | Feeling like an outsider in one's family when growing up - Relationship contentment‡ | 0.003 |
| Worse relationship with mother - Give up some happiness now for greater happiness later‡ | 0.002 | Extent of suffering§ - Give up some happiness now for greater happiness later‡ | -0.002 | Worse relationship with father - Various aspects of life are balanced§ | 0.002 |
| Act to promote good‡ - Volunteered at an organization | -0.002 | Extent of suffering§ - Feeling capable§ | 0.002 | Poorer self-rated childhood health - Forgiveness§ | 0.002 |
| Feeling capable§ - Worry about meeting normal monthly living expenses§§‡‡ | -0.002 | Feeling capable§ - Volunteered at an organization | 0.002 | Have a long list of gratitude - Worry about meeting normal monthly living expenses§§‡‡ | -0.002 |
| Not feeling loved by father - Have a long list of gratitude | -0.002 | Feel close to someone - Perceived discrimination§ | 0.002 | Not feeling loved by mother - Trust in others§ | -0.002 |
| Worse relationship with mother - Have little interest in doing things§ | -0.002 | Relationship satisfaction‡ - Volunteered at an organization | -0.002 | Not feeling loved by father - Remain hopeful for the future | 0.002 |

| Variables | rho | Variables | rho | Variables | rho |
|---|---|---|---|---|---|
| Life satisfaction‡ - Volunteered at an organization | -0.002 | Feeling like an outsider in one's family when growing up - Feeling capable§ | -0.002 | Not feeling loved by mother - Feel at peace§ | 0.002 |
| Donated to charity - Perceived support from others | 0.002 | Unable to control worry§ - Feel close to someone | -0.002 | Donated to charity - Perceived discrimination§ | 0.001 |
| Feeling like an outsider in one's family when growing up - Freedom | 0.001 | Have little interest in doing things§ - Perceived support from others | 0.001 | Ever physically or sexually abused when growing up - Show love often | 0.001 |
| Ever physically or sexually abused when growing up - Relationship satisfaction‡ | 0.001 | Feeling nervous or anxious§ - Trust in others§ | -0.001 | Freedom - Worry about meeting normal monthly living expenses§§‡‡ | 0.001 |
| Ever physically or sexually abused when growing up - Support reducing differences in income levels§ | -0.001 | Various aspects of life are balanced§ - Feeling nervous or anxious§ | -0.001 | Alcohol use - Worry about safety, food, or housing§§‡‡ | -0.001 |
| Not feeling loved by mother - Perceived discrimination§ | 0.001 | Happiness‡ - Donated to charity | 0.001 | Act to promote good‡ - Support reducing differences in income levels§ | -0.001 |
| Positive expectations - Support reducing differences in income levels§ | 0.001 | Feeling like an outsider in one's family when growing up - Worry about meeting normal monthly living expenses§§‡‡ | -0.000 | Not feeling loved by father - Worry about safety, food, or housing§§‡‡ | 0.000 |
| Feeling like an outsider in one's family when growing up - Perceived discrimination§ | -0.000 | Not feeling loved by mother - Life satisfaction‡ | -0.000 | Worse relationship with father - Worry about safety, food, or housing§§‡‡ | -0.000 |
| Have little interest in doing things§ - Feel close to someone | 0.000 | | | | |

**eTable 7.** Table of coefficients for adjusted associations between childhood abuse as predictor and 45 outcomes of interest and two flourishing composite indices.

Adjusted survey-weighted models use ANNUAL_WEIGHT_C1 with primary sampling unit (PSU) and strata and adjust for age, gender, education, and residential setting. Reference categories are shown in the table. Coefficients are on the original regression-coefficient scale: beta for continuous outcomes, log-odds for binary outcomes, and proportional-odds log coefficients for ordinal outcomes. Model intercepts are displayed for continuous and binary outcomes. Proportional-odds models do not have a single conventional intercept; their estimated ordinal cut-points/thresholds are displayed after the covariate coefficients, with standard errors (SEs), 95% CIs, and Wald p-values.

***Note:*** All results except p-values are rounded to the two decimals. P-values are rounded to three decimals. Values displayed as 0.000 indicate *P* < 0.0005 due to rounding, rather than an exact zero. Statistical significance was determined using unrounded P-values at the Bonferroni-adjusted threshold of *P* < 0.0011.

| Outcome | Model type | Analytic N | Variable | Level / comparison | Reference category | Coefficient | Standard error | 95% CI | p-value |
|---|---|---|---|---|---|---|---|---|---|
| **Belonging** | **Linear regression** | 11994 | *Intercept* | | | 8.82 | 0.12 | 8.60 to 9.05 | 0.000 |
| | | 11994 | Childhood predictor | Yes | No | -0.41 | 0.11 | -0.62 to -0.20 | 0.000 |
| | | 11994 | Age | Per 1-year increase | | -0.01 | 0.00 | -0.02 to -0.01 | 0.000 |
| | | 11994 | Gender | Female | Male | -0.23 | 0.06 | -0.35 to -0.10 | 0.000 |
| | | 11994 | Education | Secondary | Elementary or less | 0.19 | 0.08 | 0.04 to 0.34 | 0.012 |
| | | 11994 | Education | Four years beyond HS | Elementary or less | 0.30 | 0.10 | 0.10 to 0.50 | 0.003 |
| | | 11994 | Residential setting | Small town | Rural | 0.32 | 0.08 | 0.17 to 0.48 | 0.000 |
| | | 11994 | Residential setting | Large city | Rural | 0.36 | 0.11 | 0.15 to 0.57 | 0.001 |
| | | 11994 | Residential setting | Suburb | Rural | 0.01 | 0.18 | -0.34 to 0.36 | 0.946 |
| **Relationship contentment‡** | **Linear regression** | 12045 | *Intercept* | | | 8.17 | 0.13 | 7.91 to 8.43 | 0.000 |
| | | 12045 | Childhood predictor | Yes | No | -0.17 | 0.12 | -0.41 to 0.07 | 0.164 |
| | | 12045 | Age | Per 1-year increase | | -0.02 | 0.00 | -0.02 to -0.01 | 0.000 |
| | | 12045 | Gender | Female | Male | 0.38 | 0.07 | 0.24 to 0.52 | 0.000 |
| | | 12045 | Education | Secondary | Elementary or less | 0.14 | 0.07 | -0.01 to 0.28 | 0.069 |
| | | 12045 | Education | Four years beyond HS | Elementary or less | 0.30 | 0.12 | 0.07 to 0.53 | 0.010 |
| | | 12045 | Residential setting | Small town | Rural | 0.35 | 0.08 | 0.18 to 0.51 | 0.000 |
| | | 12045 | Residential setting | Large city | Rural | 0.07 | 0.10 | -0.12 to 0.26 | 0.468 |
| | | 12045 | Residential setting | Suburb | Rural | -0.29 | 0.18 | -0.64 to 0.05 | 0.093 |
| **Positive expectations** | **Linear regression** | 12020 | *Intercept* | | | 7.86 | 0.15 | 7.56 to 8.15 | 0.000 |
| | | 12020 | Childhood predictor | Yes | No | -0.24 | 0.10 | -0.43 to -0.04 | 0.020 |
| | | 12020 | Age | Per 1-year increase | | 0.00 | 0.00 | -0.01 to 0.00 | 0.113 |
| | | 12020 | Gender | Female | Male | 0.18 | 0.06 | 0.06 to 0.31 | 0.004 |
| | | 12020 | Education | Secondary | Elementary or less | 0.27 | 0.07 | 0.13 to 0.42 | 0.000 |
| | | 12020 | Education | Four years beyond HS | Elementary or less | 0.34 | 0.10 | 0.15 to 0.54 | 0.001 |
| | | 12020 | Residential setting | Small town | Rural | 0.43 | 0.09 | 0.26 to 0.60 | 0.000 |
| | | 12020 | Residential setting | Large city | Rural | 0.43 | 0.11 | 0.21 to 0.65 | 0.000 |
| | | 12020 | Residential setting | Suburb | Rural | 0.47 | 0.16 | 0.15 to 0.79 | 0.004 |
| **Worry about meeting normal monthly living expenses§§‡‡** | **Linear regression** | 12048 | *Intercept* | | | 5.68 | 0.20 | 5.30 to 6.07 | 0.000 |
| | | 12048 | Childhood predictor | Yes | No | 0.32 | 0.13 | 0.06 to 0.58 | 0.015 |
| | | 12048 | Age | Per 1-year increase | | 0.00 | 0.00 | 0.00 to 0.01 | 0.554 |
| | | 12048 | Gender | Female | Male | 0.36 | 0.09 | 0.18 to 0.54 | 0.000 |

| Outcome | Model type | Analytic N | Variable | Level / comparison | Reference category | Coefficient | Standard error | 95% CI | p-value |
|---|---|---|---|---|---|---|---|---|---|
| | | 12048 | Education | Secondary | Elementary or less | -0.46 | 0.11 | -0.67 to -0.24 | 0.000 |
| | | 12048 | Education | Four years beyond HS | Elementary or less | -0.64 | 0.15 | -0.93 to -0.35 | 0.000 |
| | | 12048 | Residential setting | Small town | Rural | 0.13 | 0.12 | -0.11 to 0.37 | 0.289 |
| | | 12048 | Residential setting | Large city | Rural | -0.34 | 0.15 | -0.63 to -0.05 | 0.023 |
| | | 12048 | Residential setting | Suburb | Rural | -0.17 | 0.26 | -0.68 to 0.35 | 0.524 |
| **Freedom** | **Linear regression** | 12033 | *Intercept* | | | 8.20 | 0.12 | 7.96 to 8.45 | 0.000 |
| | | 12033 | Childhood predictor | Yes | No | -0.10 | 0.10 | -0.30 to 0.11 | 0.360 |
| | | 12033 | Age | Per 1-year increase | | -0.01 | 0.00 | -0.01 to 0.00 | 0.006 |
| | | 12033 | Gender | Female | Male | -0.12 | 0.07 | -0.26 to 0.02 | 0.081 |
| | | 12033 | Education | Secondary | Elementary or less | 0.30 | 0.08 | 0.15 to 0.45 | 0.000 |
| | | 12033 | Education | Four years beyond HS | Elementary or less | 0.41 | 0.10 | 0.21 to 0.61 | 0.000 |
| | | 12033 | Residential setting | Small town | Rural | 0.30 | 0.08 | 0.15 to 0.45 | 0.000 |
| | | 12033 | Residential setting | Large city | Rural | 0.35 | 0.11 | 0.15 to 0.56 | 0.001 |
| | | 12033 | Residential setting | Suburb | Rural | 0.08 | 0.16 | -0.23 to 0.38 | 0.622 |
| **Give up some happiness now for greater happiness later‡** | **Linear regression** | 12000 | *Intercept* | | | 7.21 | 0.16 | 6.90 to 7.51 | 0.000 |
| | | 12000 | Childhood predictor | Yes | No | 0.09 | 0.12 | -0.14 to 0.32 | 0.462 |
| | | 12000 | Age | Per 1-year increase | | -0.01 | 0.00 | -0.01 to 0.00 | 0.001 |
| | | 12000 | Gender | Female | Male | 0.18 | 0.08 | 0.02 to 0.33 | 0.024 |
| | | 12000 | Education | Secondary | Elementary or less | 0.19 | 0.10 | 0.00 to 0.38 | 0.048 |
| | | 12000 | Education | Four years beyond HS | Elementary or less | 0.24 | 0.14 | -0.03 to 0.51 | 0.088 |
| | | 12000 | Residential setting | Small town | Rural | 0.35 | 0.10 | 0.16 to 0.54 | 0.000 |
| | | 12000 | Residential setting | Large city | Rural | 0.09 | 0.14 | -0.18 to 0.35 | 0.530 |
| | | 12000 | Residential setting | Suburb | Rural | 0.14 | 0.27 | -0.38 to 0.67 | 0.596 |
| **Have a long list of gratitude** | **Linear regression** | 11915 | *Intercept* | | | 7.44 | 0.16 | 7.14 to 7.74 | 0.000 |
| | | 11915 | Childhood predictor | Yes | No | 0.11 | 0.11 | -0.11 to 0.32 | 0.341 |
| | | 11915 | Age | Per 1-year increase | | -0.01 | 0.00 | -0.01 to 0.00 | 0.033 |
| | | 11915 | Gender | Female | Male | 0.20 | 0.07 | 0.07 to 0.34 | 0.004 |
| | | 11915 | Education | Secondary | Elementary or less | 0.29 | 0.08 | 0.13 to 0.45 | 0.000 |
| | | 11915 | Education | Four years beyond HS | Elementary or less | 0.41 | 0.12 | 0.18 to 0.64 | 0.001 |
| | | 11915 | Residential setting | Small town | Rural | 0.37 | 0.09 | 0.19 to 0.55 | 0.000 |
| | | 11915 | Residential setting | Large city | Rural | 0.35 | 0.13 | 0.09 to 0.61 | 0.009 |
| | | 11915 | Residential setting | Suburb | Rural | 0.48 | 0.18 | 0.13 to 0.83 | 0.008 |
| **Happiness‡** | **Linear regression** | 12049 | *Intercept* | | | 6.85 | 0.16 | 6.54 to 7.17 | 0.000 |
| | | 12049 | Childhood predictor | Yes | No | -0.50 | 0.13 | -0.75 to -0.24 | 0.000 |
| | | 12049 | Age | Per 1-year increase | | -0.02 | 0.00 | -0.03 to -0.01 | 0.000 |
| | | 12049 | Gender | Female | Male | 0.22 | 0.08 | 0.06 to 0.37 | 0.006 |
| | | 12049 | Education | Secondary | Elementary or less | 0.38 | 0.09 | 0.21 to 0.55 | 0.000 |
| | | 12049 | Education | Four years beyond HS | Elementary or less | 0.66 | 0.12 | 0.42 to 0.90 | 0.000 |
| | | 12049 | Residential setting | Small town | Rural | 0.37 | 0.10 | 0.17 to 0.57 | 0.000 |
| | | 12049 | Residential setting | Large city | Rural | 0.30 | 0.12 | 0.06 to 0.55 | 0.015 |

| Outcome | Model type | Analytic N | Variable | Level / comparison | Reference category | Coefficient | Standard error | 95% CI | p-value |
|---|---|---|---|---|---|---|---|---|---|
| | | 12049 | Residential setting | Suburb | Rural | -0.12 | 0.23 | -0.57 to 0.33 | 0.596 |
| **Remain hopeful for the future** | **Linear regression** | 12015 | *Intercept* | | | 8.22 | 0.13 | 7.97 to 8.47 | 0.000 |
| | | 12015 | Childhood predictor | Yes | No | -0.21 | 0.11 | -0.43 to 0.01 | 0.061 |
| | | 12015 | Age | Per 1-year increase | | -0.01 | 0.00 | -0.01 to 0.00 | 0.001 |
| | | 12015 | Gender | Female | Male | 0.00 | 0.06 | -0.11 to 0.12 | 0.971 |
| | | 12015 | Education | Secondary | Elementary or less | 0.39 | 0.07 | 0.26 to 0.52 | 0.000 |
| | | 12015 | Education | Four years beyond HS | Elementary or less | 0.59 | 0.09 | 0.40 to 0.77 | 0.000 |
| | | 12015 | Residential setting | Small town | Rural | 0.45 | 0.08 | 0.29 to 0.61 | 0.000 |
| | | 12015 | Residential setting | Large city | Rural | 0.43 | 0.10 | 0.23 to 0.63 | 0.000 |
| | | 12015 | Residential setting | Suburb | Rural | 0.38 | 0.18 | 0.02 to 0.73 | 0.038 |
| **Understand purpose in life‡** | **Linear regression** | 11994 | *Intercept* | | | 8.03 | 0.14 | 7.76 to 8.30 | 0.000 |
| | | 11994 | Childhood predictor | Yes | No | 0.07 | 0.11 | -0.14 to 0.28 | 0.525 |
| | | 11994 | Age | Per 1-year increase | | -0.01 | 0.00 | -0.02 to -0.01 | 0.000 |
| | | 11994 | Gender | Female | Male | 0.12 | 0.07 | -0.02 to 0.26 | 0.105 |
| | | 11994 | Education | Secondary | Elementary or less | 0.40 | 0.08 | 0.24 to 0.56 | 0.000 |
| | | 11994 | Education | Four years beyond HS | Elementary or less | 0.70 | 0.10 | 0.51 to 0.90 | 0.000 |
| | | 11994 | Residential setting | Small town | Rural | 0.42 | 0.09 | 0.25 to 0.60 | 0.000 |
| | | 11994 | Residential setting | Large city | Rural | 0.44 | 0.11 | 0.22 to 0.67 | 0.000 |
| | | 11994 | Residential setting | Suburb | Rural | 0.46 | 0.15 | 0.16 to 0.76 | 0.003 |
| **Life satisfaction‡** | **Linear regression** | 12034 | *Intercept* | | | 7.19 | 0.15 | 6.89 to 7.49 | 0.000 |
| | | 12034 | Childhood predictor | Yes | No | -0.29 | 0.12 | -0.53 to -0.05 | 0.020 |
| | | 12034 | Age | Per 1-year increase | | -0.01 | 0.00 | -0.02 to -0.01 | 0.000 |
| | | 12034 | Gender | Female | Male | 0.31 | 0.08 | 0.16 to 0.46 | 0.000 |
| | | 12034 | Education | Secondary | Elementary or less | 0.28 | 0.09 | 0.10 to 0.46 | 0.002 |
| | | 12034 | Education | Four years beyond HS | Elementary or less | 0.47 | 0.12 | 0.24 to 0.71 | 0.000 |
| | | 12034 | Residential setting | Small town | Rural | 0.30 | 0.09 | 0.11 to 0.48 | 0.002 |
| | | 12034 | Residential setting | Large city | Rural | 0.20 | 0.13 | -0.05 to 0.44 | 0.111 |
| | | 12034 | Residential setting | Suburb | Rural | -0.24 | 0.22 | -0.67 to 0.19 | 0.276 |
| **Loneliness§** | **Linear regression** | 12055 | *Intercept* | | | 4.14 | 0.18 | 3.80 to 4.49 | 0.000 |
| | | 12055 | Childhood predictor | Yes | No | 0.75 | 0.14 | 0.48 to 1.02 | 0.000 |
| | | 12055 | Age | Per 1-year increase | | 0.00 | 0.00 | 0.00 to 0.01 | 0.564 |
| | | 12055 | Gender | Female | Male | -0.14 | 0.09 | -0.32 to 0.04 | 0.125 |
| | | 12055 | Education | Secondary | Elementary or less | -0.33 | 0.10 | -0.52 to -0.13 | 0.001 |
| | | 12055 | Education | Four years beyond HS | Elementary or less | -0.61 | 0.15 | -0.90 to -0.32 | 0.000 |
| | | 12055 | Residential setting | Small town | Rural | -0.41 | 0.10 | -0.61 to -0.21 | 0.000 |
| | | 12055 | Residential setting | Large city | Rural | -0.51 | 0.14 | -0.79 to -0.23 | 0.000 |
| | | 12055 | Residential setting | Suburb | Rural | -0.23 | 0.23 | -0.68 to 0.23 | 0.325 |
| **Mental health‡** | **Linear regression** | 12054 | *Intercept* | | | 8.12 | 0.14 | 7.84 to 8.40 | 0.000 |
| | | 12054 | Childhood predictor | Yes | No | -0.45 | 0.11 | -0.67 to -0.23 | 0.000 |
| | | 12054 | Age | Per 1-year increase | | -0.02 | 0.00 | -0.03 to -0.02 | 0.000 |

| Outcome | Model type | Analytic N | Variable | Level / comparison | Reference category | Coefficient | Standard error | 95% CI | p-value |
|---|---|---|---|---|---|---|---|---|---|
| | | 12054 | Gender | Female | Male | -0.12 | 0.07 | -0.26 to 0.02 | 0.089 |
| | | 12054 | Education | Secondary | Elementary or less | 0.41 | 0.08 | 0.25 to 0.56 | 0.000 |
| | | 12054 | Education | Four years beyond HS | Elementary or less | 0.53 | 0.12 | 0.30 to 0.76 | 0.000 |
| | | 12054 | Residential setting | Small town | Rural | 0.42 | 0.10 | 0.23 to 0.61 | 0.000 |
| | | 12054 | Residential setting | Large city | Rural | 0.44 | 0.12 | 0.21 to 0.67 | 0.000 |
| | | 12054 | Residential setting | Suburb | Rural | 0.16 | 0.20 | -0.22 to 0.55 | 0.404 |
| **Perceived support from others** | **Linear regression** | 12038 | *Intercept* | | | 6.72 | 0.16 | 6.40 to 7.04 | 0.000 |
| | | 12038 | Childhood predictor | Yes | No | -0.08 | 0.14 | -0.35 to 0.19 | 0.554 |
| | | 12038 | Age | Per 1-year increase | | -0.01 | 0.00 | -0.02 to -0.01 | 0.000 |
| | | 12038 | Gender | Female | Male | 0.51 | 0.09 | 0.32 to 0.69 | 0.000 |
| | | 12038 | Education | Secondary | Elementary or less | 0.19 | 0.10 | -0.01 to 0.38 | 0.058 |
| | | 12038 | Education | Four years beyond HS | Elementary or less | 0.13 | 0.14 | -0.15 to 0.41 | 0.375 |
| | | 12038 | Residential setting | Small town | Rural | 0.16 | 0.11 | -0.05 to 0.38 | 0.134 |
| | | 12038 | Residential setting | Large city | Rural | -0.46 | 0.14 | -0.73 to -0.18 | 0.001 |
| | | 12038 | Residential setting | Suburb | Rural | -0.26 | 0.20 | -0.66 to 0.13 | 0.193 |
| **Physical health‡** | **Linear regression** | 12052 | *Intercept* | | | 8.44 | 0.15 | 8.15 to 8.73 | 0.000 |
| | | 12052 | Childhood predictor | Yes | No | -0.28 | 0.11 | -0.51 to -0.06 | 0.013 |
| | | 12052 | Age | Per 1-year increase | | -0.04 | 0.00 | -0.05 to -0.04 | 0.000 |
| | | 12052 | Gender | Female | Male | -0.32 | 0.08 | -0.48 to -0.17 | 0.000 |
| | | 12052 | Education | Secondary | Elementary or less | 0.35 | 0.08 | 0.19 to 0.51 | 0.000 |
| | | 12052 | Education | Four years beyond HS | Elementary or less | 0.64 | 0.11 | 0.43 to 0.86 | 0.000 |
| | | 12052 | Residential setting | Small town | Rural | 0.49 | 0.10 | 0.30 to 0.68 | 0.000 |
| | | 12052 | Residential setting | Large city | Rural | 0.64 | 0.11 | 0.43 to 0.86 | 0.000 |
| | | 12052 | Residential setting | Suburb | Rural | 0.16 | 0.18 | -0.19 to 0.51 | 0.364 |
| **Act to promote good‡** | **Linear regression** | 12020 | *Intercept* | | | 8.08 | 0.11 | 7.86 to 8.30 | 0.000 |
| | | 12020 | Childhood predictor | Yes | No | -0.13 | 0.10 | -0.32 to 0.06 | 0.193 |
| | | 12020 | Age | Per 1-year increase | | -0.01 | 0.00 | -0.01 to 0.00 | 0.000 |
| | | 12020 | Gender | Female | Male | 0.12 | 0.06 | 0.00 to 0.23 | 0.053 |
| | | 12020 | Education | Secondary | Elementary or less | 0.40 | 0.06 | 0.27 to 0.52 | 0.000 |
| | | 12020 | Education | Four years beyond HS | Elementary or less | 0.54 | 0.08 | 0.39 to 0.70 | 0.000 |
| | | 12020 | Residential setting | Small town | Rural | 0.57 | 0.07 | 0.43 to 0.72 | 0.000 |
| | | 12020 | Residential setting | Large city | Rural | 0.61 | 0.10 | 0.41 to 0.80 | 0.000 |
| | | 12020 | Residential setting | Suburb | Rural | 0.46 | 0.16 | 0.15 to 0.76 | 0.003 |
| **Relationship satisfaction‡** | **Linear regression** | 12017 | *Intercept* | | | 7.88 | 0.13 | 7.63 to 8.14 | 0.000 |
| | | 12017 | Childhood predictor | Yes | No | -0.10 | 0.11 | -0.31 to 0.11 | 0.366 |
| | | 12017 | Age | Per 1-year increase | | -0.01 | 0.00 | -0.01 to 0.00 | 0.000 |
| | | 12017 | Gender | Female | Male | 0.31 | 0.07 | 0.17 to 0.45 | 0.000 |
| | | 12017 | Education | Secondary | Elementary or less | -0.02 | 0.08 | -0.18 to 0.14 | 0.822 |
| | | 12017 | Education | Four years beyond HS | Elementary or less | 0.05 | 0.11 | -0.17 to 0.27 | 0.652 |
| | | 12017 | Residential setting | Small town | Rural | 0.42 | 0.09 | 0.25 to 0.59 | 0.000 |

| Outcome | Model type | Analytic N | Variable | Level / comparison | Reference category | Coefficient | Standard error | 95% CI | p-value |
|---|---|---|---|---|---|---|---|---|---|
| | | 12017 | Residential setting | Large city | Rural | 0.32 | 0.11 | 0.11 to 0.53 | 0.003 |
| | | 12017 | Residential setting | Suburb | Rural | 0.03 | 0.16 | -0.28 to 0.33 | 0.869 |
| **Show love often** | **Linear regression** | 12043 | *Intercept* | | | 8.30 | 0.13 | 8.04 to 8.55 | 0.000 |
| | | 12043 | Childhood predictor | Yes | No | -0.08 | 0.11 | -0.31 to 0.14 | 0.459 |
| | | 12043 | Age | Per 1-year increase | | -0.01 | 0.00 | -0.01 to 0.00 | 0.001 |
| | | 12043 | Gender | Female | Male | 0.19 | 0.07 | 0.06 to 0.33 | 0.006 |
| | | 12043 | Education | Secondary | Elementary or less | 0.17 | 0.07 | 0.03 to 0.31 | 0.021 |
| | | 12043 | Education | Four years beyond HS | Elementary or less | 0.07 | 0.11 | -0.14 to 0.29 | 0.515 |
| | | 12043 | Residential setting | Small town | Rural | 0.17 | 0.08 | 0.01 to 0.32 | 0.032 |
| | | 12043 | Residential setting | Large city | Rural | -0.03 | 0.12 | -0.26 to 0.20 | 0.779 |
| | | 12043 | Residential setting | Suburb | Rural | -0.14 | 0.20 | -0.54 to 0.26 | 0.489 |
| **Life evaluation five years from now** | **Linear regression** | 11048 | *Intercept* | | | 7.77 | 0.14 | 7.49 to 8.05 | 0.000 |
| | | 11048 | Childhood predictor | Yes | No | -0.17 | 0.12 | -0.40 to 0.06 | 0.152 |
| | | 11048 | Age | Per 1-year increase | | -0.02 | 0.00 | -0.03 to -0.02 | 0.000 |
| | | 11048 | Gender | Female | Male | 0.17 | 0.07 | 0.03 to 0.32 | 0.019 |
| | | 11048 | Education | Secondary | Elementary or less | 0.26 | 0.08 | 0.10 to 0.42 | 0.001 |
| | | 11048 | Education | Four years beyond HS | Elementary or less | 0.58 | 0.12 | 0.36 to 0.81 | 0.000 |
| | | 11048 | Residential setting | Small town | Rural | 0.43 | 0.09 | 0.25 to 0.62 | 0.000 |
| | | 11048 | Residential setting | Large city | Rural | 0.47 | 0.11 | 0.25 to 0.68 | 0.000 |
| | | 11048 | Residential setting | Suburb | Rural | 0.30 | 0.24 | -0.16 to 0.76 | 0.201 |
| **Life evaluation today** | **Linear regression** | 11973 | *Intercept* | | | 5.62 | 0.17 | 5.28 to 5.96 | 0.000 |
| | | 11973 | Childhood predictor | Yes | No | 0.15 | 0.13 | -0.10 to 0.41 | 0.238 |
| | | 11973 | Age | Per 1-year increase | | -0.01 | 0.00 | -0.02 to -0.01 | 0.001 |
| | | 11973 | Gender | Female | Male | 0.49 | 0.09 | 0.32 to 0.66 | 0.000 |
| | | 11973 | Education | Secondary | Elementary or less | 0.18 | 0.09 | 0.01 to 0.35 | 0.040 |
| | | 11973 | Education | Four years beyond HS | Elementary or less | 0.17 | 0.14 | -0.11 to 0.45 | 0.224 |
| | | 11973 | Residential setting | Small town | Rural | 0.22 | 0.11 | 0.01 to 0.43 | 0.037 |
| | | 11973 | Residential setting | Large city | Rural | 0.10 | 0.14 | -0.17 to 0.37 | 0.469 |
| | | 11973 | Residential setting | Suburb | Rural | -0.03 | 0.22 | -0.46 to 0.39 | 0.880 |
| **Worry about safety, food, or housing§§‡‡** | **Linear regression** | 12062 | *Intercept* | | | 6.14 | 0.19 | 5.77 to 6.52 | 0.000 |
| | | 12062 | Childhood predictor | Yes | No | 0.43 | 0.14 | 0.16 to 0.70 | 0.002 |
| | | 12062 | Age | Per 1-year increase | | -0.01 | 0.00 | -0.02 to 0.00 | 0.014 |
| | | 12062 | Gender | Female | Male | 0.32 | 0.09 | 0.15 to 0.50 | 0.000 |
| | | 12062 | Education | Secondary | Elementary or less | -0.56 | 0.11 | -0.77 to -0.35 | 0.000 |
| | | 12062 | Education | Four years beyond HS | Elementary or less | -0.78 | 0.17 | -1.12 to -0.45 | 0.000 |
| | | 12062 | Residential setting | Small town | Rural | 0.30 | 0.13 | 0.05 to 0.55 | 0.018 |
| | | 12062 | Residential setting | Large city | Rural | -0.21 | 0.17 | -0.55 to 0.12 | 0.215 |
| | | 12062 | Residential setting | Suburb | Rural | 0.20 | 0.26 | -0.31 to 0.71 | 0.437 |
| **Life worthwhile‡** | **Linear regression** | 11974 | *Intercept* | | | 6.70 | 0.15 | 6.41 to 6.99 | 0.000 |

| Outcome | Model type | Analytic N | Variable | Level / comparison | Reference category | Coefficient | Standard error | 95% CI | p-value |
|---|---|---|---|---|---|---|---|---|---|
| | | 11974 | Childhood predictor | Yes | No | -0.13 | 0.12 | -0.35 to 0.10 | 0.286 |
| | | 11974 | Age | Per 1-year increase | | -0.01 | 0.00 | -0.02 to 0.00 | 0.000 |
| | | 11974 | Gender | Female | Male | 0.48 | 0.07 | 0.33 to 0.62 | 0.000 |
| | | 11974 | Education | Secondary | Elementary or less | 0.42 | 0.08 | 0.26 to 0.57 | 0.000 |
| | | 11974 | Education | Four years beyond HS | Elementary or less | 0.58 | 0.12 | 0.34 to 0.81 | 0.000 |
| | | 11974 | Residential setting | Small town | Rural | 0.46 | 0.10 | 0.26 to 0.66 | 0.000 |
| | | 11974 | Residential setting | Large city | Rural | 0.45 | 0.12 | 0.21 to 0.69 | 0.000 |
| | | 11974 | Residential setting | Suburb | Rural | 0.28 | 0.22 | -0.14 to 0.70 | 0.195 |
| **Flourishing (with financial indicators)** | **Linear regression** | 11676 | *Intercept* | | | 7.04 | 0.09 | 6.88 to 7.21 | 0.000 |
| | | 11676 | Childhood predictor | Yes | No | -0.22 | 0.06 | -0.34 to -0.09 | 0.001 |
| | | 11676 | Age | Per 1-year increase | | -0.01 | 0.00 | -0.02 to -0.01 | 0.000 |
| | | 11676 | Gender | Female | Male | 0.09 | 0.04 | 0.01 to 0.18 | 0.036 |
| | | 11676 | Education | Secondary | Elementary or less | 0.32 | 0.05 | 0.23 to 0.41 | 0.000 |
| | | 11676 | Education | Four years beyond HS | Elementary or less | 0.51 | 0.07 | 0.38 to 0.64 | 0.000 |
| | | 11676 | Residential setting | Small town | Rural | 0.32 | 0.06 | 0.21 to 0.44 | 0.000 |
| | | 11676 | Residential setting | Large city | Rural | 0.36 | 0.07 | 0.22 to 0.49 | 0.000 |
| | | 11676 | Residential setting | Suburb | Rural | 0.09 | 0.14 | -0.17 to 0.36 | 0.495 |
| **Flourishing (without financial indicators)** | **Linear regression** | 11700 | *Intercept* | | | 7.65 | 0.09 | 7.47 to 7.83 | 0.000 |
| | | 11700 | Childhood predictor | Yes | No | -0.19 | 0.07 | -0.32 to -0.05 | 0.007 |
| | | 11700 | Age | Per 1-year increase | | -0.02 | 0.00 | -0.02 to -0.01 | 0.000 |
| | | 11700 | Gender | Female | Male | 0.18 | 0.05 | 0.08 to 0.27 | 0.000 |
| | | 11700 | Education | Secondary | Elementary or less | 0.29 | 0.05 | 0.19 to 0.39 | 0.000 |
| | | 11700 | Education | Four years beyond HS | Elementary or less | 0.47 | 0.07 | 0.33 to 0.61 | 0.000 |
| | | 11700 | Residential setting | Small town | Rural | 0.43 | 0.06 | 0.31 to 0.56 | 0.000 |
| | | 11700 | Residential setting | Large city | Rural | 0.38 | 0.08 | 0.23 to 0.53 | 0.000 |
| | | 11700 | Residential setting | Suburb | Rural | 0.11 | 0.14 | -0.15 to 0.38 | 0.404 |
| **Feel close to someone** | **Logistic regression** | 12055 | *Intercept* | | | 1.63 | 0.14 | 1.36 to 1.90 | 0.000 |
| | | 12055 | Childhood predictor | Yes | No | 0.07 | 0.09 | -0.10 to 0.23 | 0.450 |
| | | 12055 | Age | Per 1-year increase | | -0.01 | 0.00 | -0.01 to 0.00 | 0.009 |
| | | 12055 | Gender | Female | Male | 0.03 | 0.06 | -0.10 to 0.15 | 0.687 |
| | | 12055 | Education | Secondary | Elementary or less | 0.17 | 0.08 | 0.01 to 0.33 | 0.039 |
| | | 12055 | Education | Four years beyond HS | Elementary or less | 0.17 | 0.13 | -0.09 to 0.42 | 0.195 |
| | | 12055 | Residential setting | Small town | Rural | 0.11 | 0.11 | -0.11 to 0.33 | 0.317 |
| | | 12055 | Residential setting | Large city | Rural | -0.09 | 0.13 | -0.34 to 0.16 | 0.466 |
| | | 12055 | Residential setting | Suburb | Rural | -0.14 | 0.20 | -0.52 to 0.24 | 0.476 |
| **Donated to charity** | **Logistic regression** | 12023 | *Intercept* | | | -0.41 | 0.13 | -0.66 to -0.16 | 0.001 |
| | | 12023 | Childhood predictor | Yes | No | 0.24 | 0.07 | 0.10 to 0.38 | 0.001 |
| | | 12023 | Age | Per 1-year increase | | 0.00 | 0.00 | 0.00 to 0.00 | 0.636 |
| | | 12023 | Gender | Female | Male | 0.00 | 0.05 | -0.10 to 0.10 | 0.990 |

| Outcome | Model type | Analytic N | Variable | Level / comparison | Reference category | Coefficient | Standard error | 95% CI | p-value |
|---|---|---|---|---|---|---|---|---|---|
| | | 12023 | Education | Secondary | Elementary or less | -0.08 | 0.06 | -0.20 to 0.04 | 0.203 |
| | | 12023 | Education | Four years beyond HS | Elementary or less | -0.12 | 0.10 | -0.32 to 0.08 | 0.242 |
| | | 12023 | Residential setting | Small town | Rural | -0.04 | 0.09 | -0.22 to 0.14 | 0.648 |
| | | 12023 | Residential setting | Large city | Rural | -0.40 | 0.11 | -0.62 to -0.18 | 0.001 |
| | | 12023 | Residential setting | Suburb | Rural | -0.16 | 0.19 | -0.53 to 0.22 | 0.414 |
| **Have any health problems** | **Logistic regression** | 12031 | *Intercept* | | | -2.24 | 0.10 | -2.44 to -2.03 | 0.000 |
| | | 12031 | Childhood predictor | Yes | No | 1.08 | 0.07 | 0.93 to 1.22 | 0.000 |
| | | 12031 | Age | Per 1-year increase | | 0.03 | 0.00 | 0.02 to 0.03 | 0.000 |
| | | 12031 | Gender | Female | Male | 0.28 | 0.05 | 0.19 to 0.38 | 0.000 |
| | | 12031 | Education | Secondary | Elementary or less | -0.10 | 0.06 | -0.22 to 0.02 | 0.112 |
| | | 12031 | Education | Four years beyond HS | Elementary or less | -0.20 | 0.09 | -0.39 to -0.02 | 0.031 |
| | | 12031 | Residential setting | Small town | Rural | 0.11 | 0.07 | -0.02 to 0.24 | 0.093 |
| | | 12031 | Residential setting | Large city | Rural | -0.16 | 0.09 | -0.34 to 0.02 | 0.087 |
| | | 12031 | Residential setting | Suburb | Rural | 0.07 | 0.14 | -0.20 to 0.33 | 0.626 |
| **Helped a stranger** | **Logistic regression** | 11929 | *Intercept* | | | 0.61 | 0.12 | 0.38 to 0.84 | 0.000 |
| | | 11929 | Childhood predictor | Yes | No | 0.18 | 0.07 | 0.05 to 0.32 | 0.007 |
| | | 11929 | Age | Per 1-year increase | | -0.01 | 0.00 | -0.01 to 0.00 | 0.000 |
| | | 11929 | Gender | Female | Male | -0.30 | 0.05 | -0.39 to -0.21 | 0.000 |
| | | 11929 | Education | Secondary | Elementary or less | -0.05 | 0.06 | -0.16 to 0.07 | 0.423 |
| | | 11929 | Education | Four years beyond HS | Elementary or less | -0.08 | 0.09 | -0.25 to 0.10 | 0.385 |
| | | 11929 | Residential setting | Small town | Rural | 0.10 | 0.09 | -0.06 to 0.27 | 0.228 |
| | | 11929 | Residential setting | Large city | Rural | -0.19 | 0.10 | -0.39 to 0.00 | 0.056 |
| | | 11929 | Residential setting | Suburb | Rural | 0.08 | 0.18 | -0.27 to 0.44 | 0.643 |
| **Volunteered at an organization** | **Logistic regression** | 12007 | *Intercept* | | | -0.36 | 0.14 | -0.64 to -0.08 | 0.013 |
| | | 12007 | Childhood predictor | Yes | No | 0.27 | 0.07 | 0.13 to 0.41 | 0.000 |
| | | 12007 | Age | Per 1-year increase | | -0.01 | 0.00 | -0.01 to 0.00 | 0.001 |
| | | 12007 | Gender | Female | Male | -0.25 | 0.05 | -0.35 to -0.14 | 0.000 |
| | | 12007 | Education | Secondary | Elementary or less | -0.15 | 0.07 | -0.28 to -0.02 | 0.021 |
| | | 12007 | Education | Four years beyond HS | Elementary or less | 0.05 | 0.10 | -0.15 to 0.25 | 0.615 |
| | | 12007 | Residential setting | Small town | Rural | -0.08 | 0.11 | -0.29 to 0.13 | 0.455 |
| | | 12007 | Residential setting | Large city | Rural | -0.54 | 0.13 | -0.79 to -0.29 | 0.000 |
| | | 12007 | Residential setting | Suburb | Rural | -0.33 | 0.16 | -0.64 to -0.01 | 0.044 |
| **Cigarette smoking** | **Logistic regression** | 11625 | *Intercept* | | | -2.08 | 0.16 | -2.40 to -1.76 | 0.000 |
| | | 11625 | Childhood predictor | Yes | No | 0.43 | 0.12 | 0.19 to 0.66 | 0.000 |
| | | 11625 | Age | Per 1-year increase | | 0.01 | 0.00 | 0.00 to 0.01 | 0.012 |
| | | 11625 | Gender | Female | Male | -2.82 | 0.18 | -3.18 to -2.47 | 0.000 |
| | | 11625 | Education | Secondary | Elementary or less | -0.34 | 0.10 | -0.53 to -0.14 | 0.001 |
| | | 11625 | Education | Four years beyond HS | Elementary or less | -0.59 | 0.17 | -0.93 to -0.26 | 0.001 |
| | | 11625 | Residential setting | Small town | Rural | -0.10 | 0.11 | -0.31 to 0.11 | 0.340 |
| | | 11625 | Residential setting | Large city | Rural | 0.08 | 0.14 | -0.20 to 0.37 | 0.571 |

| Outcome | Model type | Analytic N | Variable | Level / comparison | Reference category | Coefficient | Standard error | 95% CI | p-value |
|---|---|---|---|---|---|---|---|---|---|
| | | 11625 | Residential setting | Suburb | Rural | 0.78 | 0.21 | 0.37 to 1.18 | 0.000 |
| **Alcohol use** | **Logistic regression** | 11615 | *Intercept* | | | -1.94 | 0.16 | -2.25 to -1.64 | 0.000 |
| | | 11615 | Childhood predictor | Yes | No | 0.45 | 0.12 | 0.22 to 0.68 | 0.000 |
| | | 11615 | Age | Per 1-year increase | | 0.00 | 0.00 | -0.01 to 0.01 | 0.882 |
| | | 11615 | Gender | Female | Male | -2.69 | 0.17 | -3.03 to -2.35 | 0.000 |
| | | 11615 | Education | Secondary | Elementary or less | -0.24 | 0.10 | -0.43 to -0.05 | 0.015 |
| | | 11615 | Education | Four years beyond HS | Elementary or less | -0.47 | 0.16 | -0.79 to -0.15 | 0.004 |
| | | 11615 | Residential setting | Small town | Rural | 0.16 | 0.11 | -0.05 to 0.36 | 0.145 |
| | | 11615 | Residential setting | Large city | Rural | 0.35 | 0.14 | 0.07 to 0.62 | 0.013 |
| | | 11615 | Residential setting | Suburb | Rural | 0.69 | 0.22 | 0.26 to 1.11 | 0.002 |
| **Government approval§** | **Proportional-odds regression** | 11862 | Childhood predictor | Yes | No | 0.24 | 0.07 | 0.11 to 0.38 | 0.000 |
| | | 11862 | Age | Per 1-year increase | | 0.00 | 0.00 | -0.01 to 0.00 | 0.008 |
| | | 11862 | Gender | Female | Male | 0.06 | 0.04 | -0.02 to 0.15 | 0.150 |
| | | 11862 | Education | Secondary | Elementary or less | -0.24 | 0.05 | -0.34 to -0.13 | 0.000 |
| | | 11862 | Education | Four years beyond HS | Elementary or less | -0.30 | 0.08 | -0.46 to -0.13 | 0.000 |
| | | 11862 | Residential setting | Small town | Rural | 0.07 | 0.07 | -0.06 to 0.20 | 0.265 |
| | | 11862 | Residential setting | Large city | Rural | -0.09 | 0.08 | -0.25 to 0.06 | 0.231 |
| | | 11862 | Residential setting | Suburb | Rural | -0.45 | 0.16 | -0.76 to -0.14 | 0.004 |
| | | 11862 | *Threshold / cut-point* | Strongly disapprove\|Somewhat disapprove | | -2.95 | 0.10 | -3.15 to -2.75 | 0.000 |
| | | 11862 | *Threshold / cut-point* | Somewhat disapprove\|Neither | | -2.26 | 0.10 | -2.45 to -2.07 | 0.000 |
| | | 11862 | *Threshold / cut-point* | Neither\|Somewhat approve | | -1.90 | 0.09 | -2.08 to -1.71 | 0.000 |
| | | 11862 | *Threshold / cut-point* | Somewhat approve\|Strongly approve | | -0.45 | 0.09 | -0.62 to -0.27 | 0.000 |
| **Feeling capable§** | **Proportional-odds regression** | 12023 | Childhood predictor | Yes | No | -0.07 | 0.07 | -0.21 to 0.06 | 0.263 |
| | | 12023 | Age | Per 1-year increase | | 0.00 | 0.00 | 0.00 to 0.01 | 0.188 |
| | | 12023 | Gender | Female | Male | -0.09 | 0.04 | -0.17 to 0.00 | 0.053 |
| | | 12023 | Education | Secondary | Elementary or less | 0.14 | 0.05 | 0.04 to 0.24 | 0.005 |
| | | 12023 | Education | Four years beyond HS | Elementary or less | 0.06 | 0.07 | -0.08 to 0.20 | 0.371 |
| | | 12023 | Residential setting | Small town | Rural | 0.14 | 0.05 | 0.04 to 0.24 | 0.005 |
| | | 12023 | Residential setting | Large city | Rural | 0.19 | 0.07 | 0.05 to 0.34 | 0.009 |
| | | 12023 | Residential setting | Suburb | Rural | 0.26 | 0.09 | 0.09 to 0.43 | 0.003 |
| | | 12023 | *Threshold / cut-point* | Never\|Rarely | | -2.35 | 0.09 | -2.52 to -2.17 | 0.000 |
| | | 12023 | *Threshold / cut-point* | Rarely\|Often | | -0.65 | 0.09 | -0.82 to -0.49 | 0.000 |
| | | 12023 | *Threshold / cut-point* | Often\|Always | | 0.02 | 0.09 | -0.15 to 0.18 | 0.845 |
| **Unable to control worry§** | **Proportional-odds regression** | 11982 | Childhood predictor | Yes | No | 0.78 | 0.06 | 0.65 to 0.90 | 0.000 |
| | | 11982 | Age | Per 1-year increase | | 0.01 | 0.00 | 0.00 to 0.01 | 0.001 |
| | | 11982 | Gender | Female | Male | 0.22 | 0.04 | 0.14 to 0.31 | 0.000 |

| Outcome | Model type | Analytic N | Variable | Level / comparison | Reference category | Coefficient | Standard error | 95% CI | p-value |
|---|---|---|---|---|---|---|---|---|---|
| | | 11982 | Education | Secondary | Elementary or less | -0.24 | 0.05 | -0.33 to -0.15 | 0.000 |
| | | 11982 | Education | Four years beyond HS | Elementary or less | -0.38 | 0.08 | -0.53 to -0.23 | 0.000 |
| | | 11982 | Residential setting | Small town | Rural | 0.04 | 0.05 | -0.06 to 0.13 | 0.432 |
| | | 11982 | Residential setting | Large city | Rural | -0.02 | 0.07 | -0.15 to 0.12 | 0.823 |
| | | 11982 | Residential setting | Suburb | Rural | -0.09 | 0.09 | -0.27 to 0.10 | 0.348 |
| | | 11982 | *Threshold / cut-point* | Not at all\|Several days | | 0.21 | 0.08 | 0.05 to 0.37 | 0.012 |
| | | 11982 | *Threshold / cut-point* | Several days\|More than half the days | | 1.15 | 0.08 | 0.99 to 1.31 | 0.000 |
| | | 11982 | *Threshold / cut-point* | More than half the days\|Nearly every day | | 1.97 | 0.08 | 1.81 to 2.14 | 0.000 |
| **Depressive symptoms§** | **Proportional-odds regression** | 12000 | Childhood predictor | Yes | No | 0.58 | 0.06 | 0.46 to 0.70 | 0.000 |
| | | 12000 | Age | Per 1-year increase | | 0.00 | 0.00 | 0.00 to 0.01 | 0.117 |
| | | 12000 | Gender | Female | Male | 0.13 | 0.04 | 0.05 to 0.21 | 0.001 |
| | | 12000 | Education | Secondary | Elementary or less | -0.17 | 0.05 | -0.27 to -0.08 | 0.000 |
| | | 12000 | Education | Four years beyond HS | Elementary or less | -0.35 | 0.07 | -0.48 to -0.21 | 0.000 |
| | | 12000 | Residential setting | Small town | Rural | -0.05 | 0.05 | -0.15 to 0.05 | 0.360 |
| | | 12000 | Residential setting | Large city | Rural | -0.12 | 0.07 | -0.25 to 0.02 | 0.099 |
| | | 12000 | Residential setting | Suburb | Rural | -0.23 | 0.12 | -0.46 to 0.01 | 0.055 |
| | | 12000 | *Threshold / cut-point* | Not at all\|Several days | | -0.14 | 0.08 | -0.30 to 0.02 | 0.083 |
| | | 12000 | *Threshold / cut-point* | Several days\|More than half the days | | 0.97 | 0.08 | 0.81 to 1.13 | 0.000 |
| | | 12000 | *Threshold / cut-point* | More than half the days\|Nearly every day | | 1.96 | 0.09 | 1.80 to 2.13 | 0.000 |
| **Perceived discrimination§** | **Proportional-odds regression** | 11934 | Childhood predictor | Yes | No | 0.65 | 0.06 | 0.53 to 0.77 | 0.000 |
| | | 11934 | Age | Per 1-year increase | | 0.00 | 0.00 | -0.01 to 0.00 | 0.070 |
| | | 11934 | Gender | Female | Male | -0.10 | 0.05 | -0.19 to -0.01 | 0.026 |
| | | 11934 | Education | Secondary | Elementary or less | -0.21 | 0.05 | -0.31 to -0.11 | 0.000 |
| | | 11934 | Education | Four years beyond HS | Elementary or less | -0.25 | 0.09 | -0.41 to -0.08 | 0.004 |
| | | 11934 | Residential setting | Small town | Rural | -0.21 | 0.06 | -0.32 to -0.09 | 0.000 |
| | | 11934 | Residential setting | Large city | Rural | -0.20 | 0.07 | -0.34 to -0.06 | 0.006 |
| | | 11934 | Residential setting | Suburb | Rural | -0.33 | 0.10 | -0.52 to -0.14 | 0.001 |
| | | 11934 | *Threshold / cut-point* | Never\|Rarely | | -0.02 | 0.09 | -0.20 to 0.15 | 0.814 |
| | | 11934 | *Threshold / cut-point* | Rarely\|Often | | 0.98 | 0.09 | 0.80 to 1.16 | 0.000 |
| | | 11934 | *Threshold / cut-point* | Often\|Always | | 1.62 | 0.09 | 1.44 to 1.80 | 0.000 |
| **Feeling nervous or anxious§** | **Proportional-odds regression** | 12018 | Childhood predictor | Yes | No | 0.61 | 0.07 | 0.48 to 0.74 | 0.000 |
| | | 12018 | Age | Per 1-year increase | | 0.01 | 0.00 | 0.00 to 0.01 | 0.000 |
| | | 12018 | Gender | Female | Male | 0.35 | 0.04 | 0.27 to 0.43 | 0.000 |
| | | 12018 | Education | Secondary | Elementary or less | -0.27 | 0.05 | -0.36 to -0.18 | 0.000 |
| | | 12018 | Education | Four years beyond HS | Elementary or less | -0.40 | 0.08 | -0.56 to -0.25 | 0.000 |

| Outcome | Model type | Analytic N | Variable | Level / comparison | Reference category | Coefficient | Standard error | 95% CI | p-value |
|---|---|---|---|---|---|---|---|---|---|
| | | 12018 | Residential setting | Small town | Rural | 0.05 | 0.05 | -0.05 to 0.15 | 0.362 |
| | | 12018 | Residential setting | Large city | Rural | -0.07 | 0.07 | -0.20 to 0.06 | 0.291 |
| | | 12018 | Residential setting | Suburb | Rural | -0.09 | 0.12 | -0.33 to 0.14 | 0.438 |
| | | 12018 | *Threshold / cut-point* | Not at all\|Several days | | 0.22 | 0.08 | 0.07 to 0.38 | 0.005 |
| | | 12018 | *Threshold / cut-point* | Several days\|More than half the days | | 1.25 | 0.08 | 1.10 to 1.41 | 0.000 |
| | | 12018 | *Threshold / cut-point* | More than half the days\|Nearly every day | | 2.18 | 0.08 | 2.02 to 2.34 | 0.000 |
| **Forgiveness§** | **Proportional-odds regression** | 12001 | Childhood predictor | Yes | No | -0.07 | 0.07 | -0.22 to 0.07 | 0.298 |
| | | 12001 | Age | Per 1-year increase | | 0.00 | 0.00 | 0.00 to 0.00 | 0.874 |
| | | 12001 | Gender | Female | Male | 0.07 | 0.04 | -0.02 to 0.16 | 0.126 |
| | | 12001 | Education | Secondary | Elementary or less | -0.08 | 0.05 | -0.18 to 0.03 | 0.141 |
| | | 12001 | Education | Four years beyond HS | Elementary or less | -0.21 | 0.08 | -0.36 to -0.05 | 0.008 |
| | | 12001 | Residential setting | Small town | Rural | 0.03 | 0.05 | -0.08 to 0.13 | 0.590 |
| | | 12001 | Residential setting | Large city | Rural | 0.06 | 0.08 | -0.10 to 0.21 | 0.471 |
| | | 12001 | Residential setting | Suburb | Rural | 0.02 | 0.12 | -0.21 to 0.26 | 0.847 |
| | | 12001 | *Threshold / cut-point* | Never\|Rarely | | -2.40 | 0.09 | -2.58 to -2.22 | 0.000 |
| | | 12001 | *Threshold / cut-point* | Rarely\|Often | | -1.08 | 0.09 | -1.25 to -0.90 | 0.000 |
| | | 12001 | *Threshold / cut-point* | Often\|Always | | -0.42 | 0.09 | -0.59 to -0.26 | 0.000 |
| **Support reducing differences in income levels§** | **Proportional-odds regression** | 11847 | Childhood predictor | Yes | No | -0.03 | 0.08 | -0.19 to 0.14 | 0.751 |
| | | 11847 | Age | Per 1-year increase | | 0.00 | 0.00 | -0.01 to 0.00 | 0.061 |
| | | 11847 | Gender | Female | Male | -0.09 | 0.06 | -0.20 to 0.03 | 0.135 |
| | | 11847 | Education | Secondary | Elementary or less | -0.05 | 0.07 | -0.19 to 0.09 | 0.491 |
| | | 11847 | Education | Four years beyond HS | Elementary or less | -0.26 | 0.11 | -0.48 to -0.05 | 0.014 |
| | | 11847 | Residential setting | Small town | Rural | 0.05 | 0.13 | -0.20 to 0.30 | 0.678 |
| | | 11847 | Residential setting | Large city | Rural | 0.13 | 0.15 | -0.16 to 0.41 | 0.394 |
| | | 11847 | Residential setting | Suburb | Rural | -0.04 | 0.19 | -0.40 to 0.33 | 0.837 |
| | | 11847 | *Threshold / cut-point* | Strongly disagree\|Somewhat disagree | | -3.87 | 0.18 | -4.21 to -3.52 | 0.000 |
| | | 11847 | *Threshold / cut-point* | Somewhat disagree\|Neither | | -3.32 | 0.16 | -3.65 to -3.00 | 0.000 |
| | | 11847 | *Threshold / cut-point* | Neither\|Somewhat agree | | -2.88 | 0.16 | -3.19 to -2.58 | 0.000 |
| | | 11847 | *Threshold / cut-point* | Somewhat agree\|Strongly agree | | -1.33 | 0.14 | -1.60 to -1.05 | 0.000 |
| **Have little interest in doing things§** | **Proportional-odds regression** | 11998 | Childhood predictor | Yes | No | 0.37 | 0.06 | 0.25 to 0.49 | 0.000 |
| | | 11998 | Age | Per 1-year increase | | 0.00 | 0.00 | 0.00 to 0.01 | 0.006 |
| | | 11998 | Gender | Female | Male | 0.04 | 0.04 | -0.05 to 0.12 | 0.408 |
| | | 11998 | Education | Secondary | Elementary or less | 0.08 | 0.05 | -0.02 to 0.17 | 0.105 |
| | | 11998 | Education | Four years beyond HS | Elementary or less | 0.12 | 0.08 | -0.03 to 0.27 | 0.131 |
| | | 11998 | Residential setting | Small town | Rural | 0.26 | 0.06 | 0.14 to 0.39 | 0.000 |

| Outcome | Model type | Analytic N | Variable | Level / comparison | Reference category | Coefficient | Standard error | 95% CI | p-value |
|---|---|---|---|---|---|---|---|---|---|
| | | 11998 | Residential setting | Large city | Rural | 0.09 | 0.08 | -0.07 to 0.24 | 0.267 |
| | | 11998 | Residential setting | Suburb | Rural | 0.29 | 0.15 | -0.02 to 0.59 | 0.064 |
| | | 11998 | *Threshold / cut-point* | Not at all\|Several days | | -0.39 | 0.09 | -0.56 to -0.22 | 0.000 |
| | | 11998 | *Threshold / cut-point* | Several days\|More than half the days | | 0.55 | 0.09 | 0.38 to 0.73 | 0.000 |
| | | 11998 | *Threshold / cut-point* | More than half the days\|Nearly every day | | 1.30 | 0.09 | 1.12 to 1.48 | 0.000 |
| **Various aspects of life are balanced§** | **Proportional-odds regression** | 11935 | Childhood predictor | Yes | No | 0.12 | 0.07 | -0.01 to 0.26 | 0.076 |
| | | 11935 | Age | Per 1-year increase | | 0.00 | 0.00 | 0.00 to 0.01 | 0.108 |
| | | 11935 | Gender | Female | Male | -0.02 | 0.04 | -0.10 to 0.07 | 0.699 |
| | | 11935 | Education | Secondary | Elementary or less | 0.05 | 0.05 | -0.04 to 0.13 | 0.309 |
| | | 11935 | Education | Four years beyond HS | Elementary or less | 0.11 | 0.07 | -0.03 to 0.24 | 0.122 |
| | | 11935 | Residential setting | Small town | Rural | 0.09 | 0.05 | -0.01 to 0.20 | 0.077 |
| | | 11935 | Residential setting | Large city | Rural | 0.07 | 0.07 | -0.06 to 0.20 | 0.263 |
| | | 11935 | Residential setting | Suburb | Rural | 0.36 | 0.11 | 0.15 to 0.58 | 0.001 |
| | | 11935 | *Threshold / cut-point* | Never\|Rarely | | -2.00 | 0.09 | -2.17 to -1.82 | 0.000 |
| | | 11935 | *Threshold / cut-point* | Rarely\|Often | | 0.14 | 0.08 | -0.02 to 0.30 | 0.094 |
| | | 11935 | *Threshold / cut-point* | Often\|Always | | 0.94 | 0.08 | 0.78 to 1.10 | 0.000 |
| **Obeying the law§** | **Proportional-odds regression** | 12022 | Childhood predictor | Yes | No | -0.14 | 0.11 | -0.35 to 0.07 | 0.197 |
| | | 12022 | Age | Per 1-year increase | | 0.00 | 0.00 | -0.01 to 0.00 | 0.458 |
| | | 12022 | Gender | Female | Male | -0.17 | 0.07 | -0.31 to -0.02 | 0.022 |
| | | 12022 | Education | Secondary | Elementary or less | 0.15 | 0.10 | -0.04 to 0.34 | 0.114 |
| | | 12022 | Education | Four years beyond HS | Elementary or less | -0.19 | 0.12 | -0.43 to 0.06 | 0.133 |
| | | 12022 | Residential setting | Small town | Rural | -0.03 | 0.15 | -0.32 to 0.26 | 0.847 |
| | | 12022 | Residential setting | Large city | Rural | -0.21 | 0.18 | -0.55 to 0.14 | 0.239 |
| | | 12022 | Residential setting | Suburb | Rural | -0.25 | 0.23 | -0.70 to 0.20 | 0.276 |
| | | 12022 | *Threshold / cut-point* | Strongly disagree\|Somewhat disagree | | -5.13 | 0.25 | -5.61 to -4.64 | 0.000 |
| | | 12022 | *Threshold / cut-point* | Somewhat disagree\|Neither | | -4.57 | 0.21 | -4.97 to -4.16 | 0.000 |
| | | 12022 | *Threshold / cut-point* | Neither\|Somewhat agree | | -4.05 | 0.20 | -4.44 to -3.66 | 0.000 |
| | | 12022 | *Threshold / cut-point* | Somewhat agree\|Strongly agree | | -2.20 | 0.18 | -2.56 to -1.84 | 0.000 |
| **Feel at peace§** | **Proportional-odds regression** | 11989 | Childhood predictor | Yes | No | -0.04 | 0.06 | -0.17 to 0.08 | 0.489 |
| | | 11989 | Age | Per 1-year increase | | 0.01 | 0.00 | 0.00 to 0.01 | 0.000 |
| | | 11989 | Gender | Female | Male | 0.00 | 0.04 | -0.08 to 0.08 | 0.996 |
| | | 11989 | Education | Secondary | Elementary or less | 0.00 | 0.05 | -0.09 to 0.09 | 0.976 |
| | | 11989 | Education | Four years beyond HS | Elementary or less | 0.00 | 0.06 | -0.12 to 0.13 | 0.953 |
| | | 11989 | Residential setting | Small town | Rural | 0.17 | 0.05 | 0.06 to 0.27 | 0.002 |
| | | 11989 | Residential setting | Large city | Rural | 0.13 | 0.07 | -0.01 to 0.27 | 0.060 |

| Outcome | Model type | Analytic N | Variable | Level / comparison | Reference category | Coefficient | Standard error | 95% CI | p-value |
|---|---|---|---|---|---|---|---|---|---|
| | | 11989 | Residential setting | Suburb | Rural | 0.24 | 0.10 | 0.04 to 0.43 | 0.017 |
| | | 11989 | *Threshold / cut-point* | Never\|Rarely | | -1.91 | 0.08 | -2.07 to -1.74 | 0.000 |
| | | 11989 | *Threshold / cut-point* | Rarely\|Often | | -0.11 | 0.08 | -0.26 to 0.05 | 0.190 |
| | | 11989 | *Threshold / cut-point* | Often\|Always | | 0.60 | 0.08 | 0.44 to 0.76 | 0.000 |
| **Say in government#** | **Proportional-odds regression** | 12012 | Childhood predictor | Yes | No | 0.37 | 0.09 | 0.20 to 0.54 | 0.000 |
| | | 12012 | Age | Per 1-year increase | | 0.00 | 0.00 | -0.01 to 0.00 | 0.308 |
| | | 12012 | Gender | Female | Male | 0.12 | 0.05 | 0.02 to 0.22 | 0.016 |
| | | 12012 | Education | Secondary | Elementary or less | -0.22 | 0.06 | -0.34 to -0.10 | 0.000 |
| | | 12012 | Education | Four years beyond HS | Elementary or less | -0.07 | 0.10 | -0.27 to 0.12 | 0.458 |
| | | 12012 | Residential setting | Small town | Rural | -0.04 | 0.08 | -0.19 to 0.11 | 0.600 |
| | | 12012 | Residential setting | Large city | Rural | -0.19 | 0.09 | -0.37 to -0.01 | 0.042 |
| | | 12012 | Residential setting | Suburb | Rural | -0.44 | 0.14 | -0.71 to -0.16 | 0.002 |
| | | 12012 | *Threshold / cut-point* | Disagree\|Unsure | | -1.75 | 0.11 | -1.97 to -1.53 | 0.000 |
| | | 12012 | *Threshold / cut-point* | Unsure\|Agree | | -1.35 | 0.11 | -1.57 to -1.14 | 0.000 |
| **Extent of suffering§** | **Proportional-odds regression** | 12015 | Childhood predictor | Yes | No | 0.51 | 0.07 | 0.39 to 0.64 | 0.000 |
| | | 12015 | Age | Per 1-year increase | | 0.02 | 0.00 | 0.02 to 0.02 | 0.000 |
| | | 12015 | Gender | Female | Male | 0.30 | 0.04 | 0.22 to 0.38 | 0.000 |
| | | 12015 | Education | Secondary | Elementary or less | -0.15 | 0.05 | -0.24 to -0.05 | 0.002 |
| | | 12015 | Education | Four years beyond HS | Elementary or less | -0.24 | 0.08 | -0.39 to -0.09 | 0.002 |
| | | 12015 | Residential setting | Small town | Rural | -0.01 | 0.05 | -0.12 to 0.09 | 0.787 |
| | | 12015 | Residential setting | Large city | Rural | -0.19 | 0.07 | -0.34 to -0.05 | 0.010 |
| | | 12015 | Residential setting | Suburb | Rural | 0.06 | 0.11 | -0.16 to 0.28 | 0.601 |
| | | 12015 | *Threshold / cut-point* | Not at all\|Not very much | | 0.39 | 0.09 | 0.23 to 0.56 | 0.000 |
| | | 12015 | *Threshold / cut-point* | Not very much\|Some | | 0.81 | 0.09 | 0.64 to 0.98 | 0.000 |
| | | 12015 | *Threshold / cut-point* | Some\|A lot | | 2.27 | 0.09 | 2.09 to 2.44 | 0.000 |
| **Distress from life-threatening experiences§** | **Proportional-odds regression** | 12008 | Childhood predictor | Yes | No | 0.71 | 0.07 | 0.58 to 0.84 | 0.000 |
| | | 12008 | Age | Per 1-year increase | | 0.00 | 0.00 | 0.00 to 0.01 | 0.006 |
| | | 12008 | Gender | Female | Male | 0.11 | 0.05 | 0.02 to 0.20 | 0.019 |
| | | 12008 | Education | Secondary | Elementary or less | -0.24 | 0.05 | -0.33 to -0.14 | 0.000 |
| | | 12008 | Education | Four years beyond HS | Elementary or less | -0.39 | 0.07 | -0.54 to -0.25 | 0.000 |
| | | 12008 | Residential setting | Small town | Rural | -0.20 | 0.05 | -0.30 to -0.10 | 0.000 |
| | | 12008 | Residential setting | Large city | Rural | -0.21 | 0.07 | -0.35 to -0.07 | 0.003 |
| | | 12008 | Residential setting | Suburb | Rural | -0.20 | 0.11 | -0.43 to 0.02 | 0.076 |
| | | 12008 | *Threshold / cut-point* | Not at all\|Not very much | | -0.17 | 0.08 | -0.32 to -0.01 | 0.037 |
| | | 12008 | *Threshold / cut-point* | Not very much\|Some | | 0.18 | 0.08 | 0.02 to 0.34 | 0.026 |
| | | 12008 | *Threshold / cut-point* | Some\|A lot | | 1.22 | 0.08 | 1.06 to 1.37 | 0.000 |
| **Trust in others§** | **Proportional-odds regression** | 11765 | Childhood predictor | Yes | No | 0.05 | 0.06 | -0.08 to 0.17 | 0.470 |

| Outcome | Model type | Analytic N | Variable | Level / comparison | Reference category | Coefficient | Standard error | 95% CI | p-value |
|---|---|---|---|---|---|---|---|---|---|
| | | 11765 | Age | Per 1-year increase | | 0.00 | 0.00 | 0.00 to 0.01 | 0.082 |
| | | 11765 | Gender | Female | Male | -0.03 | 0.04 | -0.11 to 0.05 | 0.417 |
| | | 11765 | Education | Secondary | Elementary or less | -0.06 | 0.05 | -0.15 to 0.04 | 0.221 |
| | | 11765 | Education | Four years beyond HS | Elementary or less | -0.08 | 0.07 | -0.22 to 0.07 | 0.292 |
| | | 11765 | Residential setting | Small town | Rural | 0.23 | 0.05 | 0.13 to 0.33 | 0.000 |
| | | 11765 | Residential setting | Large city | Rural | 0.02 | 0.07 | -0.11 to 0.16 | 0.754 |
| | | 11765 | Residential setting | Suburb | Rural | -0.20 | 0.14 | -0.48 to 0.08 | 0.163 |
| | | 11765 | *Threshold / cut-point* | None\|Not very many | | -2.16 | 0.09 | -2.35 to -1.98 | 0.000 |
| | | 11765 | *Threshold / cut-point* | Not very many\|Some | | -1.37 | 0.09 | -1.54 to -1.19 | 0.000 |
| | | 11765 | *Threshold / cut-point* | Some\|Most | | 0.82 | 0.09 | 0.65 to 0.99 | 0.000 |
| | | 11765 | *Threshold / cut-point* | Most\|All | | 1.82 | 0.09 | 1.64 to 1.99 | 0.000 |

**eTable 8.** Table of coefficients for adjusted associations between feeling like an outsider in one's family as predictor and 45 outcomes of interest and two flourishing composite indices.

Adjusted survey-weighted models use ANNUAL_WEIGHT_C1 with primary sampling unit (PSU) and strata and adjust for age, gender, education, and residential setting. Reference categories are shown in the table. Coefficients are on the original regression-coefficient scale: beta for continuous outcomes, log-odds for binary outcomes, and proportional-odds log coefficients for ordinal outcomes. Model intercepts are displayed for continuous and binary outcomes. Proportional-odds models do not have a single conventional intercept; their estimated ordinal cut-points/thresholds are displayed after the covariate coefficients, with standard errors (SEs), 95% CIs, and Wald p-values.

***Note:*** All results except p-values are rounded to the two decimals. P-values are rounded to three decimals. Values displayed as 0.000 indicate $P < 0.0005$ due to rounding, rather than an exact zero. Statistical significance was determined using unrounded P-values at the Bonferroni-adjusted threshold of $P < 0.0011$.

| Outcome | Model type | Analytic N | Variable | Level / comparison | Reference category | Coefficient | Standard error | 95% CI | p-value |
|---|---|---|---|---|---|---|---|---|---|
| **Belonging** | **Linear regression** | 12591 | *Intercept* | | | 8.66 | 0.12 | 8.43 to 8.88 | 0.000 |
| | | 12591 | Childhood predictor | Yes | No | 0.03 | 0.10 | -0.16 to 0.22 | 0.776 |
| | | 12591 | Age | Per 1-year increase | | -0.01 | 0.00 | -0.02 to -0.01 | 0.000 |
| | | 12591 | Gender | Female | Male | -0.18 | 0.06 | -0.30 to -0.06 | 0.003 |
| | | 12591 | Education | Secondary | Elementary or less | 0.20 | 0.07 | 0.06 to 0.35 | 0.006 |
| | | 12591 | Education | Four years beyond HS | Elementary or less | 0.31 | 0.10 | 0.11 to 0.52 | 0.002 |
| | | 12591 | Residential setting | Small town | Rural | 0.37 | 0.08 | 0.21 to 0.52 | 0.000 |
| | | 12591 | Residential setting | Large city | Rural | 0.42 | 0.10 | 0.21 to 0.62 | 0.000 |
| | | 12591 | Residential setting | Suburb | Rural | 0.03 | 0.18 | -0.32 to 0.38 | 0.862 |
| **Relationship contentment‡** | **Linear regression** | 12654 | *Intercept* | | | 8.11 | 0.13 | 7.87 to 8.36 | 0.000 |
| | | 12654 | Childhood predictor | Yes | No | -0.01 | 0.10 | -0.20 to 0.19 | 0.943 |
| | | 12654 | Age | Per 1-year increase | | -0.01 | 0.00 | -0.02 to -0.01 | 0.000 |
| | | 12654 | Gender | Female | Male | 0.38 | 0.07 | 0.25 to 0.52 | 0.000 |
| | | 12654 | Education | Secondary | Elementary or less | 0.16 | 0.07 | 0.01 to 0.30 | 0.034 |
| | | 12654 | Education | Four years beyond HS | Elementary or less | 0.30 | 0.12 | 0.07 to 0.53 | 0.011 |
| | | 12654 | Residential setting | Small town | Rural | 0.36 | 0.08 | 0.20 to 0.52 | 0.000 |
| | | 12654 | Residential setting | Large city | Rural | 0.07 | 0.10 | -0.12 to 0.26 | 0.466 |
| | | 12654 | Residential setting | Suburb | Rural | -0.28 | 0.17 | -0.62 to 0.06 | 0.104 |
| **Positive expectations** | **Linear regression** | 12616 | *Intercept* | | | 7.79 | 0.14 | 7.51 to 8.07 | 0.000 |
| | | 12616 | Childhood predictor | Yes | No | -0.15 | 0.10 | -0.34 to 0.03 | 0.111 |
| | | 12616 | Age | Per 1-year increase | | 0.00 | 0.00 | -0.01 to 0.00 | 0.263 |
| | | 12616 | Gender | Female | Male | 0.21 | 0.06 | 0.09 to 0.33 | 0.001 |
| | | 12616 | Education | Secondary | Elementary or less | 0.27 | 0.07 | 0.13 to 0.41 | 0.000 |
| | | 12616 | Education | Four years beyond HS | Elementary or less | 0.36 | 0.10 | 0.17 to 0.55 | 0.000 |
| | | 12616 | Residential setting | Small town | Rural | 0.45 | 0.09 | 0.28 to 0.62 | 0.000 |
| | | 12616 | Residential setting | Large city | Rural | 0.46 | 0.11 | 0.24 to 0.67 | 0.000 |
| | | 12616 | Residential setting | Suburb | Rural | 0.46 | 0.16 | 0.14 to 0.78 | 0.005 |
| **Worry about meeting normal monthly living expenses§§‡‡** | **Linear regression** | 12656 | *Intercept* | | | 5.79 | 0.19 | 5.41 to 6.16 | 0.000 |
| | | 12656 | Childhood predictor | Yes | No | -0.04 | 0.13 | -0.30 to 0.22 | 0.787 |
| | | 12656 | Age | Per 1-year increase | | 0.00 | 0.00 | -0.01 to 0.01 | 0.673 |

| Outcome | Model type | Analytic N | Variable | Level / comparison | Reference category | Coefficient | Standard error | 95% CI | p-value |
|---|---|---|---|---|---|---|---|---|---|
| | | 12656 | Gender | Female | Male | 0.36 | 0.09 | 0.18 to 0.54 | 0.000 |
| | | 12656 | Education | Secondary | Elementary or less | -0.46 | 0.11 | -0.67 to -0.25 | 0.000 |
| | | 12656 | Education | Four years beyond HS | Elementary or less | -0.66 | 0.15 | -0.95 to -0.36 | 0.000 |
| | | 12656 | Residential setting | Small town | Rural | 0.20 | 0.13 | -0.05 to 0.45 | 0.110 |
| | | 12656 | Residential setting | Large city | Rural | -0.23 | 0.15 | -0.52 to 0.07 | 0.141 |
| | | 12656 | Residential setting | Suburb | Rural | -0.24 | 0.25 | -0.72 to 0.25 | 0.343 |
| **Freedom** | **Linear regression** | 12630 | *Intercept* | | | 8.20 | 0.12 | 7.96 to 8.44 | 0.000 |
| | | 12630 | Childhood predictor | Yes | No | -0.06 | 0.09 | -0.24 to 0.12 | 0.488 |
| | | 12630 | Age | Per 1-year increase | | -0.01 | 0.00 | -0.01 to 0.00 | 0.002 |
| | | 12630 | Gender | Female | Male | -0.13 | 0.07 | -0.26 to 0.01 | 0.060 |
| | | 12630 | Education | Secondary | Elementary or less | 0.31 | 0.07 | 0.16 to 0.45 | 0.000 |
| | | 12630 | Education | Four years beyond HS | Elementary or less | 0.42 | 0.10 | 0.22 to 0.62 | 0.000 |
| | | 12630 | Residential setting | Small town | Rural | 0.35 | 0.07 | 0.20 to 0.49 | 0.000 |
| | | 12630 | Residential setting | Large city | Rural | 0.39 | 0.10 | 0.19 to 0.59 | 0.000 |
| | | 12630 | Residential setting | Suburb | Rural | 0.14 | 0.15 | -0.16 to 0.43 | 0.368 |
| **Give up some happiness now for greater happiness later‡** | **Linear regression** | 12595 | *Intercept* | | | 7.29 | 0.15 | 7.00 to 7.59 | 0.000 |
| | | 12595 | Childhood predictor | Yes | No | -0.26 | 0.11 | -0.48 to -0.04 | 0.019 |
| | | 12595 | Age | Per 1-year increase | | -0.01 | 0.00 | -0.01 to 0.00 | 0.000 |
| | | 12595 | Gender | Female | Male | 0.16 | 0.08 | 0.01 to 0.31 | 0.042 |
| | | 12595 | Education | Secondary | Elementary or less | 0.16 | 0.09 | -0.03 to 0.34 | 0.098 |
| | | 12595 | Education | Four years beyond HS | Elementary or less | 0.17 | 0.13 | -0.09 to 0.43 | 0.209 |
| | | 12595 | Residential setting | Small town | Rural | 0.39 | 0.10 | 0.20 to 0.58 | 0.000 |
| | | 12595 | Residential setting | Large city | Rural | 0.17 | 0.13 | -0.09 to 0.44 | 0.190 |
| | | 12595 | Residential setting | Suburb | Rural | 0.08 | 0.28 | -0.46 to 0.62 | 0.769 |
| **Have a long list of gratitude** | **Linear regression** | 12488 | *Intercept* | | | 7.42 | 0.15 | 7.13 to 7.71 | 0.000 |
| | | 12488 | Childhood predictor | Yes | No | 0.05 | 0.10 | -0.14 to 0.23 | 0.634 |
| | | 12488 | Age | Per 1-year increase | | -0.01 | 0.00 | -0.01 to 0.00 | 0.057 |
| | | 12488 | Gender | Female | Male | 0.21 | 0.07 | 0.07 to 0.34 | 0.003 |
| | | 12488 | Education | Secondary | Elementary or less | 0.29 | 0.08 | 0.13 to 0.44 | 0.000 |
| | | 12488 | Education | Four years beyond HS | Elementary or less | 0.39 | 0.11 | 0.17 to 0.61 | 0.001 |
| | | 12488 | Residential setting | Small town | Rural | 0.42 | 0.09 | 0.24 to 0.60 | 0.000 |
| | | 12488 | Residential setting | Large city | Rural | 0.43 | 0.13 | 0.17 to 0.69 | 0.001 |
| | | 12488 | Residential setting | Suburb | Rural | 0.51 | 0.17 | 0.18 to 0.84 | 0.003 |
| **Happiness‡** | **Linear regression** | 12654 | *Intercept* | | | 6.73 | 0.15 | 6.43 to 7.03 | 0.000 |
| | | 12654 | Childhood predictor | Yes | No | -0.34 | 0.13 | -0.58 to -0.09 | 0.008 |
| | | 12654 | Age | Per 1-year increase | | -0.02 | 0.00 | -0.02 to -0.01 | 0.000 |
| | | 12654 | Gender | Female | Male | 0.25 | 0.08 | 0.10 to 0.40 | 0.001 |
| | | 12654 | Education | Secondary | Elementary or less | 0.43 | 0.09 | 0.26 to 0.60 | 0.000 |
| | | 12654 | Education | Four years beyond HS | Elementary or less | 0.69 | 0.12 | 0.45 to 0.92 | 0.000 |
| | | 12654 | Residential setting | Small town | Rural | 0.37 | 0.10 | 0.17 to 0.57 | 0.000 |

| Outcome | Model type | Analytic N | Variable | Level / comparison | Reference category | Coefficient | Standard error | 95% CI | p-value |
|---|---|---|---|---|---|---|---|---|---|
| | | 12654 | Residential setting | Large city | Rural | 0.32 | 0.12 | 0.08 to 0.57 | 0.010 |
| | | 12654 | Residential setting | Suburb | Rural | -0.09 | 0.22 | -0.53 to 0.34 | 0.673 |
| **Remain hopeful for the future** | **Linear regression** | 12604 | *Intercept* | | | 8.16 | 0.12 | 7.92 to 8.40 | 0.000 |
| | | 12604 | Childhood predictor | Yes | No | -0.05 | 0.10 | -0.24 to 0.14 | 0.611 |
| | | 12604 | Age | Per 1-year increase | | -0.01 | 0.00 | -0.01 to 0.00 | 0.002 |
| | | 12604 | Gender | Female | Male | 0.01 | 0.05 | -0.10 to 0.12 | 0.833 |
| | | 12604 | Education | Secondary | Elementary or less | 0.39 | 0.06 | 0.26 to 0.51 | 0.000 |
| | | 12604 | Education | Four years beyond HS | Elementary or less | 0.61 | 0.09 | 0.43 to 0.79 | 0.000 |
| | | 12604 | Residential setting | Small town | Rural | 0.48 | 0.08 | 0.33 to 0.64 | 0.000 |
| | | 12604 | Residential setting | Large city | Rural | 0.47 | 0.10 | 0.27 to 0.67 | 0.000 |
| | | 12604 | Residential setting | Suburb | Rural | 0.42 | 0.18 | 0.08 to 0.77 | 0.016 |
| **Understand purpose in life‡** | **Linear regression** | 12581 | *Intercept* | | | 8.01 | 0.14 | 7.74 to 8.27 | 0.000 |
| | | 12581 | Childhood predictor | Yes | No | 0.13 | 0.10 | -0.06 to 0.32 | 0.173 |
| | | 12581 | Age | Per 1-year increase | | -0.01 | 0.00 | -0.02 to -0.01 | 0.000 |
| | | 12581 | Gender | Female | Male | 0.13 | 0.07 | -0.01 to 0.26 | 0.079 |
| | | 12581 | Education | Secondary | Elementary or less | 0.43 | 0.08 | 0.28 to 0.58 | 0.000 |
| | | 12581 | Education | Four years beyond HS | Elementary or less | 0.76 | 0.10 | 0.57 to 0.95 | 0.000 |
| | | 12581 | Residential setting | Small town | Rural | 0.42 | 0.09 | 0.24 to 0.59 | 0.000 |
| | | 12581 | Residential setting | Large city | Rural | 0.42 | 0.12 | 0.20 to 0.65 | 0.000 |
| | | 12581 | Residential setting | Suburb | Rural | 0.45 | 0.15 | 0.15 to 0.74 | 0.003 |
| **Life satisfaction‡** | **Linear regression** | 12634 | *Intercept* | | | 7.14 | 0.15 | 6.85 to 7.43 | 0.000 |
| | | 12634 | Childhood predictor | Yes | No | -0.19 | 0.11 | -0.41 to 0.04 | 0.100 |
| | | 12634 | Age | Per 1-year increase | | -0.01 | 0.00 | -0.02 to -0.01 | 0.000 |
| | | 12634 | Gender | Female | Male | 0.31 | 0.08 | 0.16 to 0.46 | 0.000 |
| | | 12634 | Education | Secondary | Elementary or less | 0.31 | 0.09 | 0.14 to 0.49 | 0.001 |
| | | 12634 | Education | Four years beyond HS | Elementary or less | 0.50 | 0.12 | 0.27 to 0.73 | 0.000 |
| | | 12634 | Residential setting | Small town | Rural | 0.27 | 0.09 | 0.09 to 0.45 | 0.003 |
| | | 12634 | Residential setting | Large city | Rural | 0.20 | 0.12 | -0.04 to 0.44 | 0.097 |
| | | 12634 | Residential setting | Suburb | Rural | -0.20 | 0.21 | -0.62 to 0.21 | 0.337 |
| **Loneliness§** | **Linear regression** | 12654 | *Intercept* | | | 4.18 | 0.17 | 3.85 to 4.51 | 0.000 |
| | | 12654 | Childhood predictor | Yes | No | 0.39 | 0.12 | 0.15 to 0.63 | 0.002 |
| | | 12654 | Age | Per 1-year increase | | 0.00 | 0.00 | -0.01 to 0.01 | 0.796 |
| | | 12654 | Gender | Female | Male | -0.15 | 0.09 | -0.31 to 0.02 | 0.090 |
| | | 12654 | Education | Secondary | Elementary or less | -0.32 | 0.10 | -0.51 to -0.13 | 0.001 |
| | | 12654 | Education | Four years beyond HS | Elementary or less | -0.66 | 0.14 | -0.94 to -0.37 | 0.000 |
| | | 12654 | Residential setting | Small town | Rural | -0.36 | 0.10 | -0.56 to -0.16 | 0.000 |
| | | 12654 | Residential setting | Large city | Rural | -0.49 | 0.14 | -0.76 to -0.22 | 0.000 |
| | | 12654 | Residential setting | Suburb | Rural | -0.14 | 0.24 | -0.60 to 0.32 | 0.555 |
| **Mental health‡** | **Linear regression** | 12659 | *Intercept* | | | 8.10 | 0.14 | 7.82 to 8.38 | 0.000 |
| | | 12659 | Childhood predictor | Yes | No | -0.23 | 0.12 | -0.46 to 0.00 | 0.049 |

| Outcome | Model type | Analytic N | Variable | Level / comparison | Reference category | Coefficient | Standard error | 95% CI | p-value |
|---|---|---|---|---|---|---|---|---|---|
| | | 12659 | Age | Per 1-year increase | | -0.02 | 0.00 | -0.03 to -0.02 | 0.000 |
| | | 12659 | Gender | Female | Male | -0.13 | 0.07 | -0.26 to 0.01 | 0.066 |
| | | 12659 | Education | Secondary | Elementary or less | 0.41 | 0.08 | 0.26 to 0.56 | 0.000 |
| | | 12659 | Education | Four years beyond HS | Elementary or less | 0.54 | 0.11 | 0.32 to 0.76 | 0.000 |
| | | 12659 | Residential setting | Small town | Rural | 0.39 | 0.09 | 0.21 to 0.57 | 0.000 |
| | | 12659 | Residential setting | Large city | Rural | 0.45 | 0.12 | 0.21 to 0.68 | 0.000 |
| | | 12659 | Residential setting | Suburb | Rural | 0.15 | 0.19 | -0.21 to 0.52 | 0.416 |
| **Perceived support from others** | **Linear regression** | 12642 | *Intercept* | | | 6.73 | 0.16 | 6.42 to 7.04 | 0.000 |
| | | 12642 | Childhood predictor | Yes | No | -0.49 | 0.12 | -0.73 to -0.24 | 0.000 |
| | | 12642 | Age | Per 1-year increase | | -0.01 | 0.00 | -0.02 to -0.01 | 0.000 |
| | | 12642 | Gender | Female | Male | 0.53 | 0.09 | 0.35 to 0.70 | 0.000 |
| | | 12642 | Education | Secondary | Elementary or less | 0.19 | 0.10 | 0.00 to 0.38 | 0.055 |
| | | 12642 | Education | Four years beyond HS | Elementary or less | 0.13 | 0.14 | -0.14 to 0.41 | 0.333 |
| | | 12642 | Residential setting | Small town | Rural | 0.16 | 0.11 | -0.05 to 0.37 | 0.124 |
| | | 12642 | Residential setting | Large city | Rural | -0.51 | 0.14 | -0.79 to -0.24 | 0.000 |
| | | 12642 | Residential setting | Suburb | Rural | -0.29 | 0.20 | -0.67 to 0.10 | 0.147 |
| **Physical health‡** | **Linear regression** | 12661 | *Intercept* | | | 8.44 | 0.14 | 8.16 to 8.72 | 0.000 |
| | | 12661 | Childhood predictor | Yes | No | -0.22 | 0.11 | -0.45 to 0.00 | 0.055 |
| | | 12661 | Age | Per 1-year increase | | -0.04 | 0.00 | -0.05 to -0.04 | 0.000 |
| | | 12661 | Gender | Female | Male | -0.34 | 0.08 | -0.48 to -0.19 | 0.000 |
| | | 12661 | Education | Secondary | Elementary or less | 0.37 | 0.08 | 0.22 to 0.52 | 0.000 |
| | | 12661 | Education | Four years beyond HS | Elementary or less | 0.67 | 0.11 | 0.46 to 0.88 | 0.000 |
| | | 12661 | Residential setting | Small town | Rural | 0.47 | 0.09 | 0.29 to 0.65 | 0.000 |
| | | 12661 | Residential setting | Large city | Rural | 0.63 | 0.11 | 0.43 to 0.84 | 0.000 |
| | | 12661 | Residential setting | Suburb | Rural | 0.16 | 0.17 | -0.19 to 0.50 | 0.373 |
| **Act to promote good‡** | **Linear regression** | 12616 | *Intercept* | | | 8.05 | 0.11 | 7.83 to 8.26 | 0.000 |
| | | 12616 | Childhood predictor | Yes | No | 0.05 | 0.09 | -0.13 to 0.23 | 0.579 |
| | | 12616 | Age | Per 1-year increase | | -0.01 | 0.00 | -0.01 to 0.00 | 0.001 |
| | | 12616 | Gender | Female | Male | 0.12 | 0.06 | 0.01 to 0.24 | 0.040 |
| | | 12616 | Education | Secondary | Elementary or less | 0.38 | 0.06 | 0.26 to 0.50 | 0.000 |
| | | 12616 | Education | Four years beyond HS | Elementary or less | 0.55 | 0.08 | 0.40 to 0.71 | 0.000 |
| | | 12616 | Residential setting | Small town | Rural | 0.60 | 0.07 | 0.46 to 0.75 | 0.000 |
| | | 12616 | Residential setting | Large city | Rural | 0.64 | 0.10 | 0.45 to 0.83 | 0.000 |
| | | 12616 | Residential setting | Suburb | Rural | 0.41 | 0.16 | 0.09 to 0.74 | 0.012 |
| **Relationship satisfaction‡** | **Linear regression** | 12622 | *Intercept* | | | 7.87 | 0.13 | 7.61 to 8.12 | 0.000 |
| | | 12622 | Childhood predictor | Yes | No | -0.09 | 0.11 | -0.31 to 0.12 | 0.377 |
| | | 12622 | Age | Per 1-year increase | | -0.01 | 0.00 | -0.01 to 0.00 | 0.001 |
| | | 12622 | Gender | Female | Male | 0.33 | 0.07 | 0.20 to 0.47 | 0.000 |
| | | 12622 | Education | Secondary | Elementary or less | -0.03 | 0.08 | -0.18 to 0.13 | 0.732 |
| | | 12622 | Education | Four years beyond HS | Elementary or less | 0.05 | 0.11 | -0.16 to 0.27 | 0.633 |

| Outcome | Model type | Analytic N | Variable | Level / comparison | Reference category | Coefficient | Standard error | 95% CI | p-value |
|---|---|---|---|---|---|---|---|---|---|
| | | 12622 | Residential setting | Small town | Rural | 0.43 | 0.09 | 0.26 to 0.60 | 0.000 |
| | | 12622 | Residential setting | Large city | Rural | 0.33 | 0.11 | 0.12 to 0.53 | 0.002 |
| | | 12622 | Residential setting | Suburb | Rural | 0.04 | 0.16 | -0.26 to 0.35 | 0.788 |
| **Show love often** | **Linear regression** | 12647 | *Intercept* | | | 8.27 | 0.13 | 8.02 to 8.51 | 0.000 |
| | | 12647 | Childhood predictor | Yes | No | 0.03 | 0.10 | -0.16 to 0.22 | 0.771 |
| | | 12647 | Age | Per 1-year increase | | -0.01 | 0.00 | -0.01 to 0.00 | 0.002 |
| | | 12647 | Gender | Female | Male | 0.22 | 0.07 | 0.09 to 0.35 | 0.001 |
| | | 12647 | Education | Secondary | Elementary or less | 0.16 | 0.07 | 0.02 to 0.30 | 0.022 |
| | | 12647 | Education | Four years beyond HS | Elementary or less | 0.04 | 0.11 | -0.17 to 0.26 | 0.682 |
| | | 12647 | Residential setting | Small town | Rural | 0.20 | 0.08 | 0.05 to 0.35 | 0.010 |
| | | 12647 | Residential setting | Large city | Rural | 0.02 | 0.11 | -0.20 to 0.24 | 0.833 |
| | | 12647 | Residential setting | Suburb | Rural | -0.14 | 0.20 | -0.53 to 0.25 | 0.481 |
| **Life evaluation five years from now** | **Linear regression** | 11524 | *Intercept* | | | 7.71 | 0.14 | 7.43 to 7.99 | 0.000 |
| | | 11524 | Childhood predictor | Yes | No | -0.13 | 0.12 | -0.36 to 0.10 | 0.261 |
| | | 11524 | Age | Per 1-year increase | | -0.02 | 0.00 | -0.03 to -0.02 | 0.000 |
| | | 11524 | Gender | Female | Male | 0.19 | 0.07 | 0.05 to 0.33 | 0.007 |
| | | 11524 | Education | Secondary | Elementary or less | 0.29 | 0.08 | 0.13 to 0.44 | 0.000 |
| | | 11524 | Education | Four years beyond HS | Elementary or less | 0.58 | 0.12 | 0.36 to 0.81 | 0.000 |
| | | 11524 | Residential setting | Small town | Rural | 0.44 | 0.09 | 0.27 to 0.62 | 0.000 |
| | | 11524 | Residential setting | Large city | Rural | 0.50 | 0.11 | 0.28 to 0.72 | 0.000 |
| | | 11524 | Residential setting | Suburb | Rural | 0.35 | 0.23 | -0.10 to 0.80 | 0.125 |
| **Life evaluation today** | **Linear regression** | 12555 | *Intercept* | | | 5.65 | 0.17 | 5.32 to 5.98 | 0.000 |
| | | 12555 | Childhood predictor | Yes | No | -0.40 | 0.12 | -0.65 to -0.16 | 0.001 |
| | | 12555 | Age | Per 1-year increase | | -0.01 | 0.00 | -0.02 to 0.00 | 0.003 |
| | | 12555 | Gender | Female | Male | 0.49 | 0.08 | 0.33 to 0.65 | 0.000 |
| | | 12555 | Education | Secondary | Elementary or less | 0.22 | 0.08 | 0.06 to 0.39 | 0.008 |
| | | 12555 | Education | Four years beyond HS | Elementary or less | 0.19 | 0.14 | -0.09 to 0.47 | 0.185 |
| | | 12555 | Residential setting | Small town | Rural | 0.22 | 0.10 | 0.02 to 0.42 | 0.033 |
| | | 12555 | Residential setting | Large city | Rural | 0.08 | 0.13 | -0.19 to 0.34 | 0.574 |
| | | 12555 | Residential setting | Suburb | Rural | -0.08 | 0.21 | -0.50 to 0.33 | 0.699 |
| **Worry about safety, food, or housing§§‡‡** | **Linear regression** | 12670 | *Intercept* | | | 6.19 | 0.19 | 5.81 to 6.57 | 0.000 |
| | | 12670 | Childhood predictor | Yes | No | 0.29 | 0.13 | 0.03 to 0.54 | 0.027 |
| | | 12670 | Age | Per 1-year increase | | -0.01 | 0.00 | -0.02 to 0.00 | 0.011 |
| | | 12670 | Gender | Female | Male | 0.34 | 0.09 | 0.16 to 0.51 | 0.000 |
| | | 12670 | Education | Secondary | Elementary or less | -0.55 | 0.10 | -0.75 to -0.34 | 0.000 |
| | | 12670 | Education | Four years beyond HS | Elementary or less | -0.80 | 0.17 | -1.12 to -0.47 | 0.000 |
| | | 12670 | Residential setting | Small town | Rural | 0.37 | 0.13 | 0.11 to 0.62 | 0.005 |
| | | 12670 | Residential setting | Large city | Rural | -0.10 | 0.18 | -0.45 to 0.25 | 0.574 |
| | | 12670 | Residential setting | Suburb | Rural | 0.17 | 0.26 | -0.33 to 0.68 | 0.497 |

| Outcome | Model type | Analytic N | Variable | Level / comparison | Reference category | Coefficient | Standard error | 95% CI | p-value |
|---|---|---|---|---|---|---|---|---|---|
| **Life worthwhile‡** | **Linear regression** | 12552 | *Intercept* | | | 6.67 | 0.15 | 6.39 to 6.95 | 0.000 |
| | | 12552 | Childhood predictor | Yes | No | -0.13 | 0.11 | -0.35 to 0.09 | 0.257 |
| | | 12552 | Age | Per 1-year increase | | -0.01 | 0.00 | -0.01 to 0.00 | 0.001 |
| | | 12552 | Gender | Female | Male | 0.45 | 0.07 | 0.30 to 0.59 | 0.000 |
| | | 12552 | Education | Secondary | Elementary or less | 0.46 | 0.08 | 0.31 to 0.61 | 0.000 |
| | | 12552 | Education | Four years beyond HS | Elementary or less | 0.62 | 0.12 | 0.38 to 0.86 | 0.000 |
| | | 12552 | Residential setting | Small town | Rural | 0.43 | 0.10 | 0.23 to 0.63 | 0.000 |
| | | 12552 | Residential setting | Large city | Rural | 0.42 | 0.13 | 0.17 to 0.67 | 0.001 |
| | | 12552 | Residential setting | Suburb | Rural | 0.34 | 0.21 | -0.07 to 0.75 | 0.102 |
| **Flourishing (with financial indicators)** | **Linear regression** | 12203 | *Intercept* | | | 7.01 | 0.08 | 6.85 to 7.17 | 0.000 |
| | | 12203 | Childhood predictor | Yes | No | -0.13 | 0.07 | -0.26 to 0.01 | 0.063 |
| | | 12203 | Age | Per 1-year increase | | -0.01 | 0.00 | -0.02 to -0.01 | 0.000 |
| | | 12203 | Gender | Female | Male | 0.10 | 0.04 | 0.01 to 0.18 | 0.021 |
| | | 12203 | Education | Secondary | Elementary or less | 0.33 | 0.04 | 0.25 to 0.42 | 0.000 |
| | | 12203 | Education | Four years beyond HS | Elementary or less | 0.52 | 0.07 | 0.39 to 0.65 | 0.000 |
| | | 12203 | Residential setting | Small town | Rural | 0.31 | 0.06 | 0.20 to 0.42 | 0.000 |
| | | 12203 | Residential setting | Large city | Rural | 0.35 | 0.07 | 0.21 to 0.48 | 0.000 |
| | | 12203 | Residential setting | Suburb | Rural | 0.11 | 0.13 | -0.15 to 0.37 | 0.414 |
| **Flourishing (without financial indicators)** | **Linear regression** | 12232 | *Intercept* | | | 7.63 | 0.09 | 7.46 to 7.80 | 0.000 |
| | | 12232 | Childhood predictor | Yes | No | -0.13 | 0.07 | -0.27 to 0.01 | 0.072 |
| | | 12232 | Age | Per 1-year increase | | -0.02 | 0.00 | -0.02 to -0.01 | 0.000 |
| | | 12232 | Gender | Female | Male | 0.18 | 0.05 | 0.09 to 0.27 | 0.000 |
| | | 12232 | Education | Secondary | Elementary or less | 0.30 | 0.05 | 0.21 to 0.40 | 0.000 |
| | | 12232 | Education | Four years beyond HS | Elementary or less | 0.48 | 0.07 | 0.35 to 0.62 | 0.000 |
| | | 12232 | Residential setting | Small town | Rural | 0.43 | 0.06 | 0.31 to 0.55 | 0.000 |
| | | 12232 | Residential setting | Large city | Rural | 0.39 | 0.08 | 0.24 to 0.54 | 0.000 |
| | | 12232 | Residential setting | Suburb | Rural | 0.12 | 0.13 | -0.14 to 0.38 | 0.359 |
| **Feel close to someone** | **Logistic regression** | 12666 | *Intercept* | | | 1.61 | 0.14 | 1.33 to 1.88 | 0.000 |
| | | 12666 | Childhood predictor | Yes | No | 0.46 | 0.10 | 0.26 to 0.66 | 0.000 |
| | | 12666 | Age | Per 1-year increase | | -0.01 | 0.00 | -0.01 to 0.00 | 0.005 |
| | | 12666 | Gender | Female | Male | 0.03 | 0.06 | -0.10 to 0.15 | 0.646 |
| | | 12666 | Education | Secondary | Elementary or less | 0.18 | 0.08 | 0.02 to 0.34 | 0.026 |
| | | 12666 | Education | Four years beyond HS | Elementary or less | 0.17 | 0.13 | -0.08 to 0.43 | 0.181 |
| | | 12666 | Residential setting | Small town | Rural | 0.11 | 0.11 | -0.11 to 0.32 | 0.335 |
| | | 12666 | Residential setting | Large city | Rural | -0.09 | 0.13 | -0.35 to 0.16 | 0.468 |
| | | 12666 | Residential setting | Suburb | Rural | -0.15 | 0.19 | -0.52 to 0.23 | 0.447 |
| **Donated to charity** | **Logistic regression** | 12631 | *Intercept* | | | -0.46 | 0.13 | -0.71 to -0.21 | 0.000 |
| | | 12631 | Childhood predictor | Yes | No | 0.59 | 0.09 | 0.42 to 0.76 | 0.000 |
| | | 12631 | Age | Per 1-year increase | | 0.00 | 0.00 | 0.00 to 0.00 | 0.590 |

| Outcome | Model type | Analytic N | Variable | Level / comparison | Reference category | Coefficient | Standard error | 95% CI | p-value |
|---|---|---|---|---|---|---|---|---|---|
| | | 12631 | Gender | Female | Male | -0.01 | 0.05 | -0.11 to 0.08 | 0.766 |
| | | 12631 | Education | Secondary | Elementary or less | -0.06 | 0.06 | -0.18 to 0.05 | 0.295 |
| | | 12631 | Education | Four years beyond HS | Elementary or less | -0.13 | 0.10 | -0.33 to 0.06 | 0.181 |
| | | 12631 | Residential setting | Small town | Rural | -0.05 | 0.09 | -0.23 to 0.13 | 0.574 |
| | | 12631 | Residential setting | Large city | Rural | -0.39 | 0.11 | -0.62 to -0.17 | 0.000 |
| | | 12631 | Residential setting | Suburb | Rural | -0.16 | 0.19 | -0.52 to 0.21 | 0.401 |
| **Have any health problems** | **Logistic regression** | 12589 | *Intercept* | | | -2.06 | 0.10 | -2.26 to -1.86 | 0.000 |
| | | 12589 | Childhood predictor | Yes | No | 0.14 | 0.07 | 0.00 to 0.28 | 0.049 |
| | | 12589 | Age | Per 1-year increase | | 0.03 | 0.00 | 0.02 to 0.03 | 0.000 |
| | | 12589 | Gender | Female | Male | 0.23 | 0.05 | 0.13 to 0.33 | 0.000 |
| | | 12589 | Education | Secondary | Elementary or less | -0.14 | 0.06 | -0.25 to -0.03 | 0.016 |
| | | 12589 | Education | Four years beyond HS | Elementary or less | -0.27 | 0.09 | -0.46 to -0.09 | 0.004 |
| | | 12589 | Residential setting | Small town | Rural | 0.12 | 0.06 | 0.00 to 0.25 | 0.053 |
| | | 12589 | Residential setting | Large city | Rural | -0.14 | 0.09 | -0.31 to 0.03 | 0.115 |
| | | 12589 | Residential setting | Suburb | Rural | 0.07 | 0.14 | -0.20 to 0.34 | 0.605 |
| **Helped a stranger** | **Logistic regression** | 12536 | *Intercept* | | | 0.60 | 0.12 | 0.37 to 0.83 | 0.000 |
| | | 12536 | Childhood predictor | Yes | No | 0.40 | 0.08 | 0.24 to 0.55 | 0.000 |
| | | 12536 | Age | Per 1-year increase | | -0.01 | 0.00 | -0.01 to 0.00 | 0.000 |
| | | 12536 | Gender | Female | Male | -0.31 | 0.05 | -0.40 to -0.22 | 0.000 |
| | | 12536 | Education | Secondary | Elementary or less | -0.06 | 0.06 | -0.17 to 0.06 | 0.336 |
| | | 12536 | Education | Four years beyond HS | Elementary or less | -0.08 | 0.09 | -0.25 to 0.10 | 0.377 |
| | | 12536 | Residential setting | Small town | Rural | 0.11 | 0.09 | -0.06 to 0.28 | 0.201 |
| | | 12536 | Residential setting | Large city | Rural | -0.14 | 0.10 | -0.34 to 0.06 | 0.168 |
| | | 12536 | Residential setting | Suburb | Rural | 0.07 | 0.18 | -0.28 to 0.42 | 0.687 |
| **Volunteered at an organization** | **Logistic regression** | 12612 | *Intercept* | | | -0.40 | 0.14 | -0.68 to -0.13 | 0.004 |
| | | 12612 | Childhood predictor | Yes | No | 0.42 | 0.09 | 0.25 to 0.60 | 0.000 |
| | | 12612 | Age | Per 1-year increase | | -0.01 | 0.00 | -0.01 to 0.00 | 0.001 |
| | | 12612 | Gender | Female | Male | -0.27 | 0.05 | -0.37 to -0.17 | 0.000 |
| | | 12612 | Education | Secondary | Elementary or less | -0.13 | 0.06 | -0.25 to 0.00 | 0.049 |
| | | 12612 | Education | Four years beyond HS | Elementary or less | 0.04 | 0.10 | -0.15 to 0.24 | 0.657 |
| | | 12612 | Residential setting | Small town | Rural | -0.09 | 0.11 | -0.30 to 0.12 | 0.403 |
| | | 12612 | Residential setting | Large city | Rural | -0.54 | 0.13 | -0.79 to -0.29 | 0.000 |
| | | 12612 | Residential setting | Suburb | Rural | -0.34 | 0.16 | -0.65 to -0.03 | 0.033 |
| **Cigarette smoking** | **Logistic regression** | 12172 | *Intercept* | | | -2.01 | 0.16 | -2.33 to -1.69 | 0.000 |
| | | 12172 | Childhood predictor | Yes | No | 0.13 | 0.11 | -0.09 to 0.36 | 0.253 |
| | | 12172 | Age | Per 1-year increase | | 0.01 | 0.00 | 0.00 to 0.01 | 0.014 |
| | | 12172 | Gender | Female | Male | -2.85 | 0.19 | -3.21 to -2.48 | 0.000 |
| | | 12172 | Education | Secondary | Elementary or less | -0.37 | 0.10 | -0.56 to -0.18 | 0.000 |
| | | 12172 | Education | Four years beyond HS | Elementary or less | -0.60 | 0.17 | -0.93 to -0.27 | 0.000 |
| | | 12172 | Residential setting | Small town | Rural | -0.07 | 0.10 | -0.27 to 0.13 | 0.491 |

| Outcome | Model type | Analytic N | Variable | Level / comparison | Reference category | Coefficient | Standard error | 95% CI | p-value |
|---|---|---|---|---|---|---|---|---|---|
| | | 12172 | Residential setting | Large city | Rural | 0.14 | 0.14 | -0.14 to 0.41 | 0.338 |
| | | 12172 | Residential setting | Suburb | Rural | 0.78 | 0.20 | 0.39 to 1.18 | 0.000 |
| **Alcohol use** | **Logistic regression** | 12158 | *Intercept* | | | -1.88 | 0.15 | -2.17 to -1.59 | 0.000 |
| | | 12158 | Childhood predictor | Yes | No | 0.12 | 0.12 | -0.11 to 0.35 | 0.316 |
| | | 12158 | Age | Per 1-year increase | | 0.00 | 0.00 | -0.01 to 0.00 | 0.828 |
| | | 12158 | Gender | Female | Male | -2.75 | 0.17 | -3.08 to -2.42 | 0.000 |
| | | 12158 | Education | Secondary | Elementary or less | -0.24 | 0.10 | -0.44 to -0.05 | 0.013 |
| | | 12158 | Education | Four years beyond HS | Elementary or less | -0.53 | 0.16 | -0.85 to -0.21 | 0.001 |
| | | 12158 | Residential setting | Small town | Rural | 0.21 | 0.10 | 0.01 to 0.42 | 0.045 |
| | | 12158 | Residential setting | Large city | Rural | 0.43 | 0.14 | 0.15 to 0.70 | 0.002 |
| | | 12158 | Residential setting | Suburb | Rural | 0.64 | 0.22 | 0.22 to 1.06 | 0.003 |
| **Government approval§** | **Proportional-odds regression** | 12449 | Childhood predictor | Yes | No | 0.11 | 0.07 | -0.02 to 0.25 | 0.094 |
| | | 12449 | Age | Per 1-year increase | | 0.00 | 0.00 | -0.01 to 0.00 | 0.032 |
| | | 12449 | Gender | Female | Male | 0.05 | 0.04 | -0.04 to 0.13 | 0.262 |
| | | 12449 | Education | Secondary | Elementary or less | -0.24 | 0.05 | -0.34 to -0.13 | 0.000 |
| | | 12449 | Education | Four years beyond HS | Elementary or less | -0.31 | 0.08 | -0.48 to -0.15 | 0.000 |
| | | 12449 | Residential setting | Small town | Rural | 0.10 | 0.07 | -0.02 to 0.23 | 0.111 |
| | | 12449 | Residential setting | Large city | Rural | -0.06 | 0.08 | -0.21 to 0.09 | 0.421 |
| | | 12449 | Residential setting | Suburb | Rural | -0.42 | 0.15 | -0.72 to -0.12 | 0.007 |
| | | 12449 | *Threshold / cut-point* | Strongly disapprove\|Somewhat disapprove | | -2.90 | 0.10 | -3.09 to -2.70 | 0.000 |
| | | 12449 | *Threshold / cut-point* | Somewhat disapprove\|Neither | | -2.23 | 0.09 | -2.41 to -2.05 | 0.000 |
| | | 12449 | *Threshold / cut-point* | Neither\|Somewhat approve | | -1.87 | 0.09 | -2.05 to -1.69 | 0.000 |
| | | 12449 | *Threshold / cut-point* | Somewhat approve\|Strongly approve | | -0.42 | 0.09 | -0.59 to -0.25 | 0.000 |
| **Feeling capable§** | **Proportional-odds regression** | 12613 | Childhood predictor | Yes | No | 0.01 | 0.06 | -0.11 to 0.12 | 0.889 |
| | | 12613 | Age | Per 1-year increase | | 0.00 | 0.00 | 0.00 to 0.01 | 0.188 |
| | | 12613 | Gender | Female | Male | -0.07 | 0.04 | -0.15 to 0.02 | 0.122 |
| | | 12613 | Education | Secondary | Elementary or less | 0.16 | 0.05 | 0.07 to 0.26 | 0.001 |
| | | 12613 | Education | Four years beyond HS | Elementary or less | 0.10 | 0.07 | -0.04 to 0.25 | 0.143 |
| | | 12613 | Residential setting | Small town | Rural | 0.17 | 0.05 | 0.07 to 0.26 | 0.001 |
| | | 12613 | Residential setting | Large city | Rural | 0.20 | 0.07 | 0.05 to 0.34 | 0.009 |
| | | 12613 | Residential setting | Suburb | Rural | 0.25 | 0.09 | 0.07 to 0.42 | 0.005 |
| | | 12613 | *Threshold / cut-point* | Never\|Rarely | | -2.34 | 0.09 | -2.51 to -2.16 | 0.000 |
| | | 12613 | *Threshold / cut-point* | Rarely\|Often | | -0.62 | 0.08 | -0.79 to -0.46 | 0.000 |
| | | 12613 | *Threshold / cut-point* | Often\|Always | | 0.04 | 0.08 | -0.12 to 0.21 | 0.616 |
| **Unable to control worry§** | **Proportional-odds regression** | 12566 | Childhood predictor | Yes | No | 0.21 | 0.06 | 0.10 to 0.33 | 0.000 |
| | | 12566 | Age | Per 1-year increase | | 0.01 | 0.00 | 0.00 to 0.01 | 0.001 |

| Outcome | Model type | Analytic N | Variable | Level / comparison | Reference category | Coefficient | Standard error | 95% CI | p-value |
|---|---|---|---|---|---|---|---|---|---|
| | | 12566 | Gender | Female | Male | 0.17 | 0.04 | 0.09 to 0.25 | 0.000 |
| | | 12566 | Education | Secondary | Elementary or less | -0.27 | 0.05 | -0.36 to -0.18 | 0.000 |
| | | 12566 | Education | Four years beyond HS | Elementary or less | -0.43 | 0.07 | -0.58 to -0.29 | 0.000 |
| | | 12566 | Residential setting | Small town | Rural | 0.05 | 0.05 | -0.04 to 0.14 | 0.298 |
| | | 12566 | Residential setting | Large city | Rural | 0.00 | 0.07 | -0.13 to 0.14 | 0.953 |
| | | 12566 | Residential setting | Suburb | Rural | -0.09 | 0.10 | -0.28 to 0.10 | 0.343 |
| | | 12566 | *Threshold / cut-point* | Not at all\|Several days | | 0.11 | 0.08 | -0.04 to 0.27 | 0.153 |
| | | 12566 | *Threshold / cut-point* | Several days\|More than half the days | | 1.02 | 0.08 | 0.87 to 1.17 | 0.000 |
| | | 12566 | *Threshold / cut-point* | More than half the days\|Nearly every day | | 1.87 | 0.08 | 1.71 to 2.03 | 0.000 |
| **Depressive symptoms§** | **Proportional-odds regression** | 12583 | Childhood predictor | Yes | No | 0.08 | 0.06 | -0.04 to 0.20 | 0.170 |
| | | 12583 | Age | Per 1-year increase | | 0.00 | 0.00 | 0.00 to 0.00 | 0.170 |
| | | 12583 | Gender | Female | Male | 0.09 | 0.04 | 0.01 to 0.17 | 0.020 |
| | | 12583 | Education | Secondary | Elementary or less | -0.20 | 0.05 | -0.29 to -0.11 | 0.000 |
| | | 12583 | Education | Four years beyond HS | Elementary or less | -0.41 | 0.07 | -0.55 to -0.28 | 0.000 |
| | | 12583 | Residential setting | Small town | Rural | -0.01 | 0.05 | -0.11 to 0.10 | 0.909 |
| | | 12583 | Residential setting | Large city | Rural | -0.09 | 0.07 | -0.22 to 0.05 | 0.217 |
| | | 12583 | Residential setting | Suburb | Rural | -0.18 | 0.11 | -0.40 to 0.04 | 0.102 |
| | | 12583 | *Threshold / cut-point* | Not at all\|Several days | | -0.24 | 0.08 | -0.40 to -0.09 | 0.002 |
| | | 12583 | *Threshold / cut-point* | Several days\|More than half the days | | 0.83 | 0.08 | 0.67 to 0.98 | 0.000 |
| | | 12583 | *Threshold / cut-point* | More than half the days\|Nearly every day | | 1.83 | 0.08 | 1.67 to 2.00 | 0.000 |
| **Perceived discrimination§** | **Proportional-odds regression** | 12516 | Childhood predictor | Yes | No | -0.03 | 0.06 | -0.15 to 0.08 | 0.563 |
| | | 12516 | Age | Per 1-year increase | | 0.00 | 0.00 | -0.01 to 0.00 | 0.042 |
| | | 12516 | Gender | Female | Male | -0.14 | 0.04 | -0.22 to -0.05 | 0.002 |
| | | 12516 | Education | Secondary | Elementary or less | -0.24 | 0.05 | -0.34 to -0.14 | 0.000 |
| | | 12516 | Education | Four years beyond HS | Elementary or less | -0.30 | 0.09 | -0.47 to -0.13 | 0.001 |
| | | 12516 | Residential setting | Small town | Rural | -0.20 | 0.06 | -0.32 to -0.09 | 0.000 |
| | | 12516 | Residential setting | Large city | Rural | -0.24 | 0.07 | -0.38 to -0.10 | 0.001 |
| | | 12516 | Residential setting | Suburb | Rural | -0.34 | 0.10 | -0.52 to -0.15 | 0.000 |
| | | 12516 | *Threshold / cut-point* | Never\|Rarely | | -0.14 | 0.09 | -0.32 to 0.03 | 0.099 |
| | | 12516 | *Threshold / cut-point* | Rarely\|Often | | 0.86 | 0.09 | 0.68 to 1.03 | 0.000 |
| | | 12516 | *Threshold / cut-point* | Often\|Always | | 1.48 | 0.09 | 1.30 to 1.65 | 0.000 |
| **Feeling nervous or anxious§** | **Proportional-odds regression** | 12614 | Childhood predictor | Yes | No | 0.29 | 0.06 | 0.17 to 0.40 | 0.000 |
| | | 12614 | Age | Per 1-year increase | | 0.01 | 0.00 | 0.00 to 0.01 | 0.000 |
| | | 12614 | Gender | Female | Male | 0.31 | 0.04 | 0.23 to 0.39 | 0.000 |
| | | 12614 | Education | Secondary | Elementary or less | -0.29 | 0.05 | -0.38 to -0.20 | 0.000 |

| Outcome | Model type | Analytic N | Variable | Level / comparison | Reference category | Coefficient | Standard error | 95% CI | p-value |
|---|---|---|---|---|---|---|---|---|---|
| | | 12614 | Education | Four years beyond HS | Elementary or less | -0.47 | 0.08 | -0.62 to -0.32 | 0.000 |
| | | 12614 | Residential setting | Small town | Rural | 0.11 | 0.05 | 0.00 to 0.21 | 0.047 |
| | | 12614 | Residential setting | Large city | Rural | -0.02 | 0.07 | -0.15 to 0.12 | 0.815 |
| | | 12614 | Residential setting | Suburb | Rural | -0.03 | 0.12 | -0.26 to 0.19 | 0.771 |
| | | 12614 | *Threshold / cut-point* | Not at all\|Several days | | 0.17 | 0.08 | 0.01 to 0.33 | 0.033 |
| | | 12614 | *Threshold / cut-point* | Several days\|More than half the days | | 1.17 | 0.08 | 1.02 to 1.33 | 0.000 |
| | | 12614 | *Threshold / cut-point* | More than half the days\|Nearly every day | | 2.11 | 0.08 | 1.95 to 2.27 | 0.000 |
| **Forgiveness§** | **Proportional-odds regression** | 12591 | Childhood predictor | Yes | No | 0.22 | 0.06 | 0.10 to 0.34 | 0.000 |
| | | 12591 | Age | Per 1-year increase | | 0.00 | 0.00 | 0.00 to 0.00 | 0.794 |
| | | 12591 | Gender | Female | Male | 0.10 | 0.04 | 0.01 to 0.18 | 0.023 |
| | | 12591 | Education | Secondary | Elementary or less | -0.07 | 0.05 | -0.17 to 0.04 | 0.200 |
| | | 12591 | Education | Four years beyond HS | Elementary or less | -0.19 | 0.08 | -0.34 to -0.04 | 0.014 |
| | | 12591 | Residential setting | Small town | Rural | 0.05 | 0.05 | -0.05 to 0.16 | 0.330 |
| | | 12591 | Residential setting | Large city | Rural | 0.04 | 0.08 | -0.11 to 0.20 | 0.595 |
| | | 12591 | Residential setting | Suburb | Rural | 0.06 | 0.12 | -0.18 to 0.29 | 0.634 |
| | | 12591 | *Threshold / cut-point* | Never\|Rarely | | -2.34 | 0.09 | -2.51 to -2.16 | 0.000 |
| | | 12591 | *Threshold / cut-point* | Rarely\|Often | | -1.00 | 0.08 | -1.17 to -0.84 | 0.000 |
| | | 12591 | *Threshold / cut-point* | Often\|Always | | -0.36 | 0.08 | -0.52 to -0.20 | 0.000 |
| **Support reducing differences in income levels§** | **Proportional-odds regression** | 12451 | Childhood predictor | Yes | No | 0.13 | 0.09 | -0.05 to 0.30 | 0.166 |
| | | 12451 | Age | Per 1-year increase | | 0.00 | 0.00 | -0.01 to 0.00 | 0.095 |
| | | 12451 | Gender | Female | Male | -0.04 | 0.06 | -0.15 to 0.07 | 0.459 |
| | | 12451 | Education | Secondary | Elementary or less | -0.06 | 0.07 | -0.19 to 0.08 | 0.408 |
| | | 12451 | Education | Four years beyond HS | Elementary or less | -0.27 | 0.11 | -0.49 to -0.06 | 0.014 |
| | | 12451 | Residential setting | Small town | Rural | 0.05 | 0.13 | -0.20 to 0.29 | 0.722 |
| | | 12451 | Residential setting | Large city | Rural | 0.10 | 0.15 | -0.19 to 0.39 | 0.493 |
| | | 12451 | Residential setting | Suburb | Rural | -0.08 | 0.18 | -0.44 to 0.27 | 0.644 |
| | | 12451 | *Threshold / cut-point* | Strongly disagree\|Somewhat disagree | | -3.82 | 0.18 | -4.17 to -3.47 | 0.000 |
| | | 12451 | *Threshold / cut-point* | Somewhat disagree\|Neither | | -3.29 | 0.17 | -3.61 to -2.96 | 0.000 |
| | | 12451 | *Threshold / cut-point* | Neither\|Somewhat agree | | -2.84 | 0.16 | -3.15 to -2.53 | 0.000 |
| | | 12451 | *Threshold / cut-point* | Somewhat agree\|Strongly agree | | -1.27 | 0.14 | -1.55 to -0.99 | 0.000 |
| **Have little interest in doing things§** | **Proportional-odds regression** | 12577 | Childhood predictor | Yes | No | 0.05 | 0.06 | -0.07 to 0.17 | 0.384 |
| | | 12577 | Age | Per 1-year increase | | 0.00 | 0.00 | 0.00 to 0.01 | 0.022 |
| | | 12577 | Gender | Female | Male | 0.03 | 0.04 | -0.05 to 0.11 | 0.502 |
| | | 12577 | Education | Secondary | Elementary or less | 0.08 | 0.05 | -0.02 to 0.17 | 0.117 |
| | | 12577 | Education | Four years beyond HS | Elementary or less | 0.10 | 0.08 | -0.05 to 0.25 | 0.194 |

| Outcome | Model type | Analytic N | Variable | Level / comparison | Reference category | Coefficient | Standard error | 95% CI | p-value |
|---|---|---|---|---|---|---|---|---|---|
| | | 12577 | Residential setting | Small town | Rural | 0.28 | 0.06 | 0.15 to 0.40 | 0.000 |
| | | 12577 | Residential setting | Large city | Rural | 0.11 | 0.08 | -0.04 to 0.27 | 0.154 |
| | | 12577 | Residential setting | Suburb | Rural | 0.28 | 0.15 | -0.01 to 0.57 | 0.060 |
| | | 12577 | *Threshold / cut-point* | Not at all\|Several days | | -0.47 | 0.09 | -0.64 to -0.29 | 0.000 |
| | | 12577 | *Threshold / cut-point* | Several days\|More than half the days | | 0.46 | 0.09 | 0.29 to 0.64 | 0.000 |
| | | 12577 | *Threshold / cut-point* | More than half the days\|Nearly every day | | 1.24 | 0.09 | 1.06 to 1.41 | 0.000 |
| **Various aspects of life are balanced§** | **Proportional-odds regression** | 12505 | Childhood predictor | Yes | No | -0.03 | 0.06 | -0.16 to 0.10 | 0.648 |
| | | 12505 | Age | Per 1-year increase | | 0.00 | 0.00 | 0.00 to 0.01 | 0.077 |
| | | 12505 | Gender | Female | Male | -0.03 | 0.04 | -0.11 to 0.05 | 0.478 |
| | | 12505 | Education | Secondary | Elementary or less | 0.05 | 0.04 | -0.04 to 0.13 | 0.275 |
| | | 12505 | Education | Four years beyond HS | Elementary or less | 0.11 | 0.07 | -0.03 to 0.24 | 0.117 |
| | | 12505 | Residential setting | Small town | Rural | 0.11 | 0.05 | 0.00 to 0.21 | 0.045 |
| | | 12505 | Residential setting | Large city | Rural | 0.09 | 0.07 | -0.04 to 0.22 | 0.167 |
| | | 12505 | Residential setting | Suburb | Rural | 0.36 | 0.11 | 0.14 to 0.58 | 0.001 |
| | | 12505 | *Threshold / cut-point* | Never\|Rarely | | -2.03 | 0.09 | -2.20 to -1.86 | 0.000 |
| | | 12505 | *Threshold / cut-point* | Rarely\|Often | | 0.15 | 0.08 | -0.01 to 0.31 | 0.072 |
| | | 12505 | *Threshold / cut-point* | Often\|Always | | 0.94 | 0.08 | 0.77 to 1.10 | 0.000 |
| **Obeying the law§** | **Proportional-odds regression** | 12639 | Childhood predictor | Yes | No | 0.09 | 0.12 | -0.15 to 0.33 | 0.449 |
| | | 12639 | Age | Per 1-year increase | | 0.00 | 0.00 | -0.01 to 0.00 | 0.475 |
| | | 12639 | Gender | Female | Male | -0.15 | 0.07 | -0.29 to -0.01 | 0.042 |
| | | 12639 | Education | Secondary | Elementary or less | 0.11 | 0.10 | -0.08 to 0.30 | 0.241 |
| | | 12639 | Education | Four years beyond HS | Elementary or less | -0.20 | 0.12 | -0.44 to 0.04 | 0.110 |
| | | 12639 | Residential setting | Small town | Rural | -0.04 | 0.15 | -0.33 to 0.25 | 0.782 |
| | | 12639 | Residential setting | Large city | Rural | -0.23 | 0.18 | -0.58 to 0.13 | 0.214 |
| | | 12639 | Residential setting | Suburb | Rural | -0.26 | 0.22 | -0.70 to 0.18 | 0.241 |
| | | 12639 | *Threshold / cut-point* | Strongly disagree\|Somewhat disagree | | -5.13 | 0.25 | -5.62 to -4.64 | 0.000 |
| | | 12639 | *Threshold / cut-point* | Somewhat disagree\|Neither | | -4.51 | 0.21 | -4.92 to -4.09 | 0.000 |
| | | 12639 | *Threshold / cut-point* | Neither\|Somewhat agree | | -3.99 | 0.21 | -4.40 to -3.59 | 0.000 |
| | | 12639 | *Threshold / cut-point* | Somewhat agree\|Strongly agree | | -2.17 | 0.19 | -2.54 to -1.81 | 0.000 |
| **Feel at peace§** | **Proportional-odds regression** | 12576 | Childhood predictor | Yes | No | -0.09 | 0.06 | -0.21 to 0.04 | 0.174 |
| | | 12576 | Age | Per 1-year increase | | 0.01 | 0.00 | 0.00 to 0.01 | 0.000 |
| | | 12576 | Gender | Female | Male | 0.01 | 0.04 | -0.06 to 0.08 | 0.814 |
| | | 12576 | Education | Secondary | Elementary or less | 0.02 | 0.05 | -0.07 to 0.11 | 0.683 |
| | | 12576 | Education | Four years beyond HS | Elementary or less | 0.05 | 0.06 | -0.07 to 0.18 | 0.416 |
| | | 12576 | Residential setting | Small town | Rural | 0.20 | 0.05 | 0.09 to 0.30 | 0.000 |

| Outcome | Model type | Analytic N | Variable | Level / comparison | Reference category | Coefficient | Standard error | 95% CI | p-value |
|---|---|---|---|---|---|---|---|---|---|
| | | 12576 | Residential setting | Large city | Rural | 0.15 | 0.07 | 0.01 to 0.29 | 0.038 |
| | | 12576 | Residential setting | Suburb | Rural | 0.24 | 0.10 | 0.05 to 0.44 | 0.014 |
| | | 12576 | *Threshold / cut-point* | Never\|Rarely | | -1.89 | 0.08 | -2.05 to -1.73 | 0.000 |
| | | 12576 | *Threshold / cut-point* | Rarely\|Often | | -0.06 | 0.08 | -0.21 to 0.10 | 0.457 |
| | | 12576 | *Threshold / cut-point* | Often\|Always | | 0.63 | 0.08 | 0.48 to 0.79 | 0.000 |
| **Say in government#** | **Proportional-odds regression** | 12615 | Childhood predictor | Yes | No | 0.13 | 0.08 | -0.03 to 0.29 | 0.117 |
| | | 12615 | Age | Per 1-year increase | | 0.00 | 0.00 | -0.01 to 0.00 | 0.389 |
| | | 12615 | Gender | Female | Male | 0.10 | 0.05 | 0.00 to 0.20 | 0.042 |
| | | 12615 | Education | Secondary | Elementary or less | -0.23 | 0.06 | -0.35 to -0.11 | 0.000 |
| | | 12615 | Education | Four years beyond HS | Elementary or less | -0.11 | 0.10 | -0.31 to 0.09 | 0.273 |
| | | 12615 | Residential setting | Small town | Rural | 0.01 | 0.08 | -0.14 to 0.16 | 0.871 |
| | | 12615 | Residential setting | Large city | Rural | -0.16 | 0.09 | -0.34 to 0.02 | 0.089 |
| | | 12615 | Residential setting | Suburb | Rural | -0.44 | 0.13 | -0.70 to -0.18 | 0.001 |
| | | 12615 | *Threshold / cut-point* | Disagree\|Unsure | | -1.77 | 0.11 | -1.99 to -1.55 | 0.000 |
| | | 12615 | *Threshold / cut-point* | Unsure\|Agree | | -1.37 | 0.11 | -1.58 to -1.16 | 0.000 |
| **Extent of suffering§** | **Proportional-odds regression** | 12591 | Childhood predictor | Yes | No | 0.07 | 0.06 | -0.05 to 0.19 | 0.250 |
| | | 12591 | Age | Per 1-year increase | | 0.02 | 0.00 | 0.01 to 0.02 | 0.000 |
| | | 12591 | Gender | Female | Male | 0.27 | 0.04 | 0.19 to 0.35 | 0.000 |
| | | 12591 | Education | Secondary | Elementary or less | -0.15 | 0.05 | -0.24 to -0.06 | 0.001 |
| | | 12591 | Education | Four years beyond HS | Elementary or less | -0.28 | 0.07 | -0.42 to -0.13 | 0.000 |
| | | 12591 | Residential setting | Small town | Rural | 0.05 | 0.06 | -0.06 to 0.16 | 0.405 |
| | | 12591 | Residential setting | Large city | Rural | -0.13 | 0.08 | -0.28 to 0.02 | 0.084 |
| | | 12591 | Residential setting | Suburb | Rural | 0.06 | 0.11 | -0.15 to 0.27 | 0.559 |
| | | 12591 | *Threshold / cut-point* | Not at all\|Not very much | | 0.30 | 0.08 | 0.14 to 0.47 | 0.000 |
| | | 12591 | *Threshold / cut-point* | Not very much\|Some | | 0.72 | 0.08 | 0.55 to 0.88 | 0.000 |
| | | 12591 | *Threshold / cut-point* | Some\|A lot | | 2.15 | 0.09 | 1.98 to 2.32 | 0.000 |
| **Distress from life-threatening experiences§** | **Proportional-odds regression** | 12609 | Childhood predictor | Yes | No | 0.15 | 0.06 | 0.03 to 0.28 | 0.015 |
| | | 12609 | Age | Per 1-year increase | | 0.00 | 0.00 | 0.00 to 0.01 | 0.019 |
| | | 12609 | Gender | Female | Male | 0.08 | 0.04 | -0.01 to 0.17 | 0.070 |
| | | 12609 | Education | Secondary | Elementary or less | -0.26 | 0.05 | -0.35 to -0.17 | 0.000 |
| | | 12609 | Education | Four years beyond HS | Elementary or less | -0.44 | 0.07 | -0.59 to -0.30 | 0.000 |
| | | 12609 | Residential setting | Small town | Rural | -0.17 | 0.05 | -0.27 to -0.08 | 0.000 |
| | | 12609 | Residential setting | Large city | Rural | -0.23 | 0.07 | -0.37 to -0.09 | 0.001 |
| | | 12609 | Residential setting | Suburb | Rural | -0.19 | 0.12 | -0.41 to 0.04 | 0.100 |
| | | 12609 | *Threshold / cut-point* | Not at all\|Not very much | | -0.27 | 0.08 | -0.42 to -0.12 | 0.000 |
| | | 12609 | *Threshold / cut-point* | Not very much\|Some | | 0.07 | 0.08 | -0.08 to 0.22 | 0.362 |
| | | 12609 | *Threshold / cut-point* | Some\|A lot | | 1.08 | 0.08 | 0.93 to 1.23 | 0.000 |

| Outcome | Model type | Analytic N | Variable | Level / comparison | Reference category | Coefficient | Standard error | 95% CI | p-value |
|---|---|---|---|---|---|---|---|---|---|
| **Trust in others§** | **Proportional-odds regression** | 12327 | Childhood predictor | Yes | No | -0.10 | 0.06 | -0.21 to 0.01 | 0.071 |
| | | 12327 | Age | Per 1-year increase | | 0.00 | 0.00 | 0.00 to 0.01 | 0.017 |
| | | 12327 | Gender | Female | Male | -0.05 | 0.04 | -0.12 to 0.03 | 0.248 |
| | | 12327 | Education | Secondary | Elementary or less | -0.04 | 0.05 | -0.14 to 0.05 | 0.331 |
| | | 12327 | Education | Four years beyond HS | Elementary or less | -0.07 | 0.07 | -0.22 to 0.07 | 0.304 |
| | | 12327 | Residential setting | Small town | Rural | 0.24 | 0.05 | 0.14 to 0.34 | 0.000 |
| | | 12327 | Residential setting | Large city | Rural | 0.01 | 0.07 | -0.12 to 0.14 | 0.896 |
| | | 12327 | Residential setting | Suburb | Rural | -0.17 | 0.14 | -0.45 to 0.11 | 0.222 |
| | | 12327 | *Threshold / cut-point* | None\|Not very many | | -2.15 | 0.09 | -2.33 to -1.97 | 0.000 |
| | | 12327 | *Threshold / cut-point* | Not very many\|Some | | -1.36 | 0.09 | -1.54 to -1.18 | 0.000 |
| | | 12327 | *Threshold / cut-point* | Some\|Most | | 0.83 | 0.09 | 0.66 to 1.00 | 0.000 |
| | | 12327 | *Threshold / cut-point* | Most\|All | | 1.81 | 0.09 | 1.63 to 1.98 | 0.000 |

**eTable 9.** Table of coefficients for adjusted associations between not feeling loved by mother as predictor and 45 outcomes of interest and two flourishing composite indices.

Adjusted survey-weighted models use ANNUAL_WEIGHT_C1 with primary sampling unit (PSU) and strata and adjust for age, gender, education, and residential setting. Reference categories are shown in the table. Coefficients are on the original regression-coefficient scale: beta for continuous outcomes, log-odds for binary outcomes, and proportional-odds log coefficients for ordinal outcomes. Model intercepts are displayed for continuous and binary outcomes. Proportional-odds models do not have a single conventional intercept; their estimated ordinal cut-points/thresholds are displayed after the covariate coefficients, with standard errors (SEs), 95% CIs, and Wald p-values.

***Note:*** All results except p-values are rounded to the two decimals. P-values are rounded to three decimals. Values displayed as 0.000 indicate $P < 0.0005$ due to rounding, rather than an exact zero. Statistical significance was determined using unrounded P-values at the Bonferroni-adjusted threshold of $P < 0.0011$.

| Outcome | Model type | Analytic N | Variable | Level / comparison | Reference category | Coefficient | Standard error | 95% CI | p-value |
|---|---|---|---|---|---|---|---|---|---|
| **Belonging** | **Linear regression** | 12326 | *Intercept* | | | 8.64 | 0.11 | 8.42 to 8.87 | 0.000 |
| | | 12326 | Childhood predictor | No | Yes | -0.31 | 0.28 | -0.86 to 0.24 | 0.265 |
| | | 12326 | Age | Per 1-year increase | | -0.01 | 0.00 | -0.02 to -0.01 | 0.000 |
| | | 12326 | Gender | Female | Male | -0.19 | 0.06 | -0.31 to -0.07 | 0.003 |
| | | 12326 | Education | Secondary | Elementary or less | 0.22 | 0.07 | 0.07 to 0.37 | 0.003 |
| | | 12326 | Education | Four years beyond HS | Elementary or less | 0.30 | 0.10 | 0.10 to 0.51 | 0.004 |
| | | 12326 | Residential setting | Small town | Rural | 0.38 | 0.08 | 0.23 to 0.54 | 0.000 |
| | | 12326 | Residential setting | Large city | Rural | 0.45 | 0.10 | 0.25 to 0.66 | 0.000 |
| | | 12326 | Residential setting | Suburb | Rural | 0.01 | 0.18 | -0.34 to 0.37 | 0.934 |
| **Relationship contentment‡** | **Linear regression** | 12391 | *Intercept* | | | 8.11 | 0.13 | 7.86 to 8.37 | 0.000 |
| | | 12391 | Childhood predictor | No | Yes | -0.55 | 0.32 | -1.17 to 0.07 | 0.082 |
| | | 12391 | Age | Per 1-year increase | | -0.01 | 0.00 | -0.02 to -0.01 | 0.000 |
| | | 12391 | Gender | Female | Male | 0.37 | 0.07 | 0.23 to 0.51 | 0.000 |
| | | 12391 | Education | Secondary | Elementary or less | 0.15 | 0.07 | 0.01 to 0.30 | 0.038 |
| | | 12391 | Education | Four years beyond HS | Elementary or less | 0.28 | 0.12 | 0.05 to 0.51 | 0.017 |
| | | 12391 | Residential setting | Small town | Rural | 0.37 | 0.08 | 0.21 to 0.54 | 0.000 |
| | | 12391 | Residential setting | Large city | Rural | 0.09 | 0.10 | -0.10 to 0.29 | 0.343 |
| | | 12391 | Residential setting | Suburb | Rural | -0.27 | 0.17 | -0.61 to 0.07 | 0.117 |
| **Positive expectations** | **Linear regression** | 12351 | *Intercept* | | | 7.78 | 0.15 | 7.50 to 8.07 | 0.000 |
| | | 12351 | Childhood predictor | No | Yes | 0.00 | 0.28 | -0.55 to 0.55 | 0.996 |
| | | 12351 | Age | Per 1-year increase | | 0.00 | 0.00 | -0.01 to 0.00 | 0.147 |
| | | 12351 | Gender | Female | Male | 0.23 | 0.06 | 0.11 to 0.36 | 0.000 |
| | | 12351 | Education | Secondary | Elementary or less | 0.29 | 0.07 | 0.14 to 0.43 | 0.000 |
| | | 12351 | Education | Four years beyond HS | Elementary or less | 0.36 | 0.10 | 0.17 to 0.55 | 0.000 |
| | | 12351 | Residential setting | Small town | Rural | 0.45 | 0.09 | 0.28 to 0.62 | 0.000 |
| | | 12351 | Residential setting | Large city | Rural | 0.45 | 0.11 | 0.23 to 0.67 | 0.000 |
| | | 12351 | Residential setting | Suburb | Rural | 0.44 | 0.16 | 0.12 to 0.75 | 0.007 |
| **Worry about meeting normal monthly living expenses§§‡‡** | **Linear regression** | 12392 | *Intercept* | | | 5.77 | 0.19 | 5.39 to 6.15 | 0.000 |
| | | 12392 | Childhood predictor | No | Yes | 0.31 | 0.33 | -0.33 to 0.95 | 0.343 |
| | | 12392 | Age | Per 1-year increase | | 0.00 | 0.00 | -0.01 to 0.01 | 0.591 |
| | | 12392 | Gender | Female | Male | 0.34 | 0.09 | 0.16 to 0.52 | 0.000 |

| Outcome | Model type | Analytic N | Variable | Level / comparison | Reference category | Coefficient | Standard error | 95% CI | p-value |
|---|---|---|---|---|---|---|---|---|---|
| | | 12392 | Education | Secondary | Elementary or less | -0.47 | 0.11 | -0.68 to -0.26 | 0.000 |
| | | 12392 | Education | Four years beyond HS | Elementary or less | -0.64 | 0.15 | -0.94 to -0.35 | 0.000 |
| | | 12392 | Residential setting | Small town | Rural | 0.22 | 0.13 | -0.03 to 0.47 | 0.080 |
| | | 12392 | Residential setting | Large city | Rural | -0.24 | 0.15 | -0.54 to 0.06 | 0.114 |
| | | 12392 | Residential setting | Suburb | Rural | -0.17 | 0.24 | -0.64 to 0.31 | 0.487 |
| **Freedom** | **Linear regression** | 12364 | *Intercept* | | | 8.18 | 0.12 | 7.94 to 8.43 | 0.000 |
| | | 12364 | Childhood predictor | No | Yes | -0.50 | 0.29 | -1.07 to 0.06 | 0.080 |
| | | 12364 | Age | Per 1-year increase | | -0.01 | 0.00 | -0.01 to 0.00 | 0.003 |
| | | 12364 | Gender | Female | Male | -0.13 | 0.07 | -0.27 to 0.00 | 0.051 |
| | | 12364 | Education | Secondary | Elementary or less | 0.30 | 0.08 | 0.15 to 0.45 | 0.000 |
| | | 12364 | Education | Four years beyond HS | Elementary or less | 0.40 | 0.10 | 0.20 to 0.61 | 0.000 |
| | | 12364 | Residential setting | Small town | Rural | 0.37 | 0.08 | 0.22 to 0.52 | 0.000 |
| | | 12364 | Residential setting | Large city | Rural | 0.40 | 0.10 | 0.20 to 0.61 | 0.000 |
| | | 12364 | Residential setting | Suburb | Rural | 0.13 | 0.16 | -0.19 to 0.45 | 0.419 |
| **Give up some happiness now for greater happiness later‡** | **Linear regression** | 12332 | *Intercept* | | | 7.25 | 0.15 | 6.95 to 7.55 | 0.000 |
| | | 12332 | Childhood predictor | No | Yes | 0.10 | 0.33 | -0.55 to 0.75 | 0.769 |
| | | 12332 | Age | Per 1-year increase | | -0.01 | 0.00 | -0.02 to 0.00 | 0.000 |
| | | 12332 | Gender | Female | Male | 0.16 | 0.08 | 0.00 to 0.31 | 0.044 |
| | | 12332 | Education | Secondary | Elementary or less | 0.18 | 0.10 | -0.01 to 0.36 | 0.067 |
| | | 12332 | Education | Four years beyond HS | Elementary or less | 0.17 | 0.13 | -0.09 to 0.43 | 0.201 |
| | | 12332 | Residential setting | Small town | Rural | 0.42 | 0.10 | 0.22 to 0.61 | 0.000 |
| | | 12332 | Residential setting | Large city | Rural | 0.19 | 0.13 | -0.08 to 0.45 | 0.168 |
| | | 12332 | Residential setting | Suburb | Rural | 0.07 | 0.27 | -0.47 to 0.60 | 0.810 |
| **Have a long list of gratitude** | **Linear regression** | 12228 | *Intercept* | | | 7.40 | 0.15 | 7.11 to 7.70 | 0.000 |
| | | 12228 | Childhood predictor | No | Yes | 0.13 | 0.29 | -0.43 to 0.70 | 0.644 |
| | | 12228 | Age | Per 1-year increase | | -0.01 | 0.00 | -0.01 to 0.00 | 0.055 |
| | | 12228 | Gender | Female | Male | 0.21 | 0.07 | 0.07 to 0.35 | 0.004 |
| | | 12228 | Education | Secondary | Elementary or less | 0.31 | 0.08 | 0.15 to 0.47 | 0.000 |
| | | 12228 | Education | Four years beyond HS | Elementary or less | 0.41 | 0.11 | 0.18 to 0.63 | 0.000 |
| | | 12228 | Residential setting | Small town | Rural | 0.44 | 0.09 | 0.25 to 0.62 | 0.000 |
| | | 12228 | Residential setting | Large city | Rural | 0.44 | 0.13 | 0.18 to 0.70 | 0.001 |
| | | 12228 | Residential setting | Suburb | Rural | 0.49 | 0.17 | 0.16 to 0.82 | 0.004 |
| **Happiness‡** | **Linear regression** | 12388 | *Intercept* | | | 6.63 | 0.16 | 6.32 to 6.93 | 0.000 |
| | | 12388 | Childhood predictor | No | Yes | -0.04 | 0.31 | -0.66 to 0.57 | 0.887 |
| | | 12388 | Age | Per 1-year increase | | -0.02 | 0.00 | -0.02 to -0.01 | 0.000 |
| | | 12388 | Gender | Female | Male | 0.27 | 0.08 | 0.11 to 0.42 | 0.001 |
| | | 12388 | Education | Secondary | Elementary or less | 0.45 | 0.09 | 0.27 to 0.62 | 0.000 |
| | | 12388 | Education | Four years beyond HS | Elementary or less | 0.68 | 0.12 | 0.44 to 0.92 | 0.000 |
| | | 12388 | Residential setting | Small town | Rural | 0.39 | 0.10 | 0.19 to 0.60 | 0.000 |
| | | 12388 | Residential setting | Large city | Rural | 0.37 | 0.13 | 0.12 to 0.61 | 0.004 |

| Outcome | Model type | Analytic N | Variable | Level / comparison | Reference category | Coefficient | Standard error | 95% CI | p-value |
|---|---|---|---|---|---|---|---|---|---|
| | | 12388 | Residential setting | Suburb | Rural | -0.08 | 0.22 | -0.50 to 0.34 | 0.715 |
| **Remain hopeful for the future** | **Linear regression** | 12338 | *Intercept* | | | 8.15 | 0.12 | 7.91 to 8.40 | 0.000 |
| | | 12338 | Childhood predictor | No | Yes | 0.32 | 0.20 | -0.07 to 0.71 | 0.112 |
| | | 12338 | Age | Per 1-year increase | | -0.01 | 0.00 | -0.01 to 0.00 | 0.001 |
| | | 12338 | Gender | Female | Male | 0.01 | 0.06 | -0.10 to 0.12 | 0.901 |
| | | 12338 | Education | Secondary | Elementary or less | 0.40 | 0.06 | 0.27 to 0.52 | 0.000 |
| | | 12338 | Education | Four years beyond HS | Elementary or less | 0.62 | 0.09 | 0.43 to 0.80 | 0.000 |
| | | 12338 | Residential setting | Small town | Rural | 0.49 | 0.08 | 0.33 to 0.65 | 0.000 |
| | | 12338 | Residential setting | Large city | Rural | 0.47 | 0.10 | 0.27 to 0.68 | 0.000 |
| | | 12338 | Residential setting | Suburb | Rural | 0.41 | 0.18 | 0.06 to 0.76 | 0.021 |
| **Understand purpose in life‡** | **Linear regression** | 12317 | *Intercept* | | | 8.00 | 0.13 | 7.74 to 8.27 | 0.000 |
| | | 12317 | Childhood predictor | No | Yes | 0.15 | 0.25 | -0.34 to 0.64 | 0.556 |
| | | 12317 | Age | Per 1-year increase | | -0.01 | 0.00 | -0.02 to -0.01 | 0.000 |
| | | 12317 | Gender | Female | Male | 0.12 | 0.07 | -0.02 to 0.26 | 0.095 |
| | | 12317 | Education | Secondary | Elementary or less | 0.44 | 0.08 | 0.29 to 0.60 | 0.000 |
| | | 12317 | Education | Four years beyond HS | Elementary or less | 0.74 | 0.10 | 0.55 to 0.93 | 0.000 |
| | | 12317 | Residential setting | Small town | Rural | 0.42 | 0.09 | 0.24 to 0.60 | 0.000 |
| | | 12317 | Residential setting | Large city | Rural | 0.42 | 0.12 | 0.18 to 0.65 | 0.000 |
| | | 12317 | Residential setting | Suburb | Rural | 0.43 | 0.15 | 0.14 to 0.72 | 0.004 |
| **Life satisfaction‡** | **Linear regression** | 12368 | *Intercept* | | | 7.07 | 0.15 | 6.77 to 7.36 | 0.000 |
| | | 12368 | Childhood predictor | No | Yes | -0.04 | 0.31 | -0.64 to 0.57 | 0.906 |
| | | 12368 | Age | Per 1-year increase | | -0.01 | 0.00 | -0.02 to -0.01 | 0.000 |
| | | 12368 | Gender | Female | Male | 0.31 | 0.08 | 0.16 to 0.46 | 0.000 |
| | | 12368 | Education | Secondary | Elementary or less | 0.31 | 0.09 | 0.13 to 0.49 | 0.001 |
| | | 12368 | Education | Four years beyond HS | Elementary or less | 0.47 | 0.12 | 0.24 to 0.71 | 0.000 |
| | | 12368 | Residential setting | Small town | Rural | 0.31 | 0.09 | 0.12 to 0.49 | 0.001 |
| | | 12368 | Residential setting | Large city | Rural | 0.26 | 0.12 | 0.02 to 0.50 | 0.035 |
| | | 12368 | Residential setting | Suburb | Rural | -0.19 | 0.21 | -0.60 to 0.22 | 0.371 |
| **Loneliness§** | **Linear regression** | 12389 | *Intercept* | | | 4.24 | 0.17 | 3.91 to 4.57 | 0.000 |
| | | 12389 | Childhood predictor | No | Yes | 0.83 | 0.35 | 0.16 to 1.51 | 0.016 |
| | | 12389 | Age | Per 1-year increase | | 0.00 | 0.00 | -0.01 to 0.01 | 0.816 |
| | | 12389 | Gender | Female | Male | -0.14 | 0.09 | -0.31 to 0.03 | 0.102 |
| | | 12389 | Education | Secondary | Elementary or less | -0.32 | 0.10 | -0.51 to -0.13 | 0.001 |
| | | 12389 | Education | Four years beyond HS | Elementary or less | -0.62 | 0.15 | -0.91 to -0.34 | 0.000 |
| | | 12389 | Residential setting | Small town | Rural | -0.37 | 0.10 | -0.58 to -0.17 | 0.000 |
| | | 12389 | Residential setting | Large city | Rural | -0.54 | 0.14 | -0.81 to -0.27 | 0.000 |
| | | 12389 | Residential setting | Suburb | Rural | -0.15 | 0.23 | -0.60 to 0.30 | 0.509 |
| **Mental health‡** | **Linear regression** | 12393 | *Intercept* | | | 8.02 | 0.14 | 7.75 to 8.30 | 0.000 |
| | | 12393 | Childhood predictor | No | Yes | -0.56 | 0.30 | -1.15 to 0.03 | 0.062 |
| | | 12393 | Age | Per 1-year increase | | -0.02 | 0.00 | -0.03 to -0.02 | 0.000 |

| Outcome | Model type | Analytic N | Variable | Level / comparison | Reference category | Coefficient | Standard error | 95% CI | p-value |
|---|---|---|---|---|---|---|---|---|---|
| | | 12393 | Gender | Female | Male | -0.13 | 0.07 | -0.26 to 0.01 | 0.064 |
| | | 12393 | Education | Secondary | Elementary or less | 0.42 | 0.08 | 0.27 to 0.57 | 0.000 |
| | | 12393 | Education | Four years beyond HS | Elementary or less | 0.52 | 0.11 | 0.29 to 0.74 | 0.000 |
| | | 12393 | Residential setting | Small town | Rural | 0.41 | 0.09 | 0.23 to 0.60 | 0.000 |
| | | 12393 | Residential setting | Large city | Rural | 0.48 | 0.12 | 0.25 to 0.71 | 0.000 |
| | | 12393 | Residential setting | Suburb | Rural | 0.20 | 0.18 | -0.15 to 0.55 | 0.262 |
| **Perceived support from others** | **Linear regression** | 12379 | *Intercept* | | | 6.64 | 0.16 | 6.33 to 6.95 | 0.000 |
| | | 12379 | Childhood predictor | No | Yes | -1.42 | 0.41 | -2.23 to -0.62 | 0.001 |
| | | 12379 | Age | Per 1-year increase | | -0.01 | 0.00 | -0.02 to -0.01 | 0.000 |
| | | 12379 | Gender | Female | Male | 0.57 | 0.09 | 0.39 to 0.74 | 0.000 |
| | | 12379 | Education | Secondary | Elementary or less | 0.20 | 0.10 | 0.00 to 0.39 | 0.045 |
| | | 12379 | Education | Four years beyond HS | Elementary or less | 0.12 | 0.14 | -0.15 to 0.39 | 0.392 |
| | | 12379 | Residential setting | Small town | Rural | 0.20 | 0.11 | -0.01 to 0.41 | 0.068 |
| | | 12379 | Residential setting | Large city | Rural | -0.45 | 0.14 | -0.73 to -0.18 | 0.001 |
| | | 12379 | Residential setting | Suburb | Rural | -0.20 | 0.21 | -0.62 to 0.22 | 0.344 |
| **Physical health‡** | **Linear regression** | 12396 | *Intercept* | | | 8.39 | 0.14 | 8.11 to 8.67 | 0.000 |
| | | 12396 | Childhood predictor | No | Yes | -0.33 | 0.29 | -0.89 to 0.24 | 0.261 |
| | | 12396 | Age | Per 1-year increase | | -0.04 | 0.00 | -0.05 to -0.04 | 0.000 |
| | | 12396 | Gender | Female | Male | -0.34 | 0.08 | -0.49 to -0.19 | 0.000 |
| | | 12396 | Education | Secondary | Elementary or less | 0.37 | 0.08 | 0.22 to 0.52 | 0.000 |
| | | 12396 | Education | Four years beyond HS | Elementary or less | 0.66 | 0.11 | 0.45 to 0.87 | 0.000 |
| | | 12396 | Residential setting | Small town | Rural | 0.50 | 0.09 | 0.32 to 0.68 | 0.000 |
| | | 12396 | Residential setting | Large city | Rural | 0.66 | 0.11 | 0.44 to 0.87 | 0.000 |
| | | 12396 | Residential setting | Suburb | Rural | 0.17 | 0.18 | -0.17 to 0.52 | 0.332 |
| **Act to promote good‡** | **Linear regression** | 12351 | *Intercept* | | | 8.06 | 0.11 | 7.85 to 8.27 | 0.000 |
| | | 12351 | Childhood predictor | No | Yes | -0.11 | 0.25 | -0.60 to 0.38 | 0.658 |
| | | 12351 | Age | Per 1-year increase | | -0.01 | 0.00 | -0.01 to 0.00 | 0.001 |
| | | 12351 | Gender | Female | Male | 0.11 | 0.06 | -0.01 to 0.23 | 0.067 |
| | | 12351 | Education | Secondary | Elementary or less | 0.39 | 0.06 | 0.26 to 0.51 | 0.000 |
| | | 12351 | Education | Four years beyond HS | Elementary or less | 0.54 | 0.08 | 0.38 to 0.70 | 0.000 |
| | | 12351 | Residential setting | Small town | Rural | 0.62 | 0.07 | 0.47 to 0.76 | 0.000 |
| | | 12351 | Residential setting | Large city | Rural | 0.63 | 0.10 | 0.44 to 0.83 | 0.000 |
| | | 12351 | Residential setting | Suburb | Rural | 0.42 | 0.16 | 0.10 to 0.74 | 0.010 |
| **Relationship satisfaction‡** | **Linear regression** | 12357 | *Intercept* | | | 7.87 | 0.13 | 7.61 to 8.12 | 0.000 |
| | | 12357 | Childhood predictor | No | Yes | -1.13 | 0.34 | -1.80 to -0.45 | 0.001 |
| | | 12357 | Age | Per 1-year increase | | -0.01 | 0.00 | -0.01 to 0.00 | 0.000 |
| | | 12357 | Gender | Female | Male | 0.34 | 0.07 | 0.20 to 0.48 | 0.000 |
| | | 12357 | Education | Secondary | Elementary or less | -0.02 | 0.08 | -0.18 to 0.13 | 0.779 |
| | | 12357 | Education | Four years beyond HS | Elementary or less | 0.04 | 0.11 | -0.18 to 0.25 | 0.745 |
| | | 12357 | Residential setting | Small town | Rural | 0.46 | 0.09 | 0.29 to 0.63 | 0.000 |

| Outcome | Model type | Analytic N | Variable | Level / comparison | Reference category | Coefficient | Standard error | 95% CI | p-value |
|---|---|---|---|---|---|---|---|---|---|
| | | 12357 | Residential setting | Large city | Rural | 0.36 | 0.11 | 0.15 to 0.57 | 0.001 |
| | | 12357 | Residential setting | Suburb | Rural | 0.07 | 0.16 | -0.25 to 0.39 | 0.655 |
| **Show love often** | **Linear regression** | 12381 | *Intercept* | | | 8.29 | 0.12 | 8.04 to 8.53 | 0.000 |
| | | 12381 | Childhood predictor | No | Yes | -0.15 | 0.28 | -0.70 to 0.41 | 0.608 |
| | | 12381 | Age | Per 1-year increase | | -0.01 | 0.00 | -0.01 to 0.00 | 0.001 |
| | | 12381 | Gender | Female | Male | 0.21 | 0.07 | 0.08 to 0.34 | 0.002 |
| | | 12381 | Education | Secondary | Elementary or less | 0.17 | 0.07 | 0.04 to 0.31 | 0.013 |
| | | 12381 | Education | Four years beyond HS | Elementary or less | 0.04 | 0.11 | -0.18 to 0.26 | 0.724 |
| | | 12381 | Residential setting | Small town | Rural | 0.20 | 0.08 | 0.05 to 0.36 | 0.010 |
| | | 12381 | Residential setting | Large city | Rural | 0.02 | 0.11 | -0.20 to 0.25 | 0.853 |
| | | 12381 | Residential setting | Suburb | Rural | -0.08 | 0.20 | -0.47 to 0.31 | 0.693 |
| **Life evaluation five years from now** | **Linear regression** | 11293 | *Intercept* | | | 7.65 | 0.14 | 7.38 to 7.93 | 0.000 |
| | | 11293 | Childhood predictor | No | Yes | -0.37 | 0.26 | -0.88 to 0.13 | 0.151 |
| | | 11293 | Age | Per 1-year increase | | -0.02 | 0.00 | -0.03 to -0.02 | 0.000 |
| | | 11293 | Gender | Female | Male | 0.20 | 0.07 | 0.06 to 0.34 | 0.007 |
| | | 11293 | Education | Secondary | Elementary or less | 0.31 | 0.08 | 0.15 to 0.46 | 0.000 |
| | | 11293 | Education | Four years beyond HS | Elementary or less | 0.58 | 0.12 | 0.35 to 0.81 | 0.000 |
| | | 11293 | Residential setting | Small town | Rural | 0.47 | 0.09 | 0.29 to 0.64 | 0.000 |
| | | 11293 | Residential setting | Large city | Rural | 0.52 | 0.11 | 0.31 to 0.74 | 0.000 |
| | | 11293 | Residential setting | Suburb | Rural | 0.39 | 0.22 | -0.03 to 0.81 | 0.072 |
| **Life evaluation today** | **Linear regression** | 12293 | *Intercept* | | | 5.53 | 0.17 | 5.20 to 5.86 | 0.000 |
| | | 12293 | Childhood predictor | No | Yes | -0.80 | 0.35 | -1.48 to -0.12 | 0.022 |
| | | 12293 | Age | Per 1-year increase | | -0.01 | 0.00 | -0.02 to 0.00 | 0.009 |
| | | 12293 | Gender | Female | Male | 0.50 | 0.08 | 0.33 to 0.66 | 0.000 |
| | | 12293 | Education | Secondary | Elementary or less | 0.24 | 0.09 | 0.07 to 0.41 | 0.006 |
| | | 12293 | Education | Four years beyond HS | Elementary or less | 0.16 | 0.14 | -0.12 to 0.44 | 0.257 |
| | | 12293 | Residential setting | Small town | Rural | 0.27 | 0.10 | 0.07 to 0.48 | 0.010 |
| | | 12293 | Residential setting | Large city | Rural | 0.13 | 0.13 | -0.13 to 0.39 | 0.324 |
| | | 12293 | Residential setting | Suburb | Rural | 0.02 | 0.21 | -0.39 to 0.42 | 0.935 |
| **Worry about safety, food, or housing§§‡‡** | **Linear regression** | 12405 | *Intercept* | | | 6.22 | 0.19 | 5.85 to 6.59 | 0.000 |
| | | 12405 | Childhood predictor | No | Yes | 0.31 | 0.37 | -0.41 to 1.03 | 0.394 |
| | | 12405 | Age | Per 1-year increase | | -0.01 | 0.00 | -0.02 to 0.00 | 0.014 |
| | | 12405 | Gender | Female | Male | 0.32 | 0.09 | 0.15 to 0.49 | 0.000 |
| | | 12405 | Education | Secondary | Elementary or less | -0.55 | 0.11 | -0.76 to -0.34 | 0.000 |
| | | 12405 | Education | Four years beyond HS | Elementary or less | -0.76 | 0.17 | -1.09 to -0.44 | 0.000 |
| | | 12405 | Residential setting | Small town | Rural | 0.39 | 0.13 | 0.13 to 0.65 | 0.004 |
| | | 12405 | Residential setting | Large city | Rural | -0.09 | 0.18 | -0.44 to 0.26 | 0.601 |
| | | 12405 | Residential setting | Suburb | Rural | 0.16 | 0.26 | -0.35 to 0.68 | 0.538 |
| **Life worthwhile‡** | **Linear regression** | 12292 | *Intercept* | | | 6.63 | 0.15 | 6.34 to 6.91 | 0.000 |

| Outcome | Model type | Analytic N | Variable | Level / comparison | Reference category | Coefficient | Standard error | 95% CI | p-value |
|---|---|---|---|---|---|---|---|---|---|
| | | 12292 | Childhood predictor | No | Yes | -0.54 | 0.38 | -1.28 to 0.21 | 0.160 |
| | | 12292 | Age | Per 1-year increase | | -0.01 | 0.00 | -0.01 to 0.00 | 0.002 |
| | | 12292 | Gender | Female | Male | 0.44 | 0.07 | 0.29 to 0.58 | 0.000 |
| | | 12292 | Education | Secondary | Elementary or less | 0.46 | 0.08 | 0.31 to 0.61 | 0.000 |
| | | 12292 | Education | Four years beyond HS | Elementary or less | 0.59 | 0.12 | 0.35 to 0.83 | 0.000 |
| | | 12292 | Residential setting | Small town | Rural | 0.47 | 0.10 | 0.28 to 0.67 | 0.000 |
| | | 12292 | Residential setting | Large city | Rural | 0.47 | 0.13 | 0.22 to 0.72 | 0.000 |
| | | 12292 | Residential setting | Suburb | Rural | 0.40 | 0.20 | 0.01 to 0.80 | 0.043 |
| **Flourishing (with financial indicators)** | **Linear regression** | 11948 | *Intercept* | | | 6.98 | 0.08 | 6.82 to 7.14 | 0.000 |
| | | 11948 | Childhood predictor | No | Yes | -0.28 | 0.16 | -0.58 to 0.03 | 0.081 |
| | | 11948 | Age | Per 1-year increase | | -0.01 | 0.00 | -0.02 to -0.01 | 0.000 |
| | | 11948 | Gender | Female | Male | 0.10 | 0.04 | 0.01 to 0.18 | 0.024 |
| | | 11948 | Education | Secondary | Elementary or less | 0.34 | 0.05 | 0.25 to 0.43 | 0.000 |
| | | 11948 | Education | Four years beyond HS | Elementary or less | 0.50 | 0.06 | 0.38 to 0.63 | 0.000 |
| | | 11948 | Residential setting | Small town | Rural | 0.33 | 0.06 | 0.21 to 0.44 | 0.000 |
| | | 11948 | Residential setting | Large city | Rural | 0.37 | 0.07 | 0.23 to 0.50 | 0.000 |
| | | 11948 | Residential setting | Suburb | Rural | 0.12 | 0.13 | -0.13 to 0.37 | 0.358 |
| **Flourishing (without financial indicators)** | **Linear regression** | 11975 | *Intercept* | | | 7.59 | 0.09 | 7.42 to 7.77 | 0.000 |
| | | 11975 | Childhood predictor | No | Yes | -0.28 | 0.18 | -0.63 to 0.08 | 0.126 |
| | | 11975 | Age | Per 1-year increase | | -0.02 | 0.00 | -0.02 to -0.01 | 0.000 |
| | | 11975 | Gender | Female | Male | 0.18 | 0.05 | 0.09 to 0.27 | 0.000 |
| | | 11975 | Education | Secondary | Elementary or less | 0.31 | 0.05 | 0.21 to 0.40 | 0.000 |
| | | 11975 | Education | Four years beyond HS | Elementary or less | 0.46 | 0.07 | 0.33 to 0.60 | 0.000 |
| | | 11975 | Residential setting | Small town | Rural | 0.45 | 0.06 | 0.33 to 0.58 | 0.000 |
| | | 11975 | Residential setting | Large city | Rural | 0.41 | 0.08 | 0.26 to 0.56 | 0.000 |
| | | 11975 | Residential setting | Suburb | Rural | 0.14 | 0.13 | -0.11 to 0.38 | 0.279 |
| **Feel close to someone** | **Logistic regression** | 12402 | *Intercept* | | | 1.66 | 0.14 | 1.40 to 1.93 | 0.000 |
| | | 12402 | Childhood predictor | No | Yes | -0.16 | 0.22 | -0.59 to 0.27 | 0.466 |
| | | 12402 | Age | Per 1-year increase | | -0.01 | 0.00 | -0.01 to 0.00 | 0.017 |
| | | 12402 | Gender | Female | Male | 0.02 | 0.07 | -0.10 to 0.15 | 0.706 |
| | | 12402 | Education | Secondary | Elementary or less | 0.16 | 0.08 | 0.00 to 0.32 | 0.051 |
| | | 12402 | Education | Four years beyond HS | Elementary or less | 0.14 | 0.13 | -0.11 to 0.40 | 0.277 |
| | | 12402 | Residential setting | Small town | Rural | 0.10 | 0.11 | -0.12 to 0.31 | 0.381 |
| | | 12402 | Residential setting | Large city | Rural | -0.11 | 0.13 | -0.36 to 0.14 | 0.404 |
| | | 12402 | Residential setting | Suburb | Rural | -0.19 | 0.19 | -0.57 to 0.19 | 0.335 |
| **Donated to charity** | **Logistic regression** | 12368 | *Intercept* | | | -0.34 | 0.13 | -0.59 to -0.09 | 0.007 |
| | | 12368 | Childhood predictor | No | Yes | -0.19 | 0.20 | -0.57 to 0.20 | 0.343 |
| | | 12368 | Age | Per 1-year increase | | 0.00 | 0.00 | 0.00 to 0.00 | 0.571 |
| | | 12368 | Gender | Female | Male | -0.03 | 0.05 | -0.13 to 0.07 | 0.519 |

| Outcome | Model type | Analytic N | Variable | Level / comparison | Reference category | Coefficient | Standard error | 95% CI | p-value |
|---|---|---|---|---|---|---|---|---|---|
| | | 12368 | Education | Secondary | Elementary or less | -0.07 | 0.06 | -0.19 to 0.04 | 0.213 |
| | | 12368 | Education | Four years beyond HS | Elementary or less | -0.15 | 0.10 | -0.35 to 0.04 | 0.129 |
| | | 12368 | Residential setting | Small town | Rural | -0.05 | 0.09 | -0.23 to 0.13 | 0.570 |
| | | 12368 | Residential setting | Large city | Rural | -0.42 | 0.11 | -0.64 to -0.20 | 0.000 |
| | | 12368 | Residential setting | Suburb | Rural | -0.16 | 0.19 | -0.53 to 0.20 | 0.374 |
| **Have any health problems** | **Logistic regression** | 12324 | *Intercept* | | | -2.03 | 0.10 | -2.23 to -1.83 | 0.000 |
| | | 12324 | Childhood predictor | No | Yes | -0.12 | 0.21 | -0.53 to 0.30 | 0.589 |
| | | 12324 | Age | Per 1-year increase | | 0.03 | 0.00 | 0.02 to 0.03 | 0.000 |
| | | 12324 | Gender | Female | Male | 0.23 | 0.05 | 0.13 to 0.33 | 0.000 |
| | | 12324 | Education | Secondary | Elementary or less | -0.15 | 0.06 | -0.27 to -0.04 | 0.010 |
| | | 12324 | Education | Four years beyond HS | Elementary or less | -0.27 | 0.09 | -0.45 to -0.08 | 0.005 |
| | | 12324 | Residential setting | Small town | Rural | 0.13 | 0.06 | 0.00 to 0.26 | 0.048 |
| | | 12324 | Residential setting | Large city | Rural | -0.14 | 0.09 | -0.31 to 0.04 | 0.122 |
| | | 12324 | Residential setting | Suburb | Rural | 0.09 | 0.14 | -0.18 to 0.36 | 0.503 |
| **Helped a stranger** | **Logistic regression** | 12280 | *Intercept* | | | 0.66 | 0.12 | 0.43 to 0.89 | 0.000 |
| | | 12280 | Childhood predictor | No | Yes | -0.09 | 0.20 | -0.48 to 0.31 | 0.665 |
| | | 12280 | Age | Per 1-year increase | | -0.01 | 0.00 | -0.01 to 0.00 | 0.000 |
| | | 12280 | Gender | Female | Male | -0.33 | 0.05 | -0.42 to -0.24 | 0.000 |
| | | 12280 | Education | Secondary | Elementary or less | -0.07 | 0.06 | -0.18 to 0.05 | 0.250 |
| | | 12280 | Education | Four years beyond HS | Elementary or less | -0.07 | 0.09 | -0.25 to 0.10 | 0.413 |
| | | 12280 | Residential setting | Small town | Rural | 0.10 | 0.09 | -0.07 to 0.27 | 0.229 |
| | | 12280 | Residential setting | Large city | Rural | -0.16 | 0.10 | -0.36 to 0.04 | 0.108 |
| | | 12280 | Residential setting | Suburb | Rural | 0.05 | 0.18 | -0.30 to 0.40 | 0.785 |
| **Volunteered at an organization** | **Logistic regression** | 12350 | *Intercept* | | | -0.31 | 0.14 | -0.57 to -0.04 | 0.026 |
| | | 12350 | Childhood predictor | No | Yes | 0.01 | 0.21 | -0.40 to 0.42 | 0.959 |
| | | 12350 | Age | Per 1-year increase | | -0.01 | 0.00 | -0.01 to 0.00 | 0.001 |
| | | 12350 | Gender | Female | Male | -0.28 | 0.05 | -0.38 to -0.17 | 0.000 |
| | | 12350 | Education | Secondary | Elementary or less | -0.14 | 0.06 | -0.26 to -0.01 | 0.033 |
| | | 12350 | Education | Four years beyond HS | Elementary or less | 0.02 | 0.10 | -0.18 to 0.22 | 0.833 |
| | | 12350 | Residential setting | Small town | Rural | -0.10 | 0.11 | -0.31 to 0.11 | 0.355 |
| | | 12350 | Residential setting | Large city | Rural | -0.57 | 0.13 | -0.82 to -0.32 | 0.000 |
| | | 12350 | Residential setting | Suburb | Rural | -0.39 | 0.17 | -0.71 to -0.06 | 0.021 |
| **Cigarette smoking** | **Logistic regression** | 11917 | *Intercept* | | | -2.00 | 0.16 | -2.32 to -1.69 | 0.000 |
| | | 11917 | Childhood predictor | No | Yes | 0.50 | 0.34 | -0.17 to 1.16 | 0.148 |
| | | 11917 | Age | Per 1-year increase | | 0.01 | 0.00 | 0.00 to 0.01 | 0.016 |
| | | 11917 | Gender | Female | Male | -2.82 | 0.19 | -3.19 to -2.46 | 0.000 |
| | | 11917 | Education | Secondary | Elementary or less | -0.39 | 0.10 | -0.58 to -0.20 | 0.000 |
| | | 11917 | Education | Four years beyond HS | Elementary or less | -0.56 | 0.17 | -0.89 to -0.24 | 0.001 |
| | | 11917 | Residential setting | Small town | Rural | -0.06 | 0.11 | -0.27 to 0.14 | 0.559 |
| | | 11917 | Residential setting | Large city | Rural | 0.12 | 0.15 | -0.17 to 0.41 | 0.408 |

| Outcome | Model type | Analytic N | Variable | Level / comparison | Reference category | Coefficient | Standard error | 95% CI | p-value |
|---|---|---|---|---|---|---|---|---|---|
| | | 11917 | Residential setting | Suburb | Rural | 0.83 | 0.20 | 0.43 to 1.23 | 0.000 |
| **Alcohol use** | **Logistic regression** | 11904 | *Intercept* | | | -1.87 | 0.15 | -2.16 to -1.58 | 0.000 |
| | | 11904 | Childhood predictor | No | Yes | 0.75 | 0.29 | 0.17 to 1.33 | 0.012 |
| | | 11904 | Age | Per 1-year increase | | 0.00 | 0.00 | -0.01 to 0.00 | 0.680 |
| | | 11904 | Gender | Female | Male | -2.79 | 0.16 | -3.10 to -2.48 | 0.000 |
| | | 11904 | Education | Secondary | Elementary or less | -0.23 | 0.10 | -0.42 to -0.03 | 0.025 |
| | | 11904 | Education | Four years beyond HS | Elementary or less | -0.48 | 0.16 | -0.79 to -0.16 | 0.003 |
| | | 11904 | Residential setting | Small town | Rural | 0.23 | 0.11 | 0.03 to 0.44 | 0.027 |
| | | 11904 | Residential setting | Large city | Rural | 0.36 | 0.14 | 0.08 to 0.63 | 0.012 |
| | | 11904 | Residential setting | Suburb | Rural | 0.65 | 0.22 | 0.21 to 1.09 | 0.004 |
| **Government approval§** | **Proportional-odds regression** | 12192 | Childhood predictor | No | Yes | -0.21 | 0.17 | -0.54 to 0.13 | 0.222 |
| | | 12192 | Age | Per 1-year increase | | 0.00 | 0.00 | -0.01 to 0.00 | 0.020 |
| | | 12192 | Gender | Female | Male | 0.06 | 0.04 | -0.03 to 0.14 | 0.199 |
| | | 12192 | Education | Secondary | Elementary or less | -0.25 | 0.05 | -0.36 to -0.15 | 0.000 |
| | | 12192 | Education | Four years beyond HS | Elementary or less | -0.34 | 0.08 | -0.51 to -0.18 | 0.000 |
| | | 12192 | Residential setting | Small town | Rural | 0.11 | 0.07 | -0.02 to 0.24 | 0.101 |
| | | 12192 | Residential setting | Large city | Rural | -0.06 | 0.08 | -0.21 to 0.09 | 0.417 |
| | | 12192 | Residential setting | Suburb | Rural | -0.44 | 0.16 | -0.75 to -0.14 | 0.005 |
| | | 12192 | *Threshold / cut-point* | Strongly disapprove\|Somewhat disapprove | | -2.93 | 0.10 | -3.13 to -2.74 | 0.000 |
| | | 12192 | *Threshold / cut-point* | Somewhat disapprove\|Neither | | -2.27 | 0.09 | -2.45 to -2.08 | 0.000 |
| | | 12192 | *Threshold / cut-point* | Neither\|Somewhat approve | | -1.91 | 0.09 | -2.09 to -1.73 | 0.000 |
| | | 12192 | *Threshold / cut-point* | Somewhat approve\|Strongly approve | | -0.46 | 0.09 | -0.63 to -0.28 | 0.000 |
| **Feeling capable§** | **Proportional-odds regression** | 12348 | Childhood predictor | No | Yes | 0.11 | 0.20 | -0.28 to 0.50 | 0.576 |
| | | 12348 | Age | Per 1-year increase | | 0.00 | 0.00 | 0.00 to 0.01 | 0.173 |
| | | 12348 | Gender | Female | Male | -0.06 | 0.04 | -0.15 to 0.02 | 0.142 |
| | | 12348 | Education | Secondary | Elementary or less | 0.16 | 0.05 | 0.07 to 0.26 | 0.001 |
| | | 12348 | Education | Four years beyond HS | Elementary or less | 0.10 | 0.07 | -0.04 to 0.24 | 0.170 |
| | | 12348 | Residential setting | Small town | Rural | 0.17 | 0.05 | 0.07 to 0.27 | 0.001 |
| | | 12348 | Residential setting | Large city | Rural | 0.19 | 0.08 | 0.04 to 0.34 | 0.014 |
| | | 12348 | Residential setting | Suburb | Rural | 0.23 | 0.09 | 0.05 to 0.40 | 0.010 |
| | | 12348 | *Threshold / cut-point* | Never\|Rarely | | -2.35 | 0.09 | -2.52 to -2.17 | 0.000 |
| | | 12348 | *Threshold / cut-point* | Rarely\|Often | | -0.62 | 0.08 | -0.79 to -0.46 | 0.000 |
| | | 12348 | *Threshold / cut-point* | Often\|Always | | 0.05 | 0.08 | -0.12 to 0.21 | 0.576 |
| **Unable to control worry§** | **Proportional-odds regression** | 12298 | Childhood predictor | No | Yes | 0.03 | 0.16 | -0.28 to 0.35 | 0.833 |
| | | 12298 | Age | Per 1-year increase | | 0.01 | 0.00 | 0.00 to 0.01 | 0.001 |
| | | 12298 | Gender | Female | Male | 0.16 | 0.04 | 0.08 to 0.24 | 0.000 |

| Outcome | Model type | Analytic N | Variable | Level / comparison | Reference category | Coefficient | Standard error | 95% CI | p-value |
|---|---|---|---|---|---|---|---|---|---|
| | | 12298 | Education | Secondary | Elementary or less | -0.27 | 0.05 | -0.36 to -0.18 | 0.000 |
| | | 12298 | Education | Four years beyond HS | Elementary or less | -0.44 | 0.07 | -0.58 to -0.29 | 0.000 |
| | | 12298 | Residential setting | Small town | Rural | 0.05 | 0.05 | -0.05 to 0.14 | 0.337 |
| | | 12298 | Residential setting | Large city | Rural | -0.01 | 0.07 | -0.14 to 0.13 | 0.930 |
| | | 12298 | Residential setting | Suburb | Rural | -0.07 | 0.10 | -0.26 to 0.13 | 0.490 |
| | | 12298 | *Threshold / cut-point* | Not at all\|Several days | | 0.08 | 0.08 | -0.08 to 0.23 | 0.339 |
| | | 12298 | *Threshold / cut-point* | Several days\|More than half the days | | 0.98 | 0.08 | 0.83 to 1.14 | 0.000 |
| | | 12298 | *Threshold / cut-point* | More than half the days\|Nearly every day | | 1.83 | 0.08 | 1.67 to 1.99 | 0.000 |
| **Depressive symptoms§** | **Proportional-odds regression** | 12324 | Childhood predictor | No | Yes | 0.22 | 0.17 | -0.12 to 0.55 | 0.205 |
| | | 12324 | Age | Per 1-year increase | | 0.00 | 0.00 | 0.00 to 0.01 | 0.117 |
| | | 12324 | Gender | Female | Male | 0.09 | 0.04 | 0.01 to 0.16 | 0.025 |
| | | 12324 | Education | Secondary | Elementary or less | -0.21 | 0.05 | -0.30 to -0.11 | 0.000 |
| | | 12324 | Education | Four years beyond HS | Elementary or less | -0.42 | 0.07 | -0.55 to -0.28 | 0.000 |
| | | 12324 | Residential setting | Small town | Rural | 0.00 | 0.05 | -0.10 to 0.11 | 0.978 |
| | | 12324 | Residential setting | Large city | Rural | -0.09 | 0.07 | -0.23 to 0.05 | 0.202 |
| | | 12324 | Residential setting | Suburb | Rural | -0.17 | 0.11 | -0.39 to 0.06 | 0.143 |
| | | 12324 | *Threshold / cut-point* | Not at all\|Several days | | -0.24 | 0.08 | -0.39 to -0.08 | 0.003 |
| | | 12324 | *Threshold / cut-point* | Several days\|More than half the days | | 0.83 | 0.08 | 0.68 to 0.99 | 0.000 |
| | | 12324 | *Threshold / cut-point* | More than half the days\|Nearly every day | | 1.83 | 0.08 | 1.67 to 1.99 | 0.000 |
| **Perceived discrimination§** | **Proportional-odds regression** | 12257 | Childhood predictor | No | Yes | 0.02 | 0.16 | -0.29 to 0.33 | 0.899 |
| | | 12257 | Age | Per 1-year increase | | 0.00 | 0.00 | -0.01 to 0.00 | 0.077 |
| | | 12257 | Gender | Female | Male | -0.14 | 0.04 | -0.23 to -0.06 | 0.001 |
| | | 12257 | Education | Secondary | Elementary or less | -0.24 | 0.05 | -0.34 to -0.14 | 0.000 |
| | | 12257 | Education | Four years beyond HS | Elementary or less | -0.29 | 0.09 | -0.46 to -0.12 | 0.001 |
| | | 12257 | Residential setting | Small town | Rural | -0.22 | 0.06 | -0.33 to -0.11 | 0.000 |
| | | 12257 | Residential setting | Large city | Rural | -0.23 | 0.07 | -0.37 to -0.09 | 0.001 |
| | | 12257 | Residential setting | Suburb | Rural | -0.34 | 0.10 | -0.54 to -0.15 | 0.001 |
| | | 12257 | *Threshold / cut-point* | Never\|Rarely | | -0.13 | 0.09 | -0.30 to 0.04 | 0.138 |
| | | 12257 | *Threshold / cut-point* | Rarely\|Often | | 0.87 | 0.09 | 0.70 to 1.05 | 0.000 |
| | | 12257 | *Threshold / cut-point* | Often\|Always | | 1.49 | 0.09 | 1.32 to 1.67 | 0.000 |
| **Feeling nervous or anxious§** | **Proportional-odds regression** | 12350 | Childhood predictor | No | Yes | 0.11 | 0.18 | -0.24 to 0.46 | 0.535 |
| | | 12350 | Age | Per 1-year increase | | 0.01 | 0.00 | 0.00 to 0.01 | 0.000 |
| | | 12350 | Gender | Female | Male | 0.31 | 0.04 | 0.23 to 0.38 | 0.000 |
| | | 12350 | Education | Secondary | Elementary or less | -0.29 | 0.05 | -0.38 to -0.19 | 0.000 |
| | | 12350 | Education | Four years beyond HS | Elementary or less | -0.48 | 0.08 | -0.63 to -0.33 | 0.000 |

| Outcome | Model type | Analytic N | Variable | Level / comparison | Reference category | Coefficient | Standard error | 95% CI | p-value |
|---|---|---|---|---|---|---|---|---|---|
| | | 12350 | Residential setting | Small town | Rural | 0.11 | 0.05 | 0.00 to 0.22 | 0.043 |
| | | 12350 | Residential setting | Large city | Rural | -0.03 | 0.07 | -0.17 to 0.10 | 0.637 |
| | | 12350 | Residential setting | Suburb | Rural | -0.04 | 0.12 | -0.27 to 0.19 | 0.728 |
| | | 12350 | *Threshold / cut-point* | Not at all\|Several days | | 0.14 | 0.08 | -0.02 to 0.29 | 0.082 |
| | | 12350 | *Threshold / cut-point* | Several days\|More than half the days | | 1.13 | 0.08 | 0.98 to 1.28 | 0.000 |
| | | 12350 | *Threshold / cut-point* | More than half the days\|Nearly every day | | 2.07 | 0.08 | 1.91 to 2.23 | 0.000 |
| **Forgiveness§** | **Proportional-odds regression** | 12324 | Childhood predictor | No | Yes | -0.09 | 0.19 | -0.46 to 0.28 | 0.636 |
| | | 12324 | Age | Per 1-year increase | | 0.00 | 0.00 | 0.00 to 0.00 | 0.891 |
| | | 12324 | Gender | Female | Male | 0.09 | 0.04 | 0.01 to 0.17 | 0.038 |
| | | 12324 | Education | Secondary | Elementary or less | -0.07 | 0.05 | -0.17 to 0.03 | 0.191 |
| | | 12324 | Education | Four years beyond HS | Elementary or less | -0.20 | 0.08 | -0.35 to -0.05 | 0.010 |
| | | 12324 | Residential setting | Small town | Rural | 0.04 | 0.05 | -0.07 to 0.14 | 0.504 |
| | | 12324 | Residential setting | Large city | Rural | 0.02 | 0.08 | -0.14 to 0.17 | 0.850 |
| | | 12324 | Residential setting | Suburb | Rural | 0.04 | 0.12 | -0.19 to 0.27 | 0.758 |
| | | 12324 | *Threshold / cut-point* | Never\|Rarely | | -2.39 | 0.09 | -2.57 to -2.22 | 0.000 |
| | | 12324 | *Threshold / cut-point* | Rarely\|Often | | -1.06 | 0.08 | -1.23 to -0.89 | 0.000 |
| | | 12324 | *Threshold / cut-point* | Often\|Always | | -0.41 | 0.08 | -0.57 to -0.25 | 0.000 |
| **Support reducing differences in income levels§** | **Proportional-odds regression** | 12186 | Childhood predictor | No | Yes | -0.64 | 0.24 | -1.10 to -0.18 | 0.006 |
| | | 12186 | Age | Per 1-year increase | | 0.00 | 0.00 | -0.01 to 0.00 | 0.089 |
| | | 12186 | Gender | Female | Male | -0.05 | 0.06 | -0.16 to 0.06 | 0.405 |
| | | 12186 | Education | Secondary | Elementary or less | -0.07 | 0.07 | -0.21 to 0.07 | 0.309 |
| | | 12186 | Education | Four years beyond HS | Elementary or less | -0.29 | 0.11 | -0.50 to -0.07 | 0.010 |
| | | 12186 | Residential setting | Small town | Rural | 0.03 | 0.13 | -0.22 to 0.28 | 0.816 |
| | | 12186 | Residential setting | Large city | Rural | 0.09 | 0.14 | -0.19 to 0.37 | 0.521 |
| | | 12186 | Residential setting | Suburb | Rural | -0.09 | 0.18 | -0.44 to 0.26 | 0.615 |
| | | 12186 | *Threshold / cut-point* | Strongly disagree\|Somewhat disagree | | -3.90 | 0.17 | -4.24 to -3.57 | 0.000 |
| | | 12186 | *Threshold / cut-point* | Somewhat disagree\|Neither | | -3.35 | 0.16 | -3.67 to -3.03 | 0.000 |
| | | 12186 | *Threshold / cut-point* | Neither\|Somewhat agree | | -2.90 | 0.16 | -3.21 to -2.59 | 0.000 |
| | | 12186 | *Threshold / cut-point* | Somewhat agree\|Strongly agree | | -1.32 | 0.14 | -1.59 to -1.04 | 0.000 |
| **Have little interest in doing things§** | **Proportional-odds regression** | 12319 | Childhood predictor | No | Yes | 0.11 | 0.15 | -0.17 to 0.40 | 0.442 |
| | | 12319 | Age | Per 1-year increase | | 0.00 | 0.00 | 0.00 to 0.01 | 0.053 |
| | | 12319 | Gender | Female | Male | 0.04 | 0.04 | -0.04 to 0.12 | 0.380 |
| | | 12319 | Education | Secondary | Elementary or less | 0.07 | 0.05 | -0.02 to 0.17 | 0.126 |
| | | 12319 | Education | Four years beyond HS | Elementary or less | 0.08 | 0.08 | -0.07 to 0.24 | 0.280 |
| | | 12319 | Residential setting | Small town | Rural | 0.28 | 0.06 | 0.15 to 0.40 | 0.000 |

| Outcome | Model type | Analytic N | Variable | Level / comparison | Reference category | Coefficient | Standard error | 95% CI | p-value |
|---|---|---|---|---|---|---|---|---|---|
| | | 12319 | Residential setting | Large city | Rural | 0.10 | 0.08 | -0.06 to 0.26 | 0.208 |
| | | 12319 | Residential setting | Suburb | Rural | 0.25 | 0.14 | -0.03 to 0.53 | 0.080 |
| | | 12319 | *Threshold / cut-point* | Not at all\|Several days | | -0.49 | 0.09 | -0.66 to -0.32 | 0.000 |
| | | 12319 | *Threshold / cut-point* | Several days\|More than half the days | | 0.44 | 0.09 | 0.27 to 0.62 | 0.000 |
| | | 12319 | *Threshold / cut-point* | More than half the days\|Nearly every day | | 1.22 | 0.09 | 1.05 to 1.40 | 0.000 |
| **Various aspects of life are balanced§** | **Proportional-odds regression** | 12245 | Childhood predictor | No | Yes | 0.33 | 0.16 | 0.02 to 0.63 | 0.035 |
| | | 12245 | Age | Per 1-year increase | | 0.00 | 0.00 | 0.00 to 0.01 | 0.098 |
| | | 12245 | Gender | Female | Male | -0.03 | 0.04 | -0.11 to 0.05 | 0.512 |
| | | 12245 | Education | Secondary | Elementary or less | 0.04 | 0.04 | -0.05 to 0.13 | 0.375 |
| | | 12245 | Education | Four years beyond HS | Elementary or less | 0.11 | 0.07 | -0.03 to 0.24 | 0.122 |
| | | 12245 | Residential setting | Small town | Rural | 0.10 | 0.05 | 0.00 to 0.20 | 0.061 |
| | | 12245 | Residential setting | Large city | Rural | 0.09 | 0.07 | -0.04 to 0.22 | 0.166 |
| | | 12245 | Residential setting | Suburb | Rural | 0.35 | 0.11 | 0.14 to 0.57 | 0.001 |
| | | 12245 | *Threshold / cut-point* | Never\|Rarely | | -2.03 | 0.09 | -2.20 to -1.86 | 0.000 |
| | | 12245 | *Threshold / cut-point* | Rarely\|Often | | 0.15 | 0.08 | -0.01 to 0.31 | 0.068 |
| | | 12245 | *Threshold / cut-point* | Often\|Always | | 0.94 | 0.08 | 0.78 to 1.10 | 0.000 |
| **Obeying the law§** | **Proportional-odds regression** | 12373 | Childhood predictor | No | Yes | -0.78 | 0.25 | -1.27 to -0.30 | 0.002 |
| | | 12373 | Age | Per 1-year increase | | 0.00 | 0.00 | -0.01 to 0.00 | 0.397 |
| | | 12373 | Gender | Female | Male | -0.14 | 0.07 | -0.29 to 0.00 | 0.056 |
| | | 12373 | Education | Secondary | Elementary or less | 0.10 | 0.10 | -0.08 to 0.29 | 0.279 |
| | | 12373 | Education | Four years beyond HS | Elementary or less | -0.23 | 0.12 | -0.47 to 0.01 | 0.066 |
| | | 12373 | Residential setting | Small town | Rural | -0.06 | 0.15 | -0.35 to 0.23 | 0.689 |
| | | 12373 | Residential setting | Large city | Rural | -0.24 | 0.18 | -0.60 to 0.12 | 0.194 |
| | | 12373 | Residential setting | Suburb | Rural | -0.29 | 0.22 | -0.73 to 0.15 | 0.202 |
| | | 12373 | *Threshold / cut-point* | Strongly disagree\|Somewhat disagree | | -5.21 | 0.26 | -5.72 to -4.71 | 0.000 |
| | | 12373 | *Threshold / cut-point* | Somewhat disagree\|Neither | | -4.60 | 0.22 | -5.02 to -4.17 | 0.000 |
| | | 12373 | *Threshold / cut-point* | Neither\|Somewhat agree | | -4.08 | 0.21 | -4.50 to -3.67 | 0.000 |
| | | 12373 | *Threshold / cut-point* | Somewhat agree\|Strongly agree | | -2.23 | 0.19 | -2.60 to -1.86 | 0.000 |
| **Feel at peace§** | **Proportional-odds regression** | 12319 | Childhood predictor | No | Yes | 0.00 | 0.14 | -0.27 to 0.28 | 0.986 |
| | | 12319 | Age | Per 1-year increase | | 0.01 | 0.00 | 0.00 to 0.01 | 0.000 |
| | | 12319 | Gender | Female | Male | 0.01 | 0.04 | -0.06 to 0.08 | 0.799 |
| | | 12319 | Education | Secondary | Elementary or less | 0.02 | 0.05 | -0.07 to 0.11 | 0.721 |
| | | 12319 | Education | Four years beyond HS | Elementary or less | 0.05 | 0.07 | -0.08 to 0.18 | 0.441 |
| | | 12319 | Residential setting | Small town | Rural | 0.21 | 0.05 | 0.10 to 0.32 | 0.000 |
| | | 12319 | Residential setting | Large city | Rural | 0.16 | 0.07 | 0.01 to 0.30 | 0.033 |

| Outcome | Model type | Analytic N | Variable | Level / comparison | Reference category | Coefficient | Standard error | 95% CI | p-value |
|---|---|---|---|---|---|---|---|---|---|
| | | 12319 | Residential setting | Suburb | Rural | 0.24 | 0.10 | 0.04 to 0.44 | 0.016 |
| | | 12319 | *Threshold / cut-point* | Never\|Rarely | | -1.88 | 0.08 | -2.04 to -1.72 | 0.000 |
| | | 12319 | *Threshold / cut-point* | Rarely\|Often | | -0.05 | 0.08 | -0.21 to 0.10 | 0.509 |
| | | 12319 | *Threshold / cut-point* | Often\|Always | | 0.64 | 0.08 | 0.48 to 0.80 | 0.000 |
| **Say in government#** | **Proportional-odds regression** | 12351 | Childhood predictor | No | Yes | -0.41 | 0.21 | -0.82 to 0.00 | 0.048 |
| | | 12351 | Age | Per 1-year increase | | 0.00 | 0.00 | -0.01 to 0.00 | 0.446 |
| | | 12351 | Gender | Female | Male | 0.10 | 0.05 | 0.00 to 0.20 | 0.045 |
| | | 12351 | Education | Secondary | Elementary or less | -0.24 | 0.06 | -0.36 to -0.12 | 0.000 |
| | | 12351 | Education | Four years beyond HS | Elementary or less | -0.14 | 0.10 | -0.33 to 0.06 | 0.159 |
| | | 12351 | Residential setting | Small town | Rural | 0.01 | 0.08 | -0.14 to 0.16 | 0.898 |
| | | 12351 | Residential setting | Large city | Rural | -0.14 | 0.09 | -0.32 to 0.04 | 0.132 |
| | | 12351 | Residential setting | Suburb | Rural | -0.45 | 0.13 | -0.70 to -0.19 | 0.001 |
| | | 12351 | *Threshold / cut-point* | Disagree\|Unsure | | -1.79 | 0.11 | -2.01 to -1.57 | 0.000 |
| | | 12351 | *Threshold / cut-point* | Unsure\|Agree | | -1.39 | 0.11 | -1.61 to -1.18 | 0.000 |
| **Extent of suffering§** | **Proportional-odds regression** | 12326 | Childhood predictor | No | Yes | 0.36 | 0.19 | -0.02 to 0.73 | 0.065 |
| | | 12326 | Age | Per 1-year increase | | 0.02 | 0.00 | 0.02 to 0.02 | 0.000 |
| | | 12326 | Gender | Female | Male | 0.27 | 0.04 | 0.19 to 0.35 | 0.000 |
| | | 12326 | Education | Secondary | Elementary or less | -0.15 | 0.05 | -0.23 to -0.06 | 0.001 |
| | | 12326 | Education | Four years beyond HS | Elementary or less | -0.27 | 0.07 | -0.42 to -0.12 | 0.000 |
| | | 12326 | Residential setting | Small town | Rural | 0.05 | 0.06 | -0.06 to 0.16 | 0.393 |
| | | 12326 | Residential setting | Large city | Rural | -0.13 | 0.08 | -0.28 to 0.02 | 0.095 |
| | | 12326 | Residential setting | Suburb | Rural | 0.07 | 0.11 | -0.15 to 0.29 | 0.530 |
| | | 12326 | *Threshold / cut-point* | Not at all\|Not very much | | 0.31 | 0.08 | 0.15 to 0.47 | 0.000 |
| | | 12326 | *Threshold / cut-point* | Not very much\|Some | | 0.72 | 0.08 | 0.56 to 0.89 | 0.000 |
| | | 12326 | *Threshold / cut-point* | Some\|A lot | | 2.17 | 0.09 | 2.00 to 2.34 | 0.000 |
| **Distress from life-threatening experiences§** | **Proportional-odds regression** | 12345 | Childhood predictor | No | Yes | 0.11 | 0.17 | -0.23 to 0.45 | 0.539 |
| | | 12345 | Age | Per 1-year increase | | 0.00 | 0.00 | 0.00 to 0.01 | 0.021 |
| | | 12345 | Gender | Female | Male | 0.08 | 0.05 | -0.01 to 0.16 | 0.096 |
| | | 12345 | Education | Secondary | Elementary or less | -0.27 | 0.05 | -0.36 to -0.18 | 0.000 |
| | | 12345 | Education | Four years beyond HS | Elementary or less | -0.45 | 0.07 | -0.59 to -0.30 | 0.000 |
| | | 12345 | Residential setting | Small town | Rural | -0.17 | 0.05 | -0.27 to -0.08 | 0.000 |
| | | 12345 | Residential setting | Large city | Rural | -0.23 | 0.07 | -0.37 to -0.09 | 0.002 |
| | | 12345 | Residential setting | Suburb | Rural | -0.17 | 0.12 | -0.39 to 0.06 | 0.151 |
| | | 12345 | *Threshold / cut-point* | Not at all\|Not very much | | -0.30 | 0.08 | -0.45 to -0.15 | 0.000 |
| | | 12345 | *Threshold / cut-point* | Not very much\|Some | | 0.04 | 0.08 | -0.11 to 0.19 | 0.580 |
| | | 12345 | *Threshold / cut-point* | Some\|A lot | | 1.06 | 0.08 | 0.91 to 1.21 | 0.000 |
| **Trust in others§** | **Proportional-odds regression** | 12068 | Childhood predictor | No | Yes | -0.03 | 0.18 | -0.38 to 0.31 | 0.847 |

| Outcome | Model type | Analytic N | Variable | Level / comparison | Reference category | Coefficient | Standard error | 95% CI | p-value |
|---|---|---|---|---|---|---|---|---|---|
| | | 12068 | Age | Per 1-year increase | | 0.00 | 0.00 | 0.00 to 0.01 | 0.034 |
| | | 12068 | Gender | Female | Male | -0.04 | 0.04 | -0.12 to 0.04 | 0.311 |
| | | 12068 | Education | Secondary | Elementary or less | -0.04 | 0.05 | -0.13 to 0.05 | 0.403 |
| | | 12068 | Education | Four years beyond HS | Elementary or less | -0.05 | 0.07 | -0.19 to 0.09 | 0.468 |
| | | 12068 | Residential setting | Small town | Rural | 0.24 | 0.05 | 0.14 to 0.34 | 0.000 |
| | | 12068 | Residential setting | Large city | Rural | 0.00 | 0.07 | -0.13 to 0.13 | 0.976 |
| | | 12068 | Residential setting | Suburb | Rural | -0.23 | 0.13 | -0.49 to 0.04 | 0.092 |
| | | 12068 | *Threshold / cut-point* | None\|Not very many | | -2.15 | 0.09 | -2.33 to -1.96 | 0.000 |
| | | 12068 | *Threshold / cut-point* | Not very many\|Some | | -1.36 | 0.09 | -1.54 to -1.18 | 0.000 |
| | | 12068 | *Threshold / cut-point* | Some\|Most | | 0.84 | 0.09 | 0.67 to 1.01 | 0.000 |
| | | 12068 | *Threshold / cut-point* | Most\|All | | 1.82 | 0.09 | 1.64 to 1.99 | 0.000 |

**eTable 10.** Table of coefficients for adjusted associations between not feeling loved by father as predictor and 45 outcomes of interest and two flourishing composite indices.

Adjusted survey-weighted models use ANNUAL_WEIGHT_C1 with primary sampling unit (PSU) and strata and adjust for age, gender, education, and residential setting. Reference categories are shown in the table. Coefficients are on the original regression-coefficient scale: beta for continuous outcomes, log-odds for binary outcomes, and proportional-odds log coefficients for ordinal outcomes. Model intercepts are displayed for continuous and binary outcomes. Proportional-odds models do not have a single conventional intercept; their estimated ordinal cut-points/thresholds are displayed after the covariate coefficients, with standard errors (SEs), 95% CIs, and Wald p-values.

***Note:*** All results except p-values are rounded to the two decimals. P-values are rounded to three decimals. Values displayed as 0.000 indicate $P < 0.0005$ due to rounding, rather than an exact zero. Statistical significance was determined using unrounded P-values at the Bonferroni-adjusted threshold of $P < 0.0011$.

| Outcome | Model type | Analytic N | Variable | Level / comparison | Reference category | Coefficient | Standard error | 95% CI | p-value |
|---|---|---|---|---|---|---|---|---|---|
| **Belonging** | **Linear regression** | 12052 | *Intercept* | | | 8.68 | 0.11 | 8.45 to 8.90 | 0.000 |
| | | 12052 | Childhood predictor | No | Yes | -0.49 | 0.22 | -0.92 to -0.05 | 0.029 |
| | | 12052 | Age | Per 1-year increase | | -0.01 | 0.00 | -0.02 to -0.01 | 0.000 |
| | | 12052 | Gender | Female | Male | -0.19 | 0.06 | -0.31 to -0.07 | 0.002 |
| | | 12052 | Education | Secondary | Elementary or less | 0.20 | 0.08 | 0.05 to 0.35 | 0.008 |
| | | 12052 | Education | Four years beyond HS | Elementary or less | 0.32 | 0.11 | 0.12 to 0.53 | 0.002 |
| | | 12052 | Residential setting | Small town | Rural | 0.37 | 0.08 | 0.21 to 0.53 | 0.000 |
| | | 12052 | Residential setting | Large city | Rural | 0.44 | 0.11 | 0.23 to 0.64 | 0.000 |
| | | 12052 | Residential setting | Suburb | Rural | 0.01 | 0.18 | -0.35 to 0.38 | 0.936 |
| **Relationship contentment‡** | **Linear regression** | 12114 | *Intercept* | | | 8.14 | 0.13 | 7.89 to 8.39 | 0.000 |
| | | 12114 | Childhood predictor | No | Yes | -0.88 | 0.25 | -1.36 to -0.39 | 0.000 |
| | | 12114 | Age | Per 1-year increase | | -0.01 | 0.00 | -0.02 to -0.01 | 0.000 |
| | | 12114 | Gender | Female | Male | 0.38 | 0.07 | 0.24 to 0.51 | 0.000 |
| | | 12114 | Education | Secondary | Elementary or less | 0.16 | 0.07 | 0.01 to 0.30 | 0.035 |
| | | 12114 | Education | Four years beyond HS | Elementary or less | 0.29 | 0.12 | 0.06 to 0.52 | 0.015 |
| | | 12114 | Residential setting | Small town | Rural | 0.40 | 0.08 | 0.24 to 0.57 | 0.000 |
| | | 12114 | Residential setting | Large city | Rural | 0.09 | 0.10 | -0.10 to 0.29 | 0.346 |
| | | 12114 | Residential setting | Suburb | Rural | -0.26 | 0.18 | -0.61 to 0.09 | 0.140 |
| **Positive expectations** | **Linear regression** | 12076 | *Intercept* | | | 7.79 | 0.15 | 7.50 to 8.08 | 0.000 |
| | | 12076 | Childhood predictor | No | Yes | -0.29 | 0.23 | -0.74 to 0.15 | 0.197 |
| | | 12076 | Age | Per 1-year increase | | 0.00 | 0.00 | -0.01 to 0.00 | 0.182 |
| | | 12076 | Gender | Female | Male | 0.22 | 0.07 | 0.09 to 0.35 | 0.001 |
| | | 12076 | Education | Secondary | Elementary or less | 0.29 | 0.07 | 0.14 to 0.43 | 0.000 |
| | | 12076 | Education | Four years beyond HS | Elementary or less | 0.36 | 0.10 | 0.17 to 0.56 | 0.000 |
| | | 12076 | Residential setting | Small town | Rural | 0.45 | 0.09 | 0.28 to 0.61 | 0.000 |
| | | 12076 | Residential setting | Large city | Rural | 0.46 | 0.11 | 0.24 to 0.68 | 0.000 |
| | | 12076 | Residential setting | Suburb | Rural | 0.41 | 0.16 | 0.09 to 0.73 | 0.012 |
| **Worry about meeting normal monthly living expenses§§‡‡** | **Linear regression** | 12118 | *Intercept* | | | 5.72 | 0.19 | 5.34 to 6.10 | 0.000 |
| | | 12118 | Childhood predictor | No | Yes | 0.35 | 0.28 | -0.19 to 0.90 | 0.203 |
| | | 12118 | Age | Per 1-year increase | | 0.00 | 0.00 | 0.00 to 0.01 | 0.431 |
| | | 12118 | Gender | Female | Male | 0.37 | 0.09 | 0.19 to 0.54 | 0.000 |

| Outcome | Model type | Analytic N | Variable | Level / comparison | Reference category | Coefficient | Standard error | 95% CI | p-value |
|---|---|---|---|---|---|---|---|---|---|
| | | 12118 | Education | Secondary | Elementary or less | -0.42 | 0.11 | -0.64 to -0.21 | 0.000 |
| | | 12118 | Education | Four years beyond HS | Elementary or less | -0.62 | 0.15 | -0.92 to -0.32 | 0.000 |
| | | 12118 | Residential setting | Small town | Rural | 0.21 | 0.13 | -0.04 to 0.46 | 0.104 |
| | | 12118 | Residential setting | Large city | Rural | -0.21 | 0.15 | -0.50 to 0.09 | 0.171 |
| | | 12118 | Residential setting | Suburb | Rural | -0.20 | 0.25 | -0.69 to 0.29 | 0.427 |
| **Freedom** | **Linear regression** | 12090 | *Intercept* | | | 8.17 | 0.12 | 7.93 to 8.41 | 0.000 |
| | | 12090 | Childhood predictor | No | Yes | -0.57 | 0.22 | -1.00 to -0.14 | 0.010 |
| | | 12090 | Age | Per 1-year increase | | -0.01 | 0.00 | -0.01 to 0.00 | 0.004 |
| | | 12090 | Gender | Female | Male | -0.11 | 0.07 | -0.25 to 0.02 | 0.097 |
| | | 12090 | Education | Secondary | Elementary or less | 0.29 | 0.08 | 0.14 to 0.44 | 0.000 |
| | | 12090 | Education | Four years beyond HS | Elementary or less | 0.41 | 0.11 | 0.20 to 0.62 | 0.000 |
| | | 12090 | Residential setting | Small town | Rural | 0.39 | 0.08 | 0.24 to 0.54 | 0.000 |
| | | 12090 | Residential setting | Large city | Rural | 0.42 | 0.10 | 0.22 to 0.63 | 0.000 |
| | | 12090 | Residential setting | Suburb | Rural | 0.16 | 0.16 | -0.16 to 0.47 | 0.328 |
| **Give up some happiness now for greater happiness later‡** | **Linear regression** | 12058 | *Intercept* | | | 7.26 | 0.15 | 6.96 to 7.56 | 0.000 |
| | | 12058 | Childhood predictor | No | Yes | -0.18 | 0.23 | -0.64 to 0.28 | 0.444 |
| | | 12058 | Age | Per 1-year increase | | -0.01 | 0.00 | -0.02 to 0.00 | 0.000 |
| | | 12058 | Gender | Female | Male | 0.17 | 0.08 | 0.01 to 0.32 | 0.034 |
| | | 12058 | Education | Secondary | Elementary or less | 0.18 | 0.10 | -0.01 to 0.37 | 0.062 |
| | | 12058 | Education | Four years beyond HS | Elementary or less | 0.17 | 0.13 | -0.09 to 0.44 | 0.198 |
| | | 12058 | Residential setting | Small town | Rural | 0.38 | 0.10 | 0.19 to 0.58 | 0.000 |
| | | 12058 | Residential setting | Large city | Rural | 0.20 | 0.13 | -0.07 to 0.46 | 0.141 |
| | | 12058 | Residential setting | Suburb | Rural | 0.04 | 0.28 | -0.51 to 0.59 | 0.885 |
| **Have a long list of gratitude** | **Linear regression** | 11956 | *Intercept* | | | 7.48 | 0.15 | 7.18 to 7.77 | 0.000 |
| | | 11956 | Childhood predictor | No | Yes | -0.02 | 0.21 | -0.42 to 0.39 | 0.926 |
| | | 11956 | Age | Per 1-year increase | | -0.01 | 0.00 | -0.01 to 0.00 | 0.025 |
| | | 11956 | Gender | Female | Male | 0.19 | 0.07 | 0.05 to 0.33 | 0.007 |
| | | 11956 | Education | Secondary | Elementary or less | 0.28 | 0.08 | 0.12 to 0.44 | 0.001 |
| | | 11956 | Education | Four years beyond HS | Elementary or less | 0.37 | 0.12 | 0.15 to 0.60 | 0.001 |
| | | 11956 | Residential setting | Small town | Rural | 0.43 | 0.09 | 0.25 to 0.62 | 0.000 |
| | | 11956 | Residential setting | Large city | Rural | 0.44 | 0.14 | 0.17 to 0.71 | 0.001 |
| | | 11956 | Residential setting | Suburb | Rural | 0.48 | 0.17 | 0.14 to 0.81 | 0.006 |
| **Happiness‡** | **Linear regression** | 12115 | *Intercept* | | | 6.66 | 0.16 | 6.35 to 6.97 | 0.000 |
| | | 12115 | Childhood predictor | No | Yes | -0.70 | 0.28 | -1.26 to -0.15 | 0.014 |
| | | 12115 | Age | Per 1-year increase | | -0.02 | 0.00 | -0.02 to -0.01 | 0.000 |
| | | 12115 | Gender | Female | Male | 0.25 | 0.08 | 0.10 to 0.40 | 0.001 |
| | | 12115 | Education | Secondary | Elementary or less | 0.43 | 0.09 | 0.26 to 0.61 | 0.000 |
| | | 12115 | Education | Four years beyond HS | Elementary or less | 0.65 | 0.12 | 0.41 to 0.89 | 0.000 |
| | | 12115 | Residential setting | Small town | Rural | 0.40 | 0.11 | 0.19 to 0.61 | 0.000 |
| | | 12115 | Residential setting | Large city | Rural | 0.37 | 0.13 | 0.13 to 0.62 | 0.003 |

| Outcome | Model type | Analytic N | Variable | Level / comparison | Reference category | Coefficient | Standard error | 95% CI | p-value |
|---|---|---|---|---|---|---|---|---|---|
| | | 12115 | Residential setting | Suburb | Rural | -0.04 | 0.22 | -0.48 to 0.39 | 0.851 |
| **Remain hopeful for the future** | **Linear regression** | 12063 | *Intercept* | | | 8.15 | 0.12 | 7.91 to 8.39 | 0.000 |
| | | 12063 | Childhood predictor | No | Yes | 0.12 | 0.16 | -0.20 to 0.44 | 0.470 |
| | | 12063 | Age | Per 1-year increase | | -0.01 | 0.00 | -0.01 to 0.00 | 0.001 |
| | | 12063 | Gender | Female | Male | 0.01 | 0.06 | -0.11 to 0.12 | 0.922 |
| | | 12063 | Education | Secondary | Elementary or less | 0.39 | 0.07 | 0.26 to 0.52 | 0.000 |
| | | 12063 | Education | Four years beyond HS | Elementary or less | 0.62 | 0.10 | 0.43 to 0.81 | 0.000 |
| | | 12063 | Residential setting | Small town | Rural | 0.50 | 0.08 | 0.34 to 0.66 | 0.000 |
| | | 12063 | Residential setting | Large city | Rural | 0.49 | 0.11 | 0.28 to 0.70 | 0.000 |
| | | 12063 | Residential setting | Suburb | Rural | 0.42 | 0.18 | 0.07 to 0.77 | 0.019 |
| **Understand purpose in life‡** | **Linear regression** | 12042 | *Intercept* | | | 8.05 | 0.14 | 7.78 to 8.31 | 0.000 |
| | | 12042 | Childhood predictor | No | Yes | -0.13 | 0.22 | -0.55 to 0.30 | 0.557 |
| | | 12042 | Age | Per 1-year increase | | -0.01 | 0.00 | -0.02 to -0.01 | 0.000 |
| | | 12042 | Gender | Female | Male | 0.13 | 0.07 | -0.02 to 0.27 | 0.083 |
| | | 12042 | Education | Secondary | Elementary or less | 0.42 | 0.08 | 0.27 to 0.58 | 0.000 |
| | | 12042 | Education | Four years beyond HS | Elementary or less | 0.75 | 0.10 | 0.55 to 0.94 | 0.000 |
| | | 12042 | Residential setting | Small town | Rural | 0.43 | 0.09 | 0.25 to 0.61 | 0.000 |
| | | 12042 | Residential setting | Large city | Rural | 0.43 | 0.12 | 0.20 to 0.66 | 0.000 |
| | | 12042 | Residential setting | Suburb | Rural | 0.45 | 0.15 | 0.15 to 0.75 | 0.003 |
| **Life satisfaction‡** | **Linear regression** | 12093 | *Intercept* | | | 7.08 | 0.15 | 6.79 to 7.37 | 0.000 |
| | | 12093 | Childhood predictor | No | Yes | -0.68 | 0.29 | -1.25 to -0.11 | 0.019 |
| | | 12093 | Age | Per 1-year increase | | -0.01 | 0.00 | -0.02 to -0.01 | 0.000 |
| | | 12093 | Gender | Female | Male | 0.29 | 0.08 | 0.14 to 0.44 | 0.000 |
| | | 12093 | Education | Secondary | Elementary or less | 0.32 | 0.09 | 0.14 to 0.50 | 0.001 |
| | | 12093 | Education | Four years beyond HS | Elementary or less | 0.47 | 0.12 | 0.24 to 0.71 | 0.000 |
| | | 12093 | Residential setting | Small town | Rural | 0.31 | 0.09 | 0.12 to 0.49 | 0.001 |
| | | 12093 | Residential setting | Large city | Rural | 0.24 | 0.13 | 0.00 to 0.49 | 0.053 |
| | | 12093 | Residential setting | Suburb | Rural | -0.20 | 0.21 | -0.62 to 0.21 | 0.336 |
| **Loneliness§** | **Linear regression** | 12114 | *Intercept* | | | 4.24 | 0.17 | 3.90 to 4.57 | 0.000 |
| | | 12114 | Childhood predictor | No | Yes | 0.69 | 0.30 | 0.10 to 1.27 | 0.022 |
| | | 12114 | Age | Per 1-year increase | | 0.00 | 0.00 | -0.01 to 0.01 | 0.858 |
| | | 12114 | Gender | Female | Male | -0.14 | 0.09 | -0.31 to 0.04 | 0.119 |
| | | 12114 | Education | Secondary | Elementary or less | -0.31 | 0.10 | -0.50 to -0.11 | 0.002 |
| | | 12114 | Education | Four years beyond HS | Elementary or less | -0.65 | 0.15 | -0.94 to -0.36 | 0.000 |
| | | 12114 | Residential setting | Small town | Rural | -0.38 | 0.10 | -0.59 to -0.18 | 0.000 |
| | | 12114 | Residential setting | Large city | Rural | -0.54 | 0.14 | -0.81 to -0.27 | 0.000 |
| | | 12114 | Residential setting | Suburb | Rural | -0.16 | 0.24 | -0.63 to 0.31 | 0.506 |
| **Mental health‡** | **Linear regression** | 12119 | *Intercept* | | | 8.06 | 0.14 | 7.78 to 8.34 | 0.000 |
| | | 12119 | Childhood predictor | No | Yes | -0.77 | 0.29 | -1.33 to -0.21 | 0.007 |
| | | 12119 | Age | Per 1-year increase | | -0.02 | 0.00 | -0.03 to -0.02 | 0.000 |

| Outcome | Model type | Analytic N | Variable | Level / comparison | Reference category | Coefficient | Standard error | 95% CI | p-value |
|---|---|---|---|---|---|---|---|---|---|
| | | 12119 | Gender | Female | Male | -0.13 | 0.07 | -0.27 to 0.01 | 0.060 |
| | | 12119 | Education | Secondary | Elementary or less | 0.41 | 0.08 | 0.26 to 0.57 | 0.000 |
| | | 12119 | Education | Four years beyond HS | Elementary or less | 0.53 | 0.11 | 0.31 to 0.76 | 0.000 |
| | | 12119 | Residential setting | Small town | Rural | 0.39 | 0.09 | 0.21 to 0.58 | 0.000 |
| | | 12119 | Residential setting | Large city | Rural | 0.47 | 0.12 | 0.23 to 0.70 | 0.000 |
| | | 12119 | Residential setting | Suburb | Rural | 0.17 | 0.18 | -0.18 to 0.53 | 0.329 |
| **Perceived support from others** | **Linear regression** | 12106 | *Intercept* | | | 6.66 | 0.16 | 6.35 to 6.98 | 0.000 |
| | | 12106 | Childhood predictor | No | Yes | -1.12 | 0.33 | -1.76 to -0.48 | 0.001 |
| | | 12106 | Age | Per 1-year increase | | -0.01 | 0.00 | -0.02 to -0.01 | 0.000 |
| | | 12106 | Gender | Female | Male | 0.53 | 0.09 | 0.35 to 0.71 | 0.000 |
| | | 12106 | Education | Secondary | Elementary or less | 0.17 | 0.10 | -0.02 to 0.37 | 0.079 |
| | | 12106 | Education | Four years beyond HS | Elementary or less | 0.07 | 0.14 | -0.21 to 0.34 | 0.630 |
| | | 12106 | Residential setting | Small town | Rural | 0.19 | 0.11 | -0.02 to 0.41 | 0.078 |
| | | 12106 | Residential setting | Large city | Rural | -0.41 | 0.15 | -0.70 to -0.12 | 0.005 |
| | | 12106 | Residential setting | Suburb | Rural | -0.23 | 0.21 | -0.65 to 0.18 | 0.265 |
| **Physical health‡** | **Linear regression** | 12122 | *Intercept* | | | 8.37 | 0.15 | 8.09 to 8.66 | 0.000 |
| | | 12122 | Childhood predictor | No | Yes | -0.22 | 0.24 | -0.69 to 0.24 | 0.347 |
| | | 12122 | Age | Per 1-year increase | | -0.04 | 0.00 | -0.05 to -0.04 | 0.000 |
| | | 12122 | Gender | Female | Male | -0.36 | 0.08 | -0.51 to -0.21 | 0.000 |
| | | 12122 | Education | Secondary | Elementary or less | 0.38 | 0.08 | 0.22 to 0.53 | 0.000 |
| | | 12122 | Education | Four years beyond HS | Elementary or less | 0.67 | 0.11 | 0.46 to 0.89 | 0.000 |
| | | 12122 | Residential setting | Small town | Rural | 0.50 | 0.10 | 0.31 to 0.69 | 0.000 |
| | | 12122 | Residential setting | Large city | Rural | 0.66 | 0.11 | 0.45 to 0.88 | 0.000 |
| | | 12122 | Residential setting | Suburb | Rural | 0.21 | 0.18 | -0.13 to 0.56 | 0.229 |
| **Act to promote good‡** | **Linear regression** | 12077 | *Intercept* | | | 8.06 | 0.11 | 7.85 to 8.28 | 0.000 |
| | | 12077 | Childhood predictor | No | Yes | -0.35 | 0.20 | -0.74 to 0.05 | 0.087 |
| | | 12077 | Age | Per 1-year increase | | -0.01 | 0.00 | -0.01 to 0.00 | 0.001 |
| | | 12077 | Gender | Female | Male | 0.12 | 0.06 | 0.00 to 0.24 | 0.057 |
| | | 12077 | Education | Secondary | Elementary or less | 0.38 | 0.06 | 0.26 to 0.51 | 0.000 |
| | | 12077 | Education | Four years beyond HS | Elementary or less | 0.56 | 0.08 | 0.40 to 0.72 | 0.000 |
| | | 12077 | Residential setting | Small town | Rural | 0.60 | 0.08 | 0.46 to 0.75 | 0.000 |
| | | 12077 | Residential setting | Large city | Rural | 0.65 | 0.10 | 0.45 to 0.84 | 0.000 |
| | | 12077 | Residential setting | Suburb | Rural | 0.41 | 0.17 | 0.09 to 0.74 | 0.013 |
| **Relationship satisfaction‡** | **Linear regression** | 12083 | *Intercept* | | | 7.81 | 0.13 | 7.56 to 8.07 | 0.000 |
| | | 12083 | Childhood predictor | No | Yes | -0.88 | 0.26 | -1.38 to -0.37 | 0.001 |
| | | 12083 | Age | Per 1-year increase | | -0.01 | 0.00 | -0.01 to 0.00 | 0.003 |
| | | 12083 | Gender | Female | Male | 0.34 | 0.07 | 0.20 to 0.48 | 0.000 |
| | | 12083 | Education | Secondary | Elementary or less | -0.01 | 0.08 | -0.17 to 0.15 | 0.913 |
| | | 12083 | Education | Four years beyond HS | Elementary or less | 0.05 | 0.11 | -0.17 to 0.27 | 0.662 |
| | | 12083 | Residential setting | Small town | Rural | 0.47 | 0.09 | 0.29 to 0.64 | 0.000 |

| Outcome | Model type | Analytic N | Variable | Level / comparison | Reference category | Coefficient | Standard error | 95% CI | p-value |
|---|---|---|---|---|---|---|---|---|---|
| | | 12083 | Residential setting | Large city | Rural | 0.35 | 0.11 | 0.13 to 0.56 | 0.002 |
| | | 12083 | Residential setting | Suburb | Rural | 0.07 | 0.16 | -0.23 to 0.38 | 0.633 |
| **Show love often** | **Linear regression** | 12105 | *Intercept* | | | 8.30 | 0.13 | 8.05 to 8.54 | 0.000 |
| | | 12105 | Childhood predictor | No | Yes | -0.27 | 0.23 | -0.72 to 0.17 | 0.225 |
| | | 12105 | Age | Per 1-year increase | | -0.01 | 0.00 | -0.01 to 0.00 | 0.002 |
| | | 12105 | Gender | Female | Male | 0.19 | 0.07 | 0.06 to 0.32 | 0.005 |
| | | 12105 | Education | Secondary | Elementary or less | 0.15 | 0.07 | 0.02 to 0.29 | 0.029 |
| | | 12105 | Education | Four years beyond HS | Elementary or less | 0.02 | 0.11 | -0.20 to 0.24 | 0.860 |
| | | 12105 | Residential setting | Small town | Rural | 0.20 | 0.08 | 0.04 to 0.35 | 0.014 |
| | | 12105 | Residential setting | Large city | Rural | 0.03 | 0.11 | -0.19 to 0.26 | 0.766 |
| | | 12105 | Residential setting | Suburb | Rural | -0.03 | 0.21 | -0.43 to 0.38 | 0.899 |
| **Life evaluation five years from now** | **Linear regression** | 11029 | *Intercept* | | | 7.68 | 0.14 | 7.40 to 7.96 | 0.000 |
| | | 11029 | Childhood predictor | No | Yes | -0.35 | 0.25 | -0.83 to 0.13 | 0.152 |
| | | 11029 | Age | Per 1-year increase | | -0.02 | 0.00 | -0.03 to -0.02 | 0.000 |
| | | 11029 | Gender | Female | Male | 0.17 | 0.07 | 0.03 to 0.31 | 0.019 |
| | | 11029 | Education | Secondary | Elementary or less | 0.29 | 0.08 | 0.13 to 0.44 | 0.000 |
| | | 11029 | Education | Four years beyond HS | Elementary or less | 0.58 | 0.12 | 0.35 to 0.81 | 0.000 |
| | | 11029 | Residential setting | Small town | Rural | 0.45 | 0.09 | 0.27 to 0.62 | 0.000 |
| | | 11029 | Residential setting | Large city | Rural | 0.52 | 0.11 | 0.30 to 0.73 | 0.000 |
| | | 11029 | Residential setting | Suburb | Rural | 0.41 | 0.22 | -0.02 to 0.84 | 0.064 |
| **Life evaluation today** | **Linear regression** | 12016 | *Intercept* | | | 5.56 | 0.17 | 5.23 to 5.90 | 0.000 |
| | | 12016 | Childhood predictor | No | Yes | -0.57 | 0.25 | -1.07 to -0.07 | 0.027 |
| | | 12016 | Age | Per 1-year increase | | -0.01 | 0.00 | -0.02 to 0.00 | 0.006 |
| | | 12016 | Gender | Female | Male | 0.49 | 0.08 | 0.33 to 0.66 | 0.000 |
| | | 12016 | Education | Secondary | Elementary or less | 0.24 | 0.09 | 0.07 to 0.42 | 0.006 |
| | | 12016 | Education | Four years beyond HS | Elementary or less | 0.14 | 0.14 | -0.14 to 0.42 | 0.316 |
| | | 12016 | Residential setting | Small town | Rural | 0.26 | 0.10 | 0.05 to 0.46 | 0.015 |
| | | 12016 | Residential setting | Large city | Rural | 0.12 | 0.13 | -0.13 to 0.38 | 0.350 |
| | | 12016 | Residential setting | Suburb | Rural | -0.04 | 0.21 | -0.46 to 0.37 | 0.844 |
| **Worry about safety, food, or housing§§‡‡** | **Linear regression** | 12130 | *Intercept* | | | 6.16 | 0.19 | 5.79 to 6.53 | 0.000 |
| | | 12130 | Childhood predictor | No | Yes | -0.06 | 0.32 | -0.68 to 0.56 | 0.845 |
| | | 12130 | Age | Per 1-year increase | | -0.01 | 0.00 | -0.01 to 0.00 | 0.046 |
| | | 12130 | Gender | Female | Male | 0.32 | 0.09 | 0.14 to 0.50 | 0.000 |
| | | 12130 | Education | Secondary | Elementary or less | -0.53 | 0.11 | -0.74 to -0.33 | 0.000 |
| | | 12130 | Education | Four years beyond HS | Elementary or less | -0.78 | 0.17 | -1.10 to -0.45 | 0.000 |
| | | 12130 | Residential setting | Small town | Rural | 0.39 | 0.13 | 0.13 to 0.65 | 0.003 |
| | | 12130 | Residential setting | Large city | Rural | -0.04 | 0.18 | -0.39 to 0.31 | 0.836 |
| | | 12130 | Residential setting | Suburb | Rural | 0.19 | 0.26 | -0.32 to 0.70 | 0.460 |
| **Life worthwhile‡** | **Linear regression** | 12018 | *Intercept* | | | 6.63 | 0.15 | 6.35 to 6.92 | 0.000 |

| Outcome | Model type | Analytic N | Variable | Level / comparison | Reference category | Coefficient | Standard error | 95% CI | p-value |
|---|---|---|---|---|---|---|---|---|---|
| | | 12018 | Childhood predictor | No | Yes | -0.25 | 0.28 | -0.80 to 0.29 | 0.365 |
| | | 12018 | Age | Per 1-year increase | | -0.01 | 0.00 | -0.01 to 0.00 | 0.002 |
| | | 12018 | Gender | Female | Male | 0.45 | 0.08 | 0.30 to 0.60 | 0.000 |
| | | 12018 | Education | Secondary | Elementary or less | 0.47 | 0.08 | 0.31 to 0.63 | 0.000 |
| | | 12018 | Education | Four years beyond HS | Elementary or less | 0.61 | 0.13 | 0.36 to 0.85 | 0.000 |
| | | 12018 | Residential setting | Small town | Rural | 0.46 | 0.10 | 0.26 to 0.66 | 0.000 |
| | | 12018 | Residential setting | Large city | Rural | 0.44 | 0.13 | 0.19 to 0.70 | 0.001 |
| | | 12018 | Residential setting | Suburb | Rural | 0.40 | 0.21 | -0.01 to 0.80 | 0.054 |
| **Flourishing (with financial indicators)** | **Linear regression** | 11680 | *Intercept* | | | 7.00 | 0.08 | 6.84 to 7.16 | 0.000 |
| | | 11680 | Childhood predictor | No | Yes | -0.43 | 0.14 | -0.70 to -0.16 | 0.002 |
| | | 11680 | Age | Per 1-year increase | | -0.01 | 0.00 | -0.02 to -0.01 | 0.000 |
| | | 11680 | Gender | Female | Male | 0.10 | 0.04 | 0.01 to 0.18 | 0.024 |
| | | 11680 | Education | Secondary | Elementary or less | 0.33 | 0.05 | 0.24 to 0.42 | 0.000 |
| | | 11680 | Education | Four years beyond HS | Elementary or less | 0.51 | 0.07 | 0.38 to 0.63 | 0.000 |
| | | 11680 | Residential setting | Small town | Rural | 0.33 | 0.06 | 0.21 to 0.44 | 0.000 |
| | | 11680 | Residential setting | Large city | Rural | 0.36 | 0.07 | 0.22 to 0.49 | 0.000 |
| | | 11680 | Residential setting | Suburb | Rural | 0.12 | 0.13 | -0.14 to 0.38 | 0.359 |
| **Flourishing (without financial indicators)** | **Linear regression** | 11707 | *Intercept* | | | 7.60 | 0.09 | 7.43 to 7.77 | 0.000 |
| | | 11707 | Childhood predictor | No | Yes | -0.52 | 0.16 | -0.82 to -0.21 | 0.001 |
| | | 11707 | Age | Per 1-year increase | | -0.02 | 0.00 | -0.02 to -0.01 | 0.000 |
| | | 11707 | Gender | Female | Male | 0.18 | 0.05 | 0.09 to 0.27 | 0.000 |
| | | 11707 | Education | Secondary | Elementary or less | 0.31 | 0.05 | 0.21 to 0.40 | 0.000 |
| | | 11707 | Education | Four years beyond HS | Elementary or less | 0.47 | 0.07 | 0.33 to 0.61 | 0.000 |
| | | 11707 | Residential setting | Small town | Rural | 0.45 | 0.06 | 0.32 to 0.58 | 0.000 |
| | | 11707 | Residential setting | Large city | Rural | 0.41 | 0.08 | 0.25 to 0.56 | 0.000 |
| | | 11707 | Residential setting | Suburb | Rural | 0.14 | 0.13 | -0.11 to 0.40 | 0.278 |
| **Feel close to someone** | **Logistic regression** | 12128 | *Intercept* | | | 1.69 | 0.14 | 1.42 to 1.97 | 0.000 |
| | | 12128 | Childhood predictor | No | Yes | -0.16 | 0.18 | -0.52 to 0.19 | 0.365 |
| | | 12128 | Age | Per 1-year increase | | -0.01 | 0.00 | -0.01 to 0.00 | 0.009 |
| | | 12128 | Gender | Female | Male | 0.01 | 0.07 | -0.12 to 0.14 | 0.930 |
| | | 12128 | Education | Secondary | Elementary or less | 0.16 | 0.08 | 0.00 to 0.32 | 0.056 |
| | | 12128 | Education | Four years beyond HS | Elementary or less | 0.12 | 0.13 | -0.14 to 0.37 | 0.365 |
| | | 12128 | Residential setting | Small town | Rural | 0.09 | 0.11 | -0.13 to 0.30 | 0.432 |
| | | 12128 | Residential setting | Large city | Rural | -0.08 | 0.13 | -0.33 to 0.18 | 0.541 |
| | | 12128 | Residential setting | Suburb | Rural | -0.16 | 0.19 | -0.53 to 0.22 | 0.405 |
| **Donated to charity** | **Logistic regression** | 12098 | *Intercept* | | | -0.33 | 0.13 | -0.58 to -0.08 | 0.010 |
| | | 12098 | Childhood predictor | No | Yes | -0.06 | 0.16 | -0.38 to 0.25 | 0.690 |
| | | 12098 | Age | Per 1-year increase | | 0.00 | 0.00 | 0.00 to 0.00 | 0.577 |
| | | 12098 | Gender | Female | Male | -0.03 | 0.05 | -0.13 to 0.07 | 0.545 |

| Outcome | Model type | Analytic N | Variable | Level / comparison | Reference category | Coefficient | Standard error | 95% CI | p-value |
|---|---|---|---|---|---|---|---|---|---|
| | | 12098 | Education | Secondary | Elementary or less | -0.09 | 0.06 | -0.21 to 0.03 | 0.129 |
| | | 12098 | Education | Four years beyond HS | Elementary or less | -0.16 | 0.10 | -0.36 to 0.04 | 0.119 |
| | | 12098 | Residential setting | Small town | Rural | -0.05 | 0.09 | -0.24 to 0.13 | 0.560 |
| | | 12098 | Residential setting | Large city | Rural | -0.40 | 0.12 | -0.63 to -0.18 | 0.000 |
| | | 12098 | Residential setting | Suburb | Rural | -0.17 | 0.19 | -0.55 to 0.20 | 0.363 |
| **Have any health problems** | **Logistic regression** | 12050 | *Intercept* | | | -2.02 | 0.10 | -2.23 to -1.82 | 0.000 |
| | | 12050 | Childhood predictor | No | Yes | 0.14 | 0.16 | -0.17 to 0.46 | 0.382 |
| | | 12050 | Age | Per 1-year increase | | 0.03 | 0.00 | 0.02 to 0.03 | 0.000 |
| | | 12050 | Gender | Female | Male | 0.21 | 0.05 | 0.11 to 0.30 | 0.000 |
| | | 12050 | Education | Secondary | Elementary or less | -0.15 | 0.06 | -0.26 to -0.03 | 0.016 |
| | | 12050 | Education | Four years beyond HS | Elementary or less | -0.27 | 0.10 | -0.46 to -0.08 | 0.005 |
| | | 12050 | Residential setting | Small town | Rural | 0.12 | 0.07 | -0.01 to 0.25 | 0.071 |
| | | 12050 | Residential setting | Large city | Rural | -0.12 | 0.09 | -0.30 to 0.05 | 0.161 |
| | | 12050 | Residential setting | Suburb | Rural | 0.10 | 0.14 | -0.18 to 0.38 | 0.483 |
| **Helped a stranger** | **Logistic regression** | 12009 | *Intercept* | | | 0.70 | 0.12 | 0.47 to 0.93 | 0.000 |
| | | 12009 | Childhood predictor | No | Yes | 0.22 | 0.16 | -0.10 to 0.54 | 0.183 |
| | | 12009 | Age | Per 1-year increase | | -0.01 | 0.00 | -0.01 to 0.00 | 0.000 |
| | | 12009 | Gender | Female | Male | -0.35 | 0.05 | -0.44 to -0.25 | 0.000 |
| | | 12009 | Education | Secondary | Elementary or less | -0.10 | 0.06 | -0.21 to 0.02 | 0.090 |
| | | 12009 | Education | Four years beyond HS | Elementary or less | -0.09 | 0.09 | -0.27 to 0.09 | 0.326 |
| | | 12009 | Residential setting | Small town | Rural | 0.11 | 0.09 | -0.06 to 0.28 | 0.211 |
| | | 12009 | Residential setting | Large city | Rural | -0.15 | 0.10 | -0.35 to 0.05 | 0.144 |
| | | 12009 | Residential setting | Suburb | Rural | 0.03 | 0.18 | -0.32 to 0.38 | 0.866 |
| **Volunteered at an organization** | **Logistic regression** | 12079 | *Intercept* | | | -0.28 | 0.14 | -0.56 to -0.01 | 0.041 |
| | | 12079 | Childhood predictor | No | Yes | -0.02 | 0.17 | -0.35 to 0.30 | 0.888 |
| | | 12079 | Age | Per 1-year increase | | -0.01 | 0.00 | -0.01 to 0.00 | 0.001 |
| | | 12079 | Gender | Female | Male | -0.29 | 0.05 | -0.39 to -0.18 | 0.000 |
| | | 12079 | Education | Secondary | Elementary or less | -0.15 | 0.07 | -0.28 to -0.02 | 0.022 |
| | | 12079 | Education | Four years beyond HS | Elementary or less | 0.02 | 0.10 | -0.18 to 0.22 | 0.860 |
| | | 12079 | Residential setting | Small town | Rural | -0.10 | 0.11 | -0.32 to 0.11 | 0.350 |
| | | 12079 | Residential setting | Large city | Rural | -0.56 | 0.13 | -0.82 to -0.31 | 0.000 |
| | | 12079 | Residential setting | Suburb | Rural | -0.40 | 0.17 | -0.74 to -0.07 | 0.019 |
| **Cigarette smoking** | **Logistic regression** | 11660 | *Intercept* | | | -1.99 | 0.16 | -2.31 to -1.67 | 0.000 |
| | | 11660 | Childhood predictor | No | Yes | -0.27 | 0.31 | -0.87 to 0.34 | 0.392 |
| | | 11660 | Age | Per 1-year increase | | 0.01 | 0.00 | 0.00 to 0.01 | 0.022 |
| | | 11660 | Gender | Female | Male | -2.82 | 0.19 | -3.19 to -2.45 | 0.000 |
| | | 11660 | Education | Secondary | Elementary or less | -0.41 | 0.10 | -0.60 to -0.21 | 0.000 |
| | | 11660 | Education | Four years beyond HS | Elementary or less | -0.56 | 0.17 | -0.89 to -0.23 | 0.001 |
| | | 11660 | Residential setting | Small town | Rural | -0.05 | 0.11 | -0.26 to 0.16 | 0.633 |
| | | 11660 | Residential setting | Large city | Rural | 0.14 | 0.14 | -0.14 to 0.41 | 0.325 |

| Outcome | Model type | Analytic N | Variable | Level / comparison | Reference category | Coefficient | Standard error | 95% CI | p-value |
|---|---|---|---|---|---|---|---|---|---|
| | | 11660 | Residential setting | Suburb | Rural | 0.85 | 0.20 | 0.46 to 1.25 | 0.000 |
| **Alcohol use** | **Logistic regression** | 11647 | *Intercept* | | | -1.90 | 0.15 | -2.19 to -1.61 | 0.000 |
| | | 11647 | Childhood predictor | No | Yes | 0.69 | 0.24 | 0.22 to 1.17 | 0.004 |
| | | 11647 | Age | Per 1-year increase | | 0.00 | 0.00 | -0.01 to 0.01 | 0.830 |
| | | 11647 | Gender | Female | Male | -2.79 | 0.18 | -3.14 to -2.43 | 0.000 |
| | | 11647 | Education | Secondary | Elementary or less | -0.24 | 0.10 | -0.43 to -0.04 | 0.017 |
| | | 11647 | Education | Four years beyond HS | Elementary or less | -0.47 | 0.16 | -0.79 to -0.15 | 0.004 |
| | | 11647 | Residential setting | Small town | Rural | 0.21 | 0.11 | 0.00 to 0.42 | 0.052 |
| | | 11647 | Residential setting | Large city | Rural | 0.40 | 0.14 | 0.13 to 0.68 | 0.004 |
| | | 11647 | Residential setting | Suburb | Rural | 0.61 | 0.23 | 0.17 to 1.06 | 0.007 |
| **Government approval§** | **Proportional-odds regression** | 11922 | Childhood predictor | No | Yes | -0.53 | 0.14 | -0.80 to -0.26 | 0.000 |
| | | 11922 | Age | Per 1-year increase | | 0.00 | 0.00 | -0.01 to 0.00 | 0.071 |
| | | 11922 | Gender | Female | Male | 0.05 | 0.04 | -0.03 to 0.14 | 0.216 |
| | | 11922 | Education | Secondary | Elementary or less | -0.24 | 0.05 | -0.35 to -0.14 | 0.000 |
| | | 11922 | Education | Four years beyond HS | Elementary or less | -0.32 | 0.09 | -0.49 to -0.16 | 0.000 |
| | | 11922 | Residential setting | Small town | Rural | 0.10 | 0.07 | -0.03 to 0.23 | 0.140 |
| | | 11922 | Residential setting | Large city | Rural | -0.08 | 0.08 | -0.23 to 0.08 | 0.333 |
| | | 11922 | Residential setting | Suburb | Rural | -0.44 | 0.16 | -0.76 to -0.13 | 0.005 |
| | | 11922 | *Threshold / cut-point* | Strongly disapprove\|Somewhat disapprove | | -2.91 | 0.10 | -3.10 to -2.71 | 0.000 |
| | | 11922 | *Threshold / cut-point* | Somewhat disapprove\|Neither | | -2.25 | 0.10 | -2.44 to -2.07 | 0.000 |
| | | 11922 | *Threshold / cut-point* | Neither\|Somewhat approve | | -1.89 | 0.09 | -2.08 to -1.71 | 0.000 |
| | | 11922 | *Threshold / cut-point* | Somewhat approve\|Strongly approve | | -0.44 | 0.09 | -0.61 to -0.26 | 0.000 |
| **Feeling capable§** | **Proportional-odds regression** | 12073 | Childhood predictor | No | Yes | -0.15 | 0.14 | -0.42 to 0.12 | 0.283 |
| | | 12073 | Age | Per 1-year increase | | 0.00 | 0.00 | 0.00 to 0.01 | 0.246 |
| | | 12073 | Gender | Female | Male | -0.07 | 0.04 | -0.15 to 0.02 | 0.127 |
| | | 12073 | Education | Secondary | Elementary or less | 0.15 | 0.05 | 0.05 to 0.24 | 0.003 |
| | | 12073 | Education | Four years beyond HS | Elementary or less | 0.11 | 0.07 | -0.04 to 0.25 | 0.145 |
| | | 12073 | Residential setting | Small town | Rural | 0.16 | 0.05 | 0.06 to 0.26 | 0.001 |
| | | 12073 | Residential setting | Large city | Rural | 0.19 | 0.08 | 0.04 to 0.34 | 0.013 |
| | | 12073 | Residential setting | Suburb | Rural | 0.22 | 0.09 | 0.05 to 0.40 | 0.012 |
| | | 12073 | *Threshold / cut-point* | Never\|Rarely | | -2.35 | 0.09 | -2.53 to -2.17 | 0.000 |
| | | 12073 | *Threshold / cut-point* | Rarely\|Often | | -0.64 | 0.08 | -0.81 to -0.48 | 0.000 |
| | | 12073 | *Threshold / cut-point* | Often\|Always | | 0.02 | 0.08 | -0.14 to 0.19 | 0.796 |
| **Unable to control worry§** | **Proportional-odds regression** | 12025 | Childhood predictor | No | Yes | 0.19 | 0.13 | -0.06 to 0.45 | 0.128 |
| | | 12025 | Age | Per 1-year increase | | 0.00 | 0.00 | 0.00 to 0.01 | 0.001 |
| | | 12025 | Gender | Female | Male | 0.17 | 0.04 | 0.08 to 0.25 | 0.000 |

| Outcome | Model type | Analytic N | Variable | Level / comparison | Reference category | Coefficient | Standard error | 95% CI | p-value |
|---|---|---|---|---|---|---|---|---|---|
| | | 12025 | Education | Secondary | Elementary or less | -0.28 | 0.05 | -0.37 to -0.19 | 0.000 |
| | | 12025 | Education | Four years beyond HS | Elementary or less | -0.41 | 0.08 | -0.56 to -0.27 | 0.000 |
| | | 12025 | Residential setting | Small town | Rural | 0.03 | 0.05 | -0.06 to 0.13 | 0.501 |
| | | 12025 | Residential setting | Large city | Rural | -0.01 | 0.07 | -0.15 to 0.12 | 0.835 |
| | | 12025 | Residential setting | Suburb | Rural | -0.08 | 0.10 | -0.27 to 0.12 | 0.440 |
| | | 12025 | *Threshold / cut-point* | Not at all\|Several days | | 0.06 | 0.08 | -0.10 to 0.22 | 0.468 |
| | | 12025 | *Threshold / cut-point* | Several days\|More than half the days | | 0.96 | 0.08 | 0.80 to 1.12 | 0.000 |
| | | 12025 | *Threshold / cut-point* | More than half the days\|Nearly every day | | 1.82 | 0.08 | 1.66 to 1.98 | 0.000 |
| **Depressive symptoms§** | **Proportional-odds regression** | 12050 | Childhood predictor | No | Yes | 0.35 | 0.13 | 0.10 to 0.60 | 0.006 |
| | | 12050 | Age | Per 1-year increase | | 0.00 | 0.00 | 0.00 to 0.01 | 0.155 |
| | | 12050 | Gender | Female | Male | 0.07 | 0.04 | 0.00 to 0.15 | 0.060 |
| | | 12050 | Education | Secondary | Elementary or less | -0.21 | 0.05 | -0.30 to -0.12 | 0.000 |
| | | 12050 | Education | Four years beyond HS | Elementary or less | -0.41 | 0.07 | -0.55 to -0.28 | 0.000 |
| | | 12050 | Residential setting | Small town | Rural | 0.00 | 0.05 | -0.10 to 0.11 | 0.969 |
| | | 12050 | Residential setting | Large city | Rural | -0.10 | 0.07 | -0.24 to 0.05 | 0.184 |
| | | 12050 | Residential setting | Suburb | Rural | -0.16 | 0.11 | -0.38 to 0.06 | 0.164 |
| | | 12050 | *Threshold / cut-point* | Not at all\|Several days | | -0.25 | 0.08 | -0.41 to -0.09 | 0.002 |
| | | 12050 | *Threshold / cut-point* | Several days\|More than half the days | | 0.82 | 0.08 | 0.67 to 0.98 | 0.000 |
| | | 12050 | *Threshold / cut-point* | More than half the days\|Nearly every day | | 1.83 | 0.08 | 1.67 to 2.00 | 0.000 |
| **Perceived discrimination§** | **Proportional-odds regression** | 11985 | Childhood predictor | No | Yes | 0.15 | 0.14 | -0.13 to 0.43 | 0.288 |
| | | 11985 | Age | Per 1-year increase | | 0.00 | 0.00 | -0.01 to 0.00 | 0.021 |
| | | 11985 | Gender | Female | Male | -0.15 | 0.04 | -0.24 to -0.07 | 0.001 |
| | | 11985 | Education | Secondary | Elementary or less | -0.26 | 0.05 | -0.36 to -0.16 | 0.000 |
| | | 11985 | Education | Four years beyond HS | Elementary or less | -0.31 | 0.09 | -0.49 to -0.14 | 0.000 |
| | | 11985 | Residential setting | Small town | Rural | -0.22 | 0.06 | -0.33 to -0.10 | 0.000 |
| | | 11985 | Residential setting | Large city | Rural | -0.22 | 0.07 | -0.36 to -0.08 | 0.002 |
| | | 11985 | Residential setting | Suburb | Rural | -0.35 | 0.10 | -0.55 to -0.14 | 0.001 |
| | | 11985 | *Threshold / cut-point* | Never\|Rarely | | -0.17 | 0.09 | -0.34 to 0.00 | 0.053 |
| | | 11985 | *Threshold / cut-point* | Rarely\|Often | | 0.83 | 0.09 | 0.66 to 1.01 | 0.000 |
| | | 11985 | *Threshold / cut-point* | Often\|Always | | 1.44 | 0.09 | 1.26 to 1.62 | 0.000 |
| **Feeling nervous or anxious§** | **Proportional-odds regression** | 12075 | Childhood predictor | No | Yes | 0.23 | 0.11 | 0.01 to 0.46 | 0.042 |
| | | 12075 | Age | Per 1-year increase | | 0.01 | 0.00 | 0.00 to 0.01 | 0.000 |
| | | 12075 | Gender | Female | Male | 0.29 | 0.04 | 0.21 to 0.37 | 0.000 |
| | | 12075 | Education | Secondary | Elementary or less | -0.28 | 0.05 | -0.37 to -0.19 | 0.000 |
| | | 12075 | Education | Four years beyond HS | Elementary or less | -0.45 | 0.08 | -0.60 to -0.30 | 0.000 |

| Outcome | Model type | Analytic N | Variable | Level / comparison | Reference category | Coefficient | Standard error | 95% CI | p-value |
|---|---|---|---|---|---|---|---|---|---|
| | | 12075 | Residential setting | Small town | Rural | 0.11 | 0.05 | 0.01 to 0.22 | 0.034 |
| | | 12075 | Residential setting | Large city | Rural | -0.04 | 0.07 | -0.17 to 0.10 | 0.602 |
| | | 12075 | Residential setting | Suburb | Rural | -0.04 | 0.12 | -0.28 to 0.19 | 0.725 |
| | | 12075 | *Threshold / cut-point* | Not at all\|Several days | | 0.14 | 0.08 | -0.01 to 0.29 | 0.072 |
| | | 12075 | *Threshold / cut-point* | Several days\|More than half the days | | 1.14 | 0.08 | 0.99 to 1.29 | 0.000 |
| | | 12075 | *Threshold / cut-point* | More than half the days\|Nearly every day | | 2.08 | 0.08 | 1.92 to 2.23 | 0.000 |
| **Forgiveness§** | **Proportional-odds regression** | 12053 | Childhood predictor | No | Yes | -0.42 | 0.14 | -0.71 to -0.14 | 0.003 |
| | | 12053 | Age | Per 1-year increase | | 0.00 | 0.00 | 0.00 to 0.00 | 0.917 |
| | | 12053 | Gender | Female | Male | 0.09 | 0.04 | 0.01 to 0.18 | 0.031 |
| | | 12053 | Education | Secondary | Elementary or less | -0.08 | 0.05 | -0.19 to 0.03 | 0.136 |
| | | 12053 | Education | Four years beyond HS | Elementary or less | -0.19 | 0.08 | -0.35 to -0.04 | 0.014 |
| | | 12053 | Residential setting | Small town | Rural | 0.02 | 0.05 | -0.08 to 0.13 | 0.663 |
| | | 12053 | Residential setting | Large city | Rural | 0.03 | 0.08 | -0.13 to 0.19 | 0.716 |
| | | 12053 | Residential setting | Suburb | Rural | 0.05 | 0.12 | -0.19 to 0.28 | 0.699 |
| | | 12053 | *Threshold / cut-point* | Never\|Rarely | | -2.42 | 0.09 | -2.60 to -2.24 | 0.000 |
| | | 12053 | *Threshold / cut-point* | Rarely\|Often | | -1.08 | 0.09 | -1.25 to -0.92 | 0.000 |
| | | 12053 | *Threshold / cut-point* | Often\|Always | | -0.43 | 0.08 | -0.60 to -0.27 | 0.000 |
| **Support reducing differences in income levels§** | **Proportional-odds regression** | 11918 | Childhood predictor | No | Yes | -0.32 | 0.17 | -0.65 to 0.01 | 0.059 |
| | | 11918 | Age | Per 1-year increase | | 0.00 | 0.00 | -0.01 to 0.00 | 0.089 |
| | | 11918 | Gender | Female | Male | -0.05 | 0.06 | -0.16 to 0.07 | 0.420 |
| | | 11918 | Education | Secondary | Elementary or less | -0.07 | 0.07 | -0.21 to 0.07 | 0.334 |
| | | 11918 | Education | Four years beyond HS | Elementary or less | -0.28 | 0.11 | -0.50 to -0.06 | 0.012 |
| | | 11918 | Residential setting | Small town | Rural | 0.03 | 0.13 | -0.21 to 0.28 | 0.787 |
| | | 11918 | Residential setting | Large city | Rural | 0.06 | 0.14 | -0.22 to 0.34 | 0.653 |
| | | 11918 | Residential setting | Suburb | Rural | -0.08 | 0.18 | -0.43 to 0.27 | 0.666 |
| | | 11918 | *Threshold / cut-point* | Strongly disagree\|Somewhat disagree | | -3.88 | 0.17 | -4.22 to -3.55 | 0.000 |
| | | 11918 | *Threshold / cut-point* | Somewhat disagree\|Neither | | -3.33 | 0.16 | -3.65 to -3.01 | 0.000 |
| | | 11918 | *Threshold / cut-point* | Neither\|Somewhat agree | | -2.88 | 0.16 | -3.18 to -2.57 | 0.000 |
| | | 11918 | *Threshold / cut-point* | Somewhat agree\|Strongly agree | | -1.32 | 0.14 | -1.59 to -1.04 | 0.000 |
| **Have little interest in doing things§** | **Proportional-odds regression** | 12043 | Childhood predictor | No | Yes | 0.05 | 0.13 | -0.22 to 0.31 | 0.729 |
| | | 12043 | Age | Per 1-year increase | | 0.00 | 0.00 | 0.00 to 0.01 | 0.055 |
| | | 12043 | Gender | Female | Male | 0.03 | 0.04 | -0.05 to 0.11 | 0.480 |
| | | 12043 | Education | Secondary | Elementary or less | 0.07 | 0.05 | -0.03 to 0.16 | 0.164 |
| | | 12043 | Education | Four years beyond HS | Elementary or less | 0.10 | 0.08 | -0.05 to 0.26 | 0.188 |
| | | 12043 | Residential setting | Small town | Rural | 0.26 | 0.06 | 0.13 to 0.38 | 0.000 |

| Outcome | Model type | Analytic N | Variable | Level / comparison | Reference category | Coefficient | Standard error | 95% CI | p-value |
|---|---|---|---|---|---|---|---|---|---|
| | | 12043 | Residential setting | Large city | Rural | 0.11 | 0.08 | -0.06 to 0.27 | 0.197 |
| | | 12043 | Residential setting | Suburb | Rural | 0.26 | 0.15 | -0.03 to 0.55 | 0.079 |
| | | 12043 | *Threshold / cut-point* | Not at all\|Several days | | -0.51 | 0.09 | -0.68 to -0.34 | 0.000 |
| | | 12043 | *Threshold / cut-point* | Several days\|More than half the days | | 0.43 | 0.09 | 0.26 to 0.60 | 0.000 |
| | | 12043 | *Threshold / cut-point* | More than half the days\|Nearly every day | | 1.21 | 0.09 | 1.03 to 1.38 | 0.000 |
| **Various aspects of life are balanced§** | **Proportional-odds regression** | 11972 | Childhood predictor | No | Yes | -0.09 | 0.15 | -0.37 to 0.20 | 0.560 |
| | | 11972 | Age | Per 1-year increase | | 0.00 | 0.00 | 0.00 to 0.01 | 0.232 |
| | | 11972 | Gender | Female | Male | -0.03 | 0.04 | -0.11 to 0.05 | 0.448 |
| | | 11972 | Education | Secondary | Elementary or less | 0.03 | 0.05 | -0.06 to 0.12 | 0.480 |
| | | 11972 | Education | Four years beyond HS | Elementary or less | 0.10 | 0.07 | -0.03 to 0.24 | 0.123 |
| | | 11972 | Residential setting | Small town | Rural | 0.09 | 0.05 | -0.02 to 0.19 | 0.095 |
| | | 11972 | Residential setting | Large city | Rural | 0.10 | 0.07 | -0.03 to 0.23 | 0.125 |
| | | 11972 | Residential setting | Suburb | Rural | 0.38 | 0.11 | 0.15 to 0.60 | 0.001 |
| | | 11972 | *Threshold / cut-point* | Never\|Rarely | | -2.08 | 0.09 | -2.25 to -1.90 | 0.000 |
| | | 11972 | *Threshold / cut-point* | Rarely\|Often | | 0.11 | 0.08 | -0.05 to 0.27 | 0.184 |
| | | 11972 | *Threshold / cut-point* | Often\|Always | | 0.90 | 0.08 | 0.73 to 1.07 | 0.000 |
| **Obeying the law§** | **Proportional-odds regression** | 12100 | Childhood predictor | No | Yes | -0.29 | 0.20 | -0.68 to 0.09 | 0.137 |
| | | 12100 | Age | Per 1-year increase | | 0.00 | 0.00 | -0.01 to 0.00 | 0.440 |
| | | 12100 | Gender | Female | Male | -0.16 | 0.08 | -0.31 to 0.00 | 0.044 |
| | | 12100 | Education | Secondary | Elementary or less | 0.11 | 0.10 | -0.08 to 0.31 | 0.242 |
| | | 12100 | Education | Four years beyond HS | Elementary or less | -0.20 | 0.13 | -0.44 to 0.05 | 0.123 |
| | | 12100 | Residential setting | Small town | Rural | -0.07 | 0.15 | -0.36 to 0.22 | 0.649 |
| | | 12100 | Residential setting | Large city | Rural | -0.27 | 0.18 | -0.62 to 0.09 | 0.140 |
| | | 12100 | Residential setting | Suburb | Rural | -0.24 | 0.23 | -0.68 to 0.21 | 0.296 |
| | | 12100 | *Threshold / cut-point* | Strongly disagree\|Somewhat disagree | | -5.18 | 0.25 | -5.68 to -4.69 | 0.000 |
| | | 12100 | *Threshold / cut-point* | Somewhat disagree\|Neither | | -4.56 | 0.21 | -4.97 to -4.14 | 0.000 |
| | | 12100 | *Threshold / cut-point* | Neither\|Somewhat agree | | -4.06 | 0.20 | -4.45 to -3.66 | 0.000 |
| | | 12100 | *Threshold / cut-point* | Somewhat agree\|Strongly agree | | -2.23 | 0.18 | -2.58 to -1.88 | 0.000 |
| **Feel at peace§** | **Proportional-odds regression** | 12041 | Childhood predictor | No | Yes | -0.17 | 0.14 | -0.43 to 0.10 | 0.219 |
| | | 12041 | Age | Per 1-year increase | | 0.01 | 0.00 | 0.00 to 0.01 | 0.000 |
| | | 12041 | Gender | Female | Male | 0.00 | 0.04 | -0.07 to 0.08 | 0.899 |
| | | 12041 | Education | Secondary | Elementary or less | 0.02 | 0.05 | -0.08 to 0.11 | 0.706 |
| | | 12041 | Education | Four years beyond HS | Elementary or less | 0.06 | 0.06 | -0.07 to 0.19 | 0.351 |
| | | 12041 | Residential setting | Small town | Rural | 0.20 | 0.05 | 0.09 to 0.30 | 0.000 |
| | | 12041 | Residential setting | Large city | Rural | 0.16 | 0.07 | 0.01 to 0.30 | 0.035 |

| Outcome | Model type | Analytic N | Variable | Level / comparison | Reference category | Coefficient | Standard error | 95% CI | p-value |
|---|---|---|---|---|---|---|---|---|---|
| | | 12041 | Residential setting | Suburb | Rural | 0.24 | 0.10 | 0.04 to 0.44 | 0.018 |
| | | 12041 | *Threshold / cut-point* | Never\|Rarely | | -1.86 | 0.08 | -2.03 to -1.70 | 0.000 |
| | | 12041 | *Threshold / cut-point* | Rarely\|Often | | -0.04 | 0.08 | -0.20 to 0.12 | 0.615 |
| | | 12041 | *Threshold / cut-point* | Often\|Always | | 0.66 | 0.08 | 0.50 to 0.82 | 0.000 |
| **Say in government#** | **Proportional-odds regression** | 12076 | Childhood predictor | No | Yes | -0.49 | 0.15 | -0.78 to -0.19 | 0.001 |
| | | 12076 | Age | Per 1-year increase | | 0.00 | 0.00 | -0.01 to 0.00 | 0.348 |
| | | 12076 | Gender | Female | Male | 0.08 | 0.05 | -0.02 to 0.18 | 0.105 |
| | | 12076 | Education | Secondary | Elementary or less | -0.25 | 0.06 | -0.37 to -0.14 | 0.000 |
| | | 12076 | Education | Four years beyond HS | Elementary or less | -0.15 | 0.10 | -0.35 to 0.04 | 0.129 |
| | | 12076 | Residential setting | Small town | Rural | 0.04 | 0.08 | -0.11 to 0.19 | 0.596 |
| | | 12076 | Residential setting | Large city | Rural | -0.11 | 0.09 | -0.29 to 0.07 | 0.236 |
| | | 12076 | Residential setting | Suburb | Rural | -0.42 | 0.13 | -0.68 to -0.16 | 0.001 |
| | | 12076 | *Threshold / cut-point* | Disagree\|Unsure | | -1.81 | 0.11 | -2.03 to -1.59 | 0.000 |
| | | 12076 | *Threshold / cut-point* | Unsure\|Agree | | -1.41 | 0.11 | -1.63 to -1.20 | 0.000 |
| **Extent of suffering§** | **Proportional-odds regression** | 12053 | Childhood predictor | No | Yes | 0.18 | 0.16 | -0.13 to 0.49 | 0.244 |
| | | 12053 | Age | Per 1-year increase | | 0.02 | 0.00 | 0.01 to 0.02 | 0.000 |
| | | 12053 | Gender | Female | Male | 0.25 | 0.04 | 0.17 to 0.33 | 0.000 |
| | | 12053 | Education | Secondary | Elementary or less | -0.15 | 0.05 | -0.24 to -0.06 | 0.001 |
| | | 12053 | Education | Four years beyond HS | Elementary or less | -0.27 | 0.08 | -0.42 to -0.12 | 0.000 |
| | | 12053 | Residential setting | Small town | Rural | 0.04 | 0.06 | -0.07 to 0.15 | 0.465 |
| | | 12053 | Residential setting | Large city | Rural | -0.14 | 0.08 | -0.29 to 0.01 | 0.070 |
| | | 12053 | Residential setting | Suburb | Rural | 0.06 | 0.11 | -0.17 to 0.28 | 0.626 |
| | | 12053 | *Threshold / cut-point* | Not at all\|Not very much | | 0.29 | 0.08 | 0.13 to 0.46 | 0.000 |
| | | 12053 | *Threshold / cut-point* | Not very much\|Some | | 0.71 | 0.08 | 0.55 to 0.88 | 0.000 |
| | | 12053 | *Threshold / cut-point* | Some\|A lot | | 2.15 | 0.09 | 1.98 to 2.32 | 0.000 |
| **Distress from life-threatening experiences§** | **Proportional-odds regression** | 12071 | Childhood predictor | No | Yes | 0.23 | 0.13 | -0.03 to 0.49 | 0.079 |
| | | 12071 | Age | Per 1-year increase | | 0.00 | 0.00 | 0.00 to 0.01 | 0.029 |
| | | 12071 | Gender | Female | Male | 0.07 | 0.05 | -0.02 to 0.16 | 0.107 |
| | | 12071 | Education | Secondary | Elementary or less | -0.27 | 0.05 | -0.36 to -0.18 | 0.000 |
| | | 12071 | Education | Four years beyond HS | Elementary or less | -0.45 | 0.08 | -0.59 to -0.30 | 0.000 |
| | | 12071 | Residential setting | Small town | Rural | -0.16 | 0.05 | -0.26 to -0.06 | 0.001 |
| | | 12071 | Residential setting | Large city | Rural | -0.22 | 0.07 | -0.36 to -0.08 | 0.003 |
| | | 12071 | Residential setting | Suburb | Rural | -0.17 | 0.12 | -0.40 to 0.07 | 0.162 |
| | | 12071 | *Threshold / cut-point* | Not at all\|Not very much | | -0.30 | 0.08 | -0.45 to -0.15 | 0.000 |
| | | 12071 | *Threshold / cut-point* | Not very much\|Some | | 0.05 | 0.08 | -0.11 to 0.20 | 0.556 |
| | | 12071 | *Threshold / cut-point* | Some\|A lot | | 1.06 | 0.08 | 0.91 to 1.22 | 0.000 |
| **Trust in others§** | **Proportional-odds regression** | 11804 | Childhood predictor | No | Yes | -0.17 | 0.14 | -0.45 to 0.10 | 0.214 |

| Outcome | Model type | Analytic N | Variable | Level / comparison | Reference category | Coefficient | Standard error | 95% CI | p-value |
|---|---|---|---|---|---|---|---|---|---|
| | | 11804 | Age | Per 1-year increase | | 0.00 | 0.00 | 0.00 to 0.01 | 0.013 |
| | | 11804 | Gender | Female | Male | -0.04 | 0.04 | -0.12 to 0.04 | 0.305 |
| | | 11804 | Education | Secondary | Elementary or less | -0.03 | 0.05 | -0.12 to 0.07 | 0.571 |
| | | 11804 | Education | Four years beyond HS | Elementary or less | -0.04 | 0.07 | -0.18 to 0.10 | 0.575 |
| | | 11804 | Residential setting | Small town | Rural | 0.24 | 0.05 | 0.14 to 0.34 | 0.000 |
| | | 11804 | Residential setting | Large city | Rural | 0.00 | 0.07 | -0.13 to 0.13 | 0.988 |
| | | 11804 | Residential setting | Suburb | Rural | -0.21 | 0.14 | -0.48 to 0.06 | 0.136 |
| | | 11804 | *Threshold / cut-point* | None\|Not very many | | -2.12 | 0.09 | -2.31 to -1.94 | 0.000 |
| | | 11804 | *Threshold / cut-point* | Not very many\|Some | | -1.33 | 0.09 | -1.51 to -1.15 | 0.000 |
| | | 11804 | *Threshold / cut-point* | Some\|Most | | 0.87 | 0.09 | 0.69 to 1.04 | 0.000 |
| | | 11804 | *Threshold / cut-point* | Most\|All | | 1.84 | 0.09 | 1.67 to 2.02 | 0.000 |

**eTable 11.** Table of coefficients for adjusted associations between bad or very bad quality of relationship with mother as predictor (very good quality as reference category) and 45 outcomes of interest and two flourishing composite indices.

Adjusted survey-weighted models use ANNUAL_WEIGHT_C1 with primary sampling unit (PSU) and strata and adjust for age, gender, education, and residential setting. Reference categories are shown in the table. Coefficients are on the original regression-coefficient scale: beta for continuous outcomes, log-odds for binary outcomes, and proportional-odds log coefficients for ordinal outcomes. Model intercepts are displayed for continuous and binary outcomes. Proportional-odds models do not have a single conventional intercept; their estimated ordinal cut-points/thresholds are displayed after the covariate coefficients, with standard errors (SEs), 95% CIs, and Wald p-values.

***Note:*** All results except p-values are rounded to the two decimals. P-values are rounded to three decimals. Values displayed as 0.000 indicate $P < 0.0005$ due to rounding, rather than an exact zero. Statistical significance was determined using unrounded P-values at the Bonferroni-adjusted threshold of $P < 0.0011$.

| Outcome | Model type | Analytic N | Variable | Level / comparison | Reference category | Coefficient | Standard error | 95% CI | p-value |
|---|---|---|---|---|---|---|---|---|---|
| **Belonging** | **Linear regression** | 12327 | *Intercept* | | | 8.68 | 0.11 | 8.46 to 8.90 | 0.000 |
| | | 12327 | Childhood predictor | Somewhat good | Very good | -0.26 | 0.13 | -0.53 to 0.00 | 0.048 |
| | | 12327 | Childhood predictor | Somewhat bad or very bad | Very good | -0.43 | 0.32 | -1.05 to 0.19 | 0.176 |
| | | 12327 | Age | Per 1-year increase | | -0.01 | 0.00 | -0.02 to -0.01 | 0.000 |
| | | 12327 | Gender | Female | Male | -0.18 | 0.06 | -0.31 to -0.06 | 0.003 |
| | | 12327 | Education | Secondary | Elementary or less | 0.21 | 0.07 | 0.06 to 0.35 | 0.006 |
| | | 12327 | Education | Four years beyond HS | Elementary or less | 0.29 | 0.10 | 0.08 to 0.49 | 0.006 |
| | | 12327 | Residential setting | Small town | Rural | 0.37 | 0.08 | 0.21 to 0.52 | 0.000 |
| | | 12327 | Residential setting | Large city | Rural | 0.43 | 0.11 | 0.23 to 0.64 | 0.000 |
| | | 12327 | Residential setting | Suburb | Rural | 0.02 | 0.19 | -0.34 to 0.38 | 0.918 |
| **Relationship contentment‡** | **Linear regression** | 12395 | *Intercept* | | | 8.15 | 0.13 | 7.90 to 8.40 | 0.000 |
| | | 12395 | Childhood predictor | Somewhat good | Very good | -0.45 | 0.17 | -0.78 to -0.12 | 0.008 |
| | | 12395 | Childhood predictor | Somewhat bad or very bad | Very good | -1.01 | 0.36 | -1.72 to -0.29 | 0.006 |
| | | 12395 | Age | Per 1-year increase | | -0.01 | 0.00 | -0.02 to -0.01 | 0.000 |
| | | 12395 | Gender | Female | Male | 0.37 | 0.07 | 0.23 to 0.51 | 0.000 |
| | | 12395 | Education | Secondary | Elementary or less | 0.14 | 0.08 | -0.01 to 0.29 | 0.065 |
| | | 12395 | Education | Four years beyond HS | Elementary or less | 0.27 | 0.12 | 0.04 to 0.50 | 0.022 |
| | | 12395 | Residential setting | Small town | Rural | 0.36 | 0.08 | 0.20 to 0.52 | 0.000 |
| | | 12395 | Residential setting | Large city | Rural | 0.08 | 0.10 | -0.12 to 0.27 | 0.433 |
| | | 12395 | Residential setting | Suburb | Rural | -0.27 | 0.18 | -0.62 to 0.07 | 0.123 |
| **Positive expectations** | **Linear regression** | 12352 | *Intercept* | | | 7.80 | 0.15 | 7.51 to 8.08 | 0.000 |
| | | 12352 | Childhood predictor | Somewhat good | Very good | -0.13 | 0.13 | -0.39 to 0.12 | 0.315 |
| | | 12352 | Childhood predictor | Somewhat bad or very bad | Very good | -0.05 | 0.30 | -0.64 to 0.53 | 0.857 |
| | | 12352 | Age | Per 1-year increase | | 0.00 | 0.00 | -0.01 to 0.00 | 0.149 |
| | | 12352 | Gender | Female | Male | 0.24 | 0.06 | 0.12 to 0.37 | 0.000 |
| | | 12352 | Education | Secondary | Elementary or less | 0.28 | 0.07 | 0.14 to 0.43 | 0.000 |
| | | 12352 | Education | Four years beyond HS | Elementary or less | 0.36 | 0.10 | 0.16 to 0.55 | 0.000 |
| | | 12352 | Residential setting | Small town | Rural | 0.44 | 0.09 | 0.27 to 0.61 | 0.000 |
| | | 12352 | Residential setting | Large city | Rural | 0.44 | 0.11 | 0.22 to 0.66 | 0.000 |
| | | 12352 | Residential setting | Suburb | Rural | 0.45 | 0.16 | 0.12 to 0.77 | 0.007 |

| Outcome | Model type | Analytic N | Variable | Level / comparison | Reference category | Coefficient | Standard error | 95% CI | p-value |
|---|---|---|---|---|---|---|---|---|---|
| **Worry about meeting normal monthly living expenses§§‡‡** | **Linear regression** | 12394 | *Intercept* | | | 5.78 | 0.19 | 5.40 to 6.16 | 0.000 |
| | | 12394 | Childhood predictor | Somewhat good | Very good | -0.12 | 0.17 | -0.46 to 0.21 | 0.467 |
| | | 12394 | Childhood predictor | Somewhat bad or very bad | Very good | -0.07 | 0.42 | -0.89 to 0.76 | 0.873 |
| | | 12394 | Age | Per 1-year increase | | 0.00 | 0.00 | 0.00 to 0.01 | 0.558 |
| | | 12394 | Gender | Female | Male | 0.34 | 0.09 | 0.16 to 0.52 | 0.000 |
| | | 12394 | Education | Secondary | Elementary or less | -0.47 | 0.11 | -0.68 to -0.26 | 0.000 |
| | | 12394 | Education | Four years beyond HS | Elementary or less | -0.64 | 0.15 | -0.94 to -0.35 | 0.000 |
| | | 12394 | Residential setting | Small town | Rural | 0.21 | 0.13 | -0.04 to 0.45 | 0.097 |
| | | 12394 | Residential setting | Large city | Rural | -0.24 | 0.15 | -0.54 to 0.06 | 0.122 |
| | | 12394 | Residential setting | Suburb | Rural | -0.24 | 0.25 | -0.73 to 0.25 | 0.339 |
| **Freedom** | **Linear regression** | 12366 | *Intercept* | | | 8.23 | 0.12 | 7.99 to 8.47 | 0.000 |
| | | 12366 | Childhood predictor | Somewhat good | Very good | -0.45 | 0.15 | -0.75 to -0.15 | 0.004 |
| | | 12366 | Childhood predictor | Somewhat bad or very bad | Very good | -0.55 | 0.40 | -1.33 to 0.23 | 0.168 |
| | | 12366 | Age | Per 1-year increase | | -0.01 | 0.00 | -0.01 to 0.00 | 0.003 |
| | | 12366 | Gender | Female | Male | -0.13 | 0.07 | -0.26 to 0.01 | 0.060 |
| | | 12366 | Education | Secondary | Elementary or less | 0.29 | 0.07 | 0.14 to 0.43 | 0.000 |
| | | 12366 | Education | Four years beyond HS | Elementary or less | 0.40 | 0.10 | 0.20 to 0.60 | 0.000 |
| | | 12366 | Residential setting | Small town | Rural | 0.36 | 0.07 | 0.22 to 0.51 | 0.000 |
| | | 12366 | Residential setting | Large city | Rural | 0.39 | 0.10 | 0.19 to 0.59 | 0.000 |
| | | 12366 | Residential setting | Suburb | Rural | 0.14 | 0.15 | -0.16 to 0.44 | 0.370 |
| **Give up some happiness now for greater happiness later‡** | **Linear regression** | 12335 | *Intercept* | | | 7.26 | 0.15 | 6.96 to 7.56 | 0.000 |
| | | 12335 | Childhood predictor | Somewhat good | Very good | 0.17 | 0.18 | -0.18 to 0.51 | 0.350 |
| | | 12335 | Childhood predictor | Somewhat bad or very bad | Very good | -0.10 | 0.37 | -0.83 to 0.64 | 0.799 |
| | | 12335 | Age | Per 1-year increase | | -0.01 | 0.00 | -0.02 to -0.01 | 0.000 |
| | | 12335 | Gender | Female | Male | 0.16 | 0.08 | 0.01 to 0.32 | 0.040 |
| | | 12335 | Education | Secondary | Elementary or less | 0.17 | 0.10 | -0.02 to 0.36 | 0.085 |
| | | 12335 | Education | Four years beyond HS | Elementary or less | 0.16 | 0.13 | -0.10 to 0.42 | 0.219 |
| | | 12335 | Residential setting | Small town | Rural | 0.42 | 0.10 | 0.22 to 0.61 | 0.000 |
| | | 12335 | Residential setting | Large city | Rural | 0.18 | 0.13 | -0.08 to 0.45 | 0.173 |
| | | 12335 | Residential setting | Suburb | Rural | 0.10 | 0.28 | -0.44 to 0.64 | 0.708 |
| **Have a long list of gratitude** | **Linear regression** | 12230 | *Intercept* | | | 7.41 | 0.15 | 7.11 to 7.71 | 0.000 |
| | | 12230 | Childhood predictor | Somewhat good | Very good | -0.16 | 0.16 | -0.48 to 0.15 | 0.308 |
| | | 12230 | Childhood predictor | Somewhat bad or very bad | Very good | -0.07 | 0.35 | -0.75 to 0.62 | 0.850 |
| | | 12230 | Age | Per 1-year increase | | -0.01 | 0.00 | -0.01 to 0.00 | 0.070 |
| | | 12230 | Gender | Female | Male | 0.21 | 0.07 | 0.07 to 0.35 | 0.003 |
| | | 12230 | Education | Secondary | Elementary or less | 0.31 | 0.08 | 0.15 to 0.47 | 0.000 |
| | | 12230 | Education | Four years beyond HS | Elementary or less | 0.40 | 0.11 | 0.18 to 0.63 | 0.000 |
| | | 12230 | Residential setting | Small town | Rural | 0.43 | 0.09 | 0.25 to 0.62 | 0.000 |
| | | 12230 | Residential setting | Large city | Rural | 0.43 | 0.13 | 0.17 to 0.70 | 0.001 |

| Outcome | Model type | Analytic N | Variable | Level / comparison | Reference category | Coefficient | Standard error | 95% CI | p-value |
|---|---|---|---|---|---|---|---|---|---|
| | | 12230 | Residential setting | Suburb | Rural | 0.52 | 0.17 | 0.19 to 0.86 | 0.002 |
| **Happiness‡** | **Linear regression** | 12390 | *Intercept* | | | 6.66 | 0.16 | 6.36 to 6.96 | 0.000 |
| | | 12390 | Childhood predictor | Somewhat good | Very good | -0.44 | 0.18 | -0.79 to -0.09 | 0.015 |
| | | 12390 | Childhood predictor | Somewhat bad or very bad | Very good | -0.64 | 0.35 | -1.33 to 0.05 | 0.071 |
| | | 12390 | Age | Per 1-year increase | | -0.02 | 0.00 | -0.02 to -0.01 | 0.000 |
| | | 12390 | Gender | Female | Male | 0.27 | 0.08 | 0.12 to 0.42 | 0.001 |
| | | 12390 | Education | Secondary | Elementary or less | 0.44 | 0.09 | 0.27 to 0.61 | 0.000 |
| | | 12390 | Education | Four years beyond HS | Elementary or less | 0.67 | 0.12 | 0.43 to 0.91 | 0.000 |
| | | 12390 | Residential setting | Small town | Rural | 0.40 | 0.10 | 0.20 to 0.60 | 0.000 |
| | | 12390 | Residential setting | Large city | Rural | 0.37 | 0.13 | 0.12 to 0.61 | 0.004 |
| | | 12390 | Residential setting | Suburb | Rural | -0.05 | 0.22 | -0.48 to 0.38 | 0.813 |
| **Remain hopeful for the future** | **Linear regression** | 12339 | *Intercept* | | | 8.17 | 0.12 | 7.93 to 8.41 | 0.000 |
| | | 12339 | Childhood predictor | Somewhat good | Very good | -0.11 | 0.14 | -0.38 to 0.16 | 0.434 |
| | | 12339 | Childhood predictor | Somewhat bad or very bad | Very good | -0.22 | 0.32 | -0.85 to 0.41 | 0.495 |
| | | 12339 | Age | Per 1-year increase | | -0.01 | 0.00 | -0.01 to 0.00 | 0.001 |
| | | 12339 | Gender | Female | Male | 0.01 | 0.06 | -0.10 to 0.12 | 0.866 |
| | | 12339 | Education | Secondary | Elementary or less | 0.39 | 0.06 | 0.27 to 0.52 | 0.000 |
| | | 12339 | Education | Four years beyond HS | Elementary or less | 0.61 | 0.09 | 0.42 to 0.79 | 0.000 |
| | | 12339 | Residential setting | Small town | Rural | 0.49 | 0.08 | 0.33 to 0.65 | 0.000 |
| | | 12339 | Residential setting | Large city | Rural | 0.47 | 0.10 | 0.27 to 0.68 | 0.000 |
| | | 12339 | Residential setting | Suburb | Rural | 0.43 | 0.18 | 0.08 to 0.78 | 0.017 |
| **Understand purpose in life‡** | **Linear regression** | 12318 | *Intercept* | | | 8.02 | 0.13 | 7.76 to 8.28 | 0.000 |
| | | 12318 | Childhood predictor | Somewhat good | Very good | -0.16 | 0.17 | -0.49 to 0.17 | 0.344 |
| | | 12318 | Childhood predictor | Somewhat bad or very bad | Very good | -0.10 | 0.30 | -0.69 to 0.50 | 0.747 |
| | | 12318 | Age | Per 1-year increase | | -0.01 | 0.00 | -0.02 to -0.01 | 0.000 |
| | | 12318 | Gender | Female | Male | 0.12 | 0.07 | -0.02 to 0.26 | 0.102 |
| | | 12318 | Education | Secondary | Elementary or less | 0.44 | 0.08 | 0.28 to 0.59 | 0.000 |
| | | 12318 | Education | Four years beyond HS | Elementary or less | 0.74 | 0.10 | 0.55 to 0.93 | 0.000 |
| | | 12318 | Residential setting | Small town | Rural | 0.42 | 0.09 | 0.24 to 0.60 | 0.000 |
| | | 12318 | Residential setting | Large city | Rural | 0.42 | 0.12 | 0.19 to 0.65 | 0.000 |
| | | 12318 | Residential setting | Suburb | Rural | 0.46 | 0.15 | 0.17 to 0.75 | 0.002 |
| **Life satisfaction‡** | **Linear regression** | 12371 | *Intercept* | | | 7.10 | 0.15 | 6.81 to 7.39 | 0.000 |
| | | 12371 | Childhood predictor | Somewhat good | Very good | -0.43 | 0.17 | -0.77 to -0.10 | 0.012 |
| | | 12371 | Childhood predictor | Somewhat bad or very bad | Very good | -0.51 | 0.33 | -1.15 to 0.13 | 0.122 |
| | | 12371 | Age | Per 1-year increase | | -0.01 | 0.00 | -0.02 to -0.01 | 0.000 |
| | | 12371 | Gender | Female | Male | 0.30 | 0.08 | 0.15 to 0.45 | 0.000 |
| | | 12371 | Education | Secondary | Elementary or less | 0.30 | 0.09 | 0.13 to 0.48 | 0.001 |
| | | 12371 | Education | Four years beyond HS | Elementary or less | 0.47 | 0.12 | 0.24 to 0.70 | 0.000 |
| | | 12371 | Residential setting | Small town | Rural | 0.31 | 0.09 | 0.13 to 0.49 | 0.001 |
| | | 12371 | Residential setting | Large city | Rural | 0.26 | 0.12 | 0.02 to 0.50 | 0.036 |

| Outcome | Model type | Analytic N | Variable | Level / comparison | Reference category | Coefficient | Standard error | 95% CI | p-value |
|---|---|---|---|---|---|---|---|---|---|
| | | 12371 | Residential setting | Suburb | Rural | -0.17 | 0.21 | -0.58 to 0.24 | 0.427 |
| **Loneliness§** | **Linear regression** | 12391 | *Intercept* | | | 4.19 | 0.17 | 3.86 to 4.51 | 0.000 |
| | | 12391 | Childhood predictor | Somewhat good | Very good | 0.58 | 0.16 | 0.25 to 0.90 | 0.001 |
| | | 12391 | Childhood predictor | Somewhat bad or very bad | Very good | 1.30 | 0.42 | 0.47 to 2.12 | 0.002 |
| | | 12391 | Age | Per 1-year increase | | 0.00 | 0.00 | -0.01 to 0.01 | 0.803 |
| | | 12391 | Gender | Female | Male | -0.15 | 0.09 | -0.32 to 0.02 | 0.079 |
| | | 12391 | Education | Secondary | Elementary or less | -0.30 | 0.10 | -0.49 to -0.11 | 0.002 |
| | | 12391 | Education | Four years beyond HS | Elementary or less | -0.62 | 0.15 | -0.90 to -0.33 | 0.000 |
| | | 12391 | Residential setting | Small town | Rural | -0.36 | 0.10 | -0.56 to -0.16 | 0.000 |
| | | 12391 | Residential setting | Large city | Rural | -0.52 | 0.14 | -0.79 to -0.25 | 0.000 |
| | | 12391 | Residential setting | Suburb | Rural | -0.18 | 0.23 | -0.63 to 0.28 | 0.454 |
| **Mental health‡** | **Linear regression** | 12395 | *Intercept* | | | 8.05 | 0.14 | 7.78 to 8.33 | 0.000 |
| | | 12395 | Childhood predictor | Somewhat good | Very good | -0.59 | 0.17 | -0.91 to -0.27 | 0.000 |
| | | 12395 | Childhood predictor | Somewhat bad or very bad | Very good | -0.56 | 0.33 | -1.22 to 0.10 | 0.094 |
| | | 12395 | Age | Per 1-year increase | | -0.02 | 0.00 | -0.03 to -0.02 | 0.000 |
| | | 12395 | Gender | Female | Male | -0.12 | 0.07 | -0.26 to 0.02 | 0.087 |
| | | 12395 | Education | Secondary | Elementary or less | 0.41 | 0.08 | 0.26 to 0.56 | 0.000 |
| | | 12395 | Education | Four years beyond HS | Elementary or less | 0.51 | 0.11 | 0.29 to 0.73 | 0.000 |
| | | 12395 | Residential setting | Small town | Rural | 0.40 | 0.09 | 0.22 to 0.58 | 0.000 |
| | | 12395 | Residential setting | Large city | Rural | 0.46 | 0.12 | 0.23 to 0.69 | 0.000 |
| | | 12395 | Residential setting | Suburb | Rural | 0.20 | 0.18 | -0.16 to 0.55 | 0.276 |
| **Perceived support from others** | **Linear regression** | 12381 | *Intercept* | | | 6.66 | 0.16 | 6.35 to 6.97 | 0.000 |
| | | 12381 | Childhood predictor | Somewhat good | Very good | -0.27 | 0.19 | -0.64 to 0.09 | 0.145 |
| | | 12381 | Childhood predictor | Somewhat bad or very bad | Very good | -1.61 | 0.39 | -2.37 to -0.85 | 0.000 |
| | | 12381 | Age | Per 1-year increase | | -0.01 | 0.00 | -0.02 to -0.01 | 0.000 |
| | | 12381 | Gender | Female | Male | 0.56 | 0.09 | 0.39 to 0.74 | 0.000 |
| | | 12381 | Education | Secondary | Elementary or less | 0.19 | 0.10 | -0.01 to 0.38 | 0.057 |
| | | 12381 | Education | Four years beyond HS | Elementary or less | 0.12 | 0.14 | -0.15 to 0.39 | 0.395 |
| | | 12381 | Residential setting | Small town | Rural | 0.19 | 0.11 | -0.02 to 0.40 | 0.078 |
| | | 12381 | Residential setting | Large city | Rural | -0.47 | 0.14 | -0.75 to -0.20 | 0.001 |
| | | 12381 | Residential setting | Suburb | Rural | -0.27 | 0.20 | -0.67 to 0.12 | 0.174 |
| **Physical health‡** | **Linear regression** | 12398 | *Intercept* | | | 8.43 | 0.14 | 8.15 to 8.71 | 0.000 |
| | | 12398 | Childhood predictor | Somewhat good | Very good | -0.40 | 0.16 | -0.72 to -0.08 | 0.014 |
| | | 12398 | Childhood predictor | Somewhat bad or very bad | Very good | -1.40 | 0.36 | -2.11 to -0.70 | 0.000 |
| | | 12398 | Age | Per 1-year increase | | -0.04 | 0.00 | -0.05 to -0.04 | 0.000 |
| | | 12398 | Gender | Female | Male | -0.34 | 0.08 | -0.49 to -0.19 | 0.000 |
| | | 12398 | Education | Secondary | Elementary or less | 0.36 | 0.08 | 0.21 to 0.51 | 0.000 |
| | | 12398 | Education | Four years beyond HS | Elementary or less | 0.65 | 0.11 | 0.44 to 0.86 | 0.000 |
| | | 12398 | Residential setting | Small town | Rural | 0.48 | 0.09 | 0.30 to 0.66 | 0.000 |
| | | 12398 | Residential setting | Large city | Rural | 0.64 | 0.11 | 0.43 to 0.86 | 0.000 |

| Outcome | Model type | Analytic N | Variable | Level / comparison | Reference category | Coefficient | Standard error | 95% CI | p-value |
|---|---|---|---|---|---|---|---|---|---|
| | | 12398 | Residential setting | Suburb | Rural | 0.18 | 0.18 | -0.17 to 0.52 | 0.311 |
| **Act to promote good‡** | **Linear regression** | 12355 | *Intercept* | | | 8.07 | 0.11 | 7.86 to 8.28 | 0.000 |
| | | 12355 | Childhood predictor | Somewhat good | Very good | -0.16 | 0.15 | -0.45 to 0.13 | 0.285 |
| | | 12355 | Childhood predictor | Somewhat bad or very bad | Very good | -0.19 | 0.28 | -0.75 to 0.36 | 0.492 |
| | | 12355 | Age | Per 1-year increase | | -0.01 | 0.00 | -0.01 to 0.00 | 0.001 |
| | | 12355 | Gender | Female | Male | 0.12 | 0.06 | 0.00 to 0.23 | 0.057 |
| | | 12355 | Education | Secondary | Elementary or less | 0.38 | 0.06 | 0.26 to 0.50 | 0.000 |
| | | 12355 | Education | Four years beyond HS | Elementary or less | 0.54 | 0.08 | 0.38 to 0.69 | 0.000 |
| | | 12355 | Residential setting | Small town | Rural | 0.61 | 0.07 | 0.46 to 0.75 | 0.000 |
| | | 12355 | Residential setting | Large city | Rural | 0.63 | 0.10 | 0.43 to 0.82 | 0.000 |
| | | 12355 | Residential setting | Suburb | Rural | 0.44 | 0.16 | 0.13 to 0.75 | 0.006 |
| **Relationship satisfaction‡** | **Linear regression** | 12360 | *Intercept* | | | 7.91 | 0.13 | 7.66 to 8.17 | 0.000 |
| | | 12360 | Childhood predictor | Somewhat good | Very good | -0.41 | 0.15 | -0.71 to -0.11 | 0.008 |
| | | 12360 | Childhood predictor | Somewhat bad or very bad | Very good | -0.96 | 0.39 | -1.72 to -0.20 | 0.014 |
| | | 12360 | Age | Per 1-year increase | | -0.01 | 0.00 | -0.01 to 0.00 | 0.000 |
| | | 12360 | Gender | Female | Male | 0.35 | 0.07 | 0.21 to 0.49 | 0.000 |
| | | 12360 | Education | Secondary | Elementary or less | -0.04 | 0.08 | -0.19 to 0.12 | 0.623 |
| | | 12360 | Education | Four years beyond HS | Elementary or less | 0.03 | 0.11 | -0.18 to 0.25 | 0.758 |
| | | 12360 | Residential setting | Small town | Rural | 0.44 | 0.09 | 0.27 to 0.61 | 0.000 |
| | | 12360 | Residential setting | Large city | Rural | 0.33 | 0.11 | 0.12 to 0.55 | 0.002 |
| | | 12360 | Residential setting | Suburb | Rural | 0.04 | 0.16 | -0.27 to 0.35 | 0.782 |
| **Show love often** | **Linear regression** | 12383 | *Intercept* | | | 8.30 | 0.12 | 8.05 to 8.54 | 0.000 |
| | | 12383 | Childhood predictor | Somewhat good | Very good | -0.25 | 0.16 | -0.56 to 0.05 | 0.103 |
| | | 12383 | Childhood predictor | Somewhat bad or very bad | Very good | 0.15 | 0.32 | -0.47 to 0.77 | 0.637 |
| | | 12383 | Age | Per 1-year increase | | -0.01 | 0.00 | -0.01 to 0.00 | 0.002 |
| | | 12383 | Gender | Female | Male | 0.21 | 0.07 | 0.08 to 0.34 | 0.002 |
| | | 12383 | Education | Secondary | Elementary or less | 0.17 | 0.07 | 0.03 to 0.31 | 0.015 |
| | | 12383 | Education | Four years beyond HS | Elementary or less | 0.04 | 0.11 | -0.18 to 0.25 | 0.728 |
| | | 12383 | Residential setting | Small town | Rural | 0.19 | 0.08 | 0.04 to 0.35 | 0.016 |
| | | 12383 | Residential setting | Large city | Rural | 0.01 | 0.12 | -0.22 to 0.23 | 0.950 |
| | | 12383 | Residential setting | Suburb | Rural | -0.11 | 0.20 | -0.50 to 0.27 | 0.561 |
| **Life evaluation five years from now** | **Linear regression** | 11296 | *Intercept* | | | 7.69 | 0.14 | 7.42 to 7.97 | 0.000 |
| | | 11296 | Childhood predictor | Somewhat good | Very good | -0.31 | 0.18 | -0.65 to 0.03 | 0.077 |
| | | 11296 | Childhood predictor | Somewhat bad or very bad | Very good | -0.96 | 0.37 | -1.69 to -0.22 | 0.011 |
| | | 11296 | Age | Per 1-year increase | | -0.02 | 0.00 | -0.03 to -0.02 | 0.000 |
| | | 11296 | Gender | Female | Male | 0.20 | 0.07 | 0.06 to 0.34 | 0.007 |
| | | 11296 | Education | Secondary | Elementary or less | 0.29 | 0.08 | 0.14 to 0.45 | 0.000 |
| | | 11296 | Education | Four years beyond HS | Elementary or less | 0.57 | 0.12 | 0.34 to 0.79 | 0.000 |
| | | 11296 | Residential setting | Small town | Rural | 0.47 | 0.09 | 0.29 to 0.64 | 0.000 |
| | | 11296 | Residential setting | Large city | Rural | 0.53 | 0.11 | 0.31 to 0.75 | 0.000 |

| Outcome | Model type | Analytic N | Variable | Level / comparison | Reference category | Coefficient | Standard error | 95% CI | p-value |
|---|---|---|---|---|---|---|---|---|---|
| | | 11296 | Residential setting | Suburb | Rural | 0.40 | 0.22 | -0.04 to 0.84 | 0.073 |
| **Life evaluation today** | **Linear regression** | 12295 | *Intercept* | | | 5.55 | 0.17 | 5.22 to 5.88 | 0.000 |
| | | 12295 | Childhood predictor | Somewhat good | Very good | -0.20 | 0.17 | -0.52 to 0.13 | 0.240 |
| | | 12295 | Childhood predictor | Somewhat bad or very bad | Very good | -1.20 | 0.36 | -1.91 to -0.49 | 0.001 |
| | | 12295 | Age | Per 1-year increase | | -0.01 | 0.00 | -0.01 to 0.00 | 0.011 |
| | | 12295 | Gender | Female | Male | 0.49 | 0.08 | 0.33 to 0.66 | 0.000 |
| | | 12295 | Education | Secondary | Elementary or less | 0.23 | 0.09 | 0.06 to 0.40 | 0.008 |
| | | 12295 | Education | Four years beyond HS | Elementary or less | 0.16 | 0.14 | -0.11 to 0.44 | 0.249 |
| | | 12295 | Residential setting | Small town | Rural | 0.26 | 0.10 | 0.06 to 0.47 | 0.011 |
| | | 12295 | Residential setting | Large city | Rural | 0.12 | 0.13 | -0.14 to 0.38 | 0.376 |
| | | 12295 | Residential setting | Suburb | Rural | -0.04 | 0.21 | -0.45 to 0.38 | 0.856 |
| **Worry about safety, food, or housing§§‡‡** | **Linear regression** | 12407 | *Intercept* | | | 6.25 | 0.19 | 5.88 to 6.62 | 0.000 |
| | | 12407 | Childhood predictor | Somewhat good | Very good | -0.13 | 0.17 | -0.47 to 0.20 | 0.437 |
| | | 12407 | Childhood predictor | Somewhat bad or very bad | Very good | 0.23 | 0.44 | -0.62 to 1.09 | 0.595 |
| | | 12407 | Age | Per 1-year increase | | -0.01 | 0.00 | -0.02 to 0.00 | 0.011 |
| | | 12407 | Gender | Female | Male | 0.33 | 0.09 | 0.15 to 0.50 | 0.000 |
| | | 12407 | Education | Secondary | Elementary or less | -0.56 | 0.11 | -0.77 to -0.36 | 0.000 |
| | | 12407 | Education | Four years beyond HS | Elementary or less | -0.78 | 0.17 | -1.10 to -0.45 | 0.000 |
| | | 12407 | Residential setting | Small town | Rural | 0.38 | 0.13 | 0.12 to 0.64 | 0.005 |
| | | 12407 | Residential setting | Large city | Rural | -0.08 | 0.18 | -0.43 to 0.27 | 0.642 |
| | | 12407 | Residential setting | Suburb | Rural | 0.17 | 0.26 | -0.34 to 0.68 | 0.516 |
| **Life worthwhile‡** | **Linear regression** | 12294 | *Intercept* | | | 6.65 | 0.15 | 6.37 to 6.94 | 0.000 |
| | | 12294 | Childhood predictor | Somewhat good | Very good | -0.28 | 0.16 | -0.59 to 0.02 | 0.072 |
| | | 12294 | Childhood predictor | Somewhat bad or very bad | Very good | -1.12 | 0.43 | -1.97 to -0.27 | 0.010 |
| | | 12294 | Age | Per 1-year increase | | -0.01 | 0.00 | -0.01 to 0.00 | 0.002 |
| | | 12294 | Gender | Female | Male | 0.43 | 0.07 | 0.29 to 0.58 | 0.000 |
| | | 12294 | Education | Secondary | Elementary or less | 0.45 | 0.08 | 0.29 to 0.60 | 0.000 |
| | | 12294 | Education | Four years beyond HS | Elementary or less | 0.58 | 0.12 | 0.34 to 0.83 | 0.000 |
| | | 12294 | Residential setting | Small town | Rural | 0.48 | 0.10 | 0.28 to 0.68 | 0.000 |
| | | 12294 | Residential setting | Large city | Rural | 0.47 | 0.13 | 0.22 to 0.72 | 0.000 |
| | | 12294 | Residential setting | Suburb | Rural | 0.44 | 0.20 | 0.04 to 0.84 | 0.033 |
| **Flourishing (with financial indicators)** | **Linear regression** | 11951 | *Intercept* | | | 7.00 | 0.08 | 6.84 to 7.16 | 0.000 |
| | | 11951 | Childhood predictor | Somewhat good | Very good | -0.24 | 0.10 | -0.43 to -0.05 | 0.013 |
| | | 11951 | Childhood predictor | Somewhat bad or very bad | Very good | -0.60 | 0.19 | -0.97 to -0.24 | 0.001 |
| | | 11951 | Age | Per 1-year increase | | -0.01 | 0.00 | -0.02 to -0.01 | 0.000 |
| | | 11951 | Gender | Female | Male | 0.10 | 0.04 | 0.01 to 0.18 | 0.024 |
| | | 11951 | Education | Secondary | Elementary or less | 0.33 | 0.05 | 0.24 to 0.42 | 0.000 |
| | | 11951 | Education | Four years beyond HS | Elementary or less | 0.50 | 0.06 | 0.37 to 0.62 | 0.000 |
| | | 11951 | Residential setting | Small town | Rural | 0.32 | 0.06 | 0.21 to 0.43 | 0.000 |

| Outcome | Model type | Analytic N | Variable | Level / comparison | Reference category | Coefficient | Standard error | 95% CI | p-value |
|---|---|---|---|---|---|---|---|---|---|
| | | 11951 | Residential setting | Large city | Rural | 0.36 | 0.07 | 0.23 to 0.50 | 0.000 |
| | | 11951 | Residential setting | Suburb | Rural | 0.14 | 0.13 | -0.12 to 0.39 | 0.301 |
| **Flourishing (without financial indicators)** | **Linear regression** | 11978 | *Intercept* | | | 7.62 | 0.09 | 7.45 to 7.79 | 0.000 |
| | | 11978 | Childhood predictor | Somewhat good | Very good | -0.32 | 0.11 | -0.55 to -0.10 | 0.005 |
| | | 11978 | Childhood predictor | Somewhat bad or very bad | Very good | -0.71 | 0.21 | -1.13 to -0.29 | 0.001 |
| | | 11978 | Age | Per 1-year increase | | -0.02 | 0.00 | -0.02 to -0.01 | 0.000 |
| | | 11978 | Gender | Female | Male | 0.18 | 0.05 | 0.09 to 0.27 | 0.000 |
| | | 11978 | Education | Secondary | Elementary or less | 0.30 | 0.05 | 0.20 to 0.39 | 0.000 |
| | | 11978 | Education | Four years beyond HS | Elementary or less | 0.46 | 0.07 | 0.32 to 0.59 | 0.000 |
| | | 11978 | Residential setting | Small town | Rural | 0.45 | 0.06 | 0.32 to 0.57 | 0.000 |
| | | 11978 | Residential setting | Large city | Rural | 0.40 | 0.08 | 0.25 to 0.55 | 0.000 |
| | | 11978 | Residential setting | Suburb | Rural | 0.15 | 0.13 | -0.10 to 0.41 | 0.245 |
| **Feel close to someone** | **Logistic regression** | 12408 | *Intercept* | | | 1.69 | 0.14 | 1.42 to 1.96 | 0.000 |
| | | 12408 | Childhood predictor | Somewhat good | Very good | -0.32 | 0.15 | -0.61 to -0.03 | 0.034 |
| | | 12408 | Childhood predictor | Somewhat bad or very bad | Very good | -0.36 | 0.25 | -0.86 to 0.14 | 0.158 |
| | | 12408 | Age | Per 1-year increase | | -0.01 | 0.00 | -0.01 to 0.00 | 0.018 |
| | | 12408 | Gender | Female | Male | 0.03 | 0.07 | -0.10 to 0.16 | 0.635 |
| | | 12408 | Education | Secondary | Elementary or less | 0.16 | 0.08 | -0.01 to 0.32 | 0.060 |
| | | 12408 | Education | Four years beyond HS | Elementary or less | 0.14 | 0.13 | -0.11 to 0.40 | 0.273 |
| | | 12408 | Residential setting | Small town | Rural | 0.09 | 0.11 | -0.12 to 0.31 | 0.400 |
| | | 12408 | Residential setting | Large city | Rural | -0.11 | 0.13 | -0.36 to 0.14 | 0.382 |
| | | 12408 | Residential setting | Suburb | Rural | -0.19 | 0.19 | -0.56 to 0.18 | 0.314 |
| **Donated to charity** | **Logistic regression** | 12371 | *Intercept* | | | -0.31 | 0.13 | -0.56 to -0.06 | 0.014 |
| | | 12371 | Childhood predictor | Somewhat good | Very good | -0.31 | 0.14 | -0.58 to -0.04 | 0.028 |
| | | 12371 | Childhood predictor | Somewhat bad or very bad | Very good | -0.10 | 0.23 | -0.55 to 0.35 | 0.655 |
| | | 12371 | Age | Per 1-year increase | | 0.00 | 0.00 | 0.00 to 0.00 | 0.615 |
| | | 12371 | Gender | Female | Male | -0.03 | 0.05 | -0.13 to 0.06 | 0.497 |
| | | 12371 | Education | Secondary | Elementary or less | -0.08 | 0.06 | -0.19 to 0.04 | 0.193 |
| | | 12371 | Education | Four years beyond HS | Elementary or less | -0.15 | 0.10 | -0.34 to 0.05 | 0.143 |
| | | 12371 | Residential setting | Small town | Rural | -0.06 | 0.09 | -0.24 to 0.12 | 0.496 |
| | | 12371 | Residential setting | Large city | Rural | -0.43 | 0.11 | -0.66 to -0.21 | 0.000 |
| | | 12371 | Residential setting | Suburb | Rural | -0.18 | 0.19 | -0.55 to 0.18 | 0.328 |
| **Have any health problems** | **Logistic regression** | 12326 | *Intercept* | | | -2.05 | 0.10 | -2.25 to -1.85 | 0.000 |
| | | 12326 | Childhood predictor | Somewhat good | Very good | 0.18 | 0.10 | -0.02 to 0.39 | 0.079 |
| | | 12326 | Childhood predictor | Somewhat bad or very bad | Very good | 0.09 | 0.20 | -0.30 to 0.49 | 0.637 |
| | | 12326 | Age | Per 1-year increase | | 0.03 | 0.00 | 0.02 to 0.03 | 0.000 |
| | | 12326 | Gender | Female | Male | 0.23 | 0.05 | 0.13 to 0.33 | 0.000 |
| | | 12326 | Education | Secondary | Elementary or less | -0.14 | 0.06 | -0.26 to -0.03 | 0.016 |
| | | 12326 | Education | Four years beyond HS | Elementary or less | -0.27 | 0.09 | -0.45 to -0.08 | 0.005 |
| | | 12326 | Residential setting | Small town | Rural | 0.13 | 0.06 | 0.00 to 0.26 | 0.042 |

| Outcome | Model type | Analytic N | Variable | Level / comparison | Reference category | Coefficient | Standard error | 95% CI | p-value |
|---|---|---|---|---|---|---|---|---|---|
| | | 12326 | Residential setting | Large city | Rural | -0.12 | 0.09 | -0.29 to 0.05 | 0.162 |
| | | 12326 | Residential setting | Suburb | Rural | 0.08 | 0.14 | -0.20 to 0.35 | 0.586 |
| **Helped a stranger** | **Logistic regression** | 12289 | *Intercept* | | | 0.66 | 0.12 | 0.43 to 0.89 | 0.000 |
| | | 12289 | Childhood predictor | Somewhat good | Very good | -0.03 | 0.12 | -0.26 to 0.20 | 0.776 |
| | | 12289 | Childhood predictor | Somewhat bad or very bad | Very good | -0.07 | 0.22 | -0.50 to 0.37 | 0.769 |
| | | 12289 | Age | Per 1-year increase | | -0.01 | 0.00 | -0.01 to 0.00 | 0.000 |
| | | 12289 | Gender | Female | Male | -0.32 | 0.05 | -0.41 to -0.23 | 0.000 |
| | | 12289 | Education | Secondary | Elementary or less | -0.06 | 0.06 | -0.18 to 0.05 | 0.263 |
| | | 12289 | Education | Four years beyond HS | Elementary or less | -0.08 | 0.09 | -0.25 to 0.10 | 0.377 |
| | | 12289 | Residential setting | Small town | Rural | 0.11 | 0.09 | -0.06 to 0.28 | 0.225 |
| | | 12289 | Residential setting | Large city | Rural | -0.16 | 0.10 | -0.36 to 0.04 | 0.114 |
| | | 12289 | Residential setting | Suburb | Rural | 0.04 | 0.18 | -0.31 to 0.38 | 0.828 |
| **Volunteered at an organization** | **Logistic regression** | 12357 | *Intercept* | | | -0.27 | 0.14 | -0.54 to 0.00 | 0.048 |
| | | 12357 | Childhood predictor | Somewhat good | Very good | -0.50 | 0.15 | -0.80 to -0.20 | 0.001 |
| | | 12357 | Childhood predictor | Somewhat bad or very bad | Very good | -0.09 | 0.25 | -0.57 to 0.39 | 0.717 |
| | | 12357 | Age | Per 1-year increase | | -0.01 | 0.00 | -0.01 to 0.00 | 0.001 |
| | | 12357 | Gender | Female | Male | -0.28 | 0.05 | -0.38 to -0.17 | 0.000 |
| | | 12357 | Education | Secondary | Elementary or less | -0.15 | 0.06 | -0.27 to -0.02 | 0.023 |
| | | 12357 | Education | Four years beyond HS | Elementary or less | 0.02 | 0.10 | -0.17 to 0.22 | 0.816 |
| | | 12357 | Residential setting | Small town | Rural | -0.11 | 0.11 | -0.32 to 0.10 | 0.312 |
| | | 12357 | Residential setting | Large city | Rural | -0.58 | 0.13 | -0.83 to -0.33 | 0.000 |
| | | 12357 | Residential setting | Suburb | Rural | -0.38 | 0.16 | -0.70 to -0.07 | 0.019 |
| **Cigarette smoking** | **Logistic regression** | 11917 | *Intercept* | | | -2.03 | 0.16 | -2.35 to -1.71 | 0.000 |
| | | 11917 | Childhood predictor | Somewhat good | Very good | 0.11 | 0.17 | -0.22 to 0.44 | 0.509 |
| | | 11917 | Childhood predictor | Somewhat bad or very bad | Very good | 0.01 | 0.38 | -0.73 to 0.75 | 0.985 |
| | | 11917 | Age | Per 1-year increase | | 0.01 | 0.00 | 0.00 to 0.01 | 0.010 |
| | | 11917 | Gender | Female | Male | -2.83 | 0.19 | -3.20 to -2.46 | 0.000 |
| | | 11917 | Education | Secondary | Elementary or less | -0.37 | 0.10 | -0.56 to -0.18 | 0.000 |
| | | 11917 | Education | Four years beyond HS | Elementary or less | -0.57 | 0.17 | -0.90 to -0.24 | 0.001 |
| | | 11917 | Residential setting | Small town | Rural | -0.06 | 0.10 | -0.26 to 0.15 | 0.568 |
| | | 11917 | Residential setting | Large city | Rural | 0.13 | 0.15 | -0.16 to 0.42 | 0.368 |
| | | 11917 | Residential setting | Suburb | Rural | 0.84 | 0.20 | 0.44 to 1.23 | 0.000 |
| **Alcohol use** | **Logistic regression** | 11904 | *Intercept* | | | -1.88 | 0.15 | -2.17 to -1.58 | 0.000 |
| | | 11904 | Childhood predictor | Somewhat good | Very good | -0.07 | 0.19 | -0.44 to 0.30 | 0.714 |
| | | 11904 | Childhood predictor | Somewhat bad or very bad | Very good | 0.80 | 0.34 | 0.14 to 1.47 | 0.018 |
| | | 11904 | Age | Per 1-year increase | | 0.00 | 0.00 | -0.01 to 0.00 | 0.769 |
| | | 11904 | Gender | Female | Male | -2.80 | 0.16 | -3.11 to -2.48 | 0.000 |
| | | 11904 | Education | Secondary | Elementary or less | -0.21 | 0.10 | -0.41 to -0.02 | 0.032 |
| | | 11904 | Education | Four years beyond HS | Elementary or less | -0.49 | 0.16 | -0.80 to -0.17 | 0.003 |
| | | 11904 | Residential setting | Small town | Rural | 0.23 | 0.11 | 0.02 to 0.44 | 0.031 |

| Outcome | Model type | Analytic N | Variable | Level / comparison | Reference category | Coefficient | Standard error | 95% CI | p-value |
|---|---|---|---|---|---|---|---|---|---|
| | | 11904 | Residential setting | Large city | Rural | 0.36 | 0.14 | 0.09 to 0.64 | 0.011 |
| | | 11904 | Residential setting | Suburb | Rural | 0.65 | 0.22 | 0.21 to 1.08 | 0.004 |
| **Government approval§** | **Proportional-odds regression** | 12194 | Childhood predictor | Somewhat good | Very good | -0.25 | 0.10 | -0.44 to -0.06 | 0.010 |
| | | 12194 | Childhood predictor | Somewhat bad or very bad | Very good | -0.27 | 0.17 | -0.59 to 0.06 | 0.113 |
| | | 12194 | Age | Per 1-year increase | | 0.00 | 0.00 | -0.01 to 0.00 | 0.021 |
| | | 12194 | Gender | Female | Male | 0.05 | 0.04 | -0.03 to 0.14 | 0.218 |
| | | 12194 | Education | Secondary | Elementary or less | -0.26 | 0.05 | -0.36 to -0.16 | 0.000 |
| | | 12194 | Education | Four years beyond HS | Elementary or less | -0.35 | 0.08 | -0.51 to -0.18 | 0.000 |
| | | 12194 | Residential setting | Small town | Rural | 0.10 | 0.07 | -0.03 to 0.23 | 0.121 |
| | | 12194 | Residential setting | Large city | Rural | -0.07 | 0.08 | -0.22 to 0.08 | 0.346 |
| | | 12194 | Residential setting | Suburb | Rural | -0.43 | 0.16 | -0.74 to -0.12 | 0.006 |
| | | 12194 | *Threshold / cut-point* | Strongly disapprove\|Somewhat disapprove | | -2.95 | 0.10 | -3.14 to -2.76 | 0.000 |
| | | 12194 | *Threshold / cut-point* | Somewhat disapprove\|Neither | | -2.29 | 0.09 | -2.47 to -2.10 | 0.000 |
| | | 12194 | *Threshold / cut-point* | Neither\|Somewhat approve | | -1.93 | 0.09 | -2.11 to -1.75 | 0.000 |
| | | 12194 | *Threshold / cut-point* | Somewhat approve\|Strongly approve | | -0.48 | 0.09 | -0.65 to -0.30 | 0.000 |
| **Feeling capable§** | **Proportional-odds regression** | 12350 | Childhood predictor | Somewhat good | Very good | -0.16 | 0.08 | -0.33 to 0.00 | 0.054 |
| | | 12350 | Childhood predictor | Somewhat bad or very bad | Very good | 0.27 | 0.22 | -0.15 to 0.69 | 0.210 |
| | | 12350 | Age | Per 1-year increase | | 0.00 | 0.00 | 0.00 to 0.01 | 0.189 |
| | | 12350 | Gender | Female | Male | -0.06 | 0.04 | -0.15 to 0.02 | 0.154 |
| | | 12350 | Education | Secondary | Elementary or less | 0.16 | 0.05 | 0.07 to 0.25 | 0.001 |
| | | 12350 | Education | Four years beyond HS | Elementary or less | 0.10 | 0.07 | -0.05 to 0.24 | 0.184 |
| | | 12350 | Residential setting | Small town | Rural | 0.17 | 0.05 | 0.07 to 0.27 | 0.001 |
| | | 12350 | Residential setting | Large city | Rural | 0.18 | 0.08 | 0.03 to 0.33 | 0.016 |
| | | 12350 | Residential setting | Suburb | Rural | 0.23 | 0.09 | 0.05 to 0.41 | 0.010 |
| | | 12350 | *Threshold / cut-point* | Never\|Rarely | | -2.36 | 0.09 | -2.54 to -2.18 | 0.000 |
| | | 12350 | *Threshold / cut-point* | Rarely\|Often | | -0.64 | 0.08 | -0.81 to -0.48 | 0.000 |
| | | 12350 | *Threshold / cut-point* | Often\|Always | | 0.03 | 0.08 | -0.13 to 0.19 | 0.715 |
| **Unable to control worry§** | **Proportional-odds regression** | 12300 | Childhood predictor | Somewhat good | Very good | 0.14 | 0.08 | -0.02 to 0.30 | 0.079 |
| | | 12300 | Childhood predictor | Somewhat bad or very bad | Very good | 0.43 | 0.21 | 0.02 to 0.84 | 0.040 |
| | | 12300 | Age | Per 1-year increase | | 0.00 | 0.00 | 0.00 to 0.01 | 0.001 |
| | | 12300 | Gender | Female | Male | 0.16 | 0.04 | 0.08 to 0.24 | 0.000 |
| | | 12300 | Education | Secondary | Elementary or less | -0.27 | 0.05 | -0.36 to -0.18 | 0.000 |
| | | 12300 | Education | Four years beyond HS | Elementary or less | -0.44 | 0.07 | -0.58 to -0.29 | 0.000 |
| | | 12300 | Residential setting | Small town | Rural | 0.05 | 0.05 | -0.05 to 0.15 | 0.313 |
| | | 12300 | Residential setting | Large city | Rural | -0.01 | 0.07 | -0.15 to 0.13 | 0.879 |
| | | 12300 | Residential setting | Suburb | Rural | -0.07 | 0.10 | -0.27 to 0.12 | 0.449 |

| Outcome | Model type | Analytic N | Variable | Level / comparison | Reference category | Coefficient | Standard error | 95% CI | p-value |
|---|---|---|---|---|---|---|---|---|---|
| | | 12300 | *Threshold / cut-point* | Not at all\|Several days | | 0.08 | 0.08 | -0.07 to 0.24 | 0.300 |
| | | 12300 | *Threshold / cut-point* | Several days\|More than half the days | | 0.99 | 0.08 | 0.84 to 1.15 | 0.000 |
| | | 12300 | *Threshold / cut-point* | More than half the days\|Nearly every day | | 1.84 | 0.08 | 1.68 to 2.00 | 0.000 |
| **Depressive symptoms§** | **Proportional-odds regression** | 12325 | Childhood predictor | Somewhat good | Very good | 0.04 | 0.09 | -0.13 to 0.21 | 0.639 |
| | | 12325 | Childhood predictor | Somewhat bad or very bad | Very good | 0.49 | 0.18 | 0.15 to 0.84 | 0.005 |
| | | 12325 | Age | Per 1-year increase | | 0.00 | 0.00 | 0.00 to 0.01 | 0.147 |
| | | 12325 | Gender | Female | Male | 0.09 | 0.04 | 0.01 to 0.17 | 0.024 |
| | | 12325 | Education | Secondary | Elementary or less | -0.21 | 0.05 | -0.30 to -0.12 | 0.000 |
| | | 12325 | Education | Four years beyond HS | Elementary or less | -0.42 | 0.07 | -0.56 to -0.29 | 0.000 |
| | | 12325 | Residential setting | Small town | Rural | 0.00 | 0.05 | -0.10 to 0.10 | 0.989 |
| | | 12325 | Residential setting | Large city | Rural | -0.09 | 0.07 | -0.22 to 0.05 | 0.227 |
| | | 12325 | Residential setting | Suburb | Rural | -0.18 | 0.11 | -0.40 to 0.04 | 0.118 |
| | | 12325 | *Threshold / cut-point* | Not at all\|Several days | | -0.24 | 0.08 | -0.39 to -0.08 | 0.003 |
| | | 12325 | *Threshold / cut-point* | Several days\|More than half the days | | 0.83 | 0.08 | 0.68 to 0.99 | 0.000 |
| | | 12325 | *Threshold / cut-point* | More than half the days\|Nearly every day | | 1.83 | 0.08 | 1.67 to 1.99 | 0.000 |
| **Perceived discrimination§** | **Proportional-odds regression** | 12258 | Childhood predictor | Somewhat good | Very good | 0.00 | 0.09 | -0.19 to 0.18 | 0.958 |
| | | 12258 | Childhood predictor | Somewhat bad or very bad | Very good | 0.18 | 0.19 | -0.19 to 0.56 | 0.336 |
| | | 12258 | Age | Per 1-year increase | | 0.00 | 0.00 | -0.01 to 0.00 | 0.075 |
| | | 12258 | Gender | Female | Male | -0.14 | 0.04 | -0.23 to -0.05 | 0.002 |
| | | 12258 | Education | Secondary | Elementary or less | -0.24 | 0.05 | -0.33 to -0.14 | 0.000 |
| | | 12258 | Education | Four years beyond HS | Elementary or less | -0.29 | 0.09 | -0.46 to -0.11 | 0.001 |
| | | 12258 | Residential setting | Small town | Rural | -0.22 | 0.06 | -0.33 to -0.11 | 0.000 |
| | | 12258 | Residential setting | Large city | Rural | -0.24 | 0.07 | -0.37 to -0.10 | 0.001 |
| | | 12258 | Residential setting | Suburb | Rural | -0.33 | 0.10 | -0.52 to -0.14 | 0.001 |
| | | 12258 | *Threshold / cut-point* | Never\|Rarely | | -0.13 | 0.09 | -0.30 to 0.04 | 0.144 |
| | | 12258 | *Threshold / cut-point* | Rarely\|Often | | 0.87 | 0.09 | 0.70 to 1.05 | 0.000 |
| | | 12258 | *Threshold / cut-point* | Often\|Always | | 1.49 | 0.09 | 1.32 to 1.67 | 0.000 |
| **Feeling nervous or anxious§** | **Proportional-odds regression** | 12352 | Childhood predictor | Somewhat good | Very good | 0.16 | 0.08 | 0.00 to 0.32 | 0.056 |
| | | 12352 | Childhood predictor | Somewhat bad or very bad | Very good | 0.43 | 0.23 | -0.03 to 0.88 | 0.066 |
| | | 12352 | Age | Per 1-year increase | | 0.01 | 0.00 | 0.00 to 0.01 | 0.000 |
| | | 12352 | Gender | Female | Male | 0.31 | 0.04 | 0.23 to 0.39 | 0.000 |
| | | 12352 | Education | Secondary | Elementary or less | -0.29 | 0.05 | -0.38 to -0.20 | 0.000 |
| | | 12352 | Education | Four years beyond HS | Elementary or less | -0.48 | 0.08 | -0.63 to -0.33 | 0.000 |
| | | 12352 | Residential setting | Small town | Rural | 0.11 | 0.05 | 0.01 to 0.22 | 0.040 |
| | | 12352 | Residential setting | Large city | Rural | -0.03 | 0.07 | -0.16 to 0.10 | 0.646 |

| Outcome | Model type | Analytic N | Variable | Level / comparison | Reference category | Coefficient | Standard error | 95% CI | p-value |
|---|---|---|---|---|---|---|---|---|---|
| | | 12352 | Residential setting | Suburb | Rural | -0.03 | 0.12 | -0.26 to 0.21 | 0.828 |
| | | 12352 | *Threshold / cut-point* | Not at all\|Several days | | 0.14 | 0.08 | -0.01 to 0.30 | 0.064 |
| | | 12352 | *Threshold / cut-point* | Several days\|More than half the days | | 1.14 | 0.08 | 0.99 to 1.30 | 0.000 |
| | | 12352 | *Threshold / cut-point* | More than half the days\|Nearly every day | | 2.08 | 0.08 | 1.92 to 2.24 | 0.000 |
| **Forgiveness§** | **Proportional-odds regression** | 12327 | Childhood predictor | Somewhat good | Very good | -0.02 | 0.10 | -0.21 to 0.18 | 0.855 |
| | | 12327 | Childhood predictor | Somewhat bad or very bad | Very good | 0.38 | 0.24 | -0.08 to 0.85 | 0.107 |
| | | 12327 | Age | Per 1-year increase | | 0.00 | 0.00 | 0.00 to 0.00 | 0.888 |
| | | 12327 | Gender | Female | Male | 0.09 | 0.04 | 0.01 to 0.18 | 0.034 |
| | | 12327 | Education | Secondary | Elementary or less | -0.07 | 0.05 | -0.18 to 0.03 | 0.172 |
| | | 12327 | Education | Four years beyond HS | Elementary or less | -0.20 | 0.08 | -0.35 to -0.05 | 0.009 |
| | | 12327 | Residential setting | Small town | Rural | 0.03 | 0.05 | -0.07 to 0.14 | 0.526 |
| | | 12327 | Residential setting | Large city | Rural | 0.00 | 0.08 | -0.16 to 0.16 | 0.989 |
| | | 12327 | Residential setting | Suburb | Rural | 0.04 | 0.12 | -0.19 to 0.28 | 0.719 |
| | | 12327 | *Threshold / cut-point* | Never\|Rarely | | -2.40 | 0.09 | -2.57 to -2.22 | 0.000 |
| | | 12327 | *Threshold / cut-point* | Rarely\|Often | | -1.06 | 0.09 | -1.23 to -0.89 | 0.000 |
| | | 12327 | *Threshold / cut-point* | Often\|Always | | -0.41 | 0.08 | -0.57 to -0.25 | 0.000 |
| **Support reducing differences in income levels§** | **Proportional-odds regression** | 12192 | Childhood predictor | Somewhat good | Very good | -1.12 | 0.11 | -1.33 to -0.91 | 0.000 |
| | | 12192 | Childhood predictor | Somewhat bad or very bad | Very good | -0.73 | 0.25 | -1.22 to -0.24 | 0.003 |
| | | 12192 | Age | Per 1-year increase | | 0.00 | 0.00 | -0.01 to 0.00 | 0.124 |
| | | 12192 | Gender | Female | Male | -0.05 | 0.06 | -0.16 to 0.07 | 0.432 |
| | | 12192 | Education | Secondary | Elementary or less | -0.09 | 0.07 | -0.23 to 0.04 | 0.176 |
| | | 12192 | Education | Four years beyond HS | Elementary or less | -0.29 | 0.11 | -0.51 to -0.08 | 0.008 |
| | | 12192 | Residential setting | Small town | Rural | 0.00 | 0.12 | -0.24 to 0.24 | 1.000 |
| | | 12192 | Residential setting | Large city | Rural | 0.05 | 0.14 | -0.22 to 0.32 | 0.710 |
| | | 12192 | Residential setting | Suburb | Rural | -0.12 | 0.17 | -0.46 to 0.22 | 0.478 |
| | | 12192 | *Threshold / cut-point* | Strongly disagree\|Somewhat disagree | | -4.04 | 0.18 | -4.38 to -3.69 | 0.000 |
| | | 12192 | *Threshold / cut-point* | Somewhat disagree\|Neither | | -3.49 | 0.17 | -3.81 to -3.16 | 0.000 |
| | | 12192 | *Threshold / cut-point* | Neither\|Somewhat agree | | -3.03 | 0.16 | -3.34 to -2.72 | 0.000 |
| | | 12192 | *Threshold / cut-point* | Somewhat agree\|Strongly agree | | -1.42 | 0.14 | -1.69 to -1.16 | 0.000 |
| **Have little interest in doing things§** | **Proportional-odds regression** | 12320 | Childhood predictor | Somewhat good | Very good | -0.02 | 0.09 | -0.20 to 0.16 | 0.824 |
| | | 12320 | Childhood predictor | Somewhat bad or very bad | Very good | 0.10 | 0.15 | -0.19 to 0.40 | 0.493 |
| | | 12320 | Age | Per 1-year increase | | 0.00 | 0.00 | 0.00 to 0.01 | 0.044 |
| | | 12320 | Gender | Female | Male | 0.04 | 0.04 | -0.04 to 0.12 | 0.384 |
| | | 12320 | Education | Secondary | Elementary or less | 0.07 | 0.05 | -0.02 to 0.17 | 0.132 |
| | | 12320 | Education | Four years beyond HS | Elementary or less | 0.08 | 0.08 | -0.07 to 0.24 | 0.283 |

| Outcome | Model type | Analytic N | Variable | Level / comparison | Reference category | Coefficient | Standard error | 95% CI | p-value |
|---|---|---|---|---|---|---|---|---|---|
| | | 12320 | Residential setting | Small town | Rural | 0.28 | 0.06 | 0.15 to 0.40 | 0.000 |
| | | 12320 | Residential setting | Large city | Rural | 0.11 | 0.08 | -0.05 to 0.27 | 0.178 |
| | | 12320 | Residential setting | Suburb | Rural | 0.28 | 0.15 | -0.02 to 0.57 | 0.065 |
| | | 12320 | *Threshold / cut-point* | Not at all\|Several days | | -0.48 | 0.09 | -0.65 to -0.31 | 0.000 |
| | | 12320 | *Threshold / cut-point* | Several days\|More than half the days | | 0.45 | 0.09 | 0.27 to 0.62 | 0.000 |
| | | 12320 | *Threshold / cut-point* | More than half the days\|Nearly every day | | 1.22 | 0.09 | 1.05 to 1.40 | 0.000 |
| **Various aspects of life are balanced§** | **Proportional-odds regression** | 12246 | Childhood predictor | Somewhat good | Very good | -0.03 | 0.08 | -0.19 to 0.12 | 0.685 |
| | | 12246 | Childhood predictor | Somewhat bad or very bad | Very good | 0.39 | 0.19 | 0.03 to 0.76 | 0.034 |
| | | 12246 | Age | Per 1-year increase | | 0.00 | 0.00 | 0.00 to 0.01 | 0.075 |
| | | 12246 | Gender | Female | Male | -0.03 | 0.04 | -0.11 to 0.05 | 0.455 |
| | | 12246 | Education | Secondary | Elementary or less | 0.04 | 0.04 | -0.05 to 0.13 | 0.351 |
| | | 12246 | Education | Four years beyond HS | Elementary or less | 0.11 | 0.07 | -0.02 to 0.24 | 0.108 |
| | | 12246 | Residential setting | Small town | Rural | 0.10 | 0.05 | 0.00 to 0.21 | 0.053 |
| | | 12246 | Residential setting | Large city | Rural | 0.09 | 0.07 | -0.04 to 0.22 | 0.155 |
| | | 12246 | Residential setting | Suburb | Rural | 0.37 | 0.11 | 0.15 to 0.58 | 0.001 |
| | | 12246 | *Threshold / cut-point* | Never\|Rarely | | -2.02 | 0.09 | -2.19 to -1.85 | 0.000 |
| | | 12246 | *Threshold / cut-point* | Rarely\|Often | | 0.15 | 0.08 | -0.01 to 0.31 | 0.058 |
| | | 12246 | *Threshold / cut-point* | Often\|Always | | 0.95 | 0.08 | 0.78 to 1.11 | 0.000 |
| **Obeying the law§** | **Proportional-odds regression** | 12377 | Childhood predictor | Somewhat good | Very good | -1.44 | 0.13 | -1.69 to -1.19 | 0.000 |
| | | 12377 | Childhood predictor | Somewhat bad or very bad | Very good | -1.16 | 0.26 | -1.67 to -0.66 | 0.000 |
| | | 12377 | Age | Per 1-year increase | | 0.00 | 0.00 | -0.01 to 0.00 | 0.469 |
| | | 12377 | Gender | Female | Male | -0.13 | 0.07 | -0.28 to 0.01 | 0.077 |
| | | 12377 | Education | Secondary | Elementary or less | 0.06 | 0.09 | -0.12 to 0.25 | 0.501 |
| | | 12377 | Education | Four years beyond HS | Elementary or less | -0.25 | 0.12 | -0.48 to -0.01 | 0.037 |
| | | 12377 | Residential setting | Small town | Rural | -0.12 | 0.14 | -0.39 to 0.14 | 0.363 |
| | | 12377 | Residential setting | Large city | Rural | -0.30 | 0.18 | -0.64 to 0.05 | 0.093 |
| | | 12377 | Residential setting | Suburb | Rural | -0.36 | 0.21 | -0.78 to 0.06 | 0.094 |
| | | 12377 | *Threshold / cut-point* | Strongly disagree\|Somewhat disagree | | -5.42 | 0.25 | -5.92 to -4.92 | 0.000 |
| | | 12377 | *Threshold / cut-point* | Somewhat disagree\|Neither | | -4.81 | 0.21 | -5.23 to -4.39 | 0.000 |
| | | 12377 | *Threshold / cut-point* | Neither\|Somewhat agree | | -4.29 | 0.21 | -4.70 to -3.88 | 0.000 |
| | | 12377 | *Threshold / cut-point* | Somewhat agree\|Strongly agree | | -2.42 | 0.18 | -2.76 to -2.07 | 0.000 |
| **Feel at peace§** | **Proportional-odds regression** | 12318 | Childhood predictor | Somewhat good | Very good | -0.25 | 0.08 | -0.40 to -0.09 | 0.002 |
| | | 12318 | Childhood predictor | Somewhat bad or very bad | Very good | 0.23 | 0.19 | -0.15 to 0.61 | 0.231 |
| | | 12318 | Age | Per 1-year increase | | 0.01 | 0.00 | 0.00 to 0.01 | 0.000 |
| | | 12318 | Gender | Female | Male | 0.01 | 0.04 | -0.06 to 0.09 | 0.754 |

| Outcome | Model type | Analytic N | Variable | Level / comparison | Reference category | Coefficient | Standard error | 95% CI | p-value |
|---|---|---|---|---|---|---|---|---|---|
| | | 12318 | Education | Secondary | Elementary or less | 0.01 | 0.05 | -0.08 to 0.11 | 0.752 |
| | | 12318 | Education | Four years beyond HS | Elementary or less | 0.05 | 0.06 | -0.08 to 0.18 | 0.441 |
| | | 12318 | Residential setting | Small town | Rural | 0.21 | 0.05 | 0.11 to 0.31 | 0.000 |
| | | 12318 | Residential setting | Large city | Rural | 0.15 | 0.07 | 0.01 to 0.30 | 0.039 |
| | | 12318 | Residential setting | Suburb | Rural | 0.23 | 0.10 | 0.04 to 0.43 | 0.018 |
| | | 12318 | *Threshold / cut-point* | Never\|Rarely | | -1.89 | 0.08 | -2.05 to -1.73 | 0.000 |
| | | 12318 | *Threshold / cut-point* | Rarely\|Often | | -0.06 | 0.08 | -0.22 to 0.09 | 0.433 |
| | | 12318 | *Threshold / cut-point* | Often\|Always | | 0.63 | 0.08 | 0.47 to 0.79 | 0.000 |
| **Say in government#** | **Proportional-odds regression** | 12353 | Childhood predictor | Somewhat good | Very good | -0.20 | 0.11 | -0.41 to 0.01 | 0.064 |
| | | 12353 | Childhood predictor | Somewhat bad or very bad | Very good | 0.08 | 0.25 | -0.40 to 0.57 | 0.732 |
| | | 12353 | Age | Per 1-year increase | | 0.00 | 0.00 | -0.01 to 0.00 | 0.389 |
| | | 12353 | Gender | Female | Male | 0.10 | 0.05 | 0.00 to 0.20 | 0.049 |
| | | 12353 | Education | Secondary | Elementary or less | -0.25 | 0.06 | -0.37 to -0.13 | 0.000 |
| | | 12353 | Education | Four years beyond HS | Elementary or less | -0.14 | 0.10 | -0.33 to 0.05 | 0.158 |
| | | 12353 | Residential setting | Small town | Rural | 0.02 | 0.08 | -0.13 to 0.16 | 0.842 |
| | | 12353 | Residential setting | Large city | Rural | -0.14 | 0.09 | -0.32 to 0.04 | 0.137 |
| | | 12353 | Residential setting | Suburb | Rural | -0.44 | 0.13 | -0.69 to -0.18 | 0.001 |
| | | 12353 | *Threshold / cut-point* | Disagree\|Unsure | | -1.81 | 0.11 | -2.03 to -1.58 | 0.000 |
| | | 12353 | *Threshold / cut-point* | Unsure\|Agree | | -1.41 | 0.11 | -1.62 to -1.19 | 0.000 |
| **Extent of suffering§** | **Proportional-odds regression** | 12329 | Childhood predictor | Somewhat good | Very good | 0.16 | 0.09 | -0.01 to 0.32 | 0.067 |
| | | 12329 | Childhood predictor | Somewhat bad or very bad | Very good | 0.60 | 0.21 | 0.19 to 1.01 | 0.004 |
| | | 12329 | Age | Per 1-year increase | | 0.02 | 0.00 | 0.01 to 0.02 | 0.000 |
| | | 12329 | Gender | Female | Male | 0.27 | 0.04 | 0.19 to 0.35 | 0.000 |
| | | 12329 | Education | Secondary | Elementary or less | -0.14 | 0.05 | -0.23 to -0.05 | 0.002 |
| | | 12329 | Education | Four years beyond HS | Elementary or less | -0.26 | 0.07 | -0.41 to -0.12 | 0.000 |
| | | 12329 | Residential setting | Small town | Rural | 0.06 | 0.06 | -0.06 to 0.17 | 0.332 |
| | | 12329 | Residential setting | Large city | Rural | -0.12 | 0.08 | -0.27 to 0.03 | 0.119 |
| | | 12329 | Residential setting | Suburb | Rural | 0.07 | 0.11 | -0.14 to 0.29 | 0.497 |
| | | 12329 | *Threshold / cut-point* | Not at all\|Not very much | | 0.32 | 0.08 | 0.16 to 0.49 | 0.000 |
| | | 12329 | *Threshold / cut-point* | Not very much\|Some | | 0.74 | 0.08 | 0.58 to 0.90 | 0.000 |
| | | 12329 | *Threshold / cut-point* | Some\|A lot | | 2.18 | 0.09 | 2.02 to 2.35 | 0.000 |
| **Distress from life-threatening experiences§** | **Proportional-odds regression** | 12346 | Childhood predictor | Somewhat good | Very good | 0.08 | 0.09 | -0.09 to 0.26 | 0.357 |
| | | 12346 | Childhood predictor | Somewhat bad or very bad | Very good | 0.30 | 0.19 | -0.06 to 0.67 | 0.106 |
| | | 12346 | Age | Per 1-year increase | | 0.00 | 0.00 | 0.00 to 0.01 | 0.018 |
| | | 12346 | Gender | Female | Male | 0.07 | 0.05 | -0.02 to 0.16 | 0.105 |
| | | 12346 | Education | Secondary | Elementary or less | -0.26 | 0.05 | -0.35 to -0.17 | 0.000 |
| | | 12346 | Education | Four years beyond HS | Elementary or less | -0.44 | 0.07 | -0.59 to -0.30 | 0.000 |
| | | 12346 | Residential setting | Small town | Rural | -0.17 | 0.05 | -0.27 to -0.08 | 0.000 |

| Outcome | Model type | Analytic N | Variable | Level / comparison | Reference category | Coefficient | Standard error | 95% CI | p-value |
|---|---|---|---|---|---|---|---|---|---|
| | | 12346 | Residential setting | Large city | Rural | -0.23 | 0.07 | -0.37 to -0.09 | 0.001 |
| | | 12346 | Residential setting | Suburb | Rural | -0.18 | 0.12 | -0.42 to 0.05 | 0.125 |
| | | 12346 | *Threshold / cut-point* | Not at all\|Not very much | | -0.29 | 0.08 | -0.44 to -0.14 | 0.000 |
| | | 12346 | *Threshold / cut-point* | Not very much\|Some | | 0.05 | 0.08 | -0.10 to 0.20 | 0.500 |
| | | 12346 | *Threshold / cut-point* | Some\|A lot | | 1.07 | 0.08 | 0.92 to 1.22 | 0.000 |
| **Trust in others§** | **Proportional-odds regression** | 12067 | Childhood predictor | Somewhat good | Very good | -0.25 | 0.10 | -0.46 to -0.05 | 0.016 |
| | | 12067 | Childhood predictor | Somewhat bad or very bad | Very good | 0.46 | 0.20 | 0.07 to 0.84 | 0.020 |
| | | 12067 | Age | Per 1-year increase | | 0.00 | 0.00 | 0.00 to 0.01 | 0.035 |
| | | 12067 | Gender | Female | Male | -0.04 | 0.04 | -0.12 to 0.04 | 0.325 |
| | | 12067 | Education | Secondary | Elementary or less | -0.04 | 0.05 | -0.13 to 0.05 | 0.364 |
| | | 12067 | Education | Four years beyond HS | Elementary or less | -0.05 | 0.07 | -0.20 to 0.09 | 0.443 |
| | | 12067 | Residential setting | Small town | Rural | 0.23 | 0.05 | 0.13 to 0.33 | 0.000 |
| | | 12067 | Residential setting | Large city | Rural | -0.01 | 0.07 | -0.14 to 0.12 | 0.880 |
| | | 12067 | Residential setting | Suburb | Rural | -0.20 | 0.14 | -0.48 to 0.08 | 0.154 |
| | | 12067 | *Threshold / cut-point* | None\|Not very many | | -2.16 | 0.09 | -2.34 to -1.98 | 0.000 |
| | | 12067 | *Threshold / cut-point* | Not very many\|Some | | -1.38 | 0.09 | -1.55 to -1.20 | 0.000 |
| | | 12067 | *Threshold / cut-point* | Some\|Most | | 0.83 | 0.09 | 0.66 to 1.00 | 0.000 |
| | | 12067 | *Threshold / cut-point* | Most\|All | | 1.80 | 0.09 | 1.62 to 1.97 | 0.000 |

**eTable 12.** Table of coefficients for adjusted associations between bad or very bad quality of relationship with father as predictor (very good quality as reference category) and 45 outcomes of interest and two flourishing composite indices.

Adjusted survey-weighted models use ANNUAL_WEIGHT_C1 with primary sampling unit (PSU) and strata and adjust for age, gender, education, and residential setting. Reference categories are shown in the table. Coefficients are on the original regression-coefficient scale: beta for continuous outcomes, log-odds for binary outcomes, and proportional-odds log coefficients for ordinal outcomes. Model intercepts are displayed for continuous and binary outcomes. Proportional-odds models do not have a single conventional intercept; their estimated ordinal cut-points/thresholds are displayed after the covariate coefficients, with standard errors (SEs), 95% CIs, and Wald p-values.

***Note:*** All results except p-values are rounded to the two decimals. P-values are rounded to three decimals. Values displayed as 0.000 indicate $P < 0.0005$ due to rounding, rather than an exact zero. Statistical significance was determined using unrounded P-values at the Bonferroni-adjusted threshold of $P < 0.0011$.

| Outcome | Model type | Analytic N | Variable | Level / comparison | Reference category | Coefficient | Standard error | 95% CI | p-value |
|---|---|---|---|---|---|---|---|---|---|
| **Belonging** | **Linear regression** | 12084 | *Intercept* | | | 8.72 | 0.11 | 8.50 to 8.95 | 0.000 |
| | | 12084 | Childhood predictor | Somewhat good | Very good | -0.36 | 0.13 | -0.62 to -0.10 | 0.007 |
| | | 12084 | Childhood predictor | Somewhat bad or very bad | Very good | -0.03 | 0.24 | -0.49 to 0.44 | 0.910 |
| | | 12084 | Age | Per 1-year increase | | -0.01 | 0.00 | -0.02 to -0.01 | 0.000 |
| | | 12084 | Gender | Female | Male | -0.19 | 0.06 | -0.31 to -0.07 | 0.003 |
| | | 12084 | Education | Secondary | Elementary or less | 0.19 | 0.07 | 0.05 to 0.34 | 0.010 |
| | | 12084 | Education | Four years beyond HS | Elementary or less | 0.32 | 0.11 | 0.12 to 0.53 | 0.002 |
| | | 12084 | Residential setting | Small town | Rural | 0.36 | 0.08 | 0.20 to 0.51 | 0.000 |
| | | 12084 | Residential setting | Large city | Rural | 0.42 | 0.10 | 0.21 to 0.62 | 0.000 |
| | | 12084 | Residential setting | Suburb | Rural | -0.01 | 0.18 | -0.37 to 0.36 | 0.972 |
| **Relationship contentment‡** | **Linear regression** | 12146 | *Intercept* | | | 8.19 | 0.13 | 7.93 to 8.44 | 0.000 |
| | | 12146 | Childhood predictor | Somewhat good | Very good | -0.35 | 0.15 | -0.63 to -0.06 | 0.018 |
| | | 12146 | Childhood predictor | Somewhat bad or very bad | Very good | -0.95 | 0.28 | -1.49 to -0.41 | 0.001 |
| | | 12146 | Age | Per 1-year increase | | -0.01 | 0.00 | -0.02 to -0.01 | 0.000 |
| | | 12146 | Gender | Female | Male | 0.37 | 0.07 | 0.23 to 0.51 | 0.000 |
| | | 12146 | Education | Secondary | Elementary or less | 0.16 | 0.07 | 0.01 to 0.30 | 0.037 |
| | | 12146 | Education | Four years beyond HS | Elementary or less | 0.29 | 0.12 | 0.06 to 0.52 | 0.015 |
| | | 12146 | Residential setting | Small town | Rural | 0.38 | 0.08 | 0.22 to 0.54 | 0.000 |
| | | 12146 | Residential setting | Large city | Rural | 0.07 | 0.10 | -0.12 to 0.26 | 0.453 |
| | | 12146 | Residential setting | Suburb | Rural | -0.29 | 0.18 | -0.65 to 0.06 | 0.104 |
| **Positive expectations** | **Linear regression** | 12107 | *Intercept* | | | 7.79 | 0.15 | 7.51 to 8.08 | 0.000 |
| | | 12107 | Childhood predictor | Somewhat good | Very good | 0.00 | 0.11 | -0.22 to 0.21 | 0.977 |
| | | 12107 | Childhood predictor | Somewhat bad or very bad | Very good | -0.07 | 0.26 | -0.57 to 0.44 | 0.793 |
| | | 12107 | Age | Per 1-year increase | | 0.00 | 0.00 | -0.01 to 0.00 | 0.165 |
| | | 12107 | Gender | Female | Male | 0.24 | 0.06 | 0.11 to 0.36 | 0.000 |
| | | 12107 | Education | Secondary | Elementary or less | 0.29 | 0.07 | 0.15 to 0.43 | 0.000 |
| | | 12107 | Education | Four years beyond HS | Elementary or less | 0.37 | 0.10 | 0.18 to 0.57 | 0.000 |
| | | 12107 | Residential setting | Small town | Rural | 0.43 | 0.08 | 0.27 to 0.60 | 0.000 |
| | | 12107 | Residential setting | Large city | Rural | 0.43 | 0.11 | 0.21 to 0.65 | 0.000 |
| | | 12107 | Residential setting | Suburb | Rural | 0.41 | 0.16 | 0.09 to 0.73 | 0.013 |

| Outcome | Model type | Analytic N | Variable | Level / comparison | Reference category | Coefficient | Standard error | 95% CI | p-value |
|---|---|---|---|---|---|---|---|---|---|
| **Worry about meeting normal monthly living expenses§§‡‡** | **Linear regression** | 12149 | *Intercept* | | | 5.73 | 0.19 | 5.35 to 6.11 | 0.000 |
| | | 12149 | Childhood predictor | Somewhat good | Very good | 0.04 | 0.17 | -0.29 to 0.36 | 0.823 |
| | | 12149 | Childhood predictor | Somewhat bad or very bad | Very good | 0.32 | 0.36 | -0.39 to 1.03 | 0.370 |
| | | 12149 | Age | Per 1-year increase | | 0.00 | 0.00 | 0.00 to 0.01 | 0.464 |
| | | 12149 | Gender | Female | Male | 0.37 | 0.09 | 0.19 to 0.55 | 0.000 |
| | | 12149 | Education | Secondary | Elementary or less | -0.44 | 0.11 | -0.66 to -0.22 | 0.000 |
| | | 12149 | Education | Four years beyond HS | Elementary or less | -0.63 | 0.15 | -0.93 to -0.34 | 0.000 |
| | | 12149 | Residential setting | Small town | Rural | 0.21 | 0.13 | -0.04 to 0.46 | 0.098 |
| | | 12149 | Residential setting | Large city | Rural | -0.20 | 0.15 | -0.50 to 0.10 | 0.188 |
| | | 12149 | Residential setting | Suburb | Rural | -0.23 | 0.25 | -0.72 to 0.27 | 0.373 |
| **Freedom** | **Linear regression** | 12121 | *Intercept* | | | 8.21 | 0.12 | 7.96 to 8.45 | 0.000 |
| | | 12121 | Childhood predictor | Somewhat good | Very good | -0.35 | 0.13 | -0.60 to -0.10 | 0.007 |
| | | 12121 | Childhood predictor | Somewhat bad or very bad | Very good | -0.18 | 0.27 | -0.70 to 0.34 | 0.504 |
| | | 12121 | Age | Per 1-year increase | | -0.01 | 0.00 | -0.01 to 0.00 | 0.004 |
| | | 12121 | Gender | Female | Male | -0.11 | 0.07 | -0.24 to 0.02 | 0.094 |
| | | 12121 | Education | Secondary | Elementary or less | 0.29 | 0.08 | 0.15 to 0.44 | 0.000 |
| | | 12121 | Education | Four years beyond HS | Elementary or less | 0.41 | 0.10 | 0.21 to 0.62 | 0.000 |
| | | 12121 | Residential setting | Small town | Rural | 0.37 | 0.08 | 0.22 to 0.52 | 0.000 |
| | | 12121 | Residential setting | Large city | Rural | 0.40 | 0.10 | 0.20 to 0.60 | 0.000 |
| | | 12121 | Residential setting | Suburb | Rural | 0.14 | 0.16 | -0.16 to 0.45 | 0.358 |
| **Give up some happiness now for greater happiness later‡** | **Linear regression** | 12090 | *Intercept* | | | 7.26 | 0.16 | 6.96 to 7.57 | 0.000 |
| | | 12090 | Childhood predictor | Somewhat good | Very good | 0.06 | 0.15 | -0.22 to 0.35 | 0.669 |
| | | 12090 | Childhood predictor | Somewhat bad or very bad | Very good | 0.17 | 0.29 | -0.40 to 0.74 | 0.560 |
| | | 12090 | Age | Per 1-year increase | | -0.01 | 0.00 | -0.02 to 0.00 | 0.000 |
| | | 12090 | Gender | Female | Male | 0.16 | 0.08 | 0.00 to 0.31 | 0.046 |
| | | 12090 | Education | Secondary | Elementary or less | 0.18 | 0.10 | -0.01 to 0.36 | 0.070 |
| | | 12090 | Education | Four years beyond HS | Elementary or less | 0.18 | 0.14 | -0.08 to 0.45 | 0.181 |
| | | 12090 | Residential setting | Small town | Rural | 0.37 | 0.10 | 0.18 to 0.57 | 0.000 |
| | | 12090 | Residential setting | Large city | Rural | 0.19 | 0.14 | -0.08 to 0.46 | 0.173 |
| | | 12090 | Residential setting | Suburb | Rural | 0.06 | 0.28 | -0.50 to 0.61 | 0.838 |
| **Have a long list of gratitude** | **Linear regression** | 11987 | *Intercept* | | | 7.47 | 0.15 | 7.17 to 7.77 | 0.000 |
| | | 11987 | Childhood predictor | Somewhat good | Very good | -0.12 | 0.14 | -0.39 to 0.16 | 0.396 |
| | | 11987 | Childhood predictor | Somewhat bad or very bad | Very good | -0.11 | 0.32 | -0.73 to 0.51 | 0.727 |
| | | 11987 | Age | Per 1-year increase | | -0.01 | 0.00 | -0.01 to 0.00 | 0.034 |
| | | 11987 | Gender | Female | Male | 0.19 | 0.07 | 0.05 to 0.33 | 0.008 |
| | | 11987 | Education | Secondary | Elementary or less | 0.29 | 0.08 | 0.13 to 0.45 | 0.000 |
| | | 11987 | Education | Four years beyond HS | Elementary or less | 0.38 | 0.12 | 0.15 to 0.60 | 0.001 |
| | | 11987 | Residential setting | Small town | Rural | 0.43 | 0.09 | 0.25 to 0.62 | 0.000 |
| | | 11987 | Residential setting | Large city | Rural | 0.45 | 0.14 | 0.18 to 0.72 | 0.001 |

| Outcome | Model type | Analytic N | Variable | Level / comparison | Reference category | Coefficient | Standard error | 95% CI | p-value |
|---|---|---|---|---|---|---|---|---|---|
| | | 11987 | Residential setting | Suburb | Rural | 0.50 | 0.17 | 0.16 to 0.83 | 0.004 |
| **Happiness‡** | **Linear regression** | 12146 | *Intercept* | | | 6.67 | 0.16 | 6.36 to 6.98 | 0.000 |
| | | 12146 | Childhood predictor | Somewhat good | Very good | -0.36 | 0.14 | -0.64 to -0.08 | 0.013 |
| | | 12146 | Childhood predictor | Somewhat bad or very bad | Very good | -0.30 | 0.31 | -0.91 to 0.31 | 0.340 |
| | | 12146 | Age | Per 1-year increase | | -0.02 | 0.00 | -0.02 to -0.01 | 0.000 |
| | | 12146 | Gender | Female | Male | 0.25 | 0.08 | 0.10 to 0.40 | 0.001 |
| | | 12146 | Education | Secondary | Elementary or less | 0.45 | 0.09 | 0.27 to 0.62 | 0.000 |
| | | 12146 | Education | Four years beyond HS | Elementary or less | 0.67 | 0.12 | 0.43 to 0.91 | 0.000 |
| | | 12146 | Residential setting | Small town | Rural | 0.39 | 0.11 | 0.18 to 0.60 | 0.000 |
| | | 12146 | Residential setting | Large city | Rural | 0.36 | 0.13 | 0.11 to 0.60 | 0.005 |
| | | 12146 | Residential setting | Suburb | Rural | -0.05 | 0.22 | -0.49 to 0.38 | 0.812 |
| **Remain hopeful for the future** | **Linear regression** | 12094 | *Intercept* | | | 8.14 | 0.13 | 7.89 to 8.39 | 0.000 |
| | | 12094 | Childhood predictor | Somewhat good | Very good | 0.07 | 0.12 | -0.16 to 0.30 | 0.537 |
| | | 12094 | Childhood predictor | Somewhat bad or very bad | Very good | 0.10 | 0.24 | -0.36 to 0.56 | 0.675 |
| | | 12094 | Age | Per 1-year increase | | -0.01 | 0.00 | -0.01 to 0.00 | 0.002 |
| | | 12094 | Gender | Female | Male | 0.01 | 0.06 | -0.10 to 0.13 | 0.804 |
| | | 12094 | Education | Secondary | Elementary or less | 0.40 | 0.06 | 0.27 to 0.53 | 0.000 |
| | | 12094 | Education | Four years beyond HS | Elementary or less | 0.63 | 0.10 | 0.44 to 0.81 | 0.000 |
| | | 12094 | Residential setting | Small town | Rural | 0.48 | 0.08 | 0.32 to 0.64 | 0.000 |
| | | 12094 | Residential setting | Large city | Rural | 0.48 | 0.11 | 0.27 to 0.69 | 0.000 |
| | | 12094 | Residential setting | Suburb | Rural | 0.43 | 0.18 | 0.08 to 0.78 | 0.017 |
| **Understand purpose in life‡** | **Linear regression** | 12073 | *Intercept* | | | 8.06 | 0.14 | 7.79 to 8.33 | 0.000 |
| | | 12073 | Childhood predictor | Somewhat good | Very good | -0.11 | 0.14 | -0.38 to 0.16 | 0.412 |
| | | 12073 | Childhood predictor | Somewhat bad or very bad | Very good | -0.28 | 0.27 | -0.81 to 0.26 | 0.308 |
| | | 12073 | Age | Per 1-year increase | | -0.01 | 0.00 | -0.02 to -0.01 | 0.000 |
| | | 12073 | Gender | Female | Male | 0.12 | 0.07 | -0.02 to 0.27 | 0.090 |
| | | 12073 | Education | Secondary | Elementary or less | 0.43 | 0.08 | 0.27 to 0.58 | 0.000 |
| | | 12073 | Education | Four years beyond HS | Elementary or less | 0.75 | 0.10 | 0.55 to 0.94 | 0.000 |
| | | 12073 | Residential setting | Small town | Rural | 0.42 | 0.09 | 0.25 to 0.60 | 0.000 |
| | | 12073 | Residential setting | Large city | Rural | 0.43 | 0.12 | 0.21 to 0.66 | 0.000 |
| | | 12073 | Residential setting | Suburb | Rural | 0.46 | 0.15 | 0.16 to 0.76 | 0.003 |
| **Life satisfaction‡** | **Linear regression** | 12125 | *Intercept* | | | 7.13 | 0.15 | 6.84 to 7.42 | 0.000 |
| | | 12125 | Childhood predictor | Somewhat good | Very good | -0.38 | 0.16 | -0.69 to -0.08 | 0.014 |
| | | 12125 | Childhood predictor | Somewhat bad or very bad | Very good | -0.48 | 0.29 | -1.05 to 0.08 | 0.093 |
| | | 12125 | Age | Per 1-year increase | | -0.01 | 0.00 | -0.02 to -0.01 | 0.000 |
| | | 12125 | Gender | Female | Male | 0.29 | 0.08 | 0.14 to 0.44 | 0.000 |
| | | 12125 | Education | Secondary | Elementary or less | 0.32 | 0.09 | 0.14 to 0.50 | 0.000 |
| | | 12125 | Education | Four years beyond HS | Elementary or less | 0.48 | 0.12 | 0.24 to 0.71 | 0.000 |
| | | 12125 | Residential setting | Small town | Rural | 0.28 | 0.09 | 0.10 to 0.46 | 0.003 |
| | | 12125 | Residential setting | Large city | Rural | 0.22 | 0.13 | -0.02 to 0.47 | 0.076 |

| Outcome | Model type | Analytic N | Variable | Level / comparison | Reference category | Coefficient | Standard error | 95% CI | p-value |
|---|---|---|---|---|---|---|---|---|---|
| | | 12125 | Residential setting | Suburb | Rural | -0.22 | 0.21 | -0.63 to 0.20 | 0.308 |
| **Loneliness§** | **Linear regression** | 12145 | *Intercept* | | | 4.19 | 0.17 | 3.85 to 4.52 | 0.000 |
| | | 12145 | Childhood predictor | Somewhat good | Very good | 0.44 | 0.17 | 0.12 to 0.76 | 0.008 |
| | | 12145 | Childhood predictor | Somewhat bad or very bad | Very good | 0.66 | 0.35 | -0.03 to 1.35 | 0.063 |
| | | 12145 | Age | Per 1-year increase | | 0.00 | 0.00 | -0.01 to 0.01 | 0.765 |
| | | 12145 | Gender | Female | Male | -0.15 | 0.09 | -0.33 to 0.02 | 0.089 |
| | | 12145 | Education | Secondary | Elementary or less | -0.30 | 0.10 | -0.50 to -0.11 | 0.002 |
| | | 12145 | Education | Four years beyond HS | Elementary or less | -0.64 | 0.15 | -0.93 to -0.35 | 0.000 |
| | | 12145 | Residential setting | Small town | Rural | -0.38 | 0.10 | -0.58 to -0.18 | 0.000 |
| | | 12145 | Residential setting | Large city | Rural | -0.50 | 0.14 | -0.77 to -0.24 | 0.000 |
| | | 12145 | Residential setting | Suburb | Rural | -0.14 | 0.24 | -0.62 to 0.33 | 0.561 |
| **Mental health‡** | **Linear regression** | 12150 | *Intercept* | | | 8.13 | 0.14 | 7.85 to 8.40 | 0.000 |
| | | 12150 | Childhood predictor | Somewhat good | Very good | -0.61 | 0.14 | -0.88 to -0.34 | 0.000 |
| | | 12150 | Childhood predictor | Somewhat bad or very bad | Very good | -0.70 | 0.40 | -1.49 to 0.09 | 0.081 |
| | | 12150 | Age | Per 1-year increase | | -0.02 | 0.00 | -0.03 to -0.02 | 0.000 |
| | | 12150 | Gender | Female | Male | -0.14 | 0.07 | -0.27 to 0.00 | 0.051 |
| | | 12150 | Education | Secondary | Elementary or less | 0.42 | 0.08 | 0.26 to 0.57 | 0.000 |
| | | 12150 | Education | Four years beyond HS | Elementary or less | 0.53 | 0.11 | 0.30 to 0.75 | 0.000 |
| | | 12150 | Residential setting | Small town | Rural | 0.38 | 0.09 | 0.20 to 0.56 | 0.000 |
| | | 12150 | Residential setting | Large city | Rural | 0.45 | 0.12 | 0.22 to 0.69 | 0.000 |
| | | 12150 | Residential setting | Suburb | Rural | 0.15 | 0.18 | -0.20 to 0.51 | 0.399 |
| **Perceived support from others** | **Linear regression** | 12136 | *Intercept* | | | 6.75 | 0.16 | 6.44 to 7.07 | 0.000 |
| | | 12136 | Childhood predictor | Somewhat good | Very good | -0.62 | 0.16 | -0.94 to -0.30 | 0.000 |
| | | 12136 | Childhood predictor | Somewhat bad or very bad | Very good | -1.39 | 0.38 | -2.13 to -0.64 | 0.000 |
| | | 12136 | Age | Per 1-year increase | | -0.01 | 0.00 | -0.02 to -0.01 | 0.000 |
| | | 12136 | Gender | Female | Male | 0.51 | 0.09 | 0.33 to 0.70 | 0.000 |
| | | 12136 | Education | Secondary | Elementary or less | 0.17 | 0.10 | -0.02 to 0.36 | 0.083 |
| | | 12136 | Education | Four years beyond HS | Elementary or less | 0.07 | 0.14 | -0.21 to 0.34 | 0.631 |
| | | 12136 | Residential setting | Small town | Rural | 0.17 | 0.11 | -0.04 to 0.38 | 0.120 |
| | | 12136 | Residential setting | Large city | Rural | -0.43 | 0.14 | -0.71 to -0.15 | 0.003 |
| | | 12136 | Residential setting | Suburb | Rural | -0.31 | 0.21 | -0.72 to 0.10 | 0.135 |
| **Physical health‡** | **Linear regression** | 12153 | *Intercept* | | | 8.44 | 0.14 | 8.16 to 8.72 | 0.000 |
| | | 12153 | Childhood predictor | Somewhat good | Very good | -0.50 | 0.14 | -0.79 to -0.22 | 0.000 |
| | | 12153 | Childhood predictor | Somewhat bad or very bad | Very good | -0.51 | 0.33 | -1.17 to 0.14 | 0.122 |
| | | 12153 | Age | Per 1-year increase | | -0.04 | 0.00 | -0.05 to -0.04 | 0.000 |
| | | 12153 | Gender | Female | Male | -0.36 | 0.08 | -0.51 to -0.21 | 0.000 |
| | | 12153 | Education | Secondary | Elementary or less | 0.38 | 0.08 | 0.22 to 0.53 | 0.000 |
| | | 12153 | Education | Four years beyond HS | Elementary or less | 0.66 | 0.11 | 0.45 to 0.87 | 0.000 |
| | | 12153 | Residential setting | Small town | Rural | 0.48 | 0.09 | 0.30 to 0.67 | 0.000 |
| | | 12153 | Residential setting | Large city | Rural | 0.66 | 0.11 | 0.44 to 0.87 | 0.000 |

| Outcome | Model type | Analytic N | Variable | Level / comparison | Reference category | Coefficient | Standard error | 95% CI | p-value |
|---|---|---|---|---|---|---|---|---|---|
| | | 12153 | Residential setting | Suburb | Rural | 0.20 | 0.18 | -0.15 to 0.54 | 0.267 |
| **Act to promote good‡** | **Linear regression** | 12110 | *Intercept* | | | 8.07 | 0.11 | 7.86 to 8.28 | 0.000 |
| | | 12110 | Childhood predictor | Somewhat good | Very good | -0.07 | 0.11 | -0.30 to 0.15 | 0.535 |
| | | 12110 | Childhood predictor | Somewhat bad or very bad | Very good | -0.07 | 0.25 | -0.56 to 0.41 | 0.766 |
| | | 12110 | Age | Per 1-year increase | | -0.01 | 0.00 | -0.01 to 0.00 | 0.001 |
| | | 12110 | Gender | Female | Male | 0.12 | 0.06 | 0.00 to 0.24 | 0.050 |
| | | 12110 | Education | Secondary | Elementary or less | 0.38 | 0.06 | 0.26 to 0.51 | 0.000 |
| | | 12110 | Education | Four years beyond HS | Elementary or less | 0.57 | 0.08 | 0.41 to 0.72 | 0.000 |
| | | 12110 | Residential setting | Small town | Rural | 0.59 | 0.07 | 0.45 to 0.74 | 0.000 |
| | | 12110 | Residential setting | Large city | Rural | 0.64 | 0.10 | 0.45 to 0.83 | 0.000 |
| | | 12110 | Residential setting | Suburb | Rural | 0.42 | 0.17 | 0.09 to 0.75 | 0.012 |
| **Relationship satisfaction‡** | **Linear regression** | 12115 | *Intercept* | | | 7.87 | 0.13 | 7.62 to 8.13 | 0.000 |
| | | 12115 | Childhood predictor | Somewhat good | Very good | -0.42 | 0.14 | -0.69 to -0.15 | 0.002 |
| | | 12115 | Childhood predictor | Somewhat bad or very bad | Very good | -0.83 | 0.35 | -1.51 to -0.16 | 0.016 |
| | | 12115 | Age | Per 1-year increase | | -0.01 | 0.00 | -0.01 to 0.00 | 0.002 |
| | | 12115 | Gender | Female | Male | 0.34 | 0.07 | 0.20 to 0.48 | 0.000 |
| | | 12115 | Education | Secondary | Elementary or less | -0.02 | 0.08 | -0.18 to 0.14 | 0.832 |
| | | 12115 | Education | Four years beyond HS | Elementary or less | 0.05 | 0.11 | -0.17 to 0.27 | 0.671 |
| | | 12115 | Residential setting | Small town | Rural | 0.45 | 0.09 | 0.28 to 0.62 | 0.000 |
| | | 12115 | Residential setting | Large city | Rural | 0.33 | 0.11 | 0.12 to 0.54 | 0.002 |
| | | 12115 | Residential setting | Suburb | Rural | 0.04 | 0.16 | -0.28 to 0.35 | 0.817 |
| **Show love often** | **Linear regression** | 12136 | *Intercept* | | | 8.30 | 0.13 | 8.06 to 8.55 | 0.000 |
| | | 12136 | Childhood predictor | Somewhat good | Very good | -0.12 | 0.12 | -0.36 to 0.12 | 0.318 |
| | | 12136 | Childhood predictor | Somewhat bad or very bad | Very good | 0.04 | 0.23 | -0.42 to 0.50 | 0.863 |
| | | 12136 | Age | Per 1-year increase | | -0.01 | 0.00 | -0.01 to 0.00 | 0.002 |
| | | 12136 | Gender | Female | Male | 0.19 | 0.07 | 0.06 to 0.32 | 0.005 |
| | | 12136 | Education | Secondary | Elementary or less | 0.16 | 0.07 | 0.02 to 0.29 | 0.028 |
| | | 12136 | Education | Four years beyond HS | Elementary or less | 0.02 | 0.11 | -0.20 to 0.24 | 0.830 |
| | | 12136 | Residential setting | Small town | Rural | 0.19 | 0.08 | 0.04 to 0.35 | 0.015 |
| | | 12136 | Residential setting | Large city | Rural | 0.04 | 0.12 | -0.19 to 0.27 | 0.752 |
| | | 12136 | Residential setting | Suburb | Rural | -0.09 | 0.20 | -0.49 to 0.30 | 0.637 |
| **Life evaluation five years from now** | **Linear regression** | 11060 | *Intercept* | | | 7.72 | 0.14 | 7.44 to 8.00 | 0.000 |
| | | 11060 | Childhood predictor | Somewhat good | Very good | -0.17 | 0.14 | -0.44 to 0.09 | 0.208 |
| | | 11060 | Childhood predictor | Somewhat bad or very bad | Very good | -0.04 | 0.28 | -0.59 to 0.52 | 0.896 |
| | | 11060 | Age | Per 1-year increase | | -0.02 | 0.00 | -0.03 to -0.02 | 0.000 |
| | | 11060 | Gender | Female | Male | 0.17 | 0.07 | 0.03 to 0.31 | 0.021 |
| | | 11060 | Education | Secondary | Elementary or less | 0.27 | 0.08 | 0.11 to 0.43 | 0.001 |
| | | 11060 | Education | Four years beyond HS | Elementary or less | 0.57 | 0.12 | 0.34 to 0.80 | 0.000 |
| | | 11060 | Residential setting | Small town | Rural | 0.43 | 0.09 | 0.26 to 0.61 | 0.000 |
| | | 11060 | Residential setting | Large city | Rural | 0.51 | 0.11 | 0.29 to 0.72 | 0.000 |

| Outcome | Model type | Analytic N | Variable | Level / comparison | Reference category | Coefficient | Standard error | 95% CI | p-value |
|---|---|---|---|---|---|---|---|---|---|
| | | 11060 | Residential setting | Suburb | Rural | 0.38 | 0.22 | -0.05 to 0.82 | 0.085 |
| **Life evaluation today** | **Linear regression** | 12047 | *Intercept* | | | 5.61 | 0.17 | 5.28 to 5.94 | 0.000 |
| | | 12047 | Childhood predictor | Somewhat good | Very good | -0.42 | 0.16 | -0.72 to -0.11 | 0.009 |
| | | 12047 | Childhood predictor | Somewhat bad or very bad | Very good | -0.37 | 0.37 | -1.09 to 0.35 | 0.313 |
| | | 12047 | Age | Per 1-year increase | | -0.01 | 0.00 | -0.02 to 0.00 | 0.006 |
| | | 12047 | Gender | Female | Male | 0.48 | 0.08 | 0.32 to 0.65 | 0.000 |
| | | 12047 | Education | Secondary | Elementary or less | 0.24 | 0.09 | 0.07 to 0.41 | 0.006 |
| | | 12047 | Education | Four years beyond HS | Elementary or less | 0.15 | 0.14 | -0.13 to 0.42 | 0.304 |
| | | 12047 | Residential setting | Small town | Rural | 0.24 | 0.10 | 0.04 to 0.44 | 0.022 |
| | | 12047 | Residential setting | Large city | Rural | 0.10 | 0.13 | -0.16 to 0.36 | 0.438 |
| | | 12047 | Residential setting | Suburb | Rural | -0.10 | 0.22 | -0.52 to 0.32 | 0.641 |
| **Worry about safety, food, or housing§§‡‡** | **Linear regression** | 12161 | *Intercept* | | | 6.19 | 0.19 | 5.82 to 6.56 | 0.000 |
| | | 12161 | Childhood predictor | Somewhat good | Very good | 0.11 | 0.16 | -0.21 to 0.43 | 0.489 |
| | | 12161 | Childhood predictor | Somewhat bad or very bad | Very good | -0.13 | 0.41 | -0.94 to 0.68 | 0.756 |
| | | 12161 | Age | Per 1-year increase | | -0.01 | 0.00 | -0.01 to 0.00 | 0.026 |
| | | 12161 | Gender | Female | Male | 0.33 | 0.09 | 0.15 to 0.50 | 0.000 |
| | | 12161 | Education | Secondary | Elementary or less | -0.56 | 0.10 | -0.76 to -0.35 | 0.000 |
| | | 12161 | Education | Four years beyond HS | Elementary or less | -0.81 | 0.17 | -1.13 to -0.48 | 0.000 |
| | | 12161 | Residential setting | Small town | Rural | 0.38 | 0.13 | 0.12 to 0.64 | 0.004 |
| | | 12161 | Residential setting | Large city | Rural | -0.03 | 0.18 | -0.38 to 0.32 | 0.863 |
| | | 12161 | Residential setting | Suburb | Rural | 0.20 | 0.26 | -0.30 to 0.71 | 0.431 |
| **Life worthwhile‡** | **Linear regression** | 12048 | *Intercept* | | | 6.67 | 0.15 | 6.38 to 6.96 | 0.000 |
| | | 12048 | Childhood predictor | Somewhat good | Very good | -0.36 | 0.14 | -0.64 to -0.08 | 0.013 |
| | | 12048 | Childhood predictor | Somewhat bad or very bad | Very good | -0.39 | 0.34 | -1.07 to 0.28 | 0.253 |
| | | 12048 | Age | Per 1-year increase | | -0.01 | 0.00 | -0.01 to 0.00 | 0.002 |
| | | 12048 | Gender | Female | Male | 0.45 | 0.08 | 0.30 to 0.60 | 0.000 |
| | | 12048 | Education | Secondary | Elementary or less | 0.47 | 0.08 | 0.31 to 0.63 | 0.000 |
| | | 12048 | Education | Four years beyond HS | Elementary or less | 0.61 | 0.13 | 0.36 to 0.85 | 0.000 |
| | | 12048 | Residential setting | Small town | Rural | 0.44 | 0.10 | 0.24 to 0.65 | 0.000 |
| | | 12048 | Residential setting | Large city | Rural | 0.44 | 0.13 | 0.19 to 0.70 | 0.001 |
| | | 12048 | Residential setting | Suburb | Rural | 0.39 | 0.21 | -0.01 to 0.80 | 0.059 |
| **Flourishing (with financial indicators)** | **Linear regression** | 11711 | *Intercept* | | | 7.02 | 0.08 | 6.86 to 7.19 | 0.000 |
| | | 11711 | Childhood predictor | Somewhat good | Very good | -0.27 | 0.08 | -0.42 to -0.12 | 0.001 |
| | | 11711 | Childhood predictor | Somewhat bad or very bad | Very good | -0.37 | 0.16 | -0.68 to -0.05 | 0.024 |
| | | 11711 | Age | Per 1-year increase | | -0.01 | 0.00 | -0.02 to -0.01 | 0.000 |
| | | 11711 | Gender | Female | Male | 0.09 | 0.04 | 0.01 to 0.18 | 0.029 |
| | | 11711 | Education | Secondary | Elementary or less | 0.34 | 0.05 | 0.24 to 0.43 | 0.000 |
| | | 11711 | Education | Four years beyond HS | Elementary or less | 0.51 | 0.07 | 0.38 to 0.64 | 0.000 |
| | | 11711 | Residential setting | Small town | Rural | 0.31 | 0.06 | 0.20 to 0.42 | 0.000 |

| Outcome | Model type | Analytic N | Variable | Level / comparison | Reference category | Coefficient | Standard error | 95% CI | p-value |
|---|---|---|---|---|---|---|---|---|---|
| | | 11711 | Residential setting | Large city | Rural | 0.35 | 0.07 | 0.21 to 0.48 | 0.000 |
| | | 11711 | Residential setting | Suburb | Rural | 0.11 | 0.13 | -0.15 to 0.37 | 0.399 |
| **Flourishing (without financial indicators)** | **Linear regression** | 11738 | *Intercept* | | | 7.64 | 0.09 | 7.46 to 7.81 | 0.000 |
| | | 11738 | Childhood predictor | Somewhat good | Very good | -0.31 | 0.09 | -0.49 to -0.13 | 0.001 |
| | | 11738 | Childhood predictor | Somewhat bad or very bad | Very good | -0.47 | 0.18 | -0.82 to -0.11 | 0.010 |
| | | 11738 | Age | Per 1-year increase | | -0.02 | 0.00 | -0.02 to -0.01 | 0.000 |
| | | 11738 | Gender | Female | Male | 0.18 | 0.05 | 0.09 to 0.27 | 0.000 |
| | | 11738 | Education | Secondary | Elementary or less | 0.31 | 0.05 | 0.21 to 0.41 | 0.000 |
| | | 11738 | Education | Four years beyond HS | Elementary or less | 0.47 | 0.07 | 0.33 to 0.61 | 0.000 |
| | | 11738 | Residential setting | Small town | Rural | 0.43 | 0.06 | 0.31 to 0.56 | 0.000 |
| | | 11738 | Residential setting | Large city | Rural | 0.40 | 0.08 | 0.24 to 0.55 | 0.000 |
| | | 11738 | Residential setting | Suburb | Rural | 0.13 | 0.13 | -0.13 to 0.39 | 0.328 |
| **Feel close to someone** | **Logistic regression** | 12158 | *Intercept* | | | 1.72 | 0.14 | 1.45 to 2.00 | 0.000 |
| | | 12158 | Childhood predictor | Somewhat good | Very good | -0.24 | 0.13 | -0.50 to 0.01 | 0.061 |
| | | 12158 | Childhood predictor | Somewhat bad or very bad | Very good | 0.00 | 0.25 | -0.48 to 0.49 | 0.989 |
| | | 12158 | Age | Per 1-year increase | | -0.01 | 0.00 | -0.01 to 0.00 | 0.009 |
| | | 12158 | Gender | Female | Male | 0.00 | 0.07 | -0.13 to 0.13 | 0.969 |
| | | 12158 | Education | Secondary | Elementary or less | 0.17 | 0.08 | 0.01 to 0.33 | 0.044 |
| | | 12158 | Education | Four years beyond HS | Elementary or less | 0.13 | 0.13 | -0.13 to 0.38 | 0.326 |
| | | 12158 | Residential setting | Small town | Rural | 0.07 | 0.11 | -0.14 to 0.29 | 0.496 |
| | | 12158 | Residential setting | Large city | Rural | -0.10 | 0.13 | -0.36 to 0.15 | 0.428 |
| | | 12158 | Residential setting | Suburb | Rural | -0.16 | 0.19 | -0.54 to 0.21 | 0.387 |
| **Donated to charity** | **Logistic regression** | 12128 | *Intercept* | | | -0.33 | 0.13 | -0.58 to -0.07 | 0.011 |
| | | 12128 | Childhood predictor | Somewhat good | Very good | -0.22 | 0.12 | -0.46 to 0.02 | 0.070 |
| | | 12128 | Childhood predictor | Somewhat bad or very bad | Very good | 0.06 | 0.20 | -0.33 to 0.46 | 0.754 |
| | | 12128 | Age | Per 1-year increase | | 0.00 | 0.00 | 0.00 to 0.01 | 0.463 |
| | | 12128 | Gender | Female | Male | -0.03 | 0.05 | -0.13 to 0.07 | 0.515 |
| | | 12128 | Education | Secondary | Elementary or less | -0.09 | 0.06 | -0.21 to 0.03 | 0.130 |
| | | 12128 | Education | Four years beyond HS | Elementary or less | -0.15 | 0.10 | -0.35 to 0.04 | 0.130 |
| | | 12128 | Residential setting | Small town | Rural | -0.05 | 0.09 | -0.24 to 0.13 | 0.557 |
| | | 12128 | Residential setting | Large city | Rural | -0.40 | 0.11 | -0.62 to -0.17 | 0.001 |
| | | 12128 | Residential setting | Suburb | Rural | -0.17 | 0.19 | -0.54 to 0.20 | 0.366 |
| **Have any health problems** | **Logistic regression** | 12081 | *Intercept* | | | -2.04 | 0.10 | -2.24 to -1.83 | 0.000 |
| | | 12081 | Childhood predictor | Somewhat good | Very good | 0.08 | 0.09 | -0.09 to 0.26 | 0.351 |
| | | 12081 | Childhood predictor | Somewhat bad or very bad | Very good | 0.29 | 0.19 | -0.08 to 0.65 | 0.129 |
| | | 12081 | Age | Per 1-year increase | | 0.03 | 0.00 | 0.02 to 0.03 | 0.000 |
| | | 12081 | Gender | Female | Male | 0.21 | 0.05 | 0.11 to 0.31 | 0.000 |
| | | 12081 | Education | Secondary | Elementary or less | -0.14 | 0.06 | -0.25 to -0.02 | 0.023 |
| | | 12081 | Education | Four years beyond HS | Elementary or less | -0.27 | 0.10 | -0.46 to -0.08 | 0.006 |
| | | 12081 | Residential setting | Small town | Rural | 0.13 | 0.07 | 0.00 to 0.26 | 0.057 |

| Outcome | Model type | Analytic N | Variable | Level / comparison | Reference category | Coefficient | Standard error | 95% CI | p-value |
|---|---|---|---|---|---|---|---|---|---|
| | | 12081 | Residential setting | Large city | Rural | -0.13 | 0.09 | -0.31 to 0.04 | 0.137 |
| | | 12081 | Residential setting | Suburb | Rural | 0.09 | 0.14 | -0.19 to 0.37 | 0.535 |
| **Helped a stranger** | **Logistic regression** | 12040 | *Intercept* | | | 0.71 | 0.12 | 0.48 to 0.94 | 0.000 |
| | | 12040 | Childhood predictor | Somewhat good | Very good | -0.16 | 0.11 | -0.37 to 0.05 | 0.133 |
| | | 12040 | Childhood predictor | Somewhat bad or very bad | Very good | 0.27 | 0.20 | -0.12 to 0.66 | 0.170 |
| | | 12040 | Age | Per 1-year increase | | -0.01 | 0.00 | -0.01 to 0.00 | 0.000 |
| | | 12040 | Gender | Female | Male | -0.35 | 0.05 | -0.44 to -0.25 | 0.000 |
| | | 12040 | Education | Secondary | Elementary or less | -0.09 | 0.06 | -0.21 to 0.02 | 0.105 |
| | | 12040 | Education | Four years beyond HS | Elementary or less | -0.10 | 0.09 | -0.27 to 0.08 | 0.289 |
| | | 12040 | Residential setting | Small town | Rural | 0.11 | 0.09 | -0.06 to 0.28 | 0.224 |
| | | 12040 | Residential setting | Large city | Rural | -0.15 | 0.10 | -0.35 to 0.05 | 0.147 |
| | | 12040 | Residential setting | Suburb | Rural | 0.03 | 0.18 | -0.31 to 0.38 | 0.845 |
| **Volunteered at an organization** | **Logistic regression** | 12111 | *Intercept* | | | -0.24 | 0.14 | -0.51 to 0.03 | 0.084 |
| | | 12111 | Childhood predictor | Somewhat good | Very good | -0.50 | 0.14 | -0.78 to -0.23 | 0.000 |
| | | 12111 | Childhood predictor | Somewhat bad or very bad | Very good | -0.48 | 0.22 | -0.91 to -0.06 | 0.027 |
| | | 12111 | Age | Per 1-year increase | | -0.01 | 0.00 | -0.01 to 0.00 | 0.002 |
| | | 12111 | Gender | Female | Male | -0.29 | 0.05 | -0.40 to -0.18 | 0.000 |
| | | 12111 | Education | Secondary | Elementary or less | -0.16 | 0.07 | -0.29 to -0.03 | 0.018 |
| | | 12111 | Education | Four years beyond HS | Elementary or less | 0.01 | 0.10 | -0.19 to 0.21 | 0.915 |
| | | 12111 | Residential setting | Small town | Rural | -0.11 | 0.11 | -0.32 to 0.10 | 0.313 |
| | | 12111 | Residential setting | Large city | Rural | -0.56 | 0.13 | -0.81 to -0.30 | 0.000 |
| | | 12111 | Residential setting | Suburb | Rural | -0.37 | 0.16 | -0.69 to -0.05 | 0.023 |
| **Cigarette smoking** | **Logistic regression** | 11690 | *Intercept* | | | -2.03 | 0.17 | -2.35 to -1.70 | 0.000 |
| | | 11690 | Childhood predictor | Somewhat good | Very good | 0.27 | 0.14 | 0.00 to 0.54 | 0.050 |
| | | 11690 | Childhood predictor | Somewhat bad or very bad | Very good | 0.63 | 0.31 | 0.03 to 1.23 | 0.041 |
| | | 11690 | Age | Per 1-year increase | | 0.01 | 0.00 | 0.00 to 0.01 | 0.026 |
| | | 11690 | Gender | Female | Male | -2.83 | 0.19 | -3.20 to -2.46 | 0.000 |
| | | 11690 | Education | Secondary | Elementary or less | -0.40 | 0.10 | -0.60 to -0.21 | 0.000 |
| | | 11690 | Education | Four years beyond HS | Elementary or less | -0.56 | 0.17 | -0.89 to -0.22 | 0.001 |
| | | 11690 | Residential setting | Small town | Rural | -0.04 | 0.11 | -0.25 to 0.17 | 0.696 |
| | | 11690 | Residential setting | Large city | Rural | 0.14 | 0.14 | -0.14 to 0.41 | 0.331 |
| | | 11690 | Residential setting | Suburb | Rural | 0.86 | 0.20 | 0.47 to 1.26 | 0.000 |
| **Alcohol use** | **Logistic regression** | 11677 | *Intercept* | | | -1.90 | 0.15 | -2.19 to -1.60 | 0.000 |
| | | 11677 | Childhood predictor | Somewhat good | Very good | 0.02 | 0.15 | -0.28 to 0.33 | 0.878 |
| | | 11677 | Childhood predictor | Somewhat bad or very bad | Very good | 0.58 | 0.30 | 0.00 to 1.16 | 0.051 |
| | | 11677 | Age | Per 1-year increase | | 0.00 | 0.00 | -0.01 to 0.01 | 0.864 |
| | | 11677 | Gender | Female | Male | -2.78 | 0.18 | -3.14 to -2.42 | 0.000 |
| | | 11677 | Education | Secondary | Elementary or less | -0.22 | 0.10 | -0.42 to -0.03 | 0.024 |
| | | 11677 | Education | Four years beyond HS | Elementary or less | -0.47 | 0.16 | -0.79 to -0.15 | 0.005 |
| | | 11677 | Residential setting | Small town | Rural | 0.21 | 0.11 | 0.00 to 0.42 | 0.049 |

| Outcome | Model type | Analytic N | Variable | Level / comparison | Reference category | Coefficient | Standard error | 95% CI | p-value |
|---|---|---|---|---|---|---|---|---|---|
| | | 11677 | Residential setting | Large city | Rural | 0.38 | 0.14 | 0.10 to 0.67 | 0.008 |
| | | 11677 | Residential setting | Suburb | Rural | 0.62 | 0.22 | 0.19 to 1.06 | 0.005 |
| **Government approval§** | **Proportional-odds regression** | 11950 | Childhood predictor | Somewhat good | Very good | -0.10 | 0.08 | -0.26 to 0.07 | 0.252 |
| | | 11950 | Childhood predictor | Somewhat bad or very bad | Very good | -0.49 | 0.17 | -0.81 to -0.16 | 0.003 |
| | | 11950 | Age | Per 1-year increase | | 0.00 | 0.00 | -0.01 to 0.00 | 0.075 |
| | | 11950 | Gender | Female | Male | 0.05 | 0.04 | -0.03 to 0.14 | 0.235 |
| | | 11950 | Education | Secondary | Elementary or less | -0.25 | 0.05 | -0.35 to -0.14 | 0.000 |
| | | 11950 | Education | Four years beyond HS | Elementary or less | -0.32 | 0.09 | -0.48 to -0.15 | 0.000 |
| | | 11950 | Residential setting | Small town | Rural | 0.09 | 0.07 | -0.04 to 0.22 | 0.195 |
| | | 11950 | Residential setting | Large city | Rural | -0.09 | 0.08 | -0.24 to 0.06 | 0.242 |
| | | 11950 | Residential setting | Suburb | Rural | -0.43 | 0.16 | -0.75 to -0.12 | 0.007 |
| | | 11950 | *Threshold / cut-point* | Strongly disapprove\|Somewhat disapprove | | -2.91 | 0.10 | -3.11 to -2.72 | 0.000 |
| | | 11950 | *Threshold / cut-point* | Somewhat disapprove\|Neither | | -2.26 | 0.10 | -2.45 to -2.07 | 0.000 |
| | | 11950 | *Threshold / cut-point* | Neither\|Somewhat approve | | -1.90 | 0.09 | -2.09 to -1.72 | 0.000 |
| | | 11950 | *Threshold / cut-point* | Somewhat approve\|Strongly approve | | -0.45 | 0.09 | -0.63 to -0.28 | 0.000 |
| **Feeling capable§** | **Proportional-odds regression** | 12104 | Childhood predictor | Somewhat good | Very good | -0.19 | 0.08 | -0.35 to -0.04 | 0.013 |
| | | 12104 | Childhood predictor | Somewhat bad or very bad | Very good | -0.04 | 0.20 | -0.43 to 0.36 | 0.855 |
| | | 12104 | Age | Per 1-year increase | | 0.00 | 0.00 | 0.00 to 0.01 | 0.188 |
| | | 12104 | Gender | Female | Male | -0.07 | 0.04 | -0.16 to 0.01 | 0.096 |
| | | 12104 | Education | Secondary | Elementary or less | 0.15 | 0.05 | 0.06 to 0.25 | 0.002 |
| | | 12104 | Education | Four years beyond HS | Elementary or less | 0.11 | 0.07 | -0.03 to 0.25 | 0.139 |
| | | 12104 | Residential setting | Small town | Rural | 0.16 | 0.05 | 0.06 to 0.26 | 0.001 |
| | | 12104 | Residential setting | Large city | Rural | 0.18 | 0.08 | 0.03 to 0.33 | 0.019 |
| | | 12104 | Residential setting | Suburb | Rural | 0.23 | 0.09 | 0.05 to 0.41 | 0.012 |
| | | 12104 | *Threshold / cut-point* | Never\|Rarely | | -2.36 | 0.09 | -2.54 to -2.18 | 0.000 |
| | | 12104 | *Threshold / cut-point* | Rarely\|Often | | -0.65 | 0.09 | -0.82 to -0.48 | 0.000 |
| | | 12104 | *Threshold / cut-point* | Often\|Always | | 0.02 | 0.08 | -0.15 to 0.18 | 0.842 |
| **Unable to control worry§** | **Proportional-odds regression** | 12056 | Childhood predictor | Somewhat good | Very good | 0.05 | 0.07 | -0.09 to 0.19 | 0.478 |
| | | 12056 | Childhood predictor | Somewhat bad or very bad | Very good | 0.25 | 0.15 | -0.05 to 0.55 | 0.098 |
| | | 12056 | Age | Per 1-year increase | | 0.00 | 0.00 | 0.00 to 0.01 | 0.002 |
| | | 12056 | Gender | Female | Male | 0.17 | 0.04 | 0.09 to 0.25 | 0.000 |
| | | 12056 | Education | Secondary | Elementary or less | -0.28 | 0.05 | -0.37 to -0.19 | 0.000 |
| | | 12056 | Education | Four years beyond HS | Elementary or less | -0.42 | 0.08 | -0.56 to -0.27 | 0.000 |
| | | 12056 | Residential setting | Small town | Rural | 0.03 | 0.05 | -0.06 to 0.13 | 0.494 |
| | | 12056 | Residential setting | Large city | Rural | -0.01 | 0.07 | -0.15 to 0.13 | 0.901 |
| | | 12056 | Residential setting | Suburb | Rural | -0.08 | 0.10 | -0.28 to 0.11 | 0.406 |

| Outcome | Model type | Analytic N | Variable | Level / comparison | Reference category | Coefficient | Standard error | 95% CI | p-value |
|---|---|---|---|---|---|---|---|---|---|
| | | 12056 | *Threshold / cut-point* | Not at all\|Several days | | 0.07 | 0.08 | -0.09 to 0.23 | 0.409 |
| | | 12056 | *Threshold / cut-point* | Several days\|More than half the days | | 0.97 | 0.08 | 0.81 to 1.13 | 0.000 |
| | | 12056 | *Threshold / cut-point* | More than half the days\|Nearly every day | | 1.83 | 0.08 | 1.66 to 1.99 | 0.000 |
| **Depressive symptoms§** | **Proportional-odds regression** | 12080 | Childhood predictor | Somewhat good | Very good | 0.03 | 0.07 | -0.12 to 0.17 | 0.721 |
| | | 12080 | Childhood predictor | Somewhat bad or very bad | Very good | 0.19 | 0.14 | -0.09 to 0.46 | 0.185 |
| | | 12080 | Age | Per 1-year increase | | 0.00 | 0.00 | 0.00 to 0.01 | 0.162 |
| | | 12080 | Gender | Female | Male | 0.08 | 0.04 | 0.00 to 0.15 | 0.053 |
| | | 12080 | Education | Secondary | Elementary or less | -0.21 | 0.05 | -0.31 to -0.12 | 0.000 |
| | | 12080 | Education | Four years beyond HS | Elementary or less | -0.42 | 0.07 | -0.55 to -0.28 | 0.000 |
| | | 12080 | Residential setting | Small town | Rural | 0.00 | 0.05 | -0.10 to 0.11 | 0.957 |
| | | 12080 | Residential setting | Large city | Rural | -0.09 | 0.07 | -0.24 to 0.05 | 0.214 |
| | | 12080 | Residential setting | Suburb | Rural | -0.17 | 0.11 | -0.39 to 0.06 | 0.141 |
| | | 12080 | *Threshold / cut-point* | Not at all\|Several days | | -0.25 | 0.08 | -0.41 to -0.09 | 0.002 |
| | | 12080 | *Threshold / cut-point* | Several days\|More than half the days | | 0.82 | 0.08 | 0.66 to 0.98 | 0.000 |
| | | 12080 | *Threshold / cut-point* | More than half the days\|Nearly every day | | 1.83 | 0.08 | 1.66 to 1.99 | 0.000 |
| **Perceived discrimination§** | **Proportional-odds regression** | 12015 | Childhood predictor | Somewhat good | Very good | 0.00 | 0.08 | -0.15 to 0.16 | 0.961 |
| | | 12015 | Childhood predictor | Somewhat bad or very bad | Very good | 0.19 | 0.17 | -0.14 to 0.52 | 0.256 |
| | | 12015 | Age | Per 1-year increase | | 0.00 | 0.00 | -0.01 to 0.00 | 0.027 |
| | | 12015 | Gender | Female | Male | -0.16 | 0.04 | -0.25 to -0.07 | 0.000 |
| | | 12015 | Education | Secondary | Elementary or less | -0.25 | 0.05 | -0.36 to -0.15 | 0.000 |
| | | 12015 | Education | Four years beyond HS | Elementary or less | -0.31 | 0.09 | -0.48 to -0.13 | 0.001 |
| | | 12015 | Residential setting | Small town | Rural | -0.21 | 0.06 | -0.33 to -0.10 | 0.000 |
| | | 12015 | Residential setting | Large city | Rural | -0.22 | 0.07 | -0.36 to -0.08 | 0.002 |
| | | 12015 | Residential setting | Suburb | Rural | -0.32 | 0.10 | -0.51 to -0.12 | 0.002 |
| | | 12015 | *Threshold / cut-point* | Never\|Rarely | | -0.16 | 0.09 | -0.34 to 0.01 | 0.063 |
| | | 12015 | *Threshold / cut-point* | Rarely\|Often | | 0.84 | 0.09 | 0.66 to 1.01 | 0.000 |
| | | 12015 | *Threshold / cut-point* | Often\|Always | | 1.45 | 0.09 | 1.27 to 1.63 | 0.000 |
| **Feeling nervous or anxious§** | **Proportional-odds regression** | 12106 | Childhood predictor | Somewhat good | Very good | 0.09 | 0.07 | -0.04 to 0.23 | 0.183 |
| | | 12106 | Childhood predictor | Somewhat bad or very bad | Very good | 0.22 | 0.17 | -0.11 to 0.55 | 0.198 |
| | | 12106 | Age | Per 1-year increase | | 0.01 | 0.00 | 0.00 to 0.01 | 0.000 |
| | | 12106 | Gender | Female | Male | 0.29 | 0.04 | 0.21 to 0.37 | 0.000 |
| | | 12106 | Education | Secondary | Elementary or less | -0.28 | 0.05 | -0.37 to -0.19 | 0.000 |
| | | 12106 | Education | Four years beyond HS | Elementary or less | -0.46 | 0.08 | -0.61 to -0.30 | 0.000 |
| | | 12106 | Residential setting | Small town | Rural | 0.11 | 0.05 | 0.00 to 0.22 | 0.044 |
| | | 12106 | Residential setting | Large city | Rural | -0.03 | 0.07 | -0.16 to 0.11 | 0.680 |

| Outcome | Model type | Analytic N | Variable | Level / comparison | Reference category | Coefficient | Standard error | 95% CI | p-value |
|---|---|---|---|---|---|---|---|---|---|
| | | 12106 | Residential setting | Suburb | Rural | -0.03 | 0.12 | -0.27 to 0.20 | 0.775 |
| | | 12106 | *Threshold / cut-point* | Not at all\|Several days | | 0.15 | 0.08 | 0.00 to 0.30 | 0.054 |
| | | 12106 | *Threshold / cut-point* | Several days\|More than half the days | | 1.15 | 0.08 | 0.99 to 1.30 | 0.000 |
| | | 12106 | *Threshold / cut-point* | More than half the days\|Nearly every day | | 2.08 | 0.08 | 1.92 to 2.24 | 0.000 |
| **Forgiveness§** | **Proportional-odds regression** | 12085 | Childhood predictor | Somewhat good | Very good | -0.10 | 0.08 | -0.25 to 0.05 | 0.203 |
| | | 12085 | Childhood predictor | Somewhat bad or very bad | Very good | -0.29 | 0.18 | -0.65 to 0.06 | 0.107 |
| | | 12085 | Age | Per 1-year increase | | 0.00 | 0.00 | 0.00 to 0.00 | 0.875 |
| | | 12085 | Gender | Female | Male | 0.09 | 0.04 | 0.01 to 0.18 | 0.032 |
| | | 12085 | Education | Secondary | Elementary or less | -0.08 | 0.05 | -0.18 to 0.03 | 0.143 |
| | | 12085 | Education | Four years beyond HS | Elementary or less | -0.20 | 0.08 | -0.35 to -0.04 | 0.012 |
| | | 12085 | Residential setting | Small town | Rural | 0.02 | 0.05 | -0.09 to 0.12 | 0.751 |
| | | 12085 | Residential setting | Large city | Rural | 0.02 | 0.08 | -0.14 to 0.18 | 0.815 |
| | | 12085 | Residential setting | Suburb | Rural | 0.04 | 0.12 | -0.19 to 0.28 | 0.719 |
| | | 12085 | *Threshold / cut-point* | Never\|Rarely | | -2.43 | 0.09 | -2.61 to -2.26 | 0.000 |
| | | 12085 | *Threshold / cut-point* | Rarely\|Often | | -1.10 | 0.09 | -1.26 to -0.93 | 0.000 |
| | | 12085 | *Threshold / cut-point* | Often\|Always | | -0.45 | 0.08 | -0.61 to -0.28 | 0.000 |
| **Support reducing differences in income levels§** | **Proportional-odds regression** | 11950 | Childhood predictor | Somewhat good | Very good | -1.06 | 0.10 | -1.26 to -0.86 | 0.000 |
| | | 11950 | Childhood predictor | Somewhat bad or very bad | Very good | -0.48 | 0.22 | -0.92 to -0.05 | 0.030 |
| | | 11950 | Age | Per 1-year increase | | 0.00 | 0.00 | -0.01 to 0.00 | 0.166 |
| | | 11950 | Gender | Female | Male | -0.07 | 0.06 | -0.19 to 0.04 | 0.213 |
| | | 11950 | Education | Secondary | Elementary or less | -0.07 | 0.07 | -0.21 to 0.07 | 0.350 |
| | | 11950 | Education | Four years beyond HS | Elementary or less | -0.30 | 0.11 | -0.52 to -0.08 | 0.007 |
| | | 11950 | Residential setting | Small town | Rural | 0.02 | 0.12 | -0.22 to 0.25 | 0.879 |
| | | 11950 | Residential setting | Large city | Rural | 0.07 | 0.14 | -0.21 to 0.34 | 0.627 |
| | | 11950 | Residential setting | Suburb | Rural | -0.10 | 0.17 | -0.44 to 0.24 | 0.566 |
| | | 11950 | *Threshold / cut-point* | Strongly disagree\|Somewhat disagree | | -4.03 | 0.18 | -4.38 to -3.68 | 0.000 |
| | | 11950 | *Threshold / cut-point* | Somewhat disagree\|Neither | | -3.48 | 0.17 | -3.81 to -3.15 | 0.000 |
| | | 11950 | *Threshold / cut-point* | Neither\|Somewhat agree | | -3.02 | 0.16 | -3.33 to -2.71 | 0.000 |
| | | 11950 | *Threshold / cut-point* | Somewhat agree\|Strongly agree | | -1.43 | 0.14 | -1.69 to -1.16 | 0.000 |
| **Have little interest in doing things§** | **Proportional-odds regression** | 12074 | Childhood predictor | Somewhat good | Very good | 0.08 | 0.09 | -0.09 to 0.25 | 0.345 |
| | | 12074 | Childhood predictor | Somewhat bad or very bad | Very good | 0.49 | 0.15 | 0.20 to 0.78 | 0.001 |
| | | 12074 | Age | Per 1-year increase | | 0.00 | 0.00 | 0.00 to 0.01 | 0.041 |
| | | 12074 | Gender | Female | Male | 0.03 | 0.04 | -0.05 to 0.11 | 0.490 |
| | | 12074 | Education | Secondary | Elementary or less | 0.07 | 0.05 | -0.02 to 0.17 | 0.141 |
| | | 12074 | Education | Four years beyond HS | Elementary or less | 0.11 | 0.08 | -0.04 to 0.27 | 0.155 |

| Outcome | Model type | Analytic N | Variable | Level / comparison | Reference category | Coefficient | Standard error | 95% CI | p-value |
|---|---|---|---|---|---|---|---|---|---|
| | | 12074 | Residential setting | Small town | Rural | 0.26 | 0.06 | 0.14 to 0.39 | 0.000 |
| | | 12074 | Residential setting | Large city | Rural | 0.10 | 0.08 | -0.06 to 0.26 | 0.234 |
| | | 12074 | Residential setting | Suburb | Rural | 0.28 | 0.15 | -0.02 to 0.58 | 0.065 |
| | | 12074 | *Threshold / cut-point* | Not at all\|Several days | | -0.48 | 0.09 | -0.65 to -0.31 | 0.000 |
| | | 12074 | *Threshold / cut-point* | Several days\|More than half the days | | 0.45 | 0.09 | 0.28 to 0.63 | 0.000 |
| | | 12074 | *Threshold / cut-point* | More than half the days\|Nearly every day | | 1.23 | 0.09 | 1.05 to 1.41 | 0.000 |
| **Various aspects of life are balanced§** | **Proportional-odds regression** | 12003 | Childhood predictor | Somewhat good | Very good | 0.00 | 0.08 | -0.15 to 0.15 | 0.985 |
| | | 12003 | Childhood predictor | Somewhat bad or very bad | Very good | 0.09 | 0.20 | -0.30 to 0.49 | 0.641 |
| | | 12003 | Age | Per 1-year increase | | 0.00 | 0.00 | 0.00 to 0.01 | 0.196 |
| | | 12003 | Gender | Female | Male | -0.03 | 0.04 | -0.11 to 0.04 | 0.392 |
| | | 12003 | Education | Secondary | Elementary or less | 0.04 | 0.04 | -0.05 to 0.13 | 0.354 |
| | | 12003 | Education | Four years beyond HS | Elementary or less | 0.12 | 0.07 | -0.02 to 0.25 | 0.082 |
| | | 12003 | Residential setting | Small town | Rural | 0.09 | 0.05 | -0.02 to 0.19 | 0.099 |
| | | 12003 | Residential setting | Large city | Rural | 0.09 | 0.07 | -0.04 to 0.22 | 0.156 |
| | | 12003 | Residential setting | Suburb | Rural | 0.38 | 0.11 | 0.16 to 0.59 | 0.001 |
| | | 12003 | *Threshold / cut-point* | Never\|Rarely | | -2.07 | 0.09 | -2.24 to -1.90 | 0.000 |
| | | 12003 | *Threshold / cut-point* | Rarely\|Often | | 0.12 | 0.08 | -0.04 to 0.28 | 0.147 |
| | | 12003 | *Threshold / cut-point* | Often\|Always | | 0.91 | 0.08 | 0.74 to 1.07 | 0.000 |
| **Obeying the law§** | **Proportional-odds regression** | 12130 | Childhood predictor | Somewhat good | Very good | -1.21 | 0.12 | -1.45 to -0.98 | 0.000 |
| | | 12130 | Childhood predictor | Somewhat bad or very bad | Very good | -0.87 | 0.25 | -1.36 to -0.38 | 0.001 |
| | | 12130 | Age | Per 1-year increase | | 0.00 | 0.00 | -0.01 to 0.00 | 0.496 |
| | | 12130 | Gender | Female | Male | -0.18 | 0.08 | -0.34 to -0.03 | 0.017 |
| | | 12130 | Education | Secondary | Elementary or less | 0.11 | 0.10 | -0.08 to 0.30 | 0.239 |
| | | 12130 | Education | Four years beyond HS | Elementary or less | -0.21 | 0.13 | -0.46 to 0.05 | 0.114 |
| | | 12130 | Residential setting | Small town | Rural | -0.09 | 0.14 | -0.36 to 0.19 | 0.536 |
| | | 12130 | Residential setting | Large city | Rural | -0.25 | 0.18 | -0.60 to 0.09 | 0.146 |
| | | 12130 | Residential setting | Suburb | Rural | -0.29 | 0.22 | -0.71 to 0.14 | 0.192 |
| | | 12130 | *Threshold / cut-point* | Strongly disagree\|Somewhat disagree | | -5.35 | 0.25 | -5.84 to -4.86 | 0.000 |
| | | 12130 | *Threshold / cut-point* | Somewhat disagree\|Neither | | -4.74 | 0.21 | -5.15 to -4.34 | 0.000 |
| | | 12130 | *Threshold / cut-point* | Neither\|Somewhat agree | | -4.25 | 0.20 | -4.63 to -3.86 | 0.000 |
| | | 12130 | *Threshold / cut-point* | Somewhat agree\|Strongly agree | | -2.40 | 0.17 | -2.74 to -2.07 | 0.000 |
| **Feel at peace§** | **Proportional-odds regression** | 12071 | Childhood predictor | Somewhat good | Very good | -0.25 | 0.08 | -0.41 to -0.09 | 0.002 |
| | | 12071 | Childhood predictor | Somewhat bad or very bad | Very good | 0.01 | 0.17 | -0.33 to 0.35 | 0.963 |
| | | 12071 | Age | Per 1-year increase | | 0.01 | 0.00 | 0.00 to 0.01 | 0.000 |
| | | 12071 | Gender | Female | Male | 0.00 | 0.04 | -0.08 to 0.07 | 0.989 |

| Outcome | Model type | Analytic N | Variable | Level / comparison | Reference category | Coefficient | Standard error | 95% CI | p-value |
|---|---|---|---|---|---|---|---|---|---|
| | | 12071 | Education | Secondary | Elementary or less | 0.02 | 0.05 | -0.07 to 0.11 | 0.683 |
| | | 12071 | Education | Four years beyond HS | Elementary or less | 0.06 | 0.06 | -0.07 to 0.19 | 0.344 |
| | | 12071 | Residential setting | Small town | Rural | 0.19 | 0.05 | 0.08 to 0.30 | 0.000 |
| | | 12071 | Residential setting | Large city | Rural | 0.14 | 0.07 | 0.00 to 0.28 | 0.056 |
| | | 12071 | Residential setting | Suburb | Rural | 0.24 | 0.10 | 0.04 to 0.44 | 0.018 |
| | | 12071 | *Threshold / cut-point* | Never\|Rarely | | -1.89 | 0.08 | -2.06 to -1.73 | 0.000 |
| | | 12071 | *Threshold / cut-point* | Rarely\|Often | | -0.07 | 0.08 | -0.23 to 0.09 | 0.399 |
| | | 12071 | *Threshold / cut-point* | Often\|Always | | 0.63 | 0.08 | 0.47 to 0.79 | 0.000 |
| **Say in government#** | **Proportional-odds regression** | 12107 | Childhood predictor | Somewhat good | Very good | -0.07 | 0.10 | -0.26 to 0.13 | 0.494 |
| | | 12107 | Childhood predictor | Somewhat bad or very bad | Very good | -0.01 | 0.21 | -0.41 to 0.39 | 0.963 |
| | | 12107 | Age | Per 1-year increase | | 0.00 | 0.00 | -0.01 to 0.00 | 0.298 |
| | | 12107 | Gender | Female | Male | 0.08 | 0.05 | -0.02 to 0.18 | 0.115 |
| | | 12107 | Education | Secondary | Elementary or less | -0.26 | 0.06 | -0.37 to -0.14 | 0.000 |
| | | 12107 | Education | Four years beyond HS | Elementary or less | -0.14 | 0.10 | -0.34 to 0.06 | 0.164 |
| | | 12107 | Residential setting | Small town | Rural | 0.03 | 0.08 | -0.11 to 0.18 | 0.654 |
| | | 12107 | Residential setting | Large city | Rural | -0.13 | 0.09 | -0.31 to 0.06 | 0.173 |
| | | 12107 | Residential setting | Suburb | Rural | -0.43 | 0.13 | -0.69 to -0.16 | 0.001 |
| | | 12107 | *Threshold / cut-point* | Disagree\|Unsure | | -1.82 | 0.11 | -2.04 to -1.59 | 0.000 |
| | | 12107 | *Threshold / cut-point* | Unsure\|Agree | | -1.42 | 0.11 | -1.64 to -1.21 | 0.000 |
| **Extent of suffering§** | **Proportional-odds regression** | 12085 | Childhood predictor | Somewhat good | Very good | 0.12 | 0.08 | -0.04 to 0.27 | 0.141 |
| | | 12085 | Childhood predictor | Somewhat bad or very bad | Very good | 0.29 | 0.21 | -0.12 to 0.69 | 0.164 |
| | | 12085 | Age | Per 1-year increase | | 0.02 | 0.00 | 0.01 to 0.02 | 0.000 |
| | | 12085 | Gender | Female | Male | 0.26 | 0.04 | 0.18 to 0.34 | 0.000 |
| | | 12085 | Education | Secondary | Elementary or less | -0.15 | 0.05 | -0.24 to -0.06 | 0.001 |
| | | 12085 | Education | Four years beyond HS | Elementary or less | -0.26 | 0.08 | -0.41 to -0.12 | 0.000 |
| | | 12085 | Residential setting | Small town | Rural | 0.05 | 0.06 | -0.07 to 0.16 | 0.419 |
| | | 12085 | Residential setting | Large city | Rural | -0.13 | 0.08 | -0.28 to 0.02 | 0.095 |
| | | 12085 | Residential setting | Suburb | Rural | 0.08 | 0.11 | -0.14 to 0.30 | 0.496 |
| | | 12085 | *Threshold / cut-point* | Not at all\|Not very much | | 0.31 | 0.08 | 0.15 to 0.48 | 0.000 |
| | | 12085 | *Threshold / cut-point* | Not very much\|Some | | 0.73 | 0.08 | 0.56 to 0.89 | 0.000 |
| | | 12085 | *Threshold / cut-point* | Some\|A lot | | 2.17 | 0.09 | 2.00 to 2.34 | 0.000 |
| **Distress from life-threatening experiences§** | **Proportional-odds regression** | 12101 | Childhood predictor | Somewhat good | Very good | 0.05 | 0.07 | -0.10 to 0.19 | 0.529 |
| | | 12101 | Childhood predictor | Somewhat bad or very bad | Very good | 0.28 | 0.17 | -0.06 to 0.61 | 0.105 |
| | | 12101 | Age | Per 1-year increase | | 0.00 | 0.00 | 0.00 to 0.01 | 0.021 |
| | | 12101 | Gender | Female | Male | 0.07 | 0.05 | -0.02 to 0.16 | 0.127 |
| | | 12101 | Education | Secondary | Elementary or less | -0.26 | 0.05 | -0.36 to -0.17 | 0.000 |
| | | 12101 | Education | Four years beyond HS | Elementary or less | -0.44 | 0.08 | -0.59 to -0.29 | 0.000 |
| | | 12101 | Residential setting | Small town | Rural | -0.15 | 0.05 | -0.25 to -0.06 | 0.002 |

| Outcome | Model type | Analytic N | Variable | Level / comparison | Reference category | Coefficient | Standard error | 95% CI | p-value |
|---|---|---|---|---|---|---|---|---|---|
| | | 12101 | Residential setting | Large city | Rural | -0.21 | 0.07 | -0.35 to -0.06 | 0.004 |
| | | 12101 | Residential setting | Suburb | Rural | -0.16 | 0.12 | -0.40 to 0.08 | 0.186 |
| | | 12101 | *Threshold / cut-point* | Not at all\|Not very much | | -0.28 | 0.08 | -0.44 to -0.13 | 0.000 |
| | | 12101 | *Threshold / cut-point* | Not very much\|Some | | 0.06 | 0.08 | -0.09 to 0.21 | 0.438 |
| | | 12101 | *Threshold / cut-point* | Some\|A lot | | 1.08 | 0.08 | 0.92 to 1.23 | 0.000 |
| **Trust in others§** | **Proportional-odds regression** | 11831 | Childhood predictor | Somewhat good | Very good | -0.27 | 0.10 | -0.46 to -0.07 | 0.009 |
| | | 11831 | Childhood predictor | Somewhat bad or very bad | Very good | 0.01 | 0.16 | -0.30 to 0.32 | 0.948 |
| | | 11831 | Age | Per 1-year increase | | 0.00 | 0.00 | 0.00 to 0.01 | 0.012 |
| | | 11831 | Gender | Female | Male | -0.04 | 0.04 | -0.12 to 0.04 | 0.306 |
| | | 11831 | Education | Secondary | Elementary or less | -0.03 | 0.05 | -0.12 to 0.06 | 0.507 |
| | | 11831 | Education | Four years beyond HS | Elementary or less | -0.04 | 0.07 | -0.18 to 0.10 | 0.613 |
| | | 11831 | Residential setting | Small town | Rural | 0.23 | 0.05 | 0.13 to 0.34 | 0.000 |
| | | 11831 | Residential setting | Large city | Rural | -0.01 | 0.07 | -0.14 to 0.12 | 0.904 |
| | | 11831 | Residential setting | Suburb | Rural | -0.20 | 0.14 | -0.49 to 0.08 | 0.158 |
| | | 11831 | *Threshold / cut-point* | None\|Not very many | | -2.14 | 0.09 | -2.32 to -1.96 | 0.000 |
| | | 11831 | *Threshold / cut-point* | Not very many\|Some | | -1.35 | 0.09 | -1.53 to -1.17 | 0.000 |
| | | 11831 | *Threshold / cut-point* | Some\|Most | | 0.85 | 0.09 | 0.67 to 1.02 | 0.000 |
| | | 11831 | *Threshold / cut-point* | Most\|All | | 1.82 | 0.09 | 1.64 to 2.00 | 0.000 |

**eTable 13.** Table of coefficients for adjusted associations between poorly self-rated childhood health as predictor (excellent self-rated childhood health as reference category) and 45 outcomes of interest and two flourishing composite indices.

Adjusted survey-weighted models use ANNUAL_WEIGHT_C1 with primary sampling unit (PSU) and strata and adjust for age, gender, education, and residential setting. Reference categories are shown in the table. Coefficients are on the original regression-coefficient scale: beta for continuous outcomes, log-odds for binary outcomes, and proportional-odds log coefficients for ordinal outcomes. Model intercepts are displayed for continuous and binary outcomes. Proportional-odds models do not have a single conventional intercept; their estimated ordinal cut-points/thresholds are displayed after the covariate coefficients, with standard errors (SEs), 95% CIs, and Wald p-values.

***Note:*** All results except p-values are rounded to the two decimals. P-values are rounded to three decimals. Values displayed as 0.000 indicate $P < 0.0005$ due to rounding, rather than an exact zero. Statistical significance was determined using unrounded P-values at the Bonferroni-adjusted threshold of $P < 0.0011$.

| Outcome | Model type | Analytic N | Variable | Level / comparison | Reference category | Coefficient | Standard error | 95% CI | p-value |
|---|---|---|---|---|---|---|---|---|---|
| **Belonging** | **Linear regression** | 12616 | *Intercept* | | | 8.80 | 0.14 | 8.53 to 9.08 | 0.000 |
| | | 12616 | Childhood predictor | Very good | Excellent | 0.00 | 0.10 | -0.19 to 0.19 | 0.989 |
| | | 12616 | Childhood predictor | Good | Excellent | -0.12 | 0.10 | -0.32 to 0.07 | 0.219 |
| | | 12616 | Childhood predictor | Fair | Excellent | -0.30 | 0.11 | -0.51 to -0.09 | 0.005 |
| | | 12616 | Childhood predictor | Poor | Excellent | -0.47 | 0.22 | -0.91 to -0.04 | 0.033 |
| | | 12616 | Age | Per 1-year increase | | -0.01 | 0.00 | -0.02 to -0.01 | 0.000 |
| | | 12616 | Gender | Female | Male | -0.18 | 0.06 | -0.30 to -0.06 | 0.003 |
| | | 12616 | Education | Secondary | Elementary or less | 0.19 | 0.07 | 0.04 to 0.33 | 0.011 |
| | | 12616 | Education | Four years beyond HS | Elementary or less | 0.29 | 0.10 | 0.09 to 0.50 | 0.005 |
| | | 12616 | Residential setting | Small town | Rural | 0.34 | 0.08 | 0.19 to 0.49 | 0.000 |
| | | 12616 | Residential setting | Large city | Rural | 0.38 | 0.10 | 0.18 to 0.59 | 0.000 |
| | | 12616 | Residential setting | Suburb | Rural | -0.01 | 0.18 | -0.35 to 0.34 | 0.973 |
| **Relationship contentment‡** | **Linear regression** | 12679 | *Intercept* | | | 8.17 | 0.15 | 7.87 to 8.47 | 0.000 |
| | | 12679 | Childhood predictor | Very good | Excellent | -0.03 | 0.11 | -0.24 to 0.19 | 0.808 |
| | | 12679 | Childhood predictor | Good | Excellent | -0.03 | 0.11 | -0.24 to 0.18 | 0.775 |
| | | 12679 | Childhood predictor | Fair | Excellent | -0.17 | 0.12 | -0.41 to 0.06 | 0.149 |
| | | 12679 | Childhood predictor | Poor | Excellent | -0.02 | 0.19 | -0.40 to 0.36 | 0.921 |
| | | 12679 | Age | Per 1-year increase | | -0.01 | 0.00 | -0.02 to -0.01 | 0.000 |
| | | 12679 | Gender | Female | Male | 0.38 | 0.07 | 0.25 to 0.52 | 0.000 |
| | | 12679 | Education | Secondary | Elementary or less | 0.15 | 0.07 | 0.01 to 0.30 | 0.038 |
| | | 12679 | Education | Four years beyond HS | Elementary or less | 0.29 | 0.12 | 0.06 to 0.52 | 0.012 |
| | | 12679 | Residential setting | Small town | Rural | 0.36 | 0.08 | 0.20 to 0.52 | 0.000 |
| | | 12679 | Residential setting | Large city | Rural | 0.06 | 0.10 | -0.13 to 0.25 | 0.519 |
| | | 12679 | Residential setting | Suburb | Rural | -0.29 | 0.17 | -0.62 to 0.05 | 0.095 |
| **Positive expectations** | **Linear regression** | 12640 | *Intercept* | | | 7.80 | 0.18 | 7.44 to 8.16 | 0.000 |
| | | 12640 | Childhood predictor | Very good | Excellent | -0.04 | 0.10 | -0.24 to 0.16 | 0.697 |
| | | 12640 | Childhood predictor | Good | Excellent | 0.05 | 0.11 | -0.17 to 0.27 | 0.681 |
| | | 12640 | Childhood predictor | Fair | Excellent | -0.20 | 0.12 | -0.44 to 0.04 | 0.104 |
| | | 12640 | Childhood predictor | Poor | Excellent | 0.01 | 0.20 | -0.37 to 0.40 | 0.942 |
| | | 12640 | Age | Per 1-year increase | | 0.00 | 0.00 | -0.01 to 0.00 | 0.261 |
| | | 12640 | Gender | Female | Male | 0.22 | 0.06 | 0.09 to 0.34 | 0.001 |

| Outcome | Model type | Analytic N | Variable | Level / comparison | Reference category | Coefficient | Standard error | 95% CI | p-value |
|---|---|---|---|---|---|---|---|---|---|
| | | 12640 | Education | Secondary | Elementary or less | 0.28 | 0.07 | 0.13 to 0.42 | 0.000 |
| | | 12640 | Education | Four years beyond HS | Elementary or less | 0.36 | 0.10 | 0.17 to 0.55 | 0.000 |
| | | 12640 | Residential setting | Small town | Rural | 0.45 | 0.09 | 0.28 to 0.62 | 0.000 |
| | | 12640 | Residential setting | Large city | Rural | 0.45 | 0.11 | 0.23 to 0.67 | 0.000 |
| | | 12640 | Residential setting | Suburb | Rural | 0.46 | 0.16 | 0.15 to 0.78 | 0.004 |
| **Worry about meeting normal monthly living expenses§§‡‡** | **Linear regression** | 12681 | *Intercept* | | | 5.46 | 0.23 | 5.01 to 5.91 | 0.000 |
| | | 12681 | Childhood predictor | Very good | Excellent | 0.11 | 0.16 | -0.19 to 0.42 | 0.472 |
| | | 12681 | Childhood predictor | Good | Excellent | 0.51 | 0.16 | 0.20 to 0.81 | 0.001 |
| | | 12681 | Childhood predictor | Fair | Excellent | 0.43 | 0.16 | 0.10 to 0.75 | 0.010 |
| | | 12681 | Childhood predictor | Poor | Excellent | 0.97 | 0.28 | 0.42 to 1.51 | 0.001 |
| | | 12681 | Age | Per 1-year increase | | 0.00 | 0.00 | -0.01 to 0.01 | 0.746 |
| | | 12681 | Gender | Female | Male | 0.36 | 0.09 | 0.18 to 0.53 | 0.000 |
| | | 12681 | Education | Secondary | Elementary or less | -0.41 | 0.11 | -0.63 to -0.20 | 0.000 |
| | | 12681 | Education | Four years beyond HS | Elementary or less | -0.60 | 0.15 | -0.89 to -0.30 | 0.000 |
| | | 12681 | Residential setting | Small town | Rural | 0.24 | 0.13 | -0.01 to 0.48 | 0.062 |
| | | 12681 | Residential setting | Large city | Rural | -0.17 | 0.15 | -0.47 to 0.12 | 0.253 |
| | | 12681 | Residential setting | Suburb | Rural | -0.16 | 0.24 | -0.63 to 0.32 | 0.520 |
| **Freedom** | **Linear regression** | 12654 | *Intercept* | | | 8.38 | 0.14 | 8.10 to 8.66 | 0.000 |
| | | 12654 | Childhood predictor | Very good | Excellent | -0.16 | 0.10 | -0.34 to 0.03 | 0.107 |
| | | 12654 | Childhood predictor | Good | Excellent | -0.14 | 0.09 | -0.32 to 0.04 | 0.123 |
| | | 12654 | Childhood predictor | Fair | Excellent | -0.34 | 0.11 | -0.54 to -0.13 | 0.002 |
| | | 12654 | Childhood predictor | Poor | Excellent | -0.56 | 0.21 | -0.97 to -0.15 | 0.007 |
| | | 12654 | Age | Per 1-year increase | | -0.01 | 0.00 | -0.01 to 0.00 | 0.001 |
| | | 12654 | Gender | Female | Male | -0.12 | 0.07 | -0.25 to 0.01 | 0.074 |
| | | 12654 | Education | Secondary | Elementary or less | 0.28 | 0.07 | 0.14 to 0.43 | 0.000 |
| | | 12654 | Education | Four years beyond HS | Elementary or less | 0.39 | 0.10 | 0.19 to 0.59 | 0.000 |
| | | 12654 | Residential setting | Small town | Rural | 0.34 | 0.08 | 0.19 to 0.49 | 0.000 |
| | | 12654 | Residential setting | Large city | Rural | 0.37 | 0.10 | 0.17 to 0.58 | 0.000 |
| | | 12654 | Residential setting | Suburb | Rural | 0.11 | 0.15 | -0.18 to 0.41 | 0.451 |
| **Give up some happiness now for greater happiness later‡** | **Linear regression** | 12621 | *Intercept* | | | 7.22 | 0.19 | 6.85 to 7.59 | 0.000 |
| | | 12621 | Childhood predictor | Very good | Excellent | 0.16 | 0.13 | -0.09 to 0.42 | 0.216 |
| | | 12621 | Childhood predictor | Good | Excellent | 0.10 | 0.13 | -0.15 to 0.35 | 0.446 |
| | | 12621 | Childhood predictor | Fair | Excellent | -0.18 | 0.14 | -0.45 to 0.09 | 0.184 |
| | | 12621 | Childhood predictor | Poor | Excellent | -0.27 | 0.22 | -0.71 to 0.16 | 0.216 |
| | | 12621 | Age | Per 1-year increase | | -0.01 | 0.00 | -0.02 to 0.00 | 0.000 |
| | | 12621 | Gender | Female | Male | 0.16 | 0.08 | 0.01 to 0.31 | 0.039 |
| | | 12621 | Education | Secondary | Elementary or less | 0.16 | 0.10 | -0.03 to 0.35 | 0.092 |
| | | 12621 | Education | Four years beyond HS | Elementary or less | 0.17 | 0.13 | -0.09 to 0.44 | 0.198 |
| | | 12621 | Residential setting | Small town | Rural | 0.38 | 0.10 | 0.19 to 0.57 | 0.000 |

| Outcome | Model type | Analytic N | Variable | Level / comparison | Reference category | Coefficient | Standard error | 95% CI | p-value |
|---|---|---|---|---|---|---|---|---|---|
| | | 12621 | Residential setting | Large city | Rural | 0.16 | 0.13 | -0.10 to 0.42 | 0.226 |
| | | 12621 | Residential setting | Suburb | Rural | 0.08 | 0.27 | -0.46 to 0.62 | 0.775 |
| **Have a long list of gratitude** | **Linear regression** | 12510 | *Intercept* | | | 7.58 | 0.18 | 7.23 to 7.93 | 0.000 |
| | | 12510 | Childhood predictor | Very good | Excellent | -0.09 | 0.11 | -0.30 to 0.12 | 0.412 |
| | | 12510 | Childhood predictor | Good | Excellent | -0.17 | 0.11 | -0.40 to 0.05 | 0.127 |
| | | 12510 | Childhood predictor | Fair | Excellent | -0.26 | 0.12 | -0.50 to -0.01 | 0.040 |
| | | 12510 | Childhood predictor | Poor | Excellent | -0.31 | 0.22 | -0.75 to 0.12 | 0.157 |
| | | 12510 | Age | Per 1-year increase | | -0.01 | 0.00 | -0.01 to 0.00 | 0.058 |
| | | 12510 | Gender | Female | Male | 0.21 | 0.07 | 0.08 to 0.35 | 0.002 |
| | | 12510 | Education | Secondary | Elementary or less | 0.27 | 0.08 | 0.11 to 0.42 | 0.001 |
| | | 12510 | Education | Four years beyond HS | Elementary or less | 0.37 | 0.11 | 0.14 to 0.59 | 0.001 |
| | | 12510 | Residential setting | Small town | Rural | 0.41 | 0.09 | 0.22 to 0.59 | 0.000 |
| | | 12510 | Residential setting | Large city | Rural | 0.41 | 0.13 | 0.15 to 0.68 | 0.002 |
| | | 12510 | Residential setting | Suburb | Rural | 0.48 | 0.17 | 0.15 to 0.81 | 0.005 |
| **Happiness‡** | **Linear regression** | 12679 | *Intercept* | | | 6.81 | 0.18 | 6.45 to 7.16 | 0.000 |
| | | 12679 | Childhood predictor | Very good | Excellent | -0.04 | 0.12 | -0.28 to 0.20 | 0.749 |
| | | 12679 | Childhood predictor | Good | Excellent | -0.03 | 0.13 | -0.28 to 0.22 | 0.810 |
| | | 12679 | Childhood predictor | Fair | Excellent | -0.45 | 0.15 | -0.74 to -0.16 | 0.003 |
| | | 12679 | Childhood predictor | Poor | Excellent | -0.82 | 0.23 | -1.27 to -0.36 | 0.001 |
| | | 12679 | Age | Per 1-year increase | | -0.02 | 0.00 | -0.02 to -0.01 | 0.000 |
| | | 12679 | Gender | Female | Male | 0.25 | 0.08 | 0.10 to 0.40 | 0.001 |
| | | 12679 | Education | Secondary | Elementary or less | 0.43 | 0.09 | 0.25 to 0.60 | 0.000 |
| | | 12679 | Education | Four years beyond HS | Elementary or less | 0.67 | 0.12 | 0.43 to 0.91 | 0.000 |
| | | 12679 | Residential setting | Small town | Rural | 0.35 | 0.10 | 0.15 to 0.55 | 0.001 |
| | | 12679 | Residential setting | Large city | Rural | 0.30 | 0.12 | 0.06 to 0.54 | 0.016 |
| | | 12679 | Residential setting | Suburb | Rural | -0.10 | 0.22 | -0.52 to 0.32 | 0.634 |
| **Remain hopeful for the future** | **Linear regression** | 12627 | *Intercept* | | | 8.21 | 0.15 | 7.92 to 8.50 | 0.000 |
| | | 12627 | Childhood predictor | Very good | Excellent | 0.06 | 0.09 | -0.12 to 0.24 | 0.540 |
| | | 12627 | Childhood predictor | Good | Excellent | -0.06 | 0.10 | -0.25 to 0.12 | 0.504 |
| | | 12627 | Childhood predictor | Fair | Excellent | -0.14 | 0.10 | -0.34 to 0.06 | 0.170 |
| | | 12627 | Childhood predictor | Poor | Excellent | -0.42 | 0.20 | -0.82 to -0.02 | 0.039 |
| | | 12627 | Age | Per 1-year increase | | -0.01 | 0.00 | -0.01 to 0.00 | 0.001 |
| | | 12627 | Gender | Female | Male | 0.02 | 0.06 | -0.09 to 0.13 | 0.754 |
| | | 12627 | Education | Secondary | Elementary or less | 0.37 | 0.06 | 0.25 to 0.50 | 0.000 |
| | | 12627 | Education | Four years beyond HS | Elementary or less | 0.59 | 0.09 | 0.41 to 0.77 | 0.000 |
| | | 12627 | Residential setting | Small town | Rural | 0.48 | 0.08 | 0.32 to 0.63 | 0.000 |
| | | 12627 | Residential setting | Large city | Rural | 0.45 | 0.10 | 0.25 to 0.66 | 0.000 |
| | | 12627 | Residential setting | Suburb | Rural | 0.40 | 0.17 | 0.06 to 0.74 | 0.021 |
| **Understand purpose in life‡** | **Linear regression** | 12604 | *Intercept* | | | 8.20 | 0.15 | 7.90 to 8.50 | 0.000 |
| | | 12604 | Childhood predictor | Very good | Excellent | -0.16 | 0.09 | -0.34 to 0.03 | 0.093 |

| Outcome | Model type | Analytic N | Variable | Level / comparison | Reference category | Coefficient | Standard error | 95% CI | p-value |
|---|---|---|---|---|---|---|---|---|---|
| | | 12604 | Childhood predictor | Good | Excellent | -0.19 | 0.10 | -0.39 to 0.01 | 0.066 |
| | | 12604 | Childhood predictor | Fair | Excellent | -0.24 | 0.11 | -0.46 to -0.02 | 0.033 |
| | | 12604 | Childhood predictor | Poor | Excellent | -0.29 | 0.21 | -0.71 to 0.12 | 0.169 |
| | | 12604 | Age | Per 1-year increase | | -0.01 | 0.00 | -0.02 to -0.01 | 0.000 |
| | | 12604 | Gender | Female | Male | 0.13 | 0.07 | -0.01 to 0.27 | 0.063 |
| | | 12604 | Education | Secondary | Elementary or less | 0.41 | 0.08 | 0.26 to 0.57 | 0.000 |
| | | 12604 | Education | Four years beyond HS | Elementary or less | 0.73 | 0.10 | 0.54 to 0.91 | 0.000 |
| | | 12604 | Residential setting | Small town | Rural | 0.41 | 0.09 | 0.23 to 0.59 | 0.000 |
| | | 12604 | Residential setting | Large city | Rural | 0.42 | 0.12 | 0.19 to 0.65 | 0.000 |
| | | 12604 | Residential setting | Suburb | Rural | 0.43 | 0.15 | 0.14 to 0.72 | 0.004 |
| **Life satisfaction‡** | **Linear regression** | 12659 | *Intercept* | | | 7.19 | 0.17 | 6.85 to 7.53 | 0.000 |
| | | 12659 | Childhood predictor | Very good | Excellent | 0.06 | 0.11 | -0.15 to 0.28 | 0.568 |
| | | 12659 | Childhood predictor | Good | Excellent | -0.03 | 0.12 | -0.26 to 0.20 | 0.772 |
| | | 12659 | Childhood predictor | Fair | Excellent | -0.39 | 0.14 | -0.66 to -0.11 | 0.006 |
| | | 12659 | Childhood predictor | Poor | Excellent | -0.73 | 0.24 | -1.19 to -0.27 | 0.002 |
| | | 12659 | Age | Per 1-year increase | | -0.01 | 0.00 | -0.02 to -0.01 | 0.000 |
| | | 12659 | Gender | Female | Male | 0.31 | 0.08 | 0.16 to 0.46 | 0.000 |
| | | 12659 | Education | Secondary | Elementary or less | 0.31 | 0.09 | 0.13 to 0.48 | 0.001 |
| | | 12659 | Education | Four years beyond HS | Elementary or less | 0.49 | 0.12 | 0.25 to 0.72 | 0.000 |
| | | 12659 | Residential setting | Small town | Rural | 0.26 | 0.09 | 0.08 to 0.44 | 0.005 |
| | | 12659 | Residential setting | Large city | Rural | 0.18 | 0.12 | -0.06 to 0.42 | 0.146 |
| | | 12659 | Residential setting | Suburb | Rural | -0.22 | 0.21 | -0.63 to 0.19 | 0.290 |
| **Loneliness§** | **Linear regression** | 12679 | *Intercept* | | | 4.19 | 0.20 | 3.79 to 4.58 | 0.000 |
| | | 12679 | Childhood predictor | Very good | Excellent | -0.06 | 0.14 | -0.33 to 0.21 | 0.645 |
| | | 12679 | Childhood predictor | Good | Excellent | 0.03 | 0.14 | -0.24 to 0.30 | 0.822 |
| | | 12679 | Childhood predictor | Fair | Excellent | 0.28 | 0.15 | -0.02 to 0.57 | 0.065 |
| | | 12679 | Childhood predictor | Poor | Excellent | 0.67 | 0.25 | 0.17 to 1.16 | 0.009 |
| | | 12679 | Age | Per 1-year increase | | 0.00 | 0.00 | -0.01 to 0.01 | 0.883 |
| | | 12679 | Gender | Female | Male | -0.15 | 0.09 | -0.32 to 0.02 | 0.088 |
| | | 12679 | Education | Secondary | Elementary or less | -0.32 | 0.10 | -0.51 to -0.13 | 0.001 |
| | | 12679 | Education | Four years beyond HS | Elementary or less | -0.65 | 0.14 | -0.93 to -0.36 | 0.000 |
| | | 12679 | Residential setting | Small town | Rural | -0.35 | 0.10 | -0.55 to -0.15 | 0.001 |
| | | 12679 | Residential setting | Large city | Rural | -0.47 | 0.14 | -0.74 to -0.20 | 0.001 |
| | | 12679 | Residential setting | Suburb | Rural | -0.13 | 0.24 | -0.60 to 0.34 | 0.597 |
| **Mental health‡** | **Linear regression** | 12683 | *Intercept* | | | 8.16 | 0.19 | 7.80 to 8.53 | 0.000 |
| | | 12683 | Childhood predictor | Very good | Excellent | 0.08 | 0.12 | -0.16 to 0.32 | 0.524 |
| | | 12683 | Childhood predictor | Good | Excellent | -0.07 | 0.12 | -0.31 to 0.16 | 0.542 |
| | | 12683 | Childhood predictor | Fair | Excellent | -0.35 | 0.14 | -0.62 to -0.08 | 0.011 |
| | | 12683 | Childhood predictor | Poor | Excellent | -0.73 | 0.23 | -1.17 to -0.28 | 0.001 |
| | | 12683 | Age | Per 1-year increase | | -0.02 | 0.00 | -0.03 to -0.02 | 0.000 |

| Outcome | Model type | Analytic N | Variable | Level / comparison | Reference category | Coefficient | Standard error | 95% CI | p-value |
|---|---|---|---|---|---|---|---|---|---|
| | | 12683 | Gender | Female | Male | -0.12 | 0.07 | -0.26 to 0.01 | 0.078 |
| | | 12683 | Education | Secondary | Elementary or less | 0.39 | 0.08 | 0.24 to 0.55 | 0.000 |
| | | 12683 | Education | Four years beyond HS | Elementary or less | 0.52 | 0.12 | 0.29 to 0.75 | 0.000 |
| | | 12683 | Residential setting | Small town | Rural | 0.38 | 0.09 | 0.19 to 0.56 | 0.000 |
| | | 12683 | Residential setting | Large city | Rural | 0.42 | 0.12 | 0.19 to 0.65 | 0.000 |
| | | 12683 | Residential setting | Suburb | Rural | 0.11 | 0.19 | -0.26 to 0.48 | 0.555 |
| **Perceived support from others** | **Linear regression** | 12668 | *Intercept* | | | 6.67 | 0.20 | 6.27 to 7.07 | 0.000 |
| | | 12668 | Childhood predictor | Very good | Excellent | 0.04 | 0.14 | -0.24 to 0.31 | 0.800 |
| | | 12668 | Childhood predictor | Good | Excellent | 0.14 | 0.14 | -0.13 to 0.42 | 0.309 |
| | | 12668 | Childhood predictor | Fair | Excellent | -0.30 | 0.15 | -0.60 to -0.01 | 0.046 |
| | | 12668 | Childhood predictor | Poor | Excellent | -0.34 | 0.25 | -0.83 to 0.16 | 0.183 |
| | | 12668 | Age | Per 1-year increase | | -0.01 | 0.00 | -0.02 to -0.01 | 0.000 |
| | | 12668 | Gender | Female | Male | 0.53 | 0.09 | 0.36 to 0.71 | 0.000 |
| | | 12668 | Education | Secondary | Elementary or less | 0.19 | 0.10 | 0.00 to 0.38 | 0.052 |
| | | 12668 | Education | Four years beyond HS | Elementary or less | 0.14 | 0.14 | -0.14 to 0.41 | 0.329 |
| | | 12668 | Residential setting | Small town | Rural | 0.16 | 0.11 | -0.05 to 0.37 | 0.141 |
| | | 12668 | Residential setting | Large city | Rural | -0.53 | 0.14 | -0.80 to -0.25 | 0.000 |
| | | 12668 | Residential setting | Suburb | Rural | -0.29 | 0.20 | -0.69 to 0.11 | 0.152 |
| **Physical health‡** | **Linear regression** | 12686 | *Intercept* | | | 8.69 | 0.17 | 8.36 to 9.03 | 0.000 |
| | | 12686 | Childhood predictor | Very good | Excellent | -0.04 | 0.10 | -0.24 to 0.17 | 0.734 |
| | | 12686 | Childhood predictor | Good | Excellent | -0.24 | 0.11 | -0.45 to -0.04 | 0.022 |
| | | 12686 | Childhood predictor | Fair | Excellent | -0.69 | 0.13 | -0.93 to -0.44 | 0.000 |
| | | 12686 | Childhood predictor | Poor | Excellent | -1.41 | 0.22 | -1.83 to -0.98 | 0.000 |
| | | 12686 | Age | Per 1-year increase | | -0.04 | 0.00 | -0.05 to -0.04 | 0.000 |
| | | 12686 | Gender | Female | Male | -0.33 | 0.07 | -0.48 to -0.19 | 0.000 |
| | | 12686 | Education | Secondary | Elementary or less | 0.33 | 0.08 | 0.18 to 0.48 | 0.000 |
| | | 12686 | Education | Four years beyond HS | Elementary or less | 0.62 | 0.11 | 0.40 to 0.83 | 0.000 |
| | | 12686 | Residential setting | Small town | Rural | 0.44 | 0.09 | 0.26 to 0.62 | 0.000 |
| | | 12686 | Residential setting | Large city | Rural | 0.58 | 0.11 | 0.37 to 0.79 | 0.000 |
| | | 12686 | Residential setting | Suburb | Rural | 0.09 | 0.18 | -0.25 to 0.44 | 0.594 |
| **Act to promote good‡** | **Linear regression** | 12641 | *Intercept* | | | 8.08 | 0.13 | 7.82 to 8.35 | 0.000 |
| | | 12641 | Childhood predictor | Very good | Excellent | 0.06 | 0.09 | -0.11 to 0.24 | 0.485 |
| | | 12641 | Childhood predictor | Good | Excellent | 0.02 | 0.09 | -0.16 to 0.19 | 0.855 |
| | | 12641 | Childhood predictor | Fair | Excellent | -0.20 | 0.10 | -0.40 to 0.00 | 0.056 |
| | | 12641 | Childhood predictor | Poor | Excellent | -0.06 | 0.17 | -0.40 to 0.28 | 0.739 |
| | | 12641 | Age | Per 1-year increase | | -0.01 | 0.00 | -0.01 to 0.00 | 0.001 |
| | | 12641 | Gender | Female | Male | 0.12 | 0.06 | 0.01 to 0.24 | 0.041 |
| | | 12641 | Education | Secondary | Elementary or less | 0.38 | 0.06 | 0.26 to 0.50 | 0.000 |
| | | 12641 | Education | Four years beyond HS | Elementary or less | 0.55 | 0.08 | 0.39 to 0.70 | 0.000 |
| | | 12641 | Residential setting | Small town | Rural | 0.59 | 0.07 | 0.45 to 0.73 | 0.000 |

| Outcome | Model type | Analytic N | Variable | Level / comparison | Reference category | Coefficient | Standard error | 95% CI | p-value |
|---|---|---|---|---|---|---|---|---|---|
| | | 12641 | Residential setting | Large city | Rural | 0.62 | 0.10 | 0.43 to 0.82 | 0.000 |
| | | 12641 | Residential setting | Suburb | Rural | 0.40 | 0.16 | 0.08 to 0.72 | 0.016 |
| **Relationship satisfaction‡** | **Linear regression** | 12647 | *Intercept* | | | 7.76 | 0.16 | 7.46 to 8.07 | 0.000 |
| | | 12647 | Childhood predictor | Very good | Excellent | 0.10 | 0.11 | -0.11 to 0.31 | 0.348 |
| | | 12647 | Childhood predictor | Good | Excellent | 0.21 | 0.11 | -0.01 to 0.42 | 0.058 |
| | | 12647 | Childhood predictor | Fair | Excellent | -0.02 | 0.12 | -0.25 to 0.21 | 0.834 |
| | | 12647 | Childhood predictor | Poor | Excellent | -0.18 | 0.22 | -0.62 to 0.25 | 0.412 |
| | | 12647 | Age | Per 1-year increase | | -0.01 | 0.00 | -0.01 to 0.00 | 0.001 |
| | | 12647 | Gender | Female | Male | 0.33 | 0.07 | 0.19 to 0.47 | 0.000 |
| | | 12647 | Education | Secondary | Elementary or less | -0.02 | 0.08 | -0.17 to 0.14 | 0.844 |
| | | 12647 | Education | Four years beyond HS | Elementary or less | 0.06 | 0.11 | -0.15 to 0.28 | 0.574 |
| | | 12647 | Residential setting | Small town | Rural | 0.44 | 0.09 | 0.27 to 0.61 | 0.000 |
| | | 12647 | Residential setting | Large city | Rural | 0.32 | 0.11 | 0.11 to 0.53 | 0.003 |
| | | 12647 | Residential setting | Suburb | Rural | 0.07 | 0.16 | -0.24 to 0.38 | 0.677 |
| **Show love often** | **Linear regression** | 12670 | *Intercept* | | | 8.25 | 0.15 | 7.96 to 8.54 | 0.000 |
| | | 12670 | Childhood predictor | Very good | Excellent | 0.04 | 0.11 | -0.16 to 0.25 | 0.675 |
| | | 12670 | Childhood predictor | Good | Excellent | 0.07 | 0.11 | -0.14 to 0.28 | 0.514 |
| | | 12670 | Childhood predictor | Fair | Excellent | -0.12 | 0.11 | -0.34 to 0.11 | 0.312 |
| | | 12670 | Childhood predictor | Poor | Excellent | 0.07 | 0.20 | -0.32 to 0.45 | 0.730 |
| | | 12670 | Age | Per 1-year increase | | -0.01 | 0.00 | -0.01 to 0.00 | 0.002 |
| | | 12670 | Gender | Female | Male | 0.21 | 0.07 | 0.09 to 0.34 | 0.001 |
| | | 12670 | Education | Secondary | Elementary or less | 0.16 | 0.07 | 0.03 to 0.30 | 0.021 |
| | | 12670 | Education | Four years beyond HS | Elementary or less | 0.05 | 0.11 | -0.16 to 0.27 | 0.633 |
| | | 12670 | Residential setting | Small town | Rural | 0.20 | 0.08 | 0.05 to 0.35 | 0.009 |
| | | 12670 | Residential setting | Large city | Rural | 0.02 | 0.11 | -0.20 to 0.24 | 0.838 |
| | | 12670 | Residential setting | Suburb | Rural | -0.13 | 0.20 | -0.52 to 0.26 | 0.503 |
| **Life evaluation five years from now** | **Linear regression** | 11547 | *Intercept* | | | 7.82 | 0.15 | 7.52 to 8.12 | 0.000 |
| | | 11547 | Childhood predictor | Very good | Excellent | -0.03 | 0.11 | -0.24 to 0.18 | 0.779 |
| | | 11547 | Childhood predictor | Good | Excellent | -0.05 | 0.11 | -0.26 to 0.16 | 0.646 |
| | | 11547 | Childhood predictor | Fair | Excellent | -0.43 | 0.12 | -0.66 to -0.19 | 0.000 |
| | | 11547 | Childhood predictor | Poor | Excellent | -0.38 | 0.20 | -0.78 to 0.02 | 0.064 |
| | | 11547 | Age | Per 1-year increase | | -0.02 | 0.00 | -0.03 to -0.02 | 0.000 |
| | | 11547 | Gender | Female | Male | 0.20 | 0.07 | 0.06 to 0.34 | 0.006 |
| | | 11547 | Education | Secondary | Elementary or less | 0.27 | 0.08 | 0.11 to 0.42 | 0.001 |
| | | 11547 | Education | Four years beyond HS | Elementary or less | 0.56 | 0.11 | 0.34 to 0.79 | 0.000 |
| | | 11547 | Residential setting | Small town | Rural | 0.44 | 0.09 | 0.26 to 0.62 | 0.000 |
| | | 11547 | Residential setting | Large city | Rural | 0.48 | 0.11 | 0.26 to 0.70 | 0.000 |
| | | 11547 | Residential setting | Suburb | Rural | 0.32 | 0.23 | -0.13 to 0.76 | 0.161 |
| **Life evaluation today** | **Linear regression** | 12580 | *Intercept* | | | 5.61 | 0.19 | 5.24 to 5.97 | 0.000 |
| | | 12580 | Childhood predictor | Very good | Excellent | 0.09 | 0.12 | -0.15 to 0.33 | 0.463 |

| Outcome | Model type | Analytic N | Variable | Level / comparison | Reference category | Coefficient | Standard error | 95% CI | p-value |
|---|---|---|---|---|---|---|---|---|---|
| | | 12580 | Childhood predictor | Good | Excellent | 0.17 | 0.12 | -0.07 to 0.41 | 0.172 |
| | | 12580 | Childhood predictor | Fair | Excellent | -0.33 | 0.13 | -0.59 to -0.07 | 0.013 |
| | | 12580 | Childhood predictor | Poor | Excellent | -0.98 | 0.27 | -1.50 to -0.45 | 0.000 |
| | | 12580 | Age | Per 1-year increase | | -0.01 | 0.00 | -0.02 to 0.00 | 0.002 |
| | | 12580 | Gender | Female | Male | 0.49 | 0.08 | 0.32 to 0.65 | 0.000 |
| | | 12580 | Education | Secondary | Elementary or less | 0.23 | 0.08 | 0.06 to 0.39 | 0.007 |
| | | 12580 | Education | Four years beyond HS | Elementary or less | 0.19 | 0.14 | -0.09 to 0.47 | 0.190 |
| | | 12580 | Residential setting | Small town | Rural | 0.21 | 0.10 | 0.01 to 0.41 | 0.045 |
| | | 12580 | Residential setting | Large city | Rural | 0.06 | 0.13 | -0.20 to 0.32 | 0.647 |
| | | 12580 | Residential setting | Suburb | Rural | -0.07 | 0.21 | -0.48 to 0.34 | 0.739 |
| **Worry about safety, food, or housing§§‡‡** | **Linear regression** | 12695 | *Intercept* | | | 5.96 | 0.22 | 5.52 to 6.40 | 0.000 |
| | | 12695 | Childhood predictor | Very good | Excellent | 0.14 | 0.16 | -0.18 to 0.47 | 0.387 |
| | | 12695 | Childhood predictor | Good | Excellent | 0.41 | 0.17 | 0.08 to 0.74 | 0.015 |
| | | 12695 | Childhood predictor | Fair | Excellent | 0.28 | 0.17 | -0.06 to 0.61 | 0.103 |
| | | 12695 | Childhood predictor | Poor | Excellent | 1.07 | 0.27 | 0.55 to 1.59 | 0.000 |
| | | 12695 | Age | Per 1-year increase | | -0.01 | 0.00 | -0.02 to 0.00 | 0.010 |
| | | 12695 | Gender | Female | Male | 0.33 | 0.09 | 0.16 to 0.50 | 0.000 |
| | | 12695 | Education | Secondary | Elementary or less | -0.51 | 0.10 | -0.72 to -0.31 | 0.000 |
| | | 12695 | Education | Four years beyond HS | Elementary or less | -0.76 | 0.17 | -1.08 to -0.43 | 0.000 |
| | | 12695 | Residential setting | Small town | Rural | 0.39 | 0.13 | 0.14 to 0.65 | 0.003 |
| | | 12695 | Residential setting | Large city | Rural | -0.07 | 0.18 | -0.42 to 0.27 | 0.680 |
| | | 12695 | Residential setting | Suburb | Rural | 0.22 | 0.25 | -0.28 to 0.71 | 0.389 |
| **Life worthwhile‡** | **Linear regression** | 12577 | *Intercept* | | | 6.77 | 0.16 | 6.45 to 7.09 | 0.000 |
| | | 12577 | Childhood predictor | Very good | Excellent | 0.04 | 0.11 | -0.18 to 0.26 | 0.734 |
| | | 12577 | Childhood predictor | Good | Excellent | -0.04 | 0.11 | -0.26 to 0.18 | 0.700 |
| | | 12577 | Childhood predictor | Fair | Excellent | -0.42 | 0.13 | -0.67 to -0.17 | 0.001 |
| | | 12577 | Childhood predictor | Poor | Excellent | -0.88 | 0.25 | -1.36 to -0.40 | 0.000 |
| | | 12577 | Age | Per 1-year increase | | -0.01 | 0.00 | -0.01 to 0.00 | 0.001 |
| | | 12577 | Gender | Female | Male | 0.45 | 0.07 | 0.31 to 0.59 | 0.000 |
| | | 12577 | Education | Secondary | Elementary or less | 0.44 | 0.08 | 0.29 to 0.60 | 0.000 |
| | | 12577 | Education | Four years beyond HS | Elementary or less | 0.60 | 0.12 | 0.36 to 0.84 | 0.000 |
| | | 12577 | Residential setting | Small town | Rural | 0.40 | 0.10 | 0.21 to 0.60 | 0.000 |
| | | 12577 | Residential setting | Large city | Rural | 0.39 | 0.12 | 0.14 to 0.63 | 0.002 |
| | | 12577 | Residential setting | Suburb | Rural | 0.32 | 0.20 | -0.08 to 0.72 | 0.121 |
| **Flourishing (with financial indicators)** | **Linear regression** | 12226 | *Intercept* | | | 7.11 | 0.10 | 6.92 to 7.30 | 0.000 |
| | | 12226 | Childhood predictor | Very good | Excellent | 0.01 | 0.06 | -0.12 to 0.13 | 0.903 |
| | | 12226 | Childhood predictor | Good | Excellent | -0.10 | 0.06 | -0.23 to 0.02 | 0.107 |
| | | 12226 | Childhood predictor | Fair | Excellent | -0.31 | 0.07 | -0.45 to -0.17 | 0.000 |
| | | 12226 | Childhood predictor | Poor | Excellent | -0.63 | 0.12 | -0.87 to -0.39 | 0.000 |

| Outcome | Model type | Analytic N | Variable | Level / comparison | Reference category | Coefficient | Standard error | 95% CI | p-value |
|---|---|---|---|---|---|---|---|---|---|
| | | 12226 | Age | Per 1-year increase | | -0.01 | 0.00 | -0.02 to -0.01 | 0.000 |
| | | 12226 | Gender | Female | Male | 0.10 | 0.04 | 0.02 to 0.18 | 0.018 |
| | | 12226 | Education | Secondary | Elementary or less | 0.32 | 0.04 | 0.23 to 0.41 | 0.000 |
| | | 12226 | Education | Four years beyond HS | Elementary or less | 0.50 | 0.07 | 0.37 to 0.63 | 0.000 |
| | | 12226 | Residential setting | Small town | Rural | 0.29 | 0.06 | 0.18 to 0.41 | 0.000 |
| | | 12226 | Residential setting | Large city | Rural | 0.32 | 0.07 | 0.19 to 0.45 | 0.000 |
| | | 12226 | Residential setting | Suburb | Rural | 0.08 | 0.13 | -0.17 to 0.33 | 0.521 |
| **Flourishing (without financial indicators)** | **Linear regression** | 12255 | *Intercept* | | | 7.69 | 0.10 | 7.48 to 7.89 | 0.000 |
| | | 12255 | Childhood predictor | Very good | Excellent | 0.04 | 0.07 | -0.09 to 0.17 | 0.575 |
| | | 12255 | Childhood predictor | Good | Excellent | -0.03 | 0.07 | -0.17 to 0.11 | 0.665 |
| | | 12255 | Childhood predictor | Fair | Excellent | -0.31 | 0.08 | -0.46 to -0.16 | 0.000 |
| | | 12255 | Childhood predictor | Poor | Excellent | -0.54 | 0.13 | -0.80 to -0.29 | 0.000 |
| | | 12255 | Age | Per 1-year increase | | -0.02 | 0.00 | -0.02 to -0.01 | 0.000 |
| | | 12255 | Gender | Female | Male | 0.18 | 0.05 | 0.10 to 0.27 | 0.000 |
| | | 12255 | Education | Secondary | Elementary or less | 0.29 | 0.05 | 0.20 to 0.39 | 0.000 |
| | | 12255 | Education | Four years beyond HS | Elementary or less | 0.47 | 0.07 | 0.33 to 0.60 | 0.000 |
| | | 12255 | Residential setting | Small town | Rural | 0.42 | 0.06 | 0.29 to 0.54 | 0.000 |
| | | 12255 | Residential setting | Large city | Rural | 0.36 | 0.08 | 0.21 to 0.51 | 0.000 |
| | | 12255 | Residential setting | Suburb | Rural | 0.10 | 0.13 | -0.15 to 0.36 | 0.431 |
| **Feel close to someone** | **Logistic regression** | 12688 | *Intercept* | | | 2.02 | 0.16 | 1.71 to 2.33 | 0.000 |
| | | 12688 | Childhood predictor | Very good | Excellent | -0.12 | 0.10 | -0.33 to 0.08 | 0.231 |
| | | 12688 | Childhood predictor | Good | Excellent | -0.43 | 0.11 | -0.63 to -0.22 | 0.000 |
| | | 12688 | Childhood predictor | Fair | Excellent | -0.51 | 0.11 | -0.73 to -0.29 | 0.000 |
| | | 12688 | Childhood predictor | Poor | Excellent | -0.63 | 0.17 | -0.95 to -0.30 | 0.000 |
| | | 12688 | Age | Per 1-year increase | | -0.01 | 0.00 | -0.01 to 0.00 | 0.003 |
| | | 12688 | Gender | Female | Male | 0.03 | 0.06 | -0.09 to 0.15 | 0.634 |
| | | 12688 | Education | Secondary | Elementary or less | 0.13 | 0.08 | -0.02 to 0.29 | 0.096 |
| | | 12688 | Education | Four years beyond HS | Elementary or less | 0.12 | 0.13 | -0.14 to 0.38 | 0.356 |
| | | 12688 | Residential setting | Small town | Rural | 0.05 | 0.11 | -0.16 to 0.27 | 0.634 |
| | | 12688 | Residential setting | Large city | Rural | -0.15 | 0.13 | -0.41 to 0.10 | 0.242 |
| | | 12688 | Residential setting | Suburb | Rural | -0.25 | 0.20 | -0.63 to 0.14 | 0.208 |
| **Donated to charity** | **Logistic regression** | 12656 | *Intercept* | | | -0.24 | 0.14 | -0.52 to 0.05 | 0.103 |
| | | 12656 | Childhood predictor | Very good | Excellent | 0.15 | 0.09 | -0.02 to 0.32 | 0.090 |
| | | 12656 | Childhood predictor | Good | Excellent | -0.31 | 0.09 | -0.50 to -0.13 | 0.001 |
| | | 12656 | Childhood predictor | Fair | Excellent | -0.15 | 0.11 | -0.37 to 0.06 | 0.169 |
| | | 12656 | Childhood predictor | Poor | Excellent | 0.23 | 0.14 | -0.04 to 0.50 | 0.095 |
| | | 12656 | Age | Per 1-year increase | | 0.00 | 0.00 | 0.00 to 0.00 | 0.791 |
| | | 12656 | Gender | Female | Male | -0.02 | 0.05 | -0.11 to 0.08 | 0.750 |
| | | 12656 | Education | Secondary | Elementary or less | -0.10 | 0.06 | -0.22 to 0.01 | 0.088 |
| | | 12656 | Education | Four years beyond HS | Elementary or less | -0.17 | 0.10 | -0.37 to 0.03 | 0.091 |

| Outcome | Model type | Analytic N | Variable | Level / comparison | Reference category | Coefficient | Standard error | 95% CI | p-value |
|---|---|---|---|---|---|---|---|---|---|
| | | 12656 | Residential setting | Small town | Rural | -0.09 | 0.09 | -0.27 to 0.09 | 0.325 |
| | | 12656 | Residential setting | Large city | Rural | -0.44 | 0.11 | -0.67 to -0.22 | 0.000 |
| | | 12656 | Residential setting | Suburb | Rural | -0.25 | 0.19 | -0.62 to 0.12 | 0.182 |
| **Have any health problems** | **Logistic regression** | 12613 | *Intercept* | | | -2.12 | 0.12 | -2.36 to -1.88 | 0.000 |
| | | 12613 | Childhood predictor | Very good | Excellent | -0.06 | 0.09 | -0.23 to 0.11 | 0.475 |
| | | 12613 | Childhood predictor | Good | Excellent | 0.06 | 0.08 | -0.10 to 0.23 | 0.447 |
| | | 12613 | Childhood predictor | Fair | Excellent | 0.22 | 0.09 | 0.04 to 0.39 | 0.016 |
| | | 12613 | Childhood predictor | Poor | Excellent | 0.41 | 0.14 | 0.13 to 0.69 | 0.004 |
| | | 12613 | Age | Per 1-year increase | | 0.03 | 0.00 | 0.02 to 0.03 | 0.000 |
| | | 12613 | Gender | Female | Male | 0.23 | 0.05 | 0.13 to 0.32 | 0.000 |
| | | 12613 | Education | Secondary | Elementary or less | -0.12 | 0.06 | -0.24 to -0.01 | 0.035 |
| | | 12613 | Education | Four years beyond HS | Elementary or less | -0.26 | 0.09 | -0.44 to -0.07 | 0.006 |
| | | 12613 | Residential setting | Small town | Rural | 0.14 | 0.06 | 0.02 to 0.27 | 0.027 |
| | | 12613 | Residential setting | Large city | Rural | -0.12 | 0.09 | -0.29 to 0.05 | 0.160 |
| | | 12613 | Residential setting | Suburb | Rural | 0.10 | 0.14 | -0.18 to 0.37 | 0.492 |
| **Helped a stranger** | **Logistic regression** | 12557 | *Intercept* | | | 0.95 | 0.14 | 0.69 to 1.22 | 0.000 |
| | | 12557 | Childhood predictor | Very good | Excellent | -0.22 | 0.09 | -0.39 to -0.05 | 0.010 |
| | | 12557 | Childhood predictor | Good | Excellent | -0.34 | 0.09 | -0.52 to -0.16 | 0.000 |
| | | 12557 | Childhood predictor | Fair | Excellent | -0.41 | 0.11 | -0.62 to -0.20 | 0.000 |
| | | 12557 | Childhood predictor | Poor | Excellent | -0.17 | 0.15 | -0.45 to 0.12 | 0.248 |
| | | 12557 | Age | Per 1-year increase | | -0.01 | 0.00 | -0.01 to 0.00 | 0.000 |
| | | 12557 | Gender | Female | Male | -0.31 | 0.05 | -0.40 to -0.22 | 0.000 |
| | | 12557 | Education | Secondary | Elementary or less | -0.09 | 0.06 | -0.20 to 0.02 | 0.114 |
| | | 12557 | Education | Four years beyond HS | Elementary or less | -0.13 | 0.09 | -0.30 to 0.05 | 0.156 |
| | | 12557 | Residential setting | Small town | Rural | 0.09 | 0.09 | -0.08 to 0.26 | 0.327 |
| | | 12557 | Residential setting | Large city | Rural | -0.18 | 0.10 | -0.37 to 0.02 | 0.079 |
| | | 12557 | Residential setting | Suburb | Rural | 0.00 | 0.18 | -0.35 to 0.36 | 0.984 |
| **Volunteered at an organization** | **Logistic regression** | 12637 | *Intercept* | | | -0.28 | 0.14 | -0.56 to -0.01 | 0.045 |
| | | 12637 | Childhood predictor | Very good | Excellent | 0.14 | 0.09 | -0.04 to 0.32 | 0.123 |
| | | 12637 | Childhood predictor | Good | Excellent | -0.24 | 0.10 | -0.44 to -0.05 | 0.014 |
| | | 12637 | Childhood predictor | Fair | Excellent | 0.01 | 0.11 | -0.21 to 0.23 | 0.912 |
| | | 12637 | Childhood predictor | Poor | Excellent | 0.17 | 0.15 | -0.13 to 0.47 | 0.266 |
| | | 12637 | Age | Per 1-year increase | | -0.01 | 0.00 | -0.01 to 0.00 | 0.001 |
| | | 12637 | Gender | Female | Male | -0.27 | 0.05 | -0.37 to -0.17 | 0.000 |
| | | 12637 | Education | Secondary | Elementary or less | -0.15 | 0.06 | -0.28 to -0.03 | 0.016 |
| | | 12637 | Education | Four years beyond HS | Elementary or less | 0.03 | 0.10 | -0.17 to 0.22 | 0.797 |
| | | 12637 | Residential setting | Small town | Rural | -0.10 | 0.11 | -0.32 to 0.11 | 0.330 |
| | | 12637 | Residential setting | Large city | Rural | -0.56 | 0.13 | -0.81 to -0.31 | 0.000 |
| | | 12637 | Residential setting | Suburb | Rural | -0.40 | 0.16 | -0.71 to -0.09 | 0.013 |
| **Cigarette smoking** | **Logistic regression** | 12199 | *Intercept* | | | -2.00 | 0.19 | -2.37 to -1.63 | 0.000 |

| Outcome | Model type | Analytic N | Variable | Level / comparison | Reference category | Coefficient | Standard error | 95% CI | p-value |
|---|---|---|---|---|---|---|---|---|---|
| | | 12199 | Childhood predictor | Very good | Excellent | 0.03 | 0.13 | -0.22 to 0.29 | 0.796 |
| | | 12199 | Childhood predictor | Good | Excellent | -0.06 | 0.12 | -0.29 to 0.18 | 0.632 |
| | | 12199 | Childhood predictor | Fair | Excellent | 0.15 | 0.14 | -0.12 to 0.42 | 0.274 |
| | | 12199 | Childhood predictor | Poor | Excellent | -0.44 | 0.30 | -1.03 to 0.16 | 0.149 |
| | | 12199 | Age | Per 1-year increase | | 0.01 | 0.00 | 0.00 to 0.01 | 0.012 |
| | | 12199 | Gender | Female | Male | -2.85 | 0.19 | -3.21 to -2.48 | 0.000 |
| | | 12199 | Education | Secondary | Elementary or less | -0.37 | 0.10 | -0.56 to -0.19 | 0.000 |
| | | 12199 | Education | Four years beyond HS | Elementary or less | -0.60 | 0.17 | -0.93 to -0.27 | 0.000 |
| | | 12199 | Residential setting | Small town | Rural | -0.07 | 0.10 | -0.27 to 0.13 | 0.495 |
| | | 12199 | Residential setting | Large city | Rural | 0.14 | 0.14 | -0.13 to 0.41 | 0.319 |
| | | 12199 | Residential setting | Suburb | Rural | 0.77 | 0.20 | 0.38 to 1.16 | 0.000 |
| **Alcohol use** | **Logistic regression** | 12184 | *Intercept* | | | -1.91 | 0.19 | -2.27 to -1.54 | 0.000 |
| | | 12184 | Childhood predictor | Very good | Excellent | -0.09 | 0.14 | -0.36 to 0.18 | 0.504 |
| | | 12184 | Childhood predictor | Good | Excellent | 0.01 | 0.14 | -0.26 to 0.28 | 0.960 |
| | | 12184 | Childhood predictor | Fair | Excellent | 0.14 | 0.16 | -0.17 to 0.44 | 0.380 |
| | | 12184 | Childhood predictor | Poor | Excellent | 0.16 | 0.25 | -0.32 to 0.65 | 0.508 |
| | | 12184 | Age | Per 1-year increase | | 0.00 | 0.00 | -0.01 to 0.01 | 0.933 |
| | | 12184 | Gender | Female | Male | -2.75 | 0.17 | -3.08 to -2.42 | 0.000 |
| | | 12184 | Education | Secondary | Elementary or less | -0.22 | 0.10 | -0.41 to -0.03 | 0.023 |
| | | 12184 | Education | Four years beyond HS | Elementary or less | -0.52 | 0.16 | -0.84 to -0.20 | 0.002 |
| | | 12184 | Residential setting | Small town | Rural | 0.22 | 0.10 | 0.01 to 0.42 | 0.039 |
| | | 12184 | Residential setting | Large city | Rural | 0.45 | 0.14 | 0.18 to 0.72 | 0.001 |
| | | 12184 | Residential setting | Suburb | Rural | 0.68 | 0.21 | 0.26 to 1.10 | 0.002 |
| **Government approval§** | **Proportional-odds regression** | 12473 | Childhood predictor | Very good | Excellent | 0.28 | 0.07 | 0.14 to 0.41 | 0.000 |
| | | 12473 | Childhood predictor | Good | Excellent | 0.17 | 0.07 | 0.03 to 0.30 | 0.019 |
| | | 12473 | Childhood predictor | Fair | Excellent | 0.06 | 0.08 | -0.09 to 0.22 | 0.415 |
| | | 12473 | Childhood predictor | Poor | Excellent | 0.01 | 0.13 | -0.25 to 0.26 | 0.949 |
| | | 12473 | Age | Per 1-year increase | | 0.00 | 0.00 | -0.01 to 0.00 | 0.036 |
| | | 12473 | Gender | Female | Male | 0.04 | 0.04 | -0.05 to 0.12 | 0.375 |
| | | 12473 | Education | Secondary | Elementary or less | -0.25 | 0.05 | -0.35 to -0.15 | 0.000 |
| | | 12473 | Education | Four years beyond HS | Elementary or less | -0.31 | 0.09 | -0.48 to -0.14 | 0.000 |
| | | 12473 | Residential setting | Small town | Rural | 0.10 | 0.07 | -0.03 to 0.23 | 0.135 |
| | | 12473 | Residential setting | Large city | Rural | -0.08 | 0.08 | -0.23 to 0.07 | 0.300 |
| | | 12473 | Residential setting | Suburb | Rural | -0.43 | 0.16 | -0.74 to -0.13 | 0.005 |
| | | 12473 | *Threshold / cut-point* | Strongly disapprove\|Somewhat disapprove | | -2.79 | 0.11 | -3.01 to -2.56 | 0.000 |
| | | 12473 | *Threshold / cut-point* | Somewhat disapprove\|Neither | | -2.12 | 0.11 | -2.33 to -1.90 | 0.000 |
| | | 12473 | *Threshold / cut-point* | Neither\|Somewhat approve | | -1.76 | 0.11 | -1.97 to -1.55 | 0.000 |
| | | 12473 | *Threshold / cut-point* | Somewhat approve\|Strongly approve | | -0.30 | 0.10 | -0.51 to -0.10 | 0.003 |

| Outcome | Model type | Analytic N | Variable | Level / comparison | Reference category | Coefficient | Standard error | 95% CI | p-value |
|---|---|---|---|---|---|---|---|---|---|
| **Feeling capable§** | **Proportional-odds regression** | 12637 | Childhood predictor | Very good | Excellent | -0.14 | 0.06 | -0.27 to -0.01 | 0.031 |
| | | 12637 | Childhood predictor | Good | Excellent | -0.17 | 0.07 | -0.29 to -0.04 | 0.011 |
| | | 12637 | Childhood predictor | Fair | Excellent | -0.22 | 0.07 | -0.36 to -0.08 | 0.002 |
| | | 12637 | Childhood predictor | Poor | Excellent | -0.07 | 0.12 | -0.30 to 0.16 | 0.566 |
| | | 12637 | Age | Per 1-year increase | | 0.00 | 0.00 | 0.00 to 0.01 | 0.226 |
| | | 12637 | Gender | Female | Male | -0.06 | 0.04 | -0.15 to 0.02 | 0.145 |
| | | 12637 | Education | Secondary | Elementary or less | 0.15 | 0.05 | 0.05 to 0.24 | 0.003 |
| | | 12637 | Education | Four years beyond HS | Elementary or less | 0.08 | 0.07 | -0.06 to 0.22 | 0.250 |
| | | 12637 | Residential setting | Small town | Rural | 0.15 | 0.05 | 0.05 to 0.25 | 0.003 |
| | | 12637 | Residential setting | Large city | Rural | 0.18 | 0.08 | 0.03 to 0.32 | 0.019 |
| | | 12637 | Residential setting | Suburb | Rural | 0.21 | 0.09 | 0.04 to 0.38 | 0.017 |
| | | 12637 | *Threshold / cut-point* | Never\|Rarely | | -2.50 | 0.10 | -2.70 to -2.30 | 0.000 |
| | | 12637 | *Threshold / cut-point* | Rarely\|Often | | -0.78 | 0.10 | -0.98 to -0.59 | 0.000 |
| | | 12637 | *Threshold / cut-point* | Often\|Always | | -0.12 | 0.10 | -0.31 to 0.07 | 0.219 |
| **Unable to control worry§** | **Proportional-odds regression** | 12588 | Childhood predictor | Very good | Excellent | -0.05 | 0.07 | -0.17 to 0.08 | 0.483 |
| | | 12588 | Childhood predictor | Good | Excellent | 0.09 | 0.06 | -0.03 to 0.22 | 0.130 |
| | | 12588 | Childhood predictor | Fair | Excellent | 0.10 | 0.07 | -0.03 to 0.23 | 0.133 |
| | | 12588 | Childhood predictor | Poor | Excellent | 0.36 | 0.13 | 0.11 to 0.61 | 0.005 |
| | | 12588 | Age | Per 1-year increase | | 0.00 | 0.00 | 0.00 to 0.01 | 0.001 |
| | | 12588 | Gender | Female | Male | 0.17 | 0.04 | 0.09 to 0.25 | 0.000 |
| | | 12588 | Education | Secondary | Elementary or less | -0.26 | 0.05 | -0.35 to -0.17 | 0.000 |
| | | 12588 | Education | Four years beyond HS | Elementary or less | -0.42 | 0.08 | -0.57 to -0.28 | 0.000 |
| | | 12588 | Residential setting | Small town | Rural | 0.06 | 0.05 | -0.04 to 0.15 | 0.244 |
| | | 12588 | Residential setting | Large city | Rural | 0.01 | 0.07 | -0.12 to 0.15 | 0.854 |
| | | 12588 | Residential setting | Suburb | Rural | -0.08 | 0.10 | -0.27 to 0.11 | 0.436 |
| | | 12588 | *Threshold / cut-point* | Not at all\|Several days | | 0.12 | 0.10 | -0.07 to 0.31 | 0.202 |
| | | 12588 | *Threshold / cut-point* | Several days\|More than half the days | | 1.03 | 0.09 | 0.85 to 1.22 | 0.000 |
| | | 12588 | *Threshold / cut-point* | More than half the days\|Nearly every day | | 1.88 | 0.10 | 1.69 to 2.07 | 0.000 |
| **Depressive symptoms§** | **Proportional-odds regression** | 12609 | Childhood predictor | Very good | Excellent | -0.02 | 0.07 | -0.17 to 0.12 | 0.746 |
| | | 12609 | Childhood predictor | Good | Excellent | 0.12 | 0.07 | -0.02 to 0.26 | 0.095 |
| | | 12609 | Childhood predictor | Fair | Excellent | 0.15 | 0.08 | 0.00 to 0.30 | 0.051 |
| | | 12609 | Childhood predictor | Poor | Excellent | 0.33 | 0.13 | 0.08 to 0.58 | 0.009 |
| | | 12609 | Age | Per 1-year increase | | 0.00 | 0.00 | 0.00 to 0.00 | 0.223 |
| | | 12609 | Gender | Female | Male | 0.09 | 0.04 | 0.01 to 0.17 | 0.020 |
| | | 12609 | Education | Secondary | Elementary or less | -0.20 | 0.05 | -0.29 to -0.11 | 0.000 |
| | | 12609 | Education | Four years beyond HS | Elementary or less | -0.40 | 0.07 | -0.53 to -0.27 | 0.000 |
| | | 12609 | Residential setting | Small town | Rural | 0.01 | 0.05 | -0.10 to 0.11 | 0.916 |

| Outcome | Model type | Analytic N | Variable | Level / comparison | Reference category | Coefficient | Standard error | 95% CI | p-value |
|---|---|---|---|---|---|---|---|---|---|
| | | 12609 | Residential setting | Large city | Rural | -0.07 | 0.07 | -0.21 to 0.07 | 0.340 |
| | | 12609 | Residential setting | Suburb | Rural | -0.16 | 0.11 | -0.38 to 0.06 | 0.152 |
| | | 12609 | *Threshold / cut-point* | Not at all\|Several days | | -0.18 | 0.10 | -0.37 to 0.01 | 0.062 |
| | | 12609 | *Threshold / cut-point* | Several days\|More than half the days | | 0.90 | 0.10 | 0.71 to 1.08 | 0.000 |
| | | 12609 | *Threshold / cut-point* | More than half the days\|Nearly every day | | 1.90 | 0.10 | 1.70 to 2.10 | 0.000 |
| **Perceived discrimination§** | **Proportional-odds regression** | 12540 | Childhood predictor | Very good | Excellent | 0.11 | 0.07 | -0.03 to 0.25 | 0.113 |
| | | 12540 | Childhood predictor | Good | Excellent | 0.10 | 0.07 | -0.05 to 0.24 | 0.185 |
| | | 12540 | Childhood predictor | Fair | Excellent | 0.16 | 0.08 | 0.01 to 0.31 | 0.037 |
| | | 12540 | Childhood predictor | Poor | Excellent | 0.17 | 0.13 | -0.08 to 0.43 | 0.180 |
| | | 12540 | Age | Per 1-year increase | | 0.00 | 0.00 | -0.01 to 0.00 | 0.040 |
| | | 12540 | Gender | Female | Male | -0.14 | 0.04 | -0.22 to -0.05 | 0.002 |
| | | 12540 | Education | Secondary | Elementary or less | -0.23 | 0.05 | -0.33 to -0.13 | 0.000 |
| | | 12540 | Education | Four years beyond HS | Elementary or less | -0.28 | 0.09 | -0.46 to -0.11 | 0.001 |
| | | 12540 | Residential setting | Small town | Rural | -0.20 | 0.06 | -0.31 to -0.09 | 0.001 |
| | | 12540 | Residential setting | Large city | Rural | -0.22 | 0.07 | -0.36 to -0.08 | 0.002 |
| | | 12540 | Residential setting | Suburb | Rural | -0.32 | 0.10 | -0.51 to -0.13 | 0.001 |
| | | 12540 | *Threshold / cut-point* | Never\|Rarely | | -0.03 | 0.11 | -0.24 to 0.18 | 0.758 |
| | | 12540 | *Threshold / cut-point* | Rarely\|Often | | 0.97 | 0.11 | 0.76 to 1.18 | 0.000 |
| | | 12540 | *Threshold / cut-point* | Often\|Always | | 1.59 | 0.11 | 1.37 to 1.81 | 0.000 |
| **Feeling nervous or anxious§** | **Proportional-odds regression** | 12639 | Childhood predictor | Very good | Excellent | 0.03 | 0.07 | -0.11 to 0.17 | 0.668 |
| | | 12639 | Childhood predictor | Good | Excellent | 0.20 | 0.07 | 0.06 to 0.34 | 0.004 |
| | | 12639 | Childhood predictor | Fair | Excellent | 0.17 | 0.08 | 0.02 to 0.32 | 0.030 |
| | | 12639 | Childhood predictor | Poor | Excellent | 0.64 | 0.12 | 0.41 to 0.88 | 0.000 |
| | | 12639 | Age | Per 1-year increase | | 0.01 | 0.00 | 0.00 to 0.01 | 0.000 |
| | | 12639 | Gender | Female | Male | 0.31 | 0.04 | 0.23 to 0.39 | 0.000 |
| | | 12639 | Education | Secondary | Elementary or less | -0.28 | 0.05 | -0.37 to -0.19 | 0.000 |
| | | 12639 | Education | Four years beyond HS | Elementary or less | -0.45 | 0.08 | -0.60 to -0.30 | 0.000 |
| | | 12639 | Residential setting | Small town | Rural | 0.12 | 0.05 | 0.01 to 0.22 | 0.031 |
| | | 12639 | Residential setting | Large city | Rural | 0.00 | 0.07 | -0.14 to 0.13 | 0.943 |
| | | 12639 | Residential setting | Suburb | Rural | -0.01 | 0.12 | -0.24 to 0.22 | 0.930 |
| | | 12639 | *Threshold / cut-point* | Not at all\|Several days | | 0.25 | 0.09 | 0.06 to 0.44 | 0.008 |
| | | 12639 | *Threshold / cut-point* | Several days\|More than half the days | | 1.26 | 0.10 | 1.07 to 1.44 | 0.000 |
| | | 12639 | *Threshold / cut-point* | More than half the days\|Nearly every day | | 2.19 | 0.10 | 2.00 to 2.38 | 0.000 |
| **Forgiveness§** | **Proportional-odds regression** | 12617 | Childhood predictor | Very good | Excellent | 0.15 | 0.07 | 0.01 to 0.29 | 0.034 |
| | | 12617 | Childhood predictor | Good | Excellent | 0.08 | 0.07 | -0.06 to 0.22 | 0.284 |

| Outcome | Model type | Analytic N | Variable | Level / comparison | Reference category | Coefficient | Standard error | 95% CI | p-value |
|---|---|---|---|---|---|---|---|---|---|
| | | 12617 | Childhood predictor | Fair | Excellent | 0.06 | 0.08 | -0.09 to 0.22 | 0.439 |
| | | 12617 | Childhood predictor | Poor | Excellent | 0.03 | 0.13 | -0.22 to 0.28 | 0.815 |
| | | 12617 | Age | Per 1-year increase | | 0.00 | 0.00 | 0.00 to 0.00 | 0.786 |
| | | 12617 | Gender | Female | Male | 0.08 | 0.04 | 0.00 to 0.17 | 0.049 |
| | | 12617 | Education | Secondary | Elementary or less | -0.08 | 0.05 | -0.18 to 0.03 | 0.150 |
| | | 12617 | Education | Four years beyond HS | Elementary or less | -0.19 | 0.08 | -0.34 to -0.04 | 0.015 |
| | | 12617 | Residential setting | Small town | Rural | 0.05 | 0.05 | -0.06 to 0.15 | 0.354 |
| | | 12617 | Residential setting | Large city | Rural | 0.03 | 0.08 | -0.13 to 0.19 | 0.713 |
| | | 12617 | Residential setting | Suburb | Rural | 0.05 | 0.12 | -0.18 to 0.28 | 0.683 |
| | | 12617 | *Threshold / cut-point* | Never\|Rarely | | -2.30 | 0.11 | -2.52 to -2.09 | 0.000 |
| | | 12617 | *Threshold / cut-point* | Rarely\|Often | | -0.97 | 0.10 | -1.17 to -0.76 | 0.000 |
| | | 12617 | *Threshold / cut-point* | Often\|Always | | -0.32 | 0.10 | -0.52 to -0.12 | 0.002 |
| **Support reducing differences in income levels§** | **Proportional-odds regression** | 12472 | Childhood predictor | Very good | Excellent | -0.12 | 0.11 | -0.33 to 0.09 | 0.265 |
| | | 12472 | Childhood predictor | Good | Excellent | -0.23 | 0.12 | -0.47 to 0.00 | 0.050 |
| | | 12472 | Childhood predictor | Fair | Excellent | -0.17 | 0.13 | -0.42 to 0.07 | 0.170 |
| | | 12472 | Childhood predictor | Poor | Excellent | -0.30 | 0.21 | -0.71 to 0.10 | 0.142 |
| | | 12472 | Age | Per 1-year increase | | 0.00 | 0.00 | -0.01 to 0.00 | 0.084 |
| | | 12472 | Gender | Female | Male | -0.04 | 0.06 | -0.15 to 0.07 | 0.463 |
| | | 12472 | Education | Secondary | Elementary or less | -0.08 | 0.07 | -0.21 to 0.05 | 0.245 |
| | | 12472 | Education | Four years beyond HS | Elementary or less | -0.30 | 0.11 | -0.51 to -0.09 | 0.006 |
| | | 12472 | Residential setting | Small town | Rural | 0.04 | 0.13 | -0.21 to 0.29 | 0.748 |
| | | 12472 | Residential setting | Large city | Rural | 0.09 | 0.15 | -0.20 to 0.38 | 0.539 |
| | | 12472 | Residential setting | Suburb | Rural | -0.11 | 0.18 | -0.46 to 0.24 | 0.544 |
| | | 12472 | *Threshold / cut-point* | Strongly disagree\|Somewhat disagree | | -4.02 | 0.20 | -4.42 to -3.61 | 0.000 |
| | | 12472 | *Threshold / cut-point* | Somewhat disagree\|Neither | | -3.47 | 0.20 | -3.85 to -3.09 | 0.000 |
| | | 12472 | *Threshold / cut-point* | Neither\|Somewhat agree | | -3.02 | 0.19 | -3.40 to -2.65 | 0.000 |
| | | 12472 | *Threshold / cut-point* | Somewhat agree\|Strongly agree | | -1.46 | 0.17 | -1.79 to -1.13 | 0.000 |
| **Have little interest in doing things§** | **Proportional-odds regression** | 12602 | Childhood predictor | Very good | Excellent | -0.19 | 0.07 | -0.32 to -0.05 | 0.006 |
| | | 12602 | Childhood predictor | Good | Excellent | -0.17 | 0.07 | -0.30 to -0.04 | 0.009 |
| | | 12602 | Childhood predictor | Fair | Excellent | -0.30 | 0.08 | -0.45 to -0.15 | 0.000 |
| | | 12602 | Childhood predictor | Poor | Excellent | -0.19 | 0.11 | -0.41 to 0.03 | 0.088 |
| | | 12602 | Age | Per 1-year increase | | 0.00 | 0.00 | 0.00 to 0.01 | 0.025 |
| | | 12602 | Gender | Female | Male | 0.03 | 0.04 | -0.05 to 0.11 | 0.475 |
| | | 12602 | Education | Secondary | Elementary or less | 0.06 | 0.05 | -0.03 to 0.15 | 0.213 |
| | | 12602 | Education | Four years beyond HS | Elementary or less | 0.08 | 0.08 | -0.08 to 0.23 | 0.323 |
| | | 12602 | Residential setting | Small town | Rural | 0.26 | 0.06 | 0.13 to 0.38 | 0.000 |
| | | 12602 | Residential setting | Large city | Rural | 0.09 | 0.08 | -0.06 to 0.25 | 0.251 |
| | | 12602 | Residential setting | Suburb | Rural | 0.24 | 0.15 | -0.04 to 0.53 | 0.092 |

| Outcome | Model type | Analytic N | Variable | Level / comparison | Reference category | Coefficient | Standard error | 95% CI | p-value |
|---|---|---|---|---|---|---|---|---|---|
| | | 12602 | *Threshold / cut-point* | Not at all\|Several days | | -0.67 | 0.10 | -0.87 to -0.46 | 0.000 |
| | | 12602 | *Threshold / cut-point* | Several days\|More than half the days | | 0.26 | 0.10 | 0.06 to 0.47 | 0.012 |
| | | 12602 | *Threshold / cut-point* | More than half the days\|Nearly every day | | 1.04 | 0.11 | 0.83 to 1.25 | 0.000 |
| **Various aspects of life are balanced§** | **Proportional-odds regression** | 12529 | Childhood predictor | Very good | Excellent | -0.04 | 0.06 | -0.17 to 0.08 | 0.503 |
| | | 12529 | Childhood predictor | Good | Excellent | -0.13 | 0.07 | -0.26 to 0.00 | 0.051 |
| | | 12529 | Childhood predictor | Fair | Excellent | -0.19 | 0.07 | -0.33 to -0.05 | 0.007 |
| | | 12529 | Childhood predictor | Poor | Excellent | -0.22 | 0.12 | -0.45 to 0.01 | 0.063 |
| | | 12529 | Age | Per 1-year increase | | 0.00 | 0.00 | 0.00 to 0.01 | 0.085 |
| | | 12529 | Gender | Female | Male | -0.03 | 0.04 | -0.11 to 0.05 | 0.459 |
| | | 12529 | Education | Secondary | Elementary or less | 0.03 | 0.04 | -0.05 to 0.12 | 0.433 |
| | | 12529 | Education | Four years beyond HS | Elementary or less | 0.09 | 0.07 | -0.04 to 0.22 | 0.184 |
| | | 12529 | Residential setting | Small town | Rural | 0.09 | 0.05 | -0.02 to 0.19 | 0.097 |
| | | 12529 | Residential setting | Large city | Rural | 0.07 | 0.07 | -0.06 to 0.20 | 0.265 |
| | | 12529 | Residential setting | Suburb | Rural | 0.33 | 0.11 | 0.12 to 0.55 | 0.002 |
| | | 12529 | *Threshold / cut-point* | Never\|Rarely | | -2.14 | 0.10 | -2.34 to -1.94 | 0.000 |
| | | 12529 | *Threshold / cut-point* | Rarely\|Often | | 0.04 | 0.10 | -0.15 to 0.23 | 0.680 |
| | | 12529 | *Threshold / cut-point* | Often\|Always | | 0.83 | 0.10 | 0.63 to 1.02 | 0.000 |
| **Obeying the law§** | **Proportional-odds regression** | 12658 | Childhood predictor | Very good | Excellent | -0.29 | 0.13 | -0.55 to -0.04 | 0.025 |
| | | 12658 | Childhood predictor | Good | Excellent | -0.39 | 0.13 | -0.66 to -0.13 | 0.004 |
| | | 12658 | Childhood predictor | Fair | Excellent | -0.37 | 0.14 | -0.64 to -0.09 | 0.009 |
| | | 12658 | Childhood predictor | Poor | Excellent | -0.49 | 0.21 | -0.90 to -0.09 | 0.017 |
| | | 12658 | Age | Per 1-year increase | | 0.00 | 0.00 | -0.01 to 0.00 | 0.461 |
| | | 12658 | Gender | Female | Male | -0.15 | 0.07 | -0.30 to -0.01 | 0.037 |
| | | 12658 | Education | Secondary | Elementary or less | 0.08 | 0.10 | -0.11 to 0.27 | 0.406 |
| | | 12658 | Education | Four years beyond HS | Elementary or less | -0.24 | 0.12 | -0.48 to 0.00 | 0.052 |
| | | 12658 | Residential setting | Small town | Rural | -0.06 | 0.15 | -0.35 to 0.23 | 0.680 |
| | | 12658 | Residential setting | Large city | Rural | -0.24 | 0.18 | -0.60 to 0.12 | 0.188 |
| | | 12658 | Residential setting | Suburb | Rural | -0.31 | 0.22 | -0.75 to 0.13 | 0.169 |
| | | 12658 | *Threshold / cut-point* | Strongly disagree\|Somewhat disagree | | -5.47 | 0.27 | -6.00 to -4.95 | 0.000 |
| | | 12658 | *Threshold / cut-point* | Somewhat disagree\|Neither | | -4.85 | 0.24 | -5.32 to -4.38 | 0.000 |
| | | 12658 | *Threshold / cut-point* | Neither\|Somewhat agree | | -4.34 | 0.24 | -4.81 to -3.86 | 0.000 |
| | | 12658 | *Threshold / cut-point* | Somewhat agree\|Strongly agree | | -2.51 | 0.21 | -2.92 to -2.10 | 0.000 |
| **Feel at peace§** | **Proportional-odds regression** | 12601 | Childhood predictor | Very good | Excellent | -0.16 | 0.07 | -0.29 to -0.03 | 0.016 |
| | | 12601 | Childhood predictor | Good | Excellent | -0.13 | 0.07 | -0.26 to 0.00 | 0.056 |
| | | 12601 | Childhood predictor | Fair | Excellent | -0.35 | 0.08 | -0.49 to -0.20 | 0.000 |

| Outcome | Model type | Analytic N | Variable | Level / comparison | Reference category | Coefficient | Standard error | 95% CI | p-value |
|---|---|---|---|---|---|---|---|---|---|
| | | 12601 | Childhood predictor | Poor | Excellent | -0.35 | 0.12 | -0.58 to -0.12 | 0.002 |
| | | 12601 | Age | Per 1-year increase | | 0.01 | 0.00 | 0.00 to 0.01 | 0.000 |
| | | 12601 | Gender | Female | Male | 0.01 | 0.04 | -0.06 to 0.09 | 0.757 |
| | | 12601 | Education | Secondary | Elementary or less | 0.01 | 0.05 | -0.09 to 0.10 | 0.901 |
| | | 12601 | Education | Four years beyond HS | Elementary or less | 0.03 | 0.06 | -0.09 to 0.16 | 0.617 |
| | | 12601 | Residential setting | Small town | Rural | 0.17 | 0.05 | 0.07 to 0.27 | 0.001 |
| | | 12601 | Residential setting | Large city | Rural | 0.12 | 0.07 | -0.02 to 0.27 | 0.085 |
| | | 12601 | Residential setting | Suburb | Rural | 0.22 | 0.10 | 0.02 to 0.41 | 0.030 |
| | | 12601 | *Threshold / cut-point* | Never\|Rarely | | -2.06 | 0.10 | -2.25 to -1.87 | 0.000 |
| | | 12601 | *Threshold / cut-point* | Rarely\|Often | | -0.23 | 0.09 | -0.41 to -0.05 | 0.014 |
| | | 12601 | *Threshold / cut-point* | Often\|Always | | 0.46 | 0.09 | 0.28 to 0.65 | 0.000 |
| **Say in government#** | **Proportional-odds regression** | 12640 | Childhood predictor | Very good | Excellent | 0.11 | 0.08 | -0.05 to 0.26 | 0.182 |
| | | 12640 | Childhood predictor | Good | Excellent | 0.20 | 0.08 | 0.04 to 0.35 | 0.013 |
| | | 12640 | Childhood predictor | Fair | Excellent | 0.13 | 0.09 | -0.04 to 0.31 | 0.132 |
| | | 12640 | Childhood predictor | Poor | Excellent | 0.12 | 0.17 | -0.20 to 0.45 | 0.453 |
| | | 12640 | Age | Per 1-year increase | | 0.00 | 0.00 | -0.01 to 0.00 | 0.385 |
| | | 12640 | Gender | Female | Male | 0.10 | 0.05 | 0.00 to 0.19 | 0.053 |
| | | 12640 | Education | Secondary | Elementary or less | -0.22 | 0.06 | -0.34 to -0.10 | 0.000 |
| | | 12640 | Education | Four years beyond HS | Elementary or less | -0.09 | 0.10 | -0.29 to 0.10 | 0.348 |
| | | 12640 | Residential setting | Small town | Rural | 0.02 | 0.08 | -0.13 to 0.17 | 0.773 |
| | | 12640 | Residential setting | Large city | Rural | -0.15 | 0.09 | -0.33 to 0.03 | 0.104 |
| | | 12640 | Residential setting | Suburb | Rural | -0.42 | 0.13 | -0.68 to -0.17 | 0.001 |
| | | 12640 | *Threshold / cut-point* | Disagree\|Unsure | | -1.66 | 0.13 | -1.91 to -1.41 | 0.000 |
| | | 12640 | *Threshold / cut-point* | Unsure\|Agree | | -1.26 | 0.12 | -1.51 to -1.02 | 0.000 |
| **Extent of suffering§** | **Proportional-odds regression** | 12616 | Childhood predictor | Very good | Excellent | -0.05 | 0.07 | -0.20 to 0.09 | 0.480 |
| | | 12616 | Childhood predictor | Good | Excellent | 0.17 | 0.07 | 0.03 to 0.31 | 0.015 |
| | | 12616 | Childhood predictor | Fair | Excellent | 0.26 | 0.07 | 0.12 to 0.41 | 0.000 |
| | | 12616 | Childhood predictor | Poor | Excellent | 0.53 | 0.13 | 0.28 to 0.78 | 0.000 |
| | | 12616 | Age | Per 1-year increase | | 0.02 | 0.00 | 0.01 to 0.02 | 0.000 |
| | | 12616 | Gender | Female | Male | 0.28 | 0.04 | 0.20 to 0.36 | 0.000 |
| | | 12616 | Education | Secondary | Elementary or less | -0.13 | 0.05 | -0.22 to -0.04 | 0.005 |
| | | 12616 | Education | Four years beyond HS | Elementary or less | -0.25 | 0.08 | -0.40 to -0.10 | 0.001 |
| | | 12616 | Residential setting | Small town | Rural | 0.06 | 0.06 | -0.05 to 0.18 | 0.257 |
| | | 12616 | Residential setting | Large city | Rural | -0.10 | 0.08 | -0.25 to 0.05 | 0.188 |
| | | 12616 | Residential setting | Suburb | Rural | 0.11 | 0.11 | -0.10 to 0.32 | 0.321 |
| | | 12616 | *Threshold / cut-point* | Not at all\|Not very much | | 0.41 | 0.10 | 0.22 to 0.61 | 0.000 |
| | | 12616 | *Threshold / cut-point* | Not very much\|Some | | 0.83 | 0.10 | 0.63 to 1.03 | 0.000 |
| | | 12616 | *Threshold / cut-point* | Some\|A lot | | 2.27 | 0.10 | 2.06 to 2.47 | 0.000 |

| Outcome | Model type | Analytic N | Variable | Level / comparison | Reference category | Coefficient | Standard error | 95% CI | p-value |
|---|---|---|---|---|---|---|---|---|---|
| **Distress from life-threatening experiences§** | **Proportional-odds regression** | 12634 | Childhood predictor | Very good | Excellent | -0.04 | 0.06 | -0.15 to 0.08 | 0.553 |
| | | 12634 | Childhood predictor | Good | Excellent | 0.07 | 0.06 | -0.05 to 0.19 | 0.270 |
| | | 12634 | Childhood predictor | Fair | Excellent | 0.13 | 0.07 | -0.01 to 0.27 | 0.060 |
| | | 12634 | Childhood predictor | Poor | Excellent | 0.32 | 0.12 | 0.09 to 0.56 | 0.007 |
| | | 12634 | Age | Per 1-year increase | | 0.00 | 0.00 | 0.00 to 0.01 | 0.016 |
| | | 12634 | Gender | Female | Male | 0.08 | 0.04 | -0.01 to 0.17 | 0.066 |
| | | 12634 | Education | Secondary | Elementary or less | -0.25 | 0.05 | -0.34 to -0.16 | 0.000 |
| | | 12634 | Education | Four years beyond HS | Elementary or less | -0.44 | 0.07 | -0.58 to -0.29 | 0.000 |
| | | 12634 | Residential setting | Small town | Rural | -0.16 | 0.05 | -0.26 to -0.07 | 0.001 |
| | | 12634 | Residential setting | Large city | Rural | -0.22 | 0.07 | -0.36 to -0.08 | 0.002 |
| | | 12634 | Residential setting | Suburb | Rural | -0.18 | 0.12 | -0.41 to 0.05 | 0.127 |
| | | 12634 | *Threshold / cut-point* | Not at all\|Not very much | | -0.24 | 0.09 | -0.42 to -0.06 | 0.008 |
| | | 12634 | *Threshold / cut-point* | Not very much\|Some | | 0.10 | 0.09 | -0.09 to 0.28 | 0.297 |
| | | 12634 | *Threshold / cut-point* | Some\|A lot | | 1.11 | 0.09 | 0.93 to 1.29 | 0.000 |
| **Trust in others§** | **Proportional-odds regression** | 12347 | Childhood predictor | Very good | Excellent | -0.06 | 0.07 | -0.19 to 0.07 | 0.366 |
| | | 12347 | Childhood predictor | Good | Excellent | -0.04 | 0.07 | -0.17 to 0.10 | 0.569 |
| | | 12347 | Childhood predictor | Fair | Excellent | -0.24 | 0.08 | -0.39 to -0.09 | 0.001 |
| | | 12347 | Childhood predictor | Poor | Excellent | -0.14 | 0.11 | -0.35 to 0.07 | 0.200 |
| | | 12347 | Age | Per 1-year increase | | 0.00 | 0.00 | 0.00 to 0.01 | 0.016 |
| | | 12347 | Gender | Female | Male | -0.04 | 0.04 | -0.12 to 0.03 | 0.279 |
| | | 12347 | Education | Secondary | Elementary or less | -0.05 | 0.05 | -0.15 to 0.04 | 0.239 |
| | | 12347 | Education | Four years beyond HS | Elementary or less | -0.08 | 0.07 | -0.22 to 0.06 | 0.260 |
| | | 12347 | Residential setting | Small town | Rural | 0.23 | 0.05 | 0.13 to 0.33 | 0.000 |
| | | 12347 | Residential setting | Large city | Rural | 0.00 | 0.07 | -0.14 to 0.13 | 0.947 |
| | | 12347 | Residential setting | Suburb | Rural | -0.19 | 0.14 | -0.46 to 0.09 | 0.180 |
| | | 12347 | *Threshold / cut-point* | None\|Not very many | | -2.22 | 0.11 | -2.43 to -2.01 | 0.000 |
| | | 12347 | *Threshold / cut-point* | Not very many\|Some | | -1.43 | 0.10 | -1.64 to -1.23 | 0.000 |
| | | 12347 | *Threshold / cut-point* | Some\|Most | | 0.76 | 0.10 | 0.57 to 0.96 | 0.000 |
| | | 12347 | *Threshold / cut-point* | Most\|All | | 1.74 | 0.10 | 1.53 to 1.94 | 0.000 |

**eTable 14.** Table of coefficients for adjusted associations between finding it very difficult in perceived family financial situation as predictor (comfortable perceived family financial situation reference category) and 45 outcomes of interest and two flourishing composite indices.

Adjusted survey-weighted models use ANNUAL_WEIGHT_C1 with primary sampling unit (PSU) and strata and adjust for age, gender, education, and residential setting. Reference categories are shown in the table. Coefficients are on the original regression-coefficient scale: beta for continuous outcomes, log-odds for binary outcomes, and proportional-odds log coefficients for ordinal outcomes. Model intercepts are displayed for continuous and binary outcomes. Proportional-odds models do not have a single conventional intercept; their estimated ordinal cut-points/thresholds are displayed after the covariate coefficients, with standard errors (SEs), 95% CIs, and Wald p-values.

***Note:*** All results except p-values are rounded to the two decimals. P-values are rounded to three decimals. Values displayed as 0.000 indicate $P < 0.0005$ due to rounding, rather than an exact zero. Statistical significance was determined using unrounded P-values at the Bonferroni-adjusted threshold of $P < 0.0011$.

| Outcome | Model type | Analytic N | Variable | Level / comparison | Reference category | Coefficient | Standard error | 95% CI | p-value |
|---|---|---|---|---|---|---|---|---|---|
| **Belonging** | **Linear regression** | 12590 | *Intercept* | | | 8.81 | 0.12 | 8.58 to 9.04 | 0.000 |
| | | 12590 | Childhood predictor | Got by | Lived comfortably | -0.08 | 0.08 | -0.24 to 0.07 | 0.294 |
| | | 12590 | Childhood predictor | Found it difficult | Lived comfortably | -0.17 | 0.09 | -0.35 to 0.01 | 0.066 |
| | | 12590 | Childhood predictor | Found it very difficult | Lived comfortably | -0.51 | 0.10 | -0.71 to -0.31 | 0.000 |
| | | 12590 | Age | Per 1-year increase | | -0.01 | 0.00 | -0.02 to -0.01 | 0.000 |
| | | 12590 | Gender | Female | Male | -0.20 | 0.06 | -0.32 to -0.08 | 0.001 |
| | | 12590 | Education | Secondary | Elementary or less | 0.17 | 0.07 | 0.02 to 0.31 | 0.024 |
| | | 12590 | Education | Four years beyond HS | Elementary or less | 0.25 | 0.10 | 0.05 to 0.46 | 0.013 |
| | | 12590 | Residential setting | Small town | Rural | 0.35 | 0.08 | 0.20 to 0.50 | 0.000 |
| | | 12590 | Residential setting | Large city | Rural | 0.38 | 0.10 | 0.18 to 0.58 | 0.000 |
| | | 12590 | Residential setting | Suburb | Rural | 0.03 | 0.17 | -0.31 to 0.37 | 0.869 |
| **Relationship contentment‡** | **Linear regression** | 12653 | *Intercept* | | | 8.25 | 0.13 | 7.99 to 8.50 | 0.000 |
| | | 12653 | Childhood predictor | Got by | Lived comfortably | -0.03 | 0.09 | -0.20 to 0.14 | 0.747 |
| | | 12653 | Childhood predictor | Found it difficult | Lived comfortably | -0.22 | 0.09 | -0.40 to -0.03 | 0.022 |
| | | 12653 | Childhood predictor | Found it very difficult | Lived comfortably | -0.54 | 0.11 | -0.75 to -0.32 | 0.000 |
| | | 12653 | Age | Per 1-year increase | | -0.01 | 0.00 | -0.02 to -0.01 | 0.000 |
| | | 12653 | Gender | Female | Male | 0.35 | 0.07 | 0.22 to 0.49 | 0.000 |
| | | 12653 | Education | Secondary | Elementary or less | 0.10 | 0.07 | -0.04 to 0.25 | 0.158 |
| | | 12653 | Education | Four years beyond HS | Elementary or less | 0.23 | 0.12 | 0.00 to 0.46 | 0.048 |
| | | 12653 | Residential setting | Small town | Rural | 0.35 | 0.08 | 0.19 to 0.51 | 0.000 |
| | | 12653 | Residential setting | Large city | Rural | 0.04 | 0.10 | -0.15 to 0.23 | 0.672 |
| | | 12653 | Residential setting | Suburb | Rural | -0.27 | 0.17 | -0.61 to 0.06 | 0.106 |
| **Positive expectations** | **Linear regression** | 12615 | *Intercept* | | | 7.73 | 0.15 | 7.44 to 8.02 | 0.000 |
| | | 12615 | Childhood predictor | Got by | Lived comfortably | 0.14 | 0.08 | -0.01 to 0.30 | 0.069 |
| | | 12615 | Childhood predictor | Found it difficult | Lived comfortably | 0.00 | 0.09 | -0.18 to 0.18 | 0.993 |
| | | 12615 | Childhood predictor | Found it very difficult | Lived comfortably | 0.00 | 0.10 | -0.20 to 0.20 | 0.997 |
| | | 12615 | Age | Per 1-year increase | | 0.00 | 0.00 | -0.01 to 0.00 | 0.236 |
| | | 12615 | Gender | Female | Male | 0.23 | 0.06 | 0.10 to 0.35 | 0.000 |
| | | 12615 | Education | Secondary | Elementary or less | 0.27 | 0.07 | 0.13 to 0.42 | 0.000 |
| | | 12615 | Education | Four years beyond HS | Elementary or less | 0.36 | 0.10 | 0.16 to 0.55 | 0.000 |
| | | 12615 | Residential setting | Small town | Rural | 0.46 | 0.09 | 0.29 to 0.63 | 0.000 |

| Outcome | Model type | Analytic N | Variable | Level / comparison | Reference category | Coefficient | Standard error | 95% CI | p-value |
|---|---|---|---|---|---|---|---|---|---|
| | | 12615 | Residential setting | Large city | Rural | 0.47 | 0.11 | 0.25 to 0.68 | 0.000 |
| | | 12615 | Residential setting | Suburb | Rural | 0.47 | 0.16 | 0.15 to 0.79 | 0.004 |
| **Worry about meeting normal monthly living expenses§§‡‡** | **Linear regression** | 12655 | *Intercept* | | | 5.42 | 0.20 | 5.03 to 5.81 | 0.000 |
| | | 12655 | Childhood predictor | Got by | Lived comfortably | 0.54 | 0.12 | 0.31 to 0.77 | 0.000 |
| | | 12655 | Childhood predictor | Found it difficult | Lived comfortably | 0.60 | 0.13 | 0.34 to 0.85 | 0.000 |
| | | 12655 | Childhood predictor | Found it very difficult | Lived comfortably | 0.49 | 0.15 | 0.19 to 0.79 | 0.001 |
| | | 12655 | Age | Per 1-year increase | | 0.00 | 0.00 | -0.01 to 0.01 | 0.799 |
| | | 12655 | Gender | Female | Male | 0.42 | 0.09 | 0.24 to 0.59 | 0.000 |
| | | 12655 | Education | Secondary | Elementary or less | -0.41 | 0.11 | -0.63 to -0.20 | 0.000 |
| | | 12655 | Education | Four years beyond HS | Elementary or less | -0.59 | 0.15 | -0.89 to -0.30 | 0.000 |
| | | 12655 | Residential setting | Small town | Rural | 0.20 | 0.13 | -0.05 to 0.45 | 0.117 |
| | | 12655 | Residential setting | Large city | Rural | -0.19 | 0.15 | -0.49 to 0.10 | 0.204 |
| | | 12655 | Residential setting | Suburb | Rural | -0.22 | 0.25 | -0.71 to 0.26 | 0.367 |
| **Freedom** | **Linear regression** | 12628 | *Intercept* | | | 8.29 | 0.13 | 8.04 to 8.54 | 0.000 |
| | | 12628 | Childhood predictor | Got by | Lived comfortably | 0.02 | 0.08 | -0.14 to 0.18 | 0.844 |
| | | 12628 | Childhood predictor | Found it difficult | Lived comfortably | -0.11 | 0.09 | -0.28 to 0.06 | 0.219 |
| | | 12628 | Childhood predictor | Found it very difficult | Lived comfortably | -0.45 | 0.10 | -0.66 to -0.25 | 0.000 |
| | | 12628 | Age | Per 1-year increase | | -0.01 | 0.00 | -0.01 to 0.00 | 0.004 |
| | | 12628 | Gender | Female | Male | -0.14 | 0.07 | -0.27 to -0.01 | 0.036 |
| | | 12628 | Education | Secondary | Elementary or less | 0.26 | 0.07 | 0.12 to 0.41 | 0.000 |
| | | 12628 | Education | Four years beyond HS | Elementary or less | 0.36 | 0.10 | 0.15 to 0.56 | 0.001 |
| | | 12628 | Residential setting | Small town | Rural | 0.33 | 0.07 | 0.19 to 0.48 | 0.000 |
| | | 12628 | Residential setting | Large city | Rural | 0.36 | 0.10 | 0.16 to 0.56 | 0.000 |
| | | 12628 | Residential setting | Suburb | Rural | 0.14 | 0.15 | -0.15 to 0.43 | 0.354 |
| **Give up some happiness now for greater happiness later‡** | **Linear regression** | 12594 | *Intercept* | | | 7.33 | 0.16 | 7.02 to 7.64 | 0.000 |
| | | 12594 | Childhood predictor | Got by | Lived comfortably | -0.16 | 0.10 | -0.36 to 0.04 | 0.121 |
| | | 12594 | Childhood predictor | Found it difficult | Lived comfortably | -0.14 | 0.10 | -0.35 to 0.06 | 0.164 |
| | | 12594 | Childhood predictor | Found it very difficult | Lived comfortably | -0.33 | 0.12 | -0.57 to -0.10 | 0.006 |
| | | 12594 | Age | Per 1-year increase | | -0.01 | 0.00 | -0.01 to 0.00 | 0.002 |
| | | 12594 | Gender | Female | Male | 0.17 | 0.08 | 0.01 to 0.32 | 0.033 |
| | | 12594 | Education | Secondary | Elementary or less | 0.15 | 0.10 | -0.04 to 0.34 | 0.116 |
| | | 12594 | Education | Four years beyond HS | Elementary or less | 0.14 | 0.13 | -0.12 to 0.40 | 0.281 |
| | | 12594 | Residential setting | Small town | Rural | 0.39 | 0.10 | 0.20 to 0.58 | 0.000 |
| | | 12594 | Residential setting | Large city | Rural | 0.18 | 0.13 | -0.08 to 0.44 | 0.177 |
| | | 12594 | Residential setting | Suburb | Rural | 0.13 | 0.27 | -0.40 to 0.66 | 0.633 |
| **Have a long list of gratitude** | **Linear regression** | 12483 | *Intercept* | | | 7.39 | 0.15 | 7.09 to 7.69 | 0.000 |
| | | 12483 | Childhood predictor | Got by | Lived comfortably | 0.23 | 0.09 | 0.06 to 0.40 | 0.008 |
| | | 12483 | Childhood predictor | Found it difficult | Lived comfortably | -0.09 | 0.10 | -0.28 to 0.10 | 0.354 |
| | | 12483 | Childhood predictor | Found it very difficult | Lived comfortably | -0.07 | 0.10 | -0.27 to 0.13 | 0.496 |

| Outcome | Model type | Analytic N | Variable | Level / comparison | Reference category | Coefficient | Standard error | 95% CI | p-value |
|---|---|---|---|---|---|---|---|---|---|
| | | 12483 | Age | Per 1-year increase | | -0.01 | 0.00 | -0.01 to 0.00 | 0.073 |
| | | 12483 | Gender | Female | Male | 0.21 | 0.07 | 0.08 to 0.35 | 0.002 |
| | | 12483 | Education | Secondary | Elementary or less | 0.27 | 0.08 | 0.11 to 0.42 | 0.001 |
| | | 12483 | Education | Four years beyond HS | Elementary or less | 0.35 | 0.11 | 0.13 to 0.58 | 0.002 |
| | | 12483 | Residential setting | Small town | Rural | 0.43 | 0.09 | 0.25 to 0.61 | 0.000 |
| | | 12483 | Residential setting | Large city | Rural | 0.44 | 0.13 | 0.18 to 0.70 | 0.001 |
| | | 12483 | Residential setting | Suburb | Rural | 0.52 | 0.17 | 0.19 to 0.85 | 0.002 |
| **Happiness‡** | **Linear regression** | 12653 | *Intercept* | | | 6.97 | 0.16 | 6.66 to 7.28 | 0.000 |
| | | 12653 | Childhood predictor | Got by | Lived comfortably | -0.28 | 0.11 | -0.49 to -0.07 | 0.009 |
| | | 12653 | Childhood predictor | Found it difficult | Lived comfortably | -0.64 | 0.11 | -0.86 to -0.42 | 0.000 |
| | | 12653 | Childhood predictor | Found it very difficult | Lived comfortably | -0.67 | 0.12 | -0.91 to -0.43 | 0.000 |
| | | 12653 | Age | Per 1-year increase | | -0.02 | 0.00 | -0.02 to -0.01 | 0.000 |
| | | 12653 | Gender | Female | Male | 0.20 | 0.08 | 0.05 to 0.35 | 0.009 |
| | | 12653 | Education | Secondary | Elementary or less | 0.37 | 0.09 | 0.20 to 0.54 | 0.000 |
| | | 12653 | Education | Four years beyond HS | Elementary or less | 0.58 | 0.12 | 0.34 to 0.82 | 0.000 |
| | | 12653 | Residential setting | Small town | Rural | 0.38 | 0.10 | 0.18 to 0.59 | 0.000 |
| | | 12653 | Residential setting | Large city | Rural | 0.32 | 0.13 | 0.07 to 0.56 | 0.013 |
| | | 12653 | Residential setting | Suburb | Rural | -0.06 | 0.21 | -0.47 to 0.35 | 0.781 |
| **Remain hopeful for the future** | **Linear regression** | 12600 | *Intercept* | | | 8.14 | 0.13 | 7.89 to 8.39 | 0.000 |
| | | 12600 | Childhood predictor | Got by | Lived comfortably | 0.24 | 0.07 | 0.09 to 0.38 | 0.001 |
| | | 12600 | Childhood predictor | Found it difficult | Lived comfortably | 0.01 | 0.09 | -0.16 to 0.18 | 0.928 |
| | | 12600 | Childhood predictor | Found it very difficult | Lived comfortably | -0.23 | 0.10 | -0.43 to -0.04 | 0.021 |
| | | 12600 | Age | Per 1-year increase | | -0.01 | 0.00 | -0.01 to 0.00 | 0.002 |
| | | 12600 | Gender | Female | Male | 0.01 | 0.06 | -0.10 to 0.12 | 0.839 |
| | | 12600 | Education | Secondary | Elementary or less | 0.36 | 0.06 | 0.23 to 0.48 | 0.000 |
| | | 12600 | Education | Four years beyond HS | Elementary or less | 0.56 | 0.09 | 0.37 to 0.75 | 0.000 |
| | | 12600 | Residential setting | Small town | Rural | 0.48 | 0.08 | 0.32 to 0.64 | 0.000 |
| | | 12600 | Residential setting | Large city | Rural | 0.46 | 0.10 | 0.26 to 0.66 | 0.000 |
| | | 12600 | Residential setting | Suburb | Rural | 0.43 | 0.17 | 0.09 to 0.77 | 0.013 |
| **Understand purpose in life‡** | **Linear regression** | 12577 | *Intercept* | | | 8.11 | 0.14 | 7.84 to 8.38 | 0.000 |
| | | 12577 | Childhood predictor | Got by | Lived comfortably | 0.04 | 0.08 | -0.12 to 0.21 | 0.588 |
| | | 12577 | Childhood predictor | Found it difficult | Lived comfortably | -0.15 | 0.09 | -0.31 to 0.02 | 0.084 |
| | | 12577 | Childhood predictor | Found it very difficult | Lived comfortably | -0.41 | 0.10 | -0.62 to -0.21 | 0.000 |
| | | 12577 | Age | Per 1-year increase | | -0.01 | 0.00 | -0.02 to -0.01 | 0.000 |
| | | 12577 | Gender | Female | Male | 0.10 | 0.07 | -0.04 to 0.24 | 0.148 |
| | | 12577 | Education | Secondary | Elementary or less | 0.39 | 0.08 | 0.23 to 0.54 | 0.000 |
| | | 12577 | Education | Four years beyond HS | Elementary or less | 0.69 | 0.10 | 0.50 to 0.88 | 0.000 |
| | | 12577 | Residential setting | Small town | Rural | 0.42 | 0.09 | 0.24 to 0.59 | 0.000 |
| | | 12577 | Residential setting | Large city | Rural | 0.40 | 0.11 | 0.18 to 0.63 | 0.000 |
| | | 12577 | Residential setting | Suburb | Rural | 0.46 | 0.15 | 0.16 to 0.76 | 0.003 |

| Outcome | Model type | Analytic N | Variable | Level / comparison | Reference category | Coefficient | Standard error | 95% CI | p-value |
|---|---|---|---|---|---|---|---|---|---|
| **Life satisfaction‡** | **Linear regression** | 12633 | *Intercept* | | | 7.39 | 0.15 | 7.09 to 7.68 | 0.000 |
| | | 12633 | Childhood predictor | Got by | Lived comfortably | -0.25 | 0.10 | -0.44 to -0.06 | 0.011 |
| | | 12633 | Childhood predictor | Found it difficult | Lived comfortably | -0.57 | 0.10 | -0.77 to -0.37 | 0.000 |
| | | 12633 | Childhood predictor | Found it very difficult | Lived comfortably | -0.71 | 0.11 | -0.94 to -0.49 | 0.000 |
| | | 12633 | Age | Per 1-year increase | | -0.01 | 0.00 | -0.02 to -0.01 | 0.000 |
| | | 12633 | Gender | Female | Male | 0.27 | 0.08 | 0.12 to 0.42 | 0.000 |
| | | 12633 | Education | Secondary | Elementary or less | 0.24 | 0.09 | 0.07 to 0.42 | 0.007 |
| | | 12633 | Education | Four years beyond HS | Elementary or less | 0.39 | 0.12 | 0.16 to 0.63 | 0.001 |
| | | 12633 | Residential setting | Small town | Rural | 0.28 | 0.09 | 0.11 to 0.46 | 0.002 |
| | | 12633 | Residential setting | Large city | Rural | 0.18 | 0.12 | -0.06 to 0.43 | 0.134 |
| | | 12633 | Residential setting | Suburb | Rural | -0.13 | 0.20 | -0.52 to 0.25 | 0.502 |
| **Loneliness§** | **Linear regression** | 12653 | *Intercept* | | | 4.11 | 0.17 | 3.77 to 4.45 | 0.000 |
| | | 12653 | Childhood predictor | Got by | Lived comfortably | 0.05 | 0.11 | -0.16 to 0.26 | 0.644 |
| | | 12653 | Childhood predictor | Found it difficult | Lived comfortably | 0.36 | 0.12 | 0.13 to 0.59 | 0.002 |
| | | 12653 | Childhood predictor | Found it very difficult | Lived comfortably | 0.44 | 0.12 | 0.21 to 0.67 | 0.000 |
| | | 12653 | Age | Per 1-year increase | | 0.00 | 0.00 | -0.01 to 0.01 | 0.923 |
| | | 12653 | Gender | Female | Male | -0.14 | 0.09 | -0.31 to 0.03 | 0.110 |
| | | 12653 | Education | Secondary | Elementary or less | -0.28 | 0.10 | -0.47 to -0.09 | 0.003 |
| | | 12653 | Education | Four years beyond HS | Elementary or less | -0.60 | 0.14 | -0.88 to -0.31 | 0.000 |
| | | 12653 | Residential setting | Small town | Rural | -0.37 | 0.10 | -0.57 to -0.18 | 0.000 |
| | | 12653 | Residential setting | Large city | Rural | -0.48 | 0.14 | -0.75 to -0.21 | 0.001 |
| | | 12653 | Residential setting | Suburb | Rural | -0.20 | 0.24 | -0.68 to 0.27 | 0.399 |
| **Mental health‡** | **Linear regression** | 12657 | *Intercept* | | | 8.30 | 0.14 | 8.01 to 8.58 | 0.000 |
| | | 12657 | Childhood predictor | Got by | Lived comfortably | -0.20 | 0.09 | -0.38 to -0.01 | 0.037 |
| | | 12657 | Childhood predictor | Found it difficult | Lived comfortably | -0.42 | 0.10 | -0.62 to -0.22 | 0.000 |
| | | 12657 | Childhood predictor | Found it very difficult | Lived comfortably | -0.73 | 0.11 | -0.94 to -0.52 | 0.000 |
| | | 12657 | Age | Per 1-year increase | | -0.02 | 0.00 | -0.03 to -0.02 | 0.000 |
| | | 12657 | Gender | Female | Male | -0.16 | 0.07 | -0.29 to -0.02 | 0.021 |
| | | 12657 | Education | Secondary | Elementary or less | 0.35 | 0.08 | 0.20 to 0.51 | 0.000 |
| | | 12657 | Education | Four years beyond HS | Elementary or less | 0.45 | 0.11 | 0.23 to 0.67 | 0.000 |
| | | 12657 | Residential setting | Small town | Rural | 0.40 | 0.09 | 0.22 to 0.57 | 0.000 |
| | | 12657 | Residential setting | Large city | Rural | 0.42 | 0.12 | 0.19 to 0.66 | 0.000 |
| | | 12657 | Residential setting | Suburb | Rural | 0.16 | 0.18 | -0.19 to 0.52 | 0.372 |
| **Perceived support from others** | **Linear regression** | 12641 | *Intercept* | | | 6.83 | 0.16 | 6.52 to 7.14 | 0.000 |
| | | 12641 | Childhood predictor | Got by | Lived comfortably | -0.16 | 0.11 | -0.37 to 0.05 | 0.129 |
| | | 12641 | Childhood predictor | Found it difficult | Lived comfortably | -0.41 | 0.12 | -0.64 to -0.19 | 0.000 |
| | | 12641 | Childhood predictor | Found it very difficult | Lived comfortably | -0.43 | 0.13 | -0.68 to -0.17 | 0.001 |
| | | 12641 | Age | Per 1-year increase | | -0.01 | 0.00 | -0.02 to -0.01 | 0.000 |
| | | 12641 | Gender | Female | Male | 0.50 | 0.09 | 0.32 to 0.68 | 0.000 |
| | | 12641 | Education | Secondary | Elementary or less | 0.17 | 0.10 | -0.02 to 0.36 | 0.083 |

| Outcome | Model type | Analytic N | Variable | Level / comparison | Reference category | Coefficient | Standard error | 95% CI | p-value |
|---|---|---|---|---|---|---|---|---|---|
| | | 12641 | Education | Four years beyond HS | Elementary or less | 0.09 | 0.14 | -0.19 to 0.36 | 0.526 |
| | | 12641 | Residential setting | Small town | Rural | 0.18 | 0.11 | -0.03 to 0.39 | 0.096 |
| | | 12641 | Residential setting | Large city | Rural | -0.52 | 0.14 | -0.79 to -0.25 | 0.000 |
| | | 12641 | Residential setting | Suburb | Rural | -0.27 | 0.20 | -0.66 to 0.12 | 0.173 |
| **Physical health‡** | **Linear regression** | 12660 | *Intercept* | | | 8.59 | 0.15 | 8.31 to 8.88 | 0.000 |
| | | 12660 | Childhood predictor | Got by | Lived comfortably | -0.20 | 0.09 | -0.38 to -0.03 | 0.025 |
| | | 12660 | Childhood predictor | Found it difficult | Lived comfortably | -0.40 | 0.09 | -0.58 to -0.21 | 0.000 |
| | | 12660 | Childhood predictor | Found it very difficult | Lived comfortably | -0.62 | 0.11 | -0.83 to -0.41 | 0.000 |
| | | 12660 | Age | Per 1-year increase | | -0.04 | 0.00 | -0.05 to -0.04 | 0.000 |
| | | 12660 | Gender | Female | Male | -0.37 | 0.08 | -0.51 to -0.22 | 0.000 |
| | | 12660 | Education | Secondary | Elementary or less | 0.32 | 0.08 | 0.17 to 0.48 | 0.000 |
| | | 12660 | Education | Four years beyond HS | Elementary or less | 0.59 | 0.11 | 0.38 to 0.81 | 0.000 |
| | | 12660 | Residential setting | Small town | Rural | 0.49 | 0.09 | 0.31 to 0.67 | 0.000 |
| | | 12660 | Residential setting | Large city | Rural | 0.64 | 0.11 | 0.43 to 0.84 | 0.000 |
| | | 12660 | Residential setting | Suburb | Rural | 0.22 | 0.17 | -0.11 to 0.56 | 0.194 |
| **Act to promote good‡** | **Linear regression** | 12615 | *Intercept* | | | 8.06 | 0.11 | 7.84 to 8.28 | 0.000 |
| | | 12615 | Childhood predictor | Got by | Lived comfortably | 0.16 | 0.07 | 0.02 to 0.30 | 0.026 |
| | | 12615 | Childhood predictor | Found it difficult | Lived comfortably | 0.01 | 0.08 | -0.14 to 0.16 | 0.919 |
| | | 12615 | Childhood predictor | Found it very difficult | Lived comfortably | -0.43 | 0.10 | -0.62 to -0.24 | 0.000 |
| | | 12615 | Age | Per 1-year increase | | -0.01 | 0.00 | -0.01 to 0.00 | 0.006 |
| | | 12615 | Gender | Female | Male | 0.12 | 0.06 | 0.00 to 0.23 | 0.045 |
| | | 12615 | Education | Secondary | Elementary or less | 0.34 | 0.06 | 0.22 to 0.46 | 0.000 |
| | | 12615 | Education | Four years beyond HS | Elementary or less | 0.49 | 0.08 | 0.33 to 0.65 | 0.000 |
| | | 12615 | Residential setting | Small town | Rural | 0.59 | 0.07 | 0.45 to 0.73 | 0.000 |
| | | 12615 | Residential setting | Large city | Rural | 0.62 | 0.10 | 0.43 to 0.81 | 0.000 |
| | | 12615 | Residential setting | Suburb | Rural | 0.46 | 0.16 | 0.14 to 0.78 | 0.005 |
| **Relationship satisfaction‡** | **Linear regression** | 12621 | *Intercept* | | | 7.93 | 0.14 | 7.67 to 8.20 | 0.000 |
| | | 12621 | Childhood predictor | Got by | Lived comfortably | 0.08 | 0.09 | -0.09 to 0.25 | 0.368 |
| | | 12621 | Childhood predictor | Found it difficult | Lived comfortably | -0.22 | 0.10 | -0.41 to -0.02 | 0.028 |
| | | 12621 | Childhood predictor | Found it very difficult | Lived comfortably | -0.41 | 0.11 | -0.63 to -0.19 | 0.000 |
| | | 12621 | Age | Per 1-year increase | | -0.01 | 0.00 | -0.01 to 0.00 | 0.003 |
| | | 12621 | Gender | Female | Male | 0.32 | 0.07 | 0.18 to 0.45 | 0.000 |
| | | 12621 | Education | Secondary | Elementary or less | -0.06 | 0.08 | -0.22 to 0.09 | 0.414 |
| | | 12621 | Education | Four years beyond HS | Elementary or less | 0.00 | 0.11 | -0.22 to 0.21 | 0.983 |
| | | 12621 | Residential setting | Small town | Rural | 0.43 | 0.09 | 0.26 to 0.60 | 0.000 |
| | | 12621 | Residential setting | Large city | Rural | 0.31 | 0.11 | 0.11 to 0.52 | 0.003 |
| | | 12621 | Residential setting | Suburb | Rural | 0.06 | 0.15 | -0.24 to 0.36 | 0.688 |
| **Show love often** | **Linear regression** | 12644 | *Intercept* | | | 8.29 | 0.13 | 8.03 to 8.55 | 0.000 |
| | | 12644 | Childhood predictor | Got by | Lived comfortably | 0.05 | 0.09 | -0.12 to 0.22 | 0.567 |
| | | 12644 | Childhood predictor | Found it difficult | Lived comfortably | -0.09 | 0.10 | -0.28 to 0.09 | 0.329 |

| Outcome | Model type | Analytic N | Variable | Level / comparison | Reference category | Coefficient | Standard error | 95% CI | p-value |
|---|---|---|---|---|---|---|---|---|---|
| | | 12644 | Childhood predictor | Found it very difficult | Lived comfortably | -0.14 | 0.11 | -0.35 to 0.08 | 0.211 |
| | | 12644 | Age | Per 1-year increase | | -0.01 | 0.00 | -0.01 to 0.00 | 0.003 |
| | | 12644 | Gender | Female | Male | 0.21 | 0.07 | 0.08 to 0.34 | 0.001 |
| | | 12644 | Education | Secondary | Elementary or less | 0.15 | 0.07 | 0.01 to 0.28 | 0.041 |
| | | 12644 | Education | Four years beyond HS | Elementary or less | 0.03 | 0.11 | -0.19 to 0.25 | 0.798 |
| | | 12644 | Residential setting | Small town | Rural | 0.20 | 0.08 | 0.05 to 0.35 | 0.010 |
| | | 12644 | Residential setting | Large city | Rural | 0.03 | 0.11 | -0.19 to 0.25 | 0.775 |
| | | 12644 | Residential setting | Suburb | Rural | -0.13 | 0.20 | -0.52 to 0.26 | 0.502 |
| **Life evaluation five years from now** | **Linear regression** | 11525 | *Intercept* | | | 7.92 | 0.14 | 7.64 to 8.20 | 0.000 |
| | | 11525 | Childhood predictor | Got by | Lived comfortably | -0.14 | 0.09 | -0.32 to 0.03 | 0.109 |
| | | 11525 | Childhood predictor | Found it difficult | Lived comfortably | -0.39 | 0.10 | -0.58 to -0.20 | 0.000 |
| | | 11525 | Childhood predictor | Found it very difficult | Lived comfortably | -0.75 | 0.11 | -0.97 to -0.54 | 0.000 |
| | | 11525 | Age | Per 1-year increase | | -0.02 | 0.00 | -0.03 to -0.02 | 0.000 |
| | | 11525 | Gender | Female | Male | 0.15 | 0.07 | 0.01 to 0.29 | 0.032 |
| | | 11525 | Education | Secondary | Elementary or less | 0.21 | 0.08 | 0.06 to 0.37 | 0.008 |
| | | 11525 | Education | Four years beyond HS | Elementary or less | 0.48 | 0.11 | 0.25 to 0.70 | 0.000 |
| | | 11525 | Residential setting | Small town | Rural | 0.45 | 0.09 | 0.27 to 0.62 | 0.000 |
| | | 11525 | Residential setting | Large city | Rural | 0.48 | 0.11 | 0.26 to 0.70 | 0.000 |
| | | 11525 | Residential setting | Suburb | Rural | 0.36 | 0.23 | -0.08 to 0.81 | 0.109 |
| **Life evaluation today** | **Linear regression** | 12554 | *Intercept* | | | 5.91 | 0.17 | 5.57 to 6.25 | 0.000 |
| | | 12554 | Childhood predictor | Got by | Lived comfortably | -0.38 | 0.10 | -0.58 to -0.17 | 0.000 |
| | | 12554 | Childhood predictor | Found it difficult | Lived comfortably | -0.69 | 0.11 | -0.90 to -0.47 | 0.000 |
| | | 12554 | Childhood predictor | Found it very difficult | Lived comfortably | -0.70 | 0.13 | -0.95 to -0.45 | 0.000 |
| | | 12554 | Age | Per 1-year increase | | -0.01 | 0.00 | -0.01 to 0.00 | 0.015 |
| | | 12554 | Gender | Female | Male | 0.44 | 0.08 | 0.28 to 0.61 | 0.000 |
| | | 12554 | Education | Secondary | Elementary or less | 0.18 | 0.08 | 0.01 to 0.34 | 0.036 |
| | | 12554 | Education | Four years beyond HS | Elementary or less | 0.10 | 0.14 | -0.18 to 0.38 | 0.481 |
| | | 12554 | Residential setting | Small town | Rural | 0.23 | 0.10 | 0.02 to 0.43 | 0.030 |
| | | 12554 | Residential setting | Large city | Rural | 0.05 | 0.13 | -0.21 to 0.32 | 0.692 |
| | | 12554 | Residential setting | Suburb | Rural | -0.04 | 0.20 | -0.43 to 0.36 | 0.857 |
| **Worry about safety, food, or housing§§‡‡** | **Linear regression** | 12669 | *Intercept* | | | 6.03 | 0.20 | 5.63 to 6.42 | 0.000 |
| | | 12669 | Childhood predictor | Got by | Lived comfortably | 0.32 | 0.13 | 0.07 to 0.56 | 0.013 |
| | | 12669 | Childhood predictor | Found it difficult | Lived comfortably | 0.39 | 0.13 | 0.14 to 0.65 | 0.003 |
| | | 12669 | Childhood predictor | Found it very difficult | Lived comfortably | 0.26 | 0.15 | -0.03 to 0.55 | 0.084 |
| | | 12669 | Age | Per 1-year increase | | -0.01 | 0.00 | -0.02 to 0.00 | 0.007 |
| | | 12669 | Gender | Female | Male | 0.36 | 0.09 | 0.19 to 0.53 | 0.000 |
| | | 12669 | Education | Secondary | Elementary or less | -0.52 | 0.11 | -0.73 to -0.31 | 0.000 |
| | | 12669 | Education | Four years beyond HS | Elementary or less | -0.76 | 0.17 | -1.09 to -0.43 | 0.000 |
| | | 12669 | Residential setting | Small town | Rural | 0.36 | 0.13 | 0.10 to 0.62 | 0.007 |

| Outcome | Model type | Analytic N | Variable | Level / comparison | Reference category | Coefficient | Standard error | 95% CI | p-value |
|---|---|---|---|---|---|---|---|---|---|
| | | 12669 | Residential setting | Large city | Rural | -0.10 | 0.18 | -0.44 to 0.25 | 0.579 |
| | | 12669 | Residential setting | Suburb | Rural | 0.13 | 0.25 | -0.36 to 0.63 | 0.597 |
| **Life worthwhile‡** | **Linear regression** | 12551 | *Intercept* | | | 6.92 | 0.15 | 6.63 to 7.20 | 0.000 |
| | | 12551 | Childhood predictor | Got by | Lived comfortably | -0.28 | 0.09 | -0.46 to -0.10 | 0.003 |
| | | 12551 | Childhood predictor | Found it difficult | Lived comfortably | -0.39 | 0.11 | -0.61 to -0.17 | 0.000 |
| | | 12551 | Childhood predictor | Found it very difficult | Lived comfortably | -0.82 | 0.12 | -1.07 to -0.58 | 0.000 |
| | | 12551 | Age | Per 1-year increase | | -0.01 | 0.00 | -0.01 to 0.00 | 0.006 |
| | | 12551 | Gender | Female | Male | 0.40 | 0.07 | 0.26 to 0.54 | 0.000 |
| | | 12551 | Education | Secondary | Elementary or less | 0.39 | 0.08 | 0.24 to 0.55 | 0.000 |
| | | 12551 | Education | Four years beyond HS | Elementary or less | 0.52 | 0.13 | 0.28 to 0.77 | 0.000 |
| | | 12551 | Residential setting | Small town | Rural | 0.43 | 0.10 | 0.23 to 0.63 | 0.000 |
| | | 12551 | Residential setting | Large city | Rural | 0.40 | 0.13 | 0.15 to 0.65 | 0.002 |
| | | 12551 | Residential setting | Suburb | Rural | 0.37 | 0.20 | -0.02 to 0.75 | 0.062 |
| **Flourishing (with financial indicators)** | **Linear regression** | 12198 | *Intercept* | | | 7.17 | 0.08 | 7.01 to 7.33 | 0.000 |
| | | 12198 | Childhood predictor | Got by | Lived comfortably | -0.16 | 0.05 | -0.25 to -0.06 | 0.002 |
| | | 12198 | Childhood predictor | Found it difficult | Lived comfortably | -0.33 | 0.06 | -0.45 to -0.21 | 0.000 |
| | | 12198 | Childhood predictor | Found it very difficult | Lived comfortably | -0.54 | 0.07 | -0.68 to -0.40 | 0.000 |
| | | 12198 | Age | Per 1-year increase | | -0.01 | 0.00 | -0.01 to -0.01 | 0.000 |
| | | 12198 | Gender | Female | Male | 0.07 | 0.04 | -0.01 to 0.15 | 0.097 |
| | | 12198 | Education | Secondary | Elementary or less | 0.29 | 0.05 | 0.20 to 0.38 | 0.000 |
| | | 12198 | Education | Four years beyond HS | Elementary or less | 0.45 | 0.07 | 0.32 to 0.58 | 0.000 |
| | | 12198 | Residential setting | Small town | Rural | 0.31 | 0.06 | 0.20 to 0.42 | 0.000 |
| | | 12198 | Residential setting | Large city | Rural | 0.33 | 0.07 | 0.20 to 0.46 | 0.000 |
| | | 12198 | Residential setting | Suburb | Rural | 0.14 | 0.13 | -0.11 to 0.39 | 0.279 |
| **Flourishing (without financial indicators)** | **Linear regression** | 12227 | *Intercept* | | | 7.77 | 0.09 | 7.59 to 7.94 | 0.000 |
| | | 12227 | Childhood predictor | Got by | Lived comfortably | -0.11 | 0.06 | -0.22 to 0.00 | 0.058 |
| | | 12227 | Childhood predictor | Found it difficult | Lived comfortably | -0.30 | 0.06 | -0.42 to -0.17 | 0.000 |
| | | 12227 | Childhood predictor | Found it very difficult | Lived comfortably | -0.57 | 0.07 | -0.71 to -0.43 | 0.000 |
| | | 12227 | Age | Per 1-year increase | | -0.01 | 0.00 | -0.02 to -0.01 | 0.000 |
| | | 12227 | Gender | Female | Male | 0.16 | 0.04 | 0.07 to 0.24 | 0.000 |
| | | 12227 | Education | Secondary | Elementary or less | 0.25 | 0.05 | 0.15 to 0.35 | 0.000 |
| | | 12227 | Education | Four years beyond HS | Elementary or less | 0.41 | 0.07 | 0.27 to 0.54 | 0.000 |
| | | 12227 | Residential setting | Small town | Rural | 0.43 | 0.06 | 0.31 to 0.55 | 0.000 |
| | | 12227 | Residential setting | Large city | Rural | 0.37 | 0.08 | 0.22 to 0.52 | 0.000 |
| | | 12227 | Residential setting | Suburb | Rural | 0.15 | 0.13 | -0.10 to 0.40 | 0.227 |
| **Feel close to someone** | **Logistic regression** | 12665 | *Intercept* | | | 1.86 | 0.14 | 1.58 to 2.14 | 0.000 |
| | | 12665 | Childhood predictor | Got by | Lived comfortably | -0.10 | 0.08 | -0.26 to 0.06 | 0.204 |
| | | 12665 | Childhood predictor | Found it difficult | Lived comfortably | -0.39 | 0.09 | -0.56 to -0.22 | 0.000 |
| | | 12665 | Childhood predictor | Found it very difficult | Lived comfortably | -0.37 | 0.10 | -0.56 to -0.18 | 0.000 |

| Outcome | Model type | Analytic N | Variable | Level / comparison | Reference category | Coefficient | Standard error | 95% CI | p-value |
|---|---|---|---|---|---|---|---|---|---|
| | | 12665 | Age | Per 1-year increase | | -0.01 | 0.00 | -0.01 to 0.00 | 0.015 |
| | | 12665 | Gender | Female | Male | 0.00 | 0.06 | -0.13 to 0.12 | 0.971 |
| | | 12665 | Education | Secondary | Elementary or less | 0.13 | 0.08 | -0.03 to 0.29 | 0.122 |
| | | 12665 | Education | Four years beyond HS | Elementary or less | 0.10 | 0.13 | -0.16 to 0.36 | 0.464 |
| | | 12665 | Residential setting | Small town | Rural | 0.09 | 0.11 | -0.12 to 0.31 | 0.395 |
| | | 12665 | Residential setting | Large city | Rural | -0.14 | 0.13 | -0.39 to 0.11 | 0.285 |
| | | 12665 | Residential setting | Suburb | Rural | -0.15 | 0.19 | -0.53 to 0.22 | 0.424 |
| **Donated to charity** | **Logistic regression** | 12631 | *Intercept* | | | -0.24 | 0.13 | -0.49 to 0.01 | 0.061 |
| | | 12631 | Childhood predictor | Got by | Lived comfortably | -0.04 | 0.07 | -0.17 to 0.09 | 0.553 |
| | | 12631 | Childhood predictor | Found it difficult | Lived comfortably | -0.19 | 0.07 | -0.33 to -0.05 | 0.009 |
| | | 12631 | Childhood predictor | Found it very difficult | Lived comfortably | -0.32 | 0.09 | -0.49 to -0.15 | 0.000 |
| | | 12631 | Age | Per 1-year increase | | 0.00 | 0.00 | 0.00 to 0.01 | 0.417 |
| | | 12631 | Gender | Female | Male | -0.05 | 0.05 | -0.14 to 0.05 | 0.357 |
| | | 12631 | Education | Secondary | Elementary or less | -0.10 | 0.06 | -0.22 to 0.01 | 0.081 |
| | | 12631 | Education | Four years beyond HS | Elementary or less | -0.19 | 0.10 | -0.39 to 0.00 | 0.056 |
| | | 12631 | Residential setting | Small town | Rural | -0.08 | 0.09 | -0.26 to 0.10 | 0.384 |
| | | 12631 | Residential setting | Large city | Rural | -0.44 | 0.11 | -0.66 to -0.22 | 0.000 |
| | | 12631 | Residential setting | Suburb | Rural | -0.19 | 0.19 | -0.56 to 0.19 | 0.326 |
| **Have any health problems** | **Logistic regression** | 12587 | *Intercept* | | | -2.12 | 0.11 | -2.33 to -1.91 | 0.000 |
| | | 12587 | Childhood predictor | Got by | Lived comfortably | 0.15 | 0.06 | 0.03 to 0.27 | 0.016 |
| | | 12587 | Childhood predictor | Found it difficult | Lived comfortably | 0.13 | 0.06 | 0.01 to 0.26 | 0.040 |
| | | 12587 | Childhood predictor | Found it very difficult | Lived comfortably | 0.19 | 0.07 | 0.05 to 0.33 | 0.010 |
| | | 12587 | Age | Per 1-year increase | | 0.02 | 0.00 | 0.02 to 0.03 | 0.000 |
| | | 12587 | Gender | Female | Male | 0.24 | 0.05 | 0.14 to 0.34 | 0.000 |
| | | 12587 | Education | Secondary | Elementary or less | -0.13 | 0.06 | -0.24 to -0.02 | 0.023 |
| | | 12587 | Education | Four years beyond HS | Elementary or less | -0.25 | 0.09 | -0.44 to -0.07 | 0.008 |
| | | 12587 | Residential setting | Small town | Rural | 0.12 | 0.06 | -0.01 to 0.24 | 0.065 |
| | | 12587 | Residential setting | Large city | Rural | -0.14 | 0.09 | -0.31 to 0.03 | 0.105 |
| | | 12587 | Residential setting | Suburb | Rural | 0.05 | 0.14 | -0.22 to 0.32 | 0.735 |
| **Helped a stranger** | **Logistic regression** | 12534 | *Intercept* | | | 0.75 | 0.12 | 0.51 to 0.98 | 0.000 |
| | | 12534 | Childhood predictor | Got by | Lived comfortably | -0.14 | 0.07 | -0.27 to -0.01 | 0.034 |
| | | 12534 | Childhood predictor | Found it difficult | Lived comfortably | -0.14 | 0.07 | -0.28 to 0.01 | 0.068 |
| | | 12534 | Childhood predictor | Found it very difficult | Lived comfortably | -0.10 | 0.08 | -0.26 to 0.06 | 0.214 |
| | | 12534 | Age | Per 1-year increase | | -0.01 | 0.00 | -0.01 to 0.00 | 0.000 |
| | | 12534 | Gender | Female | Male | -0.33 | 0.05 | -0.42 to -0.24 | 0.000 |
| | | 12534 | Education | Secondary | Elementary or less | -0.07 | 0.06 | -0.18 to 0.05 | 0.241 |
| | | 12534 | Education | Four years beyond HS | Elementary or less | -0.09 | 0.09 | -0.27 to 0.08 | 0.291 |
| | | 12534 | Residential setting | Small town | Rural | 0.10 | 0.09 | -0.07 to 0.27 | 0.243 |
| | | 12534 | Residential setting | Large city | Rural | -0.16 | 0.10 | -0.36 to 0.03 | 0.101 |
| | | 12534 | Residential setting | Suburb | Rural | 0.05 | 0.18 | -0.29 to 0.40 | 0.773 |

| Outcome | Model type | Analytic N | Variable | Level / comparison | Reference category | Coefficient | Standard error | 95% CI | p-value |
|---|---|---|---|---|---|---|---|---|---|
| **Volunteered at an organization** | **Logistic regression** | 12615 | *Intercept* | | | -0.18 | 0.14 | -0.45 to 0.10 | 0.216 |
| | | 12615 | Childhood predictor | Got by | Lived comfortably | -0.22 | 0.08 | -0.37 to -0.07 | 0.005 |
| | | 12615 | Childhood predictor | Found it difficult | Lived comfortably | -0.24 | 0.08 | -0.39 to -0.08 | 0.002 |
| | | 12615 | Childhood predictor | Found it very difficult | Lived comfortably | -0.29 | 0.10 | -0.48 to -0.10 | 0.003 |
| | | 12615 | Age | Per 1-year increase | | -0.01 | 0.00 | -0.01 to 0.00 | 0.003 |
| | | 12615 | Gender | Female | Male | -0.30 | 0.05 | -0.40 to -0.19 | 0.000 |
| | | 12615 | Education | Secondary | Elementary or less | -0.16 | 0.06 | -0.29 to -0.03 | 0.015 |
| | | 12615 | Education | Four years beyond HS | Elementary or less | 0.01 | 0.10 | -0.19 to 0.21 | 0.934 |
| | | 12615 | Residential setting | Small town | Rural | -0.11 | 0.11 | -0.32 to 0.10 | 0.322 |
| | | 12615 | Residential setting | Large city | Rural | -0.57 | 0.13 | -0.81 to -0.32 | 0.000 |
| | | 12615 | Residential setting | Suburb | Rural | -0.35 | 0.16 | -0.66 to -0.03 | 0.031 |
| **Cigarette smoking** | **Logistic regression** | 12174 | *Intercept* | | | -2.02 | 0.17 | -2.34 to -1.69 | 0.000 |
| | | 12174 | Childhood predictor | Got by | Lived comfortably | 0.01 | 0.10 | -0.19 to 0.22 | 0.895 |
| | | 12174 | Childhood predictor | Found it difficult | Lived comfortably | -0.09 | 0.12 | -0.34 to 0.15 | 0.451 |
| | | 12174 | Childhood predictor | Found it very difficult | Lived comfortably | 0.31 | 0.13 | 0.05 to 0.57 | 0.018 |
| | | 12174 | Age | Per 1-year increase | | 0.01 | 0.00 | 0.00 to 0.01 | 0.032 |
| | | 12174 | Gender | Female | Male | -2.84 | 0.19 | -3.20 to -2.47 | 0.000 |
| | | 12174 | Education | Secondary | Elementary or less | -0.34 | 0.10 | -0.53 to -0.15 | 0.000 |
| | | 12174 | Education | Four years beyond HS | Elementary or less | -0.58 | 0.17 | -0.91 to -0.25 | 0.001 |
| | | 12174 | Residential setting | Small town | Rural | -0.06 | 0.10 | -0.26 to 0.15 | 0.586 |
| | | 12174 | Residential setting | Large city | Rural | 0.15 | 0.14 | -0.13 to 0.43 | 0.304 |
| | | 12174 | Residential setting | Suburb | Rural | 0.79 | 0.20 | 0.40 to 1.17 | 0.000 |
| **Alcohol use** | **Logistic regression** | 12159 | *Intercept* | | | -1.91 | 0.15 | -2.21 to -1.61 | 0.000 |
| | | 12159 | Childhood predictor | Got by | Lived comfortably | 0.10 | 0.12 | -0.14 to 0.34 | 0.400 |
| | | 12159 | Childhood predictor | Found it difficult | Lived comfortably | 0.07 | 0.12 | -0.17 to 0.30 | 0.579 |
| | | 12159 | Childhood predictor | Found it very difficult | Lived comfortably | 0.11 | 0.13 | -0.15 to 0.36 | 0.410 |
| | | 12159 | Age | Per 1-year increase | | 0.00 | 0.00 | -0.01 to 0.00 | 0.725 |
| | | 12159 | Gender | Female | Male | -2.75 | 0.17 | -3.08 to -2.41 | 0.000 |
| | | 12159 | Education | Secondary | Elementary or less | -0.24 | 0.10 | -0.43 to -0.05 | 0.016 |
| | | 12159 | Education | Four years beyond HS | Elementary or less | -0.53 | 0.17 | -0.86 to -0.20 | 0.002 |
| | | 12159 | Residential setting | Small town | Rural | 0.21 | 0.10 | 0.01 to 0.41 | 0.043 |
| | | 12159 | Residential setting | Large city | Rural | 0.43 | 0.14 | 0.16 to 0.71 | 0.002 |
| | | 12159 | Residential setting | Suburb | Rural | 0.65 | 0.21 | 0.23 to 1.07 | 0.002 |
| **Government approval§** | **Proportional-odds regression** | 12451 | Childhood predictor | Got by | Lived comfortably | -0.04 | 0.06 | -0.15 to 0.07 | 0.466 |
| | | 12451 | Childhood predictor | Found it difficult | Lived comfortably | -0.12 | 0.06 | -0.24 to 0.00 | 0.046 |
| | | 12451 | Childhood predictor | Found it very difficult | Lived comfortably | -0.40 | 0.08 | -0.55 to -0.25 | 0.000 |
| | | 12451 | Age | Per 1-year increase | | 0.00 | 0.00 | -0.01 to 0.00 | 0.110 |
| | | 12451 | Gender | Female | Male | 0.03 | 0.04 | -0.06 to 0.11 | 0.548 |
| | | 12451 | Education | Secondary | Elementary or less | -0.29 | 0.05 | -0.39 to -0.18 | 0.000 |
| | | 12451 | Education | Four years beyond HS | Elementary or less | -0.37 | 0.09 | -0.54 to -0.20 | 0.000 |

| Outcome | Model type | Analytic N | Variable | Level / comparison | Reference category | Coefficient | Standard error | 95% CI | p-value |
|---|---|---|---|---|---|---|---|---|---|
| | | 12451 | Residential setting | Small town | Rural | 0.09 | 0.07 | -0.04 to 0.22 | 0.176 |
| | | 12451 | Residential setting | Large city | Rural | -0.08 | 0.07 | -0.23 to 0.06 | 0.273 |
| | | 12451 | Residential setting | Suburb | Rural | -0.43 | 0.15 | -0.72 to -0.13 | 0.005 |
| | | 12451 | *Threshold / cut-point* | Strongly disapprove\|Somewhat disapprove | | -3.03 | 0.10 | -3.22 to -2.83 | 0.000 |
| | | 12451 | *Threshold / cut-point* | Somewhat disapprove\|Neither | | -2.36 | 0.10 | -2.55 to -2.17 | 0.000 |
| | | 12451 | *Threshold / cut-point* | Neither\|Somewhat approve | | -2.00 | 0.09 | -2.18 to -1.82 | 0.000 |
| | | 12451 | *Threshold / cut-point* | Somewhat approve\|Strongly approve | | -0.54 | 0.09 | -0.72 to -0.37 | 0.000 |
| **Feeling capable§** | **Proportional-odds regression** | 12610 | Childhood predictor | Got by | Lived comfortably | 0.01 | 0.05 | -0.09 to 0.11 | 0.796 |
| | | 12610 | Childhood predictor | Found it difficult | Lived comfortably | -0.03 | 0.05 | -0.14 to 0.07 | 0.540 |
| | | 12610 | Childhood predictor | Found it very difficult | Lived comfortably | -0.06 | 0.06 | -0.17 to 0.06 | 0.325 |
| | | 12610 | Age | Per 1-year increase | | 0.00 | 0.00 | 0.00 to 0.01 | 0.154 |
| | | 12610 | Gender | Female | Male | -0.08 | 0.04 | -0.16 to 0.01 | 0.080 |
| | | 12610 | Education | Secondary | Elementary or less | 0.15 | 0.05 | 0.05 to 0.25 | 0.002 |
| | | 12610 | Education | Four years beyond HS | Elementary or less | 0.09 | 0.07 | -0.05 to 0.23 | 0.215 |
| | | 12610 | Residential setting | Small town | Rural | 0.17 | 0.05 | 0.08 to 0.27 | 0.000 |
| | | 12610 | Residential setting | Large city | Rural | 0.19 | 0.07 | 0.05 to 0.34 | 0.009 |
| | | 12610 | Residential setting | Suburb | Rural | 0.26 | 0.09 | 0.09 to 0.43 | 0.003 |
| | | 12610 | *Threshold / cut-point* | Never\|Rarely | | -2.35 | 0.09 | -2.52 to -2.17 | 0.000 |
| | | 12610 | *Threshold / cut-point* | Rarely\|Often | | -0.63 | 0.09 | -0.80 to -0.47 | 0.000 |
| | | 12610 | *Threshold / cut-point* | Often\|Always | | 0.03 | 0.09 | -0.14 to 0.20 | 0.720 |
| **Unable to control worry§** | **Proportional-odds regression** | 12561 | Childhood predictor | Got by | Lived comfortably | 0.05 | 0.05 | -0.05 to 0.15 | 0.348 |
| | | 12561 | Childhood predictor | Found it difficult | Lived comfortably | 0.25 | 0.06 | 0.14 to 0.36 | 0.000 |
| | | 12561 | Childhood predictor | Found it very difficult | Lived comfortably | 0.21 | 0.06 | 0.09 to 0.34 | 0.001 |
| | | 12561 | Age | Per 1-year increase | | 0.00 | 0.00 | 0.00 to 0.01 | 0.004 |
| | | 12561 | Gender | Female | Male | 0.19 | 0.04 | 0.11 to 0.27 | 0.000 |
| | | 12561 | Education | Secondary | Elementary or less | -0.25 | 0.05 | -0.34 to -0.16 | 0.000 |
| | | 12561 | Education | Four years beyond HS | Elementary or less | -0.40 | 0.07 | -0.55 to -0.26 | 0.000 |
| | | 12561 | Residential setting | Small town | Rural | 0.04 | 0.05 | -0.05 to 0.14 | 0.383 |
| | | 12561 | Residential setting | Large city | Rural | 0.01 | 0.07 | -0.13 to 0.14 | 0.906 |
| | | 12561 | Residential setting | Suburb | Rural | -0.12 | 0.10 | -0.32 to 0.07 | 0.206 |
| | | 12561 | *Threshold / cut-point* | Not at all\|Several days | | 0.16 | 0.08 | -0.01 to 0.32 | 0.060 |
| | | 12561 | *Threshold / cut-point* | Several days\|More than half the days | | 1.07 | 0.08 | 0.91 to 1.23 | 0.000 |
| | | 12561 | *Threshold / cut-point* | More than half the days\|Nearly every day | | 1.91 | 0.09 | 1.74 to 2.08 | 0.000 |
| **Depressive symptoms§** | **Proportional-odds regression** | 12584 | Childhood predictor | Got by | Lived comfortably | 0.12 | 0.05 | 0.02 to 0.23 | 0.021 |

| Outcome | Model type | Analytic N | Variable | Level / comparison | Reference category | Coefficient | Standard error | 95% CI | p-value |
|---|---|---|---|---|---|---|---|---|---|
| | | 12584 | Childhood predictor | Found it difficult | Lived comfortably | 0.24 | 0.06 | 0.13 to 0.35 | 0.000 |
| | | 12584 | Childhood predictor | Found it very difficult | Lived comfortably | 0.22 | 0.06 | 0.10 to 0.35 | 0.001 |
| | | 12584 | Age | Per 1-year increase | | 0.00 | 0.00 | 0.00 to 0.00 | 0.384 |
| | | 12584 | Gender | Female | Male | 0.11 | 0.04 | 0.04 to 0.19 | 0.004 |
| | | 12584 | Education | Secondary | Elementary or less | -0.18 | 0.05 | -0.27 to -0.09 | 0.000 |
| | | 12584 | Education | Four years beyond HS | Elementary or less | -0.38 | 0.07 | -0.52 to -0.25 | 0.000 |
| | | 12584 | Residential setting | Small town | Rural | -0.01 | 0.05 | -0.12 to 0.09 | 0.839 |
| | | 12584 | Residential setting | Large city | Rural | -0.08 | 0.07 | -0.22 to 0.06 | 0.269 |
| | | 12584 | Residential setting | Suburb | Rural | -0.21 | 0.11 | -0.43 to 0.02 | 0.069 |
| | | 12584 | *Threshold / cut-point* | Not at all\|Several days | | -0.15 | 0.08 | -0.31 to 0.01 | 0.065 |
| | | 12584 | *Threshold / cut-point* | Several days\|More than half the days | | 0.93 | 0.08 | 0.77 to 1.09 | 0.000 |
| | | 12584 | *Threshold / cut-point* | More than half the days\|Nearly every day | | 1.93 | 0.09 | 1.77 to 2.10 | 0.000 |
| **Perceived discrimination§** | **Proportional-odds regression** | 12515 | Childhood predictor | Got by | Lived comfortably | -0.11 | 0.05 | -0.22 to -0.01 | 0.034 |
| | | 12515 | Childhood predictor | Found it difficult | Lived comfortably | -0.16 | 0.06 | -0.29 to -0.04 | 0.009 |
| | | 12515 | Childhood predictor | Found it very difficult | Lived comfortably | 0.09 | 0.06 | -0.04 to 0.21 | 0.163 |
| | | 12515 | Age | Per 1-year increase | | 0.00 | 0.00 | -0.01 to 0.00 | 0.026 |
| | | 12515 | Gender | Female | Male | -0.14 | 0.04 | -0.23 to -0.05 | 0.001 |
| | | 12515 | Education | Secondary | Elementary or less | -0.23 | 0.05 | -0.33 to -0.13 | 0.000 |
| | | 12515 | Education | Four years beyond HS | Elementary or less | -0.29 | 0.09 | -0.46 to -0.12 | 0.001 |
| | | 12515 | Residential setting | Small town | Rural | -0.20 | 0.06 | -0.31 to -0.09 | 0.000 |
| | | 12515 | Residential setting | Large city | Rural | -0.23 | 0.07 | -0.37 to -0.09 | 0.001 |
| | | 12515 | Residential setting | Suburb | Rural | -0.35 | 0.09 | -0.53 to -0.17 | 0.000 |
| | | 12515 | *Threshold / cut-point* | Never\|Rarely | | -0.19 | 0.09 | -0.37 to -0.02 | 0.033 |
| | | 12515 | *Threshold / cut-point* | Rarely\|Often | | 0.81 | 0.09 | 0.63 to 0.99 | 0.000 |
| | | 12515 | *Threshold / cut-point* | Often\|Always | | 1.43 | 0.09 | 1.25 to 1.61 | 0.000 |
| **Feeling nervous or anxious§** | **Proportional-odds regression** | 12613 | Childhood predictor | Got by | Lived comfortably | 0.17 | 0.05 | 0.07 to 0.27 | 0.001 |
| | | 12613 | Childhood predictor | Found it difficult | Lived comfortably | 0.29 | 0.05 | 0.19 to 0.40 | 0.000 |
| | | 12613 | Childhood predictor | Found it very difficult | Lived comfortably | 0.26 | 0.06 | 0.14 to 0.38 | 0.000 |
| | | 12613 | Age | Per 1-year increase | | 0.01 | 0.00 | 0.00 to 0.01 | 0.000 |
| | | 12613 | Gender | Female | Male | 0.33 | 0.04 | 0.25 to 0.41 | 0.000 |
| | | 12613 | Education | Secondary | Elementary or less | -0.27 | 0.05 | -0.36 to -0.18 | 0.000 |
| | | 12613 | Education | Four years beyond HS | Elementary or less | -0.44 | 0.08 | -0.59 to -0.29 | 0.000 |
| | | 12613 | Residential setting | Small town | Rural | 0.10 | 0.05 | -0.01 to 0.21 | 0.063 |
| | | 12613 | Residential setting | Large city | Rural | -0.01 | 0.07 | -0.15 to 0.12 | 0.863 |
| | | 12613 | Residential setting | Suburb | Rural | -0.07 | 0.12 | -0.30 to 0.16 | 0.539 |
| | | 12613 | *Threshold / cut-point* | Not at all\|Several days | | 0.26 | 0.08 | 0.09 to 0.42 | 0.002 |
| | | 12613 | *Threshold / cut-point* | Several days\|More than half the days | | 1.26 | 0.08 | 1.10 to 1.43 | 0.000 |

| Outcome | Model type | Analytic N | Variable | Level / comparison | Reference category | Coefficient | Standard error | 95% CI | p-value |
|---|---|---|---|---|---|---|---|---|---|
| | | 12613 | *Threshold / cut-point* | More than half the days\|Nearly every day | | 2.20 | 0.09 | 2.03 to 2.37 | 0.000 |
| **Forgiveness§** | **Proportional-odds regression** | 12590 | Childhood predictor | Got by | Lived comfortably | 0.00 | 0.05 | -0.11 to 0.10 | 0.932 |
| | | 12590 | Childhood predictor | Found it difficult | Lived comfortably | -0.02 | 0.06 | -0.13 to 0.09 | 0.762 |
| | | 12590 | Childhood predictor | Found it very difficult | Lived comfortably | -0.03 | 0.07 | -0.16 to 0.11 | 0.689 |
| | | 12590 | Age | Per 1-year increase | | 0.00 | 0.00 | 0.00 to 0.00 | 0.764 |
| | | 12590 | Gender | Female | Male | 0.09 | 0.04 | 0.01 to 0.17 | 0.038 |
| | | 12590 | Education | Secondary | Elementary or less | -0.08 | 0.05 | -0.18 to 0.02 | 0.132 |
| | | 12590 | Education | Four years beyond HS | Elementary or less | -0.20 | 0.08 | -0.35 to -0.05 | 0.009 |
| | | 12590 | Residential setting | Small town | Rural | 0.05 | 0.05 | -0.05 to 0.15 | 0.345 |
| | | 12590 | Residential setting | Large city | Rural | 0.03 | 0.08 | -0.12 to 0.19 | 0.676 |
| | | 12590 | Residential setting | Suburb | Rural | 0.04 | 0.12 | -0.19 to 0.28 | 0.718 |
| | | 12590 | *Threshold / cut-point* | Never\|Rarely | | -2.39 | 0.09 | -2.57 to -2.21 | 0.000 |
| | | 12590 | *Threshold / cut-point* | Rarely\|Often | | -1.05 | 0.09 | -1.22 to -0.88 | 0.000 |
| | | 12590 | *Threshold / cut-point* | Often\|Always | | -0.41 | 0.08 | -0.57 to -0.24 | 0.000 |
| **Support reducing differences in income levels§** | **Proportional-odds regression** | 12455 | Childhood predictor | Got by | Lived comfortably | -0.50 | 0.08 | -0.65 to -0.34 | 0.000 |
| | | 12455 | Childhood predictor | Found it difficult | Lived comfortably | -0.33 | 0.09 | -0.51 to -0.16 | 0.000 |
| | | 12455 | Childhood predictor | Found it very difficult | Lived comfortably | -0.53 | 0.11 | -0.75 to -0.31 | 0.000 |
| | | 12455 | Age | Per 1-year increase | | 0.00 | 0.00 | -0.01 to 0.00 | 0.192 |
| | | 12455 | Gender | Female | Male | -0.09 | 0.06 | -0.20 to 0.02 | 0.123 |
| | | 12455 | Education | Secondary | Elementary or less | -0.09 | 0.07 | -0.23 to 0.04 | 0.177 |
| | | 12455 | Education | Four years beyond HS | Elementary or less | -0.31 | 0.11 | -0.53 to -0.10 | 0.004 |
| | | 12455 | Residential setting | Small town | Rural | 0.03 | 0.13 | -0.21 to 0.28 | 0.782 |
| | | 12455 | Residential setting | Large city | Rural | 0.06 | 0.15 | -0.22 to 0.35 | 0.671 |
| | | 12455 | Residential setting | Suburb | Rural | -0.09 | 0.18 | -0.44 to 0.26 | 0.607 |
| | | 12455 | *Threshold / cut-point* | Strongly disagree\|Somewhat disagree | | -4.15 | 0.18 | -4.51 to -3.80 | 0.000 |
| | | 12455 | *Threshold / cut-point* | Somewhat disagree\|Neither | | -3.62 | 0.17 | -3.95 to -3.29 | 0.000 |
| | | 12455 | *Threshold / cut-point* | Neither\|Somewhat agree | | -3.16 | 0.16 | -3.48 to -2.85 | 0.000 |
| | | 12455 | *Threshold / cut-point* | Somewhat agree\|Strongly agree | | -1.59 | 0.14 | -1.87 to -1.31 | 0.000 |
| **Have little interest in doing things§** | **Proportional-odds regression** | 12578 | Childhood predictor | Got by | Lived comfortably | 0.11 | 0.05 | 0.00 to 0.22 | 0.041 |
| | | 12578 | Childhood predictor | Found it difficult | Lived comfortably | 0.15 | 0.06 | 0.03 to 0.27 | 0.013 |
| | | 12578 | Childhood predictor | Found it very difficult | Lived comfortably | 0.08 | 0.07 | -0.06 to 0.22 | 0.269 |
| | | 12578 | Age | Per 1-year increase | | 0.00 | 0.00 | 0.00 to 0.01 | 0.036 |
| | | 12578 | Gender | Female | Male | 0.04 | 0.04 | -0.04 to 0.12 | 0.322 |
| | | 12578 | Education | Secondary | Elementary or less | 0.08 | 0.05 | -0.02 to 0.17 | 0.100 |
| | | 12578 | Education | Four years beyond HS | Elementary or less | 0.11 | 0.08 | -0.04 to 0.26 | 0.162 |
| | | 12578 | Residential setting | Small town | Rural | 0.27 | 0.06 | 0.14 to 0.40 | 0.000 |

| Outcome | Model type | Analytic N | Variable | Level / comparison | Reference category | Coefficient | Standard error | 95% CI | p-value |
|---|---|---|---|---|---|---|---|---|---|
| | | 12578 | Residential setting | Large city | Rural | 0.12 | 0.08 | -0.04 to 0.27 | 0.138 |
| | | 12578 | Residential setting | Suburb | Rural | 0.26 | 0.15 | -0.03 to 0.54 | 0.082 |
| | | 12578 | *Threshold / cut-point* | Not at all\|Several days | | -0.41 | 0.09 | -0.58 to -0.24 | 0.000 |
| | | 12578 | *Threshold / cut-point* | Several days\|More than half the days | | 0.52 | 0.09 | 0.35 to 0.70 | 0.000 |
| | | 12578 | *Threshold / cut-point* | More than half the days\|Nearly every day | | 1.30 | 0.09 | 1.12 to 1.47 | 0.000 |
| **Various aspects of life are balanced§** | **Proportional-odds regression** | 12504 | Childhood predictor | Got by | Lived comfortably | 0.07 | 0.05 | -0.03 to 0.17 | 0.185 |
| | | 12504 | Childhood predictor | Found it difficult | Lived comfortably | 0.09 | 0.05 | -0.02 to 0.20 | 0.098 |
| | | 12504 | Childhood predictor | Found it very difficult | Lived comfortably | -0.01 | 0.06 | -0.13 to 0.11 | 0.811 |
| | | 12504 | Age | Per 1-year increase | | 0.00 | 0.00 | 0.00 to 0.01 | 0.067 |
| | | 12504 | Gender | Female | Male | -0.02 | 0.04 | -0.10 to 0.06 | 0.587 |
| | | 12504 | Education | Secondary | Elementary or less | 0.04 | 0.04 | -0.04 to 0.13 | 0.310 |
| | | 12504 | Education | Four years beyond HS | Elementary or less | 0.10 | 0.07 | -0.03 to 0.23 | 0.133 |
| | | 12504 | Residential setting | Small town | Rural | 0.11 | 0.05 | 0.00 to 0.21 | 0.047 |
| | | 12504 | Residential setting | Large city | Rural | 0.09 | 0.07 | -0.04 to 0.22 | 0.157 |
| | | 12504 | Residential setting | Suburb | Rural | 0.37 | 0.11 | 0.15 to 0.58 | 0.001 |
| | | 12504 | *Threshold / cut-point* | Never\|Rarely | | -1.98 | 0.09 | -2.16 to -1.81 | 0.000 |
| | | 12504 | *Threshold / cut-point* | Rarely\|Often | | 0.19 | 0.08 | 0.03 to 0.36 | 0.021 |
| | | 12504 | *Threshold / cut-point* | Often\|Always | | 0.98 | 0.08 | 0.82 to 1.15 | 0.000 |
| **Obeying the law§** | **Proportional-odds regression** | 12641 | Childhood predictor | Got by | Lived comfortably | -0.45 | 0.11 | -0.66 to -0.24 | 0.000 |
| | | 12641 | Childhood predictor | Found it difficult | Lived comfortably | -0.33 | 0.10 | -0.54 to -0.13 | 0.001 |
| | | 12641 | Childhood predictor | Found it very difficult | Lived comfortably | -0.50 | 0.14 | -0.78 to -0.23 | 0.000 |
| | | 12641 | Age | Per 1-year increase | | 0.00 | 0.00 | -0.01 to 0.00 | 0.747 |
| | | 12641 | Gender | Female | Male | -0.19 | 0.07 | -0.33 to -0.05 | 0.010 |
| | | 12641 | Education | Secondary | Elementary or less | 0.08 | 0.10 | -0.11 to 0.27 | 0.405 |
| | | 12641 | Education | Four years beyond HS | Elementary or less | -0.23 | 0.12 | -0.47 to 0.01 | 0.063 |
| | | 12641 | Residential setting | Small town | Rural | -0.06 | 0.15 | -0.34 to 0.23 | 0.703 |
| | | 12641 | Residential setting | Large city | Rural | -0.27 | 0.18 | -0.62 to 0.09 | 0.141 |
| | | 12641 | Residential setting | Suburb | Rural | -0.27 | 0.22 | -0.71 to 0.17 | 0.229 |
| | | 12641 | *Threshold / cut-point* | Strongly disagree\|Somewhat disagree | | -5.42 | 0.25 | -5.92 to -4.93 | 0.000 |
| | | 12641 | *Threshold / cut-point* | Somewhat disagree\|Neither | | -4.80 | 0.21 | -5.21 to -4.39 | 0.000 |
| | | 12641 | *Threshold / cut-point* | Neither\|Somewhat agree | | -4.28 | 0.20 | -4.68 to -3.88 | 0.000 |
| | | 12641 | *Threshold / cut-point* | Somewhat agree\|Strongly agree | | -2.46 | 0.18 | -2.82 to -2.10 | 0.000 |
| **Feel at peace§** | **Proportional-odds regression** | 12575 | Childhood predictor | Got by | Lived comfortably | 0.00 | 0.05 | -0.09 to 0.10 | 0.945 |
| | | 12575 | Childhood predictor | Found it difficult | Lived comfortably | -0.08 | 0.05 | -0.18 to 0.03 | 0.156 |
| | | 12575 | Childhood predictor | Found it very difficult | Lived comfortably | -0.08 | 0.06 | -0.20 to 0.05 | 0.238 |

| Outcome | Model type | Analytic N | Variable | Level / comparison | Reference category | Coefficient | Standard error | 95% CI | p-value |
|---|---|---|---|---|---|---|---|---|---|
| | | 12575 | Age | Per 1-year increase | | 0.01 | 0.00 | 0.00 to 0.01 | 0.000 |
| | | 12575 | Gender | Female | Male | 0.01 | 0.04 | -0.07 to 0.08 | 0.850 |
| | | 12575 | Education | Secondary | Elementary or less | 0.01 | 0.05 | -0.08 to 0.10 | 0.812 |
| | | 12575 | Education | Four years beyond HS | Elementary or less | 0.04 | 0.07 | -0.09 to 0.17 | 0.576 |
| | | 12575 | Residential setting | Small town | Rural | 0.20 | 0.05 | 0.10 to 0.30 | 0.000 |
| | | 12575 | Residential setting | Large city | Rural | 0.15 | 0.07 | 0.01 to 0.29 | 0.040 |
| | | 12575 | Residential setting | Suburb | Rural | 0.26 | 0.10 | 0.07 to 0.45 | 0.007 |
| | | 12575 | *Threshold / cut-point* | Never\|Rarely | | -1.90 | 0.09 | -2.06 to -1.73 | 0.000 |
| | | 12575 | *Threshold / cut-point* | Rarely\|Often | | -0.07 | 0.08 | -0.23 to 0.10 | 0.422 |
| | | 12575 | *Threshold / cut-point* | Often\|Always | | 0.63 | 0.08 | 0.46 to 0.79 | 0.000 |
| **Say in government#** | **Proportional-odds regression** | 12614 | Childhood predictor | Got by | Lived comfortably | 0.12 | 0.07 | -0.02 to 0.26 | 0.106 |
| | | 12614 | Childhood predictor | Found it difficult | Lived comfortably | -0.12 | 0.07 | -0.26 to 0.02 | 0.103 |
| | | 12614 | Childhood predictor | Found it very difficult | Lived comfortably | -0.26 | 0.09 | -0.43 to -0.08 | 0.003 |
| | | 12614 | Age | Per 1-year increase | | 0.00 | 0.00 | -0.01 to 0.00 | 0.580 |
| | | 12614 | Gender | Female | Male | 0.09 | 0.05 | -0.01 to 0.19 | 0.081 |
| | | 12614 | Education | Secondary | Elementary or less | -0.27 | 0.06 | -0.39 to -0.15 | 0.000 |
| | | 12614 | Education | Four years beyond HS | Elementary or less | -0.17 | 0.10 | -0.36 to 0.03 | 0.091 |
| | | 12614 | Residential setting | Small town | Rural | 0.01 | 0.08 | -0.14 to 0.15 | 0.946 |
| | | 12614 | Residential setting | Large city | Rural | -0.18 | 0.09 | -0.36 to 0.00 | 0.054 |
| | | 12614 | Residential setting | Suburb | Rural | -0.44 | 0.13 | -0.69 to -0.18 | 0.001 |
| | | 12614 | *Threshold / cut-point* | Disagree\|Unsure | | -1.84 | 0.12 | -2.07 to -1.60 | 0.000 |
| | | 12614 | *Threshold / cut-point* | Unsure\|Agree | | -1.43 | 0.11 | -1.66 to -1.21 | 0.000 |
| **Extent of suffering§** | **Proportional-odds regression** | 12589 | Childhood predictor | Got by | Lived comfortably | 0.23 | 0.05 | 0.13 to 0.34 | 0.000 |
| | | 12589 | Childhood predictor | Found it difficult | Lived comfortably | 0.24 | 0.06 | 0.13 to 0.35 | 0.000 |
| | | 12589 | Childhood predictor | Found it very difficult | Lived comfortably | 0.38 | 0.06 | 0.25 to 0.51 | 0.000 |
| | | 12589 | Age | Per 1-year increase | | 0.02 | 0.00 | 0.01 to 0.02 | 0.000 |
| | | 12589 | Gender | Female | Male | 0.30 | 0.04 | 0.22 to 0.38 | 0.000 |
| | | 12589 | Education | Secondary | Elementary or less | -0.13 | 0.05 | -0.22 to -0.04 | 0.007 |
| | | 12589 | Education | Four years beyond HS | Elementary or less | -0.25 | 0.07 | -0.39 to -0.10 | 0.001 |
| | | 12589 | Residential setting | Small town | Rural | 0.05 | 0.06 | -0.06 to 0.16 | 0.383 |
| | | 12589 | Residential setting | Large city | Rural | -0.11 | 0.08 | -0.26 to 0.04 | 0.143 |
| | | 12589 | Residential setting | Suburb | Rural | 0.06 | 0.10 | -0.14 to 0.26 | 0.561 |
| | | 12589 | *Threshold / cut-point* | Not at all\|Not very much | | 0.45 | 0.09 | 0.28 to 0.62 | 0.000 |
| | | 12589 | *Threshold / cut-point* | Not very much\|Some | | 0.87 | 0.09 | 0.70 to 1.04 | 0.000 |
| | | 12589 | *Threshold / cut-point* | Some\|A lot | | 2.31 | 0.09 | 2.13 to 2.48 | 0.000 |
| **Distress from life-threatening experiences§** | **Proportional-odds regression** | 12608 | Childhood predictor | Got by | Lived comfortably | 0.05 | 0.06 | -0.06 to 0.16 | 0.351 |
| | | 12608 | Childhood predictor | Found it difficult | Lived comfortably | 0.14 | 0.06 | 0.02 to 0.25 | 0.018 |
| | | 12608 | Childhood predictor | Found it very difficult | Lived comfortably | 0.29 | 0.06 | 0.17 to 0.41 | 0.000 |

| Outcome | Model type | Analytic N | Variable | Level / comparison | Reference category | Coefficient | Standard error | 95% CI | p-value |
|---|---|---|---|---|---|---|---|---|---|
| | | 12608 | Age | Per 1-year increase | | 0.00 | 0.00 | 0.00 to 0.01 | 0.067 |
| | | 12608 | Gender | Female | Male | 0.10 | 0.04 | 0.01 to 0.18 | 0.031 |
| | | 12608 | Education | Secondary | Elementary or less | -0.23 | 0.05 | -0.32 to -0.14 | 0.000 |
| | | 12608 | Education | Four years beyond HS | Elementary or less | -0.41 | 0.07 | -0.55 to -0.26 | 0.000 |
| | | 12608 | Residential setting | Small town | Rural | -0.18 | 0.05 | -0.27 to -0.08 | 0.000 |
| | | 12608 | Residential setting | Large city | Rural | -0.23 | 0.07 | -0.37 to -0.08 | 0.002 |
| | | 12608 | Residential setting | Suburb | Rural | -0.22 | 0.12 | -0.44 to 0.01 | 0.060 |
| | | 12608 | *Threshold / cut-point* | Not at all\|Not very much | | -0.22 | 0.08 | -0.38 to -0.06 | 0.006 |
| | | 12608 | *Threshold / cut-point* | Not very much\|Some | | 0.12 | 0.08 | -0.04 to 0.28 | 0.135 |
| | | 12608 | *Threshold / cut-point* | Some\|A lot | | 1.13 | 0.08 | 0.97 to 1.29 | 0.000 |
| **Trust in others§** | **Proportional-odds regression** | 12320 | Childhood predictor | Got by | Lived comfortably | -0.05 | 0.05 | -0.15 to 0.05 | 0.310 |
| | | 12320 | Childhood predictor | Found it difficult | Lived comfortably | 0.01 | 0.06 | -0.10 to 0.12 | 0.832 |
| | | 12320 | Childhood predictor | Found it very difficult | Lived comfortably | -0.20 | 0.07 | -0.33 to -0.07 | 0.003 |
| | | 12320 | Age | Per 1-year increase | | 0.00 | 0.00 | 0.00 to 0.01 | 0.008 |
| | | 12320 | Gender | Female | Male | -0.05 | 0.04 | -0.13 to 0.03 | 0.219 |
| | | 12320 | Education | Secondary | Elementary or less | -0.06 | 0.05 | -0.15 to 0.03 | 0.220 |
| | | 12320 | Education | Four years beyond HS | Elementary or less | -0.09 | 0.07 | -0.23 to 0.06 | 0.237 |
| | | 12320 | Residential setting | Small town | Rural | 0.23 | 0.05 | 0.13 to 0.33 | 0.000 |
| | | 12320 | Residential setting | Large city | Rural | 0.00 | 0.07 | -0.13 to 0.14 | 0.965 |
| | | 12320 | Residential setting | Suburb | Rural | -0.18 | 0.14 | -0.45 to 0.10 | 0.213 |
| | | 12320 | *Threshold / cut-point* | None\|Not very many | | -2.17 | 0.09 | -2.35 to -1.98 | 0.000 |
| | | 12320 | *Threshold / cut-point* | Not very many\|Some | | -1.38 | 0.09 | -1.56 to -1.20 | 0.000 |
| | | 12320 | *Threshold / cut-point* | Some\|Most | | 0.82 | 0.09 | 0.64 to 0.99 | 0.000 |
| | | 12320 | *Threshold / cut-point* | Most\|All | | 1.79 | 0.09 | 1.61 to 1.97 | 0.000 |

**eTable 15.** Demographic statistics of unweighted sample sizes (n) and weighted proportions (%) using responses collected in the Wave 1 survey (2023) in India and stratified by gender and survey mode (N = 12,765).

| Characteristics | Wave 1 | Female | Male | Telephone | Web |
|---|---|---|---|---|---|
| **Age group (years)** | n (%) | n (%) | n (%) | n (%) | n (%) |
| 18-24 | 2,335 (19.9) | 1,228 (19.4) | 1,107 (20.4) | 2,248 (19.7) | 87 (33.7) |
| 25-29 | 1,933 (12.8) | 1,102 (12.5) | 831 (13.2) | 1,899 (12.8) | 34 (15.3) |
| 30-39 | 3,598 (24.4) | 2,120 (24.9) | 1,478 (23.8) | 3,546 (24.4) | 52 (24.3) |
| 40-49 | 2,365 (17.8) | 1,261 (18.1) | 1,104 (17.5) | 2,340 (17.9) | 25 (13.6) |
| 50-59 | 1,326 (12.3) | 621 (12.5) | 705 (12.2) | 1,310 (12.3) | 16 (11.0) |
| 60-69 | 876 (9.3) | 380 (9.5) | 496 (9.1) | 874 (9.4) | 2 (2.1) |
| 70-79 | 285 (2.9) | 108 (2.5) | 177 (3.2) | 285 (2.9) | 0 (0.0) |
| 80 or older | 47 (0.5) | 23 (0.6) | 24 (0.5) | 47 (0.5) | 0 (0.0) |
| **Gender** | | | | | |
| Female | 6,843 (49.3) | | | 6,762 (49.6) | 81 (28.0) |
| Male | 5,922 (50.7) | | | 5,787 (50.4) | 135 (72.0) |
| **Current marital status** | | | | | |
| Single, never married | 1,970 (16.2) | 630 (8.5) | 1,340 (23.6) | 1,890 (16.0) | 80 (32.7) |
| Married | 10,082 (77.1) | 5,696 (81.7) | 4,386 (72.7) | 9,966 (77.4) | 116 (56.3) |
| Separated | 36 (0.4) | 23 (0.4) | 13 (0.3) | 34 (0.3) | 2 (2.7) |
| Divorced | 20 (0.2) | 11 (0.2) | 9 (0.2) | 19 (0.2) | 1 (0.5) |
| Widowed | 368 (3.5) | 307 (6.2) | 61 (0.9) | 365 (3.5) | 3 (0.9) |
| Domestic partner | 229 (2.1) | 143 (2.4) | 86 (1.8) | 222 (2.1) | 7 (2.3) |
| Saw, skipped | 4 (0.0) | 1 (0.0) | 3 (0.0) | 0 (0.0) | 4 (1.8) |
| Don't know | 30 (0.3) | 18 (0.3) | 12 (0.3) | 30 (0.3) | 0 (0.0) |
| Refused | 23 (0.2) | 13 (0.2) | 10 (0.1) | 23 (0.2) | 0 (0.0) |
| Missing (NAs) | 3 (0.0) | 1 (0.0) | 2 (0.1) | 0 (0.0) | 3 (2.8) |
| **Employment status** | | | | | |
| Employed for an employer | 2,365 (20.8) | 849 (13.5) | 1,516 (28.0) | 2,316 (20.7) | 49 (28.5) |
| Self-employed | 3,145 (26.6) | 852 (13.2) | 2,293 (39.7) | 3,088 (26.6) | 57 (29.3) |
| Retired | 214 (2.2) | 51 (1.2) | 163 (3.2) | 213 (2.3) | 1 (1.1) |
| Student | 722 (4.2) | 319 (3.3) | 403 (5.0) | 676 (4.0) | 46 (15.5) |
| Homemaker | 4,624 (33.1) | 4,261 (61.0) | 363 (5.9) | 4,584 (33.3) | 40 (15.8) |
| Unemployed and looking for a job | 966 (7.1) | 286 (3.8) | 680 (10.2) | 945 (7.0) | 21 (8.6) |
| None of these/other | 683 (5.6) | 197 (3.5) | 486 (7.7) | 681 (5.7) | 2 (1.1) |
| Don't know | 39 (0.3) | 26 (0.4) | 13 (0.2) | 39 (0.3) | 0 (0.0) |
| Refused | 7 (0.1) | 2 (0.0) | 5 (0.1) | 7 (0.1) | 0 (0.0) |
| **Education (in years)** | | | | | |
| Up to eight years of basic education | 7,684 (68.6) | 4,555 (75.3) | 3,129 (62.1) | 7,648 (69.2) | 36 (25.9) |
| 9-15 years of education | 3,886 (25.3) | 1,777 (20.0) | 2,109 (30.4) | 3,780 (24.9) | 106 (50.6) |
| 16+ years of education (4-year degree or beyond) | 1,189 (6.1) | 508 (4.7) | 681 (7.5) | 1,115 (5.9) | 74 (23.5) |
| Don't know | 6 (0.0) | 3 (0.0) | 3 (0.0) | 6 (0.0) | 0 (0.0) |
| **Birth country** | | | | | |
| Born in this country | 12,637 (98.9) | 6,761 (98.7) | 5,876 (99.2) | 12,421 (98.9) | 216 (100.0) |
| Born in another country | 101 (0.9) | 57 (0.9) | 44 (0.8) | 101 (0.9) | 0 (0.0) |

| | | | | | |
|---|---|---|---|---|---|
| Don't know | 25 (0.2) | 24 (0.3) | 1 (0.0) | 25 (0.2) | 0 (0.0) |
| Refused | 2 (0.0) | 1 (0.0) | 1 (0.0) | 2 (0.0) | 0 (0.0) |
| **Residential setting** | | | | | |
| A rural area or on a farm | 5,003 (34.9) | 2,594 (34.2) | 2,409 (35.6) | 4,932 (35.1) | 71 (23.1) |
| A small town or village | 5,181 (41.4) | 2,711 (40.9) | 2,470 (41.9) | 5,127 (41.6) | 54 (23.1) |
| A large city | 1,996 (18.3) | 1,189 (19.2) | 807 (17.5) | 1,933 (18.1) | 63 (35.8) |
| A suburb of a large city | 585 (5.4) | 349 (5.7) | 236 (5.0) | 557 (5.2) | 28 (18.0) |

***Note:*** Cross-sectional Wave 1 weights were used for weight-adjusted proportions (%).

**eTable 16.** Table of coefficients for adjusted associations between childhood abuse and 14 outcomes from India in the mid-year survey (February-May 2024).

Survey-weighted models use RETENTION_WEIGHT_C with primary sampling unit (PSU) and strata and adjust for Wave 1 age, gender, education, and residential setting. Childhood abuse is coded Yes versus No (reference). Coefficients are beta coefficients for continuous outcomes, log-odds coefficients for binary outcomes, and proportional-odds log coefficients for ordinal outcomes. Model intercepts are displayed for continuous and binary outcomes. Proportional-odds models do not have a single conventional intercept; their estimated ordinal cut-points/thresholds are displayed after the covariate coefficients, with standard errors (SEs), 95% CIs, and Wald p-values.

***Note:*** All results except p-values are rounded to the two decimals. P-values are rounded to three decimals. Values displayed as 0.000 indicate *P* < 0.0005 due to rounding, rather than an exact zero. Statistical significance was determined using unrounded P-values at the Bonferroni-adjusted threshold of *P* < 0.0036.

| Outcome | Model type | Analytic N | Variable | Level / comparison | Reference category | Coefficient | Standard error | 95% CI | p-value |
|---|---|---|---|---|---|---|---|---|---|
| **Is diligent** | **Linear regression** | 7760 | *Intercept* | | | 7.97 | 0.18 | 7.61 to 8.32 | 0.000 |
| | | 7760 | Childhood abuse | Yes | No | -0.28 | 0.15 | -0.58 to 0.01 | 0.062 |
| | | 7760 | Age | Per 1-year increase | | -0.01 | 0.00 | -0.02 to 0.00 | 0.014 |
| | | 7760 | Gender | Female | Male | -0.28 | 0.09 | -0.46 to -0.09 | 0.003 |
| | | 7760 | Education | Secondary | Elementary or less | 0.25 | 0.11 | 0.03 to 0.47 | 0.027 |
| | | 7760 | Education | Four years beyond HS | Elementary or less | 0.58 | 0.13 | 0.33 to 0.83 | 0.000 |
| | | 7760 | Residential setting | Small town | Rural | 0.29 | 0.11 | 0.07 to 0.51 | 0.010 |
| | | 7760 | Residential setting | Large city | Rural | 0.33 | 0.13 | 0.07 to 0.59 | 0.014 |
| | | 7760 | Residential setting | Suburb | Rural | 0.46 | 0.24 | -0.01 to 0.94 | 0.056 |
| **Is important to be a good person** | **Linear regression** | 7765 | *Intercept* | | | 8.31 | 0.17 | 7.99 to 8.63 | 0.000 |
| | | 7765 | Childhood abuse | Yes | No | -0.37 | 0.15 | -0.66 to -0.08 | 0.013 |
| | | 7765 | Age | Per 1-year increase | | -0.01 | 0.00 | -0.01 to 0.00 | 0.057 |
| | | 7765 | Gender | Female | Male | 0.02 | 0.08 | -0.15 to 0.18 | 0.818 |
| | | 7765 | Education | Secondary | Elementary or less | 0.55 | 0.10 | 0.35 to 0.74 | 0.000 |
| | | 7765 | Education | Four years beyond HS | Elementary or less | 0.90 | 0.12 | 0.67 to 1.13 | 0.000 |
| | | 7765 | Residential setting | Small town | Rural | -0.04 | 0.10 | -0.25 to 0.17 | 0.704 |
| | | 7765 | Residential setting | Large city | Rural | 0.09 | 0.16 | -0.21 to 0.40 | 0.551 |
| | | 7765 | Residential setting | Suburb | Rural | -0.01 | 0.17 | -0.35 to 0.32 | 0.936 |
| **Is important to have good relationships** | **Linear regression** | 7768 | *Intercept* | | | 8.08 | 0.18 | 7.73 to 8.43 | 0.000 |
| | | 7768 | Childhood abuse | Yes | No | -0.48 | 0.15 | -0.77 to -0.20 | 0.001 |
| | | 7768 | Age | Per 1-year increase | | 0.00 | 0.00 | -0.01 to 0.00 | 0.187 |
| | | 7768 | Gender | Female | Male | 0.21 | 0.09 | 0.03 to 0.38 | 0.021 |
| | | 7768 | Education | Secondary | Elementary or less | 0.44 | 0.11 | 0.23 to 0.65 | 0.000 |
| | | 7768 | Education | Four years beyond HS | Elementary or less | 0.78 | 0.13 | 0.54 to 1.03 | 0.000 |
| | | 7768 | Residential setting | Small town | Rural | -0.01 | 0.11 | -0.22 to 0.20 | 0.935 |
| | | 7768 | Residential setting | Large city | Rural | 0.22 | 0.14 | -0.04 to 0.49 | 0.102 |
| | | 7768 | Residential setting | Suburb | Rural | -0.32 | 0.23 | -0.78 to 0.14 | 0.175 |
| **Is important to be happy** | **Linear regression** | 7764 | *Intercept* | | | 7.91 | 0.17 | 7.59 to 8.24 | 0.000 |
| | | 7764 | Childhood abuse | Yes | No | -0.44 | 0.13 | -0.70 to -0.18 | 0.001 |
| | | 7764 | Age | Per 1-year increase | | -0.02 | 0.00 | -0.02 to -0.01 | 0.000 |
| | | 7764 | Gender | Female | Male | 0.09 | 0.09 | -0.09 to 0.28 | 0.314 |

| Outcome | Model type | Analytic N | Variable | Level / comparison | Reference category | Coefficient | Standard error | 95% CI | p-value |
|---|---|---|---|---|---|---|---|---|---|
| | | 7764 | Education | Secondary | Elementary or less | 0.42 | 0.09 | 0.24 to 0.60 | 0.000 |
| | | 7764 | Education | Four years beyond HS | Elementary or less | 0.71 | 0.11 | 0.48 to 0.93 | 0.000 |
| | | 7764 | Residential setting | Small town | Rural | 0.26 | 0.10 | 0.07 to 0.46 | 0.009 |
| | | 7764 | Residential setting | Large city | Rural | 0.41 | 0.13 | 0.16 to 0.66 | 0.001 |
| | | 7764 | Residential setting | Suburb | Rural | 0.52 | 0.19 | 0.15 to 0.88 | 0.006 |
| **Is important to be healthy** | **Linear regression** | 7758 | *Intercept* | | | 8.16 | 0.19 | 7.78 to 8.53 | 0.000 |
| | | 7758 | Childhood abuse | Yes | No | -0.57 | 0.13 | -0.82 to -0.32 | 0.000 |
| | | 7758 | Age | Per 1-year increase | | -0.02 | 0.00 | -0.02 to -0.01 | 0.000 |
| | | 7758 | Gender | Female | Male | -0.16 | 0.09 | -0.33 to 0.00 | 0.056 |
| | | 7758 | Education | Secondary | Elementary or less | 0.61 | 0.10 | 0.43 to 0.80 | 0.000 |
| | | 7758 | Education | Four years beyond HS | Elementary or less | 1.09 | 0.12 | 0.86 to 1.32 | 0.000 |
| | | 7758 | Residential setting | Small town | Rural | 0.06 | 0.12 | -0.17 to 0.28 | 0.633 |
| | | 7758 | Residential setting | Large city | Rural | 0.38 | 0.14 | 0.12 to 0.65 | 0.005 |
| | | 7758 | Residential setting | Suburb | Rural | 0.13 | 0.19 | -0.25 to 0.51 | 0.509 |
| **Is important to have a meaningful life** | **Linear regression** | 7729 | *Intercept* | | | 7.88 | 0.17 | 7.54 to 8.21 | 0.000 |
| | | 7729 | Childhood abuse | Yes | No | -0.26 | 0.15 | -0.56 to 0.03 | 0.080 |
| | | 7729 | Age | Per 1-year increase | | -0.01 | 0.00 | -0.02 to -0.01 | 0.000 |
| | | 7729 | Gender | Female | Male | -0.05 | 0.10 | -0.24 to 0.14 | 0.627 |
| | | 7729 | Education | Secondary | Elementary or less | 0.49 | 0.11 | 0.28 to 0.70 | 0.000 |
| | | 7729 | Education | Four years beyond HS | Elementary or less | 0.85 | 0.13 | 0.60 to 1.09 | 0.000 |
| | | 7729 | Residential setting | Small town | Rural | -0.04 | 0.12 | -0.27 to 0.19 | 0.732 |
| | | 7729 | Residential setting | Large city | Rural | 0.20 | 0.14 | -0.08 to 0.48 | 0.166 |
| | | 7729 | Residential setting | Suburb | Rural | 0.15 | 0.22 | -0.28 to 0.57 | 0.498 |
| **Mind is focused on what to do** | **Linear regression** | 7743 | *Intercept* | | | 8.14 | 0.16 | 7.83 to 8.44 | 0.000 |
| | | 7743 | Childhood abuse | Yes | No | -0.33 | 0.15 | -0.62 to -0.04 | 0.025 |
| | | 7743 | Age | Per 1-year increase | | -0.02 | 0.00 | -0.02 to -0.01 | 0.000 |
| | | 7743 | Gender | Female | Male | -0.08 | 0.10 | -0.27 to 0.11 | 0.412 |
| | | 7743 | Education | Secondary | Elementary or less | 0.23 | 0.09 | 0.04 to 0.42 | 0.016 |
| | | 7743 | Education | Four years beyond HS | Elementary or less | 0.48 | 0.13 | 0.22 to 0.73 | 0.000 |
| | | 7743 | Residential setting | Small town | Rural | 0.13 | 0.12 | -0.10 to 0.36 | 0.270 |
| | | 7743 | Residential setting | Large city | Rural | 0.41 | 0.13 | 0.15 to 0.67 | 0.002 |
| | | 7743 | Residential setting | Suburb | Rural | 0.46 | 0.21 | 0.05 to 0.86 | 0.027 |
| **Is important to have plenty of money to have what is wanted** | **Linear regression** | 7744 | *Intercept* | | | 7.17 | 0.19 | 6.79 to 7.55 | 0.000 |
| | | 7744 | Childhood abuse | Yes | No | -0.25 | 0.17 | -0.58 to 0.09 | 0.148 |
| | | 7744 | Age | Per 1-year increase | | -0.01 | 0.00 | -0.02 to 0.00 | 0.003 |
| | | 7744 | Gender | Female | Male | 0.26 | 0.11 | 0.04 to 0.47 | 0.019 |
| | | 7744 | Education | Secondary | Elementary or less | 0.19 | 0.12 | -0.05 to 0.43 | 0.116 |
| | | 7744 | Education | Four years beyond HS | Elementary or less | 0.37 | 0.16 | 0.05 to 0.69 | 0.023 |
| | | 7744 | Residential setting | Small town | Rural | 0.00 | 0.14 | -0.29 to 0.28 | 0.974 |

| Outcome | Model type | Analytic N | Variable | Level / comparison | Reference category | Coefficient | Standard error | 95% CI | p-value |
|---|---|---|---|---|---|---|---|---|---|
| | | 7744 | Residential setting | Large city | Rural | 0.23 | 0.15 | -0.06 to 0.52 | 0.119 |
| | | 7744 | Residential setting | Suburb | Rural | 0.25 | 0.28 | -0.31 to 0.80 | 0.384 |
| **Feel very connected to nature** | **Linear regression** | 7719 | *Intercept* | | | 6.81 | 0.22 | 6.38 to 7.24 | 0.000 |
| | | 7719 | Childhood abuse | Yes | No | -0.28 | 0.17 | -0.62 to 0.06 | 0.104 |
| | | 7719 | Age | Per 1-year increase | | 0.00 | 0.00 | -0.01 to 0.01 | 0.943 |
| | | 7719 | Gender | Female | Male | -0.23 | 0.10 | -0.42 to -0.04 | 0.020 |
| | | 7719 | Education | Secondary | Elementary or less | 0.47 | 0.12 | 0.24 to 0.70 | 0.000 |
| | | 7719 | Education | Four years beyond HS | Elementary or less | 0.72 | 0.16 | 0.42 to 1.02 | 0.000 |
| | | 7719 | Residential setting | Small town | Rural | 0.05 | 0.14 | -0.22 to 0.31 | 0.720 |
| | | 7719 | Residential setting | Large city | Rural | 0.19 | 0.17 | -0.14 to 0.52 | 0.261 |
| | | 7719 | Residential setting | Suburb | Rural | -0.12 | 0.29 | -0.69 to 0.46 | 0.691 |
| **Happy with what you achieve in life** | **Logistic regression** | 7768 | *Intercept* | | | 2.28 | 0.18 | 1.92 to 2.63 | 0.000 |
| | | 7768 | Childhood abuse | Yes | No | -0.09 | 0.14 | -0.37 to 0.20 | 0.544 |
| | | 7768 | Age | Per 1-year increase | | -0.01 | 0.00 | -0.02 to -0.01 | 0.000 |
| | | 7768 | Gender | Female | Male | 0.21 | 0.09 | 0.04 to 0.38 | 0.019 |
| | | 7768 | Education | Secondary | Elementary or less | 0.14 | 0.10 | -0.07 to 0.34 | 0.190 |
| | | 7768 | Education | Four years beyond HS | Elementary or less | 0.13 | 0.16 | -0.19 to 0.45 | 0.420 |
| | | 7768 | Residential setting | Small town | Rural | -0.01 | 0.12 | -0.24 to 0.22 | 0.930 |
| | | 7768 | Residential setting | Large city | Rural | -0.18 | 0.14 | -0.45 to 0.09 | 0.194 |
| | | 7768 | Residential setting | Suburb | Rural | -0.58 | 0.24 | -1.06 to -0.11 | 0.016 |
| **Regularly experience things considered beautiful** | **Logistic regression** | 7737 | *Intercept* | | | 1.12 | 0.14 | 0.85 to 1.39 | 0.000 |
| | | 7737 | Childhood abuse | Yes | No | 0.19 | 0.10 | -0.01 to 0.40 | 0.064 |
| | | 7737 | Age | Per 1-year increase | | -0.01 | 0.00 | -0.02 to -0.01 | 0.000 |
| | | 7737 | Gender | Female | Male | -0.36 | 0.07 | -0.49 to -0.22 | 0.000 |
| | | 7737 | Education | Secondary | Elementary or less | 0.40 | 0.07 | 0.26 to 0.54 | 0.000 |
| | | 7737 | Education | Four years beyond HS | Elementary or less | 0.54 | 0.12 | 0.31 to 0.78 | 0.000 |
| | | 7737 | Residential setting | Small town | Rural | -0.13 | 0.08 | -0.29 to 0.03 | 0.107 |
| | | 7737 | Residential setting | Large city | Rural | -0.29 | 0.12 | -0.52 to -0.06 | 0.014 |
| | | 7737 | Residential setting | Suburb | Rural | -0.20 | 0.16 | -0.52 to 0.12 | 0.222 |
| **Often engage in arts or cultural activities** | **Proportional-odds regression** | 7731 | Childhood abuse | Yes | No | -0.04 | 0.09 | -0.23 to 0.14 | 0.653 |
| | | 7731 | Age | Per 1-year increase | | -0.01 | 0.00 | -0.02 to -0.01 | 0.000 |
| | | 7731 | Gender | Female | Male | -0.47 | 0.07 | -0.60 to -0.34 | 0.000 |
| | | 7731 | Education | Secondary | Elementary or less | 0.26 | 0.07 | 0.13 to 0.39 | 0.000 |
| | | 7731 | Education | Four years beyond HS | Elementary or less | 0.52 | 0.09 | 0.35 to 0.70 | 0.000 |
| | | 7731 | Residential setting | Small town | Rural | -0.28 | 0.07 | -0.42 to -0.14 | 0.000 |
| | | 7731 | Residential setting | Large city | Rural | -0.30 | 0.11 | -0.50 to -0.09 | 0.005 |
| | | 7731 | Residential setting | Suburb | Rural | -0.26 | 0.21 | -0.66 to 0.15 | 0.213 |
| | | 7731 | *Threshold / cut-point* | Never\|A few times a year | | -0.34 | 0.12 | -0.58 to -0.10 | 0.006 |

| Outcome | Model type | Analytic N | Variable | Level / comparison | Reference category | Coefficient | Standard error | 95% CI | p-value |
|---|---|---|---|---|---|---|---|---|---|
| | | 7731 | *Threshold / cut-point* | A few times a year\|One to three times a month | | 0.49 | 0.12 | 0.26 to 0.72 | 0.000 |
| | | 7731 | *Threshold / cut-point* | One to three times a month\|Once a week | | 1.29 | 0.12 | 1.05 to 1.52 | 0.000 |
| | | 7731 | *Threshold / cut-point* | Once a week\|More than once a week | | 2.14 | 0.14 | 1.87 to 2.40 | 0.000 |
| **Household ran out of food in last 12 months** | **Proportional-odds regression** | 7775 | Childhood abuse | Yes | No | -0.06 | 0.10 | -0.24 to 0.13 | 0.563 |
| | | 7775 | Age | Per 1-year increase | | 0.00 | 0.00 | 0.00 to 0.01 | 0.419 |
| | | 7775 | Gender | Female | Male | 0.14 | 0.07 | 0.02 to 0.27 | 0.028 |
| | | 7775 | Education | Secondary | Elementary or less | -0.28 | 0.07 | -0.43 to -0.14 | 0.000 |
| | | 7775 | Education | Four years beyond HS | Elementary or less | -0.54 | 0.10 | -0.74 to -0.34 | 0.000 |
| | | 7775 | Residential setting | Small town | Rural | 0.09 | 0.08 | -0.06 to 0.24 | 0.244 |
| | | 7775 | Residential setting | Large city | Rural | -0.12 | 0.10 | -0.32 to 0.09 | 0.270 |
| | | 7775 | Residential setting | Suburb | Rural | 0.17 | 0.19 | -0.21 to 0.55 | 0.378 |
| | | 7775 | *Threshold / cut-point* | Never\|Sometimes | | 0.02 | 0.14 | -0.24 to 0.29 | 0.858 |
| | | 7775 | *Threshold / cut-point* | Sometimes\|Often | | 1.99 | 0.15 | 1.71 to 2.28 | 0.000 |
| **Time spent on using social media each day** | **Proportional-odds regression** | 7615 | Childhood abuse | Yes | No | 0.04 | 0.09 | -0.13 to 0.20 | 0.679 |
| | | 7615 | Age | Per 1-year increase | | -0.06 | 0.00 | -0.07 to -0.06 | 0.000 |
| | | 7615 | Gender | Female | Male | -1.41 | 0.07 | -1.54 to -1.28 | 0.000 |
| | | 7615 | Education | Secondary | Elementary or less | 0.33 | 0.07 | 0.19 to 0.46 | 0.000 |
| | | 7615 | Education | Four years beyond HS | Elementary or less | 0.96 | 0.09 | 0.78 to 1.14 | 0.000 |
| | | 7615 | Residential setting | Small town | Rural | -0.17 | 0.07 | -0.31 to -0.02 | 0.024 |
| | | 7615 | Residential setting | Large city | Rural | 0.19 | 0.11 | -0.03 to 0.40 | 0.093 |
| | | 7615 | Residential setting | Suburb | Rural | 0.37 | 0.12 | 0.14 to 0.59 | 0.002 |
| | | 7615 | *Threshold / cut-point* | None\|Less than 30 minutes | | -2.90 | 0.13 | -3.16 to -2.63 | 0.000 |
| | | 7615 | *Threshold / cut-point* | Less than 30 minutes\|30 minutes to less than an hour | | -2.05 | 0.13 | -2.31 to -1.79 | 0.000 |
| | | 7615 | *Threshold / cut-point* | 30 minutes to less than an hour\|1 to 2 hours | | -1.42 | 0.13 | -1.69 to -1.16 | 0.000 |
| | | 7615 | *Threshold / cut-point* | 1 to 2 hours\|More than 2 hours | | -0.15 | 0.13 | -0.41 to 0.11 | 0.254 |

**eTable 17.** Distribution of childhood predictors collected in Wave 1 survey (2023) in India and stratified by survey mode and residential location (N = 12,765).

| Childhood Predictors (8) | Wave 1 | Telephone | Web | Large city or suburb of a large city | Small town or village | Rural area or on a farm |
|---|---|---|---|---|---|---|
| ***Ever physically or sexually abused when growing up*** | n (%) | n (%) | n (%) | n (%) | n (%) | n (%) |
| Yes | 1446 (11.5) | 1422 (11.4) | 24 (15.4) | 248 (10.0) | 611 (11.8) | 587 (12.1) |
| No | 10637 (82.5) | 10456 (82.5) | 181 (79.0) | 2211 (83.8) | 4255 (81.4) | 4171 (82.8) |
| Saw, skipped | 2 (0.0) | 0 (0.0) | 2 (1.0) | 1 (0.0) | 1 (0.0) | 0 (0.0) |
| Don't know | 598 (5.4) | 598 (5.5) | 0 (0.0) | 103 (5.5) | 282 (6.1) | 213 (4.6) |
| Refused | 73 (0.6) | 73 (0.6) | 0 (0.0) | 13 (0.4) | 30 (0.6) | 30 (0.5) |
| Missing (NAs) | 9 (0.1) | 0 (0.0) | 9 (4.6) | 5 (0.2) | 2 (0.0) | 2 (0.0) |
| ***Feeling like an outsider in one's family when growing up*** | | | | | | |
| Yes | 1903 (15.1) | 1870 (15.2) | 33 (10.2) | 324 (12.8) | 721 (14.6) | 858 (17.2) |
| No | 10792 (84.5) | 10609 (84.4) | 183 (89.8) | 2253 (87.1) | 4428 (85.0) | 4111 (82.0) |
| Does not apply | 18 (0.1) | 18 (0.1) | 0 (0.0) | 1 (0.0) | 11 (0.2) | 6 (0.1) |
| Don't know | 43 (0.3) | 43 (0.3) | 0 (0.0) | 2 (0.0) | 17 (0.2) | 24 (0.5) |
| Refused | 9 (0.0) | 9 (0.0) | 0 (0.0) | 1 (0.0) | 4 (0.0) | 4 (0.1) |
| ***Feeling loved by mother*** | | | | | | |
| Yes | 12264 (95.8) | 12054 (95.8) | 210 (98.0) | 2475 (95.3) | 4965 (95.7) | 4824 (96.3) |
| No | 165 (1.4) | 161 (1.4) | 4 (1.1) | 47 (2.0) | 61 (1.3) | 57 (1.1) |
| Does not apply | 270 (2.3) | 270 (2.4) | 0 (0.0) | 53 (2.5) | 119 (2.4) | 98 (2.1) |
| Don't know | 60 (0.5) | 59 (0.5) | 1 (0.3) | 5 (0.2) | 36 (0.7) | 19 (0.4) |
| Refused | 6 (0.0) | 5 (0.0) | 1 (0.5) | 1 (0.0) | 0 (0.0) | 5 (0.1) |
| ***Feeling loved by father*** | | | | | | |
| Yes | 11884 (92.5) | 11678 (92.5) | 206 (95.4) | 2364 (90.8) | 4824 (92.8) | 4696 (93.3) |
| No | 270 (2.3) | 264 (2.3) | 6 (2.7) | 89 (3.7) | 100 (2.1) | 81 (1.6) |
| Does not apply | 493 (4.2) | 491 (4.2) | 2 (0.7) | 104 (4.6) | 204 (4.0) | 185 (4.1) |
| Don't know | 102 (0.9) | 100 (0.9) | 2 (1.2) | 21 (0.8) | 48 (1.0) | 33 (0.7) |
| Refused | 16 (0.1) | 16 (0.1) | 0 (0.0) | 3 (0.1) | 5 (0.0) | 8 (0.2) |
| ***Quality of relationship with mother*** | | | | | | |
| Very good | 11530 (89.8) | 11338 (89.8) | 192 (90.8) | 2364 (90.8) | 4719 (90.4) | 4447 (88.5) |
| Somewhat good | 761 (6.2) | 742 (6.2) | 19 (7.4) | 129 (5.3) | 263 (5.7) | 369 (7.3) |
| Somewhat bad | 84 (0.7) | 82 (0.7) | 2 (0.6) | 11 (0.5) | 34 (0.6) | 39 (1.0) |
| Very bad | 56 (0.6) | 55 (0.6) | 1 (0.3) | 20 (0.9) | 22 (0.6) | 14 (0.3) |
| Does not apply | 241 (2.1) | 241 (2.1) | 0 (0.0) | 47 (2.2) | 104 (2.1) | 90 (2.0) |
| Don't know | 87 (0.6) | 86 (0.6) | 1 (0.3) | 9 (0.3) | 39 (0.6) | 39 (0.8) |
| Refused | 6 (0.0) | 5 (0.0) | 1 (0.5) | 1 (0.0) | 0 (0.0) | 5 (0.1) |
| ***Quality of relationship with father*** | | | | | | |
| Very good | 11022 (85.6) | 10834 (85.5) | 188 (89.6) | 2227 (84.9) | 4512 (86.2) | 4283 (85.2) |
| Somewhat good | 985 (7.8) | 964 (7.8) | 21 (7.5) | 177 (7.4) | 369 (7.5) | 439 (8.3) |
| Somewhat bad | 104 (1.0) | 101 (1.0) | 3 (1.0) | 26 (1.1) | 42 (1.0) | 36 (0.9) |
| Very bad | 74 (0.8) | 74 (0.8) | 0 (0.0) | 29 (1.4) | 27 (0.7) | 18 (0.5) |
| Does not apply | 450 (3.8) | 448 (3.8) | 2 (0.7) | 96 (4.2) | 178 (3.5) | 176 (3.9) |

| | | | | | | |
|---|---|---|---|---|---|---|
| Don't know | 117 (1.0) | 115 (1.0) | 2 (1.2) | 23 (1.0) | 50 (1.0) | 44 (1.0) |
| Refused | 13 (0.1) | 13 (0.1) | 0 (0.0) | 3 (0.1) | 3 (0.1) | 7 (0.2) |
| ***Self-rated health while growing up*** | | | | | | |
| Excellent | 2092 (17.1) | 2040 (17.0) | 52 (25.5) | 536 (20.3) | 914 (18.1) | 642 (13.7) |
| Very good | 3857 (30.4) | 3774 (30.3) | 83 (37.8) | 892 (34.2) | 1628 (31.2) | 1,337 (26.9) |
| Good | 3988 (31.6) | 3931 (31.6) | 57 (26.8) | 717 (29.5) | 1668 (32.1) | 1,603 (32.3) |
| Fair | 2357 (17.2) | 2337 (17.4) | 20 (7.9) | 369 (13.3) | 795 (15.2) | 1,193 (22.4) |
| Poor | 426 (3.3) | 422 (3.3) | 4 (2.0) | 62 (2.6) | 158 (3.0) | 206 (4.2) |
| Don't know | 42 (0.3) | 42 (0.3) | 0 (0.0) | 4 (0.1) | 18 (0.4) | 20 (0.4) |
| Refused | 3 (0.0) | 3 (0.0) | 0 (0.0) | 1 (0.1) | 0 (0.0) | 2 (0.0) |
| ***Perceived family financial situation while growing up*** | | | | | | |
| Lived comfortably | 5275 (38.7) | 5152 (38.5) | 123 (57.1) | 1183 (42.9) | 2138 (38.2) | 1954 (36.6) |
| Got by | 2940 (23.6) | 2881 (23.6) | 59 (23.1) | 592 (23.4) | 1161 (23.1) | 1187 (24.3) |
| Found it difficult | 2539 (21.2) | 2520 (21.3) | 19 (12.0) | 473 (19.5) | 1168 (23.8) | 898 (19.2) |
| Found it very difficult | 1940 (15.9) | 1925 (16.0) | 15 (7.8) | 326 (13.8) | 695 (14.5) | 919 (19.1) |
| Don't know | 67 (0.5) | 67 (0.5) | 0 (0.0) | 6 (0.4) | 19 (0.4) | 42 (0.8) |
| Refused | 4 (0.0) | 4 (0.0) | 0 (0.0) | 1 (0.0) | 0 (0.0) | 3 (0.0) |

***Note:*** Cross-sectional Wave 1 weights were used for weight-adjusted proportions (%).

**eTable 18.** Distribution of outcome variables collected in Wave 1 survey (2023) in India and stratified by survey mode and residential location (N = 12,765).

| **Outcomes and flourishing composite indices** | **Wave 1** | **Telephone** | **Web** | **Large city or suburb of a large city** | **Small town or village** | **Rural area or on a farm** |
|---|---|---|---|---|---|---|
| | Mean (sd) | Mean (sd) | Mean (sd) | Mean (sd) | Mean (sd) | Mean (sd) |
| ***Belonging*** | 8.41 (2.9) | 8.40 (2.9) | 9.04 (1.8) | 8.53 (2.8) | 8.54 (2.8) | 8.16 (3.0) |
| ***Flourishing (with financial indicators)*** | 6.89 (1.9) | 6.88 (1.9) | 7.65 (1.6) | 7.03 (1.9) | 7.00 (1.9) | 6.65 (1.9) |
| ***Flourishing (without financial indicators)*** | 7.45 (2.0) | 7.44 (2.0) | 8.10 (1.7) | 7.57 (2.0) | 7.63 (2.0) | 7.16 (2.1) |
| ***Freedom*** | 8.15 (3.0) | 8.15 (3.0) | 8.64 (2.1) | 8.30 (2.9) | 8.28 (2.9) | 7.90 (3.0) |
| ***Happiness***‡ | 6.48 (3.6) | 6.47 (3.6) | 7.32 (2.6) | 6.57 (3.5) | 6.64 (3.6) | 6.22 (3.6) |
| ***Life evaluation five years from now*** | 7.30 (3.1) | 7.29 (3.1) | 8.15 (2.5) | 7.52 (3.0) | 7.46 (3.1) | 6.97 (3.1) |
| ***Life evaluation today*** | 5.63 (3.6) | 5.61 (3.6) | 6.56 (3.0) | 5.62 (3.5) | 5.75 (3.6) | 5.49 (3.6) |
| ***Life satisfaction***‡ | 7.00 (3.5) | 7.00 (3.5) | 7.63 (2.4) | 7.03 (3.5) | 7.14 (3.5) | 6.82 (3.4) |
| ***Life worthwhile***‡ | 6.93 (3.4) | 6.92 (3.4) | 7.76 (2.5) | 7.13 (3.3) | 7.09 (3.4) | 6.61 (3.4) |
| ***Loneliness***§ | 3.84 (4.0) | 3.84 (4.0) | 3.28 (3.3) | 3.62 (4.0) | 3.72 (4.0) | 4.13 (4.0) |
| ***Positive expectations*** | 8.14 (2.9) | 8.14 (2.9) | 8.46 (2.2) | 8.34 (2.8) | 8.30 (2.8) | 7.82 (3.0) |
| ***Relationship contentment***‡ | 7.96 (3.1) | 7.95 (3.1) | 8.63 (2.0) | 7.84 (3.3) | 8.17 (3.0) | 7.79 (3.1) |
| ***Relationship satisfaction***‡ | 7.92 (3.1) | 7.91 (3.1) | 8.47 (2.1) | 7.96 (3.1) | 8.11 (3.0) | 7.67 (3.1) |
| ***Remain hopeful for the future*** | 8.32 (2.8) | 8.31 (2.8) | 8.73 (2.0) | 8.51 (2.7) | 8.49 (2.6) | 7.98 (3.0) |
| ***Understand purpose in life***‡ | 8.04 (3.0) | 8.03 (3.0) | 8.68 (1.9) | 8.25 (2.9) | 8.18 (3.0) | 7.73 (3.1) |
| ***Mental health***‡ | 7.44 (3.3) | 7.43 (3.3) | 8.11 (2.3) | 7.62 (3.2) | 7.58 (3.3) | 7.14 (3.3) |
| ***Physical health***‡ | 7.00 (3.4) | 6.99 (3.4) | 7.86 (2.4) | 7.26 (3.2) | 7.16 (3.4) | 6.63 (3.5) |
| ***Act to promote good***‡ | 8.35 (2.7) | 8.35 (2.7) | 8.61 (2.1) | 8.59 (2.5) | 8.56 (2.5) | 7.93 (2.9) |
| ***Give up some happiness now for greater happiness later***‡ | 7.21 (3.5) | 7.19 (3.5) | 8.35 (2.3) | 7.20 (3.6) | 7.40 (3.4) | 6.98 (3.5) |
| ***Have a long list of gratitude*** | 7.69 (3.1) | 7.68 (3.2) | 8.35 (2.2) | 7.89 (3.0) | 7.82 (3.1) | 7.38 (3.2) |
| ***Show love often*** | 8.20 (3.0) | 8.21 (3.0) | 8.00 (2.9) | 8.14 (3.2) | 8.32 (3.0) | 8.11 (3.0) |
| ***Perceived support from others*** | 6.46 (3.9) | 6.45 (3.9) | 7.61 (2.7) | 6.09 (4.0) | 6.67 (3.9) | 6.47 (3.8) |
| ***Worry about meeting normal monthly living expenses***§§‡‡ | 5.89 (4.1) | 5.91 (4.1) | 4.34 (3.7) | 5.60 (4.1) | 6.05 (4.1) | 5.89 (4.0) |
| ***Worry about safety, food, or housing***§§‡‡ | 6.02 (4.1) | 6.04 (4.1) | 4.06 (3.7) | 5.79 (4.2) | 6.24 (4.1) | 5.91 (4.1) |
| | n (%) | n (%) | n (%) | n (%) | n (%) | n (%) |
| ***Alcohol use*** | | | | | | |
| Yes | 828 (7.5) | 807 (7.5) | 21 (7.8) | 180 (8.8) | 351 (7.6) | 297 (6.6) |
| No | 11404 (88.3) | 11246 (88.5) | 158 (77.4) | 2255 (86.3) | 4595 (87.9) | 4554 (90.2) |
| Saw, skipped | 36 (0.2) | 0 (0.0) | 36 (13.5) | 18 (0.4) | 8 (0.1) | 10 (0.1) |
| Don't know | 111 (0.9) | 111 (0.9) | 0 (0.0) | 30 (1.1) | 45 (1.0) | 36 (0.7) |
| Refused | 385 (3.0) | 385 (3.1) | 0 (0.0) | 97 (3.3) | 182 (3.4) | 106 (2.5) |
| Missing (NAs) | 1 (0.0) | 0 (0.0) | 1 (1.3) | 1 (0.1) | 0 (0.0) | 0 (0.0) |
| ***Cigarette smoking*** | | | | | | |
| Yes | 843 (7.6) | 823 (7.6) | 20 (10.5) | 170 (8.6) | 315 (6.9) | 358 (7.7) |
| No | 11404 (88.4) | 11242 (88.5) | 162 (76.0) | 2268 (86.5) | 4636 (88.8) | 4500 (89.2) |
| Saw, skipped | 33 (0.2) | 0 (0.0) | 33 (12.1) | 15 (0.3) | 8 (0.1) | 10 (0.1) |
| Don't know | 87 (0.7) | 87 (0.8) | 0 (0.0) | 28 (1.0) | 36 (0.8) | 23 (0.5) |

| | | | | | | |
|---|---|---|---|---|---|---|
| Refused | 397 (3.1) | 397 (3.1) | 0 (0.0) | 99 (3.4) | 186 (3.4) | 112 (2.5) |
| Missing (NAs) | 1 (0.0) | 0 (0.0) | 1 (1.3) | 1 (0.1) | 0 (0.0) | 0 (0.0) |
| ***Have any health problems*** | | | | | | |
| Yes | 3476 (28.1) | 3415 (28.1) | 61 (29.5) | 652 (25.3) | 1489 (29.9) | 1335 (27.9) |
| No | 9188 (71.0) | 9039 (71.1) | 149 (66.6) | 1898 (73.5) | 3657 (69.2) | 3633 (71.5) |
| Saw, skipped | 1 (0.0) | 0 (0.0) | 1 (0.3) | 0 (0.0) | 0 (0.0) | 1 (0.0) |
| Don't know | 77 (0.7) | 77 (0.7) | 0 (0.0) | 20 (0.8) | 31 (0.8) | 26 (0.5) |
| Refused | 18 (0.1) | 18 (0.1) | 0 (0.0) | 8 (0.2) | 3 (0.1) | 7 (0.1) |
| Missing (NAs) | 5 (0.0) | 0 (0.0) | 5 (3.5) | 3 (0.2) | 1 (0.0) | 1 (0.0) |
| ***Donated to charity*** | | | | | | |
| Yes | 4972 (38.6) | 4889 (38.6) | 83 (41.7) | 828 (32.6) | 2107 (39.6) | 2037 (41.5) |
| No | 7732 (61.0) | 7599 (61.0) | 133 (58.3) | 1739 (66.9) | 3050 (60.0) | 2943 (58.1) |
| Don't know | 60 (0.4) | 60 (0.4) | 0 (0.0) | 14 (0.5) | 24 (0.4) | 22 (0.4) |
| Refused | 1 (0.0) | 1 (0.0) | 0 (0.0) | 0 (0.0) | 0 (0.0) | 1 (0.0) |
| ***Helped a stranger*** | | | | | | |
| Yes | 6889 (55.1) | 6760 (55.0) | 129 (62.0) | 1309 (51.8) | 2909 (57.2) | 2671 (54.7) |
| No | 5716 (43.8) | 5631 (43.9) | 85 (37.1) | 1243 (47.0) | 2213 (41.8) | 2260 (43.9) |
| Don't know | 159 (1.1) | 157 (1.1) | 2 (0.9) | 29 (1.1) | 59 (1.0) | 71 (1.3) |
| Refused | 1 (0.0) | 1 (0.0) | 0 (0.0) | 0 (0.0) | 0 (0.0) | 1 (0.0) |
| ***Volunteered at an organization*** | | | | | | |
| Yes | 3921 (29.0) | 3833 (28.8) | 88 (43.0) | 591 (22.3) | 1695 (30.1) | 1635 (32.3) |
| No | 8766 (70.4) | 8639 (70.5) | 127 (56.6) | 1986 (77.6) | 3453 (69.2) | 3327 (66.8) |
| Don't know | 76 (0.6) | 75 (0.6) | 1 (0.4) | 4 (0.1) | 32 (0.7) | 40 (0.8) |
| Refused | 2 (0.0) | 2 (0.0) | 0 (0.0) | 0 (0.0) | 1 (0.0) | 1 (0.0) |
| ***Feel close to someone*** | | | | | | |
| Yes | 10392 (81.6) | 10216 (81.6) | 176 (79.7) | 2061 (80.1) | 4254 (82.7) | 4077 (81.3) |
| No | 2347 (18.2) | 2307 (18.2) | 40 (20.3) | 519 (19.9) | 912 (17.0) | 916 (18.5) |
| Don't know | 24 (0.2) | 24 (0.2) | 0 (0.0) | 0 (0.0) | 15 (0.3) | 9 (0.2) |
| Refused | 2 (0.0) | 2 (0.0) | 0 (0.0) | 1 (0.0) | 0 (0.0) | 1 (0.0) |
| ***Feel at peace***§ | | | | | | |
| Always | 5334 (43.3) | 5241 (43.3) | 93 (45.2) | 1070 (43.8) | 2324 (45.5) | 1940 (40.3) |
| Often | 2196 (16.8) | 2115 (16.6) | 81 (33.7) | 511 (18.4) | 820 (16.2) | 865 (16.4) |
| Rarely | 3966 (29.5) | 3930 (29.7) | 36 (19.0) | 763 (28.2) | 1542 (27.9) | 1661 (32.4) |
| Never | 1154 (9.4) | 1150 (9.5) | 4 (1.3) | 217 (8.7) | 450 (9.4) | 487 (9.9) |
| Saw, skipped | 2 (0.0) | 0 (0.0) | 2 (0.8) | 1 (0.0) | 1 (0.0) | 0 (0.0) |
| Don't know | 106 (0.9) | 106 (0.9) | 0 (0.0) | 17 (0.9) | 41 (0.9) | 48 (1.0) |
| Refused | 7 (0.0) | 7 (0.0) | 0 (0.0) | 2 (0.0) | 3 (0.0) | 2 (0.0) |
| ***Various aspects of life are balanced***§ | | | | | | |
| Always | 3942 (31.8) | 3862 (31.7) | 80 (38.6) | 818 (33.0) | 1628 (32.1) | 1496 (30.6) |
| Often | 2346 (18.5) | 2256 (18.3) | 90 (39.3) | 545 (20.0) | 935 (19.0) | 866 (17.0) |
| Rarely | 5082 (38.5) | 5045 (38.8) | 37 (16.4) | 957 (36.3) | 2066 (38.1) | 2059 (40.4) |
| Never | 1210 (9.7) | 1203 (9.8) | 7 (3.5) | 229 (9.4) | 483 (9.4) | 498 (10.3) |
| Saw, skipped | 2 (0.0) | 0 (0.0) | 2 (2.1) | 1 (0.1) | 0 (0.0) | 1 (0.0) |
| Don't know | 169 (1.4) | 169 (1.4) | 0 (0.0) | 27 (1.0) | 64 (1.3) | 78 (1.7) |
| Refused | 14 (0.1) | 14 (0.1) | 0 (0.0) | 4 (0.1) | 5 (0.1) | 5 (0.1) |

| | | | | | | |
|---|---|---|---|---|---|---|
| ***Depressive symptoms*** § | | | | | | |
| Nearly every day | 1757 (14.1) | 1717 (14.0) | 40 (20.6) | 324 (12.8) | 731 (14.7) | 702 (14.3) |
| More than half the days | 2114 (16.7) | 2066 (16.6) | 48 (21.1) | 419 (16.5) | 935 (17.8) | 760 (15.5) |
| Several days | 3422 (25.5) | 3363 (25.5) | 59 (22.7) | 682 (24.3) | 1254 (23.4) | 1486 (28.8) |
| Not at all | 5366 (42.9) | 5297 (43.0) | 69 (35.5) | 1138 (45.7) | 2217 (43.4) | 2011 (40.5) |
| Don't know | 88 (0.7) | 88 (0.7) | 0 (0.0) | 13 (0.6) | 38 (0.7) | 37 (0.8) |
| Refused | 18 (0.1) | 18 (0.1) | 0 (0.0) | 5 (0.2) | 6 (0.1) | 7 (0.2) |
| ***Distress from life-threatening experiences*** § | | | | | | |
| A lot | 3219 (25.1) | 3178 (25.2) | 41 (19.7) | 595 (23.4) | 1248 (23.6) | 1376 (28.0) |
| Some | 2955 (22.5) | 2877 (22.3) | 78 (35.2) | 582 (21.6) | 1209 (23.2) | 1164 (22.3) |
| Not very much | 1111 (8.3) | 1071 (8.2) | 40 (16.4) | 190 (6.7) | 438 (8.1) | 483 (9.7) |
| Not at all | 5400 (43.4) | 5343 (43.6) | 57 (28.8) | 1193 (47.4) | 2255 (44.4) | 1952 (39.4) |
| Don't know | 64 (0.5) | 64 (0.5) | 0 (0.0) | 13 (0.6) | 27 (0.6) | 24 (0.5) |
| Refused | 16 (0.1) | 16 (0.1) | 0 (0.0) | 8 (0.4) | 4 (0.1) | 4 (0.1) |
| ***Extent of suffering*** § | | | | | | |
| A lot | 2418 (20.4) | 2383 (20.5) | 35 (16.0) | 476 (19.4) | 973 (20.7) | 969 (20.8) |
| Some | 4000 (30.4) | 3930 (30.4) | 70 (33.3) | 792 (29.3) | 1664 (31.6) | 1544 (29.8) |
| Not very much | 1342 (9.8) | 1288 (9.7) | 54 (24.1) | 241 (8.9) | 512 (9.7) | 589 (10.6) |
| Not at all | 4906 (38.5) | 4850 (38.7) | 56 (26.4) | 1056 (41.7) | 1999 (37.3) | 1851 (37.9) |
| Saw, skipped | 1 (0.0) | 0 (0.0) | 1 (0.3) | 0 (0.0) | 1 (0.0) | 0 (0.0) |
| Don't know | 86 (0.6) | 86 (0.6) | 0 (0.0) | 12 (0.5) | 29 (0.6) | 45 (0.8) |
| Refused | 12 (0.1) | 12 (0.1) | 0 (0.0) | 4 (0.3) | 3 (0.1) | 5 (0.1) |
| ***Feeling nervous or anxious*** § | | | | | | |
| Nearly every day | 1890 (15.3) | 1854 (15.2) | 36 (20.3) | 356 (14.0) | 815 (16.4) | 719 (14.8) |
| More than half the days | 1982 (15.9) | 1939 (15.8) | 43 (20.6) | 410 (16.1) | 895 (17.3) | 677 (13.9) |
| Several days | 3158 (23.6) | 3103 (23.6) | 55 (20.9) | 602 (21.9) | 1165 (21.6) | 1391 (27.0) |
| Not at all | 5660 (44.6) | 5580 (44.7) | 80 (37.7) | 1197 (47.1) | 2277 (44.0) | 2186 (43.7) |
| Saw, skipped | 2 (0.0) | 0 (0.0) | 2 (0.6) | 1 (0.0) | 0 (0.0) | 1 (0.0) |
| Don't know | 56 (0.5) | 56 (0.5) | 0 (0.0) | 8 (0.4) | 23 (0.6) | 25 (0.5) |
| Refused | 17 (0.2) | 17 (0.2) | 0 (0.0) | 7 (0.4) | 6 (0.1) | 4 (0.1) |
| ***Have little interest in doing things*** § | | | | | | |
| Nearly every day | 3428 (28.6) | 3361 (28.5) | 67 (33.3) | 686 (28.7) | 1526 (31.3) | 1216 (25.4) |
| More than half the days | 2147 (17.7) | 2090 (17.6) | 57 (23.3) | 435 (17.6) | 942 (19.1) | 770 (16.2) |
| Several days | 2959 (21.9) | 2910 (21.9) | 49 (20.3) | 597 (21.9) | 1109 (20.0) | 1253 (24.2) |
| Not at all | 4119 (30.9) | 4079 (31.0) | 40 (22.3) | 838 (30.8) | 1564 (29.0) | 1717 (33.3) |
| Saw, skipped | 3 (0.0) | 0 (0.0) | 3 (0.8) | 1 (0.0) | 0 (0.0) | 2 (0.0) |
| Don't know | 91 (0.7) | 91 (0.8) | 0 (0.0) | 17 (0.8) | 37 (0.7) | 37 (0.8) |
| Refused | 18 (0.1) | 18 (0.1) | 0 (0.0) | 7 (0.2) | 3 (0.0) | 8 (0.2) |
| ***Unable to control worry*** § | | | | | | |
| Nearly every day | 2071 (16.4) | 2024 (16.4) | 47 (21.8) | 402 (15.4) | 873 (17.0) | 796 (16.4) |
| More than half the days | 1873 (14.8) | 1820 (14.7) | 53 (23.6) | 405 (15.6) | 828 (16.1) | 640 (12.6) |
| Several days | 2852 (21.4) | 2811 (21.4) | 41 (17.8) | 527 (19.5) | 1052 (19.4) | 1273 (25.0) |
| Not at all | 5842 (46.4) | 5769 (46.5) | 73 (36.3) | 1218 (48.2) | 2380 (46.5) | 2244 (45.0) |
| Saw, skipped | 2 (0.0) | 0 (0.0) | 2 (0.6) | 0 (0.0) | 1 (0.0) | 1 (0.0) |
| Don't know | 83 (0.7) | 83 (0.7) | 0 (0.0) | 15 (0.6) | 34 (0.8) | 34 (0.6) |

| | | | | | | |
|---|---|---|---|---|---|---|
| Refused | 42 (0.4) | 42 (0.4) | 0 (0.0) | 14 (0.6) | 13 (0.2) | 15 (0.4) |
| ***Feeling capable*** § | | | | | | |
| Always | 6778 (54.1) | 6675 (54.1) | 103 (50.9) | 1418 (56.5) | 2852 (55.3) | 2508 (51.1) |
| Often | 2002 (15.3) | 1927 (15.1) | 75 (28.4) | 428 (14.7) | 767 (15.3) | 807 (15.6) |
| Rarely | 3012 (22.7) | 2987 (22.9) | 25 (12.1) | 552 (21.3) | 1189 (21.7) | 1271 (24.8) |
| Never | 895 (7.2) | 886 (7.2) | 9 (6.6) | 167 (6.7) | 341 (6.9) | 387 (7.9) |
| Saw, skipped | 4 (0.0) | 0 (0.0) | 4 (2.1) | 1 (0.0) | 3 (0.1) | 0 (0.0) |
| Don't know | 66 (0.6) | 66 (0.7) | 0 (0.0) | 14 (0.7) | 28 (0.7) | 24 (0.5) |
| Refused | 8 (0.0) | 8 (0.0) | 0 (0.0) | 1 (0.1) | 1 (0.0) | 6 (0.1) |
| ***Forgiveness*** § | | | | | | |
| Always | 7691 (60.7) | 7580 (60.8) | 111 (55.1) | 1549 (61.2) | 3179 (61.1) | 2963 (60.0) |
| Often | 1786 (13.7) | 1719 (13.5) | 67 (27.0) | 382 (13.6) | 704 (13.6) | 700 (13.8) |
| Rarely | 2258 (16.8) | 2230 (16.8) | 28 (15.0) | 428 (15.9) | 872 (15.9) | 958 (18.3) |
| Never | 931 (8.0) | 923 (8.0) | 8 (1.7) | 208 (8.8) | 375 (8.2) | 348 (7.1) |
| Saw, skipped | 2 (0.0) | 0 (0.0) | 2 (1.2) | 0 (0.0) | 1 (0.0) | 1 (0.0) |
| Don't know | 48 (0.4) | 48 (0.4) | 0 (0.0) | 7 (0.2) | 19 (0.4) | 22 (0.4) |
| Refused | 49 (0.5) | 49 (0.5) | 0 (0.0) | 7 (0.3) | 31 (0.8) | 11 (0.3) |
| ***Government approval*** § | | | | | | |
| Strongly approve | 7024 (54.9) | 6934 (55.1) | 90 (41.2) | 1300 (50.6) | 3035 (57.6) | 2689 (54.7) |
| Somewhat approve | 3640 (27.6) | 3571 (27.5) | 69 (32.8) | 748 (27.6) | 1315 (25.6) | 1577 (29.9) |
| Neither approve nor disapprove | 477 (4.1) | 447 (3.9) | 30 (16.6) | 128 (5.4) | 196 (3.9) | 153 (3.3) |
| Somewhat disapprove | 648 (5.1) | 633 (5.1) | 15 (5.4) | 178 (6.5) | 257 (4.9) | 213 (4.4) |
| Strongly disapprove | 735 (6.1) | 724 (6.1) | 11 (3.2) | 165 (6.8) | 284 (5.9) | 286 (6.0) |
| Saw, skipped | 1 (0.0) | 0 (0.0) | 1 (0.8) | 0 (0.0) | 1 (0.0) | 0 (0.0) |
| Don't know | 195 (1.9) | 195 (1.9) | 0 (0.0) | 53 (2.8) | 74 (1.7) | 68 (1.4) |
| Refused | 45 (0.3) | 45 (0.4) | 0 (0.0) | 9 (0.4) | 19 (0.3) | 17 (0.3) |
| ***Obeying the law*** § | | | | | | |
| Strongly agree | 11179 (87.3) | 11003 (87.3) | 176 (85.2) | 2235 (85.7) | 4558 (87.7) | 4386 (87.8) |
| Somewhat agree | 1270 (10.0) | 1235 (9.9) | 35 (13.4) | 279 (11.2) | 489 (9.5) | 502 (9.7) |
| Neither agree nor disagree | 98 (0.9) | 97 (0.9) | 1 (0.3) | 25 (1.0) | 34 (0.8) | 39 (0.8) |
| Somewhat disagree | 67 (0.6) | 65 (0.6) | 2 (0.8) | 11 (0.5) | 31 (0.7) | 25 (0.6) |
| Strongly disagree | 88 (0.7) | 86 (0.7) | 2 (0.4) | 26 (1.3) | 45 (0.8) | 17 (0.2) |
| Don't know | 61 (0.5) | 61 (0.5) | 0 (0.0) | 4 (0.2) | 24 (0.5) | 33 (0.8) |
| Refused | 2 (0.0) | 2 (0.0) | 0 (0.0) | 1 (0.1) | 0 (0.0) | 1 (0.0) |
| ***Perceived discrimination*** § | | | | | | |
| Always | 1653 (12.9) | 1601 (12.7) | 52 (25.2) | 290 (10.8) | 667 (13.2) | 696 (13.9) |
| Often | 1129 (8.6) | 1079 (8.4) | 50 (18.1) | 241 (8.7) | 405 (7.8) | 483 (9.4) |
| Rarely | 2897 (20.9) | 2842 (20.8) | 55 (27.4) | 516 (19.4) | 1121 (19.4) | 1260 (23.8) |
| Never | 6912 (56.1) | 6857 (56.5) | 55 (26.9) | 1494 (59.3) | 2916 (58.0) | 2502 (51.7) |
| Saw, skipped | 4 (0.0) | 0 (0.0) | 4 (2.4) | 0 (0.0) | 2 (0.0) | 2 (0.0) |
| Don't know | 137 (1.2) | 137 (1.3) | 0 (0.0) | 26 (1.3) | 58 (1.4) | 53 (1.0) |
| Refused | 33 (0.3) | 33 (0.3) | 0 (0.0) | 14 (0.5) | 12 (0.2) | 7 (0.2) |
| ***Say in government*** # | | | | | | |
| Agree | 9873 (77.3) | 9732 (77.4) | 141 (63.3) | 1891 (73.4) | 4074 (78.3) | 3908 (78.6) |
| Disagree | 2077 (16.1) | 2040 (16.1) | 37 (16.6) | 479 (18.3) | 786 (14.9) | 812 (16.0) |

| | | | | | | |
|---|---|---|---|---|---|---|
| Unsure | 741 (6.1) | 707 (5.9) | 34 (17.9) | 187 (7.4) | 298 (6.3) | 256 (5.0) |
| Saw, skipped | 4 (0.0) | 0 (0.0) | 4 (2.2) | 4 (0.1) | 0 (0.0) | 0 (0.0) |
| Refused | 70 (0.6) | 70 (0.6) | 0 (0.0) | 20 (0.8) | 23 (0.5) | 27 (0.5) |
| ***Support reducing differences in income levels*** § | | | | | | |
| Strongly agree | 9548 (74.1) | 9392 (74.1) | 156 (74.4) | 1957 (75.2) | 3924 (74.4) | 3667 (73.0) |
| Somewhat agree | 2250 (17.8) | 2197 (17.7) | 53 (22.5) | 451 (17.4) | 884 (17.8) | 915 (18.0) |
| Neither agree nor disagree | 237 (2.2) | 234 (2.2) | 3 (1.3) | 37 (1.6) | 102 (2.5) | 98 (2.3) |
| Somewhat disagree | 217 (1.7) | 215 (1.7) | 2 (1.3) | 40 (1.5) | 93 (1.6) | 84 (1.9) |
| Strongly disagree | 260 (2.4) | 259 (2.5) | 1 (0.2) | 72 (3.4) | 96 (2.1) | 92 (2.2) |
| Don't know | 248 (1.8) | 247 (1.8) | 1 (0.4) | 22 (0.9) | 81 (1.6) | 145 (2.7) |
| Refused | 5 (0.0) | 5 (0.0) | 0 (0.0) | 2 (0.1) | 1 (0.0) | 2 (0.0) |
| ***Trust in others*** § | | | | | | |
| All | 1919 (16.2) | 1878 (16.1) | 41 (22.8) | 339 (14.5) | 877 (18.3) | 703 (14.8) |
| Most | 2225 (17.3) | 2154 (17.1) | 71 (29.0) | 427 (16.1) | 974 (18.1) | 824 (17.0) |
| Some | 6001 (46.3) | 5929 (46.5) | 72 (31.8) | 1226 (47.0) | 2375 (45.3) | 2400 (47.0) |
| Not very much | 1154 (8.5) | 1127 (8.4) | 27 (14.3) | 260 (9.3) | 407 (7.3) | 487 (9.2) |
| None at all | 1097 (8.6) | 1093 (8.7) | 4 (1.8) | 250 (9.5) | 388 (7.5) | 459 (9.3) |
| Saw, skipped | 1 (0.0) | 0 (0.0) | 1 (0.3) | 0 (0.0) | 1 (0.0) | 0 (0.0) |
| Don't know | 321 (2.8) | 321 (2.9) | 0 (0.0) | 70 (3.2) | 135 (2.9) | 116 (2.5) |
| Refused | 47 (0.4) | 47 (0.4) | 0 (0.0) | 9 (0.4) | 24 (0.6) | 14 (0.2) |

***Note:***

- Cross-sectional Wave 1 weights were used for weight-adjusted proportions (%).
- Those marked with ‡ are included in the flourishing composite without financial indicators, and ‡‡ are the financial indicators included in the flourishing composite with financial indicators.
- The outcome variables marked with § were reverse coded in the association analysis.
- The outcome variable *say in government* marked with # was coded in the order by level 1 = disagree, level 2 = unsure, and level 3 = agree.
- The outcome variables (*worry about meeting normal monthly living expenses* and *worry about safety, food, or housing*) marked §§ were reverse coded in the association analysis and they are reverse coded for the table standalone reporting here. Within flourishing index calculations, *worry about meeting normal monthly living expenses* and *worry about safety, food, or housing* are on original scales. For binary outcomes, responses with “No” were recoded as 0; responses with “Yes” were recoded as 1 in the association analysis.

**eTable 19.** Distribution of childhood predictors collected in Wave 1 survey (2023) in India and stratified by age groups 18 to 24 inclusive, 25 to 29 inclusive, 30 to 39 inclusive, 40 to 49 inclusive, 50 to 59 inclusive, and 60+ (N = 12,765).

| **Childhood Predictors (8)** | **Wave 1** | **18 to 24** | **25 to 29** | **30 to 39** | **40 to 49** | **50 to 59** | **60+** |
|---|---|---|---|---|---|---|---|
| ***Ever physically or sexually abused when growing up*** | n (%) | n (%) | n (%) | n (%) | n (%) | n (%) | n (%) |
| Yes | 1446 (11.5) | 253 (11.9) | 180 (9.9) | 402 (11.8) | 291 (12.1) | 175 (11.6) | 145 (11.0) |
| No | 10637 (82.5) | 1973 (82.8) | 1649 (84.1) | 3038 (83.3) | 1931 (81.2) | 1070 (81.4) | 976 (81.5) |
| Saw, skipped | 2 (0.0) | 0 (0.0) | 1 (0.1) | 1 (0.0) | 0 (0.0) | 0 (0.0) | 0 (0.0) |
| Don't know | 598 (5.4) | 91 (4.6) | 88 (5.2) | 138 (4.5) | 131 (6.2) | 73 (6.2) | 77 (6.8) |
| Refused | 73 (0.6) | 14 (0.7) | 13 (0.6) | 18 (0.3) | 10 (0.4) | 8 (0.8) | 10 (0.7) |
| Missing (NAs) | 9 (0.1) | 4 (0.1) | 2 (0.1) | 1 (0.1) | 2 (0.1) | 0 (0.0) | 0 (0.0) |
| ***Feeling like an outsider in one's family when growing up*** | | | | | | | |
| Yes | 1903 (15.1) | 347 (14.6) | 333 (17.6) | 525 (14.3) | 315 (13.9) | 203 (16.9) | 180 (14.9) |
| No | 10792 (84.5) | 1982 (85.1) | 1585 (81.7) | 3049 (85.1) | 2035 (85.6) | 1120 (83.0) | 1021 (84.6) |
| Does not apply | 18 (0.1) | 0 (0.0) | 2 (0.1) | 8 (0.2) | 6 (0.2) | 1 (0.1) | 1 (0.1) |
| Don't know | 43 (0.3) | 5 (0.2) | 11 (0.6) | 13 (0.3) | 8 (0.3) | 1 (0.1) | 5 (0.4) |
| Refused | 9 (0.0) | 1 (0.0) | 2 (0.0) | 3 (0.1) | 1 (0.0) | 1 (0.0) | 1 (0.0) |
| ***Feeling loved by mother*** | | | | | | | |
| Yes | 12264 (95.8) | 2283 (97.5) | 1858 (96.1) | 3485 (97.0) | 2263 (95.4) | 1241 (93.0) | 1134 (93.7) |
| No | 165 (1.4) | 26 (1.1) | 28 (1.5) | 38 (1.1) | 36 (1.6) | 21 (1.9) | 16 (1.2) |
| Does not apply | 270 (2.3) | 19 (1.0) | 32 (1.6) | 60 (1.5) | 54 (2.3) | 53 (4.5) | 52 (4.7) |
| Don't know | 60 (0.5) | 6 (0.3) | 13 (0.7) | 14 (0.4) | 12 (0.6) | 10 (0.6) | 5 (0.4) |
| Refused | 6 (0.0) | 1 (0.0) | 2 (0.1) | 1 (0.0) | 0 (0.0) | 1 (0.0) | 1 (0.0) |
| ***Feeling loved by father*** | | | | | | | |
| Yes | 11884 (92.5) | 2217 (94.3) | 1815 (93.2) | 3377 (93.7) | 2205 (93.0) | 1194 (89.6) | 1076 (88.9) |
| No | 270 (2.3) | 49 (2.3) | 42 (2.5) | 66 (2.2) | 43 (1.7) | 39 (3.3) | 31 (2.6) |
| Does not apply | 493 (4.2) | 50 (2.6) | 61 (3.4) | 130 (3.4) | 90 (4.0) | 76 (6.1) | 86 (7.3) |
| Don't know | 102 (0.9) | 19 (0.8) | 12 (0.8) | 21 (0.6) | 22 (1.1) | 17 (1.0) | 11 (1.0) |
| Refused | 16 (0.1) | 0 (0.0) | 3 (0.1) | 4 (0.1) | 5 (0.1) | 0 (0.0) | 4 (0.3) |
| ***Quality of relationship with mother*** | | | | | | | |
| Very good | 11530 (89.8) | 2162 (91.6) | 1753 (90.4) | 3284 (91.6) | 2128 (89.8) | 1168 (87.2) | 1035 (85.6) |
| Somewhat good | 761 (6.2) | 121 (5.8) | 117 (6.3) | 191 (5.2) | 150 (6.4) | 82 (6.4) | 100 (7.9) |
| Somewhat bad | 84 (0.7) | 17 (0.8) | 10 (0.4) | 28 (0.8) | 13 (0.5) | 7 (0.6) | 9 (1.0) |
| Very bad | 56 (0.6) | 7 (0.5) | 10 (0.6) | 12 (0.4) | 11 (0.5) | 11 (1.1) | 5 (0.5) |
| Does not apply | 241 (2.1) | 17 (0.8) | 29 (1.6) | 57 (1.4) | 45 (2.0) | 44 (3.9) | 49 (4.4) |
| Don't know | 87 (0.6) | 10 (0.4) | 12 (0.6) | 25 (0.6) | 18 (0.9) | 13 (0.6) | 9 (0.6) |
| Refused | 6 (0.0) | 1 (0.0) | 2 (0.1) | 1 (0.0) | 0 (0.0) | 1 (0.0) | 1 (0.0) |
| ***Quality of relationship with father*** | | | | | | | |
| Very good | 11022 (85.6) | 2074 (87.3) | 1669 (85.7) | 3151 (87.2) | 2027 (85.6) | 1114 (83.7) | 987 (81.4) |
| Somewhat good | 985 (7.8) | 167 (7.7) | 160 (8.2) | 251 (7.0) | 201 (8.3) | 104 (7.8) | 102 (8.3) |
| Somewhat bad | 104 (1.0) | 17 (0.8) | 18 (0.9) | 30 (1.3) | 18 (0.9) | 8 (0.6) | 13 (1.2) |
| Very bad | 74 (0.8) | 14 (0.9) | 15 (1.2) | 18 (0.6) | 9 (0.4) | 13 (1.1) | 5 (0.6) |
| Does not apply | 450 (3.8) | 42 (2.1) | 53 (3.0) | 124 (3.2) | 81 (3.6) | 66 (5.2) | 84 (7.1) |

| | | | | | | | |
|---|---|---|---|---|---|---|---|
| Don't know | 117 (1.0) | 21 (1.1) | 17 (1.0) | 21 (0.6) | 25 (1.2) | 20 (1.3) | 13 (1.1) |
| Refused | 13 (0.1) | 0 (0.0) | 1 (0.0) | 3 (0.1) | 4 (0.1) | 1 (0.2) | 4 (0.3) |
| ***Self-rated health while growing up*** | | | | | | | |
| Excellent | 2092 (17.1) | 400 (16.2) | 292 (16.6) | 619 (18.4) | 385 (18.3) | 210 (16.2) | 186 (15.6) |
| Very good | 3857 (30.4) | 710 (31.0) | 613 (31.9) | 1069 (29.5) | 687 (29.5) | 402 (29.8) | 376 (31.6) |
| Good | 3988 (31.6) | 737 (32.5) | 609 (30.8) | 1092 (31.4) | 741 (30.5) | 406 (31.1) | 403 (33.0) |
| Fair | 2357 (17.2) | 425 (17.7) | 363 (17.5) | 688 (17.3) | 437 (16.7) | 251 (18.3) | 193 (16.0) |
| Poor | 426 (3.3) | 57 (2.3) | 53 (2.9) | 115 (2.9) | 103 (4.5) | 54 (4.4) | 44 (3.3) |
| Don't know | 42 (0.3) | 5 (0.2) | 3 (0.2) | 13 (0.4) | 12 (0.5) | 3 (0.1) | 6 (0.5) |
| Refused | 3 (0.0) | 1 (0.0) | 0 (0.0) | 2 (0.1) | 0 (0.0) | 0 (0.0) | 0 (0.0) |
| ***Perceived family financial situation while growing up*** | | | | | | | |
| Lived comfortably | 5275 (38.7) | 1170 (46.3) | 848 (40.8) | 1508 (38.8) | 879 (36.2) | 455 (33.6) | 415 (33.2) |
| Got by | 2940 (23.6) | 539 (23.7) | 461 (24.3) | 866 (24.9) | 554 (24.1) | 286 (21.8) | 234 (21.2) |
| Found it difficult | 2539 (21.2) | 388 (18.3) | 386 (22.6) | 713 (21.0) | 494 (21.5) | 289 (22.7) | 269 (22.7) |
| Found it very difficult | 1940 (15.9) | 227 (11.0) | 229 (11.9) | 492 (14.8) | 420 (17.6) | 287 (21.2) | 285 (22.5) |
| Don't know | 67 (0.5) | 11 (0.6) | 8 (0.4) | 18 (0.5) | 17 (0.6) | 9 (0.7) | 4 (0.4) |
| Refused | 4 (0.0) | 0 (0.0) | 1 (0.0) | 1 (0.0) | 1 (0.0) | 0 (0.0) | 1 (0.0) |

***Note:*** Cross-sectional Wave 1 weights were used for weight-adjusted proportions (%).

**eTable 20.** Distribution of outcome variables collected in Wave 1 survey (2023) in India and stratified by age groups 18 to 24 inclusive, 25 to 29 inclusive, 30 to 39 inclusive, 40 to 49 inclusive, 50 to 59 inclusive, and 60+ (N = 12,765).

| Outcomes and flourishing composite indices | Wave 1 | 18 to 24 | 25 to 29 | 30 to 39 | 40 to 49 | 50 to 59 | 60+ |
|---|---|---|---|---|---|---|---|
| | Mean (sd) | Mean (sd) | Mean (sd) | Mean (sd) | Mean (sd) | Mean (sd) | Mean (sd) |
| ***Belonging*** | 8.41 (2.9) | 8.68 (2.6) | 8.54 (2.8) | 8.49 (2.8) | 8.27 (3.0) | 8.12 (3.0) | 8.14 (3.0) |
| ***Flourishing (with financial indicators)*** | 6.89 (1.9) | 7.29 (1.7) | 7.06 (1.9) | 6.83 (1.9) | 6.74 (1.9) | 6.66 (1.9) | 6.58 (2.1) |
| ***Flourishing (without financial indicators)*** | 7.45 (2.0) | 7.92 (1.9) | 7.64 (2.0) | 7.42 (2.0) | 7.31 (2.0) | 7.18 (2.0) | 7.05 (2.2) |
| ***Freedom*** | 8.15 (3.0) | 8.34 (2.9) | 8.33 (2.9) | 8.14 (3.0) | 8.11 (3.0) | 8.06 (2.9) | 7.84 (3.1) |
| ***Happiness***‡ | 6.48 (3.6) | 7.17 (3.4) | 6.63 (3.5) | 6.43 (3.5) | 6.23 (3.6) | 5.99 (3.6) | 6.14 (3.7) |
| ***Life evaluation five years from now*** | 7.30 (3.1) | 7.80 (2.8) | 7.67 (2.9) | 7.34 (3.0) | 7.21 (3.1) | 6.72 (3.3) | 6.70 (3.3) |
| ***Life evaluation today*** | 5.63 (3.6) | 6.11 (3.6) | 5.64 (3.6) | 5.55 (3.6) | 5.41 (3.5) | 5.32 (3.6) | 5.59 (3.6) |
| ***Life satisfaction***‡ | 7.00 (3.5) | 7.51 (3.3) | 7.13 (3.5) | 6.94 (3.5) | 6.81 (3.5) | 6.62 (3.5) | 6.83 (3.5) |
| ***Life worthwhile***‡ | 6.93 (3.4) | 7.38 (3.3) | 7.07 (3.4) | 6.79 (3.5) | 6.90 (3.4) | 6.69 (3.4) | 6.67 (3.5) |
| ***Loneliness***§ | 3.84 (4.0) | 3.59 (4.0) | 3.89 (4.0) | 3.86 (4.0) | 4.05 (4.1) | 3.80 (3.9) | 3.86 (4.0) |
| ***Positive expectations*** | 8.14 (2.9) | 8.24 (2.9) | 8.16 (3.0) | 8.13 (2.9) | 8.17 (2.9) | 8.33 (2.6) | 7.78 (3.0) |
| ***Relationship contentment***‡ | 7.96 (3.1) | 8.38 (2.9) | 8.17 (3.0) | 7.84 (3.2) | 7.81 (3.2) | 7.83 (3.1) | 7.62 (3.2) |
| ***Relationship satisfaction***‡ | 7.92 (3.1) | 8.15 (3.0) | 8.01 (3.1) | 7.84 (3.1) | 7.85 (3.1) | 7.93 (3.0) | 7.72 (3.1) |
| ***Remain hopeful for the future*** | 8.32 (2.8) | 8.47 (2.7) | 8.51 (2.6) | 8.35 (2.7) | 8.27 (2.8) | 8.24 (2.8) | 7.95 (3.0) |
| ***Understand purpose in life***‡ | 8.04 (3.0) | 8.31 (2.9) | 8.23 (2.9) | 8.13 (2.9) | 7.99 (3.1) | 7.79 (3.1) | 7.58 (3.2) |
| ***Mental health***‡ | 7.44 (3.3) | 8.12 (2.9) | 7.77 (3.2) | 7.39 (3.3) | 7.28 (3.3) | 6.90 (3.5) | 6.87 (3.5) |
| ***Physical health***‡ | 7.00 (3.4) | 7.94 (3.0) | 7.52 (3.2) | 7.15 (3.3) | 6.85 (3.4) | 6.19 (3.6) | 5.74 (3.7) |
| ***Act to promote good***‡ | 8.35 (2.7) | 8.59 (2.5) | 8.50 (2.7) | 8.36 (2.7) | 8.22 (2.8) | 8.21 (2.7) | 8.13 (2.8) |
| ***Give up some happiness now for greater happiness later***‡ | 7.21 (3.5) | 7.51 (3.4) | 7.32 (3.6) | 7.20 (3.5) | 6.97 (3.5) | 7.19 (3.4) | 6.97 (3.4) |
| ***Have a long list of gratitude*** | 7.69 (3.1) | 7.81 (3.1) | 7.79 (3.2) | 7.68 (3.2) | 7.74 (3.1) | 7.66 (3.0) | 7.34 (3.2) |
| ***Show love often*** | 8.20 (3.0) | 8.38 (3.0) | 8.36 (3.0) | 8.22 (3.0) | 8.13 (3.0) | 8.03 (3.1) | 8.01 (3.0) |
| ***Perceived support from others*** | 6.46 (3.9) | 6.89 (3.8) | 6.55 (3.9) | 6.40 (3.9) | 6.27 (3.9) | 6.36 (3.9) | 6.19 (3.9) |
| ***Worry about meeting normal monthly living expenses***§§‡‡ | 5.89 (4.1) | 5.62 (4.2) | 5.69 (4.1) | 6.06 (4.0) | 6.05 (4.0) | 5.99 (4.0) | 5.88 (4.0) |
| ***Worry about safety, food, or housing***§§‡‡ | 6.02 (4.1) | 5.98 (4.2) | 6.02 (4.1) | 6.11 (4.1) | 6.13 (4.1) | 5.98 (4.1) | 5.76 (4.1) |
| | n (%) | n (%) | n (%) | n (%) | n (%) | n (%) | n (%) |
| ***Alcohol use*** | | | | | | | |
| Yes | 828 (7.5) | 89 (5.0) | 143 (8.8) | 254 (8.5) | 192 (9.4) | 91 (7.8) | 59 (5.4) |
| No | 11404 (88.3) | 2155 (91.8) | 1732 (88.4) | 3190 (87.0) | 2064 (86.0) | 1174 (87.5) | 1089 (89.5) |
| Saw, skipped | 36 (0.2) | 19 (0.3) | 5 (0.3) | 5 (0.1) | 3 (0.1) | 4 (0.3) | 0 (0.0) |
| Don't know | 111 (0.9) | 16 (0.4) | 12 (0.6) | 37 (1.4) | 16 (0.7) | 17 (1.3) | 13 (1.1) |
| Refused | 385 (3.0) | 56 (2.4) | 41 (1.9) | 111 (3.0) | 90 (3.8) | 40 (3.2) | 47 (4.1) |
| Missing (NAs) | 1 (0.0) | 0 (0.0) | 0 (0.0) | 1 (0.1) | 0 (0.0) | 0 (0.0) | 0 (0.0) |
| ***Cigarette smoking*** | | | | | | | |
| Yes | 843 (7.6) | 103 (5.8) | 116 (7.6) | 203 (6.6) | 196 (9.4) | 123 (9.4) | 102 (8.0) |
| No | 11404 (88.4) | 2140 (90.9) | 1755 (89.3) | 3239 (88.9) | 2061 (86.0) | 1154 (86.4) | 1055 (87.7) |
| Saw, skipped | 33 (0.2) | 18 (0.3) | 5 (0.3) | 5 (0.1) | 3 (0.1) | 2 (0.1) | 0 (0.0) |
| Don't know | 87 (0.7) | 14 (0.6) | 13 (0.6) | 31 (1.1) | 11 (0.6) | 14 (1.2) | 4 (0.3) |

| | | | | | | | |
|---|---|---|---|---|---|---|---|
| Refused | 397 (3.1) | 60 (2.4) | 44 (2.1) | 119 (3.2) | 94 (3.9) | 33 (2.9) | 47 (3.9) |
| Missing (NAs) | 1 (0.0) | 0 (0.0) | 0 (0.0) | 1 (0.1) | 0 (0.0) | 0 (0.0) | 0 (0.0) |
| ***Have any health problems*** | | | | | | | |
| Yes | 3476 (28.1) | 398 (18.2) | 437 (21.8) | 889 (24.9) | 768 (31.9) | 471 (35.7) | 513 (43.6) |
| No | 9188 (71.0) | 1915 (80.7) | 1483 (77.6) | 2679 (74.3) | 1576 (67.2) | 848 (63.5) | 687 (55.7) |
| Saw, skipped | 1 (0.0) | 0 (0.0) | 0 (0.0) | 1 (0.0) | 0 (0.0) | 0 (0.0) | 0 (0.0) |
| Don't know | 77 (0.7) | 17 (0.8) | 8 (0.4) | 21 (0.6) | 18 (0.8) | 6 (0.7) | 7 (0.7) |
| Refused | 18 (0.1) | 3 (0.2) | 3 (0.1) | 7 (0.2) | 3 (0.1) | 1 (0.1) | 1 (0.1) |
| Missing (NAs) | 5 (0.0) | 2 (0.1) | 2 (0.1) | 1 (0.1) | 0 (0.0) | 0 (0.0) | 0 (0.0) |
| ***Donated to charity*** | | | | | | | |
| Yes | 4972 (38.6) | 834 (36.8) | 782 (40.0) | 1418 (38.0) | 961 (40.1) | 513 (38.4) | 464 (39.5) |
| No | 7732 (61.0) | 1492 (62.8) | 1142 (59.5) | 2162 (61.6) | 1389 (59.4) | 809 (61.4) | 738 (60.2) |
| Don't know | 60 (0.4) | 9 (0.4) | 8 (0.5) | 18 (0.3) | 15 (0.6) | 4 (0.2) | 6 (0.4) |
| Refused | 1 (0.0) | 0 (0.0) | 1 (0.0) | 0 (0.0) | 0 (0.0) | 0 (0.0) | 0 (0.0) |
| ***Helped a stranger*** | | | | | | | |
| Yes | 6889 (55.1) | 1256 (56.3) | 1097 (56.7) | 1974 (57.1) | 1291 (55.8) | 699 (53.6) | 572 (48.1) |
| No | 5716 (43.8) | 1057 (42.9) | 815 (42.3) | 1577 (41.9) | 1038 (42.8) | 613 (45.5) | 616 (50.2) |
| Don't know | 159 (1.1) | 22 (0.8) | 20 (1.0) | 47 (1.1) | 36 (1.4) | 14 (1.0) | 20 (1.6) |
| Refused | 1 (0.0) | 0 (0.0) | 1 (0.0) | 0 (0.0) | 0 (0.0) | 0 (0.0) | 0 (0.0) |
| ***Volunteered at an organization*** | | | | | | | |
| Yes | 3921 (29.0) | 703 (29.5) | 626 (30.2) | 1168 (30.7) | 734 (30.1) | 376 (26.6) | 314 (24.8) |
| No | 8766 (70.4) | 1619 (69.9) | 1298 (69.4) | 2409 (68.7) | 1615 (69.2) | 938 (72.7) | 887 (74.6) |
| Don't know | 76 (0.6) | 12 (0.6) | 9 (0.4) | 20 (0.6) | 16 (0.6) | 12 (0.8) | 7 (0.6) |
| Refused | 2 (0.0) | 1 (0.0) | 0 (0.0) | 1 (0.1) | 0 (0.0) | 0 (0.0) | 0 (0.0) |
| ***Feel close to someone*** | | | | | | | |
| Yes | 10392 (81.6) | 1965 (83.9) | 1612 (83.4) | 2967 (82.4) | 1854 (79.1) | 1053 (80.1) | 941 (79.8) |
| No | 2347 (18.2) | 365 (15.9) | 319 (16.6) | 620 (17.4) | 508 (20.7) | 270 (19.8) | 265 (20.2) |
| Don't know | 24 (0.2) | 5 (0.2) | 2 (0.0) | 10 (0.2) | 2 (0.2) | 3 (0.2) | 2 (0.1) |
| Refused | 2 (0.0) | 0 (0.0) | 0 (0.0) | 1 (0.0) | 1 (0.0) | 0 (0.0) | 0 (0.0) |
| ***Feel at peace***§ | | | | | | | |
| Always | 5334 (43.3) | 932 (42.7) | 768 (40.6) | 1494 (41.4) | 979 (43.0) | 595 (46.0) | 566 (48.3) |
| Often | 2196 (16.8) | 394 (16.2) | 296 (15.8) | 626 (17.1) | 417 (16.5) | 249 (18.2) | 214 (17.3) |
| Rarely | 3966 (29.5) | 784 (30.7) | 674 (33.0) | 1135 (31.1) | 734 (30.9) | 345 (25.3) | 294 (23.5) |
| Never | 1154 (9.4) | 213 (9.8) | 178 (9.7) | 315 (9.6) | 208 (8.3) | 124 (9.5) | 116 (9.6) |
| Saw, skipped | 2 (0.0) | 0 (0.0) | 0 (0.0) | 2 (0.0) | 0 (0.0) | 0 (0.0) | 0 (0.0) |
| Don't know | 106 (0.9) | 12 (0.6) | 15 (0.9) | 25 (0.7) | 24 (1.2) | 12 (1.0) | 18 (1.3) |
| Refused | 7 (0.0) | 0 (0.0) | 2 (0.1) | 1 (0.0) | 3 (0.1) | 1 (0.1) | 0 (0.0) |
| ***Various aspects of life are balanced***§ | | | | | | | |
| Always | 3942 (31.8) | 735 (33.6) | 545 (29.3) | 1116 (30.7) | 740 (32.0) | 404 (30.8) | 402 (34.1) |
| Often | 2346 (18.5) | 400 (16.5) | 352 (18.3) | 658 (19.4) | 451 (18.8) | 263 (20.5) | 222 (18.1) |
| Rarely | 5082 (38.5) | 951 (38.1) | 818 (41.2) | 1433 (39.0) | 942 (39.3) | 507 (37.2) | 431 (35.5) |
| Never | 1210 (9.7) | 223 (10.8) | 195 (9.9) | 332 (9.4) | 198 (8.2) | 131 (9.6) | 131 (10.7) |
| Saw, skipped | 2 (0.0) | 1 (0.1) | 0 (0.0) | 1 (0.0) | 0 (0.0) | 0 (0.0) | 0 (0.0) |
| Don't know | 169 (1.4) | 22 (0.8) | 19 (1.1) | 56 (1.5) | 29 (1.4) | 21 (1.8) | 22 (1.6) |
| Refused | 14 (0.1) | 3 (0.1) | 4 (0.2) | 2 (0.0) | 5 (0.2) | 0 (0.0) | 0 (0.0) |

| | | | | | | | |
|---|---|---|---|---|---|---|---|
| ***Depressive symptoms*§** | | | | | | | |
| Nearly every day | 1757 (14.1) | 272 (12.5) | 247 (13.0) | 499 (14.1) | 333 (14.1) | 206 (15.6) | 200 (16.2) |
| More than half the days | 2114 (16.7) | 374 (16.1) | 332 (18.0) | 623 (17.5) | 395 (16.4) | 200 (15.6) | 190 (15.9) |
| Several days | 3422 (25.5) | 613 (25.9) | 510 (24.6) | 918 (23.9) | 667 (26.1) | 390 (27.4) | 324 (25.8) |
| Not at all | 5366 (42.9) | 1062 (44.9) | 828 (43.5) | 1532 (43.8) | 943 (42.2) | 519 (40.4) | 482 (41.2) |
| Don't know | 88 (0.7) | 12 (0.4) | 12 (0.8) | 24 (0.7) | 22 (1.0) | 10 (0.8) | 8 (0.7) |
| Refused | 18 (0.1) | 2 (0.2) | 4 (0.2) | 2 (0.0) | 5 (0.2) | 1 (0.2) | 4 (0.2) |
| ***Distress from life-threatening experiences*§** | | | | | | | |
| A lot | 3219 (25.1) | 494 (21.8) | 469 (24.6) | 913 (24.9) | 618 (26.2) | 396 (28.4) | 329 (26.6) |
| Some | 2955 (22.5) | 518 (21.8) | 442 (21.0) | 824 (22.6) | 600 (24.5) | 309 (23.4) | 262 (21.2) |
| Not very much | 1111 (8.3) | 223 (8.4) | 159 (7.8) | 299 (8.1) | 187 (7.5) | 128 (9.6) | 115 (9.1) |
| Not at all | 5400 (43.4) | 1092 (47.7) | 852 (46.1) | 1536 (43.6) | 943 (40.9) | 484 (37.7) | 493 (42.4) |
| Don't know | 64 (0.5) | 7 (0.3) | 10 (0.4) | 20 (0.7) | 11 (0.5) | 9 (0.9) | 7 (0.6) |
| Refused | 16 (0.1) | 1 (0.0) | 1 (0.1) | 6 (0.1) | 6 (0.4) | 0 (0.0) | 2 (0.1) |
| ***Extent of suffering*§** | | | | | | | |
| A lot | 2418 (20.4) | 318 (15.1) | 282 (15.5) | 655 (19.2) | 502 (22.0) | 340 (27.2) | 321 (27.6) |
| Some | 4000 (30.4) | 634 (26.4) | 556 (28.6) | 1157 (31.3) | 796 (32.0) | 449 (32.0) | 408 (33.1) |
| Not very much | 1342 (9.8) | 246 (9.5) | 220 (10.3) | 367 (9.7) | 242 (9.4) | 148 (11.1) | 119 (9.7) |
| Not at all | 4906 (38.5) | 1124 (48.4) | 858 (44.9) | 1387 (38.9) | 810 (36.0) | 381 (29.1) | 346 (28.7) |
| Saw, skipped | 1 (0.0) | 0 (0.0) | 0 (0.0) | 1 (0.0) | 0 (0.0) | 0 (0.0) | 0 (0.0) |
| Don't know | 86 (0.6) | 13 (0.6) | 16 (0.7) | 28 (0.8) | 11 (0.4) | 7 (0.6) | 11 (0.7) |
| Refused | 12 (0.1) | 0 (0.0) | 1 (0.1) | 3 (0.1) | 4 (0.3) | 1 (0.1) | 3 (0.3) |
| ***Feeling nervous or anxious*§** | | | | | | | |
| Nearly every day | 1890 (15.3) | 243 (11.0) | 263 (14.6) | 545 (15.2) | 376 (15.9) | 237 (18.6) | 226 (18.8) |
| More than half the days | 1982 (15.9) | 322 (14.6) | 321 (17.4) | 573 (16.5) | 387 (16.4) | 207 (16.1) | 172 (14.2) |
| Several days | 3158 (23.6) | 579 (23.9) | 450 (20.7) | 855 (22.2) | 603 (24.7) | 357 (25.3) | 314 (25.0) |
| Not at all | 5660 (44.6) | 1179 (49.9) | 889 (46.9) | 1603 (45.3) | 984 (42.1) | 519 (39.4) | 486 (41.2) |
| Saw, skipped | 2 (0.0) | 1 (0.0) | 0 (0.0) | 1 (0.0) | 0 (0.0) | 0 (0.0) | 0 (0.0) |
| Don't know | 56 (0.5) | 7 (0.3) | 6 (0.2) | 18 (0.6) | 11 (0.6) | 6 (0.6) | 8 (0.8) |
| Refused | 17 (0.2) | 4 (0.2) | 4 (0.2) | 3 (0.1) | 4 (0.3) | 0 (0.0) | 2 (0.1) |
| ***Have little interest in doing things*§** | | | | | | | |
| Nearly every day | 3428 (28.6) | 591 (27.2) | 483 (26.6) | 998 (29.5) | 662 (29.7) | 365 (29.7) | 329 (28.5) |
| More than half the days | 2147 (17.7) | 372 (16.1) | 322 (17.6) | 597 (18.2) | 413 (17.8) | 247 (19.9) | 196 (17.2) |
| Several days | 2959 (21.9) | 559 (23.0) | 465 (22.5) | 799 (20.8) | 548 (21.7) | 316 (22.7) | 272 (21.0) |
| Not at all | 4119 (30.9) | 801 (33.1) | 652 (32.8) | 1181 (30.9) | 713 (29.5) | 379 (26.4) | 393 (31.8) |
| Saw, skipped | 3 (0.0) | 1 (0.0) | 1 (0.0) | 0 (0.0) | 0 (0.0) | 1 (0.0) | 0 (0.0) |
| Don't know | 91 (0.7) | 9 (0.4) | 8 (0.4) | 21 (0.5) | 25 (1.2) | 12 (0.9) | 16 (1.4) |
| Refused | 18 (0.1) | 2 (0.1) | 2 (0.1) | 2 (0.0) | 4 (0.1) | 6 (0.4) | 2 (0.1) |
| ***Unable to control worry*§** | | | | | | | |
| Nearly every day | 2071 (16.4) | 317 (14.3) | 277 (14.4) | 591 (15.9) | 427 (17.4) | 240 (19.3) | 219 (18.6) |
| More than half the days | 1873 (14.8) | 317 (13.2) | 300 (15.8) | 541 (15.9) | 349 (14.7) | 210 (16.1) | 156 (12.7) |
| Several days | 2852 (21.4) | 483 (20.1) | 422 (20.2) | 777 (20.4) | 544 (22.4) | 327 (23.0) | 299 (23.3) |
| Not at all | 5842 (46.4) | 1197 (51.5) | 922 (48.9) | 1651 (46.7) | 1015 (43.9) | 541 (41.0) | 516 (43.8) |
| Saw, skipped | 2 (0.0) | 0 (0.0) | 0 (0.0) | 2 (0.0) | 0 (0.0) | 0 (0.0) | 0 (0.0) |

| | | | | | | | |
|---|---|---|---|---|---|---|---|
| Don't know | 83 (0.7) | 17 (0.7) | 5 (0.2) | 24 (0.7) | 21 (1.0) | 5 (0.4) | 11 (1.1) |
| Refused | 42 (0.4) | 4 (0.2) | 7 (0.5) | 12 (0.3) | 9 (0.5) | 3 (0.2) | 7 (0.5) |
| ***Feeling capable*§** | | | | | | | |
| Always | 6778 (54.1) | 1242 (54.5) | 1003 (52.3) | 1933 (53.9) | 1275 (55.2) | 723 (55.9) | 602 (52.4) |
| Often | 2002 (15.3) | 362 (14.5) | 272 (13.9) | 568 (16.1) | 381 (15.7) | 223 (15.9) | 196 (15.1) |
| Rarely | 3012 (22.7) | 552 (22.8) | 511 (25.9) | 856 (22.9) | 534 (22.0) | 281 (20.4) | 278 (22.3) |
| Never | 895 (7.2) | 164 (7.5) | 139 (7.4) | 223 (6.6) | 161 (6.4) | 89 (6.8) | 119 (9.1) |
| Saw, skipped | 4 (0.0) | 1 (0.1) | 2 (0.1) | 1 (0.0) | 0 (0.0) | 0 (0.0) | 0 (0.0) |
| Don't know | 66 (0.6) | 12 (0.7) | 6 (0.3) | 16 (0.4) | 11 (0.6) | 10 (1.0) | 11 (1.1) |
| Refused | 8 (0.0) | 2 (0.1) | 0 (0.0) | 1 (0.0) | 3 (0.1) | 0 (0.0) | 2 (0.1) |
| ***Forgiveness*§** | | | | | | | |
| Always | 7691 (60.7) | 1374 (60.4) | 1195 (61.9) | 2188 (60.4) | 1419 (60.8) | 789 (59.7) | 726 (61.6) |
| Often | 1786 (13.7) | 296 (11.8) | 255 (13.4) | 498 (14.0) | 331 (14.1) | 213 (14.1) | 193 (15.2) |
| Rarely | 2258 (16.8) | 480 (19.2) | 324 (15.5) | 621 (16.8) | 431 (16.7) | 217 (17.1) | 185 (14.1) |
| Never | 931 (8.0) | 170 (7.9) | 150 (8.7) | 259 (7.8) | 162 (7.3) | 94 (8.0) | 96 (8.5) |
| Saw, skipped | 2 (0.0) | 1 (0.1) | 0 (0.0) | 1 (0.0) | 0 (0.0) | 0 (0.0) | 0 (0.0) |
| Don't know | 48 (0.4) | 7 (0.2) | 4 (0.2) | 15 (0.4) | 10 (0.5) | 7 (0.5) | 5 (0.4) |
| Refused | 49 (0.5) | 7 (0.4) | 5 (0.4) | 16 (0.6) | 12 (0.7) | 6 (0.5) | 3 (0.3) |
| ***Government approval*§** | | | | | | | |
| Strongly approve | 7024 (54.9) | 1304 (57.3) | 1046 (52.6) | 2022 (55.9) | 1259 (53.4) | 726 (53.7) | 667 (55.2) |
| Somewhat approve | 3640 (27.6) | 685 (27.8) | 570 (29.0) | 993 (26.7) | 699 (27.9) | 368 (28.0) | 325 (26.5) |
| Neither approve nor disapprove | 477 (4.1) | 73 (2.8) | 62 (3.7) | 133 (4.2) | 98 (4.6) | 59 (5.0) | 52 (4.5) |
| Somewhat disapprove | 648 (5.1) | 115 (4.6) | 99 (5.7) | 185 (5.1) | 111 (4.7) | 72 (5.1) | 66 (5.5) |
| Strongly disapprove | 735 (6.1) | 122 (5.5) | 123 (6.8) | 204 (6.2) | 147 (7.0) | 70 (5.4) | 69 (5.7) |
| Saw, skipped | 1 (0.0) | 1 (0.1) | 0 (0.0) | 0 (0.0) | 0 (0.0) | 0 (0.0) | 0 (0.0) |
| Don't know | 195 (1.9) | 29 (1.7) | 27 (1.7) | 50 (1.7) | 36 (1.8) | 27 (2.4) | 26 (2.5) |
| Refused | 45 (0.3) | 6 (0.2) | 6 (0.4) | 11 (0.3) | 15 (0.6) | 4 (0.4) | 3 (0.2) |
| ***Obeying the law*§** | | | | | | | |
| Strongly agree | 11179 (87.3) | 2069 (88.9) | 1683 (86.3) | 3150 (86.9) | 2067 (87.3) | 1164 (87.2) | 1046 (86.5) |
| Somewhat agree | 1270 (10.0) | 216 (8.8) | 200 (10.3) | 363 (10.5) | 231 (9.5) | 127 (9.9) | 133 (11.0) |
| Neither agree nor disagree | 98 (0.9) | 13 (0.6) | 13 (0.8) | 26 (0.9) | 23 (1.2) | 11 (0.7) | 12 (1.1) |
| Somewhat disagree | 67 (0.6) | 13 (0.6) | 8 (0.5) | 20 (0.6) | 17 (1.0) | 4 (0.4) | 5 (0.4) |
| Strongly disagree | 88 (0.7) | 18 (0.7) | 17 (1.1) | 23 (0.8) | 15 (0.5) | 12 (0.9) | 3 (0.3) |
| Don't know | 61 (0.5) | 6 (0.3) | 12 (0.9) | 15 (0.3) | 11 (0.4) | 8 (0.8) | 9 (0.7) |
| Refused | 2 (0.0) | 0 (0.0) | 0 (0.0) | 1 (0.0) | 1 (0.1) | 0 (0.0) | 0 (0.0) |
| ***Perceived discrimination*§** | | | | | | | |
| Always | 1653 (12.9) | 291 (13.1) | 246 (12.8) | 471 (12.6) | 322 (13.5) | 175 (12.9) | 148 (12.3) |
| Often | 1129 (8.6) | 191 (8.4) | 148 (7.9) | 341 (9.2) | 206 (8.5) | 121 (8.2) | 122 (8.8) |
| Rarely | 2897 (20.9) | 548 (22.0) | 472 (22.2) | 804 (21.0) | 516 (19.7) | 305 (20.7) | 252 (19.5) |
| Never | 6912 (56.1) | 1279 (55.4) | 1048 (56.2) | 1930 (55.6) | 1293 (57.0) | 701 (55.9) | 661 (57.1) |
| Saw, skipped | 4 (0.0) | 2 (0.1) | 1 (0.1) | 1 (0.0) | 0 (0.0) | 0 (0.0) | 0 (0.0) |
| Don't know | 137 (1.2) | 20 (0.8) | 10 (0.6) | 42 (1.3) | 23 (1.0) | 20 (1.9) | 22 (2.1) |
| Refused | 33 (0.3) | 4 (0.2) | 8 (0.4) | 9 (0.3) | 5 (0.2) | 4 (0.3) | 3 (0.2) |
| ***Say in government*#** | | | | | | | |
| Agree | 9873 (77.3) | 1792 (77.1) | 1503 (76.3) | 2806 (78.3) | 1826 (76.5) | 1020 (76.9) | 926 (77.7) |

| | | | | | | | |
|---|---|---|---|---|---|---|---|
| Disagree | 2077 (16.1) | 387 (16.1) | 316 (17.1) | 576 (15.5) | 385 (16.4) | 217 (16.5) | 196 (15.3) |
| Unsure | 741 (6.1) | 141 (6.1) | 104 (6.1) | 199 (5.8) | 135 (6.1) | 85 (6.3) | 77 (6.3) |
| Saw, skipped | 4 (0.0) | 4 (0.1) | 0 (0.0) | 0 (0.0) | 0 (0.0) | 0 (0.0) | 0 (0.0) |
| Refused | 70 (0.6) | 11 (0.4) | 10 (0.6) | 17 (0.4) | 19 (1.0) | 4 (0.2) | 9 (0.7) |
| ***Support reducing differences in income levels***§ | | | | | | | |
| Strongly agree | 9548 (74.1) | 1773 (75.0) | 1470 (74.2) | 2699 (74.8) | 1758 (74.4) | 975 (72.6) | 873 (72.2) |
| Somewhat agree | 2250 (17.8) | 398 (17.7) | 340 (18.7) | 632 (17.4) | 422 (17.3) | 236 (18.1) | 222 (18.1) |
| Neither agree nor disagree | 237 (2.2) | 34 (1.6) | 31 (1.8) | 69 (2.3) | 46 (2.5) | 26 (2.4) | 31 (2.7) |
| Somewhat disagree | 217 (1.7) | 56 (2.2) | 23 (1.2) | 64 (1.7) | 35 (1.4) | 22 (1.6) | 17 (1.7) |
| Strongly disagree | 260 (2.4) | 37 (2.0) | 39 (2.5) | 65 (2.1) | 54 (2.5) | 32 (2.9) | 33 (3.0) |
| Don't know | 248 (1.8) | 37 (1.4) | 29 (1.5) | 67 (1.6) | 48 (1.8) | 35 (2.5) | 32 (2.3) |
| Refused | 5 (0.0) | 0 (0.0) | 1 (0.1) | 2 (0.0) | 2 (0.0) | 0 (0.0) | 0 (0.0) |
| ***Trust in others***§ | | | | | | | |
| All | 1919 (16.2) | 313 (15.5) | 271 (14.4) | 571 (16.6) | 345 (15.2) | 210 (17.3) | 209 (18.5) |
| Most | 2225 (17.3) | 438 (17.7) | 309 (14.3) | 580 (16.2) | 435 (19.0) | 238 (17.9) | 225 (18.5) |
| Some | 6001 (46.3) | 1126 (46.4) | 965 (51.3) | 1685 (46.2) | 1113 (46.5) | 599 (44.3) | 513 (43.1) |
| Not very much | 1154 (8.5) | 232 (9.5) | 175 (8.7) | 344 (8.8) | 200 (8.0) | 113 (8.0) | 90 (6.8) |
| None at all | 1097 (8.6) | 186 (8.9) | 170 (8.7) | 317 (8.9) | 203 (8.1) | 108 (8.1) | 113 (8.5) |
| Saw, skipped | 1 (0.0) | 0 (0.0) | 0 (0.0) | 1 (0.0) | 0 (0.0) | 0 (0.0) | 0 (0.0) |
| Don't know | 321 (2.8) | 35 (1.6) | 34 (2.1) | 89 (3.0) | 59 (2.8) | 52 (3.9) | 52 (4.2) |
| Refused | 47 (0.4) | 5 (0.4) | 9 (0.6) | 11 (0.3) | 10 (0.4) | 6 (0.5) | 6 (0.4) |

***Note*:**

- Cross-sectional Wave 1 weights were used for weight-adjusted proportions (%).
- Those marked with ‡ are included in the flourishing composite without financial indicators, and ‡‡ are the financial indicators included in the flourishing composite with financial indicators.
- The outcome variables marked with § were reverse coded in the association analysis.
- The outcome variable *say in government* marked with # was coded in the order by level 1 = disagree, level 2 = unsure, and level 3 = agree.
- The outcome variables (*worry about meeting normal monthly living expenses* and *worry about safety, food, or housing*) marked §§ were reverse coded in the association analysis and they are reverse coded for the table standalone reporting here. Within flourishing index calculations, *worry about meeting normal monthly living expenses* and *worry about safety, food, or housing* are on original scales. For binary outcomes, responses with “No” were recoded as 0; responses with “Yes” were recoded as 1 in the association analysis.

**eTable 21.** Parental marital status at approximately age 12 among respondents selecting "does not apply" for parental experience items, reporting unweighted sample size and weighted proportions.

| **Parents' marital status at age 12** | **Feeling loved by mother (N=270)** | **Feeling loved by father (N=493)** | **Relationship quality with mother (N=241)** | **Relationship quality with father (N=450)** |
|---|---|---|---|---|
| Yes, married | 101 (36.1) | 166 (31.6) | 90 (36.4) | 156 (33.3) |
| No, divorced | 5 (2.0) | 10 (1.9) | 5 (2.3) | 9 (2.0) |
| No, never married | 20 (7.5) | 37 (6.9) | 15 (6.0) | 31 (6.3) |
| No, one or both had died | 47 (18.1) | 101 (21.4) | 45 (19.8) | 93 (20.9) |
| Unsure | 0 (0.0) | 1 (0.1) | 0 (0.0) | 1 (0.1) |
| Don't know | 58 (24.5) | 78 (19.1) | 49 (22.9) | 66 (17.9) |
| Refused | 39 (11.9) | 100 (19.1) | 37 (12.7) | 94 (19.4) |

***Note:*** Values are unweighted n (survey-weighted %), calculated among respondents who selected "does not apply" for the corresponding parental experience item. In the Global Flourishing Study survey questionnaire, "does not apply" was instructed for respondents who did not know the respective parent or whose parent had died.
